# Multi-molecular scores map process-specific polygenic diabetes risk to atherosclerosis, cardiometabolic diseases, and vascular complications

**DOI:** 10.64898/2026.01.15.26344187

**Authors:** Hui Li, Jakub Morze, Martin Adiels, Joel Kullberg, Jordi Merino, Magdalena Sevilla-Gonzalez, Göran Bergström, Marju Orho-Melander, Stefan Söderberg, Carl Johan Östgren, Tuuli Lappalainen, Tove Fall, Miriam S Udler, Ida Häggström, Clemens Wittenbecher

## Abstract

Type 2 diabetes (T2D) is etiologically heterogeneous, and process-specific polygenic scores (pPSs) only partly resolve this complexity. Here we integrate fourteen published T2D pPSs with plasma biochemistry, proteomics and NMR metabolomics in 29,425 SCAPIS participants to derive process-specific, polygenic-informed multi-molecular scores (pMMSs) that map genetic risk onto multi-omic molecular signatures. We then test the associations of these scores with subclinical coronary atherosclerosis, incident T2D, macrovascular disease, and microvascular complications in SCAPIS and 458,905 participants from UK Biobank. The novel pMMSs recapitulate the mechanistic interpretability of their underlying pPSs, yet show substantially stronger and more granular associations with molecular traits and disease risk. For example, per standard deviation, the *Proinsulin pMMS* indicates 4- to 5-fold increased risk of incident T2D and diabetic microvascular complications, whereas several lipid-related pMMSs highlight pleiotropic gene clusters with distinct roles in lipid metabolism and cardiometabolic disease pathogenesis. These process-specific molecular endophenotypes operationalize pleiotropy dissection for T2D risk and illustrate how polygenic risk propagates through molecular layers to shape complex cardiometabolic traits.

## Introduction

Type 2 diabetes (T2D) is a multifactorial disease driven by a complex genetic predisposition underlying several pathophysiological processes.(*1, 2*) Previous studies addressed this T2D heterogeneity by clustering selected biomarkers to identify diabetes subgroups with distinct disease progression and risk of diabetic complications.(*3*) These diabetes subtypes capture part of the disease etiology, but scalable precision-health strategies to monitor and target biological process-specific risk for primary prevention remain elusive.

Recent genome-wide association studies (GWAS) have enabled the development total polygenic T2D risk scores (GRS), which aggregate the effects of multiple risk-raising single nucleotide polymorphisms (SNPs) into a continuous score to estimate cumulative genetic risk.(*4*) Building on this, T2D-related process-specific polygenic scores (pPSs) have been developed to decompose total polygenic T2D risk into more nuanced scores that reflect specific genetic risk components, for example, related to glucose metabolism, lipid metabolism, and body fat distribution.(*5, 6*)

In line with their biological interpretation, individual risk load in T2D pPSs has been linked to pPS-specific patterns of body composition, cardiometabolic traits, and diabetes-related clinical outcomes.(*5–7*) These genetic subtyping studies map diverse genetic components of T2D risk and provide valuable insights into the biological processes underlying disease heterogeneity. However, genetics alone explains only a small fraction of interindividual variance in risk factor profiles. Moreover, the associations between pPSs and T2D incidence or complications are typically precise, but the effect sizes are small and not meaningfully differentiate among pPSs.

Integrating genetic data with dynamic, non-genetic information from clinical, imaging, and multi-omics biomarkers may preserve the compelling biological interpretability produced by clustering genetic risk variants, while providing a more granular assessment of accumulated risk across diabetes-relevant biological processes. Such comprehensive, cross-layer multi-omics integrations are not yet established.

Individuals with T2D are at increased risk for developing microvascular complications, including retinopathy and nephropathy, as well as macrovascular complications such as atherosclerotic plaques in the coronary and carotid arteries. (*8–11*) Beyond associations with T2D risk and subclinical risk factors, the clinical utility of new T2D risk subtyping tools must be evaluated against the strength and precision of their associations with diabetes complications.

In this study, we leverage the deeply phenotyped Swedish Cardiopulmonary Bioimage Study (SCAPIS; n ∼ 30,000) (*12*), integrating genetics, clinical biochemistry, proteomics, metabolomics, and imaging data, together with the molecular profiling and disease incidence data from the UK Biobank (UKBB; n ∼ 500,000)(*13*), to derive process-specific, polygenic-informed multi-molecular scores (pMMSs). We reconstructed 14 established T2D pPSs and a summed Total GRS (*5*), built multiple pMMSs within and across omics layers, and evaluated their associations with a broad array of molecular markers, subclinical atherosclerosis, incident T2D and its complications, and cardiovascular disease outcomes.

## Results

### Reconstruction and characterization of T2D-related pPSs

Using 352 T2D-associated variants and weights derived from a European-ancestry clustering approach described previously (*5*), we reconstructed 14 pPSs and a process-agnostic Total GRS in SCAPIS (n = 29,425) and UKBB (n = 458,905) (**Table 1**, Table S1). Detailed information, including definitions, abbreviations, and available data for each pPS, all the clinical variables included in this study (around 30,000 for biochemistry and imaging markers; 5000 for proteomics and metabolomics markers from SCAPIS) is summarized in Table S2.

**Table 1.** Baseline Characteristics of the SCAPIS and UKBB.

| Sex | SCAPIS |  | UKBB |  |
| --- | --- | --- | --- | --- |
|  | Female | Male | Female | Male |
| No. of participants | 15,045<br>(51.1 %) | 14,380<br>(48.9 %) | 249,099<br>(54.3 %) | 209,806<br>(45.7 %) |
| Age, y | 57.4 (53.7, 61.2) | 57.5 (53.7, 61.3) | 58.0 (50.0, 63.0) | 58.0 (51.0, 64.0) |
| Body mass index, kg/m<sup>2</sup> | 25.6 (23.1, 29.1) | 26.9 (24.8, 29.6) | 26.1 (23.4, 29.6) | 27.3 (25.0, 30.1) |
| Waist to hip ratio | 0.86 (0.81, 0.91) | 0.98 (0.93, 1.02) | 0.81 (0.77, 0.86) | 0.93 (0.89, 0.98) |
| Systolic blood pressure, mmHg | 121 (110, 134) | 127 (118, 138) | 134 (122, 148) | 140 (129, 152) |
| Diastolic blood pressure, mmHg | 76 (69, 84) | 78 (72, 85) | 80 (74, 87) | 84 (78, 91) |
| Glucose, mmol/L* | 5.4 (5.1, 5.8) | 5.7 (5.4, 6.2) | 4.9 (4.6, 5.3) | 5.0 (4.6, 5.4) |
| HbA<sub>1c</sub>, mmol/mol | 36.0 (33.0, 38.0) | 36.0 (33.0, 38.0) | 35.1 (32.7, 37.6) | 35.2 (32.7, 38.0) |
| Insulin, mIE/L | 4.86 (3.53, 7.0) | 5.89 (4.09, 8.85) | — | — |
| Total cholesterol, mmol/L | 5.6 (4.9, 6.3) | 5.3 (4.6, 6.0) | 5.8 (5.1, 6.6) | 5.5 (4.7, 6.2) |
| HDL cholesterol, mmol/L | 1.8 (1.5, 2.1) | 1.4 (1.1, 1.6) | 1.6 (1.3, 1.8) | 1.2 (1.1, 1.5) |
| LDL cholesterol, mmol/L | 3.4 (2.8, 4.0) | 3.4 (2.8, 4.1) | 3.6 (3.0, 4.2) | 3.5 (2.9, 4.1) |
| Triglycerides, mmol/L | 0.9 (0.7, 1.3) | 1.2 (0.8, 1.7) | 1.3 (1.0, 1.9) | 1.7 (1.2, 2.5) |
| Screening-detected Atherosclerosis – n (%) | 3,950 (29.4 %) <sup>+</sup> | 7,564 (56.4 %) <sup>+</sup> | — | — |
| Clinically diagnosed Atherosclerosis – n (%) | — | — | 2,976 (1.2 %) <sup>#</sup> | 12,139 (5.8 %) <sup>#</sup> |
| Diabetes – n (%) | 461 (3.2 %) | 795 (5.8 %) | 3,474 (1.4 %) | 5,768 (2.7 %) |
| Hypertension – n (%) | 3,062 (21.0 %) | 3,398 (24.7 %) | 46,515 (18.7 %) | 53,024 (25.3 %) |
| Hyperlipidemia – n (%) | 1,393 (9.6 %) | 1,957 (14.2 %) | 39,948 (16.0 %) | 51,828 (24.7 %) |

Continuous data are presented as median and interquartile range (25th, 75th percentile).
\* SCAPIS: overnight fasted plasma glucose; UKBB: random glucose.
<sup>+</sup> Atherosclerosis found in at least one coronary segment based on the visual assessment of coronary arteries in the Coronary CT angiography (CCTA) images for SCAPIS.
<sup>#</sup> Prevalent coronary artery disease based on hospital registries for UKBB.

The pPSs consist of unique weighted SNP compositions and are linked to distinct pathophysiological process (**Fig. 1a**, Table S1). The Cholesterol, Lipodystrophy-2, Liver-Lipid, ALP-Negative, SHBG-LpA, and Bilirubin pPSs were non-normally distributed (**Fig. 1b**). Accordingly, we applied rank-based inverse normal transformation to each pPS across the entire cohort to enable parametric statistical approaches such as regression analysis (Fig. S1).

**Fig. 1.**
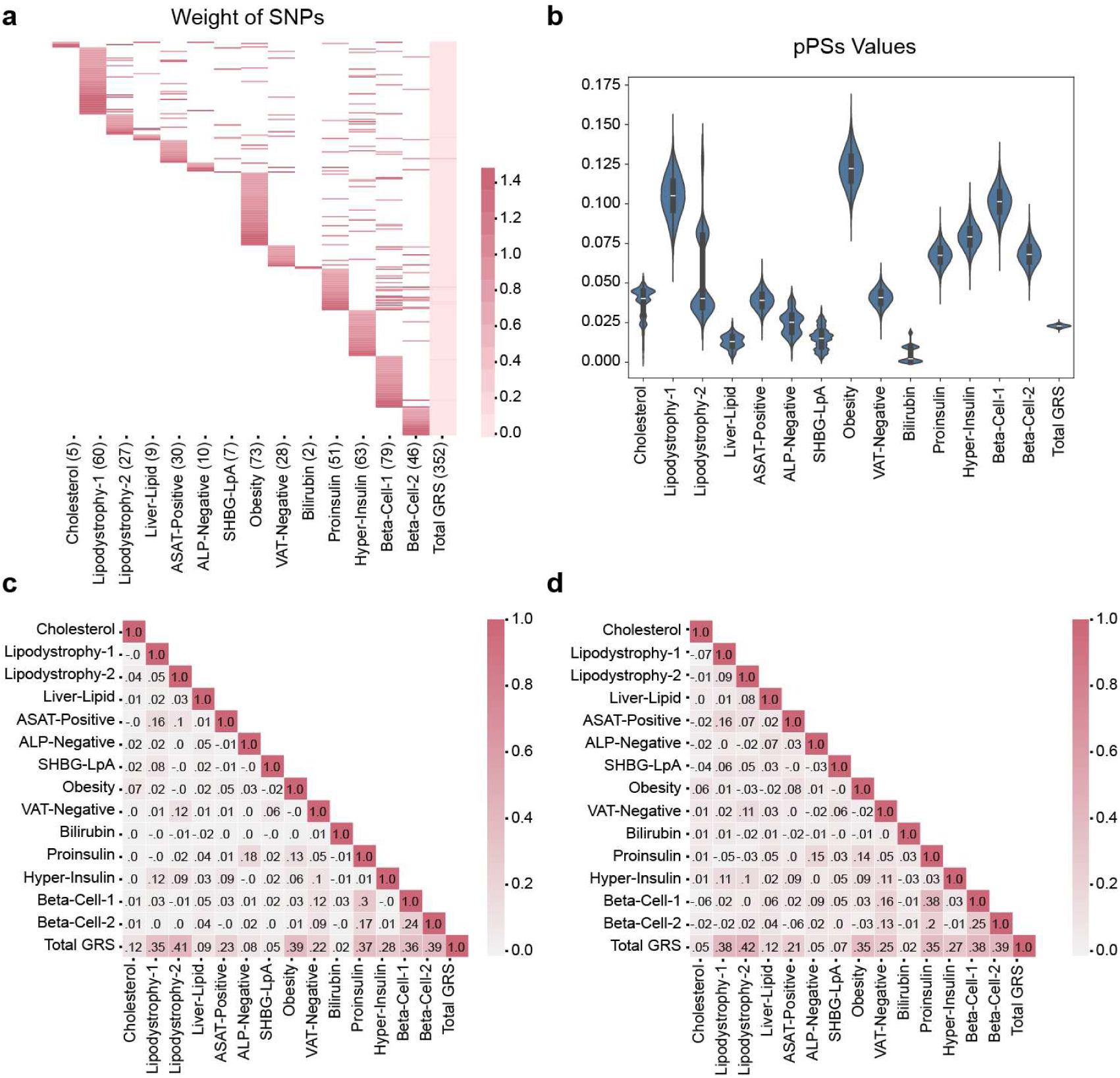
Reconstruction and characterization of European ancestry-based pPSs in SCAPIS. (**a**) Weight matrix of SNPs used to construct pPSs. The number of SNPs included in each pPS is shown in parentheses. (**b**) Distribution of individual values across the 14 pPSs and total GRS. (**c**) Spearman correlation matrix of pPSs in participants without diabetes. (**d**) Spearman correlation matrix of pPSs in participants with diabetes.

In participants without diabetes, pPSs showed predominantly low cross-correlation. Only the glucose metabolism-related Beta-Cell-1, Beta-Cell-2, and Proinsulin pPSs were moderately inter-correlated. In addition, most pPSs were moderately correlated with the Total GRS (**Fig. 1c**). The pPS correlations in participants with prevalent diabetes were comparable (**Fig. 1d**). The sample size, correlation coefficients, and false discovery rate (FDR)-corrected p-values of correlation analysis are reported in Table S3.

### Associations between pPSs, anthropometrics, and clinical biomarkers in SCAPIS

Consistent with previous studies (*14, 15*), among the associations with anthropometric traits, the Lipodystrophy-1 and Obesity pPSs showed opposite associations with BMI, hip circumference, and body weight, but convergent positive associations with waist-to-hip ratio. The Obesity and Hyper-Insulin pPSs were associated with modestly but significantly higher blood pressure (Fig. S2a, Table S4).

The Total GRS was broadly associated with clinical biochemistry markers of immune function, metabolism, and organ health. Subsets of pPSs decomposed these biomarker associations into distinct biological processes. Glucose metabolism-related pPSs (Proinsulin, Hyper-Insulin, Beta-Cell-1, and Beta-Cell-2) showed particularly strong and consistent associations with fasting glucose and HbA1_c_ levels. The Beta-Cell-2 pPS showed the strongest associations with fasting glucose and HbA1_c_ levels and was one of few pPSs significantly associated with lower insulin levels (**Fig. 2a**, Table S5).

**Fig. 2.**
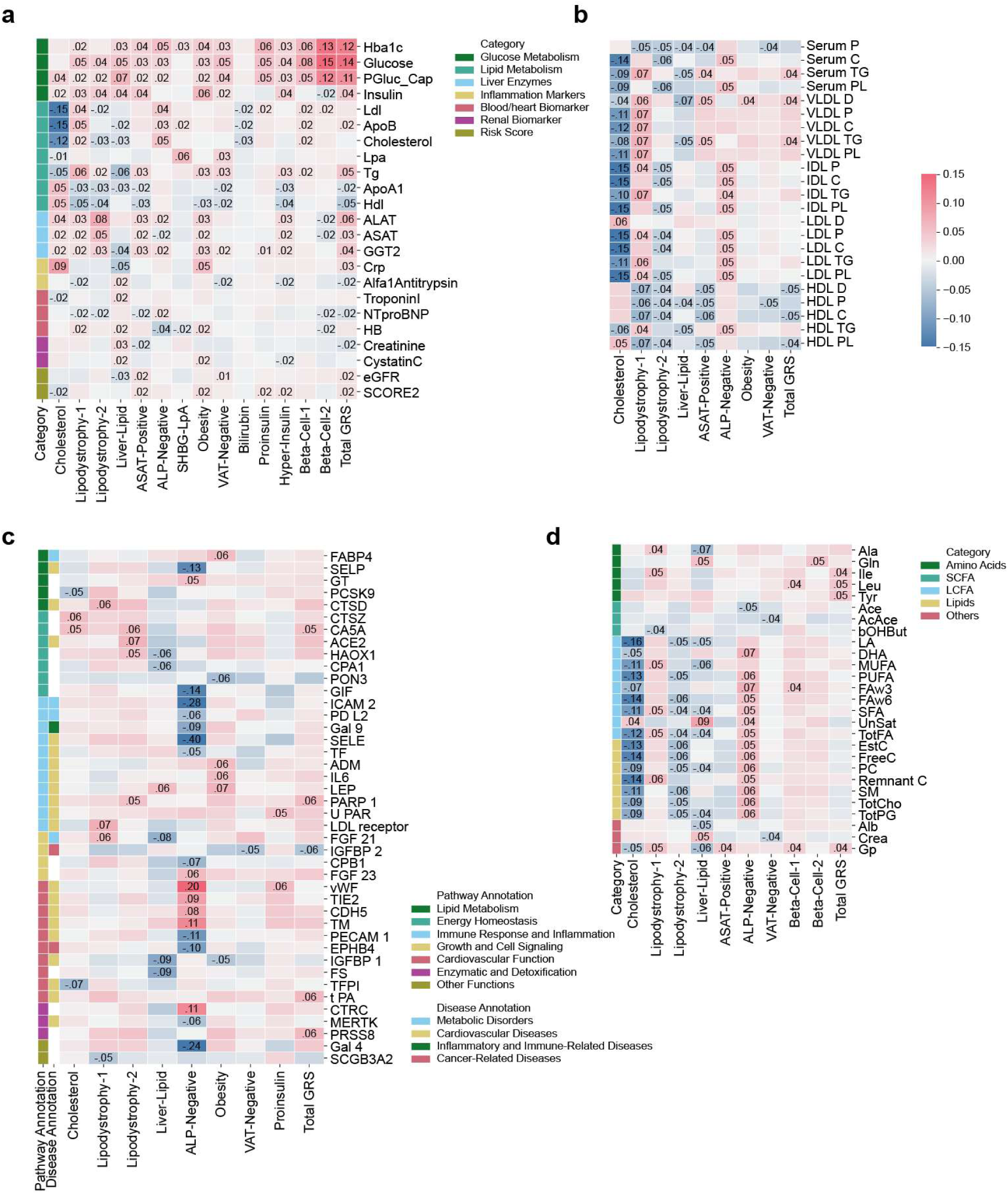
Associations of pPSs with plasma molecular features in SCAPIS. (**a**) Correlation of pPSs with clinical biomarkers. (**b**) Correlation of pPSs with NMR-detected lipoprotein traits. (**c**) Correlation of pPSs with proteins. (**d**) Correlation of pPSs with NMR-detected small molecules. All plots show only molecular features with at least one significant pPS association; significant coefficients are annotated with two decimal places. _P: Particle concentration; _C: Total cholesterol; _TG: Triglycerides; _PL: Phospholipids; _D: Mean particle diameter; SCFA: Short-chain fatty acids; LCFA: Long-chain fatty acids.

Lipid metabolism-related pPSs (Cholesterol, Lipodystrophy-1, Lipodystrophy-2, and Liver-Lipid) were linked to biomarkers of lipid metabolism and liver health but displayed divergent biomarker signatures. The Cholesterol pPS was associated with a favourable lipid profile, including positive correlations with high-density lipoprotein (HDL) content (as approximated by apolipoprotein A1 [apoA1] and HDL cholesterol [HDL-C]) and inverse correlations with apolipoprotein B (apoB)-containing lipoproteins (approximated by low density lipoprotein [LDL] cholesterol [LDL-C], triglycerides [TG], apoB, and lipoprotein(a) [Lp(a)]). The Lipodystrophy-1 pPS was correlated with the same lipid biomarkers in the opposite direction, reflecting an adverse lipid profile. The Lipodystrophy-2 pPS was correlated with lower total cholesterol and related markers (LDL-C, HDL-C, apoA1) but higher triglycerides. In addition, the ALP-Negative, VAT-Negative, and Beta-Cell-1 pPSs were correlated with biomarkers of lipid metabolism (**Fig. 2a**, **2b**, Table S6).

The Cholesterol, Lipodystrophy-1, and especially Lipodystrophy-2 pPSs were linked to higher levels of liver damage-related biomarkers (aspartate aminotransferase [ASAT], alanine aminotransferase [ALAT], gamma-glutamyl transferase [GGT2]). Other pPSs related to biomarkers of hepatic stress included the ASAT-Positive, ALP-Negative, Obesity, and Hyper-Insulin pPSs. In contrast, the Liver-Lipid pPS was correlated with lower GGT2 levels, whereas the Beta-Cell-2 pPS correlated with lower ASAT and ALAT levels.

Even though the Cholesterol pPS coincided with a favourable lipid profile, it correlated with higher levels of the inflammation marker inflammatory marker high-sensitivity C-reactive protein (hs-CRP). Markers of inflammation, kidney function (estimated glomerular filtration rate), and predicted cardiovascular risk score (SCORE2) also showed distinct correlation patterns with other pPSs, including the ASAT-Positive, Obesity, VAT-Negative, Hyper-Insulin, and Beta-Cell-2 pPSs. Several correlations of pPSs with biomarker of kidney function and predicted cardiovascular risk were statistically significant but weak. As components of T2D polygenic risk, pPSs demonstrated process-specific associations with T2D-related clinical markers and captured opposing effect directions for related traits.

Partial correlation analyses, adjusting for all other pPSs (excluding the total GRS) revealed consistent correlation patterns between pPSs and anthropometric traits and clinical biomarkers (Fig. S2, Table S4-S6).

### Proteomics, lipoprotein subfraction, and metabolomics associations of pPSs in SCAPIS

Correlations of T2D pPSs with 184 proteomic markers included on the Olink CVD panels II and III in a subset of SCAPIS participants (n ∼ 4,000) provided further insights into pPS-specific molecular patterns (**Fig. 2c**, Table S7). Full protein function annotation table based on the UniProt database(*16*) and gene set annotation engine Enrichr(*17*) are provided in Table S8.

The ALP-Negative pPS directly correlated with coagulation-related proteins such as von Willebrand factor (vWF) and thrombomodulin (TM) and inversely correlated with immune-regulatory and inflammatory proteins such as intercellular adhesion molecule 2 (ICAM2) and E-selectin (SELE). Other notable proteomics associations include direct links of the Obesity pPS with inflammatory regulators (interleukin-1 receptor antagonist, interleukin-6) and adipokines (adipocyte fatty acid-binding protein [FABP4], leptin [LEP]) and correlations of the Lipodystrophy-1 pPS with liver-derived proteins involved in lipid metabolism and cardiovascular health (proprotein convertase subtilisin/kexin type 9 [PCSK9], cathepsin D [CTSD], LDL receptor, and fibroblast growth factor 21 [FGF21]).

In addition to standard lipid measures, we examined how lipid-related pPSs relate to specific lipoprotein particle subclasses measured by nuclear magnetic resonance (NMR) spectroscopy (**Fig. 2b**, **2d**; Table S6, S9). We confirmed that the Cholesterol pPS aligns with a favourable lipoprotein pattern, characterized by lower levels of very-low density lipoprotein (VLDL) and LDL particles, triglyceride content in HDL particles, and long-chain fatty acids (LCFA), but a higher degree of unsaturation. Conversely, the Lipodystrophy-1 pPS was linked to an unfavourable profile, including higher levels of apoB-containing lipoproteins, enrichment in triglyceride-rich particles, and reduced levels of HDL particles. The Lipodystrophy-2 and Liver-Lipid pPSs tended toward lower overall lipoprotein levels, whereas the ALP-Negative pPS was generally associated with higher lipoprotein particle levels. Data on pPS corelations with additional lipoprotein subfractions is available in Fig. S3 and Table S9.

These associations persisted in partial correlation analyses adjusted for all other pPSs (Fig. S2-S3, Table S7-S9).

### Elastic Net regression identifies key multi-omics predictors of pPSs in SCAPIS

To integrate genetic pPSs with dynamic molecular markers, we constructed multiple Elastic Net regression models in which each pPS served as the dependent variable and markers from multiple omics layers served as predictors. Participants from SCAPIS without diabetes, hyperlipidemia, or corresponding therapy were randomly split into training and testing sets in a 7:3 ratio. Details of model construction and evaluation are provided in the Materials and methods section. Elastic net model performance (R²), sample sizes, numbers of selected variables, and permutation-based p-values (FDR-corrected) are reported in Table S10, and full model coefficients for each omics layer are reported in Table S11.

Elastic Net models showed significant associations between genetic pPSs and specific molecular signatures across multiple omics layers. The strength of these associations varied by biological process and omics layers, with the biochemical layer capturing the largest proportion of pPS-related variation based on the permutation-based p-values. (**Fig. 3a**).

**Fig. 3.**
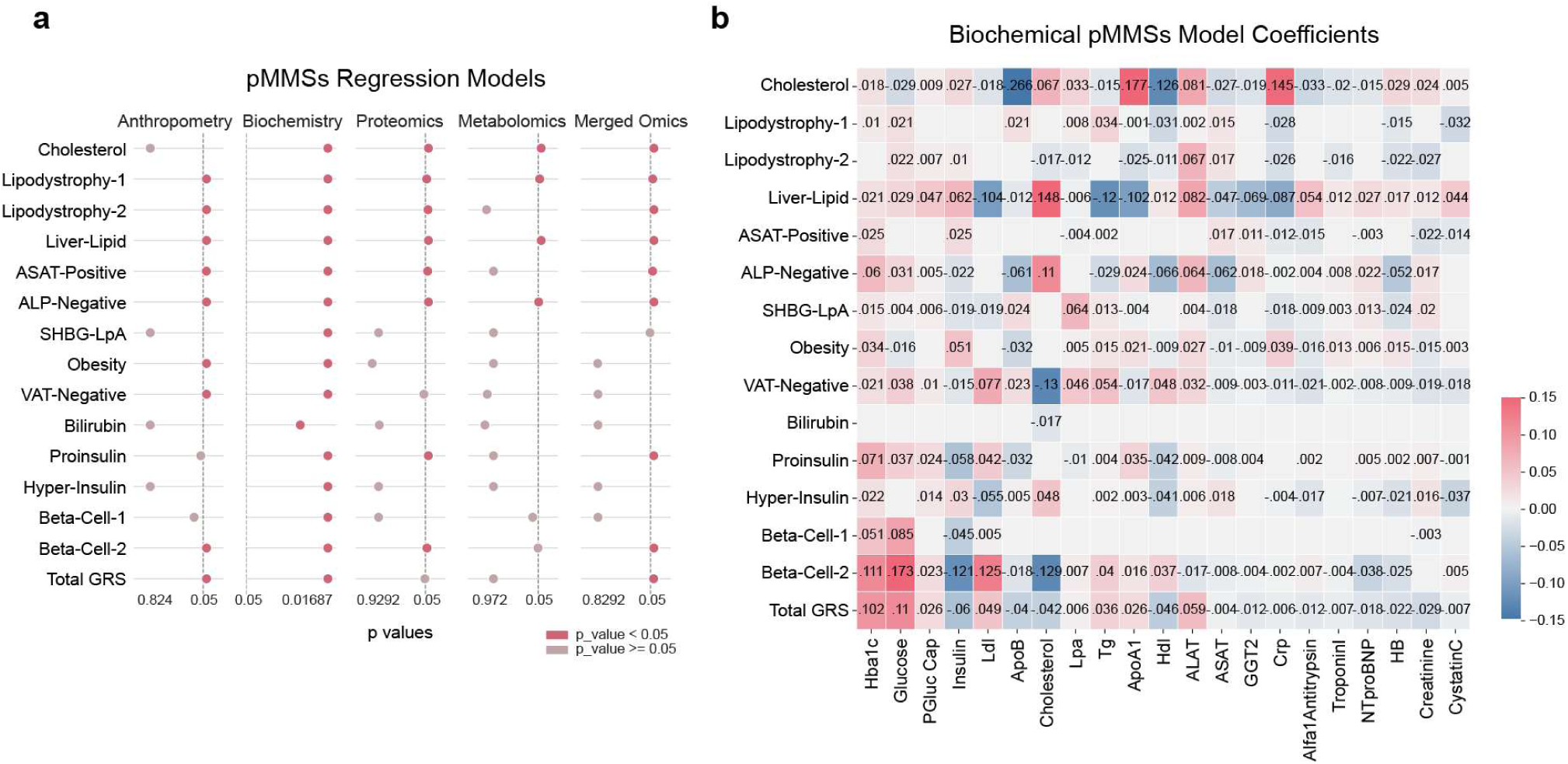
Key molecular predictors of pPSs identified by Elastic Net regression models in SCAPIS. (**a**) Significance of Elastic Net regression models across anthropometry and different molecular layers according to permutation-based p-values (FDR corrected). (**b**) Coefficient matrix from Elastic Net models predicting pPS values (y-axis) from clinical biomarker data (x-axis). Coefficients with absolute values that remained non-zero after rounding to three decimal places are annotated.

The coefficients derived from the Elastic Net regression models were generally consistent with the association patterns observed in the single-marker correlation analyses. In multivariable anthropometric models, the Lipodystrophy-1 and Obesity pPSs captured opposite genetic influence on BMI and weight, while waist-to-hip ratio was positively associated with both. We also observed slight sex differences in Lipodystrophy-1, Liver-Lipid, and Obesity pPSs (higher in woman) as well as VAT-Negative and Beta-Cell-2 pPSs (higher in men) (Fig. S4a).

Multivariable modelling of biochemical markers revealed that the Beta-Cell-2 pPS was linked to a pattern of high fasting glucose, HbA1_c_, and LDL-C levels, alongside lower levels of insulin and cholesterol. (**Fig. 3b**) The molecular imprint of high T2D risk load in the Cholesterol pPS was dominated by high relative levels of apoA1, hs-CRP, and ALAT, and low apoB levels. Interestingly, the Cholesterol pPS-related coefficients changed from negative univariable correlations to positive coefficients in the multivariable model for total cholesterol, Lp(a), HDL-C, ASAT, and GGT2. Other strong biomarker predictors in multivariable models included high total cholesterol combined with relatively low LDL, triglycerides, and apoA1 for the Liver-Lipid pPS; low total cholesterol for the VAT-Negative pPS; and high plasma glucose, HbA1_c_, and LDL-C combined with relatively low insulin and total cholesterol for the Beta-Cell-2 pPS.

Among the 184 proteins included in the proteomics analyses, we observed, for example, proteome signatures of ALP-Negative pPS consistent with subclinical vascular damage and proinflammatory processes, and links of the Cholesterol, Lipodystrophy-1, Lipodystrophy-2, and ASAT-positive pPSs to signatures of growth factors, cytokines and metabolic regulators (Fig. S4b). Multivariable modelling of 143 NMR-based lipoprotein metrics and metabolites yielded strong links with hepatic lipid metabolism-related pPSs (Fig. S4c). The Cholesterol pPS contributed to high saturated fatty acids (SFA), cholesterol esters in medium VLDL, phosphatidylcholine and other cholines (PC) in combination with relatively monounsaturated fatty acids (MUFA), linoleic acid (LA), and cholesteryl esters in small LDL. High genetic T2D risk load in the Liver-Lipid pPS was linked to higher levels of TG-rich lipoprotein fractions (including triglycerides in small HDL, chylomicrons, and extremely large VLDL particles), phospholipids (PL) in very large VLDL, and SFA.

Merged molecular models based on all omics and biomarker layers provided unique molecular signatures for most pPSs (**Fig. 4**). The Cholesterol pPS was linked to a pattern of high hs-CRP, apoA1, and insulin, alongside negative weights for apoB and PCSK9, reflecting a profile an inflammatory profile combined with a cardioprotective lipoprotein pattern. The molecular signatures of the Lipodystrophy pPSs included markers of cardiovascular function (tissue-type plasminogen activator [t PA]), liver function (ALAT), lipid metabolism (PCSK9), and metabolic disorders (CA5A, CTSD, SERPINA12). The Liver-Lipid pPS molecular signature was driven by growth and cell signaling markers (IGFBP2), immune response and inflammation markers (interleukin-16 [IL16], LEP), and glucose metabolism marker (capillary blood glucose [PGluc Cap]). The molecular signature of the ALP-Negative pPS showed prominent negative coefficients for immune response and inflammation markers (SELE, ICAM2), as well as positive coefficients for cardiovascular function and disease-related proteins (CDH5, vWF, TIE2), suggesting vascular and immune system dysregulation. Molecular signatures of Beta-cell-related Proinsulin and Beta-Cell-2 pPSs were marked by strong contributions from insulin, HbA1_c_, glucose, and proteomics markers involved in metabolic and immune regulation (PARP-1, TNFRSF10C), indicative of impaired insulin secretion and glycemic control. Compared to pPSs, the Total GRS exhibited less diverse associations in merged-omics model, largely driven by glycemic markers such as HbA1_c_ and glucose.

**Fig. 4.**
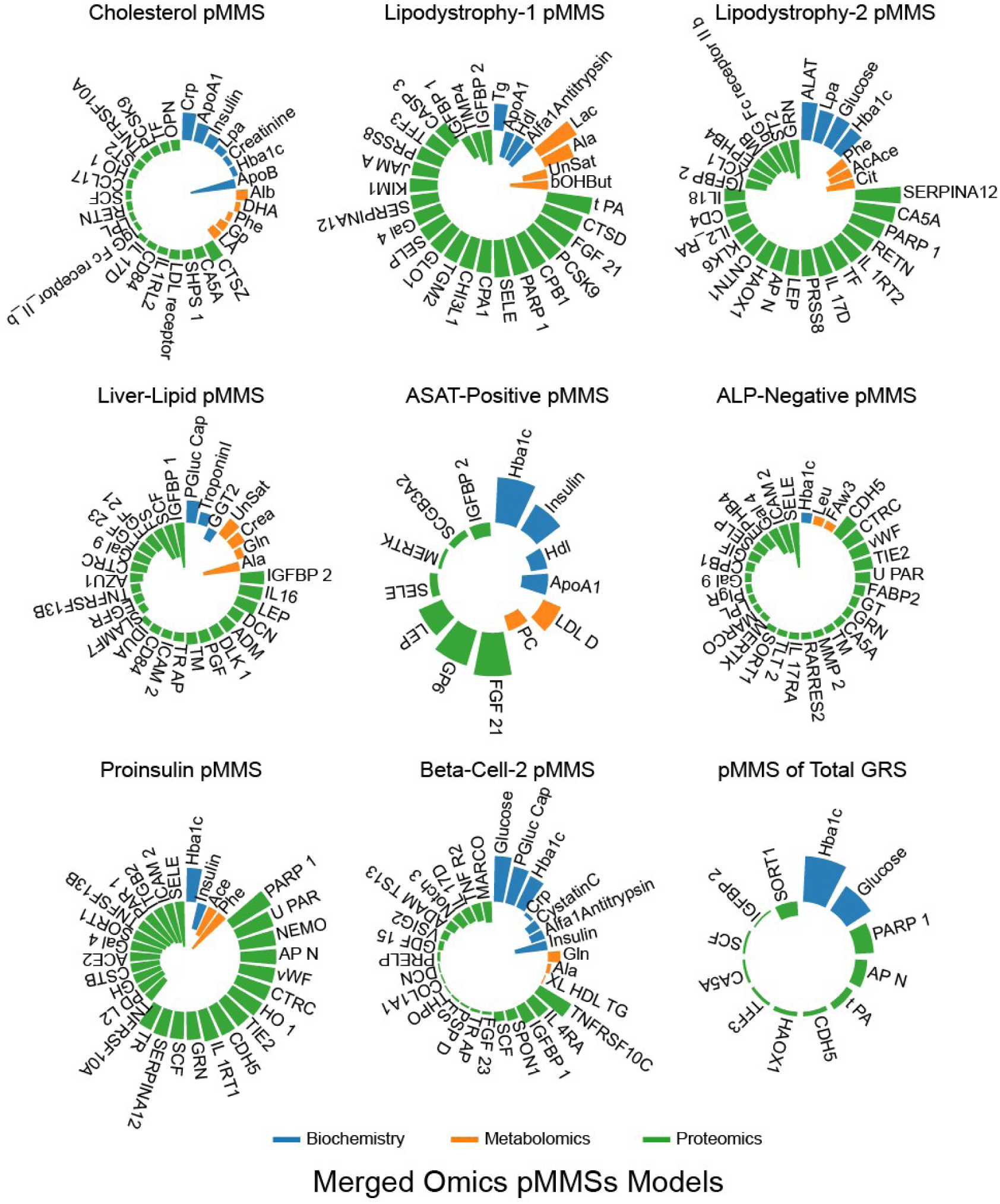
Coefficients of markers selected from merged molecular Elastic Net regression models in SCAPIS. The coefficients of merged omics pMMS Elastic Net models including more than one marker and significant in permutation testing are shown in circular plot with up to top 30 markers.

### Molecular signatures of pPSs and imaging-derived atherosclerosis load in SCAPIS

Using the Elastic Net models fitted within each omics layer and across combined omics layers, we calculated pMMSs based on the selected markers and their corresponding weights in SCAPIS. We then compared the associations of genetic pPSs and model-derived pMMSs with imaging-based markers of subclinical atherosclerosis, including the coronary artery calcium score (CACS), segment involvement score (SIS), and Duke coronary artery disease (CAD) index (**Fig. 5**, Fig. S5a, and Table S12).

**Fig. 5.**
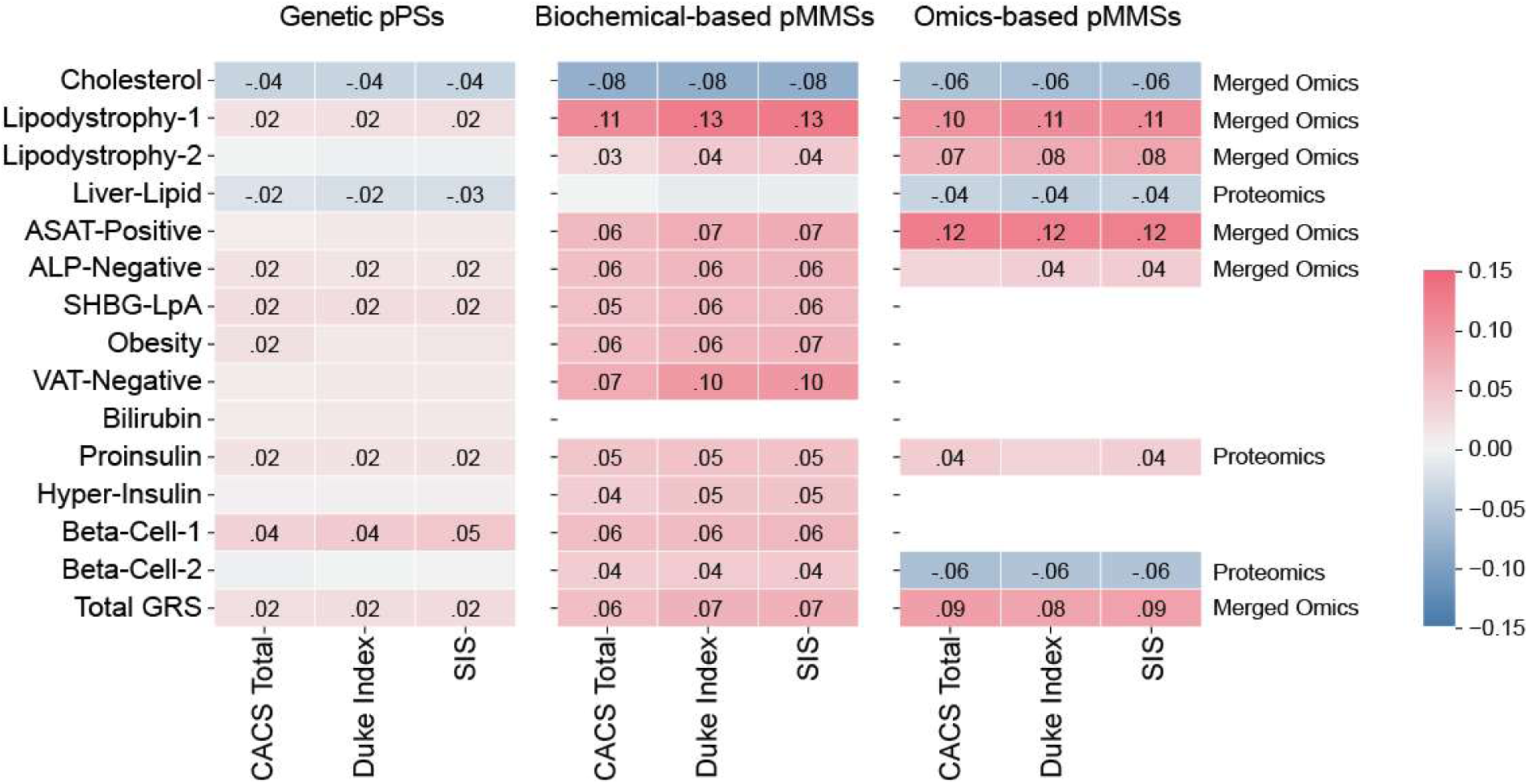
Associations between pPSs, pMMSs, and subclinical atherosclerosis indices in SCAPIS. Matrix of association coefficients between genetically derived pPSs and pMMSs generated from Elastic Net models based on biochemical data layer and selected layers from proteomics, metabolomics, and merged omics layers. Only statistically significant associations with atherosclerosis markers are numerically annotated and only pMMSs with more than one marker and significant in permutation testing were analyzed.

The genetic pPSs linked to lower levels of apoB-containing lipoproteins and cholesterol (Cholesterol and Liver-Lipid pPSs) were inversely associated with atherosclerosis indices, whereas the pPSs associated with higher levels of apoB and cholesterol (for example, Lipodystrophy-1 and ALP-Negative pPSs) were linked to higher atherosclerosis burden. Higher atherosclerosis burden was also observed for the SHBG-LpA, Proinsulin, and Beta-Cell-1 pPSs.

We observed similar association patterns in partial correlation analyses and logistic regression models using categorical CACS as the outcome (Fig. S5b, S5c, and Table S12, S13). Notably, in categorical analyses stratifying individuals by CACS severity, both genetic Obesity and Beta-Cell-1 pPSs demonstrated progressively stronger associations with increasing CACS categories.

Compared with purely genetic pPSs, Lipodystrophy-1, Lipodystrophy-2, ASAT-Positive, and Total GRS pMMSs derived from biochemistry and merged omics layers displayed consistent but stronger associations with imaging-derived atherosclerosis markers. For the Liver-Lipid pMMS, the proteomics signature showed a clear negative association with imaging markers of atherosclerosis. The atherosclerosis association of the genetic Beta-Cell-2 pPS was inconclusive, whereas its biochemistry signature was associated with higher atherosclerosis load and its proteomics signature with lower load. (**Fig. 5**, Fig. S5a)

Similar association analyses with other cardiac-based, binary-transformed CT phenotypes (e.g., presence of any coronary plaque) in logistic regression showed consistent results (Fig. S6, Table S14).

### Molecular signatures of pPSs and risk of T2D and related diseases in SCAPIS and UKBB

Cross-sectional associations of pPSs with prevalent disease (self-reported health history) in SCAPIS are shown in Fig. S7 and Table S15. Overall, most genetic pPSs showed modest associations with multiple cardiometabolic diseases, including hyperlipidemia and hypertension.

We also compared the associations of the genetic T2D pPSs and the newly SCAPIS-derived molecular pMMSs with the incidence of T2D and related clinical outcomes (microvascular diabetes complications and vascular diseases) in UKBB using Cox proportional hazards regression models. pPSs and corresponding pMMSs showed directionally consistent, statistically significant associations with the risk of developing T2D and its complications (**Fig. 6**), and related cardiometabolic diseases (**Fig. 7**). However, the pMMSs (especially those derived from biochemical data) demonstrated much larger hazard ratios (HRs) than their genetic counterparts.

**Fig. 6.**
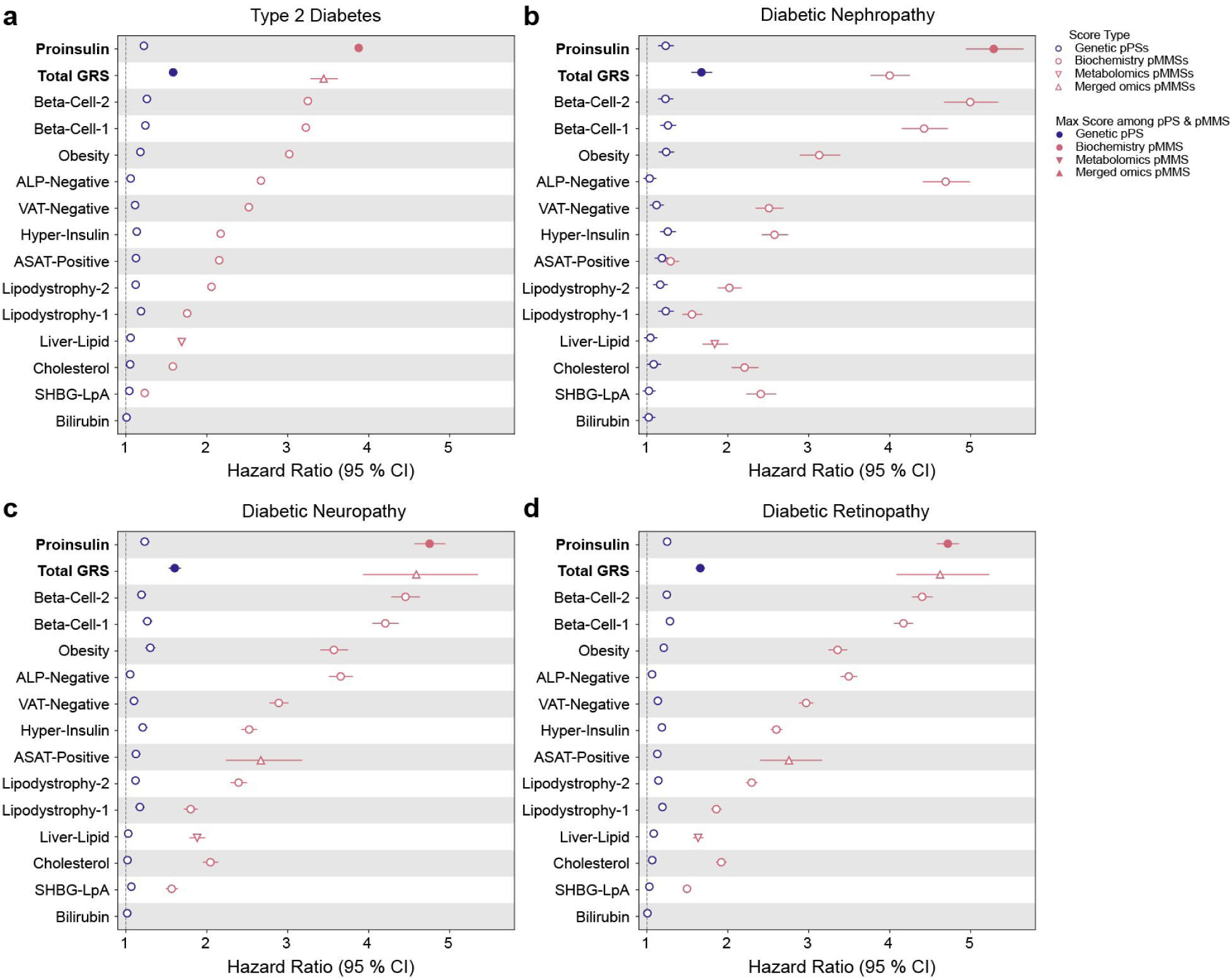
Associations of pPSs and pMMSs with incidence of T2D and microvascular complications in the UKBB. HRs per SD for genetic pPSs and the top-performing corresponding pMMS (either biochemistry, proteomics, metabolomics, or merged omics) with: (**a**) T2D, (**b**) diabetic nephropathy, (**c**) diabetic neuropathy, (**d**) diabetic retinopathy. The pMMSs were derived using Elastic Net Regression, predicting the pPS with the indicated molecular data; only FDR-significant models (permutation test) with more than one maker are retained. Biological processes in all panels are sorted by descending pMMS HRs for T2D. The score with the strongest association among genetic and omics layer is annotated with a solid dot. Points denote HRs per SD with 95% confidence intervals (CI) from Cox proportional hazards models.

**Fig. 7.**
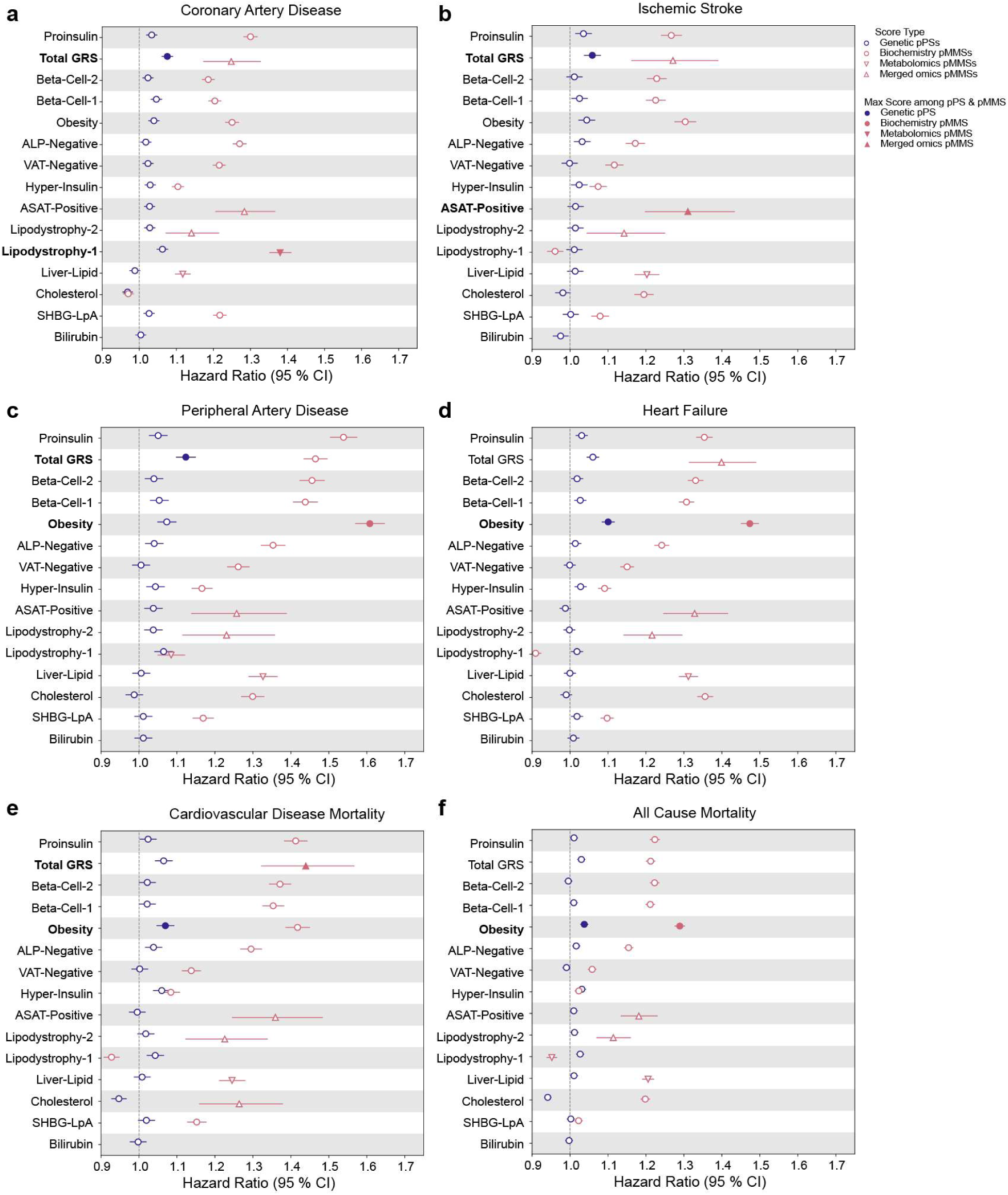
HRs for associations of pPSs and pMMSs with T2D-related diseases in the general UKBB population. HRs per SD for genetic pPSs and the top-performing corresponding pMMS (either biochemistry, proteomics, metabolomics, or merged omics) with: (**a**) CAD, (**b**) ischemic stroke, (**c**) PAD, (**d**) heart failure, (**e**) cardiovascular disease mortality, (**f**) all-cause mortality. The pMMSs were derived using Elastic Net Regression, predicting the pPS with the indicated molecular data; only FDR-significant models (permutation test) with more than one maker are retained. Biological processes in all panels are sorted by descending pMMS HRs for T2D (see **Fig 6A**). The highest score among genetic and omics layer is annotated with a solid dot. Points denote HRs per SD with 95% confidence intervals (CI) from Cox proportional hazards models.

Among genetic scores, the Total GRS, which aggregates all T2D-associated SNPs, showed the strongest association with incident T2D (HR per standard deviation [SD] 1.58, 95 % confidence interval [CI] 1.57 – 1.60). The partitioned genetic pPSs were also linked to the risk of T2D with up to ∼1.3-fold higher risk per SD (Beta-Cell-2 pPS: HR per SD 1.26, 95 % CI 1.25 – 1.27). However, most pMMSs exhibited substantially larger effect sizes. The strongest T2D risk was related to the biochemistry-derived Proinsulin pMMS, with an almost 4-fold higher T2D risk per SD higher score (HR per SD 3.88, 95 % CI 3.84 – 3.92). (**Fig. 6a**, Table S16)

A similar pattern was observed for diabetic microvascular complications. For example, the biochemistry-derived Proinsulin pMMS was also very strongly related to the risk of diabetic nephropathy (**Fig. 6b**, HR per SD 5.28, 95 % CI 4.94 – 5.65), diabetic neuropathy (**Fig. 6c**, HR per SD 4.75, 95 % CI 4.57 – 4.94), and diabetic retinopathy (**Fig. 6d**, HR per SD 4.72, 95 % CI 4.56 – 4.85). None of the genetic risk scores showed comparable associations with diabetic complications. The per-SD relative risk of microvascular diabetic complications was below 1.7 for the Total GRS as top genetic benchmark (diabetic nephropathy HR per SD 1.67, 95 % CI 1.55 – 1.80; diabetic neuropathy HR per SD 1.60, 95 % CI 1.53 – 1.68; and diabetic retinopathy HR per SD 1.66, 95 % CI 1.61 – 1.71) and below 1.3 for the corresponding genetic Proinsulin pPS (diabetic nephropathy HR per SD 1.23, 95 % CI 1.14 – 1.33; diabetic neuropathy HR per SD 1.23, 95 % CI 1.18 – 1.29; and diabetic retinopathy HR per SD 1.25, 95 % CI 1.21 – 1.29).

To understand the driving factors underlying the increased risk of T2D and microvascular complications associated with biochemical data based pMMSs, we repeated the Cox regression analysis of biochemical pMMSs with T2D and related clinicals outcomes in UKBB, additionally adjusting for HbA1_c_ and glucose (excluding the Bilirubin pMMS, which comprised only a single biomarker) (Fig. S7, Table S17). As expected, the HRs for glucose metabolism-related pMMSs (Beta-Cell-1, Beta-Cell-2, and Proinsulin) were attenuated for T2D and related clinical outcomes, particularly for Beta-Cell-1 and Beta-Cell-2 pMMSs, consistent with their strong associations with glycemic traits. However, the strong associations of the biochemical Proinsulin pMMS with the risk of T2D and its microvascular complications persisted after adjustment for glycemic traits, suggesting additional contributions from lipid metabolism related markers, such as LDL-C, HDL-C, and apoA1 (**Fig. 2a, 3b**). For lipid metabolism-related pMMSs such as the biochemical Cholesterol pMMS, HRs were similar before and after adjustment for HbA1_c_ and glucose.

We also examined the associations of the genetic pPSs and derived pMMSs with clinical endpoints for which diabetes constitutes an important risk factor. On the genetic level, the Total GRS was the strongest predictor of CAD, ischemic stroke, peripheral artery disease (PAD), and cardiovascular mortality, and the Obesity pPS was the top predictor of heart failure and total mortality. In general, the effect sizes of genetic pPSs on diabetes-related diseases in UKBB population were relatively small. (**Fig. 7**, Table S16) The newly derived pMMSs preserved the association structure but showed markedly stronger disease risk associations compared to genetic pPSs. The molecular layer (biochemistry, NMR metabolomics, proteomics, multi-omics) and the biological process that yielded the strongest risk associations with clinical outcomes varied across diseases.

For example, associations with incident CAD highlight distinct roles of pPS and pMMS in diabetes-related risk (**Fig. 7a**). Anthropometry-related (Lipodystrophy-1, ASAT-Positive, Obesity, and VAT-Negative) and glucose metabolism-related (Proinsulin, Hyper-Insulin, Beta-Cell-1, and Beta-Cell-2) pPSs and pMMSs were consistently associated with higher CAD risk. The NMR-derived Lipodystrophy-1 pMMS was the strongest predictor of CAD incidence (HR per SD, 1.38; 95% CI, 1.35–1.41), much higher than other lipid-metabolism pMMSs (Lipodystrophy-2, Liver Lipid). The Cholesterol pMMS, consistent with its genetic counterpart, was associated with slightly lower CAD risk (Cholesterol pPS HR per SD, 0.97; 95% CI, 0.96–0.98; biochemistry-derived Cholesterol pMMS HR per SD, 0.97; 95% CI, 0.96–0.98). Notably, all lipid scores, including the Cholesterol, were positively associated with T2D risk, underscoring that specific lipid metabolic processes diverge while other converge on T2D versus CAD risk.

## Discussion

In a large cohort of middle-aged individuals without prevalent diabetes, we examined how process-specific polygenic T2D risk scores relate to detailed molecular profiles and subclinical atherosclerosis. Our analyses revealed molecular alterations that closely align with the current biological interpretation of T2D pPSs. We further examined the associations of genetic pPSs and multi-layer molecular pMMSs with incident T2D and T2D-related clinical endpoints in an independent study population. Compared to genetic-based pPSs, integrating molecular biomarkers markedly strengthened the associations with T2D incidence, diabetic complications, and atherosclerosis, while maintaining the clear biological interpretation provided by genetic clustering.

The Total GRS, as a summary metric of overall genetic risk of T2D, showed broad but weak associations with biomarkers of beta cell function, insulin resistance, lipid metabolism, and hepatic function. Genetic pPSs derived from soft clustering methods decomposed the complex interplay of genetic background risk into different biological processes and showed distinct association patterns across clinical biomarkers.

Dysregulated glucose metabolism is a hallmark of diabetes and clinical biomarkers of blood glucose concentration (HbA1_c_, fasting, or 2-hour plasma glucose) are central to T2D diagnosis. The insulin production-related Beta-Cell-1, Beta-Cell-2, and Proinsulin pPSs were most strongly linked to glycemic control markers and were also related to circulating immune-regulatory proteins. These findings support the notion that immune-metabolic crosstalk is a critical component of beta-cell function decline and to impaired glucose-stimulated insulin secretion in T2D.(*18, 19*) Notably, the two beta cell-related pPSs showed partially distinct downstream associations: Beta-Cell-1 pPS was more strongly associated with dyslipidemia markers (apoB, cholesterol) and indices of subclinical atherosclerosis, whereas Beta-Cell-2 pPS was more strongly associated with glycemic traits, including low insulin levels.

Lipid metabolism-related pPSs demonstrated distinct but complementary associations with lipoprotein subclasses and fatty acid profiles. The Cholesterol pPS was linked to a cardioprotective lipid profile but a higher T2D risk, consistent with the evidence from Mendelian randomization analyses and pharmacological interventions, showing that LDL-C lowering via PCSK9 inhibition reduces atherosclerotic cardiovascular disease (ASCVD) risk while increasing T2D risk, highlighting opposing causal effects of specific lipid pathways.(*20*) Conversely, the Lipodystrophy-1 pPS was characterized by TG-enrichment across all lipoprotein classes and higher remnant cholesterol, in line with the genetic and epidemiological evidence that triglyceride-rich lipoproteins and remnant cholesterol causally increase both ASCVD and T2D risk, underscoring shared pathogenic components of lipid metabolism that contribute to both diabetogenesis and atherogenesis.(*21*)

Despite its cardioprotective lipid profile, the Cholesterol pPS was enriched for pro-inflammatory markers (hs-CRP, SFAs and PCs), and all lipid metabolism-related pPSs were linked to higher liver enzymes levels. The Liver-lipid pPS was additionally associated with inflammatory proteins (alfa-1 antitrypsin, IL16), suggesting chronic systemic inflammation, while other pPSs, particularly ALP-Negative pPS demonstrated links to proteomic markers of cardiovascular health (e.g., vWF, TIE2), indicating potential vascular dysfunction. Together, these patterns show that T2D pPSs capture the complex interplay between lipoprotein metabolism, systemic inflammation, T2D development, and CAD risk.

These association patterns were further validated and extended through integrative Elastic Net regression using markers from single-omic and multi-omics marker sets. The molecular predictors in each pMMS model were largely consistent with the pPS-biomarker associations. The associations of both, genetic pPSs and newly derived pMMSs with imaging-based atherosclerosis markers in SCAPIS revealed notable differences among the dissected processes; for example, the apoB and the cholesterol associated scores showed opposite associations with atherosclerosis indices. Importantly, the pMMSs preserved the direction and pattern of the genetic associations but with markedly larger effect sizes.

We further compared the association of pPSs and pMMSs with the risk of T2D incidence and occurrence of T2D complications in the independent UKBB study. The Total GRS was the strongest genetic predictor of incident T2D with 60 % higher T2D risk per SD higher score, also we observed comparable genetic Total GRS-related risks for diabetic nephropathy, diabetic neuropathy, and diabetic retinopathy. Genetic pPSs were also linked to the risk of T2D and complications, with up to 30 % higher risk per SD higher individual score. However, the biomarker-based pMMSs showed much larger effects compared to genetic benchmarks. In the UKBB analysis, one SD higher biochemistry-derived Proinsulin pMMS was related to a 388 % higher risk of T2D incidence, 528 % higher risk of diabetic nephropathy, 475 % higher risk of diabetic neuropathy, and 470 % higher risk of diabetic retinopathy. The exceptionally strong associations with T2D and its complications highlight the capacity of our pMMSs to capture genetic risk architecture while integrating accumulated, dynamic pathophysiology through biomarkers integration.

We further associated the established pPSs and our pMMSs with incidence of several T2D-related diseases and mortality. Once again, integrated pMMSs preserved the association patterns but yielded a marked increase in effect sizes. For instance, we observed 61 % higher PAD risk, 47 % higher heart failure risk, and 29 % higher total mortality per SD higher for biochemistry-derived Obesity pMMS. The NMR-derived Lipodystrophy-1 pMMS was linked to 38 % higher CAD risk per SD increase, while the Cholesterol pMMS was associated with a slight but statistically significant reduction in CAD risk. Together, these results support that our biomarker-based pMMSs provide process-specific insights into the diverse preclinical processes that increase T2D risk yet contribute differently to broader cardiometabolic health trajectories.

To our knowledge, this is the most comprehensive study on the associations of established T2D pPSs with a broad spectrum of molecular markers across multiple omics layers. A key finding of our research is that extended biomarkers in much large samples, allow meaningful stratification of overall T2D risk, and in several cases the integration of NMR metabolomics and proteomics further strengthened outcome associations. However, our analyses were limited by the availability of proteomics and NMR-metabolomics data in approximately 5,000 participants and by the restricted set of CVD-related proteins on the available Olink panels in SCAPIS, which may reduce the power and coverage for multi-omics discovery. A major strength of our study is that the pMMSs derivation and clinical outcome analyses were conducted in independent cohorts. One of the cohorts, SCAPIS has not contributed to any of the GWAS or genetic clustering studies used, which reduced risk of overfitting via information leakage. However, the included study populations were largely of European genetic ancestry; further studies to evaluate the generalizability and refine the pMMSs in more diverse population datasets are warranted.

In conclusion, our study corroborates that process-specific polygenic scores capture biologically distinct and clinically relevant processes underlying the development of T2D and its complications. Projecting the genetic clusters into comprehensive molecular data revealed clear and distinct association that we summarized in pMMSs. Using atherosclerosis measures from advanced imaging screens and comprehensive disease incidence data in a large population study, we show that these pMMSs are strong process-specific predictors of T2D and its complications. These findings suggest that combining genetic and molecular data can identify distinct, dynamic risk groups of T2D with distinct cardiometabolic health trajectories well before clinical outcomes occur. Notably, standard biochemistry or NMR-derived scores effectively capture major processes underlying polygenic diabetes predisposition, which opens cost-effective paths towards nuanced, process-specific T2D risk assessment. Our findings could enable stratified preventive strategies that target the precise biological processes driving T2D and its complications in individual patients.

## Methods

### Study population, sample collection of SCAPIS

Our primary molecular and phenotypical association analyses were based on SCAPIS, a population-based cohort comprising 30,154 predominantly healthy individuals (aged 50-64 years). Each participant underwent extensive phenotyping over two to three visits within 2 weeks, including clinical assessments, anthropometry, venous blood sampling for immediate analysis and biobank storage, advanced imaging such as computed tomography (CT) and carotid ultrasound, and standardized lifestyle questionnaires.

A venous blood sample (100 mL) was collected from participants after an overnight fast. Immediate analyses of cholesterol, HDL, LDL, triglycerides, glucose, HbA1c, hs-CRP and creatininel, the remaining blood was biobanked for further assays. SCAPIS genotyping was performed in 29,425 participants in 10 batches using a customized version of the Illumina GSA-MDv3 at the SNP&SEQ Technology Platform (Uppsala, Sweden). Genotypes were called with the GenomeStudio 2.0.3 software and imputed based on the Haplotype Reference Consortium (HRC) r1.1 reference panel. Plasma proteomics was generated at SciLifeLab laboratories (Uppsala, Sweden) using Olink Proseek Multiplex CVD II and III panels, targeting 184 cardiovascular proteins in ∼5,000 participants. NMR-based metabolomics was produced by Nightingale Health (Helsinki, Finland) in ∼5,000 participants.

SCAPIS medical imaging technologies including CT and carotid ultrasound in ∼30,000 participants. Coronary CT angiography (CCTA) was performed on a dedicated dual-source scanner with a Stellar Detector, and coronary artery calcium was quantified from non-contrast scans to derive total CACS. Coronary atherosclerosis was assessed based on the visual assessment of coronary arteries in the CCTA images with 18 coronary segment model (*22*). SIS was calculated as the total number of clinically relevant segments with atherosclerotic plaque.

Detailed study procedures were described in previous publications.(*12, 23*)

### Study population of UKBB

UKBB is a large, population-based prospective cohort study with over 500,000 participants (aged 40-69 years, 2006 - 2010). Baseline assessments included standardized questionnaires, physical measures and biospecimen collection. Non-fasting venous blood (∼ 45 – 50 mL), spot urine, and saliva were collected. Core hematology/biochemistry and extended biomarker panels were assayed according to UKBB protocols. Genome-wide genotype calling was performed by Affymetrix on two closely related purpose-designed arrays for ∼ 488 k participants. Detailed study design and methods were published previously. (*13*)

### Generation of T2D pPS

We constructed 14 European subpopulation-based pPSs based on a previously published multi-ancestry clustering approach using 352 genetic variants associated with T2D in SCAPIS and UKBB. Each pPS was calculated as the sum of multiplying a variant’s genotype dosage by its respective cluster weight using PLINK v2.0. A Total GRS was computed using the effect sizes of all 352 variants. All SNPs and weights for genetic scores were extracted from Smith et al(*5*), and are provided in Table S1.

### Clinical and imaging risk scores

We extracted clinical and imaging-based cardiovascular risk scores from the SCAPIS database, including the CACS according to Agatston from the three major coronary arteries, the modified Duke prognostic coronary artery disease (CAD) index, the segment involvement score (SIS). CACS values were categorized into four clinically relevant categories to support risk evaluation and guide therapy decisions: CACS = 0; 0 < CACS <= 99; 99 < CACS <= 300; CACS > 300.(*24, 25*) Three dimensions one-hot encoding was subsequently used to incorporate these categories into downstream analyses.

### Metabolite aggregation

To facilitate interpretation, we derived composite variables for lipoprotein-related measures by summing metabolite concentrations across particle sizes. Specifically, we aggregated values for HDL, VLDL, LDL, total serum particle count, and total serum phospholipids by summing the values of all subclasses.(*26*)

### Disease Adjudication

For SCAPIS, health status at baseline was obtained via a standardized questionnaire (self-reported, doctor-diagnosed conditions) and linkage to Swedish national health registers. Prevalent conditions included atrial fibrillation, heart failure, stroke, hypertension, hyperlipidemia, diabetes (including type 1 diabetes and T2D), sleep apnea, rheumatic disease, cancer, and CAD (defined as a composite of myocardial infarction, angina pectoris, or coronary revascularization by Coronary Artery Bypass Grafting [CABG] or Percutaneous Coronary Intervention [PCI]) were included in this study.

For UKBB, incident outcomes were derived from national death and hospital admissions data. Endpoints include T2D, diabetic nephropathy, diabetic neuropathy, diabetic retinopathy, CAD, ischemic stroke, PAD, heart failure, cardiovascular disease mortality, and all-cause mortality were included in this study. Participants with relevant condition at baseline were excluded from the corresponding incident risk analyses. Follow-up accrued from baseline assessment until the first qualifying event, death, loss to follow-up, or end of registry linkage.

Detailed information, including the abbreviations, the available data of the clinical variables from SCAPIS and the endpoints from UKBB included in this study is summarized in Table S2.

### Molecular markers

In total, we evaluated associations between the T2D pPSs and a broad set of clinical and biochemical parameters derived from physical examination, blood analysis, imaging, and core questionnaires. These included 9 anthropometric traits, 21 biochemical markers, 184 plasma proteins, 143 plasma metabolites, 8 calculated metabolite sums, 10 imaging-derived variables, 5 established clinical risk scores, and 10 cardiometabolic outcomes from SCAPIS, as well as 10 T2D and T2D-relevant clinical endpoints from UKBB. The detailed definitions, abbreviations, and available data of clinical variables included in this study were summarized in Table S2.

### Statistical analysis Correlation analysis

Spearman correlation was used to evaluate the intercorrelation of associations pPS and the relation between each pPS and anthropometric, biochemical, proteomic, metabolomic variables, as well as clinical risk scores. The intercorrelation between pPSs was examined also using Spearman coefficients and stratified by diabetes status. Correlation of pPSs with molecular markers was examined in participants without diabetes. To minimize medication-related biases, individuals with diagnosed or self-reported diabetes or hyperlipidemia, or those receiving insulin or hyperlipidemia therapies were excluded from multi-omics correlation analyses. To account for potential pleiotropic effects, partial correlation analyses were conducted, adjusting for all other pPSs (excluding the Total GRS). Except for the pPS intercorrelation analyses, all correlation analyses were adjusted for age, sex, and principal components (PCs) 1-10. The sample size, correlation coefficients, 95% confidence intervals (CIs), and FDR-corrected p-values from the association analyses are summarized in the corresponding supplementary tables.

### Multivariable regression

Binary-transformed CT imaging variables, carotid ultrasound findings, and health history variables in SCAPIS were modelled as dependent variables in logistic regression analyses, restricted to outcomes with at least 100 events to ensure model stability. Each pPS was included as an independent variable. For one-hot–encoded categorical CACS, we applied categorical linear regression with the three encoded CACS features as predictors and each pPS as the response variable. To enable cross-score comparisons, all pPSs were rank-based inverse normal transformed across the cohort prior to analysis. Associations were evaluated in participants without diabetes. All models were adjusted for age, sex, and genetic PCs 1–10. Sample size, number of events, regression coefficients, 95% CIs, and FDR-corrected p-values are summarized in the corresponding supplementary tables.

### Elastic Net linear regression

We constructed Elastic Net linear regression models in participants without diabetes and hyperlipidemia, or corresponding therapy in SCAPIS, with each pPS as the dependent variable and anthropometric, biochemical, proteomic, and metabolomic markers considered separately as predictors. Merged-omics Elastic Net models were also implemented, combining markers from biochemical, proteomic, and metabolomic layers into a single predictor set. All variables and pPSs were rank-based inverse normal transformed across whole cohort to compare coefficient across models.

The dataset was randomly split into a 7:3 ratio for training and testing. We used 10-fold cross-validation on the training set to identify the optimal α (L1 vs. L2 ratio) and λ (regularization strength). To evaluate the performance of each Elastic Net regression models, we calculated the coefficient of determination (R^2^) of each model on test set and performed permutation test where we shuffled the order of response variables (pPS) on training set and retrain the Elastic Net linear regression models 1000 times. We compared the R^2^ of the original and retrained models, calculated permutation-based p-value on how many of shuffled models performed better than original models on the same test set.

### Predicted molecular signature-based pMMSs and clinical and imaging risk factors

For each omics layer (biochemic, proteomic, metabolomic, and merged multi-omics), we retained Elastic Net models that selected more than one marker and significant in permutation testing. For each pPS, we then derived pMMSs across multiple omics layers by computing the weighted sum of the features selected by the corresponding model. Scores were constructed independently in SCAPIS and UKBB. For UKBB, insulin was excluded from Elastic Net models.

In participants without diabetes from the full SCAPIS cohort, we evaluated the associations between pMMSs and binary imaging outcomes using logistic regression, and with continuous clinical risk scores using Spearman correlation. All association analyses were adjusted for age, sex, and genetic PCs 1-10.

### Cox proportional hazards regression

We used Cox proportional hazards regression to evaluate associations of established genetic pPSs and newly derived pMMSs with incident T2D, incident T2D microvascular complications (diabetic nephropathy, neuropathy, and retinopathy), and T2D-related diseases and mortality (CAD, ischemic stroke, PAD, heart failure, cardiovascular disease mortality, all-cause mortality) in UKBB. All models were adjusted for age, sex, and genetic PCs 1-10.

All pPSs, pMMSs, and molecular markers were rank based inverse-normal transformed in UKBB prior to analysis. For biochemical pMMSs, we additionally fitted Cox models for T2D related clinical outcomes with further adjustment for HbA1_c_ and glucose.

### Multiple testing correction

All p-values were adjusted using false discovery rate (FDR) correction with a threshold of q < 0.05 within each analysis set.

### Protein functional annotation

Proteins were annotated using UniProt database(*16*) and gene sets annotation engine Enrichr(*17*), where the pathway relevant annotations were extracted from UniProt protein atlas, KEGG 2021 Human, and the MSigDB Hallmark 2020 libraries; the disease annotations were extracted from Elsevier pathway collection. The annotation for the whole protein set is provided in Table S8.

## Acknowledgments

We are grateful to all participants for generously dedicating their time to take part in the SCAPIS and UK Biobank study. This research was conducted under ethical approval number 2023-07277-01, using the SCAPIS resource under application number Petition-581 and the UK Biobank Resource under application number 554976. The authors acknowledge support from InfraVis for providing application expertise for visualization through the Swedish Research Council grant 2021-00181.

## Author contributions

Conceptualization: Clemens Wittenbecher, Hui Li

Methodology: Clemens Wittenbecher, Hui Li, Jakub Morze

Investigation: Hui Li, Jakub Morze

Visualization: Hui Li

Funding acquisition and supervision: Clemens Wittenbecher

Writing – original draft: Hui Li & Clemens Wittenbecher

Writing – review & editing: Jakub Morze, Martin Adiels, Joel Kullberg, Jordi Merino, Magdalena Sevilla-Gonzalez, Göran Bergström, Marju Orho-Melander, Stefan Söderberg, Carl Johan Östgren, Tuuli Lappalainen, Tove Fall, Miriam S Udler, Ida Häggström

## Data availability

The research data supporting the findings of this study comprise sensitive human participant data from the SCAPIS study (https://www.scapis.org) and the UK Biobank (https://www.ukbiobank.ac.uk). Due to ethical and legal restrictions related to participant privacy and data protection, access to SCAPIS and UK Biobank data is available for research and validation purposes through application to the respective data access committees and is subject to approval in accordance with institutional data access policies, ethical approvals, and data security requirements.

## Code availability

All code used to generate the results presented in this manuscript will be made publicly available upon completion of the revision process.

## References and Notes

1. R. A. DeFronzo, E. Ferrannini, L. Groop, R. R. Henry, W. H. Herman, J. J. Holst, F. B. Hu, C. R. Kahn, I. Raz, G. I. Shulman, D. C. Simonson, M. A. Testa, R. Weiss, Type 2 diabetes mellitus. Nat Rev Dis Primers 1 (2015), doi:10.1038/nrdp.2015.19.

2. S. E. Kahn, R. L. Hull, K. M. Utzschneider, Mechanisms linking obesity to insulin resistance and type 2 diabetes. Nature 444, 840–846 (2006).

3. E. Ahlqvist, P. Storm, A. Käräjämäki, M. Martinell, M. Dorkhan, A. Carlsson, P. Vikman, R. B. Prasad, D. M. Aly, P. Almgren, Y. Wessman, N. Shaat, P. Spégel, H. Mulder, E. Lindholm, O. Melander, O. Hansson, U. Malmqvist, Å. Lernmark, K. Lahti, T. Forsén, T. Tuomi, A. H. Rosengren, L. Groop, Novel subgroups of adult-onset diabetes and their association with outcomes: a data-driven cluster analysis of six variables. Lancet Diabetes Endocrinol 6, 361–369 (2018).

4. J. B. Meigs, The Genetic Epidemiology of Type 2 Diabetes: Opportunities for Health Translation. Curr Diab Rep 19 (2019), doi:10.1007/S11892-019-1173-Y.

5. K. Smith, A. J. Deutsch, C. McGrail, H. Kim, S. Hsu, A. Huerta-Chagoya, R. Mandla, P. H. Schroeder, K. E. Westerman, L. Szczerbinski, T. D. Majarian, V. Kaur, A. Williamson, N. Zaitlen, M. Claussnitzer, J. C. Florez, A. K. Manning, J. M. Mercader, K. J. Gaulton, M. S. Udler, Multi-ancestry polygenic mechanisms of type 2 diabetes. Nat Med (2024), doi:10.1038/s41591-024-02865-3.

6. M. S. Udler, J. Kim, M. von Grotthuss, S. Bonàs-Guarch, J. B. Cole, J. Chiou, M. Boehnke, M. Laakso, G. Atzmon, B. Glaser, J. M. Mercader, K. Gaulton, J. Flannick, G. Getz, J. C. Florez, Type 2 diabetes genetic loci informed by multi-trait associations point to disease mechanisms and subtypes: A soft clustering analysis. PLoS Med 15 (2018), doi:10.1371/journal.pmed.1002654.

7. D. Dicorpo, J. Leclair, J. B. Cole, C. Sarnowski, F. Ahmadizar, L. F. Bielak, A. Blokstra, E. P. Bottinger, L. Chaker, Y. D. I. Chen, Y. Chen, P. S. de Vries, T. Faquih, M. Ghanbari, V. Gudmundsdottir, X. Guo, N. R. Hasbani, D. Ibi, M. A. Ikram, M. Kavousi, H. L. Leonard, A. Leong, J. M. Mercader, A. C. Morrison, G. N. Nadkarni, M. A. Nalls, R. Noordam, M. Preuss, J. A. Smith, S. Trompet, P. Vissink, J. Yao, W. Zhao, E. Boerwinkle, M. O. Goodarzi, V. Gudnason, J. Wouter Jukema, S. L. R. Kardia, R. J. F. Loos, C. T. Liu, A. K. Manning, D. Mook-Kanamori, J. S. Pankow, H. S. J. Picavet, N. Sattar, E. M. Simonsick, W. M. M. Verschuren, K. W. van Dijk, J. C. Florez, J. I. Rotter, J. B. Meigs, J. Dupuis, M. S. Udler, Type 2 diabetes partitioned polygenic scores associate with disease outcomes in 454,193 individuals across 13 cohorts. Diabetes Care 45, 674–683 (2022).

8. M. Yapanis, S. James, M. E. Craig, D. O’Neal, E. I. Ekinci, Complications of Diabetes and Metrics of Glycemic Management Derived From Continuous Glucose Monitoring. J Clin Endocrinol Metab 107, e2221 (2022).

9. N. Esser, S. Legrand-Poels, J. Piette, A. J. Scheen, N. Paquot, Inflammation as a link between obesity, metabolic syndrome and type 2 diabetes. Diabetes Res Clin Pract 105, 141–150 (2014).

10. K. Yahagi, F. D. Kolodgie, C. Lutter, H. Mori, M. E. Romero, A. V. Finn, R. Virmani, Pathology of human coronary and carotid artery atherosclerosis and vascular calcification in diabetes mellitus. Arterioscler Thromb Vasc Biol 37, 191–204 (2017).

11. C. Faselis, A. Katsimardou, K. Imprialos, P. Deligkaris, M. Kallistratos, K. Dimitriadis, Microvascular Complications of Type 2 Diabetes Mellitus. Curr Vasc Pharmacol 18, 117–124 (2019).

12. G. Bergström, G. Berglund, A. Blomberg, J. Brandberg, G. Engström, J. Engvall, M. Eriksson, U. de Faire, A. Flinck, M. G. Hansson, B. Hedblad, O. Hjelmgren, C. Janson, T. Jernberg, A. Johnsson, L. Johansson, L. Lind, C. G. Löfdahl, O. Melander, C. J. Östgren, A. Persson, M. Persson, A. Sandström, C. Schmidt, S. Söderberg, J. Sundström, K. Toren, A. Waldenström, H. Wedel, J. Vikgren, B. Fagerberg, A. Rosengren, The Swedish CArdioPulmonary BioImage Study: objectives and design. J Intern Med 278, 645–659 (2015).

13. C. Sudlow, J. Gallacher, N. Allen, V. Beral, P. Burton, J. Danesh, P. Downey, P. Elliott, J. Green, M. Landray, B. Liu, P. Matthews, G. Ong, J. Pell, A. Silman, A. Young, T. Sprosen, T. Peakman, R. Collins, UK biobank: an open access resource for identifying the causes of a wide range of complex diseases of middle and old age. PLoS Med 12, e1001779–e1001779 (2015).

14. M. S. Udler, J. Kim, M. von Grotthuss, S. Bonàs-Guarch, J. B. Cole, J. Chiou, M. Boehnke, M. Laakso, G. Atzmon, B. Glaser, J. M. Mercader, K. Gaulton, J. Flannick, G. Getz, J. C. Florez, Type 2 diabetes genetic loci informed by multi-trait associations point to disease mechanisms and subtypes: A soft clustering analysis. PLoS Med 15, e1002654 (2018).

15. K. Suzuki, K. Hatzikotoulas, L. Southam, H. J. Taylor, X. Yin, K. M. Lorenz, R. Mandla, A. Huerta-Chagoya, G. E. M. Melloni, S. Kanoni, N. W. Rayner, O. Bocher, A. L. Arruda, K. Sonehara, S. Namba, S. S. K. Lee, M. H. Preuss, L. E. Petty, P. Schroeder, B. Vanderwerff, M. Kals, F. Bragg, K. Lin, X. Guo, W. Zhang, J. Yao, Y. J. Kim, M. Graff, F. Takeuchi, J. Nano, A. Lamri, M. Nakatochi, S. Moon, R. A. Scott, J. P. Cook, J. J. Lee, I. Pan, D. Taliun, E. J. Parra, J. F. Chai, L. F. Bielak, Y. Tabara, Y. Hai, G. Thorleifsson, N. Grarup, T. Sofer, M. Wuttke, C. Sarnowski, C. Gieger, D. Nousome, S. Trompet, S. H. Kwak, J. Long, M. Sun, L. Tong, W. M. Chen, S. S. Nongmaithem, R. Noordam, V. J. Y. Lim, C. H. T. Tam, Y. Y. Joo, C. H. Chen, L. M. Raffield, B. P. Prins, A. Nicolas, L. R. Yanek, G. Chen, J. A. Brody, E. Kabagambe, P. An, A. H. Xiang, H. S. Choi, B. E. Cade, J. Tan, K. A. Broadaway, A. Williamson, Z. Kamali, J. Cui, M. Thangam, L. S. Adair, A. Adeyemo, C. A. Aguilar-Salinas, T. S. Ahluwalia, S. S. Anand, A. Bertoni, J. Bork-Jensen, I. Brandslund, T. A. Buchanan, C. F. Burant, A. S. Butterworth, M. Canouil, J. C. N. Chan, L. C. Chang, M. L. Chee, J. Chen, S. H. Chen, Y. T. Chen, Z. Chen, L. M. Chuang, M. Cushman, J. Danesh, S. K. Das, H. J. de Silva, G. Dedoussis, L. Dimitrov, A. P. Doumatey, S. Du, Q. Duan, K. U. Eckardt, L. S. Emery, D. S. Evans, M. K. Evans, K. Fischer, J. S. Floyd, I. Ford, O. H. Franco, T. M. Frayling, B. I. Freedman, P. Genter, H. C. Gerstein, V. Giedraitis, C. González-Villalpando, M. E. González-Villalpando, P. Gordon-Larsen, M. Gross, L. A. Guare, S. Hackinger, L. Hakaste, S. Han, A. T. Hattersley, C. Herder, M. Horikoshi, A. G. Howard, W. Hsueh, M. Huang, W. Huang, Y. J. Hung, M. Y. Hwang, C. M. Hwu, S. Ichihara, M. A. Ikram, M. Ingelsson, M. T. Islam, M. Isono, H. M. Jang, F. Jasmine, G. Jiang, J. B. Jonas, T. Jørgensen, F. K. Kamanu, F. R. Kandeel, A. Kasturiratne, T. Katsuya, V. Kaur, T. Kawaguchi, J. M. Keaton, A. N. Kho, C. C. Khor, M. G. Kibriya, D. H. Kim, F. Kronenberg, J. Kuusisto, K. Läll, L. A. Lange, K. M. Lee, M. S. Lee, N. R. Lee, A. Leong, L. Li, Y. Li, R. Li-Gao, S. Ligthart, C. M. Lindgren, A. Linneberg, C. T. Liu, J. Liu, A. E. Locke, T. Louie, J. Luan, A. O. Luk, X. Luo, J. Lv, J. A. Lynch, V. Lyssenko, S. Maeda, V. Mamakou, S. R. Mansuri, K. Matsuda, T. Meitinger, O. Melander, A. Metspalu, H. Mo, A. D. Morris, F. A. Moura, J. L. Nadler, M. A. Nalls, U. Nayak, I. Ntalla, Y. Okada, L. Orozco, S. R. Patel, S. Patil, P. Pei, M. A. Pereira, A. Peters, F. J. Pirie, H. G. Polikowsky, B. Porneala, G. Prasad, L. J. Rasmussen-Torvik, A. P. Reiner, M. Roden, R. Rohde, K. Roll, C. Sabanayagam, K. Sandow, A. Sankareswaran, N. Sattar, S. Schönherr, M. Shahriar, B. Shen, J. Shi, D. M. Shin, N. Shojima, J. A. Smith, W. Y. So, A. Stančáková, V. Steinthorsdottir, A. M. Stilp, K. Strauch, K. D. Taylor, B. Thorand, U. Thorsteinsdottir, B. Tomlinson, T. C. Tran, F. J. Tsai, J. Tuomilehto, T. Tusie-Luna, M. S. Udler, A. Valladares-Salgado, R. M. van Dam, J. B. van Klinken, R. Varma, N. Wacher-Rodarte, E. Wheeler, A. R. Wickremasinghe, K. W. van Dijk, D. R. Witte, C. S. Yajnik, K. Yamamoto, K. Yamamoto, K. Yoon, C. Yu, J. M. Yuan, S. Yusuf, M. Zawistowski, L. Zhang, W. Zheng, S. Kanona, D. A. van Heel, L. J. Raffel, M. Igase, E. Ipp, S. Redline, Y. S. Cho, L. Lind, M. A. Province, M. Fornage, C. L. Hanis, E. Ingelsson, A. B. Zonderman, B. M. Psaty, Y. X. Wang, C. N. Rotimi, D. M. Becker, F. Matsuda, Y. Liu, M. Yokota, S. L. R. Kardia, P. A. Peyser, J. S. Pankow, J. C. Engert, A. Bonnefond, P. Froguel, J. G. Wilson, W. H. H. Sheu, J. Y. Wu, M. G. Hayes, R. C. W. Ma, T. Y. Wong, D. O. Mook-Kanamori, T. Tuomi, G. R. Chandak, F. S. Collins, D. Bharadwaj, G. Paré, M. M. Sale, H. Ahsan, A. A. Motala, X. O. Shu, K. S. Park, J. W. Jukema, M. Cruz, Y. D. I. Chen, S. S. Rich, R. McKean-Cowdin, H. Grallert, C. Y. Cheng, M. Ghanbari, E. S. Tai, J. Dupuis, N. Kato, M. Laakso, A. Köttgen, W. P. Koh, D. W. Bowden, C. N. A. Palmer, J. S. Kooner, C. Kooperberg, S. Liu, K. E. North, D. Saleheen, T. Hansen, O. Pedersen, N. J. Wareham, J. Lee, B. J. Kim, I. Y. Millwood, R. G. Walters, K. Stefansson, E. Ahlqvist, M. O. Goodarzi, K. L. Mohlke, C. Langenberg, C. A. Haiman, R. J. F. Loos, J. C. Florez, D. J. Rader, M. D. Ritchie, S. Zöllner, R. Mägi, N. A. Marston, C. T. Ruff, D. A. van Heel, S. Finer, J. C. Denny, T. Yamauchi, T. Kadowaki, J. C. Chambers, M. C. Y. Ng, X. Sim, J. E. Below, P. S. Tsao, K. M. Chang, M. I. McCarthy, J. B. Meigs, A. Mahajan, C. N. Spracklen, J. M. Mercader, M. Boehnke, J. I. Rotter, M. Vujkovic, B. F. Voight, A. P. Morris, E. Zeggini, Genetic drivers of heterogeneity in type 2 diabetes pathophysiology. Nature 2024 627:8003 627, 347–357 (2024).

16. T. U. Consortium, A. Bateman, M.-J. Martin, S. Orchard, M. Magrane, A. Adesina, S. Ahmad, E. H. Bowler-Barnett, H. Bye-A-Jee, D. Carpentier, P. Denny, J. Fan, P. Garmiri, L. J. da C. Gonzales, A. Hussein, A. Ignatchenko, G. Insana, R. Ishtiaq, V. Joshi, D. Jyothi, S. Kandasaamy, A. Lock, A. Luciani, J. Luo, Y. Lussi, J. S. M. Marin, P. Raposo, D. L. Rice, R. Santos, E. Speretta, J. Stephenson, P. Totoo, N. Tyagi, N. Urakova, P. Vasudev, K. Warner, S. Wijerathne, C. W.-H. Yu, R. Zaru, A. J. Bridge, L. Aimo, G. Argoud-Puy, A. H. Auchincloss, K. B. Axelsen, P. Bansal, D. Baratin, T. M. Batista Neto, M.-C. Blatter, J. T. Bolleman, E. Boutet, L. Breuza, B. C. Gil, C. Casals-Casas, K. C. Echioukh, E. Coudert, B. Cuche, E. de Castro, A. Estreicher, M. L. Famiglietti, M. Feuermann, E. Gasteiger, P. Gaudet, S. Gehant, V. Gerritsen, A. Gos, N. Gruaz, C. Hulo, N. Hyka-Nouspikel, F. Jungo, A. Kerhornou, P. Le Mercier, D. Lieberherr, P. Masson, A. Morgat, S. Paesano, I. Pedruzzi, S. Pilbout, L. Pourcel, S. Poux, M. Pozzato, M. Pruess, N. Redaschi, C. Rivoire, C. J. A. Sigrist, K. Sonesson, S. Sundaram, A. Sveshnikova, C. H. Wu, C. N. Arighi, C. Chen, Y. Chen, H. Huang, K. Laiho, M. Lehvaslaiho, P. McGarvey, D. A. Natale, K. Ross, C. R. Vinayaka, Y. Wang, J. Zhang, UniProt: the Universal Protein Knowledgebase in 2025. Nucleic Acids Res 53, D609–D617 (2025).

17. Z. Xie, A. Bailey, M. V. Kuleshov, D. J. B. Clarke, J. E. Evangelista, S. L. Jenkins, A. Lachmann, M. L. Wojciechowicz, E. Kropiwnicki, K. M. Jagodnik, M. Jeon, A. Ma’ayan, Gene Set Knowledge Discovery with Enrichr. Curr Protoc 1, e90 (2021).

18. E. K. Sokolowski, R. Kursawe, V. Selvam, R. M. Bhuiyan, A. Thibodeau, C. Zhao, C. N. Spracklen, D. Ucar, M. L. Stitzel, Multi-omic human pancreatic islet endoplasmic reticulum and cytokine stress response mapping provides type 2 diabetes genetic insights. Cell Metab 36, 2468–2488.e7 (2024).

19. Y. S. Lee, J. Wollam, J. M. Olefsky, An Integrated View of Immunometabolism. Cell 172, 22–40 (2018).

20. B. A. Ference, J. G. Robinson, R. D. Brook, A. L. Catapano, M. J. Chapman, D. R. Neff, S. Voros, R. P. Giugliano, G. Davey Smith, S. Fazio, M. S. Sabatine, Variation in PCSK9 and HMGCR and Risk of Cardiovascular Disease and Diabetes. New England Journal of Medicine 375, 2144–2153 (2016).

21. A. Varbo, M. Benn, A. Tybjærg-Hansen, A. B. Jørgensen, R. Frikke-Schmidt, B. G. Nordestgaard, Remnant cholesterol as a causal risk factor for ischemic heart disease. J Am Coll Cardiol 61, 427–436 (2013).

22. J. Leipsic, S. Abbara, S. Achenbach, R. Cury, J. P. Earls, G. B. J. Mancini, K. Nieman, G. Pontone, G. L. Raff, SCCT guidelines for the interpretation and reporting of coronary CT angiography: A report of the Society of Cardiovascular Computed Tomography Guidelines Committee. J Cardiovasc Comput Tomogr 8, 342–358 (2014).

23. G. Bergström, M. Persson, M. Adiels, E. Björnson, C. Bonander, H. Ahlström, J. Alfredsson, O. Angerås, G. Berglund, A. Blomberg, J. Brandberg, M. Börjesson, K. Cederlund, U. De Faire, O. Duvernoy, Ö. Ekblom, G. Engström, J. E. Engvall, E. Fagman, M. Eriksson, D. Erlinge, B. Fagerberg, A. Flinck, I. Gonçalves, E. Hagström, O. Hjelmgren, L. Lind, E. Lindberg, P. Lindqvist, J. Ljungberg, M. Magnusson, M. Mannila, H. Markstad, M. A. Mohammad, F. H. Nystrom, E. Ostenfeld, A. Persson, A. Rosengren, A. Sandström, A. Själander, M. C. Sköld, J. Sundström, E. Swahn, S. Söderberg, K. Torén, C. J. Östgren, T. Jernberg, Prevalence of Subclinical Coronary Artery Atherosclerosis in the General Population. Circulation 144, 916 (2021).

24. S. M. Grundy, N. J. Stone, A. L. Bailey, C. Beam, K. K. Birtcher, R. S. Blumenthal, L. T. Braun, S. de Ferranti, J. Faiella-Tommasino, D. E. Forman, R. Goldberg, P. A. Heidenreich, M. A. Hlatky, D. W. Jones, D. Lloyd-Jones, N. Lopez-Pajares, C. E. Ndumele, C. E. Orringer, C. A. Peralta, J. J. Saseen, S. C. Smith, L. Sperling, S. S. Virani, J. Yeboah, 2018 AHA/ACC/AACVPR/AAPA/ABC/ACPM/ADA/AGS/APhA/ASPC/NLA/PCNA Guideline on the Management of Blood Cholesterol: Executive Summary: A Report of the American College of Cardiology/American Heart Association Task Force on Clinical Practice Guidelines. J Am Coll Cardiol 73, 3168–3209 (2019).

25. R. Detrano, A. D. Guerci, J. J. Carr, D. E. Bild, G. Burke, A. R. Folsom, K. Liu, S. Shea, M. Szklo, D. A. Bluemke, D. H. O’Leary, R. Tracy, K. Watson, N. D. Wong, R. A. Kronmal, Coronary calcium as a predictor of coronary events in four racial or ethnic groups. N Engl J Med 358, 1336–1345 (2008).

26. S. C. Ritchie, P. Surendran, S. Karthikeyan, S. A. Lambert, T. Bolton, L. Pennells, J. Danesh, E. Di Angelantonio, A. S. Butterworth, M. Inouye, Quality control and removal of technical variation of NMR metabolic biomarker data in ∼120,000 UK Biobank participants. Scientific Data 2023 10:1 10, 1–15 (2023).

