## Supplementary material for "Multi-molecular scores map process-specific polygenic diabetes risk to atherosclerosis, cardiometabolic diseases, and vascular complications"

**Index of Supplemental Materials**


| Item | Title | Page(s) |
|---|---|---|
| **Supplementary Figures** | | |
| Fig. S1 | Rank-based inverse normal transformation of European ancestry-based pPSs in SCAPIS cohort. | 2 |
| Fig. S2 | Associations between pPSs, anthropometric traits, and clinical biomarkers in SCAPIS. | 3-4 |
| Fig. S3 | Associations between pPSs and lipoprotein subfraction in SCAPIS. | 5-6 |
| Fig. S4 | Key molecular predictors of pPSs identified by Elastic Net regression models in SCAPIS. | 7-8 |
| Fig. S5 | Associations between pPSs, pMMSs, and subclinical atherosclerosis indices in SCAPIS. | 9 |
| Fig. S6 | Logistic regression coefficients of pPSs and pMMSs with binary transformed atherosclerosis and stenosis indices in SCAPIS. | 10 |
| Fig. S7 | Associations of pPSs with health history in SCAPIS. | 11 |
| Fig. S8 | Hazard ratios for the associations of biochemical pMMSs with incident T2D and related outcomes in UKBB, before and after adjustment for $HbA1_c$ and glucose. | 12-13 |
| **Supplementary Tables** | | |
| Table S1 | SNP weight matrix for pPS construction. | 14-18 |
| Table S2 | Summary of clinical, molecular, and imaging-derived variables in SCAPIS, and disease outcomes in UKBB. | 19-23 |
| Table S3 | pPS cross-correlation in SCAPIS. | 24-26 |
| Table S4 | Associations between pPSs and anthropometric traits in SCAPIS. | 27-28 |
| Table S5 | Associations between pPSs and biochemical traits in SCAPIS. | 29-32 |
| Table S6 | Associations between pPSs and selected metabolites in SCAPIS. | 33-36 |
| Table S7 | Associations between pPSs and plasma proteins in SCAPIS. | 37-62 |
| Table S8 | Protein functional and pathway annotations. | 63-64 |
| Table S9 | Associations between pPSs and global metabolomic profile in SCAPIS. | 65-87 |
| Table S10 | Elastic Net regression summary statistics (generated from SCAPIS). | 88-89 |
| Table S11 | Elastic Net model coefficients (generated from SCAPIS). | 90-100 |
| Table S12 | Associations between pPSs, pMMSs and imaging risk scores in SCAPIS. | 101-103 |
| Table S13 | pPS associations with Categorical Coronary Artery Calcium Scores (CACS) in SCAPIS. | 104 |
| Table S14 | Associations between pPSs, pMMSs and imaging-derived phenotypes in SCAPIS. | 105-108 |
| Table S15 | Associations between pPSs and health history in SCAPIS. | 109-111 |
| Table S16 | Associations between pPSs, pMMSs and T2D-related disease outcomes in UKBB. | 112-117 |
| Table S17 | Associations of biochemical pMMSs with T2D-related disease outcomes in UKBB, before and after adjustment for HbA1c and glucose. | 118-120 |

### Supplementary Materials

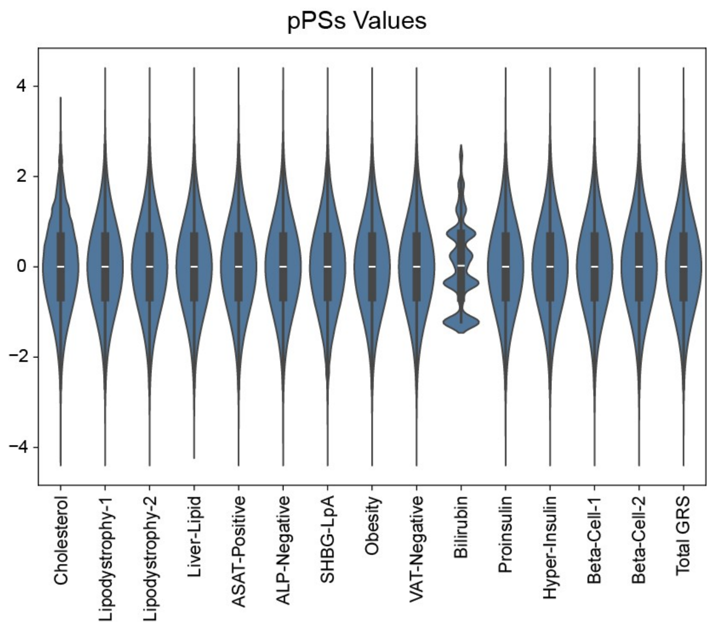


**Fig. S1. Rank-based inverse normal transformation of European ancestry-based pPSs in SCAPIS cohort.** Distribution of individual values across the 14 pPSs and total GRS after normal transformation.

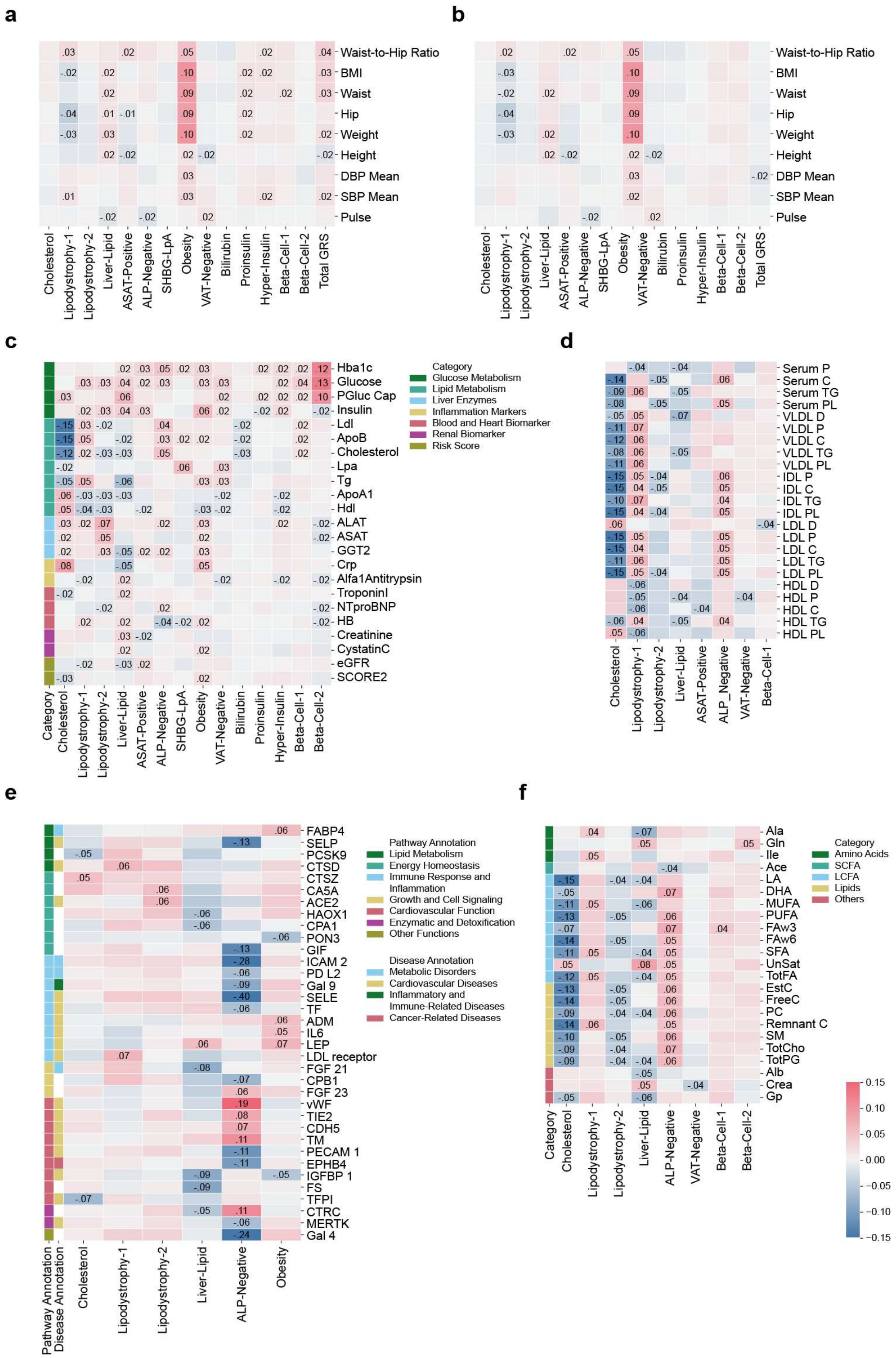
a
Waist-to-Hip Ratio
BMI
Waist
Hip
Weight
Height
DBP Mean
SBP Mean
Pulse
Cholesterol
Lipodystrophy-1
Lipodystrophy-2
Liver-Lipid
ASAT-Positive
ALP-Negative
SHBG-LpA
Obesity
VAT-Negative
Bilirubin
Proinsulin
Hyper-Insulin
Beta-Cell-1
Beta-Cell-2
Total GRS
b
Waist-to-Hip Ratio
BMI
Waist
Hip
Weight
Height
DBP Mean
SBP Mean
Pulse
Cholesterol
Lipodystrophy-1
Lipodystrophy-2
Liver-Lipid
ASAT-Positive
ALP-Negative
SHBG-LpA
Obesity
VAT-Negative
Bilirubin
Proinsulin
Hyper-Insulin
Beta-Cell-1
Beta-Cell-2
Total GRS
c
Hba1c
Glucose
PGluc Cap
Insulin
Ldl
ApoB
Cholesterol
Lpa
Tg
ApoA1
Hdl
ALAT
ASAT
GGT2
Crp
Alfa1Antitrypsin
TroponinI
NTproBNP
HB
Creatinine
CystatinC
eGFR
SCORE2
Category
Glucose Metabolism
Lipid Metabolism
Liver Enzymes
Inflammation Markers
Blood and Heart Biomarker
Renal Biomarker
Risk Score
Category
Cholesterol
Lipodystrophy-1
Lipodystrophy-2
Liver-Lipid
ASAT-Positive
ALP-Negative
SHBG-LpA
Obesity
VAT-Negative
Bilirubin
Proinsulin
Hyper-Insulin
Beta-Cell-1
Beta-Cell-2
d
Serum P
Serum C
Serum TG
Serum PL
VLDL D
VLDL P
VLDL C
VLDL TG
VLDL PL
IDL P
IDL C
IDL TG
IDL PL
LDL D
LDL P
LDL C
LDL TG
LDL PL
HDL D
HDL P
HDL C
HDL TG
HDL PL
Cholesterol
Lipodystrophy-1
Lipodystrophy-2
Liver-Lipid
ASAT-Positive
ALP_Negative
VAT-Negative
Beta-Cell-1
e
FABP4
SELP
PCSK9
CTSD
CTSZ
CA5A
ACE2
HAOX1
CPA1
PON3
GIF
ICAM 2
PD L2
Gal 9
SELE
TF
ADM
IL6
LEP
LDL receptor
FGF 21
CPB1
FGF 23
vWF
TIE2
CDH5
TM
PECAM 1
EPHB4
IGFBP 1
FS
TFPI
CTRC
MERTK
Gal 4
Pathway Annotation
Lipid Metabolism
Energy Homeostasis
Immune Response and Inflammation
Growth and Cell Signaling
Cardiovascular Function
Enzymatic and Detoxification
Other Functions
Disease Annotation
Metabolic Disorders
Cardiovascular Diseases
Inflammatory and Immune-Related Diseases
Cancer-Related Diseases
Pathway Annotation
Disease Annotation
Cholesterol
Lipodystrophy-1
Lipodystrophy-2
Liver-Lipid
ALP-Negative
Obesity
f
Ala
Gln
Ile
Ace
LA
DHA
MUFA
PUFA
FAw3
FAw6
SFA
UnSat
TotFA
EstC
FreeC
PC
Remnant C
SM
TotCho
TotPG
Alb
Crea
Gp
Category
Amino Acids
SCFA
LCFA
Lipids
Others
Category
Cholesterol
Lipodystrophy-1
Lipodystrophy-2
Liver-Lipid
ALP-Negative
VAT-Negative
Beta-Cell-1
Beta-Cell-2
0.15
0.10
0.05
0.00
−0.05
−0.10
−0.15

**Fig. S2. Associations between pPSs, anthropometric traits, and clinical biomarkers in SCAPIS.**

(**a**) Correlation of pPSs with anthropometric traits. (**b**) Partial correlations of pPSs with anthropometric traits. (**c**) Partial correlations of pPSs with biochemical markers. (**d**) Partial correlations of pPSs with proteins. (**e**) Partial correlations of pPSs with NMR-detected lipoprotein traits. (**f**) Partial correlations between pPSs and the remaining NMR-metabolomics markers. All plots show only molecular features with at least one significant pPS association; significant coefficients are annotated with two decimal places. _P: Particle concentration; _C: Total cholesterol; _TG: Triglycerides; _PL: Phospholipids; _D: Mean particle diameter; SCFA: Short-chain fatty acids; LCFA: Long-chain fatty acids.

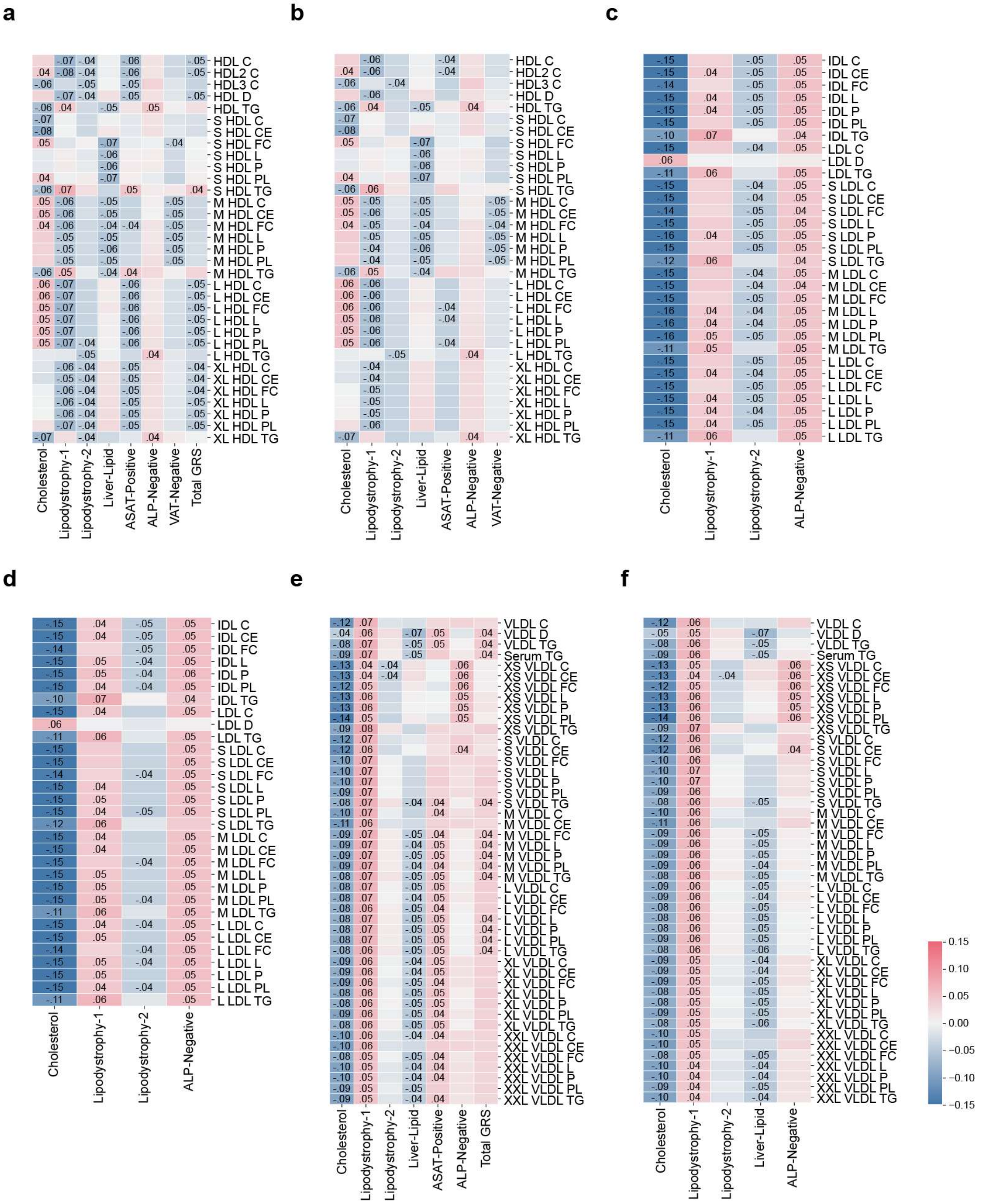


**Fig. S3. Associations between pPSs and lipoprotein subfraction in SCAPIS.**

(**a**) Correlation of pPSs with HDL related particles. (**b**) Partial correlations of pPSs with HDL related particles. (**c**) Correlation of pPSs with IDL related particles. (**d**) Partial correlations of pPSs with IDL related particles. (**e**) Correlation of pPSs with VLDL related particles. (**f**) Partial correlations of pPSs with VLDL related particles. All plots show only molecular features with at least one significant pPS association; significant coefficients are annotated with two decimal places. XS : very small; S: small; M: medium; L: large; XL: very large; XXL VLDL: chylomicrons and extremely large VLDL; _P: particle concentration; _PL: phospholipids; _TG: triglycerides; _FC: free cholesterol; _CE: cholesterol esters; _C: total cholesterol; _L: total lipids; _D: mean particle diameter.

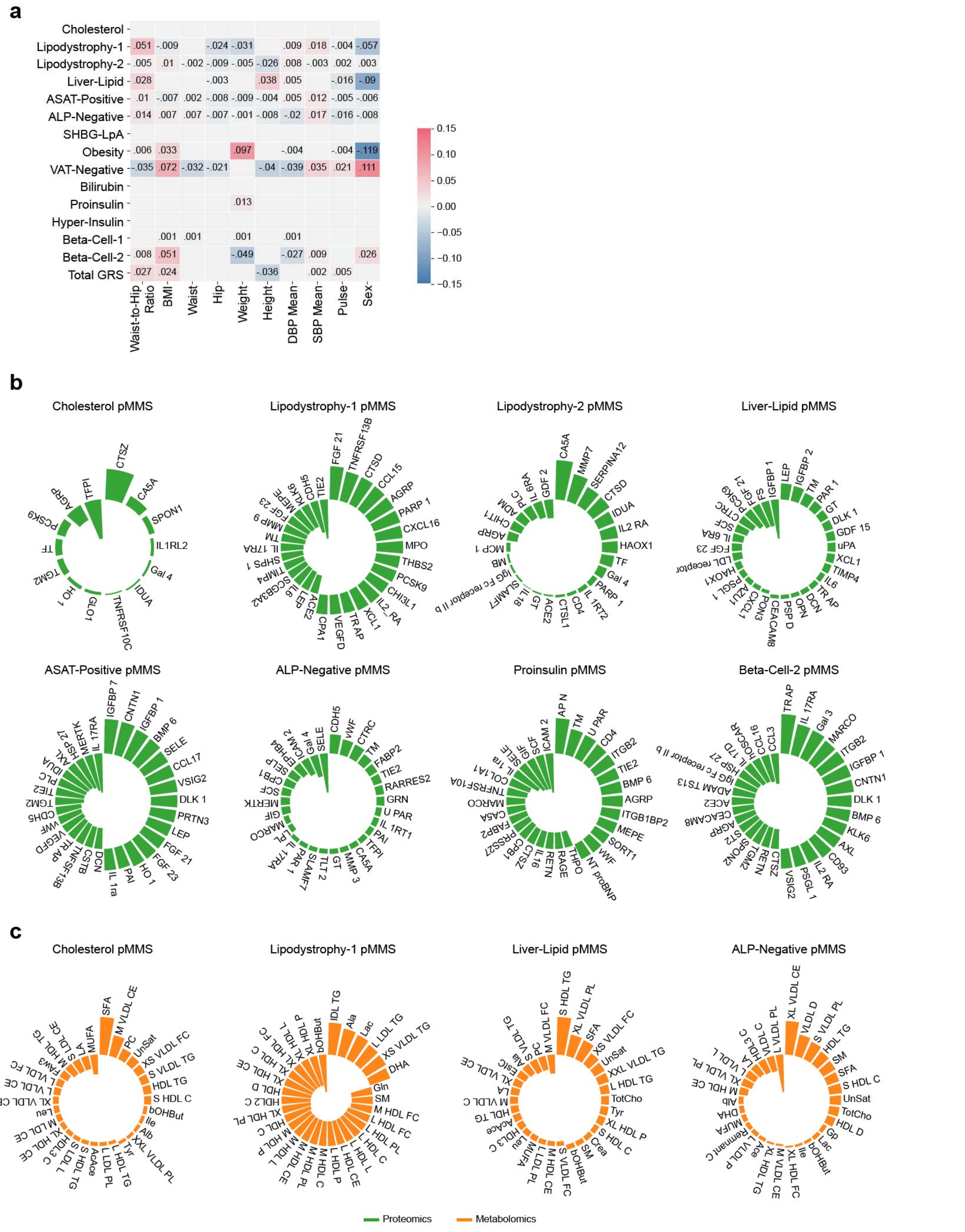


**Fig. S4. Key molecular predictors of pPSs identified by Elastic Net regression models in SCAPIS.**

(**a**) Coefficient matrix from Elastic Net models predicting pPS values (y-axis) from anthropometric traits (x-axis, sex coded as 0 for female and 1 for male). Coefficients with absolute values that remained non-zero after rounding to three decimal places are annotated for all models. (**b**) The coefficients of proteomic-based Elastic Net models including more than one marker and significant in permutation testing are shown in circular plot with up to top 30 markers. (**c**) The coefficients of NMR-metabolomic-based Elastic Net models including more than one marker and significant in permutation testing are shown in circular plot with up to top 30 markers. XS: very small; S: small; M: medium; L: large; XL: very large; XXL VLDL: chylomicrons and extremely large VLDL; _P: particle concentration; _PL: phospholipids; _TG: triglycerides; _FC: free cholesterol; _CE: cholesterol esters; _C: total cholesterol; _L: total lipids; _D: mean particle diameter.

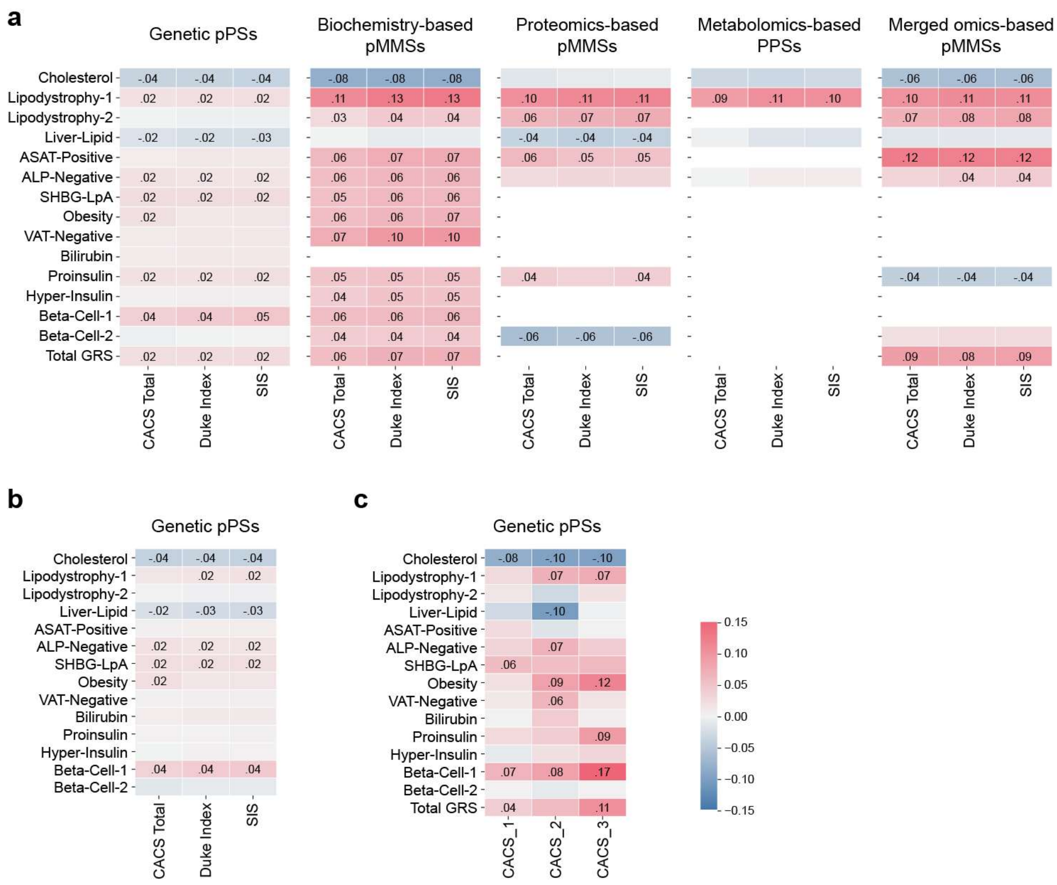


**Fig. S5. Associations between pPSs, pMMSs, and subclinical atherosclerosis indices in SCAPIS.**

(**a**) Matrix of association coefficients between genetically derived pPSs and multi-biomarker pMMs. pMMSs were derived using Elastic Net; only models include more than one marker and significant in permutation-testing were retained. (**b**) Partial correlation coefficients between genetically derived pPSs and imaging-based subclinical atherosclerosis indices. (**c**) Regression analysis of genetically derived pPSs and categorized coronary artery calcium score (CACS). CACS categories were encoded as follows: CACS = 0 coded as [0, 0, 0]; 0 < CACS ≤ 99 coded as [1, 0, 0]; 99 < CACS ≤ 300 coded as [0, 1, 0]; and CACS > 300 coded as [0, 0, 1]. CACS_1, CACS_2, and CACS_3 represent the first, second, and third dimensions of the encoded CACS, respectively. Significant association coefficients are annotated with two decimal places.

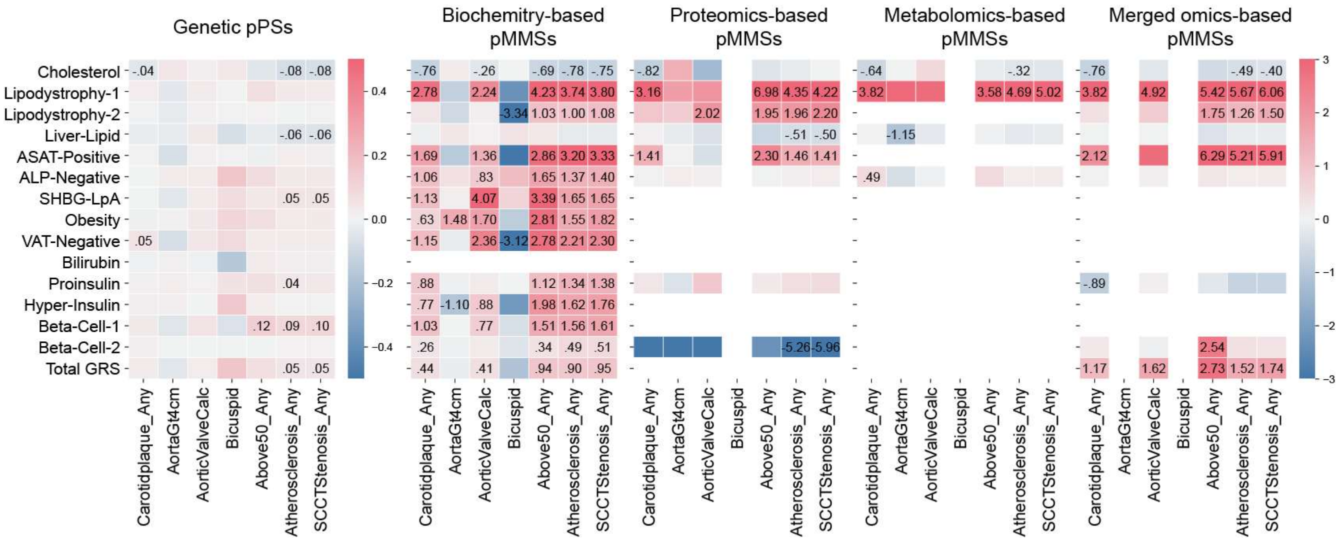


**Fig. S6. Logistic regression coefficients of pPSs and pMMSs with binary transformed atherosclerosis and stenosis indices in SCAPIS.**

Significant regression coefficients are annotated with two decimal places. pMMSs were derived using Elastic Net; only models include more than one marker and significant in permutation-testing were retained. Logistic regression analyses were conducted only for outcomes with > 100 events.

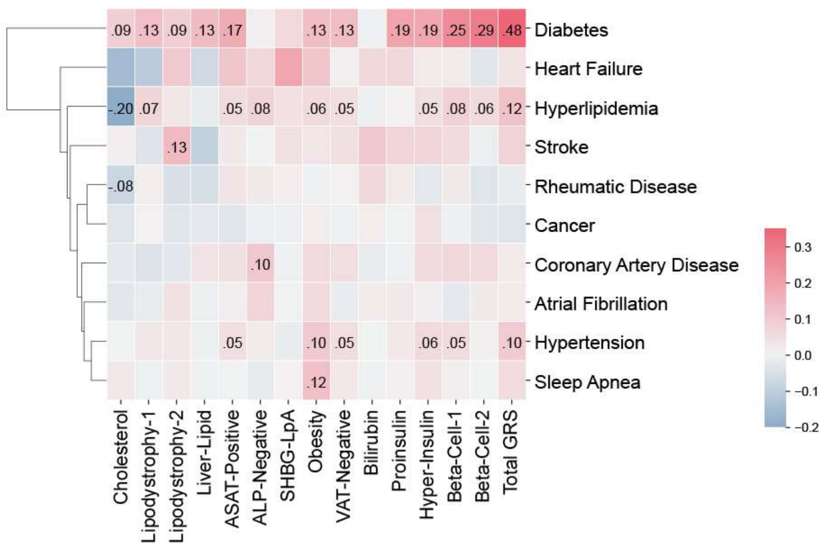


**Fig. S7. Associations of pPSs with health history in SCAPIS.**

Logistic regression between genetic pPSs and health history in SCAPIS. Significant regression coefficients are annotated with two decimal places. Most of the genetic T2D-pPSs were associated with the prevalence of at least one clinical diagnosis, including hyperlipidemia, stroke, CAD, hypertension, or sleep apnea. The Cholesterol pPS was inversely associated with hyperlipidemia. The ALP-Negative pPS showed particularly strong associations with hyperlipidemia and CAD, consistent with its molecular signatures. The ASAT-Positive, Obesity, VAT-Negative, Hyper-Insulin, and Beta-Cell-1 pPSs were linked to the prevalence of multiple cardiometabolic diseases.

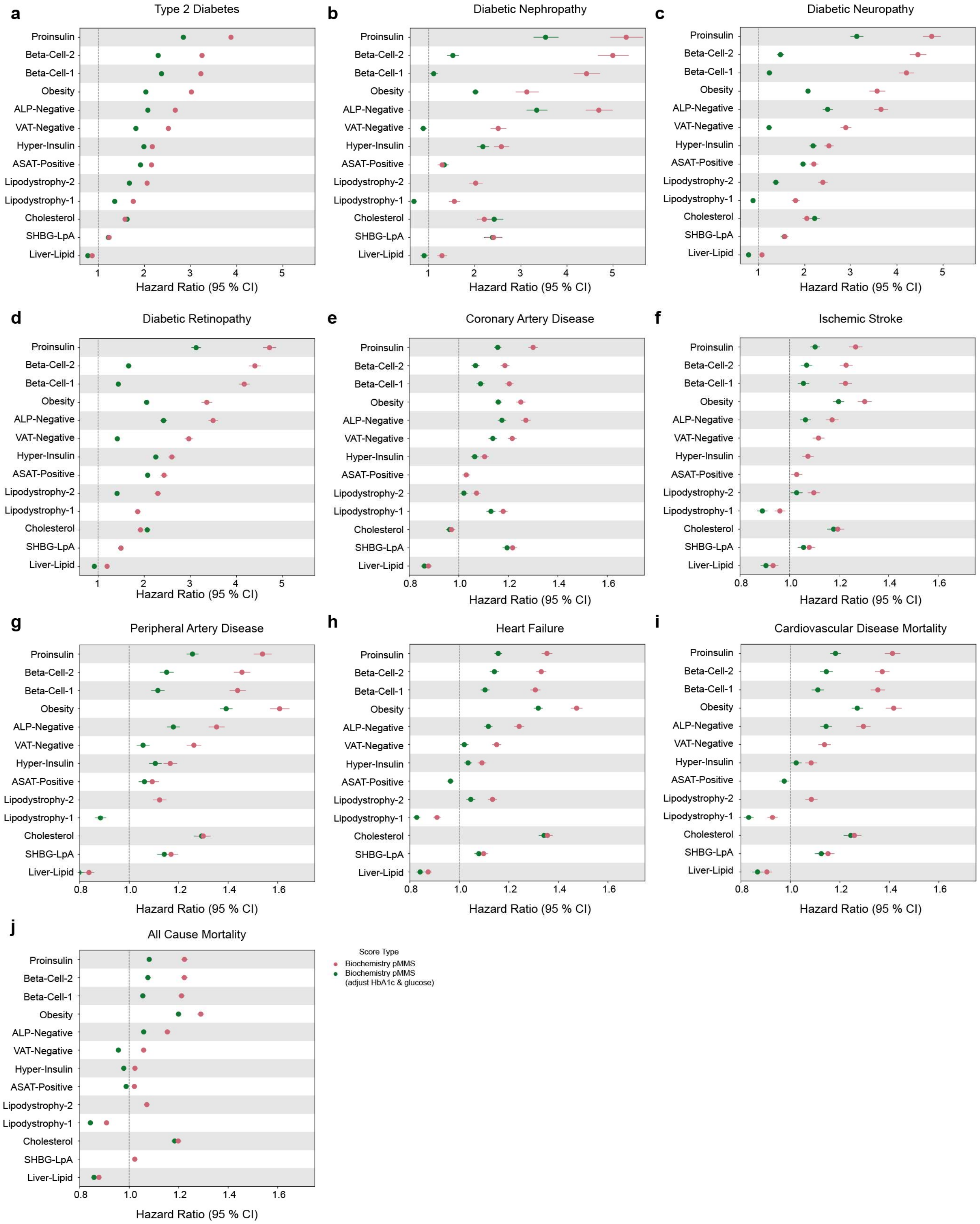
a
Type 2 Diabetes
b
Diabetic Nephropathy
c
Diabetic Neuropathy
d
Diabetic Retinopathy
e
Coronary Artery Disease
f
Ischemic Stroke
g
Peripheral Artery Disease
h
Heart Failure
i
Cardiovascular Disease Mortality
j
All Cause Mortality
Proinsulin
Beta-Cell-2
Beta-Cell-1
Obesity
ALP-Negative
VAT-Negative
Hyper-Insulin
ASAT-Positive
Lipodystrophy-2
Lipodystrophy-1
Cholesterol
SHBG-LpA
Liver-Lipid
Hazard Ratio (95 % CI)
Score Type
Biochemistry pMMS
Biochemistry pMMS
(adjust HbA1c & glucose)

**Fig. S8. Hazard ratios for the associations of biochemical pMMSs with incident T2D and related outcomes in UKBB, before and after adjustment for $HbA1_c$ and glucose.** HRs per SD for biochemical pMMSs and for the same models additionally adjusted for $HbA1_c$ and glucose with: (**a**) type 2 diabetes (T2D), (**b**) diabetic nephropathy, (**c**) diabetic neuropathy, (**d**) diabetic retinopathy, (**e**) coronary artery disease, (**f**) ischemic stroke, (**g**) peripheral artery disease, (**h**) heart failure, (**i**) cardiovascular disease mortality, and (**j**) all-cause mortality. The pMMSs were derived using Elastic Net Regression, predicting the pPS with the biochemistry markers; only FDR-significant models (permutation test) with more than one maker are retained. Biological processes in all panels are sorted by descending biochemical-based pMMS HRs for T2D. Points denote HRs per SD with 95% confidence intervals (CI) from Cox proportional hazards models.

**Table S1: SNP weight matrix for pPS construction**

| RSID | Effect_Allele | Gene | Cholesterol_pPS | Lipodystrophy_1_pPS | Lipodystrophy_2_pPS | Liver-Lipid_pPS | ASAT_Pos_pPS | ALP_Neg_pPS | SHBG-LpA_pPS | Obesity_pPS | VAT_Neg_pPS | Bilirubin_pPS | Proinsulin_pPS | Hyper_Insulin_pPS | Beta_Cell_1_pPS | Beta_Cell_2_pPS | Total_GRS |
|---|---|---|---|---|---|---|---|---|---|---|---|---|---|---|---|---|---|
| rs174541 | T | FADS1 | 0.739696069 | | | 1.502964071 | | 2.171701449 | | | | | | | 1.590062273 | | 0.0292 |
| rs253412 | A | ANKDD1B | 0.783887678 | | | | | | | 1.153291238 | | | 0.785428512 | | | | 0.0459 |
| rs58542926 | T | TM6SF2 | 1.886152842 | | 1.822228601 | | | | | | | | | | | | 0.089 |
| rs4803764 | T | NECTIN2 | 2.883512587 | | | | | | | | | | | | | | 0.0342 |
| rs429358 | T | APOE | 7.768353506 | | | | | | | 1.005399437 | | | | | | | 0.073 |
| rs1641523 | C | ATP1B2 | | 0.740951629 | | | | | 5.634739671 | | | | | | | | 0.0255 |
| rs17622532 | A | LINC02915 | | 0.747724121 | | | | | | | | | | 0.997518161 | | | 0.026 |
| rs17175860 | G | INSR | | 0.754031453 | | | | | | | | | | | | | 0.0445 |
| rs890940 | T | EBF1 | | 0.759547663 | | | 0.838594967 | | | | 0.76040755 | | | | 0.773259704 | | 0.0484 |
| rs11642655 | A | CMIP | | 0.775114585 | | | | | | | | | | 0.858903665 | | | 0.0288 |
| rs12185610 | A | SPRED2 | | 0.786637993 | | | | | | | | | | | | | 0.0451 |
| rs7609422 | G | TEX41 | | 0.787122369 | | | | | | | | | | | | | 0.0295 |
| rs25643 | C | P2RX4 | | 0.789629638 | | | | | | | | | | | | | 0.0248 |
| rs1513272 | C | JAZF1 | | 0.802795869 | | | | | | | | | | | | 0.796075849 | 0.0811 |
| rs1596972 | G | KLF14 | | 0.805694198 | 0.998770029 | | | | | | 1.033251674 | | | | | | 0.0403 |
| rs1705263 | C | TSPAN8 | | 0.808572961 | | | | | | | | | | | | | 0.0398 |
| rs12454712 | T | BCL2 | | 0.811591683 | 0.845392523 | | 1.252358877 | | | | | | | | | | 0.0412 |
| rs8756 | A | HMGA2 | | 0.831178432 | | | 1.535972739 | | | | | | | | | | 0.0432 |
| rs116843064 | G | ANGPTL4 | | 0.831761421 | | | | | | | | | | | | | 0.0965 |
| rs2908797 | C | MPHOSPH10P1 | | 0.832470382 | | | | | | | | | | | | | 0.0363 |
| rs8126001 | C | OPRL1 | | 0.837788373 | | | | | | | | | | | | | 0.0281 |
| rs9826367 | A | PPARG | | 0.846334489 | 0.928307718 | | | | | | | | | | | | 0.0402 |
| rs73175562 | A | IGF2BP2 | | 0.866375715 | | | | | | | | | | | | | 0.0531 |
| rs2426439 | C | ZFP64 | | 0.872502922 | | | | | | 0.967245183 | | | | | | | 0.0365 |
| rs12263348 | T | REEP3 | | 0.875831655 | | | | | 2.200270423 | | | | | 1.33440212 | | | 0.0282 |
| rs2242186 | A | BMF | | 0.883526717 | | | | | | | | | | | | | 0.0643 |
| rs2268078 | A | RALY | | 0.886389371 | | | | | | | | | | | | | 0.0387 |
| rs891368 | G | RFT1 | | 0.898953308 | | | | | | | | | | | | | 0.0247 |
| rs61779275 | T | MACF1 | | 0.90565653 | | 0.745213025 | | | | | | | | | | | 0.0753 |
| rs56008051 | C | BHLHE41 | | 0.907557365 | | | | | | | | | | | 0.830199393 | | 0.032 |
| rs355799 | C | COBLL1 | | 0.959670397 | | | | | | | | | | 0.995210581 | | | 0.0439 |
| rs4976033 | G | PIK3R1 | | 0.970214727 | 0.927069934 | | | | | | | | | 1.046372952 | | | 0.028 |
| rs10937208 | G | EHHADH-AS1 | | 0.971056303 | | | | | | | | | | | | | 0.0443 |
| rs34048824 | T | DNMT3A | | 0.97762937 | | | | | | | | | | | | | 0.0323 |
| rs12820906 | A | PITPNM2 | | 0.994715587 | | | | | | | | | | | | | 0.0433 |
| rs62262091 | T | SMARCC1 | | 1.013882886 | | | | | | 0.851811628 | | | | | | | 0.0557 |
| rs7660000 | C | FAM13A | | 1.045643164 | | | | | | | | | | | | | 0.0309 |
| rs7544489 | T | MTOR | | 1.062952593 | | | 0.840788 | | | | | | | | | | 0.027 |
| rs60710264 | C | PPARG | | 1.11936111 | | | | | | | | | | | 1.040503029 | | 0.0374 |
| rs2306363 | G | SIPA1 | | 1.120359335 | | | | | | | | | | | | | 0.0438 |
| rs17231538 | A | LOC646736 | | 1.128336547 | | | | | | | | | | | | | 0.076 |
| rs4925114 | A | RAI1 | | 1.142238582 | | | | | | | | | | | | | 0.033 |
| rs9854769 | G | IGF2BP2 | | 1.156443427 | | | | | | | | | | | | 0.918261576 | 0.1075 |
| rs4709746 | C | QKI | | 1.164061963 | | | | | | | | | | | | | 0.0581 |
| rs12491473 | G | CCDC12 | | 1.211150669 | | | 0.85654076 | | | | | | | | | | 0.0215 |
| rs2267373 | T | MAFF | | 1.234841845 | | | | | | | | | | | | | 0.0284 |
| rs28819812 | C | PDGFC | | 1.280808732 | | | 1.176842623 | | | | | | | 1.329658907 | | | 0.0384 |
| rs10096633 | C | LPL | | 1.332956393 | 1.4733489 | | | | | | | | | | | | 0.0404 |
| rs11057376 | T | DNAH10 | | 1.391594518 | | | | | | | | | | | | | 0.0394 |
| rs17036160 | C | PPARG | | 1.460900547 | 2.029112536 | | | | | | 0.969348805 | | | | | | 0.1028 |
| rs4916253 | G | DNM3 | | 1.50024674 | | | | | | | | | | | | | 0.0274 |
| rs9687846 | A | MAP3K1 | | 1.539219108 | | | 2.090455283 | | | | | | | | | | 0.0659 |
| rs4132228 | C | MIR548A2 | | 1.74665907 | | | | | | | | | | | | | 0.0471 |
| rs77136196 | T | RPS10-NUDT3 | | 1.825515886 | | | | | | | | | | | | | 0.0662 |
| rs13131633 | T | FAM13A | | 1.919157022 | | | | | | | | | | | | | 0.0231 |
| rs2925979 | T | CMIP | | 2.094870208 | | | | | | | | | | 0.827427027 | | | 0.0448 |
| rs35169799 | T | PLCB3 | | 2.101718928 | | | | | | | | | | | | | 0.0504 |
| rs2820444 | G | SLC30A10 | | 2.178492439 | | | | | | | | | | | | | 0.0472 |
| rs11048458 | T | ITPR2 | | 2.204558141 | | | | | | | | | | | | | 0.0458 |
| rs10773051 | C | ZNF664-RFLNA | | 2.341734578 | | | | | | | | | | | | | 0.035 |
| rs2972145 | C | LOC646736 | | 2.561065798 | 1.890151609 | | | | | | | | | | | | 0.09 |
| rs11155073 | T | LINC01625 | | 2.747243184 | | | | 1.701221845 | | | | | | | | | 0.0304 |
| rs13389219 | C | COBLL1 | | 2.855698825 | 1.140631041 | | | | | | | | | | | | 0.0648 |
| rs12823740 | C | ZNF664 | | 2.987252062 | | | | | | | | | | 1.211205048 | | | 0.041 |
| rs998584 | A | VEGFA | | 3.28618196 | | | 0.974305567 | | | | | | | | 1.368077306 | | 0.0365 |
| rs2287821 | C | PEPD | | | 0.745393408 | | | | | | 0.84528604 | | | | | | 0.0239 |
| rs62496027 | A | PRAG1 | | | 0.74940589 | | | | | | | | | | | | 0.0276 |
| rs8033609 | A | RORA | | | 0.751884212 | | | | | | | | | | | | 0.0265 |
| rs67611724 | T | PXMP4 | | | 0.775716634 | | 0.880527158 | | | | | | | | | | 0.0443 |
| rs756145 | A | DMRT2 | | | 0.780012722 | | | | | | 0.851590616 | | | 0.852062118 | | | 0.0286 |
| rs34990153 | A | MSRA | | | 0.789724774 | | | | | | | | 0.77916001 | | | | 0.0384 |
| rs2115107 | A | MAP2K7 | | | 0.813833592 | | | | | | | | | 1.386353037 | | | 0.0382 |
| rs11604324 | C | NUP160 | | | 0.824748485 | | | | | | | | | | | | 0.0736 |
| rs1009358 | T | CEP68 | | | 0.862733719 | | | | | | | | | 1.228929318 | | | 0.0493 |

**Table S1: SNP weight matrix for pPS construction**

| RSID | Effect_Allele | Gene | Cholesterol_pPS | Lipodystrophy_1_pPS | Lipodystrophy_2_pPS | Liver-Lipid_pPS | ASAT_Pos_pPS | ALP_Neg_pPS | SHBG-LpA_pPS | Obesity_pPS | VAT_Neg_pPS | Bilirubin_pPS | Proinsulin_pPS | Hyper_Insulin_pPS | Beta_Cell_1_pPS | Beta_Cell_2_pPS | Total_GRS |
|---|---|---|---|---|---|---|---|---|---|---|---|---|---|---|---|---|---|
| rs79598313 | T | KDF1 | | | 0.888015136 | | | | 1.769887989 | | | | | | | | 0.084 |
| rs6066138 | G | EYA2 | | | 0.919852488 | | | | | | | | | | | | 0.0451 |
| rs4865796 | A | ARL15 | | | 0.929463279 | | 1.087421244 | | | | | | | | | | 0.0466 |
| rs1800961 | T | HNF4A | | | 1.110082382 | 1.133214397 | | | | | | | | | | | 0.1077 |
| rs2126263 | G | LOC157273 | | | 1.170448828 | 2.399613907 | | | | | | | | 1.073785505 | | | 0.0608 |
| rs9852406 | T | PPP2R3A | | | 1.229956693 | | | | | | | | | | | | 0.0375 |
| rs157843 | A | ANKRD55 | | | 1.514382359 | | | | | | 1.88634886 | | | 1.021353335 | | | 0.0688 |
| rs2862954 | T | ERLIN1 | | | 2.450363977 | | | | | | | | | | | | 0.0291 |
| rs3747207 | A | PNPLA3 | | | 3.845927174 | | | | | | | | | | | | 0.0471 |
| rs2737226 | T | TRPS1 | | | | 0.746153082 | | | | 0.866919138 | | | | | | | 0.0383 |
| rs11759026 | G | CENPW | | | | 0.782090005 | | | | | 1.101231732 | | | | | | 0.0646 |
| rs1800574 | T | HNF1A | | | | 0.913991554 | | | 0.765632283 | | | | | | | | 0.147 |
| rs56158042 | G | C12orf43 | | | | 1.356189443 | | | 0.748315577 | | | | 1.347903571 | | 1.085947684 | | 0.0566 |
| rs1260326 | C | GCKR | | | | 7.41431942 | | | | | | | | | | 0.785198312 | 0.0638 |
| rs38221 | T | MEOX2 | | | | | 0.735247067 | | | | | | | | | | 0.033 |
| rs17818197 | G | EBF2 | | | | | 0.736947612 | | | | | | | | | | 0.0346 |
| rs2066827 | G | CDKN1B | | | | | 0.738867416 | | | | | | | | | | 0.0351 |
| rs72926932 | C | TCF4 | | | | | 0.766823158 | | | | | | | | | | 0.0749 |
| rs7188071 | T | RABEP2 | | | | | 0.78118693 | | | 1.679624536 | | | | | | | 0.0292 |
| rs10821311 | A | LINC02603 | | | | | 0.800887049 | | | | | | | 0.758765389 | | | 0.0358 |
| rs3020781 | G | PKLR | | | | | 0.823621158 | | | | | | | | | | 0.0333 |
| rs2046323 | G | BPTF | | | | | 0.827085259 | | | 0.90855968 | | | | | | | 0.0484 |
| rs17439448 | T | SUGCT | | | | | 0.83480232 | | | | | | | | | | 0.0404 |
| rs10998304 | C | TET1 | | | | | 0.835291215 | | | | | | | 1.112797152 | | | 0.0306 |
| rs5219 | T | KCNJ11 | | | | | 0.891800699 | | | | | | 1.066534884 | | 0.751504045 | | 0.0685 |
| rs62450857 | A | FOXK1 | | | | | 0.911446673 | | | | | | | | | | 0.0388 |
| rs2280141 | T | PLEKHA1 | | | | | 0.940558955 | | | | | | | | 0.771338775 | | 0.0453 |
| rs7558413 | A | NT5C1B-RDH14 | | | | | 0.954927244 | | | | | | | | | | 0.0286 |
| rs3176466 | C | CDKN2C | | | | | 1.036553093 | | | | | | | | | | 0.0642 |
| rs6495182 | C | PTPN9 | | | | | 1.113922042 | | | | | | | | | | 0.0406 |
| rs76895963 | T | CCND2 | | | | | 1.128015822 | | 1.040774465 | | | | | | | 0.889676394 | 0.4388 |
| rs484943 | T | BMF | | | | | 1.160592288 | | | | | | | | | | 0.0332 |
| rs2011603 | A | LCORL | | | | | 1.555861007 | | | | | | | | | | 0.0383 |
| rs62370480 | A | FST | | | | | 1.740522935 | | | | | | | | | | 0.0335 |
| rs8054556 | A | LOC613038 | | | | | | 0.747157985 | | 1.310404847 | | | | | | | 0.0364 |
| rs534043 | G | EPO | | | | | | 0.764667991 | | | | | | | | | 0.0447 |
| rs7619708 | T | TFRC | | | | | | 0.811268375 | | | | | | | | | 0.0334 |
| rs2894536 | C | VEGFA | | | | | | 0.848664475 | | | | | | | | | 0.0384 |
| rs9369425 | G | LINC01512 | | | | | | 1.155447956 | | | 1.433387669 | | 1.436999791 | | | | 0.0384 |
| rs2732480 | C | ZNF641 | | | | | | 1.418781907 | | | | | | | | | 0.0337 |
| rs10159477 | G | HK1 | | | | | | 3.691084837 | | | | | | | | 1.241988693 | 0.0408 |
| rs495203 | T | ABO | | | | | | 5.324443417 | | | | | 0.979071076 | | | | 0.0493 |
| rs543159 | C | SLC22A3 | | | | | | | 2.586728114 | | 1.595149252 | | | | | | 0.0324 |
| rs9927842 | T | PDXDC1 | | | | | | | | 0.748716107 | | | | | | | 0.0375 |
| rs35895680 | C | GIP | | | | | | | | 0.760610664 | | | 0.956498495 | | | | 0.0559 |
| rs1880368 | A | AUTS2 | | | | | | | | 0.763004685 | | | | | | | 0.0311 |
| rs2008027 | G | LINC02523 | | | | | | | | 0.776067846 | | | | | | | 0.0272 |
| rs917195 | C | CRHR2 | | | | | | | | 0.77875085 | | | | | 1.252755395 | | 0.0467 |
| rs3872707 | A | SETD5 | | | | | | | | 0.780789589 | | | | | 1.272073964 | | 0.0446 |
| rs2876826 | G | DDC | | | | | | | | 0.782367212 | | | | 0.754799359 | | | 0.0306 |
| rs1783541 | T | SCYL1 | | | | | | | | 0.783478574 | | | | 0.758466326 | | | 0.0484 |
| rs1569339 | C | LINC00536 | | | | | | | | 0.789808625 | | | | | | | 0.0347 |
| rs11716527 | C | EPHA3 | | | | | | | | 0.804234987 | | | | | | | 0.0492 |
| rs12140153 | G | PATJ | | | | | | | | 0.806830055 | | | | | | | 0.0461 |
| rs2933211 | A | MDGA2 | | | | | | | | 0.814660384 | | | | | | | 0.0268 |
| rs2410763 | T | TMEM161B-DT | | | | | | | | 0.81722549 | | | | 1.119906177 | | | 0.031 |
| rs10498536 | A | NRXN3 | | | | | | | | 0.825727844 | | | | | | | 0.043 |
| rs7656001 | A | CCSER1 | | | | | | | | 0.832106478 | | | | | | | 0.026 |
| rs12992995 | C | LINC01305 | | | | | | | | 0.832156963 | | | | | | | 0.0312 |
| rs36051838 | C | LINC00838 | | | | | | | | 0.834743288 | | | | | | | 0.044 |
| rs567185 | T | NAV1 | | | | | | | | 0.836071559 | | | | | | | 0.037 |
| rs2604918 | G | MAML3 | | | | | | | | 0.859608018 | | | | | | | 0.0313 |
| rs6432613 | G | RBMS1 | | | | | | | | 0.862847697 | | | | | | | 0.0393 |
| rs1056387 | T | AGBL2 | | | | | | | | 0.866516283 | | | | | | | 0.0352 |
| rs11667244 | G | ZC3H4 | | | | | | | | 0.877008931 | | | | | | | 0.0354 |
| rs12519500 | C | BHMT | | | | | | | | 0.877663146 | | | | | | | 0.038 |
| rs7099048 | A | LRMDA | | | | | | | | 0.880274244 | | | 0.999874939 | | | | 0.0282 |
| rs4796224 | G | ZNHIT3 | | | | | | | | 0.891280449 | | | | | 0.762622711 | | 0.0252 |
| rs2733289 | C | PDZRN4 | | | | | | | | 0.894063868 | | | | | | | 0.0301 |
| rs7561798 | G | SPHKAP | | | | | | | | 0.897638893 | | | | | | | 0.0281 |
| rs4899280 | T | DCAF5 | | | | | | | | 0.904985376 | | | | | | | 0.0276 |
| rs197379 | C | INKA2 | | | | | | | | 0.906686938 | | | | | | | 0.0269 |
| rs10769936 | C | TRIM66 | | | | | | | | 0.916255412 | | | | | | | 0.0349 |
| rs12890750 | G | MARK3 | | | | | | | | 0.926623482 | | | | | | | 0.0284 |

**Table S1: SNP weight matrix for pPS construction**

| RSID | Effect_Allele | Gene | Cholesterol_pPS | Lipodystrophy_1_pPS | Lipodystrophy_2_pPS | Liver-Lipid_pPS | ASAT_Pos_pPS | ALP_Neg_pPS | SHBG-LpA_pPS | Obesity_pPS | VAT_Neg_pPS | Bilirubin_pPS | Proinsulin_pPS | Hyper_Insulin_pPS | Beta_Cell_1_pPS | Beta_Cell_2_pPS | Total_GRS |
|---|---|---|---|---|---|---|---|---|---|---|---|---|---|---|---|---|---|
| rs1225052 | G | CPNE4 | | | | | | | | 0.943140675 | | | | | | | 0.0271 |
| rs17522122 | T | AKAP6 | | | | | | | | 0.951612313 | | | | | | | 0.0343 |
| rs4465929 | T | ANKRD28 | | | | | | | | 0.966285047 | | | | 0.888925538 | | | 0.0304 |
| rs686998 | G | NLGN1 | | | | | | | | 1.013813523 | | | | | | | 0.0267 |
| rs73612051 | C | FTO | | | | | | | | 1.019615058 | | | | | | | 0.0565 |
| rs9563574 | T | PCDH17 | | | | | | | | 1.021854321 | | | | | | | 0.0406 |
| rs1752169 | A | DENND1A | | | | | | | | 1.02267197 | | | | | | | 0.0317 |
| rs11842871 | G | HMGB1 | | | | | | | | 1.023201622 | | | | | | | 0.0299 |
| rs2613503 | A | NEGR1 | | | | | | | | 1.041281343 | | | 0.817293538 | | | | 0.0392 |
| rs34298980 | T | LRFN2 | | | | | | | | 1.053218731 | | | | | | | 0.0381 |
| rs1296328 | A | PCDH18 | | | | | | | | 1.057976038 | | | | | | | 0.0251 |
| rs10133111 | A | TRAF3 | | | | | | | | 1.077497202 | | | | 1.275531828 | | | 0.0341 |
| rs303760 | T | RMC1 | | | | | | | | 1.111007435 | | | | | | | 0.0342 |
| rs6741676 | A | UBE2E3 | | | | | | | | 1.11660443 | | | | | | | 0.0324 |
| rs9302652 | C | FTO | | | | | | | | 1.125010072 | | | | | | | 0.044 |
| rs980183 | G | LINC01122 | | | | | | | | 1.145259436 | | | | | 0.750786192 | | 0.0363 |
| rs10490871 | G | ARPP21 | | | | | | | | 1.152788676 | | | 1.226256696 | | 0.8958059 | | 0.0272 |
| rs10404726 | C | CRTC1 | | | | | | | | 1.162844317 | | | | | | | 0.0277 |
| rs2725371 | A | PURG | | | | | | | | 1.179044855 | | | | | | | 0.0371 |
| rs11555762 | T | HSD17B12 | | | | | | | | 1.244297233 | | | | | 0.949710819 | | 0.0407 |
| rs10145154 | T | NRXN3 | | | | | | | | 1.245311157 | | | | | | | 0.0546 |
| rs4688760 | T | RBM6 | | | | | | | | 1.269998687 | | | | 1.057428558 | | | 0.034 |
| rs1412234 | C | LINGO2 | | | | | | | | 1.283604022 | | | | | | | 0.044 |
| rs2032912 | G | MIR1538 | | | | | | | | 1.345943257 | | | | | | | 0.0421 |
| rs3798519 | C | TFAP2B | | | | | | | | 1.383533879 | | | | | | | 0.0495 |
| rs1861410 | C | LINC01122 | | | | | | | | 1.437995701 | | | 1.150133559 | | | | 0.0353 |
| rs79688165 | C | MC4R | | | | | | | | 1.579555244 | | | | | | | 0.1216 |
| rs7132908 | A | FAIM2 | | | | | | | | 1.593947199 | | | | | | | 0.033 |
| rs10938398 | A | GNPDA2 | | | | | | | | 1.691508573 | | | | | | | 0.0425 |
| rs490689 | A | SEC16B | | | | | | | | 1.723285696 | | | | | | | 0.0335 |
| rs10188334 | C | TMEM18 | | | | | | | | 1.781729037 | | | | | | | 0.0503 |
| rs62107261 | T | ALKAL2 | | | | | | | | 1.982980691 | | | | | | | 0.0749 |
| rs663640 | T | MC4R | | | | | | | | 2.121364391 | | | | | | | 0.0503 |
| rs1421085 | C | FTO | | | | | | | | 4.069967742 | | | 0.764129991 | | | | 0.1177 |
| rs75912292 | T | TP53INP1 | | | | | | | | | 0.734691384 | | | | | | 0.0783 |
| rs7163757 | C | C2CD4A | | | | | | | | | 0.76686057 | | | | 2.479604841 | 0.814269258 | 0.0405 |
| rs11063069 | G | CCND2 | | | | | | | | | 0.776917146 | | | | | | 0.0458 |
| rs2334499 | T | MOB2 | | | | | | | | | 0.801373807 | | | | 1.163499525 | | 0.0283 |
| rs11870735 | T | VPS53 | | | | | | | | | 0.803746718 | | | | | | 0.0339 |
| rs1359790 | G | SPRY2 | | | | | | | | | 0.806173341 | | | | | | 0.0795 |
| rs2033159 | C | ZEB2 | | | | | | | | | 0.807589705 | | | | | | 0.0352 |
| rs6819331 | C | TMEM154 | | | | | | | | | 0.821925217 | | | | | | 0.0404 |
| rs9873618 | G | SLC2A2 | | | | | | | | | 0.830976372 | | | | | 2.020595723 | 0.0581 |
| rs4959424 | G | RREB1 | | | | | | | | | 0.883157584 | | | | | | 0.0343 |
| rs2189301 | G | HNF1B | | | | | | | | | 0.890959696 | | | | | | 0.0462 |
| rs4923543 | A | METTL15 | | | | | | | | | 0.924268516 | | | | | | 0.0238 |
| rs329124 | G | JADE2 | | | | | | | | | 0.933352163 | | | 1.001307654 | | | 0.0259 |
| rs9358361 | A | CDKAL1 | | | | | | | | | 0.964379583 | | 0.979907798 | | | | 0.043 |
| rs12694695 | A | LOC646736 | | | | | | | | | 1.058661051 | | | | | | 0.0291 |
| rs719727 | A | RSPO3 | | | | | | | | | 1.138083793 | | | | | | 0.056 |
| rs7834679 | C | EBF2 | | | | | | | | | 1.212081272 | | | | | | 0.0306 |
| rs11603334 | G | ARAP1 | | | | | | | | | 1.77517413 | | 1.201943712 | | | 1.395650821 | 0.0899 |
| rs889138 | C | PEPD | | | | | | | | | 1.780667259 | | | | | | 0.0312 |
| rs838733 | G | DGKD | | | | | | | | | | 1.523820383 | | | 0.967759168 | | 0.0289 |
| rs2602374 | T | UGT1A8 | | | | | | | | | | 12.17989184 | | | | | 0.0291 |
| rs17354348 | A | IKZF2 | | | | | | | | | | | 0.73210331 | | 1.01652074 | | 0.0294 |
| rs6011033 | G | RTEL1 | | | | | | | | | | | 0.733908926 | | | | 0.0338 |
| rs9523299 | G | MIR17HG | | | | | | | | | | | 0.734808663 | | | | 0.0418 |
| rs72695645 | G | ACSL1 | | | | | | | | | | | 0.749231424 | | | 0.821010028 | 0.0611 |
| rs13262861 | C | NKX6-3 | | | | | | | | | | | 0.752403657 | | | | 0.1016 |
| rs827237 | T | PCBD1 | | | | | | | | | | | 0.762494268 | | | | 0.0365 |
| rs10882891 | C | ARHGAP19 | | | | | | | | | | | 0.768376636 | | 0.921215874 | | 0.0313 |
| rs7568172 | G | CYTIP | | | | | | | | | | | 0.77594787 | | | | 0.0674 |
| rs10963942 | G | HAUS6 | | | | | | | | | | | 0.788975887 | | 1.202893658 | | 0.0371 |
| rs12625671 | C | HNF4A | | | | | | | | | | | 0.795998518 | | | | 0.0653 |
| rs4809369 | G | ZBTB46 | | | | | | | | | | | 0.801811875 | | | | 0.0337 |
| rs7943372 | G | DEUP1 | | | | | | | | | | | 0.82025363 | | | | 0.0382 |
| rs2056857 | C | LRRC74A | | | | | | | | | | | 0.821735191 | | 0.793738038 | | 0.0261 |
| rs78503878 | C | GPSM1 | | | | | | | | | | | 0.861585019 | | | | 0.0614 |
| rs684214 | T | NAGLU | | | | | | | | | | | 0.872152702 | | | | 0.0424 |
| rs55974245 | C | MYEOV | | | | | | | | | | | 0.878253974 | | | | 0.037 |
| rs35011184 | A | TCF7L2 | | | | | | | | | | | 0.909537761 | | 1.094982696 | 2.624728482 | 0.2608 |
| rs10174764 | A | ZFP36L2 | | | | | | | | | | | 0.915262864 | | | | 0.0392 |
| rs11196187 | A | TCF7L2 | | | | | | | | | | | 0.977230037 | | 0.998489858 | 0.793197108 | 0.1943 |

**Table S1: SNP weight matrix for pPS construction**

| RSID | Effect_Allele | Gene | Cholesterol_pPS | Lipodystrophy_1_pPS | Lipodystrophy_2_pPS | Liver-Lipid_pPS | ASAT_Pos_pPS | ALP_Neg_pPS | SHBG-LpA_pPS | Obesity_pPS | VAT_Neg_pPS | Bilirubin_pPS | Proinsulin_pPS | Hyper_Insulin_pPS | Beta_Cell_1_pPS | Beta_Cell_2_pPS | Total_GRS |
|---|---|---|---|---|---|---|---|---|---|---|---|---|---|---|---|---|---|
| rs8031576 | C | AP3S2 | | | | | | | | | | | 0.978977845 | | 1.016138912 | | 0.057 |
| rs7071943 | G | CPEB3 | | | | | | | | | | | 0.99083651 | | | | 0.0443 |
| rs7596678 | T | SH3YL1 | | | | | | | | | | | 1.011889366 | | | | 0.0271 |
| rs9316500 | T | DLEU1 | | | | | | | | | | | 1.019552534 | | | | 0.0473 |
| rs7798816 | A | UBE3C | | | | | | | | | | | 1.045507408 | | | | 0.0489 |
| rs11583755 | C | PHF13 | | | | | | | | | | | 1.05467736 | | 1.253570977 | | 0.0387 |
| rs7756992 | G | CDKAL1 | | | | | | | | | | | 1.057831011 | | 2.415885131 | 1.286265689 | 0.1217 |
| rs41276588 | A | TENT5C | | | | | | | | | | | 1.208561891 | | | 0.748458007 | 0.0375 |
| rs2237895 | C | KCNQ1 | | | | | | | | | | | 1.2430368 | | 0.885974644 | 0.856599374 | 0.0725 |
| rs6485644 | C | CRY2 | | | | | | | | | | | 1.246484134 | | 1.260053614 | 0.893196007 | 0.0294 |
| rs11196181 | G | TCF7L2 | | | | | | | | | | | 1.323234788 | | | | 0.1381 |
| rs1111875 | C | HHEX | | | | | | | | | | | 1.342881551 | | 2.198509737 | | 0.0923 |
| rs75432112 | A | MACIR | | | | | | | | | | | 1.346735116 | | | | 0.1344 |
| rs3887925 | T | ST6GAL1 | | | | | | | | | | | 1.348176196 | | 1.521879309 | | 0.0424 |
| rs3217792 | C | CCND2 | | | | | | | | | | | 1.363025493 | | | | 0.1108 |
| rs576674 | G | KL | | | | | | | | | | | 1.394844685 | | 0.769566338 | 1.183685995 | 0.0612 |
| rs11257655 | T | CDC123 | | | | | | | | | | | 1.413302104 | | 0.754770534 | 0.96363826 | 0.0907 |
| rs59191643 | T | NKX6-3 | | | | | | | | | | | 1.913782251 | | 1.589811602 | | 0.0444 |
| rs2513523 | T | EMSY | | | | | | | | | | | | 0.735228219 | | | 0.0255 |
| rs4886092 | C | PCDH17 | | | | | | | | | | | | 0.744234357 | | | 0.0198 |
| rs12977104 | A | UHRF1 | | | | | | | | | | | | 0.745338342 | | | 0.0412 |
| rs79489938 | G | 1:147121000 | | | | | | | | | | | | 0.758406347 | | | 0.1131 |
| rs62493853 | A | LOC157273 | | | | | | | | | | | | 0.765689615 | | | 0.0563 |
| rs4984980 | A | LMF1 | | | | | | | | | | | | 0.771560527 | | | 0.0351 |
| rs4397977 | A | KBTBD6 | | | | | | | | | | | | 0.786186211 | | | 0.0291 |
| rs35824707 | G | MIR4432 | | | | | | | | | | | | 0.792662983 | | | 0.0685 |
| rs11652572 | T | CYB5D2 | | | | | | | | | | | | 0.795253836 | | | 0.1151 |
| rs12489177 | T | SYN2 | | | | | | | | | | | | 0.812081074 | | | 0.0745 |
| rs73872717 | C | ZBTB38 | | | | | | | | | | | | 0.822404305 | | 0.89288354 | 0.0856 |
| rs73195303 | C | MSRA | | | | | | | | | | | | 0.839306815 | | | 0.0314 |
| rs113780182 | T | LDLRAD4 | | | | | | | | | | | | 0.854306244 | | | 0.0309 |
| rs243021 | A | MIR4432 | | | | | | | | | | | | 0.865499332 | | | 0.054 |
| rs7943101 | T | WT1-AS | | | | | | | | | | | | 0.882817815 | | | 0.0358 |
| rs2675662 | A | CAMK2G | | | | | | | | | | | | 0.88512431 | | | 0.0267 |
| rs62075585 | G | CYTH1 | | | | | | | | | | | | 0.928655402 | | | 0.0295 |
| rs66485956 | T | MAP3K1 | | | | | | | | | | | | 0.946921309 | | | 0.0385 |
| rs62492368 | A | AOC1 | | | | | | | | | | | | 0.956430788 | | | 0.0342 |
| rs8005994 | A | MIR548AI | | | | | | | | | | | | 0.959587336 | | | 0.0265 |
| rs10305420 | C | GLP1R | | | | | | | | | | | | 0.963576282 | | | 0.0316 |
| rs9296291 | T | GLP1R | | | | | | | | | | | | 0.964564282 | | | 0.0325 |
| rs1431841 | T | LINC00907 | | | | | | | | | | | | 0.965322255 | | | 0.0309 |
| rs31229 | G | ARL15 | | | | | | | | | | | | 0.965661923 | | | 0.0391 |
| rs7599488 | T | BCL11A | | | | | | | | | | | | 0.988819438 | | | 0.0255 |
| rs34965774 | A | KSR2 | | | | | | | | | | | | 1.006214682 | | | 0.0521 |
| rs76365198 | C | TRA2A | | | | | | | | | | | | 1.021257476 | | | 0.0496 |
| rs7828303 | T | BOP1 | | | | | | | | | | | | 1.030316492 | | 0.830068774 | 0.0427 |
| rs11108094 | A | USP44 | | | | | | | | | | | | 1.039814948 | | | 0.0601 |
| rs75332279 | C | ONECUT1 | | | | | | | | | | | | 1.053350914 | | | 0.0559 |
| rs798549 | C | AMZ1 | | | | | | | | | | | | 1.05903011 | | | 0.0295 |
| rs2767323 | G | MAN1A2 | | | | | | | | | | | | 1.06850776 | | | 0.0339 |
| rs1018942 | T | COBL | | | | | | | | | | | | 1.185600125 | | | 0.0711 |
| rs844215 | C | FOXP1 | | | | | | | | | | | | 1.187060707 | | | 0.0262 |
| rs28566988 | C | XKR6 | | | | | | | | | | | | 1.194986196 | | | 0.0392 |
| rs2289065 | G | OCIAD2 | | | | | | | | | | | | 1.214248595 | | | 0.0272 |
| rs12602834 | G | NF1 | | | | | | | | | | | | 1.254451708 | | | 0.0289 |
| rs34845373 | A | DTNB | | | | | | | | | | | | 1.392007917 | | | 0.0372 |
| rs11129735 | A | TRANK1 | | | | | | | | | | | | 1.450118735 | | | 0.026 |
| rs1656794 | G | SEPTIN9 | | | | | | | | | | | | 1.505103387 | | | 0.0311 |
| rs1572993 | A | CNTN2 | | | | | | | | | | | | 1.578130778 | | | 0.031 |
| rs10916785 | T | LINC01141 | | | | | | | | | | | | | 0.741404206 | | 0.0271 |
| rs13095782 | T | ST6GAL1 | | | | | | | | | | | | | 0.752962035 | | 0.0285 |
| rs17294565 | C | SGCZ | | | | | | | | | | | | | 0.772226911 | | 0.0274 |
| rs11020132 | A | MTNR1B | | | | | | | | | | | | | 0.775554598 | | 0.1424 |
| rs9379084 | G | RREB1 | | | | | | | | | | | | | 0.7775043 | | 0.0747 |
| rs4714422 | G | UNC5CL | | | | | | | | | | | | | 0.785686444 | | 0.0293 |
| rs11967298 | A | CDKAL1 | | | | | | | | | | | | | 0.786669457 | | 0.0378 |
| rs10794522 | C | FAM110D | | | | | | | | | | | | | 0.805212778 | | 0.0261 |
| rs11657964 | A | HNF1B | | | | | | | | | | | | | 0.806954667 | | 0.0589 |
| rs67013744 | G | CHD4 | | | | | | | | | | | | | 0.809827436 | | 0.035 |
| rs9383649 | G | RGS17 | | | | | | | | | | | | | 0.812825108 | | 0.0329 |
| rs17122776 | G | SLC7A7 | | | | | | | | | | | | | 0.820396792 | | 0.029 |
| rs9784137 | G | LINC01101 | | | | | | | | | | | | | 0.820403467 | | 0.0608 |
| rs80105613 | T | ANK1 | | | | | | | | | | | | | 0.836685732 | | 0.0641 |
| rs56269620 | A | FBXL13 | | | | | | | | | | | | | 0.841545229 | | 0.0316 |

**Table S1: SNP weight matrix for pPS construction**

| RSID | Effect_Allele | Gene | Cholesterol_pPS | Lipodystrophy_1_pPS | Lipodystrophy_2_pPS | Liver-Lipid_pPS | ASAT_Pos_pPS | ALP_Neg_pPS | SHBG-LpA_pPS | Obesity_pPS | VAT_Neg_pPS | Bilirubin_pPS | Proinsulin_pPS | Hyper_Insulin_pPS | Beta_Cell_1_pPS | Beta_Cell_2_pPS | Total_GRS |
|---|---|---|---|---|---|---|---|---|---|---|---|---|---|---|---|---|---|
| rs2812533 | T | FAM241B | | | | | | | | | | | | | 0.848763744 | | 0.0383 |
| rs9828639 | T | P2RY1 | | | | | | | | | | | | | 0.903080833 | | 0.0204 |
| rs1431819 | G | COL27A1 | | | | | | | | | | | | | 0.917711405 | | 0.0293 |
| rs11187152 | G | HHEX | | | | | | | | | | | | | 0.919680097 | | 0.0537 |
| rs1430780 | T | ETAA1 | | | | | | | | | | | | | 0.953168181 | | 0.0273 |
| rs12969494 | G | WDR7 | | | | | | | | | | | | | 0.96631744 | | 0.0292 |
| rs17265513 | C | ZHX3 | | | | | | | | | | | | | 0.986958227 | | 0.0325 |
| rs4838049 | C | CRB2 | | | | | | | | | | | | | 0.988468433 | | 0.027 |
| rs9895554 | A | NFE2L1 | | | | | | | | | | | | | 0.990992464 | | 0.0549 |
| rs11940813 | G | SLIT2 | | | | | | | | | | | | | 0.992971725 | | 0.0371 |
| rs67755137 | A | GTF2I | | | | | | | | | | | | | 1.005616254 | | 0.0333 |
| rs12192642 | G | CDKAL1 | | | | | | | | | | | | | 1.017925908 | | 0.0545 |
| rs55857387 | T | FAM234A | | | | | | | | | | | | | 1.033043419 | | 0.0524 |
| rs2290203 | A | PRC1 | | | | | | | | | | | | | 1.05530558 | | 0.0557 |
| rs11600952 | A | IGF2 | | | | | | | | | | | | | 1.070559907 | | 0.0356 |
| rs450563 | G | SLC22A18AS | | | | | | | | | | | | | 1.107866685 | | 0.0341 |
| rs2427363 | C | SLCO4A1 | | | | | | | | | | | | | 1.108923771 | | 0.027 |
| rs1531583 | T | PCGF3 | | | | | | | | | | | | | 1.120568807 | | 0.0994 |
| rs4929965 | A | MIR4686 | | | | | | | | | | | | | 1.130831535 | | 0.0619 |
| rs10841868 | G | LDHB | | | | | | | | | | | | | 1.140154825 | | 0.0315 |
| rs10407429 | G | EML2 | | | | | | | | | | | | | 1.148643219 | | 0.0539 |
| rs35352848 | T | UBE2E2 | | | | | | | | | | | | | 1.216430753 | | 0.0623 |
| rs72807217 | T | CPEB3 | | | | | | | | | | | | | 1.218630582 | | 0.0478 |
| rs7977709 | C | KDM2B | | | | | | | | | | | | | 1.259736259 | | 0.0261 |
| rs10811661 | T | CDKN2B-AS1 | | | | | | | | | | | | | 1.359358766 | 1.385442078 | 0.1379 |
| rs31931 | T | ANKH | | | | | | | | | | | | | 1.45529761 | | 0.038 |
| rs1063192 | A | MTAP | | | | | | | | | | | | | 1.467135031 | | 0.0434 |
| rs17035289 | C | TET2 | | | | | | | | | | | | | 1.507315546 | | 0.0428 |
| rs7219033 | A | GLP2R | | | | | | | | | | | | | 1.539435291 | | 0.0288 |
| rs3764002 | C | WSCD2 | | | | | | | | | | | | | 1.552706495 | | 0.0396 |
| rs11558471 | A | SLC30A8 | | | | | | | | | | | | | 1.570214503 | 2.613645214 | 0.1031 |
| rs231360 | T | KCNQ1 | | | | | | | | | | | | | | 0.741101218 | 0.0506 |
| rs10010131 | G | WFS1 | | | | | | | | | | | | | | 0.743525812 | 0.0816 |
| rs10885414 | A | TCF7L2 | | | | | | | | | | | | | | 0.756106327 | 0.0787 |
| rs10885410 | G | TCF7L2 | | | | | | | | | | | | | | 0.75762406 | 0.0895 |
| rs348330 | G | ABCB10 | | | | | | | | | | | | | | 0.761342767 | 0.0525 |
| rs116913033 | C | CAMK2B | | | | | | | | | | | | | | 0.790033277 | 0.0408 |
| rs9873519 | T | SLC12A8 | | | | | | | | | | | | | | 0.790351147 | 0.0375 |
| rs911300 | G | MIR296 | | | | | | | | | | | | | | 0.791727162 | 0.0348 |
| rs11786992 | A | ESRP1 | | | | | | | | | | | | | | 0.813994537 | 0.0319 |
| rs11614914 | T | FBRSL1 | | | | | | | | | | | | | | 0.89531812 | 0.0389 |
| rs11243150 | T | SSR1 | | | | | | | | | | | | | | 0.928903505 | 0.0447 |
| rs12910361 | G | HMG20A | | | | | | | | | | | | | | 0.964617012 | 0.0716 |
| rs6777684 | G | LPP-AS2 | | | | | | | | | | | | | | 1.019198131 | 0.0569 |
| rs113036477 | C | RMST | | | | | | | | | | | | | | 1.055798053 | 0.0715 |
| rs7018475 | G | CDKN2B-AS1 | | | | | | | | | | | | | | 1.066692714 | 0.1094 |
| rs6878122 | G | ZBED3-AS1 | | | | | | | | | | | | | | 1.143891482 | 0.0554 |
| rs10885123 | C | ADRA2A | | | | | | | | | | | | | | 1.284552573 | 0.0316 |
| rs13414140 | C | THADA | | | | | | | | | | | | | | 1.32635212 | 0.1183 |
| rs10974438 | C | GLIS3 | | | | | | | | | | | | | | 1.374976901 | 0.047 |
| rs340874 | C | PROX1 | | | | | | | | | | | | | | 1.507686703 | 0.0668 |
| rs17168486 | T | DGKB | | | | | | | | | | | | | | 1.577377 | 0.069 |
| rs2041547 | C | GCK | | | | | | | | | | | | | | 1.799511274 | 0.0385 |
| rs11708067 | A | ADCY5 | | | | | | | | | | | | | | 2.191656735 | 0.0782 |
| rs2191349 | T | DGKB | | | | | | | | | | | | | | 2.314437915 | 0.0661 |
| rs2908286 | T | GCK | | | | | | | | | | | | | | 3.961336475 | 0.0683 |

**Table S2: Summary of clinical, molecular, and imaging-derived variables in SCAPIS, and disease outcomes in UKBB**

| Category | Variable_Name | Data_Available | Summary | Unit | Heading | Context | Method | Source | Cohort |
|---|---|---|---|---|---|---|---|---|---|
| Derived - PPS | Cholesterol_pPS | 29425 | 0.04 (0.03, 0.04) | | Cholesterol process-specific polygenic score | Process-specific polygenic score | The PPS were calculated based on genotype data by multiplying a variant's genotype dosage by its | Derived | SCAPIS |
| Derived - PPS | Lipodystrophy_1_pPS | 29425 | 0.11 (0.1, 0.11) | | Lipodystrophy 1 process-specific polygenic score | Process-specific polygenic score | The PPS were calculated based on genotype data by multiplying a variant's genotype dosage by its | Derived | SCAPIS |
| Derived - PPS | Lipodystrophy_2_pPS | 29425 | 0.04 (0.03, 0.08) | | Lipodystrophy 2 process-specific polygenic score | Process-specific polygenic score | The PPS were calculated based on genotype data by multiplying a variant's genotype dosage by its | Derived | SCAPIS |
| Derived - PPS | Liver-Lipid_pPS | 29425 | 0.01 (0.01, 0.02) | | Liver-Lipid process-specific polygenic score | Process-specific polygenic score | The PPS were calculated based on genotype data by multiplying a variant's genotype dosage by its | Derived | SCAPIS |
| Derived - PPS | ASAT_Pos_pPS | 29425 | 0.04 (0.04, 0.04) | | Abdominal subcutaneous adipose tissue positive process-specific polygenic score | Process-specific polygenic score | The PPS were calculated based on genotype data by multiplying a variant's genotype dosage by its | Derived | SCAPIS |
| Derived - PPS | ALP_Neg_pPS | 29425 | 0.03 (0.02, 0.03) | | Alanine transaminase negative process-specific polygenic score | Process-specific polygenic score | The PPS were calculated based on genotype data by multiplying a variant's genotype dosage by its | Derived | SCAPIS |
| Derived - PPS | SHBG-LpA_pPS | 29425 | 0.02 (0.01, 0.02) | | Sex hormone binding globulin-lipoprotein A process-specific polygenic score | Process-specific polygenic score | The PPS were calculated based on genotype data by multiplying a variant's genotype dosage by its | Derived | SCAPIS |
| Derived - PPS | Obesity_pPS | 29425 | 0.12 (0.11, 0.13) | | Obesity process-specific polygenic score | Process-specific polygenic score | The PPS were calculated based on genotype data by multiplying a variant's genotype dosage by its | Derived | SCAPIS |
| Derived - PPS | VAT_Neg_pPS | 29425 | 0.04 (0.04, 0.04) | | Visceral adipose tissue negative process-specific polygenic score | Process-specific polygenic score | The PPS were calculated based on genotype data by multiplying a variant's genotype dosage by its | Derived | SCAPIS |
| Derived - PPS | Bilirubin_pPS | 29425 | 0.0 (0.0, 0.01) | | Bilirubin process-specific polygenic score | Process-specific polygenic score | The PPS were calculated based on genotype data by multiplying a variant's genotype dosage by its | Derived | SCAPIS |
| Derived - PPS | Proinsulin_pPS | 29425 | 0.07 (0.06, 0.07) | | Proinsulin process-specific polygenic score | Process-specific polygenic score | The PPS were calculated based on genotype data by multiplying a variant's genotype dosage by its | Derived | SCAPIS |
| Derived - PPS | Hyper_Insulin_pPS | 29425 | 0.08 (0.07, 0.08) | | Hyper insulin process-specific polygenic score | Process-specific polygenic score | The PPS were calculated based on genotype data by multiplying a variant's genotype dosage by its | Derived | SCAPIS |
| Derived - PPS | Beta_Cell_1_pPS | 29425 | 0.1 (0.09, 0.11) | | Beta Cell 1 process-specific polygenic score | Process-specific polygenic score | The PPS were calculated based on genotype data by multiplying a variant's genotype dosage by its | Derived | SCAPIS |
| Derived - PPS | Beta_Cell_2_pPS | 29425 | 0.07 (0.06, 0.07) | | Beta Cell 2 process-specific polygenic score | Process-specific polygenic score | The PPS were calculated based on genotype data by multiplying a variant's genotype dosage by its | Derived | SCAPIS |
| Derived - GRS | Total_GRS | 29425 | 0.02 (0.02, 0.02) | | Total genetic risk score for type 2 diabetes | Total genetic risk score | The GRS were calculated based on genotype data by multiplying a variant's genotype dosage by its | Derived | SCAPIS |
| Anthropometry | BMI | 29424 | 26.3 (23.9, 29.4) | kg/m2 | Body mass index | Height, weight, waist and hip circumference were me | BMI (kg/m2) = Weight (kg) divided by Height (m) squared. | Physical examination | SCAPIS |
| Anthropometry | DBP_Mean | 29279 | 77.0 (70.0, 84.0) | mmHg | Diastolic blood pressure, mean brachial (recommended for general analysis) | | Brachial arterial blood pressure was obtained by automatic measurement in both arms. Systolic ar | Physical examination | SCAPIS |
| Anthropometry | Height | 29424 | 172.0 (165.0, 179.0) | centimeter (cm) | Body height | Height, Weight, and Waist and Hip circumferences we | The instruction according to protocol: The subject should be measured in indoor clothing to the ne | Physical examination | SCAPIS |
| Anthropometry | Hip | 29376 | 102.0 (97.0, 107.0) | centimeter (cm) | Hip circumference | Height, Weight, and Waist and Hip circumferences we | The instruction according to protocol: The subject should stand erect with arms at the sides, feet to | Physical examination | SCAPIS |
| Anthropometry | Pulse | 29273 | 62.0 (55.0, 68.0) | bpm | Pulse rate | | Heart rate was recorded during the ECG measurement. The subject was instructed to remove all cl | Physical examination | SCAPIS |
| Anthropometry | SBP_Mean | 29281 | 124.0 (114.0, 136.0) | mmHg | Systolic blood pressure, mean brachial (recommended for general analysis) | | Brachial arterial blood pressure was obtained by automatic measurement in both arms. Systolic ar | Physical examination | SCAPIS |
| Anthropometry | Waist | 29411 | 94.0 (85.0, 103.0) | centimeter (cm) | Waist circumference | Height, Weight, and Waist and Hip circumferences we | The instruction according to protocol: All clothing except underwear should be removed to ensure | Physical examination | SCAPIS |
| Anthropometry | Waist/hip_Ratio | 29372 | 0.92 (0.85, 0.98) | centimeter (cm) | Waist/hip Ratio | Height, Weight, and Waist and Hip circumferences we | The instruction according to protocol: All clothing except underwear should be removed to ensure | Physical examination | SCAPIS |
| Anthropometry | Weight | 29425 | 79.0 (68.8, 90.0) | kg | Body weight | Height, Weight, and Waist and Hip circumferences we | The instruction according to protocol: Weight (kg) should be measured on a balance beam or digit | Physical examination | SCAPIS |
| Biochemistry | ALAT | 29118 | 0.38 (0.29, 0.53) | µkat/L | ALAT, Alaninaminotransferas Numerical result | | Analysed in EDTA-plasma from frozen samples. Instrument: Alinity C, Abbott. Reagent: Alinity c A | Blood Analysis | SCAPIS |
| Biochemistry | ASAT | 29087 | 0.41 (0.35, 0.48) | µkat/L | ASAT, AspartateAminotransferase Numerical result | | Analysed in EDTA-plasma from frozen samples. Instrument: Alinity C, Abbott. Reagent: Alinity c A | Blood Analysis | SCAPIS |
| Biochemistry | Alfa1Antitrypsin | 29119 | 1.15 (1.06, 1.25) | g/L | Alfa1 Antitrypsin, #±-1 Antitrypsin Numerical result | | Analysed in EDTA-plasma from frozen samples. Instrument: Alinity C, Abbott. Reagent: Alinity c A | Blood Analysis | SCAPIS |
| Biochemistry | ApoA1 | 29116 | 1.54 (1.37, 1.75) | g/L | ApoA1, Apolipoprotein A1 Numerical result | | Analysed in EDTA-plasma from frozen samples. Instrument: Alinity C, Abbott. Reagent: Alinity c A | Blood Analysis | SCAPIS |
| Biochemistry | ApoB | 29119 | 0.96 (0.82, 1.12) | g/L | ApoB, Apolipoprotein B Numerical result | | Analysed in EDTA-plasma from frozen samples. Instrument: Alinity C, Abbott. Reagent: Alinity c A | Blood Analysis | SCAPIS |
| Biochemistry | Cholesterol | 29393 | 5.4 (4.8, 6.1) | mmol/L | Cholesterol Numerical result | | Baseline biochemistry from Day 1 performed at each site using fresh blood samples after an over-r | Blood Analysis | SCAPIS |
| Biochemistry | Creatinine | 29401 | 77.0 (67.0, 87.0) | µmol/L | Creatinine Numerical result | | Baseline biochemistry from Day 1 performed at each site using fresh blood samples after an over-r | Blood Analysis | SCAPIS |
| Biochemistry | Crp | 29393 | 1.0 (0.6, 2.2) | mg/L | hsCRP Numerical result - national variable | | Baseline biochemistry from Day 1 performed at each site using fresh blood samples after an over-r | Blood Analysis | SCAPIS |
| Biochemistry | CystatinC | 29119 | 0.8 (0.7, 0.9) | mg/L | Cyst-C, Cystatin-C Numerical result | | Analysed in EDTA-plasma from frozen samples. Instrument: Alinity C, Abbott. Reagent: Gentian C | Blood Analysis | SCAPIS |
| Biochemistry | GGT2 | 29119 | 0.38 (0.27, 0.59) | µkat/L | GGT, Gamma-glutamyltransferase, Î³-GT Numerical result | | Analysed in EDTA-plasma from frozen samples. Instrument: Alinity C, Abbott. Reagent:Alinity c Ga | Blood Analysis | SCAPIS |
| Biochemistry | Glucose | 26980 | 5.6 (5.2, 6.0) | mmol/L | Glucose, venous, day 1, numerical result | | Baseline biochemistry from Day 1 performed at each site using fresh blood samples after an over-r | Blood Analysis | SCAPIS |
| Biochemistry | HB | 29364 | 141.0 (133.0, 150.0) | g/L | Hb Numerical Result | | Baseline biochemistry from Day 1 performed at each site using fresh blood samples after an over-r | Blood Analysis | SCAPIS |
| Biochemistry | Hba1c | 29375 | 36.0 (33.0, 38.0) | mmol/mol | HbA1c Numerical result | | Baseline biochemistry from Day 1 performed at each site using fresh blood samples after an over-r | Blood Analysis | SCAPIS |
| Biochemistry | Hdl | 29386 | 1.6 (1.3, 1.9) | mmol/L | HDL Numerical result - national variable. | | Baseline biochemistry from Day 1 performed at each site using fresh blood samples after an over-r | Blood Analysis | SCAPIS |
| Biochemistry | Insulin | 29095 | 5.31 (3.75, 7.92) | mIE/L | Insulin Numerical result | | Analysed in EDTA-plasma from frozen samples. Instrument: Alinity I, Abbott. Reagent: Alinity I Ins | Blood Analysis | SCAPIS |
| Biochemistry | Ldl | 29252 | 3.4 (2.8, 4.0) | mmol/L | LDL Numerical result - national variable. | | Baseline biochemistry from Day 1 performed at each site using fresh blood samples after an over-r | Blood Analysis | SCAPIS |
| Biochemistry | Lpa | 29111 | 0.1 (0.05, 0.27) | g/L | Lp(a), Lipoprotein(a) Numerical result | | Analysed in EDTA-plasma from frozen samples. Instrument: Alinity C, Abbott. Reagent: Alinity c Lp | Blood Analysis | SCAPIS |
| Biochemistry | NTproBNP | 29062 | 48.16 (29.02, 80.82) | ng/L | NT-proBNP, N-terminal pro b-type nauretic peptide Numerical result | | Analysed in EDTA-plasma from frozen samples. Instrument: Alinity I, Abbott. Reagent: Alere NT-p | Blood Analysis | SCAPIS |
| Biochemistry | PGluc_Cap | 29396 | 5.5 (5.0, 5.9) | mmol/L | Glucose, capillary, day 1 | | Capillary plasma glucose from Day 1 performed at each site using fresh blood samples after an ove | Blood analysis | SCAPIS |
| Biochemistry | Tg | 29394 | 1.0 (0.8, 1.5) | mmol/L | Triglycerides Numerical result - national variable | | Baseline biochemistry from Day 1 performed at each site using fresh blood samples after an over-r | Blood Analysis | SCAPIS |
| Biochemistry | TroponinI | 29106 | 1.7 (1.3, 2.7) | ng/L | Trop I, Troponin I Numerical result | | Analysed in EDTA-plasma from frozen samples. Instrument: Alinity I, Abbott. Reagent: Alinity I ST | Blood Analysis | SCAPIS |
| Proteomics | ACE2 | 4933 | 2.73 (2.46, 3.08) | NPX | Angiotensin-converting enzyme 2 - Olink CVD2 | Uni-Prot ID: Q9BYF1, OLINK ID: OID00457, Limit of de | Analyses made in EDTA plasma. Data is presented as normalized protein expression (NPX) values, ( | Blood Analysis | SCAPIS |
| Proteomics | ADAM_TS13 | 4933 | 4.81 (4.72, 4.89) | NPX | A disintegrin and metalloproteinase with thrombospondin motifs 13 - Olink CVD2 | UNI-Prot ID: Q76LX8, OLINK ID: OID00385, Limit of de | Analyses made in EDTA plasma. Data is presented as normalized protein expression (NPX) values, ( | Blood Analysis | SCAPIS |
| Proteomics | ADM | 4933 | 6.9 (6.66, 7.14) | NPX | ADM - Olink CVD2 | UNI-Prot ID: P35318, OLINK ID: OID00381, Limit of de | Analyses made in EDTA plasma. Data is presented as normalized protein expression (NPX) values, ( | Blood Analysis | SCAPIS |
| Proteomics | AGRP | 4933 | 2.49 (2.28, 2.7) | NPX | Agouti-related protein - Olink CVD2 | Uni-Prot ID: O00253, OLINK ID: OID00448, Limit of de | Analyses made in EDTA plasma. Data is presented as normalized protein expression (NPX) values, ( | Blood Analysis | SCAPIS |
| Proteomics | ALCAM | 5006 | 5.25 (5.09, 5.4) | NPX | CD166 antigen - Olink CVD3 | Uni-Prot ID: Q13740, OLINK ID: OID00572, Limit of de | Analyses made in EDTA plasma. Data is presented as normalized protein expression (NPX) values, ( | Blood Analysis | SCAPIS |
| Proteomics | AMBP | 4933 | 7.14 (7.04, 7.25) | NPX | Protein AMBP - Olink CVD2 | Uni-Prot ID: P02760, OLINK ID: OID00430, Limit of de | Analyses made in EDTA plasma. Data is presented as normalized protein expression (NPX) values, ( | Blood Analysis | SCAPIS |
| Proteomics | ANG_1 | 4933 | 7.74 (7.23, 8.31) | NPX | Angiopoietin-1 - Olink CVD2 | UNI-Prot ID: Q15389, OLINK ID: OID00380, Limit of de | Analyses made in EDTA plasma. Data is presented as normalized protein expression (NPX) values, ( | Blood Analysis | SCAPIS |
| Proteomics | AP_N | 5006 | 5.09 (4.91, 5.26) | NPX | Aminopeptidase N - Olink CVD3 | Uni-Prot ID: P15144, OLINK ID: OID00611, Limit of de | Analyses made in EDTA plasma. Data is presented as normalized protein expression (NPX) values, ( | Blood Analysis | SCAPIS |
| Proteomics | AXL | 5006 | 8.74 (8.54, 8.94) | NPX | Tyrosine-protein kinase receptor UFO - Olink CVD3 | Uni-Prot ID: P30530, OLINK ID: OID00612, Limit of de | Analyses made in EDTA plasma. Data is presented as normalized protein expression (NPX) values, ( | Blood Analysis | SCAPIS |
| Proteomics | AZU1 | 4671 | 3.55 (3.32, 3.81) | NPX | Azurocidin - Olink CVD3 | Uni-Prot ID: P20160, OLINK ID: OID00597, Limit of de | Analyses made in EDTA plasma. Data is presented as normalized protein expression (NPX) values, ( | Blood Analysis | SCAPIS |
| Proteomics | BLM_hydrolase | 4830 | 5.39 (5.19, 5.6) | NPX | Bleomycin hydrolase - Olink CVD3 | Uni-Prot ID: Q13867, OLINK ID: OID00581, Limit of de | Analyses made in EDTA plasma. Data is presented as normalized protein expression (NPX) values, ( | Blood Analysis | SCAPIS |
| Proteomics | BMP_6 | 4933 | 4.35 (4.12, 4.59) | NPX | Bone morphogenetic protein 6 - Olink CVD2 | UNI-Prot ID: P22004, OLINK ID: OID00379, Limit of de | Analyses made in EDTA plasma. Data is presented as normalized protein expression (NPX) values, ( | Blood Analysis | SCAPIS |
| Proteomics | BNP | 4933 | 0.61 (0.41, 0.94) | NPX | Natriuretic peptides B - Olink CVD2 | Uni-Prot ID: P16860, OLINK ID: OID00455, Limit of de | Analyses made in EDTA plasma. Data is presented as normalized protein expression (NPX) values, ( | Blood Analysis | SCAPIS |
| Proteomics | BOC | 4933 | 3.84 (3.68, 4.0) | NPX | Brother of CDO - Olink CVD2 | UNI-Prot ID: Q9BWV1, OLINK ID: OID00386, Limit of d | Analyses made in EDTA plasma. Data is presented as normalized protein expression (NPX) values, ( | Blood Analysis | SCAPIS |
| Proteomics | CA5A | 4933 | 1.97 (1.51, 2.5) | NPX | Carbonic anhydrase 5A, mitochondrial - Olink CVD2 | Uni-Prot ID: P35218, OLINK ID: OID00464, Limit of de | Analyses made in EDTA plasma. Data is presented as normalized protein expression (NPX) values, ( | Blood Analysis | SCAPIS |
| Proteomics | CASP_3 | 5006 | 7.43 (6.91, 7.93) | NPX | Caspase-3 - Olink CVD3 | Uni-Prot ID: P42574, OLINK ID: OID00630, Limit of de | Analyses made in EDTA plasma. Data is presented as normalized protein expression (NPX) values, ( | Blood Analysis | SCAPIS |
| Proteomics | CCL15 | 5006 | 7.3 (7.07, 7.62) | NPX | C-C motif chemokine 15 - Olink CVD3 | Uni-Prot ID: Q16663, OLINK ID: OID00629, Limit of de | Analyses made in EDTA plasma. Data is presented as normalized protein expression (NPX) values, ( | Blood Analysis | SCAPIS |
| Proteomics | CCL16 | 5006 | 6.56 (6.21, 6.87) | NPX | C-C motif chemokine 16 - Olink CVD3 | Uni-Prot ID: O15467, OLINK ID: OID00654, Limit of de | Analyses made in EDTA plasma. Data is presented as normalized protein expression (NPX) values, ( | Blood Analysis | SCAPIS |
| Proteomics | CCL17 | 4932 | 7.68 (7.14, 8.28) | NPX | C-C motif chemokine 17 - Olink CVD2 | Uni-Prot ID: Q92583, OLINK ID: OID00439, Limit of de | Analyses made in EDTA plasma. Data is presented as normalized protein expression (NPX) values, ( | Blood Analysis | SCAPIS |
| Proteomics | CCL24 | 5006 | 5.7 (5.07, 6.39) | NPX | C-C motif chemokine 24 - Olink CVD3 | Uni-Prot ID: O00175, OLINK ID: OID00592, Limit of de | Analyses made in EDTA plasma. Data is presented as normalized protein expression (NPX) values, ( | Blood Analysis | SCAPIS |
| Proteomics | CCL3 | 4933 | 5.2 (4.86, 5.59) | NPX | C-C motif chemokine 3 - Olink CVD2 | Uni-Prot ID: P10147, OLINK ID: OID00440, Limit of de | Analyses made in EDTA plasma. Data is presented as normalized protein expression (NPX) values, ( | Blood Analysis | SCAPIS |
| Proteomics | CD163 | 5006 | 7.52 (7.21, 7.83) | NPX | Scavenger receptor cysteine-rich type 1 protein M130 - Olink CVD3 | Uni-Prot ID: Q86VB7, OLINK ID: OID00577, Limit of de | Analyses made in EDTA plasma. Data is presented as normalized protein expression (NPX) values, ( | Blood Analysis | SCAPIS |
| Proteomics | CD4 | 4933 | 3.81 (3.65, 3.98) | NPX | T-cell surface glycoprotein CD4 - Olink CVD2 | Uni-Prot ID: P01730, OLINK ID: OID00466, Limit of de | Analyses made in EDTA plasma. Data is presented as normalized protein expression (NPX) values, ( | Blood Analysis | SCAPIS |
| Proteomics | CD40_L | 4933 | 5.16 (4.58, 5.74) | NPX | CD40 ligand - Olink CVD2 | Uni-Prot ID: P29965, OLINK ID: OID00382, Limit of de | Analyses made in EDTA plasma. Data is presented as normalized protein expression (NPX) values, ( | Blood Analysis | SCAPIS |
| Proteomics | CD84 | 4933 | 4.75 (4.51, 5.01) | NPX | SLAM family member 5 - Olink CVD2 | Uni-Prot ID: Q9UIB8, OLINK ID: OID00420, Limit of de | Analyses made in EDTA plasma. Data is presented as normalized protein expression (NPX) values, ( | Blood Analysis | SCAPIS |
| Proteomics | CD93 | 5006 | 10.98 (10.78, 11.18) | NPX | Complement component C1q receptor - Olink CVD3 | Uni-Prot ID: Q9NPY3, OLINK ID: OID00639, Limit of de | Analyses made in EDTA plasma. Data is presented as normalized protein expression (NPX) values, ( | Blood Analysis | SCAPIS |
| Proteomics | CDH5 | 5006 | 3.72 (3.52, 3.93) | NPX | Cadherin-5 - Olink CVD3 | Uni-Prot ID: P33151, OLINK ID: OID00587, Limit of de | Analyses made in EDTA plasma. Data is presented as normalized protein expression (NPX) values, ( | Blood Analysis | SCAPIS |
| Proteomics | CEACAM8 | 4933 | 2.34 (2.07, 2.64) | NPX | Carcinoembryonic antigenrelated cell adhesion molecule 8 - Olink CVD2 | Uni-Prot ID: P31997, OLINK ID: OID00436, Limit of de | Analyses made in EDTA plasma. Data is presented as normalized protein expression (NPX) values, ( | Blood Analysis | SCAPIS |
| Proteomics | CHI3L1 | 5006 | 6.28 (5.89, 6.8) | NPX | Chitinase-3-like protein 1 - Olink CVD3 | Uni-Prot ID: P36222, OLINK ID: OID00633, Limit of de | Analyses made in EDTA plasma. Data is presented as normalized protein expression (NPX) values, ( | Blood Analysis | SCAPIS |
| Proteomics | CHIT1 | 5006 | 6.28 (5.68, 6.86) | NPX | Chitotriosidase-1 - Olink CVD3 | Uni-Prot ID: Q13231, OLINK ID: OID00605, Limit of de | Analyses made in EDTA plasma. Data is presented as normalized protein expression (NPX) values, ( | Blood Analysis | SCAPIS |
| Proteomics | CNTN1 | 5006 | 4.2 (3.99, 4.39) | NPX | Contactin-1 - Olink CVD3 | Uni-Prot ID: Q12860, OLINK ID: OID00586, Limit of de | Analyses made in EDTA plasma. Data is presented as normalized protein expression (NPX) values, ( | Blood Analysis | SCAPIS |
| Proteomics | COL1A1 | 5006 | 3.28 (3.05, 3.54) | NPX | Collagen alpha-1(I) chain - Olink CVD3 | Uni-Prot ID: P02452, OLINK ID: OID00641, Limit of de | Analyses made in EDTA plasma. Data is presented as normalized protein expression (NPX) values, ( | Blood Analysis | SCAPIS |
| Proteomics | CPA1 | 5006 | 5.81 (5.43, 6.24) | NPX | Carboxypeptidase A1 - Olink CVD3 | Uni-Prot ID: P15085, OLINK ID: OID00624, Limit of de | Analyses made in EDTA plasma. Data is presented as normalized protein expression (NPX) values, ( | Blood Analysis | SCAPIS |
| Proteomics | CPB1 | 5006 | 5.65 (5.25, 6.07) | NPX | Carboxypeptidase B - Olink CVD3 | Uni-Prot ID: P15086, OLINK ID: OID00632, Limit of de | Analyses made in EDTA plasma. Data is presented as normalized protein expression (NPX) values, ( | Blood Analysis | SCAPIS |
| Proteomics | CSTB | 4836 | 4.96 (4.72, 5.22) | NPX | Cystatin-B - Olink CVD3 | Uni-Prot ID: P04080, OLINK ID: OID00575, Limit of de | Analyses made in EDTA plasma. Data is presented as normalized protein expression (NPX) values, ( | Blood Analysis | SCAPIS |
| Proteomics | CTRC | 4931 | 9.73 (9.31, 10.18) | NPX | Chymotrypsin C - Olink CVD2 | Uni-Prot ID: Q99895, OLINK ID: OID00414, Limit of de | Analyses made in EDTA plasma. Data is presented as normalized protein expression (NPX) values, ( | Blood Analysis | SCAPIS |
| Proteomics | CTSD | 4655 | 4.31 (4.1, 4.57) | NPX | Cathepsin D - Olink CVD3 | Uni-Prot ID: P07339, OLINK ID: OID00622, Limit of de | Analyses made in EDTA plasma. Data is presented as normalized protein expression (NPX) values, ( | Blood Analysis | SCAPIS |
| Proteomics | CTSL1 | 4933 | 5.79 (5.61, 5.98) | NPX | Cathepsin L1 - Olink CVD2 | Uni-Prot ID: P07711, OLINK ID: OID00459, Limit of de | Analyses made in EDTA plasma. Data is presented as normalized protein expression (NPX) values, ( | Blood Analysis | SCAPIS |
| Proteomics | CTSZ | 5006 | 5.32 (5.08, 5.57) | NPX | Cathepsin Z - Olink CVD3 | Uni-Prot ID: Q9UBR2, OLINK ID: OID00643, Limit of de | Analyses made in EDTA plasma. Data is presented as normalized protein expression (NPX) values, ( | Blood Analysis | SCAPIS |
| Proteomics | CXCL1 | 4933 | 8.44 (7.89, 8.9) | NPX | C-X-C motif chemokine 1 - Olink CVD2 | Uni-Prot ID: P09341, OLINK ID: OID00404, Limit of de | Analyses made in EDTA plasma. Data is presented as normalized protein expression (NPX) values, ( | Blood Analysis | SCAPIS |
| Proteomics | CXCL16 | 4922 | 4.94 (4.78, 5.12) | NPX | C-X-C motif chemokine 16 - Olink CVD3 | Uni-Prot ID: Q9H2A7, OLINK ID: OID00601, Limit of de | Analyses made in EDTA plasma. Data is presented as normalized protein expression (NPX) values, ( | Blood Analysis | SCAPIS |
| Proteomics | DCN | 4933 | 4.12 (3.98, 4.26) | NPX | Decorin - Olink CVD2 | Uni-Prot ID: P07585, OLINK ID: OID00444, Limit of de | Analyses made in EDTA plasma. Data is presented as normalized protein expression (NPX) values, ( | Blood Analysis | SCAPIS |
| Proteomics | DECR1 | 4933 | 5.25 (4.67, 5.85) | NPX | 2,4-dienoyl-CoA reductase, mitochondrial - Olink CVD2 | Uni-Prot ID: Q16698, OLINK ID: OID00424, Limit of de | Analyses made in EDTA plasma. Data is presented as normalized protein expression (NPX) values, ( | Blood Analysis | SCAPIS |
| Proteomics | DLK_1 | 5006 | 6.1 (5.69, 6.49) | NPX | Protein delta homolog 1 - Olink CVD3 | Uni-Prot ID: P80370, OLINK ID: OID00598, Limit of de | Analyses made in EDTA plasma. Data is presented as normalized protein expression (NPX) values, ( | Blood Analysis | SCAPIS |
| Proteomics | Dkk_1 | 4933 | 8.2 (7.87, 8.54) | NPX | Dickkopf-related protein 1 - Olink CVD2 | Uni-Prot ID: O94907, OLINK ID: OID00445, Limit of de | Analyses made in EDTA plasma. Data is presented as normalized protein expression (NPX) values, ( | Blood Analysis | SCAPIS |
| Proteomics | EGFR | 5006 | 3.03 (2.9, 3.15) | NPX | Epidermal growth factor receptor - Olink CVD3 | Uni-Prot ID: P00533, OLINK ID: OID00637, Limit of de | Analyses made in EDTA plasma. Data is presented as normalized protein expression (NPX) values, ( | Blood Analysis | SCAPIS |
| Proteomics | EPHB4 | 5006 | 4.66 (4.49, 4.83) | NPX | Ephrin type-B receptor 4 - Olink CVD3 | Uni-Prot ID: P54760, OLINK ID: OID00569, Limit of de | Analyses made in EDTA plasma. Data is presented as normalized protein expression (NPX) values, ( | Blood Analysis | SCAPIS |
| Proteomics | Ep_CAM | 5006 | 6.49 (5.64, 7.38) | NPX | Epithelial cell adhesion molecule - Olink CVD3 | Uni-Prot ID: P16422, OLINK ID: OID00610, Limit of de | Analyses made in EDTA plasma. Data is presented as normalized protein expression (NPX) values, ( | Blood Analysis | SCAPIS |
| Proteomics | FABP2 | 4933 | 7.64 (7.2, 8.12) | NPX | Fatty acid-binding protein, intesinal - Olink CVD2 | Uni-Prot ID: P12104, OLINK ID: OID00451, Limit of de | Analyses made in EDTA plasma. Data is presented as normalized protein expression (NPX) values, ( | Blood Analysis | SCAPIS |
| Proteomics | FABP4 | 5006 | 5.89 (5.44, 6.37) | NPX | Fatty acid-binding protein, adipocyte - Olink CVD3 | Uni-Prot ID: P15090, OLINK ID: OID00589, Limit of de | Analyses made in EDTA plasma. Data is presented as normalized protein expression (NPX) values, ( | Blood Analysis | SCAPIS |
| Proteomics | FAS | 5005 | 5.61 (5.43, 5.82) | NPX | Tumor necrosis factor receptor superfamily member 6 - Olink CVD3 | Uni-Prot ID: P25445, OLINK ID: OID00615, Limit of de | Analyses made in EDTA plasma. Data is presented as normalized protein expression (NPX) values, ( | Blood Analysis | SCAPIS |
| Proteomics | FGF_21 | 4933 | 5.32 (4.46, 6.16) | NPX | Fibroblast growth factor 21 - Olink CVD2 | Uni-Prot ID: Q9NSA1, OLINK ID: OID00410, Limit of de | Analyses made in EDTA plasma. Data is presented as normalized protein expression (NPX) values, ( | Blood Analysis | SCAPIS |
| Proteomics | FGF_23 | 4933 | 2.07 (1.86, 2.29) | NPX | Fibroblast growth factor 23 - Olink CVD2 | Uni-Prot ID: Q9GZV9, OLINK ID: OID00415, Limit of de | Analyses made in EDTA plasma. Data is presented as normalized protein expression (NPX) values, ( | Blood Analysis | SCAPIS |
| Proteomics | FS | 4933 | 10.72 (10.45, 11.0) | NPX | Follistatin - Olink CVD2 | Uni-Prot ID: P19883, OLINK ID: OID00418, Limit of de | Analyses made in EDTA plasma. Data is presented as normalized protein expression (NPX) values, ( | Blood Analysis | SCAPIS |

Table S2: Summary of clinical, molecular, and imaging-derived variables in SCAPIS, and disease outcomes in UKBB

| Category | Variable_Name | Data_Available | Summary | Unit | Heading | Context | Method | Source | Cohort |
|---|---|---|---|---|---|---|---|---|---|
| Proteomics | GDF_15 | 5006 | 5.59 (5.35, 5.88) | NPX | Growth/differentiation factor 15 - Olink CVD3 | Uni-Prot ID: Q99988, OLINK ID: OID00595, Limit of de | Analyses made in EDTA plasma. Data is presented as normalized protein expression (NPX) values, ( | Blood Analysis | SCAPIS |
| Proteomics | GDF_2 | 4933 | 6.93 (6.64, 7.21) | NPX | Growth/differentiation factor 2 - Olink CVD2 | Uni-Prot ID: Q9UK05, OLINK ID: OID00450, Limit of de | Analyses made in EDTA plasma. Data is presented as normalized protein expression (NPX) values, ( | Blood Analysis | SCAPIS |
| Proteomics | GH | 4933 | 8.76 (7.08, 10.54) | NPX | Growth hormone - Olink CVD2 | Uni-Prot ID: P01241, OLINK ID: OID00417, Limit of de | Analyses made in EDTA plasma. Data is presented as normalized protein expression (NPX) values, ( | Blood Analysis | SCAPIS |
| Proteomics | GIF | 4933 | 6.07 (5.58, 6.55) | NPX | Gastric intrinsic factor - Olink CVD2 | Uni-Prot ID: P27352, OLINK ID: OID00407, Limit of de | Analyses made in EDTA plasma. Data is presented as normalized protein expression (NPX) values, ( | Blood Analysis | SCAPIS |
| Proteomics | GLO1 | 4933 | 5.71 (5.3, 6.11) | NPX | Lactoylglutathione lyase - Olink CVD2 | Uni-Prot ID: Q04760, OLINK ID: OID00419, Limit of de | Analyses made in EDTA plasma. Data is presented as normalized protein expression (NPX) values, ( | Blood Analysis | SCAPIS |
| Proteomics | GP6 | 5006 | 2.77 (2.45, 3.11) | NPX | Platelet glycoprotein VI - Olink CVD3 | Uni-Prot ID: Q9HCN6, OLINK ID: OID05026, Limit of d | Analyses made in EDTA plasma. Data is presented as normalized protein expression (NPX) values, ( | Blood Analysis | SCAPIS |
| Proteomics | GRN | 5006 | 6.27 (6.06, 6.45) | NPX | Granulins - Olink CVD3 | Uni-Prot ID: P28799, OLINK ID: OID00579, Limit of de | Analyses made in EDTA plasma. Data is presented as normalized protein expression (NPX) values, ( | Blood Analysis | SCAPIS |
| Proteomics | GT | 4933 | 1.02 (0.73, 1.35) | NPX | Gastrotropin - Olink CVD2 | Uni-Prot ID: P51161, OLINK ID: OID00454, Limit of de | Analyses made in EDTA plasma. Data is presented as normalized protein expression (NPX) values, ( | Blood Analysis | SCAPIS |
| Proteomics | Gal_3 | 5006 | 5.59 (5.38, 5.82) | NPX | Galectin-3 - Olink CVD3 | Uni-Prot ID: P17931, OLINK ID: OID00578, Limit of de | Analyses made in EDTA plasma. Data is presented as normalized protein expression (NPX) values, ( | Blood Analysis | SCAPIS |
| Proteomics | Gal_4 | 5006 | 3.63 (3.35, 3.95) | NPX | Galectin-4 - Olink CVD3 | Uni-Prot ID: P56470, OLINK ID: OID00626, Limit of de | Analyses made in EDTA plasma. Data is presented as normalized protein expression (NPX) values, ( | Blood Analysis | SCAPIS |
| Proteomics | Gal_9 | 4933 | 7.92 (7.72, 8.13) | NPX | Galectin-9 - Olink CVD2 | Uni-Prot ID: O00182, OLINK ID: OID00406, Limit of de | Analyses made in EDTA plasma. Data is presented as normalized protein expression (NPX) values, ( | Blood Analysis | SCAPIS |
| Proteomics | HAOX1 | 4933 | 2.87 (2.18, 3.73) | NPX | Hydroxyacid oxidase 1 - Olink CVD2 | Uni-Prot ID: Q9UJM8, OLINK ID: OID00470, Limit of d | Analyses made in EDTA plasma. Data is presented as normalized protein expression (NPX) values, ( | Blood Analysis | SCAPIS |
| Proteomics | HB_EGF | 4933 | 3.37 (3.05, 3.71) | NPX | Proheparin-binding EGF-like growth factor - Olink CVD2 | Uni-Prot ID: Q99075, OLINK ID: OID00449, Limit of de | Analyses made in EDTA plasma. Data is presented as normalized protein expression (NPX) values, ( | Blood Analysis | SCAPIS |
| Proteomics | HO_1 | 4933 | 11.15 (10.89, 11.41) | NPX | Heme oxygenase 1 - Olink CVD2 | Uni-Prot ID: P09601, OLINK ID: OID00432, Limit of de | Analyses made in EDTA plasma. Data is presented as normalized protein expression (NPX) values, ( | Blood Analysis | SCAPIS |
| Proteomics | HSP_27 | 4933 | 9.95 (9.84, 10.05) | NPX | Heat shock 27 kDa protein - Olink CVD2 | Uni-Prot ID: P04792, OLINK ID: OID00465, Limit of de | Analyses made in EDTA plasma. Data is presented as normalized protein expression (NPX) values, ( | Blood Analysis | SCAPIS |
| Proteomics | ICAM_2 | 5006 | 5.11 (4.86, 5.35) | NPX | Intercellular adhesion molecule 2 - Olink CVD3 | Uni-Prot ID: P13598, OLINK ID: OID00646, Limit of de | Analyses made in EDTA plasma. Data is presented as normalized protein expression (NPX) values, ( | Blood Analysis | SCAPIS |
| Proteomics | IDUA | 4933 | 4.93 (4.61, 5.27) | NPX | Alpha-L-iduronidase - Olink CVD2 | UNI-Prot ID: P35475, OLINK ID: OID00393, Limit of de | Analyses made in EDTA plasma. Data is presented as normalized protein expression (NPX) values, ( | Blood Analysis | SCAPIS |
| Proteomics | IGFBP_1 | 5006 | 5.19 (4.56, 5.81) | NPX | Insulin-like growth factor-binding protein 1 - Olink CVD3 | Uni-Prot ID: P08833, OLINK ID: OID00604, Limit of de | Analyses made in EDTA plasma. Data is presented as normalized protein expression (NPX) values, ( | Blood Analysis | SCAPIS |
| Proteomics | IGFBP_2 | 5006 | 7.68 (7.22, 8.13) | NPX | Insulin-like growth factor-binding protein 2 - Olink CVD3 | Uni-Prot ID: P18065, OLINK ID: OID00650, Limit of de | Analyses made in EDTA plasma. Data is presented as normalized protein expression (NPX) values, ( | Blood Analysis | SCAPIS |
| Proteomics | IGFBP_7 | 5006 | 7.39 (7.21, 7.57) | NPX | Insulin-like growth factor-binding protein 7 - Olink CVD3 | Uni-Prot ID: Q16270, OLINK ID: OID00638, Limit of de | Analyses made in EDTA plasma. Data is presented as normalized protein expression (NPX) values, ( | Blood Analysis | SCAPIS |
| Proteomics | IL16 | 4933 | 5.51 (5.24, 5.77) | NPX | Pro-interleukin-16 - Olink CVD2 | Uni-Prot ID: Q14005, OLINK ID: OID00434, Limit of de | Analyses made in EDTA plasma. Data is presented as normalized protein expression (NPX) values, ( | Blood Analysis | SCAPIS |
| Proteomics | IL18 | 4933 | 7.35 (7.04, 7.69) | NPX | Interleukin-18 - Olink CVD2 | Uni-Prot ID: Q14116, OLINK ID: OID00409, Limit of de | Analyses made in EDTA plasma. Data is presented as normalized protein expression (NPX) values, ( | Blood Analysis | SCAPIS |
| Proteomics | IL1RL2 | 4933 | 3.34 (3.1, 3.58) | NPX | Interleukin-1 receptor-like 2 - Olink CVD2 | Uni-Prot ID: Q9HB29, OLINK ID: OID00400, Limit of de | Analyses made in EDTA plasma. Data is presented as normalized protein expression (NPX) values, ( | Blood Analysis | SCAPIS |
| Proteomics | IL2_RA | 5006 | 4.05 (3.8, 4.31) | NPX | Interleukin-2 receptor subunit alpha - Olink CVD3 | Uni-Prot ID: P01589, OLINK ID: OID00570, Limit of de | Analyses made in EDTA plasma. Data is presented as normalized protein expression (NPX) values, ( | Blood Analysis | SCAPIS |
| Proteomics | IL6 | 4933 | 2.82 (2.49, 3.26) | NPX | Interleukin-6 - Olink CVD2 | UNI-Prot ID: P05231, OLINK ID: OID00390, Limit of de | Analyses made in EDTA plasma. Data is presented as normalized protein expression (NPX) values, ( | Blood Analysis | SCAPIS |
| Proteomics | IL_17D | 4933 | 2.27 (2.14, 2.42) | NPX | Interleukin-17D - Olink CVD2 | Uni-Prot ID: Q8TAD2, OLINK ID: OID00403, Limit of de | Analyses made in EDTA plasma. Data is presented as normalized protein expression (NPX) values, ( | Blood Analysis | SCAPIS |
| Proteomics | IL_17RA | 5006 | 3.69 (3.35, 4.01) | NPX | Interleukin-17 receptor A - Olink CVD3 | Uni-Prot ID: Q96F46, OLINK ID: OID00566, Limit of de | Analyses made in EDTA plasma. Data is presented as normalized protein expression (NPX) values, ( | Blood Analysis | SCAPIS |
| Proteomics | IL_18BP | 5006 | 5.93 (5.74, 6.14) | NPX | Interleukin-18-binding protein - Olink CVD3 | Uni-Prot ID: O95998, OLINK ID: OID00640, Limit of de | Analyses made in EDTA plasma. Data is presented as normalized protein expression (NPX) values, ( | Blood Analysis | SCAPIS |
| Proteomics | IL_1RT1 | 5006 | 6.43 (6.27, 6.61) | NPX | Interleukin-1 receptor type 1 - Olink CVD3 | Uni-Prot ID: P14778, OLINK ID: OID00613, Limit of de | Analyses made in EDTA plasma. Data is presented as normalized protein expression (NPX) values, ( | Blood Analysis | SCAPIS |
| Proteomics | IL_1RT2 | 5006 | 5.6 (5.4, 5.8) | NPX | Interleukin-1 receptor type 2 - Olink CVD3 | Uni-Prot ID: P27930, OLINK ID: OID00627, Limit of de | Analyses made in EDTA plasma. Data is presented as normalized protein expression (NPX) values, ( | Blood Analysis | SCAPIS |
| Proteomics | IL_1ra | 4933 | 3.38 (2.98, 3.84) | NPX | Interleukin-1 receptor antagonist protein - Olink CVD2 | UNI-Prot ID: P18510, OLINK ID: OID00389, Limit of de | Analyses made in EDTA plasma. Data is presented as normalized protein expression (NPX) values, ( | Blood Analysis | SCAPIS |
| Proteomics | IL_27 | 4933 | 4.16 (3.9, 4.43) | NPX | Interleukin-27 - Olink CVD2 | Uni-Prot ID: Q8NEV9,Q14213, OLINK ID: OID00402, L | Analyses made in EDTA plasma. Data is presented as normalized protein expression (NPX) values, ( | Blood Analysis | SCAPIS |
| Proteomics | IL_4RA | 4933 | 1.49 (1.35, 1.64) | NPX | Interleukin-4 receptor subunit alpha - Olink CVD2 | UNI-Prot ID:P24394, OLINK ID: OID00387, Limit of de | Analyses made in EDTA plasma. Data is presented as normalized protein expression (NPX) values, ( | Blood Analysis | SCAPIS |
| Proteomics | IL_6RA | 5006 | 12.26 (11.97, 12.51) | NPX | Interleukin-6 receptor subunit alpha - Olink CVD3 | Uni-Prot ID: P08887, OLINK ID: OID00602, Limit of de | Analyses made in EDTA plasma. Data is presented as normalized protein expression (NPX) values, ( | Blood Analysis | SCAPIS |
| Proteomics | ITGB1BP2 | 4933 | 3.95 (3.43, 4.51) | NPX | Melusin - Olink CVD2 | Uni-Prot ID: Q9UKP3, OLINK ID: OID00443, Limit of de | Analyses made in EDTA plasma. Data is presented as normalized protein expression (NPX) values, ( | Blood Analysis | SCAPIS |
| Proteomics | ITGB2 | 5006 | 5.87 (5.64, 6.09) | NPX | Integrin beta-2 - Olink CVD3 | Uni-Prot ID: P05107, OLINK ID: OID00565, Limit of de | Analyses made in EDTA plasma. Data is presented as normalized protein expression (NPX) values, ( | Blood Analysis | SCAPIS |
| Proteomics | IgG_Fc_receptor_II_b | 4933 | 1.92 (1.56, 2.24) | NPX | Low affinity immunoglobulin gamma Fc region receptor II-b - Olink CVD2 | Uni-Prot ID: P31994, OLINK ID: OID00442, Limit of de | Analyses made in EDTA plasma. Data is presented as normalized protein expression (NPX) values, ( | Blood Analysis | SCAPIS |
| Proteomics | JAM_A | 5006 | 5.76 (5.38, 6.17) | NPX | Junctional adhesion molecule A - Olink CVD3 | Uni-Prot ID: Q9Y624, OLINK ID: OID00625, Limit of de | Analyses made in EDTA plasma. Data is presented as normalized protein expression (NPX) values, ( | Blood Analysis | SCAPIS |
| Proteomics | KIM1 | 4933 | 6.88 (6.47, 7.31) | NPX | Kidney Injury Molecule - Olink CVD2 | Uni-Prot ID: Q96D42, OLINK ID: OID00426, Limit of de | Analyses made in EDTA plasma. Data is presented as normalized protein expression (NPX) values, ( | Blood Analysis | SCAPIS |
| Proteomics | KLK6 | 5006 | 5.55 (5.36, 5.76) | NPX | Kallikrein-6 - Olink CVD3 | Uni-Prot ID: Q92876, OLINK ID: OID00647, Limit of de | Analyses made in EDTA plasma. Data is presented as normalized protein expression (NPX) values, ( | Blood Analysis | SCAPIS |
| Proteomics | LDL_receptor | 5006 | 4.83 (4.44, 5.21) | NPX | Low-density lipoprotein receptor - Olink CVD3 | Uni-Prot ID: P01130, OLINK ID: OID00564, Limit of de | Analyses made in EDTA plasma. Data is presented as normalized protein expression (NPX) values, ( | Blood Analysis | SCAPIS |
| Proteomics | LEP | 4933 | 5.29 (4.48, 5.98) | NPX | Leptin - Olink CVD2 | Uni-Prot ID: P41159, OLINK ID: OID00463, Limit of de | Analyses made in EDTA plasma. Data is presented as normalized protein expression (NPX) values, ( | Blood Analysis | SCAPIS |
| Proteomics | LOX_1 | 4933 | 5.41 (5.12, 5.72) | NPX | Lectin-like oxidized LDL receptor 1 - Olink CVD2 | Uni-Prot ID: P78380, OLINK ID: OID00405, Limit of de | Analyses made in EDTA plasma. Data is presented as normalized protein expression (NPX) values, ( | Blood Analysis | SCAPIS |
| Proteomics | LPL | 4933 | 8.63 (8.3, 8.89) | NPX | Lipoprotein lipase - Olink CVD2 | Uni-Prot ID: P06858, OLINK ID: OID00446, Limit of de | Analyses made in EDTA plasma. Data is presented as normalized protein expression (NPX) values, ( | Blood Analysis | SCAPIS |
| Proteomics | LTBR | 5006 | 4.03 (3.85, 4.2) | NPX | Lymphotoxin-beta receptor - Olink CVD3 | Uni-Prot ID: P36941, OLINK ID: OID00583, Limit of de | Analyses made in EDTA plasma. Data is presented as normalized protein expression (NPX) values, ( | Blood Analysis | SCAPIS |
| Proteomics | MARCO | 4933 | 5.92 (5.78, 6.06) | NPX | Macrophage receptor MARCO - Olink CVD2 | Uni-Prot ID: Q9UEW3, OLINK ID: OID00453, Limit of d | Analyses made in EDTA plasma. Data is presented as normalized protein expression (NPX) values, ( | Blood Analysis | SCAPIS |
| Proteomics | MB | 5006 | 6.99 (6.67, 7.33) | NPX | Myoglobin - Olink CVD3 | Uni-Prot ID: P02144, OLINK ID: OID00616, Limit of de | Analyses made in EDTA plasma. Data is presented as normalized protein expression (NPX) values, ( | Blood Analysis | SCAPIS |
| Proteomics | MCP_1 | 5006 | 3.7 (3.5, 3.93) | NPX | Monocyte chemotactic protein 1 - Olink CVD3 | Uni-Prot ID: P13500, OLINK ID: OID00576, Limit of de | Analyses made in EDTA plasma. Data is presented as normalized protein expression (NPX) values, ( | Blood Analysis | SCAPIS |
| Proteomics | MEPE | 5006 | 5.7 (5.43, 5.95) | NPX | Matrix extracellular phosphoglycoprotein - Olink CVD3 | Uni-Prot ID: Q9NQ76, OLINK ID: OID00132, Limit of d | Analyses made in EDTA plasma. Data is presented as normalized protein expression (NPX) values, ( | Blood Analysis | SCAPIS |
| Proteomics | MERTK | 4933 | 4.77 (4.56, 4.98) | NPX | Tyrosine-protein kinase Mer - Olink CVD2 | Uni-Prot ID: Q12866, OLINK ID: OID00425, Limit of de | Analyses made in EDTA plasma. Data is presented as normalized protein expression (NPX) values, ( | Blood Analysis | SCAPIS |
| Proteomics | MMP12 | 4933 | 5.84 (5.49, 6.2) | NPX | Matrix metalloproteinase-12 - Olink CVD2 | Uni-Prot ID: P39900, OLINK ID: OID00456, Limit of de | Analyses made in EDTA plasma. Data is presented as normalized protein expression (NPX) values, ( | Blood Analysis | SCAPIS |
| Proteomics | MMP7 | 4933 | 8.33 (8.11, 8.54) | NPX | Matrix metalloproteinase-7 - Olink CVD2 | Uni-Prot ID: P09237, OLINK ID: OID00441, Limit of de | Analyses made in EDTA plasma. Data is presented as normalized protein expression (NPX) values, ( | Blood Analysis | SCAPIS |
| Proteomics | MMP_2 | 5006 | 4.14 (3.93, 4.33) | NPX | Matrix metalloproteinase-2 - Olink CVD3 | Uni-Prot ID: P08253, OLINK ID: OID00614, Limit of de | Analyses made in EDTA plasma. Data is presented as normalized protein expression (NPX) values, ( | Blood Analysis | SCAPIS |
| Proteomics | MMP_3 | 5006 | 6.46 (5.99, 6.94) | NPX | Matrix metalloproteinase-3 - Olink CVD3 | Uni-Prot ID: P08254, OLINK ID: OID00644, Limit of de | Analyses made in EDTA plasma. Data is presented as normalized protein expression (NPX) values, ( | Blood Analysis | SCAPIS |
| Proteomics | MMP_9 | 5006 | 4.21 (3.75, 4.71) | NPX | Matrix metalloproteinase-9 - Olink CVD3 | Uni-Prot ID: P14780, OLINK ID: OID00568, Limit of de | Analyses made in EDTA plasma. Data is presented as normalized protein expression (NPX) values, ( | Blood Analysis | SCAPIS |
| Proteomics | MPO | 4918 | 3.42 (3.21, 3.65) | NPX | Myeloperoxidase - Olink CVD3 | Uni-Prot ID: P05164, OLINK ID: OID00600, Limit of de | Analyses made in EDTA plasma. Data is presented as normalized protein expression (NPX) values, ( | Blood Analysis | SCAPIS |
| Proteomics | NEMO | 4933 | 5.1 (4.59, 5.6) | NPX | NF-kappa-B essential modulator - Olink CVD2 | Uni-Prot ID: Q9Y6K9, OLINK ID: OID00467, Limit of de | Analyses made in EDTA plasma. Data is presented as normalized protein expression (NPX) values, ( | Blood Analysis | SCAPIS |
| Proteomics | NT_proBNP | 5006 | 3.66 (3.02, 4.34) | NPX | N-terminal prohormone brain natriuretic peptide - Olink CVD3 | Uni-Prot ID: NA, OLINK ID: OID00131, Limit of detecti | Analyses made in EDTA plasma. Data is presented as normalized protein expression (NPX) values, ( | Blood Analysis | SCAPIS |
| Proteomics | Notch_3 | 5006 | 4.62 (4.41, 4.84) | NPX | Neurogenic locus notch homolog protein 3 - Olink CVD3 | Uni-Prot ID: Q9UM47, OLINK ID: OID00584, Limit of d | Analyses made in EDTA plasma. Data is presented as normalized protein expression (NPX) values, ( | Blood Analysis | SCAPIS |
| Proteomics | OPG | 5006 | 3.83 (3.64, 4.01) | NPX | Osteoprotegerin - Olink CVD3 | Uni-Prot ID: O00300, OLINK ID: OID00571, Limit of de | Analyses made in EDTA plasma. Data is presented as normalized protein expression (NPX) values, ( | Blood Analysis | SCAPIS |
| Proteomics | OPN | 5006 | 6.02 (5.74, 6.32) | NPX | Osteopontin - Olink CVD3 | Uni-Prot ID: P10451, OLINK ID: OID00621, Limit of de | Analyses made in EDTA plasma. Data is presented as normalized protein expression (NPX) values, ( | Blood Analysis | SCAPIS |
| Proteomics | PAI | 5006 | 5.66 (5.11, 6.21) | NPX | Plasminogen activator inhibitor 1 - Olink CVD3 | Uni-Prot ID: P05121, OLINK ID: OID00591, Limit of de | Analyses made in EDTA plasma. Data is presented as normalized protein expression (NPX) values, ( | Blood Analysis | SCAPIS |
| Proteomics | PAPPA | 4933 | 1.56 (1.33, 1.79) | NPX | Pappalysin-1 - Olink CVD2 | Uni-Prot ID: Q13219, OLINK ID: OID00421, Limit of de | Analyses made in EDTA plasma. Data is presented as normalized protein expression (NPX) values, ( | Blood Analysis | SCAPIS |
| Proteomics | PARP_1 | 4933 | 1.66 (1.47, 1.87) | NPX | Poly [ADP-ribose] polymerase 1 - Olink CVD2 | Uni-Prot ID: P09874, OLINK ID: OID00469, Limit of de | Analyses made in EDTA plasma. Data is presented as normalized protein expression (NPX) values, ( | Blood Analysis | SCAPIS |
| Proteomics | PAR_1 | 4933 | 7.98 (7.75, 8.21) | NPX | Proteinase-activated receptor 1 - Olink CVD2 | UNI-Prot ID: P25116, OLINK ID: OID00395, Limit of de | Analyses made in EDTA plasma. Data is presented as normalized protein expression (NPX) values, ( | Blood Analysis | SCAPIS |
| Proteomics | PCSK9 | 5006 | 2.95 (2.72, 3.2) | NPX | Proprotein convertase subtilisin/kexin type 9 - Olink CVD3 | Uni-Prot ID: Q8NBP7, OLINK ID: OID00619, Limit of de | Analyses made in EDTA plasma. Data is presented as normalized protein expression (NPX) values, ( | Blood Analysis | SCAPIS |
| Proteomics | PDGF_subunit_A | 5006 | 3.82 (3.38, 4.27) | NPX | Platelet-derived growth factor subunit A - Olink CVD3 | Uni-Prot ID: P04085, OLINK ID: OID00648, Limit of de | Analyses made in EDTA plasma. Data is presented as normalized protein expression (NPX) values, ( | Blood Analysis | SCAPIS |
| Proteomics | PDGF_subunit_B | 4933 | 9.51 (9.01, 10.0) | NPX | Platelet-derived growth factor subunit B - Olink CVD2 | Uni-Prot ID: P01127, OLINK ID: OID00401, Limit of de | Analyses made in EDTA plasma. Data is presented as normalized protein expression (NPX) values, ( | Blood Analysis | SCAPIS |
| Proteomics | PD_L2 | 4933 | 1.81 (1.63, 1.99) | NPX | Programmed cell death 1 ligand 2 - Olink CVD2 | Uni-Prot ID: Q9BQ51, OLINK ID: OID00458, Limit of de | Analyses made in EDTA plasma. Data is presented as normalized protein expression (NPX) values, ( | Blood Analysis | SCAPIS |
| Proteomics | PECAM_1 | 5006 | 4.85 (4.6, 5.12) | NPX | Platelet endothelial cell adhesion molecule - Olink CVD3 | Uni-Prot ID: P16284, OLINK ID: OID00652, Limit of de | Analyses made in EDTA plasma. Data is presented as normalized protein expression (NPX) values, ( | Blood Analysis | SCAPIS |
| Proteomics | PGF | 4933 | 7.0 (6.84, 7.15) | NPX | Placenta growth factor - Olink CVD2 | UNI-Prot ID: P49763, OLINK ID: OID00384, Limit of de | Analyses made in EDTA plasma. Data is presented as normalized protein expression (NPX) values, ( | Blood Analysis | SCAPIS |
| Proteomics | PGLYRP1 | 5006 | 7.68 (7.42, 7.96) | NPX | Peptidoglycan recognition protein 1 - Olink CVD3 | Uni-Prot ID: O75594, OLINK ID: OID00623, Limit of de | Analyses made in EDTA plasma. Data is presented as normalized protein expression (NPX) values, ( | Blood Analysis | SCAPIS |
| Proteomics | PI3 | 4671 | 3.57 (3.29, 3.88) | NPX | Elafin - Olink CVD3 | Uni-Prot ID: P19957, OLINK ID: OID00609, Limit of de | Analyses made in EDTA plasma. Data is presented as normalized protein expression (NPX) values, ( | Blood Analysis | SCAPIS |
| Proteomics | PIgR | 4933 | 5.96 (5.86, 6.06) | NPX | Polymeric immunoglobulin receptor - Olink CVD2 | Uni-Prot ID: P01833, OLINK ID: OID00411, Limit of de | Analyses made in EDTA plasma. Data is presented as normalized protein expression (NPX) values, ( | Blood Analysis | SCAPIS |
| Proteomics | PLC | 5004 | 6.76 (6.57, 6.93) | NPX | Perlecan - Olink CVD3 | Uni-Prot ID: P98160, OLINK ID: OID00582, Limit of de | Analyses made in EDTA plasma. Data is presented as normalized protein expression (NPX) values, ( | Blood Analysis | SCAPIS |
| Proteomics | PON3 | 5006 | 6.07 (5.62, 6.46) | NPX | Paraoxonase - Olink CVD3 | Uni-Prot ID: Q15166, OLINK ID: OID00642, Limit of de | Analyses made in EDTA plasma. Data is presented as normalized protein expression (NPX) values, ( | Blood Analysis | SCAPIS |
| Proteomics | PRELP | 4932 | 5.86 (5.76, 5.96) | NPX | Prolargin - Olink CVD2 | Uni-Prot ID: P51888, OLINK ID: OID00431, Limit of de | Analyses made in EDTA plasma. Data is presented as normalized protein expression (NPX) values, ( | Blood Analysis | SCAPIS |
| Proteomics | PRSS27 | 4932 | 8.07 (7.8, 8.38) | NPX | Serine protease 27 - Olink CVD2 | Uni-Prot ID: Q9BQR3, OLINK ID: OID00397, Limit of de | Analyses made in EDTA plasma. Data is presented as normalized protein expression (NPX) values, ( | Blood Analysis | SCAPIS |
| Proteomics | PRSS8 | 4933 | 8.38 (8.12, 8.66) | NPX | Prostasin - Olink CVD2 | Uni-Prot ID: Q16651, OLINK ID: OID00447, Limit of de | Analyses made in EDTA plasma. Data is presented as normalized protein expression (NPX) values, ( | Blood Analysis | SCAPIS |
| Proteomics | PRTN3 | 4671 | 4.89 (4.62, 5.16) | NPX | Myeloblastin - Olink CVD3 | Uni-Prot ID: P24158, OLINK ID: OID00618, Limit of de | Analyses made in EDTA plasma. Data is presented as normalized protein expression (NPX) values, ( | Blood Analysis | SCAPIS |
| Proteomics | PSGL_1 | 4933 | 4.03 (3.89, 4.18) | NPX | P-selectin glycoprotein ligand 1 - Olink CVD2 | Uni-Prot ID: Q14242, OLINK ID: OID00438, Limit of de | Analyses made in EDTA plasma. Data is presented as normalized protein expression (NPX) values, ( | Blood Analysis | SCAPIS |
| Proteomics | PSP_D | 5006 | 3.0 (2.57, 3.48) | NPX | Pulmonary surfactant-associated protein D - Olink CVD3 | Uni-Prot ID: P35247, OLINK ID: OID00608, Limit of de | Analyses made in EDTA plasma. Data is presented as normalized protein expression (NPX) values, ( | Blood Analysis | SCAPIS |
| Proteomics | PTX3 | 4933 | 1.95 (1.7, 2.2) | NPX | Pentraxin-related protein PTX3 - Olink CVD2 | Uni-Prot ID: P26022, OLINK ID: OID00437, Limit of de | Analyses made in EDTA plasma. Data is presented as normalized protein expression (NPX) values, ( | Blood Analysis | SCAPIS |
| Proteomics | RAGE | 4933 | 4.29 (4.07, 4.53) | NPX | Receptor for advanced glycosylation end products - Olink CVD2 | Uni-Prot ID: Q15109, OLINK ID: OID00412, Limit of de | Analyses made in EDTA plasma. Data is presented as normalized protein expression (NPX) values, ( | Blood Analysis | SCAPIS |
| Proteomics | RARRES2 | 5006 | 11.69 (11.46, 11.92) | NPX | Retinoic acid receptor responder protein 2 - Olink CVD3 | Uni-Prot ID: Q99969, OLINK ID: OID00645, Limit of de | Analyses made in EDTA plasma. Data is presented as normalized protein expression (NPX) values, ( | Blood Analysis | SCAPIS |
| Proteomics | REN | 4932 | 6.78 (6.32, 7.24) | NPX | Renin - Olink CVD2 | Uni-Prot ID: P00797, OLINK ID: OID00423, Limit of de | Analyses made in EDTA plasma. Data is presented as normalized protein expression (NPX) values, ( | Blood Analysis | SCAPIS |
| Proteomics | RETN | 5006 | 6.16 (5.89, 6.42) | NPX | Resistin - Olink CVD3 | Uni-Prot ID: Q9HD89, OLINK ID: OID00603, Limit of de | Analyses made in EDTA plasma. Data is presented as normalized protein expression (NPX) values, ( | Blood Analysis | SCAPIS |
| Proteomics | SCF | 4933 | 9.69 (9.45, 9.93) | NPX | Stem cell factor - Olink CVD2 | Uni-Prot ID: P21583, OLINK ID: OID00408, Limit of de | Analyses made in EDTA plasma. Data is presented as normalized protein expression (NPX) values, ( | Blood Analysis | SCAPIS |
| Proteomics | SCGB3A2 | 5006 | 2.78 (2.35, 3.22) | NPX | Secretoglobin family 3A member 2 - Olink CVD3 | Uni-Prot ID: Q96PL1, OLINK ID: OID00636, Limit of de | Analyses made in EDTA plasma. Data is presented as normalized protein expression (NPX) values, ( | Blood Analysis | SCAPIS |
| Proteomics | SELE | 5006 | 12.59 (12.17, 12.98) | NPX | E-selectin - Olink CVD3 | Uni-Prot ID: P16581, OLINK ID: OID00596, Limit of de | Analyses made in EDTA plasma. Data is presented as normalized protein expression (NPX) values, ( | Blood Analysis | SCAPIS |
| Proteomics | SELP | 5006 | 10.0 (9.69, 10.33) | NPX | P-selectin - Olink CVD3 | Uni-Prot ID: P16109, OLINK ID: OID00574, Limit of de | Analyses made in EDTA plasma. Data is presented as normalized protein expression (NPX) values, ( | Blood Analysis | SCAPIS |
| Proteomics | SERPINA12 | 4933 | 2.42 (1.84, 3.21) | NPX | Serpin A12 - Olink CVD2 | Uni-Prot ID: Q8IW75, OLINK ID: OID00422, Limit of de | Analyses made in EDTA plasma. Data is presented as normalized protein expression (NPX) values, ( | Blood Analysis | SCAPIS |
| Proteomics | SHPS_1 | 5006 | 3.46 (3.2, 3.71) | NPX | Tyrosine-protein phosphatase non-receptor type substrate 1 - Olink CVD3 | Uni-Prot ID: P78324, OLINK ID: OID00628, Limit of de | Analyses made in EDTA plasma. Data is presented as normalized protein expression (NPX) values, ( | Blood Analysis | SCAPIS |
| Proteomics | SLAMF7 | 4933 | 2.59 (2.32, 2.89) | NPX | SLAM family member 7 - Olink CVD2 | UNI-Prot ID: Q9NQ25, OLINK ID: OID00383, Limit of d | Analyses made in EDTA plasma. Data is presented as normalized protein expression (NPX) values, ( | Blood Analysis | SCAPIS |
| Proteomics | SOD2 | 4933 | 8.38 (8.25, 8.52) | NPX | Superoxide dismutase [Mn], mitochondrial - Olink CVD2 | Uni-Prot ID: P04179, OLINK ID: OID00413, Limit of de | Analyses made in EDTA plasma. Data is presented as normalized protein expression (NPX) values, ( | Blood Analysis | SCAPIS |
| Proteomics | SORT1 | 4933 | 7.65 (7.48, 7.83) | NPX | Sortilin - Olink CVD2 | Uni-Prot ID: Q99523, OLINK ID: OID00435, Limit of de | Analyses made in EDTA plasma. Data is presented as normalized protein expression (NPX) values, ( | Blood Analysis | SCAPIS |
| Proteomics | SPON1 | 5006 | 1.23 (1.09, 1.38) | NPX | Spondin-1 - Olink CVD3 | Uni-Prot ID: Q9HCB6, OLINK ID: OID00599, Limit of de | Analyses made in EDTA plasma. Data is presented as normalized protein expression (NPX) values, ( | Blood Analysis | SCAPIS |
| Proteomics | SPON2 | 4933 | 9.62 (9.51, 9.73) | NPX | Spondin-2 - Olink CVD2 | Uni-Prot ID: Q9BUD6, OLINK ID: OID00416, Limit of de | Analyses made in EDTA plasma. Data is presented as normalized protein expression (NPX) values, ( | Blood Analysis | SCAPIS |
| Proteomics | SRC | 4933 | 6.74 (6.49, 6.89) | NPX | Proto-oncogene tyrosine-protein kinase Src - Olink CVD2 | UNI-Prot ID: P12931, OLINK ID:OID00387, Limit of de | Analyses made in EDTA plasma. Data is presented as normalized protein expression (NPX) values, ( | Blood Analysis | SCAPIS |

**Table S2: Summary of clinical, molecular, and imaging-derived variables in SCAPIS, and disease outcomes in UKBB**

| Category | Variable_Name | Data_Available | Summary | Unit | Heading | Context | Method | Source | Cohort |
|---|---|---|---|---|---|---|---|---|---|
| Proteomics | ST2 | 5006 | 4.14 (3.78, 4.47) | NPX | ST2 protein - Olink CVD3 | Uni-Prot ID: Q01638, OLINK ID: OID00634, Limit of de | Analyses made in EDTA plasma. Data is presented as normalized protein expression (NPX) values, ( | Blood Analysis | SCAPIS |
| Proteomics | STK4 | 4933 | 4.93 (4.46, 5.28) | NPX | Serine/threonine-protein kinase 4 - Olink CVD2 | UNI-Prot ID: Q13043, OLINK ID: OID00392, Limit of de | Analyses made in EDTA plasma. Data is presented as normalized protein expression (NPX) values, ( | Blood Analysis | SCAPIS |
| Proteomics | TF | 4933 | 5.51 (5.34, 5.7) | NPX | Tissue factor - Olink CVD2 | Uni-Prot ID: P13726, OLINK ID: OID00399, Limit of de | Analyses made in EDTA plasma. Data is presented as normalized protein expression (NPX) values, ( | Blood Analysis | SCAPIS |
| Proteomics | TFF3 | 5006 | 5.07 (4.89, 5.28) | NPX | Trefoil factor 3 - Olink CVD3 | Uni-Prot ID: Q07654, OLINK ID: OID00573, Limit of de | Analyses made in EDTA plasma. Data is presented as normalized protein expression (NPX) values, ( | Blood Analysis | SCAPIS |
| Proteomics | TFPI | 5006 | 9.5 (9.29, 9.71) | NPX | Tissue factor pathway inhibitor - Olink CVD3 | Uni-Prot ID: P10646, OLINK ID: OID00590, Limit of de | Analyses made in EDTA plasma. Data is presented as normalized protein expression (NPX) values, ( | Blood Analysis | SCAPIS |
| Proteomics | TGM2 | 4933 | 7.1 (6.74, 7.44) | NPX | Protein-glutamine gamma-glutamyltransferase 2 - Olink CVD2 | Uni-Prot ID: P21980, OLINK ID: OID00462, Limit of de | Analyses made in EDTA plasma. Data is presented as normalized protein expression (NPX) values, ( | Blood Analysis | SCAPIS |
| Proteomics | THBS2 | 4933 | 5.38 (5.25, 5.51) | NPX | Thrombospondin-2 - Olink CVD2 | Uni-Prot ID: P35442, OLINK ID: OID00427, Limit of de | Analyses made in EDTA plasma. Data is presented as normalized protein expression (NPX) values, ( | Blood Analysis | SCAPIS |
| Proteomics | THPO | 4933 | 1.82 (1.67, 1.98) | NPX | Thrombopoietin - Olink CVD2 | Uni-Prot ID: P40225, OLINK ID: OID00452, Limit of de | Analyses made in EDTA plasma. Data is presented as normalized protein expression (NPX) values, ( | Blood Analysis | SCAPIS |
| Proteomics | TIE2 | 4933 | 7.15 (7.01, 7.3) | NPX | Angiopoietin-1 receptor - Olink CVD2 | Uni-Prot ID: Q02763, OLINK ID: OID00398, Limit of de | Analyses made in EDTA plasma. Data is presented as normalized protein expression (NPX) values, ( | Blood Analysis | SCAPIS |
| Proteomics | TIMP4 | 5005 | 3.39 (3.17, 3.66) | NPX | Metalloproteinase inhibitor 4 - Olink CVD3 | Uni-Prot ID: Q99727, OLINK ID: OID00585, Limit of de | Analyses made in EDTA plasma. Data is presented as normalized protein expression (NPX) values, ( | Blood Analysis | SCAPIS |
| Proteomics | TLT_2 | 5006 | 5.07 (4.83, 5.32) | NPX | Trem-like transcript 2 protein - Olink CVD3 | Uni-Prot ID: Q5T2D2, OLINK ID: OID00588, Limit of de | Analyses made in EDTA plasma. Data is presented as normalized protein expression (NPX) values, ( | Blood Analysis | SCAPIS |
| Proteomics | TM | 4933 | 8.63 (8.44, 8.81) | NPX | Thrombomodulin - Olink CVD2 | Uni-Prot ID: P07204, OLINK ID: OID00428, Limit of de | Analyses made in EDTA plasma. Data is presented as normalized protein expression (NPX) values, ( | Blood Analysis | SCAPIS |
| Proteomics | TNFRSF10A | 4933 | 2.25 (2.08, 2.44) | NPX | Tumor necrosis factor receptor superfamily member 10A - Olink CVD2 | UNI-Prot ID: O00220, OLINK ID: OID00391, Limit of de | Analyses made in EDTA plasma. Data is presented as normalized protein expression (NPX) values, ( | Blood Analysis | SCAPIS |
| Proteomics | TNFRSF10C | 5006 | 6.17 (5.75, 6.51) | NPX | Tumor necrosis factor receptor superfamily member 10C - Olink CVD3 | Uni-Prot ID: O14798, OLINK ID: OID00594, Limit of de | Analyses made in EDTA plasma. Data is presented as normalized protein expression (NPX) values, ( | Blood Analysis | SCAPIS |
| Proteomics | TNFRSF11A | 4933 | 4.55 (4.34, 4.79) | NPX | Tumor necrosis factor receptor superfamily member 11A - Olink CVD2 | UNI-Prot ID: Q9Y6Q6, OLINK ID: OID00394, Limit of d | Analyses made in EDTA plasma. Data is presented as normalized protein expression (NPX) values, ( | Blood Analysis | SCAPIS |
| Proteomics | TNFRSF13B | 4933 | 7.84 (7.63, 8.07) | NPX | Tumor necrosis factor receptor superfamily member 13B - Olink CVD2 | Uni-Prot ID: O14836, OLINK ID: OID00461, Limit of de | Analyses made in EDTA plasma. Data is presented as normalized protein expression (NPX) values, ( | Blood Analysis | SCAPIS |
| Proteomics | TNFRSF14 | 5006 | 4.49 (4.31, 4.69) | NPX | Tumor necrosis factor receptor superfamily member 14 - Olink CVD3 | Uni-Prot ID: Q92956, OLINK ID: OID00563, Limit of de | Analyses made in EDTA plasma. Data is presented as normalized protein expression (NPX) values, ( | Blood Analysis | SCAPIS |
| Proteomics | TNFSF13B | 5006 | 7.21 (7.01, 7.42) | NPX | Tumor necrosis factor ligand superfamily member 13B - Olink CVD3 | Uni-Prot ID: Q9Y275, OLINK ID: OID00617, Limit of de | Analyses made in EDTA plasma. Data is presented as normalized protein expression (NPX) values, ( | Blood Analysis | SCAPIS |
| Proteomics | TNF_R1 | 5006 | 6.13 (5.93, 6.33) | NPX | Tumor necrosis factor receptor 1 - Olink CVD3 | Uni-Prot ID: P19438, OLINK ID: OID00649, Limit of de | Analyses made in EDTA plasma. Data is presented as normalized protein expression (NPX) values, ( | Blood Analysis | SCAPIS |
| Proteomics | TNF_R2 | 5006 | 5.15 (4.93, 5.36) | NPX | Tumor necrosis factor receptor 2 - Olink CVD3 | Uni-Prot ID: P20333, OLINK ID: OID00567, Limit of de | Analyses made in EDTA plasma. Data is presented as normalized protein expression (NPX) values, ( | Blood Analysis | SCAPIS |
| Proteomics | TR | 5006 | 4.73 (4.4, 5.07) | NPX | Transferrin receptor protein 1 - Olink CVD3 | Uni-Prot ID: P02786, OLINK ID: OID00593, Limit of de | Analyses made in EDTA plasma. Data is presented as normalized protein expression (NPX) values, ( | Blood Analysis | SCAPIS |
| Proteomics | TRAIL_R2 | 4933 | 4.27 (4.1, 4.45) | NPX | TNF-related apoptosis-inducing ligand receptor 2 - Olink CVD2 | Uni-Prot ID: O14763, OLINK ID: OID00396, Limit of de | Analyses made in EDTA plasma. Data is presented as normalized protein expression (NPX) values, ( | Blood Analysis | SCAPIS |
| Proteomics | TR_AP | 5006 | 3.98 (3.77, 4.2) | NPX | Tartrate-resistant acid phosphatase type 5 - Olink CVD3 | Uni-Prot ID: P13686, OLINK ID: OID00606, Limit of de | Analyses made in EDTA plasma. Data is presented as normalized protein expression (NPX) values, ( | Blood Analysis | SCAPIS |
| Proteomics | U_PAR | 5006 | 4.92 (4.73, 5.13) | NPX | Urokinase plasminogen activator surface receptor - Olink CVD3 | Uni-Prot ID: Q03405, OLINK ID: OID00620, Limit of de | Analyses made in EDTA plasma. Data is presented as normalized protein expression (NPX) values, ( | Blood Analysis | SCAPIS |
| Proteomics | VEGFD | 4933 | 6.44 (6.17, 6.72) | NPX | Vascular endothelial growth factor D - Olink CVD2 | Uni-Prot ID: O43915, OLINK ID: OID00468, Limit of de | Analyses made in EDTA plasma. Data is presented as normalized protein expression (NPX) values, ( | Blood Analysis | SCAPIS |
| Proteomics | VSIG2 | 4933 | 2.64 (2.4, 2.92) | NPX | V-set and immunoglobulin domain-containing protein 2 - Olink CVD2 | Uni-Prot ID: Q96IQ7, OLINK ID: OID00429, Limit of de | Analyses made in EDTA plasma. Data is presented as normalized protein expression (NPX) values, ( | Blood Analysis | SCAPIS |
| Proteomics | XCL1 | 4933 | 4.61 (4.32, 4.94) | NPX | Lymphotactin - Olink CVD2 | Uni-Prot ID: P47992, OLINK ID: OID00433, Limit of de | Analyses made in EDTA plasma. Data is presented as normalized protein expression (NPX) values, ( | Blood Analysis | SCAPIS |
| Proteomics | hOSCAR | 4933 | 9.51 (9.35, 9.66) | NPX | Osteoclast-associated immunoglobulin-like receptor - Olink CVD2 | Uni-Prot ID: Q8IYS5, OLINK ID: OID00460, Limit of det | Analyses made in EDTA plasma. Data is presented as normalized protein expression (NPX) values, ( | Blood Analysis | SCAPIS |
| Proteomics | t_PA | 5006 | 6.26 (5.85, 6.7) | NPX | Tissue-type plasminogen activator - Olink CVD3 | Uni-Prot ID: P00750, OLINK ID: OID00635, Limit of de | Analyses made in EDTA plasma. Data is presented as normalized protein expression (NPX) values, ( | Blood Analysis | SCAPIS |
| Proteomics | uPA | 5006 | 5.51 (5.32, 5.71) | NPX | Urokinase-type plasminogen activator - Olink CVD3 | Uni-Prot ID: P00749, OLINK ID: OID00631, Limit of de | Analyses made in EDTA plasma. Data is presented as normalized protein expression (NPX) values, ( | Blood Analysis | SCAPIS |
| Proteomics | vWF | 5006 | 6.07 (5.6, 6.56) | NPX | von Willebrand factor - Olink CVD3 | Uni-Prot ID: P04275, OLINK ID: OID00651, Limit of de | Analyses made in EDTA plasma. Data is presented as normalized protein expression (NPX) values, ( | Blood Analysis | SCAPIS |
| Metabolomics NMR | HDL2_C | 5042 | 0.85 (0.67, 1.04) | mmol/L | Total cholesterol in HDL2 | | Analyses made in EDTA plasma. A high-throughput NMR metabolomics platform (Nightingale) was | Blood Analysis | SCAPIS |
| Metabolomics NMR | HDL3_C | 5042 | 0.47 (0.45, 0.49) | mmol/L | Total cholesterol in HDL3 | | Analyses made in EDTA plasma. A high-throughput NMR metabolomics platform (Nightingale) was | Blood Analysis | SCAPIS |
| Metabolomics NMR | HDL_C | 5042 | 1.32 (1.13, 1.52) | mmol/L | Total cholesterol in HDL | | Analyses made in EDTA plasma. A high-throughput NMR metabolomics platform (Nightingale) was | Blood Analysis | SCAPIS |
| Metabolomics NMR | HDL_D | 5042 | 9.93 (9.77, 10.12) | nm | Mean diameter for HDL particles | | Analyses made in EDTA plasma. A high-throughput NMR metabolomics platform (Nightingale) was | Blood Analysis | SCAPIS |
| Metabolomics NMR | HDL_TG | 5042 | 0.12 (0.11, 0.14) | mmol/L | Triglycerides in HDL | | Analyses made in EDTA plasma. A high-throughput NMR metabolomics platform (Nightingale) was | Blood Analysis | SCAPIS |
| Metabolomics NMR | L_HDL_C | 5044 | 0.3 (0.2, 0.41) | mmol/L | Total cholesterol in large HDL | | Analyses made in EDTA plasma. A high-throughput NMR metabolomics platform (Nightingale) was | Blood Analysis | SCAPIS |
| Metabolomics NMR | L_HDL_CE | 5044 | 0.23 (0.16, 0.32) | mmol/L | Cholesterol esters in large HDL | | Analyses made in EDTA plasma. A high-throughput NMR metabolomics platform (Nightingale) was | Blood Analysis | SCAPIS |
| Metabolomics NMR | L_HDL_FC | 5044 | 0.06 (0.04, 0.09) | mmol/L | Free cholesterol in large HDL | | Analyses made in EDTA plasma. A high-throughput NMR metabolomics platform (Nightingale) was | Blood Analysis | SCAPIS |
| Metabolomics NMR | L_HDL_L | 5044 | 0.64 (0.45, 0.85) | mmol/L | Total lipids in large HDL | | Analyses made in EDTA plasma. A high-throughput NMR metabolomics platform (Nightingale) was | Blood Analysis | SCAPIS |
| Metabolomics NMR | L_HDL_P | 5044 | 0.0 (0.0, 0.0) | µmol/L | Concentration of large HDL particles | | Analyses made in EDTA plasma. A high-throughput NMR metabolomics platform (Nightingale) was | Blood Analysis | SCAPIS |
| Metabolomics NMR | L_HDL_PL | 5044 | 0.32 (0.23, 0.41) | µmol/L | Phospholipids in large HDL | | Analyses made in EDTA plasma. A high-throughput NMR metabolomics platform (Nightingale) was | Blood Analysis | SCAPIS |
| Metabolomics NMR | L_HDL_TG | 5044 | 0.02 (0.01, 0.03) | mmol/L | Triglycerides in large HDL | | Analyses made in EDTA plasma. A high-throughput NMR metabolomics platform (Nightingale) was | Blood Analysis | SCAPIS |
| Metabolomics NMR | M_HDL_C | 5044 | 0.38 (0.33, 0.42) | mmol/L | Total cholesterol in medium HDL | | Analyses made in EDTA plasma. A high-throughput NMR metabolomics platform (Nightingale) was | Blood Analysis | SCAPIS |
| Metabolomics NMR | M_HDL_CE | 5044 | 0.31 (0.27, 0.34) | mmol/L | Cholesterol esters in medium HDL | | Analyses made in EDTA plasma. A high-throughput NMR metabolomics platform (Nightingale) was | Blood Analysis | SCAPIS |
| Metabolomics NMR | M_HDL_FC | 5044 | 0.07 (0.06, 0.08) | mmol/L | Free cholesterol in medium HDL | | Analyses made in EDTA plasma. A high-throughput NMR metabolomics platform (Nightingale) was | Blood Analysis | SCAPIS |
| Metabolomics NMR | M_HDL_L | 5044 | 0.78 (0.71, 0.86) | mmol/L | Total lipids in medium HDL | | Analyses made in EDTA plasma. A high-throughput NMR metabolomics platform (Nightingale) was | Blood Analysis | SCAPIS |
| Metabolomics NMR | M_HDL_P | 5044 | 0.0 (0.0, 0.0) | µmol/L | Concentration of medium HDL particles | | Analyses made in EDTA plasma. A high-throughput NMR metabolomics platform (Nightingale) was | Blood Analysis | SCAPIS |
| Metabolomics NMR | M_HDL_PL | 5044 | 0.37 (0.33, 0.4) | µmol/L | Phospholipids in medium HDL | | Analyses made in EDTA plasma. A high-throughput NMR metabolomics platform (Nightingale) was | Blood Analysis | SCAPIS |
| Metabolomics NMR | M_HDL_TG | 5044 | 0.04 (0.03, 0.05) | mmol/L | Triglycerides in medium HDL | | Analyses made in EDTA plasma. A high-throughput NMR metabolomics platform (Nightingale) was | Blood Analysis | SCAPIS |
| Metabolomics NMR | S_HDL_C | 5044 | 0.42 (0.4, 0.45) | mmol/L | Total cholesterol in small HDL | | Analyses made in EDTA plasma. A high-throughput NMR metabolomics platform (Nightingale) was | Blood Analysis | SCAPIS |
| Metabolomics NMR | S_HDL_CE | 5044 | 0.32 (0.3, 0.35) | mmol/L | Cholesterol esters in small HDL | | Analyses made in EDTA plasma. A high-throughput NMR metabolomics platform (Nightingale) was | Blood Analysis | SCAPIS |
| Metabolomics NMR | S_HDL_FC | 5044 | 0.1 (0.09, 0.1) | mmol/L | Free cholesterol in small HDL | | Analyses made in EDTA plasma. A high-throughput NMR metabolomics platform (Nightingale) was | Blood Analysis | SCAPIS |
| Metabolomics NMR | S_HDL_L | 5044 | 1.03 (0.98, 1.09) | mmol/L | Total lipids in small HDL | | Analyses made in EDTA plasma. A high-throughput NMR metabolomics platform (Nightingale) was | Blood Analysis | SCAPIS |
| Metabolomics NMR | S_HDL_P | 5044 | 0.0 (0.0, 0.0) | µmol/L | Concentration of small HDL particles | | Analyses made in EDTA plasma. A high-throughput NMR metabolomics platform (Nightingale) was | Blood Analysis | SCAPIS |
| Metabolomics NMR | S_HDL_PL | 5044 | 0.56 (0.53, 0.6) | mmol/L | Phospholipids in small HDL | | Analyses made in EDTA plasma. A high-throughput NMR metabolomics platform (Nightingale) was | Blood Analysis | SCAPIS |
| Metabolomics NMR | S_HDL_TG | 5044 | 0.05 (0.04, 0.05) | mmol/L | Triglycerides in small HDL | | Analyses made in EDTA plasma. A high-throughput NMR metabolomics platform (Nightingale) was | Blood Analysis | SCAPIS |
| Metabolomics NMR | XL_HDL_C | 5044 | 0.21 (0.16, 0.27) | mmol/L | Total cholesterol in very large HDL | | Analyses made in EDTA plasma. A high-throughput NMR metabolomics platform (Nightingale) was | Blood Analysis | SCAPIS |
| Metabolomics NMR | XL_HDL_CE | 5044 | 0.15 (0.12, 0.2) | mmol/L | Cholesterol esters in very large HDL | | Analyses made in EDTA plasma. A high-throughput NMR metabolomics platform (Nightingale) was | Blood Analysis | SCAPIS |
| Metabolomics NMR | XL_HDL_FC | 5044 | 0.06 (0.04, 0.08) | mmol/L | Free cholesterol in very large HDL | | Analyses made in EDTA plasma. A high-throughput NMR metabolomics platform (Nightingale) was | Blood Analysis | SCAPIS |
| Metabolomics NMR | XL_HDL_L | 5044 | 0.43 (0.32, 0.58) | mmol/L | Total lipids in very large HDL | | Analyses made in EDTA plasma. A high-throughput NMR metabolomics platform (Nightingale) was | Blood Analysis | SCAPIS |
| Metabolomics NMR | XL_HDL_P | 5044 | 0.0 (0.0, 0.0) | µmol/L | Concentration of very large HDL particles | | Analyses made in EDTA plasma. A high-throughput NMR metabolomics platform (Nightingale) was | Blood Analysis | SCAPIS |
| Metabolomics NMR | XL_HDL_PL | 5044 | 0.21 (0.15, 0.29) | mmol/L | Phospholipids in very large HDL | | Analyses made in EDTA plasma. A high-throughput NMR metabolomics platform (Nightingale) was | Blood Analysis | SCAPIS |
| Metabolomics NMR | XL_HDL_TG | 5044 | 0.01 (0.01, 0.02) | mmol/L | Triglycerides in very large HDL | | Analyses made in EDTA plasma. A high-throughput NMR metabolomics platform (Nightingale) was | Blood Analysis | SCAPIS |
| Metabolomics NMR | IDL_C | 5044 | 0.65 (0.55, 0.74) | mmol/L | Total cholesterol in IDL | | Analyses made in EDTA plasma. A high-throughput NMR metabolomics platform (Nightingale) was | Blood Analysis | SCAPIS |
| Metabolomics NMR | IDL_CE | 5044 | 0.46 (0.39, 0.53) | mmol/L | Cholesterol esters in IDL | | Analyses made in EDTA plasma. A high-throughput NMR metabolomics platform (Nightingale) was | Blood Analysis | SCAPIS |
| Metabolomics NMR | IDL_FC | 5044 | 0.19 (0.16, 0.21) | mmol/L | Free cholesterol in IDL | | Analyses made in EDTA plasma. A high-throughput NMR metabolomics platform (Nightingale) was | Blood Analysis | SCAPIS |
| Metabolomics NMR | IDL_L | 5042 | 1.03 (0.89, 1.17) | mmol/L | Total lipids in IDL | | Analyses made in EDTA plasma. A high-throughput NMR metabolomics platform (Nightingale) was | Blood Analysis | SCAPIS |
| Metabolomics NMR | IDL_P | 5042 | 0.0 (0.0, 0.0) | µmol/L | Concentration of IDL particles | | Analyses made in EDTA plasma. A high-throughput NMR metabolomics platform (Nightingale) was | Blood Analysis | SCAPIS |
| Metabolomics NMR | IDL_PL | 5044 | 0.28 (0.25, 0.32) | mmol/L | Phospholipids in IDL | | Analyses made in EDTA plasma. A high-throughput NMR metabolomics platform (Nightingale) was | Blood Analysis | SCAPIS |
| Metabolomics NMR | IDL_TG | 5044 | 0.1 (0.09, 0.11) | mmol/L | Triglycerides in IDL | | Analyses made in EDTA plasma. A high-throughput NMR metabolomics platform (Nightingale) was | Blood Analysis | SCAPIS |
| Metabolomics NMR | LDL_C | 5042 | 1.52 (1.27, 1.78) | mmol/L | Total cholesterol in LDL | | Analyses made in EDTA plasma. A high-throughput NMR metabolomics platform (Nightingale) was | Blood Analysis | SCAPIS |
| Metabolomics NMR | LDL_D | 5042 | 23.56 (23.52, 23.61) | nm | Mean diameter for LDL particles | | Analyses made in EDTA plasma. A high-throughput NMR metabolomics platform (Nightingale) was | Blood Analysis | SCAPIS |
| Metabolomics NMR | LDL_TG | 5042 | 0.16 (0.14, 0.18) | mmol/L | Triglycerides in LDL | | Analyses made in EDTA plasma. A high-throughput NMR metabolomics platform (Nightingale) was | Blood Analysis | SCAPIS |
| Metabolomics NMR | L_LDL_C | 5044 | 0.8 (0.67, 0.93) | mmol/L | Total cholesterol in large LDL | | Analyses made in EDTA plasma. A high-throughput NMR metabolomics platform (Nightingale) was | Blood Analysis | SCAPIS |
| Metabolomics NMR | L_LDL_CE | 5044 | 0.57 (0.47, 0.67) | mmol/L | Cholesterol esters in large LDL | | Analyses made in EDTA plasma. A high-throughput NMR metabolomics platform (Nightingale) was | Blood Analysis | SCAPIS |
| Metabolomics NMR | L_LDL_FC | 5044 | 0.23 (0.2, 0.26) | mmol/L | Free cholesterol in large LDL | | Analyses made in EDTA plasma. A high-throughput NMR metabolomics platform (Nightingale) was | Blood Analysis | SCAPIS |
| Metabolomics NMR | L_LDL_L | 5044 | 1.19 (1.02, 1.37) | mmol/L | Total lipids in large LDL | | Analyses made in EDTA plasma. A high-throughput NMR metabolomics platform (Nightingale) was | Blood Analysis | SCAPIS |
| Metabolomics NMR | L_LDL_P | 5044 | 0.0 (0.0, 0.0) | µmol/L | Concentration of large LDL particles | | Analyses made in EDTA plasma. A high-throughput NMR metabolomics platform (Nightingale) was | Blood Analysis | SCAPIS |
| Metabolomics NMR | L_LDL_PL | 5044 | 0.3 (0.27, 0.34) | mmol/L | Phospholipids in large LDL | | Analyses made in EDTA plasma. A high-throughput NMR metabolomics platform (Nightingale) was | Blood Analysis | SCAPIS |
| Metabolomics NMR | L_LDL_TG | 5044 | 0.09 (0.08, 0.1) | mmol/L | Triglycerides in large LDL | | Analyses made in EDTA plasma. A high-throughput NMR metabolomics platform (Nightingale) was | Blood Analysis | SCAPIS |
| Metabolomics NMR | M_LDL_C | 5044 | 0.45 (0.37, 0.53) | mmol/L | Total cholesterol in medium LDL | | Analyses made in EDTA plasma. A high-throughput NMR metabolomics platform (Nightingale) was | Blood Analysis | SCAPIS |
| Metabolomics NMR | M_LDL_CE | 5044 | 0.32 (0.25, 0.39) | mmol/L | Cholesterol esters in medium LDL | | Analyses made in EDTA plasma. A high-throughput NMR metabolomics platform (Nightingale) was | Blood Analysis | SCAPIS |
| Metabolomics NMR | M_LDL_FC | 5044 | 0.13 (0.12, 0.15) | mmol/L | Free cholesterol in medium LDL | | Analyses made in EDTA plasma. A high-throughput NMR metabolomics platform (Nightingale) was | Blood Analysis | SCAPIS |
| Metabolomics NMR | M_LDL_L | 5044 | 0.68 (0.58, 0.79) | mmol/L | Total lipids in medium LDL | | Analyses made in EDTA plasma. A high-throughput NMR metabolomics platform (Nightingale) was | Blood Analysis | SCAPIS |
| Metabolomics NMR | M_LDL_P | 5044 | 0.0 (0.0, 0.0) | µmol/L | Concentration of medium LDL particles | | Analyses made in EDTA plasma. A high-throughput NMR metabolomics platform (Nightingale) was | Blood Analysis | SCAPIS |
| Metabolomics NMR | M_LDL_PL | 5044 | 0.19 (0.17, 0.21) | mmol/L | Phospholipids in medium LDL | | Analyses made in EDTA plasma. A high-throughput NMR metabolomics platform (Nightingale) was | Blood Analysis | SCAPIS |
| Metabolomics NMR | M_LDL_TG | 5044 | 0.04 (0.04, 0.05) | mmol/L | Triglycerides in medium LDL | | Analyses made in EDTA plasma. A high-throughput NMR metabolomics platform (Nightingale) was | Blood Analysis | SCAPIS |
| Metabolomics NMR | S_LDL_C | 5044 | 0.27 (0.23, 0.32) | mmol/L | Total cholesterol in small LDL | | Analyses made in EDTA plasma. A high-throughput NMR metabolomics platform (Nightingale) was | Blood Analysis | SCAPIS |
| Metabolomics NMR | S_LDL_CE | 5044 | 0.2 (0.16, 0.24) | mmol/L | Cholesterol esters in small LDL | | Analyses made in EDTA plasma. A high-throughput NMR metabolomics platform (Nightingale) was | Blood Analysis | SCAPIS |
| Metabolomics NMR | S_LDL_FC | 5044 | 0.08 (0.07, 0.09) | mmol/L | Free cholesterol in small LDL | | Analyses made in EDTA plasma. A high-throughput NMR metabolomics platform (Nightingale) was | Blood Analysis | SCAPIS |
| Metabolomics NMR | S_LDL_L | 5044 | 0.44 (0.37, 0.5) | mmol/L | Total lipids in small LDL | | Analyses made in EDTA plasma. A high-throughput NMR metabolomics platform (Nightingale) was | Blood Analysis | SCAPIS |
| Metabolomics NMR | S_LDL_P | 5044 | 0.0 (0.0, 0.0) | µmol/L | Concentration of small LDL particles | | Analyses made in EDTA plasma. A high-throughput NMR metabolomics platform (Nightingale) was | Blood Analysis | SCAPIS |
| Metabolomics NMR | S_LDL_PL | 5044 | 0.14 (0.12, 0.15) | mmol/L | Phospholipids in small LDL | | Analyses made in EDTA plasma. A high-throughput NMR metabolomics platform (Nightingale) was | Blood Analysis | SCAPIS |
| Metabolomics NMR | S_LDL_TG | 5044 | 0.03 (0.02, 0.03) | mmol/L | Triglycerides in small LDL | | Analyses made in EDTA plasma. A high-throughput NMR metabolomics platform (Nightingale) was | Blood Analysis | SCAPIS |
| Metabolomics NMR | AcAce | 5044 | 0.03 (0.02, 0.04) | mmol/L | Acetoacetate | | Analyses made in EDTA plasma. A high-throughput NMR metabolomics platform (Nightingale) was | Blood Analysis | SCAPIS |
| Metabolomics NMR | Ace | 5043 | 0.04 (0.04, 0.05) | mmol/L | Acetate | | Analyses made in EDTA plasma. A high-throughput NMR metabolomics platform (Nightingale) was | Blood Analysis | SCAPIS |
| Metabolomics NMR | Ala | 5044 | 0.33 (0.3, 0.36) | mmol/L | Alanine | | Analyses made in EDTA plasma. A high-throughput NMR metabolomics platform (Nightingale) was | Blood Analysis | SCAPIS |
| Metabolomics NMR | Alb | 5042 | 0.08 (0.08, 0.08) | signal area | Albumin | | Analyses made in EDTA plasma. A high-throughput NMR metabolomics platform (Nightingale) was | Blood Analysis | SCAPIS |

**Table S2: Summary of clinical, molecular, and imaging-derived variables in SCAPIS, and disease outcomes in UKBB**

| Category | Variable_Name | Data_Available | Summary | Unit | Heading | Context | Method | Source | Cohort |
|---|---|---|---|---|---|---|---|---|---|
| Metabolomics NMR | Cit | 5044 | 0.13 (0.12, 0.15) | mmol/L | Citrate | | Analyses made in EDTA plasma. A high-throughput NMR metabolomics platform (Nightingale) was | Blood Analysis | SCAPIS |
| Metabolomics NMR | Crea | 5034 | 0.06 (0.06, 0.07) | mmol/L | Creatinine | | Analyses made in EDTA plasma. A high-throughput NMR metabolomics platform (Nightingale) was | Blood Analysis | SCAPIS |
| Metabolomics NMR | DHA | 4994 | 0.12 (0.1, 0.14) | mmol/L | docosahexaenoic acid | | Analyses made in EDTA plasma. A high-throughput NMR metabolomics platform (Nightingale) was | Blood Analysis | SCAPIS |
| Metabolomics NMR | EstC | 4994 | 2.85 (2.52, 3.19) | mmol/L | Esterified cholesterol | | Analyses made in EDTA plasma. A high-throughput NMR metabolomics platform (Nightingale) was | Blood Analysis | SCAPIS |
| Metabolomics NMR | FAw3 | 4994 | 0.36 (0.29, 0.43) | mmol/L | Omega-3 fatty acids | | Analyses made in EDTA plasma. A high-throughput NMR metabolomics platform (Nightingale) was | Blood Analysis | SCAPIS |
| Metabolomics NMR | FAw6 | 4994 | 3.24 (2.94, 3.57) | mmol/L | Omega-6 fatty acids | | Analyses made in EDTA plasma. A high-throughput NMR metabolomics platform (Nightingale) was | Blood Analysis | SCAPIS |
| Metabolomics NMR | FreeC | 4994 | 1.24 (1.11, 1.37) | mmol/L | Free cholesterol | | Analyses made in EDTA plasma. A high-throughput NMR metabolomics platform (Nightingale) was | Blood Analysis | SCAPIS |
| Metabolomics NMR | Gln | 5039 | 0.44 (0.41, 0.48) | mmol/L | Glutamine | | Analyses made in EDTA plasma. A high-throughput NMR metabolomics platform (Nightingale) was | Blood Analysis | SCAPIS |
| Metabolomics NMR | Gp | 5043 | 1.04 (0.96, 1.13) | mmol/L | Glycoprotein acetyls, mainly a1-acid glycoprotein | | Analyses made in EDTA plasma. A high-throughput NMR metabolomics platform (Nightingale) was | Blood Analysis | SCAPIS |
| Metabolomics NMR | His | 5035 | 0.06 (0.06, 0.06) | mmol/L | Histidine | | Analyses made in EDTA plasma. A high-throughput NMR metabolomics platform (Nightingale) was | Blood Analysis | SCAPIS |
| Metabolomics NMR | Ile | 5044 | 0.04 (0.03, 0.05) | mmol/L | Isoleucine | | Analyses made in EDTA plasma. A high-throughput NMR metabolomics platform (Nightingale) was | Blood Analysis | SCAPIS |
| Metabolomics NMR | LA | 4994 | 2.59 (2.32, 2.88) | mmol/L | linoleic acid | | Analyses made in EDTA plasma. A high-throughput NMR metabolomics platform (Nightingale) was | Blood Analysis | SCAPIS |
| Metabolomics NMR | Lac | 5044 | 0.99 (0.85, 1.18) | mmol/L | Lactate | | Analyses made in EDTA plasma. A high-throughput NMR metabolomics platform (Nightingale) was | Blood Analysis | SCAPIS |
| Metabolomics NMR | Leu | 5043 | 0.06 (0.05, 0.07) | mmol/L | Leucine | | Analyses made in EDTA plasma. A high-throughput NMR metabolomics platform (Nightingale) was | Blood Analysis | SCAPIS |
| Metabolomics NMR | MUFA | 4994 | 2.79 (2.49, 3.18) | mmol/L | Monounsaturated fatty acids | | Analyses made in EDTA plasma. A high-throughput NMR metabolomics platform (Nightingale) was | Blood Analysis | SCAPIS |
| Metabolomics NMR | PC | 4994 | 1.76 (1.61, 1.91) | mmol/L | Phosphatidylcholine and other cholines | | Analyses made in EDTA plasma. A high-throughput NMR metabolomics platform (Nightingale) was | Blood Analysis | SCAPIS |
| Metabolomics NMR | PUFA | 4994 | 3.6 (3.25, 3.98) | mmol/L | Polyunsaturated fatty acids | | Analyses made in EDTA plasma. A high-throughput NMR metabolomics platform (Nightingale) was | Blood Analysis | SCAPIS |
| Metabolomics NMR | Phe | 5043 | 0.05 (0.05, 0.06) | mmol/L | Phenylalanine | | Analyses made in EDTA plasma. A high-throughput NMR metabolomics platform (Nightingale) was | Blood Analysis | SCAPIS |
| Metabolomics NMR | Remnant_C | 5042 | 1.21 (1.01, 1.42) | mmol/L | Remnant cholesterol (non-HDL, non-LDL -cholesterol) | | Analyses made in EDTA plasma. A high-throughput NMR metabolomics platform (Nightingale) was | Blood Analysis | SCAPIS |
| Metabolomics NMR | SFA | 4994 | 3.89 (3.53, 4.3) | mmol/L | Saturated fatty acids | | Analyses made in EDTA plasma. A high-throughput NMR metabolomics platform (Nightingale) was | Blood Analysis | SCAPIS |
| Metabolomics NMR | SM | 4994 | 0.44 (0.4, 0.48) | mmol/L | Sphingomyelins | | Analyses made in EDTA plasma. A high-throughput NMR metabolomics platform (Nightingale) was | Blood Analysis | SCAPIS |
| Metabolomics NMR | Serum_C | 5042 | 4.09 (3.62, 4.55) | mmol/L | Serum total cholesterol | | Analyses made in EDTA plasma. A high-throughput NMR metabolomics platform (Nightingale) was | Blood Analysis | SCAPIS |
| Metabolomics NMR | TotCho | 4994 | 2.19 (2.02, 2.36) | mmol/L | Total cholines | | Analyses made in EDTA plasma. A high-throughput NMR metabolomics platform (Nightingale) was | Blood Analysis | SCAPIS |
| Metabolomics NMR | TotFA | 4994 | 10.29 (9.34, 11.41) | mmol/L | Total fatty acids | | Analyses made in EDTA plasma. A high-throughput NMR metabolomics platform (Nightingale) was | Blood Analysis | SCAPIS |
| Metabolomics NMR | TotPG | 4994 | 1.79 (1.63, 1.95) | mmol/L | Total phosphoglycerides | | Analyses made in EDTA plasma. A high-throughput NMR metabolomics platform (Nightingale) was | Blood Analysis | SCAPIS |
| Metabolomics NMR | Tyr | 5036 | 0.05 (0.04, 0.06) | mmol/L | Tyrosine | | Analyses made in EDTA plasma. A high-throughput NMR metabolomics platform (Nightingale) was | Blood Analysis | SCAPIS |
| Metabolomics NMR | UnSat | 4994 | 1.16 (1.13, 1.19) | | Estimated degree of unsaturation | | Analyses made in EDTA plasma. A high-throughput NMR metabolomics platform (Nightingale) was | Blood Analysis | SCAPIS |
| Metabolomics NMR | Val | 5043 | 0.15 (0.13, 0.17) | mmol/L | Valine | | Analyses made in EDTA plasma. A high-throughput NMR metabolomics platform (Nightingale) was | Blood Analysis | SCAPIS |
| Metabolomics NMR | bOHBut | 5036 | 0.14 (0.11, 0.18) | mmol/L | 3-hydroxybutyrate | | Analyses made in EDTA plasma. A high-throughput NMR metabolomics platform (Nightingale) was | Blood Analysis | SCAPIS |
| Metabolomics NMR | L_VLDL_C | 5044 | 0.04 (0.02, 0.07) | mmol/L | Total cholesterol in large VLDL | | Analyses made in EDTA plasma. A high-throughput NMR metabolomics platform (Nightingale) was | Blood Analysis | SCAPIS |
| Metabolomics NMR | L_VLDL_CE | 5044 | 0.03 (0.02, 0.04) | mmol/L | Cholesterol esters in large VLDL | | Analyses made in EDTA plasma. A high-throughput NMR metabolomics platform (Nightingale) was | Blood Analysis | SCAPIS |
| Metabolomics NMR | L_VLDL_FC | 5044 | 0.01 (0.01, 0.03) | mmol/L | Free cholesterol in large VLDL | | Analyses made in EDTA plasma. A high-throughput NMR metabolomics platform (Nightingale) was | Blood Analysis | SCAPIS |
| Metabolomics NMR | L_VLDL_L | 5044 | 0.18 (0.11, 0.29) | mmol/L | Total lipids in large VLDL | | Analyses made in EDTA plasma. A high-throughput NMR metabolomics platform (Nightingale) was | Blood Analysis | SCAPIS |
| Metabolomics NMR | L_VLDL_P | 5044 | 0.0 (0.0, 0.0) | nmol/L | Concentration of large VLDL particles | | Analyses made in EDTA plasma. A high-throughput NMR metabolomics platform (Nightingale) was | Blood Analysis | SCAPIS |
| Metabolomics NMR | L_VLDL_PL | 5044 | 0.03 (0.02, 0.05) | mmol/L | Phospholipids in large VLDL | | Analyses made in EDTA plasma. A high-throughput NMR metabolomics platform (Nightingale) was | Blood Analysis | SCAPIS |
| Metabolomics NMR | L_VLDL_TG | 5044 | 0.11 (0.07, 0.17) | mmol/L | Triglycerides in large VLDL | | Analyses made in EDTA plasma. A high-throughput NMR metabolomics platform (Nightingale) was | Blood Analysis | SCAPIS |
| Metabolomics NMR | M_VLDL_C | 5044 | 0.12 (0.08, 0.16) | mmol/L | Total cholesterol in medium VLDL | | Analyses made in EDTA plasma. A high-throughput NMR metabolomics platform (Nightingale) was | Blood Analysis | SCAPIS |
| Metabolomics NMR | M_VLDL_CE | 5044 | 0.07 (0.05, 0.09) | mmol/L | Cholesterol esters in medium VLDL | | Analyses made in EDTA plasma. A high-throughput NMR metabolomics platform (Nightingale) was | Blood Analysis | SCAPIS |
| Metabolomics NMR | M_VLDL_FC | 5044 | 0.05 (0.03, 0.07) | mmol/L | Free cholesterol in medium VLDL | | Analyses made in EDTA plasma. A high-throughput NMR metabolomics platform (Nightingale) was | Blood Analysis | SCAPIS |
| Metabolomics NMR | M_VLDL_L | 5044 | 0.44 (0.32, 0.61) | mmol/L | Total lipids in medium VLDL | | Analyses made in EDTA plasma. A high-throughput NMR metabolomics platform (Nightingale) was | Blood Analysis | SCAPIS |
| Metabolomics NMR | M_VLDL_P | 5044 | 0.0 (0.0, 0.0) | µmol/L | Concentration of medium VLDL particles | | Analyses made in EDTA plasma. A high-throughput NMR metabolomics platform (Nightingale) was | Blood Analysis | SCAPIS |
| Metabolomics NMR | M_VLDL_PL | 5044 | 0.09 (0.07, 0.12) | mmol/L | Phospholipids in medium VLDL | | Analyses made in EDTA plasma. A high-throughput NMR metabolomics platform (Nightingale) was | Blood Analysis | SCAPIS |
| Metabolomics NMR | M_VLDL_TG | 5044 | 0.23 (0.17, 0.33) | mmol/L | Triglycerides in medium VLDL | | Analyses made in EDTA plasma. A high-throughput NMR metabolomics platform (Nightingale) was | Blood Analysis | SCAPIS |
| Metabolomics NMR | S_VLDL_C | 5044 | 0.16 (0.13, 0.2) | mmol/L | Total cholesterol in small VLDL | | Analyses made in EDTA plasma. A high-throughput NMR metabolomics platform (Nightingale) was | Blood Analysis | SCAPIS |
| Metabolomics NMR | S_VLDL_CE | 5044 | 0.1 (0.08, 0.13) | mmol/L | Cholesterol esters in small VLDL | | Analyses made in EDTA plasma. A high-throughput NMR metabolomics platform (Nightingale) was | Blood Analysis | SCAPIS |
| Metabolomics NMR | S_VLDL_FC | 5044 | 0.06 (0.05, 0.08) | mmol/L | Free cholesterol in small VLDL | | Analyses made in EDTA plasma. A high-throughput NMR metabolomics platform (Nightingale) was | Blood Analysis | SCAPIS |
| Metabolomics NMR | S_VLDL_L | 5044 | 0.48 (0.38, 0.59) | mmol/L | Total lipids in small VLDL | | Analyses made in EDTA plasma. A high-throughput NMR metabolomics platform (Nightingale) was | Blood Analysis | SCAPIS |
| Metabolomics NMR | S_VLDL_P | 5044 | 0.0 (0.0, 0.0) | µmol/L | Concentration of small VLDL particles | | Analyses made in EDTA plasma. A high-throughput NMR metabolomics platform (Nightingale) was | Blood Analysis | SCAPIS |
| Metabolomics NMR | S_VLDL_PL | 5044 | 0.12 (0.1, 0.14) | mmol/L | Phospholipids in small VLDL | | Analyses made in EDTA plasma. A high-throughput NMR metabolomics platform (Nightingale) was | Blood Analysis | SCAPIS |
| Metabolomics NMR | S_VLDL_TG | 5044 | 0.2 (0.16, 0.25) | mmol/L | Triglycerides in small VLDL | | Analyses made in EDTA plasma. A high-throughput NMR metabolomics platform (Nightingale) was | Blood Analysis | SCAPIS |
| Metabolomics NMR | Serum_TG | 5042 | 1.06 (0.86, 1.35) | mmol/L | Serum total triglycerides | | Analyses made in EDTA plasma. A high-throughput NMR metabolomics platform (Nightingale) was | Blood Analysis | SCAPIS |
| Metabolomics NMR | VLDL_C | 5042 | 0.56 (0.44, 0.69) | mmol/L | Total cholesterol in VLDL | | Analyses made in EDTA plasma. A high-throughput NMR metabolomics platform (Nightingale) was | Blood Analysis | SCAPIS |
| Metabolomics NMR | VLDL_D | 5042 | 36.45 (35.76, 37.28) | nm | Mean diameter for VLDL particles | | Analyses made in EDTA plasma. A high-throughput NMR metabolomics platform (Nightingale) was | Blood Analysis | SCAPIS |
| Metabolomics NMR | VLDL_TG | 5042 | 0.68 (0.5, 0.94) | mmol/L | Triglycerides in VLDL | | Analyses made in EDTA plasma. A high-throughput NMR metabolomics platform (Nightingale) was | Blood Analysis | SCAPIS |
| Metabolomics NMR | XL_VLDL_C | 5044 | 0.01 (0.0, 0.01) | mmol/L | Total cholesterol in very large VLDL | | Analyses made in EDTA plasma. A high-throughput NMR metabolomics platform (Nightingale) was | Blood Analysis | SCAPIS |
| Metabolomics NMR | XL_VLDL_CE | 5044 | 0.01 (0.0, 0.01) | mmol/L | Cholesterol esters in very large VLDL | | Analyses made in EDTA plasma. A high-throughput NMR metabolomics platform (Nightingale) was | Blood Analysis | SCAPIS |
| Metabolomics NMR | XL_VLDL_FC | 5044 | 0.0 (0.0, 0.01) | mmol/L | Free cholesterol in very large VLDL | | Analyses made in EDTA plasma. A high-throughput NMR metabolomics platform (Nightingale) was | Blood Analysis | SCAPIS |
| Metabolomics NMR | XL_VLDL_L | 5044 | 0.04 (0.02, 0.07) | mmol/L | Total lipids in very large VLDL | | Analyses made in EDTA plasma. A high-throughput NMR metabolomics platform (Nightingale) was | Blood Analysis | SCAPIS |
| Metabolomics NMR | XL_VLDL_P | 5044 | 0.0 (0.0, 0.0) | nmol/L | Concentration of very large VLDL particles | | Analyses made in EDTA plasma. A high-throughput NMR metabolomics platform (Nightingale) was | Blood Analysis | SCAPIS |
| Metabolomics NMR | XL_VLDL_PL | 5044 | 0.01 (0.0, 0.01) | mmol/L | Phospholipids in very large VLDL | | Analyses made in EDTA plasma. A high-throughput NMR metabolomics platform (Nightingale) was | Blood Analysis | SCAPIS |
| Metabolomics NMR | XL_VLDL_TG | 5044 | 0.03 (0.02, 0.05) | mmol/L | Triglycerides in very large VLDL | | Analyses made in EDTA plasma. A high-throughput NMR metabolomics platform (Nightingale) was | Blood Analysis | SCAPIS |
| Metabolomics NMR | XS_VLDL_C | 5044 | 0.22 (0.19, 0.25) | mmol/L | Total cholesterol in very small VLDL | | Analyses made in EDTA plasma. A high-throughput NMR metabolomics platform (Nightingale) was | Blood Analysis | SCAPIS |
| Metabolomics NMR | XS_VLDL_CE | 5044 | 0.15 (0.13, 0.17) | mmol/L | Cholesterol esters in very small VLDL | | Analyses made in EDTA plasma. A high-throughput NMR metabolomics platform (Nightingale) was | Blood Analysis | SCAPIS |
| Metabolomics NMR | XS_VLDL_FC | 5044 | 0.07 (0.06, 0.08) | mmol/L | Free cholesterol in very small VLDL | | Analyses made in EDTA plasma. A high-throughput NMR metabolomics platform (Nightingale) was | Blood Analysis | SCAPIS |
| Metabolomics NMR | XS_VLDL_L | 5044 | 0.45 (0.39, 0.51) | mmol/L | Total lipids in very small VLDL | | Analyses made in EDTA plasma. A high-throughput NMR metabolomics platform (Nightingale) was | Blood Analysis | SCAPIS |
| Metabolomics NMR | XS_VLDL_P | 5044 | 0.0 (0.0, 0.0) | µmol/L | Concentration of very small VLDL particles | | Analyses made in EDTA plasma. A high-throughput NMR metabolomics platform (Nightingale) was | Blood Analysis | SCAPIS |
| Metabolomics NMR | XS_VLDL_PL | 5044 | 0.14 (0.12, 0.16) | mmol/L | Phospholipids in very small VLDL | | Analyses made in EDTA plasma. A high-throughput NMR metabolomics platform (Nightingale) was | Blood Analysis | SCAPIS |
| Metabolomics NMR | XS_VLDL_TG | 5044 | 0.09 (0.08, 0.11) | mmol/L | Triglycerides in very small VLDL | | Analyses made in EDTA plasma. A high-throughput NMR metabolomics platform (Nightingale) was | Blood Analysis | SCAPIS |
| Metabolomics NMR | XXL_VLDL_C | 5044 | 0.0 (0.0, 0.01) | mmol/L | Total cholesterol in chylomicrons and extremely large VLDL | | Analyses made in EDTA plasma. A high-throughput NMR metabolomics platform (Nightingale) was | Blood Analysis | SCAPIS |
| Metabolomics NMR | XXL_VLDL_CE | 5044 | 0.0 (0.0, 0.0) | mmol/L | Cholesterol esters in chylomicrons and extremely large VLDL | | Analyses made in EDTA plasma. A high-throughput NMR metabolomics platform (Nightingale) was | Blood Analysis | SCAPIS |
| Metabolomics NMR | XXL_VLDL_FC | 5044 | 0.0 (0.0, 0.0) | mmol/L | Free cholesterol in chylomicrons and extremely large VLDL | | Analyses made in EDTA plasma. A high-throughput NMR metabolomics platform (Nightingale) was | Blood Analysis | SCAPIS |
| Metabolomics NMR | XXL_VLDL_L | 5044 | 0.02 (0.02, 0.03) | mmol/L | Total lipids in chylomicrons and extremely large VLDL | | Analyses made in EDTA plasma. A high-throughput NMR metabolomics platform (Nightingale) was | Blood Analysis | SCAPIS |
| Metabolomics NMR | XXL_VLDL_P | 5044 | 0.0 (0.0, 0.0) | nmol/L | Concentration of chylomicrons and extremely large VLDL particles | | Analyses made in EDTA plasma. A high-throughput NMR metabolomics platform (Nightingale) was | Blood Analysis | SCAPIS |
| Metabolomics NMR | XXL_VLDL_PL | 5044 | 0.0 (0.0, 0.0) | mmol/L | Phospholipids in chylomicrons and extremely large VLDL | | Analyses made in EDTA plasma. A high-throughput NMR metabolomics platform (Nightingale) was | Blood Analysis | SCAPIS |
| Metabolomics NMR | XXL_VLDL_TG | 5044 | 0.02 (0.01, 0.03) | mmol/L | Triglycerides in chylomicrons and extremely large VLDL | | Analyses made in EDTA plasma. A high-throughput NMR metabolomics platform (Nightingale) was | Blood Analysis | SCAPIS |
| Calculated Metabolomics NMR | HDL_P | 5044 | 0.0 (0.0, 0.0) | µmol/L | Concentration of HDL particles | | XL_HDL_P + L_HDL_P + M_HDL_P + S_HDL_P | Derived | SCAPIS |
| Calculated Metabolomics NMR | HDL_PL | 5044 | 1.47 (1.29, 1.66) | mmol/L | Phospholipids in HDL | | XL_HDL_PL + L_HDL_PL + M_HDL_PL + S_HDL_PL | Derived | SCAPIS |
| Calculated Metabolomics NMR | VLDL_P | 5044 | 0.0 (0.0, 0.0) | µmol/L | Concentration of VLDL particles | | XXL_VLDL_P + XL_VLDL_P + L_VLDL_P + M_VLDL_P + S_VLDL_P + XS_VLDL_P | Derived | SCAPIS |
| Calculated Metabolomics NMR | VLDL_PL | 5044 | 0.39 (0.32, 0.48) | mmol/L | Phospholipids in VLDL | | XXL_VLDL_PL + XL_VLDL_PL + L_VLDL_PL + M_VLDL_PL + S_VLDL_PL + XS_VLDL_PL | Derived | SCAPIS |
| Calculated Metabolomics NMR | LDL_P | 5044 | 0.0 (0.0, 0.0) | µmol/L | Concentration of LDL particles | | L_LDL_P + M_LDL_P + S_LDL_P | Derived | SCAPIS |
| Calculated Metabolomics NMR | LDL_PL | 5044 | 0.63 (0.56, 0.7) | mmol/L | Phospholipids in LDL | | L_LDL_PL + M_LDL_PL + S_LDL_PL | Derived | SCAPIS |
| Calculated Metabolomics NMR | Serum_P | 5042 | 0.0 (0.0, 0.0) | µmol/L | Serum total particles | | VLDL_P + IDL_P + LDL_P + HDL_P | Derived | SCAPIS |
| Calculated Metabolomics NMR | Serum_PL | 5044 | 2.81 (2.59, 3.04) | mmol/L | Serum total phospholipids | | VLDL_PL + IDL_PL + LDL_PL + HDL_PL | Derived | SCAPIS |
| Imaging - CT Cardiac | Above50_Any | 26874 | NO: 25303, 94.15 %, YES: 1571, 5.85 % | | Any stenosis >50% in any coronary segment | | Based on the visual assessement of coronary arteries in the CCTA images. This variable is calculate | Imaging | SCAPIS |
| Imaging - CT Cardiac | AortaGt4cm | 28776 | NO: 27747, 96.42 %, YES: 1029, 3.58 % | | Aortic diameter >4 cm | | | Imaging | SCAPIS |
| Imaging - CT Cardiac | AorticValveCalc | 28679 | NO: 26598, 92.74 %, YES: 2081, 7.26 % | | Aortic valve calcification | | | Imaging | SCAPIS |
| Imaging - CT Cardiac | Atherosclerosis_Any | 26874 | NO: 15360, 57.16 %, YES: 11514, 42.84 % | | Atherosclerosis in any coronary segment | | Based on the visual assessement of coronary arteries in the CCTA images. This variable is calculate | Imaging | SCAPIS |
| Imaging - CT Cardiac | Bicuspid | 26769 | NO: 26631, 99.48 %, YES: 138, 0.52 % | | Bicuspid aortic valve | | | Imaging | SCAPIS |
| Imaging - CT Cardiac | SCCTStenosis_Any | 24811 | NO: 13443, 54.18 %, YES: 11368, 45.82 % | | Stenosis exists in any positions of segment SCCT S01-18 | Main variable describing outcome of visual assesemen | Visual assesement by individual readers. Stenosis: Degree of luminal obstruction was judged visual | Imaging | SCAPIS |
| Imaging - Carotid Ultrasound | carotidplaque | 29187 | YES: 16100, 55.16 %, NO: 13087, 44.84 % | | Presence of any carotid artery plaque | | Based on initial on-site analysis of carotid ultrasound imaging of the carotid arteries. Derived for s | Imaging | SCAPIS |
| Imaging - CT Cardiac | CACS_1 | 28261 | 0.0: 20172, 71.38 %, 1.0: 8089, 28.62 % | | The first dimension of categorical summed coronary artery calcium score (cacs). | The summed Coronary Artery Calcium Score (CACS) is | Reference: Grundy, S. M. et al. 2018 AHA/ACC/AACVPR/AAPA/ABC/ACPM/ADA/AGS/APhA/ASPC/ | Imaging | SCAPIS |
| Imaging - CT Cardiac | CACS_2 | 28261 | 0.0: 26234, 92.83 %, 1.0: 2027, 7.17 % | | The second dimension of categorical summed coronary artery calcium score (cacs). | The summed Coronary Artery Calcium Score (CACS) is | Reference: Grundy, S. M. et al. 2018 AHA/ACC/AACVPR/AAPA/ABC/ACPM/ADA/AGS/APhA/ASPC/ | Imaging | SCAPIS |
| Imaging - CT Cardiac | CACS_3 | 28261 | 0.0: 26799, 94.83 %, 1.0: 1462, 5.17 % | | The third dimension of categorical summed coronary artery calcium score (cacs). | The summed Coronary Artery Calcium Score (CACS) is | Reference: Grundy, S. M. et al. 2018 AHA/ACC/AACVPR/AAPA/ABC/ACPM/ADA/AGS/APhA/ASPC/ | Imaging | SCAPIS |
| Risk scores - imaging | CACS_Tot | 28261 | 0.0 (0.0, 21.0) | | Summed coronary artery calcium score | The summed Coronary Artery Calcium Score (CACS) a | The non-contrast CACS scan (120 kV) was planned using the lung image. Two different CACS proto | Imaging | SCAPIS |
| Risk scores - imaging | DukeIndex | 26874 | 0.0 (0.0, 2.0) | | Duke prognostic CAD index modified for SCAPIS | | 0= (No atherosclerosis) \| 1= (One stenosis <50% (Low-grade stenosis>=1)) \| 2= (Two stenoses <50% | Imaging | SCAPIS |
| Risk scores - imaging | SIS | 26874 | 0.0 (0.0, 2.0) | Number of segments | Segment Involvement Score - total number of coronary segments with any atherosclerosis | | Defined as the number of coronary segments with any atherosclerosis. A segment is defined as hav | Imaging | SCAPIS |
| Risk scores - clinic | eGFR | 29401 | 76.95 (70.81, 83.32) | mL/min/1,73m2 | Estimated glomerular filtration rate (eGFR) was calculated using the revised Lund–Malmö Study equation without adjustment for body weight, referred t | | Revised Lund-Malmö Study equation without body weight measure (LM Revised; pCr in #mol/L): e | Derived | SCAPIS |
| Risk scores - clinic | SCORE2 | 21467 | 5.0 (3.0, 7.0) | | Systematic coronary risk evaluation score 2 (SCORE2) | | Score2 was computed based on the age, sex, current smoking status, systolic blood pressure, and r | Derived | SCAPIS |
| Clinical Outcomes | cqhe009 | 28238 | NO: 27719, 98.16 %, YES: 519, 1.84 % | | Atrial fibrillation, doctor-diagnosed, self-reported | | | Core Questionnaire | SCAPIS |

**Table S2: Summary of clinical, molecular, and imaging-derived variables in SCAPIS, and disease outcomes in UKBB**

| Category | Variable_Name | Data_Available | Summary | Unit | Heading | Context | Method | Source | Cohort |
|---|---|---|---|---|---|---|---|---|---|
| Clinical Outcomes | cqhe011 | 28238 | NO: 28094, 99.49 %, YES: 144, 0.51 % | | Heart failure, self-reported, doctor-diagnosed | | | Core Questionnaire | SCAPIS |
| Clinical Outcomes | cqhe029 | 28238 | NO: 27831, 98.56 %, YES: 407, 1.44 % | | Stroke, doctor-diagnosed, self-reported | | | Core Questionnaire | SCAPIS |
| Clinical Outcomes | cqhe034 | 28313 | NO: 21853, 77.18 %, YES: 6460, 22.82 % | | Hypertension, doctor-diagnosed, self-reported | | | Core Questionnaire | SCAPIS |
| Clinical Outcomes | cqhe036 | 28313 | NO: 24963, 88.17 %, YES: 3350, 11.83 % | | Hyperlipidemia, doctor-diagnosed, self-reported | | | Core Questionnaire | SCAPIS |
| Clinical Outcomes | cqhe038 | 28313 | NO: 27057, 95.56 %, YES: 1256, 4.44 % | | Diabetes, doctor-diagnosed, self-reported | | | Core Questionnaire | SCAPIS |
| Clinical Outcomes | cqhe058 | 28238 | NO: 26972, 95.52 %, YES: 1266, 4.48 % | | Sleep apnea, doctor-diagnosed, self-reported | | | Core Questionnaire | SCAPIS |
| Clinical Outcomes | cqhe069 | 28238 | NO: 27185, 96.27 %, YES: 1053, 3.73 % | | Rheumatic disease (e.g. rheumatoid arthritis, Bechterew's disease, psoriatic arthritis, SLE, Sjögren's syndrome), doctor-diagnosed, self-reported | | | Core Questionnaire | SCAPIS |
| Clinical Outcomes | cqhe073 | 28238 | NO: 26563, 94.07 %, YES: 1675, 5.93 % | | Cancer, doctor-diagnosed, self-reported | | | Core Questionnaire | SCAPIS |
| Clinical Outcomes | cqhe_cad | 28241 | NO: 27539, 97.51 %, YES: 702, 2.49 % | | Coronary artery disease (merged from participants with myocardial infarction, angina pectoris, and participants who received CABG or PCI interventions), doctor-diagnosed, self-reported. | | | Core Questionnaire | SCAPIS |
| Diabetes Related Diseases | acmort | 458905 | ALL-YES: 41,085, 8.95%, INCIDENT-YES: 41,085, 8.95% | | All cause mortality | | | Derived | UK Biobank |
| Diabetes Related Diseases | cad | 458905 | ALL-YES: 35,095, 7.65%, INCIDENT-YES: 19,980, 4.35% | | Coronary artery disease | | | Derived | UK Biobank |
| Diabetes Related Diseases | cvdmort | 458905 | ALL-YES: 8,491, 1.85%, INCIDENT-YES: 8,491, 1.85% | | Cardiovascular disease mortality | | | Derived | UK Biobank |
| Diabetes Related Diseases | hf | 458905 | ALL-YES: 19,983, 4.35%, INCIDENT-YES: 17,352, 3.78% | | Heart failure | | | Derived | UK Biobank |
| Diabetes Related Diseases | is | 458905 | ALL-YES: 11,148, 2.43%, INCIDENT-YES: 9,186, 2.00% | | Ischemic stroke | | | Derived | UK Biobank |
| Diabetes related Diseases | pad | 458905 | ALL-YES: 10,663, 2.32%, INCIDENT-YES: 7,415, 1.62% | | Peripheral artery disease | | | Derived | UK Biobank |
| Diabetic Complications | nephrodm | 458905 | ALL-YES: 811, 0.18%, INCIDENT-YES: 692, 0.15% | | Diabetic nephropathy | | | Derived | UK Biobank |
| Diabetic Complications | neurodm | 458905 | ALL-YES: 2,328, 0.51%, INCIDENT-YES: 1,923, 0.42% | | Diabetic neuropathy | | | Derived | UK Biobank |
| Diabetic Complications | retidm | 458905 | ALL-YES: 4,865, 1.06%, INCIDENT-YES: 3,782, 0.82% | | Diabetic retinopathy | | | Derived | UK Biobank |

**Table S3: pPS cross-correlation in SCAPIS**

**a) PPS Cross-Associations in Participants without Doctor-Diagnosed or Self-Reported Diabetes**

| PPS_Column1 | PPS_Column2 | Sample_Size | r | p | fdr_p_adjusted |
|---|---|---|---|---|---|
| Cholesterol_pPS | Cholesterol_pPS | 27057 | 1 | 0 | 0 |
| Cholesterol_pPS | Lipodystrophy_1_pPS | 27057 | -0.00182 | 0.764156733 | 0.83870861 |
| Cholesterol_pPS | Lipodystrophy_2_pPS | 27057 | 0.038898 | 1.55E-10 | 3.83E-10 |
| Cholesterol_pPS | Liver-Lipid_pPS | 27057 | 0.014161 | 0.01984397 | 0.03166591 |
| Cholesterol_pPS | ASAT_Pos_pPS | 27057 | -0.00139 | 0.819181844 | 0.857283326 |
| Cholesterol_pPS | ALP_Neg_pPS | 27057 | 0.017995 | 0.003074783 | 0.005624602 |
| Cholesterol_pPS | SHBG-LpA_pPS | 27057 | 0.015843 | 0.009160626 | 0.015733899 |
| Cholesterol_pPS | Obesity_pPS | 27057 | 0.073786 | 5.52E-34 | 1.75E-33 |
| Cholesterol_pPS | VAT_Neg_pPS | 27057 | 0.00488 | 0.422114498 | 0.536586227 |
| Cholesterol_pPS | Bilirubin_pPS | 27057 | 0.002373 | 0.696293542 | 0.787266567 |
| Cholesterol_pPS | Proinsulin_pPS | 27057 | 0.003576 | 0.556411505 | 0.69167176 |
| Cholesterol_pPS | Hyper_Insulin_pPS | 27057 | 0.002679 | 0.659436852 | 0.760888675 |
| Cholesterol_pPS | Beta_Cell_1_pPS | 27057 | 0.007313 | 0.228992645 | 0.319410419 |
| Cholesterol_pPS | Beta_Cell_2_pPS | 27057 | 0.008599 | 0.157238341 | 0.232846282 |
| Cholesterol_pPS | Total_GRS | 27057 | 0.117023 | 4.05E-83 | 1.94E-82 |
| Lipodystrophy_1_pPS | Cholesterol_pPS | 27057 | -0.00182 | 0.764156733 | 0.83870861 |
| Lipodystrophy_1_pPS | Lipodystrophy_1_pPS | 27057 | 1 | 0 | 0 |
| Lipodystrophy_1_pPS | Lipodystrophy_2_pPS | 27057 | 0.052445 | 6.02E-18 | 1.76E-17 |
| Lipodystrophy_1_pPS | Liver-Lipid_pPS | 27057 | 0.015548 | 0.010539777 | 0.017566295 |
| Lipodystrophy_1_pPS | ASAT_Pos_pPS | 27057 | 0.15945 | 1.53E-153 | 8.01E-153 |
| Lipodystrophy_1_pPS | ALP_Neg_pPS | 27057 | 0.023568 | 0.00010574 | 0.000215789 |
| Lipodystrophy_1_pPS | SHBG-LpA_pPS | 27057 | 0.075664 | 1.18E-35 | 3.86E-35 |
| Lipodystrophy_1_pPS | Obesity_pPS | 27057 | 0.019175 | 0.001609031 | 0.003042286 |
| Lipodystrophy_1_pPS | VAT_Neg_pPS | 27057 | 0.014903 | 0.014228554 | 0.023031832 |
| Lipodystrophy_1_pPS | Bilirubin_pPS | 27057 | -0.00152 | 0.802247197 | 0.847444222 |
| Lipodystrophy_1_pPS | Proinsulin_pPS | 27057 | -0.0016 | 0.791883519 | 0.847444222 |
| Lipodystrophy_1_pPS | Hyper_Insulin_pPS | 27057 | 0.116239 | 5.01E-82 | 2.21E-81 |
| Lipodystrophy_1_pPS | Beta_Cell_1_pPS | 27057 | 0.026096 | 1.76E-05 | 3.71E-05 |
| Lipodystrophy_1_pPS | Beta_Cell_2_pPS | 27057 | 0.002877 | 0.636036137 | 0.757185878 |
| Lipodystrophy_1_pPS | Total_GRS | 27057 | 0.354758 | 0 | 0 |
| Lipodystrophy_2_pPS | Cholesterol_pPS | 27057 | 0.038898 | 1.55E-10 | 3.83E-10 |
| Lipodystrophy_2_pPS | Lipodystrophy_1_pPS | 27057 | 0.052445 | 6.02E-18 | 1.76E-17 |
| Lipodystrophy_2_pPS | Lipodystrophy_2_pPS | 27057 | 1 | 0 | 0 |
| Lipodystrophy_2_pPS | Liver-Lipid_pPS | 27057 | 0.030486 | 5.29E-07 | 1.20E-06 |
| Lipodystrophy_2_pPS | ASAT_Pos_pPS | 27057 | 0.098531 | 2.38E-59 | 9.38E-59 |
| Lipodystrophy_2_pPS | ALP_Neg_pPS | 27057 | 0.000942 | 0.876910813 | 0.900935767 |
| Lipodystrophy_2_pPS | SHBG-LpA_pPS | 27057 | -0.00187 | 0.757976656 | 0.83870861 |
| Lipodystrophy_2_pPS | Obesity_pPS | 27057 | -0.00049 | 0.935114587 | 0.952039738 |
| Lipodystrophy_2_pPS | VAT_Neg_pPS | 27057 | 0.116086 | 8.15E-82 | 3.46E-81 |
| Lipodystrophy_2_pPS | Bilirubin_pPS | 27057 | -0.00728 | 0.231395104 | 0.319410419 |
| Lipodystrophy_2_pPS | Proinsulin_pPS | 27057 | 0.015227 | 0.012253497 | 0.020124357 |
| Lipodystrophy_2_pPS | Hyper_Insulin_pPS | 27057 | 0.08952 | 2.89E-49 | 1.10E-48 |
| Lipodystrophy_2_pPS | Beta_Cell_1_pPS | 27057 | -0.0072 | 0.236271677 | 0.32218865 |
| Lipodystrophy_2_pPS | Beta_Cell_2_pPS | 27057 | 0.001535 | 0.8007066 | 0.847444222 |
| Lipodystrophy_2_pPS | Total_GRS | 27057 | 0.413784 | 0 | 0 |
| Liver-Lipid_pPS | Cholesterol_pPS | 27057 | 0.014161 | 0.01984397 | 0.03166591 |
| Liver-Lipid_pPS | Lipodystrophy_1_pPS | 27057 | 0.015548 | 0.010539777 | 0.017566295 |
| Liver-Lipid_pPS | Lipodystrophy_2_pPS | 27057 | 0.030486 | 5.29E-07 | 1.20E-06 |
| Liver-Lipid_pPS | Liver-Lipid_pPS | 27057 | 1 | 0 | 0 |
| Liver-Lipid_pPS | ASAT_Pos_pPS | 27057 | 0.005399 | 0.374515044 | 0.486584051 |
| Liver-Lipid_pPS | ALP_Neg_pPS | 27057 | 0.045842 | 4.55E-14 | 1.18E-13 |
| Liver-Lipid_pPS | SHBG-LpA_pPS | 27057 | 0.023128 | 0.000142011 | 0.000282764 |
| Liver-Lipid_pPS | Obesity_pPS | 27057 | 0.015771 | 0.009478933 | 0.016035789 |
| Liver-Lipid_pPS | VAT_Neg_pPS | 27057 | 0.007891 | 0.194280146 | 0.278426961 |
| Liver-Lipid_pPS | Bilirubin_pPS | 27057 | -0.01659 | 0.006347381 | 0.011245361 |
| Liver-Lipid_pPS | Proinsulin_pPS | 27057 | 0.036426 | 2.05E-09 | 4.97E-09 |
| Liver-Lipid_pPS | Hyper_Insulin_pPS | 27057 | 0.034946 | 8.94E-09 | 2.12E-08 |
| Liver-Lipid_pPS | Beta_Cell_1_pPS | 27057 | 0.048981 | 7.55E-16 | 2.10E-15 |
| Liver-Lipid_pPS | Beta_Cell_2_pPS | 27057 | 0.039018 | 1.36E-10 | 3.44E-10 |
| Liver-Lipid_pPS | Total_GRS | 27057 | 0.088281 | 5.92E-48 | 2.18E-47 |
| ASAT_Pos_pPS | Cholesterol_pPS | 27057 | -0.00139 | 0.819181844 | 0.857283326 |
| ASAT_Pos_pPS | Lipodystrophy_1_pPS | 27057 | 0.15945 | 1.53E-153 | 8.01E-153 |
| ASAT_Pos_pPS | Lipodystrophy_2_pPS | 27057 | 0.098531 | 2.38E-59 | 9.38E-59 |
| ASAT_Pos_pPS | Liver-Lipid_pPS | 27057 | 0.005399 | 0.374515044 | 0.486584051 |
| ASAT_Pos_pPS | ASAT_Pos_pPS | 27057 | 1 | 0 | 0 |
| ASAT_Pos_pPS | ALP_Neg_pPS | 27057 | -0.00946 | 0.119659388 | 0.183152124 |
| ASAT_Pos_pPS | SHBG-LpA_pPS | 27057 | -0.00608 | 0.317263145 | 0.422391761 |
| ASAT_Pos_pPS | Obesity_pPS | 27057 | 0.046476 | 2.03E-14 | 5.38E-14 |
| ASAT_Pos_pPS | VAT_Neg_pPS | 27057 | 0.008911 | 0.142723275 | 0.215521724 |
| ASAT_Pos_pPS | Bilirubin_pPS | 27057 | 0.002764 | 0.649394815 | 0.760888675 |
| ASAT_Pos_pPS | Proinsulin_pPS | 27057 | 0.012521 | 0.039439652 | 0.062055396 |
| ASAT_Pos_pPS | Hyper_Insulin_pPS | 27057 | 0.08693 | 1.51E-46 | 5.24E-46 |
| ASAT_Pos_pPS | Beta_Cell_1_pPS | 27057 | 0.034316 | 1.64E-08 | 3.81E-08 |
| ASAT_Pos_pPS | Beta_Cell_2_pPS | 27057 | -3.50E-05 | 0.995402312 | 0.995402312 |
| ASAT_Pos_pPS | Total_GRS | 27057 | 0.231962 | 0 | 0 |
| ALP_Neg_pPS | Cholesterol_pPS | 27057 | 0.017995 | 0.003074783 | 0.005624602 |
| ALP_Neg_pPS | Lipodystrophy_1_pPS | 27057 | 0.023568 | 0.00010574 | 0.000215789 |
| ALP_Neg_pPS | Lipodystrophy_2_pPS | 27057 | 0.000942 | 0.876910813 | 0.900935767 |
| ALP_Neg_pPS | Liver-Lipid_pPS | 27057 | 0.045842 | 4.55E-14 | 1.18E-13 |
| ALP_Neg_pPS | ASAT_Pos_pPS | 27057 | -0.00946 | 0.119659388 | 0.183152124 |
| ALP_Neg_pPS | ALP_Neg_pPS | 27057 | 1 | 0 | 0 |
| ALP_Neg_pPS | SHBG-LpA_pPS | 27057 | -0.00248 | 0.683359554 | 0.7804868 |
| ALP_Neg_pPS | Obesity_pPS | 27057 | 0.028239 | 3.39E-06 | 7.40E-06 |
| ALP_Neg_pPS | VAT_Neg_pPS | 27057 | 0.004249 | 0.484631806 | 0.609174058 |
| ALP_Neg_pPS | Bilirubin_pPS | 27057 | 0.001614 | 0.790583673 | 0.847444222 |
| ALP_Neg_pPS | Proinsulin_pPS | 27057 | 0.176136 | 1.95E-187 | 1.12E-186 |
| ALP_Neg_pPS | Hyper_Insulin_pPS | 27057 | -0.00291 | 0.632263419 | 0.757185878 |
| ALP_Neg_pPS | Beta_Cell_1_pPS | 27057 | 0.009835 | 0.105712804 | 0.16403711 |
| ALP_Neg_pPS | Beta_Cell_2_pPS | 27057 | 0.019244 | 0.00154727 | 0.002975518 |
| ALP_Neg_pPS | Total_GRS | 27057 | 0.084748 | 2.56E-44 | 8.60E-44 |
| SHBG-LpA_pPS | Cholesterol_pPS | 27057 | 0.015843 | 0.009160626 | 0.015733899 |
| SHBG-LpA_pPS | Lipodystrophy_1_pPS | 27057 | 0.075664 | 1.18E-35 | 3.86E-35 |

**b) PPS Cross-Associations in Participants with Doctor-Diagnosed or Self-Reported Diabetes**

| PPS_Column1 | PPS_Column2 | Sample_Size | r | p | fdr_p_adjusted |
|---|---|---|---|---|---|
| Cholesterol_pPS | Cholesterol_pPS | 1256 | 1 | 0 | 0 |
| Cholesterol_pPS | Lipodystrophy_1_pPS | 1256 | -0.06931 | 0.014016 | 0.040957044 |
| Cholesterol_pPS | Lipodystrophy_2_pPS | 1256 | -0.0075 | 0.790564 | 0.92164143 |
| Cholesterol_pPS | Liver-Lipid_pPS | 1256 | 0.002298 | 0.935162 | 0.969638401 |
| Cholesterol_pPS | ASAT_Pos_pPS | 1256 | -0.01517 | 0.591233 | 0.768945161 |
| Cholesterol_pPS | ALP_Neg_pPS | 1256 | -0.02023 | 0.473869 | 0.67911204 |
| Cholesterol_pPS | SHBG-LpA_pPS | 1256 | -0.04258 | 0.131512 | 0.271469733 |
| Cholesterol_pPS | Obesity_pPS | 1256 | 0.056595 | 0.044928 | 0.113582875 |
| Cholesterol_pPS | VAT_Neg_pPS | 1256 | 0.006029 | 0.83096 | 0.938155988 |
| Cholesterol_pPS | Bilirubin_pPS | 1256 | 0.005607 | 0.842651 | 0.938155988 |
| Cholesterol_pPS | Proinsulin_pPS | 1256 | 0.006796 | 0.809848 | 0.924952884 |
| Cholesterol_pPS | Hyper_Insulin_pPS | 1256 | 0.00924 | 0.743561 | 0.888699624 |
| Cholesterol_pPS | Beta_Cell_1_pPS | 1256 | -0.0597 | 0.03439 | 0.088940185 |
| Cholesterol_pPS | Beta_Cell_2_pPS | 1256 | -0.01938 | 0.492478 | 0.679801386 |
| Cholesterol_pPS | Total_GRS | 1256 | 0.049938 | 0.076868 | 0.178303114 |
| Lipodystrophy_1_pPS | Cholesterol_pPS | 1256 | -0.06931 | 0.014016 | 0.040957044 |
| Lipodystrophy_1_pPS | Lipodystrophy_1_pPS | 1256 | 1 | 0 | 0 |
| Lipodystrophy_1_pPS | Lipodystrophy_2_pPS | 1256 | 0.087347 | 0.001946 | 0.006948391 |
| Lipodystrophy_1_pPS | Liver-Lipid_pPS | 1256 | 0.00913 | 0.746508 | 0.888699624 |
| Lipodystrophy_1_pPS | ASAT_Pos_pPS | 1256 | 0.1555 | 3.04E-08 | 1.59E-07 |
| Lipodystrophy_1_pPS | ALP_Neg_pPS | 1256 | 0.001682 | 0.952526 | 0.972043728 |
| Lipodystrophy_1_pPS | SHBG-LpA_pPS | 1256 | 0.061548 | 0.029172 | 0.079080961 |
| Lipodystrophy_1_pPS | Obesity_pPS | 1256 | 0.01367 | 0.628383 | 0.807921029 |
| Lipodystrophy_1_pPS | VAT_Neg_pPS | 1256 | 0.015513 | 0.582823 | 0.766872431 |
| Lipodystrophy_1_pPS | Bilirubin_pPS | 1256 | 0.008787 | 0.755718 | 0.890244 |
| Lipodystrophy_1_pPS | Proinsulin_pPS | 1256 | -0.04908 | 0.082103 | 0.186597242 |
| Lipodystrophy_1_pPS | Hyper_Insulin_pPS | 1256 | 0.109084 | 0.000107 | 0.000438571 |
| Lipodystrophy_1_pPS | Beta_Cell_1_pPS | 1256 | 0.024148 | 0.3925 | 0.635342177 |
| Lipodystrophy_1_pPS | Beta_Cell_2_pPS | 1256 | -0.02138 | 0.449027 | 0.660333913 |
| Lipodystrophy_1_pPS | Total_GRS | 1256 | 0.377446 | 8.37E-44 | 8.19E-43 |
| Lipodystrophy_2_pPS | Cholesterol_pPS | 1256 | -0.0075 | 0.790564 | 0.92164143 |
| Lipodystrophy_2_pPS | Lipodystrophy_1_pPS | 1256 | 0.087347 | 0.001946 | 0.006948391 |
| Lipodystrophy_2_pPS | Lipodystrophy_2_pPS | 1256 | 1 | 0 | 0 |
| Lipodystrophy_2_pPS | Liver-Lipid_pPS | 1256 | 0.083108 | 0.003203 | 0.010446061 |
| Lipodystrophy_2_pPS | ASAT_Pos_pPS | 1256 | 0.074885 | 0.00793 | 0.024441993 |
| Lipodystrophy_2_pPS | ALP_Neg_pPS | 1256 | -0.0235 | 0.405424 | 0.639493899 |
| Lipodystrophy_2_pPS | SHBG-LpA_pPS | 1256 | 0.04585 | 0.104344 | 0.223593931 |
| Lipodystrophy_2_pPS | Obesity_pPS | 1256 | -0.0315 | 0.264567 | 0.489258376 |
| Lipodystrophy_2_pPS | VAT_Neg_pPS | 1256 | 0.106543 | 0.000155 | 0.000611836 |
| Lipodystrophy_2_pPS | Bilirubin_pPS | 1256 | -0.02034 | 0.471374 | 0.67911204 |
| Lipodystrophy_2_pPS | Proinsulin_pPS | 1256 | -0.02612 | 0.354941 | 0.591568053 |
| Lipodystrophy_2_pPS | Hyper_Insulin_pPS | 1256 | 0.104035 | 0.000221 | 0.00084399 |
| Lipodystrophy_2_pPS | Beta_Cell_1_pPS | 1256 | 0.000836 | 0.976398 | 0.97639817 |
| Lipodystrophy_2_pPS | Beta_Cell_2_pPS | 1256 | 0.021932 | 0.437398 | 0.651752464 |
| Lipodystrophy_2_pPS | Total_GRS | 1256 | 0.424215 | 5.00E-56 | 6.61E-55 |
| Liver-Lipid_pPS | Cholesterol_pPS | 1256 | 0.002298 | 0.935162 | 0.969638401 |
| Liver-Lipid_pPS | Lipodystrophy_1_pPS | 1256 | 0.00913 | 0.746508 | 0.888699624 |
| Liver-Lipid_pPS | Lipodystrophy_2_pPS | 1256 | 0.083108 | 0.003203 | 0.010446061 |
| Liver-Lipid_pPS | Liver-Lipid_pPS | 1256 | 1 | 0 | 0 |
| Liver-Lipid_pPS | ASAT_Pos_pPS | 1256 | 0.02207 | 0.434529 | 0.651752464 |
| Liver-Lipid_pPS | ALP_Neg_pPS | 1256 | 0.070968 | 0.011877 | 0.0356296 |
| Liver-Lipid_pPS | SHBG-LpA_pPS | 1256 | 0.03262 | 0.247997 | 0.48521131 |
| Liver-Lipid_pPS | Obesity_pPS | 1256 | -0.01959 | 0.487806 | 0.679801386 |
| Liver-Lipid_pPS | VAT_Neg_pPS | 1256 | 0.030191 | 0.284994 | 0.498316621 |
| Liver-Lipid_pPS | Bilirubin_pPS | 1256 | 0.011324 | 0.688468 | 0.855829823 |
| Liver-Lipid_pPS | Proinsulin_pPS | 1256 | 0.045405 | 0.107754 | 0.226585242 |
| Liver-Lipid_pPS | Hyper_Insulin_pPS | 1256 | 0.022021 | 0.435542 | 0.651752464 |
| Liver-Lipid_pPS | Beta_Cell_1_pPS | 1256 | 0.062987 | 0.025598 | 0.071104785 |
| Liver-Lipid_pPS | Beta_Cell_2_pPS | 1256 | 0.036505 | 0.196048 | 0.397394279 |
| Liver-Lipid_pPS | Total_GRS | 1256 | 0.115883 | 3.84E-05 | 0.000169576 |
| ASAT_Pos_pPS | Cholesterol_pPS | 1256 | -0.01517 | 0.591233 | 0.768945161 |
| ASAT_Pos_pPS | Lipodystrophy_1_pPS | 1256 | 0.1555 | 3.04E-08 | 1.59E-07 |
| ASAT_Pos_pPS | Lipodystrophy_2_pPS | 1256 | 0.074885 | 0.00793 | 0.024441993 |
| ASAT_Pos_pPS | Liver-Lipid_pPS | 1256 | 0.02207 | 0.434529 | 0.651752464 |
| ASAT_Pos_pPS | ASAT_Pos_pPS | 1256 | 1 | 0 | 0 |
| ASAT_Pos_pPS | ALP_Neg_pPS | 1256 | 0.028618 | 0.310856 | 0.525884238 |
| ASAT_Pos_pPS | SHBG-LpA_pPS | 1256 | -0.00089 | 0.974898 | 0.97639817 |
| ASAT_Pos_pPS | Obesity_pPS | 1256 | 0.076907 | 0.006393 | 0.020260328 |
| ASAT_Pos_pPS | VAT_Neg_pPS | 1256 | 0.003093 | 0.912807 | 0.9617738 |
| ASAT_Pos_pPS | Bilirubin_pPS | 1256 | -0.01785 | 0.527405 | 0.710575928 |
| ASAT_Pos_pPS | Proinsulin_pPS | 1256 | 0.003509 | 0.901116 | 0.960906067 |
| ASAT_Pos_pPS | Hyper_Insulin_pPS | 1256 | 0.086055 | 0.00227 | 0.007857957 |
| ASAT_Pos_pPS | Beta_Cell_1_pPS | 1256 | 0.019435 | 0.49135 | 0.679801386 |
| ASAT_Pos_pPS | Beta_Cell_2_pPS | 1256 | -0.05563 | 0.0487 | 0.120412274 |
| ASAT_Pos_pPS | Total_GRS | 1256 | 0.206737 | 1.37E-13 | 8.31E-13 |
| ALP_Neg_pPS | Cholesterol_pPS | 1256 | -0.02023 | 0.473869 | 0.67911204 |
| ALP_Neg_pPS | Lipodystrophy_1_pPS | 1256 | 0.001682 | 0.952526 | 0.972043728 |
| ALP_Neg_pPS | Lipodystrophy_2_pPS | 1256 | -0.0235 | 0.405424 | 0.639493899 |
| ALP_Neg_pPS | Liver-Lipid_pPS | 1256 | 0.070968 | 0.011877 | 0.0356296 |
| ALP_Neg_pPS | ASAT_Pos_pPS | 1256 | 0.028618 | 0.310856 | 0.525884238 |
| ALP_Neg_pPS | ALP_Neg_pPS | 1256 | 1 | 0 | 0 |
| ALP_Neg_pPS | SHBG-LpA_pPS | 1256 | -0.03266 | 0.247402 | 0.48521131 |
| ALP_Neg_pPS | Obesity_pPS | 1256 | 0.010396 | 0.712811 | 0.876407545 |
| ALP_Neg_pPS | VAT_Neg_pPS | 1256 | -0.02262 | 0.42324 | 0.651752464 |
| ALP_Neg_pPS | Bilirubin_pPS | 1256 | -0.0093 | 0.741946 | 0.888699624 |
| ALP_Neg_pPS | Proinsulin_pPS | 1256 | 0.150027 | 9.20E-08 | 4.60E-07 |
| ALP_Neg_pPS | Hyper_Insulin_pPS | 1256 | 0.003806 | 0.892799 | 0.960906067 |
| ALP_Neg_pPS | Beta_Cell_1_pPS | 1256 | 0.085133 | 0.002531 | 0.008500178 |
| ALP_Neg_pPS | Beta_Cell_2_pPS | 1256 | 0.023445 | 0.406434 | 0.639493899 |
| ALP_Neg_pPS | Total_GRS | 1256 | 0.046554 | 0.099124 | 0.21653349 |
| SHBG-LpA_pPS | Cholesterol_pPS | 1256 | -0.04258 | 0.131512 | 0.271469733 |
| SHBG-LpA_pPS | Lipodystrophy_1_pPS | 1256 | 0.061548 | 0.029172 | 0.079080961 |

**Table S3: pPS cross-correlation in SCAPIS**

**a) PPS Cross-Associations in Participants without Doctor-Diagnosed or Self-Reported Diabetes**

| PPS_Column1 | PPS_Column2 | Sample_Size | r | p | fdr_p_adjusted |
|---|---|---|---|---|---|
| SHBG-LpA_pPS | Lipodystrophy_2_pPS | 27057 | -0.00187 | 0.757976656 | 0.83870861 |
| SHBG-LpA_pPS | Liver-Lipid_pPS | 27057 | 0.023128 | 0.000142011 | 0.000282764 |
| SHBG-LpA_pPS | ASAT_Pos_pPS | 27057 | -0.00608 | 0.317263145 | 0.422391761 |
| SHBG-LpA_pPS | ALP_Neg_pPS | 27057 | -0.00248 | 0.683359554 | 0.7804868 |
| SHBG-LpA_pPS | SHBG-LpA_pPS | 27057 | 1 | 0 | 0 |
| SHBG-LpA_pPS | Obesity_pPS | 27057 | -0.01618 | 0.007779833 | 0.013569475 |
| SHBG-LpA_pPS | VAT_Neg_pPS | 27057 | 0.064789 | 1.44E-26 | 4.44E-26 |
| SHBG-LpA_pPS | Bilirubin_pPS | 27057 | -0.00219 | 0.718289633 | 0.804055559 |
| SHBG-LpA_pPS | Proinsulin_pPS | 27057 | 0.023558 | 0.000106456 | 0.000215789 |
| SHBG-LpA_pPS | Hyper_Insulin_pPS | 27057 | 0.021424 | 0.000424662 | 0.00083086 |
| SHBG-LpA_pPS | Beta_Cell_1_pPS | 27057 | 0.018041 | 0.003000099 | 0.005578697 |
| SHBG-LpA_pPS | Beta_Cell_2_pPS | 27057 | 0.003193 | 0.599484109 | 0.729102294 |
| SHBG-LpA_pPS | Total_GRS | 27057 | 0.050081 | 1.69E-16 | 4.80E-16 |
| Obesity_pPS | Cholesterol_pPS | 27057 | 0.073786 | 5.52E-34 | 1.75E-33 |
| Obesity_pPS | Lipodystrophy_1_pPS | 27057 | 0.019175 | 0.001609031 | 0.003042286 |
| Obesity_pPS | Lipodystrophy_2_pPS | 27057 | -0.00049 | 0.935114587 | 0.952039738 |
| Obesity_pPS | Liver-Lipid_pPS | 27057 | 0.015771 | 0.009478933 | 0.016035789 |
| Obesity_pPS | ASAT_Pos_pPS | 27057 | 0.046476 | 2.03E-14 | 5.38E-14 |
| Obesity_pPS | ALP_Neg_pPS | 27057 | 0.028239 | 3.39E-06 | 7.40E-06 |
| Obesity_pPS | SHBG-LpA_pPS | 27057 | -0.01618 | 0.007779833 | 0.013569475 |
| Obesity_pPS | Obesity_pPS | 27057 | 1 | 0 | 0 |
| Obesity_pPS | VAT_Neg_pPS | 27057 | -0.00268 | 0.658863696 | 0.760888675 |
| Obesity_pPS | Bilirubin_pPS | 27057 | 0.001198 | 0.843730905 | 0.874836192 |
| Obesity_pPS | Proinsulin_pPS | 27057 | 0.134275 | 4.65E-109 | 2.32E-108 |
| Obesity_pPS | Hyper_Insulin_pPS | 27057 | 0.058747 | 4.00E-22 | 1.20E-21 |
| Obesity_pPS | Beta_Cell_1_pPS | 27057 | 0.02701 | 8.85E-06 | 1.90E-05 |
| Obesity_pPS | Beta_Cell_2_pPS | 27057 | 0.005654 | 0.352408534 | 0.46369544 |
| Obesity_pPS | Total_GRS | 27057 | 0.390167 | 0 | 0 |
| VAT_Neg_pPS | Cholesterol_pPS | 27057 | 0.00488 | 0.422114498 | 0.536586227 |
| VAT_Neg_pPS | Lipodystrophy_1_pPS | 27057 | 0.014903 | 0.014228554 | 0.023031832 |
| VAT_Neg_pPS | Lipodystrophy_2_pPS | 27057 | 0.116086 | 8.15E-82 | 3.46E-81 |
| VAT_Neg_pPS | Liver-Lipid_pPS | 27057 | 0.007891 | 0.194280146 | 0.278426961 |
| VAT_Neg_pPS | ASAT_Pos_pPS | 27057 | 0.008911 | 0.142723275 | 0.215521724 |
| VAT_Neg_pPS | ALP_Neg_pPS | 27057 | 0.004249 | 0.484631806 | 0.609174058 |
| VAT_Neg_pPS | SHBG-LpA_pPS | 27057 | 0.064789 | 1.44E-26 | 4.44E-26 |
| VAT_Neg_pPS | Obesity_pPS | 27057 | -0.00268 | 0.658863696 | 0.760888675 |
| VAT_Neg_pPS | VAT_Neg_pPS | 27057 | 1 | 0 | 0 |
| VAT_Neg_pPS | Bilirubin_pPS | 27057 | 0.007412 | 0.222767314 | 0.315236765 |
| VAT_Neg_pPS | Proinsulin_pPS | 27057 | 0.047726 | 4.01E-15 | 1.09E-14 |
| VAT_Neg_pPS | Hyper_Insulin_pPS | 27057 | 0.101343 | 1.07E-62 | 4.37E-62 |
| VAT_Neg_pPS | Beta_Cell_1_pPS | 27057 | 0.116957 | 5.01E-83 | 2.30E-82 |
| VAT_Neg_pPS | Beta_Cell_2_pPS | 27057 | 0.087641 | 2.77E-47 | 9.89E-47 |
| VAT_Neg_pPS | Total_GRS | 27057 | 0.222481 | 1.25E-300 | 7.62E-300 |
| Bilirubin_pPS | Cholesterol_pPS | 27057 | 0.002373 | 0.696293542 | 0.787266567 |
| Bilirubin_pPS | Lipodystrophy_1_pPS | 27057 | -0.00152 | 0.802247197 | 0.847444222 |
| Bilirubin_pPS | Lipodystrophy_2_pPS | 27057 | -0.00728 | 0.231395104 | 0.319410419 |
| Bilirubin_pPS | Liver-Lipid_pPS | 27057 | -0.01659 | 0.006347381 | 0.011245361 |
| Bilirubin_pPS | ASAT_Pos_pPS | 27057 | 0.002764 | 0.649394815 | 0.760888675 |
| Bilirubin_pPS | ALP_Neg_pPS | 27057 | 0.001614 | 0.790583673 | 0.847444222 |
| Bilirubin_pPS | SHBG-LpA_pPS | 27057 | -0.00219 | 0.718289633 | 0.804055559 |
| Bilirubin_pPS | Obesity_pPS | 27057 | 0.001198 | 0.843730905 | 0.874836192 |
| Bilirubin_pPS | VAT_Neg_pPS | 27057 | 0.007412 | 0.222767314 | 0.315236765 |
| Bilirubin_pPS | Bilirubin_pPS | 27057 | 1 | 0 | 0 |
| Bilirubin_pPS | Proinsulin_pPS | 27057 | -0.00654 | 0.282285165 | 0.380324325 |
| Bilirubin_pPS | Hyper_Insulin_pPS | 27057 | -0.00858 | 0.158335472 | 0.232846282 |
| Bilirubin_pPS | Beta_Cell_1_pPS | 27057 | 0.028371 | 3.05E-06 | 6.79E-06 |
| Bilirubin_pPS | Beta_Cell_2_pPS | 27057 | -0.00335 | 0.581778388 | 0.715301297 |
| Bilirubin_pPS | Total_GRS | 27057 | 0.016978 | 0.005224497 | 0.009404094 |
| Proinsulin_pPS | Cholesterol_pPS | 27057 | 0.003576 | 0.556411505 | 0.69167176 |
| Proinsulin_pPS | Lipodystrophy_1_pPS | 27057 | -0.0016 | 0.791883519 | 0.847444222 |
| Proinsulin_pPS | Lipodystrophy_2_pPS | 27057 | 0.015227 | 0.012253497 | 0.020124357 |
| Proinsulin_pPS | Liver-Lipid_pPS | 27057 | 0.036426 | 2.05E-09 | 4.97E-09 |
| Proinsulin_pPS | ASAT_Pos_pPS | 27057 | 0.012521 | 0.039439652 | 0.062055396 |
| Proinsulin_pPS | ALP_Neg_pPS | 27057 | 0.176136 | 1.95E-187 | 1.12E-186 |
| Proinsulin_pPS | SHBG-LpA_pPS | 27057 | 0.023558 | 0.000106456 | 0.000215789 |
| Proinsulin_pPS | Obesity_pPS | 27057 | 0.134275 | 4.65E-109 | 2.32E-108 |
| Proinsulin_pPS | VAT_Neg_pPS | 27057 | 0.047726 | 4.01E-15 | 1.09E-14 |
| Proinsulin_pPS | Bilirubin_pPS | 27057 | -0.00654 | 0.282285165 | 0.380324325 |
| Proinsulin_pPS | Proinsulin_pPS | 27057 | 1 | 0 | 0 |
| Proinsulin_pPS | Hyper_Insulin_pPS | 27057 | 0.005355 | 0.378454262 | 0.486584051 |
| Proinsulin_pPS | Beta_Cell_1_pPS | 27057 | 0.302096 | 0 | 0 |
| Proinsulin_pPS | Beta_Cell_2_pPS | 27057 | 0.170536 | 1.16E-175 | 6.35E-175 |
| Proinsulin_pPS | Total_GRS | 27057 | 0.369098 | 0 | 0 |
| Hyper_Insulin_pPS | Cholesterol_pPS | 27057 | 0.002679 | 0.659436852 | 0.760888675 |
| Hyper_Insulin_pPS | Lipodystrophy_1_pPS | 27057 | 0.116239 | 5.01E-82 | 2.21E-81 |
| Hyper_Insulin_pPS | Lipodystrophy_2_pPS | 27057 | 0.08952 | 2.89E-49 | 1.10E-48 |
| Hyper_Insulin_pPS | Liver-Lipid_pPS | 27057 | 0.034946 | 8.94E-09 | 2.12E-08 |
| Hyper_Insulin_pPS | ASAT_Pos_pPS | 27057 | 0.08693 | 1.51E-46 | 5.24E-46 |
| Hyper_Insulin_pPS | ALP_Neg_pPS | 27057 | -0.00291 | 0.632263419 | 0.757185878 |
| Hyper_Insulin_pPS | SHBG-LpA_pPS | 27057 | 0.021424 | 0.000424662 | 0.00083086 |
| Hyper_Insulin_pPS | Obesity_pPS | 27057 | 0.058747 | 4.00E-22 | 1.20E-21 |
| Hyper_Insulin_pPS | VAT_Neg_pPS | 27057 | 0.101343 | 1.07E-62 | 4.37E-62 |
| Hyper_Insulin_pPS | Bilirubin_pPS | 27057 | -0.00858 | 0.158335472 | 0.232846282 |
| Hyper_Insulin_pPS | Proinsulin_pPS | 27057 | 0.005355 | 0.378454262 | 0.486584051 |
| Hyper_Insulin_pPS | Hyper_Insulin_pPS | 27057 | 1 | 0 | 0 |
| Hyper_Insulin_pPS | Beta_Cell_1_pPS | 27057 | -0.00019 | 0.975107548 | 0.983852907 |
| Hyper_Insulin_pPS | Beta_Cell_2_pPS | 27057 | 0.008336 | 0.170321484 | 0.247240864 |
| Hyper_Insulin_pPS | Total_GRS | 27057 | 0.277836 | 0 | 0 |
| Beta_Cell_1_pPS | Cholesterol_pPS | 27057 | 0.007313 | 0.228992645 | 0.319410419 |
| Beta_Cell_1_pPS | Lipodystrophy_1_pPS | 27057 | 0.026096 | 1.76E-05 | 3.71E-05 |
| Beta_Cell_1_pPS | Lipodystrophy_2_pPS | 27057 | -0.0072 | 0.236271677 | 0.32218865 |
| Beta_Cell_1_pPS | Liver-Lipid_pPS | 27057 | 0.048981 | 7.55E-16 | 2.10E-15 |

**b) PPS Cross-Associations in Participants with Doctor-Diagnosed or Self-Reported Diabetes**

| PPS_Column1 | PPS_Column2 | Sample_Size | r | p | fdr_p_adjusted |
|---|---|---|---|---|---|
| SHBG-LpA_pPS | Lipodystrophy_2_pPS | 1256 | 0.04585 | 0.104344 | 0.223593931 |
| SHBG-LpA_pPS | Liver-Lipid_pPS | 1256 | 0.03262 | 0.247997 | 0.48521131 |
| SHBG-LpA_pPS | ASAT_Pos_pPS | 1256 | -0.00089 | 0.974898 | 0.97639817 |
| SHBG-LpA_pPS | ALP_Neg_pPS | 1256 | -0.03266 | 0.247402 | 0.48521131 |
| SHBG-LpA_pPS | SHBG-LpA_pPS | 1256 | 1 | 0 | 0 |
| SHBG-LpA_pPS | Obesity_pPS | 1256 | -0.0016 | 0.954763 | 0.972043728 |
| SHBG-LpA_pPS | VAT_Neg_pPS | 1256 | 0.060709 | 0.031446 | 0.083240662 |
| SHBG-LpA_pPS | Bilirubin_pPS | 1256 | -0.00287 | 0.919028 | 0.9617738 |
| SHBG-LpA_pPS | Proinsulin_pPS | 1256 | 0.031966 | 0.257615 | 0.489258376 |
| SHBG-LpA_pPS | Hyper_Insulin_pPS | 1256 | 0.047724 | 0.090912 | 0.20252757 |
| SHBG-LpA_pPS | Beta_Cell_1_pPS | 1256 | 0.050627 | 0.072878 | 0.172605999 |
| SHBG-LpA_pPS | Beta_Cell_2_pPS | 1256 | 0.004879 | 0.862844 | 0.9416461 |
| SHBG-LpA_pPS | Total_GRS | 1256 | 0.068246 | 0.015561 | 0.044319786 |
| Obesity_pPS | Cholesterol_pPS | 1256 | 0.056595 | 0.044928 | 0.113582875 |
| Obesity_pPS | Lipodystrophy_1_pPS | 1256 | 0.01367 | 0.628383 | 0.807921029 |
| Obesity_pPS | Lipodystrophy_2_pPS | 1256 | -0.0315 | 0.264567 | 0.489258376 |
| Obesity_pPS | Liver-Lipid_pPS | 1256 | -0.01959 | 0.487806 | 0.679801386 |
| Obesity_pPS | ASAT_Pos_pPS | 1256 | 0.076907 | 0.006393 | 0.020260328 |
| Obesity_pPS | ALP_Neg_pPS | 1256 | 0.010396 | 0.712811 | 0.876407545 |
| Obesity_pPS | SHBG-LpA_pPS | 1256 | -0.0016 | 0.954763 | 0.972043728 |
| Obesity_pPS | Obesity_pPS | 1256 | 1 | 0 | 0 |
| Obesity_pPS | VAT_Neg_pPS | 1256 | -0.01661 | 0.55652 | 0.740928962 |
| Obesity_pPS | Bilirubin_pPS | 1256 | -0.01267 | 0.653666 | 0.83093119 |
| Obesity_pPS | Proinsulin_pPS | 1256 | 0.136604 | 1.18E-06 | 5.64E-06 |
| Obesity_pPS | Hyper_Insulin_pPS | 1256 | 0.087673 | 0.001871 | 0.006899528 |
| Obesity_pPS | Beta_Cell_1_pPS | 1256 | 0.031404 | 0.266088 | 0.489258376 |
| Obesity_pPS | Beta_Cell_2_pPS | 1256 | -0.02556 | 0.365468 | 0.600221858 |
| Obesity_pPS | Total_GRS | 1256 | 0.354286 | 1.90E-38 | 1.58E-37 |
| VAT_Neg_pPS | Cholesterol_pPS | 1256 | 0.006029 | 0.83096 | 0.938155988 |
| VAT_Neg_pPS | Lipodystrophy_1_pPS | 1256 | 0.015513 | 0.582823 | 0.766872431 |
| VAT_Neg_pPS | Lipodystrophy_2_pPS | 1256 | 0.106543 | 0.000155 | 0.000611836 |
| VAT_Neg_pPS | Liver-Lipid_pPS | 1256 | 0.030191 | 0.284994 | 0.498316621 |
| VAT_Neg_pPS | ASAT_Pos_pPS | 1256 | 0.003093 | 0.912807 | 0.9617738 |
| VAT_Neg_pPS | ALP_Neg_pPS | 1256 | -0.02262 | 0.42324 | 0.651752464 |
| VAT_Neg_pPS | SHBG-LpA_pPS | 1256 | 0.060709 | 0.031446 | 0.083240662 |
| VAT_Neg_pPS | Obesity_pPS | 1256 | -0.01661 | 0.55652 | 0.740928962 |
| VAT_Neg_pPS | VAT_Neg_pPS | 1256 | 1 | 0 | 0 |
| VAT_Neg_pPS | Bilirubin_pPS | 1256 | 0.004755 | 0.866314 | 0.9416461 |
| VAT_Neg_pPS | Proinsulin_pPS | 1256 | 0.052806 | 0.061359 | 0.148448513 |
| VAT_Neg_pPS | Hyper_Insulin_pPS | 1256 | 0.114233 | 4.96E-05 | 0.000210414 |
| VAT_Neg_pPS | Beta_Cell_1_pPS | 1256 | 0.164744 | 4.28E-09 | 2.35E-08 |
| VAT_Neg_pPS | Beta_Cell_2_pPS | 1256 | 0.13372 | 1.98E-06 | 9.08E-06 |
| VAT_Neg_pPS | Total_GRS | 1256 | 0.249207 | 3.11E-19 | 2.12E-18 |
| Bilirubin_pPS | Cholesterol_pPS | 1256 | 0.005607 | 0.842651 | 0.938155988 |
| Bilirubin_pPS | Lipodystrophy_1_pPS | 1256 | 0.008787 | 0.755718 | 0.890244 |
| Bilirubin_pPS | Lipodystrophy_2_pPS | 1256 | -0.02034 | 0.471374 | 0.67911204 |
| Bilirubin_pPS | Liver-Lipid_pPS | 1256 | 0.011324 | 0.688468 | 0.855829823 |
| Bilirubin_pPS | ASAT_Pos_pPS | 1256 | -0.01785 | 0.527405 | 0.710575928 |
| Bilirubin_pPS | ALP_Neg_pPS | 1256 | -0.0093 | 0.741946 | 0.888699624 |
| Bilirubin_pPS | SHBG-LpA_pPS | 1256 | -0.00287 | 0.919028 | 0.9617738 |
| Bilirubin_pPS | Obesity_pPS | 1256 | -0.01267 | 0.653666 | 0.83093119 |
| Bilirubin_pPS | VAT_Neg_pPS | 1256 | 0.004755 | 0.866314 | 0.9416461 |
| Bilirubin_pPS | Bilirubin_pPS | 1256 | 1 | 0 | 0 |
| Bilirubin_pPS | Proinsulin_pPS | 1256 | 0.030669 | 0.27744 | 0.498316621 |
| Bilirubin_pPS | Hyper_Insulin_pPS | 1256 | -0.03015 | 0.285702 | 0.498316621 |
| Bilirubin_pPS | Beta_Cell_1_pPS | 1256 | -0.00547 | 0.846425 | 0.938155988 |
| Bilirubin_pPS | Beta_Cell_2_pPS | 1256 | -0.00681 | 0.809442 | 0.924952884 |
| Bilirubin_pPS | Total_GRS | 1256 | 0.017987 | 0.524195 | 0.710575928 |
| Proinsulin_pPS | Cholesterol_pPS | 1256 | 0.006796 | 0.809848 | 0.924952884 |
| Proinsulin_pPS | Lipodystrophy_1_pPS | 1256 | -0.04908 | 0.082103 | 0.186597242 |
| Proinsulin_pPS | Lipodystrophy_2_pPS | 1256 | -0.02612 | 0.354941 | 0.591568053 |
| Proinsulin_pPS | Liver-Lipid_pPS | 1256 | 0.045405 | 0.107754 | 0.226585242 |
| Proinsulin_pPS | ASAT_Pos_pPS | 1256 | 0.003509 | 0.901116 | 0.960906067 |
| Proinsulin_pPS | ALP_Neg_pPS | 1256 | 0.150027 | 9.20E-08 | 4.60E-07 |
| Proinsulin_pPS | SHBG-LpA_pPS | 1256 | 0.031966 | 0.257615 | 0.489258376 |
| Proinsulin_pPS | Obesity_pPS | 1256 | 0.136604 | 1.18E-06 | 5.64E-06 |
| Proinsulin_pPS | VAT_Neg_pPS | 1256 | 0.052806 | 0.061359 | 0.148448513 |
| Proinsulin_pPS | Bilirubin_pPS | 1256 | 0.030669 | 0.27744 | 0.498316621 |
| Proinsulin_pPS | Proinsulin_pPS | 1256 | 1 | 0 | 0 |
| Proinsulin_pPS | Hyper_Insulin_pPS | 1256 | 0.031314 | 0.267461 | 0.489258376 |
| Proinsulin_pPS | Beta_Cell_1_pPS | 1256 | 0.376046 | 1.82E-43 | 1.63E-42 |
| Proinsulin_pPS | Beta_Cell_2_pPS | 1256 | 0.195767 | 2.58E-12 | 1.49E-11 |
| Proinsulin_pPS | Total_GRS | 1256 | 0.348291 | 3.93E-37 | 3.05E-36 |
| Hyper_Insulin_pPS | Cholesterol_pPS | 1256 | 0.00924 | 0.743561 | 0.888699624 |
| Hyper_Insulin_pPS | Lipodystrophy_1_pPS | 1256 | 0.109084 | 0.000107 | 0.000438571 |
| Hyper_Insulin_pPS | Lipodystrophy_2_pPS | 1256 | 0.104035 | 0.000221 | 0.00084399 |
| Hyper_Insulin_pPS | Liver-Lipid_pPS | 1256 | 0.022021 | 0.435542 | 0.651752464 |
| Hyper_Insulin_pPS | ASAT_Pos_pPS | 1256 | 0.086055 | 0.00227 | 0.007857957 |
| Hyper_Insulin_pPS | ALP_Neg_pPS | 1256 | 0.003806 | 0.892799 | 0.960906067 |
| Hyper_Insulin_pPS | SHBG-LpA_pPS | 1256 | 0.047724 | 0.090912 | 0.20252757 |
| Hyper_Insulin_pPS | Obesity_pPS | 1256 | 0.087673 | 0.001871 | 0.006899528 |
| Hyper_Insulin_pPS | VAT_Neg_pPS | 1256 | 0.114233 | 4.96E-05 | 0.000210414 |
| Hyper_Insulin_pPS | Bilirubin_pPS | 1256 | -0.03015 | 0.285702 | 0.498316621 |
| Hyper_Insulin_pPS | Proinsulin_pPS | 1256 | 0.031314 | 0.267461 | 0.489258376 |
| Hyper_Insulin_pPS | Hyper_Insulin_pPS | 1256 | 1 | 0 | 0 |
| Hyper_Insulin_pPS | Beta_Cell_1_pPS | 1256 | 0.029353 | 0.298583 | 0.512832553 |
| Hyper_Insulin_pPS | Beta_Cell_2_pPS | 1256 | -0.01199 | 0.671072 | 0.843525717 |
| Hyper_Insulin_pPS | Total_GRS | 1256 | 0.271773 | 1.05E-22 | 7.61E-22 |
| Beta_Cell_1_pPS | Cholesterol_pPS | 1256 | -0.0597 | 0.03439 | 0.088940185 |
| Beta_Cell_1_pPS | Lipodystrophy_1_pPS | 1256 | 0.024148 | 0.3925 | 0.635342177 |
| Beta_Cell_1_pPS | Lipodystrophy_2_pPS | 1256 | 0.000836 | 0.976398 | 0.97639817 |
| Beta_Cell_1_pPS | Liver-Lipid_pPS | 1256 | 0.062987 | 0.025598 | 0.071104785 |

**Table S3: pPS cross-correlation in SCAPIS**

**a) PPS Cross-Associations in Participants without Doctor-Diagnosed or Self-Reported Diabetes**

| PPS_Column1 | PPS_Column2 | Sample_Size | r | p | fdr_p_adjusted |
|---|---|---|---|---|---|
| Beta_Cell_1_pPS | ASAT_Pos_pPS | 27057 | 0.034316 | 1.64E-08 | 3.81E-08 |
| Beta_Cell_1_pPS | ALP_Neg_pPS | 27057 | 0.009835 | 0.105712804 | 0.16403711 |
| Beta_Cell_1_pPS | SHBG-LpA_pPS | 27057 | 0.018041 | 0.003000099 | 0.005578697 |
| Beta_Cell_1_pPS | Obesity_pPS | 27057 | 0.02701 | 8.85E-06 | 1.90E-05 |
| Beta_Cell_1_pPS | VAT_Neg_pPS | 27057 | 0.116957 | 5.01E-83 | 2.30E-82 |
| Beta_Cell_1_pPS | Bilirubin_pPS | 27057 | 0.028371 | 3.05E-06 | 6.79E-06 |
| Beta_Cell_1_pPS | Proinsulin_pPS | 27057 | 0.302096 | 0 | 0 |
| Beta_Cell_1_pPS | Hyper_Insulin_pPS | 27057 | -0.00019 | 0.975107548 | 0.983852907 |
| Beta_Cell_1_pPS | Beta_Cell_1_pPS | 27057 | 1 | 0 | 0 |
| Beta_Cell_1_pPS | Beta_Cell_2_pPS | 27057 | 0.235737 | 0 | 0 |
| Beta_Cell_1_pPS | Total_GRS | 27057 | 0.35974 | 0 | 0 |
| Beta_Cell_2_pPS | Cholesterol_pPS | 27057 | 0.008599 | 0.157238341 | 0.232846282 |
| Beta_Cell_2_pPS | Lipodystrophy_1_pPS | 27057 | 0.002877 | 0.636036137 | 0.757185878 |
| Beta_Cell_2_pPS | Lipodystrophy_2_pPS | 27057 | 0.001535 | 0.8007066 | 0.847444222 |
| Beta_Cell_2_pPS | Liver-Lipid_pPS | 27057 | 0.039018 | 1.36E-10 | 3.44E-10 |
| Beta_Cell_2_pPS | ASAT_Pos_pPS | 27057 | -3.50E-05 | 0.995402312 | 0.995402312 |
| Beta_Cell_2_pPS | ALP_Neg_pPS | 27057 | 0.019244 | 0.00154727 | 0.002975518 |
| Beta_Cell_2_pPS | SHBG-LpA_pPS | 27057 | 0.003193 | 0.599484109 | 0.729102294 |
| Beta_Cell_2_pPS | Obesity_pPS | 27057 | 0.005654 | 0.352408534 | 0.46369544 |
| Beta_Cell_2_pPS | VAT_Neg_pPS | 27057 | 0.087641 | 2.77E-47 | 9.89E-47 |
| Beta_Cell_2_pPS | Bilirubin_pPS | 27057 | -0.00335 | 0.581778388 | 0.715301297 |
| Beta_Cell_2_pPS | Proinsulin_pPS | 27057 | 0.170536 | 1.16E-175 | 6.35E-175 |
| Beta_Cell_2_pPS | Hyper_Insulin_pPS | 27057 | 0.008336 | 0.170321484 | 0.247240864 |
| Beta_Cell_2_pPS | Beta_Cell_1_pPS | 27057 | 0.235737 | 0 | 0 |
| Beta_Cell_2_pPS | Beta_Cell_2_pPS | 27057 | 1 | 0 | 0 |
| Beta_Cell_2_pPS | Total_GRS | 27057 | 0.38974 | 0 | 0 |
| Total_GRS | Cholesterol_pPS | 27057 | 0.117023 | 4.05E-83 | 1.94E-82 |
| Total_GRS | Lipodystrophy_1_pPS | 27057 | 0.354758 | 0 | 0 |
| Total_GRS | Lipodystrophy_2_pPS | 27057 | 0.413784 | 0 | 0 |
| Total_GRS | Liver-Lipid_pPS | 27057 | 0.088281 | 5.92E-48 | 2.18E-47 |
| Total_GRS | ASAT_Pos_pPS | 27057 | 0.231962 | 0 | 0 |
| Total_GRS | ALP_Neg_pPS | 27057 | 0.084748 | 2.56E-44 | 8.60E-44 |
| Total_GRS | SHBG-LpA_pPS | 27057 | 0.050081 | 1.69E-16 | 4.80E-16 |
| Total_GRS | Obesity_pPS | 27057 | 0.390167 | 0 | 0 |
| Total_GRS | VAT_Neg_pPS | 27057 | 0.222481 | 1.25E-300 | 7.62E-300 |
| Total_GRS | Bilirubin_pPS | 27057 | 0.016978 | 0.005224497 | 0.009404094 |
| Total_GRS | Proinsulin_pPS | 27057 | 0.369098 | 0 | 0 |
| Total_GRS | Hyper_Insulin_pPS | 27057 | 0.277836 | 0 | 0 |
| Total_GRS | Beta_Cell_1_pPS | 27057 | 0.35974 | 0 | 0 |
| Total_GRS | Beta_Cell_2_pPS | 27057 | 0.38974 | 0 | 0 |
| Total_GRS | Total_GRS | 27057 | 1 | 0 | 0 |

**b) PPS Cross-Associations in Participants with Doctor-Diagnosed or Self-Reported Diabetes**

| PPS_Column1 | PPS_Column2 | Sample_Size | r | p | fdr_p_adjusted |
|---|---|---|---|---|---|
| Beta_Cell_1_pPS | ASAT_Pos_pPS | 1256 | 0.019435 | 0.49135 | 0.679801386 |
| Beta_Cell_1_pPS | ALP_Neg_pPS | 1256 | 0.085133 | 0.002531 | 0.008500178 |
| Beta_Cell_1_pPS | SHBG-LpA_pPS | 1256 | 0.050627 | 0.072878 | 0.172605999 |
| Beta_Cell_1_pPS | Obesity_pPS | 1256 | 0.031404 | 0.266088 | 0.489258376 |
| Beta_Cell_1_pPS | VAT_Neg_pPS | 1256 | 0.164744 | 4.28E-09 | 2.35E-08 |
| Beta_Cell_1_pPS | Bilirubin_pPS | 1256 | -0.00547 | 0.846425 | 0.938155988 |
| Beta_Cell_1_pPS | Proinsulin_pPS | 1256 | 0.376046 | 1.82E-43 | 1.63E-42 |
| Beta_Cell_1_pPS | Hyper_Insulin_pPS | 1256 | 0.029353 | 0.298583 | 0.512832553 |
| Beta_Cell_1_pPS | Beta_Cell_1_pPS | 1256 | 1 | 0 | 0 |
| Beta_Cell_1_pPS | Beta_Cell_2_pPS | 1256 | 0.245659 | 1.02E-18 | 6.54E-18 |
| Beta_Cell_1_pPS | Total_GRS | 1256 | 0.378993 | 3.54E-44 | 3.79E-43 |
| Beta_Cell_2_pPS | Cholesterol_pPS | 1256 | -0.01938 | 0.492478 | 0.679801386 |
| Beta_Cell_2_pPS | Lipodystrophy_1_pPS | 1256 | -0.02138 | 0.449027 | 0.660333913 |
| Beta_Cell_2_pPS | Lipodystrophy_2_pPS | 1256 | 0.021932 | 0.437398 | 0.651752464 |
| Beta_Cell_2_pPS | Liver-Lipid_pPS | 1256 | 0.036505 | 0.196048 | 0.397394279 |
| Beta_Cell_2_pPS | ASAT_Pos_pPS | 1256 | -0.05563 | 0.0487 | 0.120412274 |
| Beta_Cell_2_pPS | ALP_Neg_pPS | 1256 | 0.023445 | 0.406434 | 0.639493899 |
| Beta_Cell_2_pPS | SHBG-LpA_pPS | 1256 | 0.004879 | 0.862844 | 0.9416461 |
| Beta_Cell_2_pPS | Obesity_pPS | 1256 | -0.02556 | 0.365468 | 0.600221858 |
| Beta_Cell_2_pPS | VAT_Neg_pPS | 1256 | 0.13372 | 1.98E-06 | 9.08E-06 |
| Beta_Cell_2_pPS | Bilirubin_pPS | 1256 | -0.00681 | 0.809442 | 0.924952884 |
| Beta_Cell_2_pPS | Proinsulin_pPS | 1256 | 0.195767 | 2.58E-12 | 1.49E-11 |
| Beta_Cell_2_pPS | Hyper_Insulin_pPS | 1256 | -0.01199 | 0.671072 | 0.843525717 |
| Beta_Cell_2_pPS | Beta_Cell_1_pPS | 1256 | 0.245659 | 1.02E-18 | 6.54E-18 |
| Beta_Cell_2_pPS | Beta_Cell_2_pPS | 1256 | 1 | 0 | 0 |
| Beta_Cell_2_pPS | Total_GRS | 1256 | 0.386217 | 5.99E-46 | 7.09E-45 |
| Total_GRS | Cholesterol_pPS | 1256 | 0.049938 | 0.076868 | 0.178303114 |
| Total_GRS | Lipodystrophy_1_pPS | 1256 | 0.377446 | 8.37E-44 | 8.19E-43 |
| Total_GRS | Lipodystrophy_2_pPS | 1256 | 0.424215 | 5.00E-56 | 6.61E-55 |
| Total_GRS | Liver-Lipid_pPS | 1256 | 0.115883 | 3.84E-05 | 0.000169576 |
| Total_GRS | ASAT_Pos_pPS | 1256 | 0.206737 | 1.37E-13 | 8.31E-13 |
| Total_GRS | ALP_Neg_pPS | 1256 | 0.046554 | 0.099124 | 0.21653349 |
| Total_GRS | SHBG-LpA_pPS | 1256 | 0.068246 | 0.015561 | 0.044319786 |
| Total_GRS | Obesity_pPS | 1256 | 0.354286 | 1.90E-38 | 1.58E-37 |
| Total_GRS | VAT_Neg_pPS | 1256 | 0.249207 | 3.11E-19 | 2.12E-18 |
| Total_GRS | Bilirubin_pPS | 1256 | 0.017987 | 0.524195 | 0.710575928 |
| Total_GRS | Proinsulin_pPS | 1256 | 0.348291 | 3.93E-37 | 3.05E-36 |
| Total_GRS | Hyper_Insulin_pPS | 1256 | 0.271773 | 1.05E-22 | 7.61E-22 |
| Total_GRS | Beta_Cell_1_pPS | 1256 | 0.378993 | 3.54E-44 | 3.79E-43 |
| Total_GRS | Beta_Cell_2_pPS | 1256 | 0.386217 | 5.99E-46 | 7.09E-45 |
| Total_GRS | Total_GRS | 1256 | 1 | 0 | 0 |

**Table S4: Associations between pPSs and anthropometric traits in SCAPIS**

**a) Spearman Correlation**

| Anthropometry | PPS | Sample_Size | r | CI95% | p | fdr_p_adjusted |
|---|---|---|---|---|---|---|
| Waist/hip_Ratio | Cholesterol_pPS | 27030 | 0.007547427 | [-0. 0.02] | 0.214774827 | 0.406330107 |
| Waist/hip_Ratio | Lipodystrophy_1_pPS | 27030 | 0.025833458 | [0.01 0.04] | 2.17E-05 | 0.000243954 |
| Waist/hip_Ratio | Lipodystrophy_2_pPS | 27030 | 0.006716191 | [-0.01 0.02] | 0.269631318 | 0.460762379 |
| Waist/hip_Ratio | Liver-Lipid_pPS | 27030 | 0.013308118 | [0. 0.03] | 0.028708133 | 0.082459531 |
| Waist/hip_Ratio | ASAT_Pos_pPS | 27030 | 0.023188931 | [0.01 0.04] | 0.000137882 | 0.001094949 |
| Waist/hip_Ratio | ALP_Neg_pPS | 27030 | 0.014812262 | [0. 0.03] | 0.014902698 | 0.050296604 |
| Waist/hip_Ratio | SHBG-LpA_pPS | 27030 | 0.006931652 | [-0. 0.02] | 0.254566709 | 0.446318256 |
| Waist/hip_Ratio | Obesity_pPS | 27030 | 0.052013708 | [0.04 0.06] | 1.18E-17 | 3.18E-16 |
| Waist/hip_Ratio | VAT_Neg_pPS | 27030 | -0.009372702 | [-0.02 0. ] | 0.123421989 | 0.252454068 |
| Waist/hip_Ratio | Bilirubin_pPS | 27030 | -0.013712615 | [-0.03 -0. ] | 0.024198355 | 0.072595066 |
| Waist/hip_Ratio | Proinsulin_pPS | 27030 | 0.007434848 | [-0. 0.02] | 0.221693347 | 0.409980847 |
| Waist/hip_Ratio | Hyper_Insulin_pPS | 27030 | 0.015320811 | [0. 0.03] | 0.011791063 | 0.045479815 |
| Waist/hip_Ratio | Beta_Cell_1_pPS | 27030 | 0.014285696 | [0. 0.03] | 0.018865762 | 0.059229717 |
| Waist/hip_Ratio | Beta_Cell_2_pPS | 27030 | 0.000570112 | [-0.01 0.01] | 0.925342874 | 0.953597618 |
| Waist/hip_Ratio | Total_GRS | 27030 | 0.036376622 | [0.02 0.05] | 2.22E-09 | 4.28E-08 |
| BMI | Cholesterol_pPS | 27055 | 0.007254391 | [-0. 0.02] | 0.232897787 | 0.419216017 |
| BMI | Lipodystrophy_1_pPS | 27055 | -0.024139871 | [-0.04 -0.01] | 7.18E-05 | 0.000606074 |
| BMI | Lipodystrophy_2_pPS | 27055 | -0.003673598 | [-0.02 0.01] | 0.545784904 | 0.708470789 |
| BMI | Liver-Lipid_pPS | 27055 | 0.017189577 | [0.01 0.03] | 0.004700655 | 0.024602931 |
| BMI | ASAT_Pos_pPS | 27055 | -0.004835066 | [-0.02 0.01] | 0.426565388 | 0.600510621 |
| BMI | ALP_Neg_pPS | 27055 | 0.008662541 | [-0. 0.02] | 0.15430335 | 0.301897859 |
| BMI | SHBG-LpA_pPS | 27055 | -0.006523273 | [-0.02 0.01] | 0.283406542 | 0.466583941 |
| BMI | Obesity_pPS | 27055 | 0.101218354 | [0.09 0.11] | 1.62E-62 | 2.09E-60 |
| BMI | VAT_Neg_pPS | 27055 | 0.005325236 | [-0.01 0.02] | 0.381199748 | 0.56551611 |
| BMI | Bilirubin_pPS | 27055 | -0.005247575 | [-0.02 0.01] | 0.388183169 | 0.569616607 |
| BMI | Proinsulin_pPS | 27055 | 0.016444583 | [0. 0.03] | 0.006844241 | 0.032387395 |
| BMI | Hyper_Insulin_pPS | 27055 | 0.016433145 | [0. 0.03] | 0.00688309 | 0.032387395 |
| BMI | Beta_Cell_1_pPS | 27055 | 0.009517202 | [-0. 0.02] | 0.117572645 | 0.244189339 |
| BMI | Beta_Cell_2_pPS | 27055 | 5.21E-05 | [-0.01 0.01] | 0.993158253 | 0.993158253 |
| BMI | Total_GRS | 27055 | 0.031377188 | [0.02 0.04] | 2.46E-07 | 4.11E-06 |
| Waist | Cholesterol_pPS | 27046 | 0.007256971 | [-0. 0.02] | 0.232809394 | 0.419216017 |
| Waist | Lipodystrophy_1_pPS | 27046 | -0.013245014 | [-0.03 -0. ] | 0.029425212 | 0.08275841 |
| Waist | Lipodystrophy_2_pPS | 27046 | -0.00555877 | [-0.02 0.01] | 0.360748954 | 0.54720347 |
| Waist | Liver-Lipid_pPS | 27046 | 0.019254444 | [0.01 0.03] | 0.001545686 | 0.009936555 |
| Waist | ASAT_Pos_pPS | 27046 | 0.006238883 | [-0.01 0.02] | 0.305004278 | 0.486618816 |
| Waist | ALP_Neg_pPS | 27046 | 0.012382934 | [0. 0.02] | 0.041751523 | 0.108393376 |
| Waist | SHBG-LpA_pPS | 27046 | -0.004081055 | [-0.02 0.01] | 0.50223355 | 0.658267274 |
| Waist | Obesity_pPS | 27046 | 0.091392732 | [0.08 0.1 ] | 3.08E-51 | 1.38E-49 |
| Waist | VAT_Neg_pPS | 27046 | -0.009018525 | [-0.02 0. ] | 0.138130409 | 0.278322465 |
| Waist | Bilirubin_pPS | 27046 | -0.009546754 | [-0.02 0. ] | 0.116498328 | 0.244189339 |
| Waist | Proinsulin_pPS | 27046 | 0.017176882 | [0.01 0.03] | 0.004738342 | 0.024602931 |
| Waist | Hyper_Insulin_pPS | 27046 | 0.011964342 | [0. 0.02] | 0.049164896 | 0.117157587 |
| Waist | Beta_Cell_1_pPS | 27046 | 0.015474925 | [0. 0.03] | 0.010945786 | 0.044778214 |
| Waist | Beta_Cell_2_pPS | 27046 | -5.37E-05 | [-0.01 0.01] | 0.99296021 | 0.993158253 |
| Waist | Total_GRS | 27046 | 0.031259384 | [0.02 0.04] | 2.74E-07 | 4.11E-06 |
| Hip | Cholesterol_pPS | 27034 | 0.001161386 | [-0.01 0.01] | 0.848600657 | 0.917447633 |
| Hip | Lipodystrophy_1_pPS | 27034 | -0.042171608 | [-0.05 -0.03] | 4.06E-12 | 9.13E-11 |
| Hip | Lipodystrophy_2_pPS | 27034 | -0.012076989 | [-0.02 -0. ] | 0.047117707 | 0.117157587 |
| Hip | Liver-Lipid_pPS | 27034 | 0.014899699 | [0. 0.03] | 0.014313807 | 0.049547794 |
| Hip | ASAT_Pos_pPS | 27034 | -0.014911942 | [-0.03 -0. ] | 0.014234004 | 0.049547794 |
| Hip | ALP_Neg_pPS | 27034 | 0.000410113 | [-0.01 0.01] | 0.946253144 | 0.961028951 |
| Hip | SHBG-LpA_pPS | 27034 | -0.010482085 | [-0.02 0. ] | 0.084879209 | 0.184817633 |
| Hip | Obesity_pPS | 27034 | 0.086904758 | [0.08 0.1 ] | 1.84E-46 | 6.20E-45 |
| Hip | VAT_Neg_pPS | 27034 | -0.004812343 | [-0.02 0.01] | 0.428921396 | 0.600510621 |
| Hip | Bilirubin_pPS | 27034 | -0.000932334 | [-0.01 0.01] | 0.878198445 | 0.933518032 |
| Hip | Proinsulin_pPS | 27034 | 0.015192824 | [0. 0.03] | 0.012507913 | 0.046904673 |
| Hip | Hyper_Insulin_pPS | 27034 | 0.004290655 | [-0.01 0.02] | 0.480633247 | 0.642430577 |
| Hip | Beta_Cell_1_pPS | 27034 | 0.007929472 | [-0. 0.02] | 0.19242591 | 0.371107112 |
| Hip | Beta_Cell_2_pPS | 27034 | -0.003133606 | [-0.02 0.01] | 0.606489301 | 0.751156474 |
| Hip | Total_GRS | 27034 | 0.01021943 | [-0. 0.02] | 0.092981872 | 0.199246868 |
| Weight | Cholesterol_pPS | 27055 | 0.003261388 | [-0.01 0.02] | 0.591748543 | 0.746598629 |
| Weight | Lipodystrophy_1_pPS | 27055 | -0.02929067 | [-0.04 -0.02] | 1.45E-06 | 1.96E-05 |
| Weight | Lipodystrophy_2_pPS | 27055 | -0.011946477 | [-0.02 -0. ] | 0.049466537 | 0.117157587 |
| Weight | Liver-Lipid_pPS | 27055 | 0.025585049 | [0.01 0.04] | 2.58E-05 | 0.000267677 |
| Weight | ASAT_Pos_pPS | 27055 | -0.014391156 | [-0.03 -0. ] | 0.017952145 | 0.058245536 |
| Weight | ALP_Neg_pPS | 27055 | 0.006551505 | [-0.01 0.02] | 0.281328028 | 0.466583941 |
| Weight | SHBG-LpA_pPS | 27055 | -0.006207437 | [-0.02 0.01] | 0.307367949 | 0.486618816 |
| Weight | Obesity_pPS | 27055 | 0.100985966 | [0.09 0.11] | 3.09E-62 | 2.09E-60 |
| Weight | VAT_Neg_pPS | 27055 | -0.004919467 | [-0.02 0.01] | 0.418537052 | 0.600510621 |
| Weight | Bilirubin_pPS | 27055 | -0.002980209 | [-0.01 0.01] | 0.62408679 | 0.75224747 |
| Weight | Proinsulin_pPS | 27055 | 0.015572244 | [0. 0.03] | 0.010441512 | 0.044050127 |
| Weight | Hyper_Insulin_pPS | 27055 | 0.012665037 | [0. 0.02] | 0.037276592 | 0.102700816 |
| Weight | Beta_Cell_1_pPS | 27055 | 0.012175891 | [0. 0.02] | 0.045254932 | 0.115271996 |
| Weight | Beta_Cell_2_pPS | 27055 | -0.001783429 | [-0.01 0.01] | 0.769317861 | 0.876847022 |
| Weight | Total_GRS | 27055 | 0.019745097 | [0.01 0.03] | 0.001165466 | 0.007866899 |
| Height | Cholesterol_pPS | 27055 | -0.004404323 | [-0.02 0.01] | 0.468912106 | 0.633031342 |
| Height | Lipodystrophy_1_pPS | 27055 | -0.012459329 | [-0.02 -0. ] | 0.04047277 | 0.107133802 |
| Height | Lipodystrophy_2_pPS | 27055 | -0.012612367 | [-0.02 -0. ] | 0.038073772 | 0.102799184 |
| Height | Liver-Lipid_pPS | 27055 | 0.017501741 | [0.01 0.03] | 0.003999439 | 0.023474966 |
| Height | ASAT_Pos_pPS | 27055 | -0.024386274 | [-0.04 -0.01] | 6.05E-05 | 0.00054491 |
| Height | ALP_Neg_pPS | 27055 | -0.000405841 | [-0.01 0.01] | 0.946791485 | 0.961028951 |
| Height | SHBG-LpA_pPS | 27055 | -0.003171884 | [-0.02 0.01] | 0.601958728 | 0.751156474 |
| Height | Obesity_pPS | 27055 | 0.016050926 | [0. 0.03] | 0.008300795 | 0.036758374 |
| Height | VAT_Neg_pPS | 27055 | -0.021160445 | [-0.03 -0.01] | 0.000501391 | 0.003760431 |
| Height | Bilirubin_pPS | 27055 | 0.002995768 | [-0.01 0.01] | 0.622277528 | 0.75224747 |
| Height | Proinsulin_pPS | 27055 | 0.004955781 | [-0.01 0.02] | 0.415110464 | 0.600510621 |
| Height | Hyper_Insulin_pPS | 27055 | -0.002556682 | [-0.01 0.01] | 0.674177632 | 0.798368249 |
| Height | Beta_Cell_1_pPS | 27055 | 0.005719669 | [-0.01 0.02] | 0.346935632 | 0.533746085 |
| Height | Beta_Cell_2_pPS | 27055 | -0.004192492 | [-0.02 0.01] | 0.49056141 | 0.649272455 |
| Height | Total_GRS | 27055 | -0.01641146 | [-0.03 -0. ] | 0.006957292 | 0.032387395 |
| DBP_Mean | Cholesterol_pPS | 26976 | -0.00340312 | [-0.02 0.01] | 0.576303336 | 0.733971229 |
| DBP_Mean | Lipodystrophy_1_pPS | 26976 | 0.011198807 | [-0. 0.02] | 0.065930328 | 0.148343239 |
| DBP_Mean | Lipodystrophy_2_pPS | 26976 | 0.006182869 | [-0.01 0.02] | 0.309994209 | 0.486618816 |
| DBP_Mean | Liver-Lipid_pPS | 26976 | 0.004494933 | [-0.01 0.02] | 0.460472046 | 0.627919168 |
| DBP_Mean | ASAT_Pos_pPS | 26976 | 0.011477387 | [-0. 0.02] | 0.059477894 | 0.136093486 |
| DBP_Mean | ALP_Neg_pPS | 26976 | -0.008745055 | [-0.02 0. ] | 0.151013201 | 0.29980562 |
| DBP_Mean | SHBG-LpA_pPS | 26976 | -0.001706671 | [-0.01 0.01] | 0.779297384 | 0.876847022 |
| DBP_Mean | Obesity_pPS | 26976 | 0.027188079 | [0.02 0.04] | 8.00E-06 | 9.82E-05 |
| DBP_Mean | VAT_Neg_pPS | 26976 | 0.005434057 | [-0.01 0.02] | 0.372244819 | 0.558367229 |
| DBP_Mean | Bilirubin_pPS | 26976 | -0.005716319 | [-0.02 0.01] | 0.347923374 | 0.533746085 |
| DBP_Mean | Proinsulin_pPS | 26976 | 0.007121493 | [-0. 0.02] | 0.24225823 | 0.430327118 |
| DBP_Mean | Hyper_Insulin_pPS | 26976 | 0.013893326 | [0. 0.03] | 0.022525483 | 0.069112279 |
| DBP_Mean | Beta_Cell_1_pPS | 26976 | 0.007523211 | [-0. 0.02] | 0.21670939 | 0.406330107 |
| DBP_Mean | Beta_Cell_2_pPS | 26976 | -0.013576551 | [-0.03 -0. ] | 0.025789519 | 0.075686633 |
| DBP_Mean | Total_GRS | 26976 | 0.01439113 | [0. 0.03] | 0.018120834 | 0.058245536 |
| SBP_Mean | Cholesterol_pPS | 26978 | -0.006259606 | [-0.02 0.01] | 0.304009691 | 0.486618816 |
| SBP_Mean | Lipodystrophy_1_pPS | 26978 | 0.014981944 | [0. 0.03] | 0.013883715 | 0.049547794 |

**b) Partial Correlation**

| Anthropometry | PPS | Sample_Size | r | CI95% | p | fdr_p_adjusted |
|---|---|---|---|---|---|---|
| Waist/hip_Ratio | Cholesterol_pPS | 27030 | 0.003069094 | [-0.01 0.01] | 0.614030856 | 0.794844847 |
| Waist/hip_Ratio | Lipodystrophy_1_pPS | 27030 | 0.019428629 | [0.01 0.03] | 0.001408524 | 0.011831602 |
| Waist/hip_Ratio | Lipodystrophy_2_pPS | 27030 | 0.004174864 | [-0.01 0.02] | 0.492691121 | 0.740718687 |
| Waist/hip_Ratio | Liver-Lipid_pPS | 27030 | 0.010562134 | [-0. 0.02] | 0.082623173 | 0.273961048 |
| Waist/hip_Ratio | ASAT_Pos_pPS | 27030 | 0.016116945 | [0. 0.03] | 0.008083098 | 0.048129329 |
| Waist/hip_Ratio | ALP_Neg_pPS | 27030 | 0.013390644 | [0. 0.03] | 0.027770887 | 0.112875218 |
| Waist/hip_Ratio | SHBG-LpA_pPS | 27030 | 0.006542795 | [-0.01 0.02] | 0.282306627 | 0.583125165 |
| Waist/hip_Ratio | Obesity_pPS | 27030 | 0.049442232 | [0.04 0.06] | 4.31E-16 | 1.09E-14 |
| Waist/hip_Ratio | VAT_Neg_pPS | 27030 | -0.012913831 | [-0.02 -0. ] | 0.0338255 | 0.129151908 |
| Waist/hip_Ratio | Bilirubin_pPS | 27030 | -0.013953337 | [-0.03 -0. ] | 0.021848865 | 0.097443079 |
| Waist/hip_Ratio | Proinsulin_pPS | 27030 | -0.00561605 | [-0.02 0.01] | 0.356079584 | 0.644518049 |
| Waist/hip_Ratio | Hyper_Insulin_pPS | 27030 | 0.008962925 | [-0. 0.02] | 0.140789717 | 0.369573008 |
| Waist/hip_Ratio | Beta_Cell_1_pPS | 27030 | 0.014121854 | [0. 0.03] | 0.020304089 | 0.094752417 |
| Waist/hip_Ratio | Beta_Cell_2_pPS | 27030 | -0.002398995 | [-0.01 0.01] | 0.693423082 | 0.848265129 |
| BMI | Cholesterol_pPS | 27055 | -0.000428808 | [-0.01 0.01] | 0.943798766 | 0.973965573 |
| BMI | Lipodystrophy_1_pPS | 27055 | -0.026309792 | [-0.04 -0.01] | 1.52E-05 | 0.000239053 |
| BMI | Lipodystrophy_2_pPS | 27055 | -0.003960369 | [-0.02 0.01] | 0.514989066 | 0.740718687 |
| BMI | Liver-Lipid_pPS | 27055 | 0.015468082 | [0. 0.03] | 0.010987021 | 0.06018977 |
| BMI | ASAT_Pos_pPS | 27055 | -0.006110272 | [-0.02 0.01] | 0.315118041 | 0.620388644 |
| BMI | ALP_Neg_pPS | 27055 | 0.005708685 | [-0.01 0.02] | 0.347978339 | 0.644518049 |
| BMI | SHBG-LpA_pPS | 27055 | -0.003979656 | [-0.02 0.01] | 0.512944506 | 0.740718687 |
| BMI | Obesity_pPS | 27055 | 0.099279138 | [0.09 0.11] | 3.57E-60 | 2.25E-58 |
| BMI | VAT_Neg_pPS | 27055 | 0.004326792 | [-0.01 0.02] | 0.476879896 | 0.740718687 |
| BMI | Bilirubin_pPS | 27055 | -0.005276106 | [-0.02 0.01] | 0.385722806 | 0.65214261 |
| BMI | Proinsulin_pPS | 27055 | -0.000222647 | [-0.01 0.01] | 0.9708013 | 0.973965573 |
| BMI | Hyper_Insulin_pPS | 27055 | 0.01342268 | [0. 0.03] | 0.027328479 | 0.112875218 |
| BMI | Beta_Cell_1_pPS | 27055 | 0.006939883 | [-0. 0.02] | 0.253897612 | 0.533184986 |
| BMI | Beta_Cell_2_pPS | 27055 | -0.003704721 | [-0.02 0.01] | 0.542484805 | 0.740718687 |
| Waist | Cholesterol_pPS | 27046 | 0.000260791 | [-0.01 0.01] | 0.965807513 | 0.973965573 |
| Waist | Lipodystrophy_1_pPS | 27046 | -0.016725173 | [-0.03 -0. ] | 0.005971176 | 0.040315172 |
| Waist | Lipodystrophy_2_pPS | 27046 | -0.005383729 | [-0.02 0.01] | 0.376185527 | 0.65214261 |
| Waist | Liver-Lipid_pPS | 27046 | 0.017246973 | [0.01 0.03] | 0.00458044 | 0.033949147 |
| Waist | ASAT_Pos_pPS | 27046 | 0.004234591 | [-0.01 0.02] | 0.486393863 | 0.740718687 |
| Waist | ALP_Neg_pPS | 27046 | 0.009433081 | [-0. 0.02] | 0.121002658 | 0.338807441 |
| Waist | SHBG-LpA_pPS | 27046 | -0.001504511 | [-0.01 0.01] | 0.804675147 | 0.897391577 |
| Waist | Obesity_pPS | 27046 | 0.088871292 | [0.08 0.1 ] | 1.63E-48 | 6.85E-47 |
| Waist | VAT_Neg_pPS | 27046 | -0.01041864 | [-0.02 0. ] | 0.086787963 | 0.280391881 |
| Waist | Bilirubin_pPS | 27046 | -0.009760011 | [-0.02 0. ] | 0.108643321 | 0.311114965 |
| Waist | Proinsulin_pPS | 27046 | -0.000356859 | [-0.01 0.01] | 0.953224531 | 0.973965573 |
| Waist | Hyper_Insulin_pPS | 27046 | 0.009142937 | [-0. 0.02] | 0.132869016 | 0.358535892 |
| Waist | Beta_Cell_1_pPS | 27046 | 0.013888777 | [0. 0.03] | 0.022427375 | 0.097443079 |
| Waist | Beta_Cell_2_pPS | 27046 | -0.004445608 | [-0.02 0.01] | 0.464936862 | 0.740718687 |
| Hip | Cholesterol_pPS | 27034 | -0.005131241 | [-0.02 0.01] | 0.399085302 | 0.661641421 |
| Hip | Lipodystrophy_1_pPS | 27034 | -0.04119039 | [-0.05 -0.03] | 1.27E-11 | 2.67E-10 |
| Hip | Lipodystrophy_2_pPS | 27034 | -0.009124481 | [-0.02 0. ] | 0.133739579 | 0.358535892 |
| Hip | Liver-Lipid_pPS | 27034 | 0.014728467 | [0. 0.03] | 0.015496762 | 0.078103679 |
| Hip | ASAT_Pos_pPS | 27034 | -0.011875408 | [-0.02 0. ] | 0.050981879 | 0.183534763 |
| Hip | ALP_Neg_pPS | 27034 | -0.002140015 | [-0.01 0.01] | 0.725075919 | 0.870091103 |
| Hip | SHBG-LpA_pPS | 27034 | -0.006322211 | [-0.02 0.01] | 0.298813741 | 0.607266636 |
| Hip | Obesity_pPS | 27034 | 0.086363742 | [0.07 0.1 ] | 6.96E-46 | 2.19E-44 |
| Hip | VAT_Neg_pPS | 27034 | -0.00376348 | [-0.02 0.01] | 0.536259294 | 0.740718687 |
| Hip | Bilirubin_pPS | 27034 | -0.001003041 | [-0.01 0.01] | 0.869072817 | 0.920989005 |
| Hip | Proinsulin_pPS | 27034 | 0.002583436 | [-0.01 0.01] | 0.671162079 | 0.837291306 |
| Hip | Hyper_Insulin_pPS | 27034 | 0.005875133 | [-0.01 0.02] | 0.334290769 | 0.631606209 |
| Hip | Beta_Cell_1_pPS | 27034 | 0.006975163 | [-0. 0.02] | 0.25167598 | 0.533184986 |
| Hip | Beta_Cell_2_pPS | 27034 | -0.006150799 | [-0.02 0.01] | 0.312107112 | 0.620388644 |
| Weight | Cholesterol_pPS | 27055 | -0.004365654 | [-0.02 0.01] | 0.472930726 | 0.740718687 |
| Weight | Lipodystrophy_1_pPS | 27055 | -0.029732919 | [-0.04 -0.02] | 1.01E-06 | 1.82E-05 |
| Weight | Lipodystrophy_2_pPS | 27055 | -0.010035897 | [-0.02 0. ] | 0.098953705 | 0.296861115 |
| Weight | Liver-Lipid_pPS | 27055 | 0.024441089 | [0.01 0.04] | 5.85E-05 | 0.000737273 |
| Weight | ASAT_Pos_pPS | 27055 | -0.014337167 | [-0.03 -0. ] | 0.018415436 | 0.089244035 |
| Weight | ALP_Neg_pPS | 27055 | 0.003503654 | [-0.01 0.02] | 0.564612484 | 0.756820989 |
| Weight | SHBG-LpA_pPS | 27055 | -0.002945802 | [-0.01 0.01] | 0.628178538 | 0.799499957 |
| Weight | Obesity_pPS | 27055 | 0.099902048 | [0.09 0.11] | 6.53E-61 | 8.23E-59 |
| Weight | VAT_Neg_pPS | 27055 | -0.00524887 | [-0.02 0.01] | 0.388180125 | 0.65214261 |
| Weight | Bilirubin_pPS | 27055 | -0.003031525 | [-0.01 0.01] | 0.618212659 | 0.794844847 |
| Weight | Proinsulin_pPS | 27055 | -0.001280415 | [-0.01 0.01] | 0.833276742 | 0.920989005 |
| Weight | Hyper_Insulin_pPS | 27055 | 0.012067598 | [0. 0.02] | 0.047256909 | 0.175128546 |
| Weight | Beta_Cell_1_pPS | 27055 | 0.011103005 | [-0. 0.02] | 0.067941436 | 0.231368135 |
| Weight | Beta_Cell_2_pPS | 27055 | -0.005854759 | [-0.02 0.01] | 0.335781912 | 0.631606209 |
| Height | Cholesterol_pPS | 27055 | -0.005590315 | [-0.02 0.01] | 0.358065583 | 0.644518049 |
| Height | Lipodystrophy_1_pPS | 27055 | -0.008666233 | [-0.02 0. ] | 0.154226844 | 0.396583312 |
| Height | Lipodystrophy_2_pPS | 27055 | -0.007926992 | [-0.02 0. ] | 0.192499236 | 0.467137484 |
| Height | Liver-Lipid_pPS | 27055 | 0.017947487 | [0.01 0.03] | 0.003169397 | 0.024958999 |
| Height | ASAT_Pos_pPS | 27055 | -0.022746407 | [-0.03 -0.01] | 0.000184018 | 0.001932191 |
| Height | ALP_Neg_pPS | 27055 | -0.00188365 | [-0.01 0.01] | 0.756809498 | 0.876029913 |
| Height | SHBG-LpA_pPS | 27055 | -0.001503251 | [-0.01 0.01] | 0.804803557 | 0.897391577 |
| Height | Obesity_pPS | 27055 | 0.016632727 | [0. 0.03] | 0.006245299 | 0.040315172 |
| Height | VAT_Neg_pPS | 27055 | -0.02036632 | [-0.03 -0.01] | 0.000812271 | 0.007872781 |
| Height | Bilirubin_pPS | 27055 | 0.003056644 | [-0.01 0.01] | 0.615305487 | 0.794844847 |
| Height | Proinsulin_pPS | 27055 | 0.002225902 | [-0.01 0.01] | 0.71441011 | 0.865535325 |
| Height | Hyper_Insulin_pPS | 27055 | 0.001852777 | [-0.01 0.01] | 0.760672617 | 0.876029913 |
| Height | Beta_Cell_1_pPS | 27055 | 0.007693696 | [-0. 0.02] | 0.205919485 | 0.467137484 |
| Height | Beta_Cell_2_pPS | 27055 | -0.005512205 | [-0.02 0.01] | 0.364821673 | 0.647430012 |
| DBP_Mean | Cholesterol_pPS | 26976 | -0.005295356 | [-0.02 0.01] | 0.384687664 | 0.65214261 |
| DBP_Mean | Lipodystrophy_1_pPS | 26976 | 0.008198337 | [-0. 0.02] | 0.17834719 | 0.44943492 |
| DBP_Mean | Lipodystrophy_2_pPS | 26976 | 0.003447382 | [-0.01 0.02] | 0.571445591 | 0.75791731 |
| DBP_Mean | Liver-Lipid_pPS | 26976 | 0.004244745 | [-0.01 0.02] | 0.485915168 | 0.740718687 |
| DBP_Mean | ASAT_Pos_pPS | 26976 | 0.007212992 | [-0. 0.02] | 0.236373434 | 0.513500908 |
| DBP_Mean | ALP_Neg_pPS | 26976 | -0.010247902 | [-0.02 0. ] | 0.092503013 | 0.291384491 |
| DBP_Mean | SHBG-LpA_pPS | 26976 | -0.002636299 | [-0.01 0.01] | 0.665177818 | 0.837291306 |
| DBP_Mean | Obesity_pPS | 26976 | 0.025529586 | [0.01 0.04] | 2.77E-05 | 0.000387758 |
| DBP_Mean | VAT_Neg_pPS | 26976 | 0.004038727 | [-0.01 0.02] | 0.507330483 | 0.740718687 |
| DBP_Mean | Bilirubin_pPS | 26976 | -0.005862458 | [-0.02 0.01] | 0.335854095 | 0.631606209 |
| DBP_Mean | Proinsulin_pPS | 26976 | 0.005068351 | [-0.01 0.02] | 0.405394438 | 0.663372717 |
| DBP_Mean | Hyper_Insulin_pPS | 26976 | 0.009860484 | [-0. 0.02] | 0.105504025 | 0.309151329 |
| DBP_Mean | Beta_Cell_1_pPS | 26976 | 0.007676216 | [-0. 0.02] | 0.20761666 | 0.467137484 |
| DBP_Mean | Beta_Cell_2_pPS | 26976 | -0.016608206 | [-0.03 -0. ] | 0.006399234 | 0.040315172 |
| SBP_Mean | Cholesterol_pPS | 26978 | -0.007858834 | [-0.02 0. ] | 0.196990326 | 0.467137484 |
| SBP_Mean | Lipodystrophy_1_pPS | 26978 | 0.01145069 | [-0. 0.02] | 0.060124926 | 0.21043724 |
| SBP_Mean | Lipodystrophy_2_pPS | 26978 | -0.002450526 | [-0.01 0.01] | 0.687467784 | 0.848265129 |
| SBP_Mean | Liver-Lipid_pPS | 26978 | -0.003798423 | [-0.02 0.01] | 0.532907152 | 0.740718687 |
| SBP_Mean | ASAT_Pos_pPS | 26978 | 0.007725508 | [-0. 0.02] | 0.204696107 | 0.467137484 |
| SBP_Mean | ALP_Neg_pPS | 26978 | -0.001174168 | [-0.01 0.01] | 0.847147985 | 0.920989005 |
| SBP_Mean | SHBG-LpA_pPS | 26978 | 0.000366315 | [-0.01 0.01] | 0.952046919 | 0.973965573 |
| SBP_Mean | Obesity_pPS | 26978 | 0.023064366 | [0.01 0.03] | 0.000152544 | 0.001747327 |
| SBP_Mean | VAT_Neg_pPS | 26978 | 0.010138098 | [-0. 0.02] | 0.096037617 | 0.295139995 |

**Table S4: Associations between pPSs and anthropometric traits in SCAPIS**

**a) Spearman Correlation**

| Anthropometry | PPS | Sample_Size | r | CI95% | p | fdr_p_adjusted |
|---|---|---|---|---|---|---|
| SBP_Mean | Lipodystrophy_2_pPS | 26978 | 0.001012459 | [-0.01 0.01] | 0.867958967 | 0.929956036 |
| SBP_Mean | Liver-Lipid_pPS | 26978 | -0.002887494 | [-0.01 0.01] | 0.635397289 | 0.759102956 |
| SBP_Mean | ASAT_Pos_pPS | 26978 | 0.012001162 | [0. 0.02] | 0.048754797 | 0.117157587 |
| SBP_Mean | ALP_Neg_pPS | 26978 | 0.000636067 | [-0.01 0.01] | 0.91681561 | 0.953597618 |
| SBP_Mean | SHBG-LpA_pPS | 26978 | 0.001859642 | [-0.01 0.01] | 0.760088872 | 0.876847022 |
| SBP_Mean | Obesity_pPS | 26978 | 0.025157128 | [0.01 0.04] | 3.60E-05 | 0.000347386 |
| SBP_Mean | VAT_Neg_pPS | 26978 | 0.011888953 | [-0. 0.02] | 0.050902319 | 0.118479536 |
| SBP_Mean | Bilirubin_pPS | 26978 | -0.001705637 | [-0.01 0.01] | 0.779419575 | 0.876847022 |
| SBP_Mean | Proinsulin_pPS | 26978 | 0.011072756 | [-0. 0.02] | 0.069023634 | 0.152757222 |
| SBP_Mean | Hyper_Insulin_pPS | 26978 | 0.017398198 | [0.01 0.03] | 0.004275327 | 0.024048713 |
| SBP_Mean | Beta_Cell_1_pPS | 26978 | 0.006830329 | [-0.01 0.02] | 0.26203605 | 0.453523933 |
| SBP_Mean | Beta_Cell_2_pPS | 26978 | -0.004790702 | [-0.02 0.01] | 0.431478002 | 0.600510621 |
| SBP_Mean | Total_GRS | 26978 | 0.018038661 | [0.01 0.03] | 0.003053587 | 0.018737921 |
| Pulse | Cholesterol_pPS | 26972 | 0.00065041 | [-0.01 0.01] | 0.914956279 | 0.953597618 |
| Pulse | Lipodystrophy_1_pPS | 26972 | -0.00242409 | [-0.01 0.01] | 0.690626003 | 0.810734873 |
| Pulse | Lipodystrophy_2_pPS | 26972 | 0.00155285 | [-0.01 0.01] | 0.79875515 | 0.891173102 |
| Pulse | Liver-Lipid_pPS | 26972 | -0.016041058 | [-0.03 -0. ] | 0.008440812 | 0.036758374 |
| Pulse | ASAT_Pos_pPS | 26972 | 0.0035229 | [-0.01 0.02] | 0.562982204 | 0.723834262 |
| Pulse | ALP_Neg_pPS | 26972 | -0.020988684 | [-0.03 -0.01] | 0.000567989 | 0.004035713 |
| Pulse | SHBG-LpA_pPS | 26972 | -0.001155819 | [-0.01 0.01] | 0.849488549 | 0.917447633 |
| Pulse | Obesity_pPS | 26972 | 0.003000665 | [-0.01 0.01] | 0.622244032 | 0.75224747 |
| Pulse | VAT_Neg_pPS | 26972 | 0.01535591 | [0. 0.03] | 0.011689041 | 0.045479815 |
| Pulse | Bilirubin_pPS | 26972 | 0.00129128 | [-0.01 0.01] | 0.832097675 | 0.915174886 |
| Pulse | Proinsulin_pPS | 26972 | -0.004495246 | [-0.02 0.01] | 0.460474057 | 0.627919168 |
| Pulse | Hyper_Insulin_pPS | 26972 | -0.002211164 | [-0.01 0.01] | 0.716570737 | 0.833940082 |
| Pulse | Beta_Cell_1_pPS | 26972 | -0.00127779 | [-0.01 0.01] | 0.833826007 | 0.915174886 |
| Pulse | Beta_Cell_2_pPS | 26972 | -0.000617271 | [-0.01 0.01] | 0.919274082 | 0.953597618 |
| Pulse | Total_GRS | 26972 | 0.006538483 | [-0.01 0.02] | 0.283025341 | 0.466583941 |

**b) Partial Correlation**

| Anthropometry | PPS | Sample_Size | r | CI95% | p | fdr_p_adjusted |
|---|---|---|---|---|---|---|
| SBP_Mean | Bilirubin_pPS | 26978 | -0.001856328 | [-0.01 0.01] | 0.760559341 | 0.876029913 |
| SBP_Mean | Proinsulin_pPS | 26978 | 0.007244962 | [-0. 0.02] | 0.234285225 | 0.513500908 |
| SBP_Mean | Hyper_Insulin_pPS | 26978 | 0.013080579 | [0. 0.03] | 0.031755151 | 0.125035909 |
| SBP_Mean | Beta_Cell_1_pPS | 26978 | 0.003843632 | [-0.01 0.02] | 0.528042957 | 0.740718687 |
| SBP_Mean | Beta_Cell_2_pPS | 26978 | -0.007894217 | [-0.02 0. ] | 0.194981409 | 0.467137484 |
| Pulse | Cholesterol_pPS | 26972 | 0.000998389 | [-0.01 0.01] | 0.869822949 | 0.920989005 |
| Pulse | Lipodystrophy_1_pPS | 26972 | -0.002054307 | [-0.01 0.01] | 0.735958308 | 0.874818366 |
| Pulse | Lipodystrophy_2_pPS | 26972 | 0.000253728 | [-0.01 0.01] | 0.966778661 | 0.973965573 |
| Pulse | Liver-Lipid_pPS | 26972 | -0.015008972 | [-0.03 -0. ] | 0.013746188 | 0.072167485 |
| Pulse | ASAT_Pos_pPS | 26972 | 0.003671563 | [-0.01 0.02] | 0.546720936 | 0.740718687 |
| Pulse | ALP_Neg_pPS | 26972 | -0.019813285 | [-0.03 -0.01] | 0.001143438 | 0.010290939 |
| Pulse | SHBG-LpA_pPS | 26972 | -0.001607279 | [-0.01 0.01] | 0.791909742 | 0.897391577 |
| Pulse | Obesity_pPS | 26972 | 0.003911809 | [-0.01 0.02] | 0.52079698 | 0.740718687 |
| Pulse | VAT_Neg_pPS | 26972 | 0.016054079 | [0. 0.03] | 0.008403534 | 0.048129329 |
| Pulse | Bilirubin_pPS | 26972 | 0.001074224 | [-0.01 0.01] | 0.860033416 | 0.920989005 |
| Pulse | Proinsulin_pPS | 26972 | -0.001046997 | [-0.01 0.01] | 0.863545797 | 0.920989005 |
| Pulse | Hyper_Insulin_pPS | 26972 | -0.003678042 | [-0.02 0.01] | 0.546013494 | 0.740718687 |
| Pulse | Beta_Cell_1_pPS | 26972 | -0.001822741 | [-0.01 0.01] | 0.764788019 | 0.876029913 |
| Pulse | Beta_Cell_2_pPS | 26972 | -0.000198815 | [-0.01 0.01] | 0.973965573 | 0.973965573 |

**Table S5: Associations between pPSs and biochemical traits in SCAPIS**

**a) Spearman Correlation**

| Biochemistry | PPS | Sample_Size | r | CI95% | p | fdr_p_adjusted |
|---|---|---|---|---|---|---|
| Hba1c | Cholesterol_pPS | 24307 | 0.010732531 | [-0. 0.02] | 0.094361552 | 0.158803587 |
| Hba1c | Lipodystrophy_1_pPS | 24307 | 0.019493338 | [0.01 0.03] | 0.002377352 | 0.006358034 |
| Hba1c | Lipodystrophy_2_pPS | 24307 | 0.007238202 | [-0.01 0.02] | 0.259250415 | 0.359202382 |
| Hba1c | Liver-Lipid_pPS | 24307 | 0.028593142 | [0.02 0.04] | 8.29E-06 | 3.82E-05 |
| Hba1c | ASAT_Pos_pPS | 24307 | 0.035962698 | [0.02 0.05] | 2.06E-08 | 1.34E-07 |
| Hba1c | ALP_Neg_pPS | 24307 | 0.054807376 | [0.04 0.07] | 1.24E-17 | 2.04E-16 |
| Hba1c | SHBG-LpA_pPS | 24307 | 0.025764391 | [0.01 0.04] | 5.91E-05 | 0.000227027 |
| Hba1c | Obesity_pPS | 24307 | 0.039569072 | [0.03 0.05] | 6.84E-10 | 5.25E-09 |
| Hba1c | VAT_Neg_pPS | 24307 | 0.033012753 | [0.02 0.05] | 2.65E-07 | 1.45E-06 |
| Hba1c | Bilirubin_pPS | 24307 | 0.007333377 | [-0.01 0.02] | 0.253038879 | 0.352009731 |
| Hba1c | Proinsulin_pPS | 24307 | 0.064927755 | [0.05 0.08] | 4.05E-24 | 9.98E-23 |
| Hba1c | Hyper_Insulin_pPS | 24307 | 0.03126682 | [0.02 0.04] | 1.09E-06 | 5.38E-06 |
| Hba1c | Beta_Cell_1_pPS | 24307 | 0.063677344 | [0.05 0.08] | 2.94E-23 | 6.75E-22 |
| Hba1c | Beta_Cell_2_pPS | 24307 | 0.134910621 | [0.12 0.15] | 4.80E-99 | 4.14E-97 |
| Hba1c | Total_GRS | 24307 | 0.122306983 | [0.11 0.13] | 1.30E-81 | 7.46E-80 |
| Glucose | Cholesterol_pPS | 22368 | 0.004552659 | [-0.01 0.02] | 0.496076796 | 0.598414317 |
| Glucose | Lipodystrophy_1_pPS | 22368 | 0.04564651 | [0.03 0.06] | 8.60E-12 | 7.81E-11 |
| Glucose | Lipodystrophy_2_pPS | 22368 | 0.037064488 | [0.02 0.05] | 2.97E-08 | 1.89E-07 |
| Glucose | Liver-Lipid_pPS | 22368 | 0.047599983 | [0.03 0.06] | 1.07E-12 | 1.00E-11 |
| Glucose | ASAT_Pos_pPS | 22368 | 0.03497706 | [0.02 0.05] | 1.69E-07 | 9.63E-07 |
| Glucose | ALP_Neg_pPS | 22368 | 0.032598365 | [0.02 0.05] | 1.09E-06 | 5.38E-06 |
| Glucose | SHBG-LpA_pPS | 22368 | 0.000972392 | [-0.01 0.01] | 0.884408284 | 0.913535503 |
| Glucose | Obesity_pPS | 22368 | 0.03496418 | [0.02 0.05] | 1.70E-07 | 9.63E-07 |
| Glucose | VAT_Neg_pPS | 22368 | 0.051593274 | [0.04 0.06] | 1.17E-14 | 1.27E-13 |
| Glucose | Bilirubin_pPS | 22368 | 0.002859011 | [-0.01 0.02] | 0.66904866 | 0.739813422 |
| Glucose | Proinsulin_pPS | 22368 | 0.053466293 | [0.04 0.07] | 1.25E-15 | 1.60E-14 |
| Glucose | Hyper_Insulin_pPS | 22368 | 0.036282463 | [0.02 0.05] | 5.75E-08 | 3.48E-07 |
| Glucose | Beta_Cell_1_pPS | 22368 | 0.078876819 | [0.07 0.09] | 3.41E-32 | 9.81E-31 |
| Glucose | Beta_Cell_2_pPS | 22368 | 0.146760241 | [0.13 0.16] | 7.36E-108 | 8.46E-106 |
| Glucose | Total_GRS | 22368 | 0.139820482 | [0.13 0.15] | 5.55E-98 | 3.83E-96 |
| PGluc_Cap | Cholesterol_pPS | 24326 | 0.035244241 | [0.02 0.05] | 3.86E-08 | 2.40E-07 |
| PGluc_Cap | Lipodystrophy_1_pPS | 24326 | 0.015237068 | [0. 0.03] | 0.017505078 | 0.037279333 |
| PGluc_Cap | Lipodystrophy_2_pPS | 24326 | 0.022196168 | [0.01 0.03] | 0.000537603 | 0.001730453 |
| PGluc_Cap | Liver-Lipid_pPS | 24326 | 0.066543132 | [0.05 0.08] | 2.84E-25 | 7.54E-24 |
| PGluc_Cap | ASAT_Pos_pPS | 24326 | 0.019200718 | [0.01 0.03] | 0.002752739 | 0.007249579 |
| PGluc_Cap | ALP_Neg_pPS | 24326 | 0.019162263 | [0.01 0.03] | 0.00280734 | 0.007282198 |
| PGluc_Cap | SHBG-LpA_pPS | 24326 | 0.003591058 | [-0.01 0.02] | 0.5755307 | 0.670804364 |
| PGluc_Cap | Obesity_pPS | 24326 | 0.02304338 | [0.01 0.04] | 0.000326348 | 0.001137273 |
| PGluc_Cap | VAT_Neg_pPS | 24326 | 0.03893212 | [0.03 0.05] | 1.26E-09 | 9.44E-09 |
| PGluc_Cap | Bilirubin_pPS | 24326 | 0.007178205 | [-0.01 0.02] | 0.263033229 | 0.362600992 |
| PGluc_Cap | Proinsulin_pPS | 24326 | 0.049514324 | [0.04 0.06] | 1.12E-14 | 1.24E-13 |
| PGluc_Cap | Hyper_Insulin_pPS | 24326 | 0.027789362 | [0.02 0.04] | 1.47E-05 | 6.24E-05 |
| PGluc_Cap | Beta_Cell_1_pPS | 24326 | 0.054605163 | [0.04 0.07] | 1.59E-17 | 2.38E-16 |
| PGluc_Cap | Beta_Cell_2_pPS | 24326 | 0.116144643 | [0.1 0.13] | 8.76E-74 | 3.78E-72 |
| PGluc_Cap | Total_GRS | 24326 | 0.105503712 | [0.09 0.12] | 3.90E-61 | 1.49E-59 |
| Insulin | Cholesterol_pPS | 24080 | 0.015459206 | [0. 0.03] | 0.016469613 | 0.035512604 |
| Insulin | Lipodystrophy_1_pPS | 24080 | 0.032674325 | [0.02 0.05] | 3.98E-07 | 2.09E-06 |
| Insulin | Lipodystrophy_2_pPS | 24080 | 0.038007331 | [0.03 0.05] | 3.67E-09 | 2.54E-08 |
| Insulin | Liver-Lipid_pPS | 24080 | 0.037685675 | [0.03 0.05] | 4.97E-09 | 3.36E-08 |
| Insulin | ASAT_Pos_pPS | 24080 | 0.041467694 | [0.03 0.05] | 1.23E-10 | 9.64E-10 |
| Insulin | ALP_Neg_pPS | 24080 | 0.000247425 | [-0.01 0.01] | 0.969382119 | 0.975034493 |
| Insulin | SHBG-LpA_pPS | 24080 | -0.003437464 | [-0.02 0.01] | 0.593855432 | 0.679843511 |
| Insulin | Obesity_pPS | 24080 | 0.060699558 | [0.05 0.07] | 4.30E-21 | 8.72E-20 |
| Insulin | VAT_Neg_pPS | 24080 | 0.021431744 | [0.01 0.03] | 0.000883865 | 0.002628737 |
| Insulin | Bilirubin_pPS | 24080 | -0.005264106 | [-0.02 0.01] | 0.414140144 | 0.521453832 |
| Insulin | Proinsulin_pPS | 24080 | -0.010576534 | [-0.02 0. ] | 0.100841034 | 0.164104513 |
| Insulin | Hyper_Insulin_pPS | 24080 | 0.038242413 | [0.03 0.05] | 2.94E-09 | 2.12E-08 |
| Insulin | Beta_Cell_1_pPS | 24080 | -0.003672062 | [-0.02 0.01] | 0.568914369 | 0.665340533 |
| Insulin | Beta_Cell_2_pPS | 24080 | -0.022296102 | [-0.03 -0.01] | 0.000541707 | 0.001730453 |
| Insulin | Total_GRS | 24080 | 0.043074788 | [0.03 0.06] | 2.30E-11 | 1.85E-10 |
| Ldl | Cholesterol_pPS | 24249 | -0.145303088 | [-0.16 -0.13] | 1.77E-114 | 3.06E-112 |
| Ldl | Lipodystrophy_1_pPS | 24249 | 0.035289263 | [0.02 0.05] | 3.90E-08 | 2.40E-07 |
| Ldl | Lipodystrophy_2_pPS | 24249 | -0.018457789 | [-0.03 -0.01] | 0.004057597 | 0.010071014 |
| Ldl | Liver-Lipid_pPS | 24249 | -0.011730611 | [-0.02 0. ] | 0.067817443 | 0.120603185 |
| Ldl | ASAT_Pos_pPS | 24249 | 0.012615178 | [0. 0.03] | 0.049537321 | 0.091392384 |
| Ldl | ALP_Neg_pPS | 24249 | 0.043134614 | [0.03 0.06] | 1.84E-11 | 1.55E-10 |
| Ldl | SHBG-LpA_pPS | 24249 | 0.010272082 | [-0. 0.02] | 0.109789101 | 0.175357591 |
| Ldl | Obesity_pPS | 24249 | 0.003433035 | [-0.01 0.02] | 0.59303922 | 0.679843511 |
| Ldl | VAT_Neg_pPS | 24249 | 0.009890918 | [-0. 0.02] | 0.123609154 | 0.191097776 |
| Ldl | Bilirubin_pPS | 24249 | -0.022094443 | [-0.03 -0.01] | 0.000581805 | 0.001841493 |
| Ldl | Proinsulin_pPS | 24249 | 0.017413126 | [0. 0.03] | 0.006708419 | 0.015744249 |
| Ldl | Hyper_Insulin_pPS | 24249 | 0.000116232 | [-0.01 0.01] | 0.985563539 | 0.988428549 |
| Ldl | Beta_Cell_1_pPS | 24249 | 0.021987603 | [0.01 0.03] | 0.000618613 | 0.001940197 |
| Ldl | Beta_Cell_2_pPS | 24249 | 0.016848805 | [0. 0.03] | 0.008713041 | 0.020310805 |
| Ldl | Total_GRS | 24249 | 0.012400465 | [-0. 0.02] | 0.053543879 | 0.097389496 |
| ApoB | Cholesterol_pPS | 24104 | -0.148592032 | [-0.16 -0.14] | 5.55E-119 | 1.91E-116 |
| ApoB | Lipodystrophy_1_pPS | 24104 | 0.048551331 | [0.04 0.06] | 4.70E-14 | 4.91E-13 |
| ApoB | Lipodystrophy_2_pPS | 24104 | -0.005874492 | [-0.02 0.01] | 0.361886881 | 0.474619933 |
| ApoB | Liver-Lipid_pPS | 24104 | -0.021767662 | [-0.03 -0.01] | 0.000727728 | 0.002202334 |
| ApoB | ASAT_Pos_pPS | 24104 | 0.01416086 | [0. 0.03] | 0.027949973 | 0.056062445 |
| ApoB | ALP_Neg_pPS | 24104 | 0.027939803 | [0.02 0.04] | 1.44E-05 | 6.22E-05 |
| ApoB | SHBG-LpA_pPS | 24104 | 0.019001787 | [0.01 0.03] | 0.003182945 | 0.008134193 |
| ApoB | Obesity_pPS | 24104 | 0.006453356 | [-0.01 0.02] | 0.316525968 | 0.426001907 |
| ApoB | VAT_Neg_pPS | 24104 | 0.014483526 | [0. 0.03] | 0.024571352 | 0.050459026 |
| ApoB | Bilirubin_pPS | 24104 | -0.020473103 | [-0.03 -0.01] | 0.001483352 | 0.004194725 |
| ApoB | Proinsulin_pPS | 24104 | 0.012366842 | [-0. 0.02] | 0.054920425 | 0.099201814 |
| ApoB | Hyper_Insulin_pPS | 24104 | 0.006138198 | [-0.01 0.02] | 0.340739092 | 0.448683155 |
| ApoB | Beta_Cell_1_pPS | 24104 | 0.020835171 | [0.01 0.03] | 0.001220119 | 0.003507842 |
| ApoB | Beta_Cell_2_pPS | 24104 | 0.008983149 | [-0. 0.02] | 0.163231864 | 0.238622851 |
| ApoB | Total_GRS | 24104 | 0.01823398 | [0.01 0.03] | 0.004650533 | 0.011141902 |
| Cholesterol | Cholesterol_pPS | 24324 | -0.121015841 | [-0.13 -0.11] | 5.57E-80 | 2.74E-78 |
| Cholesterol | Lipodystrophy_1_pPS | 24324 | 0.023439937 | [0.01 0.04] | 0.000257041 | 0.000933464 |
| Cholesterol | Lipodystrophy_2_pPS | 24324 | -0.032501549 | [-0.05 -0.02] | 4.00E-07 | 2.09E-06 |
| Cholesterol | Liver-Lipid_pPS | 24324 | -0.025002485 | [-0.04 -0.01] | 9.66E-05 | 0.000362406 |
| Cholesterol | ASAT_Pos_pPS | 24324 | 0.004611936 | [-0.01 0.02] | 0.47209528 | 0.575522514 |
| Cholesterol | ALP_Neg_pPS | 24324 | 0.04696686 | [0.03 0.06] | 2.36E-13 | 2.32E-12 |
| Cholesterol | SHBG-LpA_pPS | 24324 | 0.005652243 | [-0.01 0.02] | 0.378168068 | 0.488131215 |
| Cholesterol | Obesity_pPS | 24324 | -0.000420573 | [-0.01 0.01] | 0.947717251 | 0.959128817 |
| Cholesterol | VAT_Neg_pPS | 24324 | -0.001780172 | [-0.01 0.01] | 0.781353372 | 0.824363649 |
| Cholesterol | Bilirubin_pPS | 24324 | -0.028148411 | [-0.04 -0.02] | 1.14E-05 | 4.96E-05 |
| Cholesterol | Proinsulin_pPS | 24324 | 0.014382116 | [0. 0.03] | 0.024928809 | 0.050890173 |
| Cholesterol | Hyper_Insulin_pPS | 24324 | -0.006413285 | [-0.02 0.01] | 0.317340551 | 0.426001907 |
| Cholesterol | Beta_Cell_1_pPS | 24324 | 0.018193293 | [0.01 0.03] | 0.004556205 | 0.011001118 |
| Cholesterol | Beta_Cell_2_pPS | 24324 | 0.010585538 | [-0. 0.02] | 0.0988426 | 0.162384271 |
| Cholesterol | Total_GRS | 24324 | -0.003039689 | [-0.02 0.01] | 0.635546757 | 0.714213783 |

**b) Partial Correlation**

| Biochemistry | PPS | Sample_Size | r | CI95% | p | fdr_p_adjusted |
|---|---|---|---|---|---|---|
| Hba1c | Cholesterol_pPS | 24307 | 0.005902768 | [-0.01 0.02] | 0.357692809 | 0.489926609 |
| Hba1c | Lipodystrophy_1_pPS | 24307 | 0.007057561 | [-0.01 0.02] | 0.271456598 | 0.402806565 |
| Hba1c | Lipodystrophy_2_pPS | 24307 | 9.92E-05 | [-0.01 0.01] | 0.987670207 | 0.992414235 |
| Hba1c | Liver-Lipid_pPS | 24307 | 0.017045161 | [0. 0.03] | 0.007903855 | 0.024874651 |
| Hba1c | ASAT_Pos_pPS | 24307 | 0.029579212 | [0.02 0.04] | 4.03E-06 | 3.09E-05 |
| Hba1c | ALP_Neg_pPS | 24307 | 0.046171727 | [0.03 0.06] | 6.10E-13 | 8.93E-12 |
| Hba1c | SHBG-LpA_pPS | 24307 | 0.023314811 | [0.01 0.04] | 0.000279733 | 0.001376365 |
| Hba1c | Obesity_pPS | 24307 | 0.030383502 | [0.02 0.04] | 2.19E-06 | 1.90E-05 |
| Hba1c | VAT_Neg_pPS | 24307 | 0.014653911 | [0. 0.03] | 0.022402257 | 0.057250211 |
| Hba1c | Bilirubin_pPS | 24307 | 0.007531838 | [-0.01 0.02] | 0.240547806 | 0.360262295 |
| Hba1c | Proinsulin_pPS | 24307 | 0.020912272 | [0.01 0.03] | 0.001118523 | 0.004446476 |
| Hba1c | Hyper_Insulin_pPS | 24307 | 0.022437773 | [0.01 0.04] | 0.000471109 | 0.002078043 |
| Hba1c | Beta_Cell_1_pPS | 24307 | 0.020718244 | [0.01 0.03] | 0.00124381 | 0.004884231 |
| Hba1c | Beta_Cell_2_pPS | 24307 | 0.118541686 | [0.11 0.13] | 1.05E-76 | 6.76E-75 |
| Glucose | Cholesterol_pPS | 22368 | -0.000747178 | [-0.01 0.01] | 0.911078167 | 0.940651714 |
| Glucose | Lipodystrophy_1_pPS | 22368 | 0.03498574 | [0.02 0.05] | 1.69E-07 | 1.81E-06 |
| Glucose | Lipodystrophy_2_pPS | 22368 | 0.027814414 | [0.01 0.04] | 3.21E-05 | 0.000194921 |
| Glucose | Liver-Lipid_pPS | 22368 | 0.035414376 | [0.02 0.05] | 1.19E-07 | 1.32E-06 |
| Glucose | ASAT_Pos_pPS | 22368 | 0.020497393 | [0.01 0.03] | 0.00218389 | 0.007901264 |
| Glucose | ALP_Neg_pPS | 22368 | 0.025106639 | [0.01 0.04] | 0.000174581 | 0.000944992 |
| Glucose | SHBG-LpA_pPS | 22368 | -0.005053151 | [-0.02 0.01] | 0.450077371 | 0.577390094 |
| Glucose | Obesity_pPS | 22368 | 0.027988381 | [0.01 0.04] | 2.86E-05 | 0.000180705 |
| Glucose | VAT_Neg_pPS | 22368 | 0.02981883 | [0.02 0.04] | 8.28E-06 | 6.06E-05 |
| Glucose | Bilirubin_pPS | 22368 | 0.00284027 | [-0.01 0.02] | 0.671180327 | 0.768519872 |
| Glucose | Proinsulin_pPS | 22368 | 0.006563053 | [-0.01 0.02] | 0.326606088 | 0.455131698 |
| Glucose | Hyper_Insulin_pPS | 22368 | 0.020716906 | [0.01 0.03] | 0.001955992 | 0.0073236 |
| Glucose | Beta_Cell_1_pPS | 22368 | 0.035961618 | [0.02 0.05] | 7.58E-08 | 8.72E-07 |
| Glucose | Beta_Cell_2_pPS | 22368 | 0.127626763 | [0.11 0.14] | 8.89E-82 | 9.54E-80 |
| PGluc_Cap | Cholesterol_pPS | 24326 | 0.031857158 | [0.02 0.04] | 6.79E-07 | 6.25E-06 |
| PGluc_Cap | Lipodystrophy_1_pPS | 24326 | 0.007911981 | [-0. 0.02] | 0.217450043 | 0.335018727 |
| PGluc_Cap | Lipodystrophy_2_pPS | 24326 | 0.013457254 | [0. 0.03] | 0.035922067 | 0.083818155 |
| PGluc_Cap | Liver-Lipid_pPS | 24326 | 0.058028833 | [0.05 0.07] | 1.39E-19 | 4.47E-18 |
| PGluc_Cap | ASAT_Pos_pPS | 24326 | 0.012335469 | [-0. 0.02] | 0.054489704 | 0.116806641 |
| PGluc_Cap | ALP_Neg_pPS | 24326 | 0.009453856 | [-0. 0.02] | 0.140563001 | 0.242038965 |
| PGluc_Cap | SHBG-LpA_pPS | 24326 | -0.001137898 | [-0.01 0.01] | 0.859213606 | 0.922222604 |
| PGluc_Cap | Obesity_pPS | 24326 | 0.013736332 | [0. 0.03] | 0.032248466 | 0.078075234 |
| PGluc_Cap | VAT_Neg_pPS | 24326 | 0.022889777 | [0.01 0.04] | 0.000359003 | 0.001631717 |
| PGluc_Cap | Bilirubin_pPS | 24326 | 0.008155337 | [-0. 0.02] | 0.203631497 | 0.324791176 |
| PGluc_Cap | Proinsulin_pPS | 24326 | 0.017984581 | [0.01 0.03] | 0.005052632 | 0.016958798 |
| PGluc_Cap | Hyper_Insulin_pPS | 24326 | 0.01834893 | [0.01 0.03] | 0.004230167 | 0.014968283 |
| PGluc_Cap | Beta_Cell_1_pPS | 24326 | 0.015922989 | [0. 0.03] | 0.013056333 | 0.036878415 |
| PGluc_Cap | Beta_Cell_2_pPS | 24326 | 0.100363085 | [0.09 0.11] | 1.94E-55 | 1.04E-53 |
| Insulin | Cholesterol_pPS | 24080 | 0.009397136 | [-0. 0.02] | 0.145000033 | 0.247037093 |
| Insulin | Lipodystrophy_1_pPS | 24080 | 0.022122513 | [0.01 0.03] | 0.000600538 | 0.002511341 |
| Insulin | Lipodystrophy_2_pPS | 24080 | 0.028253828 | [0.02 0.04] | 1.17E-05 | 8.20E-05 |
| Insulin | Liver-Lipid_pPS | 24080 | 0.036007955 | [0.02 0.05] | 2.32E-08 | 2.77E-07 |
| Insulin | ASAT_Pos_pPS | 24080 | 0.029786743 | [0.02 0.04] | 3.83E-06 | 3.00E-05 |
| Insulin | ALP_Neg_pPS | 24080 | -7.82E-05 | [-0.01 0.01] | 0.990324309 | 0.992414235 |
| Insulin | SHBG-LpA_pPS | 24080 | -0.006278088 | [-0.02 0.01] | 0.330220441 | 0.456356146 |
| Insulin | Obesity_pPS | 24080 | 0.05799039 | [0.05 0.07] | 2.23E-19 | 6.53E-18 |
| Insulin | VAT_Neg_pPS | 24080 | 0.017106327 | [0. 0.03] | 0.007973453 | 0.024874651 |
| Insulin | Bilirubin_pPS | 24080 | -0.004800013 | [-0.02 0.01] | 0.456615155 | 0.581146561 |
| Insulin | Proinsulin_pPS | 24080 | -0.016087801 | [-0.03 -0. ] | 0.01258871 | 0.035872252 |
| Insulin | Hyper_Insulin_pPS | 24080 | 0.02418304 | [0.01 0.04] | 0.000176086 | 0.000944992 |
| Insulin | Beta_Cell_1_pPS | 24080 | 6.13E-05 | [-0.01 0.01] | 0.992414235 | 0.992414235 |
| Insulin | Beta_Cell_2_pPS | 24080 | -0.022415924 | [-0.04 -0.01] | 0.00050726 | 0.002207267 |
| Ldl | Cholesterol_pPS | 24249 | -0.146469792 | [-0.16 -0.13] | 2.99E-116 | 4.82E-114 |
| Ldl | Lipodystrophy_1_pPS | 24249 | 0.032679088 | [0.02 0.05] | 3.63E-07 | 3.54E-06 |
| Ldl | Lipodystrophy_2_pPS | 24249 | -0.015138089 | [-0.03 -0. ] | 0.018467211 | 0.048741328 |
| Ldl | Liver-Lipid_pPS | 24249 | -0.013803518 | [-0.03 -0. ] | 0.031683627 | 0.077288848 |
| Ldl | ASAT_Pos_pPS | 24249 | 0.008166131 | [-0. 0.02] | 0.203750986 | 0.324791176 |
| Ldl | ALP_Neg_pPS | 24249 | 0.044644624 | [0.03 0.06] | 3.61E-12 | 4.84E-11 |
| Ldl | SHBG-LpA_pPS | 24249 | 0.010165771 | [-0. 0.02] | 0.113611675 | 0.209045483 |
| Ldl | Obesity_pPS | 24249 | 0.011828174 | [-0. 0.02] | 0.065634226 | 0.137235199 |
| Ldl | VAT_Neg_pPS | 24249 | 0.008097655 | [-0. 0.02] | 0.207568527 | 0.327588555 |
| Ldl | Bilirubin_pPS | 24249 | -0.023721104 | [-0.04 -0.01] | 0.000222238 | 0.001154206 |
| Ldl | Proinsulin_pPS | 24249 | 0.001238481 | [-0.01 0.01] | 0.847156337 | 0.913074192 |
| Ldl | Hyper_Insulin_pPS | 24249 | -0.003979277 | [-0.02 0.01] | 0.535713704 | 0.653408381 |
| Ldl | Beta_Cell_1_pPS | 24249 | 0.016327038 | [0. 0.03] | 0.011047639 | 0.032636144 |
| Ldl | Beta_Cell_2_pPS | 24249 | 0.011725615 | [-0. 0.02] | 0.068008497 | 0.139482395 |
| ApoB | Cholesterol_pPS | 24104 | -0.150301816 | [-0.16 -0.14] | 1.17E-121 | 3.78E-119 |
| ApoB | Lipodystrophy_1_pPS | 24104 | 0.045077245 | [0.03 0.06] | 2.60E-12 | 3.63E-11 |
| ApoB | Lipodystrophy_2_pPS | 24104 | -0.00277486 | [-0.02 0.01] | 0.666784787 | 0.768519872 |
| ApoB | Liver-Lipid_pPS | 24104 | -0.023718644 | [-0.04 -0.01] | 0.000232462 | 0.001169572 |
| ApoB | ASAT_Pos_pPS | 24104 | 0.00614987 | [-0.01 0.02] | 0.339952057 | 0.467797275 |
| ApoB | ALP_Neg_pPS | 24104 | 0.029978387 | [0.02 0.04] | 3.28E-06 | 2.64E-05 |
| ApoB | SHBG-LpA_pPS | 24104 | 0.018065891 | [0.01 0.03] | 0.005056039 | 0.016958798 |
| ApoB | Obesity_pPS | 24104 | 0.015954084 | [0. 0.03] | 0.013297912 | 0.037234152 |
| ApoB | VAT_Neg_pPS | 24104 | 0.011104319 | [-0. 0.02] | 0.084876523 | 0.166647807 |
| ApoB | Bilirubin_pPS | 24104 | -0.022347492 | [-0.03 -0.01] | 0.000524338 | 0.002221536 |
| ApoB | Proinsulin_pPS | 24104 | -0.000806282 | [-0.01 0.01] | 0.900438401 | 0.940651714 |
| ApoB | Hyper_Insulin_pPS | 24104 | -0.000622145 | [-0.01 0.01] | 0.923095096 | 0.949637767 |
| ApoB | Beta_Cell_1_pPS | 24104 | 0.017507816 | [0. 0.03] | 0.00659111 | 0.021223375 |
| ApoB | Beta_Cell_2_pPS | 24104 | 0.004524729 | [-0.01 0.02] | 0.482624367 | 0.607050961 |
| Cholesterol | Cholesterol_pPS | 24324 | -0.121188337 | [-0.13 -0.11] | 3.66E-80 | 2.94E-78 |
| Cholesterol | Lipodystrophy_1_pPS | 24324 | 0.022863526 | [0.01 0.04] | 0.000364856 | 0.001631717 |
| Cholesterol | Lipodystrophy_2_pPS | 24324 | -0.027504065 | [-0.04 -0.01] | 1.80E-05 | 0.000120965 |
| Cholesterol | Liver-Lipid_pPS | 24324 | -0.026641855 | [-0.04 -0.01] | 3.27E-05 | 0.000195251 |
| Cholesterol | ASAT_Pos_pPS | 24324 | 0.003628282 | [-0.01 0.02] | 0.571696801 | 0.681093501 |
| Cholesterol | ALP_Neg_pPS | 24324 | 0.049141472 | [0.04 0.06] | 1.80E-14 | 3.04E-13 |
| Cholesterol | SHBG-LpA_pPS | 24324 | 0.006577295 | [-0.01 0.02] | 0.305251836 | 0.431101277 |
| Cholesterol | Obesity_pPS | 24324 | 0.006907266 | [-0.01 0.02] | 0.281627047 | 0.41505624 |
| Cholesterol | VAT_Neg_pPS | 24324 | -0.000735007 | [-0.01 0.01] | 0.908787516 | 0.940651714 |
| Cholesterol | Bilirubin_pPS | 24324 | -0.029951303 | [-0.04 -0.02] | 3.02E-06 | 2.49E-05 |
| Cholesterol | Proinsulin_pPS | 24324 | -0.000124471 | [-0.01 0.01] | 0.984520645 | 0.992414235 |
| Cholesterol | Hyper_Insulin_pPS | 24324 | -0.006276252 | [-0.02 0.01] | 0.327920975 | 0.455131698 |
| Cholesterol | Beta_Cell_1_pPS | 24324 | 0.016355694 | [0. 0.03] | 0.010785051 | 0.03215543 |
| Cholesterol | Beta_Cell_2_pPS | 24324 | 0.006861108 | [-0.01 0.02] | 0.284855036 | 0.41505624 |
| Lpa | Cholesterol_pPS | 24097 | -0.015649912 | [-0.03 -0. ] | 0.015176879 | 0.041414873 |
| Lpa | Lipodystrophy_1_pPS | 24097 | 0.005685163 | [-0.01 0.02] | 0.377764333 | 0.506371365 |
| Lpa | Lipodystrophy_2_pPS | 24097 | -0.011225221 | [-0.02 0. ] | 0.081582886 | 0.161764059 |
| Lpa | Liver-Lipid_pPS | 24097 | -0.00236509 | [-0.01 0.01] | 0.713672171 | 0.793623694 |
| Lpa | ASAT_Pos_pPS | 24097 | -0.010093917 | [-0.02 0. ] | 0.117338012 | 0.211623247 |
| Lpa | ALP_Neg_pPS | 24097 | 0.003678789 | [-0.01 0.02] | 0.568174505 | 0.680119667 |
| Lpa | SHBG-LpA_pPS | 24097 | 0.055420432 | [0.04 0.07] | 7.65E-18 | 2.05E-16 |

**Table S5: Associations between pPSs and biochemical traits in SCAPIS**

**a) Spearman Correlation**

| Biochemistry | PPS | Sample_Size | r | CI95% | p | fdr_p_adjusted |
|---|---|---|---|---|---|---|
| Lpa | Cholesterol_pPS | 24097 | -0.014679456 | [-0.03 -0. ] | 0.022717075 | 0.047213197 |
| Lpa | Lipodystrophy_1_pPS | 24097 | 0.007135056 | [-0.01 0.02] | 0.268177012 | 0.367147099 |
| Lpa | Lipodystrophy_2_pPS | 24097 | -0.010216352 | [-0.02 0. ] | 0.112859647 | 0.178608156 |
| Lpa | Liver-Lipid_pPS | 24097 | -0.001375562 | [-0.01 0.01] | 0.830962559 | 0.868733584 |
| Lpa | ASAT_Pos_pPS | 24097 | -0.011114667 | [-0.02 0. ] | 0.084547848 | 0.145119441 |
| Lpa | ALP_Neg_pPS | 24097 | 0.002418204 | [-0.01 0.02] | 0.707459332 | 0.769947853 |
| Lpa | SHBG-LpA_pPS | 24097 | 0.057572824 | [0.04 0.07] | 3.83E-19 | 6.60E-18 |
| Lpa | Obesity_pPS | 24097 | 0.001101927 | [-0.01 0.01] | 0.864221408 | 0.898061403 |
| Lpa | VAT_Neg_pPS | 24097 | 0.034471059 | [0.02 0.05] | 8.75E-08 | 5.11E-07 |
| Lpa | Bilirubin_pPS | 24097 | -0.009731323 | [-0.02 0. ] | 0.130993754 | 0.199087424 |
| Lpa | Proinsulin_pPS | 24097 | -0.002937529 | [-0.02 0.01] | 0.648488876 | 0.72639176 |
| Lpa | Hyper_Insulin_pPS | 24097 | -0.00849459 | [-0.02 0. ] | 0.187416665 | 0.264994875 |
| Lpa | Beta_Cell_1_pPS | 24097 | 0.003510823 | [-0.01 0.02] | 0.585870377 | 0.679843511 |
| Lpa | Beta_Cell_2_pPS | 24097 | 0.006848581 | [-0.01 0.02] | 0.287867158 | 0.391000667 |
| Lpa | Total_GRS | 24097 | 0.00388433 | [-0.01 0.02] | 0.546646127 | 0.645866143 |
| Tg | Cholesterol_pPS | 24324 | -0.050085167 | [-0.06 -0.04] | 5.54E-15 | 6.59E-14 |
| Tg | Lipodystrophy_1_pPS | 24324 | 0.061513445 | [0.05 0.07] | 8.00E-22 | 1.72E-20 |
| Tg | Lipodystrophy_2_pPS | 24324 | 0.015425462 | [0. 0.03] | 0.016163411 | 0.035293524 |
| Tg | Liver-Lipid_pPS | 24324 | -0.059044219 | [-0.07 -0.05] | 3.14E-20 | 5.71E-19 |
| Tg | ASAT_Pos_pPS | 24324 | 0.028183399 | [0.02 0.04] | 1.11E-05 | 4.90E-05 |
| Tg | ALP_Neg_pPS | 24324 | 0.00130523 | [-0.01 0.01] | 0.838740285 | 0.874215705 |
| Tg | SHBG-LpA_pPS | 24324 | 0.0126455 | [0. 0.03] | 0.048643581 | 0.090225996 |
| Tg | Obesity_pPS | 24324 | 0.030639565 | [0.02 0.04] | 1.77E-06 | 8.46E-06 |
| Tg | VAT_Neg_pPS | 24324 | 0.031848619 | [0.02 0.04] | 6.80E-07 | 3.45E-06 |
| Tg | Bilirubin_pPS | 24324 | -0.009163721 | [-0.02 0. ] | 0.153064035 | 0.224711031 |
| Tg | Proinsulin_pPS | 24324 | 0.009914986 | [-0. 0.02] | 0.122120641 | 0.19064082 |
| Tg | Hyper_Insulin_pPS | 24324 | 0.0262271 | [0.01 0.04] | 4.32E-05 | 0.000171134 |
| Tg | Beta_Cell_1_pPS | 24324 | 0.015822802 | [0. 0.03] | 0.013618674 | 0.029926387 |
| Tg | Beta_Cell_2_pPS | 24324 | 0.009616613 | [-0. 0.02] | 0.13376748 | 0.201051768 |
| Tg | Total_GRS | 24324 | 0.047815487 | [0.04 0.06] | 8.69E-14 | 8.82E-13 |
| ApoA1 | Cholesterol_pPS | 24101 | 0.05293248 | [0.04 0.07] | 2.02E-16 | 2.68E-15 |
| ApoA1 | Lipodystrophy_1_pPS | 24101 | -0.033541444 | [-0.05 -0.02] | 1.92E-07 | 1.07E-06 |
| ApoA1 | Lipodystrophy_2_pPS | 24101 | -0.032140553 | [-0.04 -0.02] | 6.06E-07 | 3.12E-06 |
| ApoA1 | Liver-Lipid_pPS | 24101 | -0.026970987 | [-0.04 -0.01] | 2.83E-05 | 0.000114925 |
| ApoA1 | ASAT_Pos_pPS | 24101 | -0.019859193 | [-0.03 -0.01] | 0.002053276 | 0.005577796 |
| ApoA1 | ALP_Neg_pPS | 24101 | 0.012861164 | [0. 0.03] | 0.045921641 | 0.087049265 |
| ApoA1 | SHBG-LpA_pPS | 24101 | -0.01214487 | [-0.02 0. ] | 0.059438992 | 0.106804439 |
| ApoA1 | Obesity_pPS | 24101 | -0.008060922 | [-0.02 0. ] | 0.210912094 | 0.295039568 |
| ApoA1 | VAT_Neg_pPS | 24101 | -0.021675322 | [-0.03 -0.01] | 0.000767193 | 0.002301579 |
| ApoA1 | Bilirubin_pPS | 24101 | -0.005319456 | [-0.02 0.01] | 0.409044594 | 0.518824944 |
| ApoA1 | Proinsulin_pPS | 24101 | 0.010674615 | [-0. 0.02] | 0.097574318 | 0.161067654 |
| ApoA1 | Hyper_Insulin_pPS | 24101 | -0.027762947 | [-0.04 -0.02] | 1.64E-05 | 6.72E-05 |
| ApoA1 | Beta_Cell_1_pPS | 24101 | 0.008938755 | [-0. 0.02] | 0.165348167 | 0.240696699 |
| ApoA1 | Beta_Cell_2_pPS | 24101 | 0.002110799 | [-0.01 0.01] | 0.743219669 | 0.801283706 |
| ApoA1 | Total_GRS | 24101 | -0.024300484 | [-0.04 -0.01] | 0.000161977 | 0.000594489 |
| Hdl | Cholesterol_pPS | 24319 | 0.050348899 | [0.04 0.06] | 4.02E-15 | 4.95E-14 |
| Hdl | Lipodystrophy_1_pPS | 24319 | -0.053061953 | [-0.07 -0.04] | 1.25E-16 | 1.80E-15 |
| Hdl | Lipodystrophy_2_pPS | 24319 | -0.03879609 | [-0.05 -0.03] | 1.44E-09 | 1.06E-08 |
| Hdl | Liver-Lipid_pPS | 24319 | -0.00535371 | [-0.02 0.01] | 0.403918191 | 0.516117689 |
| Hdl | ASAT_Pos_pPS | 24319 | -0.030508386 | [-0.04 -0.02] | 1.96E-06 | 9.15E-06 |
| Hdl | ALP_Neg_pPS | 24319 | 0.006341 | [-0.01 0.02] | 0.322875772 | 0.431752485 |
| Hdl | SHBG-LpA_pPS | 24319 | -0.013667559 | [-0.03 -0. ] | 0.033100813 | 0.064518534 |
| Hdl | Obesity_pPS | 24319 | -0.025773543 | [-0.04 -0.01] | 5.85E-05 | 0.000227027 |
| Hdl | VAT_Neg_pPS | 24319 | -0.024802718 | [-0.04 -0.01] | 0.000110041 | 0.000408216 |
| Hdl | Bilirubin_pPS | 24319 | -0.003949169 | [-0.02 0.01] | 0.53810975 | 0.640165047 |
| Hdl | Proinsulin_pPS | 24319 | 0.002517647 | [-0.01 0.02] | 0.694689325 | 0.762325554 |
| Hdl | Hyper_Insulin_pPS | 24319 | -0.037860114 | [-0.05 -0.03] | 3.54E-09 | 2.49E-08 |
| Hdl | Beta_Cell_1_pPS | 24319 | -0.005832626 | [-0.02 0.01] | 0.363187427 | 0.474619933 |
| Hdl | Beta_Cell_2_pPS | 24319 | 0.003397676 | [-0.01 0.02] | 0.596322882 | 0.679843511 |
| Hdl | Total_GRS | 24319 | -0.046853633 | [-0.06 -0.03] | 2.70E-13 | 2.59E-12 |
| ALAT | Cholesterol_pPS | 24103 | 0.036113393 | [0.02 0.05] | 2.06E-08 | 1.34E-07 |
| ALAT | Lipodystrophy_1_pPS | 24103 | 0.031369465 | [0.02 0.04] | 1.12E-06 | 5.43E-06 |
| ALAT | Lipodystrophy_2_pPS | 24103 | 0.076206846 | [0.06 0.09] | 2.29E-32 | 7.17E-31 |
| ALAT | Liver-Lipid_pPS | 24103 | 0.014346264 | [0. 0.03] | 0.025965842 | 0.052695386 |
| ALAT | ASAT_Pos_pPS | 24103 | 0.02651111 | [0.01 0.04] | 3.87E-05 | 0.000155071 |
| ALAT | ALP_Neg_pPS | 24103 | 0.023002203 | [0.01 0.04] | 0.00035627 | 0.001229131 |
| ALAT | SHBG-LpA_pPS | 24103 | -0.008054954 | [-0.02 0. ] | 0.211231227 | 0.295039568 |
| ALAT | Obesity_pPS | 24103 | 0.034599156 | [0.02 0.05] | 7.81E-08 | 4.64E-07 |
| ALAT | VAT_Neg_pPS | 24103 | 0.011376037 | [-0. 0.02] | 0.077450967 | 0.134274289 |
| ALAT | Bilirubin_pPS | 24103 | -0.004040567 | [-0.02 0.01] | 0.530582158 | 0.633393926 |
| ALAT | Proinsulin_pPS | 24103 | 0.001470108 | [-0.01 0.01] | 0.819515812 | 0.859370684 |
| ALAT | Hyper_Insulin_pPS | 24103 | 0.027863486 | [0.02 0.04] | 1.52E-05 | 6.33E-05 |
| ALAT | Beta_Cell_1_pPS | 24103 | 0.003405843 | [-0.01 0.02] | 0.597079953 | 0.679843511 |
| ALAT | Beta_Cell_2_pPS | 24103 | -0.018292466 | [-0.03 -0.01] | 0.004521061 | 0.011001118 |
| ALAT | Total_GRS | 24103 | 0.059635925 | [0.05 0.07] | 1.97E-20 | 3.77E-19 |
| ASAT | Cholesterol_pPS | 24080 | 0.02027467 | [0.01 0.03] | 0.001657778 | 0.004575468 |
| ASAT | Lipodystrophy_1_pPS | 24080 | 0.018283615 | [0.01 0.03] | 0.004559884 | 0.011001118 |
| ASAT | Lipodystrophy_2_pPS | 24080 | 0.053203861 | [0.04 0.07] | 1.46E-16 | 2.02E-15 |
| ASAT | Liver-Lipid_pPS | 24080 | -0.012439196 | [-0.03 0. ] | 0.053634795 | 0.097398496 |
| ASAT | ASAT_Pos_pPS | 24080 | 0.018195776 | [0.01 0.03] | 0.004758323 | 0.011321527 |
| ASAT | ALP_Neg_pPS | 24080 | -0.015464797 | [-0.03 -0. ] | 0.016430647 | 0.035512604 |
| ASAT | SHBG-LpA_pPS | 24080 | -0.012725001 | [-0.03 -0. ] | 0.048368458 | 0.090200639 |
| ASAT | Obesity_pPS | 24080 | 0.019088058 | [0.01 0.03] | 0.003062324 | 0.007884343 |
| ASAT | VAT_Neg_pPS | 24080 | -0.00377047 | [-0.02 0.01] | 0.558603324 | 0.656314795 |
| ASAT | Bilirubin_pPS | 24080 | -0.002783863 | [-0.02 0.01] | 0.665840881 | 0.737863775 |
| ASAT | Proinsulin_pPS | 24080 | -0.005668708 | [-0.02 0.01] | 0.379185987 | 0.488131215 |
| ASAT | Hyper_Insulin_pPS | 24080 | 0.021358329 | [0.01 0.03] | 0.000920676 | 0.002714813 |
| ASAT | Beta_Cell_1_pPS | 24080 | 0.000608681 | [-0.01 0.01] | 0.924771456 | 0.943923528 |
| ASAT | Beta_Cell_2_pPS | 24080 | -0.022529357 | [-0.04 -0.01] | 0.000473288 | 0.001570041 |
| ASAT | Total_GRS | 24080 | 0.028400981 | [0.02 0.04] | 1.05E-05 | 4.70E-05 |
| GGT2 | Cholesterol_pPS | 24104 | 0.018297489 | [0.01 0.03] | 0.00450919 | 0.011001118 |
| GGT2 | Lipodystrophy_1_pPS | 24104 | 0.021831052 | [0.01 0.03] | 0.000702104 | 0.002162733 |
| GGT2 | Lipodystrophy_2_pPS | 24104 | 0.032973043 | [0.02 0.05] | 3.07E-07 | 1.66E-06 |
| GGT2 | Liver-Lipid_pPS | 24104 | -0.043509409 | [-0.06 -0.03] | 1.42E-11 | 1.22E-10 |
| GGT2 | ASAT_Pos_pPS | 24104 | 0.028628404 | [0.02 0.04] | 8.82E-06 | 4.00E-05 |
| GGT2 | ALP_Neg_pPS | 24104 | 0.023525564 | [0.01 0.04] | 0.000260349 | 0.000935628 |
| GGT2 | SHBG-LpA_pPS | 24104 | -0.002828889 | [-0.02 0.01] | 0.660613043 | 0.735788776 |
| GGT2 | Obesity_pPS | 24104 | 0.030763032 | [0.02 0.04] | 1.79E-06 | 8.46E-06 |
| GGT2 | VAT_Neg_pPS | 24104 | 0.016129309 | [0. 0.03] | 0.012295104 | 0.027366521 |
| GGT2 | Bilirubin_pPS | 24104 | 0.000440836 | [-0.01 0.01] | 0.94545028 | 0.959128817 |
| GGT2 | Proinsulin_pPS | 24104 | 0.014905056 | [0. 0.03] | 0.020694546 | 0.043270414 |
| GGT2 | Hyper_Insulin_pPS | 24104 | 0.021150136 | [0.01 0.03] | 0.001026956 | 0.003002541 |
| GGT2 | Beta_Cell_1_pPS | 24104 | 0.008712392 | [-0. 0.02] | 0.176292799 | 0.254481237 |
| GGT2 | Beta_Cell_2_pPS | 24104 | -0.001840641 | [-0.01 0.01] | 0.775121407 | 0.820297194 |
| GGT2 | Total_GRS | 24104 | 0.04379231 | [0.03 0.06] | 1.04E-11 | 9.24E-11 |

**b) Partial Correlation**

| Biochemistry | PPS | Sample_Size | r | CI95% | p | fdr_p_adjusted |
|---|---|---|---|---|---|---|
| Lpa | Obesity_pPS | 24097 | 0.004824001 | [-0.01 0.02] | 0.454208918 | 0.580378061 |
| Lpa | VAT_Neg_pPS | 24097 | 0.03286778 | [0.02 0.05] | 3.39E-07 | 3.41E-06 |
| Lpa | Bilirubin_pPS | 24097 | -0.010291252 | [-0.02 0. ] | 0.110340873 | 0.205374341 |
| Lpa | Proinsulin_pPS | 24097 | -0.007244119 | [-0.02 0.01] | 0.261058568 | 0.389170643 |
| Lpa | Hyper_Insulin_pPS | 24097 | -0.011797739 | [-0.02 0. ] | 0.067189252 | 0.139482395 |
| Lpa | Beta_Cell_1_pPS | 24097 | 0.000181226 | [-0.01 0.01] | 0.977569626 | 0.992414235 |
| Lpa | Beta_Cell_2_pPS | 24097 | 0.005371024 | [-0.01 0.02] | 0.404683785 | 0.534049913 |
| Tg | Cholesterol_pPS | 24324 | -0.052652828 | [-0.07 -0.04] | 2.16E-16 | 4.09E-15 |
| Tg | Lipodystrophy_1_pPS | 24324 | 0.054789026 | [0.04 0.07] | 1.27E-17 | 2.92E-16 |
| Tg | Lipodystrophy_2_pPS | 24324 | 0.011579682 | [-0. 0.02] | 0.071070889 | 0.144301867 |
| Tg | Liver-Lipid_pPS | 24324 | -0.062046173 | [-0.07 -0.05] | 3.64E-22 | 1.30E-20 |
| Tg | ASAT_Pos_pPS | 24324 | 0.014721071 | [0. 0.03] | 0.021747629 | 0.056021893 |
| Tg | ALP_Neg_pPS | 24324 | 0.002634681 | [-0.01 0.02] | 0.681309176 | 0.768519872 |
| Tg | SHBG-LpA_pPS | 24324 | 0.008957593 | [-0. 0.02] | 0.162630774 | 0.269933554 |
| Tg | Obesity_pPS | 24324 | 0.031961307 | [0.02 0.04] | 6.25E-07 | 5.92E-06 |
| Tg | VAT_Neg_pPS | 24324 | 0.025962543 | [0.01 0.04] | 5.18E-05 | 0.000292378 |
| Tg | Bilirubin_pPS | 24324 | -0.010690965 | [-0.02 0. ] | 0.095617026 | 0.183265966 |
| Tg | Proinsulin_pPS | 24324 | 0.000713552 | [-0.01 0.01] | 0.911438927 | 0.940651714 |
| Tg | Hyper_Insulin_pPS | 24324 | 0.014511129 | [0. 0.03] | 0.023696331 | 0.059611083 |
| Tg | Beta_Cell_1_pPS | 24324 | 0.010739618 | [-0. 0.02] | 0.094117133 | 0.181471359 |
| Tg | Beta_Cell_2_pPS | 24324 | 0.006797809 | [-0.01 0.02] | 0.289322296 | 0.419647655 |
| ApoA1 | Cholesterol_pPS | 24101 | 0.055289373 | [0.04 0.07] | 9.08E-18 | 2.25E-16 |
| ApoA1 | Lipodystrophy_1_pPS | 24101 | -0.027206555 | [-0.04 -0.01] | 2.42E-05 | 0.000155905 |
| ApoA1 | Lipodystrophy_2_pPS | 24101 | -0.027365233 | [-0.04 -0.01] | 2.17E-05 | 0.000142571 |
| ApoA1 | Liver-Lipid_pPS | 24101 | -0.026869167 | [-0.04 -0.01] | 3.05E-05 | 0.000188886 |
| ApoA1 | ASAT_Pos_pPS | 24101 | -0.010684515 | [-0.02 0. ] | 0.097354212 | 0.185491457 |
| ApoA1 | ALP_Neg_pPS | 24101 | 0.012360634 | [-0. 0.02] | 0.055123134 | 0.116806641 |
| ApoA1 | SHBG-LpA_pPS | 24101 | -0.009613329 | [-0.02 0. ] | 0.135803046 | 0.235099898 |
| ApoA1 | Obesity_pPS | 24101 | -0.011609035 | [-0.02 0. ] | 0.071659315 | 0.144301867 |
| ApoA1 | VAT_Neg_pPS | 24101 | -0.01661295 | [-0.03 -0. ] | 0.009943643 | 0.02992386 |
| ApoA1 | Bilirubin_pPS | 24101 | -0.005794877 | [-0.02 0.01] | 0.368588971 | 0.500783328 |
| ApoA1 | Proinsulin_pPS | 24101 | 0.008127028 | [-0. 0.02] | 0.207315699 | 0.327588555 |
| ApoA1 | Hyper_Insulin_pPS | 24101 | -0.017939985 | [-0.03 -0.01] | 0.005373823 | 0.017838877 |
| ApoA1 | Beta_Cell_1_pPS | 24101 | 0.010017709 | [-0. 0.02] | 0.120100851 | 0.214847079 |
| ApoA1 | Beta_Cell_2_pPS | 24101 | 0.000386056 | [-0.01 0.01] | 0.952236032 | 0.976095812 |
| Hdl | Cholesterol_pPS | 24319 | 0.054006769 | [0.04 0.07] | 3.65E-17 | 7.84E-16 |
| Hdl | Lipodystrophy_1_pPS | 24319 | -0.043750621 | [-0.06 -0.03] | 8.97E-12 | 1.16E-10 |
| Hdl | Lipodystrophy_2_pPS | 24319 | -0.032952648 | [-0.05 -0.02] | 2.79E-07 | 2.89E-06 |
| Hdl | Liver-Lipid_pPS | 24319 | -0.003325869 | [-0.02 0.01] | 0.604205419 | 0.707469618 |
| Hdl | ASAT_Pos_pPS | 24319 | -0.016191735 | [-0.03 -0. ] | 0.011610531 | 0.03398719 |
| Hdl | ALP_Neg_pPS | 24319 | 0.005884344 | [-0.01 0.02] | 0.359076645 | 0.489926609 |
| Hdl | SHBG-LpA_pPS | 24319 | -0.010220778 | [-0.02 0. ] | 0.111154689 | 0.205700056 |
| Hdl | Obesity_pPS | 24319 | -0.027559696 | [-0.04 -0.01] | 1.74E-05 | 0.00011904 |
| Hdl | VAT_Neg_pPS | 24319 | -0.016852527 | [-0.03 -0. ] | 0.008619807 | 0.026434074 |
| Hdl | Bilirubin_pPS | 24319 | -0.003820119 | [-0.02 0.01] | 0.551579019 | 0.667700918 |
| Hdl | Proinsulin_pPS | 24319 | 0.006886879 | [-0.01 0.02] | 0.283099266 | 0.41505624 |
| Hdl | Hyper_Insulin_pPS | 24319 | -0.024710041 | [-0.04 -0.01] | 0.000117239 | 0.000650879 |
| Hdl | Beta_Cell_1_pPS | 24319 | -0.004414092 | [-0.02 0.01] | 0.491470534 | 0.61577242 |
| Hdl | Beta_Cell_2_pPS | 24319 | 0.004714904 | [-0.01 0.02] | 0.462427015 | 0.586226374 |
| ALAT | Cholesterol_pPS | 24103 | 0.03052401 | [0.02 0.04] | 2.17E-06 | 1.90E-05 |
| ALAT | Lipodystrophy_1_pPS | 24103 | 0.023164668 | [0.01 0.04] | 0.000324669 | 0.001560349 |
| ALAT | Lipodystrophy_2_pPS | 24103 | 0.069775794 | [0.06 0.08] | 2.23E-27 | 8.97E-26 |
| ALAT | Liver-Lipid_pPS | 24103 | 0.009843941 | [-0. 0.02] | 0.126648984 | 0.221657487 |
| ALAT | ASAT_Pos_pPS | 24103 | 0.012740125 | [0. 0.03] | 0.048055891 | 0.104622238 |
| ALAT | ALP_Neg_pPS | 24103 | 0.021789696 | [0.01 0.03] | 0.000721301 | 0.002939985 |
| ALAT | SHBG-LpA_pPS | 24103 | -0.010514087 | [-0.02 0. ] | 0.102797402 | 0.193571717 |
| ALAT | Obesity_pPS | 24103 | 0.030419559 | [0.02 0.04] | 2.35E-06 | 1.99E-05 |
| ALAT | VAT_Neg_pPS | 24103 | 0.0026395 | [-0.01 0.02] | 0.682133896 | 0.768519872 |
| ALAT | Bilirubin_pPS | 24103 | -0.003525219 | [-0.02 0.01] | 0.584389309 | 0.691813815 |
| ALAT | Proinsulin_pPS | 24103 | -0.006689828 | [-0.02 0.01] | 0.299258784 | 0.424499244 |
| ALAT | Hyper_Insulin_pPS | 24103 | 0.015444009 | [0. 0.03] | 0.016553448 | 0.04441842 |
| ALAT | Beta_Cell_1_pPS | 24103 | 0.007588883 | [-0.01 0.02] | 0.238984282 | 0.359593172 |
| ALAT | Beta_Cell_2_pPS | 24103 | -0.019856264 | [-0.03 -0.01] | 0.002061282 | 0.007629112 |
| ASAT | Cholesterol_pPS | 24080 | 0.017571255 | [0. 0.03] | 0.006424053 | 0.021107602 |
| ASAT | Lipodystrophy_1_pPS | 24080 | 0.013688914 | [0. 0.03] | 0.033745875 | 0.080609321 |
| ASAT | Lipodystrophy_2_pPS | 24080 | 0.050626046 | [0.04 0.06] | 3.95E-15 | 7.06E-14 |
| ASAT | Liver-Lipid_pPS | 24080 | -0.013685073 | [-0.03 -0. ] | 0.033795834 | 0.080609321 |
| ASAT | ASAT_Pos_pPS | 24080 | 0.008607674 | [-0. 0.02] | 0.181883678 | 0.295790628 |
| ASAT | ALP_Neg_pPS | 24080 | -0.014685822 | [-0.03 -0. ] | 0.022742813 | 0.057662881 |
| ASAT | SHBG-LpA_pPS | 24080 | -0.013207478 | [-0.03 -0. ] | 0.040518893 | 0.092532507 |
| ASAT | Obesity_pPS | 24080 | 0.017089848 | [0. 0.03] | 0.008034049 | 0.024874651 |
| ASAT | VAT_Neg_pPS | 24080 | -0.009263117 | [-0.02 0. ] | 0.150821615 | 0.254264712 |
| ASAT | Bilirubin_pPS | 24080 | -0.002672448 | [-0.02 0.01] | 0.67853203 | 0.768519872 |
| ASAT | Proinsulin_pPS | 24080 | -0.003632219 | [-0.02 0.01] | 0.573218443 | 0.681093501 |
| ASAT | Hyper_Insulin_pPS | 24080 | 0.014916239 | [0. 0.03] | 0.020696894 | 0.054182113 |
| ASAT | Beta_Cell_1_pPS | 24080 | 0.007902273 | [-0. 0.02] | 0.220359677 | 0.337884838 |
| ASAT | Beta_Cell_2_pPS | 24080 | -0.021649452 | [-0.03 -0.01] | 0.000785147 | 0.003160217 |
| GGT2 | Cholesterol_pPS | 24104 | 0.015293602 | [0. 0.03] | 0.017635361 | 0.046930465 |
| GGT2 | Lipodystrophy_1_pPS | 24104 | 0.014807216 | [0. 0.03] | 0.021578695 | 0.056021893 |
| GGT2 | Lipodystrophy_2_pPS | 24104 | 0.028550084 | [0.02 0.04] | 9.38E-06 | 6.71E-05 |
| GGT2 | Liver-Lipid_pPS | 24104 | -0.047308193 | [-0.06 -0.03] | 2.06E-13 | 3.16E-12 |
| GGT2 | ASAT_Pos_pPS | 24104 | 0.020382076 | [0.01 0.03] | 0.001561874 | 0.005965006 |
| GGT2 | ALP_Neg_pPS | 24104 | 0.023061121 | [0.01 0.04] | 0.000345204 | 0.001631717 |
| GGT2 | SHBG-LpA_pPS | 24104 | -0.004024713 | [-0.02 0.01] | 0.532298387 | 0.651711333 |
| GGT2 | Obesity_pPS | 24104 | 0.02640359 | [0.01 0.04] | 4.17E-05 | 0.000244344 |
| GGT2 | VAT_Neg_pPS | 24104 | 0.010619308 | [-0. 0.02] | 0.099393215 | 0.188262443 |
| GGT2 | Bilirubin_pPS | 24104 | -9.18E-05 | [-0.01 0.01] | 0.988631922 | 0.992414235 |
| GGT2 | Proinsulin_pPS | 24104 | 0.005559583 | [-0.01 0.02] | 0.38832162 | 0.516692404 |
| GGT2 | Hyper_Insulin_pPS | 24104 | 0.013594658 | [0. 0.03] | 0.03489914 | 0.082628847 |
| GGT2 | Beta_Cell_1_pPS | 24104 | 0.006734123 | [-0.01 0.02] | 0.296060358 | 0.421820509 |
| GGT2 | Beta_Cell_2_pPS | 24104 | -0.004075433 | [-0.02 0.01] | 0.527144177 | 0.650346456 |
| Crp | Cholesterol_pPS | 24328 | 0.083123529 | [0.07 0.1 ] | 1.58E-38 | 7.28E-37 |
| Crp | Lipodystrophy_1_pPS | 24328 | -0.013348949 | [-0.03 -0. ] | 0.037433049 | 0.086715407 |
| Crp | Lipodystrophy_2_pPS | 24328 | -0.013120767 | [-0.03 -0. ] | 0.040812073 | 0.092545686 |
| Crp | Liver-Lipid_pPS | 24328 | -0.052815109 | [-0.07 -0.04] | 1.74E-16 | 3.50E-15 |
| Crp | ASAT_Pos_pPS | 24328 | -0.00182357 | [-0.01 0.01] | 0.77620384 | 0.850128015 |
| Crp | ALP_Neg_pPS | 24328 | 0.009873732 | [-0. 0.02] | 0.123751618 | 0.218945171 |
| Crp | SHBG-LpA_pPS | 24328 | -0.00795588 | [-0.02 0. ] | 0.214889867 | 0.332666045 |
| Crp | Obesity_pPS | 24328 | 0.047915823 | [0.04 0.06] | 7.80E-14 | 1.26E-12 |
| Crp | VAT_Neg_pPS | 24328 | 0.00468505 | [-0.01 0.02] | 0.465183293 | 0.587407923 |
| Crp | Bilirubin_pPS | 24328 | -0.004935148 | [-0.02 0.01] | 0.44169979 | 0.56890933 |
| Crp | Proinsulin_pPS | 24328 | -0.000775227 | [-0.01 0.01] | 0.903812085 | 0.940651714 |
| Crp | Hyper_Insulin_pPS | 24328 | 0.010852102 | [-0. 0.02] | 0.09069507 | 0.176992803 |
| Crp | Beta_Cell_1_pPS | 24328 | 0.007969445 | [-0. 0.02] | 0.214108942 | 0.332666045 |
| Crp | Beta_Cell_2_pPS | 24328 | 0.004149368 | [-0.01 0.02] | 0.517740753 | 0.641202009 |

**Table S5: Associations between pPSs and biochemical traits in SCAPIS**

**a) Spearman Correlation**

| Biochemistry | PPS | Sample_Size | r | CI95% | p | fdr_p_adjusted |
|---|---|---|---|---|---|---|
| Crp | Cholesterol_pPS | 24328 | 0.085610216 | [0.07 0.1 ] | 8.63E-41 | 2.98E-39 |
| Crp | Lipodystrophy_1_pPS | 24328 | -0.012732452 | [-0.03 -0. ] | 0.047097253 | 0.088735127 |
| Crp | Lipodystrophy_2_pPS | 24328 | -0.011413252 | [-0.02 0. ] | 0.075124732 | 0.131563618 |
| Crp | Liver-Lipid_pPS | 24328 | -0.050049426 | [-0.06 -0.04] | 5.76E-15 | 6.63E-14 |
| Crp | ASAT_Pos_pPS | 24328 | -0.001958563 | [-0.01 0.01] | 0.760065414 | 0.811834575 |
| Crp | ALP_Neg_pPS | 24328 | 0.009862601 | [-0. 0.02] | 0.124075078 | 0.191097776 |
| Crp | SHBG-LpA_pPS | 24328 | -0.00861149 | [-0.02 0. ] | 0.179337982 | 0.256728646 |
| Crp | Obesity_pPS | 24328 | 0.054746359 | [0.04 0.07] | 1.31E-17 | 2.05E-16 |
| Crp | VAT_Neg_pPS | 24328 | 0.004663448 | [-0.01 0.02] | 0.467124573 | 0.573515935 |
| Crp | Bilirubin_pPS | 24328 | -0.004225027 | [-0.02 0.01] | 0.510022654 | 0.613093434 |
| Crp | Proinsulin_pPS | 24328 | 0.009609222 | [-0. 0.02] | 0.134034512 | 0.201051768 |
| Crp | Hyper_Insulin_pPS | 24328 | 0.010279237 | [-0. 0.02] | 0.108965146 | 0.174851049 |
| Crp | Beta_Cell_1_pPS | 24328 | 0.008472991 | [-0. 0.02] | 0.186434987 | 0.264691649 |
| Crp | Beta_Cell_2_pPS | 24328 | 0.005274547 | [-0.01 0.02] | 0.410818248 | 0.519165918 |
| Crp | Total_GRS | 24328 | 0.025750455 | [0.01 0.04] | 5.92E-05 | 0.000227027 |
| Alfa1Antitrypsin | Cholesterol_pPS | 24104 | -0.001934758 | [-0.01 0.01] | 0.763955972 | 0.813471637 |
| Alfa1Antitrypsin | Lipodystrophy_1_pPS | 24104 | -0.020369847 | [-0.03 -0.01] | 0.001567489 | 0.004361158 |
| Alfa1Antitrypsin | Lipodystrophy_2_pPS | 24104 | -0.011436424 | [-0.02 0. ] | 0.07588441 | 0.132222836 |
| Alfa1Antitrypsin | Liver-Lipid_pPS | 24104 | 0.022459011 | [0.01 0.04] | 0.000489832 | 0.001594263 |
| Alfa1Antitrypsin | ASAT_Pos_pPS | 24104 | -0.013098174 | [-0.03 -0. ] | 0.042049741 | 0.080150059 |
| Alfa1Antitrypsin | ALP_Neg_pPS | 24104 | 3.69E-05 | [-0.01 0.01] | 0.995432009 | 0.995432009 |
| Alfa1Antitrypsin | SHBG-LpA_pPS | 24104 | -0.01374799 | [-0.03 -0. ] | 0.03285115 | 0.064395721 |
| Alfa1Antitrypsin | Obesity_pPS | 24104 | 0.009833731 | [-0. 0.02] | 0.126932726 | 0.19463018 |
| Alfa1Antitrypsin | VAT_Neg_pPS | 24104 | -0.02350213 | [-0.04 -0.01] | 0.000264063 | 0.000939192 |
| Alfa1Antitrypsin | Bilirubin_pPS | 24104 | 0.004843093 | [-0.01 0.02] | 0.452236981 | 0.561229347 |
| Alfa1Antitrypsin | Proinsulin_pPS | 24104 | 0.006646245 | [-0.01 0.02] | 0.302277707 | 0.408963957 |
| Alfa1Antitrypsin | Hyper_Insulin_pPS | 24104 | -0.020396742 | [-0.03 -0.01] | 0.001545161 | 0.004333988 |
| Alfa1Antitrypsin | Beta_Cell_1_pPS | 24104 | 0.004136242 | [-0.01 0.02] | 0.520885976 | 0.623977992 |
| Alfa1Antitrypsin | Beta_Cell_2_pPS | 24104 | -0.013935405 | [-0.03 -0. ] | 0.030541922 | 0.060211218 |
| Alfa1Antitrypsin | Total_GRS | 24104 | -0.019270554 | [-0.03 -0.01] | 0.002778649 | 0.007262378 |
| TroponinI | Cholesterol_pPS | 24091 | -0.022687204 | [-0.04 -0.01] | 0.00043035 | 0.001441462 |
| TroponinI | Lipodystrophy_1_pPS | 24091 | 0.003425089 | [-0.01 0.02] | 0.595100722 | 0.679843511 |
| TroponinI | Lipodystrophy_2_pPS | 24091 | -0.009661758 | [-0.02 0. ] | 0.133818734 | 0.201051768 |
| TroponinI | Liver-Lipid_pPS | 24091 | 0.022801924 | [0.01 0.04] | 0.00040234 | 0.001374329 |
| TroponinI | ASAT_Pos_pPS | 24091 | 0.004739047 | [-0.01 0.02] | 0.462130424 | 0.569410701 |
| TroponinI | ALP_Neg_pPS | 24091 | 0.009967629 | [-0. 0.02] | 0.121941409 | 0.19064082 |
| TroponinI | SHBG-LpA_pPS | 24091 | 0.001020094 | [-0.01 0.01] | 0.874233079 | 0.905736974 |
| TroponinI | Obesity_pPS | 24091 | 0.012444625 | [-0. 0.03] | 0.053475882 | 0.097389496 |
| TroponinI | VAT_Neg_pPS | 24091 | -0.004986425 | [-0.02 0.01] | 0.439091299 | 0.548864123 |
| TroponinI | Bilirubin_pPS | 24091 | 0.002542373 | [-0.01 0.02] | 0.693218864 | 0.762325554 |
| TroponinI | Proinsulin_pPS | 24091 | 0.00389202 | [-0.01 0.02] | 0.545902379 | 0.645866143 |
| TroponinI | Hyper_Insulin_pPS | 24091 | 0.009380324 | [-0. 0.02] | 0.145519909 | 0.21639814 |
| TroponinI | Beta_Cell_1_pPS | 24091 | 0.005120305 | [-0.01 0.02] | 0.426902828 | 0.535569002 |
| TroponinI | Beta_Cell_2_pPS | 24091 | 0.003293744 | [-0.01 0.02] | 0.609296047 | 0.689203725 |
| TroponinI | Total_GRS | 24091 | 0.006286051 | [-0.01 0.02] | 0.329366225 | 0.437043645 |
| NTproBNP | Cholesterol_pPS | 24056 | -0.002436128 | [-0.02 0.01] | 0.705631096 | 0.769947853 |
| NTproBNP | Lipodystrophy_1_pPS | 24056 | -0.016385822 | [-0.03 -0. ] | 0.011058496 | 0.0252661 |
| NTproBNP | Lipodystrophy_2_pPS | 24056 | -0.019351745 | [-0.03 -0.01] | 0.002692437 | 0.007145313 |
| NTproBNP | Liver-Lipid_pPS | 24056 | 0.013283888 | [0. 0.03] | 0.039417627 | 0.075550452 |
| NTproBNP | ASAT_Pos_pPS | 24056 | -0.016763147 | [-0.03 -0. ] | 0.009339707 | 0.021625496 |
| NTproBNP | ALP_Neg_pPS | 24056 | 0.020557507 | [0.01 0.03] | 0.001433376 | 0.004086898 |
| NTproBNP | SHBG-LpA_pPS | 24056 | 0.005801932 | [-0.01 0.02] | 0.368324771 | 0.477714458 |
| NTproBNP | Obesity_pPS | 24056 | -0.002086767 | [-0.01 0.01] | 0.746270963 | 0.802066922 |
| NTproBNP | VAT_Neg_pPS | 24056 | -0.010167025 | [-0.02 0. ] | 0.114917368 | 0.18103421 |
| NTproBNP | Bilirubin_pPS | 24056 | 0.000634556 | [-0.01 0.01] | 0.921622636 | 0.943500918 |
| NTproBNP | Proinsulin_pPS | 24056 | 0.00957774 | [-0. 0.02] | 0.137519083 | 0.205385644 |
| NTproBNP | Hyper_Insulin_pPS | 24056 | -0.009794423 | [-0.02 0. ] | 0.128839731 | 0.19668012 |
| NTproBNP | Beta_Cell_1_pPS | 24056 | -0.00535402 | [-0.02 0.01] | 0.406446077 | 0.517431353 |
| NTproBNP | Beta_Cell_2_pPS | 24056 | -0.016628664 | [-0.03 -0. ] | 0.009922856 | 0.02282257 |
| NTproBNP | Total_GRS | 24056 | -0.022518912 | [-0.04 -0.01] | 0.000479288 | 0.001574804 |
| HB | Cholesterol_pPS | 24295 | 0.001866947 | [-0.01 0.01] | 0.771118847 | 0.818572315 |
| HB | Lipodystrophy_1_pPS | 24295 | 0.020855344 | [0.01 0.03] | 0.001153745 | 0.003344891 |
| HB | Lipodystrophy_2_pPS | 24295 | 0.010407177 | [-0. 0.02] | 0.104865192 | 0.169852072 |
| HB | Liver-Lipid_pPS | 24295 | 0.016268821 | [0. 0.03] | 0.011238053 | 0.025507423 |
| HB | ASAT_Pos_pPS | 24295 | 0.014198241 | [0. 0.03] | 0.026931449 | 0.054335379 |
| HB | ALP_Neg_pPS | 24295 | -0.042965988 | [-0.06 -0.03] | 2.11E-11 | 1.73E-10 |
| HB | SHBG-LpA_pPS | 24295 | -0.018602925 | [-0.03 -0.01] | 0.003743596 | 0.00935899 |
| HB | Obesity_pPS | 24295 | 0.023272699 | [0.01 0.04] | 0.000286855 | 0.001009848 |
| HB | VAT_Neg_pPS | 24295 | 0.000267295 | [-0.01 0.01] | 0.966777374 | 0.975034493 |
| HB | Bilirubin_pPS | 24295 | -0.000814731 | [-0.01 0.01] | 0.898977604 | 0.925812757 |
| HB | Proinsulin_pPS | 24295 | -0.007077566 | [-0.02 0.01] | 0.270090401 | 0.368305092 |
| HB | Hyper_Insulin_pPS | 24295 | 0.004426435 | [-0.01 0.02] | 0.490358131 | 0.593967239 |
| HB | Beta_Cell_1_pPS | 24295 | 0.005582491 | [-0.01 0.02] | 0.38436503 | 0.492958867 |
| HB | Beta_Cell_2_pPS | 24295 | -0.016144129 | [-0.03 -0. ] | 0.01187711 | 0.026781719 |
| HB | Total_GRS | 24295 | 0.013543135 | [0. 0.03] | 0.034822599 | 0.067493239 |
| Creatinine | Cholesterol_pPS | 24332 | 0.011628066 | [-0. 0.02] | 0.069777871 | 0.123453156 |
| Creatinine | Lipodystrophy_1_pPS | 24332 | 0.011017357 | [-0. 0.02] | 0.085777217 | 0.146500693 |
| Creatinine | Lipodystrophy_2_pPS | 24332 | -0.009272141 | [-0.02 0. ] | 0.148195391 | 0.218642226 |
| Creatinine | Liver-Lipid_pPS | 24332 | 0.027733892 | [0.02 0.04] | 1.52E-05 | 6.33E-05 |
| Creatinine | ASAT_Pos_pPS | 24332 | -0.021922766 | [-0.03 -0.01] | 0.000628374 | 0.001953053 |
| Creatinine | ALP_Neg_pPS | 24332 | -0.004823266 | [-0.02 0.01] | 0.451962123 | 0.561229347 |
| Creatinine | SHBG-LpA_pPS | 24332 | 0.014074666 | [0. 0.03] | 0.028169197 | 0.056175566 |
| Creatinine | Obesity_pPS | 24332 | -0.011795651 | [-0.02 0. ] | 0.065843746 | 0.117699961 |
| Creatinine | VAT_Neg_pPS | 24332 | -0.013455137 | [-0.03 -0. ] | 0.035878803 | 0.069151882 |
| Creatinine | Bilirubin_pPS | 24332 | 0.002042786 | [-0.01 0.01] | 0.750064883 | 0.803640946 |
| Creatinine | Proinsulin_pPS | 24332 | -0.008332134 | [-0.02 0. ] | 0.193826805 | 0.272939786 |
| Creatinine | Hyper_Insulin_pPS | 24332 | -0.004419901 | [-0.02 0.01] | 0.490668589 | 0.593967239 |
| Creatinine | Beta_Cell_1_pPS | 24332 | -0.0087808 | [-0.02 0. ] | 0.170902254 | 0.247736461 |
| Creatinine | Beta_Cell_2_pPS | 24332 | -0.011220759 | [-0.02 0. ] | 0.080148521 | 0.138256198 |
| Creatinine | Total_GRS | 24332 | -0.020147719 | [-0.03 -0.01] | 0.001677017 | 0.004591832 |
| CystatinC | Cholesterol_pPS | 24104 | 0.011049126 | [-0. 0.02] | 0.086351779 | 0.146755486 |
| CystatinC | Lipodystrophy_1_pPS | 24104 | -0.00582581 | [-0.02 0.01] | 0.365879023 | 0.476333067 |
| CystatinC | Lipodystrophy_2_pPS | 24104 | -0.00326743 | [-0.02 0.01] | 0.612061954 | 0.69006985 |
| CystatinC | Liver-Lipid_pPS | 24104 | 0.016164666 | [0. 0.03] | 0.012105695 | 0.0271199 |
| CystatinC | ASAT_Pos_pPS | 24104 | -0.010729766 | [-0.02 0. ] | 0.095834442 | 0.159966643 |
| CystatinC | ALP_Neg_pPS | 24104 | 0.004634237 | [-0.01 0.02] | 0.471972048 | 0.575522514 |
| CystatinC | SHBG-LpA_pPS | 24104 | 0.003452307 | [-0.01 0.02] | 0.592078147 | 0.679843511 |
| CystatinC | Obesity_pPS | 24104 | 0.017510486 | [0. 0.03] | 0.00656829 | 0.01552096 |
| CystatinC | VAT_Neg_pPS | 24104 | -0.011526642 | [-0.02 0. ] | 0.073601018 | 0.129552811 |
| CystatinC | Bilirubin_pPS | 24104 | 0.002824173 | [-0.01 0.02] | 0.661143538 | 0.735788776 |
| CystatinC | Proinsulin_pPS | 24104 | -0.000420132 | [-0.01 0.01] | 0.948008483 | 0.959128817 |
| CystatinC | Hyper_Insulin_pPS | 24104 | -0.015036557 | [-0.03 -0. ] | 0.019599666 | 0.041231004 |
| CystatinC | Beta_Cell_1_pPS | 24104 | 0.008669778 | [-0. 0.02] | 0.178417482 | 0.25647513 |
| CystatinC | Beta_Cell_2_pPS | 24104 | -0.011018304 | [-0.02 0. ] | 0.087232604 | 0.147525727 |
| CystatinC | Total_GRS | 24104 | -0.003326292 | [-0.02 0.01] | 0.605667076 | 0.687352438 |

**b) Partial Correlation**

| Biochemistry | PPS | Sample_Size | r | CI95% | p | fdr_p_adjusted |
|---|---|---|---|---|---|---|
| Alfa1Antitrypsin | Cholesterol_pPS | 24104 | -0.002355399 | [-0.01 0.01] | 0.714754259 | 0.793623694 |
| Alfa1Antitrypsin | Lipodystrophy_1_pPS | 24104 | -0.01585821 | [-0.03 -0. ] | 0.013862361 | 0.038480001 |
| Alfa1Antitrypsin | Lipodystrophy_2_pPS | 24104 | -0.006793398 | [-0.02 0.01] | 0.29182942 | 0.419749624 |
| Alfa1Antitrypsin | Liver-Lipid_pPS | 24104 | 0.023975035 | [0.01 0.04] | 0.000198714 | 0.001048948 |
| Alfa1Antitrypsin | ASAT_Pos_pPS | 24104 | -0.008895881 | [-0.02 0. ] | 0.167473985 | 0.276546786 |
| Alfa1Antitrypsin | ALP_Neg_pPS | 24104 | -0.001791227 | [-0.01 0.01] | 0.781060038 | 0.852546888 |
| Alfa1Antitrypsin | SHBG-LpA_pPS | 24104 | -0.011530188 | [-0.02 0. ] | 0.073590174 | 0.147180348 |
| Alfa1Antitrypsin | Obesity_pPS | 24104 | 0.010084104 | [-0. 0.02] | 0.117641494 | 0.211623247 |
| Alfa1Antitrypsin | VAT_Neg_pPS | 24104 | -0.019772895 | [-0.03 -0.01] | 0.002152214 | 0.007875148 |
| Alfa1Antitrypsin | Bilirubin_pPS | 24104 | 0.00500626 | [-0.01 0.02] | 0.437273034 | 0.565469546 |
| Alfa1Antitrypsin | Proinsulin_pPS | 24104 | 0.006322307 | [-0.01 0.02] | 0.326584259 | 0.455131698 |
| Alfa1Antitrypsin | Hyper_Insulin_pPS | 24104 | -0.015810482 | [-0.03 -0. ] | 0.014151164 | 0.038945938 |
| Alfa1Antitrypsin | Beta_Cell_1_pPS | 24104 | 0.0068922 | [-0.01 0.02] | 0.284867792 | 0.41505624 |
| Alfa1Antitrypsin | Beta_Cell_2_pPS | 24104 | -0.015562006 | [-0.03 -0. ] | 0.015742075 | 0.042596202 |
| TroponinI | Cholesterol_pPS | 24091 | -0.023759496 | [-0.04 -0.01] | 0.000227635 | 0.001163467 |
| TroponinI | Lipodystrophy_1_pPS | 24091 | 0.001375367 | [-0.01 0.01] | 0.831051828 | 0.901005685 |
| TroponinI | Lipodystrophy_2_pPS | 24091 | -0.009846073 | [-0.02 0. ] | 0.126661421 | 0.221657487 |
| TroponinI | Liver-Lipid_pPS | 24091 | 0.022370516 | [0.01 0.03] | 0.000519217 | 0.002221536 |
| TroponinI | ASAT_Pos_pPS | 24091 | 0.004031788 | [-0.01 0.02] | 0.53168872 | 0.651711333 |
| TroponinI | ALP_Neg_pPS | 24091 | 0.009015842 | [-0. 0.02] | 0.161931987 | 0.269933554 |
| TroponinI | SHBG-LpA_pPS | 24091 | 0.001236793 | [-0.01 0.01] | 0.847854607 | 0.913074192 |
| TroponinI | Obesity_pPS | 24091 | 0.012741514 | [0. 0.03] | 0.04808724 | 0.104622238 |
| TroponinI | VAT_Neg_pPS | 24091 | -0.005671652 | [-0.02 0.01] | 0.378958333 | 0.506371365 |
| TroponinI | Bilirubin_pPS | 24091 | 0.002671105 | [-0.01 0.02] | 0.678615186 | 0.768519872 |
| TroponinI | Proinsulin_pPS | 24091 | -0.001068783 | [-0.01 0.01] | 0.868318945 | 0.925823511 |
| TroponinI | Hyper_Insulin_pPS | 24091 | 0.008847363 | [-0. 0.02] | 0.169918288 | 0.27773446 |
| TroponinI | Beta_Cell_1_pPS | 24091 | 0.003474208 | [-0.01 0.02] | 0.589931634 | 0.695816799 |
| TroponinI | Beta_Cell_2_pPS | 24091 | 0.001671683 | [-0.01 0.01] | 0.795389466 | 0.865254757 |
| NTproBNP | Cholesterol_pPS | 24056 | -0.002076503 | [-0.01 0.01] | 0.747542292 | 0.824344582 |
| NTproBNP | Lipodystrophy_1_pPS | 24056 | -0.013552547 | [-0.03 -0. ] | 0.035650242 | 0.08379108 |
| NTproBNP | Lipodystrophy_2_pPS | 24056 | -0.016859352 | [-0.03 -0. ] | 0.008960024 | 0.027218185 |
| NTproBNP | Liver-Lipid_pPS | 24056 | 0.014004422 | [0. 0.03] | 0.029934959 | 0.073939728 |
| NTproBNP | ASAT_Pos_pPS | 24056 | -0.011782113 | [-0.02 0. ] | 0.067786439 | 0.139482395 |
| NTproBNP | ALP_Neg_pPS | 24056 | 0.018480765 | [0.01 0.03] | 0.004170463 | 0.01492099 |
| NTproBNP | SHBG-LpA_pPS | 24056 | 0.00679778 | [-0.01 0.02] | 0.291999739 | 0.419749624 |
| NTproBNP | Obesity_pPS | 24056 | -0.002637997 | [-0.02 0.01] | 0.682598396 | 0.768519872 |
| NTproBNP | VAT_Neg_pPS | 24056 | -0.006517535 | [-0.02 0.01] | 0.312350542 | 0.439200325 |
| NTproBNP | Bilirubin_pPS | 24056 | 0.000911263 | [-0.01 0.01] | 0.887667657 | 0.940226926 |
| NTproBNP | Proinsulin_pPS | 24056 | 0.010172714 | [-0. 0.02] | 0.114812488 | 0.210054665 |
| NTproBNP | Hyper_Insulin_pPS | 24056 | -0.005171969 | [-0.02 0.01] | 0.422715555 | 0.553310604 |
| NTproBNP | Beta_Cell_1_pPS | 24056 | -0.003746412 | [-0.02 0.01] | 0.561416965 | 0.674538294 |
| NTproBNP | Beta_Cell_2_pPS | 24056 | -0.017312636 | [-0.03 -0. ] | 0.007277861 | 0.023202686 |
| HB | Cholesterol_pPS | 24295 | 0.000741129 | [-0.01 0.01] | 0.90808588 | 0.940651714 |
| HB | Lipodystrophy_1_pPS | 24295 | 0.020571037 | [0.01 0.03] | 0.001351088 | 0.00524157 |
| HB | Lipodystrophy_2_pPS | 24295 | 0.008072396 | [-0. 0.02] | 0.208557931 | 0.327588555 |
| HB | Liver-Lipid_pPS | 24295 | 0.018225941 | [0.01 0.03] | 0.004518736 | 0.015815576 |
| HB | ASAT_Pos_pPS | 24295 | 0.008388612 | [-0. 0.02] | 0.191279463 | 0.309507472 |
| HB | ALP_Neg_pPS | 24295 | -0.043498793 | [-0.06 -0.03] | 1.21E-11 | 1.49E-10 |
| HB | SHBG-LpA_pPS | 24295 | -0.020286544 | [-0.03 -0.01] | 0.001574613 | 0.005965006 |
| HB | Obesity_pPS | 24295 | 0.022889409 | [0.01 0.04] | 0.000362214 | 0.001631717 |
| HB | VAT_Neg_pPS | 24295 | 0.000802129 | [-0.01 0.01] | 0.900558485 | 0.940651714 |
| HB | Bilirubin_pPS | 24295 | -0.00087797 | [-0.01 0.01] | 0.89121218 | 0.940651714 |
| HB | Proinsulin_pPS | 24295 | -0.002110448 | [-0.01 0.01] | 0.742332152 | 0.821412209 |
| HB | Hyper_Insulin_pPS | 24295 | -0.001027636 | [-0.01 0.01] | 0.872813787 | 0.927544685 |
| HB | Beta_Cell_1_pPS | 24295 | 0.008182048 | [-0. 0.02] | 0.202442634 | 0.324791176 |
| HB | Beta_Cell_2_pPS | 24295 | -0.017467902 | [-0.03 -0. ] | 0.006501341 | 0.021145775 |
| Creatinine | Cholesterol_pPS | 24332 | 0.012300979 | [-0. 0.02] | 0.055138539 | 0.116806641 |
| Creatinine | Lipodystrophy_1_pPS | 24332 | 0.01428726 | [0. 0.03] | 0.025914987 | 0.064687022 |
| Creatinine | Lipodystrophy_2_pPS | 24332 | -0.007708045 | [-0.02 0. ] | 0.229481933 | 0.346916349 |
| Creatinine | Liver-Lipid_pPS | 24332 | 0.028877189 | [0.02 0.04] | 6.70E-06 | 5.02E-05 |
| Creatinine | ASAT_Pos_pPS | 24332 | -0.021923337 | [-0.03 -0.01] | 0.000630281 | 0.00260193 |
| Creatinine | ALP_Neg_pPS | 24332 | -0.005714173 | [-0.02 0.01] | 0.373014376 | 0.504666509 |
| Creatinine | SHBG-LpA_pPS | 24332 | 0.012834216 | [0. 0.03] | 0.045400591 | 0.101520765 |
| Creatinine | Obesity_pPS | 24332 | -0.01115971 | [-0.02 0. ] | 0.081886775 | 0.161764059 |
| Creatinine | VAT_Neg_pPS | 24332 | -0.011551969 | [-0.02 0. ] | 0.071702791 | 0.144301867 |
| Creatinine | Bilirubin_pPS | 24332 | 0.002685916 | [-0.01 0.02] | 0.675411678 | 0.768519872 |
| Creatinine | Proinsulin_pPS | 24332 | -0.002551451 | [-0.02 0.01] | 0.690800266 | 0.772353075 |
| Creatinine | Hyper_Insulin_pPS | 24332 | -0.002596497 | [-0.02 0.01] | 0.685630472 | 0.769243945 |
| Creatinine | Beta_Cell_1_pPS | 24332 | -0.00506747 | [-0.02 0.01] | 0.429516939 | 0.557679251 |
| Creatinine | Beta_Cell_2_pPS | 24332 | -0.009299692 | [-0.02 0. ] | 0.147101983 | 0.249299151 |
| CystatinC | Cholesterol_pPS | 24104 | 0.009402923 | [-0. 0.02] | 0.144552289 | 0.247037093 |
| CystatinC | Lipodystrophy_1_pPS | 24104 | -0.003746233 | [-0.02 0.01] | 0.561044846 | 0.674538294 |
| CystatinC | Lipodystrophy_2_pPS | 24104 | -0.000364669 | [-0.01 0.01] | 0.954876338 | 0.976095812 |
| CystatinC | Liver-Lipid_pPS | 24104 | 0.016116545 | [0. 0.03] | 0.012387712 | 0.035872252 |
| CystatinC | ASAT_Pos_pPS | 24104 | -0.009763122 | [-0.02 0. ] | 0.129787333 | 0.225900114 |
| CystatinC | ALP_Neg_pPS | 24104 | 0.004394712 | [-0.01 0.02] | 0.49529379 | 0.618157365 |
| CystatinC | SHBG-LpA_pPS | 24104 | 0.004251174 | [-0.01 0.02] | 0.509484352 | 0.633412979 |
| CystatinC | Obesity_pPS | 24104 | 0.018089837 | [0.01 0.03] | 0.00499807 | 0.016958798 |
| CystatinC | VAT_Neg_pPS | 24104 | -0.010380824 | [-0.02 0. ] | 0.107224461 | 0.200734165 |
| CystatinC | Bilirubin_pPS | 24104 | 0.002670279 | [-0.01 0.02] | 0.678627087 | 0.768519872 |
| CystatinC | Proinsulin_pPS | 24104 | -0.005138633 | [-0.02 0.01] | 0.425249806 | 0.554374241 |
| CystatinC | Hyper_Insulin_pPS | 24104 | -0.013978028 | [-0.03 -0. ] | 0.030081069 | 0.073939728 |
| CystatinC | Beta_Cell_1_pPS | 24104 | 0.012562337 | [-0. 0.03] | 0.051256616 | 0.110769332 |
| CystatinC | Beta_Cell_2_pPS | 24104 | -0.012773421 | [-0.03 -0. ] | 0.047470061 | 0.104622238 |
| eGFR_LM_revised | Cholesterol_pPS | 24332 | -0.010806382 | [-0.02 0. ] | 0.09203615 | 0.178527954 |
| eGFR_LM_revised | Lipodystrophy_1_pPS | 24332 | -0.016022653 | [-0.03 -0. ] | 0.012486943 | 0.035872252 |
| eGFR_LM_revised | Lipodystrophy_2_pPS | 24332 | 0.008038548 | [-0. 0.02] | 0.210124977 | 0.32844778 |
| eGFR_LM_revised | Liver-Lipid_pPS | 24332 | -0.026221432 | [-0.04 -0.01] | 4.34E-05 | 0.000249543 |
| eGFR_LM_revised | ASAT_Pos_pPS | 24332 | 0.022915721 | [0.01 0.04] | 0.000352905 | 0.001631717 |
| eGFR_LM_revised | ALP_Neg_pPS | 24332 | 0.003333528 | [-0.01 0.02] | 0.603276106 | 0.707469618 |
| eGFR_LM_revised | SHBG-LpA_pPS | 24332 | -0.012771542 | [-0.03 -0. ] | 0.046464187 | 0.10318254 |
| eGFR_LM_revised | Obesity_pPS | 24332 | 0.010070476 | [-0. 0.02] | 0.11641116 | 0.211623247 |
| eGFR_LM_revised | VAT_Neg_pPS | 24332 | 0.013168647 | [0. 0.03] | 0.040065979 | 0.092151752 |
| eGFR_LM_revised | Bilirubin_pPS | 24332 | -0.003138372 | [-0.02 0.01] | 0.624650323 | 0.72875871 |
| eGFR_LM_revised | Proinsulin_pPS | 24332 | 0.001078371 | [-0.01 0.01] | 0.86649121 | 0.925823511 |
| eGFR_LM_revised | Hyper_Insulin_pPS | 24332 | 0.001831913 | [-0.01 0.01] | 0.775189344 | 0.850128015 |
| eGFR_LM_revised | Beta_Cell_1_pPS | 24332 | 0.00515188 | [-0.01 0.02] | 0.42187166 | 0.553310604 |
| eGFR_LM_revised | Beta_Cell_2_pPS | 24332 | 0.008820252 | [-0. 0.02] | 0.169101901 | 0.27773446 |
| SCORE2 | Cholesterol_pPS | 18681 | -0.026582305 | [-0.04 -0.01] | 0.000282112 | 0.001376365 |
| SCORE2 | Lipodystrophy_1_pPS | 18681 | 0.007671125 | [-0.01 0.02] | 0.294767227 | 0.421820509 |
| SCORE2 | Lipodystrophy_2_pPS | 18681 | -0.004504256 | [-0.02 0.01] | 0.538432794 | 0.65424664 |
| SCORE2 | Liver-Lipid_pPS | 18681 | -0.00644135 | [-0.02 0.01] | 0.378992233 | 0.506371365 |
| SCORE2 | ASAT_Pos_pPS | 18681 | 0.014813748 | [0. 0.03] | 0.043038066 | 0.096910889 |
| SCORE2 | ALP_Neg_pPS | 18681 | 0.008833363 | [-0.01 0.02] | 0.22763826 | 0.345752451 |
| SCORE2 | SHBG-LpA_pPS | 18681 | 9.06E-05 | [-0.01 0.01] | 0.99012549 | 0.992414235 |

**Table S5: Associations between pPSs and biochemical traits in SCAPIS**

**a) Spearman Correlation**

| Biochemistry | PPS | Sample_Size | r | CI95% | p | fdr_p_adjusted |
|---|---|---|---|---|---|---|
| eGFR_LM_revised | Cholesterol_pPS | 24332 | -0.010215499 | [-0.02 0. ] | 0.111148173 | 0.176710228 |
| eGFR_LM_revised | Lipodystrophy_1_pPS | 24332 | -0.012718272 | [-0.03 -0. ] | 0.047325401 | 0.088735127 |
| eGFR_LM_revised | Lipodystrophy_2_pPS | 24332 | 0.009874616 | [-0. 0.02] | 0.123586693 | 0.191097776 |
| eGFR_LM_revised | Liver-Lipid_pPS | 24332 | -0.02524998 | [-0.04 -0.01] | 8.21E-05 | 0.000311334 |
| eGFR_LM_revised | ASAT_Pos_pPS | 24332 | 0.022599573 | [0.01 0.04] | 0.000424054 | 0.001434299 |
| eGFR_LM_revised | ALP_Neg_pPS | 24332 | 0.002199054 | [-0.01 0.01] | 0.731656772 | 0.793778574 |
| eGFR_LM_revised | SHBG-LpA_pPS | 24332 | -0.013998571 | [-0.03 -0. ] | 0.029031848 | 0.057563148 |
| eGFR_LM_revised | Obesity_pPS | 24332 | 0.010529064 | [-0. 0.02] | 0.100599318 | 0.164104513 |
| eGFR_LM_revised | VAT_Neg_pPS | 24332 | 0.014970993 | [0. 0.03] | 0.019558162 | 0.041231004 |
| eGFR_LM_revised | Bilirubin_pPS | 24332 | -0.002505285 | [-0.02 0.01] | 0.696036375 | 0.762325554 |
| eGFR_LM_revised | Proinsulin_pPS | 24332 | 0.006296403 | [-0.01 0.02] | 0.326161373 | 0.434462061 |
| eGFR_LM_revised | Hyper_Insulin_pPS | 24332 | 0.003744295 | [-0.01 0.02] | 0.559294347 | 0.656314795 |
| eGFR_LM_revised | Beta_Cell_1_pPS | 24332 | 0.008517181 | [-0. 0.02] | 0.184112295 | 0.26247414 |
| eGFR_LM_revised | Beta_Cell_2_pPS | 24332 | 0.010664803 | [-0. 0.02] | 0.096287601 | 0.159966643 |
| eGFR_LM_revised | Total_GRS | 24332 | 0.018604943 | [0.01 0.03] | 0.003713584 | 0.009351726 |
| SCORE2 | Cholesterol_pPS | 18681 | -0.024755305 | [-0.04 -0.01] | 0.000717713 | 0.002191248 |
| SCORE2 | Lipodystrophy_1_pPS | 18681 | 0.012166722 | [-0. 0.03] | 0.096443657 | 0.159966643 |
| SCORE2 | Lipodystrophy_2_pPS | 18681 | -0.001899697 | [-0.02 0.01] | 0.7952131 | 0.836428413 |
| SCORE2 | Liver-Lipid_pPS | 18681 | -0.005446239 | [-0.02 0.01] | 0.456815943 | 0.56487993 |
| SCORE2 | ASAT_Pos_pPS | 18681 | 0.018273418 | [0. 0.03] | 0.012531202 | 0.027713234 |
| SCORE2 | ALP_Neg_pPS | 18681 | 0.010580276 | [-0. 0.02] | 0.148296466 | 0.218642226 |
| SCORE2 | SHBG-LpA_pPS | 18681 | 0.000730574 | [-0.01 0.02] | 0.920491243 | 0.943500918 |
| SCORE2 | Obesity_pPS | 18681 | 0.022312804 | [0.01 0.04] | 0.002296998 | 0.006191128 |
| SCORE2 | VAT_Neg_pPS | 18681 | 0.008178738 | [-0.01 0.02] | 0.263805359 | 0.362600992 |
| SCORE2 | Bilirubin_pPS | 18681 | -0.002475068 | [-0.02 0.01] | 0.735244421 | 0.795170299 |
| SCORE2 | Proinsulin_pPS | 18681 | 0.016653464 | [0. 0.03] | 0.022879312 | 0.047265644 |
| SCORE2 | Hyper_Insulin_pPS | 18681 | 0.017461989 | [0. 0.03] | 0.017035846 | 0.036505384 |
| SCORE2 | Beta_Cell_1_pPS | 18681 | 0.011796158 | [-0. 0.03] | 0.107024877 | 0.172540105 |
| SCORE2 | Beta_Cell_2_pPS | 18681 | -0.007060519 | [-0.02 0.01] | 0.334715975 | 0.442440657 |
| SCORE2 | Total_GRS | 18681 | 0.021444493 | [0.01 0.04] | 0.003387446 | 0.008593153 |

**b) Partial Correlation**

| Biochemistry | PPS | Sample_Size | r | CI95% | p | fdr_p_adjusted |
|---|---|---|---|---|---|---|
| SCORE2 | Obesity_pPS | 18681 | 0.020583382 | [0.01 0.03] | 0.00493063 | 0.016958798 |
| SCORE2 | VAT_Neg_pPS | 18681 | 0.006152465 | [-0.01 0.02] | 0.400741183 | 0.531023295 |
| SCORE2 | Bilirubin_pPS | 18681 | -0.003092242 | [-0.02 0.01] | 0.67278218 | 0.768519872 |
| SCORE2 | Proinsulin_pPS | 18681 | 0.010232117 | [-0. 0.02] | 0.16225824 | 0.269933554 |
| SCORE2 | Hyper_Insulin_pPS | 18681 | 0.013804223 | [-0. 0.03] | 0.059370402 | 0.124949473 |
| SCORE2 | Beta_Cell_1_pPS | 18681 | 0.008893851 | [-0.01 0.02] | 0.224470336 | 0.342556627 |
| SCORE2 | Beta_Cell_2_pPS | 18681 | -0.01132017 | [-0.03 0. ] | 0.122072311 | 0.217167316 |

**Table S6: Associations between pPSs and selected metabolites in SCAPIS**

**a) Spearman Correlation**

| Metabolomics (select | PPS | Sample_Size | r | CI95% | p | fdr_p_adjusted |
|---|---|---|---|---|---|---|
| Serum_P | Cholesterol_pPS | 4254 | 0.008670619 | [-0.02 0.04] | 0.572368238 | 0.769190689 |
| Serum_P | Lipodystrophy_1_pPS | 4254 | -0.053308191 | [-0.08 -0.02] | 0.000513826 | 0.00506486 |
| Serum_P | Lipodystrophy_2_pPS | 4254 | -0.046550842 | [-0.08 -0.02] | 0.002424274 | 0.015597931 |
| Serum_P | Liver-Lipid_pPS | 4254 | -0.042142968 | [-0.07 -0.01] | 0.006047237 | 0.028976342 |
| Serum_P | ASAT_Pos_pPS | 4254 | -0.044538502 | [-0.07 -0.01] | 0.003714923 | 0.020343628 |
| Serum_P | ALP_Neg_pPS | 4254 | 0.029036778 | [-0. 0.06] | 0.058620979 | 0.185911907 |
| Serum_P | SHBG-LpA_pPS | 4254 | 0.002485303 | [-0.03 0.03] | 0.871446079 | 0.939527804 |
| Serum_P | Obesity_pPS | 4254 | -0.017541289 | [-0.05 0.01] | 0.253359882 | 0.52340814 |
| Serum_P | VAT_Neg_pPS | 4254 | -0.043201906 | [-0.07 -0.01] | 0.004889084 | 0.02444542 |
| Serum_P | Bilirubin_pPS | 4254 | 0.002415927 | [-0.03 0.03] | 0.875004704 | 0.940425616 |
| Serum_P | Proinsulin_pPS | 4254 | 0.006663416 | [-0.02 0.04] | 0.664383254 | 0.824335 |
| Serum_P | Hyper_Insulin_pPS | 4254 | -0.01439497 | [-0.04 0.02] | 0.348590991 | 0.619916968 |
| Serum_P | Beta_Cell_1_pPS | 4254 | 0.014214592 | [-0.02 0.04] | 0.354664638 | 0.624282144 |
| Serum_P | Beta_Cell_2_pPS | 4254 | 0.003369977 | [-0.03 0.03] | 0.826319249 | 0.9137184 |
| Serum_P | Total_GRS | 4254 | -0.031735829 | [-0.06 -0. ] | 0.038744874 | 0.132346353 |
| Serum_C | Cholesterol_pPS | 4254 | -0.137575844 | [-0.17 -0.11] | 2.25E-19 | 1.11E-17 |
| Serum_C | Lipodystrophy_1_pPS | 4254 | 0.025241838 | [-0. 0.06] | 0.100218683 | 0.29551663 |
| Serum_C | Lipodystrophy_2_pPS | 4254 | -0.059085404 | [-0.09 -0.03] | 0.000117877 | 0.001311856 |
| Serum_C | Liver-Lipid_pPS | 4254 | -0.003128088 | [-0.03 0.03] | 0.838607614 | 0.917391664 |
| Serum_C | ASAT_Pos_pPS | 4254 | -0.029023404 | [-0.06 0. ] | 0.058737385 | 0.185911907 |
| Serum_C | ALP_Neg_pPS | 4254 | 0.05435364 | [0.02 0.08] | 0.000397615 | 0.004034618 |
| Serum_C | SHBG-LpA_pPS | 4254 | 0.016473679 | [-0.01 0.05] | 0.283406736 | 0.557170584 |
| Serum_C | Obesity_pPS | 4254 | -0.007461043 | [-0.04 0.02] | 0.627104701 | 0.813350082 |
| Serum_C | VAT_Neg_pPS | 4254 | 0.001811271 | [-0.03 0.03] | 0.906119498 | 0.960971434 |
| Serum_C | Bilirubin_pPS | 4254 | -0.012068435 | [-0.04 0.02] | 0.431971868 | 0.680503628 |
| Serum_C | Proinsulin_pPS | 4254 | -0.001537248 | [-0.03 0.03] | 0.920270914 | 0.96502573 |
| Serum_C | Hyper_Insulin_pPS | 4254 | -0.021392914 | [-0.05 0.01] | 0.163594712 | 0.418075376 |
| Serum_C | Beta_Cell_1_pPS | 4254 | 0.022072621 | [-0.01 0.05] | 0.150617055 | 0.397937135 |
| Serum_C | Beta_Cell_2_pPS | 4254 | 0.025732914 | [-0. 0.06] | 0.093780383 | 0.283809055 |
| Serum_C | Total_GRS | 4254 | -0.009956738 | [-0.04 0.02] | 0.516782109 | 0.734110863 |
| Serum_TG | Cholesterol_pPS | 4254 | -0.088232746 | [-0.12 -0.06] | 8.60E-09 | 2.28E-07 |
| Serum_TG | Lipodystrophy_1_pPS | 4254 | 0.069359198 | [0.04 0.1 ] | 6.14E-06 | 0.000100865 |
| Serum_TG | Lipodystrophy_2_pPS | 4254 | 0.008606302 | [-0.02 0.04] | 0.575220863 | 0.769190689 |
| Serum_TG | Liver-Lipid_pPS | 4254 | -0.04577601 | [-0.08 -0.02] | 0.002862735 | 0.016739724 |
| Serum_TG | ASAT_Pos_pPS | 4254 | 0.038712226 | [0.01 0.07] | 0.011683773 | 0.048565081 |
| Serum_TG | ALP_Neg_pPS | 4254 | 0.011034591 | [-0.02 0.04] | 0.472449034 | 0.705151034 |
| Serum_TG | SHBG-LpA_pPS | 4254 | -0.001413148 | [-0.03 0.03] | 0.92668839 | 0.965799256 |
| Serum_TG | Obesity_pPS | 4254 | 0.030960639 | [0. 0.06] | 0.043760752 | 0.148014307 |
| Serum_TG | VAT_Neg_pPS | 4254 | 0.016329217 | [-0.01 0.05] | 0.287650556 | 0.557170584 |
| Serum_TG | Bilirubin_pPS | 4254 | -0.006859188 | [-0.04 0.02] | 0.655151697 | 0.824335 |
| Serum_TG | Proinsulin_pPS | 4254 | 0.012998246 | [-0.02 0.04] | 0.397346203 | 0.663329569 |
| Serum_TG | Hyper_Insulin_pPS | 4254 | 0.013119858 | [-0.02 0.04] | 0.392944506 | 0.663329569 |
| Serum_TG | Beta_Cell_1_pPS | 4254 | 0.012279811 | [-0.02 0.04] | 0.423950495 | 0.678435595 |
| Serum_TG | Beta_Cell_2_pPS | 4254 | 0.000666744 | [-0.03 0.03] | 0.965372577 | 0.968349016 |
| Serum_TG | Total_GRS | 4254 | 0.03964044 | [0.01 0.07] | 0.009821219 | 0.04183112 |
| Serum_PL | Cholesterol_pPS | 4255 | -0.087252655 | [-0.12 -0.06] | 1.25E-08 | 3.08E-07 |
| Serum_PL | Lipodystrophy_1_pPS | 4255 | 0.003699913 | [-0.03 0.03] | 0.809604795 | 0.909816463 |
| Serum_PL | Lipodystrophy_2_pPS | 4255 | -0.061062875 | [-0.09 -0.03] | 6.89E-05 | 0.000880306 |
| Serum_PL | Liver-Lipid_pPS | 4255 | -0.03740949 | [-0.07 -0.01] | 0.014812706 | 0.06012216 |
| Serum_PL | ASAT_Pos_pPS | 4255 | -0.029303282 | [-0.06 0. ] | 0.056311928 | 0.182014793 |
| Serum_PL | ALP_Neg_pPS | 4255 | 0.052029058 | [0.02 0.08] | 0.000697951 | 0.006117046 |
| Serum_PL | SHBG-LpA_pPS | 4255 | 0.010458891 | [-0.02 0.04] | 0.495812993 | 0.721038307 |
| Serum_PL | Obesity_pPS | 4255 | -0.006851791 | [-0.04 0.02] | 0.655461549 | 0.824335 |
| Serum_PL | VAT_Neg_pPS | 4255 | -0.021806903 | [-0.05 0.01] | 0.155545027 | 0.403481461 |
| Serum_PL | Bilirubin_pPS | 4255 | -0.007233902 | [-0.04 0.02] | 0.637589671 | 0.81772653 |
| Serum_PL | Proinsulin_pPS | 4255 | 0.00902068 | [-0.02 0.04] | 0.556914467 | 0.753472514 |
| Serum_PL | Hyper_Insulin_pPS | 4255 | -0.014628632 | [-0.04 0.02] | 0.340765096 | 0.60913968 |
| Serum_PL | Beta_Cell_1_pPS | 4255 | 0.016278871 | [-0.01 0.05] | 0.289082709 | 0.557170584 |
| Serum_PL | Beta_Cell_2_pPS | 4255 | 0.010989999 | [-0.02 0.04] | 0.474188521 | 0.705151034 |
| Serum_PL | Total_GRS | 4255 | -0.013629928 | [-0.04 0.02] | 0.374750473 | 0.643228425 |
| VLDL_D | Cholesterol_pPS | 4254 | -0.043575647 | [-0.07 -0.01] | 0.004530886 | 0.023330681 |
| VLDL_D | Lipodystrophy_1_pPS | 4254 | 0.058458148 | [0.03 0.09] | 0.000139219 | 0.001455467 |
| VLDL_D | Lipodystrophy_2_pPS | 4254 | 0.022046504 | [-0.01 0.05] | 0.151100767 | 0.397937135 |
| VLDL_D | Liver-Lipid_pPS | 4254 | -0.067502375 | [-0.1 -0.04] | 1.08E-05 | 0.00016223 |
| VLDL_D | ASAT_Pos_pPS | 4254 | 0.048401954 | [0.02 0.08] | 0.001613975 | 0.011229348 |
| VLDL_D | ALP_Neg_pPS | 4254 | -0.015716317 | [-0.05 0.01] | 0.306130443 | 0.565677705 |
| VLDL_D | SHBG-LpA_pPS | 4254 | -0.009783731 | [-0.04 0.02] | 0.524093503 | 0.735009181 |
| VLDL_D | Obesity_pPS | 4254 | 0.039065794 | [0.01 0.07] | 0.010940251 | 0.046029106 |
| VLDL_D | VAT_Neg_pPS | 4254 | 0.022302452 | [-0.01 0.05] | 0.146411135 | 0.394623763 |
| VLDL_D | Bilirubin_pPS | 4254 | -0.002191545 | [-0.03 0.03] | 0.886531202 | 0.943991558 |
| VLDL_D | Proinsulin_pPS | 4254 | 0.010113358 | [-0.02 0.04] | 0.510209088 | 0.730382304 |
| VLDL_D | Hyper_Insulin_pPS | 4254 | 0.020740186 | [-0.01 0.05] | 0.176833623 | 0.436537446 |
| VLDL_D | Beta_Cell_1_pPS | 4254 | 0.008033185 | [-0.02 0.04] | 0.600931995 | 0.796736025 |
| VLDL_D | Beta_Cell_2_pPS | 4254 | -0.006712604 | [-0.04 0.02] | 0.662058966 | 0.824335 |
| VLDL_D | Total_GRS | 4254 | 0.043117866 | [0.01 0.07] | 0.004973074 | 0.024510151 |
| VLDL_P | Cholesterol_pPS | 4255 | -0.109786672 | [-0.14 -0.08] | 7.45E-13 | 2.34E-11 |
| VLDL_P | Lipodystrophy_1_pPS | 4255 | 0.071367542 | [0.04 0.1 ] | 3.27E-06 | 6.26E-05 |
| VLDL_P | Lipodystrophy_2_pPS | 4255 | -0.00407325 | [-0.03 0.03] | 0.79081878 | 0.900437225 |
| VLDL_P | Liver-Lipid_pPS | 4255 | -0.029286771 | [-0.06 0. ] | 0.056450965 | 0.182014793 |
| VLDL_P | ASAT_Pos_pPS | 4255 | 0.033357064 | [0. 0.06] | 0.029796325 | 0.108567634 |
| VLDL_P | ALP_Neg_pPS | 4255 | 0.022457597 | [-0.01 0.05] | 0.143575916 | 0.390029064 |
| VLDL_P | SHBG-LpA_pPS | 4255 | 0.004710553 | [-0.03 0.03] | 0.759034615 | 0.880438492 |
| VLDL_P | Obesity_pPS | 4255 | 0.023430363 | [-0.01 0.05] | 0.127016309 | 0.35339215 |
| VLDL_P | VAT_Neg_pPS | 4255 | 0.014669911 | [-0.02 0.04] | 0.339404211 | 0.60913968 |
| VLDL_P | Bilirubin_pPS | 4255 | -0.01293207 | [-0.04 0.02] | 0.39969825 | 0.663329569 |
| VLDL_P | Proinsulin_pPS | 4255 | 0.012553787 | [-0.02 0.04] | 0.413629169 | 0.669962738 |
| VLDL_P | Hyper_Insulin_pPS | 4255 | 0.005998518 | [-0.02 0.04] | 0.696076864 | 0.848574269 |
| VLDL_P | Beta_Cell_1_pPS | 4255 | 0.017574089 | [-0.01 0.05] | 0.252417323 | 0.52340814 |
| VLDL_P | Beta_Cell_2_pPS | 4255 | 0.005258844 | [-0.02 0.04] | 0.732009493 | 0.87083888 |
| VLDL_P | Total_GRS | 4255 | 0.032837592 | [0. 0.06] | 0.032441181 | 0.114206198 |
| VLDL_C | Cholesterol_pPS | 4254 | -0.116828527 | [-0.15 -0.09] | 2.29E-14 | 9.89E-13 |
| VLDL_C | Lipodystrophy_1_pPS | 4254 | 0.06661252 | [0.04 0.1 ] | 1.41E-05 | 0.000202907 |
| VLDL_C | Lipodystrophy_2_pPS | 4254 | -0.018406302 | [-0.05 0.01] | 0.230698133 | 0.503739594 |
| VLDL_C | Liver-Lipid_pPS | 4254 | -0.021158407 | [-0.05 0.01] | 0.168262375 | 0.420655937 |
| VLDL_C | ASAT_Pos_pPS | 4254 | 0.027613227 | [-0. 0.06] | 0.072132661 | 0.224196107 |
| VLDL_C | ALP_Neg_pPS | 4254 | 0.034253185 | [0. 0.06] | 0.025685836 | 0.097380368 |
| VLDL_C | SHBG-LpA_pPS | 4254 | 0.007665344 | [-0.02 0.04] | 0.617702625 | 0.810294318 |
| VLDL_C | Obesity_pPS | 4254 | 0.017227993 | [-0.01 0.05] | 0.261937829 | 0.531285097 |
| VLDL_C | VAT_Neg_pPS | 4254 | 0.015879415 | [-0.01 0.05] | 0.301137665 | 0.561581051 |
| VLDL_C | Bilirubin_pPS | 4254 | -0.019020356 | [-0.05 0.01] | 0.215510392 | 0.489151876 |
| VLDL_C | Proinsulin_pPS | 4254 | 0.012118038 | [-0.02 0.04] | 0.43008165 | 0.680503628 |
| VLDL_C | Hyper_Insulin_pPS | 4254 | 0.0025021 | [-0.03 0.03] | 0.870584884 | 0.939527804 |
| VLDL_C | Beta_Cell_1_pPS | 4254 | 0.02000835 | [-0.01 0.05] | 0.192607888 | 0.449271456 |
| VLDL_C | Beta_Cell_2_pPS | 4254 | 0.010828694 | [-0.02 0.04] | 0.480752625 | 0.706504658 |
| VLDL_C | Total_GRS | 4254 | 0.025588837 | [-0. 0.06] | 0.095633974 | 0.286384099 |
| VLDL_TG | Cholesterol_pPS | 4254 | -0.080753704 | [-0.11 -0.05] | 1.39E-07 | 3.20E-06 |
| VLDL_TG | Lipodystrophy_1_pPS | 4254 | 0.069031052 | [0.04 0.1 ] | 6.79E-06 | 0.000106523 |
| VLDL_TG | Lipodystrophy_2_pPS | 4254 | 0.013945075 | [-0.02 0.04] | 0.36386337 | 0.627664313 |
| VLDL_TG | Liver-Lipid_pPS | 4254 | -0.049245087 | [-0.08 -0.02] | 0.001334919 | 0.010049968 |
| VLDL_TG | ASAT_Pos_pPS | 4254 | 0.045223422 | [0.02 0.08] | 0.003218401 | 0.018505805 |
| VLDL_TG | ALP_Neg_pPS | 4254 | 0.001766122 | [-0.03 0.03] | 0.908449187 | 0.960971434 |
| VLDL_TG | SHBG-LpA_pPS | 4254 | -0.003099383 | [-0.03 0.03] | 0.840068547 | 0.917391664 |
| VLDL_TG | Obesity_pPS | 4254 | 0.032883855 | [0. 0.06] | 0.032218078 | 0.114206198 |
| VLDL_TG | VAT_Neg_pPS | 4254 | 0.018427155 | [-0.01 0.05] | 0.230170195 | 0.503739594 |

**b) Partial Correlation**

| Metabolomics (selecte | PPS | Sample_Size | r | CI95% | p | fdr_p_adjusted |
|---|---|---|---|---|---|---|
| Serum_P | Cholesterol_pPS | 4254 | 0.011475842 | [-0.02 0.04] | 0.455615062 | 0.722699753 |
| Serum_P | Lipodystrophy_1_pPS | 4254 | -0.044536667 | [-0.07 -0.01] | 0.003769485 | 0.023799491 |
| Serum_P | Lipodystrophy_2_pPS | 4254 | -0.034579897 | [-0.06 -0. ] | 0.024527464 | 0.0999727 |
| Serum_P | Liver-Lipid_pPS | 4254 | -0.041943332 | [-0.07 -0.01] | 0.006371943 | 0.034573773 |
| Serum_P | ASAT_Pos_pPS | 4254 | -0.031980804 | [-0.06 -0. ] | 0.037557358 | 0.137425786 |
| Serum_P | ALP_Neg_pPS | 4254 | 0.032232563 | [0. 0.06] | 0.036079258 | 0.133534727 |
| Serum_P | SHBG-LpA_pPS | 4254 | 0.009947983 | [-0.02 0.04] | 0.517793966 | 0.74670541 |
| Serum_P | Obesity_pPS | 4254 | -0.015926154 | [-0.05 0.01] | 0.300459069 | 0.569104824 |
| Serum_P | VAT_Neg_pPS | 4254 | -0.038768866 | [-0.07 -0.01] | 0.011689838 | 0.054552577 |
| Serum_P | Bilirubin_pPS | 4254 | 0.003471224 | [-0.03 0.03] | 0.821457843 | 0.933298923 |
| Serum_P | Proinsulin_pPS | 4254 | -0.000985887 | [-0.03 0.03] | 0.948895329 | 0.979308641 |
| Serum_P | Hyper_Insulin_pPS | 4254 | 0.001082185 | [-0.03 0.03] | 0.943911475 | 0.979048807 |
| Serum_P | Beta_Cell_1_pPS | 4254 | 0.020362326 | [-0.01 0.05] | 0.185528486 | 0.461243954 |
| Serum_P | Beta_Cell_2_pPS | 4254 | 0.00193338 | [-0.03 0.03] | 0.899975638 | 0.970866349 |
| Serum_C | Cholesterol_pPS | 4254 | -0.135714741 | [-0.17 -0.11] | 7.72E-19 | 3.55E-17 |
| Serum_C | Lipodystrophy_1_pPS | 4254 | 0.033245155 | [0. 0.06] | 0.030624637 | 0.117394442 |
| Serum_C | Lipodystrophy_2_pPS | 4254 | -0.051976309 | [-0.08 -0.02] | 0.000721318 | 0.006277419 |
| Serum_C | Liver-Lipid_pPS | 4254 | -0.001049496 | [-0.03 0.03] | 0.94560304 | 0.979048807 |
| Serum_C | ASAT_Pos_pPS | 4254 | -0.029495285 | [-0.06 0. ] | 0.055117213 | 0.186818344 |
| Serum_C | ALP_Neg_pPS | 4254 | 0.057727076 | [0.03 0.09] | 0.000172611 | 0.001985021 |
| Serum_C | SHBG-LpA_pPS | 4254 | 0.015418761 | [-0.01 0.05] | 0.316122948 | 0.587129581 |
| Serum_C | Obesity_pPS | 4254 | 0.00640931 | [-0.02 0.04] | 0.676910033 | 0.841563824 |
| Serum_C | VAT_Neg_pPS | 4254 | 0.000726391 | [-0.03 0.03] | 0.962334869 | 0.983462418 |
| Serum_C | Bilirubin_pPS | 4254 | -0.012843961 | [-0.04 0.02] | 0.403695133 | 0.684157014 |
| Serum_C | Proinsulin_pPS | 4254 | -0.018197479 | [-0.05 0.01] | 0.236752067 | 0.504862024 |
| Serum_C | Hyper_Insulin_pPS | 4254 | -0.019507043 | [-0.05 0.01] | 0.204689583 | 0.470786042 |
| Serum_C | Beta_Cell_1_pPS | 4254 | 0.017339485 | [-0.01 0.05] | 0.259593977 | 0.532415672 |
| Serum_C | Beta_Cell_2_pPS | 4254 | 0.024652357 | [-0.01 0.05] | 0.108949844 | 0.335747967 |
| Serum_TG | Cholesterol_pPS | 4254 | -0.090381545 | [-0.12 -0.06] | 3.91E-09 | 9.69E-08 |
| Serum_TG | Lipodystrophy_1_pPS | 4254 | 0.062438868 | [0.03 0.09] | 4.84E-05 | 0.000708431 |
| Serum_TG | Lipodystrophy_2_pPS | 4254 | 0.005915915 | [-0.02 0.04] | 0.700529269 | 0.847425962 |
| Serum_TG | Liver-Lipid_pPS | 4254 | -0.0464781 | [-0.08 -0.02] | 0.002500784 | 0.016433726 |
| Serum_TG | ASAT_Pos_pPS | 4254 | 0.024650697 | [-0.01 0.05] | 0.108973676 | 0.335747967 |
| Serum_TG | ALP_Neg_pPS | 4254 | 0.009527788 | [-0.02 0.04] | 0.535632985 | 0.7538823 |
| Serum_TG | SHBG-LpA_pPS | 4254 | -0.007920094 | [-0.04 0.02] | 0.606618993 | 0.810503385 |
| Serum_TG | Obesity_pPS | 4254 | 0.034750197 | [0. 0.06] | 0.023830797 | 0.09837842 |
| Serum_TG | VAT_Neg_pPS | 4254 | 0.01043812 | [-0.02 0.04] | 0.499729143 | 0.741067525 |
| Serum_TG | Bilirubin_pPS | 4254 | -0.010462226 | [-0.04 0.02] | 0.496387946 | 0.739985734 |
| Serum_TG | Proinsulin_pPS | 4254 | 0.007338008 | [-0.02 0.04] | 0.633318236 | 0.821747013 |
| Serum_TG | Hyper_Insulin_pPS | 4254 | 0.003337198 | [-0.03 0.03] | 0.828241893 | 0.933298923 |
| Serum_TG | Beta_Cell_1_pPS | 4254 | 0.006072417 | [-0.02 0.04] | 0.693004656 | 0.847101549 |
| Serum_TG | Beta_Cell_2_pPS | 4254 | -0.001632983 | [-0.03 0.03] | 0.915453198 | 0.970866349 |
| Serum_PL | Cholesterol_pPS | 4255 | -0.084990731 | [-0.11 -0.05] | 3.09E-08 | 7.12E-07 |
| Serum_PL | Lipodystrophy_1_pPS | 4255 | 0.010933134 | [-0.02 0.04] | 0.477155377 | 0.731638244 |
| Serum_PL | Lipodystrophy_2_pPS | 4255 | -0.05165074 | [-0.08 -0.02] | 0.000777919 | 0.006422814 |
| Serum_PL | Liver-Lipid_pPS | 4255 | -0.036357216 | [-0.07 -0.01] | 0.018044687 | 0.081836469 |
| Serum_PL | ASAT_Pos_pPS | 4255 | -0.025716958 | [-0.06 0. ] | 0.094450725 | 0.301120134 |
| Serum_PL | ALP_Neg_pPS | 4255 | 0.054201998 | [0.02 0.08] | 0.000420671 | 0.004104725 |
| Serum_PL | SHBG-LpA_pPS | 4255 | 0.012275532 | [-0.02 0.04] | 0.424768545 | 0.694291734 |
| Serum_PL | Obesity_pPS | 4255 | 0.001226706 | [-0.03 0.03] | 0.936428531 | 0.978993464 |
| Serum_PL | VAT_Neg_pPS | 4255 | -0.01995835 | [-0.05 0.01] | 0.194354087 | 0.467333255 |
| Serum_PL | Bilirubin_pPS | 4255 | -0.007719698 | [-0.04 0.02] | 0.61571232 | 0.813935509 |
| Serum_PL | Proinsulin_pPS | 4255 | -0.003322732 | [-0.03 0.03] | 0.828954944 | 0.933298923 |
| Serum_PL | Hyper_Insulin_pPS | 4255 | -0.007236129 | [-0.04 0.02] | 0.638002339 | 0.821747013 |
| Serum_PL | Beta_Cell_1_pPS | 4255 | 0.015520344 | [-0.01 0.05] | 0.312887579 | 0.585754653 |
| Serum_PL | Beta_Cell_2_pPS | 4255 | 0.010085021 | [-0.02 0.04] | 0.511993398 | 0.74670541 |
| VLDL_D | Cholesterol_pPS | 4254 | -0.045951039 | [-0.08 -0.02] | 0.002799595 | 0.01802939 |
| VLDL_D | Lipodystrophy_1_pPS | 4254 | 0.049059513 | [0.02 0.08] | 0.001416067 | 0.010363033 |
| VLDL_D | Lipodystrophy_2_pPS | 4254 | 0.016872524 | [-0.01 0.05] | 0.272647162 | 0.545294324 |
| VLDL_D | Liver-Lipid_pPS | 4254 | -0.068553061 | [-0.1 -0.04] | 8.12E-06 | 0.000153807 |
| VLDL_D | ASAT_Pos_pPS | 4254 | 0.035095262 | [0. 0.07] | 0.022471638 | 0.095208781 |
| VLDL_D | ALP_Neg_pPS | 4254 | -0.016795205 | [-0.05 0.01] | 0.274851068 | 0.546308914 |
| VLDL_D | SHBG-LpA_pPS | 4254 | -0.016181353 | [-0.05 0.01] | 0.292779594 | 0.561160889 |
| VLDL_D | Obesity_pPS | 4254 | 0.038973152 | [0.01 0.07] | 0.011255412 | 0.054093175 |
| VLDL_D | VAT_Neg_pPS | 4254 | 0.017477559 | [-0.01 0.05] | 0.255818576 | 0.528655484 |
| VLDL_D | Bilirubin_pPS | 4254 | -0.006088655 | [-0.04 0.02] | 0.692225634 | 0.847101549 |
| VLDL_D | Proinsulin_pPS | 4254 | 0.009923457 | [-0.02 0.04] | 0.518826699 | 0.74670541 |
| VLDL_D | Hyper_Insulin_pPS | 4254 | 0.011039156 | [-0.02 0.04] | 0.47294575 | 0.729020706 |
| VLDL_D | Beta_Cell_1_pPS | 4254 | 0.003599776 | [-0.03 0.03] | 0.814963437 | 0.933298923 |
| VLDL_D | Beta_Cell_2_pPS | 4254 | -0.008750515 | [-0.04 0.02] | 0.569427197 | 0.783570759 |
| VLDL_P | Cholesterol_pPS | 4255 | -0.11115647 | [-0.14 -0.08] | 4.17E-13 | 1.22E-11 |
| VLDL_P | Lipodystrophy_1_pPS | 4255 | 0.066219458 | [0.04 0.1 ] | 1.63E-05 | 0.000291689 |
| VLDL_P | Lipodystrophy_2_pPS | 4255 | -0.00559755 | [-0.04 0.02] | 0.715894419 | 0.860141802 |
| VLDL_P | Liver-Lipid_pPS | 4255 | -0.02949699 | [-0.06 0. ] | 0.055074391 | 0.186818344 |
| VLDL_P | ASAT_Pos_pPS | 4255 | 0.020353038 | [-0.01 0.05] | 0.185677166 | 0.461243954 |
| VLDL_P | ALP_Neg_pPS | 4255 | 0.021185589 | [-0.01 0.05] | 0.168319031 | 0.441777123 |
| VLDL_P | SHBG-LpA_pPS | 4255 | -0.001720264 | [-0.03 0.03] | 0.910942071 | 0.970866349 |
| VLDL_P | Obesity_pPS | 4255 | 0.02955484 | [-0. 0.06] | 0.05459912 | 0.186818344 |
| VLDL_P | VAT_Neg_pPS | 4255 | 0.008662212 | [-0.02 0.04] | 0.573284486 | 0.785521721 |
| VLDL_P | Bilirubin_pPS | 4255 | -0.016363445 | [-0.05 0.01] | 0.287324259 | 0.555601364 |
| VLDL_P | Proinsulin_pPS | 4255 | 0.003745779 | [-0.03 0.03] | 0.807580506 | 0.932046319 |
| VLDL_P | Hyper_Insulin_pPS | 4255 | -0.003427713 | [-0.03 0.03] | 0.823638297 | 0.933298923 |
| VLDL_P | Beta_Cell_1_pPS | 4255 | 0.010704666 | [-0.02 0.04] | 0.486410636 | 0.738793513 |
| VLDL_P | Beta_Cell_2_pPS | 4255 | 0.00220771 | [-0.03 0.03] | 0.885860727 | 0.960428128 |
| VLDL_C | Cholesterol_pPS | 4254 | -0.117253308 | [-0.15 -0.09] | 2.02E-14 | 8.14E-13 |
| VLDL_C | Lipodystrophy_1_pPS | 4254 | 0.063024583 | [0.03 0.09] | 4.11E-05 | 0.00065858 |
| VLDL_C | Lipodystrophy_2_pPS | 4254 | -0.019590303 | [-0.05 0.01] | 0.202763398 | 0.469710892 |
| VLDL_C | Liver-Lipid_pPS | 4254 | -0.021330004 | [-0.05 0.01] | 0.16548625 | 0.441777123 |
| VLDL_C | ASAT_Pos_pPS | 4254 | 0.016887866 | [-0.01 0.05] | 0.272211294 | 0.545294324 |
| VLDL_C | ALP_Neg_pPS | 4254 | 0.033515821 | [0. 0.06] | 0.029292515 | 0.117394442 |
| VLDL_C | SHBG-LpA_pPS | 4254 | 0.001794184 | [-0.03 0.03] | 0.907143131 | 0.970866349 |
| VLDL_C | Obesity_pPS | 4254 | 0.024291455 | [-0.01 0.05] | 0.114231072 | 0.340577825 |
| VLDL_C | VAT_Neg_pPS | 4254 | 0.011036961 | [-0.02 0.04] | 0.473033768 | 0.729020706 |
| VLDL_C | Bilirubin_pPS | 4254 | -0.022283305 | [-0.05 0.01] | 0.147378973 | 0.419964862 |
| VLDL_C | Proinsulin_pPS | 4254 | 0.000532179 | [-0.03 0.03] | 0.972400468 | 0.987738015 |
| VLDL_C | Hyper_Insulin_pPS | 4254 | -0.00587131 | [-0.04 0.02] | 0.702679291 | 0.847425962 |
| VLDL_C | Beta_Cell_1_pPS | 4254 | 0.012126465 | [-0.02 0.04] | 0.43046912 | 0.699530799 |
| VLDL_C | Beta_Cell_2_pPS | 4254 | 0.007696708 | [-0.02 0.04] | 0.616805977 | 0.813935509 |
| VLDL_TG | Cholesterol_pPS | 4254 | -0.083039824 | [-0.11 -0.05] | 6.38E-08 | 1.37E-06 |
| VLDL_TG | Lipodystrophy_1_pPS | 4254 | 0.060670928 | [0.03 0.09] | 7.88E-05 | 0.001015255 |
| VLDL_TG | Lipodystrophy_2_pPS | 4254 | 0.010283686 | [-0.02 0.04] | 0.50376603 | 0.741067525 |
| VLDL_TG | Liver-Lipid_pPS | 4254 | -0.050022491 | [-0.08 -0.02] | 0.001137617 | 0.008721734 |
| VLDL_TG | ASAT_Pos_pPS | 4254 | 0.030712004 | [0. 0.06] | 0.045813703 | 0.160347959 |
| VLDL_TG | ALP_Neg_pPS | 4254 | 0.000381369 | [-0.03 0.03] | 0.980219747 | 0.992549555 |
| VLDL_TG | SHBG-LpA_pPS | 4254 | -0.009853784 | [-0.04 0.02] | 0.521766203 | 0.74670541 |
| VLDL_TG | Obesity_pPS | 4254 | 0.035915492 | [0.01 0.07] | 0.019508194 | 0.084887008 |
| VLDL_TG | VAT_Neg_pPS | 4254 | 0.012077877 | [-0.02 0.04] | 0.432318724 | 0.699530799 |
| VLDL_TG | Bilirubin_pPS | 4254 | -0.009741675 | [-0.04 0.02] | 0.526514061 | 0.750166051 |
| VLDL_TG | Proinsulin_pPS | 4254 | 0.007246267 | [-0.02 0.04] | 0.63757135 | 0.821747013 |
| VLDL_TG | Hyper_Insulin_pPS | 4254 | 0.006040891 | [-0.02 0.04] | 0.69451804 | 0.847101549 |
| VLDL_TG | Beta_Cell_1_pPS | 4254 | 0.00680795 | [-0.02 0.04] | 0.65805477 | 0.830955435 |
| VLDL_TG | Beta_Cell_2_pPS | 4254 | -0.003212188 | [-0.03 0.03] | 0.834581097 | 0.93635928 |
| VLDL_PL | Cholesterol_pPS | 4255 | -0.111536893 | [-0.14 -0.08] | 3.47E-13 | 1.12E-11 |
| VLDL_PL | Lipodystrophy_1_pPS | 4255 | 0.064540457 | [0.03 0.09] | 2.66E-05 | 0.000450853 |

**Table S6: Associations between pPSs and selected metabolites in SCAPIS**

**a) Spearman Correlation**

| Metabolomics (select | PPS | Sample_Size | r | CI95% | p | fdr_p_adjusted |
|---|---|---|---|---|---|---|
| VLDL_TG | Bilirubin_pPS | 4254 | -0.005928963 | [-0.04 0.02] | 0.699462055 | 0.849698624 |
| VLDL_TG | Proinsulin_pPS | 4254 | 0.011577591 | [-0.02 0.04] | 0.450933958 | 0.694735958 |
| VLDL_TG | Hyper_Insulin_pPS | 4254 | 0.016370905 | [-0.01 0.05] | 0.28642152 | 0.557170584 |
| VLDL_TG | Beta_Cell_1_pPS | 4254 | 0.012724894 | [-0.02 0.04] | 0.407347893 | 0.663329569 |
| VLDL_TG | Beta_Cell_2_pPS | 4254 | -0.000733228 | [-0.03 0.03] | 0.961922205 | 0.968349016 |
| VLDL_TG | Total_GRS | 4254 | 0.042519548 | [0.01 0.07] | 0.005609732 | 0.027258556 |
| VLDL_PL | Cholesterol_pPS | 4255 | -0.110196088 | [-0.14 -0.08] | 6.12E-13 | 2.11E-11 |
| VLDL_PL | Lipodystrophy_1_pPS | 4255 | 0.069923932 | [0.04 0.1 ] | 5.14E-06 | 8.87E-05 |
| VLDL_PL | Lipodystrophy_2_pPS | 4255 | -0.005155205 | [-0.04 0.02] | 0.737093621 | 0.870881162 |
| VLDL_PL | Liver-Lipid_pPS | 4255 | -0.033000894 | [-0.06 -0. ] | 0.031588872 | 0.113522508 |
| VLDL_PL | ASAT_Pos_pPS | 4255 | 0.033820698 | [0. 0.06] | 0.027594443 | 0.103479161 |
| VLDL_PL | ALP_Neg_pPS | 4255 | 0.021180879 | [-0.01 0.05] | 0.167760684 | 0.420655937 |
| VLDL_PL | SHBG-LpA_pPS | 4255 | 0.004871573 | [-0.03 0.03] | 0.751065699 | 0.879631403 |
| VLDL_PL | Obesity_pPS | 4255 | 0.024597779 | [-0.01 0.05] | 0.109149382 | 0.313804472 |
| VLDL_PL | VAT_Neg_pPS | 4255 | 0.014121761 | [-0.02 0.04] | 0.3577596 | 0.625745312 |
| VLDL_PL | Bilirubin_pPS | 4255 | -0.011404872 | [-0.04 0.02] | 0.457663693 | 0.695568168 |
| VLDL_PL | Proinsulin_pPS | 4255 | 0.01178847 | [-0.02 0.04] | 0.442676429 | 0.694197128 |
| VLDL_PL | Hyper_Insulin_pPS | 4255 | 0.006616168 | [-0.02 0.04] | 0.666581972 | 0.824335 |
| VLDL_PL | Beta_Cell_1_pPS | 4255 | 0.018534451 | [-0.01 0.05] | 0.22741256 | 0.503739584 |
| VLDL_PL | Beta_Cell_2_pPS | 4255 | 0.003441874 | [-0.03 0.03] | 0.822654201 | 0.9137184 |
| VLDL_PL | Total_GRS | 4255 | 0.032509747 | [0. 0.06] | 0.034211854 | 0.118485294 |
| IDL_P | Cholesterol_pPS | 4254 | -0.148350427 | [-0.18 -0.12] | 2.65E-22 | 1.82E-20 |
| IDL_P | Lipodystrophy_1_pPS | 4254 | 0.043673381 | [0.01 0.07] | 0.004441222 | 0.02321548 |
| IDL_P | Lipodystrophy_2_pPS | 4254 | -0.046455832 | [-0.08 -0.02] | 0.002474517 | 0.015597931 |
| IDL_P | Liver-Lipid_pPS | 4254 | 0.006979655 | [-0.02 0.04] | 0.649497107 | 0.824335 |
| IDL_P | ASAT_Pos_pPS | 4254 | -0.015557313 | [-0.05 0.01] | 0.311050403 | 0.570810581 |
| IDL_P | ALP_Neg_pPS | 4254 | 0.052648352 | [0.02 0.08] | 0.000602716 | 0.005776028 |
| IDL_P | SHBG-LpA_pPS | 4254 | 0.018017468 | [-0.01 0.05] | 0.240700158 | 0.512602188 |
| IDL_P | Obesity_pPS | 4254 | -0.005650822 | [-0.04 0.02] | 0.712920876 | 0.859991966 |
| IDL_P | VAT_Neg_pPS | 4254 | 0.012717752 | [-0.02 0.04] | 0.407611213 | 0.663329569 |
| IDL_P | Bilirubin_pPS | 4254 | -0.011517082 | [-0.04 0.02] | 0.453303664 | 0.695044 |
| IDL_P | Proinsulin_pPS | 4254 | -0.002253036 | [-0.03 0.03] | 0.883369901 | 0.943991558 |
| IDL_P | Hyper_Insulin_pPS | 4254 | -0.018228132 | [-0.05 0.01] | 0.235243967 | 0.50839597 |
| IDL_P | Beta_Cell_1_pPS | 4254 | 0.015136407 | [-0.01 0.05] | 0.324324633 | 0.588905255 |
| IDL_P | Beta_Cell_2_pPS | 4254 | 0.023685433 | [-0.01 0.05] | 0.122974208 | 0.347105915 |
| IDL_P | Total_GRS | 4254 | -0.000673186 | [-0.03 0.03] | 0.965038246 | 0.968349016 |
| IDL_C | Cholesterol_pPS | 4255 | -0.148062327 | [-0.18 -0.12] | 3.16E-22 | 1.82E-20 |
| IDL_C | Lipodystrophy_1_pPS | 4255 | 0.035092257 | [0.01 0.07] | 0.022261223 | 0.086293505 |
| IDL_C | Lipodystrophy_2_pPS | 4255 | -0.050995063 | [-0.08 -0.02] | 0.000890818 | 0.007317436 |
| IDL_C | Liver-Lipid_pPS | 4255 | 0.01021588 | [-0.02 0.04] | 0.505880061 | 0.729885038 |
| IDL_C | ASAT_Pos_pPS | 4255 | -0.015906045 | [-0.05 0.01] | 0.300270617 | 0.561581051 |
| IDL_C | ALP_Neg_pPS | 4255 | 0.05029172 | [0.02 0.08] | 0.001049066 | 0.008225631 |
| IDL_C | SHBG-LpA_pPS | 4255 | 0.017754681 | [-0.01 0.05] | 0.247574679 | 0.520812587 |
| IDL_C | Obesity_pPS | 4255 | -0.01042012 | [-0.04 0.02] | 0.497411933 | 0.721038307 |
| IDL_C | VAT_Neg_pPS | 4255 | 0.011674188 | [-0.02 0.04] | 0.447111861 | 0.694735958 |
| IDL_C | Bilirubin_pPS | 4255 | -0.012814422 | [-0.04 0.02] | 0.40400032 | 0.663329569 |
| IDL_C | Proinsulin_pPS | 4255 | -0.005484084 | [-0.04 0.02] | 0.721000911 | 0.86616008 |
| IDL_C | Hyper_Insulin_pPS | 4255 | -0.020578184 | [-0.05 0.01] | 0.180188393 | 0.437781659 |
| IDL_C | Beta_Cell_1_pPS | 4255 | 0.017042976 | [-0.01 0.05] | 0.267040286 | 0.531285097 |
| IDL_C | Beta_Cell_2_pPS | 4255 | 0.024873359 | [-0.01 0.05] | 0.105236165 | 0.305096446 |
| IDL_C | Total_GRS | 4255 | -0.006283219 | [-0.04 0.02] | 0.682420964 | 0.834876711 |
| IDL_TG | Cholesterol_pPS | 4255 | -0.099760962 | [-0.13 -0.07] | 7.39E-11 | 2.12E-09 |
| IDL_TG | Lipodystrophy_1_pPS | 4255 | 0.072372473 | [0.04 0.1 ] | 2.37E-06 | 4.81E-05 |
| IDL_TG | Lipodystrophy_2_pPS | 4255 | 0.000878882 | [-0.03 0.03] | 0.954360379 | 0.968349016 |
| IDL_TG | Liver-Lipid_pPS | 4255 | -0.016426111 | [-0.05 0.01] | 0.284742631 | 0.557170584 |
| IDL_TG | ASAT_Pos_pPS | 4255 | 0.003137167 | [-0.03 0.03] | 0.838126815 | 0.917391664 |
| IDL_TG | ALP_Neg_pPS | 4255 | 0.044649429 | [0.01 0.07] | 0.003626074 | 0.020177347 |
| IDL_TG | SHBG-LpA_pPS | 4255 | 0.006615024 | [-0.02 0.04] | 0.66663613 | 0.824335 |
| IDL_TG | Obesity_pPS | 4255 | 0.011610772 | [-0.02 0.04] | 0.449584004 | 0.694735958 |
| IDL_TG | VAT_Neg_pPS | 4255 | 0.005441961 | [-0.02 0.04] | 0.723055371 | 0.86616008 |
| IDL_TG | Bilirubin_pPS | 4255 | -0.004525063 | [-0.03 0.03] | 0.768246242 | 0.880548018 |
| IDL_TG | Proinsulin_pPS | 4255 | 0.012907662 | [-0.02 0.04] | 0.400588526 | 0.663329569 |
| IDL_TG | Hyper_Insulin_pPS | 4255 | 0.000968746 | [-0.03 0.03] | 0.949699713 | 0.968349016 |
| IDL_TG | Beta_Cell_1_pPS | 4255 | -0.000961846 | [-0.03 0.03] | 0.950057498 | 0.968349016 |
| IDL_TG | Beta_Cell_2_pPS | 4255 | 0.006643997 | [-0.02 0.04] | 0.665264691 | 0.824335 |
| IDL_TG | Total_GRS | 4255 | 0.027059414 | [-0. 0.06] | 0.078000397 | 0.240269079 |
| IDL_PL | Cholesterol_pPS | 4255 | -0.149625475 | [-0.18 -0.12] | 1.14E-22 | 9.84E-21 |
| IDL_PL | Lipodystrophy_1_pPS | 4255 | 0.035842322 | [0.01 0.07] | 0.019555404 | 0.078449005 |
| IDL_PL | Lipodystrophy_2_pPS | 4255 | -0.048359062 | [-0.08 -0.02] | 0.001627442 | 0.011229348 |
| IDL_PL | Liver-Lipid_pPS | 4255 | 0.012457536 | [-0.02 0.04] | 0.4172191 | 0.672619578 |
| IDL_PL | ASAT_Pos_pPS | 4255 | -0.017294327 | [-0.05 0.01] | 0.260048864 | 0.530868983 |
| IDL_PL | ALP_Neg_pPS | 4255 | 0.051961717 | [0.02 0.08] | 0.000709223 | 0.006117046 |
| IDL_PL | SHBG-LpA_pPS | 4255 | 0.017333347 | [-0.01 0.05] | 0.258974973 | 0.530868983 |
| IDL_PL | Obesity_pPS | 4255 | -0.009697828 | [-0.04 0.02] | 0.527694773 | 0.73706355 |
| IDL_PL | VAT_Neg_pPS | 4255 | 0.010870659 | [-0.02 0.04] | 0.479001954 | 0.706504658 |
| IDL_PL | Bilirubin_pPS | 4255 | -0.011038099 | [-0.04 0.02] | 0.472256027 | 0.705151034 |
| IDL_PL | Proinsulin_pPS | 4255 | -0.004592643 | [-0.03 0.03] | 0.764886285 | 0.880438492 |
| IDL_PL | Hyper_Insulin_pPS | 4255 | -0.020227322 | [-0.05 0.01] | 0.187731305 | 0.446671035 |
| IDL_PL | Beta_Cell_1_pPS | 4255 | 0.015476113 | [-0.01 0.05] | 0.313525879 | 0.572309144 |
| IDL_PL | Beta_Cell_2_pPS | 4255 | 0.024133781 | [-0.01 0.05] | 0.115997042 | 0.330735368 |
| IDL_PL | Total_GRS | 4255 | -0.00535174 | [-0.04 0.02] | 0.727462361 | 0.868423926 |
| LDL_D | Cholesterol_pPS | 4254 | 0.059395725 | [0.03 0.09] | 0.000108497 | 0.001247716 |
| LDL_D | Lipodystrophy_1_pPS | 4254 | 0.000674526 | [-0.03 0.03] | 0.964968697 | 0.968349016 |
| LDL_D | Lipodystrophy_2_pPS | 4254 | -0.004445692 | [-0.03 0.03] | 0.772224166 | 0.882176614 |
| LDL_D | Liver-Lipid_pPS | 4254 | 0.025101157 | [-0. 0.06] | 0.102126612 | 0.298590518 |
| LDL_D | ASAT_Pos_pPS | 4254 | 0.015700624 | [-0.01 0.05] | 0.306613713 | 0.565677705 |
| LDL_D | ALP_Neg_pPS | 4254 | 0.009220964 | [-0.02 0.04] | 0.548237654 | 0.752092176 |
| LDL_D | SHBG-LpA_pPS | 4254 | -0.002204309 | [-0.03 0.03] | 0.885874869 | 0.943991558 |
| LDL_D | Obesity_pPS | 4254 | -0.009422077 | [-0.04 0.02] | 0.539546692 | 0.744574435 |
| LDL_D | VAT_Neg_pPS | 4254 | 0.004850233 | [-0.03 0.03] | 0.752148591 | 0.879631403 |
| LDL_D | Bilirubin_pPS | 4254 | -0.001204048 | [-0.03 0.03] | 0.937512016 | 0.965799256 |
| LDL_D | Proinsulin_pPS | 4254 | 0.019262108 | [-0.01 0.05] | 0.209732974 | 0.481974709 |
| LDL_D | Hyper_Insulin_pPS | 4254 | 0.021862226 | [-0.01 0.05] | 0.154547451 | 0.403481461 |
| LDL_D | Beta_Cell_1_pPS | 4254 | -0.033340827 | [-0.06 -0. ] | 0.029895436 | 0.108567634 |
| LDL_D | Beta_Cell_2_pPS | 4254 | -0.009899451 | [-0.04 0.02] | 0.519197248 | 0.734110863 |
| LDL_D | Total_GRS | 4254 | 0.001676911 | [-0.03 0.03] | 0.913054808 | 0.960971434 |
| LDL_P | Cholesterol_pPS | 4255 | -0.154747116 | [-0.18 -0.13] | 3.74E-24 | 7.26E-22 |
| LDL_P | Lipodystrophy_1_pPS | 4255 | 0.041390584 | [0.01 0.07] | 0.007007787 | 0.032017676 |
| LDL_P | Lipodystrophy_2_pPS | 4255 | -0.044032471 | [-0.07 -0.01] | 0.00412101 | 0.021907798 |
| LDL_P | Liver-Lipid_pPS | 4255 | 0.003042702 | [-0.03 0.03] | 0.842936688 | 0.917391664 |
| LDL_P | ASAT_Pos_pPS | 4255 | -0.015953379 | [-0.05 0.01] | 0.298834433 | 0.561581051 |
| LDL_P | ALP_Neg_pPS | 4255 | 0.048701008 | [0.02 0.08] | 0.001507428 | 0.010834638 |
| LDL_P | SHBG-LpA_pPS | 4255 | 0.017081819 | [-0.01 0.05] | 0.265951516 | 0.531285097 |
| LDL_P | Obesity_pPS | 4255 | -0.004778104 | [-0.03 0.03] | 0.755688353 | 0.880438492 |
| LDL_P | VAT_Neg_pPS | 4255 | 0.010171116 | [-0.02 0.04] | 0.507746113 | 0.729885038 |
| LDL_P | Bilirubin_pPS | 4255 | -0.012839872 | [-0.04 0.02] | 0.40306735 | 0.663329569 |
| LDL_P | Proinsulin_pPS | 4255 | -0.003390268 | [-0.03 0.03] | 0.825270046 | 0.9137184 |
| LDL_P | Hyper_Insulin_pPS | 4255 | -0.020066485 | [-0.05 0.01] | 0.191265883 | 0.449271456 |
| LDL_P | Beta_Cell_1_pPS | 4255 | 0.022052711 | [-0.01 0.05] | 0.150937534 | 0.397937135 |
| LDL_P | Beta_Cell_2_pPS | 4255 | 0.022829188 | [-0.01 0.05] | 0.137064886 | 0.375296711 |
| LDL_P | Total_GRS | 4255 | -0.000663398 | [-0.03 0.03] | 0.965542207 | 0.968349016 |
| LDL_C | Cholesterol_pPS | 4254 | -0.153993054 | [-0.18 -0.12] | 6.31E-24 | 7.26E-22 |
| LDL_C | Lipodystrophy_1_pPS | 4254 | 0.037590185 | [0.01 0.07] | 0.01434857 | 0.058931625 |
| LDL_C | Lipodystrophy_2_pPS | 4254 | -0.044029959 | [-0.07 -0.01] | 0.004127556 | 0.021907798 |

**b) Partial Correlation**

| Metabolomics (selecte | PPS | Sample_Size | r | CI95% | p | fdr_p_adjusted |
|---|---|---|---|---|---|---|
| VLDL_PL | Lipodystrophy_2_pPS | 4255 | -0.006439016 | [-0.04 0.02] | 0.67546159 | 0.841563824 |
| VLDL_PL | Liver-Lipid_pPS | 4255 | -0.033150835 | [-0.06 -0. ] | 0.031080992 | 0.11774211 |
| VLDL_PL | ASAT_Pos_pPS | 4255 | 0.020981213 | [-0.01 0.05] | 0.172462954 | 0.447847347 |
| VLDL_PL | ALP_Neg_pPS | 4255 | 0.020243331 | [-0.01 0.05] | 0.188059745 | 0.462253725 |
| VLDL_PL | SHBG-LpA_pPS | 4255 | -0.001352255 | [-0.03 0.03] | 0.929938187 | 0.975374906 |
| VLDL_PL | Obesity_pPS | 4255 | 0.030915791 | [0. 0.06] | 0.044367574 | 0.15873732 |
| VLDL_PL | VAT_Neg_pPS | 4255 | 0.008160781 | [-0.02 0.04] | 0.59568453 | 0.799210078 |
| VLDL_PL | Bilirubin_pPS | 4255 | -0.014866735 | [-0.04 0.02] | 0.333705512 | 0.593663949 |
| VLDL_PL | Proinsulin_pPS | 4255 | 0.002929109 | [-0.03 0.03] | 0.848957442 | 0.941866437 |
| VLDL_PL | Hyper_Insulin_pPS | 4255 | -0.002423076 | [-0.03 0.03] | 0.874813938 | 0.954881655 |
| VLDL_PL | Beta_Cell_1_pPS | 4255 | 0.012415635 | [-0.02 0.04] | 0.419501645 | 0.689181275 |
| VLDL_PL | Beta_Cell_2_pPS | 4255 | 0.000232476 | [-0.03 0.03] | 0.98794013 | 0.994114756 |
| IDL_P | Cholesterol_pPS | 4254 | -0.147002256 | [-0.18 -0.12] | 7.32E-22 | 4.71E-20 |
| IDL_P | Lipodystrophy_1_pPS | 4254 | 0.049327257 | [0.02 0.08] | 0.001332929 | 0.009981467 |
| IDL_P | Lipodystrophy_2_pPS | 4254 | -0.042528202 | [-0.07 -0.01] | 0.005673545 | 0.032076082 |
| IDL_P | Liver-Lipid_pPS | 4254 | 0.009183598 | [-0.02 0.04] | 0.55047238 | 0.770661332 |
| IDL_P | ASAT_Pos_pPS | 4254 | -0.019863832 | [-0.05 0.01] | 0.196528387 | 0.467333255 |
| IDL_P | ALP_Neg_pPS | 4254 | 0.055019958 | [0.02 0.09] | 0.000344063 | 0.003692945 |
| IDL_P | SHBG-LpA_pPS | 4254 | 0.01479852 | [-0.02 0.04] | 0.335985702 | 0.593781861 |
| IDL_P | Obesity_pPS | 4254 | 0.008229603 | [-0.02 0.04] | 0.592630364 | 0.798439235 |
| IDL_P | VAT_Neg_pPS | 4254 | 0.01051331 | [-0.02 0.04] | 0.494287597 | 0.739985734 |
| IDL_P | Bilirubin_pPS | 4254 | -0.012542019 | [-0.04 0.02] | 0.414838817 | 0.685015893 |
| IDL_P | Proinsulin_pPS | 4254 | -0.016459063 | [-0.05 0.01] | 0.284573644 | 0.555601364 |
| IDL_P | Hyper_Insulin_pPS | 4254 | -0.020831561 | [-0.05 0.01] | 0.175596425 | 0.44874642 |
| IDL_P | Beta_Cell_1_pPS | 4254 | 0.008405039 | [-0.02 0.04] | 0.584767536 | 0.794494289 |
| IDL_P | Beta_Cell_2_pPS | 4254 | 0.023816435 | [-0.01 0.05] | 0.121486882 | 0.358887852 |
| IDL_C | Cholesterol_pPS | 4255 | -0.14605175 | [-0.18 -0.12] | 1.33E-21 | 7.14E-20 |
| IDL_C | Lipodystrophy_1_pPS | 4255 | 0.040933908 | [0.01 0.07] | 0.007753791 | 0.040269688 |
| IDL_C | Lipodystrophy_2_pPS | 4255 | -0.046638845 | [-0.08 -0.02] | 0.002412763 | 0.016185619 |
| IDL_C | Liver-Lipid_pPS | 4255 | 0.012782419 | [-0.02 0.04] | 0.405896242 | 0.684285812 |
| IDL_C | ASAT_Pos_pPS | 4255 | -0.018057558 | [-0.05 0.01] | 0.240321252 | 0.509101599 |
| IDL_C | ALP_Neg_pPS | 4255 | 0.053324784 | [0.02 0.08] | 0.000521225 | 0.004795266 |
| IDL_C | SHBG-LpA_pPS | 4255 | 0.015180771 | [-0.01 0.05] | 0.323594167 | 0.589063983 |
| IDL_C | Obesity_pPS | 4255 | 0.004070716 | [-0.03 0.03] | 0.791259161 | 0.920494699 |
| IDL_C | VAT_Neg_pPS | 4255 | 0.009867591 | [-0.02 0.04] | 0.521133714 | 0.74670541 |
| IDL_C | Bilirubin_pPS | 4255 | -0.013654212 | [-0.04 0.02] | 0.374633423 | 0.641659373 |
| IDL_C | Proinsulin_pPS | 4255 | -0.019656275 | [-0.05 0.01] | 0.20119325 | 0.469450917 |
| IDL_C | Hyper_Insulin_pPS | 4255 | -0.021901711 | [-0.05 0.01] | 0.154387673 | 0.426545975 |
| IDL_C | Beta_Cell_1_pPS | 4255 | 0.011119271 | [-0.02 0.04] | 0.469686783 | 0.729020706 |
| IDL_C | Beta_Cell_2_pPS | 4255 | 0.024893932 | [-0.01 0.05] | 0.105482749 | 0.33299456 |
| IDL_TG | Cholesterol_pPS | 4255 | -0.101425728 | [-0.13 -0.07] | 3.80E-11 | 1.02E-09 |
| IDL_TG | Lipodystrophy_1_pPS | 4255 | 0.072598864 | [0.04 0.1 ] | 2.28E-06 | 4.60E-05 |
| IDL_TG | Lipodystrophy_2_pPS | 4255 | 0.001624959 | [-0.03 0.03] | 0.915857146 | 0.970866349 |
| IDL_TG | Liver-Lipid_pPS | 4255 | -0.016714773 | [-0.05 0.01] | 0.277099752 | 0.547399509 |
| IDL_TG | ASAT_Pos_pPS | 4255 | -0.01001825 | [-0.04 0.02] | 0.514791394 | 0.74670541 |
| IDL_TG | ALP_Neg_pPS | 4255 | 0.042288212 | [0.01 0.07] | 0.005945404 | 0.033007243 |
| IDL_TG | SHBG-LpA_pPS | 4255 | 0.001096745 | [-0.03 0.03] | 0.943151417 | 0.979048807 |
| IDL_TG | Obesity_pPS | 4255 | 0.017236588 | [-0.01 0.05] | 0.262376027 | 0.5347157 |
| IDL_TG | VAT_Neg_pPS | 4255 | 0.000672234 | [-0.03 0.03] | 0.965137031 | 0.983462418 |
| IDL_TG | Bilirubin_pPS | 4255 | -0.006256572 | [-0.04 0.02] | 0.68415381 | 0.844730591 |
| IDL_TG | Proinsulin_pPS | 4255 | 0.006245054 | [-0.02 0.04] | 0.684703988 | 0.844730591 |
| IDL_TG | Hyper_Insulin_pPS | 4255 | -0.006905156 | [-0.04 0.02] | 0.65345095 | 0.830955435 |
| IDL_TG | Beta_Cell_1_pPS | 4255 | -0.00723896 | [-0.04 0.02] | 0.637870893 | 0.821747013 |
| IDL_TG | Beta_Cell_2_pPS | 4255 | 0.007641389 | [-0.02 0.04] | 0.619298757 | 0.813935509 |
| IDL_PL | Cholesterol_pPS | 4255 | -0.147936875 | [-0.18 -0.12] | 3.97E-22 | 3.19E-20 |
| IDL_PL | Lipodystrophy_1_pPS | 4255 | 0.041808424 | [0.01 0.07] | 0.00653727 | 0.034573773 |
| IDL_PL | Lipodystrophy_2_pPS | 4255 | -0.043818095 | [-0.07 -0.01] | 0.004366597 | 0.02703931 |
| IDL_PL | Liver-Lipid_pPS | 4255 | 0.015037137 | [-0.02 0.05] | 0.328193879 | 0.593246997 |
| IDL_PL | ASAT_Pos_pPS | 4255 | -0.019928909 | [-0.05 0.01] | 0.19501305 | 0.467333255 |
| IDL_PL | ALP_Neg_pPS | 4255 | 0.054657737 | [0.02 0.08] | 0.000375882 | 0.003881426 |
| IDL_PL | SHBG-LpA_pPS | 4255 | 0.014751699 | [-0.02 0.04] | 0.337459877 | 0.593781861 |
| IDL_PL | Obesity_pPS | 4255 | 0.004836153 | [-0.03 0.03] | 0.753183233 | 0.888369967 |
| IDL_PL | VAT_Neg_pPS | 4255 | 0.008824423 | [-0.02 0.04] | 0.566124581 | 0.782369592 |
| IDL_PL | Bilirubin_pPS | 4255 | -0.011737424 | [-0.04 0.02] | 0.445353341 | 0.713451621 |
| IDL_PL | Proinsulin_pPS | 4255 | -0.018567524 | [-0.05 0.01] | 0.227297326 | 0.487931594 |
| IDL_PL | Hyper_Insulin_pPS | 4255 | -0.02170015 | [-0.05 0.01] | 0.158217159 | 0.431745128 |
| IDL_PL | Beta_Cell_1_pPS | 4255 | 0.009514682 | [-0.02 0.04] | 0.536146108 | 0.7538823 |
| IDL_PL | Beta_Cell_2_pPS | 4255 | 0.024328119 | [-0.01 0.05] | 0.113642797 | 0.340577825 |
| LDL_D | Cholesterol_pPS | 4254 | 0.059906279 | [0.03 0.09] | 9.70E-05 | 0.001200691 |
| LDL_D | Lipodystrophy_1_pPS | 4254 | -0.002682646 | [-0.03 0.03] | 0.861548532 | 0.950063792 |
| LDL_D | Lipodystrophy_2_pPS | 4254 | -0.012566217 | [-0.04 0.02] | 0.413939094 | 0.685015893 |
| LDL_D | Liver-Lipid_pPS | 4254 | 0.023618952 | [-0.01 0.05] | 0.124607407 | 0.364759864 |
| LDL_D | ASAT_Pos_pPS | 4254 | 0.017466527 | [-0.01 0.05] | 0.256118806 | 0.528655484 |
| LDL_D | ALP_Neg_pPS | 4254 | 0.002611912 | [-0.03 0.03] | 0.865163742 | 0.950794282 |
| LDL_D | SHBG-LpA_pPS | 4254 | -0.003408748 | [-0.03 0.03] | 0.824618571 | 0.933298923 |
| LDL_D | Obesity_pPS | 4254 | -0.020169783 | [-0.05 0.01] | 0.189721974 | 0.462806634 |
| LDL_D | VAT_Neg_pPS | 4254 | 0.010551426 | [-0.02 0.04] | 0.49272351 | 0.739985734 |
| LDL_D | Bilirubin_pPS | 4254 | -1.64E-05 | [-0.03 0.03] | 0.999150638 | 0.999150638 |
| LDL_D | Proinsulin_pPS | 4254 | 0.030750357 | [0. 0.06] | 0.045543384 | 0.160347959 |
| LDL_D | Hyper_Insulin_pPS | 4254 | 0.019683498 | [-0.01 0.05] | 0.200623065 | 0.469450917 |
| LDL_D | Beta_Cell_1_pPS | 4254 | -0.040002165 | [-0.07 -0.01] | 0.009243573 | 0.04579124 |
| LDL_D | Beta_Cell_2_pPS | 4254 | -0.005974004 | [-0.04 0.02] | 0.697732899 | 0.847425962 |
| LDL_P | Cholesterol_pPS | 4255 | -0.153425624 | [-0.18 -0.12] | 1.07E-23 | 2.36E-21 |
| LDL_P | Lipodystrophy_1_pPS | 4255 | 0.047026669 | [0.02 0.08] | 0.002218324 | 0.015528269 |
| LDL_P | Lipodystrophy_2_pPS | 4255 | -0.038783967 | [-0.07 -0.01] | 0.011647338 | 0.054552577 |
| LDL_P | Liver-Lipid_pPS | 4255 | 0.005373605 | [-0.02 0.04] | 0.726796423 | 0.866772031 |
| LDL_P | ASAT_Pos_pPS | 4255 | -0.020328113 | [-0.05 0.01] | 0.186216503 | 0.461243954 |
| LDL_P | ALP_Neg_pPS | 4255 | 0.051686062 | [0.02 0.08] | 0.000771463 | 0.006422814 |
| LDL_P | SHBG-LpA_pPS | 4255 | 0.014090008 | [-0.02 0.04] | 0.359579341 | 0.619168704 |
| LDL_P | Obesity_pPS | 4255 | 0.010276398 | [-0.02 0.04] | 0.504017975 | 0.741067525 |
| LDL_P | VAT_Neg_pPS | 4255 | 0.006723348 | [-0.02 0.04] | 0.662001244 | 0.83267344 |
| LDL_P | Bilirubin_pPS | 4255 | -0.014113206 | [-0.04 0.02] | 0.358788784 | 0.619168704 |
| LDL_P | Proinsulin_pPS | 4255 | -0.018871597 | [-0.05 0.01] | 0.219775264 | 0.487931594 |
| LDL_P | Hyper_Insulin_pPS | 4255 | -0.022061855 | [-0.05 0.01] | 0.151395596 | 0.42390767 |
| LDL_P | Beta_Cell_1_pPS | 4255 | 0.01691747 | [-0.01 0.05] | 0.271314887 | 0.545294324 |
| LDL_P | Beta_Cell_2_pPS | 4255 | 0.021526136 | [-0.01 0.05] | 0.161580721 | 0.437218422 |
| LDL_C | Cholesterol_pPS | 4254 | -0.152364411 | [-0.18 -0.12] | 2.20E-23 | 2.36E-21 |
| LDL_C | Lipodystrophy_1_pPS | 4254 | 0.043108049 | [0.01 0.07] | 0.005050049 | 0.02956574 |
| LDL_C | Lipodystrophy_2_pPS | 4254 | -0.03921673 | [-0.07 -0.01] | 0.010756204 | 0.052477238 |
| LDL_C | Liver-Lipid_pPS | 4254 | 0.011641531 | [-0.02 0.04] | 0.449134054 | 0.715946364 |
| LDL_C | ASAT_Pos_pPS | 4254 | -0.018597629 | [-0.05 0.01] | 0.226599445 | 0.487931594 |
| LDL_C | ALP_Neg_pPS | 4254 | 0.050769667 | [0.02 0.08] | 0.000957437 | 0.007519387 |
| LDL_C | SHBG-LpA_pPS | 4254 | 0.014955501 | [-0.02 0.05] | 0.330884162 | 0.593246997 |
| LDL_C | Obesity_pPS | 4254 | 0.006812328 | [-0.02 0.04] | 0.65784889 | 0.830955435 |
| LDL_C | VAT_Neg_pPS | 4254 | 0.008979624 | [-0.02 0.04] | 0.559360966 | 0.776354444 |
| LDL_C | Bilirubin_pPS | 4254 | -0.014585449 | [-0.04 0.02] | 0.342990538 | 0.596988936 |
| LDL_C | Proinsulin_pPS | 4254 | -0.022621809 | [-0.05 0.01] | 0.141327151 | 0.406315558 |
| LDL_C | Hyper_Insulin_pPS | 4254 | -0.022154209 | [-0.05 0.01] | 0.14973843 | 0.422945389 |
| LDL_C | Beta_Cell_1_pPS | 4254 | 0.015986794 | [-0.01 0.05] | 0.298622252 | 0.568972574 |
| LDL_C | Beta_Cell_2_pPS | 4254 | 0.023066486 | [-0.01 0.05] | 0.133669528 | 0.387762055 |
| LDL_TG | Cholesterol_pPS | 4254 | -0.113546745 | [-0.14 -0.08] | 1.30E-13 | 4.66E-12 |
| LDL_TG | Lipodystrophy_1_pPS | 4254 | 0.062865118 | [0.03 0.09] | 4.30E-05 | 0.00065858 |
| LDL_TG | Lipodystrophy_2_pPS | 4254 | -0.014932492 | [-0.05 0.02] | 0.331628756 | 0.593246997 |
| LDL_TG | Liver-Lipid_pPS | 4254 | -0.018858793 | [-0.05 0.01] | 0.220142915 | 0.487931594 |

**Table S6: Associations between pPSs and selected metabolites in SCAPIS**

**a) Spearman Correlation**

| Metabolomics (select | PPS | Sample_Size | r | CI95% | p | fdr_p_adjusted |
|---|---|---|---|---|---|---|
| LDL_C | Liver-Lipid_pPS | 4254 | 0.009121113 | [-0.02 0.04] | 0.552578205 | 0.752092176 |
| LDL_C | ASAT_Pos_pPS | 4254 | -0.014990235 | [-0.05 0.02] | 0.329019547 | 0.594302322 |
| LDL_C | ALP_Neg_pPS | 4254 | 0.04716263 | [0.02 0.08] | 0.002122478 | 0.014081824 |
| LDL_C | SHBG-LpA_pPS | 4254 | 0.017635798 | [-0.01 0.05] | 0.250811075 | 0.52340814 |
| LDL_C | Obesity_pPS | 4254 | -0.008640992 | [-0.04 0.02] | 0.573681438 | 0.769190689 |
| LDL_C | VAT_Neg_pPS | 4254 | 0.012155094 | [-0.02 0.04] | 0.428672736 | 0.680503628 |
| LDL_C | Bilirubin_pPS | 4254 | -0.013513749 | [-0.04 0.02] | 0.378891896 | 0.646405804 |
| LDL_C | Proinsulin_pPS | 4254 | -0.007553087 | [-0.04 0.02] | 0.622861161 | 0.813350082 |
| LDL_C | Hyper_Insulin_pPS | 4254 | -0.020285061 | [-0.05 0.01] | 0.186526198 | 0.446671035 |
| LDL_C | Beta_Cell_1_pPS | 4254 | 0.020725251 | [-0.01 0.05] | 0.17714563 | 0.436537446 |
| LDL_C | Beta_Cell_2_pPS | 4254 | 0.023636462 | [-0.01 0.05] | 0.123750805 | 0.347105915 |
| LDL_C | Total_GRS | 4254 | -0.003763164 | [-0.03 0.03] | 0.806436363 | 0.909217468 |
| LDL_TG | Cholesterol_pPS | 4254 | -0.112611992 | [-0.14 -0.08] | 1.90E-13 | 7.28E-12 |
| LDL_TG | Lipodystrophy_1_pPS | 4254 | 0.059559519 | [0.03 0.09] | 0.000103835 | 0.001247716 |
| LDL_TG | Lipodystrophy_2_pPS | 4254 | -0.01843372 | [-0.05 0.01] | 0.230004168 | 0.503739594 |
| LDL_TG | Liver-Lipid_pPS | 4254 | -0.019210734 | [-0.05 0.01] | 0.21095125 | 0.481974709 |
| LDL_TG | ASAT_Pos_pPS | 4254 | -0.009176973 | [-0.04 0.02] | 0.550147898 | 0.752092176 |
| LDL_TG | ALP_Neg_pPS | 4254 | 0.052287601 | [0.02 0.08] | 0.000657168 | 0.006117046 |
| LDL_TG | SHBG-LpA_pPS | 4254 | 0.009432665 | [-0.02 0.04] | 0.539091028 | 0.744574435 |
| LDL_TG | Obesity_pPS | 4254 | 0.009813691 | [-0.02 0.04] | 0.522823622 | 0.735009181 |
| LDL_TG | VAT_Neg_pPS | 4254 | 0.001198389 | [-0.03 0.03] | 0.937805075 | 0.965799256 |
| LDL_TG | Bilirubin_pPS | 4254 | -0.005193092 | [-0.04 0.02] | 0.735263733 | 0.870881162 |
| LDL_TG | Proinsulin_pPS | 4254 | 0.010816613 | [-0.02 0.04] | 0.481242303 | 0.706504658 |
| LDL_TG | Hyper_Insulin_pPS | 4254 | -0.007993132 | [-0.04 0.02] | 0.602748123 | 0.796736025 |
| LDL_TG | Beta_Cell_1_pPS | 4254 | 0.003373793 | [-0.03 0.03] | 0.826125735 | 0.9137184 |
| LDL_TG | Beta_Cell_2_pPS | 4254 | 0.011573986 | [-0.02 0.04] | 0.451074941 | 0.694735958 |
| LDL_TG | Total_GRS | 4254 | 0.017012523 | [-0.01 0.05] | 0.267952484 | 0.531285097 |
| LDL_PL | Cholesterol_pPS | 4255 | -0.154368027 | [-0.18 -0.12] | 4.84E-24 | 7.26E-22 |
| LDL_PL | Lipodystrophy_1_pPS | 4255 | 0.041357601 | [0.01 0.07] | 0.007053169 | 0.032017676 |
| LDL_PL | Lipodystrophy_2_pPS | 4255 | -0.049076875 | [-0.08 -0.02] | 0.001384947 | 0.010166103 |
| LDL_PL | Liver-Lipid_pPS | 4255 | -0.00459453 | [-0.03 0.03] | 0.764792508 | 0.880438492 |
| LDL_PL | ASAT_Pos_pPS | 4255 | -0.013956378 | [-0.04 0.02] | 0.363418055 | 0.627664313 |
| LDL_PL | ALP_Neg_pPS | 4255 | 0.049222551 | [0.02 0.08] | 0.001339996 | 0.010049968 |
| LDL_PL | SHBG-LpA_pPS | 4255 | 0.017900315 | [-0.01 0.05] | 0.243717085 | 0.515842909 |
| LDL_PL | Obesity_pPS | 4255 | -0.003565106 | [-0.03 0.03] | 0.816415842 | 0.9137184 |
| LDL_PL | VAT_Neg_pPS | 4255 | 0.007346124 | [-0.02 0.04] | 0.632380048 | 0.814071331 |
| LDL_PL | Bilirubin_pPS | 4255 | -0.014081751 | [-0.04 0.02] | 0.359123397 | 0.625745312 |
| LDL_PL | Proinsulin_pPS | 4255 | -0.003091387 | [-0.03 0.03] | 0.840457047 | 0.917391664 |
| LDL_PL | Hyper_Insulin_pPS | 4255 | -0.019476007 | [-0.05 0.01] | 0.204662006 | 0.473881827 |
| LDL_PL | Beta_Cell_1_pPS | 4255 | 0.023358056 | [-0.01 0.05] | 0.1281937 | 0.353814612 |
| LDL_PL | Beta_Cell_2_pPS | 4255 | 0.021184645 | [-0.01 0.05] | 0.167685105 | 0.420655937 |
| LDL_PL | Total_GRS | 4255 | -0.001423729 | [-0.03 0.03] | 0.926132349 | 0.965799256 |
| HDL_D | Cholesterol_pPS | 4254 | 0.029898792 | [-0. 0.06] | 0.051512105 | 0.169254059 |
| HDL_D | Lipodystrophy_1_pPS | 4254 | -0.065451105 | [-0.1 -0.04] | 1.99E-05 | 0.000274398 |
| HDL_D | Lipodystrophy_2_pPS | 4254 | -0.04077578 | [-0.07 -0.01] | 0.007905616 | 0.034967146 |
| HDL_D | Liver-Lipid_pPS | 4254 | 0.020380755 | [-0.01 0.05] | 0.184456344 | 0.445017054 |
| HDL_D | ASAT_Pos_pPS | 4254 | -0.052067164 | [-0.08 -0.02] | 0.000692657 | 0.006117046 |
| HDL_D | ALP_Neg_pPS | 4254 | 0.018207367 | [-0.01 0.05] | 0.235777841 | 0.50839597 |
| HDL_D | SHBG-LpA_pPS | 4254 | 0.009095043 | [-0.02 0.04] | 0.55371424 | 0.752092176 |
| HDL_D | Obesity_pPS | 4254 | -0.03429266 | [-0.06 -0. ] | 0.025515893 | 0.097380368 |
| HDL_D | VAT_Neg_pPS | 4254 | -0.016169035 | [-0.05 0.01] | 0.292406079 | 0.559363939 |
| HDL_D | Bilirubin_pPS | 4254 | -0.008134624 | [-0.04 0.02] | 0.596343595 | 0.794357298 |
| HDL_D | Proinsulin_pPS | 4254 | -0.007405235 | [-0.04 0.02] | 0.629683641 | 0.813636166 |
| HDL_D | Hyper_Insulin_pPS | 4254 | -0.012829041 | [-0.04 0.02] | 0.403519687 | 0.663329569 |
| HDL_D | Beta_Cell_1_pPS | 4254 | -0.00958714 | [-0.04 0.02] | 0.532465484 | 0.740728193 |
| HDL_D | Beta_Cell_2_pPS | 4254 | 0.011466139 | [-0.02 0.04] | 0.455304185 | 0.695044 |
| HDL_D | Total_GRS | 4254 | -0.045966767 | [-0.08 -0.02] | 0.002748542 | 0.016635909 |
| HDL_P | Cholesterol_pPS | 4255 | 0.03573556 | [0.01 0.07] | 0.019922104 | 0.079001466 |
| HDL_P | Lipodystrophy_1_pPS | 4255 | -0.063266437 | [-0.09 -0.03] | 3.72E-05 | 0.000494107 |
| HDL_P | Lipodystrophy_2_pPS | 4255 | -0.040242013 | [-0.07 -0.01] | 0.008752156 | 0.037940965 |
| HDL_P | Liver-Lipid_pPS | 4255 | -0.041632216 | [-0.07 -0.01] | 0.006683237 | 0.031158334 |
| HDL_P | ASAT_Pos_pPS | 4255 | -0.046075879 | [-0.08 -0.02] | 0.002681989 | 0.016522966 |
| HDL_P | ALP_Neg_pPS | 4255 | 0.021505058 | [-0.01 0.05] | 0.161347943 | 0.415410748 |
| HDL_P | SHBG-LpA_pPS | 4255 | 2.74E-05 | [-0.03 0.03] | 0.998576519 | 0.998576519 |
| HDL_P | Obesity_pPS | 4255 | -0.018099384 | [-0.05 0.01] | 0.238512672 | 0.511098583 |
| HDL_P | VAT_Neg_pPS | 4255 | -0.045111482 | [-0.08 -0.02] | 0.003291495 | 0.018615834 |
| HDL_P | Bilirubin_pPS | 4255 | 0.005701442 | [-0.02 0.04] | 0.710432034 | 0.859991966 |
| HDL_P | Proinsulin_pPS | 4255 | 0.006739644 | [-0.02 0.04] | 0.660745092 | 0.824335 |
| HDL_P | Hyper_Insulin_pPS | 4255 | -0.013470909 | [-0.04 0.02] | 0.380348923 | 0.646405804 |
| HDL_P | Beta_Cell_1_pPS | 4255 | 0.012256937 | [-0.02 0.04] | 0.424759677 | 0.678435595 |
| HDL_P | Beta_Cell_2_pPS | 4255 | 0.001232578 | [-0.03 0.03] | 0.936026955 | 0.965799256 |
| HDL_P | Total_GRS | 4255 | -0.033516238 | [-0.06 -0. ] | 0.029023931 | 0.107669423 |
| HDL_C | Cholesterol_pPS | 4254 | 0.032489773 | [0. 0.06] | 0.034343563 | 0.118485294 |
| HDL_C | Lipodystrophy_1_pPS | 4254 | -0.073283652 | [-0.1 -0.04] | 1.77E-06 | 3.82E-05 |
| HDL_C | Lipodystrophy_2_pPS | 4254 | -0.043489176 | [-0.07 -0.01] | 0.00461158 | 0.023396987 |
| HDL_C | Liver-Lipid_pPS | 4254 | -0.004578847 | [-0.03 0.03] | 0.765598688 | 0.880438492 |
| HDL_C | ASAT_Pos_pPS | 4254 | -0.059480862 | [-0.09 -0.03] | 0.00010605 | 0.001247716 |
| HDL_C | ALP_Neg_pPS | 4254 | 0.017080917 | [-0.01 0.05] | 0.266033131 | 0.531285097 |
| HDL_C | SHBG-LpA_pPS | 4254 | 0.007468073 | [-0.02 0.04] | 0.626780141 | 0.813350082 |
| HDL_C | Obesity_pPS | 4254 | -0.030603514 | [-0.06 -0. ] | 0.046249866 | 0.153956868 |
| HDL_C | VAT_Neg_pPS | 4254 | -0.030581072 | [-0.06 -0. ] | 0.046410186 | 0.153956868 |
| HDL_C | Bilirubin_pPS | 4254 | -0.006419203 | [-0.04 0.02] | 0.675970619 | 0.82992834 |
| HDL_C | Proinsulin_pPS | 4254 | -0.007007829 | [-0.04 0.02] | 0.64817755 | 0.824335 |
| HDL_C | Hyper_Insulin_pPS | 4254 | -0.020002818 | [-0.05 0.01] | 0.192730943 | 0.449271456 |
| HDL_C | Beta_Cell_1_pPS | 4254 | 0.002638794 | [-0.03 0.03] | 0.863582199 | 0.936905216 |
| HDL_C | Beta_Cell_2_pPS | 4254 | 0.009938247 | [-0.02 0.04] | 0.517561024 | 0.734110863 |
| HDL_C | Total_GRS | 4254 | -0.047228797 | [-0.08 -0.02] | 0.002091991 | 0.014081824 |
| HDL_TG | Cholesterol_pPS | 4254 | -0.058609632 | [-0.09 -0.03] | 0.000133755 | 0.001442043 |
| HDL_TG | Lipodystrophy_1_pPS | 4254 | 0.041711756 | [0.01 0.07] | 0.006585768 | 0.031124518 |
| HDL_TG | Lipodystrophy_2_pPS | 4254 | -0.028421297 | [-0.06 0. ] | 0.064180268 | 0.201292658 |
| HDL_TG | Liver-Lipid_pPS | 4254 | -0.045823094 | [-0.08 -0.02] | 0.002834154 | 0.016739724 |
| HDL_TG | ASAT_Pos_pPS | 4254 | 0.016003192 | [-0.01 0.05] | 0.297384953 | 0.561581051 |
| HDL_TG | ALP_Neg_pPS | 4254 | 0.046433198 | [0.02 0.08] | 0.002486627 | 0.015597931 |
| HDL_TG | SHBG-LpA_pPS | 4254 | -0.007752846 | [-0.04 0.02] | 0.613694708 | 0.808109443 |
| HDL_TG | Obesity_pPS | 4254 | 0.016133659 | [-0.01 0.05] | 0.2934634 | 0.559363939 |
| HDL_TG | VAT_Neg_pPS | 4254 | 0.004993564 | [-0.03 0.04] | 0.745074996 | 0.877306736 |
| HDL_TG | Bilirubin_pPS | 4254 | -0.010511347 | [-0.04 0.02] | 0.493705037 | 0.721038307 |
| HDL_TG | Proinsulin_pPS | 4254 | 0.026373947 | [-0. 0.06] | 0.085879733 | 0.262199185 |
| HDL_TG | Hyper_Insulin_pPS | 4254 | 0.006487163 | [-0.02 0.04] | 0.672738209 | 0.828909578 |
| HDL_TG | Beta_Cell_1_pPS | 4254 | 0.011177225 | [-0.02 0.04] | 0.466743393 | 0.705151034 |
| HDL_TG | Beta_Cell_2_pPS | 4254 | 0.011028661 | [-0.02 0.04] | 0.472687075 | 0.705151034 |
| HDL_TG | Total_GRS | 4254 | 0.020612342 | [-0.01 0.05] | 0.179517569 | 0.437781659 |
| HDL_PL | Cholesterol_pPS | 4255 | 0.050790317 | [0.02 0.08] | 0.000934438 | 0.007497234 |
| HDL_PL | Lipodystrophy_1_pPS | 4255 | -0.070407888 | [-0.1 -0.04] | 4.42E-06 | 8.03E-05 |
| HDL_PL | Lipodystrophy_2_pPS | 4255 | -0.04021475 | [-0.07 -0.01] | 0.008797905 | 0.037940965 |
| HDL_PL | Liver-Lipid_pPS | 4255 | -0.018786684 | [-0.05 0.01] | 0.221148339 | 0.495429721 |
| HDL_PL | ASAT_Pos_pPS | 4255 | -0.051454918 | [-0.08 -0.02] | 0.000799638 | 0.006728659 |
| HDL_PL | ALP_Neg_pPS | 4255 | 0.018923845 | [-0.01 0.05] | 0.217794278 | 0.491104744 |
| HDL_PL | SHBG-LpA_pPS | 4255 | 0.003795331 | [-0.03 0.03] | 0.804792515 | 0.909217468 |
| HDL_PL | Obesity_pPS | 4255 | -0.025535258 | [-0.06 0. ] | 0.096291465 | 0.286384099 |
| HDL_PL | VAT_Neg_pPS | 4255 | -0.035161798 | [-0.07 -0.01] | 0.021997352 | 0.086239618 |
| HDL_PL | Bilirubin_pPS | 4255 | 0.000667415 | [-0.03 0.03] | 0.965333648 | 0.968349016 |
| HDL_PL | Proinsulin_pPS | 4255 | 0.001207229 | [-0.03 0.03] | 0.937339895 | 0.965799256 |
| HDL_PL | Hyper_Insulin_pPS | 4255 | -0.014236445 | [-0.04 0.02] | 0.353868505 | 0.624282144 |

**b) Partial Correlation**

| Metabolomics (selecte | PPS | Sample_Size | r | CI95% | p | fdr_p_adjusted |
|---|---|---|---|---|---|---|
| LDL_TG | ASAT_Pos_pPS | 4254 | -0.018690622 | [-0.05 0.01] | 0.224285151 | 0.487931594 |
| LDL_TG | ALP_Neg_pPS | 4254 | 0.051490134 | [0.02 0.08] | 0.000809065 | 0.006512974 |
| LDL_TG | SHBG-LpA_pPS | 4254 | 0.005478573 | [-0.02 0.04] | 0.721710935 | 0.86390677 |
| LDL_TG | Obesity_pPS | 4254 | 0.017982058 | [-0.01 0.05] | 0.242348932 | 0.510041543 |
| LDL_TG | VAT_Neg_pPS | 4254 | -0.001862465 | [-0.03 0.03] | 0.903626242 | 0.970866349 |
| LDL_TG | Bilirubin_pPS | 4254 | -0.00663657 | [-0.04 0.02] | 0.666135134 | 0.834612892 |
| LDL_TG | Proinsulin_pPS | 4254 | 0.000851713 | [-0.03 0.03] | 0.95584273 | 0.980221262 |
| LDL_TG | Hyper_Insulin_pPS | 4254 | -0.012642653 | [-0.04 0.02] | 0.411104706 | 0.685015893 |
| LDL_TG | Beta_Cell_1_pPS | 4254 | -0.00249246 | [-0.03 0.03] | 0.871275209 | 0.9542538 |
| LDL_TG | Beta_Cell_2_pPS | 4254 | 0.012606189 | [-0.02 0.04] | 0.412455427 | 0.685015893 |
| LDL_PL | Cholesterol_pPS | 4255 | -0.152839429 | [-0.18 -0.12] | 1.58E-23 | 2.36E-21 |
| LDL_PL | Lipodystrophy_1_pPS | 4255 | 0.046699278 | [0.02 0.08] | 0.002381473 | 0.016185619 |
| LDL_PL | Lipodystrophy_2_pPS | 4255 | -0.04354536 | [-0.07 -0.01] | 0.004616689 | 0.027529147 |
| LDL_PL | Liver-Lipid_pPS | 4255 | -0.002362398 | [-0.03 0.03] | 0.877923924 | 0.955038863 |
| LDL_PL | ASAT_Pos_pPS | 4255 | -0.017919516 | [-0.05 0.01] | 0.243935343 | 0.510046625 |
| LDL_PL | ALP_Neg_pPS | 4255 | 0.052575498 | [0.02 0.08] | 0.000624409 | 0.005584989 |
| LDL_PL | SHBG-LpA_pPS | 4255 | 0.015174244 | [-0.01 0.05] | 0.323802251 | 0.589063983 |
| LDL_PL | Obesity_pPS | 4255 | 0.011362792 | [-0.02 0.04] | 0.460014003 | 0.726100534 |
| LDL_PL | VAT_Neg_pPS | 4255 | 0.004344965 | [-0.03 0.03] | 0.77755433 | 0.910445433 |
| LDL_PL | Bilirubin_pPS | 4255 | -0.015554303 | [-0.05 0.01] | 0.311829817 | 0.585754653 |
| LDL_PL | Proinsulin_pPS | 4255 | -0.01890463 | [-0.05 0.01] | 0.21896899 | 0.487931594 |
| LDL_PL | Hyper_Insulin_pPS | 4255 | -0.02088461 | [-0.05 0.01] | 0.174448032 | 0.44874642 |
| LDL_PL | Beta_Cell_1_pPS | 4255 | 0.018931564 | [-0.01 0.05] | 0.218313154 | 0.487931594 |
| LDL_PL | Beta_Cell_2_pPS | 4255 | 0.019823625 | [-0.01 0.05] | 0.19738299 | 0.467333255 |
| HDL_D | Cholesterol_pPS | 4254 | 0.033434261 | [0. 0.06] | 0.029688578 | 0.117394442 |
| HDL_D | Lipodystrophy_1_pPS | 4254 | -0.055494544 | [-0.09 -0.03] | 0.000305527 | 0.003392405 |
| HDL_D | Lipodystrophy_2_pPS | 4254 | -0.034843302 | [-0.06 -0. ] | 0.023457223 | 0.098093843 |
| HDL_D | Liver-Lipid_pPS | 4254 | 0.021166494 | [-0.01 0.05] | 0.168753373 | 0.441777123 |
| HDL_D | ASAT_Pos_pPS | 4254 | -0.036172083 | [-0.07 -0.01] | 0.018654082 | 0.083425201 |
| HDL_D | ALP_Neg_pPS | 4254 | 0.020589834 | [-0.01 0.05] | 0.180662266 | 0.458057084 |
| HDL_D | SHBG-LpA_pPS | 4254 | 0.015186187 | [-0.01 0.05] | 0.323478769 | 0.589063983 |
| HDL_D | Obesity_pPS | 4254 | -0.033259232 | [-0.06 -0. ] | 0.030554094 | 0.117394442 |
| HDL_D | VAT_Neg_pPS | 4254 | -0.009564529 | [-0.04 0.02] | 0.534060909 | 0.7538823 |
| HDL_D | Bilirubin_pPS | 4254 | -0.004875055 | [-0.04 0.03] | 0.75129164 | 0.888369967 |
| HDL_D | Proinsulin_pPS | 4254 | -0.007242274 | [-0.04 0.02] | 0.637756751 | 0.821747013 |
| HDL_D | Hyper_Insulin_pPS | 4254 | -0.00015229 | [-0.03 0.03] | 0.992100573 | 0.995191229 |
| HDL_D | Beta_Cell_1_pPS | 4254 | -0.006878906 | [-0.04 0.02] | 0.654720865 | 0.830955435 |
| HDL_D | Beta_Cell_2_pPS | 4254 | 0.01458622 | [-0.02 0.04] | 0.342965016 | 0.596988936 |
| HDL_P | Cholesterol_pPS | 4255 | 0.038481267 | [0.01 0.07] | 0.012316318 | 0.056655065 |
| HDL_P | Lipodystrophy_1_pPS | 4255 | -0.05455332 | [-0.08 -0.02] | 0.000385732 | 0.003881426 |
| HDL_P | Lipodystrophy_2_pPS | 4255 | -0.028679157 | [-0.06 0. ] | 0.06217016 | 0.20487562 |
| HDL_P | Liver-Lipid_pPS | 4255 | -0.041798781 | [-0.07 -0.01] | 0.00654969 | 0.034573773 |
| HDL_P | ASAT_Pos_pPS | 4255 | -0.032265283 | [-0.06 -0. ] | 0.035868936 | 0.133534727 |
| HDL_P | ALP_Neg_pPS | 4255 | 0.024612391 | [-0.01 0.05] | 0.109483033 | 0.335747967 |
| HDL_P | SHBG-LpA_pPS | 4255 | 0.008347961 | [-0.02 0.04] | 0.587276168 | 0.79455011 |
| HDL_P | Obesity_pPS | 4255 | -0.018701616 | [-0.05 0.01] | 0.223957895 | 0.487931594 |
| HDL_P | VAT_Neg_pPS | 4255 | -0.040100145 | [-0.07 -0.01] | 0.009098531 | 0.045776983 |
| HDL_P | Bilirubin_pPS | 4255 | 0.007044181 | [-0.02 0.04] | 0.646943166 | 0.829943026 |
| HDL_P | Proinsulin_pPS | 4255 | 0.000851126 | [-0.03 0.03] | 0.955867939 | 0.980221262 |
| HDL_P | Hyper_Insulin_pPS | 4255 | 0.002885302 | [-0.03 0.03] | 0.851189854 | 0.941866437 |
| HDL_P | Beta_Cell_1_pPS | 4255 | 0.019378756 | [-0.01 0.05] | 0.207629646 | 0.47416132 |
| HDL_P | Beta_Cell_2_pPS | 4255 | -0.000276945 | [-0.03 0.03] | 0.985633489 | 0.994114756 |
| HDL_C | Cholesterol_pPS | 4254 | 0.036066221 | [0.01 0.07] | 0.019002418 | 0.083818887 |
| HDL_C | Lipodystrophy_1_pPS | 4254 | -0.061716783 | [-0.09 -0.03] | 5.92E-05 | 0.000828257 |
| HDL_C | Lipodystrophy_2_pPS | 4254 | -0.033371881 | [-0.06 -0. ] | 0.029994596 | 0.117394442 |
| HDL_C | Liver-Lipid_pPS | 4254 | -0.003817207 | [-0.03 0.03] | 0.804007958 | 0.931261016 |
| HDL_C | ASAT_Pos_pPS | 4254 | -0.042524213 | [-0.07 -0.01] | 0.005678064 | 0.032076082 |
| HDL_C | ALP_Neg_pPS | 4254 | 0.021246141 | [-0.01 0.05] | 0.167155946 | 0.441777123 |
| HDL_C | SHBG-LpA_pPS | 4254 | 0.015382301 | [-0.01 0.05] | 0.31726878 | 0.587129581 |
| HDL_C | Obesity_pPS | 4254 | -0.028640007 | [-0.06 0. ] | 0.062558997 | 0.20487562 |
| HDL_C | VAT_Neg_pPS | 4254 | -0.024481395 | [-0.05 0.01] | 0.111427029 | 0.338485882 |
| HDL_C | Bilirubin_pPS | 4254 | -0.003667157 | [-0.03 0.03] | 0.811564419 | 0.933298923 |
| HDL_C | Proinsulin_pPS | 4254 | -0.01048268 | [-0.04 0.02] | 0.495546381 | 0.739985734 |
| HDL_C | Hyper_Insulin_pPS | 4254 | -0.004059319 | [-0.03 0.03] | 0.791854136 | 0.920494699 |
| HDL_C | Beta_Cell_1_pPS | 4254 | 0.009048977 | [-0.02 0.04] | 0.556330983 | 0.775491673 |
| HDL_C | Beta_Cell_2_pPS | 4254 | 0.011146317 | [-0.02 0.04] | 0.468659598 | 0.729020706 |
| HDL_TG | Cholesterol_pPS | 4254 | -0.05837337 | [-0.09 -0.03] | 0.000145762 | 0.001738343 |
| HDL_TG | Lipodystrophy_1_pPS | 4254 | 0.040204625 | [0.01 0.07] | 0.008927406 | 0.045628961 |
| HDL_TG | Lipodystrophy_2_pPS | 4254 | -0.029044954 | [-0.06 0. ] | 0.058937997 | 0.197687864 |
| HDL_TG | Liver-Lipid_pPS | 4254 | -0.047354225 | [-0.08 -0.02] | 0.002067929 | 0.014797182 |
| HDL_TG | ASAT_Pos_pPS | 4254 | 0.010736316 | [-0.02 0.04] | 0.485174359 | 0.738793513 |
| HDL_TG | ALP_Neg_pPS | 4254 | 0.043566415 | [0.01 0.07] | 0.004601755 | 0.027529147 |
| HDL_TG | SHBG-LpA_pPS | 4254 | -0.011033209 | [-0.04 0.02] | 0.473184247 | 0.729020706 |
| HDL_TG | Obesity_pPS | 4254 | 0.016337256 | [-0.01 0.05] | 0.288153502 | 0.555601364 |
| HDL_TG | VAT_Neg_pPS | 4254 | 0.005099837 | [-0.03 0.04] | 0.740228767 | 0.879533812 |
| HDL_TG | Bilirubin_pPS | 4254 | -0.013319243 | [-0.04 0.02] | 0.386521499 | 0.658518109 |
| HDL_TG | Proinsulin_pPS | 4254 | 0.016453769 | [-0.01 0.05] | 0.284728611 | 0.555601364 |
| HDL_TG | Hyper_Insulin_pPS | 4254 | 0.003022159 | [-0.03 0.03] | 0.84423789 | 0.941866437 |
| HDL_TG | Beta_Cell_1_pPS | 4254 | 0.001601219 | [-0.03 0.03] | 0.917091744 | 0.970866349 |
| HDL_TG | Beta_Cell_2_pPS | 4254 | 0.008445906 | [-0.02 0.04] | 0.582942894 | 0.794494289 |
| HDL_PL | Cholesterol_pPS | 4255 | 0.054064633 | [0.02 0.08] | 0.000435118 | 0.004120823 |
| HDL_PL | Lipodystrophy_1_pPS | 4255 | -0.060742253 | [-0.09 -0.03] | 7.72E-05 | 0.001015255 |
| HDL_PL | Lipodystrophy_2_pPS | 4255 | -0.030932279 | [-0.06 -0. ] | 0.044254287 | 0.15873732 |
| HDL_PL | Liver-Lipid_pPS | 4255 | -0.018805205 | [-0.05 0.01] | 0.221402215 | 0.487931594 |
| HDL_PL | ASAT_Pos_pPS | 4255 | -0.035519464 | [-0.07 -0.01] | 0.020878355 | 0.089637737 |
| HDL_PL | ALP_Neg_pPS | 4255 | 0.021869907 | [-0.01 0.05] | 0.154987202 | 0.426545975 |
| HDL_PL | SHBG-LpA_pPS | 4255 | 0.0118936 | [-0.02 0.04] | 0.439321155 | 0.707307059 |
| HDL_PL | Obesity_pPS | 4255 | -0.026825537 | [-0.06 0. ] | 0.081073753 | 0.261057485 |
| HDL_PL | VAT_Neg_pPS | 4255 | -0.02858976 | [-0.06 0. ] | 0.062989709 | 0.20487562 |
| HDL_PL | Bilirubin_pPS | 4255 | 0.002991632 | [-0.03 0.03] | 0.84577338 | 0.941866437 |
| HDL_PL | Proinsulin_pPS | 4255 | -0.00151416 | [-0.03 0.03] | 0.921575309 | 0.970866349 |
| HDL_PL | Hyper_Insulin_pPS | 4255 | 0.001493838 | [-0.03 0.03] | 0.922624543 | 0.970866349 |
| HDL_PL | Beta_Cell_1_pPS | 4255 | 0.007791393 | [-0.02 0.04] | 0.612436876 | 0.813935509 |
| HDL_PL | Beta_Cell_2_pPS | 4255 | 0.004486306 | [-0.03 0.03] | 0.770517809 | 0.905499031 |

**Table S6: Associations between pPSs and selected metabolites in SCAPIS**

**a) Spearman Correlation**

| Metabolomics (select | PPS | Sample_Size | r | CI95% | p | fdr_p_adjusted |
|---|---|---|---|---|---|---|
| HDL_PL | Beta_Cell_1_pPS | 4255 | 0.001665786 | [-0.03 0.03] | 0.913619218 | 0.960971434 |
| HDL_PL | Beta_Cell_2_pPS | 4255 | 0.003936897 | [-0.03 0.03] | 0.797666475 | 0.905246493 |
| HDL_PL | Total_GRS | 4255 | -0.040803563 | [-0.07 -0.01] | 0.007855945 | 0.034967146 |

**b) Partial Correlation**

| Metabolomics (selecte | PPS | Sample_Size | r | CI95% | p | fdr_p_adjusted |
|---|---|---|---|---|---|---|

**Table S7: Associations between pPSs and plasma proteins in SCAPIS**

**a) Spearman Correlation**

| Proteomics | PPS | Sample_Size | r | CI95% | p | fdr_p_adjusted |
|---|---|---|---|---|---|---|
| ANG_1 | Cholesterol_pPS | 4159 | -0.012451521 | [-0.04 0.02] | 0.42276558 | 0.824034606 |
| ANG_1 | Lipodystrophy_1_pPS | 4159 | -0.001520743 | [-0.03 0.03] | 0.922010007 | 0.990056084 |
| ANG_1 | Lipodystrophy_2_pPS | 4159 | -0.004492449 | [-0.03 0.03] | 0.772416531 | 0.95301975 |
| ANG_1 | Liver-Lipid_pPS | 4159 | -0.011847342 | [-0.04 0.02] | 0.445622968 | 0.831350198 |
| ANG_1 | ASAT_Pos_pPS | 4159 | -0.00125852 | [-0.03 0.03] | 0.935425439 | 0.99138381 |
| ANG_1 | ALP_Neg_pPS | 4159 | -0.00366241 | [-0.03 0.03] | 0.813604421 | 0.962515303 |
| ANG_1 | SHBG-LpA_pPS | 4159 | -0.00564251 | [-0.04 0.02] | 0.716414232 | 0.941449667 |
| ANG_1 | Obesity_pPS | 4159 | -0.015430168 | [-0.05 0.02] | 0.320505663 | 0.770109076 |
| ANG_1 | VAT_Neg_pPS | 4159 | -0.00796932 | [-0.04 0.02] | 0.607912595 | 0.909483337 |
| ANG_1 | Bilirubin_pPS | 4159 | 0.009617642 | [-0.02 0.04] | 0.535799861 | 0.880766894 |
| ANG_1 | Proinsulin_pPS | 4159 | -0.00219324 | [-0.03 0.03] | 0.887715006 | 0.979291846 |
| ANG_1 | Hyper_Insulin_pPS | 4159 | -0.027750819 | [-0.06 0. ] | 0.073956793 | 0.504001849 |
| ANG_1 | Beta_Cell_1_pPS | 4159 | -0.021776237 | [-0.05 0.01] | 0.160893665 | 0.664061293 |
| ANG_1 | Beta_Cell_2_pPS | 4159 | -0.015906165 | [-0.05 0.01] | 0.305802884 | 0.76365449 |
| ANG_1 | Total_GRS | 4159 | -0.02668034 | [-0.06 0. ] | 0.0858099 | 0.539488209 |
| CTSD | Cholesterol_pPS | 3924 | 0.016172283 | [-0.02 0.05] | 0.311894788 | 0.76365449 |
| CTSD | Lipodystrophy_1_pPS | 3924 | 0.063473508 | [0.03 0.09] | 7.10E-05 | 0.008170459 |
| CTSD | Lipodystrophy_2_pPS | 3924 | 0.052266162 | [0.02 0.08] | 0.001074605 | 0.051136358 |
| CTSD | Liver-Lipid_pPS | 3924 | -0.02327097 | [-0.05 0.01] | 0.145603501 | 0.641415816 |
| CTSD | ASAT_Pos_pPS | 3924 | 0.005771354 | [-0.03 0.04] | 0.718202103 | 0.941449667 |
| CTSD | ALP_Neg_pPS | 3924 | -0.021307797 | [-0.05 0.01] | 0.182714844 | 0.685180665 |
| CTSD | SHBG-LpA_pPS | 3924 | 0.006514904 | [-0.02 0.04] | 0.683747136 | 0.926969309 |
| CTSD | Obesity_pPS | 3924 | 0.030334312 | [-0. 0.06] | 0.057812156 | 0.45329986 |
| CTSD | VAT_Neg_pPS | 3924 | 0.013331217 | [-0.02 0.04] | 0.404515089 | 0.81230319 |
| CTSD | Bilirubin_pPS | 3924 | 0.000381476 | [-0.03 0.03] | 0.980970504 | 0.996862516 |
| CTSD | Proinsulin_pPS | 3924 | 0.00643094 | [-0.02 0.04] | 0.687606704 | 0.926969309 |
| CTSD | Hyper_Insulin_pPS | 3924 | 0.003680057 | [-0.03 0.04] | 0.818014044 | 0.963461584 |
| CTSD | Beta_Cell_1_pPS | 3924 | 0.017047085 | [-0.01 0.05] | 0.286440189 | 0.751497075 |
| CTSD | Beta_Cell_2_pPS | 3924 | 0.005490291 | [-0.03 0.04] | 0.731381449 | 0.942134248 |
| CTSD | Total_GRS | 3924 | 0.047202245 | [0.02 0.08] | 0.003146884 | 0.105919518 |
| DECR1 | Cholesterol_pPS | 4159 | 0.017937589 | [-0.01 0.05] | 0.248141505 | 0.737212651 |
| DECR1 | Lipodystrophy_1_pPS | 4159 | 0.005653366 | [-0.02 0.04] | 0.715892288 | 0.94141015 |
| DECR1 | Lipodystrophy_2_pPS | 4159 | 0.018705487 | [-0.01 0.05] | 0.228464637 | 0.726631332 |
| DECR1 | Liver-Lipid_pPS | 4159 | -0.008446952 | [-0.04 0.02] | 0.586575126 | 0.903925569 |
| DECR1 | ASAT_Pos_pPS | 4159 | 0.014494183 | [-0.02 0.04] | 0.350741231 | 0.793480163 |
| DECR1 | ALP_Neg_pPS | 4159 | -0.030017196 | [-0.06 0. ] | 0.053251436 | 0.442381248 |
| DECR1 | SHBG-LpA_pPS | 4159 | -0.000155305 | [-0.03 0.03] | 0.99202271 | 0.999402232 |
| DECR1 | Obesity_pPS | 4159 | -0.006226904 | [-0.04 0.02] | 0.688509813 | 0.926969309 |
| DECR1 | VAT_Neg_pPS | 4159 | 0.00531129 | [-0.03 0.04] | 0.732402421 | 0.942134248 |
| DECR1 | Bilirubin_pPS | 4159 | 0.006292599 | [-0.02 0.04] | 0.685398382 | 0.926969309 |
| DECR1 | Proinsulin_pPS | 4159 | 0.00275705 | [-0.03 0.03] | 0.859121059 | 0.977006629 |
| DECR1 | Hyper_Insulin_pPS | 4159 | 0.008536368 | [-0.02 0.04] | 0.582619483 | 0.903823705 |
| DECR1 | Beta_Cell_1_pPS | 4159 | -0.012350356 | [-0.04 0.02] | 0.426544307 | 0.82738773 |
| DECR1 | Beta_Cell_2_pPS | 4159 | -0.011806126 | [-0.04 0.02] | 0.447207498 | 0.831350198 |
| DECR1 | Total_GRS | 4159 | 0.001911145 | [-0.03 0.03] | 0.90207905 | 0.987003157 |
| FABP2 | Cholesterol_pPS | 4159 | 0.00073355 | [-0.03 0.03] | 0.962334415 | 0.99612031 |
| FABP2 | Lipodystrophy_1_pPS | 4159 | 0.008376202 | [-0.02 0.04] | 0.589713761 | 0.904476019 |
| FABP2 | Lipodystrophy_2_pPS | 4159 | -0.005525912 | [-0.04 0.02] | 0.722028744 | 0.942134248 |
| FABP2 | Liver-Lipid_pPS | 4159 | 0.015752343 | [-0.01 0.05] | 0.310504532 | 0.76365449 |
| FABP2 | ASAT_Pos_pPS | 4159 | -0.005414457 | [-0.04 0.03] | 0.727409656 | 0.942134248 |
| FABP2 | ALP_Neg_pPS | 4159 | 0.014079128 | [-0.02 0.04] | 0.364709318 | 0.798887077 |
| FABP2 | SHBG-LpA_pPS | 4159 | 0.011137546 | [-0.02 0.04] | 0.473353812 | 0.846453261 |
| FABP2 | Obesity_pPS | 4159 | 0.02891265 | [-0. 0.06] | 0.062642322 | 0.469817417 |
| FABP2 | VAT_Neg_pPS | 4159 | -0.020673288 | [-0.05 0.01] | 0.183174905 | 0.685789735 |
| FABP2 | Bilirubin_pPS | 4159 | 0.005483808 | [-0.02 0.04] | 0.724059879 | 0.942134248 |
| FABP2 | Proinsulin_pPS | 4159 | -0.012302484 | [-0.04 0.02] | 0.42833926 | 0.827882604 |
| FABP2 | Hyper_Insulin_pPS | 4159 | -0.003595224 | [-0.03 0.03] | 0.816962618 | 0.963461584 |
| FABP2 | Beta_Cell_1_pPS | 4159 | -0.008218468 | [-0.04 0.02] | 0.596739024 | 0.907623171 |
| FABP2 | Beta_Cell_2_pPS | 4159 | -0.012567453 | [-0.04 0.02] | 0.418459451 | 0.819693459 |
| FABP2 | Total_GRS | 4159 | 0.016001339 | [-0.01 0.05] | 0.302917634 | 0.76319014 |
| FABP4 | Cholesterol_pPS | 4225 | -0.012222464 | [-0.04 0.02] | 0.427704798 | 0.82738773 |
| FABP4 | Lipodystrophy_1_pPS | 4225 | 0.004398998 | [-0.03 0.03] | 0.775303122 | 0.953637204 |
| FABP4 | Lipodystrophy_2_pPS | 4225 | -0.006339195 | [-0.04 0.02] | 0.680820202 | 0.92591406 |
| FABP4 | Liver-Lipid_pPS | 4225 | 0.019887535 | [-0.01 0.05] | 0.196843362 | 0.697416791 |
| FABP4 | ASAT_Pos_pPS | 4225 | 0.024117753 | [-0.01 0.05] | 0.117538545 | 0.613244581 |
| FABP4 | ALP_Neg_pPS | 4225 | 0.028312241 | [-0. 0.06] | 0.066135617 | 0.476590869 |
| FABP4 | SHBG-LpA_pPS | 4225 | 0.007815784 | [-0.02 0.04] | 0.612041705 | 0.909483337 |
| FABP4 | Obesity_pPS | 4225 | 0.056902129 | [0.03 0.09] | 0.000219644 | 0.017829955 |
| FABP4 | VAT_Neg_pPS | 4225 | -0.024180248 | [-0.05 0.01] | 0.116590585 | 0.611872879 |
| FABP4 | Bilirubin_pPS | 4225 | -0.013885171 | [-0.04 0.02] | 0.367572656 | 0.799230784 |
| FABP4 | Proinsulin_pPS | 4225 | 0.01051919 | [-0.02 0.04] | 0.494864734 | 0.86226431 |
| FABP4 | Hyper_Insulin_pPS | 4225 | 0.004971432 | [-0.03 0.04] | 0.747005693 | 0.947292085 |
| FABP4 | Beta_Cell_1_pPS | 4225 | -0.013527437 | [-0.04 0.02] | 0.380044984 | 0.805487278 |
| FABP4 | Beta_Cell_2_pPS | 4225 | -0.025137158 | [-0.06 0.01] | 0.102812936 | 0.586842904 |
| FABP4 | Total_GRS | 4225 | 0.017387604 | [-0.01 0.05] | 0.259176978 | 0.743022111 |
| GT | Cholesterol_pPS | 4159 | -0.011833639 | [-0.04 0.02] | 0.446149394 | 0.831350198 |
| GT | Lipodystrophy_1_pPS | 4159 | 0.002532352 | [-0.03 0.03] | 0.870496806 | 0.977006629 |
| GT | Lipodystrophy_2_pPS | 4159 | 0.016405011 | [-0.01 0.05] | 0.29088168 | 0.753421877 |
| GT | Liver-Lipid_pPS | 4159 | 0.030608784 | [0. 0.06] | 0.048725283 | 0.438114317 |
| GT | ASAT_Pos_pPS | 4159 | 0.013856319 | [-0.02 0.04] | 0.372348953 | 0.799802236 |
| GT | ALP_Neg_pPS | 4159 | 0.052612546 | [0.02 0.08] | 0.000700389 | 0.039450459 |
| GT | SHBG-LpA_pPS | 4159 | 0.003606778 | [-0.03 0.03] | 0.816384908 | 0.963461584 |
| GT | Obesity_pPS | 4159 | 0.018947265 | [-0.01 0.05] | 0.222505604 | 0.722453819 |
| GT | VAT_Neg_pPS | 4159 | 0.019280806 | [-0.01 0.05] | 0.214468691 | 0.715184753 |
| GT | Bilirubin_pPS | 4159 | -0.004360194 | [-0.03 0.03] | 0.778939755 | 0.953637204 |
| GT | Proinsulin_pPS | 4159 | 0.014668489 | [-0.02 0.05] | 0.344977665 | 0.790391668 |
| GT | Hyper_Insulin_pPS | 4159 | -0.010295588 | [-0.04 0.02] | 0.507442768 | 0.868283967 |
| GT | Beta_Cell_1_pPS | 4159 | 0.001226214 | [-0.03 0.03] | 0.937079584 | 0.991487772 |
| GT | Beta_Cell_2_pPS | 4159 | 0.000937109 | [-0.03 0.03] | 0.951893572 | 0.995161462 |
| GT | Total_GRS | 4159 | 0.023136357 | [-0.01 0.05] | 0.136311821 | 0.629136669 |
| LPL | Cholesterol_pPS | 4159 | -0.016190452 | [-0.05 0.01] | 0.297238371 | 0.759609171 |
| LPL | Lipodystrophy_1_pPS | 4159 | -0.026831297 | [-0.06 0. ] | 0.084050871 | 0.534941474 |
| LPL | Lipodystrophy_2_pPS | 4159 | 0.021812096 | [-0.01 0.05] | 0.160205246 | 0.663913634 |
| LPL | Liver-Lipid_pPS | 4159 | 0.037959401 | [0.01 0.07] | 0.014500347 | 0.252355767 |
| LPL | ASAT_Pos_pPS | 4159 | -0.021460211 | [-0.05 0.01] | 0.167057661 | 0.666299341 |
| LPL | ALP_Neg_pPS | 4159 | -0.003934785 | [-0.03 0.03] | 0.800025777 | 0.958074488 |
| LPL | SHBG-LpA_pPS | 4159 | 0.014131846 | [-0.02 0.04] | 0.362916143 | 0.7979754 |
| LPL | Obesity_pPS | 4159 | 0.000579337 | [-0.03 0.03] | 0.970248652 | 0.996862516 |
| LPL | VAT_Neg_pPS | 4159 | -0.028570925 | [-0.06 0. ] | 0.065811037 | 0.476590869 |
| LPL | Bilirubin_pPS | 4159 | -0.004197754 | [-0.03 0.03] | 0.786973064 | 0.957247257 |
| LPL | Proinsulin_pPS | 4159 | 0.010472217 | [-0.02 0.04] | 0.500186283 | 0.863780476 |
| LPL | Hyper_Insulin_pPS | 4159 | -0.038419507 | [-0.07 -0.01] | 0.013350635 | 0.242414912 |
| LPL | Beta_Cell_1_pPS | 4159 | -0.010533137 | [-0.04 0.02] | 0.497696313 | 0.863780476 |
| LPL | Beta_Cell_2_pPS | 4159 | 0.002961556 | [-0.03 0.03] | 0.848792891 | 0.97528582 |
| LPL | Total_GRS | 4159 | -0.009998689 | [-0.04 0.02] | 0.519763766 | 0.872596104 |
| MMP_9 | Cholesterol_pPS | 4225 | -0.008491474 | [-0.04 0.02] | 0.581628344 | 0.903823705 |
| MMP_9 | Lipodystrophy_1_pPS | 4225 | 0.001678222 | [-0.03 0.03] | 0.913284038 | 0.990056084 |

**b) Partial Correlation**

| Proteomics | PPS | Sample_Size | r | CI95% | p | fdr_p_adjusted |
|---|---|---|---|---|---|---|
| ANG_1 | Cholesterol_pPS | 4159 | -0.01073768 | [-0.04 0.02] | 0.490066508 | 0.895803896 |
| ANG_1 | Lipodystrophy_1_pPS | 4159 | 0.002950468 | [-0.03 0.03] | 0.849585852 | 0.974254359 |
| ANG_1 | Lipodystrophy_2_pPS | 4159 | -0.002456986 | [-0.03 0.03] | 0.874514079 | 0.97781424 |
| ANG_1 | Liver-Lipid_pPS | 4159 | -0.008945389 | [-0.04 0.02] | 0.565296705 | 0.90516325 |
| ANG_1 | ASAT_Pos_pPS | 4159 | 0.001420177 | [-0.03 0.03] | 0.927266626 | 0.991547874 |
| ANG_1 | ALP_Neg_pPS | 4159 | -0.003895498 | [-0.03 0.03] | 0.802285073 | 0.967638637 |
| ANG_1 | SHBG-LpA_pPS | 4159 | -0.005524756 | [-0.04 0.02] | 0.722502645 | 0.946314812 |
| ANG_1 | Obesity_pPS | 4159 | -0.013267152 | [-0.04 0.02] | 0.393766415 | 0.85037326 |
| ANG_1 | VAT_Neg_pPS | 4159 | -0.002430971 | [-0.03 0.03] | 0.875831919 | 0.97781424 |
| ANG_1 | Bilirubin_pPS | 4159 | 0.01011587 | [-0.02 0.04] | 0.515542474 | 0.895803896 |
| ANG_1 | Proinsulin_pPS | 4159 | 0.008391286 | [-0.02 0.04] | 0.589628762 | 0.913889104 |
| ANG_1 | Hyper_Insulin_pPS | 4159 | -0.025419997 | [-0.06 0.01] | 0.102221334 | 0.659955277 |
| ANG_1 | Beta_Cell_1_pPS | 4159 | -0.019707152 | [-0.05 0.01] | 0.205214441 | 0.765145476 |
| ANG_1 | Beta_Cell_2_pPS | 4159 | -0.010107041 | [-0.04 0.02] | 0.515909102 | 0.895803896 |
| CTSD | Cholesterol_pPS | 3924 | 0.011856744 | [-0.02 0.04] | 0.459210723 | 0.874172623 |
| CTSD | Lipodystrophy_1_pPS | 3924 | 0.060506474 | [0.03 0.09] | 0.000156579 | 0.015513377 |
| CTSD | Lipodystrophy_2_pPS | 3924 | 0.0495868 | [0.02 0.08] | 0.001953514 | 0.099117562 |
| CTSD | Liver-Lipid_pPS | 3924 | -0.025056566 | [-0.06 0.01] | 0.117741192 | 0.681046037 |
| CTSD | ASAT_Pos_pPS | 3924 | -0.010535486 | [-0.04 0.02] | 0.510754163 | 0.895803896 |
| CTSD | ALP_Neg_pPS | 3924 | -0.021767633 | [-0.05 0.01] | 0.174165869 | 0.730702407 |
| CTSD | SHBG-LpA_pPS | 3924 | 0.001307312 | [-0.03 0.03] | 0.934960818 | 0.994163908 |
| CTSD | Obesity_pPS | 3924 | 0.028206214 | [-0. 0.06] | 0.078232095 | 0.602398521 |
| CTSD | VAT_Neg_pPS | 3924 | 0.005089734 | [-0.03 0.04] | 0.750702511 | 0.954135344 |
| CTSD | Bilirubin_pPS | 3924 | -0.002431561 | [-0.03 0.03] | 0.879358108 | 0.97781424 |
| CTSD | Proinsulin_pPS | 3924 | 0.001412637 | [-0.03 0.03] | 0.929733935 | 0.991797911 |
| CTSD | Hyper_Insulin_pPS | 3924 | -0.007618922 | [-0.04 0.02] | 0.634363015 | 0.931124222 |
| CTSD | Beta_Cell_1_pPS | 3924 | 0.014945622 | [-0.02 0.05] | 0.350826698 | 0.847284666 |
| CTSD | Beta_Cell_2_pPS | 3924 | 0.000539266 | [-0.03 0.03] | 0.973146647 | 0.997444743 |
| DECR1 | Cholesterol_pPS | 4159 | 0.018384458 | [-0.01 0.05] | 0.237287843 | 0.78980606 |
| DECR1 | Lipodystrophy_1_pPS | 4159 | 0.00243997 | [-0.03 0.03] | 0.875376017 | 0.97781424 |
| DECR1 | Lipodystrophy_2_pPS | 4159 | 0.015373494 | [-0.02 0.05] | 0.323046611 | 0.841027489 |
| DECR1 | Liver-Lipid_pPS | 4159 | -0.008291604 | [-0.04 0.02] | 0.594056761 | 0.917990532 |
| DECR1 | ASAT_Pos_pPS | 4159 | 0.012342935 | [-0.02 0.04] | 0.427548359 | 0.864060889 |
| DECR1 | ALP_Neg_pPS | 4159 | -0.031701563 | [-0.06 -0. ] | 0.041531383 | 0.481247748 |
| DECR1 | SHBG-LpA_pPS | 4159 | -0.002043875 | [-0.03 0.03] | 0.895479636 | 0.98328949 |
| DECR1 | Obesity_pPS | 4159 | -0.00963598 | [-0.04 0.02] | 0.535663525 | 0.898959051 |
| DECR1 | VAT_Neg_pPS | 4159 | 0.005427057 | [-0.03 0.04] | 0.727212317 | 0.946981479 |
| DECR1 | Bilirubin_pPS | 4159 | 0.00622333 | [-0.02 0.04] | 0.689142301 | 0.944989936 |
| DECR1 | Proinsulin_pPS | 4159 | 0.013838528 | [-0.02 0.04] | 0.373713924 | 0.847435798 |
| DECR1 | Hyper_Insulin_pPS | 4159 | 0.007073028 | [-0.02 0.04] | 0.649370462 | 0.935848959 |
| DECR1 | Beta_Cell_1_pPS | 4159 | -0.01294004 | [-0.04 0.02] | 0.405534 | 0.852958996 |
| DECR1 | Beta_Cell_2_pPS | 4159 | -0.010393549 | [-0.04 0.02] | 0.504081544 | 0.895803896 |
| FABP2 | Cholesterol_pPS | 4159 | -0.0028806 | [-0.03 0.03] | 0.853106701 | 0.974979087 |
| FABP2 | Lipodystrophy_1_pPS | 4159 | 0.008326846 | [-0.02 0.04] | 0.592489559 | 0.916668531 |
| FABP2 | Lipodystrophy_2_pPS | 4159 | -0.00293821 | [-0.03 0.03] | 0.850203338 | 0.974254359 |
| FABP2 | Liver-Lipid_pPS | 4159 | 0.016279869 | [-0.01 0.05] | 0.295336561 | 0.817900851 |
| FABP2 | ASAT_Pos_pPS | 4159 | -0.007392334 | [-0.04 0.02] | 0.634671736 | 0.931124222 |
| FABP2 | ALP_Neg_pPS | 4159 | 0.016130374 | [-0.01 0.05] | 0.299793931 | 0.821481367 |
| FABP2 | SHBG-LpA_pPS | 4159 | 0.013486001 | [-0.02 0.04] | 0.386010122 | 0.85037326 |
| FABP2 | Obesity_pPS | 4159 | 0.030831084 | [0. 0.06] | 0.047457649 | 0.520248282 |
| FABP2 | VAT_Neg_pPS | 4159 | -0.019722802 | [-0.05 0.01] | 0.204854797 | 0.765145476 |
| FABP2 | Bilirubin_pPS | 4159 | 0.006265204 | [-0.02 0.04] | 0.687160931 | 0.944571269 |
| FABP2 | Proinsulin_pPS | 4159 | -0.016400083 | [-0.05 0.01] | 0.291784562 | 0.81655641 |
| FABP2 | Hyper_Insulin_pPS | 4159 | -0.004875373 | [-0.04 0.03] | 0.753995776 | 0.95462436 |
| FABP2 | Beta_Cell_1_pPS | 4159 | 0.000234738 | [-0.03 0.03] | 0.987961809 | 0.998803911 |
| FABP2 | Beta_Cell_2_pPS | 4159 | -0.00884495 | [-0.04 0.02] | 0.569671269 | 0.905732577 |
| FABP4 | Cholesterol_pPS | 4225 | -0.018260359 | [-0.05 0.01] | 0.236748556 | 0.78980606 |
| FABP4 | Lipodystrophy_1_pPS | 4225 | -0.001641037 | [-0.03 0.03] | 0.915328674 | 0.989461462 |
| FABP4 | Lipodystrophy_2_pPS | 4225 | -0.005633809 | [-0.04 0.02] | 0.715106333 | 0.945970482 |
| FABP4 | Liver-Lipid_pPS | 4225 | 0.019337047 | [-0.01 0.05] | 0.21023192 | 0.767305935 |
| FABP4 | ASAT_Pos_pPS | 4225 | 0.022528973 | [-0.01 0.05] | 0.144346423 | 0.697681852 |
| FABP4 | ALP_Neg_pPS | 4225 | 0.025785342 | [-0. 0.06] | 0.094749038 | 0.64003016 |
| FABP4 | SHBG-LpA_pPS | 4225 | 0.009901826 | [-0.02 0.04] | 0.521174239 | 0.895803896 |
| FABP4 | Obesity_pPS | 4225 | 0.056316744 | [0.03 0.09] | 0.000260659 | 0.023153666 |
| FABP4 | VAT_Neg_pPS | 4225 | -0.022335465 | [-0.05 0.01] | 0.147826004 | 0.703126624 |
| FABP4 | Bilirubin_pPS | 4225 | -0.013645407 | [-0.04 0.02] | 0.376641488 | 0.849453763 |
| FABP4 | Proinsulin_pPS | 4225 | 0.005620644 | [-0.02 0.04] | 0.715743157 | 0.945970482 |
| FABP4 | Hyper_Insulin_pPS | 4225 | 0.001514218 | [-0.03 0.03] | 0.921850188 | 0.990503296 |
| FABP4 | Beta_Cell_1_pPS | 4225 | -0.008088985 | [-0.04 0.02] | 0.600223835 | 0.918301271 |
| FABP4 | Beta_Cell_2_pPS | 4225 | -0.022864668 | [-0.05 0.01] | 0.13845942 | 0.696623956 |
| GT | Cholesterol_pPS | 4159 | -0.014012176 | [-0.04 0.02] | 0.367747156 | 0.847284666 |
| GT | Lipodystrophy_1_pPS | 4159 | -0.001080501 | [-0.03 0.03] | 0.944630571 | 0.996180826 |
| GT | Lipodystrophy_2_pPS | 4159 | 0.013460574 | [-0.02 0.04] | 0.386906466 | 0.85037326 |
| GT | Liver-Lipid_pPS | 4159 | 0.028894247 | [-0. 0.06] | 0.063223564 | 0.558815902 |
| GT | ASAT_Pos_pPS | 4159 | 0.01272668 | [-0.02 0.04] | 0.413321393 | 0.854426619 |
| GT | ALP_Neg_pPS | 4159 | 0.05040951 | [0.02 0.08] | 0.001186022 | 0.07228782 |
| GT | SHBG-LpA_pPS | 4159 | 0.003694771 | [-0.03 0.03] | 0.812277687 | 0.967638637 |
| GT | Obesity_pPS | 4159 | 0.018971575 | [-0.01 0.05] | 0.222638991 | 0.780100778 |
| GT | VAT_Neg_pPS | 4159 | 0.017809817 | [-0.01 0.05] | 0.252272179 | 0.799790128 |
| GT | Bilirubin_pPS | 4159 | -0.004670869 | [-0.04 0.03] | 0.764001798 | 0.959095176 |
| GT | Proinsulin_pPS | 4159 | 0.003335286 | [-0.03 0.03] | 0.830249255 | 0.971458352 |
| GT | Hyper_Insulin_pPS | 4159 | -0.016937764 | [-0.05 0.01] | 0.276249845 | 0.808820203 |
| GT | Beta_Cell_1_pPS | 4159 | -0.002849414 | [-0.03 0.03] | 0.854679194 | 0.975477892 |
| GT | Beta_Cell_2_pPS | 4159 | -0.001772472 | [-0.03 0.03] | 0.909294237 | 0.988841471 |
| LPL | Cholesterol_pPS | 4159 | -0.019346445 | [-0.05 0.01] | 0.213631025 | 0.767305935 |
| LPL | Lipodystrophy_1_pPS | 4159 | -0.020690438 | [-0.05 0.01] | 0.183499702 | 0.738879686 |
| LPL | Lipodystrophy_2_pPS | 4159 | 0.029450571 | [-0. 0.06] | 0.058305028 | 0.558815902 |
| LPL | Liver-Lipid_pPS | 4159 | 0.04008841 | [0.01 0.07] | 0.009943477 | 0.248683454 |
| LPL | ASAT_Pos_pPS | 4159 | -0.017333542 | [-0.05 0.01] | 0.265181656 | 0.80722329 |
| LPL | ALP_Neg_pPS | 4159 | -0.005881434 | [-0.04 0.02] | 0.705398849 | 0.945970482 |
| LPL | SHBG-LpA_pPS | 4159 | 0.01664663 | [-0.01 0.05] | 0.284589894 | 0.812753402 |
| LPL | Obesity_pPS | 4159 | 0.004176846 | [-0.03 0.03] | 0.788333522 | 0.967638637 |
| LPL | VAT_Neg_pPS | 4159 | -0.029325071 | [-0.06 0. ] | 0.059385897 | 0.558815902 |
| LPL | Bilirubin_pPS | 4159 | -0.002392553 | [-0.03 0.03] | 0.877778696 | 0.97781424 |
| LPL | Proinsulin_pPS | 4159 | 0.012304952 | [-0.02 0.04] | 0.428971872 | 0.864060889 |
| LPL | Hyper_Insulin_pPS | 4159 | -0.03525226 | [-0.07 -0. ] | 0.023414365 | 0.372317304 |
| LPL | Beta_Cell_1_pPS | 4159 | -0.010659513 | [-0.04 0.02] | 0.493231449 | 0.895803896 |
| LPL | Beta_Cell_2_pPS | 4159 | 0.004365798 | [-0.03 0.03] | 0.779001394 | 0.964905159 |
| MMP_9 | Cholesterol_pPS | 4225 | -0.010829786 | [-0.04 0.02] | 0.482889955 | 0.89490973 |
| MMP_9 | Lipodystrophy_1_pPS | 4225 | 0.005327541 | [-0.02 0.04] | 0.729971231 | 0.947307754 |
| MMP_9 | Lipodystrophy_2_pPS | 4225 | 0.008528909 | [-0.02 0.04] | 0.580549685 | 0.909583681 |
| MMP_9 | Liver-Lipid_pPS | 4225 | -0.005615696 | [-0.04 0.02] | 0.71598252 | 0.945970482 |
| MMP_9 | ASAT_Pos_pPS | 4225 | -0.012718901 | [-0.04 0.02] | 0.409901501 | 0.854336605 |
| MMP_9 | ALP_Neg_pPS | 4225 | 0.023913164 | [-0.01 0.05] | 0.121259069 | 0.682614177 |
| MMP_9 | SHBG-LpA_pPS | 4225 | -0.002210537 | [-0.03 0.03] | 0.88611905 | 0.979597623 |
| MMP_9 | Obesity_pPS | 4225 | 0.005112471 | [-0.03 0.04] | 0.7404711 | 0.951867558 |
| MMP_9 | VAT_Neg_pPS | 4225 | -0.023801967 | [-0.05 0.01] | 0.122999955 | 0.684336682 |

Table S7: Associations between pPSs and plasma proteins in SCAPIS

a) Spearman Correlation

| Proteomics | PPS | Sample_Size | r | CI95% | p | fdr_p_adjusted |
|---|---|---|---|---|---|---|
| MMP_9 | Lipodystrophy_2_pPS | 4225 | 0.004326313 | [-0.03 0.03] | 0.77891849 | 0.953637204 |
| MMP_9 | Liver-Lipid_pPS | 4225 | -0.006245515 | [-0.04 0.02] | 0.68528255 | 0.926969309 |
| MMP_9 | ASAT_Pos_pPS | 4225 | -0.012251458 | [-0.04 0.02] | 0.426609474 | 0.82738773 |
| MMP_9 | ALP_Neg_pPS | 4225 | 0.030296166 | [0. 0.06] | 0.049261656 | 0.438587647 |
| MMP_9 | SHBG-LpA_pPS | 4225 | -0.002913837 | [-0.03 0.03] | 0.850034396 | 0.97528582 |
| MMP_9 | Obesity_pPS | 4225 | 0.007616738 | [-0.02 0.04] | 0.621133288 | 0.910907479 |
| MMP_9 | VAT_Neg_pPS | 4225 | -0.028229369 | [-0.06 0. ] | 0.066933136 | 0.477352596 |
| MMP_9 | Bilirubin_pPS | 4225 | 0.015154205 | [-0.02 0.05] | 0.325416476 | 0.775125841 |
| MMP_9 | Proinsulin_pPS | 4225 | 0.030961944 | [0. 0.06] | 0.044478657 | 0.424281318 |
| MMP_9 | Hyper_Insulin_pPS | 4225 | -0.012810427 | [-0.04 0.02] | 0.405814459 | 0.812806898 |
| MMP_9 | Beta_Cell_1_pPS | 4225 | -0.028431172 | [-0.06 0. ] | 0.065004777 | 0.475856102 |
| MMP_9 | Beta_Cell_2_pPS | 4225 | -0.025579015 | [-0.06 0. ] | 0.096903971 | 0.571484959 |
| MMP_9 | Total_GRS | 4225 | -0.011240262 | [-0.04 0.02] | 0.46576602 | 0.841554994 |
| PCSK9 | Cholesterol_pPS | 4225 | -0.052465331 | [-0.08 -0.02] | 0.000657447 | 0.038607533 |
| PCSK9 | Lipodystrophy_1_pPS | 4225 | 0.050195492 | [0.02 0.08] | 0.001117435 | 0.051789895 |
| PCSK9 | Lipodystrophy_2_pPS | 4225 | 0.00851935 | [-0.02 0.04] | 0.580388898 | 0.903823705 |
| PCSK9 | Liver-Lipid_pPS | 4225 | -0.043918932 | [-0.07 -0.01] | 0.00435541 | 0.130662292 |
| PCSK9 | ASAT_Pos_pPS | 4225 | 0.028466976 | [-0. 0.06] | 0.064667475 | 0.475856102 |
| PCSK9 | ALP_Neg_pPS | 4225 | 0.001330001 | [-0.03 0.03] | 0.931226664 | 0.990056084 |
| PCSK9 | SHBG-LpA_pPS | 4225 | -0.011642274 | [-0.04 0.02] | 0.4499648 | 0.831350198 |
| PCSK9 | Obesity_pPS | 4225 | 0.000397179 | [-0.03 0.03] | 0.979438977 | 0.996862516 |
| PCSK9 | VAT_Neg_pPS | 4225 | -0.009851554 | [-0.04 0.02] | 0.522649405 | 0.872980599 |
| PCSK9 | Bilirubin_pPS | 4225 | -0.002924517 | [-0.03 0.03] | 0.849491289 | 0.97528582 |
| PCSK9 | Proinsulin_pPS | 4225 | 0.023608412 | [-0.01 0.05] | 0.125491638 | 0.622944103 |
| PCSK9 | Hyper_Insulin_pPS | 4225 | -0.00022655 | [-0.03 0.03] | 0.988271168 | 0.999402232 |
| PCSK9 | Beta_Cell_1_pPS | 4225 | -0.007238209 | [-0.04 0.02] | 0.638582431 | 0.915577927 |
| PCSK9 | Beta_Cell_2_pPS | 4225 | -2.74E-05 | [-0.03 0.03] | 0.998580092 | 0.999464614 |
| PCSK9 | Total_GRS | 4225 | 0.017587932 | [-0.01 0.05] | 0.253728292 | 0.739680398 |
| RETN | Cholesterol_pPS | 4225 | -0.007095366 | [-0.04 0.02] | 0.645220184 | 0.916568313 |
| RETN | Lipodystrophy_1_pPS | 4225 | 0.008998559 | [-0.02 0.04] | 0.559278442 | 0.897093962 |
| RETN | Lipodystrophy_2_pPS | 4225 | 0.021559061 | [-0.01 0.05] | 0.161784542 | 0.664061293 |
| RETN | Liver-Lipid_pPS | 4225 | 0.02073684 | [-0.01 0.05] | 0.178391422 | 0.679661798 |
| RETN | ASAT_Pos_pPS | 4225 | 0.0041303 | [-0.03 0.03] | 0.788691954 | 0.957247257 |
| RETN | ALP_Neg_pPS | 4225 | 0.024074191 | [-0.01 0.05] | 0.118202894 | 0.615547148 |
| RETN | SHBG-LpA_pPS | 4225 | -0.003950301 | [-0.03 0.03] | 0.797696367 | 0.958074488 |
| RETN | Obesity_pPS | 4225 | 0.008786801 | [-0.02 0.04] | 0.568560952 | 0.900387238 |
| RETN | VAT_Neg_pPS | 4225 | -0.009921364 | [-0.04 0.02] | 0.519707138 | 0.872596104 |
| RETN | Bilirubin_pPS | 4225 | 0.012748672 | [-0.02 0.04] | 0.408081699 | 0.813804543 |
| RETN | Proinsulin_pPS | 4225 | 0.018268969 | [-0.01 0.05] | 0.235803038 | 0.726631332 |
| RETN | Hyper_Insulin_pPS | 4225 | -0.008299362 | [-0.04 0.02] | 0.590203374 | 0.904476019 |
| RETN | Beta_Cell_1_pPS | 4225 | 0.002071526 | [-0.03 0.03] | 0.893072 | 0.981916701 |
| RETN | Beta_Cell_2_pPS | 4225 | -0.019706341 | [-0.05 0.01] | 0.20095467 | 0.701181907 |
| RETN | Total_GRS | 4225 | 0.012222864 | [-0.02 0.04] | 0.427689696 | 0.82738773 |
| SELP | Cholesterol_pPS | 4225 | -0.021306257 | [-0.05 0.01] | 0.166761425 | 0.666299341 |
| SELP | Lipodystrophy_1_pPS | 4225 | 0.029368829 | [-0. 0.06] | 0.056636396 | 0.451781658 |
| SELP | Lipodystrophy_2_pPS | 4225 | 0.027297385 | [-0. 0.06] | 0.076458856 | 0.511887462 |
| SELP | Liver-Lipid_pPS | 4225 | -0.027601373 | [-0.06 0. ] | 0.073237264 | 0.502472104 |
| SELP | ASAT_Pos_pPS | 4225 | 0.015073895 | [-0.02 0.05] | 0.327987278 | 0.775125841 |
| SELP | ALP_Neg_pPS | 4225 | -0.128782749 | [-0.16 -0.1 ] | 4.80E-17 | 2.21E-14 |
| SELP | SHBG-LpA_pPS | 4225 | 0.008009961 | [-0.02 0.04] | 0.603229731 | 0.908795882 |
| SELP | Obesity_pPS | 4225 | -0.003923752 | [-0.03 0.03] | 0.799026765 | 0.958074488 |
| SELP | VAT_Neg_pPS | 4225 | -0.016383716 | [-0.05 0.01] | 0.287698557 | 0.751557788 |
| SELP | Bilirubin_pPS | 4225 | -0.005607707 | [-0.04 0.02] | 0.715949241 | 0.94141015 |
| SELP | Proinsulin_pPS | 4225 | 0.01624635 | [-0.01 0.05] | 0.291759923 | 0.753421877 |
| SELP | Hyper_Insulin_pPS | 4225 | 0.002184933 | [-0.03 0.03] | 0.887256248 | 0.979291846 |
| SELP | Beta_Cell_1_pPS | 4225 | -0.00273515 | [-0.03 0.03] | 0.859131569 | 0.977006629 |
| SELP | Beta_Cell_2_pPS | 4225 | 0.013921375 | [-0.02 0.04] | 0.366324754 | 0.798890546 |
| SELP | Total_GRS | 4225 | 0.015137246 | [-0.02 0.05] | 0.325958257 | 0.775125841 |
| SOD2 | Cholesterol_pPS | 4159 | 0.018263883 | [-0.01 0.05] | 0.239640219 | 0.728421813 |
| SOD2 | Lipodystrophy_1_pPS | 4159 | -0.010098199 | [-0.04 0.02] | 0.515617083 | 0.870853148 |
| SOD2 | Lipodystrophy_2_pPS | 4159 | 0.031187217 | [0. 0.06] | 0.044616456 | 0.424281318 |
| SOD2 | Liver-Lipid_pPS | 4159 | -0.013155298 | [-0.04 0.02] | 0.39702541 | 0.812191725 |
| SOD2 | ASAT_Pos_pPS | 4159 | -0.011344668 | [-0.04 0.02] | 0.465165172 | 0.841554994 |
| SOD2 | ALP_Neg_pPS | 4159 | 0.008511043 | [-0.02 0.04] | 0.583738528 | 0.903823705 |
| SOD2 | SHBG-LpA_pPS | 4159 | 0.019205379 | [-0.01 0.05] | 0.216267589 | 0.715705689 |
| SOD2 | Obesity_pPS | 4159 | -0.013499056 | [-0.04 0.02] | 0.384803925 | 0.806422804 |
| SOD2 | VAT_Neg_pPS | 4159 | 0.007376451 | [-0.02 0.04] | 0.634868185 | 0.914460114 |
| SOD2 | Bilirubin_pPS | 4159 | 0.023080308 | [-0.01 0.05] | 0.137263998 | 0.629316669 |
| SOD2 | Proinsulin_pPS | 4159 | 0.000175675 | [-0.03 0.03] | 0.990976458 | 0.999402232 |
| SOD2 | Hyper_Insulin_pPS | 4159 | 0.001490996 | [-0.03 0.03] | 0.923530814 | 0.990056084 |
| SOD2 | Beta_Cell_1_pPS | 4159 | 0.001524722 | [-0.03 0.03] | 0.92180662 | 0.990056084 |
| SOD2 | Beta_Cell_2_pPS | 4159 | -0.014781583 | [-0.05 0.02] | 0.341270621 | 0.787547587 |
| SOD2 | Total_GRS | 4159 | -0.00357271 | [-0.03 0.03] | 0.818088751 | 0.963461584 |
| SORT1 | Cholesterol_pPS | 4159 | -0.008741546 | [-0.04 0.02] | 0.573590043 | 0.903411176 |
| SORT1 | Lipodystrophy_1_pPS | 4159 | -0.001355989 | [-0.03 0.03] | 0.930436542 | 0.990056084 |
| SORT1 | Lipodystrophy_2_pPS | 4159 | 0.009618554 | [-0.02 0.04] | 0.535761178 | 0.880766894 |
| SORT1 | Liver-Lipid_pPS | 4159 | -0.020155191 | [-0.05 0.01] | 0.194397846 | 0.696574877 |
| SORT1 | ASAT_Pos_pPS | 4159 | -0.02164863 | [-0.05 0.01] | 0.163361595 | 0.664061293 |
| SORT1 | ALP_Neg_pPS | 4159 | -0.008309388 | [-0.04 0.02] | 0.59268493 | 0.905925461 |
| SORT1 | SHBG-LpA_pPS | 4159 | 0.006880593 | [-0.02 0.04] | 0.657792783 | 0.922711159 |
| SORT1 | Obesity_pPS | 4159 | 0.003824417 | [-0.03 0.03] | 0.805520901 | 0.960193229 |
| SORT1 | VAT_Neg_pPS | 4159 | 0.00792021 | [-0.02 0.04] | 0.610125982 | 0.909483337 |
| SORT1 | Bilirubin_pPS | 4159 | 0.020556979 | [-0.01 0.05] | 0.185651615 | 0.68727393 |
| SORT1 | Proinsulin_pPS | 4159 | 0.028705241 | [-0. 0.06] | 0.064550156 | 0.475856102 |
| SORT1 | Hyper_Insulin_pPS | 4159 | -0.004731606 | [-0.04 0.03] | 0.760661625 | 0.950403646 |
| SORT1 | Beta_Cell_1_pPS | 4159 | 0.008626672 | [-0.02 0.04] | 0.578637203 | 0.903823705 |
| SORT1 | Beta_Cell_2_pPS | 4159 | 0.00048094 | [-0.03 0.03] | 0.975299959 | 0.996862516 |
| SORT1 | Total_GRS | 4159 | 0.020774863 | [-0.01 0.05] | 0.181032029 | 0.680719889 |
| VSIG2 | Cholesterol_pPS | 4159 | 0.015315997 | [-0.02 0.05] | 0.324099788 | 0.774472222 |
| VSIG2 | Lipodystrophy_1_pPS | 4159 | 0.008527414 | [-0.02 0.04] | 0.583015034 | 0.903823705 |
| VSIG2 | Lipodystrophy_2_pPS | 4159 | 0.032145868 | [0. 0.06] | 0.038450949 | 0.398325363 |
| VSIG2 | Liver-Lipid_pPS | 4159 | 0.006736271 | [-0.02 0.04] | 0.664527239 | 0.924111434 |
| VSIG2 | ASAT_Pos_pPS | 4159 | 0.019897704 | [-0.01 0.05] | 0.200159579 | 0.700947446 |
| VSIG2 | ALP_Neg_pPS | 4159 | -0.019893188 | [-0.05 0.01] | 0.200261726 | 0.700947446 |
| VSIG2 | SHBG-LpA_pPS | 4159 | 0.011508899 | [-0.02 0.04] | 0.458728586 | 0.836808259 |
| VSIG2 | Obesity_pPS | 4159 | -0.006297171 | [-0.04 0.02] | 0.685182072 | 0.926969309 |
| VSIG2 | VAT_Neg_pPS | 4159 | 0.02046478 | [-0.01 0.05] | 0.187632452 | 0.689088129 |
| VSIG2 | Bilirubin_pPS | 4159 | -0.01049007 | [-0.04 0.02] | 0.499455886 | 0.863780476 |
| VSIG2 | Proinsulin_pPS | 4159 | 0.012610311 | [-0.02 0.04] | 0.416874117 | 0.817883456 |
| VSIG2 | Hyper_Insulin_pPS | 4159 | 0.004207781 | [-0.03 0.03] | 0.786476503 | 0.957247257 |
| VSIG2 | Beta_Cell_1_pPS | 4159 | 0.003900029 | [-0.03 0.03] | 0.801755209 | 0.958074488 |
| VSIG2 | Beta_Cell_2_pPS | 4159 | 0.021178597 | [-0.01 0.05] | 0.172698466 | 0.674627844 |
| VSIG2 | Total_GRS | 4159 | 0.035658704 | [0.01 0.07] | 0.021655369 | 0.31623713 |
| ACE2 | Cholesterol_pPS | 4159 | 0.01173546 | [-0.02 0.04] | 0.449931619 | 0.831350198 |
| ACE2 | Lipodystrophy_1_pPS | 4159 | 0.005458596 | [-0.02 0.04] | 0.725277042 | 0.942134248 |
| ACE2 | Lipodystrophy_2_pPS | 4159 | 0.067904808 | [0.04 0.1 ] | 1.21E-05 | 0.001663415 |
| ACE2 | Liver-Lipid_pPS | 4159 | -0.048081203 | [-0.08 -0.02] | 0.001953891 | 0.079304974 |

b) Partial Correlation

| Proteomics | PPS | Sample_Size | r | CI95% | p | fdr_p_adjusted |
|---|---|---|---|---|---|---|
| MMP_9 | Bilirubin_pPS | 4225 | 0.016247073 | [-0.01 0.05] | 0.292484589 | 0.816672929 |
| MMP_9 | Proinsulin_pPS | 4225 | 0.037740925 | [0.01 0.07] | 0.014443705 | 0.312663737 |
| MMP_9 | Hyper_Insulin_pPS | 4225 | -0.010542814 | [-0.04 0.02] | 0.494563835 | 0.895803896 |
| MMP_9 | Beta_Cell_1_pPS | 4225 | -0.029150862 | [-0.06 0. ] | 0.05888769 | 0.558815902 |
| MMP_9 | Beta_Cell_2_pPS | 4225 | -0.022166596 | [-0.05 0.01] | 0.15091458 | 0.706166997 |
| PCSK9 | Cholesterol_pPS | 4225 | -0.052642683 | [-0.08 -0.02] | 0.000642551 | 0.043558162 |
| PCSK9 | Lipodystrophy_1_pPS | 4225 | 0.048382343 | [0.02 0.08] | 0.001710138 | 0.093730125 |
| PCSK9 | Lipodystrophy_2_pPS | 4225 | 0.007935062 | [-0.02 0.04] | 0.607178021 | 0.918301271 |
| PCSK9 | Liver-Lipid_pPS | 4225 | -0.04320585 | [-0.07 -0.01] | 0.005101824 | 0.172924966 |
| PCSK9 | ASAT_Pos_pPS | 4225 | 0.019395717 | [-0.01 0.05] | 0.208851363 | 0.767305935 |
| PCSK9 | ALP_Neg_pPS | 4225 | -0.003207154 | [-0.03 0.03] | 0.835396069 | 0.972099644 |
| PCSK9 | SHBG-LpA_pPS | 4225 | -0.01653175 | [-0.05 0.01] | 0.284109122 | 0.812280908 |
| PCSK9 | Obesity_pPS | 4225 | -8.29E-05 | [-0.03 0.03] | 0.995712253 | 0.99881416 |
| PCSK9 | VAT_Neg_pPS | 4225 | -0.011215376 | [-0.04 0.02] | 0.467442899 | 0.879984873 |
| PCSK9 | Bilirubin_pPS | 4225 | -0.005620185 | [-0.04 0.02] | 0.715765336 | 0.945970482 |
| PCSK9 | Proinsulin_pPS | 4225 | 0.02937003 | [-0. 0.06] | 0.057009306 | 0.558815902 |
| PCSK9 | Hyper_Insulin_pPS | 4225 | -0.004402641 | [-0.03 0.03] | 0.775460132 | 0.963155882 |
| PCSK9 | Beta_Cell_1_pPS | 4225 | -0.015320549 | [-0.05 0.01] | 0.320881645 | 0.83927892 |
| PCSK9 | Beta_Cell_2_pPS | 4225 | 0.001754331 | [-0.03 0.03] | 0.909507392 | 0.988841471 |
| RETN | Cholesterol_pPS | 4225 | -0.009473479 | [-0.04 0.02] | 0.53935898 | 0.899241939 |
| RETN | Lipodystrophy_1_pPS | 4225 | 0.008090837 | [-0.02 0.04] | 0.600140355 | 0.918301271 |
| RETN | Lipodystrophy_2_pPS | 4225 | 0.022895762 | [-0.01 0.05] | 0.137923628 | 0.696606 |
| RETN | Liver-Lipid_pPS | 4225 | 0.020215677 | [-0.01 0.05] | 0.190239986 | 0.750471982 |
| RETN | ASAT_Pos_pPS | 4225 | 9.06E-05 | [-0.03 0.03] | 0.995314301 | 0.998803911 |
| RETN | ALP_Neg_pPS | 4225 | 0.0201907 | [-0.01 0.05] | 0.190788273 | 0.750920235 |
| RETN | SHBG-LpA_pPS | 4225 | -0.003661167 | [-0.03 0.03] | 0.812500921 | 0.967638637 |
| RETN | Obesity_pPS | 4225 | 0.00753647 | [-0.02 0.04] | 0.625351102 | 0.927058714 |
| RETN | VAT_Neg_pPS | 4225 | -0.011522309 | [-0.04 0.02] | 0.455345218 | 0.874045665 |
| RETN | Bilirubin_pPS | 4225 | 0.012651214 | [-0.02 0.04] | 0.412397903 | 0.854426619 |
| RETN | Proinsulin_pPS | 4225 | 0.015571711 | [-0.01 0.05] | 0.31301156 | 0.833834311 |
| RETN | Hyper_Insulin_pPS | 4225 | -0.010895993 | [-0.04 0.02] | 0.480218086 | 0.89059884 |
| RETN | Beta_Cell_1_pPS | 4225 | 0.003502354 | [-0.03 0.03] | 0.820492462 | 0.968668398 |
| RETN | Beta_Cell_2_pPS | 4225 | -0.023028091 | [-0.05 0.01] | 0.135661269 | 0.694065456 |
| SELP | Cholesterol_pPS | 4225 | -0.0225441 | [-0.05 0.01] | 0.144077089 | 0.697681852 |
| SELP | Lipodystrophy_1_pPS | 4225 | 0.029882396 | [-0. 0.06] | 0.052811807 | 0.544172861 |
| SELP | Lipodystrophy_2_pPS | 4225 | 0.028332669 | [-0. 0.06] | 0.066359311 | 0.563434057 |
| SELP | Liver-Lipid_pPS | 4225 | -0.023587145 | [-0.05 0.01] | 0.126418387 | 0.694065456 |
| SELP | ASAT_Pos_pPS | 4225 | 0.00788973 | [-0.02 0.04] | 0.609232923 | 0.919381376 |
| SELP | ALP_Neg_pPS | 4225 | -0.133581409 | [-0.16 -0.1 ] | 3.52E-18 | 1.81E-15 |
| SELP | SHBG-LpA_pPS | 4225 | 0.000821103 | [-0.03 0.03] | 0.957574389 | 0.996425456 |
| SELP | Obesity_pPS | 4225 | -0.005743966 | [-0.04 0.02] | 0.709785785 | 0.945970482 |
| SELP | VAT_Neg_pPS | 4225 | -0.021735553 | [-0.05 0.01] | 0.159020967 | 0.716150369 |
| SELP | Bilirubin_pPS | 4225 | -0.00723126 | [-0.04 0.02] | 0.639422891 | 0.933760412 |
| SELP | Proinsulin_pPS | 4225 | 0.040112533 | [0.01 0.07] | 0.009326132 | 0.248521918 |
| SELP | Hyper_Insulin_pPS | 4225 | 0.002336437 | [-0.03 0.03] | 0.879681041 | 0.97781424 |
| SELP | Beta_Cell_1_pPS | 4225 | -0.014333134 | [-0.04 0.02] | 0.353062994 | 0.847284666 |
| SELP | Beta_Cell_2_pPS | 4225 | 0.014046593 | [-0.02 0.04] | 0.362771263 | 0.847284666 |
| SOD2 | Cholesterol_pPS | 4159 | 0.019394554 | [-0.01 0.05] | 0.21249428 | 0.767305935 |
| SOD2 | Lipodystrophy_1_pPS | 4159 | -0.012036914 | [-0.04 0.02] | 0.439095385 | 0.865424415 |
| SOD2 | Lipodystrophy_2_pPS | 4159 | 0.031917301 | [0. 0.06] | 0.040163538 | 0.474592997 |
| SOD2 | Liver-Lipid_pPS | 4159 | -0.014115777 | [-0.04 0.02] | 0.3642157 | 0.847284666 |
| SOD2 | ASAT_Pos_pPS | 4159 | -0.012385565 | [-0.04 0.02] | 0.425953938 | 0.86398216 |
| SOD2 | ALP_Neg_pPS | 4159 | 0.009812133 | [-0.02 0.04] | 0.528232038 | 0.898959051 |
| SOD2 | SHBG-LpA_pPS | 4159 | 0.020472717 | [-0.01 0.05] | 0.188154727 | 0.746820611 |
| SOD2 | Obesity_pPS | 4159 | -0.014221798 | [-0.04 0.02] | 0.36062379 | 0.847284666 |
| SOD2 | VAT_Neg_pPS | 4159 | 0.003787844 | [-0.03 0.03] | 0.807640428 | 0.967638637 |
| SOD2 | Bilirubin_pPS | 4159 | 0.023610438 | [-0.01 0.05] | 0.129061375 | 0.694065456 |
| SOD2 | Proinsulin_pPS | 4159 | -0.000137334 | [-0.03 0.03] | 0.99295687 | 0.998803911 |
| SOD2 | Hyper_Insulin_pPS | 4159 | 0.002381107 | [-0.03 0.03] | 0.878358839 | 0.97781424 |
| SOD2 | Beta_Cell_1_pPS | 4159 | 0.006863951 | [-0.02 0.04] | 0.659069918 | 0.937489141 |
| SOD2 | Beta_Cell_2_pPS | 4159 | -0.017313602 | [-0.05 0.01] | 0.26573189 | 0.80722329 |
| SORT1 | Cholesterol_pPS | 4159 | -0.009008298 | [-0.04 0.02] | 0.562565015 | 0.904956448 |
| SORT1 | Lipodystrophy_1_pPS | 4159 | 0.001575941 | [-0.03 0.03] | 0.919315154 | 0.99041009 |
| SORT1 | Lipodystrophy_2_pPS | 4159 | 0.011728369 | [-0.02 0.04] | 0.450916882 | 0.870351501 |
| SORT1 | Liver-Lipid_pPS | 4159 | -0.019859423 | [-0.05 0.01] | 0.201734673 | 0.765145476 |
| SORT1 | ASAT_Pos_pPS | 4159 | -0.02318078 | [-0.05 0.01] | 0.136175507 | 0.694065456 |
| SORT1 | ALP_Neg_pPS | 4159 | -0.012593473 | [-0.04 0.02] | 0.418827843 | 0.858586823 |
| SORT1 | SHBG-LpA_pPS | 4159 | 0.005006765 | [-0.03 0.04] | 0.747588608 | 0.954135344 |
| SORT1 | Obesity_pPS | 4159 | 0.002287996 | [-0.03 0.03] | 0.883080638 | 0.979257737 |
| SORT1 | VAT_Neg_pPS | 4159 | 0.005705217 | [-0.02 0.04] | 0.713831028 | 0.945970482 |
| SORT1 | Bilirubin_pPS | 4159 | 0.02023458 | [-0.01 0.05] | 0.19334537 | 0.752352981 |
| SORT1 | Proinsulin_pPS | 4159 | 0.029060211 | [-0. 0.06] | 0.061721614 | 0.558815902 |
| SORT1 | Hyper_Insulin_pPS | 4159 | -0.003095643 | [-0.03 0.03] | 0.842279711 | 0.972178273 |
| SORT1 | Beta_Cell_1_pPS | 4159 | 0.001078962 | [-0.03 0.03] | 0.944709267 | 0.996180826 |
| SORT1 | Beta_Cell_2_pPS | 4159 | -0.005629358 | [-0.04 0.02] | 0.717471833 | 0.945970482 |
| VSIG2 | Cholesterol_pPS | 4159 | 0.015641938 | [-0.01 0.05] | 0.314668469 | 0.836563355 |
| VSIG2 | Lipodystrophy_1_pPS | 4159 | 0.003696958 | [-0.03 0.03] | 0.812168661 | 0.967638637 |
| VSIG2 | Lipodystrophy_2_pPS | 4159 | 0.026789233 | [-0. 0.06] | 0.085027172 | 0.614725404 |
| VSIG2 | Liver-Lipid_pPS | 4159 | 0.004982285 | [-0.03 0.04] | 0.748781055 | 0.954135344 |
| VSIG2 | ASAT_Pos_pPS | 4159 | 0.017188323 | [-0.01 0.05] | 0.269206792 | 0.808113719 |
| VSIG2 | ALP_Neg_pPS | 4159 | -0.022144632 | [-0.05 0.01] | 0.15457404 | 0.708464493 |
| VSIG2 | SHBG-LpA_pPS | 4159 | 0.008234828 | [-0.02 0.04] | 0.596585606 | 0.918301271 |
| VSIG2 | Obesity_pPS | 4159 | -0.009950364 | [-0.04 0.02] | 0.522437345 | 0.895803896 |
| VSIG2 | VAT_Neg_pPS | 4159 | 0.015501531 | [-0.01 0.05] | 0.319032595 | 0.839235059 |
| VSIG2 | Bilirubin_pPS | 4159 | -0.011255122 | [-0.04 0.02] | 0.469393935 | 0.882597647 |
| VSIG2 | Proinsulin_pPS | 4159 | 0.013686017 | [-0.02 0.04] | 0.379003514 | 0.85037326 |
| VSIG2 | Hyper_Insulin_pPS | 4159 | -0.001074507 | [-0.03 0.03] | 0.944937199 | 0.996180826 |
| VSIG2 | Beta_Cell_1_pPS | 4159 | -0.006013099 | [-0.04 0.02] | 0.699122011 | 0.945970482 |
| VSIG2 | Beta_Cell_2_pPS | 4159 | 0.018180131 | [-0.01 0.05] | 0.242542174 | 0.791874069 |
| ACE2 | Cholesterol_pPS | 4159 | 0.008199939 | [-0.02 0.04] | 0.598142012 | 0.918301271 |
| ACE2 | Lipodystrophy_1_pPS | 4159 | -0.007298133 | [-0.04 0.02] | 0.638993504 | 0.933760412 |
| ACE2 | Lipodystrophy_2_pPS | 4159 | 0.062421567 | [0.03 0.09] | 5.92E-05 | 0.006926058 |
| ACE2 | Liver-Lipid_pPS | 4159 | -0.050232467 | [-0.08 -0.02] | 0.00123436 | 0.07228782 |
| ACE2 | ASAT_Pos_pPS | 4159 | 0.005581087 | [-0.02 0.04] | 0.719791933 | 0.946314812 |
| ACE2 | ALP_Neg_pPS | 4159 | -0.030735031 | [-0.06 -0. ] | 0.048153127 | 0.520248282 |
| ACE2 | SHBG-LpA_pPS | 4159 | 0.013623141 | [-0.02 0.04] | 0.38119762 | 0.85037326 |
| ACE2 | Obesity_pPS | 4159 | 0.03976077 | [0.01 0.07] | 0.010566913 | 0.259241595 |
| ACE2 | VAT_Neg_pPS | 4159 | 0.029324403 | [-0. 0.06] | 0.059391693 | 0.558815902 |
| ACE2 | Bilirubin_pPS | 4159 | 0.016225596 | [-0.01 0.05] | 0.296949599 | 0.81802395 |
| ACE2 | Proinsulin_pPS | 4159 | -0.002776232 | [-0.03 0.03] | 0.858371541 | 0.97556306 |
| ACE2 | Hyper_Insulin_pPS | 4159 | 0.036578918 | [0.01 0.07] | 0.018674738 | 0.340663962 |
| ACE2 | Beta_Cell_1_pPS | 4159 | -0.008642816 | [-0.04 0.02] | 0.578523684 | 0.909583681 |
| ACE2 | Beta_Cell_2_pPS | 4159 | -0.036985027 | [-0.07 -0.01] | 0.017402543 | 0.33440804 |
| TR_AP | Cholesterol_pPS | 4225 | 0.013801792 | [-0.02 0.04] | 0.371196325 | 0.847284666 |
| TR_AP | Lipodystrophy_1_pPS | 4225 | 0.043280499 | [0.01 0.07] | 0.005025783 | 0.172618901 |
| TR_AP | Lipodystrophy_2_pPS | 4225 | 0.024250662 | [-0.01 0.05] | 0.116093058 | 0.681046037 |
| TR_AP | Liver-Lipid_pPS | 4225 | 0.012030827 | [-0.02 0.04] | 0.43569534 | 0.865276785 |

**Table S7: Associations between pPSs and plasma proteins in SCAPIS**

**a) Spearman Correlation**

| Proteomics | PPS | Sample_Size | r | CI95% | p | fdr_p_adjusted |
|---|---|---|---|---|---|---|
| ACE2 | ASAT_Pos_pPS | 4159 | 0.015245488 | [-0.02 0.05] | 0.326332486 | 0.775125841 |
| ACE2 | ALP_Neg_pPS | 4159 | -0.032424463 | [-0.06 -0. ] | 0.036801056 | 0.387675241 |
| ACE2 | SHBG-LpA_pPS | 4159 | 0.015778086 | [-0.01 0.05] | 0.30971438 | 0.76365449 |
| ACE2 | Obesity_pPS | 4159 | 0.040674632 | [0.01 0.07] | 0.008802598 | 0.204161103 |
| ACE2 | VAT_Neg_pPS | 4159 | 0.035847291 | [0.01 0.07] | 0.020970714 | 0.312860385 |
| ACE2 | Bilirubin_pPS | 4159 | 0.016532914 | [-0.01 0.05] | 0.28713612 | 0.751557788 |
| ACE2 | Proinsulin_pPS | 4159 | -0.01148838 | [-0.04 0.02] | 0.459530028 | 0.837163616 |
| ACE2 | Hyper_Insulin_pPS | 4159 | 0.042508697 | [0.01 0.07] | 0.006184162 | 0.156589778 |
| ACE2 | Beta_Cell_1_pPS | 4159 | -0.01869511 | [-0.05 0.01] | 0.228722921 | 0.726631332 |
| ACE2 | Beta_Cell_2_pPS | 4159 | -0.037931631 | [-0.07 -0.01] | 0.01457245 | 0.252355767 |
| ACE2 | Total_GRS | 4159 | 0.048766471 | [0.02 0.08] | 0.001681728 | 0.071408772 |
| TR_AP | Cholesterol_pPS | 4225 | 0.017207486 | [-0.01 0.05] | 0.26414473 | 0.746520328 |
| TR_AP | Lipodystrophy_1_pPS | 4225 | 0.038818954 | [0.01 0.07] | 0.011740545 | 0.23392385 |
| TR_AP | Lipodystrophy_2_pPS | 4225 | 0.022063072 | [-0.01 0.05] | 0.152197928 | 0.649125224 |
| TR_AP | Liver-Lipid_pPS | 4225 | 0.012227748 | [-0.02 0.04] | 0.427505054 | 0.82738773 |
| TR_AP | ASAT_Pos_pPS | 4225 | -0.012084686 | [-0.04 0.02] | 0.432932047 | 0.828898134 |
| TR_AP | ALP_Neg_pPS | 4225 | -0.033165056 | [-0.06 -0. ] | 0.031348681 | 0.369753673 |
| TR_AP | SHBG-LpA_pPS | 4225 | 0.001854962 | [-0.03 0.03] | 0.904193557 | 0.987954956 |
| TR_AP | Obesity_pPS | 4225 | 0.004466857 | [-0.03 0.03] | 0.771932121 | 0.95301975 |
| TR_AP | VAT_Neg_pPS | 4225 | -0.028710943 | [-0.06 0. ] | 0.062407379 | 0.469330694 |
| TR_AP | Bilirubin_pPS | 4225 | 0.001229216 | [-0.03 0.03] | 0.936426664 | 0.99138381 |
| TR_AP | Proinsulin_pPS | 4225 | 0.020241433 | [-0.01 0.05] | 0.188991593 | 0.691799467 |
| TR_AP | Hyper_Insulin_pPS | 4225 | -0.006915249 | [-0.04 0.02] | 0.653630349 | 0.919948884 |
| TR_AP | Beta_Cell_1_pPS | 4225 | 1.41E-05 | [-0.03 0.03] | 0.999271474 | 0.999464614 |
| TR_AP | Beta_Cell_2_pPS | 4225 | 0.03898559 | [0.01 0.07] | 0.011384384 | 0.232377917 |
| TR_AP | Total_GRS | 4225 | 0.033856226 | [0. 0.06] | 0.027984389 | 0.356352975 |
| AP_N | Cholesterol_pPS | 4225 | 0.009634502 | [-0.02 0.04] | 0.531851893 | 0.877412567 |
| AP_N | Lipodystrophy_1_pPS | 4225 | 0.015514682 | [-0.01 0.05] | 0.314039075 | 0.76365449 |
| AP_N | Lipodystrophy_2_pPS | 4225 | 0.04037422 | [0.01 0.07] | 0.008770165 | 0.204161103 |
| AP_N | Liver-Lipid_pPS | 4225 | -0.020212759 | [-0.05 0.01] | 0.189619055 | 0.692278961 |
| AP_N | ASAT_Pos_pPS | 4225 | -0.003728553 | [-0.03 0.03] | 0.80882625 | 0.96133624 |
| AP_N | ALP_Neg_pPS | 4225 | 0.003557835 | [-0.03 0.03] | 0.817421358 | 0.963461584 |
| AP_N | SHBG-LpA_pPS | 4225 | 0.008260232 | [-0.02 0.04] | 0.591957109 | 0.905925461 |
| AP_N | Obesity_pPS | 4225 | 0.01059415 | [-0.02 0.04] | 0.491795189 | 0.859085266 |
| AP_N | VAT_Neg_pPS | 4225 | 0.025114554 | [-0.01 0.06] | 0.103122757 | 0.586842904 |
| AP_N | Bilirubin_pPS | 4225 | 0.028003183 | [-0. 0.06] | 0.069150202 | 0.48811907 |
| AP_N | Proinsulin_pPS | 4225 | 0.04864557 | [0.02 0.08] | 0.00158637 | 0.068412199 |
| AP_N | Hyper_Insulin_pPS | 4225 | 0.018193815 | [-0.01 0.05] | 0.237735971 | 0.726631332 |
| AP_N | Beta_Cell_1_pPS | 4225 | 0.021902855 | [-0.01 0.05] | 0.155197324 | 0.650979658 |
| AP_N | Beta_Cell_2_pPS | 4225 | 0.008383917 | [-0.02 0.04] | 0.586422037 | 0.903925569 |
| AP_N | Total_GRS | 4225 | 0.047635883 | [0.02 0.08] | 0.001982975 | 0.079319011 |
| CA5A | Cholesterol_pPS | 4159 | 0.054737138 | [0.02 0.09] | 0.000421091 | 0.027671712 |
| CA5A | Lipodystrophy_1_pPS | 4159 | 0.033120709 | [0. 0.06] | 0.032939232 | 0.378159894 |
| CA5A | Lipodystrophy_2_pPS | 4159 | 0.059314619 | [0.03 0.09] | 0.000132422 | 0.013053067 |
| CA5A | Liver-Lipid_pPS | 4159 | -0.037631419 | [-0.07 -0.01] | 0.015372345 | 0.258767146 |
| CA5A | ASAT_Pos_pPS | 4159 | 0.015546921 | [-0.01 0.05] | 0.316857307 | 0.767141882 |
| CA5A | ALP_Neg_pPS | 4159 | 0.015731786 | [-0.01 0.05] | 0.311136457 | 0.76365449 |
| CA5A | SHBG-LpA_pPS | 4159 | -0.000243496 | [-0.03 0.03] | 0.987493093 | 0.99907659 |
| CA5A | Obesity_pPS | 4159 | 0.043855277 | [0.01 0.07] | 0.00473301 | 0.136074023 |
| CA5A | VAT_Neg_pPS | 4159 | 0.016841872 | [-0.01 0.05] | 0.278223207 | 0.747422497 |
| CA5A | Bilirubin_pPS | 4159 | 0.012600436 | [-0.02 0.04] | 0.417239096 | 0.817883456 |
| CA5A | Proinsulin_pPS | 4159 | -0.013465105 | [-0.04 0.02] | 0.386000633 | 0.807658934 |
| CA5A | Hyper_Insulin_pPS | 4159 | 0.016264937 | [-0.01 0.05] | 0.295021184 | 0.757449737 |
| CA5A | Beta_Cell_1_pPS | 4159 | 0.009606193 | [-0.02 0.04] | 0.536285511 | 0.881040482 |
| CA5A | Beta_Cell_2_pPS | 4159 | -0.027175815 | [-0.06 0. ] | 0.080145558 | 0.521702218 |
| CA5A | Total_GRS | 4159 | 0.052914914 | [0.02 0.08] | 0.000652174 | 0.038607533 |
| GIF | Cholesterol_pPS | 4159 | 0.010628918 | [-0.02 0.04] | 0.493794862 | 0.861487876 |
| GIF | Lipodystrophy_1_pPS | 4159 | 0.024682216 | [-0.01 0.06] | 0.112008787 | 0.604978148 |
| GIF | Lipodystrophy_2_pPS | 4159 | -0.001657288 | [-0.03 0.03] | 0.915032834 | 0.990056084 |
| GIF | Liver-Lipid_pPS | 4159 | 0.005850138 | [-0.02 0.04] | 0.706454388 | 0.937738119 |
| GIF | ASAT_Pos_pPS | 4159 | 0.004145414 | [-0.03 0.03] | 0.789566397 | 0.957247257 |
| GIF | ALP_Neg_pPS | 4159 | -0.136034288 | [-0.17 -0.11] | 1.39E-18 | 7.65E-16 |
| GIF | SHBG-LpA_pPS | 4159 | 0.023736551 | [-0.01 0.05] | 0.126432603 | 0.623533826 |
| GIF | Obesity_pPS | 4159 | -0.002226476 | [-0.03 0.03] | 0.886024987 | 0.979291846 |
| GIF | VAT_Neg_pPS | 4159 | 0.008172698 | [-0.02 0.04] | 0.598784656 | 0.907623171 |
| GIF | Bilirubin_pPS | 4159 | -0.017291762 | [-0.05 0.01] | 0.26558448 | 0.746520328 |
| GIF | Proinsulin_pPS | 4159 | -0.050927607 | [-0.08 -0.02] | 0.001035367 | 0.050992277 |
| GIF | Hyper_Insulin_pPS | 4159 | -0.024205867 | [-0.05 0.01] | 0.119103261 | 0.615837655 |
| GIF | Beta_Cell_1_pPS | 4159 | -0.000837001 | [-0.03 0.03] | 0.957027354 | 0.99612031 |
| GIF | Beta_Cell_2_pPS | 4159 | 0.023564002 | [-0.01 0.05] | 0.129213905 | 0.624571591 |
| GIF | Total_GRS | 4159 | 0.009693593 | [-0.02 0.04] | 0.532583723 | 0.878093252 |
| CHIT1 | Cholesterol_pPS | 4225 | -0.011577505 | [-0.04 0.02] | 0.452489854 | 0.834174785 |
| CHIT1 | Lipodystrophy_1_pPS | 4225 | -0.017528325 | [-0.05 0.01] | 0.255341132 | 0.74027471 |
| CHIT1 | Lipodystrophy_2_pPS | 4225 | -0.015381956 | [-0.05 0.01] | 0.318197399 | 0.76767904 |
| CHIT1 | Liver-Lipid_pPS | 4225 | 0.000166496 | [-0.03 0.03] | 0.99138012 | 0.999402232 |
| CHIT1 | ASAT_Pos_pPS | 4225 | 0.011771074 | [-0.02 0.04] | 0.444967225 | 0.831350198 |
| CHIT1 | ALP_Neg_pPS | 4225 | -0.014818342 | [-0.04 0.02] | 0.336255204 | 0.782502359 |
| CHIT1 | SHBG-LpA_pPS | 4225 | 0.005334033 | [-0.02 0.04] | 0.729253176 | 0.942134248 |
| CHIT1 | Obesity_pPS | 4225 | -0.008176003 | [-0.04 0.02] | 0.595740106 | 0.907418705 |
| CHIT1 | VAT_Neg_pPS | 4225 | -0.00409566 | [-0.03 0.03] | 0.790422644 | 0.957247257 |
| CHIT1 | Bilirubin_pPS | 4225 | -3.44E-05 | [-0.03 0.03] | 0.998220804 | 0.999464614 |
| CHIT1 | Proinsulin_pPS | 4225 | 0.003447398 | [-0.03 0.03] | 0.822993345 | 0.963461584 |
| CHIT1 | Hyper_Insulin_pPS | 4225 | -0.006484238 | [-0.04 0.02] | 0.673933245 | 0.925361755 |
| CHIT1 | Beta_Cell_1_pPS | 4225 | -0.002457717 | [-0.03 0.03] | 0.873292679 | 0.977804379 |
| CHIT1 | Beta_Cell_2_pPS | 4225 | -0.006083147 | [-0.04 0.02] | 0.693042794 | 0.930013012 |
| CHIT1 | Total_GRS | 4225 | -0.033653561 | [-0.06 -0. ] | 0.028937032 | 0.364685878 |
| CPA1 | Cholesterol_pPS | 4225 | -0.019283095 | [-0.05 0.01] | 0.210801382 | 0.710713005 |
| CPA1 | Lipodystrophy_1_pPS | 4225 | 0.034992033 | [0. 0.07] | 0.023130556 | 0.324062605 |
| CPA1 | Lipodystrophy_2_pPS | 4225 | 0.021074834 | [-0.01 0.05] | 0.171417457 | 0.674627844 |
| CPA1 | Liver-Lipid_pPS | 4225 | -0.058206968 | [-0.09 -0.03] | 0.000156728 | 0.014916138 |
| CPA1 | ASAT_Pos_pPS | 4225 | -0.006123505 | [-0.04 0.02] | 0.691110867 | 0.928659199 |
| CPA1 | ALP_Neg_pPS | 4225 | -0.027267369 | [-0.06 0. ] | 0.076783119 | 0.511887462 |
| CPA1 | SHBG-LpA_pPS | 4225 | -0.025983181 | [-0.06 0. ] | 0.091739798 | 0.556626417 |
| CPA1 | Obesity_pPS | 4225 | -0.00309106 | [-0.03 0.03] | 0.841031442 | 0.972342391 |
| CPA1 | VAT_Neg_pPS | 4225 | -0.012050024 | [-0.04 0.02] | 0.434252913 | 0.828998134 |
| CPA1 | Bilirubin_pPS | 4225 | -0.001031048 | [-0.03 0.03] | 0.946658896 | 0.993154165 |
| CPA1 | Proinsulin_pPS | 4225 | 0.006611548 | [-0.02 0.04] | 0.667910694 | 0.924640793 |
| CPA1 | Hyper_Insulin_pPS | 4225 | -0.003915276 | [-0.03 0.03] | 0.799451641 | 0.958074488 |
| CPA1 | Beta_Cell_1_pPS | 4225 | -0.002221914 | [-0.03 0.03] | 0.885361084 | 0.979291846 |
| CPA1 | Beta_Cell_2_pPS | 4225 | 0.002311728 | [-0.03 0.03] | 0.880761104 | 0.979035738 |
| CPA1 | Total_GRS | 4225 | 0.000477582 | [-0.03 0.03] | 0.975277914 | 0.996862516 |
| CTSL1 | Cholesterol_pPS | 4159 | 0.023288918 | [-0.01 0.05] | 0.133745921 | 0.629316669 |
| CTSL1 | Lipodystrophy_1_pPS | 4159 | -0.008594141 | [-0.04 0.02] | 0.580070312 | 0.903823705 |
| CTSL1 | Lipodystrophy_2_pPS | 4159 | 0.034523681 | [0. 0.06] | 0.026200899 | 0.344354675 |
| CTSL1 | Liver-Lipid_pPS | 4159 | 0.007500251 | [-0.02 0.04] | 0.62919772 | 0.914460114 |
| CTSL1 | ASAT_Pos_pPS | 4159 | -0.007729542 | [-0.04 0.02] | 0.618753123 | 0.909483337 |
| CTSL1 | ALP_Neg_pPS | 4159 | -0.000155693 | [-0.03 0.03] | 0.992002801 | 0.999402232 |

**b) Partial Correlation**

| Proteomics | PPS | Sample_Size | r | CI95% | p | fdr_p_adjusted |
|---|---|---|---|---|---|---|
| TR_AP | ASAT_Pos_pPS | 4225 | -0.01987035 | [-0.05 0.01] | 0.197923676 | 0.763097292 |
| TR_AP | ALP_Neg_pPS | 4225 | -0.038286932 | [-0.07 -0.01] | 0.01308498 | 0.297162209 |
| TR_AP | SHBG-LpA_pPS | 4225 | -0.001481751 | [-0.03 0.03] | 0.923520664 | 0.990503296 |
| TR_AP | Obesity_pPS | 4225 | -9.96E-05 | [-0.03 0.03] | 0.994850619 | 0.998803911 |
| TR_AP | VAT_Neg_pPS | 4225 | -0.033271356 | [-0.06 -0. ] | 0.031069193 | 0.419027441 |
| TR_AP | Bilirubin_pPS | 4225 | 0.000353392 | [-0.03 0.03] | 0.981733552 | 0.998803911 |
| TR_AP | Proinsulin_pPS | 4225 | 0.022168602 | [-0.01 0.05] | 0.150877615 | 0.706166997 |
| TR_AP | Hyper_Insulin_pPS | 4225 | -0.009842571 | [-0.04 0.02] | 0.523670893 | 0.896925678 |
| TR_AP | Beta_Cell_1_pPS | 4225 | -0.012290839 | [-0.04 0.02] | 0.425840039 | 0.86398216 |
| TR_AP | Beta_Cell_2_pPS | 4225 | 0.038990919 | [0.01 0.07] | 0.011500576 | 0.27431004 |
| AP_N | Cholesterol_pPS | 4225 | 0.008250949 | [-0.02 0.04] | 0.592945489 | 0.916823277 |
| AP_N | Lipodystrophy_1_pPS | 4225 | 0.011639945 | [-0.02 0.04] | 0.450755774 | 0.870351501 |
| AP_N | Lipodystrophy_2_pPS | 4225 | 0.037249336 | [0.01 0.07] | 0.015771877 | 0.32502684 |
| AP_N | Liver-Lipid_pPS | 4225 | -0.023253257 | [-0.05 0.01] | 0.131877722 | 0.694065456 |
| AP_N | ASAT_Pos_pPS | 4225 | -0.011954725 | [-0.04 0.02] | 0.438604536 | 0.865276785 |
| AP_N | ALP_Neg_pPS | 4225 | -0.003912757 | [-0.03 0.03] | 0.799880967 | 0.967638637 |
| AP_N | SHBG-LpA_pPS | 4225 | 0.003995 | [-0.03 0.03] | 0.795766745 | 0.967638637 |
| AP_N | Obesity_pPS | 4225 | 0.003386041 | [-0.03 0.03] | 0.826357265 | 0.970677755 |
| AP_N | VAT_Neg_pPS | 4225 | 0.016568325 | [-0.01 0.05] | 0.283044931 | 0.81132836 |
| AP_N | Bilirubin_pPS | 4225 | 0.026378636 | [-0. 0.06] | 0.087391774 | 0.616770438 |
| AP_N | Proinsulin_pPS | 4225 | 0.042947352 | [0.01 0.07] | 0.005373238 | 0.175208366 |
| AP_N | Hyper_Insulin_pPS | 4225 | 0.013800057 | [-0.02 0.04] | 0.371256473 | 0.847284666 |
| AP_N | Beta_Cell_1_pPS | 4225 | 0.007381096 | [-0.02 0.04] | 0.632498039 | 0.931037113 |
| AP_N | Beta_Cell_2_pPS | 4225 | -0.004009128 | [-0.03 0.03] | 0.795060551 | 0.967638637 |
| CA5A | Cholesterol_pPS | 4159 | 0.051270907 | [0.02 0.08] | 0.000974817 | 0.06438788 |
| CA5A | Lipodystrophy_1_pPS | 4159 | 0.024171821 | [-0.01 0.05] | 0.120205163 | 0.682045157 |
| CA5A | Lipodystrophy_2_pPS | 4159 | 0.055573947 | [0.03 0.09] | 0.000350395 | 0.028631564 |
| CA5A | Liver-Lipid_pPS | 4159 | -0.041886413 | [-0.07 -0.01] | 0.007070522 | 0.206973464 |
| CA5A | ASAT_Pos_pPS | 4159 | 0.003321707 | [-0.03 0.03] | 0.830929907 | 0.971458352 |
| CA5A | ALP_Neg_pPS | 4159 | 0.019490801 | [-0.01 0.05] | 0.210233274 | 0.767305935 |
| CA5A | SHBG-LpA_pPS | 4159 | -0.000258493 | [-0.03 0.03] | 0.986743661 | 0.998803911 |
| CA5A | Obesity_pPS | 4159 | 0.040308226 | [0.01 0.07] | 0.009543782 | 0.248521918 |
| CA5A | VAT_Neg_pPS | 4159 | 0.011299016 | [-0.02 0.04] | 0.467662768 | 0.879984873 |
| CA5A | Bilirubin_pPS | 4159 | 0.010564561 | [-0.02 0.04] | 0.497090612 | 0.895803896 |
| CA5A | Proinsulin_pPS | 4159 | -0.022803962 | [-0.05 0.01] | 0.142660518 | 0.697681852 |
| CA5A | Hyper_Insulin_pPS | 4159 | 0.006400101 | [-0.02 0.04] | 0.680792531 | 0.943368241 |
| CA5A | Beta_Cell_1_pPS | 4159 | 0.023434133 | [-0.01 0.05] | 0.131944844 | 0.694065456 |
| CA5A | Beta_Cell_2_pPS | 4159 | -0.030169131 | [-0.06 0. ] | 0.05242649 | 0.542372046 |
| GIF | Cholesterol_pPS | 4159 | 0.012047721 | [-0.02 0.04] | 0.438684581 | 0.865276785 |
| GIF | Lipodystrophy_1_pPS | 4159 | 0.028695728 | [-0. 0.06] | 0.065059667 | 0.562394973 |
| GIF | Lipodystrophy_2_pPS | 4159 | -0.003289498 | [-0.03 0.03] | 0.832544868 | 0.971458352 |
| GIF | Liver-Lipid_pPS | 4159 | 0.010691649 | [-0.02 0.04] | 0.491928936 | 0.895803896 |
| GIF | ASAT_Pos_pPS | 4159 | 0.001946818 | [-0.03 0.03] | 0.900416491 | 0.985399384 |
| GIF | ALP_Neg_pPS | 4159 | -0.128309448 | [-0.16 -0.1 ] | 1.22E-16 | 5.22E-14 |
| GIF | SHBG-LpA_pPS | 4159 | 0.018999819 | [-0.01 0.05] | 0.221950979 | 0.779500334 |
| GIF | Obesity_pPS | 4159 | 0.003607359 | [-0.03 0.03] | 0.81663897 | 0.968668398 |
| GIF | VAT_Neg_pPS | 4159 | 0.007747174 | [-0.02 0.04] | 0.61850477 | 0.92152711 |
| GIF | Bilirubin_pPS | 4159 | -0.017584194 | [-0.05 0.01] | 0.258332088 | 0.805645834 |
| GIF | Proinsulin_pPS | 4159 | -0.034236518 | [-0.06 -0. ] | 0.027717818 | 0.409743846 |
| GIF | Hyper_Insulin_pPS | 4159 | -0.027128185 | [-0.06 0. ] | 0.08115344 | 0.610643339 |
| GIF | Beta_Cell_1_pPS | 4159 | 0.001215441 | [-0.03 0.03] | 0.937728914 | 0.994163908 |
| GIF | Beta_Cell_2_pPS | 4159 | 0.030003486 | [-0. 0.06] | 0.053735745 | 0.547701318 |
| CHIT1 | Cholesterol_pPS | 4225 | -0.010069841 | [-0.04 0.02] | 0.514128615 | 0.895803896 |
| CHIT1 | Lipodystrophy_1_pPS | 4225 | -0.018399858 | [-0.05 0.01] | 0.233185549 | 0.786988264 |
| CHIT1 | Lipodystrophy_2_pPS | 4225 | -0.014981268 | [-0.05 0.02] | 0.331716328 | 0.844649909 |
| CHIT1 | Liver-Lipid_pPS | 4225 | 0.001840405 | [-0.03 0.03] | 0.905087995 | 0.988345348 |
| CHIT1 | ASAT_Pos_pPS | 4225 | 0.016642126 | [-0.01 0.05] | 0.280905838 | 0.808820203 |
| CHIT1 | ALP_Neg_pPS | 4225 | -0.015196162 | [-0.05 0.02] | 0.324826696 | 0.842951766 |
| CHIT1 | SHBG-LpA_pPS | 4225 | 0.006045667 | [-0.02 0.04] | 0.695286585 | 0.945970482 |
| CHIT1 | Obesity_pPS | 4225 | -0.007498186 | [-0.04 0.02] | 0.627108827 | 0.928365329 |
| CHIT1 | VAT_Neg_pPS | 4225 | -0.001549195 | [-0.03 0.03] | 0.920050988 | 0.99041009 |
| CHIT1 | Bilirubin_pPS | 4225 | 0.000292404 | [-0.03 0.03] | 0.984885555 | 0.998803911 |
| CHIT1 | Proinsulin_pPS | 4225 | 0.008403939 | [-0.02 0.04] | 0.586107776 | 0.911172983 |
| CHIT1 | Hyper_Insulin_pPS | 4225 | -0.00333863 | [-0.03 0.03] | 0.828750673 | 0.971458352 |
| CHIT1 | Beta_Cell_1_pPS | 4225 | -0.003534045 | [-0.03 0.03] | 0.818896267 | 0.968668398 |
| CHIT1 | Beta_Cell_2_pPS | 4225 | -0.005597313 | [-0.04 0.02] | 0.716872168 | 0.945970482 |
| CPA1 | Cholesterol_pPS | 4225 | -0.019596984 | [-0.05 0.01] | 0.204165286 | 0.765145476 |
| CPA1 | Lipodystrophy_1_pPS | 4225 | 0.039601231 | [0.01 0.07] | 0.010267259 | 0.254312113 |
| CPA1 | Lipodystrophy_2_pPS | 4225 | 0.024304191 | [-0.01 0.05] | 0.115289833 | 0.681046037 |
| CPA1 | Liver-Lipid_pPS | 4225 | -0.057304982 | [-0.09 -0.03] | 0.000202614 | 0.018640452 |
| CPA1 | ASAT_Pos_pPS | 4225 | -0.014200764 | [-0.04 0.02] | 0.357527213 | 0.847284666 |
| CPA1 | ALP_Neg_pPS | 4225 | -0.028636109 | [-0.06 0. ] | 0.063502026 | 0.558815902 |
| CPA1 | SHBG-LpA_pPS | 4225 | -0.029759777 | [-0.06 0. ] | 0.053792094 | 0.547701318 |
| CPA1 | Obesity_pPS | 4225 | -0.002609712 | [-0.03 0.03] | 0.865734792 | 0.97556306 |
| CPA1 | VAT_Neg_pPS | 4225 | -0.013634419 | [-0.04 0.02] | 0.377025925 | 0.849453763 |
| CPA1 | Bilirubin_pPS | 4225 | -0.003105701 | [-0.03 0.03] | 0.840532035 | 0.972178273 |
| CPA1 | Proinsulin_pPS | 4225 | 0.014763024 | [-0.02 0.04] | 0.338809253 | 0.844649909 |
| CPA1 | Hyper_Insulin_pPS | 4225 | -0.004200198 | [-0.03 0.03] | 0.785526576 | 0.966801939 |
| CPA1 | Beta_Cell_1_pPS | 4225 | -0.00395242 | [-0.03 0.03] | 0.797896144 | 0.967638637 |
| CPA1 | Beta_Cell_2_pPS | 4225 | 0.003910443 | [-0.03 0.03] | 0.79999682 | 0.967638637 |
| CTSL1 | Cholesterol_pPS | 4159 | 0.018457234 | [-0.01 0.05] | 0.235435948 | 0.788664502 |
| CTSL1 | Lipodystrophy_1_pPS | 4159 | -0.006327574 | [-0.04 0.02] | 0.684213716 | 0.944323894 |
| CTSL1 | Lipodystrophy_2_pPS | 4159 | 0.034234087 | [0. 0.06] | 0.027728886 | 0.409743846 |
| CTSL1 | Liver-Lipid_pPS | 4159 | 0.006637613 | [-0.02 0.04] | 0.669635101 | 0.937489141 |
| CTSL1 | ASAT_Pos_pPS | 4159 | -0.007977557 | [-0.04 0.02] | 0.608105519 | 0.918301271 |
| CTSL1 | ALP_Neg_pPS | 4159 | -0.003791293 | [-0.03 0.03] | 0.807468691 | 0.967638637 |
| CTSL1 | SHBG-LpA_pPS | 4159 | -0.021635794 | [-0.05 0.01] | 0.164273579 | 0.718997225 |
| CTSL1 | Obesity_pPS | 4159 | 0.021245208 | [-0.01 0.05] | 0.172024677 | 0.727559563 |
| CTSL1 | VAT_Neg_pPS | 4159 | -0.003491607 | [-0.03 0.03] | 0.822422869 | 0.968668398 |
| CTSL1 | Bilirubin_pPS | 4159 | -0.01034609 | [-0.04 0.02] | 0.506030782 | 0.895803896 |
| CTSL1 | Proinsulin_pPS | 4159 | 0.014337461 | [-0.02 0.04] | 0.356730645 | 0.847284666 |
| CTSL1 | Hyper_Insulin_pPS | 4159 | -0.006655887 | [-0.04 0.02] | 0.668779643 | 0.937489141 |
| CTSL1 | Beta_Cell_1_pPS | 4159 | -0.03732857 | [-0.07 -0.01] | 0.016386318 | 0.33440804 |
| CTSL1 | Beta_Cell_2_pPS | 4159 | 0.013051078 | [-0.02 0.04] | 0.401516141 | 0.85241904 |
| CTSZ | Cholesterol_pPS | 4225 | 0.054414705 | [0.02 0.08] | 0.000418649 | 0.031718795 |
| CTSZ | Lipodystrophy_1_pPS | 4225 | 0.029770697 | [-0. 0.06] | 0.053704179 | 0.547701318 |
| CTSZ | Lipodystrophy_2_pPS | 4225 | 0.031287929 | [0. 0.06] | 0.042602893 | 0.489933267 |
| CTSZ | Liver-Lipid_pPS | 4225 | -0.018946088 | [-0.05 0.01] | 0.219600354 | 0.775610053 |
| CTSZ | ASAT_Pos_pPS | 4225 | 0.000108687 | [-0.03 0.03] | 0.99438165 | 0.998803911 |
| CTSZ | ALP_Neg_pPS | 4225 | 0.007913375 | [-0.02 0.04] | 0.608160702 | 0.918301271 |
| CTSZ | SHBG-LpA_pPS | 4225 | -0.022880557 | [-0.05 0.01] | 0.138185429 | 0.696606 |
| CTSZ | Obesity_pPS | 4225 | 0.01859634 | [-0.01 0.05] | 0.228231854 | 0.786988264 |
| CTSZ | VAT_Neg_pPS | 4225 | -0.000939268 | [-0.03 0.03] | 0.951475928 | 0.996425456 |
| CTSZ | Bilirubin_pPS | 4225 | 0.015471867 | [-0.01 0.05] | 0.316124799 | 0.838658582 |
| CTSZ | Proinsulin_pPS | 4225 | -0.017424078 | [-0.05 0.01] | 0.258915188 | 0.80598779 |
| CTSZ | Hyper_Insulin_pPS | 4225 | -0.007605006 | [-0.04 0.02] | 0.62220964 | 0.924343732 |
| CTSZ | Beta_Cell_1_pPS | 4225 | 0.013822001 | [-0.02 0.04] | 0.370496243 | 0.847284666 |

**Table S7: Associations between pPSs and plasma proteins in SCAPIS**

**a) Spearman Correlation**

| Proteomics | PPS | Sample_Size | r | CI95% | p | fdr_p_adjusted |
|---|---|---|---|---|---|---|
| CTSL1 | SHBG-LpA_pPS | 4159 | -0.021598898 | [-0.05 0.01] | 0.16433111 | 0.664061293 |
| CTSL1 | Obesity_pPS | 4159 | 0.02386562 | [-0.01 0.05] | 0.124382783 | 0.61966874 |
| CTSL1 | VAT_Neg_pPS | 4159 | -0.005566501 | [-0.04 0.02] | 0.720072554 | 0.941449667 |
| CTSL1 | Bilirubin_pPS | 4159 | -0.011137671 | [-0.04 0.02] | 0.473348844 | 0.846453261 |
| CTSL1 | Proinsulin_pPS | 4159 | 0.007424895 | [-0.02 0.04] | 0.632646709 | 0.914460114 |
| CTSL1 | Hyper_Insulin_pPS | 4159 | -0.004402237 | [-0.03 0.03] | 0.77686433 | 0.953637204 |
| CTSL1 | Beta_Cell_1_pPS | 4159 | -0.034273335 | [-0.06 -0. ] | 0.027307774 | 0.352193725 |
| CTSL1 | Beta_Cell_2_pPS | 4159 | 0.006257546 | [-0.02 0.04] | 0.687057891 | 0.926969309 |
| CTSL1 | Total_GRS | 4159 | 0.015889712 | [-0.01 0.05] | 0.306303528 | 0.76365449 |
| CTSZ | Cholesterol_pPS | 4225 | 0.056639411 | [0.03 0.09] | 0.000234893 | 0.01852303 |
| CTSZ | Lipodystrophy_1_pPS | 4225 | 0.031588372 | [0. 0.06] | 0.040342048 | 0.407062052 |
| CTSZ | Lipodystrophy_2_pPS | 4225 | 0.033218315 | [0. 0.06] | 0.031077642 | 0.369452038 |
| CTSZ | Liver-Lipid_pPS | 4225 | -0.016763434 | [-0.05 0.01] | 0.276670929 | 0.747422497 |
| CTSZ | ASAT_Pos_pPS | 4225 | 0.008656226 | [-0.02 0.04] | 0.574321211 | 0.903411176 |
| CTSZ | ALP_Neg_pPS | 4225 | 0.005791788 | [-0.02 0.04] | 0.707048623 | 0.937738119 |
| CTSZ | SHBG-LpA_pPS | 4225 | -0.0227784 | [-0.05 0.01] | 0.139342222 | 0.634028726 |
| CTSZ | Obesity_pPS | 4225 | 0.022216173 | [-0.01 0.05] | 0.149373163 | 0.64822316 |
| CTSZ | VAT_Neg_pPS | 4225 | -1.39E-05 | [-0.03 0.03] | 0.999281428 | 0.999464614 |
| CTSZ | Bilirubin_pPS | 4225 | 0.016952951 | [-0.01 0.05] | 0.271276203 | 0.746520328 |
| CTSZ | Proinsulin_pPS | 4225 | -0.01343744 | [-0.04 0.02] | 0.383223152 | 0.806170656 |
| CTSZ | Hyper_Insulin_pPS | 4225 | -0.00109895 | [-0.03 0.03] | 0.94315174 | 0.992903797 |
| CTSZ | Beta_Cell_1_pPS | 4225 | 0.004967158 | [-0.03 0.04] | 0.74721577 | 0.947292085 |
| CTSZ | Beta_Cell_2_pPS | 4225 | -0.014156598 | [-0.04 0.02] | 0.35828151 | 0.7979754 |
| CTSZ | Total_GRS | 4225 | 0.024383905 | [-0.01 0.05] | 0.113543 | 0.604978148 |
| EGFR | Cholesterol_pPS | 4225 | -0.005271138 | [-0.04 0.02] | 0.73232238 | 0.942134248 |
| EGFR | Lipodystrophy_1_pPS | 4225 | 0.024917498 | [-0.01 0.06] | 0.105855237 | 0.595837331 |
| EGFR | Lipodystrophy_2_pPS | 4225 | 0.011701332 | [-0.02 0.04] | 0.447669333 | 0.831350198 |
| EGFR | Liver-Lipid_pPS | 4225 | -0.035601687 | [-0.07 -0.01] | 0.020840157 | 0.312602357 |
| EGFR | ASAT_Pos_pPS | 4225 | 0.000999994 | [-0.03 0.03] | 0.948263181 | 0.993154165 |
| EGFR | ALP_Neg_pPS | 4225 | -0.023833629 | [-0.05 0.01] | 0.12192477 | 0.615837655 |
| EGFR | SHBG-LpA_pPS | 4225 | 0.004476448 | [-0.03 0.03] | 0.771456033 | 0.95301975 |
| EGFR | Obesity_pPS | 4225 | -0.007796719 | [-0.04 0.02] | 0.612909967 | 0.909483337 |
| EGFR | VAT_Neg_pPS | 4225 | -0.016303665 | [-0.05 0.01] | 0.2900607 | 0.753421877 |
| EGFR | Bilirubin_pPS | 4225 | -0.006222603 | [-0.04 0.02] | 0.686375641 | 0.926969309 |
| EGFR | Proinsulin_pPS | 4225 | 0.008876066 | [-0.02 0.04] | 0.564639009 | 0.899863296 |
| EGFR | Hyper_Insulin_pPS | 4225 | -0.002650407 | [-0.03 0.03] | 0.863452549 | 0.977006629 |
| EGFR | Beta_Cell_1_pPS | 4225 | 0.012997867 | [-0.02 0.04] | 0.398979083 | 0.81230319 |
| EGFR | Beta_Cell_2_pPS | 4225 | 0.012333822 | [-0.02 0.04] | 0.423506893 | 0.824896983 |
| EGFR | Total_GRS | 4225 | 0.001357639 | [-0.03 0.03] | 0.929801167 | 0.990056084 |
| GLO1 | Cholesterol_pPS | 4159 | -0.015738478 | [-0.05 0.01] | 0.310930623 | 0.76365449 |
| GLO1 | Lipodystrophy_1_pPS | 4159 | 0.029017509 | [-0. 0.06] | 0.061695676 | 0.466520728 |
| GLO1 | Lipodystrophy_2_pPS | 4159 | 0.003206198 | [-0.03 0.03] | 0.836471837 | 0.970026164 |
| GLO1 | Liver-Lipid_pPS | 4159 | -0.004790556 | [-0.04 0.03] | 0.757772513 | 0.950403646 |
| GLO1 | ASAT_Pos_pPS | 4159 | -0.007976851 | [-0.04 0.02] | 0.607573491 | 0.909483337 |
| GLO1 | ALP_Neg_pPS | 4159 | -0.009129782 | [-0.04 0.02] | 0.556688396 | 0.896417721 |
| GLO1 | SHBG-LpA_pPS | 4159 | -0.015514706 | [-0.05 0.01] | 0.317861261 | 0.767539002 |
| GLO1 | Obesity_pPS | 4159 | 0.014469081 | [-0.02 0.04] | 0.351576234 | 0.794717776 |
| GLO1 | VAT_Neg_pPS | 4159 | 0.012038129 | [-0.02 0.04] | 0.43833008 | 0.830394412 |
| GLO1 | Bilirubin_pPS | 4159 | 0.003178759 | [-0.03 0.03] | 0.837851823 | 0.971218409 |
| GLO1 | Proinsulin_pPS | 4159 | 0.017575609 | [-0.01 0.05] | 0.257816807 | 0.740941247 |
| GLO1 | Hyper_Insulin_pPS | 4159 | -0.009006169 | [-0.04 0.02] | 0.562043423 | 0.897748415 |
| GLO1 | Beta_Cell_1_pPS | 4159 | -0.026632973 | [-0.06 0. ] | 0.086367937 | 0.54140702 |
| GLO1 | Beta_Cell_2_pPS | 4159 | -0.022236934 | [-0.05 0.01] | 0.152217743 | 0.649125224 |
| GLO1 | Total_GRS | 4159 | 0.007112577 | [-0.02 0.04] | 0.64702587 | 0.91814468 |
| HAOX1 | Cholesterol_pPS | 4159 | 0.024911446 | [-0.01 0.06] | 0.10871565 | 0.600110386 |
| HAOX1 | Lipodystrophy_1_pPS | 4159 | 0.029485647 | [-0. 0.06] | 0.057612666 | 0.45329986 |
| HAOX1 | Lipodystrophy_2_pPS | 4159 | 0.051725514 | [0.02 0.08] | 0.000861608 | 0.047560737 |
| HAOX1 | Liver-Lipid_pPS | 4159 | -0.057535042 | [-0.09 -0.03] | 0.000209688 | 0.017829955 |
| HAOX1 | ASAT_Pos_pPS | 4159 | 0.01025919 | [-0.02 0.04] | 0.508944968 | 0.868660083 |
| HAOX1 | ALP_Neg_pPS | 4159 | 0.006895609 | [-0.02 0.04] | 0.657093675 | 0.922711159 |
| HAOX1 | SHBG-LpA_pPS | 4159 | -0.001256699 | [-0.03 0.03] | 0.935518667 | 0.99138381 |
| HAOX1 | Obesity_pPS | 4159 | 0.03903266 | [0.01 0.07] | 0.011944006 | 0.234493393 |
| HAOX1 | VAT_Neg_pPS | 4159 | 0.025474653 | [-0. 0.06] | 0.100949447 | 0.586731767 |
| HAOX1 | Bilirubin_pPS | 4159 | 0.035935561 | [0.01 0.07] | 0.020656782 | 0.312602357 |
| HAOX1 | Proinsulin_pPS | 4159 | -0.013172953 | [-0.04 0.02] | 0.396392074 | 0.812191725 |
| HAOX1 | Hyper_Insulin_pPS | 4159 | 0.015418277 | [-0.02 0.05] | 0.320878782 | 0.770109076 |
| HAOX1 | Beta_Cell_1_pPS | 4159 | -0.016238865 | [-0.05 0.01] | 0.295796005 | 0.75802876 |
| HAOX1 | Beta_Cell_2_pPS | 4159 | -0.033604379 | [-0.06 -0. ] | 0.030465362 | 0.369452038 |
| HAOX1 | Total_GRS | 4159 | 0.044936731 | [0.01 0.08] | 0.003798965 | 0.117810613 |
| HO_1 | Cholesterol_pPS | 4159 | -0.000690656 | [-0.03 0.03] | 0.964535416 | 0.99612031 |
| HO_1 | Lipodystrophy_1_pPS | 4159 | -0.026859328 | [-0.06 0. ] | 0.083727478 | 0.534925551 |
| HO_1 | Lipodystrophy_2_pPS | 4159 | 0.021611579 | [-0.01 0.05] | 0.164083497 | 0.664061293 |
| HO_1 | Liver-Lipid_pPS | 4159 | -0.013869939 | [-0.04 0.02] | 0.371879146 | 0.799802236 |
| HO_1 | ASAT_Pos_pPS | 4159 | -0.004090723 | [-0.03 0.03] | 0.792278647 | 0.957813871 |
| HO_1 | ALP_Neg_pPS | 4159 | -0.006389619 | [-0.04 0.02] | 0.680813144 | 0.92591406 |
| HO_1 | SHBG-LpA_pPS | 4159 | 0.013962625 | [-0.02 0.04] | 0.368691665 | 0.799230784 |
| HO_1 | Obesity_pPS | 4159 | -0.007648657 | [-0.04 0.02] | 0.622428925 | 0.911354818 |
| HO_1 | VAT_Neg_pPS | 4159 | -0.008384471 | [-0.04 0.02] | 0.589346546 | 0.904476019 |
| HO_1 | Bilirubin_pPS | 4159 | 0.020914574 | [-0.01 0.05] | 0.178115095 | 0.679661798 |
| HO_1 | Proinsulin_pPS | 4159 | 0.006728519 | [-0.02 0.04] | 0.664889756 | 0.924111434 |
| HO_1 | Hyper_Insulin_pPS | 4159 | -0.009748624 | [-0.04 0.02] | 0.530259579 | 0.877258512 |
| HO_1 | Beta_Cell_1_pPS | 4159 | -0.019046421 | [-0.05 0.01] | 0.220094169 | 0.719739225 |
| HO_1 | Beta_Cell_2_pPS | 4159 | -0.012857446 | [-0.04 0.02] | 0.407801802 | 0.813804543 |
| HO_1 | Total_GRS | 4159 | -0.003377526 | [-0.03 0.03] | 0.82786679 | 0.966003345 |
| IDUA | Cholesterol_pPS | 4159 | 0.042893148 | [0.01 0.07] | 0.005733641 | 0.152834705 |
| IDUA | Lipodystrophy_1_pPS | 4159 | 0.009864825 | [-0.02 0.04] | 0.525369001 | 0.873942847 |
| IDUA | Lipodystrophy_2_pPS | 4159 | 0.048266083 | [0.02 0.08] | 0.001876735 | 0.078291058 |
| IDUA | Liver-Lipid_pPS | 4159 | -0.001767002 | [-0.03 0.03] | 0.909431422 | 0.990056084 |
| IDUA | ASAT_Pos_pPS | 4159 | -0.011810516 | [-0.04 0.02] | 0.447038569 | 0.831350198 |
| IDUA | ALP_Neg_pPS | 4159 | 0.012220189 | [-0.02 0.04] | 0.43143518 | 0.827992564 |
| IDUA | SHBG-LpA_pPS | 4159 | 0.025864083 | [-0. 0.06] | 0.095842729 | 0.567652214 |
| IDUA | Obesity_pPS | 4159 | -0.003579233 | [-0.03 0.03] | 0.817762429 | 0.963461584 |
| IDUA | VAT_Neg_pPS | 4159 | 0.021872172 | [-0.01 0.05] | 0.159056892 | 0.661140091 |
| IDUA | Bilirubin_pPS | 4159 | -0.008563962 | [-0.04 0.02] | 0.581401276 | 0.903823705 |
| IDUA | Proinsulin_pPS | 4159 | -0.011064285 | [-0.04 0.02] | 0.476269116 | 0.84751951 |
| IDUA | Hyper_Insulin_pPS | 4159 | 0.011864807 | [-0.02 0.04] | 0.444952497 | 0.831350198 |
| IDUA | Beta_Cell_1_pPS | 4159 | -0.006418954 | [-0.04 0.02] | 0.679429053 | 0.925579558 |
| IDUA | Beta_Cell_2_pPS | 4159 | -0.0068704 | [-0.04 0.02] | 0.65826749 | 0.922711159 |
| IDUA | Total_GRS | 4159 | 0.021765174 | [-0.01 0.05] | 0.161106498 | 0.664061293 |
| MPO | Cholesterol_pPS | 4154 | -0.015954057 | [-0.05 0.01] | 0.304640633 | 0.76365449 |
| MPO | Lipodystrophy_1_pPS | 4154 | 0.012505397 | [-0.02 0.04] | 0.421041566 | 0.821254221 |
| MPO | Lipodystrophy_2_pPS | 4154 | 0.020843221 | [-0.01 0.05] | 0.179863135 | 0.680030484 |
| MPO | Liver-Lipid_pPS | 4154 | 0.003732319 | [-0.03 0.03] | 0.81022613 | 0.961816825 |
| MPO | ASAT_Pos_pPS | 4154 | 0.013702909 | [-0.02 0.04] | 0.377954127 | 0.803170077 |
| MPO | ALP_Neg_pPS | 4154 | 0.015167822 | [-0.02 0.05] | 0.329095289 | 0.775125841 |
| MPO | SHBG-LpA_pPS | 4154 | 0.004543074 | [-0.03 0.03] | 0.770058721 | 0.95301975 |
| MPO | Obesity_pPS | 4154 | -0.000403776 | [-0.03 0.03] | 0.979274458 | 0.996862516 |

**b) Partial Correlation**

| Proteomics | PPS | Sample_Size | r | CI95% | p | fdr_p_adjusted |
|---|---|---|---|---|---|---|
| CTSZ | Beta_Cell_2_pPS | 4225 | -0.014782833 | [-0.05 0.02] | 0.33816147 | 0.844649909 |
| EGFR | Cholesterol_pPS | 4225 | -0.004786149 | [-0.04 0.03] | 0.756494514 | 0.95462436 |
| EGFR | Lipodystrophy_1_pPS | 4225 | 0.025949615 | [-0. 0.06] | 0.092663933 | 0.638855661 |
| EGFR | Lipodystrophy_2_pPS | 4225 | 0.014416598 | [-0.02 0.04] | 0.350266367 | 0.847284666 |
| EGFR | Liver-Lipid_pPS | 4225 | -0.035499492 | [-0.07 -0.01] | 0.021410273 | 0.355824931 |
| EGFR | ASAT_Pos_pPS | 4225 | -0.004109903 | [-0.03 0.03] | 0.790028209 | 0.967638637 |
| EGFR | ALP_Neg_pPS | 4225 | -0.024163163 | [-0.05 0.01] | 0.11741549 | 0.681046037 |
| EGFR | SHBG-LpA_pPS | 4225 | 0.002404686 | [-0.03 0.03] | 0.876194369 | 0.97781424 |
| EGFR | Obesity_pPS | 4225 | -0.008560354 | [-0.04 0.02] | 0.579155023 | 0.909583681 |
| EGFR | VAT_Neg_pPS | 4225 | -0.02042445 | [-0.05 0.01] | 0.185702488 | 0.74165831 |
| EGFR | Bilirubin_pPS | 4225 | -0.007960932 | [-0.04 0.02] | 0.606006694 | 0.918301271 |
| EGFR | Proinsulin_pPS | 4225 | 0.009760024 | [-0.02 0.04] | 0.527159106 | 0.898959051 |
| EGFR | Hyper_Insulin_pPS | 4225 | -0.002535173 | [-0.03 0.03] | 0.869534842 | 0.977511154 |
| EGFR | Beta_Cell_1_pPS | 4225 | 0.010244205 | [-0.02 0.04] | 0.506869452 | 0.895803896 |
| EGFR | Beta_Cell_2_pPS | 4225 | 0.010313899 | [-0.02 0.04] | 0.503983054 | 0.895803896 |
| GLO1 | Cholesterol_pPS | 4159 | -0.017390847 | [-0.05 0.01] | 0.263604747 | 0.80722329 |
| GLO1 | Lipodystrophy_1_pPS | 4159 | 0.033339578 | [0. 0.06] | 0.032068524 | 0.428023414 |
| GLO1 | Lipodystrophy_2_pPS | 4159 | 0.00119861 | [-0.03 0.03] | 0.938589528 | 0.994163908 |
| GLO1 | Liver-Lipid_pPS | 4159 | -0.002754744 | [-0.03 0.03] | 0.859456325 | 0.97556306 |
| GLO1 | ASAT_Pos_pPS | 4159 | -0.013010447 | [-0.04 0.02] | 0.40298356 | 0.85241904 |
| GLO1 | ALP_Neg_pPS | 4159 | -0.015032812 | [-0.05 0.02] | 0.333886678 | 0.844649909 |
| GLO1 | SHBG-LpA_pPS | 4159 | -0.020181108 | [-0.05 0.01] | 0.194525212 | 0.755802332 |
| GLO1 | Obesity_pPS | 4159 | 0.013004176 | [-0.02 0.04] | 0.403210357 | 0.85241904 |
| GLO1 | VAT_Neg_pPS | 4159 | 0.016831741 | [-0.01 0.05] | 0.279267561 | 0.808820203 |
| GLO1 | Bilirubin_pPS | 4159 | 0.003062671 | [-0.03 0.03] | 0.843937902 | 0.972178273 |
| GLO1 | Proinsulin_pPS | 4159 | 0.030347486 | [-0. 0.06] | 0.051046697 | 0.537121778 |
| GLO1 | Hyper_Insulin_pPS | 4159 | -0.012722894 | [-0.04 0.02] | 0.41346037 | 0.854426619 |
| GLO1 | Beta_Cell_1_pPS | 4159 | -0.031134454 | [-0.06 -0. ] | 0.045316293 | 0.512674866 |
| GLO1 | Beta_Cell_2_pPS | 4159 | -0.019076223 | [-0.05 0.01] | 0.220097418 | 0.775610053 |
| HAOX1 | Cholesterol_pPS | 4159 | 0.022315273 | [-0.01 0.05] | 0.151420571 | 0.706166997 |
| HAOX1 | Lipodystrophy_1_pPS | 4159 | 0.022138289 | [-0.01 0.05] | 0.154692219 | 0.708464493 |
| HAOX1 | Lipodystrophy_2_pPS | 4159 | 0.047565849 | [0.02 0.08] | 0.00221996 | 0.105900329 |
| HAOX1 | Liver-Lipid_pPS | 4159 | -0.059377021 | [-0.09 -0.03] | 0.000133488 | 0.013754644 |
| HAOX1 | ASAT_Pos_pPS | 4159 | -0.000624699 | [-0.03 0.03] | 0.967970655 | 0.996924797 |
| HAOX1 | ALP_Neg_pPS | 4159 | 0.009621778 | [-0.02 0.04] | 0.53626497 | 0.898959051 |
| HAOX1 | SHBG-LpA_pPS | 4159 | -0.002488246 | [-0.03 0.03] | 0.872931035 | 0.97781424 |
| HAOX1 | Obesity_pPS | 4159 | 0.037755137 | [0.01 0.07] | 0.015197301 | 0.317679992 |
| HAOX1 | VAT_Neg_pPS | 4159 | 0.023184926 | [-0.01 0.05] | 0.136105456 | 0.694065456 |
| HAOX1 | Bilirubin_pPS | 4159 | 0.034747189 | [0. 0.07] | 0.025475782 | 0.392967754 |
| HAOX1 | Proinsulin_pPS | 4159 | -0.011646417 | [-0.04 0.02] | 0.454086786 | 0.872801276 |
| HAOX1 | Hyper_Insulin_pPS | 4159 | 0.00777258 | [-0.02 0.04] | 0.617354136 | 0.921525431 |
| HAOX1 | Beta_Cell_1_pPS | 4159 | -0.005598714 | [-0.04 0.02] | 0.718944406 | 0.946314812 |
| HAOX1 | Beta_Cell_2_pPS | 4159 | -0.030824048 | [-0.06 -0. ] | 0.04750831 | 0.520248282 |
| HO_1 | Cholesterol_pPS | 4159 | -0.000685146 | [-0.03 0.03] | 0.964873363 | 0.996924797 |
| HO_1 | Lipodystrophy_1_pPS | 4159 | -0.027369381 | [-0.06 0. ] | 0.078485102 | 0.602398521 |
| HO_1 | Lipodystrophy_2_pPS | 4159 | 0.024230768 | [-0.01 0.05] | 0.119303521 | 0.681046037 |
| HO_1 | Liver-Lipid_pPS | 4159 | -0.012785567 | [-0.04 0.02] | 0.4111633 | 0.854426619 |
| HO_1 | ASAT_Pos_pPS | 4159 | -0.000587796 | [-0.03 0.03] | 0.969861777 | 0.996924797 |
| HO_1 | ALP_Neg_pPS | 4159 | -0.007109518 | [-0.04 0.02] | 0.647683661 | 0.935848959 |
| HO_1 | SHBG-LpA_pPS | 4159 | 0.015888231 | [-0.01 0.05] | 0.307108431 | 0.830127301 |
| HO_1 | Obesity_pPS | 4159 | -0.007223521 | [-0.04 0.02] | 0.642425324 | 0.933976497 |
| HO_1 | VAT_Neg_pPS | 4159 | -0.008130504 | [-0.04 0.02] | 0.601244991 | 0.918301271 |
| HO_1 | Bilirubin_pPS | 4159 | 0.022141754 | [-0.01 0.05] | 0.154627666 | 0.708464493 |
| HO_1 | Proinsulin_pPS | 4159 | 0.014642782 | [-0.02 0.05] | 0.346581461 | 0.847284666 |
| HO_1 | Hyper_Insulin_pPS | 4159 | -0.005829461 | [-0.04 0.02] | 0.707882133 | 0.945970482 |
| HO_1 | Beta_Cell_1_pPS | 4159 | -0.017410303 | [-0.05 0.01] | 0.263070857 | 0.80722329 |
| HO_1 | Beta_Cell_2_pPS | 4159 | -0.010056914 | [-0.04 0.02] | 0.51799312 | 0.895803896 |
| IDUA | Cholesterol_pPS | 4159 | 0.042714896 | [0.01 0.07] | 0.006017466 | 0.186758931 |
| IDUA | Lipodystrophy_1_pPS | 4159 | 0.00642709 | [-0.02 0.04] | 0.679521138 | 0.943176659 |
| IDUA | Lipodystrophy_2_pPS | 4159 | 0.044920036 | [0.01 0.08] | 0.003867496 | 0.142323847 |
| IDUA | Liver-Lipid_pPS | 4159 | -0.004826105 | [-0.04 0.03] | 0.756402665 | 0.95462436 |
| IDUA | ASAT_Pos_pPS | 4159 | -0.01687956 | [-0.05 0.01] | 0.277903727 | 0.808820203 |
| IDUA | ALP_Neg_pPS | 4159 | 0.014781265 | [-0.02 0.05] | 0.342039327 | 0.847284666 |
| IDUA | SHBG-LpA_pPS | 4159 | 0.025232478 | [-0.01 0.06] | 0.104777701 | 0.663720785 |
| IDUA | Obesity_pPS | 4159 | -0.005782115 | [-0.04 0.02] | 0.710147035 | 0.945970482 |
| IDUA | VAT_Neg_pPS | 4159 | 0.016626576 | [-0.01 0.05] | 0.285170573 | 0.813509852 |
| IDUA | Bilirubin_pPS | 4159 | -0.008038507 | [-0.04 0.02] | 0.605367413 | 0.918301271 |
| IDUA | Proinsulin_pPS | 4159 | -0.012984035 | [-0.04 0.02] | 0.403939209 | 0.85241904 |
| IDUA | Hyper_Insulin_pPS | 4159 | 0.006785113 | [-0.02 0.04] | 0.662742419 | 0.937489141 |
| IDUA | Beta_Cell_1_pPS | 4159 | 0.000189295 | [-0.03 0.03] | 0.990292173 | 0.998803911 |
| IDUA | Beta_Cell_2_pPS | 4159 | -0.007091491 | [-0.04 0.02] | 0.648516781 | 0.935848959 |
| MPO | Cholesterol_pPS | 4154 | -0.01676992 | [-0.05 0.01] | 0.281328766 | 0.808820203 |
| MPO | Lipodystrophy_1_pPS | 4154 | 0.009246098 | [-0.02 0.04] | 0.552537512 | 0.899707098 |
| MPO | Lipodystrophy_2_pPS | 4154 | 0.01974073 | [-0.01 0.05] | 0.204717459 | 0.765145476 |
| MPO | Liver-Lipid_pPS | 4154 | 0.002129313 | [-0.03 0.03] | 0.891202616 | 0.981230818 |
| MPO | ASAT_Pos_pPS | 4154 | 0.009178402 | [-0.02 0.04] | 0.555450042 | 0.899898936 |
| MPO | ALP_Neg_pPS | 4154 | 0.010001501 | [-0.02 0.04] | 0.520554486 | 0.895803896 |
| MPO | SHBG-LpA_pPS | 4154 | 0.002730545 | [-0.03 0.03] | 0.86076172 | 0.97556306 |
| MPO | Obesity_pPS | 4154 | -0.003584198 | [-0.03 0.03] | 0.817903848 | 0.968668398 |
| MPO | VAT_Neg_pPS | 4154 | -0.002918353 | [-0.03 0.03] | 0.851292842 | 0.974635716 |
| MPO | Bilirubin_pPS | 4154 | 0.004481117 | [-0.03 0.03] | 0.773454901 | 0.962056893 |
| MPO | Proinsulin_pPS | 4154 | 0.027786132 | [-0. 0.06] | 0.074218218 | 0.588265012 |
| MPO | Hyper_Insulin_pPS | 4154 | -0.000812305 | [-0.03 0.03] | 0.958384682 | 0.996425456 |
| MPO | Beta_Cell_1_pPS | 4154 | 0.003319019 | [-0.03 0.03] | 0.831165374 | 0.971458352 |
| MPO | Beta_Cell_2_pPS | 4154 | 0.000676291 | [-0.03 0.03] | 0.965348016 | 0.996924797 |
| PON3 | Cholesterol_pPS | 4225 | -0.012411715 | [-0.04 0.02] | 0.421303057 | 0.860518922 |
| PON3 | Lipodystrophy_1_pPS | 4225 | -0.009950551 | [-0.04 0.02] | 0.519125887 | 0.895803896 |
| PON3 | Lipodystrophy_2_pPS | 4225 | -0.008019148 | [-0.04 0.02] | 0.603374576 | 0.918301271 |
| PON3 | Liver-Lipid_pPS | 4225 | -0.01735244 | [-0.05 0.01] | 0.260878763 | 0.807171938 |
| PON3 | ASAT_Pos_pPS | 4225 | -0.033757153 | [-0.06 -0. ] | 0.028692496 | 0.414433192 |
| PON3 | ALP_Neg_pPS | 4225 | -0.002647636 | [-0.03 0.03] | 0.863802581 | 0.97556306 |
| PON3 | SHBG-LpA_pPS | 4225 | -0.001189496 | [-0.03 0.03] | 0.938571644 | 0.994163908 |
| PON3 | Obesity_pPS | 4225 | -0.059428102 | [-0.09 -0.03] | 0.000116374 | 0.012490839 |
| PON3 | VAT_Neg_pPS | 4225 | -0.0448605 | [-0.08 -0.01] | 0.003638749 | 0.137844373 |
| PON3 | Bilirubin_pPS | 4225 | 0.010280663 | [-0.02 0.04] | 0.505358438 | 0.895803896 |
| PON3 | Proinsulin_pPS | 4225 | 0.009779694 | [-0.02 0.04] | 0.526326806 | 0.898776289 |
| PON3 | Hyper_Insulin_pPS | 4225 | -0.029560382 | [-0.06 0. ] | 0.055418572 | 0.557649379 |
| PON3 | Beta_Cell_1_pPS | 4225 | -0.005717875 | [-0.04 0.02] | 0.711044708 | 0.945970482 |
| PON3 | Beta_Cell_2_pPS | 4225 | 0.039197443 | [0.01 0.07] | 0.011069258 | 0.267922059 |
| TR | Cholesterol_pPS | 4225 | -0.009353764 | [-0.04 0.02] | 0.54449747 | 0.899241939 |
| TR | Lipodystrophy_1_pPS | 4225 | 0.022820334 | [-0.01 0.05] | 0.139226121 | 0.697681852 |
| TR | Lipodystrophy_2_pPS | 4225 | 0.007350322 | [-0.02 0.04] | 0.633917704 | 0.931124222 |
| TR | Liver-Lipid_pPS | 4225 | -0.009156389 | [-0.04 0.02] | 0.553022127 | 0.899898936 |
| TR | ASAT_Pos_pPS | 4225 | 0.009697529 | [-0.02 0.04] | 0.529807833 | 0.898959051 |
| TR | ALP_Neg_pPS | 4225 | -0.034226361 | [-0.06 -0. ] | 0.026547245 | 0.399916393 |
| TR | SHBG-LpA_pPS | 4225 | 0.004216485 | [-0.03 0.03] | 0.78471536 | 0.966727291 |
| TR | Obesity_pPS | 4225 | 0.029416166 | [-0. 0.06] | 0.05662031 | 0.558815902 |

**Table S7: Associations between pPSs and plasma proteins in SCAPIS**

**a) Spearman Correlation**

| Proteomics | PPS | Sample_Size | r | CI95% | p | fdr_p_adjusted |
|---|---|---|---|---|---|---|
| MPO | VAT_Neg_pPS | 4154 | 0.002187435 | [-0.03 0.03] | 0.888077374 | 0.979291846 |
| MPO | Bilirubin_pPS | 4154 | 0.005820104 | [-0.02 0.04] | 0.708060232 | 0.937738119 |
| MPO | Proinsulin_pPS | 4154 | 0.032510135 | [0. 0.06] | 0.036418763 | 0.386599178 |
| MPO | Hyper_Insulin_pPS | 4154 | 0.00235715 | [-0.03 0.03] | 0.879457652 | 0.979035738 |
| MPO | Beta_Cell_1_pPS | 4154 | 0.012823572 | [-0.02 0.04] | 0.409321034 | 0.814510493 |
| MPO | Beta_Cell_2_pPS | 4154 | 0.00724998 | [-0.02 0.04] | 0.640884699 | 0.916400472 |
| MPO | Total_GRS | 4154 | 0.016866505 | [-0.01 0.05] | 0.277810795 | 0.747422497 |
| PON3 | Cholesterol_pPS | 4225 | -0.016777222 | [-0.05 0.01] | 0.276275995 | 0.747422497 |
| PON3 | Lipodystrophy_1_pPS | 4225 | -0.024405794 | [-0.05 0.01] | 0.113219209 | 0.604978148 |
| PON3 | Lipodystrophy_2_pPS | 4225 | -0.019917086 | [-0.05 0.01] | 0.196178723 | 0.697416791 |
| PON3 | Liver-Lipid_pPS | 4225 | -0.019990543 | [-0.05 0.01] | 0.194533729 | 0.696574877 |
| PON3 | ASAT_Pos_pPS | 4225 | -0.04250328 | [-0.07 -0.01] | 0.005794037 | 0.152834705 |
| PON3 | ALP_Neg_pPS | 4225 | -0.003447891 | [-0.03 0.03] | 0.822968431 | 0.963461584 |
| PON3 | SHBG-LpA_pPS | 4225 | -0.004456841 | [-0.03 0.03] | 0.772429413 | 0.95301975 |
| PON3 | Obesity_pPS | 4225 | -0.062983299 | [-0.09 -0.03] | 4.30E-05 | 0.005339807 |
| PON3 | VAT_Neg_pPS | 4225 | -0.04747872 | [-0.08 -0.02] | 0.002052319 | 0.080919995 |
| PON3 | Bilirubin_pPS | 4225 | 0.00927584 | [-0.02 0.04] | 0.547236028 | 0.8909229 |
| PON3 | Proinsulin_pPS | 4225 | 0.005458822 | [-0.02 0.04] | 0.723176524 | 0.942134248 |
| PON3 | Hyper_Insulin_pPS | 4225 | -0.042410994 | [-0.07 -0.01] | 0.005901244 | 0.152834705 |
| PON3 | Beta_Cell_1_pPS | 4225 | -0.000714646 | [-0.03 0.03] | 0.963013575 | 0.99612031 |
| PON3 | Beta_Cell_2_pPS | 4225 | 0.035092966 | [0. 0.07] | 0.022736899 | 0.320646716 |
| PON3 | Total_GRS | 4225 | -0.050120665 | [-0.08 -0.02] | 0.001136748 | 0.051789895 |
| TR | Cholesterol_pPS | 4225 | -0.006483575 | [-0.04 0.02] | 0.673964673 | 0.925361755 |
| TR | Lipodystrophy_1_pPS | 4225 | 0.023549039 | [-0.01 0.05] | 0.126445367 | 0.623533826 |
| TR | Lipodystrophy_2_pPS | 4225 | 0.006426827 | [-0.02 0.04] | 0.676656038 | 0.925579558 |
| TR | Liver-Lipid_pPS | 4225 | -0.010260415 | [-0.04 0.02] | 0.505539549 | 0.867177846 |
| TR | ASAT_Pos_pPS | 4225 | 0.014108862 | [-0.02 0.04] | 0.359904767 | 0.7979754 |
| TR | ALP_Neg_pPS | 4225 | -0.033263144 | [-0.06 -0. ] | 0.030851062 | 0.369452038 |
| TR | SHBG-LpA_pPS | 4225 | 0.006343502 | [-0.02 0.04] | 0.68061533 | 0.92591406 |
| TR | Obesity_pPS | 4225 | 0.02946366 | [-0. 0.06] | 0.055842361 | 0.451781658 |
| TR | VAT_Neg_pPS | 4225 | -0.011344262 | [-0.04 0.02] | 0.461648961 | 0.83934244 |
| TR | Bilirubin_pPS | 4225 | -0.007364941 | [-0.04 0.02] | 0.632717504 | 0.914460114 |
| TR | Proinsulin_pPS | 4225 | 0.006279311 | [-0.02 0.04] | 0.683671461 | 0.926969309 |
| TR | Hyper_Insulin_pPS | 4225 | -0.012873244 | [-0.04 0.02] | 0.40351594 | 0.81230319 |
| TR | Beta_Cell_1_pPS | 4225 | 0.002727694 | [-0.03 0.03] | 0.859511559 | 0.977006629 |
| TR | Beta_Cell_2_pPS | 4225 | 0.007354627 | [-0.02 0.04] | 0.63319396 | 0.914460114 |
| TR | Total_GRS | 4225 | 0.000761023 | [-0.03 0.03] | 0.960615223 | 0.99612031 |
| TGM2 | Cholesterol_pPS | 4159 | -0.023146085 | [-0.05 0.01] | 0.136147091 | 0.629316669 |
| TGM2 | Lipodystrophy_1_pPS | 4159 | 0.020942052 | [-0.01 0.05] | 0.177545552 | 0.678995855 |
| TGM2 | Lipodystrophy_2_pPS | 4159 | -0.012987048 | [-0.04 0.02] | 0.40309149 | 0.81230319 |
| TGM2 | Liver-Lipid_pPS | 4159 | 7.64E-05 | [-0.03 0.03] | 0.996077072 | 0.999464614 |
| TGM2 | ASAT_Pos_pPS | 4159 | -0.023566842 | [-0.05 0.01] | 0.129167757 | 0.624571591 |
| TGM2 | ALP_Neg_pPS | 4159 | -0.002085872 | [-0.03 0.03] | 0.893178022 | 0.981916701 |
| TGM2 | SHBG-LpA_pPS | 4159 | 0.013186287 | [-0.02 0.04] | 0.395914139 | 0.812191725 |
| TGM2 | Obesity_pPS | 4159 | 0.002345959 | [-0.03 0.03] | 0.879953722 | 0.979035738 |
| TGM2 | VAT_Neg_pPS | 4159 | 0.009071715 | [-0.02 0.04] | 0.55920079 | 0.897093962 |
| TGM2 | Bilirubin_pPS | 4159 | 0.009743231 | [-0.02 0.04] | 0.530487122 | 0.877258512 |
| TGM2 | Proinsulin_pPS | 4159 | 0.002284764 | [-0.03 0.03] | 0.88306237 | 0.979291846 |
| TGM2 | Hyper_Insulin_pPS | 4159 | -0.002512199 | [-0.03 0.03] | 0.871518433 | 0.977006629 |
| TGM2 | Beta_Cell_1_pPS | 4159 | -0.012824039 | [-0.04 0.02] | 0.409021227 | 0.814501144 |
| TGM2 | Beta_Cell_2_pPS | 4159 | -0.020847104 | [-0.05 0.01] | 0.179519366 | 0.679661798 |
| TGM2 | Total_GRS | 4159 | -0.009405359 | [-0.04 0.02] | 0.544840409 | 0.888221812 |
| ADM | Cholesterol_pPS | 4159 | 0.00017687 | [-0.03 0.03] | 0.99091509 | 0.999402232 |
| ADM | Lipodystrophy_1_pPS | 4159 | -0.008579714 | [-0.04 0.02] | 0.580706387 | 0.903823705 |
| ADM | Lipodystrophy_2_pPS | 4159 | -0.018358657 | [-0.05 0.01] | 0.237209912 | 0.726631332 |
| ADM | Liver-Lipid_pPS | 4159 | 0.013985136 | [-0.02 0.04] | 0.367920073 | 0.799230784 |
| ADM | ASAT_Pos_pPS | 4159 | 0.005116885 | [-0.03 0.04] | 0.741841334 | 0.945527195 |
| ADM | ALP_Neg_pPS | 4159 | 0.030390812 | [-0. 0.06] | 0.050353895 | 0.440215663 |
| ADM | SHBG-LpA_pPS | 4159 | 0.017947562 | [-0.01 0.05] | 0.247878595 | 0.737212651 |
| ADM | Obesity_pPS | 4159 | 0.056769319 | [0.03 0.09] | 0.000254555 | 0.019515886 |
| ADM | VAT_Neg_pPS | 4159 | -0.031796748 | [-0.06 -0. ] | 0.040606851 | 0.407062052 |
| ADM | Bilirubin_pPS | 4159 | 0.001402385 | [-0.03 0.03] | 0.928062745 | 0.990056084 |
| ADM | Proinsulin_pPS | 4159 | 0.020780074 | [-0.01 0.05] | 0.180922614 | 0.680719889 |
| ADM | Hyper_Insulin_pPS | 4159 | -0.014526128 | [-0.04 0.02] | 0.349680416 | 0.793215848 |
| ADM | Beta_Cell_1_pPS | 4159 | -0.01637333 | [-0.05 0.01] | 0.291814488 | 0.753421877 |
| ADM | Beta_Cell_2_pPS | 4159 | -0.036569961 | [-0.07 -0.01] | 0.018518667 | 0.290406363 |
| ADM | Total_GRS | 4159 | 0.013726995 | [-0.02 0.04] | 0.376828342 | 0.803170077 |
| RAGE | Cholesterol_pPS | 4159 | -0.001344947 | [-0.03 0.03] | 0.931001577 | 0.990056084 |
| RAGE | Lipodystrophy_1_pPS | 4159 | 0.008439697 | [-0.02 0.04] | 0.586896601 | 0.903925569 |
| RAGE | Lipodystrophy_2_pPS | 4159 | 0.000306299 | [-0.03 0.03] | 0.984267671 | 0.998375146 |
| RAGE | Liver-Lipid_pPS | 4159 | -0.0008728 | [-0.03 0.03] | 0.955191311 | 0.99612031 |
| RAGE | ASAT_Pos_pPS | 4159 | -0.010699224 | [-0.04 0.02] | 0.490941522 | 0.859085266 |
| RAGE | ALP_Neg_pPS | 4159 | -0.007376616 | [-0.04 0.02] | 0.634860593 | 0.914460114 |
| RAGE | SHBG-LpA_pPS | 4159 | 0.021432718 | [-0.01 0.05] | 0.167602187 | 0.666544721 |
| RAGE | Obesity_pPS | 4159 | -0.013191838 | [-0.04 0.02] | 0.395715286 | 0.812191725 |
| RAGE | VAT_Neg_pPS | 4159 | -0.012112012 | [-0.04 0.02] | 0.435524382 | 0.828998134 |
| RAGE | Bilirubin_pPS | 4159 | 0.010075474 | [-0.02 0.04] | 0.516562562 | 0.870853148 |
| RAGE | Proinsulin_pPS | 4159 | -0.004304065 | [-0.03 0.03] | 0.781712934 | 0.955139045 |
| RAGE | Hyper_Insulin_pPS | 4159 | -0.016773822 | [-0.05 0.01] | 0.280169982 | 0.748127084 |
| RAGE | Beta_Cell_1_pPS | 4159 | -0.000400964 | [-0.03 0.03] | 0.979406357 | 0.996862516 |
| RAGE | Beta_Cell_2_pPS | 4159 | 0.022765674 | [-0.01 0.05] | 0.14270448 | 0.640622936 |
| RAGE | Total_GRS | 4159 | 0.004257715 | [-0.03 0.03] | 0.784005055 | 0.955343908 |
| AGRP | Cholesterol_pPS | 4159 | -0.038403564 | [-0.07 -0.01] | 0.013389083 | 0.242414912 |
| AGRP | Lipodystrophy_1_pPS | 4159 | 0.045966448 | [0.02 0.08] | 0.003068577 | 0.105623173 |
| AGRP | Lipodystrophy_2_pPS | 4159 | -0.009080178 | [-0.04 0.02] | 0.558834289 | 0.897093962 |
| AGRP | Liver-Lipid_pPS | 4159 | -0.023184817 | [-0.05 0.01] | 0.135492696 | 0.629316669 |
| AGRP | ASAT_Pos_pPS | 4159 | -0.000777633 | [-0.03 0.03] | 0.960072743 | 0.99612031 |
| AGRP | ALP_Neg_pPS | 4159 | 0.00054459 | [-0.03 0.03] | 0.972032276 | 0.996862516 |
| AGRP | SHBG-LpA_pPS | 4159 | 0.013628225 | [-0.02 0.04] | 0.380271711 | 0.805487278 |
| AGRP | Obesity_pPS | 4159 | 0.001395655 | [-0.03 0.03] | 0.928407014 | 0.990056084 |
| AGRP | VAT_Neg_pPS | 4159 | 0.006719779 | [-0.02 0.04] | 0.665298545 | 0.924111434 |
| AGRP | Bilirubin_pPS | 4159 | 0.022862543 | [-0.01 0.05] | 0.141012132 | 0.638159106 |
| AGRP | Proinsulin_pPS | 4159 | 0.021606189 | [-0.01 0.05] | 0.16418871 | 0.664061293 |
| AGRP | Hyper_Insulin_pPS | 4159 | -0.019624938 | [-0.05 0.01] | 0.206398105 | 0.709139138 |
| AGRP | Beta_Cell_1_pPS | 4159 | 0.02629014 | [-0. 0.06] | 0.090494736 | 0.55302075 |
| AGRP | Beta_Cell_2_pPS | 4159 | -0.014645908 | [-0.05 0.02] | 0.3457209 | 0.790391668 |
| AGRP | Total_GRS | 4159 | 0.006949775 | [-0.02 0.04] | 0.654574302 | 0.920338805 |
| ALCAM | Cholesterol_pPS | 4225 | 0.003286336 | [-0.03 0.03] | 0.831135423 | 0.967496317 |
| ALCAM | Lipodystrophy_1_pPS | 4225 | 0.032199537 | [0. 0.06] | 0.036625237 | 0.387301355 |
| ALCAM | Lipodystrophy_2_pPS | 4225 | 0.024756079 | [-0.01 0.05] | 0.108136055 | 0.599333113 |
| ALCAM | Liver-Lipid_pPS | 4225 | 0.013215454 | [-0.02 0.04] | 0.391131783 | 0.81106215 |
| ALCAM | ASAT_Pos_pPS | 4225 | 0.004742852 | [-0.03 0.03] | 0.758266735 | 0.950403646 |
| ALCAM | ALP_Neg_pPS | 4225 | -0.013274784 | [-0.04 0.02] | 0.389008425 | 0.810650617 |
| ALCAM | SHBG-LpA_pPS | 4225 | 0.020958715 | [-0.01 0.05] | 0.173789999 | 0.674627844 |
| ALCAM | Obesity_pPS | 4225 | 0.00387858 | [-0.03 0.03] | 0.801291779 | 0.958074488 |
| ALCAM | VAT_Neg_pPS | 4225 | -0.026200014 | [-0.06 0. ] | 0.089061737 | 0.546245321 |
| ALCAM | Bilirubin_pPS | 4225 | 0.00149562 | [-0.03 0.03] | 0.922687939 | 0.990056084 |

**b) Partial Correlation**

| Proteomics | PPS | Sample_Size | r | CI95% | p | fdr_p_adjusted |
|---|---|---|---|---|---|---|
| TR | VAT_Neg_pPS | 4225 | -0.012609366 | [-0.04 0.02] | 0.413945815 | 0.854426619 |
| TR | Bilirubin_pPS | 4225 | -0.008819677 | [-0.04 0.02] | 0.567714027 | 0.905613993 |
| TR | Proinsulin_pPS | 4225 | 0.007277777 | [-0.02 0.04] | 0.637269667 | 0.933760412 |
| TR | Hyper_Insulin_pPS | 4225 | -0.016361737 | [-0.05 0.01] | 0.289091348 | 0.81655641 |
| TR | Beta_Cell_1_pPS | 4225 | -0.000498944 | [-0.03 0.03] | 0.974212335 | 0.997444743 |
| TR | Beta_Cell_2_pPS | 4225 | 0.007347816 | [-0.02 0.04] | 0.634033371 | 0.931124222 |
| TGM2 | Cholesterol_pPS | 4159 | -0.022996916 | [-0.05 0.01] | 0.139310792 | 0.697681852 |
| TGM2 | Lipodystrophy_1_pPS | 4159 | 0.025141095 | [-0.01 0.06] | 0.106041742 | 0.665922758 |
| TGM2 | Lipodystrophy_2_pPS | 4159 | -0.012579292 | [-0.04 0.02] | 0.41875219 | 0.858842071 |
| TGM2 | Liver-Lipid_pPS | 4159 | 0.00224785 | [-0.03 0.03] | 0.885117765 | 0.979466202 |
| TGM2 | ASAT_Pos_pPS | 4159 | -0.026864063 | [-0.06 0. ] | 0.084159361 | 0.614725404 |
| TGM2 | ALP_Neg_pPS | 4159 | -0.003484737 | [-0.03 0.03] | 0.822766481 | 0.968668398 |
| TGM2 | SHBG-LpA_pPS | 4159 | 0.010601643 | [-0.02 0.04] | 0.495581569 | 0.895803896 |
| TGM2 | Obesity_pPS | 4159 | 0.004184139 | [-0.03 0.03] | 0.787972775 | 0.967638637 |
| TGM2 | VAT_Neg_pPS | 4159 | 0.011193934 | [-0.02 0.04] | 0.471813063 | 0.883720482 |
| TGM2 | Bilirubin_pPS | 4159 | 0.010527581 | [-0.02 0.04] | 0.498597981 | 0.895803896 |
| TGM2 | Proinsulin_pPS | 4159 | 0.008574502 | [-0.02 0.04] | 0.581530013 | 0.909583681 |
| TGM2 | Hyper_Insulin_pPS | 4159 | -0.00297158 | [-0.03 0.03] | 0.848522516 | 0.974061498 |
| TGM2 | Beta_Cell_1_pPS | 4159 | -0.011537823 | [-0.04 0.02] | 0.458306457 | 0.874172623 |
| TGM2 | Beta_Cell_2_pPS | 4159 | -0.019437836 | [-0.05 0.01] | 0.211475361 | 0.767305935 |
| ADM | Cholesterol_pPS | 4159 | -0.005690756 | [-0.04 0.02] | 0.714524598 | 0.945970482 |
| ADM | Lipodystrophy_1_pPS | 4159 | -0.010179315 | [-0.04 0.02] | 0.512911973 | 0.895803896 |
| ADM | Lipodystrophy_2_pPS | 4159 | -0.014437938 | [-0.04 0.02] | 0.3533702 | 0.847284666 |
| ADM | Liver-Lipid_pPS | 4159 | 0.014873345 | [-0.02 0.05] | 0.339040375 | 0.844649909 |
| ADM | ASAT_Pos_pPS | 4159 | 0.007149954 | [-0.02 0.04] | 0.645816548 | 0.935271696 |
| ADM | ALP_Neg_pPS | 4159 | 0.027087063 | [-0. 0.06] | 0.081615632 | 0.61116822 |
| ADM | SHBG-LpA_pPS | 4159 | 0.021640142 | [-0.01 0.05] | 0.164188802 | 0.718997225 |
| ADM | Obesity_pPS | 4159 | 0.055513217 | [0.03 0.09] | 0.000355672 | 0.028631564 |
| ADM | VAT_Neg_pPS | 4159 | -0.026257508 | [-0.06 0. ] | 0.091403897 | 0.63465347 |
| ADM | Bilirubin_pPS | 4159 | 0.002428195 | [-0.03 0.03] | 0.875972553 | 0.97781424 |
| ADM | Proinsulin_pPS | 4159 | 0.017545537 | [-0.01 0.05] | 0.259380387 | 0.80598779 |
| ADM | Hyper_Insulin_pPS | 4159 | -0.014561903 | [-0.05 0.02] | 0.349251886 | 0.847284666 |
| ADM | Beta_Cell_1_pPS | 4159 | -0.010616513 | [-0.04 0.02] | 0.494977133 | 0.895803896 |
| ADM | Beta_Cell_2_pPS | 4159 | -0.034819422 | [-0.07 -0. ] | 0.025171669 | 0.392967754 |
| RAGE | Cholesterol_pPS | 4159 | -0.00043429 | [-0.03 0.03] | 0.977730141 | 0.998803911 |
| RAGE | Lipodystrophy_1_pPS | 4159 | 0.011521955 | [-0.02 0.04] | 0.45892486 | 0.874172623 |
| RAGE | Lipodystrophy_2_pPS | 4159 | 0.002580932 | [-0.03 0.03] | 0.86824024 | 0.977511154 |
| RAGE | Liver-Lipid_pPS | 4159 | 5.82E-05 | [-0.03 0.03] | 0.997013399 | 0.999305801 |
| RAGE | ASAT_Pos_pPS | 4159 | -0.010702282 | [-0.04 0.02] | 0.491498419 | 0.895803896 |
| RAGE | ALP_Neg_pPS | 4159 | -0.005635978 | [-0.04 0.02] | 0.717153844 | 0.945970482 |
| RAGE | SHBG-LpA_pPS | 4159 | 0.02144494 | [-0.01 0.05] | 0.16802754 | 0.722132896 |
| RAGE | Obesity_pPS | 4159 | -0.011498933 | [-0.04 0.02] | 0.459822944 | 0.874172623 |
| RAGE | VAT_Neg_pPS | 4159 | -0.013977303 | [-0.04 0.02] | 0.36894067 | 0.847284666 |
| RAGE | Bilirubin_pPS | 4159 | 0.010460136 | [-0.02 0.04] | 0.501353341 | 0.895803896 |
| RAGE | Proinsulin_pPS | 4159 | -0.005483121 | [-0.04 0.03] | 0.724508443 | 0.946981479 |
| RAGE | Hyper_Insulin_pPS | 4159 | -0.015950622 | [-0.05 0.01] | 0.305212559 | 0.8267377 |
| RAGE | Beta_Cell_1_pPS | 4159 | -0.003490153 | [-0.03 0.03] | 0.822495599 | 0.968668398 |
| RAGE | Beta_Cell_2_pPS | 4159 | 0.024882381 | [-0.01 0.06] | 0.10968599 | 0.676681438 |
| AGRP | Cholesterol_pPS | 4159 | -0.036831396 | [-0.07 -0.01] | 0.017874592 | 0.33440804 |
| AGRP | Lipodystrophy_1_pPS | 4159 | 0.047838095 | [0.02 0.08] | 0.00209349 | 0.101751522 |
| AGRP | Lipodystrophy_2_pPS | 4159 | -0.007785971 | [-0.04 0.02] | 0.616748085 | 0.921525431 |
| AGRP | Liver-Lipid_pPS | 4159 | -0.021289976 | [-0.05 0.01] | 0.171122634 | 0.726214013 |
| AGRP | ASAT_Pos_pPS | 4159 | -0.008791659 | [-0.04 0.02] | 0.571998849 | 0.907308519 |
| AGRP | ALP_Neg_pPS | 4159 | -0.001825658 | [-0.03 0.03] | 0.90658474 | 0.988841471 |
| AGRP | SHBG-LpA_pPS | 4159 | 0.009568961 | [-0.02 0.04] | 0.538504679 | 0.899241939 |
| AGRP | Obesity_pPS | 4159 | 0.002642304 | [-0.03 0.03] | 0.865136767 | 0.97556306 |
| AGRP | VAT_Neg_pPS | 4159 | 0.004508715 | [-0.03 0.03] | 0.771963904 | 0.962056893 |
| AGRP | Bilirubin_pPS | 4159 | 0.020857953 | [-0.01 0.05] | 0.179976569 | 0.738879686 |
| AGRP | Proinsulin_pPS | 4159 | 0.017653324 | [-0.01 0.05] | 0.256464725 | 0.802737705 |
| AGRP | Hyper_Insulin_pPS | 4159 | -0.022775759 | [-0.05 0.01] | 0.143155267 | 0.697681852 |
| AGRP | Beta_Cell_1_pPS | 4159 | 0.022840965 | [-0.01 0.05] | 0.142013397 | 0.697681852 |
| AGRP | Beta_Cell_2_pPS | 4159 | -0.022668195 | [-0.05 0.01] | 0.145054275 | 0.697681852 |
| ALCAM | Cholesterol_pPS | 4225 | -3.69E-05 | [-0.03 0.03] | 0.998092445 | 0.999305801 |
| ALCAM | Lipodystrophy_1_pPS | 4225 | 0.028871066 | [-0. 0.06] | 0.061360057 | 0.558815902 |
| ALCAM | Lipodystrophy_2_pPS | 4225 | 0.024353865 | [-0.01 0.05] | 0.114548357 | 0.681046037 |
| ALCAM | Liver-Lipid_pPS | 4225 | 0.011763477 | [-0.02 0.04] | 0.445964595 | 0.869647841 |
| ALCAM | ASAT_Pos_pPS | 4225 | -0.003095678 | [-0.03 0.03] | 0.841039824 | 0.972178273 |
| ALCAM | ALP_Neg_pPS | 4225 | -0.01768699 | [-0.05 0.01] | 0.251796634 | 0.799790128 |
| ALCAM | SHBG-LpA_pPS | 4225 | 0.018692094 | [-0.01 0.05] | 0.225845052 | 0.784416549 |
| ALCAM | Obesity_pPS | 4225 | -0.000777305 | [-0.03 0.03] | 0.959835411 | 0.996425456 |
| ALCAM | VAT_Neg_pPS | 4225 | -0.033765156 | [-0.06 -0. ] | 0.028654689 | 0.414433192 |
| ALCAM | Bilirubin_pPS | 4225 | 0.00132063 | [-0.03 0.03] | 0.931815245 | 0.993113807 |
| ALCAM | Proinsulin_pPS | 4225 | 0.018141853 | [-0.01 0.05] | 0.239805449 | 0.790085251 |
| ALCAM | Hyper_Insulin_pPS | 4225 | 0.018141949 | [-0.01 0.05] | 0.239802955 | 0.790085251 |
| ALCAM | Beta_Cell_1_pPS | 4225 | -0.016862209 | [-0.05 0.01] | 0.274592107 | 0.808820203 |
| ALCAM | Beta_Cell_2_pPS | 4225 | 0.034400934 | [0. 0.06] | 0.025785205 | 0.395373145 |
| AXL | Cholesterol_pPS | 4225 | 0.024101984 | [-0.01 0.05] | 0.118347137 | 0.681046037 |
| AXL | Lipodystrophy_1_pPS | 4225 | 0.015897542 | [-0.01 0.05] | 0.302992679 | 0.826182442 |
| AXL | Lipodystrophy_2_pPS | 4225 | 0.025234122 | [-0.01 0.06] | 0.102021471 | 0.659955277 |
| AXL | Liver-Lipid_pPS | 4225 | 0.03285794 | [0. 0.06] | 0.03322292 | 0.436493301 |
| AXL | ASAT_Pos_pPS | 4225 | -0.023941698 | [-0.05 0.01] | 0.120815469 | 0.682614177 |
| AXL | ALP_Neg_pPS | 4225 | 0.029245379 | [-0. 0.06] | 0.058071424 | 0.558815902 |
| AXL | SHBG-LpA_pPS | 4225 | 0.004626511 | [-0.03 0.03] | 0.764372048 | 0.959095176 |
| AXL | Obesity_pPS | 4225 | 0.000569415 | [-0.03 0.03] | 0.970571662 | 0.996924797 |
| AXL | VAT_Neg_pPS | 4225 | -0.013432197 | [-0.04 0.02] | 0.384144124 | 0.85037326 |
| AXL | Bilirubin_pPS | 4225 | -0.010286859 | [-0.04 0.02] | 0.505101874 | 0.895803896 |
| AXL | Proinsulin_pPS | 4225 | 0.009654356 | [-0.02 0.04] | 0.531641545 | 0.898959051 |
| AXL | Hyper_Insulin_pPS | 4225 | -0.010143171 | [-0.04 0.02] | 0.511069131 | 0.895803896 |
| AXL | Beta_Cell_1_pPS | 4225 | 0.01028362 | [-0.02 0.04] | 0.50523599 | 0.895803896 |
| AXL | Beta_Cell_2_pPS | 4225 | 0.02358246 | [-0.01 0.05] | 0.126493749 | 0.694065456 |
| BMP_6 | Cholesterol_pPS | 4159 | -0.013911871 | [-0.04 0.02] | 0.371186489 | 0.847284666 |
| BMP_6 | Lipodystrophy_1_pPS | 4159 | -0.010347145 | [-0.04 0.02] | 0.505987438 | 0.895803896 |
| BMP_6 | Lipodystrophy_2_pPS | 4159 | 0.024326553 | [-0.01 0.05] | 0.117849728 | 0.681046037 |
| BMP_6 | Liver-Lipid_pPS | 4159 | 0.020732212 | [-0.01 0.05] | 0.182616378 | 0.738879686 |
| BMP_6 | ASAT_Pos_pPS | 4159 | 0.022824367 | [-0.01 0.05] | 0.142303401 | 0.697681852 |
| BMP_6 | ALP_Neg_pPS | 4159 | -0.003217259 | [-0.03 0.03] | 0.836169604 | 0.972099644 |
| BMP_6 | SHBG-LpA_pPS | 4159 | 0.002810491 | [-0.03 0.03] | 0.856642662 | 0.97556306 |
| BMP_6 | Obesity_pPS | 4159 | 0.007735297 | [-0.02 0.04] | 0.619042969 | 0.92152711 |
| BMP_6 | VAT_Neg_pPS | 4159 | 0.013808654 | [-0.02 0.04] | 0.374746465 | 0.849029811 |
| BMP_6 | Bilirubin_pPS | 4159 | -0.014388488 | [-0.04 0.02] | 0.355021553 | 0.847284666 |
| BMP_6 | Proinsulin_pPS | 4159 | 0.035356471 | [0. 0.07] | 0.02300753 | 0.369842052 |
| BMP_6 | Hyper_Insulin_pPS | 4159 | -0.007862852 | [-0.04 0.02] | 0.613273481 | 0.920963261 |
| BMP_6 | Beta_Cell_1_pPS | 4159 | -0.038669599 | [-0.07 -0.01] | 0.012901167 | 0.296726834 |
| BMP_6 | Beta_Cell_2_pPS | 4159 | 0.017100037 | [-0.01 0.05] | 0.271674257 | 0.808820203 |
| CASP_3 | Cholesterol_pPS | 4225 | -0.005223438 | [-0.04 0.03] | 0.735047451 | 0.948434512 |
| CASP_3 | Lipodystrophy_1_pPS | 4225 | 0.017564637 | [-0.01 0.05] | 0.25509228 | 0.801363065 |
| CASP_3 | Lipodystrophy_2_pPS | 4225 | 0.018954754 | [-0.01 0.05] | 0.219389476 | 0.775610053 |

**Table S7: Associations between pPSs and plasma proteins in SCAPIS**

**a) Spearman Correlation**

| Proteomics | PPS | Sample_Size | r | CI95% | p | fdr_p_adjusted |
|---|---|---|---|---|---|---|
| ALCAM | Proinsulin_pPS | 4225 | 0.018005899 | [-0.01 0.05] | 0.242617994 | 0.730235185 |
| ALCAM | Hyper_Insulin_pPS | 4225 | 0.021445547 | [-0.01 0.05] | 0.164005222 | 0.664061293 |
| ALCAM | Beta_Cell_1_pPS | 4225 | -0.006617913 | [-0.04 0.02] | 0.667610156 | 0.924640793 |
| ALCAM | Beta_Cell_2_pPS | 4225 | 0.032946008 | [0. 0.06] | 0.032484854 | 0.376562033 |
| ALCAM | Total_GRS | 4225 | 0.02883458 | [-0. 0.06] | 0.061287181 | 0.466331669 |
| AXL | Cholesterol_pPS | 4225 | 0.026991499 | [-0. 0.06] | 0.07981622 | 0.521702218 |
| AXL | Lipodystrophy_1_pPS | 4225 | 0.013112762 | [-0.02 0.04] | 0.394823628 | 0.812191725 |
| AXL | Lipodystrophy_2_pPS | 4225 | 0.02399901 | [-0.01 0.05] | 0.119356373 | 0.615837655 |
| AXL | Liver-Lipid_pPS | 4225 | 0.036384317 | [0.01 0.07] | 0.018191317 | 0.290017524 |
| AXL | ASAT_Pos_pPS | 4225 | -0.019976874 | [-0.05 0.01] | 0.19483906 | 0.696574877 |
| AXL | ALP_Neg_pPS | 4225 | 0.033245503 | [0. 0.06] | 0.030940057 | 0.369452038 |
| AXL | SHBG-LpA_pPS | 4225 | 0.004745902 | [-0.03 0.03] | 0.758116167 | 0.950403646 |
| AXL | Obesity_pPS | 4225 | 0.004811903 | [-0.03 0.04] | 0.754859388 | 0.950403646 |
| AXL | VAT_Neg_pPS | 4225 | -0.007666735 | [-0.04 0.02] | 0.618844097 | 0.909483337 |
| AXL | Bilirubin_pPS | 4225 | -0.009961703 | [-0.04 0.02] | 0.518010859 | 0.872306267 |
| AXL | Proinsulin_pPS | 4225 | 0.023904529 | [-0.01 0.05] | 0.120818434 | 0.615837655 |
| AXL | Hyper_Insulin_pPS | 4225 | -0.006335069 | [-0.04 0.02] | 0.681016501 | 0.92591406 |
| AXL | Beta_Cell_1_pPS | 4225 | 0.01820385 | [-0.01 0.05] | 0.237477238 | 0.726631332 |
| AXL | Beta_Cell_2_pPS | 4225 | 0.03007747 | [-0. 0.06] | 0.050924019 | 0.440215663 |
| AXL | Total_GRS | 4225 | 0.025306655 | [-0. 0.06] | 0.10051325 | 0.586504377 |
| BMP_6 | Cholesterol_pPS | 4159 | -0.011427528 | [-0.04 0.02] | 0.461911441 | 0.83934244 |
| BMP_6 | Lipodystrophy_1_pPS | 4159 | -0.006212788 | [-0.04 0.02] | 0.68917902 | 0.927417892 |
| BMP_6 | Lipodystrophy_2_pPS | 4159 | 0.029054269 | [-0. 0.06] | 0.061366621 | 0.466331669 |
| BMP_6 | Liver-Lipid_pPS | 4159 | 0.021555361 | [-0.01 0.05] | 0.165183396 | 0.664413675 |
| BMP_6 | ASAT_Pos_pPS | 4159 | 0.022243769 | [-0.01 0.05] | 0.15209177 | 0.649125224 |
| BMP_6 | ALP_Neg_pPS | 4159 | 0.003502937 | [-0.03 0.03] | 0.821580971 | 0.963461584 |
| BMP_6 | SHBG-LpA_pPS | 4159 | 0.005476291 | [-0.02 0.04] | 0.7244227 | 0.942134248 |
| BMP_6 | Obesity_pPS | 4159 | 0.010869307 | [-0.02 0.04] | 0.484075564 | 0.854586077 |
| BMP_6 | VAT_Neg_pPS | 4159 | 0.014886614 | [-0.02 0.05] | 0.337850808 | 0.782928826 |
| BMP_6 | Bilirubin_pPS | 4159 | -0.014686294 | [-0.05 0.02] | 0.344392351 | 0.790391668 |
| BMP_6 | Proinsulin_pPS | 4159 | 0.030338317 | [-0. 0.06] | 0.050752859 | 0.440215663 |
| BMP_6 | Hyper_Insulin_pPS | 4159 | -0.002915552 | [-0.03 0.03] | 0.851114027 | 0.97528582 |
| BMP_6 | Beta_Cell_1_pPS | 4159 | -0.023957182 | [-0.05 0.01] | 0.122944401 | 0.615837655 |
| BMP_6 | Beta_Cell_2_pPS | 4159 | 0.015345093 | [-0.02 0.05] | 0.323181355 | 0.773617121 |
| BMP_6 | Total_GRS | 4159 | 0.030096667 | [-0. 0.06] | 0.05262372 | 0.441027526 |
| CASP_3 | Cholesterol_pPS | 4225 | -0.005258056 | [-0.04 0.02] | 0.732961291 | 0.942134248 |
| CASP_3 | Lipodystrophy_1_pPS | 4225 | 0.020817403 | [-0.01 0.05] | 0.176710275 | 0.678995855 |
| CASP_3 | Lipodystrophy_2_pPS | 4225 | 0.020990945 | [-0.01 0.05] | 0.173129033 | 0.674627844 |
| CASP_3 | Liver-Lipid_pPS | 4225 | -0.020299243 | [-0.05 0.01] | 0.187731241 | 0.689088129 |
| CASP_3 | ASAT_Pos_pPS | 4225 | 0.02014277 | [-0.01 0.05] | 0.191157012 | 0.693468715 |
| CASP_3 | ALP_Neg_pPS | 4225 | -0.016749038 | [-0.05 0.01] | 0.277083686 | 0.747422497 |
| CASP_3 | SHBG-LpA_pPS | 4225 | 0.008615178 | [-0.02 0.04] | 0.576137706 | 0.903823705 |
| CASP_3 | Obesity_pPS | 4225 | -0.003687866 | [-0.03 0.03] | 0.810872626 | 0.962155826 |
| CASP_3 | VAT_Neg_pPS | 4225 | -0.000557368 | [-0.03 0.03] | 0.971149416 | 0.996862516 |
| CASP_3 | Bilirubin_pPS | 4225 | 0.003657687 | [-0.03 0.03] | 0.812391375 | 0.96231768 |
| CASP_3 | Proinsulin_pPS | 4225 | 0.024607883 | [-0.01 0.05] | 0.110264123 | 0.603827339 |
| CASP_3 | Hyper_Insulin_pPS | 4225 | 0.000261998 | [-0.03 0.03] | 0.986436105 | 0.998703927 |
| CASP_3 | Beta_Cell_1_pPS | 4225 | 0.000372838 | [-0.03 0.03] | 0.980698781 | 0.996862516 |
| CASP_3 | Beta_Cell_2_pPS | 4225 | 0.01132144 | [-0.02 0.04] | 0.46255067 | 0.83934244 |
| CASP_3 | Total_GRS | 4225 | 0.019462949 | [-0.01 0.05] | 0.206575314 | 0.709139138 |
| CCL15 | Cholesterol_pPS | 4225 | 0.01663268 | [-0.01 0.05] | 0.280435256 | 0.748127084 |
| CCL15 | Lipodystrophy_1_pPS | 4225 | 0.029428501 | [-0. 0.06] | 0.056135663 | 0.451781658 |
| CCL15 | Lipodystrophy_2_pPS | 4225 | 0.01145543 | [-0.02 0.04] | 0.457270707 | 0.835860883 |
| CCL15 | Liver-Lipid_pPS | 4225 | -0.009635931 | [-0.04 0.02] | 0.531791006 | 0.877412567 |
| CCL15 | ASAT_Pos_pPS | 4225 | 0.018443966 | [-0.01 0.05] | 0.231345285 | 0.726631332 |
| CCL15 | ALP_Neg_pPS | 4225 | -0.041102069 | [-0.07 -0.01] | 0.007626485 | 0.191355453 |
| CCL15 | SHBG-LpA_pPS | 4225 | -0.008186009 | [-0.04 0.02] | 0.59529013 | 0.907233991 |
| CCL15 | Obesity_pPS | 4225 | 0.025228481 | [-0. 0.06] | 0.101568745 | 0.586731767 |
| CCL15 | VAT_Neg_pPS | 4225 | -0.002232928 | [-0.03 0.03] | 0.884796753 | 0.979291846 |
| CCL15 | Bilirubin_pPS | 4225 | 0.010836618 | [-0.02 0.04] | 0.48193669 | 0.853751773 |
| CCL15 | Proinsulin_pPS | 4225 | 0.010408064 | [-0.02 0.04] | 0.49943402 | 0.863780476 |
| CCL15 | Hyper_Insulin_pPS | 4225 | -0.00949169 | [-0.04 0.02] | 0.537951182 | 0.88195499 |
| CCL15 | Beta_Cell_1_pPS | 4225 | 0.004124832 | [-0.03 0.03] | 0.78896509 | 0.957247257 |
| CCL15 | Beta_Cell_2_pPS | 4225 | -0.016207896 | [-0.05 0.01] | 0.292903731 | 0.753479147 |
| CCL15 | Total_GRS | 4225 | 0.007908747 | [-0.02 0.04] | 0.60781583 | 0.909483337 |
| CCL16 | Cholesterol_pPS | 4225 | 0.018009412 | [-0.01 0.05] | 0.242526087 | 0.730235185 |
| CCL16 | Lipodystrophy_1_pPS | 4225 | 0.02823816 | [-0. 0.06] | 0.066848164 | 0.477352596 |
| CCL16 | Lipodystrophy_2_pPS | 4225 | 0.023466935 | [-0.01 0.05] | 0.12777352 | 0.623575746 |
| CCL16 | Liver-Lipid_pPS | 4225 | -0.035190582 | [-0.07 -0. ] | 0.022361729 | 0.319784317 |
| CCL16 | ASAT_Pos_pPS | 4225 | 0.031181816 | [0. 0.06] | 0.042987732 | 0.419244309 |
| CCL16 | ALP_Neg_pPS | 4225 | 0.00241501 | [-0.03 0.03] | 0.875476325 | 0.978250438 |
| CCL16 | SHBG-LpA_pPS | 4225 | 0.016932959 | [-0.01 0.05] | 0.27184188 | 0.746520328 |
| CCL16 | Obesity_pPS | 4225 | 0.024219152 | [-0.01 0.05] | 0.116003499 | 0.611872879 |
| CCL16 | VAT_Neg_pPS | 4225 | -0.011738469 | [-0.04 0.02] | 0.446229327 | 0.831350198 |
| CCL16 | Bilirubin_pPS | 4225 | -0.002820646 | [-0.03 0.03] | 0.854776447 | 0.977006629 |
| CCL16 | Proinsulin_pPS | 4225 | -0.004678187 | [-0.03 0.03] | 0.761461948 | 0.950403646 |
| CCL16 | Hyper_Insulin_pPS | 4225 | 0.016591365 | [-0.01 0.05] | 0.281631899 | 0.748127084 |
| CCL16 | Beta_Cell_1_pPS | 4225 | -0.001315037 | [-0.03 0.03] | 0.931998522 | 0.990113904 |
| CCL16 | Beta_Cell_2_pPS | 4225 | -0.036696267 | [-0.07 -0.01] | 0.017220626 | 0.282910286 |
| CCL16 | Total_GRS | 4225 | 0.022238036 | [-0.01 0.05] | 0.148973084 | 0.648099837 |
| CCL17 | Cholesterol_pPS | 4158 | -0.017505116 | [-0.05 0.01] | 0.259788528 | 0.743022111 |
| CCL17 | Lipodystrophy_1_pPS | 4158 | 0.007854213 | [-0.02 0.04] | 0.613148949 | 0.909483337 |
| CCL17 | Lipodystrophy_2_pPS | 4158 | 0.026449491 | [-0. 0.06] | 0.088595723 | 0.546245321 |
| CCL17 | Liver-Lipid_pPS | 4158 | -0.022863559 | [-0.05 0.01] | 0.141042411 | 0.638159106 |
| CCL17 | ASAT_Pos_pPS | 4158 | 0.014259688 | [-0.02 0.04] | 0.358648628 | 0.7979754 |
| CCL17 | ALP_Neg_pPS | 4158 | 0.011743222 | [-0.02 0.04] | 0.449686657 | 0.831350198 |
| CCL17 | SHBG-LpA_pPS | 4158 | 0.003210693 | [-0.03 0.03] | 0.836265292 | 0.970026164 |
| CCL17 | Obesity_pPS | 4158 | -0.010952952 | [-0.04 0.02] | 0.480771082 | 0.852690632 |
| CCL17 | VAT_Neg_pPS | 4158 | -0.013516714 | [-0.04 0.02] | 0.384239758 | 0.806422804 |
| CCL17 | Bilirubin_pPS | 4158 | -0.006623482 | [-0.04 0.02] | 0.669846721 | 0.925160508 |
| CCL17 | Proinsulin_pPS | 4158 | 0.017707009 | [-0.01 0.05] | 0.254332137 | 0.739680398 |
| CCL17 | Hyper_Insulin_pPS | 4158 | -0.005588994 | [-0.04 0.02] | 0.719021762 | 0.941449667 |
| CCL17 | Beta_Cell_1_pPS | 4158 | -0.039364267 | [-0.07 -0.01] | 0.011249187 | 0.232377917 |
| CCL17 | Beta_Cell_2_pPS | 4158 | -0.012998891 | [-0.04 0.02] | 0.402719445 | 0.81230319 |
| CCL17 | Total_GRS | 4158 | 0.003794175 | [-0.03 0.03] | 0.807051127 | 0.960526567 |
| MCP_1 | Cholesterol_pPS | 4225 | -0.000255685 | [-0.03 0.03] | 0.986762902 | 0.998703927 |
| MCP_1 | Lipodystrophy_1_pPS | 4225 | 0.008663238 | [-0.02 0.04] | 0.574011151 | 0.903411176 |
| MCP_1 | Lipodystrophy_2_pPS | 4225 | -0.019011808 | [-0.05 0.01] | 0.217293587 | 0.716955357 |
| MCP_1 | Liver-Lipid_pPS | 4225 | -0.001605484 | [-0.03 0.03] | 0.917028635 | 0.990056084 |
| MCP_1 | ASAT_Pos_pPS | 4225 | 0.013116062 | [-0.02 0.04] | 0.394704659 | 0.812191725 |
| MCP_1 | ALP_Neg_pPS | 4225 | 0.024390315 | [-0.01 0.05] | 0.113448098 | 0.604978148 |
| MCP_1 | SHBG-LpA_pPS | 4225 | -0.020866565 | [-0.05 0.01] | 0.17569021 | 0.678995855 |
| MCP_1 | Obesity_pPS | 4225 | 0.032945836 | [0. 0.06] | 0.032485757 | 0.376562033 |
| MCP_1 | VAT_Neg_pPS | 4225 | -0.040852209 | [-0.07 -0.01] | 0.008003073 | 0.197592038 |
| MCP_1 | Bilirubin_pPS | 4225 | 0.018410065 | [-0.01 0.05] | 0.232204162 | 0.726631332 |
| MCP_1 | Proinsulin_pPS | 4225 | 0.005266546 | [-0.02 0.04] | 0.732546602 | 0.942134248 |
| MCP_1 | Hyper_Insulin_pPS | 4225 | -0.024752413 | [-0.05 0.01] | 0.108188307 | 0.599333113 |

**b) Partial Correlation**

| Proteomics | PPS | Sample_Size | r | CI95% | p | fdr_p_adjusted |
|---|---|---|---|---|---|---|
| CASP_3 | Liver-Lipid_pPS | 4225 | -0.0205663 | [-0.05 0.01] | 0.182665479 | 0.738879686 |
| CASP_3 | ASAT_Pos_pPS | 4225 | 0.015578286 | [-0.01 0.05] | 0.31280727 | 0.833834311 |
| CASP_3 | ALP_Neg_pPS | 4225 | -0.020820479 | [-0.05 0.01] | 0.177315822 | 0.736718642 |
| CASP_3 | SHBG-LpA_pPS | 4225 | 0.005347102 | [-0.02 0.04] | 0.729018708 | 0.946981479 |
| CASP_3 | Obesity_pPS | 4225 | -0.007332814 | [-0.04 0.02] | 0.634725984 | 0.931124222 |
| CASP_3 | VAT_Neg_pPS | 4225 | -0.00409709 | [-0.03 0.03] | 0.790667545 | 0.967638637 |
| CASP_3 | Bilirubin_pPS | 4225 | 0.002227107 | [-0.03 0.03] | 0.885271255 | 0.979466202 |
| CASP_3 | Proinsulin_pPS | 4225 | 0.028296075 | [-0. 0.06] | 0.066710942 | 0.563434057 |
| CASP_3 | Hyper_Insulin_pPS | 4225 | -0.002658268 | [-0.03 0.03] | 0.863261054 | 0.97556306 |
| CASP_3 | Beta_Cell_1_pPS | 4225 | -0.009747552 | [-0.04 0.02] | 0.527687154 | 0.898959051 |
| CASP_3 | Beta_Cell_2_pPS | 4225 | 0.009445729 | [-0.02 0.04] | 0.540547916 | 0.899241939 |
| CCL15 | Cholesterol_pPS | 4225 | 0.014585512 | [-0.02 0.04] | 0.344649632 | 0.847284666 |
| CCL15 | Lipodystrophy_1_pPS | 4225 | 0.02817061 | [-0. 0.06] | 0.067928167 | 0.568910429 |
| CCL15 | Lipodystrophy_2_pPS | 4225 | 0.009922136 | [-0.02 0.04] | 0.520319944 | 0.895803896 |
| CCL15 | Liver-Lipid_pPS | 4225 | -0.008380803 | [-0.04 0.02] | 0.587139452 | 0.912226314 |
| CCL15 | ASAT_Pos_pPS | 4225 | 0.012090356 | [-0.02 0.04] | 0.433427466 | 0.864060889 |
| CCL15 | ALP_Neg_pPS | 4225 | -0.043885173 | [-0.07 -0.01] | 0.004446609 | 0.15594322 |
| CCL15 | SHBG-LpA_pPS | 4225 | -0.011357878 | [-0.04 0.02] | 0.461804172 | 0.875999666 |
| CCL15 | Obesity_pPS | 4225 | 0.02190268 | [-0.01 0.05] | 0.155839671 | 0.708464493 |
| CCL15 | VAT_Neg_pPS | 4225 | -0.001441973 | [-0.03 0.03] | 0.925567733 | 0.990553585 |
| CCL15 | Bilirubin_pPS | 4225 | 0.009133804 | [-0.02 0.04] | 0.554001749 | 0.899898936 |
| CCL15 | Proinsulin_pPS | 4225 | 0.016533816 | [-0.01 0.05] | 0.284048947 | 0.812280908 |
| CCL15 | Hyper_Insulin_pPS | 4225 | -0.014000327 | [-0.04 0.02] | 0.364354293 | 0.847284666 |
| CCL15 | Beta_Cell_1_pPS | 4225 | 0.003072535 | [-0.03 0.03] | 0.842212503 | 0.972178273 |
| CCL15 | Beta_Cell_2_pPS | 4225 | -0.018942171 | [-0.05 0.01] | 0.219695695 | 0.775610053 |
| CCL16 | Cholesterol_pPS | 4225 | 0.016412627 | [-0.01 0.05] | 0.287593886 | 0.81609738 |
| CCL16 | Lipodystrophy_1_pPS | 4225 | 0.019042429 | [-0.01 0.05] | 0.217264409 | 0.771962922 |
| CCL16 | Lipodystrophy_2_pPS | 4225 | 0.021940983 | [-0.01 0.05] | 0.155117398 | 0.708464493 |
| CCL16 | Liver-Lipid_pPS | 4225 | -0.036561171 | [-0.07 -0.01] | 0.017811117 | 0.33440804 |
| CCL16 | ASAT_Pos_pPS | 4225 | 0.023410591 | [-0.01 0.05] | 0.129282849 | 0.694065456 |
| CCL16 | ALP_Neg_pPS | 4225 | 0.004268937 | [-0.03 0.03] | 0.782104435 | 0.964905159 |
| CCL16 | SHBG-LpA_pPS | 4225 | 0.016937688 | [-0.01 0.05] | 0.272449272 | 0.808820203 |
| CCL16 | Obesity_pPS | 4225 | 0.021899724 | [-0.01 0.05] | 0.155895507 | 0.708464493 |
| CCL16 | VAT_Neg_pPS | 4225 | -0.014674631 | [-0.04 0.02] | 0.341709519 | 0.847284666 |
| CCL16 | Bilirubin_pPS | 4225 | -0.004013719 | [-0.03 0.03] | 0.79483112 | 0.967638637 |
| CCL16 | Proinsulin_pPS | 4225 | -0.004540728 | [-0.03 0.03] | 0.768615221 | 0.960245786 |
| CCL16 | Hyper_Insulin_pPS | 4225 | 0.0119594 | [-0.02 0.04] | 0.438425505 | 0.865276785 |
| CCL16 | Beta_Cell_1_pPS | 4225 | 0.010981138 | [-0.02 0.04] | 0.476793852 | 0.886891048 |
| CCL16 | Beta_Cell_2_pPS | 4225 | -0.036321662 | [-0.07 -0.01] | 0.018573174 | 0.340663962 |
| CCL17 | Cholesterol_pPS | 4158 | -0.017784902 | [-0.05 0.01] | 0.252993896 | 0.799790128 |
| CCL17 | Lipodystrophy_1_pPS | 4158 | 0.006362711 | [-0.02 0.04] | 0.68259178 | 0.943830609 |
| CCL17 | Lipodystrophy_2_pPS | 4158 | 0.026112384 | [-0. 0.06] | 0.093249118 | 0.638855661 |
| CCL17 | Liver-Lipid_pPS | 4158 | -0.02217155 | [-0.05 0.01] | 0.154123169 | 0.708464493 |
| CCL17 | ASAT_Pos_pPS | 4158 | 0.012973722 | [-0.02 0.04] | 0.404369591 | 0.85241904 |
| CCL17 | ALP_Neg_pPS | 4158 | 0.007456301 | [-0.02 0.04] | 0.63178538 | 0.930939769 |
| CCL17 | SHBG-LpA_pPS | 4158 | 0.001927748 | [-0.03 0.03] | 0.901398824 | 0.985399384 |
| CCL17 | Obesity_pPS | 4158 | -0.012446839 | [-0.04 0.02] | 0.423724436 | 0.860815573 |
| CCL17 | VAT_Neg_pPS | 4158 | -0.01198121 | [-0.04 0.02] | 0.441271652 | 0.866107938 |
| CCL17 | Bilirubin_pPS | 4158 | -0.006138786 | [-0.04 0.02] | 0.693184498 | 0.945970482 |
| CCL17 | Proinsulin_pPS | 4158 | 0.030936171 | [0. 0.06] | 0.046733035 | 0.518671587 |
| CCL17 | Hyper_Insulin_pPS | 4158 | -0.006063752 | [-0.04 0.02] | 0.696747564 | 0.945970482 |
| CCL17 | Beta_Cell_1_pPS | 4158 | -0.041974662 | [-0.07 -0.01] | 0.006957795 | 0.206973464 |
| CCL17 | Beta_Cell_2_pPS | 4158 | -0.004993299 | [-0.04 0.03] | 0.74827392 | 0.954135344 |
| MCP_1 | Cholesterol_pPS | 4225 | -0.003769065 | [-0.03 0.03] | 0.807082498 | 0.967638637 |
| MCP_1 | Lipodystrophy_1_pPS | 4225 | 0.010206831 | [-0.02 0.04] | 0.508420846 | 0.895803896 |
| MCP_1 | Lipodystrophy_2_pPS | 4225 | -0.014196868 | [-0.04 0.02] | 0.357659138 | 0.847284666 |
| MCP_1 | Liver-Lipid_pPS | 4225 | -0.000774768 | [-0.03 0.03] | 0.95996641 | 0.996425456 |
| MCP_1 | ASAT_Pos_pPS | 4225 | 0.013221603 | [-0.02 0.04] | 0.391643772 | 0.85037326 |
| MCP_1 | ALP_Neg_pPS | 4225 | 0.023508359 | [-0.01 0.05] | 0.12769047 | 0.694065456 |
| MCP_1 | SHBG-LpA_pPS | 4225 | -0.017595725 | [-0.05 0.01] | 0.254252082 | 0.801363065 |
| MCP_1 | Obesity_pPS | 4225 | 0.03296796 | [0. 0.06] | 0.03263764 | 0.433374027 |
| MCP_1 | VAT_Neg_pPS | 4225 | -0.035407592 | [-0.07 -0.01] | 0.021749711 | 0.358391511 |
| MCP_1 | Bilirubin_pPS | 4225 | 0.017652284 | [-0.01 0.05] | 0.252728431 | 0.799790128 |
| MCP_1 | Proinsulin_pPS | 4225 | 2.88E-05 | [-0.03 0.03] | 0.998512928 | 0.999305801 |
| MCP_1 | Hyper_Insulin_pPS | 4225 | -0.024466726 | [-0.05 0.01] | 0.112877661 | 0.681046037 |
| MCP_1 | Beta_Cell_1_pPS | 4225 | -0.002666243 | [-0.03 0.03] | 0.862854865 | 0.97556306 |
| MCP_1 | Beta_Cell_2_pPS | 4225 | -0.001483152 | [-0.03 0.03] | 0.923448535 | 0.990503296 |
| CCL24 | Cholesterol_pPS | 4225 | -0.01374068 | [-0.04 0.02] | 0.37331836 | 0.847284666 |
| CCL24 | Lipodystrophy_1_pPS | 4225 | -9.91E-05 | [-0.03 0.03] | 0.994876122 | 0.998803911 |
| CCL24 | Lipodystrophy_2_pPS | 4225 | 0.005342403 | [-0.02 0.04] | 0.729247487 | 0.946981479 |
| CCL24 | Liver-Lipid_pPS | 4225 | 0.005727013 | [-0.02 0.04] | 0.710603719 | 0.945970482 |
| CCL24 | ASAT_Pos_pPS | 4225 | 0.011082466 | [-0.02 0.04] | 0.472736238 | 0.883721733 |
| CCL24 | ALP_Neg_pPS | 4225 | 0.022983512 | [-0.01 0.05] | 0.136420199 | 0.694065456 |
| CCL24 | SHBG-LpA_pPS | 4225 | 0.016641303 | [-0.01 0.05] | 0.28092965 | 0.808820203 |
| CCL24 | Obesity_pPS | 4225 | 0.00659622 | [-0.02 0.04] | 0.669116191 | 0.937489141 |
| CCL24 | VAT_Neg_pPS | 4225 | -0.010436306 | [-0.04 0.02] | 0.498934598 | 0.895803896 |
| CCL24 | Bilirubin_pPS | 4225 | -0.008585336 | [-0.04 0.02] | 0.578048167 | 0.909583681 |
| CCL24 | Proinsulin_pPS | 4225 | -0.00090794 | [-0.03 0.03] | 0.953092501 | 0.996425456 |
| CCL24 | Hyper_Insulin_pPS | 4225 | 0.00410542 | [-0.03 0.03] | 0.790251888 | 0.967638637 |
| CCL24 | Beta_Cell_1_pPS | 4225 | -0.000186083 | [-0.03 0.03] | 0.990380948 | 0.998803911 |
| CCL24 | Beta_Cell_2_pPS | 4225 | 0.009403388 | [-0.02 0.04] | 0.542364552 | 0.899241939 |
| CCL3 | Cholesterol_pPS | 4159 | 0.01772108 | [-0.01 0.05] | 0.25464359 | 0.801363065 |
| CCL3 | Lipodystrophy_1_pPS | 4159 | -0.000146512 | [-0.03 0.03] | 0.992486168 | 0.998803911 |
| CCL3 | Lipodystrophy_2_pPS | 4159 | 0.017323544 | [-0.01 0.05] | 0.265457448 | 0.80722329 |
| CCL3 | Liver-Lipid_pPS | 4159 | -0.011974834 | [-0.04 0.02] | 0.44145952 | 0.866107938 |
| CCL3 | ASAT_Pos_pPS | 4159 | -0.007987529 | [-0.04 0.02] | 0.607657181 | 0.918301271 |
| CCL3 | ALP_Neg_pPS | 4159 | 0.009332159 | [-0.02 0.04] | 0.548603644 | 0.899241939 |
| CCL3 | SHBG-LpA_pPS | 4159 | -0.001980378 | [-0.03 0.03] | 0.898708976 | 0.985399384 |
| CCL3 | Obesity_pPS | 4159 | 0.029653924 | [-0. 0.06] | 0.056588354 | 0.558815902 |
| CCL3 | VAT_Neg_pPS | 4159 | 0.006960174 | [-0.02 0.04] | 0.654598727 | 0.936605382 |
| CCL3 | Bilirubin_pPS | 4159 | -0.005832602 | [-0.04 0.02] | 0.707731947 | 0.945970482 |
| CCL3 | Proinsulin_pPS | 4159 | -0.000815592 | [-0.03 0.03] | 0.958191164 | 0.996425456 |
| CCL3 | Hyper_Insulin_pPS | 4159 | -0.004309518 | [-0.03 0.03] | 0.781777764 | 0.964905159 |
| CCL3 | Beta_Cell_1_pPS | 4159 | 0.01979021 | [-0.01 0.05] | 0.203311003 | 0.765145476 |
| CCL3 | Beta_Cell_2_pPS | 4159 | -0.043099756 | [-0.07 -0.01] | 0.005578155 | 0.177399096 |
| CD163 | Cholesterol_pPS | 4225 | 0.006556849 | [-0.02 0.04] | 0.670974861 | 0.938344866 |
| CD163 | Lipodystrophy_1_pPS | 4225 | 0.024878099 | [-0.01 0.06] | 0.106950048 | 0.668697388 |
| CD163 | Lipodystrophy_2_pPS | 4225 | 0.0143427 | [-0.02 0.04] | 0.352741768 | 0.847284666 |
| CD163 | Liver-Lipid_pPS | 4225 | 0.019496611 | [-0.01 0.05] | 0.206492612 | 0.765359665 |
| CD163 | ASAT_Pos_pPS | 4225 | 0.013064312 | [-0.02 0.04] | 0.397302707 | 0.85037326 |
| CD163 | ALP_Neg_pPS | 4225 | 0.007251527 | [-0.02 0.04] | 0.638484388 | 0.933760412 |
| CD163 | SHBG-LpA_pPS | 4225 | 0.013299874 | [-0.02 0.04] | 0.388846088 | 0.85037326 |
| CD163 | Obesity_pPS | 4225 | 0.037513577 | [0.01 0.07] | 0.015045179 | 0.317679992 |
| CD163 | VAT_Neg_pPS | 4225 | -0.011453232 | [-0.04 0.02] | 0.458052387 | 0.874172623 |
| CD163 | Bilirubin_pPS | 4225 | 0.026588017 | [-0. 0.06] | 0.08490804 | 0.614725404 |
| CD163 | Proinsulin_pPS | 4225 | 0.018251301 | [-0.01 0.05] | 0.236981222 | 0.78980606 |
| CD163 | Hyper_Insulin_pPS | 4225 | 0.015326788 | [-0.01 0.05] | 0.320684605 | 0.83927892 |

**Table S7: Associations between pPSs and plasma proteins in SCAPIS**

**a) Spearman Correlation**

| Proteomics | PPS | Sample_Size | r | CI95% | p | fdr_p_adjusted |
|---|---|---|---|---|---|---|
| MCP_1 | Beta_Cell_1_pPS | 4225 | -0.005340785 | [-0.04 0.02] | 0.728923935 | 0.942134248 |
| MCP_1 | Beta_Cell_2_pPS | 4225 | -0.004860929 | [-0.04 0.03] | 0.752443077 | 0.950019622 |
| MCP_1 | Total_GRS | 4225 | -0.002787011 | [-0.03 0.03] | 0.856489285 | 0.977006629 |
| CCL24 | Cholesterol_pPS | 4225 | -0.012590262 | [-0.04 0.02] | 0.413931903 | 0.817598568 |
| CCL24 | Lipodystrophy_1_pPS | 4225 | 0.00402054 | [-0.03 0.03] | 0.794179386 | 0.958013595 |
| CCL24 | Lipodystrophy_2_pPS | 4225 | 0.005581993 | [-0.02 0.04] | 0.71719568 | 0.941449667 |
| CCL24 | Liver-Lipid_pPS | 4225 | 0.007095916 | [-0.02 0.04] | 0.645194535 | 0.916568313 |
| CCL24 | ASAT_Pos_pPS | 4225 | 0.01223189 | [-0.02 0.04] | 0.427348535 | 0.82738773 |
| CCL24 | ALP_Neg_pPS | 4225 | 0.02330584 | [-0.01 0.05] | 0.130410966 | 0.626855379 |
| CCL24 | SHBG-LpA_pPS | 4225 | 0.015899156 | [-0.01 0.05] | 0.302196072 | 0.76319014 |
| CCL24 | Obesity_pPS | 4225 | 0.006564009 | [-0.02 0.04] | 0.670157115 | 0.925160508 |
| CCL24 | VAT_Neg_pPS | 4225 | -0.007109658 | [-0.04 0.02] | 0.64455476 | 0.916568313 |
| CCL24 | Bilirubin_pPS | 4225 | -0.008470148 | [-0.04 0.02] | 0.582577358 | 0.903823705 |
| CCL24 | Proinsulin_pPS | 4225 | 0.005810393 | [-0.02 0.04] | 0.706151222 | 0.937738119 |
| CCL24 | Hyper_Insulin_pPS | 4225 | 0.006035733 | [-0.02 0.04] | 0.695315039 | 0.930772879 |
| CCL24 | Beta_Cell_1_pPS | 4225 | 0.001431827 | [-0.03 0.03] | 0.9259759 | 0.990056084 |
| CCL24 | Beta_Cell_2_pPS | 4225 | 0.009363253 | [-0.02 0.04] | 0.543466403 | 0.886924725 |
| CCL24 | Total_GRS | 4225 | 0.009154351 | [-0.02 0.04] | 0.552496518 | 0.893839618 |
| CCL3 | Cholesterol_pPS | 4159 | 0.02009299 | [-0.01 0.05] | 0.195778426 | 0.697416791 |
| CCL3 | Lipodystrophy_1_pPS | 4159 | 0.00118158 | [-0.03 0.03] | 0.939365352 | 0.992211394 |
| CCL3 | Lipodystrophy_2_pPS | 4159 | 0.016241515 | [-0.01 0.05] | 0.295717204 | 0.75802876 |
| CCL3 | Liver-Lipid_pPS | 4159 | -0.010996497 | [-0.04 0.02] | 0.478975331 | 0.850689777 |
| CCL3 | ASAT_Pos_pPS | 4159 | -0.004866084 | [-0.04 0.03] | 0.754075851 | 0.950403646 |
| CCL3 | ALP_Neg_pPS | 4159 | 0.009167363 | [-0.02 0.04] | 0.555065276 | 0.896216632 |
| CCL3 | SHBG-LpA_pPS | 4159 | -0.002764319 | [-0.03 0.03] | 0.858753516 | 0.977006629 |
| CCL3 | Obesity_pPS | 4159 | 0.030682563 | [0. 0.06] | 0.048184145 | 0.438114317 |
| CCL3 | VAT_Neg_pPS | 4159 | 0.006520078 | [-0.02 0.04] | 0.674666145 | 0.925361755 |
| CCL3 | Bilirubin_pPS | 4159 | -0.005206622 | [-0.04 0.03] | 0.737479415 | 0.942470055 |
| CCL3 | Proinsulin_pPS | 4159 | 0.002761813 | [-0.03 0.03] | 0.858880244 | 0.977006629 |
| CCL3 | Hyper_Insulin_pPS | 4159 | -0.00209607 | [-0.03 0.03] | 0.892658892 | 0.981916701 |
| CCL3 | Beta_Cell_1_pPS | 4159 | 0.009109351 | [-0.02 0.04] | 0.557571751 | 0.896793725 |
| CCL3 | Beta_Cell_2_pPS | 4159 | -0.03839897 | [-0.07 -0.01] | 0.013400182 | 0.242414912 |
| CCL3 | Total_GRS | 4159 | 0.001483401 | [-0.03 0.03] | 0.923919199 | 0.990056084 |
| CD163 | Cholesterol_pPS | 4225 | 0.012174542 | [-0.02 0.04] | 0.429518786 | 0.827992564 |
| CD163 | Lipodystrophy_1_pPS | 4225 | 0.033218454 | [0. 0.06] | 0.031076938 | 0.369452038 |
| CD163 | Lipodystrophy_2_pPS | 4225 | 0.018647163 | [-0.01 0.05] | 0.226244699 | 0.724402982 |
| CD163 | Liver-Lipid_pPS | 4225 | 0.021466308 | [-0.01 0.05] | 0.163597371 | 0.664061293 |
| CD163 | ASAT_Pos_pPS | 4225 | 0.022079678 | [-0.01 0.05] | 0.151889588 | 0.649125224 |
| CD163 | ALP_Neg_pPS | 4225 | 0.012762905 | [-0.02 0.04] | 0.407558506 | 0.813804543 |
| CD163 | SHBG-LpA_pPS | 4225 | 0.015038061 | [-0.02 0.05] | 0.329138616 | 0.775125841 |
| CD163 | Obesity_pPS | 4225 | 0.043775633 | [0.01 0.07] | 0.004484675 | 0.131677681 |
| CD163 | VAT_Neg_pPS | 4225 | -0.007338757 | [-0.04 0.02] | 0.633927389 | 0.914460114 |
| CD163 | Bilirubin_pPS | 4225 | 0.02699475 | [-0. 0.06] | 0.079779919 | 0.521702218 |
| CD163 | Proinsulin_pPS | 4225 | 0.022880986 | [-0.01 0.05] | 0.137569216 | 0.629670042 |
| CD163 | Hyper_Insulin_pPS | 4225 | 0.023215807 | [-0.01 0.05] | 0.13190326 | 0.629316669 |
| CD163 | Beta_Cell_1_pPS | 4225 | -0.009197334 | [-0.04 0.02] | 0.550632489 | 0.89239323 |
| CD163 | Beta_Cell_2_pPS | 4225 | 0.00470258 | [-0.03 0.03] | 0.760256177 | 0.950403646 |
| CD163 | Total_GRS | 4225 | 0.03632768 | [0.01 0.07] | 0.018372599 | 0.290017524 |
| CD4 | Cholesterol_pPS | 4159 | 0.018542016 | [-0.01 0.05] | 0.232557512 | 0.726631332 |
| CD4 | Lipodystrophy_1_pPS | 4159 | -0.00428627 | [-0.03 0.03] | 0.782592733 | 0.955139045 |
| CD4 | Lipodystrophy_2_pPS | 4159 | 0.028511801 | [-0. 0.06] | 0.066372451 | 0.477051992 |
| CD4 | Liver-Lipid_pPS | 4159 | 0.011625601 | [-0.02 0.04] | 0.454185275 | 0.834640464 |
| CD4 | ASAT_Pos_pPS | 4159 | -0.00838413 | [-0.04 0.02] | 0.589361705 | 0.904476019 |
| CD4 | ALP_Neg_pPS | 4159 | -0.007948308 | [-0.04 0.02] | 0.608859167 | 0.909483337 |
| CD4 | SHBG-LpA_pPS | 4159 | 0.0397288 | [0.01 0.07] | 0.010507569 | 0.223701034 |
| CD4 | Obesity_pPS | 4159 | 0.020372793 | [-0.01 0.05] | 0.189624237 | 0.692278961 |
| CD4 | VAT_Neg_pPS | 4159 | 0.029514424 | [-0. 0.06] | 0.057369186 | 0.45329986 |
| CD4 | Bilirubin_pPS | 4159 | 0.007797989 | [-0.02 0.04] | 0.615649992 | 0.909483337 |
| CD4 | Proinsulin_pPS | 4159 | 0.026500794 | [-0. 0.06] | 0.087940659 | 0.544206768 |
| CD4 | Hyper_Insulin_pPS | 4159 | -0.029655491 | [-0.06 0. ] | 0.056187963 | 0.451781658 |
| CD4 | Beta_Cell_1_pPS | 4159 | 0.008528779 | [-0.02 0.04] | 0.582954696 | 0.903823705 |
| CD4 | Beta_Cell_2_pPS | 4159 | -0.037868166 | [-0.07 -0.01] | 0.014738423 | 0.252355767 |
| CD4 | Total_GRS | 4159 | 0.022605289 | [-0.01 0.05] | 0.145540691 | 0.641415816 |
| CD40_L | Cholesterol_pPS | 4159 | -0.026187335 | [-0.06 0. ] | 0.091762688 | 0.556626417 |
| CD40_L | Lipodystrophy_1_pPS | 4159 | -0.000144219 | [-0.03 0.03] | 0.992592165 | 0.999464614 |
| CD40_L | Lipodystrophy_2_pPS | 4159 | 0.018971473 | [-0.01 0.05] | 0.221915127 | 0.722207328 |
| CD40_L | Liver-Lipid_pPS | 4159 | -0.005112334 | [-0.04 0.03] | 0.742062796 | 0.945527195 |
| CD40_L | ASAT_Pos_pPS | 4159 | 0.014308245 | [-0.02 0.04] | 0.356956257 | 0.7979754 |
| CD40_L | ALP_Neg_pPS | 4159 | -0.011931356 | [-0.04 0.02] | 0.442403006 | 0.831350198 |
| CD40_L | SHBG-LpA_pPS | 4159 | -0.005427797 | [-0.04 0.03] | 0.726764883 | 0.942134248 |
| CD40_L | Obesity_pPS | 4159 | -0.008742316 | [-0.04 0.02] | 0.573556287 | 0.903411176 |
| CD40_L | VAT_Neg_pPS | 4159 | -0.003475362 | [-0.03 0.03] | 0.8229621 | 0.963461584 |
| CD40_L | Bilirubin_pPS | 4159 | -0.00501637 | [-0.04 0.03] | 0.746737029 | 0.947292085 |
| CD40_L | Proinsulin_pPS | 4159 | 0.004471444 | [-0.03 0.03] | 0.773451519 | 0.953322233 |
| CD40_L | Hyper_Insulin_pPS | 4159 | -0.024846875 | [-0.06 0.01] | 0.10963543 | 0.603827339 |
| CD40_L | Beta_Cell_1_pPS | 4159 | -0.011919001 | [-0.04 0.02] | 0.44287569 | 0.831350198 |
| CD40_L | Beta_Cell_2_pPS | 4159 | -0.007315366 | [-0.04 0.02] | 0.637674025 | 0.915577927 |
| CD40_L | Total_GRS | 4159 | -0.008109164 | [-0.04 0.02] | 0.601629513 | 0.907623171 |
| CD84 | Cholesterol_pPS | 4159 | 0.028820631 | [-0. 0.06] | 0.063482923 | 0.473548288 |
| CD84 | Lipodystrophy_1_pPS | 4159 | 0.006063145 | [-0.02 0.04] | 0.696288484 | 0.930772879 |
| CD84 | Lipodystrophy_2_pPS | 4159 | 0.029132333 | [-0. 0.06] | 0.060672671 | 0.463868622 |
| CD84 | Liver-Lipid_pPS | 4159 | 0.013573309 | [-0.02 0.04] | 0.382194566 | 0.806027092 |
| CD84 | ASAT_Pos_pPS | 4159 | 0.001016183 | [-0.03 0.03] | 0.947839899 | 0.993154165 |
| CD84 | ALP_Neg_pPS | 4159 | -0.006529741 | [-0.04 0.02] | 0.674211683 | 0.925361755 |
| CD84 | SHBG-LpA_pPS | 4159 | 0.010485003 | [-0.02 0.04] | 0.499663131 | 0.863780476 |
| CD84 | Obesity_pPS | 4159 | -0.015662801 | [-0.05 0.01] | 0.313263246 | 0.76365449 |
| CD84 | VAT_Neg_pPS | 4159 | -0.007218275 | [-0.04 0.02] | 0.642144388 | 0.916400472 |
| CD84 | Bilirubin_pPS | 4159 | 0.005782985 | [-0.02 0.04] | 0.709670305 | 0.938946716 |
| CD84 | Proinsulin_pPS | 4159 | 0.03502252 | [0. 0.07] | 0.024110468 | 0.331069119 |
| CD84 | Hyper_Insulin_pPS | 4159 | -0.015404329 | [-0.05 0.02] | 0.321316793 | 0.770490312 |
| CD84 | Beta_Cell_1_pPS | 4159 | -0.016620952 | [-0.05 0.01] | 0.284576955 | 0.749111537 |
| CD84 | Beta_Cell_2_pPS | 4159 | -0.012733402 | [-0.04 0.02] | 0.412340664 | 0.8164748 |
| CD84 | Total_GRS | 4159 | 0.007984088 | [-0.02 0.04] | 0.60724769 | 0.909483337 |
| CHI3L1 | Cholesterol_pPS | 4225 | 0.019537556 | [-0.01 0.05] | 0.204840424 | 0.70858232 |
| CHI3L1 | Lipodystrophy_1_pPS | 4225 | 0.027425042 | [-0. 0.06] | 0.075092175 | 0.507976478 |
| CHI3L1 | Lipodystrophy_2_pPS | 4225 | 0.00175196 | [-0.03 0.03] | 0.909489966 | 0.990056084 |
| CHI3L1 | Liver-Lipid_pPS | 4225 | -0.010142573 | [-0.04 0.02] | 0.510440518 | 0.870176547 |
| CHI3L1 | ASAT_Pos_pPS | 4225 | 0.028005219 | [-0. 0.06] | 0.069129975 | 0.48811907 |
| CHI3L1 | ALP_Neg_pPS | 4225 | -0.019313684 | [-0.05 0.01] | 0.21007826 | 0.709689105 |
| CHI3L1 | SHBG-LpA_pPS | 4225 | -0.001383774 | [-0.03 0.03] | 0.928453375 | 0.990056084 |
| CHI3L1 | Obesity_pPS | 4225 | 0.029531571 | [-0. 0.06] | 0.05527944 | 0.451394246 |
| CHI3L1 | VAT_Neg_pPS | 4225 | -0.010819776 | [-0.04 0.02] | 0.482617972 | 0.853862566 |
| CHI3L1 | Bilirubin_pPS | 4225 | -0.008146115 | [-0.04 0.02] | 0.59708513 | 0.907623171 |
| CHI3L1 | Proinsulin_pPS | 4225 | 0.019997862 | [-0.01 0.05] | 0.194370381 | 0.696574877 |
| CHI3L1 | Hyper_Insulin_pPS | 4225 | 0.010791211 | [-0.02 0.04] | 0.483774656 | 0.854586077 |
| CHI3L1 | Beta_Cell_1_pPS | 4225 | -0.004805547 | [-0.03 0.03] | 0.755172849 | 0.950403646 |
| CHI3L1 | Beta_Cell_2_pPS | 4225 | -0.024123857 | [-0.05 0.01] | 0.117445693 | 0.613244581 |

**b) Partial Correlation**

| Proteomics | PPS | Sample_Size | r | CI95% | p | fdr_p_adjusted |
|---|---|---|---|---|---|---|
| CD163 | Beta_Cell_1_pPS | 4225 | -0.01670678 | [-0.05 0.01] | 0.279040907 | 0.808820203 |
| CD163 | Beta_Cell_2_pPS | 4225 | 0.004028873 | [-0.03 0.03] | 0.794073882 | 0.967638637 |
| CD4 | Cholesterol_pPS | 4159 | 0.017709945 | [-0.01 0.05] | 0.254942256 | 0.801363065 |
| CD4 | Lipodystrophy_1_pPS | 4159 | -0.005227373 | [-0.04 0.03] | 0.736870266 | 0.949205935 |
| CD4 | Lipodystrophy_2_pPS | 4159 | 0.027864224 | [-0. 0.06] | 0.073234028 | 0.58769737 |
| CD4 | Liver-Lipid_pPS | 4159 | 0.012081402 | [-0.02 0.04] | 0.437405678 | 0.865276785 |
| CD4 | ASAT_Pos_pPS | 4159 | -0.009643282 | [-0.04 0.02] | 0.535354447 | 0.898959051 |
| CD4 | ALP_Neg_pPS | 4159 | -0.011140106 | [-0.04 0.02] | 0.473946907 | 0.88406027 |
| CD4 | SHBG-LpA_pPS | 4159 | 0.038048066 | [0.01 0.07] | 0.014425412 | 0.312663737 |
| CD4 | Obesity_pPS | 4159 | 0.018464759 | [-0.01 0.05] | 0.235245055 | 0.788664502 |
| CD4 | VAT_Neg_pPS | 4159 | 0.028720672 | [-0. 0.06] | 0.06482657 | 0.562394973 |
| CD4 | Bilirubin_pPS | 4159 | 0.007995319 | [-0.02 0.04] | 0.607307016 | 0.918301271 |
| CD4 | Proinsulin_pPS | 4159 | 0.026669558 | [-0. 0.06] | 0.086430085 | 0.615938909 |
| CD4 | Hyper_Insulin_pPS | 4159 | -0.033927392 | [-0.06 -0. ] | 0.029156039 | 0.414433192 |
| CD4 | Beta_Cell_1_pPS | 4159 | 0.009462662 | [-0.02 0.04] | 0.543026509 | 0.899241939 |
| CD4 | Beta_Cell_2_pPS | 4159 | -0.047355489 | [-0.08 -0.02] | 0.002322433 | 0.108774327 |
| CD40_L | Cholesterol_pPS | 4159 | -0.025857563 | [-0.06 0. ] | 0.096449051 | 0.645331829 |
| CD40_L | Lipodystrophy_1_pPS | 4159 | 0.000631734 | [-0.03 0.03] | 0.96761016 | 0.996924797 |
| CD40_L | Lipodystrophy_2_pPS | 4159 | 0.020107758 | [-0.01 0.05] | 0.196152225 | 0.758690888 |
| CD40_L | Liver-Lipid_pPS | 4159 | -0.003101019 | [-0.03 0.03] | 0.842009413 | 0.972178273 |
| CD40_L | ASAT_Pos_pPS | 4159 | 0.014567321 | [-0.02 0.05] | 0.349072595 | 0.847284666 |
| CD40_L | ALP_Neg_pPS | 4159 | -0.013153983 | [-0.04 0.02] | 0.397813994 | 0.85037326 |
| CD40_L | SHBG-LpA_pPS | 4159 | -0.006474287 | [-0.04 0.02] | 0.677299916 | 0.942803089 |
| CD40_L | Obesity_pPS | 4159 | -0.006212258 | [-0.04 0.02] | 0.689666568 | 0.944989936 |
| CD40_L | VAT_Neg_pPS | 4159 | -0.002634235 | [-0.03 0.03] | 0.86554465 | 0.97556306 |
| CD40_L | Bilirubin_pPS | 4159 | -0.005316361 | [-0.04 0.03] | 0.732561015 | 0.947803704 |
| CD40_L | Proinsulin_pPS | 4159 | 0.012346116 | [-0.02 0.04] | 0.427429274 | 0.864060889 |
| CD40_L | Hyper_Insulin_pPS | 4159 | -0.025387316 | [-0.06 0.01] | 0.102663247 | 0.660510098 |
| CD40_L | Beta_Cell_1_pPS | 4159 | -0.012889972 | [-0.04 0.02] | 0.407353492 | 0.853674654 |
| CD40_L | Beta_Cell_2_pPS | 4159 | -0.004552533 | [-0.04 0.03] | 0.769809911 | 0.96077051 |
| CD84 | Cholesterol_pPS | 4159 | 0.02870256 | [-0. 0.06] | 0.064995753 | 0.562394973 |
| CD84 | Lipodystrophy_1_pPS | 4159 | 0.007094846 | [-0.02 0.04] | 0.648361701 | 0.935848959 |
| CD84 | Lipodystrophy_2_pPS | 4159 | 0.028034362 | [-0. 0.06] | 0.07149637 | 0.578232608 |
| CD84 | Liver-Lipid_pPS | 4159 | 0.013587769 | [-0.02 0.04] | 0.382435349 | 0.85037326 |
| CD84 | ASAT_Pos_pPS | 4159 | -0.000284808 | [-0.03 0.03] | 0.985394312 | 0.998803911 |
| CD84 | ALP_Neg_pPS | 4159 | -0.01403011 | [-0.04 0.02] | 0.367134326 | 0.847284666 |
| CD84 | SHBG-LpA_pPS | 4159 | 0.008474438 | [-0.02 0.04] | 0.585946752 | 0.911172983 |
| CD84 | Obesity_pPS | 4159 | -0.022666647 | [-0.05 0.01] | 0.145081749 | 0.697681852 |
| CD84 | VAT_Neg_pPS | 4159 | -0.006429328 | [-0.04 0.02] | 0.679415741 | 0.943176659 |
| CD84 | Bilirubin_pPS | 4159 | 0.006368332 | [-0.02 0.04] | 0.682290302 | 0.943830609 |
| CD84 | Proinsulin_pPS | 4159 | 0.045667116 | [0.02 0.08] | 0.003315476 | 0.133452752 |
| CD84 | Hyper_Insulin_pPS | 4159 | -0.016938365 | [-0.05 0.01] | 0.27623278 | 0.808820203 |
| CD84 | Beta_Cell_1_pPS | 4159 | -0.023678083 | [-0.05 0.01] | 0.127968123 | 0.694065456 |
| CD84 | Beta_Cell_2_pPS | 4159 | -0.014055682 | [-0.04 0.02] | 0.366261591 | 0.847284666 |
| CHI3L1 | Cholesterol_pPS | 4225 | 0.017067508 | [-0.01 0.05] | 0.268790469 | 0.808113719 |
| CHI3L1 | Lipodystrophy_1_pPS | 4225 | 0.022514499 | [-0.01 0.05] | 0.144604499 | 0.697681852 |
| CHI3L1 | Lipodystrophy_2_pPS | 4225 | -0.001207773 | [-0.03 0.03] | 0.937629706 | 0.994163908 |
| CHI3L1 | Liver-Lipid_pPS | 4225 | -0.010345559 | [-0.04 0.02] | 0.502674717 | 0.895803896 |
| CHI3L1 | ASAT_Pos_pPS | 4225 | 0.022529558 | [-0.01 0.05] | 0.144335995 | 0.697681852 |
| CHI3L1 | ALP_Neg_pPS | 4225 | -0.024067743 | [-0.05 0.01] | 0.118871086 | 0.681046037 |
| CHI3L1 | SHBG-LpA_pPS | 4225 | -0.003925487 | [-0.03 0.03] | 0.799243782 | 0.967638637 |
| CHI3L1 | Obesity_pPS | 4225 | 0.023426757 | [-0.01 0.05] | 0.129018491 | 0.694065456 |
| CHI3L1 | VAT_Neg_pPS | 4225 | -0.009204958 | [-0.04 0.02] | 0.550918386 | 0.899241939 |
| CHI3L1 | Bilirubin_pPS | 4225 | -0.00943838 | [-0.04 0.02] | 0.540863013 | 0.899241939 |
| CHI3L1 | Proinsulin_pPS | 4225 | 0.025960566 | [-0. 0.06] | 0.092526257 | 0.638855661 |
| CHI3L1 | Hyper_Insulin_pPS | 4225 | 0.006717313 | [-0.02 0.04] | 0.663412193 | 0.937489141 |
| CHI3L1 | Beta_Cell_1_pPS | 4225 | -0.005371167 | [-0.04 0.02] | 0.727847479 | 0.946981479 |
| CHI3L1 | Beta_Cell_2_pPS | 4225 | -0.025555446 | [-0.06 0. ] | 0.097729999 | 0.64755296 |
| CXCL1 | Cholesterol_pPS | 4159 | -0.010147997 | [-0.04 0.02] | 0.514209605 | 0.895803896 |
| CXCL1 | Lipodystrophy_1_pPS | 4159 | -0.00497178 | [-0.04 0.03] | 0.749292923 | 0.954135344 |
| CXCL1 | Lipodystrophy_2_pPS | 4159 | 0.013120091 | [-0.02 0.04] | 0.399031032 | 0.85161884 |
| CXCL1 | Liver-Lipid_pPS | 4159 | -0.022428485 | [-0.05 0.01] | 0.149355585 | 0.704651991 |
| CXCL1 | ASAT_Pos_pPS | 4159 | -0.006455075 | [-0.04 0.02] | 0.67820372 | 0.943176659 |
| CXCL1 | ALP_Neg_pPS | 4159 | -0.016252337 | [-0.05 0.01] | 0.296154085 | 0.817900851 |
| CXCL1 | SHBG-LpA_pPS | 4159 | -0.000296054 | [-0.03 0.03] | 0.98481767 | 0.998803911 |
| CXCL1 | Obesity_pPS | 4159 | -0.024079345 | [-0.05 0.01] | 0.121630399 | 0.682614177 |
| CXCL1 | VAT_Neg_pPS | 4159 | -0.000627011 | [-0.03 0.03] | 0.967852201 | 0.996924797 |
| CXCL1 | Bilirubin_pPS | 4159 | 0.00400841 | [-0.03 0.03] | 0.796678134 | 0.967638637 |
| CXCL1 | Proinsulin_pPS | 4159 | 0.022008079 | [-0.01 0.05] | 0.157133291 | 0.710132208 |
| CXCL1 | Hyper_Insulin_pPS | 4159 | 0.002882782 | [-0.03 0.03] | 0.8529967 | 0.974979087 |
| CXCL1 | Beta_Cell_1_pPS | 4159 | -0.006664097 | [-0.04 0.02] | 0.668395449 | 0.937489141 |
| CXCL1 | Beta_Cell_2_pPS | 4159 | -0.003737046 | [-0.03 0.03] | 0.810170563 | 0.967638637 |
| CXCL16 | Cholesterol_pPS | 4159 | -0.005438259 | [-0.04 0.03] | 0.726671785 | 0.946981479 |
| CXCL16 | Lipodystrophy_1_pPS | 4159 | 0.02529657 | [-0.01 0.06] | 0.103898305 | 0.662566294 |
| CXCL16 | Lipodystrophy_2_pPS | 4159 | 0.00785786 | [-0.02 0.04] | 0.613498819 | 0.920963261 |
| CXCL16 | Liver-Lipid_pPS | 4159 | -0.001731177 | [-0.03 0.03] | 0.911398651 | 0.989461462 |
| CXCL16 | ASAT_Pos_pPS | 4159 | 0.009710499 | [-0.02 0.04] | 0.53251331 | 0.898959051 |
| CXCL16 | ALP_Neg_pPS | 4159 | 0.013089801 | [-0.02 0.04] | 0.400120594 | 0.85241904 |
| CXCL16 | SHBG-LpA_pPS | 4159 | 0.01866669 | [-0.01 0.05] | 0.230163189 | 0.786988264 |
| CXCL16 | Obesity_pPS | 4159 | 0.018248794 | [-0.01 0.05] | 0.240767439 | 0.790085251 |
| CXCL16 | VAT_Neg_pPS | 4159 | -0.037872441 | [-0.07 -0.01] | 0.014883938 | 0.317679992 |
| CXCL16 | Bilirubin_pPS | 4159 | -0.028290822 | [-0.06 0. ] | 0.06894103 | 0.571035672 |
| CXCL16 | Proinsulin_pPS | 4159 | -0.004871976 | [-0.04 0.03] | 0.754161662 | 0.95462436 |
| CXCL16 | Hyper_Insulin_pPS | 4159 | 0.026767456 | [-0. 0.06] | 0.085281078 | 0.614725404 |
| CXCL16 | Beta_Cell_1_pPS | 4159 | 0.012759396 | [-0.02 0.04] | 0.412121576 | 0.854426619 |
| CXCL16 | Beta_Cell_2_pPS | 4159 | 0.000408116 | [-0.03 0.03] | 0.979071979 | 0.998803911 |
| TF | Cholesterol_pPS | 4159 | -0.015801268 | [-0.05 0.01] | 0.30976395 | 0.830335001 |
| TF | Lipodystrophy_1_pPS | 4159 | 0.018672886 | [-0.01 0.05] | 0.230008495 | 0.786988264 |
| TF | Lipodystrophy_2_pPS | 4159 | 0.027506123 | [-0. 0.06] | 0.077004307 | 0.599284271 |
| TF | Liver-Lipid_pPS | 4159 | 0.03282801 | [0. 0.06] | 0.034802707 | 0.443820658 |
| TF | ASAT_Pos_pPS | 4159 | -0.003268365 | [-0.03 0.03] | 0.833604885 | 0.971458352 |
| TF | ALP_Neg_pPS | 4159 | -0.058032862 | [-0.09 -0.03] | 0.00018898 | 0.018030072 |
| TF | SHBG-LpA_pPS | 4159 | -0.000786225 | [-0.03 0.03] | 0.95969528 | 0.996425456 |
| TF | Obesity_pPS | 4159 | -0.010265012 | [-0.04 0.02] | 0.509370013 | 0.895803896 |
| TF | VAT_Neg_pPS | 4159 | -0.040491918 | [-0.07 -0.01] | 0.009220823 | 0.248521918 |
| TF | Bilirubin_pPS | 4159 | -0.014944551 | [-0.05 0.02] | 0.336732824 | 0.844649909 |
| TF | Proinsulin_pPS | 4159 | 0.021854975 | [-0.01 0.05] | 0.160040776 | 0.717834051 |
| TF | Hyper_Insulin_pPS | 4159 | -0.011129946 | [-0.04 0.02] | 0.474350255 | 0.884172399 |
| TF | Beta_Cell_1_pPS | 4159 | -0.024097961 | [-0.05 0.01] | 0.121342429 | 0.682614177 |
| TF | Beta_Cell_2_pPS | 4159 | 0.00964921 | [-0.02 0.04] | 0.535103555 | 0.898959051 |
| FAS | Cholesterol_pPS | 4224 | 0.01917111 | [-0.01 0.05] | 0.214226854 | 0.767522081 |
| FAS | Lipodystrophy_1_pPS | 4224 | 0.023200507 | [-0.01 0.05] | 0.132802846 | 0.694065456 |
| FAS | Lipodystrophy_2_pPS | 4224 | 0.032285459 | [0. 0.06] | 0.036437601 | 0.454495981 |
| FAS | Liver-Lipid_pPS | 4224 | 0.029509034 | [-0. 0.06] | 0.055873198 | 0.558815902 |
| FAS | ASAT_Pos_pPS | 4224 | 0.018746231 | [-0.01 0.05] | 0.224558736 | 0.781707168 |
| FAS | ALP_Neg_pPS | 4224 | -0.03331407 | [-0.06 -0. ] | 0.030873612 | 0.419027441 |
| FAS | SHBG-LpA_pPS | 4224 | 0.010598058 | [-0.02 0.04] | 0.492356438 | 0.895803896 |

**Table S7: Associations between pPSs and plasma proteins in SCAPIS**

**a) Spearman Correlation**

| Proteomics | PPS | Sample_Size | r | CI95% | p | fdr_p_adjusted |
|---|---|---|---|---|---|---|
| CHI3L1 | Total_GRS | 4225 | 0.029568121 | [-0. 0.06] | 0.054978439 | 0.450599046 |
| CXCL1 | Cholesterol_pPS | 4159 | -0.012239706 | [-0.04 0.02] | 0.43069979 | 0.827992564 |
| CXCL1 | Lipodystrophy_1_pPS | 4159 | -0.006498104 | [-0.04 0.02] | 0.675700007 | 0.925361755 |
| CXCL1 | Lipodystrophy_2_pPS | 4159 | 0.011756247 | [-0.02 0.04] | 0.449129342 | 0.831350198 |
| CXCL1 | Liver-Lipid_pPS | 4159 | -0.02311065 | [-0.05 0.01] | 0.136747911 | 0.629316669 |
| CXCL1 | ASAT_Pos_pPS | 4159 | -0.006991259 | [-0.04 0.02] | 0.652647494 | 0.919948884 |
| CXCL1 | ALP_Neg_pPS | 4159 | -0.013830401 | [-0.04 0.02] | 0.373244025 | 0.80093559 |
| CXCL1 | SHBG-LpA_pPS | 4159 | 0.001074651 | [-0.03 0.03] | 0.944843401 | 0.993154165 |
| CXCL1 | Obesity_pPS | 4159 | -0.023529918 | [-0.05 0.01] | 0.129768891 | 0.626157587 |
| CXCL1 | VAT_Neg_pPS | 4159 | 0.000540125 | [-0.03 0.03] | 0.972261484 | 0.996862516 |
| CXCL1 | Bilirubin_pPS | 4159 | 0.003993166 | [-0.03 0.03] | 0.79712303 | 0.958074488 |
| CXCL1 | Proinsulin_pPS | 4159 | 0.014428328 | [-0.02 0.04] | 0.352934555 | 0.796092727 |
| CXCL1 | Hyper_Insulin_pPS | 4159 | -0.000486568 | [-0.03 0.03] | 0.975011006 | 0.996862516 |
| CXCL1 | Beta_Cell_1_pPS | 4159 | -0.002988086 | [-0.03 0.03] | 0.847454905 | 0.97528582 |
| CXCL1 | Beta_Cell_2_pPS | 4159 | -0.00252482 | [-0.03 0.03] | 0.870878571 | 0.977006629 |
| CXCL1 | Total_GRS | 4159 | -0.007363045 | [-0.04 0.02] | 0.635483514 | 0.914460114 |
| CXCL16 | Cholesterol_pPS | 4159 | -0.002248392 | [-0.03 0.03] | 0.884910893 | 0.979291846 |
| CXCL16 | Lipodystrophy_1_pPS | 4159 | 0.032202904 | [0. 0.06] | 0.038108143 | 0.396899905 |
| CXCL16 | Lipodystrophy_2_pPS | 4159 | 0.007154788 | [-0.02 0.04] | 0.645074587 | 0.916568313 |
| CXCL16 | Liver-Lipid_pPS | 4159 | 0.000833173 | [-0.03 0.03] | 0.957223727 | 0.99612031 |
| CXCL16 | ASAT_Pos_pPS | 4159 | 0.017419656 | [-0.01 0.05] | 0.262064847 | 0.746520328 |
| CXCL16 | ALP_Neg_pPS | 4159 | 0.013400009 | [-0.02 0.04] | 0.388301473 | 0.810062031 |
| CXCL16 | SHBG-LpA_pPS | 4159 | 0.018328433 | [-0.01 0.05] | 0.237983043 | 0.726631332 |
| CXCL16 | Obesity_pPS | 4159 | 0.020328518 | [-0.01 0.05] | 0.190588434 | 0.693468715 |
| CXCL16 | VAT_Neg_pPS | 4159 | -0.031531869 | [-0.06 -0. ] | 0.042310116 | 0.414099011 |
| CXCL16 | Bilirubin_pPS | 4159 | -0.02760382 | [-0.06 0. ] | 0.075500537 | 0.508247516 |
| CXCL16 | Proinsulin_pPS | 4159 | 0.002696954 | [-0.03 0.03] | 0.862160763 | 0.977006629 |
| CXCL16 | Hyper_Insulin_pPS | 4159 | 0.030131121 | [-0. 0.06] | 0.052353501 | 0.441027526 |
| CXCL16 | Beta_Cell_1_pPS | 4159 | 0.008196687 | [-0.02 0.04] | 0.597712088 | 0.907623171 |
| CXCL16 | Beta_Cell_2_pPS | 4159 | 0.00075612 | [-0.03 0.03] | 0.961176449 | 0.99612031 |
| CXCL16 | Total_GRS | 4159 | 0.027789947 | [-0. 0.06] | 0.073550264 | 0.502472104 |
| TF | Cholesterol_pPS | 4159 | -0.012890285 | [-0.04 0.02] | 0.406605191 | 0.813210382 |
| TF | Lipodystrophy_1_pPS | 4159 | 0.015068824 | [-0.02 0.05] | 0.331970377 | 0.778452202 |
| TF | Lipodystrophy_2_pPS | 4159 | 0.024206182 | [-0.01 0.05] | 0.119098448 | 0.615837655 |
| TF | Liver-Lipid_pPS | 4159 | 0.03013046 | [-0. 0.06] | 0.052358674 | 0.441027526 |
| TF | ASAT_Pos_pPS | 4159 | 0.000318633 | [-0.03 0.03] | 0.983634229 | 0.998099438 |
| TF | ALP_Neg_pPS | 4159 | -0.053939024 | [-0.08 -0.02] | 0.000510844 | 0.032789046 |
| TF | SHBG-LpA_pPS | 4159 | 0.001418204 | [-0.03 0.03] | 0.927253509 | 0.990056084 |
| TF | Obesity_pPS | 4159 | -0.010071034 | [-0.04 0.02] | 0.516747372 | 0.870853148 |
| TF | VAT_Neg_pPS | 4159 | -0.03892727 | [-0.07 -0.01] | 0.012176009 | 0.236660449 |
| TF | Bilirubin_pPS | 4159 | -0.015879176 | [-0.05 0.01] | 0.306624401 | 0.76365449 |
| TF | Proinsulin_pPS | 4159 | 0.005610088 | [-0.02 0.04] | 0.717973909 | 0.941449667 |
| TF | Hyper_Insulin_pPS | 4159 | -0.011327046 | [-0.04 0.02] | 0.465858793 | 0.841554994 |
| TF | Beta_Cell_1_pPS | 4159 | -0.02154528 | [-0.05 0.01] | 0.16538123 | 0.664413675 |
| TF | Beta_Cell_2_pPS | 4159 | 0.004054177 | [-0.03 0.03] | 0.794092477 | 0.958013595 |
| TF | Total_GRS | 4159 | 0.017110606 | [-0.01 0.05] | 0.270625322 | 0.746520328 |
| FAS | Cholesterol_pPS | 4224 | 0.022394049 | [-0.01 0.05] | 0.146189634 | 0.642489472 |
| FAS | Lipodystrophy_1_pPS | 4224 | 0.029395596 | [-0. 0.06] | 0.056440622 | 0.451781658 |
| FAS | Lipodystrophy_2_pPS | 4224 | 0.03656613 | [0.01 0.07] | 0.017633362 | 0.286282817 |
| FAS | Liver-Lipid_pPS | 4224 | 0.029895274 | [-0. 0.06] | 0.052372553 | 0.441027526 |
| FAS | ASAT_Pos_pPS | 4224 | 0.02668341 | [-0. 0.06] | 0.083354508 | 0.534925551 |
| FAS | ALP_Neg_pPS | 4224 | -0.030024863 | [-0.06 0. ] | 0.051358494 | 0.440215663 |
| FAS | SHBG-LpA_pPS | 4224 | 0.01336901 | [-0.02 0.04] | 0.38570703 | 0.807658934 |
| FAS | Obesity_pPS | 4224 | 0.017061554 | [-0.01 0.05] | 0.268274289 | 0.746520328 |
| FAS | VAT_Neg_pPS | 4224 | -0.001976214 | [-0.03 0.03] | 0.897976312 | 0.984642786 |
| FAS | Bilirubin_pPS | 4224 | 0.002316474 | [-0.03 0.03] | 0.880532205 | 0.979035738 |
| FAS | Proinsulin_pPS | 4224 | 0.007522284 | [-0.02 0.04] | 0.625508998 | 0.913767992 |
| FAS | Hyper_Insulin_pPS | 4224 | 0.005167654 | [-0.03 0.04] | 0.737411603 | 0.942470055 |
| FAS | Beta_Cell_1_pPS | 4224 | -0.002876292 | [-0.03 0.03] | 0.851961635 | 0.97528582 |
| FAS | Beta_Cell_2_pPS | 4224 | -0.006499813 | [-0.04 0.02] | 0.673231876 | 0.925361755 |
| FAS | Total_GRS | 4224 | 0.043807478 | [0.01 0.07] | 0.004460377 | 0.131677681 |
| GDF_15 | Cholesterol_pPS | 4225 | 0.02833914 | [-0. 0.06] | 0.065878444 | 0.476590869 |
| GDF_15 | Lipodystrophy_1_pPS | 4225 | 0.023436974 | [-0.01 0.05] | 0.128260885 | 0.623575746 |
| GDF_15 | Lipodystrophy_2_pPS | 4225 | 0.023040243 | [-0.01 0.05] | 0.134851253 | 0.629316669 |
| GDF_15 | Liver-Lipid_pPS | 4225 | 0.00770672 | [-0.02 0.04] | 0.617016024 | 0.909483337 |
| GDF_15 | ASAT_Pos_pPS | 4225 | 0.005234736 | [-0.02 0.04] | 0.734100696 | 0.94220178 |
| GDF_15 | ALP_Neg_pPS | 4225 | -0.026395846 | [-0.06 0. ] | 0.086697495 | 0.54140702 |
| GDF_15 | SHBG-LpA_pPS | 4225 | 0.015167745 | [-0.02 0.05] | 0.324984333 | 0.775125841 |
| GDF_15 | Obesity_pPS | 4225 | 0.015017737 | [-0.02 0.05] | 0.329792781 | 0.77532204 |
| GDF_15 | VAT_Neg_pPS | 4225 | -0.016614721 | [-0.05 0.01] | 0.280954987 | 0.748127084 |
| GDF_15 | Bilirubin_pPS | 4225 | 0.013858762 | [-0.02 0.04] | 0.368484569 | 0.799230784 |
| GDF_15 | Proinsulin_pPS | 4225 | 0.005770846 | [-0.02 0.04] | 0.708059244 | 0.937738119 |
| GDF_15 | Hyper_Insulin_pPS | 4225 | 0.013748068 | [-0.02 0.04] | 0.372322278 | 0.799802236 |
| GDF_15 | Beta_Cell_1_pPS | 4225 | -0.008508029 | [-0.04 0.02] | 0.580892125 | 0.903823705 |
| GDF_15 | Beta_Cell_2_pPS | 4225 | -0.009859968 | [-0.04 0.02] | 0.522294317 | 0.872980599 |
| GDF_15 | Total_GRS | 4225 | 0.022314696 | [-0.01 0.05] | 0.14757666 | 0.645439059 |
| GDF_2 | Cholesterol_pPS | 4159 | -0.001438669 | [-0.03 0.03] | 0.92620672 | 0.990056084 |
| GDF_2 | Lipodystrophy_1_pPS | 4159 | 0.012707464 | [-0.02 0.04] | 0.41329352 | 0.817113262 |
| GDF_2 | Lipodystrophy_2_pPS | 4159 | -0.020851471 | [-0.05 0.01] | 0.179428237 | 0.679661798 |
| GDF_2 | Liver-Lipid_pPS | 4159 | -0.043002594 | [-0.07 -0.01] | 0.005610912 | 0.15182468 |
| GDF_2 | ASAT_Pos_pPS | 4159 | -0.02045926 | [-0.05 0.01] | 0.187751548 | 0.689088129 |
| GDF_2 | ALP_Neg_pPS | 4159 | -0.004323077 | [-0.03 0.03] | 0.78077329 | 0.955139045 |
| GDF_2 | SHBG-LpA_pPS | 4159 | 0.023572713 | [-0.01 0.05] | 0.129072372 | 0.624571591 |
| GDF_2 | Obesity_pPS | 4159 | -0.037840162 | [-0.07 -0.01] | 0.014812186 | 0.252355767 |
| GDF_2 | VAT_Neg_pPS | 4159 | -0.005834808 | [-0.04 0.02] | 0.70718808 | 0.937738119 |
| GDF_2 | Bilirubin_pPS | 4159 | 0.010279925 | [-0.02 0.04] | 0.508088917 | 0.868660083 |
| GDF_2 | Proinsulin_pPS | 4159 | -0.006608068 | [-0.04 0.02] | 0.670532365 | 0.925160508 |
| GDF_2 | Hyper_Insulin_pPS | 4159 | 0.004926982 | [-0.03 0.04] | 0.751099343 | 0.948757065 |
| GDF_2 | Beta_Cell_1_pPS | 4159 | 0.039107822 | [0.01 0.07] | 0.011780948 | 0.23392385 |
| GDF_2 | Beta_Cell_2_pPS | 4159 | 0.011624867 | [-0.02 0.04] | 0.45421376 | 0.834640464 |
| GDF_2 | Total_GRS | 4159 | -0.0166029 | [-0.05 0.01] | 0.285100439 | 0.749406868 |
| GH | Cholesterol_pPS | 4159 | -0.013744771 | [-0.04 0.02] | 0.376210685 | 0.803170077 |
| GH | Lipodystrophy_1_pPS | 4159 | -0.013331699 | [-0.04 0.02] | 0.390724911 | 0.81106215 |
| GH | Lipodystrophy_2_pPS | 4159 | -1.04E-05 | [-0.03 0.03] | 0.999464614 | 0.999464614 |
| GH | Liver-Lipid_pPS | 4159 | -0.008928768 | [-0.04 0.02] | 0.565409119 | 0.899863296 |
| GH | ASAT_Pos_pPS | 4159 | -0.003839638 | [-0.03 0.03] | 0.804762494 | 0.960193229 |
| GH | ALP_Neg_pPS | 4159 | -0.021749845 | [-0.05 0.01] | 0.161401766 | 0.664061293 |
| GH | SHBG-LpA_pPS | 4159 | 0.006872499 | [-0.02 0.04] | 0.65816972 | 0.922711159 |
| GH | Obesity_pPS | 4159 | -0.027023224 | [-0.06 0. ] | 0.081856703 | 0.528738199 |
| GH | VAT_Neg_pPS | 4159 | 0.008366311 | [-0.02 0.04] | 0.590153204 | 0.904476019 |
| GH | Bilirubin_pPS | 4159 | 0.003333121 | [-0.03 0.03] | 0.830095106 | 0.966693035 |
| GH | Proinsulin_pPS | 4159 | 0.00088384 | [-0.03 0.03] | 0.95462513 | 0.99612031 |
| GH | Hyper_Insulin_pPS | 4159 | 0.00060039 | [-0.03 0.03] | 0.969168026 | 0.996862516 |
| GH | Beta_Cell_1_pPS | 4159 | -0.004026879 | [-0.03 0.03] | 0.79544806 | 0.958074488 |
| GH | Beta_Cell_2_pPS | 4159 | -0.023639385 | [-0.05 0.01] | 0.127993031 | 0.623575746 |
| GH | Total_GRS | 4159 | -0.030273933 | [-0.06 0. ] | 0.051245796 | 0.440215663 |
| GRN | Cholesterol_pPS | 4225 | 0.015064211 | [-0.02 0.05] | 0.328298182 | 0.775125841 |

**b) Partial Correlation**

| Proteomics | PPS | Sample_Size | r | CI95% | p | fdr_p_adjusted |
|---|---|---|---|---|---|---|
| FAS | Obesity_pPS | 4224 | 0.012619544 | [-0.02 0.04] | 0.413624688 | 0.854426619 |
| FAS | VAT_Neg_pPS | 4224 | -0.006429492 | [-0.04 0.02] | 0.677037323 | 0.942803089 |
| FAS | Bilirubin_pPS | 4224 | 0.001814692 | [-0.03 0.03] | 0.90641901 | 0.988841471 |
| FAS | Proinsulin_pPS | 4224 | 0.011607593 | [-0.02 0.04] | 0.452069219 | 0.870351501 |
| FAS | Hyper_Insulin_pPS | 4224 | -0.002509091 | [-0.03 0.03] | 0.870880473 | 0.977511154 |
| FAS | Beta_Cell_1_pPS | 4224 | -0.004618533 | [-0.03 0.03] | 0.764793553 | 0.959156861 |
| FAS | Beta_Cell_2_pPS | 4224 | -0.007927185 | [-0.04 0.02] | 0.607577629 | 0.918301271 |
| GDF_15 | Cholesterol_pPS | 4225 | 0.025389436 | [-0. 0.06] | 0.099928799 | 0.651687563 |
| GDF_15 | Lipodystrophy_1_pPS | 4225 | 0.019992232 | [-0.01 0.05] | 0.195186238 | 0.757228538 |
| GDF_15 | Lipodystrophy_2_pPS | 4225 | 0.022096465 | [-0.01 0.05] | 0.152211611 | 0.70648128 |
| GDF_15 | Liver-Lipid_pPS | 4225 | 0.007490991 | [-0.02 0.04] | 0.627439456 | 0.928365329 |
| GDF_15 | ASAT_Pos_pPS | 4225 | -0.001503863 | [-0.03 0.03] | 0.92238295 | 0.990503296 |
| GDF_15 | ALP_Neg_pPS | 4225 | -0.028793016 | [-0.06 0. ] | 0.062064863 | 0.558815902 |
| GDF_15 | SHBG-LpA_pPS | 4225 | 0.013811846 | [-0.02 0.04] | 0.370847951 | 0.847284666 |
| GDF_15 | Obesity_pPS | 4225 | 0.010467707 | [-0.02 0.04] | 0.497643895 | 0.895803896 |
| GDF_15 | VAT_Neg_pPS | 4225 | -0.019602567 | [-0.05 0.01] | 0.204036403 | 0.765145476 |
| GDF_15 | Bilirubin_pPS | 4225 | 0.014072774 | [-0.02 0.04] | 0.361877329 | 0.847284666 |
| GDF_15 | Proinsulin_pPS | 4225 | 0.011265928 | [-0.02 0.04] | 0.465438224 | 0.879924083 |
| GDF_15 | Hyper_Insulin_pPS | 4225 | 0.010747422 | [-0.02 0.04] | 0.486225112 | 0.895803896 |
| GDF_15 | Beta_Cell_1_pPS | 4225 | -0.007149895 | [-0.04 0.02] | 0.643196547 | 0.933976497 |
| GDF_15 | Beta_Cell_2_pPS | 4225 | -0.009309185 | [-0.04 0.02] | 0.546417144 | 0.899241939 |
| GDF_2 | Cholesterol_pPS | 4159 | 0.004324458 | [-0.03 0.03] | 0.78104048 | 0.964905159 |
| GDF_2 | Lipodystrophy_1_pPS | 4159 | 0.015155104 | [-0.02 0.05] | 0.329968803 | 0.84456682 |
| GDF_2 | Lipodystrophy_2_pPS | 4159 | -0.017024539 | [-0.05 0.01] | 0.273796553 | 0.808820203 |
| GDF_2 | Liver-Lipid_pPS | 4159 | -0.043326915 | [-0.07 -0.01] | 0.005332643 | 0.175208366 |
| GDF_2 | ASAT_Pos_pPS | 4159 | -0.021660429 | [-0.05 0.01] | 0.163793672 | 0.718997225 |
| GDF_2 | ALP_Neg_pPS | 4159 | 0.000333857 | [-0.03 0.03] | 0.982879317 | 0.998803911 |
| GDF_2 | SHBG-LpA_pPS | 4159 | 0.023695818 | [-0.01 0.05] | 0.127682688 | 0.694065456 |
| GDF_2 | Obesity_pPS | 4159 | -0.036394845 | [-0.07 -0.01] | 0.019277678 | 0.340663962 |
| GDF_2 | VAT_Neg_pPS | 4159 | -0.01071363 | [-0.04 0.02] | 0.491039125 | 0.895803896 |
| GDF_2 | Bilirubin_pPS | 4159 | 0.009296875 | [-0.02 0.04] | 0.550116338 | 0.899241939 |
| GDF_2 | Proinsulin_pPS | 4159 | -0.014713493 | [-0.05 0.02] | 0.34425741 | 0.847284666 |
| GDF_2 | Hyper_Insulin_pPS | 4159 | 0.010320777 | [-0.02 0.04] | 0.507072077 | 0.895803896 |
| GDF_2 | Beta_Cell_1_pPS | 4159 | 0.041940321 | [0.01 0.07] | 0.006997275 | 0.206973464 |
| GDF_2 | Beta_Cell_2_pPS | 4159 | 0.004608912 | [-0.03 0.04] | 0.767041148 | 0.960245786 |
| GH | Cholesterol_pPS | 4159 | -0.010561087 | [-0.04 0.02] | 0.497232108 | 0.895803896 |
| GH | Lipodystrophy_1_pPS | 4159 | -0.01282379 | [-0.04 0.02] | 0.409766079 | 0.854336605 |
| GH | Lipodystrophy_2_pPS | 4159 | 0.000429239 | [-0.03 0.03] | 0.977989105 | 0.998803911 |
| GH | Liver-Lipid_pPS | 4159 | -0.007169872 | [-0.04 0.02] | 0.644897629 | 0.934865668 |
| GH | ASAT_Pos_pPS | 4159 | -0.001579139 | [-0.03 0.03] | 0.919151997 | 0.99041009 |
| GH | ALP_Neg_pPS | 4159 | -0.022124485 | [-0.05 0.01] | 0.154949632 | 0.708464493 |
| GH | SHBG-LpA_pPS | 4159 | 0.006049131 | [-0.02 0.04] | 0.697407831 | 0.945970482 |
| GH | Obesity_pPS | 4159 | -0.025919359 | [-0.06 0. ] | 0.095655272 | 0.643037244 |
| GH | VAT_Neg_pPS | 4159 | 0.009362964 | [-0.02 0.04] | 0.547284621 | 0.899241939 |
| GH | Bilirubin_pPS | 4159 | 0.003977694 | [-0.03 0.03] | 0.798202412 | 0.967638637 |
| GH | Proinsulin_pPS | 4159 | 0.01170955 | [-0.02 0.04] | 0.451643704 | 0.870351501 |
| GH | Hyper_Insulin_pPS | 4159 | 0.004479205 | [-0.03 0.03] | 0.773415514 | 0.962056893 |
| GH | Beta_Cell_1_pPS | 4159 | -0.001149717 | [-0.03 0.03] | 0.941089861 | 0.995263376 |
| GH | Beta_Cell_2_pPS | 4159 | -0.024603912 | [-0.06 0.01] | 0.113718349 | 0.681046037 |
| GRN | Cholesterol_pPS | 4225 | 0.013184216 | [-0.02 0.04] | 0.39298442 | 0.85037326 |
| GRN | Lipodystrophy_1_pPS | 4225 | 0.000791945 | [-0.03 0.03] | 0.959079599 | 0.996425456 |
| GRN | Lipodystrophy_2_pPS | 4225 | 0.009877916 | [-0.02 0.04] | 0.522110928 | 0.895803896 |
| GRN | Liver-Lipid_pPS | 4225 | -0.016712652 | [-0.05 0.01] | 0.278871973 | 0.808820203 |
| GRN | ASAT_Pos_pPS | 4225 | -0.001079177 | [-0.03 0.03] | 0.944259039 | 0.996180826 |
| GRN | ALP_Neg_pPS | 4225 | 0.020277702 | [-0.01 0.05] | 0.188883484 | 0.747409915 |
| GRN | SHBG-LpA_pPS | 4225 | 0.000196336 | [-0.03 0.03] | 0.989850998 | 0.998803911 |
| GRN | Obesity_pPS | 4225 | 0.009445505 | [-0.02 0.04] | 0.540557501 | 0.899241939 |
| GRN | VAT_Neg_pPS | 4225 | -0.023245586 | [-0.05 0.01] | 0.132005256 | 0.694065456 |
| GRN | Bilirubin_pPS | 4225 | -0.002221659 | [-0.03 0.03] | 0.885549994 | 0.979466202 |
| GRN | Proinsulin_pPS | 4225 | -0.005037631 | [-0.04 0.03] | 0.74413636 | 0.952731244 |
| GRN | Hyper_Insulin_pPS | 4225 | 0.015111639 | [-0.02 0.05] | 0.32752536 | 0.84456682 |
| GRN | Beta_Cell_1_pPS | 4225 | 0.012165951 | [-0.02 0.04] | 0.430557362 | 0.864060889 |
| GRN | Beta_Cell_2_pPS | 4225 | 0.018843432 | [-0.01 0.05] | 0.222109179 | 0.779500334 |
| HB_EGF | Cholesterol_pPS | 4159 | -0.009547766 | [-0.04 0.02] | 0.539404783 | 0.899241939 |
| HB_EGF | Lipodystrophy_1_pPS | 4159 | 0.00773094 | [-0.02 0.04] | 0.619240461 | 0.92152711 |
| HB_EGF | Lipodystrophy_2_pPS | 4159 | 0.008744558 | [-0.02 0.04] | 0.57405981 | 0.908340339 |
| HB_EGF | Liver-Lipid_pPS | 4159 | -0.006781337 | [-0.04 0.02] | 0.662918496 | 0.937489141 |
| HB_EGF | ASAT_Pos_pPS | 4159 | -0.005128511 | [-0.04 0.03] | 0.74166735 | 0.951867558 |
| HB_EGF | ALP_Neg_pPS | 4159 | 0.004787185 | [-0.03 0.04] | 0.758305741 | 0.956140767 |
| HB_EGF | SHBG-LpA_pPS | 4159 | -0.003267535 | [-0.03 0.03] | 0.833646537 | 0.971458352 |
| HB_EGF | Obesity_pPS | 4159 | -0.009960487 | [-0.04 0.02] | 0.522014265 | 0.895803896 |
| HB_EGF | VAT_Neg_pPS | 4159 | -0.0156974 | [-0.05 0.01] | 0.312955481 | 0.833834311 |
| HB_EGF | Bilirubin_pPS | 4159 | 0.002085506 | [-0.03 0.03] | 0.893363276 | 0.983043058 |
| HB_EGF | Proinsulin_pPS | 4159 | 0.010249032 | [-0.02 0.04] | 0.510029502 | 0.895803896 |
| HB_EGF | Hyper_Insulin_pPS | 4159 | -0.043405722 | [-0.07 -0.01] | 0.005249775 | 0.175208366 |
| HB_EGF | Beta_Cell_1_pPS | 4159 | -0.010145654 | [-0.04 0.02] | 0.514306726 | 0.895803896 |
| HB_EGF | Beta_Cell_2_pPS | 4159 | -0.012937695 | [-0.04 0.02] | 0.405619088 | 0.852958996 |
| ICAM_2 | Cholesterol_pPS | 4225 | 0.01646746 | [-0.01 0.05] | 0.285986258 | 0.814934294 |
| ICAM_2 | Lipodystrophy_1_pPS | 4225 | 0.026754219 | [-0. 0.06] | 0.082977405 | 0.614725404 |
| ICAM_2 | Lipodystrophy_2_pPS | 4225 | 0.02352722 | [-0.01 0.05] | 0.12738503 | 0.694065456 |
| ICAM_2 | Liver-Lipid_pPS | 4225 | 0.022762279 | [-0.01 0.05] | 0.14023506 | 0.697681852 |
| ICAM_2 | ASAT_Pos_pPS | 4225 | -0.010582713 | [-0.04 0.02] | 0.492931794 | 0.895803896 |
| ICAM_2 | ALP_Neg_pPS | 4225 | -0.27691736 | [-0.3 -0.25] | 8.28E-75 | 1.07E-71 |
| ICAM_2 | SHBG-LpA_pPS | 4225 | -0.017674856 | [-0.05 0.01] | 0.252122152 | 0.799790128 |
| ICAM_2 | Obesity_pPS | 4225 | 0.025173246 | [-0.01 0.06] | 0.102851149 | 0.660510098 |
| ICAM_2 | VAT_Neg_pPS | 4225 | -0.00816225 | [-0.04 0.02] | 0.596926432 | 0.918301271 |
| ICAM_2 | Bilirubin_pPS | 4225 | -0.004421303 | [-0.03 0.03] | 0.774534048 | 0.962510816 |
| ICAM_2 | Proinsulin_pPS | 4225 | -0.000503224 | [-0.03 0.03] | 0.973991236 | 0.997444743 |
| ICAM_2 | Hyper_Insulin_pPS | 4225 | 0.006241128 | [-0.02 0.04] | 0.685951784 | 0.944421056 |
| ICAM_2 | Beta_Cell_1_pPS | 4225 | 0.014976128 | [-0.02 0.05] | 0.331882255 | 0.844649909 |
| ICAM_2 | Beta_Cell_2_pPS | 4225 | 0.015677825 | [-0.01 0.05] | 0.309725018 | 0.830335001 |
| NEMO | Cholesterol_pPS | 4159 | -0.028723225 | [-0.06 0. ] | 0.064802758 | 0.562394973 |
| NEMO | Lipodystrophy_1_pPS | 4159 | 0.003145881 | [-0.03 0.03] | 0.839754525 | 0.972178273 |
| NEMO | Lipodystrophy_2_pPS | 4159 | 0.019394243 | [-0.01 0.05] | 0.212501627 | 0.767305935 |
| NEMO | Liver-Lipid_pPS | 4159 | -0.007585763 | [-0.04 0.02] | 0.625836608 | 0.927058714 |
| NEMO | ASAT_Pos_pPS | 4159 | 0.013463596 | [-0.02 0.04] | 0.386799881 | 0.85037326 |
| NEMO | ALP_Neg_pPS | 4159 | -0.026722626 | [-0.06 0. ] | 0.085805694 | 0.61569768 |
| NEMO | SHBG-LpA_pPS | 4159 | 0.006551611 | [-0.02 0.04] | 0.673666892 | 0.939698114 |
| NEMO | Obesity_pPS | 4159 | -0.009180442 | [-0.04 0.02] | 0.555122701 | 0.899898936 |
| NEMO | VAT_Neg_pPS | 4159 | 0.00706273 | [-0.02 0.04] | 0.649846835 | 0.935848959 |
| NEMO | Bilirubin_pPS | 4159 | -0.002636861 | [-0.03 0.03] | 0.865411915 | 0.97556306 |
| NEMO | Proinsulin_pPS | 4159 | 0.018311875 | [-0.01 0.05] | 0.239145046 | 0.790085251 |
| NEMO | Hyper_Insulin_pPS | 4159 | -0.01228664 | [-0.04 0.02] | 0.429659141 | 0.864060889 |
| NEMO | Beta_Cell_1_pPS | 4159 | -0.007250895 | [-0.04 0.02] | 0.641165338 | 0.933976497 |
| NEMO | Beta_Cell_2_pPS | 4159 | -0.002675622 | [-0.03 0.03] | 0.863452794 | 0.97556306 |
| IL16 | Cholesterol_pPS | 4159 | -0.000859386 | [-0.03 0.03] | 0.955948425 | 0.996425456 |
| IL16 | Lipodystrophy_1_pPS | 4159 | 0.016911946 | [-0.01 0.05] | 0.276982622 | 0.808820203 |

**Table S7: Associations between pPSs and plasma proteins in SCAPIS**

**a) Spearman Correlation**

| Proteomics | PPS | Sample_Size | r | CI95% | p | fdr_p_adjusted |
|---|---|---|---|---|---|---|
| GRN | Lipodystrophy_1_pPS | 4225 | 0.003346142 | [-0.03 0.03] | 0.8281099 | 0.966003345 |
| GRN | Lipodystrophy_2_pPS | 4225 | 0.008085243 | [-0.02 0.04] | 0.599828781 | 0.907623171 |
| GRN | Liver-Lipid_pPS | 4225 | -0.01394208 | [-0.04 0.02] | 0.365612265 | 0.798890546 |
| GRN | ASAT_Pos_pPS | 4225 | 0.001235179 | [-0.03 0.03] | 0.936118909 | 0.99138381 |
| GRN | ALP_Neg_pPS | 4225 | 0.019959945 | [-0.01 0.05] | 0.19521771 | 0.697025716 |
| GRN | SHBG-LpA_pPS | 4225 | -0.001615828 | [-0.03 0.03] | 0.916496026 | 0.990056084 |
| GRN | Obesity_pPS | 4225 | 0.01174263 | [-0.02 0.04] | 0.446068145 | 0.831350198 |
| GRN | VAT_Neg_pPS | 4225 | -0.018755672 | [-0.05 0.01] | 0.223554024 | 0.7224931 |
| GRN | Bilirubin_pPS | 4225 | -0.001407566 | [-0.03 0.03] | 0.927226656 | 0.990056084 |
| GRN | Proinsulin_pPS | 4225 | 0.005724709 | [-0.02 0.04] | 0.710287439 | 0.938946716 |
| GRN | Hyper_Insulin_pPS | 4225 | 0.015083565 | [-0.02 0.05] | 0.327677065 | 0.775125841 |
| GRN | Beta_Cell_1_pPS | 4225 | 0.013235071 | [-0.02 0.04] | 0.390428937 | 0.81106215 |
| GRN | Beta_Cell_2_pPS | 4225 | 0.020790103 | [-0.01 0.05] | 0.177278629 | 0.678995855 |
| GRN | Total_GRS | 4225 | 0.016024631 | [-0.01 0.05] | 0.298396165 | 0.759754073 |
| HB_EGF | Cholesterol_pPS | 4159 | -0.010416823 | [-0.04 0.02] | 0.502456096 | 0.864575328 |
| HB_EGF | Lipodystrophy_1_pPS | 4159 | 0.000774146 | [-0.03 0.03] | 0.960251629 | 0.99612031 |
| HB_EGF | Lipodystrophy_2_pPS | 4159 | 0.003471985 | [-0.03 0.03] | 0.82313131 | 0.963461584 |
| HB_EGF | Liver-Lipid_pPS | 4159 | -0.00965526 | [-0.04 0.02] | 0.53420569 | 0.879718201 |
| HB_EGF | ASAT_Pos_pPS | 4159 | -0.007342093 | [-0.04 0.02] | 0.636445721 | 0.915367478 |
| HB_EGF | ALP_Neg_pPS | 4159 | 0.005308741 | [-0.03 0.04] | 0.732525925 | 0.942134248 |
| HB_EGF | SHBG-LpA_pPS | 4159 | -0.004165176 | [-0.03 0.03] | 0.788586945 | 0.957247257 |
| HB_EGF | Obesity_pPS | 4159 | -0.012633456 | [-0.04 0.02] | 0.416019436 | 0.817860704 |
| HB_EGF | VAT_Neg_pPS | 4159 | -0.020540143 | [-0.05 0.01] | 0.186012183 | 0.68727393 |
| HB_EGF | Bilirubin_pPS | 4159 | 0.001878964 | [-0.03 0.03] | 0.903719831 | 0.987828409 |
| HB_EGF | Proinsulin_pPS | 4159 | 0.003765575 | [-0.03 0.03] | 0.80845452 | 0.96133624 |
| HB_EGF | Hyper_Insulin_pPS | 4159 | -0.04571639 | [-0.08 -0.02] | 0.0032331 | 0.106782928 |
| HB_EGF | Beta_Cell_1_pPS | 4159 | -0.01385566 | [-0.04 0.02] | 0.372371693 | 0.799802236 |
| HB_EGF | Beta_Cell_2_pPS | 4159 | -0.016834908 | [-0.05 0.01] | 0.278421996 | 0.747422497 |
| HB_EGF | Total_GRS | 4159 | -0.02410746 | [-0.05 0.01] | 0.120611842 | 0.615837655 |
| ICAM_2 | Cholesterol_pPS | 4225 | 0.019205766 | [-0.01 0.05] | 0.212637509 | 0.712633026 |
| ICAM_2 | Lipodystrophy_1_pPS | 4225 | 0.021091959 | [-0.01 0.05] | 0.171069613 | 0.674627844 |
| ICAM_2 | Lipodystrophy_2_pPS | 4225 | 0.022269226 | [-0.01 0.05] | 0.148403708 | 0.647068301 |
| ICAM_2 | Liver-Lipid_pPS | 4225 | 0.012942832 | [-0.02 0.04] | 0.400978806 | 0.81230319 |
| ICAM_2 | ASAT_Pos_pPS | 4225 | -0.002478582 | [-0.03 0.03] | 0.872226208 | 0.977006629 |
| ICAM_2 | ALP_Neg_pPS | 4225 | -0.278154147 | [-0.31 -0.25] | 1.02E-75 | 1.41E-72 |
| ICAM_2 | SHBG-LpA_pPS | 4225 | -0.007596399 | [-0.04 0.02] | 0.622065574 | 0.911342425 |
| ICAM_2 | Obesity_pPS | 4225 | 0.022489329 | [-0.01 0.05] | 0.144432942 | 0.641415816 |
| ICAM_2 | VAT_Neg_pPS | 4225 | -0.002557552 | [-0.03 0.03] | 0.868191867 | 0.977006629 |
| ICAM_2 | Bilirubin_pPS | 4225 | -0.003108778 | [-0.03 0.03] | 0.840132505 | 0.972342391 |
| ICAM_2 | Proinsulin_pPS | 4225 | -0.036956761 | [-0.07 -0.01] | 0.016445155 | 0.271788193 |
| ICAM_2 | Hyper_Insulin_pPS | 4225 | 0.005213096 | [-0.02 0.04] | 0.735158535 | 0.942470055 |
| ICAM_2 | Beta_Cell_1_pPS | 4225 | 0.017615033 | [-0.01 0.05] | 0.252997335 | 0.739680398 |
| ICAM_2 | Beta_Cell_2_pPS | 4225 | 0.017062051 | [-0.01 0.05] | 0.268203506 | 0.746520328 |
| ICAM_2 | Total_GRS | 4225 | 0.006216454 | [-0.02 0.04] | 0.686669106 | 0.926969309 |
| NEMO | Cholesterol_pPS | 4159 | -0.02979757 | [-0.06 0. ] | 0.055018797 | 0.450599046 |
| NEMO | Lipodystrophy_1_pPS | 4159 | 0.004943229 | [-0.03 0.04] | 0.750305827 | 0.948757065 |
| NEMO | Lipodystrophy_2_pPS | 4159 | 0.020256499 | [-0.01 0.05] | 0.192164565 | 0.694207068 |
| NEMO | Liver-Lipid_pPS | 4159 | -0.009023533 | [-0.04 0.02] | 0.561289702 | 0.897748415 |
| NEMO | ASAT_Pos_pPS | 4159 | 0.015054885 | [-0.02 0.05] | 0.332417892 | 0.778839882 |
| NEMO | ALP_Neg_pPS | 4159 | -0.024806189 | [-0.06 0.01] | 0.110218144 | 0.603827339 |
| NEMO | SHBG-LpA_pPS | 4159 | 0.009431492 | [-0.02 0.04] | 0.543723419 | 0.886924725 |
| NEMO | Obesity_pPS | 4159 | -0.010612607 | [-0.04 0.02] | 0.494458079 | 0.862099999 |
| NEMO | VAT_Neg_pPS | 4159 | 0.009530669 | [-0.02 0.04] | 0.539494667 | 0.882112133 |
| NEMO | Bilirubin_pPS | 4159 | -0.002084979 | [-0.03 0.03] | 0.893223469 | 0.981916701 |
| NEMO | Proinsulin_pPS | 4159 | 0.011185416 | [-0.02 0.04] | 0.471454258 | 0.846453261 |
| NEMO | Hyper_Insulin_pPS | 4159 | -0.010914296 | [-0.04 0.02] | 0.482268178 | 0.853791001 |
| NEMO | Beta_Cell_1_pPS | 4159 | -0.002185588 | [-0.03 0.03] | 0.888104163 | 0.979291846 |
| NEMO | Beta_Cell_2_pPS | 4159 | -0.00168932 | [-0.03 0.03] | 0.913397023 | 0.990056084 |
| NEMO | Total_GRS | 4159 | -0.000987828 | [-0.03 0.03] | 0.949293352 | 0.993154165 |
| IL16 | Cholesterol_pPS | 4159 | 0.00312073 | [-0.03 0.03] | 0.840771918 | 0.972342391 |
| IL16 | Lipodystrophy_1_pPS | 4159 | 0.019157109 | [-0.01 0.05] | 0.217424505 | 0.716955357 |
| IL16 | Lipodystrophy_2_pPS | 4159 | 0.019103995 | [-0.01 0.05] | 0.218702636 | 0.718175854 |
| IL16 | Liver-Lipid_pPS | 4159 | 0.036633363 | [0.01 0.07] | 0.01831602 | 0.290017524 |
| IL16 | ASAT_Pos_pPS | 4159 | 0.007491054 | [-0.02 0.04] | 0.629618209 | 0.914460114 |
| IL16 | ALP_Neg_pPS | 4159 | 0.007922165 | [-0.02 0.04] | 0.610037793 | 0.909483337 |
| IL16 | SHBG-LpA_pPS | 4159 | 0.006712735 | [-0.02 0.04] | 0.665628091 | 0.924111434 |
| IL16 | Obesity_pPS | 4159 | 0.016879218 | [-0.01 0.05] | 0.277158729 | 0.747422497 |
| IL16 | VAT_Neg_pPS | 4159 | -0.017755184 | [-0.05 0.01] | 0.252984783 | 0.739680398 |
| IL16 | Bilirubin_pPS | 4159 | -0.015522686 | [-0.05 0.01] | 0.317612366 | 0.767539002 |
| IL16 | Proinsulin_pPS | 4159 | 0.003533565 | [-0.03 0.03] | 0.820047559 | 0.963461584 |
| IL16 | Hyper_Insulin_pPS | 4159 | 0.003365844 | [-0.03 0.03] | 0.828452869 | 0.966003345 |
| IL16 | Beta_Cell_1_pPS | 4159 | -0.015812005 | [-0.05 0.01] | 0.308675311 | 0.76365449 |
| IL16 | Beta_Cell_2_pPS | 4159 | -0.024936692 | [-0.06 0.01] | 0.10835769 | 0.599333113 |
| IL16 | Total_GRS | 4159 | 0.021670941 | [-0.01 0.05] | 0.162928065 | 0.664061293 |
| IL_17D | Cholesterol_pPS | 4159 | 0.001376344 | [-0.03 0.03] | 0.929395041 | 0.990056084 |
| IL_17D | Lipodystrophy_1_pPS | 4159 | -0.013283032 | [-0.04 0.02] | 0.392457065 | 0.812191725 |
| IL_17D | Lipodystrophy_2_pPS | 4159 | 0.01952791 | [-0.01 0.05] | 0.208650965 | 0.709689105 |
| IL_17D | Liver-Lipid_pPS | 4159 | -0.000488337 | [-0.03 0.03] | 0.974920185 | 0.996862516 |
| IL_17D | ASAT_Pos_pPS | 4159 | -0.007764868 | [-0.04 0.02] | 0.61715071 | 0.909483337 |
| IL_17D | ALP_Neg_pPS | 4159 | -0.002558036 | [-0.03 0.03] | 0.869195087 | 0.977006629 |
| IL_17D | SHBG-LpA_pPS | 4159 | -0.005594884 | [-0.04 0.02] | 0.71870575 | 0.941449667 |
| IL_17D | Obesity_pPS | 4159 | -0.032672101 | [-0.06 -0. ] | 0.03538548 | 0.386023413 |
| IL_17D | VAT_Neg_pPS | 4159 | 0.00338616 | [-0.03 0.03] | 0.827433708 | 0.966003345 |
| IL_17D | Bilirubin_pPS | 4159 | 0.005217117 | [-0.03 0.04] | 0.736969859 | 0.942470055 |
| IL_17D | Proinsulin_pPS | 4159 | -0.007261753 | [-0.04 0.02] | 0.640140902 | 0.916384279 |
| IL_17D | Hyper_Insulin_pPS | 4159 | -0.011004435 | [-0.04 0.02] | 0.478657975 | 0.850673542 |
| IL_17D | Beta_Cell_1_pPS | 4159 | -0.034466725 | [-0.06 -0. ] | 0.026449259 | 0.344676278 |
| IL_17D | Beta_Cell_2_pPS | 4159 | -0.017900508 | [-0.05 0.01] | 0.249120807 | 0.738661262 |
| IL_17D | Total_GRS | 4159 | -0.008209982 | [-0.04 0.02] | 0.597118079 | 0.907623171 |
| IL_17RA | Cholesterol_pPS | 4225 | -0.018271384 | [-0.05 0.01] | 0.235741108 | 0.726631332 |
| IL_17RA | Lipodystrophy_1_pPS | 4225 | -0.007206215 | [-0.04 0.02] | 0.640066688 | 0.916384279 |
| IL_17RA | Lipodystrophy_2_pPS | 4225 | 0.03233396 | [0. 0.06] | 0.035848073 | 0.386599178 |
| IL_17RA | Liver-Lipid_pPS | 4225 | -0.006179294 | [-0.04 0.02] | 0.688443589 | 0.926969309 |
| IL_17RA | ASAT_Pos_pPS | 4225 | -0.015954253 | [-0.05 0.01] | 0.300523551 | 0.76319014 |
| IL_17RA | ALP_Neg_pPS | 4225 | 0.029765774 | [-0. 0.06] | 0.053374259 | 0.442381248 |
| IL_17RA | SHBG-LpA_pPS | 4225 | -0.025838115 | [-0.06 0. ] | 0.093567317 | 0.560186108 |
| IL_17RA | Obesity_pPS | 4225 | 0.00111301 | [-0.03 0.03] | 0.942425719 | 0.992903797 |
| IL_17RA | VAT_Neg_pPS | 4225 | -0.002249107 | [-0.03 0.03] | 0.883967898 | 0.979291846 |
| IL_17RA | Bilirubin_pPS | 4225 | -0.018370066 | [-0.05 0.01] | 0.233220412 | 0.726631332 |
| IL_17RA | Proinsulin_pPS | 4225 | 0.012851456 | [-0.02 0.04] | 0.404312292 | 0.81230319 |
| IL_17RA | Hyper_Insulin_pPS | 4225 | -0.009820864 | [-0.04 0.02] | 0.523945605 | 0.872980599 |
| IL_17RA | Beta_Cell_1_pPS | 4225 | 0.024251046 | [-0.01 0.05] | 0.115523932 | 0.610816194 |
| IL_17RA | Beta_Cell_2_pPS | 4225 | 0.030250174 | [0. 0.06] | 0.049607421 | 0.440215663 |
| IL_17RA | Total_GRS | 4225 | 0.02962412 | [-0. 0.06] | 0.05451991 | 0.449178958 |
| IL18 | Cholesterol_pPS | 4159 | 0.006271246 | [-0.02 0.04] | 0.686409116 | 0.926969309 |
| IL18 | Lipodystrophy_1_pPS | 4159 | 0.02331964 | [-0.01 0.05] | 0.133233766 | 0.629316669 |
| IL18 | Lipodystrophy_2_pPS | 4159 | 0.039283993 | [0.01 0.07] | 0.011406471 | 0.232377917 |

**b) Partial Correlation**

| Proteomics | PPS | Sample_Size | r | CI95% | p | fdr_p_adjusted |
|---|---|---|---|---|---|---|
| IL16 | Lipodystrophy_2_pPS | 4159 | 0.018774015 | [-0.01 0.05] | 0.227494171 | 0.786988264 |
| IL16 | Liver-Lipid_pPS | 4159 | 0.036498329 | [0.01 0.07] | 0.018936651 | 0.340663962 |
| IL16 | ASAT_Pos_pPS | 4159 | 0.002220636 | [-0.03 0.03] | 0.886499117 | 0.979597623 |
| IL16 | ALP_Neg_pPS | 4159 | 0.005260654 | [-0.03 0.04] | 0.735257655 | 0.948434512 |
| IL16 | SHBG-LpA_pPS | 4159 | 0.006425217 | [-0.02 0.04] | 0.679609316 | 0.943176659 |
| IL16 | Obesity_pPS | 4159 | 0.015241375 | [-0.02 0.05] | 0.327222937 | 0.84456682 |
| IL16 | VAT_Neg_pPS | 4159 | -0.018192688 | [-0.05 0.01] | 0.242216905 | 0.791815669 |
| IL16 | Bilirubin_pPS | 4159 | -0.014768973 | [-0.05 0.02] | 0.342440941 | 0.847284666 |
| IL16 | Proinsulin_pPS | 4159 | 0.006929205 | [-0.02 0.04] | 0.656036405 | 0.936605382 |
| IL16 | Hyper_Insulin_pPS | 4159 | -0.001122255 | [-0.03 0.03] | 0.942494514 | 0.99584326 |
| IL16 | Beta_Cell_1_pPS | 4159 | -0.009881309 | [-0.04 0.02] | 0.52532805 | 0.898044266 |
| IL16 | Beta_Cell_2_pPS | 4159 | -0.022502714 | [-0.05 0.01] | 0.14801337 | 0.703126624 |
| IL_17D | Cholesterol_pPS | 4159 | 0.003537066 | [-0.03 0.03] | 0.8201502 | 0.968668398 |
| IL_17D | Lipodystrophy_1_pPS | 4159 | -0.010286152 | [-0.04 0.02] | 0.508498269 | 0.895803896 |
| IL_17D | Lipodystrophy_2_pPS | 4159 | 0.019336411 | [-0.01 0.05] | 0.213868657 | 0.767305935 |
| IL_17D | Liver-Lipid_pPS | 4159 | 0.001148092 | [-0.03 0.03] | 0.941172976 | 0.995263376 |
| IL_17D | ASAT_Pos_pPS | 4159 | -0.00491831 | [-0.04 0.03] | 0.751900135 | 0.954135344 |
| IL_17D | ALP_Neg_pPS | 4159 | -0.002986991 | [-0.03 0.03] | 0.847746543 | 0.97401221 |
| IL_17D | SHBG-LpA_pPS | 4159 | -0.005565635 | [-0.04 0.02] | 0.720535165 | 0.946314812 |
| IL_17D | Obesity_pPS | 4159 | -0.031803747 | [-0.06 -0. ] | 0.040878665 | 0.4797351 |
| IL_17D | VAT_Neg_pPS | 4159 | 0.00706583 | [-0.02 0.04] | 0.649703438 | 0.935848959 |
| IL_17D | Bilirubin_pPS | 4159 | 0.006775347 | [-0.02 0.04] | 0.66319791 | 0.937489141 |
| IL_17D | Proinsulin_pPS | 4159 | 0.008236526 | [-0.02 0.04] | 0.59650991 | 0.918301271 |
| IL_17D | Hyper_Insulin_pPS | 4159 | -0.008690293 | [-0.04 0.02] | 0.576438627 | 0.909113996 |
| IL_17D | Beta_Cell_1_pPS | 4159 | -0.031068534 | [-0.06 -0. ] | 0.045774542 | 0.512674866 |
| IL_17D | Beta_Cell_2_pPS | 4159 | -0.010571081 | [-0.04 0.02] | 0.496825114 | 0.895803896 |
| IL_17RA | Cholesterol_pPS | 4225 | -0.020230552 | [-0.05 0.01] | 0.189914018 | 0.750335139 |
| IL_17RA | Lipodystrophy_1_pPS | 4225 | -0.003358923 | [-0.03 0.03] | 0.827726027 | 0.971458352 |
| IL_17RA | Lipodystrophy_2_pPS | 4225 | 0.036840858 | [0.01 0.07] | 0.016956005 | 0.33440804 |
| IL_17RA | Liver-Lipid_pPS | 4225 | -0.00890457 | [-0.04 0.02] | 0.5639923 | 0.904956448 |
| IL_17RA | ASAT_Pos_pPS | 4225 | -0.017922935 | [-0.05 0.01] | 0.245525415 | 0.79432283 |
| IL_17RA | ALP_Neg_pPS | 4225 | 0.02895174 | [-0. 0.06] | 0.060638537 | 0.558815902 |
| IL_17RA | SHBG-LpA_pPS | 4225 | -0.025057853 | [-0.06 0.01] | 0.104438564 | 0.663720785 |
| IL_17RA | Obesity_pPS | 4225 | 0.003258526 | [-0.03 0.03] | 0.832798071 | 0.971458352 |
| IL_17RA | VAT_Neg_pPS | 4225 | -0.008826687 | [-0.04 0.02] | 0.567406262 | 0.905613993 |
| IL_17RA | Bilirubin_pPS | 4225 | -0.019677894 | [-0.05 0.01] | 0.202303179 | 0.765145476 |
| IL_17RA | Proinsulin_pPS | 4225 | -0.001009073 | [-0.03 0.03] | 0.947874693 | 0.996425456 |
| IL_17RA | Hyper_Insulin_pPS | 4225 | -0.010188867 | [-0.04 0.02] | 0.509167433 | 0.895803896 |
| IL_17RA | Beta_Cell_1_pPS | 4225 | 0.019770092 | [-0.01 0.05] | 0.200196392 | 0.765145476 |
| IL_17RA | Beta_Cell_2_pPS | 4225 | 0.024677908 | [-0.01 0.05] | 0.109803122 | 0.676681438 |
| IL18 | Cholesterol_pPS | 4159 | 0.007052963 | [-0.02 0.04] | 0.650298772 | 0.935848959 |
| IL18 | Lipodystrophy_1_pPS | 4159 | 0.021830491 | [-0.01 0.05] | 0.160509477 | 0.717834051 |
| IL18 | Lipodystrophy_2_pPS | 4159 | 0.033748346 | [0. 0.06] | 0.030018066 | 0.416452728 |
| IL18 | Liver-Lipid_pPS | 4159 | -0.00563741 | [-0.04 0.02] | 0.71708506 | 0.945970482 |
| IL18 | ASAT_Pos_pPS | 4159 | -0.002798931 | [-0.03 0.03] | 0.857225971 | 0.97556306 |
| IL18 | ALP_Neg_pPS | 4159 | -0.015999867 | [-0.05 0.01] | 0.30372164 | 0.826182442 |
| IL18 | SHBG-LpA_pPS | 4159 | -0.015263884 | [-0.05 0.02] | 0.326508947 | 0.84456682 |
| IL18 | Obesity_pPS | 4159 | -0.007772594 | [-0.04 0.02] | 0.617353535 | 0.921525431 |
| IL18 | VAT_Neg_pPS | 4159 | 0.036385776 | [0.01 0.07] | 0.019307818 | 0.340663962 |
| IL18 | Bilirubin_pPS | 4159 | 0.007934133 | [-0.02 0.04] | 0.610059657 | 0.919490897 |
| IL18 | Proinsulin_pPS | 4159 | 0.011748665 | [-0.02 0.04] | 0.450133765 | 0.870351501 |
| IL18 | Hyper_Insulin_pPS | 4159 | -0.005763399 | [-0.04 0.02] | 0.711043066 | 0.945970482 |
| IL18 | Beta_Cell_1_pPS | 4159 | 0.000472308 | [-0.03 0.03] | 0.975781208 | 0.998477998 |
| IL18 | Beta_Cell_2_pPS | 4159 | -0.007895085 | [-0.04 0.02] | 0.61181927 | 0.920050462 |
| IL_1RT1 | Cholesterol_pPS | 4225 | 0.010467576 | [-0.02 0.04] | 0.497649248 | 0.895803896 |
| IL_1RT1 | Lipodystrophy_1_pPS | 4225 | 0.001430995 | [-0.03 0.03] | 0.926132723 | 0.990746634 |
| IL_1RT1 | Lipodystrophy_2_pPS | 4225 | 0.035382564 | [0.01 0.07] | 0.021842961 | 0.358391511 |
| IL_1RT1 | Liver-Lipid_pPS | 4225 | -0.026710598 | [-0.06 0. ] | 0.083480639 | 0.614725404 |
| IL_1RT1 | ASAT_Pos_pPS | 4225 | -0.010988533 | [-0.04 0.02] | 0.476497076 | 0.886891048 |
| IL_1RT1 | ALP_Neg_pPS | 4225 | -0.026690873 | [-0.06 0. ] | 0.08370901 | 0.614725404 |
| IL_1RT1 | SHBG-LpA_pPS | 4225 | -0.00663833 | [-0.04 0.02] | 0.667130416 | 0.937489141 |
| IL_1RT1 | Obesity_pPS | 4225 | 0.001761879 | [-0.03 0.03] | 0.909119766 | 0.988841471 |
| IL_1RT1 | VAT_Neg_pPS | 4225 | -0.021153044 | [-0.05 0.01] | 0.170493683 | 0.726214013 |
| IL_1RT1 | Bilirubin_pPS | 4225 | -0.000203108 | [-0.03 0.03] | 0.989500975 | 0.998803911 |
| IL_1RT1 | Proinsulin_pPS | 4225 | 0.022590982 | [-0.01 0.05] | 0.143244785 | 0.697681852 |
| IL_1RT1 | Hyper_Insulin_pPS | 4225 | -0.00644972 | [-0.04 0.02] | 0.676042485 | 0.942362252 |
| IL_1RT1 | Beta_Cell_1_pPS | 4225 | 0.017301973 | [-0.01 0.05] | 0.262268211 | 0.807171938 |
| IL_1RT1 | Beta_Cell_2_pPS | 4225 | 0.001761039 | [-0.03 0.03] | 0.909162893 | 0.988841471 |
| IL_1RT2 | Cholesterol_pPS | 4225 | -0.006796602 | [-0.04 0.02] | 0.659687862 | 0.937489141 |
| IL_1RT2 | Lipodystrophy_1_pPS | 4225 | 0.00665672 | [-0.02 0.04] | 0.666263965 | 0.937489141 |
| IL_1RT2 | Lipodystrophy_2_pPS | 4225 | 0.047768684 | [0.02 0.08] | 0.001957467 | 0.099117562 |
| IL_1RT2 | Liver-Lipid_pPS | 4225 | -0.030474245 | [-0.06 -0. ] | 0.048287719 | 0.520248282 |
| IL_1RT2 | ASAT_Pos_pPS | 4225 | 0.017965231 | [-0.01 0.05] | 0.244412895 | 0.79432283 |
| IL_1RT2 | ALP_Neg_pPS | 4225 | -0.003162904 | [-0.03 0.03] | 0.837635333 | 0.972099644 |
| IL_1RT2 | SHBG-LpA_pPS | 4225 | 0.002959688 | [-0.03 0.03] | 0.847935669 | 0.97401221 |
| IL_1RT2 | Obesity_pPS | 4225 | -0.015421986 | [-0.05 0.01] | 0.317687718 | 0.839009314 |
| IL_1RT2 | VAT_Neg_pPS | 4225 | 0.00301765 | [-0.03 0.03] | 0.844995013 | 0.972178273 |
| IL_1RT2 | Bilirubin_pPS | 4225 | 0.025328082 | [-0. 0.06] | 0.100751341 | 0.653741699 |
| IL_1RT2 | Proinsulin_pPS | 4225 | 0.014368806 | [-0.02 0.04] | 0.351866025 | 0.847284666 |
| IL_1RT2 | Hyper_Insulin_pPS | 4225 | 0.008432064 | [-0.02 0.04] | 0.584854736 | 0.911131843 |
| IL_1RT2 | Beta_Cell_1_pPS | 4225 | -0.000890627 | [-0.03 0.03] | 0.953985946 | 0.996425456 |
| IL_1RT2 | Beta_Cell_2_pPS | 4225 | -0.002696025 | [-0.03 0.03] | 0.861338399 | 0.97556306 |
| ST2 | Cholesterol_pPS | 4225 | 0.000219071 | [-0.03 0.03] | 0.988675826 | 0.998803911 |
| ST2 | Lipodystrophy_1_pPS | 4225 | 0.018055711 | [-0.01 0.05] | 0.242044887 | 0.791815669 |
| ST2 | Lipodystrophy_2_pPS | 4225 | 0.013116445 | [-0.02 0.04] | 0.395421662 | 0.85037326 |
| ST2 | Liver-Lipid_pPS | 4225 | -0.022956996 | [-0.05 0.01] | 0.136873157 | 0.694065456 |
| ST2 | ASAT_Pos_pPS | 4225 | -0.040077749 | [-0.07 -0.01] | 0.009387628 | 0.248521918 |
| ST2 | ALP_Neg_pPS | 4225 | -0.025345941 | [-0.06 0. ] | 0.10051136 | 0.653741699 |
| ST2 | SHBG-LpA_pPS | 4225 | 0.004604859 | [-0.03 0.03] | 0.765442368 | 0.959503426 |
| ST2 | Obesity_pPS | 4225 | -0.005019766 | [-0.04 0.03] | 0.745012135 | 0.953378669 |
| ST2 | VAT_Neg_pPS | 4225 | 0.014945985 | [-0.02 0.05] | 0.332856462 | 0.844649909 |
| ST2 | Bilirubin_pPS | 4225 | 0.000217365 | [-0.03 0.03] | 0.988764024 | 0.998803911 |
| ST2 | Proinsulin_pPS | 4225 | 0.021393966 | [-0.01 0.05] | 0.165675654 | 0.720912981 |
| ST2 | Hyper_Insulin_pPS | 4225 | 0.008284259 | [-0.02 0.04] | 0.59145363 | 0.915615716 |
| ST2 | Beta_Cell_1_pPS | 4225 | 0.003253109 | [-0.03 0.03] | 0.833071919 | 0.971458352 |
| ST2 | Beta_Cell_2_pPS | 4225 | -0.026411287 | [-0.06 0. ] | 0.087000651 | 0.616770438 |
| IL1RL2 | Cholesterol_pPS | 4159 | 0.000530199 | [-0.03 0.03] | 0.972813782 | 0.997444743 |
| IL1RL2 | Lipodystrophy_1_pPS | 4159 | -0.002869332 | [-0.03 0.03] | 0.853674818 | 0.975195713 |
| IL1RL2 | Lipodystrophy_2_pPS | 4159 | -0.005732455 | [-0.04 0.02] | 0.712525351 | 0.945970482 |
| IL1RL2 | Liver-Lipid_pPS | 4159 | 0.009778324 | [-0.02 0.04] | 0.529654254 | 0.898959051 |
| IL1RL2 | ASAT_Pos_pPS | 4159 | 0.012053784 | [-0.02 0.04] | 0.438454195 | 0.865276785 |
| IL1RL2 | ALP_Neg_pPS | 4159 | -0.006022567 | [-0.04 0.02] | 0.698671407 | 0.945970482 |
| IL1RL2 | SHBG-LpA_pPS | 4159 | 0.005014113 | [-0.03 0.04] | 0.747230816 | 0.954135344 |
| IL1RL2 | Obesity_pPS | 4159 | -0.03407654 | [-0.06 -0. ] | 0.028454298 | 0.414433192 |
| IL1RL2 | VAT_Neg_pPS | 4159 | 0.030691607 | [0. 0.06] | 0.048470337 | 0.520248282 |
| IL1RL2 | Bilirubin_pPS | 4159 | 0.011457462 | [-0.02 0.04] | 0.461443159 | 0.875959895 |
| IL1RL2 | Proinsulin_pPS | 4159 | 0.013469836 | [-0.02 0.04] | 0.386579833 | 0.85037326 |

**Table S7: Associations between pPSs and plasma proteins in SCAPIS**

**a) Spearman Correlation**

| Proteomics | PPS | Sample_Size | r | CI95% | p | fdr_p_adjusted |
|---|---|---|---|---|---|---|
| IL18 | Liver-Lipid_pPS | 4159 | -0.004735991 | [-0.04 0.03] | 0.760446638 | 0.950403646 |
| IL18 | ASAT_Pos_pPS | 4159 | 0.004280269 | [-0.03 0.03] | 0.782889472 | 0.955139045 |
| IL18 | ALP_Neg_pPS | 4159 | -0.013590356 | [-0.04 0.02] | 0.381597031 | 0.806027092 |
| IL18 | SHBG-LpA_pPS | 4159 | -0.010356314 | [-0.04 0.02] | 0.504941718 | 0.867177846 |
| IL18 | Obesity_pPS | 4159 | -0.005572062 | [-0.04 0.02] | 0.719804673 | 0.941449667 |
| IL18 | VAT_Neg_pPS | 4159 | 0.040000825 | [0.01 0.07] | 0.009989406 | 0.222344847 |
| IL18 | Bilirubin_pPS | 4159 | 0.009301567 | [-0.02 0.04] | 0.549288053 | 0.891638418 |
| IL18 | Proinsulin_pPS | 4159 | 0.008546013 | [-0.02 0.04] | 0.582193525 | 0.903823705 |
| IL18 | Hyper_Insulin_pPS | 4159 | 0.001027641 | [-0.03 0.03] | 0.947252597 | 0.993154165 |
| IL18 | Beta_Cell_1_pPS | 4159 | 0.005674712 | [-0.02 0.04] | 0.714866302 | 0.94141015 |
| IL18 | Beta_Cell_2_pPS | 4159 | -0.003061453 | [-0.03 0.03] | 0.843757082 | 0.973549297 |
| IL18 | Total_GRS | 4159 | 0.024725959 | [-0.01 0.06] | 0.111374365 | 0.604978148 |
| IL_1RT1 | Cholesterol_pPS | 4225 | 0.012040863 | [-0.02 0.04] | 0.434602414 | 0.828998134 |
| IL_1RT1 | Lipodystrophy_1_pPS | 4225 | -0.000396245 | [-0.03 0.03] | 0.979487321 | 0.996862516 |
| IL_1RT1 | Lipodystrophy_2_pPS | 4225 | 0.030976849 | [0. 0.06] | 0.044376229 | 0.424281318 |
| IL_1RT1 | Liver-Lipid_pPS | 4225 | -0.025866368 | [-0.06 0. ] | 0.093209123 | 0.559254737 |
| IL_1RT1 | ASAT_Pos_pPS | 4225 | -0.007774889 | [-0.04 0.02] | 0.613904828 | 0.909483337 |
| IL_1RT1 | ALP_Neg_pPS | 4225 | -0.023782181 | [-0.05 0.01] | 0.122732505 | 0.615837655 |
| IL_1RT1 | SHBG-LpA_pPS | 4225 | -0.006009873 | [-0.04 0.02] | 0.696555489 | 0.930772879 |
| IL_1RT1 | Obesity_pPS | 4225 | 0.0043904 | [-0.03 0.03] | 0.775730512 | 0.953637204 |
| IL_1RT1 | VAT_Neg_pPS | 4225 | -0.015834745 | [-0.05 0.01] | 0.304159133 | 0.76365449 |
| IL_1RT1 | Bilirubin_pPS | 4225 | 0.001186714 | [-0.03 0.03] | 0.938620397 | 0.992111394 |
| IL_1RT1 | Proinsulin_pPS | 4225 | 0.024486339 | [-0.01 0.05] | 0.112034025 | 0.604978148 |
| IL_1RT1 | Hyper_Insulin_pPS | 4225 | -0.007878281 | [-0.04 0.02] | 0.609199333 | 0.909483337 |
| IL_1RT1 | Beta_Cell_1_pPS | 4225 | 0.021276114 | [-0.01 0.05] | 0.16736244 | 0.666544721 |
| IL_1RT1 | Beta_Cell_2_pPS | 4225 | 0.008778181 | [-0.02 0.04] | 0.568940342 | 0.900387238 |
| IL_1RT1 | Total_GRS | 4225 | 0.011810331 | [-0.02 0.04] | 0.443450351 | 0.831350198 |
| IL_1RT2 | Cholesterol_pPS | 4225 | -0.007396253 | [-0.04 0.02] | 0.631271941 | 0.914460114 |
| IL_1RT2 | Lipodystrophy_1_pPS | 4225 | 0.014051213 | [-0.02 0.04] | 0.361871229 | 0.7979754 |
| IL_1RT2 | Lipodystrophy_2_pPS | 4225 | 0.05071965 | [0.02 0.08] | 0.000990417 | 0.050992277 |
| IL_1RT2 | Liver-Lipid_pPS | 4225 | -0.029117496 | [-0.06 0. ] | 0.058786402 | 0.458334656 |
| IL_1RT2 | ASAT_Pos_pPS | 4225 | 0.024907271 | [-0.01 0.06] | 0.105998598 | 0.595837331 |
| IL_1RT2 | ALP_Neg_pPS | 4225 | -0.001675435 | [-0.03 0.03] | 0.913427487 | 0.990056084 |
| IL_1RT2 | SHBG-LpA_pPS | 4225 | 0.004942071 | [-0.03 0.04] | 0.748449223 | 0.948012784 |
| IL_1RT2 | Obesity_pPS | 4225 | -0.013262017 | [-0.04 0.02] | 0.389464753 | 0.810650617 |
| IL_1RT2 | VAT_Neg_pPS | 4225 | 0.010742504 | [-0.02 0.04] | 0.485750452 | 0.854965648 |
| IL_1RT2 | Bilirubin_pPS | 4225 | 0.026547276 | [-0. 0.06] | 0.084904258 | 0.536237419 |
| IL_1RT2 | Proinsulin_pPS | 4225 | 0.01276445 | [-0.02 0.04] | 0.407501732 | 0.813804543 |
| IL_1RT2 | Hyper_Insulin_pPS | 4225 | 0.012195326 | [-0.02 0.04] | 0.428731506 | 0.827992564 |
| IL_1RT2 | Beta_Cell_1_pPS | 4225 | 0.002951601 | [-0.03 0.03] | 0.84811432 | 0.97528582 |
| IL_1RT2 | Beta_Cell_2_pPS | 4225 | -0.000181498 | [-0.03 0.03] | 0.990603449 | 0.999402232 |
| IL_1RT2 | Total_GRS | 4225 | 0.022153683 | [-0.01 0.05] | 0.150521259 | 0.649125224 |
| ST2 | Cholesterol_pPS | 4225 | -0.000328817 | [-0.03 0.03] | 0.982977297 | 0.99799074 |
| ST2 | Lipodystrophy_1_pPS | 4225 | 0.013218219 | [-0.02 0.04] | 0.39103269 | 0.81106215 |
| ST2 | Lipodystrophy_2_pPS | 4225 | 0.012061155 | [-0.02 0.04] | 0.433828491 | 0.828998134 |
| ST2 | Liver-Lipid_pPS | 4225 | -0.023440301 | [-0.05 0.01] | 0.128206688 | 0.623575746 |
| ST2 | ASAT_Pos_pPS | 4225 | -0.035333289 | [-0.07 -0.01] | 0.021822948 | 0.317007038 |
| ST2 | ALP_Neg_pPS | 4225 | -0.022814945 | [-0.05 0.01] | 0.138708616 | 0.633834075 |
| ST2 | SHBG-LpA_pPS | 4225 | 0.008526144 | [-0.02 0.04] | 0.580087046 | 0.903823705 |
| ST2 | Obesity_pPS | 4225 | -0.003934943 | [-0.03 0.03] | 0.798465917 | 0.958074488 |
| ST2 | VAT_Neg_pPS | 4225 | 0.016535754 | [-0.01 0.05] | 0.283248047 | 0.749111537 |
| ST2 | Bilirubin_pPS | 4225 | 0.000121712 | [-0.03 0.03] | 0.993698603 | 0.999464614 |
| ST2 | Proinsulin_pPS | 4225 | 0.014000827 | [-0.02 0.04] | 0.363595465 | 0.798348037 |
| ST2 | Hyper_Insulin_pPS | 4225 | 0.007533731 | [-0.02 0.04] | 0.624941945 | 0.913580386 |
| ST2 | Beta_Cell_1_pPS | 4225 | 0.002697008 | [-0.03 0.03] | 0.861075889 | 0.977006629 |
| ST2 | Beta_Cell_2_pPS | 4225 | -0.021706433 | [-0.05 0.01] | 0.15893546 | 0.661140091 |
| ST2 | Total_GRS | 4225 | 0.001901474 | [-0.03 0.03] | 0.901803321 | 0.987003157 |
| IL1RL2 | Cholesterol_pPS | 4159 | -0.004627385 | [-0.04 0.03] | 0.765777655 | 0.952160028 |
| IL1RL2 | Lipodystrophy_1_pPS | 4159 | -0.002551529 | [-0.03 0.03] | 0.869524855 | 0.977006629 |
| IL1RL2 | Lipodystrophy_2_pPS | 4159 | -0.002859158 | [-0.03 0.03] | 0.853961166 | 0.976764533 |
| IL1RL2 | Liver-Lipid_pPS | 4159 | 0.008530791 | [-0.02 0.04] | 0.58286582 | 0.903823705 |
| IL1RL2 | ASAT_Pos_pPS | 4159 | 0.010165023 | [-0.02 0.04] | 0.512842151 | 0.870690038 |
| IL1RL2 | ALP_Neg_pPS | 4159 | -0.004845568 | [-0.04 0.03] | 0.755079448 | 0.950403646 |
| IL1RL2 | SHBG-LpA_pPS | 4159 | 0.006989957 | [-0.02 0.04] | 0.652707906 | 0.919948884 |
| IL1RL2 | Obesity_pPS | 4159 | -0.033654688 | [-0.06 -0. ] | 0.030217447 | 0.369452038 |
| IL1RL2 | VAT_Neg_pPS | 4159 | 0.02952359 | [-0. 0.06] | 0.057291808 | 0.45329986 |
| IL1RL2 | Bilirubin_pPS | 4159 | 0.011956364 | [-0.02 0.04] | 0.441447119 | 0.831350198 |
| IL1RL2 | Proinsulin_pPS | 4159 | 0.010068991 | [-0.02 0.04] | 0.516832412 | 0.870853148 |
| IL1RL2 | Hyper_Insulin_pPS | 4159 | -0.018120713 | [-0.05 0.01] | 0.243344756 | 0.731624757 |
| IL1RL2 | Beta_Cell_1_pPS | 4159 | 0.015868647 | [-0.01 0.05] | 0.306945278 | 0.76365449 |
| IL1RL2 | Beta_Cell_2_pPS | 4159 | -0.02060039 | [-0.05 0.01] | 0.184724341 | 0.686251968 |
| IL1RL2 | Total_GRS | 4159 | -0.031024628 | [-0.06 -0. ] | 0.045740745 | 0.430868454 |
| IL_1ra | Cholesterol_pPS | 4159 | 0.017587586 | [-0.01 0.05] | 0.257492553 | 0.740941247 |
| IL_1ra | Lipodystrophy_1_pPS | 4159 | 0.013894558 | [-0.02 0.04] | 0.371030831 | 0.799802236 |
| IL_1ra | Lipodystrophy_2_pPS | 4159 | 0.023512698 | [-0.01 0.05] | 0.130049974 | 0.626418724 |
| IL_1ra | Liver-Lipid_pPS | 4159 | 0.020221612 | [-0.01 0.05] | 0.192931492 | 0.695157855 |
| IL_1ra | ASAT_Pos_pPS | 4159 | 0.023158944 | [-0.01 0.05] | 0.135929561 | 0.629316669 |
| IL_1ra | ALP_Neg_pPS | 4159 | 0.000292978 | [-0.03 0.03] | 0.984951753 | 0.998701998 |
| IL_1ra | SHBG-LpA_pPS | 4159 | 0.001603047 | [-0.03 0.03] | 0.917803703 | 0.990056084 |
| IL_1ra | Obesity_pPS | 4159 | 0.047673484 | [0.02 0.08] | 0.002134435 | 0.082972391 |
| IL_1ra | VAT_Neg_pPS | 4159 | -0.008720804 | [-0.04 0.02] | 0.574499809 | 0.903411176 |
| IL_1ra | Bilirubin_pPS | 4159 | 0.010263729 | [-0.02 0.04] | 0.508757491 | 0.868660083 |
| IL_1ra | Proinsulin_pPS | 4159 | -0.020875945 | [-0.05 0.01] | 0.178918097 | 0.679661798 |
| IL_1ra | Hyper_Insulin_pPS | 4159 | 0.021229245 | [-0.01 0.05] | 0.171673609 | 0.674627844 |
| IL_1ra | Beta_Cell_1_pPS | 4159 | -0.013954068 | [-0.04 0.02] | 0.368985242 | 0.799230784 |
| IL_1ra | Beta_Cell_2_pPS | 4159 | -0.038224356 | [-0.07 -0.01] | 0.013828059 | 0.246172756 |
| IL_1ra | Total_GRS | 4159 | 0.027383553 | [-0. 0.06] | 0.077862839 | 0.514945086 |
| IL_27 | Cholesterol_pPS | 4159 | 0.023709811 | [-0.01 0.05] | 0.126860546 | 0.623575746 |
| IL_27 | Lipodystrophy_1_pPS | 4159 | 0.019266269 | [-0.01 0.05] | 0.214814551 | 0.715184753 |
| IL_27 | Lipodystrophy_2_pPS | 4159 | 0.013772105 | [-0.02 0.04] | 0.375262103 | 0.80226445 |
| IL_27 | Liver-Lipid_pPS | 4159 | -0.011374591 | [-0.04 0.02] | 0.463988682 | 0.841398661 |
| IL_27 | ASAT_Pos_pPS | 4159 | -0.011549787 | [-0.04 0.02] | 0.457133876 | 0.835860883 |
| IL_27 | ALP_Neg_pPS | 4159 | 0.009140186 | [-0.02 0.04] | 0.556238801 | 0.896216632 |
| IL_27 | SHBG-LpA_pPS | 4159 | 0.005272081 | [-0.03 0.04] | 0.734302909 | 0.94220178 |
| IL_27 | Obesity_pPS | 4159 | -0.007656101 | [-0.04 0.02] | 0.622090264 | 0.911342425 |
| IL_27 | VAT_Neg_pPS | 4159 | 0.017272693 | [-0.01 0.05] | 0.266112027 | 0.746520328 |
| IL_27 | Bilirubin_pPS | 4159 | -0.00071379 | [-0.03 0.03] | 0.963348324 | 0.99612031 |
| IL_27 | Proinsulin_pPS | 4159 | 0.021164635 | [-0.01 0.05] | 0.172981772 | 0.674627844 |
| IL_27 | Hyper_Insulin_pPS | 4159 | -0.000675819 | [-0.03 0.03] | 0.96529682 | 0.99612031 |
| IL_27 | Beta_Cell_1_pPS | 4159 | -0.013570907 | [-0.04 0.02] | 0.382278791 | 0.806027092 |
| IL_27 | Beta_Cell_2_pPS | 4159 | -0.034872844 | [-0.07 -0. ] | 0.024721968 | 0.333340061 |
| IL_27 | Total_GRS | 4159 | -0.011937023 | [-0.04 0.02] | 0.442186279 | 0.831350198 |
| IL2_RA | Cholesterol_pPS | 4225 | -0.002805833 | [-0.03 0.03] | 0.855530736 | 0.977006629 |
| IL2_RA | Lipodystrophy_1_pPS | 4225 | 0.03526066 | [0.01 0.07] | 0.022095728 | 0.31789781 |
| IL2_RA | Lipodystrophy_2_pPS | 4225 | 0.032013546 | [0. 0.06] | 0.03772417 | 0.394389052 |
| IL2_RA | Liver-Lipid_pPS | 4225 | 0.0080976 | [-0.02 0.04] | 0.599271356 | 0.907623171 |
| IL2_RA | ASAT_Pos_pPS | 4225 | 0.02178358 | [-0.01 0.05] | 0.157459225 | 0.657469682 |

**b) Partial Correlation**

| Proteomics | PPS | Sample_Size | r | CI95% | p | fdr_p_adjusted |
|---|---|---|---|---|---|---|
| IL1RL2 | Hyper_Insulin_pPS | 4159 | -0.018646936 | [-0.05 0.01] | 0.230656833 | 0.786988264 |
| IL1RL2 | Beta_Cell_1_pPS | 4159 | 0.013900541 | [-0.02 0.04] | 0.371576262 | 0.847284666 |
| IL1RL2 | Beta_Cell_2_pPS | 4159 | -0.027741013 | [-0.06 0. ] | 0.074513853 | 0.58879658 |
| IL_1ra | Cholesterol_pPS | 4159 | 0.011816215 | [-0.02 0.04] | 0.44753296 | 0.870351501 |
| IL_1ra | Lipodystrophy_1_pPS | 4159 | 0.005135451 | [-0.03 0.04] | 0.741330266 | 0.951867558 |
| IL_1ra | Lipodystrophy_2_pPS | 4159 | 0.021103403 | [-0.01 0.05] | 0.174905442 | 0.732083915 |
| IL_1ra | Liver-Lipid_pPS | 4159 | 0.019500479 | [-0.01 0.05] | 0.210006881 | 0.767305935 |
| IL_1ra | ASAT_Pos_pPS | 4159 | 0.016238937 | [-0.01 0.05] | 0.29655256 | 0.817900851 |
| IL_1ra | ALP_Neg_pPS | 4159 | 0.002441717 | [-0.03 0.03] | 0.875287494 | 0.97781424 |
| IL_1ra | SHBG-LpA_pPS | 4159 | 0.003425691 | [-0.03 0.03] | 0.825720919 | 0.970677755 |
| IL_1ra | Obesity_pPS | 4159 | 0.047063177 | [0.02 0.08] | 0.002472024 | 0.113713115 |
| IL_1ra | VAT_Neg_pPS | 4159 | -0.009532247 | [-0.04 0.02] | 0.540064296 | 0.899241939 |
| IL_1ra | Bilirubin_pPS | 4159 | 0.010586224 | [-0.02 0.04] | 0.49620876 | 0.895803896 |
| IL_1ra | Proinsulin_pPS | 4159 | -0.020663293 | [-0.05 0.01] | 0.184075365 | 0.738879686 |
| IL_1ra | Hyper_Insulin_pPS | 4159 | 0.015016904 | [-0.02 0.05] | 0.334398521 | 0.844649909 |
| IL_1ra | Beta_Cell_1_pPS | 4159 | 0.001064061 | [-0.03 0.03] | 0.945471669 | 0.996180826 |
| IL_1ra | Beta_Cell_2_pPS | 4159 | -0.034445834 | [-0.06 -0. ] | 0.026779062 | 0.401063155 |
| IL_27 | Cholesterol_pPS | 4159 | 0.024910552 | [-0.01 0.06] | 0.109284433 | 0.676681438 |
| IL_27 | Lipodystrophy_1_pPS | 4159 | 0.020741551 | [-0.01 0.05] | 0.182419333 | 0.738879686 |
| IL_27 | Lipodystrophy_2_pPS | 4159 | 0.010984757 | [-0.02 0.04] | 0.480134841 | 0.89059884 |
| IL_27 | Liver-Lipid_pPS | 4159 | -0.011765812 | [-0.04 0.02] | 0.449472763 | 0.870351501 |
| IL_27 | ASAT_Pos_pPS | 4159 | -0.015337005 | [-0.05 0.02] | 0.324196533 | 0.842714702 |
| IL_27 | ALP_Neg_pPS | 4159 | 0.004394493 | [-0.03 0.03] | 0.777586946 | 0.964867039 |
| IL_27 | SHBG-LpA_pPS | 4159 | 0.002155508 | [-0.03 0.03] | 0.889806366 | 0.981230818 |
| IL_27 | Obesity_pPS | 4159 | -0.012882749 | [-0.04 0.02] | 0.407616391 | 0.853674654 |
| IL_27 | VAT_Neg_pPS | 4159 | 0.019943802 | [-0.01 0.05] | 0.199825033 | 0.765145476 |
| IL_27 | Bilirubin_pPS | 4159 | -0.000597559 | [-0.03 0.03] | 0.969361462 | 0.996924797 |
| IL_27 | Proinsulin_pPS | 4159 | 0.029898139 | [-0. 0.06] | 0.054582518 | 0.551390458 |
| IL_27 | Hyper_Insulin_pPS | 4159 | -0.003183902 | [-0.03 0.03] | 0.837844504 | 0.972099644 |
| IL_27 | Beta_Cell_1_pPS | 4159 | -0.013022304 | [-0.04 0.02] | 0.402555018 | 0.85241904 |
| IL_27 | Beta_Cell_2_pPS | 4159 | -0.036818502 | [-0.07 -0.01] | 0.017914716 | 0.33440804 |
| IL2_RA | Cholesterol_pPS | 4225 | -0.006917878 | [-0.04 0.02] | 0.654007651 | 0.936605382 |
| IL2_RA | Lipodystrophy_1_pPS | 4225 | 0.033940765 | [0. 0.06] | 0.027835859 | 0.409743846 |
| IL2_RA | Lipodystrophy_2_pPS | 4225 | 0.03367398 | [0. 0.06] | 0.029087947 | 0.414433192 |
| IL2_RA | Liver-Lipid_pPS | 4225 | 0.007543563 | [-0.02 0.04] | 0.625025653 | 0.927058714 |
| IL2_RA | ASAT_Pos_pPS | 4225 | 0.013786085 | [-0.02 0.04] | 0.371741013 | 0.847284666 |
| IL2_RA | ALP_Neg_pPS | 4225 | 0.005885929 | [-0.02 0.04] | 0.70294983 | 0.945970482 |
| IL2_RA | SHBG-LpA_pPS | 4225 | -0.011923737 | [-0.04 0.02] | 0.439792328 | 0.866107938 |
| IL2_RA | Obesity_pPS | 4225 | 0.013560901 | [-0.02 0.04] | 0.379604289 | 0.85037326 |
| IL2_RA | VAT_Neg_pPS | 4225 | -0.034240216 | [-0.06 -0. ] | 0.026486063 | 0.399916393 |
| IL2_RA | Bilirubin_pPS | 4225 | 0.014371795 | [-0.02 0.04] | 0.351765849 | 0.847284666 |
| IL2_RA | Proinsulin_pPS | 4225 | 0.016834941 | [-0.01 0.05] | 0.275369079 | 0.808820203 |
| IL2_RA | Hyper_Insulin_pPS | 4225 | -0.022959527 | [-0.05 0.01] | 0.136829864 | 0.694065456 |
| IL2_RA | Beta_Cell_1_pPS | 4225 | -0.009136565 | [-0.04 0.02] | 0.553881936 | 0.899898936 |
| IL2_RA | Beta_Cell_2_pPS | 4225 | 0.013804896 | [-0.02 0.04] | 0.371088742 | 0.847284666 |
| IL_4RA | Cholesterol_pPS | 4159 | 0.020760385 | [-0.01 0.05] | 0.182022436 | 0.738879686 |
| IL_4RA | Lipodystrophy_1_pPS | 4159 | -0.00403413 | [-0.03 0.03] | 0.795402383 | 0.967638637 |
| IL_4RA | Lipodystrophy_2_pPS | 4159 | 0.011705071 | [-0.02 0.04] | 0.451816769 | 0.870351501 |
| IL_4RA | Liver-Lipid_pPS | 4159 | -0.013853815 | [-0.04 0.02] | 0.373186276 | 0.847284666 |
| IL_4RA | ASAT_Pos_pPS | 4159 | -0.027808673 | [-0.06 0. ] | 0.073808811 | 0.588265012 |
| IL_4RA | ALP_Neg_pPS | 4159 | 0.003494777 | [-0.03 0.03] | 0.822264323 | 0.968668398 |
| IL_4RA | SHBG-LpA_pPS | 4159 | 0.013198513 | [-0.02 0.04] | 0.396218343 | 0.85037326 |
| IL_4RA | Obesity_pPS | 4159 | -0.025356856 | [-0.06 0.01] | 0.103076498 | 0.660510098 |
| IL_4RA | VAT_Neg_pPS | 4159 | -0.002830229 | [-0.03 0.03] | 0.855646878 | 0.97556306 |
| IL_4RA | Bilirubin_pPS | 4159 | 0.004997811 | [-0.03 0.04] | 0.748024719 | 0.954135344 |
| IL_4RA | Proinsulin_pPS | 4159 | 0.011996488 | [-0.02 0.04] | 0.440634043 | 0.866107938 |
| IL_4RA | Hyper_Insulin_pPS | 4159 | 0.007124301 | [-0.02 0.04] | 0.647000799 | 0.935848959 |
| IL_4RA | Beta_Cell_1_pPS | 4159 | 0.024936958 | [-0.01 0.06] | 0.108909089 | 0.676681438 |
| IL_4RA | Beta_Cell_2_pPS | 4159 | -0.010024704 | [-0.04 0.02] | 0.519334524 | 0.895803896 |
| IL6 | Cholesterol_pPS | 4159 | -0.003665132 | [-0.03 0.03] | 0.813755843 | 0.967790883 |
| IL6 | Lipodystrophy_1_pPS | 4159 | -0.029594083 | [-0.06 0. ] | 0.057089095 | 0.558815902 |
| IL6 | Lipodystrophy_2_pPS | 4159 | 0.005619946 | [-0.02 0.04] | 0.717924027 | 0.945970482 |
| IL6 | Liver-Lipid_pPS | 4159 | 0.005648765 | [-0.02 0.04] | 0.716539786 | 0.945970482 |
| IL6 | ASAT_Pos_pPS | 4159 | 0.015579817 | [-0.01 0.05] | 0.316594451 | 0.839009314 |
| IL6 | ALP_Neg_pPS | 4159 | -0.006876806 | [-0.04 0.02] | 0.658471887 | 0.937489141 |
| IL6 | SHBG-LpA_pPS | 4159 | 0.001243681 | [-0.03 0.03] | 0.936285154 | 0.994163908 |
| IL6 | Obesity_pPS | 4159 | 0.053606966 | [0.02 0.08] | 0.000564418 | 0.041541171 |
| IL6 | VAT_Neg_pPS | 4159 | 0.005382738 | [-0.03 0.04] | 0.729352195 | 0.946981479 |
| IL6 | Bilirubin_pPS | 4159 | 0.013911634 | [-0.02 0.04] | 0.371194671 | 0.847284666 |
| IL6 | Proinsulin_pPS | 4159 | 0.014258593 | [-0.02 0.04] | 0.359382406 | 0.847284666 |
| IL6 | Hyper_Insulin_pPS | 4159 | 0.023314628 | [-0.01 0.05] | 0.133927573 | 0.694065456 |
| IL6 | Beta_Cell_1_pPS | 4159 | -0.002685506 | [-0.03 0.03] | 0.862953334 | 0.97556306 |
| IL6 | Beta_Cell_2_pPS | 4159 | -0.021496834 | [-0.05 0.01] | 0.167000499 | 0.722132896 |
| IL_6RA | Cholesterol_pPS | 4225 | -0.005513398 | [-0.04 0.02] | 0.720938043 | 0.946314812 |
| IL_6RA | Lipodystrophy_1_pPS | 4225 | -0.003467494 | [-0.03 0.03] | 0.822249187 | 0.968668398 |
| IL_6RA | Lipodystrophy_2_pPS | 4225 | -0.010260886 | [-0.04 0.02] | 0.506177792 | 0.895803896 |
| IL_6RA | Liver-Lipid_pPS | 4225 | -0.029281149 | [-0.06 0. ] | 0.057764967 | 0.558815902 |
| IL_6RA | ASAT_Pos_pPS | 4225 | 0.005867311 | [-0.02 0.04] | 0.70384496 | 0.945970482 |
| IL_6RA | ALP_Neg_pPS | 4225 | -0.002614184 | [-0.03 0.03] | 0.865506906 | 0.97556306 |
| IL_6RA | SHBG-LpA_pPS | 4225 | -0.00408003 | [-0.03 0.03] | 0.791519056 | 0.967638637 |
| IL_6RA | Obesity_pPS | 4225 | 0.024967957 | [-0.01 0.06] | 0.105688657 | 0.665922758 |
| IL_6RA | VAT_Neg_pPS | 4225 | -0.030464066 | [-0.06 -0. ] | 0.048362683 | 0.520248282 |
| IL_6RA | Bilirubin_pPS | 4225 | 0.01747722 | [-0.01 0.05] | 0.257465181 | 0.80473167 |
| IL_6RA | Proinsulin_pPS | 4225 | 0.0035826 | [-0.03 0.03] | 0.816452067 | 0.968668398 |
| IL_6RA | Hyper_Insulin_pPS | 4225 | -0.016122788 | [-0.05 0.01] | 0.296192639 | 0.817900851 |
| IL_6RA | Beta_Cell_1_pPS | 4225 | 0.021064674 | [-0.01 0.05] | 0.172287008 | 0.727559563 |
| IL_6RA | Beta_Cell_2_pPS | 4225 | 0.007848816 | [-0.02 0.04] | 0.611090224 | 0.919490897 |
| ITGB2 | Cholesterol_pPS | 4225 | -0.006110304 | [-0.04 0.02] | 0.692194461 | 0.945970482 |
| ITGB2 | Lipodystrophy_1_pPS | 4225 | 0.004642327 | [-0.03 0.03] | 0.763590457 | 0.959095176 |
| ITGB2 | Lipodystrophy_2_pPS | 4225 | 0.004930677 | [-0.03 0.04] | 0.749384363 | 0.954135344 |
| ITGB2 | Liver-Lipid_pPS | 4225 | 0.023451265 | [-0.01 0.05] | 0.128618522 | 0.694065456 |
| ITGB2 | ASAT_Pos_pPS | 4225 | 0.020152559 | [-0.01 0.05] | 0.191627756 | 0.750920235 |
| ITGB2 | ALP_Neg_pPS | 4225 | -0.004047732 | [-0.03 0.03] | 0.793131791 | 0.967638637 |
| ITGB2 | SHBG-LpA_pPS | 4225 | 0.020622469 | [-0.01 0.05] | 0.181473129 | 0.738879686 |
| ITGB2 | Obesity_pPS | 4225 | 0.009217458 | [-0.02 0.04] | 0.550377596 | 0.899241939 |
| ITGB2 | VAT_Neg_pPS | 4225 | -0.006969996 | [-0.04 0.02] | 0.65157274 | 0.936605382 |
| ITGB2 | Bilirubin_pPS | 4225 | -0.008012079 | [-0.04 0.02] | 0.603693932 | 0.918301271 |
| ITGB2 | Proinsulin_pPS | 4225 | 0.012492294 | [-0.02 0.04] | 0.418294434 | 0.858586823 |
| ITGB2 | Hyper_Insulin_pPS | 4225 | 0.010555228 | [-0.02 0.04] | 0.494055759 | 0.895803896 |
| ITGB2 | Beta_Cell_1_pPS | 4225 | 0.017850895 | [-0.01 0.05] | 0.247428459 | 0.796719639 |
| ITGB2 | Beta_Cell_2_pPS | 4225 | 0.022570681 | [-0.01 0.05] | 0.143604745 | 0.697681852 |
| LDL_receptor | Cholesterol_pPS | 4225 | -0.024266179 | [-0.05 0.01] | 0.115859768 | 0.681046037 |
| LDL_receptor | Lipodystrophy_1_pPS | 4225 | 0.066788538 | [0.04 0.1 ] | 1.48E-05 | 0.002003866 |
| LDL_receptor | Lipodystrophy_2_pPS | 4225 | 0.002332525 | [-0.03 0.03] | 0.879880982 | 0.97781424 |
| LDL_receptor | Liver-Lipid_pPS | 4225 | -0.039957262 | [-0.07 -0.01] | 0.009603452 | 0.248521918 |
| LDL_receptor | ASAT_Pos_pPS | 4225 | 0.01671185 | [-0.01 0.05] | 0.278895035 | 0.808820203 |
| LDL_receptor | ALP_Neg_pPS | 4225 | -0.019227724 | [-0.05 0.01] | 0.212821982 | 0.767305935 |

**Table S7: Associations between pPSs and plasma proteins in SCAPIS**

**a) Spearman Correlation**

| Proteomics | PPS | Sample_Size | r | CI95% | p | fdr_p_adjusted |
|---|---|---|---|---|---|---|
| IL2_RA | ALP_Neg_pPS | 4225 | 0.010264591 | [-0.02 0.04] | 0.505366299 | 0.867177846 |
| IL2_RA | SHBG-LpA_pPS | 4225 | -0.011176653 | [-0.04 0.02] | 0.468294115 | 0.843663028 |
| IL2_RA | Obesity_pPS | 4225 | 0.01623469 | [-0.01 0.05] | 0.292106421 | 0.75346579 |
| IL2_RA | VAT_Neg_pPS | 4225 | -0.031157997 | [-0.06 -0. ] | 0.043147178 | 0.419317643 |
| IL2_RA | Bilirubin_pPS | 4225 | 0.015861574 | [-0.01 0.05] | 0.303340429 | 0.76319014 |
| IL2_RA | Proinsulin_pPS | 4225 | 0.019721493 | [-0.01 0.05] | 0.200608474 | 0.700947446 |
| IL2_RA | Hyper_Insulin_pPS | 4225 | -0.017455207 | [-0.05 0.01] | 0.257329268 | 0.740941247 |
| IL2_RA | Beta_Cell_1_pPS | 4225 | -0.003399962 | [-0.03 0.03] | 0.825389414 | 0.964468579 |
| IL2_RA | Beta_Cell_2_pPS | 4225 | 0.012854794 | [-0.02 0.04] | 0.40419025 | 0.81230319 |
| IL2_RA | Total_GRS | 4225 | 0.02720762 | [-0. 0.06] | 0.077431952 | 0.514390694 |
| IL_4RA | Cholesterol_pPS | 4159 | 0.018498529 | [-0.01 0.05] | 0.233655028 | 0.726631332 |
| IL_4RA | Lipodystrophy_1_pPS | 4159 | -0.006672722 | [-0.04 0.02] | 0.667501275 | 0.924640793 |
| IL_4RA | Lipodystrophy_2_pPS | 4159 | 0.008797529 | [-0.02 0.04] | 0.571137895 | 0.902829662 |
| IL_4RA | Liver-Lipid_pPS | 4159 | -0.012372209 | [-0.04 0.02] | 0.425726392 | 0.82738773 |
| IL_4RA | ASAT_Pos_pPS | 4159 | -0.027494231 | [-0.06 0. ] | 0.076668434 | 0.511887462 |
| IL_4RA | ALP_Neg_pPS | 4159 | 0.004701182 | [-0.03 0.04] | 0.762154041 | 0.95054006 |
| IL_4RA | SHBG-LpA_pPS | 4159 | 0.013085116 | [-0.02 0.04] | 0.399549028 | 0.81230319 |
| IL_4RA | Obesity_pPS | 4159 | -0.022787544 | [-0.05 0.01] | 0.142321048 | 0.640622936 |
| IL_4RA | VAT_Neg_pPS | 4159 | 0.00131214 | [-0.03 0.03] | 0.932680586 | 0.990208704 |
| IL_4RA | Bilirubin_pPS | 4159 | 0.005097055 | [-0.03 0.04] | 0.742806345 | 0.945527195 |
| IL_4RA | Proinsulin_pPS | 4159 | 0.016857196 | [-0.01 0.05] | 0.277786089 | 0.747422497 |
| IL_4RA | Hyper_Insulin_pPS | 4159 | 0.003526277 | [-0.03 0.03] | 0.820412387 | 0.963461584 |
| IL_4RA | Beta_Cell_1_pPS | 4159 | 0.025390738 | [-0.01 0.06] | 0.102077733 | 0.586842904 |
| IL_4RA | Beta_Cell_2_pPS | 4159 | -0.000986785 | [-0.03 0.03] | 0.949346824 | 0.993154165 |
| IL_4RA | Total_GRS | 4159 | -0.002534958 | [-0.03 0.03] | 0.870364683 | 0.977006629 |
| IL6 | Cholesterol_pPS | 4159 | 0.001985137 | [-0.03 0.03] | 0.898308165 | 0.984642786 |
| IL6 | Lipodystrophy_1_pPS | 4159 | -0.021514469 | [-0.05 0.01] | 0.165986947 | 0.664911429 |
| IL6 | Lipodystrophy_2_pPS | 4159 | 0.007476947 | [-0.02 0.04] | 0.630263471 | 0.914460114 |
| IL6 | Liver-Lipid_pPS | 4159 | 0.006887029 | [-0.02 0.04] | 0.657493072 | 0.922711159 |
| IL6 | ASAT_Pos_pPS | 4159 | 0.015886676 | [-0.01 0.05] | 0.306395949 | 0.76365449 |
| IL6 | ALP_Neg_pPS | 4159 | -0.003551971 | [-0.03 0.03] | 0.81912635 | 0.963461584 |
| IL6 | SHBG-LpA_pPS | 4159 | -4.80E-05 | [-0.03 0.03] | 0.997532449 | 0.999464614 |
| IL6 | Obesity_pPS | 4159 | 0.056427162 | [0.03 0.09] | 0.000277385 | 0.019630339 |
| IL6 | VAT_Neg_pPS | 4159 | 0.006115392 | [-0.02 0.04] | 0.693803185 | 0.930013012 |
| IL6 | Bilirubin_pPS | 4159 | 0.01378337 | [-0.02 0.04] | 0.374871624 | 0.80226445 |
| IL6 | Proinsulin_pPS | 4159 | 0.016817149 | [-0.01 0.05] | 0.27892941 | 0.747422497 |
| IL6 | Hyper_Insulin_pPS | 4159 | 0.025021369 | [-0.01 0.06] | 0.107163885 | 0.597786172 |
| IL6 | Beta_Cell_1_pPS | 4159 | -0.00213099 | [-0.03 0.03] | 0.890881757 | 0.981178631 |
| IL6 | Beta_Cell_2_pPS | 4159 | -0.018221921 | [-0.05 0.01] | 0.240721845 | 0.729918816 |
| IL6 | Total_GRS | 4159 | 0.032512378 | [0. 0.06] | 0.036293083 | 0.386599178 |
| IL_6RA | Cholesterol_pPS | 4225 | -0.003892524 | [-0.03 0.03] | 0.800592435 | 0.958074488 |
| IL_6RA | Lipodystrophy_1_pPS | 4225 | -0.00460984 | [-0.03 0.03] | 0.764843448 | 0.952160028 |
| IL_6RA | Lipodystrophy_2_pPS | 4225 | -0.016613403 | [-0.05 0.01] | 0.280993166 | 0.748127084 |
| IL_6RA | Liver-Lipid_pPS | 4225 | -0.029837676 | [-0.06 0. ] | 0.052800471 | 0.441027526 |
| IL_6RA | ASAT_Pos_pPS | 4225 | 0.004673366 | [-0.03 0.03] | 0.761700313 | 0.950403646 |
| IL_6RA | ALP_Neg_pPS | 4225 | -0.002996483 | [-0.03 0.03] | 0.845833451 | 0.974332356 |
| IL_6RA | SHBG-LpA_pPS | 4225 | -0.00666531 | [-0.04 0.02] | 0.665373775 | 0.924111434 |
| IL_6RA | Obesity_pPS | 4225 | 0.024555259 | [-0.01 0.05] | 0.111027699 | 0.604978148 |
| IL_6RA | VAT_Neg_pPS | 4225 | -0.03066748 | [-0.06 -0. ] | 0.046543503 | 0.434802524 |
| IL_6RA | Bilirubin_pPS | 4225 | 0.019190985 | [-0.01 0.05] | 0.212989763 | 0.712633026 |
| IL_6RA | Proinsulin_pPS | 4225 | 0.012814525 | [-0.02 0.04] | 0.405664279 | 0.812806898 |
| IL_6RA | Hyper_Insulin_pPS | 4225 | -0.019465222 | [-0.05 0.01] | 0.206522291 | 0.709139138 |
| IL_6RA | Beta_Cell_1_pPS | 4225 | 0.022798676 | [-0.01 0.05] | 0.138990401 | 0.634028726 |
| IL_6RA | Beta_Cell_2_pPS | 4225 | 0.011737531 | [-0.02 0.04] | 0.446265681 | 0.831350198 |
| IL_6RA | Total_GRS | 4225 | 7.23E-05 | [-0.03 0.03] | 0.99625583 | 0.999464614 |
| ITGB2 | Cholesterol_pPS | 4225 | -0.004325606 | [-0.03 0.03] | 0.778953707 | 0.953637204 |
| ITGB2 | Lipodystrophy_1_pPS | 4225 | 0.011998072 | [-0.02 0.04] | 0.436237016 | 0.829210857 |
| ITGB2 | Lipodystrophy_2_pPS | 4225 | 0.007819901 | [-0.02 0.04] | 0.611854314 | 0.909483337 |
| ITGB2 | Liver-Lipid_pPS | 4225 | 0.02580035 | [-0. 0.06] | 0.094047841 | 0.560630757 |
| ITGB2 | ASAT_Pos_pPS | 4225 | 0.023824316 | [-0.01 0.05] | 0.122070671 | 0.615837655 |
| ITGB2 | ALP_Neg_pPS | 4225 | -0.000502191 | [-0.03 0.03] | 0.97400444 | 0.996862516 |
| ITGB2 | SHBG-LpA_pPS | 4225 | 0.022117988 | [-0.01 0.05] | 0.15118008 | 0.649125224 |
| ITGB2 | Obesity_pPS | 4225 | 0.013046738 | [-0.02 0.04] | 0.397208344 | 0.812191725 |
| ITGB2 | VAT_Neg_pPS | 4225 | 0.001423679 | [-0.03 0.03] | 0.926395937 | 0.990056084 |
| ITGB2 | Bilirubin_pPS | 4225 | -0.006487127 | [-0.04 0.02] | 0.673796347 | 0.925361755 |
| ITGB2 | Proinsulin_pPS | 4225 | 0.025221458 | [-0. 0.06] | 0.101663998 | 0.586731767 |
| ITGB2 | Hyper_Insulin_pPS | 4225 | 0.015167673 | [-0.02 0.05] | 0.324986655 | 0.775125841 |
| ITGB2 | Beta_Cell_1_pPS | 4225 | 0.03014534 | [-0. 0.06] | 0.050403168 | 0.440215663 |
| ITGB2 | Beta_Cell_2_pPS | 4225 | 0.031495649 | [0. 0.06] | 0.040932971 | 0.407062052 |
| ITGB2 | Total_GRS | 4225 | 0.040618668 | [0.01 0.07] | 0.008370026 | 0.197592038 |
| LDL_receptor | Cholesterol_pPS | 4225 | -0.022703433 | [-0.05 0.01] | 0.140648988 | 0.638159106 |
| LDL_receptor | Lipodystrophy_1_pPS | 4225 | 0.072518999 | [0.04 0.1 ] | 2.45E-06 | 0.000376411 |
| LDL_receptor | Lipodystrophy_2_pPS | 4225 | 0.005661957 | [-0.02 0.04] | 0.71332211 | 0.940644541 |
| LDL_receptor | Liver-Lipid_pPS | 4225 | -0.039403882 | [-0.07 -0.01] | 0.010532247 | 0.223701034 |
| LDL_receptor | ASAT_Pos_pPS | 4225 | 0.030540731 | [0. 0.06] | 0.047456807 | 0.436063541 |
| LDL_receptor | ALP_Neg_pPS | 4225 | -0.018214217 | [-0.05 0.01] | 0.237210126 | 0.726631332 |
| LDL_receptor | SHBG-LpA_pPS | 4225 | 0.012584882 | [-0.02 0.04] | 0.414131449 | 0.817598568 |
| LDL_receptor | Obesity_pPS | 4225 | 0.027156732 | [-0. 0.06] | 0.07798806 | 0.514945086 |
| LDL_receptor | VAT_Neg_pPS | 4225 | 0.001438224 | [-0.03 0.03] | 0.925646103 | 0.990056084 |
| LDL_receptor | Bilirubin_pPS | 4225 | -0.001981019 | [-0.03 0.03] | 0.897717533 | 0.984642786 |
| LDL_receptor | Proinsulin_pPS | 4225 | 0.00974755 | [-0.02 0.04] | 0.527048694 | 0.873942847 |
| LDL_receptor | Hyper_Insulin_pPS | 4225 | 0.011825744 | [-0.02 0.04] | 0.442855599 | 0.831350198 |
| LDL_receptor | Beta_Cell_1_pPS | 4225 | 0.014542576 | [-0.02 0.04] | 0.345326088 | 0.790391668 |
| LDL_receptor | Beta_Cell_2_pPS | 4225 | 0.013473107 | [-0.02 0.04] | 0.381961657 | 0.806027092 |
| LDL_receptor | Total_GRS | 4225 | 0.038411162 | [0.01 0.07] | 0.012654132 | 0.242414912 |
| LEP | Cholesterol_pPS | 4159 | -0.003486295 | [-0.03 0.03] | 0.822414435 | 0.963461584 |
| LEP | Lipodystrophy_1_pPS | 4159 | -0.01565503 | [-0.05 0.01] | 0.313503425 | 0.76365449 |
| LEP | Lipodystrophy_2_pPS | 4159 | 0.014025506 | [-0.02 0.04] | 0.366538899 | 0.798890546 |
| LEP | Liver-Lipid_pPS | 4159 | 0.055818909 | [0.03 0.09] | 0.000322773 | 0.022271342 |
| LEP | ASAT_Pos_pPS | 4159 | 0.050939227 | [0.02 0.08] | 0.001032619 | 0.050992277 |
| LEP | ALP_Neg_pPS | 4159 | 0.010417551 | [-0.02 0.04] | 0.502426258 | 0.864575328 |
| LEP | SHBG-LpA_pPS | 4159 | 0.000822536 | [-0.03 0.03] | 0.957769287 | 0.99612031 |
| LEP | Obesity_pPS | 4159 | 0.067067359 | [0.04 0.1 ] | 1.54E-05 | 0.002027419 |
| LEP | VAT_Neg_pPS | 4159 | 0.001616991 | [-0.03 0.03] | 0.917091287 | 0.990056084 |
| LEP | Bilirubin_pPS | 4159 | -0.015487831 | [-0.05 0.01] | 0.318700369 | 0.768220977 |
| LEP | Proinsulin_pPS | 4159 | 5.21E-05 | [-0.03 0.03] | 0.997324258 | 0.999464614 |
| LEP | Hyper_Insulin_pPS | 4159 | 0.011721452 | [-0.02 0.04] | 0.450472744 | 0.831350198 |
| LEP | Beta_Cell_1_pPS | 4159 | -0.017010052 | [-0.05 0.01] | 0.273451457 | 0.746520328 |
| LEP | Beta_Cell_2_pPS | 4159 | -0.030607834 | [-0.06 -0. ] | 0.048732281 | 0.438114317 |
| LEP | Total_GRS | 4159 | 0.018577878 | [-0.01 0.05] | 0.231655205 | 0.726631332 |
| Gal_3 | Cholesterol_pPS | 4225 | -0.014149928 | [-0.04 0.02] | 0.358508051 | 0.7979754 |
| Gal_3 | Lipodystrophy_1_pPS | 4225 | 0.017015117 | [-0.01 0.05] | 0.269522414 | 0.746520328 |
| Gal_3 | Lipodystrophy_2_pPS | 4225 | 0.01005873 | [-0.02 0.04] | 0.513942574 | 0.870853148 |
| Gal_3 | Liver-Lipid_pPS | 4225 | -0.030885104 | [-0.06 -0. ] | 0.04500988 | 0.425435853 |
| Gal_3 | ASAT_Pos_pPS | 4225 | 0.002296065 | [-0.03 0.03] | 0.881563042 | 0.979291846 |
| Gal_3 | ALP_Neg_pPS | 4225 | -0.045557122 | [-0.08 -0.02] | 0.003099811 | 0.105623173 |
| Gal_3 | SHBG-LpA_pPS | 4225 | 0.014785938 | [-0.02 0.04] | 0.337313055 | 0.782502359 |

**b) Partial Correlation**

| Proteomics | PPS | Sample_Size | r | CI95% | p | fdr_p_adjusted |
|---|---|---|---|---|---|---|
| LDL_receptor | SHBG-LpA_pPS | 4225 | 0.006672337 | [-0.02 0.04] | 0.665528494 | 0.937489141 |
| LDL_receptor | Obesity_pPS | 4225 | 0.025483232 | [-0. 0.06] | 0.098681665 | 0.648957106 |
| LDL_receptor | VAT_Neg_pPS | 4225 | -0.004264825 | [-0.03 0.03] | 0.782309006 | 0.964905159 |
| LDL_receptor | Bilirubin_pPS | 4225 | -0.005680042 | [-0.04 0.02] | 0.712871622 | 0.945970482 |
| LDL_receptor | Proinsulin_pPS | 4225 | 0.005910098 | [-0.02 0.04] | 0.70178837 | 0.945970482 |
| LDL_receptor | Hyper_Insulin_pPS | 4225 | 0.002731338 | [-0.03 0.03] | 0.859540951 | 0.97556306 |
| LDL_receptor | Beta_Cell_1_pPS | 4225 | 0.007778876 | [-0.02 0.04] | 0.614270951 | 0.921048876 |
| LDL_receptor | Beta_Cell_2_pPS | 4225 | 0.011226871 | [-0.02 0.04] | 0.466986629 | 0.879984873 |
| LEP | Cholesterol_pPS | 4159 | -0.011145603 | [-0.04 0.02] | 0.473728741 | 0.88406027 |
| LEP | Lipodystrophy_1_pPS | 4159 | -0.028306608 | [-0.06 0. ] | 0.068786228 | 0.571035672 |
| LEP | Lipodystrophy_2_pPS | 4159 | 0.00926062 | [-0.02 0.04] | 0.551672831 | 0.899241939 |
| LEP | Liver-Lipid_pPS | 4159 | 0.055137553 | [0.02 0.09] | 0.000389999 | 0.030443542 |
| LEP | ASAT_Pos_pPS | 4159 | 0.051006385 | [0.02 0.08] | 0.001035645 | 0.066695564 |
| LEP | ALP_Neg_pPS | 4159 | 0.008707642 | [-0.02 0.04] | 0.575677603 | 0.909113996 |
| LEP | SHBG-LpA_pPS | 4159 | 0.003061057 | [-0.03 0.03] | 0.84401906 | 0.972178273 |
| LEP | Obesity_pPS | 4159 | 0.066457504 | [0.04 0.1 ] | 1.90E-05 | 0.002446424 |
| LEP | VAT_Neg_pPS | 4159 | 0.0030529 | [-0.03 0.03] | 0.844429424 | 0.972178273 |
| LEP | Bilirubin_pPS | 4159 | -0.015047119 | [-0.05 0.02] | 0.33342676 | 0.844649909 |
| LEP | Proinsulin_pPS | 4159 | -0.002321056 | [-0.03 0.03] | 0.881403609 | 0.978445416 |
| LEP | Hyper_Insulin_pPS | 4159 | 0.003998744 | [-0.03 0.03] | 0.797157739 | 0.967638637 |
| LEP | Beta_Cell_1_pPS | 4159 | -0.011512048 | [-0.04 0.02] | 0.459311229 | 0.874172623 |
| LEP | Beta_Cell_2_pPS | 4159 | -0.028551222 | [-0.06 0. ] | 0.066423676 | 0.563434057 |
| Gal_3 | Cholesterol_pPS | 4225 | -0.015029718 | [-0.05 0.02] | 0.330154809 | 0.84456682 |
| Gal_3 | Lipodystrophy_1_pPS | 4225 | 0.016891005 | [-0.01 0.05] | 0.273773248 | 0.808820203 |
| Gal_3 | Lipodystrophy_2_pPS | 4225 | 0.011232342 | [-0.02 0.04] | 0.466769566 | 0.879984873 |
| Gal_3 | Liver-Lipid_pPS | 4225 | -0.030373768 | [-0.06 -0. ] | 0.049032001 | 0.524093094 |
| Gal_3 | ASAT_Pos_pPS | 4225 | -0.001671727 | [-0.03 0.03] | 0.913751298 | 0.989461462 |
| Gal_3 | ALP_Neg_pPS | 4225 | -0.050307601 | [-0.08 -0.02] | 0.001108636 | 0.069654785 |
| Gal_3 | SHBG-LpA_pPS | 4225 | 0.010706568 | [-0.02 0.04] | 0.487883988 | 0.895803896 |
| Gal_3 | Obesity_pPS | 4225 | 0.000742894 | [-0.03 0.03] | 0.961612062 | 0.996924797 |
| Gal_3 | VAT_Neg_pPS | 4225 | -0.015435356 | [-0.05 0.01] | 0.317268317 | 0.839009314 |
| Gal_3 | Bilirubin_pPS | 4225 | -0.014645855 | [-0.04 0.02] | 0.34265711 | 0.847284666 |
| Gal_3 | Proinsulin_pPS | 4225 | 0.035863764 | [0.01 0.07] | 0.020109775 | 0.347669668 |
| Gal_3 | Hyper_Insulin_pPS | 4225 | 0.007408003 | [-0.02 0.04] | 0.631257846 | 0.930807219 |
| Gal_3 | Beta_Cell_1_pPS | 4225 | -0.007612508 | [-0.04 0.02] | 0.621866168 | 0.924343732 |
| Gal_3 | Beta_Cell_2_pPS | 4225 | 0.015072788 | [-0.02 0.05] | 0.328770691 | 0.84456682 |
| Gal_9 | Cholesterol_pPS | 4159 | 0.014599366 | [-0.02 0.05] | 0.348013334 | 0.847284666 |
| Gal_9 | Lipodystrophy_1_pPS | 4159 | 0.009264613 | [-0.02 0.04] | 0.551501301 | 0.899241939 |
| Gal_9 | Lipodystrophy_2_pPS | 4159 | 0.013404113 | [-0.02 0.04] | 0.388901375 | 0.85037326 |
| Gal_9 | Liver-Lipid_pPS | 4159 | 0.003973948 | [-0.03 0.03] | 0.798388326 | 0.967638637 |
| Gal_9 | ASAT_Pos_pPS | 4159 | 0.001988191 | [-0.03 0.03] | 0.898311544 | 0.985399384 |
| Gal_9 | ALP_Neg_pPS | 4159 | -0.089527369 | [-0.12 -0.06] | 8.11E-09 | 1.61E-06 |
| Gal_9 | SHBG-LpA_pPS | 4159 | 0.013232529 | [-0.02 0.04] | 0.395002069 | 0.85037326 |
| Gal_9 | Obesity_pPS | 4159 | 0.03947991 | [0.01 0.07] | 0.01112875 | 0.267922059 |
| Gal_9 | VAT_Neg_pPS | 4159 | -0.001111951 | [-0.03 0.03] | 0.943021645 | 0.9959917 |
| Gal_9 | Bilirubin_pPS | 4159 | 0.009353871 | [-0.02 0.04] | 0.547673798 | 0.899241939 |
| Gal_9 | Proinsulin_pPS | 4159 | 0.009328271 | [-0.02 0.04] | 0.548770208 | 0.899241939 |
| Gal_9 | Hyper_Insulin_pPS | 4159 | -0.000173148 | [-0.03 0.03] | 0.991120236 | 0.998803911 |
| Gal_9 | Beta_Cell_1_pPS | 4159 | 0.004200222 | [-0.03 0.03] | 0.787177317 | 0.967638637 |
| Gal_9 | Beta_Cell_2_pPS | 4159 | -0.024260358 | [-0.05 0.01] | 0.118852925 | 0.681046037 |
| LTBR | Cholesterol_pPS | 4225 | 0.005426878 | [-0.02 0.04] | 0.725138431 | 0.946981479 |
| LTBR | Lipodystrophy_1_pPS | 4225 | 0.027921738 | [-0. 0.06] | 0.070396713 | 0.573866874 |
| LTBR | Lipodystrophy_2_pPS | 4225 | 0.004741511 | [-0.03 0.03] | 0.758694722 | 0.956163211 |
| LTBR | Liver-Lipid_pPS | 4225 | 0.017080636 | [-0.01 0.05] | 0.268422386 | 0.808113719 |
| LTBR | ASAT_Pos_pPS | 4225 | -0.006075596 | [-0.04 0.02] | 0.693854176 | 0.945970482 |
| LTBR | ALP_Neg_pPS | 4225 | 0.007413447 | [-0.02 0.04] | 0.631007078 | 0.930807219 |
| LTBR | SHBG-LpA_pPS | 4225 | -0.004714846 | [-0.03 0.03] | 0.760009985 | 0.956884516 |
| LTBR | Obesity_pPS | 4225 | 0.005228455 | [-0.03 0.04] | 0.734802527 | 0.948434512 |
| LTBR | VAT_Neg_pPS | 4225 | -0.039803257 | [-0.07 -0.01] | 0.009885746 | 0.248683454 |
| LTBR | Bilirubin_pPS | 4225 | 0.001604461 | [-0.03 0.03] | 0.917208983 | 0.99041009 |
| LTBR | Proinsulin_pPS | 4225 | 0.015421447 | [-0.01 0.05] | 0.317704639 | 0.839009314 |
| LTBR | Hyper_Insulin_pPS | 4225 | -0.000861117 | [-0.03 0.03] | 0.955508951 | 0.996425456 |
| LTBR | Beta_Cell_1_pPS | 4225 | -0.005047277 | [-0.04 0.03] | 0.743663621 | 0.952599446 |
| LTBR | Beta_Cell_2_pPS | 4225 | -0.002259542 | [-0.03 0.03] | 0.883612208 | 0.979373526 |
| MARCO | Cholesterol_pPS | 4159 | 0.014063671 | [-0.02 0.04] | 0.3659892 | 0.847284666 |
| MARCO | Lipodystrophy_1_pPS | 4159 | 0.026990913 | [-0. 0.06] | 0.082704656 | 0.614725404 |
| MARCO | Lipodystrophy_2_pPS | 4159 | 0.021388359 | [-0.01 0.05] | 0.169152712 | 0.723816256 |
| MARCO | Liver-Lipid_pPS | 4159 | 0.001763236 | [-0.03 0.03] | 0.909764863 | 0.988841471 |
| MARCO | ASAT_Pos_pPS | 4159 | -0.012259467 | [-0.04 0.02] | 0.43068017 | 0.864060889 |
| MARCO | ALP_Neg_pPS | 4159 | -0.027319037 | [-0.06 0. ] | 0.079036075 | 0.603132929 |
| MARCO | SHBG-LpA_pPS | 4159 | 0.014566792 | [-0.02 0.05] | 0.349090093 | 0.847284666 |
| MARCO | Obesity_pPS | 4159 | 0.024411099 | [-0.01 0.05] | 0.116578092 | 0.681046037 |
| MARCO | VAT_Neg_pPS | 4159 | 0.000864653 | [-0.03 0.03] | 0.955678712 | 0.996425456 |
| MARCO | Bilirubin_pPS | 4159 | 0.008279327 | [-0.02 0.04] | 0.594603186 | 0.918284057 |
| MARCO | Proinsulin_pPS | 4159 | -0.006664088 | [-0.04 0.02] | 0.668395866 | 0.937489141 |
| MARCO | Hyper_Insulin_pPS | 4159 | 0.003347141 | [-0.03 0.03] | 0.829655127 | 0.971458352 |
| MARCO | Beta_Cell_1_pPS | 4159 | 0.014582786 | [-0.02 0.05] | 0.348561126 | 0.847284666 |
| MARCO | Beta_Cell_2_pPS | 4159 | 0.014492745 | [-0.02 0.04] | 0.351545654 | 0.847284666 |
| MMP_3 | Cholesterol_pPS | 4225 | -0.000708766 | [-0.03 0.03] | 0.963374312 | 0.996924797 |
| MMP_3 | Lipodystrophy_1_pPS | 4225 | 0.033518457 | [0. 0.06] | 0.029839976 | 0.416452728 |
| MMP_3 | Lipodystrophy_2_pPS | 4225 | 0.012990062 | [-0.02 0.04] | 0.399991075 | 0.85241904 |
| MMP_3 | Liver-Lipid_pPS | 4225 | -0.014100672 | [-0.04 0.02] | 0.360926321 | 0.847284666 |
| MMP_3 | ASAT_Pos_pPS | 4225 | -0.003665212 | [-0.03 0.03] | 0.812297622 | 0.967638637 |
| MMP_3 | ALP_Neg_pPS | 4225 | 0.013260532 | [-0.02 0.04] | 0.390250785 | 0.85037326 |
| MMP_3 | SHBG-LpA_pPS | 4225 | 0.016840336 | [-0.01 0.05] | 0.275215242 | 0.808820203 |
| MMP_3 | Obesity_pPS | 4225 | -0.004181917 | [-0.03 0.03] | 0.786437392 | 0.967460707 |
| MMP_3 | VAT_Neg_pPS | 4225 | -0.021489958 | [-0.05 0.01] | 0.163784762 | 0.718997225 |
| MMP_3 | Bilirubin_pPS | 4225 | -0.003710141 | [-0.03 0.03] | 0.81004044 | 0.967638637 |
| MMP_3 | Proinsulin_pPS | 4225 | 0.011753582 | [-0.02 0.04] | 0.446347274 | 0.869735686 |
| MMP_3 | Hyper_Insulin_pPS | 4225 | 0.015405314 | [-0.01 0.05] | 0.318211219 | 0.839009314 |
| MMP_3 | Beta_Cell_1_pPS | 4225 | -0.021301575 | [-0.05 0.01] | 0.167511057 | 0.722132896 |
| MMP_3 | Beta_Cell_2_pPS | 4225 | -0.000765362 | [-0.03 0.03] | 0.960452022 | 0.996425456 |
| LOX_1 | Cholesterol_pPS | 4159 | -0.037130681 | [-0.07 -0.01] | 0.016965131 | 0.33440804 |
| LOX_1 | Lipodystrophy_1_pPS | 4159 | -0.008546731 | [-0.04 0.02] | 0.582754262 | 0.909803018 |
| LOX_1 | Lipodystrophy_2_pPS | 4159 | 0.01324808 | [-0.02 0.04] | 0.394446789 | 0.85037326 |
| LOX_1 | Liver-Lipid_pPS | 4159 | -0.018074735 | [-0.05 0.01] | 0.245284204 | 0.79432283 |
| LOX_1 | ASAT_Pos_pPS | 4159 | -0.007591192 | [-0.04 0.02] | 0.625589415 | 0.927058714 |
| LOX_1 | ALP_Neg_pPS | 4159 | 0.014951183 | [-0.02 0.05] | 0.336518436 | 0.844649909 |
| LOX_1 | SHBG-LpA_pPS | 4159 | 0.005337477 | [-0.03 0.04] | 0.73153973 | 0.947604067 |
| LOX_1 | Obesity_pPS | 4159 | 0.008711507 | [-0.02 0.04] | 0.575508115 | 0.909113996 |
| LOX_1 | VAT_Neg_pPS | 4159 | -0.013765946 | [-0.04 0.02] | 0.376225598 | 0.849453763 |
| LOX_1 | Bilirubin_pPS | 4159 | 0.017319326 | [-0.01 0.05] | 0.26557386 | 0.80722329 |
| LOX_1 | Proinsulin_pPS | 4159 | 0.038918335 | [0.01 0.07] | 0.012332243 | 0.288798701 |
| LOX_1 | Hyper_Insulin_pPS | 4159 | -0.010844715 | [-0.04 0.02] | 0.485750531 | 0.895803896 |
| LOX_1 | Beta_Cell_1_pPS | 4159 | -0.009701723 | [-0.04 0.02] | 0.532883818 | 0.898959051 |
| LOX_1 | Beta_Cell_2_pPS | 4159 | -0.011190989 | [-0.04 0.02] | 0.471929668 | 0.883720482 |
| PARP_1 | Cholesterol_pPS | 4159 | 0.015186093 | [-0.02 0.05] | 0.328980749 | 0.84456682 |

**Table S7: Associations between pPSs and plasma proteins in SCAPIS**

**a) Spearman Correlation**

| Proteomics | PPS | Sample_Size | r | CI95% | p | fdr_p_adjusted |
|---|---|---|---|---|---|---|
| Gal_3 | Obesity_pPS | 4225 | 0.003764389 | [-0.03 0.03] | 0.8070249 | 0.960526567 |
| Gal_3 | VAT_Neg_pPS | 4225 | -0.011148193 | [-0.04 0.02] | 0.469427728 | 0.844602693 |
| Gal_3 | Bilirubin_pPS | 4225 | -0.013343321 | [-0.04 0.02] | 0.386564294 | 0.807658934 |
| Gal_3 | Proinsulin_pPS | 4225 | 0.029325784 | [-0. 0.06] | 0.056999904 | 0.45329986 |
| Gal_3 | Hyper_Insulin_pPS | 4225 | 0.006926671 | [-0.02 0.04] | 0.653095719 | 0.919948884 |
| Gal_3 | Beta_Cell_1_pPS | 4225 | 0.003915739 | [-0.03 0.03] | 0.799428438 | 0.958074488 |
| Gal_3 | Beta_Cell_2_pPS | 4225 | 0.017714252 | [-0.01 0.05] | 0.25033377 | 0.738661262 |
| Gal_3 | Total_GRS | 4225 | 0.027394582 | [-0. 0.06] | 0.075416453 | 0.508247516 |
| Gal_9 | Cholesterol_pPS | 4159 | 0.018456601 | [-0.01 0.05] | 0.234716648 | 0.726631332 |
| Gal_9 | Lipodystrophy_1_pPS | 4159 | 0.012010314 | [-0.02 0.04] | 0.439389024 | 0.830625826 |
| Gal_9 | Lipodystrophy_2_pPS | 4159 | 0.014244873 | [-0.02 0.04] | 0.359090244 | 0.7979754 |
| Gal_9 | Liver-Lipid_pPS | 4159 | 0.00185249 | [-0.03 0.03] | 0.905069924 | 0.988521167 |
| Gal_9 | ASAT_Pos_pPS | 4159 | 0.007368504 | [-0.02 0.04] | 0.635232885 | 0.914460114 |
| Gal_9 | ALP_Neg_pPS | 4159 | -0.088738614 | [-0.12 -0.06] | 1.04E-08 | 2.21E-06 |
| Gal_9 | SHBG-LpA_pPS | 4159 | 0.016046474 | [-0.01 0.05] | 0.301555675 | 0.76319014 |
| Gal_9 | Obesity_pPS | 4159 | 0.040977228 | [0.01 0.07] | 0.008311864 | 0.197592038 |
| Gal_9 | VAT_Neg_pPS | 4159 | -1.52E-05 | [-0.03 0.03] | 0.999216837 | 0.999464614 |
| Gal_9 | Bilirubin_pPS | 4159 | 0.009547069 | [-0.02 0.04] | 0.538796984 | 0.88195499 |
| Gal_9 | Proinsulin_pPS | 4159 | -0.002378717 | [-0.03 0.03] | 0.87829043 | 0.979031336 |
| Gal_9 | Hyper_Insulin_pPS | 4159 | 0.002708871 | [-0.03 0.03] | 0.861557828 | 0.977006629 |
| Gal_9 | Beta_Cell_1_pPS | 4159 | 0.001144747 | [-0.03 0.03] | 0.94125204 | 0.992903797 |
| Gal_9 | Beta_Cell_2_pPS | 4159 | -0.022089577 | [-0.05 0.01] | 0.154953188 | 0.650979658 |
| Gal_9 | Total_GRS | 4159 | 0.021175133 | [-0.01 0.05] | 0.172768714 | 0.674627844 |
| LTBR | Cholesterol_pPS | 4225 | 0.008526197 | [-0.02 0.04] | 0.580084669 | 0.903823705 |
| LTBR | Lipodystrophy_1_pPS | 4225 | 0.026503414 | [-0. 0.06] | 0.08542056 | 0.538266545 |
| LTBR | Lipodystrophy_2_pPS | 4225 | 0.001676665 | [-0.03 0.03] | 0.913364203 | 0.990056084 |
| LTBR | Liver-Lipid_pPS | 4225 | 0.017500377 | [-0.01 0.05] | 0.256099804 | 0.740941247 |
| LTBR | ASAT_Pos_pPS | 4225 | -0.001048393 | [-0.03 0.03] | 0.945762927 | 0.993154165 |
| LTBR | ALP_Neg_pPS | 4225 | 0.011682395 | [-0.02 0.04] | 0.448404658 | 0.831350198 |
| LTBR | SHBG-LpA_pPS | 4225 | -0.004960623 | [-0.04 0.03] | 0.747537015 | 0.947292085 |
| LTBR | Obesity_pPS | 4225 | 0.008807936 | [-0.02 0.04] | 0.567631159 | 0.899863296 |
| LTBR | VAT_Neg_pPS | 4225 | -0.039855811 | [-0.07 -0.01] | 0.009675761 | 0.21711464 |
| LTBR | Bilirubin_pPS | 4225 | 0.001740028 | [-0.03 0.03] | 0.910103742 | 0.990056084 |
| LTBR | Proinsulin_pPS | 4225 | 0.015643823 | [-0.01 0.05] | 0.310027553 | 0.76365449 |
| LTBR | Hyper_Insulin_pPS | 4225 | 1.50E-05 | [-0.03 0.03] | 0.99922415 | 0.999464614 |
| LTBR | Beta_Cell_1_pPS | 4225 | -0.005077079 | [-0.04 0.03] | 0.741818999 | 0.945527195 |
| LTBR | Beta_Cell_2_pPS | 4225 | -0.003200178 | [-0.03 0.03] | 0.835498481 | 0.970026164 |
| LTBR | Total_GRS | 4225 | 0.007141737 | [-0.02 0.04] | 0.64306222 | 0.916568313 |
| MARCO | Cholesterol_pPS | 4159 | 0.016987158 | [-0.01 0.05] | 0.274097704 | 0.746520328 |
| MARCO | Lipodystrophy_1_pPS | 4159 | 0.029358163 | [-0. 0.06] | 0.058701657 | 0.458334656 |
| MARCO | Lipodystrophy_2_pPS | 4159 | 0.023158681 | [-0.01 0.05] | 0.135934012 | 0.629316669 |
| MARCO | Liver-Lipid_pPS | 4159 | 0.003119998 | [-0.03 0.03] | 0.840808792 | 0.972342391 |
| MARCO | ASAT_Pos_pPS | 4159 | -0.003813294 | [-0.03 0.03] | 0.806075258 | 0.960193229 |
| MARCO | ALP_Neg_pPS | 4159 | -0.027796522 | [-0.06 0. ] | 0.073482127 | 0.502472104 |
| MARCO | SHBG-LpA_pPS | 4159 | 0.017220854 | [-0.01 0.05] | 0.267549806 | 0.746520328 |
| MARCO | Obesity_pPS | 4159 | 0.02602591 | [-0. 0.06] | 0.093782386 | 0.560258408 |
| MARCO | VAT_Neg_pPS | 4159 | 0.007927466 | [-0.02 0.04] | 0.609798721 | 0.909483337 |
| MARCO | Bilirubin_pPS | 4159 | 0.009262073 | [-0.02 0.04] | 0.550985112 | 0.892440674 |
| MARCO | Proinsulin_pPS | 4159 | -0.000209382 | [-0.03 0.03] | 0.989245221 | 0.999402232 |
| MARCO | Hyper_Insulin_pPS | 4159 | 0.009296151 | [-0.02 0.04] | 0.549520634 | 0.891638418 |
| MARCO | Beta_Cell_1_pPS | 4159 | 0.017943946 | [-0.01 0.05] | 0.247973894 | 0.737212651 |
| MARCO | Beta_Cell_2_pPS | 4159 | 0.019014848 | [-0.01 0.05] | 0.220859961 | 0.721388749 |
| MARCO | Total_GRS | 4159 | 0.039750325 | [0.01 0.07] | 0.010465714 | 0.223701034 |
| MMP_3 | Cholesterol_pPS | 4225 | 0.000513894 | [-0.03 0.03] | 0.973398889 | 0.996862516 |
| MMP_3 | Lipodystrophy_1_pPS | 4225 | 0.036181189 | [0.01 0.07] | 0.018848839 | 0.29257652 |
| MMP_3 | Lipodystrophy_2_pPS | 4225 | 0.013707655 | [-0.02 0.04] | 0.373729507 | 0.80093559 |
| MMP_3 | Liver-Lipid_pPS | 4225 | -0.013048527 | [-0.04 0.02] | 0.3971436 | 0.812191725 |
| MMP_3 | ASAT_Pos_pPS | 4225 | 0.003477489 | [-0.03 0.03] | 0.821474243 | 0.963461584 |
| MMP_3 | ALP_Neg_pPS | 4225 | 0.015394354 | [-0.01 0.05] | 0.317807455 | 0.767539002 |
| MMP_3 | SHBG-LpA_pPS | 4225 | 0.018522027 | [-0.01 0.05] | 0.22937624 | 0.726631332 |
| MMP_3 | Obesity_pPS | 4225 | -0.001173775 | [-0.03 0.03] | 0.939288312 | 0.992211394 |
| MMP_3 | VAT_Neg_pPS | 4225 | -0.019420143 | [-0.05 0.01] | 0.207575519 | 0.709689105 |
| MMP_3 | Bilirubin_pPS | 4225 | -0.003891439 | [-0.03 0.03] | 0.800646833 | 0.958074488 |
| MMP_3 | Proinsulin_pPS | 4225 | 0.007312725 | [-0.02 0.04] | 0.635131176 | 0.914460114 |
| MMP_3 | Hyper_Insulin_pPS | 4225 | 0.018202128 | [-0.01 0.05] | 0.237521616 | 0.726631332 |
| MMP_3 | Beta_Cell_1_pPS | 4225 | -0.022095961 | [-0.05 0.01] | 0.151587723 | 0.649125224 |
| MMP_3 | Beta_Cell_2_pPS | 4225 | -0.006022552 | [-0.04 0.02] | 0.695947216 | 0.930772879 |
| MMP_3 | Total_GRS | 4225 | 0.013452463 | [-0.02 0.04] | 0.382691506 | 0.806170656 |
| LOX_1 | Cholesterol_pPS | 4159 | -0.035535089 | [-0.07 -0.01] | 0.02211463 | 0.31789781 |
| LOX_1 | Lipodystrophy_1_pPS | 4159 | -0.010085979 | [-0.04 0.02] | 0.516125363 | 0.870853148 |
| LOX_1 | Lipodystrophy_2_pPS | 4159 | 0.008538616 | [-0.02 0.04] | 0.582520191 | 0.903823705 |
| LOX_1 | Liver-Lipid_pPS | 4159 | -0.018905377 | [-0.05 0.01] | 0.223529955 | 0.7224931 |
| LOX_1 | ASAT_Pos_pPS | 4159 | -0.008121272 | [-0.04 0.02] | 0.60108688 | 0.907623171 |
| LOX_1 | ALP_Neg_pPS | 4159 | 0.02093839 | [-0.01 0.05] | 0.177621379 | 0.678995855 |
| LOX_1 | SHBG-LpA_pPS | 4159 | 0.005093508 | [-0.03 0.04] | 0.742978991 | 0.945527195 |
| LOX_1 | Obesity_pPS | 4159 | 0.008880887 | [-0.02 0.04] | 0.567495976 | 0.899863296 |
| LOX_1 | VAT_Neg_pPS | 4159 | -0.013012058 | [-0.04 0.02] | 0.402186274 | 0.81230319 |
| LOX_1 | Bilirubin_pPS | 4159 | 0.017350712 | [-0.01 0.05] | 0.263958164 | 0.746520328 |
| LOX_1 | Proinsulin_pPS | 4159 | 0.039306812 | [0.01 0.07] | 0.011358747 | 0.232377917 |
| LOX_1 | Hyper_Insulin_pPS | 4159 | -0.013779285 | [-0.04 0.02] | 0.375013195 | 0.80226445 |
| LOX_1 | Beta_Cell_1_pPS | 4159 | -0.002908583 | [-0.03 0.03] | 0.851465771 | 0.97528582 |
| LOX_1 | Beta_Cell_2_pPS | 4159 | -0.008055998 | [-0.04 0.02] | 0.604014776 | 0.908869305 |
| LOX_1 | Total_GRS | 4159 | -0.000680056 | [-0.03 0.03] | 0.965079364 | 0.99612031 |
| PARP_1 | Cholesterol_pPS | 4159 | 0.020475727 | [-0.01 0.05] | 0.187396465 | 0.689088129 |
| PARP_1 | Lipodystrophy_1_pPS | 4159 | 0.028052875 | [-0. 0.06] | 0.07086562 | 0.495162304 |
| PARP_1 | Lipodystrophy_2_pPS | 4159 | 0.051447019 | [0.02 0.08] | 0.000918925 | 0.049730068 |
| PARP_1 | Liver-Lipid_pPS | 4159 | 0.005091823 | [-0.03 0.04] | 0.743061046 | 0.945527195 |
| PARP_1 | ASAT_Pos_pPS | 4159 | 0.015603175 | [-0.01 0.05] | 0.315109214 | 0.764236758 |
| PARP_1 | ALP_Neg_pPS | 4159 | 0.001662538 | [-0.03 0.03] | 0.914764692 | 0.990056084 |
| PARP_1 | SHBG-LpA_pPS | 4159 | 0.02004709 | [-0.01 0.05] | 0.196801812 | 0.697416791 |
| PARP_1 | Obesity_pPS | 4159 | 0.045416954 | [0.02 0.08] | 0.003440661 | 0.110421206 |
| PARP_1 | VAT_Neg_pPS | 4159 | 0.016711748 | [-0.01 0.05] | 0.281953842 | 0.748262118 |
| PARP_1 | Bilirubin_pPS | 4159 | 0.004928647 | [-0.03 0.04] | 0.751018006 | 0.948757065 |
| PARP_1 | Proinsulin_pPS | 4159 | 0.019048105 | [-0.01 0.05] | 0.220053391 | 0.719739225 |
| PARP_1 | Hyper_Insulin_pPS | 4159 | -0.006420421 | [-0.04 0.02] | 0.679359866 | 0.925579558 |
| PARP_1 | Beta_Cell_1_pPS | 4159 | -0.015197286 | [-0.05 0.02] | 0.327864546 | 0.775125841 |
| PARP_1 | Beta_Cell_2_pPS | 4159 | -0.000456624 | [-0.03 0.03] | 0.976548383 | 0.996862516 |
| PARP_1 | Total_GRS | 4159 | 0.058185747 | [0.03 0.09] | 0.000177508 | 0.015803968 |
| PD_L2 | Cholesterol_pPS | 4159 | 0.004097761 | [-0.03 0.03] | 0.791929503 | 0.957811318 |
| PD_L2 | Lipodystrophy_1_pPS | 4159 | 0.011970735 | [-0.02 0.04] | 0.440898344 | 0.831350198 |
| PD_L2 | Lipodystrophy_2_pPS | 4159 | 0.0275683 | [-0. 0.06] | 0.075877481 | 0.509542206 |
| PD_L2 | Liver-Lipid_pPS | 4159 | 0.003299086 | [-0.03 0.03] | 0.831803978 | 0.967866348 |
| PD_L2 | ASAT_Pos_pPS | 4159 | -0.007569083 | [-0.04 0.02] | 0.626054369 | 0.913767992 |
| PD_L2 | ALP_Neg_pPS | 4159 | -0.063355502 | [-0.09 -0.03] | 4.45E-05 | 0.005339807 |
| PD_L2 | SHBG-LpA_pPS | 4159 | 0.025965505 | [-0. 0.06] | 0.094547247 | 0.562393109 |
| PD_L2 | Obesity_pPS | 4159 | 0.002459424 | [-0.03 0.03] | 0.874194796 | 0.978250438 |
| PD_L2 | VAT_Neg_pPS | 4159 | 0.034656702 | [0. 0.07] | 0.025628692 | 0.338445886 |

**b) Partial Correlation**

| Proteomics | PPS | Sample_Size | r | CI95% | p | fdr_p_adjusted |
|---|---|---|---|---|---|---|
| PARP_1 | Lipodystrophy_1_pPS | 4159 | 0.022370735 | [-0.01 0.05] | 0.15040626 | 0.706166997 |
| PARP_1 | Lipodystrophy_2_pPS | 4159 | 0.046655988 | [0.02 0.08] | 0.00269511 | 0.119687959 |
| PARP_1 | Liver-Lipid_pPS | 4159 | 0.003350082 | [-0.03 0.03] | 0.829507728 | 0.971458352 |
| PARP_1 | ASAT_Pos_pPS | 4159 | 0.00721145 | [-0.02 0.04] | 0.642981251 | 0.933976497 |
| PARP_1 | ALP_Neg_pPS | 4159 | -0.002056231 | [-0.03 0.03] | 0.894851454 | 0.98328949 |
| PARP_1 | SHBG-LpA_pPS | 4159 | 0.017011891 | [-0.01 0.05] | 0.27415322 | 0.808820203 |
| PARP_1 | Obesity_pPS | 4159 | 0.041460643 | [0.01 0.07] | 0.007673652 | 0.217223368 |
| PARP_1 | VAT_Neg_pPS | 4159 | 0.012888498 | [-0.02 0.04] | 0.407407159 | 0.853674654 |
| PARP_1 | Bilirubin_pPS | 4159 | 0.004295147 | [-0.03 0.03] | 0.782487142 | 0.964905159 |
| PARP_1 | Proinsulin_pPS | 4159 | 0.017660401 | [-0.01 0.05] | 0.256274094 | 0.802737705 |
| PARP_1 | Hyper_Insulin_pPS | 4159 | -0.017367066 | [-0.05 0.01] | 0.264258376 | 0.80722329 |
| PARP_1 | Beta_Cell_1_pPS | 4159 | -0.02088565 | [-0.05 0.01] | 0.179398918 | 0.738879686 |
| PARP_1 | Beta_Cell_2_pPS | 4159 | -0.000399044 | [-0.03 0.03] | 0.97953708 | 0.998803911 |
| PD_L2 | Cholesterol_pPS | 4159 | 0.004307056 | [-0.03 0.03] | 0.781899256 | 0.964905159 |
| PD_L2 | Lipodystrophy_1_pPS | 4159 | 0.009635708 | [-0.02 0.04] | 0.535675078 | 0.898959051 |
| PD_L2 | Lipodystrophy_2_pPS | 4159 | 0.023838656 | [-0.01 0.05] | 0.125401807 | 0.694065456 |
| PD_L2 | Liver-Lipid_pPS | 4159 | 0.003416558 | [-0.03 0.03] | 0.826178125 | 0.970677755 |
| PD_L2 | ASAT_Pos_pPS | 4159 | -0.01288699 | [-0.04 0.02] | 0.407462016 | 0.853674654 |
| PD_L2 | ALP_Neg_pPS | 4159 | -0.061549178 | [-0.09 -0.03] | 7.50E-05 | 0.008396643 |
| PD_L2 | SHBG-LpA_pPS | 4159 | 0.021064746 | [-0.01 0.05] | 0.175696971 | 0.732083915 |
| PD_L2 | Obesity_pPS | 4159 | 0.00363674 | [-0.03 0.03] | 0.815172395 | 0.968668398 |
| PD_L2 | VAT_Neg_pPS | 4159 | 0.029577584 | [-0. 0.06] | 0.057227802 | 0.558815902 |
| PD_L2 | Bilirubin_pPS | 4159 | -0.031927262 | [-0.06 -0. ] | 0.040101316 | 0.474592997 |
| PD_L2 | Proinsulin_pPS | 4159 | -0.005154834 | [-0.04 0.03] | 0.740389053 | 0.951867558 |
| PD_L2 | Hyper_Insulin_pPS | 4159 | 0.012669102 | [-0.02 0.04] | 0.415437969 | 0.854508456 |
| PD_L2 | Beta_Cell_1_pPS | 4159 | 0.008126579 | [-0.02 0.04] | 0.601420621 | 0.918301271 |
| PD_L2 | Beta_Cell_2_pPS | 4159 | -0.015418176 | [-0.05 0.02] | 0.321642076 | 0.83927892 |
| PGLYRP1 | Cholesterol_pPS | 4225 | 0.003532606 | [-0.03 0.03] | 0.818968717 | 0.968668398 |
| PGLYRP1 | Lipodystrophy_1_pPS | 4225 | 0.003947053 | [-0.03 0.03] | 0.798164634 | 0.967638637 |
| PGLYRP1 | Lipodystrophy_2_pPS | 4225 | 0.007476075 | [-0.02 0.04] | 0.628125018 | 0.928846181 |
| PGLYRP1 | Liver-Lipid_pPS | 4225 | 0.008775843 | [-0.02 0.04] | 0.569640314 | 0.905732577 |
| PGLYRP1 | ASAT_Pos_pPS | 4225 | 0.013144453 | [-0.02 0.04] | 0.394413285 | 0.85037326 |
| PGLYRP1 | ALP_Neg_pPS | 4225 | 0.009775697 | [-0.02 0.04] | 0.526495893 | 0.898776289 |
| PGLYRP1 | SHBG-LpA_pPS | 4225 | -0.002669581 | [-0.03 0.03] | 0.862684864 | 0.97556306 |
| PGLYRP1 | Obesity_pPS | 4225 | 0.006168665 | [-0.02 0.04] | 0.689406974 | 0.944989936 |
| PGLYRP1 | VAT_Neg_pPS | 4225 | -0.01585995 | [-0.05 0.01] | 0.3041376 | 0.826182442 |
| PGLYRP1 | Bilirubin_pPS | 4225 | 0.010345401 | [-0.02 0.04] | 0.502681219 | 0.895803896 |
| PGLYRP1 | Proinsulin_pPS | 4225 | 0.036471731 | [0.01 0.07] | 0.018092425 | 0.335295597 |
| PGLYRP1 | Hyper_Insulin_pPS | 4225 | 0.004346792 | [-0.03 0.03] | 0.778233528 | 0.964905159 |
| PGLYRP1 | Beta_Cell_1_pPS | 4225 | 0.014384209 | [-0.02 0.04] | 0.351349966 | 0.847284666 |
| PGLYRP1 | Beta_Cell_2_pPS | 4225 | -0.037430974 | [-0.07 -0.01] | 0.015269113 | 0.317679992 |
| PIgR | Cholesterol_pPS | 4159 | 0.022819507 | [-0.01 0.05] | 0.142388385 | 0.697681852 |
| PIgR | Lipodystrophy_1_pPS | 4159 | -0.015425977 | [-0.05 0.02] | 0.321397289 | 0.83927892 |
| PIgR | Lipodystrophy_2_pPS | 4159 | 0.013337649 | [-0.02 0.04] | 0.391257658 | 0.85037326 |
| PIgR | Liver-Lipid_pPS | 4159 | 0.008700719 | [-0.02 0.04] | 0.575981226 | 0.909113996 |
| PIgR | ASAT_Pos_pPS | 4159 | -0.013160185 | [-0.04 0.02] | 0.397591536 | 0.85037326 |
| PIgR | ALP_Neg_pPS | 4159 | -0.045573162 | [-0.08 -0.02] | 0.003380716 | 0.133980364 |
| PIgR | SHBG-LpA_pPS | 4159 | 0.030906995 | [0. 0.06] | 0.046914006 | 0.518671587 |
| PIgR | Obesity_pPS | 4159 | 0.013736662 | [-0.02 0.04] | 0.377241888 | 0.849453763 |
| PIgR | VAT_Neg_pPS | 4159 | -0.01174744 | [-0.04 0.02] | 0.450181029 | 0.870351501 |
| PIgR | Bilirubin_pPS | 4159 | -0.006776501 | [-0.04 0.02] | 0.663144059 | 0.937489141 |
| PIgR | Proinsulin_pPS | 4159 | 0.008838551 | [-0.02 0.04] | 0.569950507 | 0.905732577 |
| PIgR | Hyper_Insulin_pPS | 4159 | -0.006982146 | [-0.04 0.02] | 0.653579466 | 0.936605382 |
| PIgR | Beta_Cell_1_pPS | 4159 | -0.009634133 | [-0.04 0.02] | 0.535741741 | 0.898959051 |
| PIgR | Beta_Cell_2_pPS | 4159 | -0.013410412 | [-0.04 0.02] | 0.388678479 | 0.85037326 |
| U_PAR | Cholesterol_pPS | 4225 | -0.003518252 | [-0.03 0.03] | 0.81969161 | 0.968668398 |
| U_PAR | Lipodystrophy_1_pPS | 4225 | 0.011954401 | [-0.02 0.04] | 0.43861695 | 0.865276785 |
| U_PAR | Lipodystrophy_2_pPS | 4225 | 0.016154613 | [-0.01 0.05] | 0.295240172 | 0.817900851 |
| U_PAR | Liver-Lipid_pPS | 4225 | 0.026111936 | [-0. 0.06] | 0.090639948 | 0.634479633 |
| U_PAR | ASAT_Pos_pPS | 4225 | -0.003800868 | [-0.03 0.03] | 0.8054872 | 0.967638637 |
| U_PAR | ALP_Neg_pPS | 4225 | 0.029079266 | [-0. 0.06] | 0.059512335 | 0.558815902 |
| U_PAR | SHBG-LpA_pPS | 4225 | 0.011622945 | [-0.02 0.04] | 0.451417368 | 0.870351501 |
| U_PAR | Obesity_pPS | 4225 | 0.026736831 | [-0. 0.06] | 0.083177704 | 0.614725404 |
| U_PAR | VAT_Neg_pPS | 4225 | -0.016036516 | [-0.05 0.01] | 0.298784973 | 0.820509533 |
| U_PAR | Bilirubin_pPS | 4225 | 0.014952752 | [-0.02 0.05] | 0.332637613 | 0.844649909 |
| U_PAR | Proinsulin_pPS | 4225 | 0.046186667 | [0.02 0.08] | 0.002753957 | 0.119687959 |
| U_PAR | Hyper_Insulin_pPS | 4225 | 0.014243969 | [-0.02 0.04] | 0.356066242 | 0.847284666 |
| U_PAR | Beta_Cell_1_pPS | 4225 | -0.019398581 | [-0.05 0.01] | 0.208784129 | 0.767305935 |
| U_PAR | Beta_Cell_2_pPS | 4225 | -0.003962681 | [-0.03 0.03] | 0.797382872 | 0.967638637 |
| RARRES2 | Cholesterol_pPS | 4225 | 0.009866351 | [-0.02 0.04] | 0.522668189 | 0.895803896 |
| RARRES2 | Lipodystrophy_1_pPS | 4225 | 0.022542015 | [-0.01 0.05] | 0.144114189 | 0.697681852 |
| RARRES2 | Lipodystrophy_2_pPS | 4225 | 0.027363818 | [-0. 0.06] | 0.076198513 | 0.597342987 |
| RARRES2 | Liver-Lipid_pPS | 4225 | -0.017415317 | [-0.05 0.01] | 0.259154772 | 0.80598779 |
| RARRES2 | ASAT_Pos_pPS | 4225 | 0.019653182 | [-0.01 0.05] | 0.202870607 | 0.765145476 |
| RARRES2 | ALP_Neg_pPS | 4225 | 0.00432717 | [-0.03 0.03] | 0.779208618 | 0.964905159 |
| RARRES2 | SHBG-LpA_pPS | 4225 | -0.006819819 | [-0.04 0.02] | 0.658598941 | 0.937489141 |
| RARRES2 | Obesity_pPS | 4225 | 0.036566935 | [0.01 0.07] | 0.017793117 | 0.33440804 |
| RARRES2 | VAT_Neg_pPS | 4225 | -0.016421545 | [-0.05 0.01] | 0.287332003 | 0.81609738 |
| RARRES2 | Bilirubin_pPS | 4225 | 0.010103618 | [-0.02 0.04] | 0.512718174 | 0.895803896 |
| RARRES2 | Proinsulin_pPS | 4225 | 0.002253027 | [-0.03 0.03] | 0.883945438 | 0.979373526 |
| RARRES2 | Hyper_Insulin_pPS | 4225 | -0.005306105 | [-0.04 0.02] | 0.731015506 | 0.947604067 |
| RARRES2 | Beta_Cell_1_pPS | 4225 | 0.021378652 | [-0.01 0.05] | 0.165978817 | 0.721014222 |
| RARRES2 | Beta_Cell_2_pPS | 4225 | -0.013151394 | [-0.04 0.02] | 0.394163642 | 0.85037326 |
| SELE | Cholesterol_pPS | 4225 | 0.008155082 | [-0.02 0.04] | 0.59724869 | 0.918301271 |
| SELE | Lipodystrophy_1_pPS | 4225 | 0.042072878 | [0.01 0.07] | 0.006390723 | 0.193676513 |
| SELE | Lipodystrophy_2_pPS | 4225 | 0.0412176 | [0.01 0.07] | 0.007550274 | 0.216105607 |
| SELE | Liver-Lipid_pPS | 4225 | 0.027094299 | [-0. 0.06] | 0.079137783 | 0.603132929 |
| SELE | ASAT_Pos_pPS | 4225 | 0.022893248 | [-0.01 0.05] | 0.137966893 | 0.696606 |
| SELE | ALP_Neg_pPS | 4225 | -0.398880265 | [-0.42 -0.37] | 3.32E-160 | 8.56E-157 |
| SELE | SHBG-LpA_pPS | 4225 | -0.015383925 | [-0.05 0.01] | 0.318883699 | 0.839235059 |
| SELE | Obesity_pPS | 4225 | 0.032112068 | [0. 0.06] | 0.037432348 | 0.459170134 |
| SELE | VAT_Neg_pPS | 4225 | -0.01858091 | [-0.05 0.01] | 0.228618139 | 0.786988264 |
| SELE | Bilirubin_pPS | 4225 | -0.021961315 | [-0.05 0.01] | 0.154735039 | 0.708464493 |
| SELE | Proinsulin_pPS | 4225 | 0.008891972 | [-0.02 0.04] | 0.564543871 | 0.904956448 |
| SELE | Hyper_Insulin_pPS | 4225 | 0.018391104 | [-0.01 0.05] | 0.23340801 | 0.786988264 |
| SELE | Beta_Cell_1_pPS | 4225 | 0.002609679 | [-0.03 0.03] | 0.865736473 | 0.97556306 |
| SELE | Beta_Cell_2_pPS | 4225 | 0.020520811 | [-0.01 0.05] | 0.183635376 | 0.738879686 |
| PAI | Cholesterol_pPS | 4225 | 0.010584207 | [-0.02 0.04] | 0.492870726 | 0.895803896 |
| PAI | Lipodystrophy_1_pPS | 4225 | 0.029287554 | [-0. 0.06] | 0.057710234 | 0.558815902 |
| PAI | Lipodystrophy_2_pPS | 4225 | 0.012764132 | [-0.02 0.04] | 0.408238326 | 0.853713988 |
| PAI | Liver-Lipid_pPS | 4225 | -0.01321906 | [-0.04 0.02] | 0.39173486 | 0.85037326 |
| PAI | ASAT_Pos_pPS | 4225 | 0.02738923 | [-0. 0.06] | 0.075926032 | 0.597342987 |
| PAI | ALP_Neg_pPS | 4225 | 6.53E-05 | [-0.03 0.03] | 0.996626759 | 0.999305801 |
| PAI | SHBG-LpA_pPS | 4225 | -0.005283737 | [-0.04 0.02] | 0.732105763 | 0.947690676 |
| PAI | Obesity_pPS | 4225 | 0.017305132 | [-0.01 0.05] | 0.262181094 | 0.807171938 |
| PAI | VAT_Neg_pPS | 4225 | 0.011666269 | [-0.02 0.04] | 0.449732369 | 0.870351501 |
| PAI | Bilirubin_pPS | 4225 | 0.003806537 | [-0.03 0.03] | 0.80520289 | 0.967638637 |

**Table S7: Associations between pPSs and plasma proteins in SCAPIS**

**a) Spearman Correlation**

| Proteomics | PPS | Sample_Size | r | CI95% | p | fdr_p_adjusted |
|---|---|---|---|---|---|---|
| PD_L2 | Bilirubin_pPS | 4159 | -0.031973291 | [-0.06 -0. ] | 0.039504183 | 0.40532173 |
| PD_L2 | Proinsulin_pPS | 4159 | -0.014158885 | [-0.04 0.02] | 0.361998574 | 0.7979754 |
| PD_L2 | Hyper_Insulin_pPS | 4159 | 0.01658376 | [-0.01 0.05] | 0.285656189 | 0.750153265 |
| PD_L2 | Beta_Cell_1_pPS | 4159 | 0.00436419 | [-0.03 0.03] | 0.778742424 | 0.953637204 |
| PD_L2 | Beta_Cell_2_pPS | 4159 | -0.012205314 | [-0.04 0.02] | 0.43199612 | 0.827992564 |
| PD_L2 | Total_GRS | 4159 | 0.013148548 | [-0.02 0.04] | 0.397267692 | 0.812191725 |
| PGLYRP1 | Cholesterol_pPS | 4225 | 0.005248173 | [-0.02 0.04] | 0.73344411 | 0.942134248 |
| PGLYRP1 | Lipodystrophy_1_pPS | 4225 | 0.00804443 | [-0.02 0.04] | 0.601671482 | 0.907623171 |
| PGLYRP1 | Lipodystrophy_2_pPS | 4225 | 0.007263865 | [-0.02 0.04] | 0.637393268 | 0.915577927 |
| PGLYRP1 | Liver-Lipid_pPS | 4225 | 0.009938848 | [-0.02 0.04] | 0.518971557 | 0.872596104 |
| PGLYRP1 | ASAT_Pos_pPS | 4225 | 0.016854897 | [-0.01 0.05] | 0.274058307 | 0.746520328 |
| PGLYRP1 | ALP_Neg_pPS | 4225 | 0.016859876 | [-0.01 0.05] | 0.273916572 | 0.746520328 |
| PGLYRP1 | SHBG-LpA_pPS | 4225 | -0.002408336 | [-0.03 0.03] | 0.875817693 | 0.978250438 |
| PGLYRP1 | Obesity_pPS | 4225 | 0.012121922 | [-0.02 0.04] | 0.431515707 | 0.827992564 |
| PGLYRP1 | VAT_Neg_pPS | 4225 | -0.014498247 | [-0.04 0.02] | 0.346798663 | 0.790391668 |
| PGLYRP1 | Bilirubin_pPS | 4225 | 0.011322225 | [-0.02 0.04] | 0.462519657 | 0.83934244 |
| PGLYRP1 | Proinsulin_pPS | 4225 | 0.039401648 | [0.01 0.07] | 0.010536643 | 0.223701034 |
| PGLYRP1 | Hyper_Insulin_pPS | 4225 | 0.005611057 | [-0.02 0.04] | 0.715786953 | 0.94141015 |
| PGLYRP1 | Beta_Cell_1_pPS | 4225 | 0.016142394 | [-0.01 0.05] | 0.294858982 | 0.757449737 |
| PGLYRP1 | Beta_Cell_2_pPS | 4225 | -0.028400919 | [-0.06 0. ] | 0.065290913 | 0.475856102 |
| PGLYRP1 | Total_GRS | 4225 | 0.010779102 | [-0.02 0.04] | 0.484265443 | 0.854586077 |
| PIgR | Cholesterol_pPS | 4159 | 0.024936807 | [-0.01 0.06] | 0.108356057 | 0.599333113 |
| PIgR | Lipodystrophy_1_pPS | 4159 | -0.016897169 | [-0.05 0.01] | 0.276648041 | 0.747422497 |
| PIgR | Lipodystrophy_2_pPS | 4159 | 0.010569612 | [-0.02 0.04] | 0.496208624 | 0.863780476 |
| PIgR | Liver-Lipid_pPS | 4159 | 0.007297879 | [-0.02 0.04] | 0.638478189 | 0.915577927 |
| PIgR | ASAT_Pos_pPS | 4159 | -0.015377232 | [-0.05 0.02] | 0.322168863 | 0.771862902 |
| PIgR | ALP_Neg_pPS | 4159 | -0.045570978 | [-0.08 -0.02] | 0.003332428 | 0.108205887 |
| PIgR | SHBG-LpA_pPS | 4159 | 0.030289913 | [-0. 0.06] | 0.051123082 | 0.440215663 |
| PIgR | Obesity_pPS | 4159 | 0.014324683 | [-0.02 0.04] | 0.35640405 | 0.7979754 |
| PIgR | VAT_Neg_pPS | 4159 | -0.012607247 | [-0.04 0.02] | 0.416987341 | 0.817883456 |
| PIgR | Bilirubin_pPS | 4159 | -0.008427595 | [-0.04 0.02] | 0.587433064 | 0.904020128 |
| PIgR | Proinsulin_pPS | 4159 | -0.000776437 | [-0.03 0.03] | 0.960134101 | 0.99612031 |
| PIgR | Hyper_Insulin_pPS | 4159 | -0.009303246 | [-0.04 0.02] | 0.549215979 | 0.891638418 |
| PIgR | Beta_Cell_1_pPS | 4159 | -0.014179895 | [-0.04 0.02] | 0.361286606 | 0.7979754 |
| PIgR | Beta_Cell_2_pPS | 4159 | -0.015148232 | [-0.05 0.02] | 0.329428483 | 0.775125841 |
| PIgR | Total_GRS | 4159 | 0.001900785 | [-0.03 0.03] | 0.902607234 | 0.987003157 |
| U_PAR | Cholesterol_pPS | 4225 | 0.002146791 | [-0.03 0.03] | 0.889211564 | 0.980121373 |
| U_PAR | Lipodystrophy_1_pPS | 4225 | 0.016327363 | [-0.01 0.05] | 0.289360045 | 0.753421877 |
| U_PAR | Lipodystrophy_2_pPS | 4225 | 0.017591813 | [-0.01 0.05] | 0.253623515 | 0.739680398 |
| U_PAR | Liver-Lipid_pPS | 4225 | 0.028919144 | [-0. 0.06] | 0.06053063 | 0.463868622 |
| U_PAR | ASAT_Pos_pPS | 4225 | 0.002329136 | [-0.03 0.03] | 0.879870009 | 0.979035738 |
| U_PAR | ALP_Neg_pPS | 4225 | 0.039529383 | [0.01 0.07] | 0.010287885 | 0.223701034 |
| U_PAR | SHBG-LpA_pPS | 4225 | 0.012960333 | [-0.02 0.04] | 0.400342247 | 0.81230319 |
| U_PAR | Obesity_pPS | 4225 | 0.034551988 | [0. 0.06] | 0.024916706 | 0.333835472 |
| U_PAR | VAT_Neg_pPS | 4225 | -0.012412709 | [-0.04 0.02] | 0.420547675 | 0.821148791 |
| U_PAR | Bilirubin_pPS | 4225 | 0.014710247 | [-0.02 0.04] | 0.339792382 | 0.78610811 |
| U_PAR | Proinsulin_pPS | 4225 | 0.052278191 | [0.02 0.08] | 0.000687371 | 0.039450459 |
| U_PAR | Hyper_Insulin_pPS | 4225 | 0.018812369 | [-0.01 0.05] | 0.222157254 | 0.722207328 |
| U_PAR | Beta_Cell_1_pPS | 4225 | -0.007358368 | [-0.04 0.02] | 0.633021124 | 0.914460114 |
| U_PAR | Beta_Cell_2_pPS | 4225 | 0.000694239 | [-0.03 0.03] | 0.964069014 | 0.99612031 |
| U_PAR | Total_GRS | 4225 | 0.037652905 | [0.01 0.07] | 0.014521315 | 0.252355767 |
| RARRES2 | Cholesterol_pPS | 4225 | 0.013826987 | [-0.02 0.04] | 0.369583665 | 0.799412943 |
| RARRES2 | Lipodystrophy_1_pPS | 4225 | 0.028976132 | [-0. 0.06] | 0.060025162 | 0.463868622 |
| RARRES2 | Lipodystrophy_2_pPS | 4225 | 0.027600195 | [-0. 0.06] | 0.073249533 | 0.502472104 |
| RARRES2 | Liver-Lipid_pPS | 4225 | -0.01553478 | [-0.05 0.01] | 0.313412556 | 0.76365449 |
| RARRES2 | ASAT_Pos_pPS | 4225 | 0.028521005 | [-0. 0.06] | 0.06416122 | 0.475856102 |
| RARRES2 | ALP_Neg_pPS | 4225 | 0.005438331 | [-0.02 0.04] | 0.724173126 | 0.942134248 |
| RARRES2 | SHBG-LpA_pPS | 4225 | -0.006544569 | [-0.04 0.02] | 0.671076571 | 0.925160508 |
| RARRES2 | Obesity_pPS | 4225 | 0.039576513 | [0.01 0.07] | 0.01019743 | 0.223701034 |
| RARRES2 | VAT_Neg_pPS | 4225 | -0.011707221 | [-0.04 0.02] | 0.447440807 | 0.831350198 |
| RARRES2 | Bilirubin_pPS | 4225 | 0.012547227 | [-0.02 0.04] | 0.415529762 | 0.817860704 |
| RARRES2 | Proinsulin_pPS | 4225 | 0.011643179 | [-0.02 0.04] | 0.449929555 | 0.831350198 |
| RARRES2 | Hyper_Insulin_pPS | 4225 | 0.001096881 | [-0.03 0.03] | 0.943258607 | 0.992903797 |
| RARRES2 | Beta_Cell_1_pPS | 4225 | 0.019314986 | [-0.01 0.05] | 0.210047502 | 0.709689105 |
| RARRES2 | Beta_Cell_2_pPS | 4225 | -0.007841917 | [-0.04 0.02] | 0.610852454 | 0.909483337 |
| RARRES2 | Total_GRS | 4225 | 0.037889535 | [0.01 0.07] | 0.013914112 | 0.246172756 |
| SELE | Cholesterol_pPS | 4225 | 0.01198945 | [-0.02 0.04] | 0.436566783 | 0.829266566 |
| SELE | Lipodystrophy_1_pPS | 4225 | 0.038966507 | [0.01 0.07] | 0.011424679 | 0.232377917 |
| SELE | Lipodystrophy_2_pPS | 4225 | 0.040614828 | [0.01 0.07] | 0.008376184 | 0.197592038 |
| SELE | Liver-Lipid_pPS | 4225 | 0.011942343 | [-0.02 0.04] | 0.438371139 | 0.830394412 |
| SELE | ASAT_Pos_pPS | 4225 | 0.033053535 | [0. 0.06] | 0.031922791 | 0.373334335 |
| SELE | ALP_Neg_pPS | 4225 | -0.398757162 | [-0.42 -0.37] | 1.38E-160 | 3.80E-157 |
| SELE | SHBG-LpA_pPS | 4225 | 0.000197308 | [-0.03 0.03] | 0.989784993 | 0.999402232 |
| SELE | Obesity_pPS | 4225 | 0.027906272 | [-0. 0.06] | 0.070118391 | 0.491184667 |
| SELE | VAT_Neg_pPS | 4225 | -0.008927787 | [-0.04 0.02] | 0.562372543 | 0.897748415 |
| SELE | Bilirubin_pPS | 4225 | -0.018296599 | [-0.05 0.01] | 0.235095189 | 0.726631332 |
| SELE | Proinsulin_pPS | 4225 | -0.050545746 | [-0.08 -0.02] | 0.001030997 | 0.050992277 |
| SELE | Hyper_Insulin_pPS | 4225 | 0.018020288 | [-0.01 0.05] | 0.242241713 | 0.730235185 |
| SELE | Beta_Cell_1_pPS | 4225 | 0.007556062 | [-0.02 0.04] | 0.623916309 | 0.91287224 |
| SELE | Beta_Cell_2_pPS | 4225 | 0.01696397 | [-0.01 0.05] | 0.270964777 | 0.746520328 |
| SELE | Total_GRS | 4225 | 0.023807446 | [-0.01 0.05] | 0.122335314 | 0.615837655 |
| PAI | Cholesterol_pPS | 4225 | 0.011629271 | [-0.02 0.04] | 0.450471083 | 0.831350198 |
| PAI | Lipodystrophy_1_pPS | 4225 | 0.035084276 | [0. 0.07] | 0.022770564 | 0.320646716 |
| PAI | Lipodystrophy_2_pPS | 4225 | 0.018250197 | [-0.01 0.05] | 0.236284786 | 0.726631332 |
| PAI | Liver-Lipid_pPS | 4225 | -0.013176363 | [-0.04 0.02] | 0.392534669 | 0.812191725 |
| PAI | ASAT_Pos_pPS | 4225 | 0.034058326 | [0. 0.06] | 0.027061439 | 0.350655265 |
| PAI | ALP_Neg_pPS | 4225 | -0.000896006 | [-0.03 0.03] | 0.953636821 | 0.99612031 |
| PAI | SHBG-LpA_pPS | 4225 | -0.003135859 | [-0.03 0.03] | 0.838758868 | 0.97186166 |
| PAI | Obesity_pPS | 4225 | 0.018662791 | [-0.01 0.05] | 0.225855756 | 0.724402982 |
| PAI | VAT_Neg_pPS | 4225 | 0.010231266 | [-0.02 0.04] | 0.506749507 | 0.867976055 |
| PAI | Bilirubin_pPS | 4225 | 0.004889709 | [-0.03 0.04] | 0.751025755 | 0.948757065 |
| PAI | Proinsulin_pPS | 4225 | -0.011478135 | [-0.04 0.02] | 0.456379341 | 0.835860883 |
| PAI | Hyper_Insulin_pPS | 4225 | -0.000103852 | [-0.03 0.03] | 0.99462327 | 0.999464614 |
| PAI | Beta_Cell_1_pPS | 4225 | -0.01752889 | [-0.05 0.01] | 0.255325819 | 0.74027471 |
| PAI | Beta_Cell_2_pPS | 4225 | -0.019626683 | [-0.05 0.01] | 0.202781779 | 0.705772649 |
| PAI | Total_GRS | 4225 | 0.008516753 | [-0.02 0.04] | 0.580504322 | 0.903823705 |
| PSP_D | Cholesterol_pPS | 4225 | 0.018652202 | [-0.01 0.05] | 0.226119219 | 0.724402982 |
| PSP_D | Lipodystrophy_1_pPS | 4225 | -0.000834887 | [-0.03 0.03] | 0.956796191 | 0.99612031 |
| PSP_D | Lipodystrophy_2_pPS | 4225 | 0.017267409 | [-0.01 0.05] | 0.262484796 | 0.746520328 |
| PSP_D | Liver-Lipid_pPS | 4225 | 0.012052944 | [-0.02 0.04] | 0.434141548 | 0.828998134 |
| PSP_D | ASAT_Pos_pPS | 4225 | 0.010893365 | [-0.02 0.04] | 0.479645034 | 0.851331379 |
| PSP_D | ALP_Neg_pPS | 4225 | 0.002218462 | [-0.03 0.03] | 0.885537917 | 0.979291846 |
| PSP_D | SHBG-LpA_pPS | 4225 | 0.013068481 | [-0.02 0.04] | 0.396422051 | 0.812191725 |
| PSP_D | Obesity_pPS | 4225 | 0.022925118 | [-0.01 0.05] | 0.136811831 | 0.629316669 |
| PSP_D | VAT_Neg_pPS | 4225 | 0.017041775 | [-0.01 0.05] | 0.268772728 | 0.746520328 |
| PSP_D | Bilirubin_pPS | 4225 | -0.002274447 | [-0.03 0.03] | 0.88267005 | 0.979291846 |
| PSP_D | Proinsulin_pPS | 4225 | 0.015085289 | [-0.02 0.05] | 0.327621773 | 0.775125841 |

**b) Partial Correlation**

| Proteomics | PPS | Sample_Size | r | CI95% | p | fdr_p_adjusted |
|---|---|---|---|---|---|---|
| PAI | Proinsulin_pPS | 4225 | -0.006715196 | [-0.04 0.02] | 0.663511738 | 0.937489141 |
| PAI | Hyper_Insulin_pPS | 4225 | -0.00798278 | [-0.04 0.02] | 0.605018282 | 0.918301271 |
| PAI | Beta_Cell_1_pPS | 4225 | -0.013168419 | [-0.04 0.02] | 0.393551705 | 0.85037326 |
| PAI | Beta_Cell_2_pPS | 4225 | -0.014804064 | [-0.05 0.02] | 0.337468079 | 0.844649909 |
| PSP_D | Cholesterol_pPS | 4225 | 0.016425498 | [-0.01 0.05] | 0.287215982 | 0.81609738 |
| PSP_D | Lipodystrophy_1_pPS | 4225 | -0.007921185 | [-0.04 0.02] | 0.60780674 | 0.918301271 |
| PSP_D | Lipodystrophy_2_pPS | 4225 | 0.013517933 | [-0.02 0.04] | 0.381116254 | 0.85037326 |
| PSP_D | Liver-Lipid_pPS | 4225 | 0.008456404 | [-0.02 0.04] | 0.583771356 | 0.91071482 |
| PSP_D | ASAT_Pos_pPS | 4225 | 0.007816275 | [-0.02 0.04] | 0.61256921 | 0.920640773 |
| PSP_D | ALP_Neg_pPS | 4225 | 0.00087004 | [-0.03 0.03] | 0.955048422 | 0.996425456 |
| PSP_D | SHBG-LpA_pPS | 4225 | 0.012352425 | [-0.02 0.04] | 0.423524911 | 0.860815573 |
| PSP_D | Obesity_pPS | 4225 | 0.018926403 | [-0.01 0.05] | 0.220079852 | 0.775610053 |
| PSP_D | VAT_Neg_pPS | 4225 | 0.008518626 | [-0.02 0.04] | 0.581006088 | 0.909583681 |
| PSP_D | Bilirubin_pPS | 4225 | -0.003472226 | [-0.03 0.03] | 0.822010659 | 0.968668398 |
| PSP_D | Proinsulin_pPS | 4225 | 0.000336268 | [-0.03 0.03] | 0.982618532 | 0.998803911 |
| PSP_D | Hyper_Insulin_pPS | 4225 | 0.019758924 | [-0.01 0.05] | 0.200450742 | 0.765145476 |
| PSP_D | Beta_Cell_1_pPS | 4225 | 0.021318139 | [-0.01 0.05] | 0.16718089 | 0.722132896 |
| PSP_D | Beta_Cell_2_pPS | 4225 | 0.020662785 | [-0.01 0.05] | 0.180620867 | 0.738879686 |
| SLAMF7 | Cholesterol_pPS | 4159 | 0.024566113 | [-0.01 0.06] | 0.114274579 | 0.681046037 |
| SLAMF7 | Lipodystrophy_1_pPS | 4159 | -0.000969116 | [-0.03 0.03] | 0.950330603 | 0.996425456 |
| SLAMF7 | Lipodystrophy_2_pPS | 4159 | -0.001018975 | [-0.03 0.03] | 0.947778764 | 0.996425456 |
| SLAMF7 | Liver-Lipid_pPS | 4159 | -0.000398083 | [-0.03 0.03] | 0.979586378 | 0.998803911 |
| SLAMF7 | ASAT_Pos_pPS | 4159 | 0.014421987 | [-0.02 0.04] | 0.35390235 | 0.847284666 |
| SLAMF7 | ALP_Neg_pPS | 4159 | 0.013947702 | [-0.02 0.04] | 0.36995563 | 0.847284666 |
| SLAMF7 | SHBG-LpA_pPS | 4159 | -0.012216491 | [-0.04 0.02] | 0.432297873 | 0.864060889 |
| SLAMF7 | Obesity_pPS | 4159 | 0.007080304 | [-0.02 0.04] | 0.64903397 | 0.935848959 |
| SLAMF7 | VAT_Neg_pPS | 4159 | 0.015980562 | [-0.01 0.05] | 0.304305518 | 0.826182442 |
| SLAMF7 | Bilirubin_pPS | 4159 | -0.000701686 | [-0.03 0.03] | 0.964025913 | 0.996924797 |
| SLAMF7 | Proinsulin_pPS | 4159 | -0.018459777 | [-0.05 0.01] | 0.235371425 | 0.788664502 |
| SLAMF7 | Hyper_Insulin_pPS | 4159 | 0.017192185 | [-0.01 0.05] | 0.269099196 | 0.808113719 |
| SLAMF7 | Beta_Cell_1_pPS | 4159 | 0.010586894 | [-0.02 0.04] | 0.496181505 | 0.895803896 |
| SLAMF7 | Beta_Cell_2_pPS | 4159 | -0.01271425 | [-0.04 0.02] | 0.413777783 | 0.854426619 |
| SPON2 | Cholesterol_pPS | 4159 | 0.013257927 | [-0.02 0.04] | 0.394095406 | 0.85037326 |
| SPON2 | Lipodystrophy_1_pPS | 4159 | 0.015493795 | [-0.01 0.05] | 0.319274207 | 0.839235059 |
| SPON2 | Lipodystrophy_2_pPS | 4159 | 0.011639417 | [-0.02 0.04] | 0.454358118 | 0.872801276 |
| SPON2 | Liver-Lipid_pPS | 4159 | -0.009764964 | [-0.04 0.02] | 0.530216827 | 0.898959051 |
| SPON2 | ASAT_Pos_pPS | 4159 | -0.001246428 | [-0.03 0.03] | 0.93614473 | 0.994163908 |
| SPON2 | ALP_Neg_pPS | 4159 | 0.015154365 | [-0.02 0.05] | 0.3299924 | 0.84456682 |
| SPON2 | SHBG-LpA_pPS | 4159 | 0.014683033 | [-0.02 0.05] | 0.345257324 | 0.847284666 |
| SPON2 | Obesity_pPS | 4159 | 0.025977652 | [-0. 0.06] | 0.094911305 | 0.64003016 |
| SPON2 | VAT_Neg_pPS | 4159 | 0.018772704 | [-0.01 0.05] | 0.227526643 | 0.786988264 |
| SPON2 | Bilirubin_pPS | 4159 | -0.009428875 | [-0.04 0.02] | 0.544467712 | 0.899241939 |
| SPON2 | Proinsulin_pPS | 4159 | -0.00471434 | [-0.04 0.03] | 0.76187149 | 0.958675598 |
| SPON2 | Hyper_Insulin_pPS | 4159 | -0.009424737 | [-0.04 0.02] | 0.544644337 | 0.899241939 |
| SPON2 | Beta_Cell_1_pPS | 4159 | -0.004941561 | [-0.04 0.03] | 0.750766093 | 0.954135344 |
| SPON2 | Beta_Cell_2_pPS | 4159 | -0.0232885 | [-0.05 0.01] | 0.134364118 | 0.694065456 |
| OPN | Cholesterol_pPS | 4225 | -0.013939059 | [-0.04 0.02] | 0.366457285 | 0.847284666 |
| OPN | Lipodystrophy_1_pPS | 4225 | -0.019515965 | [-0.05 0.01] | 0.20604236 | 0.765145476 |
| OPN | Lipodystrophy_2_pPS | 4225 | 0.001939235 | [-0.03 0.03] | 0.900017275 | 0.985399384 |
| OPN | Liver-Lipid_pPS | 4225 | 0.03559183 | [0.01 0.07] | 0.02107387 | 0.354719791 |
| OPN | ASAT_Pos_pPS | 4225 | -0.017039674 | [-0.05 0.01] | 0.269572094 | 0.808113719 |
| OPN | ALP_Neg_pPS | 4225 | -0.022442747 | [-0.05 0.01] | 0.145889075 | 0.69983288 |
| OPN | SHBG-LpA_pPS | 4225 | 0.022516189 | [-0.01 0.05] | 0.144574344 | 0.697681852 |
| OPN | Obesity_pPS | 4225 | -0.02306569 | [-0.05 0.01] | 0.135023707 | 0.694065456 |
| OPN | VAT_Neg_pPS | 4225 | -0.024763693 | [-0.05 0.01] | 0.108573248 | 0.676681438 |
| OPN | Bilirubin_pPS | 4225 | 0.006190935 | [-0.02 0.04] | 0.688344382 | 0.944989936 |
| OPN | Proinsulin_pPS | 4225 | 0.029978024 | [-0. 0.06] | 0.052057704 | 0.540728413 |
| OPN | Hyper_Insulin_pPS | 4225 | 0.01031473 | [-0.02 0.04] | 0.503948666 | 0.895803896 |
| OPN | Beta_Cell_1_pPS | 4225 | -0.016694086 | [-0.05 0.01] | 0.279406419 | 0.808820203 |
| OPN | Beta_Cell_2_pPS | 4225 | 0.017570692 | [-0.01 0.05] | 0.254928496 | 0.801363065 |
| SRC | Cholesterol_pPS | 4159 | 0.005571613 | [-0.02 0.04] | 0.720247563 | 0.946314812 |
| SRC | Lipodystrophy_1_pPS | 4159 | 0.000991812 | [-0.03 0.03] | 0.949168899 | 0.996425456 |
| SRC | Lipodystrophy_2_pPS | 4159 | 0.001130408 | [-0.03 0.03] | 0.942077505 | 0.995811101 |
| SRC | Liver-Lipid_pPS | 4159 | -0.011553141 | [-0.04 0.02] | 0.45770991 | 0.874172623 |
| SRC | ASAT_Pos_pPS | 4159 | -0.006863482 | [-0.04 0.02] | 0.65909174 | 0.937489141 |
| SRC | ALP_Neg_pPS | 4159 | -0.036416634 | [-0.07 -0.01] | 0.019205431 | 0.340663962 |
| SRC | SHBG-LpA_pPS | 4159 | 0.005641603 | [-0.02 0.04] | 0.716883709 | 0.945970482 |
| SRC | Obesity_pPS | 4159 | -0.015410046 | [-0.05 0.02] | 0.32189735 | 0.83927892 |
| SRC | VAT_Neg_pPS | 4159 | -0.009715405 | [-0.04 0.02] | 0.532306271 | 0.898959051 |
| SRC | Bilirubin_pPS | 4159 | 0.006645079 | [-0.02 0.04] | 0.669285555 | 0.937489141 |
| SRC | Proinsulin_pPS | 4159 | 0.011222486 | [-0.02 0.04] | 0.47068338 | 0.883720482 |
| SRC | Hyper_Insulin_pPS | 4159 | -0.004886859 | [-0.04 0.03] | 0.753434999 | 0.95462436 |
| SRC | Beta_Cell_1_pPS | 4159 | 0.006990029 | [-0.02 0.04] | 0.653213942 | 0.936605382 |
| SRC | Beta_Cell_2_pPS | 4159 | -0.004489492 | [-0.03 0.03] | 0.772909412 | 0.962056893 |
| THPO | Cholesterol_pPS | 4159 | -0.014302785 | [-0.04 0.02] | 0.357895015 | 0.847284666 |
| THPO | Lipodystrophy_1_pPS | 4159 | -0.015431839 | [-0.05 0.02] | 0.321213411 | 0.83927892 |
| THPO | Lipodystrophy_2_pPS | 4159 | 0.020134325 | [-0.01 0.05] | 0.195561791 | 0.757544623 |
| THPO | Liver-Lipid_pPS | 4159 | -0.035943395 | [-0.07 -0.01] | 0.020828922 | 0.354719791 |
| THPO | ASAT_Pos_pPS | 4159 | -0.016360972 | [-0.05 0.01] | 0.292937029 | 0.816672929 |
| THPO | ALP_Neg_pPS | 4159 | 0.005394501 | [-0.03 0.04] | 0.72878405 | 0.946981479 |
| THPO | SHBG-LpA_pPS | 4159 | -0.001480896 | [-0.03 0.03] | 0.924166112 | 0.990503296 |
| THPO | Obesity_pPS | 4159 | 0.010055026 | [-0.02 0.04] | 0.518071709 | 0.895803896 |
| THPO | VAT_Neg_pPS | 4159 | -0.017970249 | [-0.05 0.01] | 0.248023925 | 0.797639988 |
| THPO | Bilirubin_pPS | 4159 | -0.013363569 | [-0.04 0.02] | 0.390337724 | 0.85037326 |
| THPO | Proinsulin_pPS | 4159 | 0.003824771 | [-0.03 0.03] | 0.805802404 | 0.967638637 |
| THPO | Hyper_Insulin_pPS | 4159 | -0.01134323 | [-0.04 0.02] | 0.465922535 | 0.879924083 |
| THPO | Beta_Cell_1_pPS | 4159 | -0.00862485 | [-0.04 0.02] | 0.579313613 | 0.909583681 |
| THPO | Beta_Cell_2_pPS | 4159 | 0.002138419 | [-0.03 0.03] | 0.890674484 | 0.981230818 |
| TNFRSF10A | Cholesterol_pPS | 4159 | -0.005793693 | [-0.04 0.02] | 0.709592928 | 0.945970482 |
| TNFRSF10A | Lipodystrophy_1_pPS | 4159 | 0.027114369 | [-0. 0.06] | 0.081308488 | 0.610643339 |
| TNFRSF10A | Lipodystrophy_2_pPS | 4159 | 0.006901475 | [-0.02 0.04] | 0.657324829 | 0.937489141 |
| TNFRSF10A | Liver-Lipid_pPS | 4159 | -0.019252641 | [-0.05 0.01] | 0.215860029 | 0.768032367 |
| TNFRSF10A | ASAT_Pos_pPS | 4159 | 0.022538154 | [-0.01 0.05] | 0.14737578 | 0.703126624 |
| TNFRSF10A | ALP_Neg_pPS | 4159 | -0.000805161 | [-0.03 0.03] | 0.958725391 | 0.996425456 |
| TNFRSF10A | SHBG-LpA_pPS | 4159 | 0.014417023 | [-0.02 0.04] | 0.354068062 | 0.847284666 |
| TNFRSF10A | Obesity_pPS | 4159 | -0.000977992 | [-0.03 0.03] | 0.949876272 | 0.996425456 |
| TNFRSF10A | VAT_Neg_pPS | 4159 | 0.009440021 | [-0.02 0.04] | 0.543992063 | 0.899241939 |
| TNFRSF10A | Bilirubin_pPS | 4159 | -0.000120842 | [-0.03 0.03] | 0.993802623 | 0.998803911 |
| TNFRSF10A | Proinsulin_pPS | 4159 | -0.004831606 | [-0.04 0.03] | 0.75613383 | 0.95462436 |
| TNFRSF10A | Hyper_Insulin_pPS | 4159 | -0.002751233 | [-0.03 0.03] | 0.859633544 | 0.97556306 |
| TNFRSF10A | Beta_Cell_1_pPS | 4159 | -0.013336028 | [-0.04 0.02] | 0.391315245 | 0.85037326 |
| TNFRSF10A | Beta_Cell_2_pPS | 4159 | -0.023817191 | [-0.05 0.01] | 0.125742528 | 0.694065456 |
| TRAIL_R2 | Cholesterol_pPS | 4159 | 0.017674545 | [-0.01 0.05] | 0.255893376 | 0.802737705 |
| TRAIL_R2 | Lipodystrophy_1_pPS | 4159 | -0.002370789 | [-0.03 0.03] | 0.878881873 | 0.97781424 |
| TRAIL_R2 | Lipodystrophy_2_pPS | 4159 | 0.026282013 | [-0. 0.06] | 0.091101802 | 0.63465347 |
| TRAIL_R2 | Liver-Lipid_pPS | 4159 | 0.000541368 | [-0.03 0.03] | 0.972241339 | 0.997444743 |
| TRAIL_R2 | ASAT_Pos_pPS | 4159 | 0.001562257 | [-0.03 0.03] | 0.920013392 | 0.99041009 |

**Table S7: Associations between pPSs and plasma proteins in SCAPIS**

**a) Spearman Correlation**

| Proteomics | PPS | Sample_Size | r | CI95% | p | fdr_p_adjusted |
|---|---|---|---|---|---|---|
| PSP_D | Hyper_Insulin_pPS | 4225 | 0.024185699 | [-0.01 0.05] | 0.11650818 | 0.611872879 |
| PSP_D | Beta_Cell_1_pPS | 4225 | 0.030123081 | [-0. 0.06] | 0.050573493 | 0.440215663 |
| PSP_D | Beta_Cell_2_pPS | 4225 | 0.029842389 | [-0. 0.06] | 0.05276305 | 0.441027526 |
| PSP_D | Total_GRS | 4225 | 0.042390652 | [0.01 0.07] | 0.005925114 | 0.152834705 |
| SLAMF7 | Cholesterol_pPS | 4159 | 0.024623697 | [-0.01 0.06] | 0.112861955 | 0.604978148 |
| SLAMF7 | Lipodystrophy_1_pPS | 4159 | 0.004397417 | [-0.03 0.03] | 0.777102205 | 0.953637204 |
| SLAMF7 | Lipodystrophy_2_pPS | 4159 | 0.00351663 | [-0.03 0.03] | 0.820895336 | 0.963461584 |
| SLAMF7 | Liver-Lipid_pPS | 4159 | 0.001576191 | [-0.03 0.03] | 0.91917595 | 0.990056084 |
| SLAMF7 | ASAT_Pos_pPS | 4159 | 0.016445042 | [-0.01 0.05] | 0.289705867 | 0.753421877 |
| SLAMF7 | ALP_Neg_pPS | 4159 | 0.011654771 | [-0.02 0.04] | 0.453053624 | 0.834174785 |
| SLAMF7 | SHBG-LpA_pPS | 4159 | -0.012779505 | [-0.04 0.02] | 0.410650241 | 0.815978881 |
| SLAMF7 | Obesity_pPS | 4159 | 0.008990612 | [-0.02 0.04] | 0.562719115 | 0.897748415 |
| SLAMF7 | VAT_Neg_pPS | 4159 | 0.015806343 | [-0.01 0.05] | 0.308848584 | 0.76365449 |
| SLAMF7 | Bilirubin_pPS | 4159 | -0.000106403 | [-0.03 0.03] | 0.994534548 | 0.999464614 |
| SLAMF7 | Proinsulin_pPS | 4159 | -0.015022242 | [-0.05 0.02] | 0.333467399 | 0.779518406 |
| SLAMF7 | Hyper_Insulin_pPS | 4159 | 0.020663886 | [-0.01 0.05] | 0.183374212 | 0.685789735 |
| SLAMF7 | Beta_Cell_1_pPS | 4159 | 0.00444259 | [-0.03 0.03] | 0.774873868 | 0.953637204 |
| SLAMF7 | Beta_Cell_2_pPS | 4159 | -0.012098734 | [-0.04 0.02] | 0.436027851 | 0.829210857 |
| SLAMF7 | Total_GRS | 4159 | 0.006072943 | [-0.02 0.04] | 0.695822175 | 0.930772879 |
| SPON2 | Cholesterol_pPS | 4159 | 0.014797032 | [-0.02 0.05] | 0.340766239 | 0.787547587 |
| SPON2 | Lipodystrophy_1_pPS | 4159 | 0.018094785 | [-0.01 0.05] | 0.244019938 | 0.732214246 |
| SPON2 | Lipodystrophy_2_pPS | 4159 | 0.014408402 | [-0.02 0.04] | 0.353599918 | 0.796092727 |
| SPON2 | Liver-Lipid_pPS | 4159 | -0.009050201 | [-0.04 0.02] | 0.560133078 | 0.897253218 |
| SPON2 | ASAT_Pos_pPS | 4159 | 0.002731984 | [-0.03 0.03] | 0.860388677 | 0.977006629 |
| SPON2 | ALP_Neg_pPS | 4159 | 0.014138122 | [-0.02 0.04] | 0.362703042 | 0.7979754 |
| SPON2 | SHBG-LpA_pPS | 4159 | 0.015779309 | [-0.01 0.05] | 0.309676866 | 0.76365449 |
| SPON2 | Obesity_pPS | 4159 | 0.026432289 | [-0. 0.06] | 0.088764791 | 0.546245321 |
| SPON2 | VAT_Neg_pPS | 4159 | 0.017744627 | [-0.01 0.05] | 0.253267088 | 0.739680398 |
| SPON2 | Bilirubin_pPS | 4159 | -0.009271403 | [-0.04 0.02] | 0.550583986 | 0.89239323 |
| SPON2 | Proinsulin_pPS | 4159 | -0.004039845 | [-0.03 0.03] | 0.794804103 | 0.958074488 |
| SPON2 | Hyper_Insulin_pPS | 4159 | -0.003678994 | [-0.03 0.03] | 0.812776017 | 0.962360278 |
| SPON2 | Beta_Cell_1_pPS | 4159 | -0.011215898 | [-0.04 0.02] | 0.470246855 | 0.845525291 |
| SPON2 | Beta_Cell_2_pPS | 4159 | -0.024634621 | [-0.06 0.01] | 0.112702295 | 0.604978148 |
| SPON2 | Total_GRS | 4159 | 0.023688619 | [-0.01 0.05] | 0.127200502 | 0.623575746 |
| OPN | Cholesterol_pPS | 4225 | -0.013413323 | [-0.04 0.02] | 0.384077598 | 0.806422804 |
| OPN | Lipodystrophy_1_pPS | 4225 | -0.022312669 | [-0.05 0.01] | 0.147613447 | 0.645439059 |
| OPN | Lipodystrophy_2_pPS | 4225 | -0.001555609 | [-0.03 0.03] | 0.919597314 | 0.990056084 |
| OPN | Liver-Lipid_pPS | 4225 | 0.034589732 | [0. 0.06] | 0.024758954 | 0.333340061 |
| OPN | ASAT_Pos_pPS | 4225 | -0.02099961 | [-0.05 0.01] | 0.172951661 | 0.674627844 |
| OPN | ALP_Neg_pPS | 4225 | -0.017105698 | [-0.05 0.01] | 0.266980936 | 0.746520328 |
| OPN | SHBG-LpA_pPS | 4225 | 0.022140613 | [-0.01 0.05] | 0.15076224 | 0.649125224 |
| OPN | Obesity_pPS | 4225 | -0.021400668 | [-0.05 0.01] | 0.1648895 | 0.664372291 |
| OPN | VAT_Neg_pPS | 4225 | -0.022322087 | [-0.05 0.01] | 0.147442547 | 0.645439059 |
| OPN | Bilirubin_pPS | 4225 | 0.004127902 | [-0.03 0.03] | 0.788811728 | 0.957247257 |
| OPN | Proinsulin_pPS | 4225 | 0.024479948 | [-0.01 0.05] | 0.112127708 | 0.604978148 |
| OPN | Hyper_Insulin_pPS | 4225 | 0.004525434 | [-0.03 0.03] | 0.769025723 | 0.952961063 |
| OPN | Beta_Cell_1_pPS | 4225 | -0.006957607 | [-0.04 0.02] | 0.651648508 | 0.919948884 |
| OPN | Beta_Cell_2_pPS | 4225 | 0.018246773 | [-0.01 0.05] | 0.236372756 | 0.726631332 |
| OPN | Total_GRS | 4225 | -0.013904769 | [-0.04 0.02] | 0.366896808 | 0.798890546 |
| SRC | Cholesterol_pPS | 4159 | 0.003959274 | [-0.03 0.03] | 0.798807795 | 0.958074488 |
| SRC | Lipodystrophy_1_pPS | 4159 | -0.001668401 | [-0.03 0.03] | 0.914465286 | 0.990056084 |
| SRC | Lipodystrophy_2_pPS | 4159 | -0.001352166 | [-0.03 0.03] | 0.930632158 | 0.990056084 |
| SRC | Liver-Lipid_pPS | 4159 | -0.013007386 | [-0.04 0.02] | 0.402355302 | 0.81230319 |
| SRC | ASAT_Pos_pPS | 4159 | -0.007386123 | [-0.04 0.02] | 0.634424362 | 0.914460114 |
| SRC | ALP_Neg_pPS | 4159 | -0.035913874 | [-0.07 -0.01] | 0.020733532 | 0.312602357 |
| SRC | SHBG-LpA_pPS | 4159 | 0.006432678 | [-0.02 0.04] | 0.678781911 | 0.925579558 |
| SRC | Obesity_pPS | 4159 | -0.014919003 | [-0.05 0.02] | 0.336800662 | 0.782502359 |
| SRC | VAT_Neg_pPS | 4159 | -0.009378432 | [-0.04 0.02] | 0.545992571 | 0.889574673 |
| SRC | Bilirubin_pPS | 4159 | 0.006742558 | [-0.02 0.04] | 0.664233297 | 0.924111434 |
| SRC | Proinsulin_pPS | 4159 | 0.005051237 | [-0.03 0.04] | 0.745037622 | 0.947005534 |
| SRC | Hyper_Insulin_pPS | 4159 | -0.008301499 | [-0.04 0.02] | 0.59303618 | 0.905925461 |
| SRC | Beta_Cell_1_pPS | 4159 | 0.007747062 | [-0.02 0.04] | 0.617958193 | 0.909483337 |
| SRC | Beta_Cell_2_pPS | 4159 | -0.00220286 | [-0.03 0.03] | 0.887225788 | 0.979291846 |
| SRC | Total_GRS | 4159 | -0.009902694 | [-0.04 0.02] | 0.523780176 | 0.872980599 |
| THPO | Cholesterol_pPS | 4159 | -0.013005448 | [-0.04 0.02] | 0.40242541 | 0.81230319 |
| THPO | Lipodystrophy_1_pPS | 4159 | -0.019939791 | [-0.05 0.01] | 0.199209378 | 0.700947446 |
| THPO | Lipodystrophy_2_pPS | 4159 | 0.013529904 | [-0.02 0.04] | 0.383718534 | 0.806422804 |
| THPO | Liver-Lipid_pPS | 4159 | -0.036610524 | [-0.07 -0.01] | 0.018388792 | 0.290017524 |
| THPO | ASAT_Pos_pPS | 4159 | -0.019336444 | [-0.05 0.01] | 0.21314868 | 0.712633026 |
| THPO | ALP_Neg_pPS | 4159 | 0.004597474 | [-0.03 0.04] | 0.767247791 | 0.952160028 |
| THPO | SHBG-LpA_pPS | 4159 | -0.00353577 | [-0.03 0.03] | 0.819937186 | 0.963461584 |
| THPO | Obesity_pPS | 4159 | 0.006742654 | [-0.02 0.04] | 0.664228798 | 0.924111434 |
| THPO | VAT_Neg_pPS | 4159 | -0.018778607 | [-0.05 0.01] | 0.226650623 | 0.724711501 |
| THPO | Bilirubin_pPS | 4159 | -0.013710511 | [-0.04 0.02] | 0.377401666 | 0.803170077 |
| THPO | Proinsulin_pPS | 4159 | 0.001797045 | [-0.03 0.03] | 0.907898332 | 0.990056084 |
| THPO | Hyper_Insulin_pPS | 4159 | -0.016209447 | [-0.05 0.01] | 0.29667189 | 0.759568105 |
| THPO | Beta_Cell_1_pPS | 4159 | -0.012553584 | [-0.04 0.02] | 0.418973251 | 0.820117853 |
| THPO | Beta_Cell_2_pPS | 4159 | -0.001715335 | [-0.03 0.03] | 0.912068736 | 0.990056084 |
| THPO | Total_GRS | 4159 | -0.007100901 | [-0.04 0.02] | 0.647566038 | 0.918420039 |
| TNFRSF10A | Cholesterol_pPS | 4159 | -0.007093999 | [-0.04 0.02] | 0.647885441 | 0.918420039 |
| TNFRSF10A | Lipodystrophy_1_pPS | 4159 | 0.032992989 | [0. 0.06] | 0.033620496 | 0.381862422 |
| TNFRSF10A | Lipodystrophy_2_pPS | 4159 | 0.011325889 | [-0.02 0.04] | 0.465904359 | 0.841554994 |
| TNFRSF10A | Liver-Lipid_pPS | 4159 | -0.020180632 | [-0.05 0.01] | 0.193835223 | 0.696574877 |
| TNFRSF10A | ASAT_Pos_pPS | 4159 | 0.028143788 | [-0. 0.06] | 0.069956246 | 0.491184667 |
| TNFRSF10A | ALP_Neg_pPS | 4159 | -0.002751624 | [-0.03 0.03] | 0.859395412 | 0.977006629 |
| TNFRSF10A | SHBG-LpA_pPS | 4159 | 0.016629659 | [-0.01 0.05] | 0.284324704 | 0.749111537 |
| TNFRSF10A | Obesity_pPS | 4159 | -0.001498439 | [-0.03 0.03] | 0.923150276 | 0.990056084 |
| TNFRSF10A | VAT_Neg_pPS | 4159 | 0.00845188 | [-0.02 0.04] | 0.586356793 | 0.903925569 |
| TNFRSF10A | Bilirubin_pPS | 4159 | 0.000271349 | [-0.03 0.03] | 0.986062606 | 0.998703927 |
| TNFRSF10A | Proinsulin_pPS | 4159 | -0.0135721 | [-0.04 0.02] | 0.382236954 | 0.806027092 |
| TNFRSF10A | Hyper_Insulin_pPS | 4159 | 0.002441876 | [-0.03 0.03] | 0.875085017 | 0.978250438 |
| TNFRSF10A | Beta_Cell_1_pPS | 4159 | -0.020527923 | [-0.05 0.01] | 0.186274196 | 0.687321901 |
| TNFRSF10A | Beta_Cell_2_pPS | 4159 | -0.029630622 | [-0.06 0. ] | 0.056394719 | 0.451781658 |
| TNFRSF10A | Total_GRS | 4159 | 0.01967916 | [-0.01 0.05] | 0.205146876 | 0.708632128 |
| TRAIL_R2 | Cholesterol_pPS | 4159 | 0.021744867 | [-0.01 0.05] | 0.16149774 | 0.664061293 |
| TRAIL_R2 | Lipodystrophy_1_pPS | 4159 | 0.000554193 | [-0.03 0.03] | 0.971539332 | 0.996862516 |
| TRAIL_R2 | Lipodystrophy_2_pPS | 4159 | 0.027011204 | [-0. 0.06] | 0.081992735 | 0.528738199 |
| TRAIL_R2 | Liver-Lipid_pPS | 4159 | 0.00138113 | [-0.03 0.03] | 0.929150167 | 0.990056084 |
| TRAIL_R2 | ASAT_Pos_pPS | 4159 | 0.004067996 | [-0.03 0.03] | 0.793406471 | 0.958013595 |
| TRAIL_R2 | ALP_Neg_pPS | 4159 | 0.011810388 | [-0.02 0.04] | 0.447043502 | 0.831350198 |
| TRAIL_R2 | SHBG-LpA_pPS | 4159 | 0.00829228 | [-0.02 0.04] | 0.593446824 | 0.905925461 |
| TRAIL_R2 | Obesity_pPS | 4159 | 0.032907652 | [0. 0.06] | 0.034082367 | 0.382561193 |
| TRAIL_R2 | VAT_Neg_pPS | 4159 | -0.002415161 | [-0.03 0.03] | 0.876440607 | 0.978550193 |
| TRAIL_R2 | Bilirubin_pPS | 4159 | 0.016153461 | [-0.01 0.05] | 0.298343614 | 0.759754073 |
| TRAIL_R2 | Proinsulin_pPS | 4159 | 0.019087706 | [-0.01 0.05] | 0.219095677 | 0.718175854 |
| TRAIL_R2 | Hyper_Insulin_pPS | 4159 | -0.004721261 | [-0.04 0.03] | 0.761169 | 0.950403646 |
| TRAIL_R2 | Beta_Cell_1_pPS | 4159 | -0.015926291 | [-0.05 0.01] | 0.305191252 | 0.76365449 |

**b) Partial Correlation**

| Proteomics | PPS | Sample_Size | r | CI95% | p | fdr_p_adjusted |
|---|---|---|---|---|---|---|
| TRAIL_R2 | ALP_Neg_pPS | 4159 | 0.008147151 | [-0.02 0.04] | 0.600500405 | 0.918301271 |
| TRAIL_R2 | SHBG-LpA_pPS | 4159 | 0.008768653 | [-0.02 0.04] | 0.573005053 | 0.907786603 |
| TRAIL_R2 | Obesity_pPS | 4159 | 0.029066748 | [-0. 0.06] | 0.061663063 | 0.558815902 |
| TRAIL_R2 | VAT_Neg_pPS | 4159 | -0.002049288 | [-0.03 0.03] | 0.895204436 | 0.98328949 |
| TRAIL_R2 | Bilirubin_pPS | 4159 | 0.016445101 | [-0.01 0.05] | 0.290461808 | 0.81655641 |
| TRAIL_R2 | Proinsulin_pPS | 4159 | 0.019853732 | [-0.01 0.05] | 0.20186396 | 0.765145476 |
| TRAIL_R2 | Hyper_Insulin_pPS | 4159 | -0.008243431 | [-0.04 0.02] | 0.596202141 | 0.918301271 |
| TRAIL_R2 | Beta_Cell_1_pPS | 4159 | -0.016120728 | [-0.05 0.01] | 0.300083061 | 0.821481367 |
| TRAIL_R2 | Beta_Cell_2_pPS | 4159 | -0.017367607 | [-0.05 0.01] | 0.264243495 | 0.80722329 |
| TNFRSF10C | Cholesterol_pPS | 4225 | -0.045896651 | [-0.08 -0.02] | 0.002928688 | 0.123677069 |
| TNFRSF10C | Lipodystrophy_1_pPS | 4225 | 0.001473417 | [-0.03 0.03] | 0.923949483 | 0.990503296 |
| TNFRSF10C | Lipodystrophy_2_pPS | 4225 | 0.018480987 | [-0.01 0.05] | 0.23113097 | 0.786988264 |
| TNFRSF10C | Liver-Lipid_pPS | 4225 | 0.006337618 | [-0.02 0.04] | 0.681361149 | 0.943648559 |
| TNFRSF10C | ASAT_Pos_pPS | 4225 | 0.003115494 | [-0.03 0.03] | 0.84003596 | 0.972178273 |
| TNFRSF10C | ALP_Neg_pPS | 4225 | 0.015765967 | [-0.01 0.05] | 0.307012513 | 0.830127301 |
| TNFRSF10C | SHBG-LpA_pPS | 4225 | 0.002027428 | [-0.03 0.03] | 0.895495786 | 0.98328949 |
| TNFRSF10C | Obesity_pPS | 4225 | -0.013753469 | [-0.04 0.02] | 0.372873642 | 0.847284666 |
| TNFRSF10C | VAT_Neg_pPS | 4225 | -0.010288968 | [-0.04 0.02] | 0.505014563 | 0.895803896 |
| TNFRSF10C | Bilirubin_pPS | 4225 | 0.012624462 | [-0.02 0.04] | 0.413387061 | 0.854426619 |
| TNFRSF10C | Proinsulin_pPS | 4225 | 0.028161026 | [-0. 0.06] | 0.068021899 | 0.568910429 |
| TNFRSF10C | Hyper_Insulin_pPS | 4225 | 0.006292155 | [-0.02 0.04] | 0.683522663 | 0.944323894 |
| TNFRSF10C | Beta_Cell_1_pPS | 4225 | -0.012583555 | [-0.04 0.02] | 0.414902278 | 0.854508456 |
| TNFRSF10C | Beta_Cell_2_pPS | 4225 | -0.018788033 | [-0.05 0.01] | 0.223471564 | 0.780100778 |
| TNFRSF11A | Cholesterol_pPS | 4159 | 0.024788408 | [-0.01 0.06] | 0.111033941 | 0.680721977 |
| TNFRSF11A | Lipodystrophy_1_pPS | 4159 | 0.002409822 | [-0.03 0.03] | 0.876903533 | 0.97781424 |
| TNFRSF11A | Lipodystrophy_2_pPS | 4159 | 0.023664329 | [-0.01 0.05] | 0.128189816 | 0.694065456 |
| TNFRSF11A | Liver-Lipid_pPS | 4159 | 0.028702557 | [-0. 0.06] | 0.064995784 | 0.562394973 |
| TNFRSF11A | ASAT_Pos_pPS | 4159 | -0.010842238 | [-0.04 0.02] | 0.485850207 | 0.895803896 |
| TNFRSF11A | ALP_Neg_pPS | 4159 | -0.026523394 | [-0.06 0. ] | 0.088168805 | 0.620554211 |
| TNFRSF11A | SHBG-LpA_pPS | 4159 | 0.018615824 | [-0.01 0.05] | 0.231435879 | 0.786988264 |
| TNFRSF11A | Obesity_pPS | 4159 | 0.033568146 | [0. 0.06] | 0.030907669 | 0.419027441 |
| TNFRSF11A | VAT_Neg_pPS | 4159 | -0.020452384 | [-0.05 0.01] | 0.188593878 | 0.747409915 |
| TNFRSF11A | Bilirubin_pPS | 4159 | -0.002316132 | [-0.03 0.03] | 0.881653331 | 0.978445416 |
| TNFRSF11A | Proinsulin_pPS | 4159 | 0.028912596 | [-0. 0.06] | 0.063056038 | 0.558815902 |
| TNFRSF11A | Hyper_Insulin_pPS | 4159 | -0.020843391 | [-0.05 0.01] | 0.180280817 | 0.738879686 |
| TNFRSF11A | Beta_Cell_1_pPS | 4159 | 0.007004171 | [-0.02 0.04] | 0.652558435 | 0.936605382 |
| TNFRSF11A | Beta_Cell_2_pPS | 4159 | -0.013596687 | [-0.04 0.02] | 0.382123062 | 0.85037326 |
| OPG | Cholesterol_pPS | 4225 | 0.017883345 | [-0.01 0.05] | 0.246569959 | 0.795945131 |
| OPG | Lipodystrophy_1_pPS | 4225 | 0.011211208 | [-0.02 0.04] | 0.467608363 | 0.879984873 |
| OPG | Lipodystrophy_2_pPS | 4225 | 0.002137063 | [-0.03 0.03] | 0.889879667 | 0.981230818 |
| OPG | Liver-Lipid_pPS | 4225 | -0.009973899 | [-0.04 0.02] | 0.518145807 | 0.895803896 |
| OPG | ASAT_Pos_pPS | 4225 | 0.024386449 | [-0.01 0.05] | 0.114064032 | 0.681046037 |
| OPG | ALP_Neg_pPS | 4225 | -0.028785957 | [-0.06 0. ] | 0.062128934 | 0.558815902 |
| OPG | SHBG-LpA_pPS | 4225 | -0.006224046 | [-0.04 0.02] | 0.686765714 | 0.944571269 |
| OPG | Obesity_pPS | 4225 | 0.001762153 | [-0.03 0.03] | 0.909105664 | 0.988841471 |
| OPG | VAT_Neg_pPS | 4225 | -0.015623407 | [-0.05 0.01] | 0.311407589 | 0.833007217 |
| OPG | Bilirubin_pPS | 4225 | 0.00100384 | [-0.03 0.03] | 0.948144618 | 0.996425456 |
| OPG | Proinsulin_pPS | 4225 | 0.029031519 | [-0. 0.06] | 0.059931941 | 0.558815902 |
| OPG | Hyper_Insulin_pPS | 4225 | 0.00650737 | [-0.02 0.04] | 0.673313546 | 0.939698114 |
| OPG | Beta_Cell_1_pPS | 4225 | -0.022689943 | [-0.05 0.01] | 0.141500038 | 0.697681852 |
| OPG | Beta_Cell_2_pPS | 4225 | -0.007150948 | [-0.04 0.02] | 0.643147664 | 0.933976497 |
| TNFRSF13B | Cholesterol_pPS | 4159 | 0.027209195 | [-0. 0.06] | 0.080249137 | 0.609798752 |
| TNFRSF13B | Lipodystrophy_1_pPS | 4159 | 0.031986945 | [0. 0.06] | 0.039730214 | 0.473819584 |
| TNFRSF13B | Lipodystrophy_2_pPS | 4159 | 0.000353007 | [-0.03 0.03] | 0.981897416 | 0.998803911 |
| TNFRSF13B | Liver-Lipid_pPS | 4159 | -0.000786941 | [-0.03 0.03] | 0.959658614 | 0.996425456 |
| TNFRSF13B | ASAT_Pos_pPS | 4159 | -0.016197823 | [-0.05 0.01] | 0.297777334 | 0.818649319 |
| TNFRSF13B | ALP_Neg_pPS | 4159 | -0.001622825 | [-0.03 0.03] | 0.916923362 | 0.99041009 |
| TNFRSF13B | SHBG-LpA_pPS | 4159 | 0.038563505 | [0.01 0.07] | 0.013150812 | 0.297162209 |
| TNFRSF13B | Obesity_pPS | 4159 | 0.02033321 | [-0.01 0.05] | 0.191182922 | 0.750920235 |
| TNFRSF13B | VAT_Neg_pPS | 4159 | 0.011996723 | [-0.02 0.04] | 0.440625094 | 0.866107938 |
| TNFRSF13B | Bilirubin_pPS | 4159 | 0.010091876 | [-0.02 0.04] | 0.516539127 | 0.895803896 |
| TNFRSF13B | Proinsulin_pPS | 4159 | -0.013335304 | [-0.04 0.02] | 0.391340969 | 0.85037326 |
| TNFRSF13B | Hyper_Insulin_pPS | 4159 | -0.005656256 | [-0.04 0.02] | 0.716180148 | 0.945970482 |
| TNFRSF13B | Beta_Cell_1_pPS | 4159 | 0.031072187 | [0. 0.06] | 0.045749042 | 0.512674866 |
| TNFRSF13B | Beta_Cell_2_pPS | 4159 | 0.002370681 | [-0.03 0.03] | 0.878887318 | 0.97781424 |
| TNFRSF14 | Cholesterol_pPS | 4225 | 0.005960305 | [-0.02 0.04] | 0.699377922 | 0.945970482 |
| TNFRSF14 | Lipodystrophy_1_pPS | 4225 | 0.01663831 | [-0.01 0.05] | 0.281016192 | 0.808820203 |
| TNFRSF14 | Lipodystrophy_2_pPS | 4225 | 0.014144093 | [-0.02 0.04] | 0.359449295 | 0.847284666 |
| TNFRSF14 | Liver-Lipid_pPS | 4225 | 0.017133609 | [-0.01 0.05] | 0.266940552 | 0.808113719 |
| TNFRSF14 | ASAT_Pos_pPS | 4225 | 0.008728503 | [-0.02 0.04] | 0.57172418 | 0.907308519 |
| TNFRSF14 | ALP_Neg_pPS | 4225 | -0.004539691 | [-0.03 0.03] | 0.768666545 | 0.960245786 |
| TNFRSF14 | SHBG-LpA_pPS | 4225 | 0.012568731 | [-0.02 0.04] | 0.415452167 | 0.854508456 |
| TNFRSF14 | Obesity_pPS | 4225 | 0.005233843 | [-0.03 0.04] | 0.734539578 | 0.948434512 |
| TNFRSF14 | VAT_Neg_pPS | 4225 | -0.009088931 | [-0.04 0.02] | 0.555950566 | 0.899977567 |
| TNFRSF14 | Bilirubin_pPS | 4225 | 0.014265987 | [-0.02 0.04] | 0.355323123 | 0.847284666 |
| TNFRSF14 | Proinsulin_pPS | 4225 | 0.017101498 | [-0.01 0.05] | 0.267838124 | 0.808113719 |
| TNFRSF14 | Hyper_Insulin_pPS | 4225 | -0.006879275 | [-0.04 0.02] | 0.65581358 | 0.936605382 |
| TNFRSF14 | Beta_Cell_1_pPS | 4225 | -0.017940965 | [-0.05 0.01] | 0.245050752 | 0.79432283 |
| TNFRSF14 | Beta_Cell_2_pPS | 4225 | 0.014386964 | [-0.02 0.04] | 0.351257687 | 0.847284666 |
| TNF_R1 | Cholesterol_pPS | 4225 | 0.010514362 | [-0.02 0.04] | 0.495729417 | 0.895803896 |
| TNF_R1 | Lipodystrophy_1_pPS | 4225 | 0.005100209 | [-0.03 0.04] | 0.741071222 | 0.951867558 |
| TNF_R1 | Lipodystrophy_2_pPS | 4225 | 0.010166091 | [-0.02 0.04] | 0.510114813 | 0.895803896 |
| TNF_R1 | Liver-Lipid_pPS | 4225 | 0.029022514 | [-0. 0.06] | 0.06001136 | 0.558815902 |
| TNF_R1 | ASAT_Pos_pPS | 4225 | 0.009944217 | [-0.02 0.04] | 0.519391944 | 0.895803896 |
| TNF_R1 | ALP_Neg_pPS | 4225 | 0.001384558 | [-0.03 0.03] | 0.928523271 | 0.991797911 |
| TNF_R1 | SHBG-LpA_pPS | 4225 | 0.017112725 | [-0.01 0.05] | 0.267524072 | 0.808113719 |
| TNF_R1 | Obesity_pPS | 4225 | 0.016749279 | [-0.01 0.05] | 0.277819652 | 0.808820203 |
| TNF_R1 | VAT_Neg_pPS | 4225 | -0.027576326 | [-0.06 0. ] | 0.073944273 | 0.588265012 |
| TNF_R1 | Bilirubin_pPS | 4225 | 0.009650689 | [-0.02 0.04] | 0.531797461 | 0.898959051 |
| TNF_R1 | Proinsulin_pPS | 4225 | 0.012762508 | [-0.02 0.04] | 0.408297994 | 0.853713988 |
| TNF_R1 | Hyper_Insulin_pPS | 4225 | 0.004315495 | [-0.03 0.03] | 0.779788951 | 0.964905159 |
| TNF_R1 | Beta_Cell_1_pPS | 4225 | 0.000622068 | [-0.03 0.03] | 0.967851865 | 0.996924797 |
| TNF_R1 | Beta_Cell_2_pPS | 4225 | -0.00219704 | [-0.03 0.03] | 0.886809649 | 0.979597623 |
| TNF_R2 | Cholesterol_pPS | 4225 | 0.02052709 | [-0.01 0.05] | 0.183501265 | 0.738879686 |
| TNF_R2 | Lipodystrophy_1_pPS | 4225 | 0.01347946 | [-0.02 0.04] | 0.382473164 | 0.85037326 |
| TNF_R2 | Lipodystrophy_2_pPS | 4225 | 0.010728163 | [-0.02 0.04] | 0.48700677 | 0.895803896 |
| TNF_R2 | Liver-Lipid_pPS | 4225 | 0.031953536 | [0. 0.06] | 0.038383271 | 0.46420332 |
| TNF_R2 | ASAT_Pos_pPS | 4225 | -0.006719355 | [-0.04 0.02] | 0.663316207 | 0.937489141 |
| TNF_R2 | ALP_Neg_pPS | 4225 | 0.003030353 | [-0.03 0.03] | 0.844350836 | 0.972178273 |
| TNF_R2 | SHBG-LpA_pPS | 4225 | 0.013863057 | [-0.02 0.04] | 0.369076515 | 0.847284666 |
| TNF_R2 | Obesity_pPS | 4225 | 0.024009615 | [-0.01 0.05] | 0.119764695 | 0.681046037 |
| TNF_R2 | VAT_Neg_pPS | 4225 | -0.014821892 | [-0.05 0.02] | 0.336886528 | 0.844649909 |
| TNF_R2 | Bilirubin_pPS | 4225 | 0.011663323 | [-0.02 0.04] | 0.449846814 | 0.870351501 |
| TNF_R2 | Proinsulin_pPS | 4225 | 0.012235985 | [-0.02 0.04] | 0.427908277 | 0.864060889 |
| TNF_R2 | Hyper_Insulin_pPS | 4225 | -0.002322759 | [-0.03 0.03] | 0.880380093 | 0.977947012 |
| TNF_R2 | Beta_Cell_1_pPS | 4225 | -0.003724761 | [-0.03 0.03] | 0.809306259 | 0.967638637 |
| TNF_R2 | Beta_Cell_2_pPS | 4225 | -0.000464409 | [-0.03 0.03] | 0.975996738 | 0.998477998 |

**Table S7: Associations between pPSs and plasma proteins in SCAPIS**

**a) Spearman Correlation**

| Proteomics | PPS | Sample_Size | r | CI95% | p | fdr_p_adjusted |
|---|---|---|---|---|---|---|
| TRAIL_R2 | Beta_Cell_2_pPS | 4159 | -0.017669349 | [-0.05 0.01] | 0.255286524 | 0.74027471 |
| TRAIL_R2 | Total_GRS | 4159 | 0.030898256 | [0. 0.06] | 0.046630995 | 0.434802524 |
| TNFRSF10C | Cholesterol_pPS | 4225 | -0.045637197 | [-0.08 -0.02] | 0.003047884 | 0.105623173 |
| TNFRSF10C | Lipodystrophy_1_pPS | 4225 | 0.003407199 | [-0.03 0.03] | 0.825023747 | 0.964449615 |
| TNFRSF10C | Lipodystrophy_2_pPS | 4225 | 0.01734527 | [-0.01 0.05] | 0.260338736 | 0.743824961 |
| TNFRSF10C | Liver-Lipid_pPS | 4225 | 0.005840743 | [-0.02 0.04] | 0.704688221 | 0.937320237 |
| TNFRSF10C | ASAT_Pos_pPS | 4225 | 0.005937167 | [-0.02 0.04] | 0.70004741 | 0.93384768 |
| TNFRSF10C | ALP_Neg_pPS | 4225 | 0.020781053 | [-0.01 0.05] | 0.177467353 | 0.678995855 |
| TNFRSF10C | SHBG-LpA_pPS | 4225 | 0.002893513 | [-0.03 0.03] | 0.851068124 | 0.97528582 |
| TNFRSF10C | Obesity_pPS | 4225 | -0.01409589 | [-0.04 0.02] | 0.360346688 | 0.7979754 |
| TNFRSF10C | VAT_Neg_pPS | 4225 | -0.007879015 | [-0.04 0.02] | 0.609165987 | 0.909483337 |
| TNFRSF10C | Bilirubin_pPS | 4225 | 0.012164942 | [-0.02 0.04] | 0.429882678 | 0.827992564 |
| TNFRSF10C | Proinsulin_pPS | 4225 | 0.023466686 | [-0.01 0.05] | 0.127777562 | 0.623575746 |
| TNFRSF10C | Hyper_Insulin_pPS | 4225 | 0.005885196 | [-0.02 0.04] | 0.702547349 | 0.936277491 |
| TNFRSF10C | Beta_Cell_1_pPS | 4225 | -0.010172395 | [-0.04 0.02] | 0.509197885 | 0.868660083 |
| TNFRSF10C | Beta_Cell_2_pPS | 4225 | -0.018259856 | [-0.05 0.01] | 0.23603681 | 0.726631332 |
| TNFRSF10C | Total_GRS | 4225 | -0.002283092 | [-0.03 0.03] | 0.882227287 | 0.979291846 |
| TNFRSF11A | Cholesterol_pPS | 4159 | 0.030191944 | [-0. 0.06] | 0.051879312 | 0.441027526 |
| TNFRSF11A | Lipodystrophy_1_pPS | 4159 | 0.001411527 | [-0.03 0.03] | 0.927595041 | 0.990056084 |
| TNFRSF11A | Lipodystrophy_2_pPS | 4159 | 0.020716454 | [-0.01 0.05] | 0.182261983 | 0.684412345 |
| TNFRSF11A | Liver-Lipid_pPS | 4159 | 0.029039158 | [-0. 0.06] | 0.061501713 | 0.466331669 |
| TNFRSF11A | ASAT_Pos_pPS | 4159 | -0.008269946 | [-0.04 0.02] | 0.594442096 | 0.906943165 |
| TNFRSF11A | ALP_Neg_pPS | 4159 | -0.020964491 | [-0.05 0.01] | 0.177081431 | 0.678995855 |
| TNFRSF11A | SHBG-LpA_pPS | 4159 | 0.018817306 | [-0.01 0.05] | 0.225694701 | 0.724402982 |
| TNFRSF11A | Obesity_pPS | 4159 | 0.038293392 | [0.01 0.07] | 0.01365747 | 0.244770243 |
| TNFRSF11A | VAT_Neg_pPS | 4159 | -0.018414604 | [-0.05 0.01] | 0.235783439 | 0.726631332 |
| TNFRSF11A | Bilirubin_pPS | 4159 | -0.00240231 | [-0.03 0.03] | 0.877092812 | 0.978705778 |
| TNFRSF11A | Proinsulin_pPS | 4159 | 0.031711485 | [0. 0.06] | 0.041148664 | 0.407062052 |
| TNFRSF11A | Hyper_Insulin_pPS | 4159 | -0.018236216 | [-0.05 0.01] | 0.240353008 | 0.729784711 |
| TNFRSF11A | Beta_Cell_1_pPS | 4159 | 0.010454262 | [-0.02 0.04] | 0.500921409 | 0.863780476 |
| TNFRSF11A | Beta_Cell_2_pPS | 4159 | -0.006654511 | [-0.04 0.02] | 0.668354487 | 0.924640793 |
| TNFRSF11A | Total_GRS | 4159 | 0.026418596 | [-0. 0.06] | 0.088930272 | 0.546245321 |
| OPG | Cholesterol_pPS | 4225 | 0.019089471 | [-0.01 0.05] | 0.215420522 | 0.715705689 |
| OPG | Lipodystrophy_1_pPS | 4225 | 0.014792092 | [-0.02 0.04] | 0.337111985 | 0.782502359 |
| OPG | Lipodystrophy_2_pPS | 4225 | 0.004566458 | [-0.03 0.03] | 0.766992184 | 0.952160028 |
| OPG | Liver-Lipid_pPS | 4225 | -0.010429417 | [-0.04 0.02] | 0.498554303 | 0.863780476 |
| OPG | ASAT_Pos_pPS | 4225 | 0.026661692 | [-0. 0.06] | 0.083569332 | 0.534925551 |
| OPG | ALP_Neg_pPS | 4225 | -0.02397231 | [-0.05 0.01] | 0.119768129 | 0.615837655 |
| OPG | SHBG-LpA_pPS | 4225 | -0.00452042 | [-0.03 0.03] | 0.769274365 | 0.952961063 |
| OPG | Obesity_pPS | 4225 | 0.007760803 | [-0.02 0.04] | 0.614547146 | 0.909483337 |
| OPG | VAT_Neg_pPS | 4225 | -0.01813099 | [-0.05 0.01] | 0.239360392 | 0.728373408 |
| OPG | Bilirubin_pPS | 4225 | 0.001440441 | [-0.03 0.03] | 0.925531864 | 0.990056084 |
| OPG | Proinsulin_pPS | 4225 | 0.017718558 | [-0.01 0.05] | 0.250218608 | 0.738661262 |
| OPG | Hyper_Insulin_pPS | 4225 | 0.007693576 | [-0.02 0.04] | 0.617616673 | 0.909483337 |
| OPG | Beta_Cell_1_pPS | 4225 | -0.01866463 | [-0.05 0.01] | 0.225810005 | 0.724402982 |
| OPG | Beta_Cell_2_pPS | 4225 | -0.010106705 | [-0.04 0.02] | 0.511937171 | 0.870690038 |
| OPG | Total_GRS | 4225 | 0.015684039 | [-0.01 0.05] | 0.308785264 | 0.76365449 |
| TNFRSF13B | Cholesterol_pPS | 4159 | 0.027716441 | [-0. 0.06] | 0.074315496 | 0.505198939 |
| TNFRSF13B | Lipodystrophy_1_pPS | 4159 | 0.034235869 | [0. 0.06] | 0.027476847 | 0.352726036 |
| TNFRSF13B | Lipodystrophy_2_pPS | 4159 | 0.002518342 | [-0.03 0.03] | 0.871206991 | 0.977006629 |
| TNFRSF13B | Liver-Lipid_pPS | 4159 | 0.000982532 | [-0.03 0.03] | 0.949564822 | 0.993154165 |
| TNFRSF13B | ASAT_Pos_pPS | 4159 | -0.009067495 | [-0.04 0.02] | 0.55938359 | 0.897093962 |
| TNFRSF13B | ALP_Neg_pPS | 4159 | -0.003919763 | [-0.03 0.03] | 0.800773118 | 0.958074488 |
| TNFRSF13B | SHBG-LpA_pPS | 4159 | 0.040605743 | [0.01 0.07] | 0.008917872 | 0.205111057 |
| TNFRSF13B | Obesity_pPS | 4159 | 0.022117051 | [-0.01 0.05] | 0.154440382 | 0.650979658 |
| TNFRSF13B | VAT_Neg_pPS | 4159 | 0.017995017 | [-0.01 0.05] | 0.246630184 | 0.736582988 |
| TNFRSF13B | Bilirubin_pPS | 4159 | 0.0111489 | [-0.02 0.04] | 0.472902913 | 0.846453261 |
| TNFRSF13B | Proinsulin_pPS | 4159 | 0.000802874 | [-0.03 0.03] | 0.958777892 | 0.99612031 |
| TNFRSF13B | Hyper_Insulin_pPS | 4159 | 0.000646665 | [-0.03 0.03] | 0.966792942 | 0.996862516 |
| TNFRSF13B | Beta_Cell_1_pPS | 4159 | 0.032132173 | [0. 0.06] | 0.038533649 | 0.398325363 |
| TNFRSF13B | Beta_Cell_2_pPS | 4159 | 0.010795163 | [-0.02 0.04] | 0.487062208 | 0.855691721 |
| TNFRSF13B | Total_GRS | 4159 | 0.032289262 | [0. 0.06] | 0.037594053 | 0.394389052 |
| TNFRSF14 | Cholesterol_pPS | 4225 | 0.0081324 | [-0.02 0.04] | 0.597702793 | 0.907623171 |
| TNFRSF14 | Lipodystrophy_1_pPS | 4225 | 0.019372315 | [-0.01 0.05] | 0.208697224 | 0.709689105 |
| TNFRSF14 | Lipodystrophy_2_pPS | 4225 | 0.016266993 | [-0.01 0.05] | 0.291147152 | 0.753421877 |
| TNFRSF14 | Liver-Lipid_pPS | 4225 | 0.017583319 | [-0.01 0.05] | 0.253852836 | 0.739680398 |
| TNFRSF14 | ASAT_Pos_pPS | 4225 | 0.01262419 | [-0.02 0.04] | 0.412674763 | 0.8164748 |
| TNFRSF14 | ALP_Neg_pPS | 4225 | -0.00071364 | [-0.03 0.03] | 0.963065614 | 0.99612031 |
| TNFRSF14 | SHBG-LpA_pPS | 4225 | 0.014096388 | [-0.02 0.04] | 0.360329708 | 0.7979754 |
| TNFRSF14 | Obesity_pPS | 4225 | 0.008919978 | [-0.02 0.04] | 0.562714494 | 0.897748415 |
| TNFRSF14 | VAT_Neg_pPS | 4225 | -0.006984908 | [-0.04 0.02] | 0.650372477 | 0.919948884 |
| TNFRSF14 | Bilirubin_pPS | 4225 | 0.014386072 | [-0.02 0.04] | 0.350542782 | 0.793480163 |
| TNFRSF14 | Proinsulin_pPS | 4225 | 0.016693776 | [-0.01 0.05] | 0.278672048 | 0.747422497 |
| TNFRSF14 | Hyper_Insulin_pPS | 4225 | -0.00259857 | [-0.03 0.03] | 0.866097719 | 0.977006629 |
| TNFRSF14 | Beta_Cell_1_pPS | 4225 | -0.009648486 | [-0.04 0.02] | 0.531256518 | 0.877412567 |
| TNFRSF14 | Beta_Cell_2_pPS | 4225 | 0.013523956 | [-0.02 0.04] | 0.380167591 | 0.805487278 |
| TNFRSF14 | Total_GRS | 4225 | 0.016490537 | [-0.01 0.05] | 0.284566759 | 0.749111537 |
| TNF_R1 | Cholesterol_pPS | 4225 | 0.014182514 | [-0.02 0.04] | 0.357402208 | 0.7979754 |
| TNF_R1 | Lipodystrophy_1_pPS | 4225 | 0.009744363 | [-0.02 0.04] | 0.527183813 | 0.873942847 |
| TNF_R1 | Lipodystrophy_2_pPS | 4225 | 0.009877753 | [-0.02 0.04] | 0.521544179 | 0.872980599 |
| TNF_R1 | Liver-Lipid_pPS | 4225 | 0.030083723 | [-0. 0.06] | 0.050875842 | 0.440215663 |
| TNF_R1 | ASAT_Pos_pPS | 4225 | 0.013264481 | [-0.02 0.04] | 0.389376639 | 0.810650617 |
| TNF_R1 | ALP_Neg_pPS | 4225 | 0.005007123 | [-0.03 0.04] | 0.745252181 | 0.947005534 |
| TNF_R1 | SHBG-LpA_pPS | 4225 | 0.016225575 | [-0.01 0.05] | 0.292377486 | 0.75346579 |
| TNF_R1 | Obesity_pPS | 4225 | 0.020842205 | [-0.01 0.05] | 0.176195116 | 0.678995855 |
| TNF_R1 | VAT_Neg_pPS | 4225 | -0.024475564 | [-0.05 0.01] | 0.112192013 | 0.604978148 |
| TNF_R1 | Bilirubin_pPS | 4225 | 0.009517983 | [-0.02 0.04] | 0.536825635 | 0.881309122 |
| TNF_R1 | Proinsulin_pPS | 4225 | 0.01716539 | [-0.01 0.05] | 0.265315165 | 0.746520328 |
| TNF_R1 | Hyper_Insulin_pPS | 4225 | 0.006782145 | [-0.02 0.04] | 0.659873852 | 0.924024268 |
| TNF_R1 | Beta_Cell_1_pPS | 4225 | 0.002533062 | [-0.03 0.03] | 0.869442611 | 0.977006629 |
| TNF_R1 | Beta_Cell_2_pPS | 4225 | 2.74E-05 | [-0.03 0.03] | 0.998582022 | 0.999464614 |
| TNF_R1 | Total_GRS | 4225 | 0.019279083 | [-0.01 0.05] | 0.210896359 | 0.710713005 |
| TNF_R2 | Cholesterol_pPS | 4225 | 0.024821173 | [-0.01 0.05] | 0.10721165 | 0.597786172 |
| TNF_R2 | Lipodystrophy_1_pPS | 4225 | 0.015035397 | [-0.02 0.05] | 0.329224309 | 0.775125841 |
| TNF_R2 | Lipodystrophy_2_pPS | 4225 | 0.011038031 | [-0.02 0.04] | 0.473829815 | 0.846453261 |
| TNF_R2 | Liver-Lipid_pPS | 4225 | 0.033196336 | [0. 0.06] | 0.031189248 | 0.369452038 |
| TNF_R2 | ASAT_Pos_pPS | 4225 | -0.002487644 | [-0.03 0.03] | 0.871763092 | 0.977006629 |
| TNF_R2 | ALP_Neg_pPS | 4225 | 0.00697024 | [-0.02 0.04] | 0.651057929 | 0.919948884 |
| TNF_R2 | SHBG-LpA_pPS | 4225 | 0.014014221 | [-0.02 0.04] | 0.363136632 | 0.7979754 |
| TNF_R2 | Obesity_pPS | 4225 | 0.028432807 | [-0. 0.06] | 0.064989339 | 0.475856102 |
| TNF_R2 | VAT_Neg_pPS | 4225 | -0.012841404 | [-0.04 0.02] | 0.404680031 | 0.81230319 |
| TNF_R2 | Bilirubin_pPS | 4225 | 0.011348211 | [-0.02 0.04] | 0.461493048 | 0.83934244 |
| TNF_R2 | Proinsulin_pPS | 4225 | 0.017080664 | [-0.01 0.05] | 0.267681664 | 0.746520328 |
| TNF_R2 | Hyper_Insulin_pPS | 4225 | 0.001613385 | [-0.03 0.03] | 0.916621809 | 0.990056084 |
| TNF_R2 | Beta_Cell_1_pPS | 4225 | -0.000589896 | [-0.03 0.03] | 0.969466511 | 0.996862516 |
| TNF_R2 | Beta_Cell_2_pPS | 4225 | 0.001890767 | [-0.03 0.03] | 0.902353467 | 0.987003157 |
| TNF_R2 | Total_GRS | 4225 | 0.023013318 | [-0.01 0.05] | 0.135307826 | 0.629316669 |

**b) Partial Correlation**

| Proteomics | PPS | Sample_Size | r | CI95% | p | fdr_p_adjusted |
|---|---|---|---|---|---|---|
| TNFSF13B | Cholesterol_pPS | 4225 | -0.009248819 | [-0.04 0.02] | 0.549021927 | 0.899241939 |
| TNFSF13B | Lipodystrophy_1_pPS | 4225 | 0.014452496 | [-0.02 0.04] | 0.349067851 | 0.847284666 |
| TNFSF13B | Lipodystrophy_2_pPS | 4225 | 0.027181607 | [-0. 0.06] | 0.078175708 | 0.602398521 |
| TNFSF13B | Liver-Lipid_pPS | 4225 | 0.008898259 | [-0.02 0.04] | 0.564268593 | 0.904956448 |
| TNFSF13B | ASAT_Pos_pPS | 4225 | -0.016329958 | [-0.05 0.01] | 0.290029109 | 0.81655641 |
| TNFSF13B | ALP_Neg_pPS | 4225 | 0.012082994 | [-0.02 0.04] | 0.433707581 | 0.864060889 |
| TNFSF13B | SHBG-LpA_pPS | 4225 | -0.014291741 | [-0.04 0.02] | 0.354455211 | 0.847284666 |
| TNFSF13B | Obesity_pPS | 4225 | 0.020076746 | [-0.01 0.05] | 0.193304436 | 0.752352981 |
| TNFSF13B | VAT_Neg_pPS | 4225 | 0.008015488 | [-0.02 0.04] | 0.603539895 | 0.918301271 |
| TNFSF13B | Bilirubin_pPS | 4225 | 0.005576725 | [-0.02 0.04] | 0.717868966 | 0.945970482 |
| TNFSF13B | Proinsulin_pPS | 4225 | 0.02660061 | [-0. 0.06] | 0.0847605 | 0.614725404 |
| TNFSF13B | Hyper_Insulin_pPS | 4225 | 0.002519035 | [-0.03 0.03] | 0.870358005 | 0.977511154 |
| TNFSF13B | Beta_Cell_1_pPS | 4225 | 2.06E-05 | [-0.03 0.03] | 0.998934312 | 0.999322248 |
| TNFSF13B | Beta_Cell_2_pPS | 4225 | 0.009279106 | [-0.02 0.04] | 0.547714296 | 0.899241939 |
| XCL1 | Cholesterol_pPS | 4159 | 0.028857413 | [-0. 0.06] | 0.06356097 | 0.558815902 |
| XCL1 | Lipodystrophy_1_pPS | 4159 | 0.011042789 | [-0.02 0.04] | 0.477818131 | 0.888066022 |
| XCL1 | Lipodystrophy_2_pPS | 4159 | -0.000664396 | [-0.03 0.03] | 0.965936523 | 0.996924797 |
| XCL1 | Liver-Lipid_pPS | 4159 | 0.036457922 | [0.01 0.07] | 0.019069177 | 0.340663962 |
| XCL1 | ASAT_Pos_pPS | 4159 | 0.016032071 | [-0.01 0.05] | 0.302749277 | 0.826182442 |
| XCL1 | ALP_Neg_pPS | 4159 | -0.016216105 | [-0.05 0.01] | 0.297232305 | 0.81802395 |
| XCL1 | SHBG-LpA_pPS | 4159 | 0.006750197 | [-0.02 0.04] | 0.664371494 | 0.937489141 |
| XCL1 | Obesity_pPS | 4159 | 0.00996689 | [-0.02 0.04] | 0.521746745 | 0.895803896 |
| XCL1 | VAT_Neg_pPS | 4159 | 0.028605737 | [-0. 0.06] | 0.065906363 | 0.563434057 |
| XCL1 | Bilirubin_pPS | 4159 | -0.015697505 | [-0.05 0.01] | 0.312952245 | 0.833834311 |
| XCL1 | Proinsulin_pPS | 4159 | -0.00063452 | [-0.03 0.03] | 0.967467408 | 0.996924797 |
| XCL1 | Hyper_Insulin_pPS | 4159 | 0.035838079 | [0.01 0.07] | 0.021206074 | 0.354719791 |
| XCL1 | Beta_Cell_1_pPS | 4159 | -0.005880873 | [-0.04 0.02] | 0.705425637 | 0.945970482 |
| XCL1 | Beta_Cell_2_pPS | 4159 | -0.025747559 | [-0.06 0. ] | 0.097875072 | 0.64755296 |
| BOC | Cholesterol_pPS | 4159 | -0.001376372 | [-0.03 0.03] | 0.929504109 | 0.991797911 |
| BOC | Lipodystrophy_1_pPS | 4159 | 0.012467203 | [-0.02 0.04] | 0.422910309 | 0.860518922 |
| BOC | Lipodystrophy_2_pPS | 4159 | 0.022042283 | [-0.01 0.05] | 0.156489246 | 0.708464493 |
| BOC | Liver-Lipid_pPS | 4159 | 0.032122159 | [0. 0.06] | 0.038900227 | 0.467187506 |
| BOC | ASAT_Pos_pPS | 4159 | -0.023435226 | [-0.05 0.01] | 0.131926821 | 0.694065456 |
| BOC | ALP_Neg_pPS | 4159 | 0.010578461 | [-0.02 0.04] | 0.496524697 | 0.895803896 |
| BOC | SHBG-LpA_pPS | 4159 | 0.010798001 | [-0.02 0.04] | 0.487631652 | 0.895803896 |
| BOC | Obesity_pPS | 4159 | -0.009970832 | [-0.04 0.02] | 0.521582097 | 0.895803896 |
| BOC | VAT_Neg_pPS | 4159 | 0.015253947 | [-0.02 0.05] | 0.326824013 | 0.84456682 |
| BOC | Bilirubin_pPS | 4159 | -0.012826631 | [-0.04 0.02] | 0.40966233 | 0.854336605 |
| BOC | Proinsulin_pPS | 4159 | -0.006369209 | [-0.04 0.02] | 0.682248935 | 0.943830609 |
| BOC | Hyper_Insulin_pPS | 4159 | -0.00031239 | [-0.03 0.03] | 0.98398002 | 0.998803911 |
| BOC | Beta_Cell_1_pPS | 4159 | -0.025735688 | [-0.06 0. ] | 0.098029969 | 0.64755296 |
| BOC | Beta_Cell_2_pPS | 4159 | 0.02339365 | [-0.01 0.05] | 0.132613952 | 0.694065456 |
| CNTN1 | Cholesterol_pPS | 4225 | -0.006569831 | [-0.04 0.02] | 0.670361777 | 0.937996706 |
| CNTN1 | Lipodystrophy_1_pPS | 4225 | 0.017192625 | [-0.01 0.05] | 0.265296286 | 0.80722329 |
| CNTN1 | Lipodystrophy_2_pPS | 4225 | 0.007944778 | [-0.02 0.04] | 0.606738004 | 0.918301271 |
| CNTN1 | Liver-Lipid_pPS | 4225 | 0.00426315 | [-0.03 0.03] | 0.782392391 | 0.964905159 |
| CNTN1 | ASAT_Pos_pPS | 4225 | 0.01895935 | [-0.01 0.05] | 0.219277719 | 0.775610053 |
| CNTN1 | ALP_Neg_pPS | 4225 | 0.011953355 | [-0.02 0.04] | 0.438657019 | 0.865276785 |
| CNTN1 | SHBG-LpA_pPS | 4225 | 0.012063764 | [-0.02 0.04] | 0.434439693 | 0.864182741 |
| CNTN1 | Obesity_pPS | 4225 | -0.013173419 | [-0.04 0.02] | 0.393372086 | 0.85037326 |
| CNTN1 | VAT_Neg_pPS | 4225 | 0.019740096 | [-0.01 0.05] | 0.200880064 | 0.765145476 |
| CNTN1 | Bilirubin_pPS | 4225 | -0.026457176 | [-0.06 0. ] | 0.086453327 | 0.615938909 |
| CNTN1 | Proinsulin_pPS | 4225 | -0.003537199 | [-0.03 0.03] | 0.818737404 | 0.968668398 |
| CNTN1 | Hyper_Insulin_pPS | 4225 | 0.002858894 | [-0.03 0.03] | 0.85305435 | 0.974979087 |
| CNTN1 | Beta_Cell_1_pPS | 4225 | -0.01075787 | [-0.04 0.02] | 0.485801357 | 0.895803896 |
| CNTN1 | Beta_Cell_2_pPS | 4225 | 0.033438793 | [0. 0.06] | 0.030231623 | 0.416452728 |
| CPB1 | Cholesterol_pPS | 4225 | -0.014846692 | [-0.05 0.02] | 0.336078634 | 0.844649909 |
| CPB1 | Lipodystrophy_1_pPS | 4225 | 0.04983516 | [0.02 0.08] | 0.00123473 | 0.07228782 |
| CPB1 | Lipodystrophy_2_pPS | 4225 | 0.021412992 | [-0.01 0.05] | 0.165299554 | 0.720493489 |
| CPB1 | Liver-Lipid_pPS | 4225 | -0.044114557 | [-0.07 -0.01] | 0.004243213 | 0.153950943 |
| CPB1 | ASAT_Pos_pPS | 4225 | -0.021241237 | [-0.05 0.01] | 0.168717945 | 0.723157113 |
| CPB1 | ALP_Neg_pPS | 4225 | -0.072413725 | [-0.1 -0.04] | 2.63E-06 | 0.000398652 |
| CPB1 | SHBG-LpA_pPS | 4225 | -0.037859462 | [-0.07 -0.01] | 0.014138589 | 0.312663737 |
| CPB1 | Obesity_pPS | 4225 | -0.001016948 | [-0.03 0.03] | 0.947468475 | 0.996425456 |
| CPB1 | VAT_Neg_pPS | 4225 | -0.009117738 | [-0.04 0.02] | 0.554699083 | 0.899898936 |
| CPB1 | Bilirubin_pPS | 4225 | 0.001871902 | [-0.03 0.03] | 0.903471552 | 0.986998608 |
| CPB1 | Proinsulin_pPS | 4225 | 0.003476629 | [-0.03 0.03] | 0.821788746 | 0.968668398 |
| CPB1 | Hyper_Insulin_pPS | 4225 | -0.003768188 | [-0.03 0.03] | 0.807126514 | 0.967638637 |
| CPB1 | Beta_Cell_1_pPS | 4225 | 0.003924368 | [-0.03 0.03] | 0.799299819 | 0.967638637 |
| CPB1 | Beta_Cell_2_pPS | 4225 | 0.00026287 | [-0.03 0.03] | 0.986411993 | 0.998803911 |
| DCN | Cholesterol_pPS | 4159 | 0.006957321 | [-0.02 0.04] | 0.654731113 | 0.936605382 |
| DCN | Lipodystrophy_1_pPS | 4159 | -0.01681723 | [-0.05 0.01] | 0.279682333 | 0.808820203 |
| DCN | Lipodystrophy_2_pPS | 4159 | 0.006951215 | [-0.02 0.04] | 0.655014492 | 0.936605382 |
| DCN | Liver-Lipid_pPS | 4159 | 0.042847958 | [0.01 0.07] | 0.005862173 | 0.184158005 |
| DCN | ASAT_Pos_pPS | 4159 | -0.019877256 | [-0.05 0.01] | 0.201329989 | 0.765145476 |
| DCN | ALP_Neg_pPS | 4159 | -0.027954727 | [-0.06 0. ] | 0.072305445 | 0.582058829 |
| DCN | SHBG-LpA_pPS | 4159 | 0.018258378 | [-0.01 0.05] | 0.240520449 | 0.790085251 |
| DCN | Obesity_pPS | 4159 | 0.017427299 | [-0.01 0.05] | 0.262605056 | 0.80722329 |
| DCN | VAT_Neg_pPS | 4159 | 0.021292532 | [-0.01 0.05] | 0.17107123 | 0.726214013 |
| DCN | Bilirubin_pPS | 4159 | -0.010692823 | [-0.04 0.02] | 0.491881402 | 0.895803896 |
| DCN | Proinsulin_pPS | 4159 | 0.017175811 | [-0.01 0.05] | 0.269555541 | 0.808113719 |
| DCN | Hyper_Insulin_pPS | 4159 | 0.016320993 | [-0.01 0.05] | 0.294118216 | 0.817312323 |
| DCN | Beta_Cell_1_pPS | 4159 | -0.019306605 | [-0.05 0.01] | 0.214575688 | 0.767704128 |
| DCN | Beta_Cell_2_pPS | 4159 | -0.000970326 | [-0.03 0.03] | 0.950268662 | 0.996425456 |
| Dkk_1 | Cholesterol_pPS | 4159 | -0.007781787 | [-0.04 0.02] | 0.616937423 | 0.921525431 |
| Dkk_1 | Lipodystrophy_1_pPS | 4159 | 0.003774301 | [-0.03 0.03] | 0.808314765 | 0.967638637 |
| Dkk_1 | Lipodystrophy_2_pPS | 4159 | 0.014663823 | [-0.02 0.05] | 0.34588887 | 0.847284666 |
| Dkk_1 | Liver-Lipid_pPS | 4159 | -0.000922801 | [-0.03 0.03] | 0.952701465 | 0.996425456 |
| Dkk_1 | ASAT_Pos_pPS | 4159 | 0.004273893 | [-0.03 0.03] | 0.783536945 | 0.965736945 |
| Dkk_1 | ALP_Neg_pPS | 4159 | 0.001775345 | [-0.03 0.03] | 0.909147822 | 0.988841471 |
| Dkk_1 | SHBG-LpA_pPS | 4159 | -0.014174425 | [-0.04 0.02] | 0.362225987 | 0.847284666 |
| Dkk_1 | Obesity_pPS | 4159 | -0.024518176 | [-0.05 0.01] | 0.114983056 | 0.681046037 |
| Dkk_1 | VAT_Neg_pPS | 4159 | -0.023882022 | [-0.05 0.01] | 0.12471565 | 0.692386883 |
| Dkk_1 | Bilirubin_pPS | 4159 | 0.009115985 | [-0.02 0.04] | 0.557903729 | 0.901040756 |
| Dkk_1 | Proinsulin_pPS | 4159 | 0.014350407 | [-0.02 0.04] | 0.356296535 | 0.847284666 |
| Dkk_1 | Hyper_Insulin_pPS | 4159 | -0.028027816 | [-0.06 0. ] | 0.071562591 | 0.578232608 |
| Dkk_1 | Beta_Cell_1_pPS | 4159 | -0.01615385 | [-0.05 0.01] | 0.299091014 | 0.820509533 |
| Dkk_1 | Beta_Cell_2_pPS | 4159 | 0.002065275 | [-0.03 0.03] | 0.894391644 | 0.98328949 |
| IgG_Fc_receptor_II_b | Cholesterol_pPS | 4159 | -0.014639175 | [-0.05 0.02] | 0.34670028 | 0.847284666 |
| IgG_Fc_receptor_II_b | Lipodystrophy_1_pPS | 4159 | -0.000355416 | [-0.03 0.03] | 0.981773905 | 0.998803911 |
| IgG_Fc_receptor_II_b | Lipodystrophy_2_pPS | 4159 | 0.001677723 | [-0.03 0.03] | 0.914123672 | 0.989461462 |
| IgG_Fc_receptor_II_b | Liver-Lipid_pPS | 4159 | 0.001925755 | [-0.03 0.03] | 0.901488372 | 0.985399384 |
| IgG_Fc_receptor_II_b | ASAT_Pos_pPS | 4159 | 0.000910721 | [-0.03 0.03] | 0.953319936 | 0.996425456 |
| IgG_Fc_receptor_II_b | ALP_Neg_pPS | 4159 | 0.015037956 | [-0.02 0.05] | 0.333721252 | 0.844649909 |
| IgG_Fc_receptor_II_b | SHBG-LpA_pPS | 4159 | 0.004028304 | [-0.03 0.03] | 0.795691332 | 0.967638637 |
| IgG_Fc_receptor_II_b | Obesity_pPS | 4159 | 0.006123987 | [-0.02 0.04] | 0.693851551 | 0.945970482 |
| IgG_Fc_receptor_II_b | VAT_Neg_pPS | 4159 | 0.014784067 | [-0.02 0.05] | 0.341947837 | 0.847284666 |

**Table S7: Associations between pPSs and plasma proteins in SCAPIS**

**a) Spearman Correlation**

| Proteomics | PPS | Sample_Size | r | CI95% | p | fdr_p_adjusted |
|---|---|---|---|---|---|---|
| TNFSF13B | Cholesterol_pPS | 4225 | -0.005409268 | [-0.04 0.02] | 0.725587508 | 0.942134248 |
| TNFSF13B | Lipodystrophy_1_pPS | 4225 | 0.014326562 | [-0.02 0.04] | 0.352539446 | 0.796092727 |
| TNFSF13B | Lipodystrophy_2_pPS | 4225 | 0.028395317 | [-0. 0.06] | 0.065344008 | 0.475856102 |
| TNFSF13B | Liver-Lipid_pPS | 4225 | 0.011454666 | [-0.02 0.04] | 0.457300701 | 0.835860883 |
| TNFSF13B | ASAT_Pos_pPS | 4225 | -0.009770074 | [-0.04 0.02] | 0.526094345 | 0.873942847 |
| TNFSF13B | ALP_Neg_pPS | 4225 | 0.018775925 | [-0.01 0.05] | 0.223054348 | 0.7224931 |
| TNFSF13B | SHBG-LpA_pPS | 4225 | -0.011439679 | [-0.04 0.02] | 0.457889641 | 0.83593406 |
| TNFSF13B | Obesity_pPS | 4225 | 0.023810403 | [-0.01 0.05] | 0.122288906 | 0.615837655 |
| TNFSF13B | VAT_Neg_pPS | 4225 | 0.01343775 | [-0.02 0.04] | 0.383212177 | 0.806170656 |
| TNFSF13B | Bilirubin_pPS | 4225 | 0.006624006 | [-0.02 0.04] | 0.667322464 | 0.924640793 |
| TNFSF13B | Proinsulin_pPS | 4225 | 0.034939652 | [0. 0.07] | 0.023337173 | 0.325306047 |
| TNFSF13B | Hyper_Insulin_pPS | 4225 | 0.007502129 | [-0.02 0.04] | 0.62639458 | 0.913767992 |
| TNFSF13B | Beta_Cell_1_pPS | 4225 | 0.012128967 | [-0.02 0.04] | 0.43124802 | 0.827992564 |
| TNFSF13B | Beta_Cell_2_pPS | 4225 | 0.016772077 | [-0.01 0.05] | 0.27642331 | 0.747422497 |
| TNFSF13B | Total_GRS | 4225 | 0.040049677 | [0.01 0.07] | 0.00932778 | 0.212765894 |
| XCL1 | Cholesterol_pPS | 4159 | 0.03035727 | [-0. 0.06] | 0.050608514 | 0.440215663 |
| XCL1 | Lipodystrophy_1_pPS | 4159 | 0.020314144 | [-0.01 0.05] | 0.190902257 | 0.693468715 |
| XCL1 | Lipodystrophy_2_pPS | 4159 | 0.00933181 | [-0.02 0.04] | 0.547990275 | 0.891251125 |
| XCL1 | Liver-Lipid_pPS | 4159 | 0.038554634 | [0.01 0.07] | 0.013028646 | 0.242414912 |
| XCL1 | ASAT_Pos_pPS | 4159 | 0.02199486 | [-0.01 0.05] | 0.156731056 | 0.655420779 |
| XCL1 | ALP_Neg_pPS | 4159 | -0.014558733 | [-0.04 0.02] | 0.348599755 | 0.793186581 |
| XCL1 | SHBG-LpA_pPS | 4159 | 0.00994927 | [-0.02 0.04] | 0.521829476 | 0.872980599 |
| XCL1 | Obesity_pPS | 4159 | 0.01554674 | [-0.01 0.05] | 0.316862951 | 0.767141882 |
| XCL1 | VAT_Neg_pPS | 4159 | 0.029691866 | [-0. 0.06] | 0.055886678 | 0.451781658 |
| XCL1 | Bilirubin_pPS | 4159 | -0.016108958 | [-0.05 0.01] | 0.299676938 | 0.762311842 |
| XCL1 | Proinsulin_pPS | 4159 | -0.006053566 | [-0.04 0.02] | 0.696744499 | 0.930772879 |
| XCL1 | Hyper_Insulin_pPS | 4159 | 0.04375201 | [0.01 0.07] | 0.004832274 | 0.137495623 |
| XCL1 | Beta_Cell_1_pPS | 4159 | -0.008714442 | [-0.04 0.02] | 0.574778994 | 0.903411176 |
| XCL1 | Beta_Cell_2_pPS | 4159 | -0.024624066 | [-0.06 0.01] | 0.112856556 | 0.604978148 |
| XCL1 | Total_GRS | 4159 | 0.019668145 | [-0.01 0.05] | 0.205400617 | 0.708632128 |
| BOC | Cholesterol_pPS | 4159 | -0.000524395 | [-0.03 0.03] | 0.973069036 | 0.996862516 |
| BOC | Lipodystrophy_1_pPS | 4159 | 0.010140755 | [-0.02 0.04] | 0.513848974 | 0.870853148 |
| BOC | Lipodystrophy_2_pPS | 4159 | 0.024829831 | [-0.01 0.06] | 0.109879247 | 0.603827339 |
| BOC | Liver-Lipid_pPS | 4159 | 0.032904809 | [0. 0.06] | 0.034097845 | 0.382561193 |
| BOC | ASAT_Pos_pPS | 4159 | -0.020956753 | [-0.05 0.01] | 0.177241386 | 0.678995855 |
| BOC | ALP_Neg_pPS | 4159 | 0.010692472 | [-0.02 0.04] | 0.491215166 | 0.859085266 |
| BOC | SHBG-LpA_pPS | 4159 | 0.012952861 | [-0.02 0.04] | 0.404330805 | 0.81230319 |
| BOC | Obesity_pPS | 4159 | -0.01109482 | [-0.04 0.02] | 0.475052829 | 0.84751951 |
| BOC | VAT_Neg_pPS | 4159 | 0.017984354 | [-0.01 0.05] | 0.246910317 | 0.736582988 |
| BOC | Bilirubin_pPS | 4159 | -0.014333612 | [-0.04 0.02] | 0.356104311 | 0.7979754 |
| BOC | Proinsulin_pPS | 4159 | -0.008489236 | [-0.04 0.02] | 0.58470299 | 0.903925569 |
| BOC | Hyper_Insulin_pPS | 4159 | 0.003784114 | [-0.03 0.03] | 0.807529952 | 0.960682184 |
| BOC | Beta_Cell_1_pPS | 4159 | -0.022191071 | [-0.05 0.01] | 0.153065148 | 0.649125224 |
| BOC | Beta_Cell_2_pPS | 4159 | 0.018670272 | [-0.01 0.05] | 0.229341981 | 0.726631332 |
| BOC | Total_GRS | 4159 | 0.023963007 | [-0.01 0.05] | 0.122853341 | 0.615837655 |
| CNTN1 | Cholesterol_pPS | 4225 | -0.007818199 | [-0.04 0.02] | 0.61193179 | 0.909483337 |
| CNTN1 | Lipodystrophy_1_pPS | 4225 | 0.022022261 | [-0.01 0.05] | 0.152957733 | 0.649125224 |
| CNTN1 | Lipodystrophy_2_pPS | 4225 | 0.014507302 | [-0.02 0.04] | 0.346497535 | 0.790391668 |
| CNTN1 | Liver-Lipid_pPS | 4225 | 0.006327438 | [-0.02 0.04] | 0.681379652 | 0.925951669 |
| CNTN1 | ASAT_Pos_pPS | 4225 | 0.021987415 | [-0.01 0.05] | 0.153608743 | 0.649249818 |
| CNTN1 | ALP_Neg_pPS | 4225 | 0.011971502 | [-0.02 0.04] | 0.43725376 | 0.829429813 |
| CNTN1 | SHBG-LpA_pPS | 4225 | 0.015211077 | [-0.01 0.05] | 0.323603904 | 0.773957344 |
| CNTN1 | Obesity_pPS | 4225 | -0.012136604 | [-0.04 0.02] | 0.430957988 | 0.827992564 |
| CNTN1 | VAT_Neg_pPS | 4225 | 0.024321814 | [-0.01 0.05] | 0.114465427 | 0.607547265 |
| CNTN1 | Bilirubin_pPS | 4225 | -0.025528969 | [-0.06 0. ] | 0.097559294 | 0.572901389 |
| CNTN1 | Proinsulin_pPS | 4225 | 0.000421811 | [-0.03 0.03] | 0.978164141 | 0.996862516 |
| CNTN1 | Hyper_Insulin_pPS | 4225 | 0.009511859 | [-0.02 0.04] | 0.537087661 | 0.881309122 |
| CNTN1 | Beta_Cell_1_pPS | 4225 | -0.001100585 | [-0.03 0.03] | 0.943067323 | 0.992903797 |
| CNTN1 | Beta_Cell_2_pPS | 4225 | 0.032358943 | [0. 0.06] | 0.03570519 | 0.386599178 |
| CNTN1 | Total_GRS | 4225 | 0.016732837 | [-0.01 0.05] | 0.277548702 | 0.747422497 |
| CPB1 | Cholesterol_pPS | 4225 | -0.014511056 | [-0.04 0.02] | 0.346372729 | 0.790391668 |
| CPB1 | Lipodystrophy_1_pPS | 4225 | 0.04323617 | [0.01 0.07] | 0.005003056 | 0.140763349 |
| CPB1 | Lipodystrophy_2_pPS | 4225 | 0.018498228 | [-0.01 0.05] | 0.229975288 | 0.726631332 |
| CPB1 | Liver-Lipid_pPS | 4225 | -0.046819038 | [-0.08 -0.02] | 0.002368314 | 0.089966861 |
| CPB1 | ASAT_Pos_pPS | 4225 | -0.011247729 | [-0.04 0.02] | 0.465469753 | 0.841554994 |
| CPB1 | ALP_Neg_pPS | 4225 | -0.072661826 | [-0.1 -0.04] | 2.35E-06 | 0.000376411 |
| CPB1 | SHBG-LpA_pPS | 4225 | -0.032474992 | [-0.06 -0. ] | 0.035047854 | 0.386023413 |
| CPB1 | Obesity_pPS | 4225 | -0.002876644 | [-0.03 0.03] | 0.851926362 | 0.97528582 |
| CPB1 | VAT_Neg_pPS | 4225 | -0.007696112 | [-0.04 0.02] | 0.617500775 | 0.909483337 |
| CPB1 | Bilirubin_pPS | 4225 | 0.003978933 | [-0.03 0.03] | 0.796262211 | 0.958074488 |
| CPB1 | Proinsulin_pPS | 4225 | -0.010656994 | [-0.04 0.02] | 0.489229659 | 0.858689586 |
| CPB1 | Hyper_Insulin_pPS | 4225 | -0.00319284 | [-0.03 0.03] | 0.835870303 | 0.970026164 |
| CPB1 | Beta_Cell_1_pPS | 4225 | 0.00270983 | [-0.03 0.03] | 0.860422209 | 0.977006629 |
| CPB1 | Beta_Cell_2_pPS | 4225 | -0.001149884 | [-0.03 0.03] | 0.940521719 | 0.992673019 |
| CPB1 | Total_GRS | 4225 | -0.005279787 | [-0.04 0.02] | 0.731900037 | 0.942134248 |
| DCN | Cholesterol_pPS | 4159 | 0.010179265 | [-0.02 0.04] | 0.512251736 | 0.870690038 |
| DCN | Lipodystrophy_1_pPS | 4159 | -0.016509311 | [-0.05 0.01] | 0.287824849 | 0.751557788 |
| DCN | Lipodystrophy_2_pPS | 4159 | 0.009934076 | [-0.02 0.04] | 0.522465439 | 0.872980599 |
| DCN | Liver-Lipid_pPS | 4159 | 0.043388851 | [0.01 0.07] | 0.005196487 | 0.14342303 |
| DCN | ASAT_Pos_pPS | 4159 | -0.020615784 | [-0.05 0.01] | 0.184396334 | 0.686251968 |
| DCN | ALP_Neg_pPS | 4159 | -0.023957352 | [-0.05 0.01] | 0.122941755 | 0.615837655 |
| DCN | SHBG-LpA_pPS | 4159 | 0.02030962 | [-0.01 0.05] | 0.191001103 | 0.693468715 |
| DCN | Obesity_pPS | 4159 | 0.01969885 | [-0.01 0.05] | 0.204693858 | 0.70858232 |
| DCN | VAT_Neg_pPS | 4159 | 0.022647705 | [-0.01 0.05] | 0.144786464 | 0.641415816 |
| DCN | Bilirubin_pPS | 4159 | -0.012625659 | [-0.04 0.02] | 0.416307249 | 0.817860704 |
| DCN | Proinsulin_pPS | 4159 | 0.012124547 | [-0.02 0.04] | 0.435049395 | 0.828998134 |
| DCN | Hyper_Insulin_pPS | 4159 | 0.017976038 | [-0.01 0.05] | 0.24712893 | 0.736582988 |
| DCN | Beta_Cell_1_pPS | 4159 | -0.013231828 | [-0.04 0.02] | 0.394284487 | 0.812191725 |
| DCN | Beta_Cell_2_pPS | 4159 | 0.000858685 | [-0.03 0.03] | 0.955915212 | 0.99612031 |
| DCN | Total_GRS | 4159 | 0.026563379 | [-0. 0.06] | 0.087193134 | 0.542011375 |
| Dkk_1 | Cholesterol_pPS | 4159 | -0.008737933 | [-0.04 0.02] | 0.57374845 | 0.903411176 |
| Dkk_1 | Lipodystrophy_1_pPS | 4159 | -0.000994043 | [-0.03 0.03] | 0.948974778 | 0.993154165 |
| Dkk_1 | Lipodystrophy_2_pPS | 4159 | 0.01082487 | [-0.02 0.04] | 0.485864358 | 0.854965648 |
| Dkk_1 | Liver-Lipid_pPS | 4159 | -0.002949898 | [-0.03 0.03] | 0.849380972 | 0.97528582 |
| Dkk_1 | ASAT_Pos_pPS | 4159 | 0.002538604 | [-0.03 0.03] | 0.870179898 | 0.977006629 |
| Dkk_1 | ALP_Neg_pPS | 4159 | 0.003666812 | [-0.03 0.03] | 0.813384512 | 0.962515303 |
| Dkk_1 | SHBG-LpA_pPS | 4159 | -0.015100964 | [-0.05 0.02] | 0.330940043 | 0.777357036 |
| Dkk_1 | Obesity_pPS | 4159 | -0.025202873 | [-0.06 0.01] | 0.104640069 | 0.593032011 |
| Dkk_1 | VAT_Neg_pPS | 4159 | -0.026797686 | [-0.06 0. ] | 0.084439981 | 0.535757118 |
| Dkk_1 | Bilirubin_pPS | 4159 | 0.008927999 | [-0.02 0.04] | 0.565442603 | 0.899863296 |
| Dkk_1 | Proinsulin_pPS | 4159 | 0.006503259 | [-0.02 0.04] | 0.675457404 | 0.925361765 |
| Dkk_1 | Hyper_Insulin_pPS | 4159 | -0.030654316 | [-0.06 -0. ] | 0.048390724 | 0.438114317 |
| Dkk_1 | Beta_Cell_1_pPS | 4159 | -0.015644109 | [-0.05 0.01] | 0.313841164 | 0.76365449 |
| Dkk_1 | Beta_Cell_2_pPS | 4159 | -0.002606339 | [-0.03 0.03] | 0.866747991 | 0.977006629 |
| Dkk_1 | Total_GRS | 4159 | -0.019821895 | [-0.05 0.01] | 0.201879455 | 0.703519315 |
| IgG_Fc_receptor_II_b | Cholesterol_pPS | 4159 | -0.014187236 | [-0.04 0.02] | 0.361038064 | 0.7979754 |
| IgG_Fc_receptor_II_b | Lipodystrophy_1_pPS | 4159 | 0.002284859 | [-0.03 0.03] | 0.883057548 | 0.979291846 |

**b) Partial Correlation**

| Proteomics | PPS | Sample_Size | r | CI95% | p | fdr_p_adjusted |
|---|---|---|---|---|---|---|
| IgG_Fc_receptor_II_b | Bilirubin_pPS | 4159 | -0.004240813 | [-0.03 0.03] | 0.785170782 | 0.966801939 |
| IgG_Fc_receptor_II_b | Proinsulin_pPS | 4159 | 0.014919594 | [-0.02 0.05] | 0.337540456 | 0.844649909 |
| IgG_Fc_receptor_II_b | Hyper_Insulin_pPS | 4159 | 0.008982002 | [-0.02 0.04] | 0.563706098 | 0.904956448 |
| IgG_Fc_receptor_II_b | Beta_Cell_1_pPS | 4159 | -0.007377522 | [-0.04 0.02] | 0.635350464 | 0.931509843 |
| IgG_Fc_receptor_II_b | Beta_Cell_2_pPS | 4159 | -0.025680369 | [-0.06 0. ] | 0.098754342 | 0.648957106 |
| FGF_21 | Cholesterol_pPS | 4159 | 0.016800316 | [-0.01 0.05] | 0.2801663 | 0.808820203 |
| FGF_21 | Lipodystrophy_1_pPS | 4159 | 0.046495869 | [0.02 0.08] | 0.002787763 | 0.119687959 |
| FGF_21 | Lipodystrophy_2_pPS | 4159 | -0.008995227 | [-0.04 0.02] | 0.563132046 | 0.904956448 |
| FGF_21 | Liver-Lipid_pPS | 4159 | -0.083630018 | [-0.11 -0.05] | 7.25E-08 | 1.33E-05 |
| FGF_21 | ASAT_Pos_pPS | 4159 | 0.023430418 | [-0.01 0.05] | 0.132006139 | 0.694065456 |
| FGF_21 | ALP_Neg_pPS | 4159 | 0.001534947 | [-0.03 0.03] | 0.921407096 | 0.990503296 |
| FGF_21 | SHBG-LpA_pPS | 4159 | 0.013147023 | [-0.02 0.04] | 0.398063724 | 0.85037326 |
| FGF_21 | Obesity_pPS | 4159 | 0.013864019 | [-0.02 0.04] | 0.372834295 | 0.847284666 |
| FGF_21 | VAT_Neg_pPS | 4159 | 0.04502156 | [0.01 0.08] | 0.003787874 | 0.141413949 |
| FGF_21 | Bilirubin_pPS | 4159 | -0.00869027 | [-0.04 0.02] | 0.57643963 | 0.909113996 |
| FGF_21 | Proinsulin_pPS | 4159 | -0.012787703 | [-0.04 0.02] | 0.411085141 | 0.854426619 |
| FGF_21 | Hyper_Insulin_pPS | 4159 | 0.018252115 | [-0.01 0.05] | 0.240681813 | 0.790085251 |
| FGF_21 | Beta_Cell_1_pPS | 4159 | 0.022461913 | [-0.01 0.05] | 0.148749997 | 0.703126624 |
| FGF_21 | Beta_Cell_2_pPS | 4159 | -0.014088416 | [-0.04 0.02] | 0.365146286 | 0.847284666 |
| FGF_23 | Cholesterol_pPS | 4159 | 0.016434507 | [-0.01 0.05] | 0.290772721 | 0.81655641 |
| FGF_23 | Lipodystrophy_1_pPS | 4159 | -0.015133967 | [-0.05 0.02] | 0.330643834 | 0.844649909 |
| FGF_23 | Lipodystrophy_2_pPS | 4159 | 0.005478677 | [-0.03 0.04] | 0.724722668 | 0.946981479 |
| FGF_23 | Liver-Lipid_pPS | 4159 | -0.040178728 | [-0.07 -0.01] | 0.009777481 | 0.248683454 |
| FGF_23 | ASAT_Pos_pPS | 4159 | 0.034778282 | [0. 0.07] | 0.025344487 | 0.392967754 |
| FGF_23 | ALP_Neg_pPS | 4159 | 0.056561241 | [0.03 0.09] | 0.000274246 | 0.023548612 |
| FGF_23 | SHBG-LpA_pPS | 4159 | 0.003315968 | [-0.03 0.03] | 0.831217618 | 0.971458352 |
| FGF_23 | Obesity_pPS | 4159 | 0.028909415 | [-0. 0.06] | 0.063085059 | 0.558815902 |
| FGF_23 | VAT_Neg_pPS | 4159 | 0.015531439 | [-0.01 0.05] | 0.318099701 | 0.839009314 |
| FGF_23 | Bilirubin_pPS | 4159 | 0.014416599 | [-0.02 0.04] | 0.354082197 | 0.847284666 |
| FGF_23 | Proinsulin_pPS | 4159 | 0.007517669 | [-0.02 0.04] | 0.628940858 | 0.929162059 |
| FGF_23 | Hyper_Insulin_pPS | 4159 | -0.008745426 | [-0.04 0.02] | 0.574021833 | 0.908340339 |
| FGF_23 | Beta_Cell_1_pPS | 4159 | 0.015953071 | [-0.01 0.05] | 0.305138299 | 0.8267377 |
| FGF_23 | Beta_Cell_2_pPS | 4159 | -0.00102149 | [-0.03 0.03] | 0.947650076 | 0.996425456 |
| HSP_27 | Cholesterol_pPS | 4159 | -0.005121998 | [-0.04 0.03] | 0.741983717 | 0.951867558 |
| HSP_27 | Lipodystrophy_1_pPS | 4159 | -0.012220081 | [-0.04 0.02] | 0.432162609 | 0.864060889 |
| HSP_27 | Lipodystrophy_2_pPS | 4159 | 0.017781053 | [-0.01 0.05] | 0.25303919 | 0.799790128 |
| HSP_27 | Liver-Lipid_pPS | 4159 | 0.001942825 | [-0.03 0.03] | 0.900619688 | 0.985399384 |
| HSP_27 | ASAT_Pos_pPS | 4159 | -0.023385523 | [-0.05 0.01] | 0.13274858 | 0.694065456 |
| HSP_27 | ALP_Neg_pPS | 4159 | 0.010771223 | [-0.02 0.04] | 0.488711769 | 0.895803896 |
| HSP_27 | SHBG-LpA_pPS | 4159 | 0.024739552 | [-0.01 0.06] | 0.111739877 | 0.680721977 |
| HSP_27 | Obesity_pPS | 4159 | -0.019252829 | [-0.05 0.01] | 0.215855544 | 0.768032367 |
| HSP_27 | VAT_Neg_pPS | 4159 | 0.001476178 | [-0.03 0.03] | 0.92440701 | 0.990503296 |
| HSP_27 | Bilirubin_pPS | 4159 | -0.00192528 | [-0.03 0.03] | 0.901512539 | 0.985399384 |
| HSP_27 | Proinsulin_pPS | 4159 | 0.005722307 | [-0.02 0.04] | 0.713011707 | 0.945970482 |
| HSP_27 | Hyper_Insulin_pPS | 4159 | 0.007058622 | [-0.02 0.04] | 0.65003692 | 0.935848959 |
| HSP_27 | Beta_Cell_1_pPS | 4159 | 0.019588559 | [-0.01 0.05] | 0.207954614 | 0.767305935 |
| HSP_27 | Beta_Cell_2_pPS | 4159 | -0.030964027 | [-0.06 -0. ] | 0.046509016 | 0.518645994 |
| IGFBP_2 | Cholesterol_pPS | 4225 | 0.009222483 | [-0.02 0.04] | 0.550160248 | 0.899241939 |
| IGFBP_2 | Lipodystrophy_1_pPS | 4225 | -0.032570594 | [-0.06 -0. ] | 0.034794028 | 0.443820658 |
| IGFBP_2 | Lipodystrophy_2_pPS | 4225 | -0.027950349 | [-0.06 0. ] | 0.070109238 | 0.573866874 |
| IGFBP_2 | Liver-Lipid_pPS | 4225 | 0.017690135 | [-0.01 0.05] | 0.251712315 | 0.799790128 |
| IGFBP_2 | ASAT_Pos_pPS | 4225 | -0.024010974 | [-0.05 0.01] | 0.119743742 | 0.681046037 |
| IGFBP_2 | ALP_Neg_pPS | 4225 | -0.008498326 | [-0.04 0.02] | 0.581907573 | 0.909583681 |
| IGFBP_2 | SHBG-LpA_pPS | 4225 | -0.023089382 | [-0.05 0.01] | 0.134623159 | 0.694065456 |
| IGFBP_2 | Obesity_pPS | 4225 | -0.04773923 | [-0.08 -0.02] | 0.001970126 | 0.099117562 |
| IGFBP_2 | VAT_Neg_pPS | 4225 | -0.045306865 | [-0.08 -0.02] | 0.003315596 | 0.133452752 |
| IGFBP_2 | Bilirubin_pPS | 4225 | 0.013041782 | [-0.02 0.04] | 0.398117295 | 0.85037326 |
| IGFBP_2 | Proinsulin_pPS | 4225 | 0.028694613 | [-0. 0.06] | 0.062962988 | 0.558815902 |
| IGFBP_2 | Hyper_Insulin_pPS | 4225 | -0.018406111 | [-0.05 0.01] | 0.23302673 | 0.786982264 |
| IGFBP_2 | Beta_Cell_1_pPS | 4225 | -0.018781623 | [-0.05 0.01] | 0.223629603 | 0.780100778 |
| IGFBP_2 | Beta_Cell_2_pPS | 4225 | 0.015096978 | [-0.02 0.05] | 0.327994952 | 0.84456682 |
| ITGB1BP2 | Cholesterol_pPS | 4159 | -0.000680047 | [-0.03 0.03] | 0.965134581 | 0.996924797 |
| ITGB1BP2 | Lipodystrophy_1_pPS | 4159 | -0.010681236 | [-0.04 0.02] | 0.49235079 | 0.895803896 |
| ITGB1BP2 | Lipodystrophy_2_pPS | 4159 | 0.016413262 | [-0.01 0.05] | 0.291396914 | 0.81655641 |
| ITGB1BP2 | Liver-Lipid_pPS | 4159 | -0.028532239 | [-0.06 0. ] | 0.066604593 | 0.563434057 |
| ITGB1BP2 | ASAT_Pos_pPS | 4159 | 0.014214381 | [-0.02 0.04] | 0.360874338 | 0.847284666 |
| ITGB1BP2 | ALP_Neg_pPS | 4159 | -0.016717129 | [-0.05 0.01] | 0.282554875 | 0.81132836 |
| ITGB1BP2 | SHBG-LpA_pPS | 4159 | 0.007811891 | [-0.02 0.04] | 0.615575658 | 0.921525431 |
| ITGB1BP2 | Obesity_pPS | 4159 | -0.017469998 | [-0.05 0.01] | 0.261437362 | 0.807171938 |
| ITGB1BP2 | VAT_Neg_pPS | 4159 | 0.010067717 | [-0.02 0.04] | 0.517543612 | 0.895803896 |
| ITGB1BP2 | Bilirubin_pPS | 4159 | 0.006087575 | [-0.02 0.04] | 0.695580553 | 0.945970482 |
| ITGB1BP2 | Proinsulin_pPS | 4159 | 0.032242363 | [0. 0.06] | 0.038174783 | 0.463859628 |
| ITGB1BP2 | Hyper_Insulin_pPS | 4159 | 0.002949066 | [-0.03 0.03] | 0.849656438 | 0.974254359 |
| ITGB1BP2 | Beta_Cell_1_pPS | 4159 | -0.010504462 | [-0.04 0.02] | 0.499541546 | 0.895803896 |
| ITGB1BP2 | Beta_Cell_2_pPS | 4159 | 0.001358391 | [-0.03 0.03] | 0.930422752 | 0.992040153 |
| SCF | Cholesterol_pPS | 4159 | -0.006576468 | [-0.04 0.02] | 0.672500634 | 0.939458586 |
| SCF | Lipodystrophy_1_pPS | 4159 | -0.013387048 | [-0.04 0.02] | 0.389505531 | 0.85037326 |
| SCF | Lipodystrophy_2_pPS | 4159 | -0.00121451 | [-0.03 0.03] | 0.93777653 | 0.994163908 |
| SCF | Liver-Lipid_pPS | 4159 | -0.030342487 | [-0.06 0. ] | 0.051084952 | 0.537121778 |
| SCF | ASAT_Pos_pPS | 4159 | -0.024085233 | [-0.05 0.01] | 0.121539266 | 0.682614177 |
| SCF | ALP_Neg_pPS | 4159 | -0.007031557 | [-0.04 0.02] | 0.651289722 | 0.936605382 |
| SCF | SHBG-LpA_pPS | 4159 | 0.008851274 | [-0.02 0.04] | 0.569395351 | 0.905732577 |
| SCF | Obesity_pPS | 4159 | -0.031681462 | [-0.06 -0. ] | 0.041660811 | 0.481247748 |
| SCF | VAT_Neg_pPS | 4159 | -0.004932297 | [-0.04 0.03] | 0.751217866 | 0.954135344 |
| SCF | Bilirubin_pPS | 4159 | 0.005178438 | [-0.03 0.04] | 0.739243459 | 0.95166974 |
| SCF | Proinsulin_pPS | 4159 | -0.013176589 | [-0.04 0.02] | 0.397003475 | 0.85037326 |
| SCF | Hyper_Insulin_pPS | 4159 | -0.005701763 | [-0.04 0.02] | 0.713996671 | 0.945970482 |
| SCF | Beta_Cell_1_pPS | 4159 | 0.006081137 | [-0.02 0.04] | 0.695886458 | 0.945970482 |
| SCF | Beta_Cell_2_pPS | 4159 | 0.019412989 | [-0.01 0.05] | 0.21205986 | 0.767305935 |
| Notch_3 | Cholesterol_pPS | 4225 | 0.014706611 | [-0.02 0.04] | 0.34065839 | 0.847042484 |
| Notch_3 | Lipodystrophy_1_pPS | 4225 | -0.013103392 | [-0.04 0.02] | 0.395892143 | 0.85037326 |
| Notch_3 | Lipodystrophy_2_pPS | 4225 | 0.001534366 | [-0.03 0.03] | 0.920813746 | 0.990503296 |
| Notch_3 | Liver-Lipid_pPS | 4225 | 0.032457972 | [0. 0.06] | 0.035426882 | 0.449078407 |
| Notch_3 | ASAT_Pos_pPS | 4225 | 0.004790244 | [-0.03 0.04] | 0.756292764 | 0.95462436 |
| Notch_3 | ALP_Neg_pPS | 4225 | -0.038481903 | [-0.07 -0.01] | 0.012627917 | 0.293058678 |
| Notch_3 | SHBG-LpA_pPS | 4225 | 0.004822758 | [-0.03 0.03] | 0.754691556 | 0.95462436 |
| Notch_3 | Obesity_pPS | 4225 | 0.005864906 | [-0.02 0.04] | 0.703960638 | 0.945970482 |
| Notch_3 | VAT_Neg_pPS | 4225 | 0.006800893 | [-0.02 0.04] | 0.659486572 | 0.937489141 |
| Notch_3 | Bilirubin_pPS | 4225 | -0.014361183 | [-0.04 0.02] | 0.352121594 | 0.847284666 |
| Notch_3 | Proinsulin_pPS | 4225 | 0.021580673 | [-0.01 0.05] | 0.16201279 | 0.718997225 |
| Notch_3 | Hyper_Insulin_pPS | 4225 | 0.027802458 | [-0. 0.06] | 0.071605668 | 0.578232608 |
| Notch_3 | Beta_Cell_1_pPS | 4225 | -0.014897605 | [-0.05 0.02] | 0.33442396 | 0.844649909 |
| Notch_3 | Beta_Cell_2_pPS | 4225 | 0.027569529 | [-0. 0.06] | 0.074015522 | 0.588265012 |
| PTX3 | Cholesterol_pPS | 4159 | 0.006840941 | [-0.02 0.04] | 0.660140934 | 0.937489141 |
| PTX3 | Lipodystrophy_1_pPS | 4159 | -0.023305658 | [-0.05 0.01] | 0.134077314 | 0.694065456 |
| PTX3 | Lipodystrophy_2_pPS | 4159 | 0.005989831 | [-0.02 0.04] | 0.700229783 | 0.945970482 |
| PTX3 | Liver-Lipid_pPS | 4159 | -0.009473358 | [-0.04 0.02] | 0.542570641 | 0.899241939 |

**Table S7: Associations between pPSs and plasma proteins in SCAPIS**

**a) Spearman Correlation**

| Proteomics | PPS | Sample_Size | r | CI95% | p | fdr_p_adjusted |
|---|---|---|---|---|---|---|
| IgG_Fc_receptor_II_b | Lipodystrophy_2_pPS | 4159 | 0.003739779 | [-0.03 0.03] | 0.809741489 | 0.961655124 |
| IgG_Fc_receptor_II_b | Liver-Lipid_pPS | 4159 | 0.002241383 | [-0.03 0.03] | 0.885267161 | 0.979291846 |
| IgG_Fc_receptor_II_b | ASAT_Pos_pPS | 4159 | 0.002452566 | [-0.03 0.03] | 0.874542675 | 0.978250438 |
| IgG_Fc_receptor_II_b | ALP_Neg_pPS | 4159 | 0.017848362 | [-0.01 0.05] | 0.250502515 | 0.738661262 |
| IgG_Fc_receptor_II_b | SHBG-LpA_pPS | 4159 | 0.005218936 | [-0.03 0.04] | 0.736881552 | 0.942470055 |
| IgG_Fc_receptor_II_b | Obesity_pPS | 4159 | 0.007032758 | [-0.02 0.04] | 0.650722272 | 0.919948884 |
| IgG_Fc_receptor_II_b | VAT_Neg_pPS | 4159 | 0.014308981 | [-0.02 0.04] | 0.356931539 | 0.7979754 |
| IgG_Fc_receptor_II_b | Bilirubin_pPS | 4159 | -0.004506746 | [-0.03 0.03] | 0.77171233 | 0.95301975 |
| IgG_Fc_receptor_II_b | Proinsulin_pPS | 4159 | 0.012765785 | [-0.02 0.04] | 0.411152878 | 0.816389887 |
| IgG_Fc_receptor_II_b | Hyper_Insulin_pPS | 4159 | 0.010685787 | [-0.02 0.04] | 0.491486171 | 0.859085266 |
| IgG_Fc_receptor_II_b | Beta_Cell_1_pPS | 4159 | -0.008116159 | [-0.04 0.02] | 0.60131598 | 0.907623171 |
| IgG_Fc_receptor_II_b | Beta_Cell_2_pPS | 4159 | -0.024355866 | [-0.05 0.01] | 0.116832249 | 0.611872879 |
| IgG_Fc_receptor_II_b | Total_GRS | 4159 | 0.016665689 | [-0.01 0.05] | 0.283282444 | 0.749111537 |
| FGF_21 | Cholesterol_pPS | 4159 | 0.013034505 | [-0.02 0.04] | 0.401374872 | 0.81230319 |
| FGF_21 | Lipodystrophy_1_pPS | 4159 | 0.056616081 | [0.03 0.09] | 0.000264553 | 0.019630339 |
| FGF_21 | Lipodystrophy_2_pPS | 4159 | 0.000414619 | [-0.03 0.03] | 0.978705189 | 0.996862516 |
| FGF_21 | Liver-Lipid_pPS | 4159 | -0.080776179 | [-0.11 -0.05] | 1.90E-07 | 3.28E-05 |
| FGF_21 | ASAT_Pos_pPS | 4159 | 0.033621118 | [0. 0.06] | 0.030382681 | 0.369452038 |
| FGF_21 | ALP_Neg_pPS | 4159 | -0.002437617 | [-0.03 0.03] | 0.875301099 | 0.978250438 |
| FGF_21 | SHBG-LpA_pPS | 4159 | 0.018569023 | [-0.01 0.05] | 0.231877777 | 0.726631332 |
| FGF_21 | Obesity_pPS | 4159 | 0.016536166 | [-0.01 0.05] | 0.287041293 | 0.751557788 |
| FGF_21 | VAT_Neg_pPS | 4159 | 0.0482083 | [0.02 0.08] | 0.001900544 | 0.078291058 |
| FGF_21 | Bilirubin_pPS | 4159 | -0.005109647 | [-0.04 0.03] | 0.742193518 | 0.945527195 |
| FGF_21 | Proinsulin_pPS | 4159 | -0.006954449 | [-0.04 0.02] | 0.654357114 | 0.920338805 |
| FGF_21 | Hyper_Insulin_pPS | 4159 | 0.027120148 | [-0. 0.06] | 0.080766404 | 0.5244628 |
| FGF_21 | Beta_Cell_1_pPS | 4159 | 0.021128947 | [-0.01 0.05] | 0.173707551 | 0.674627844 |
| FGF_21 | Beta_Cell_2_pPS | 4159 | -0.009559606 | [-0.04 0.02] | 0.538263935 | 0.88195499 |
| FGF_21 | Total_GRS | 4159 | 0.028458769 | [-0. 0.06] | 0.066879367 | 0.477352596 |
| FGF_23 | Cholesterol_pPS | 4159 | 0.017037751 | [-0.01 0.05] | 0.272670956 | 0.746520328 |
| FGF_23 | Lipodystrophy_1_pPS | 4159 | -0.006325342 | [-0.04 0.02] | 0.683849617 | 0.926969309 |
| FGF_23 | Lipodystrophy_2_pPS | 4159 | 0.008508187 | [-0.02 0.04] | 0.583864799 | 0.903823705 |
| FGF_23 | Liver-Lipid_pPS | 4159 | -0.036369942 | [-0.07 -0.01] | 0.019170881 | 0.29559571 |
| FGF_23 | ASAT_Pos_pPS | 4159 | 0.03404514 | [0. 0.06] | 0.028351617 | 0.358947074 |
| FGF_23 | ALP_Neg_pPS | 4159 | 0.057466628 | [0.03 0.09] | 0.000213372 | 0.017829955 |
| FGF_23 | SHBG-LpA_pPS | 4159 | 0.001292175 | [-0.03 0.03] | 0.933702545 | 0.990399318 |
| FGF_23 | Obesity_pPS | 4159 | 0.032870952 | [0. 0.06] | 0.034282663 | 0.383006082 |
| FGF_23 | VAT_Neg_pPS | 4159 | 0.016985949 | [-0.01 0.05] | 0.27413186 | 0.746520328 |
| FGF_23 | Bilirubin_pPS | 4159 | 0.016732425 | [-0.01 0.05] | 0.281358776 | 0.748127084 |
| FGF_23 | Proinsulin_pPS | 4159 | 0.026507596 | [-0. 0.06] | 0.087859161 | 0.544206768 |
| FGF_23 | Hyper_Insulin_pPS | 4159 | -0.004355952 | [-0.03 0.03] | 0.779149237 | 0.953637204 |
| FGF_23 | Beta_Cell_1_pPS | 4159 | 0.021675074 | [-0.01 0.05] | 0.162847832 | 0.664061293 |
| FGF_23 | Beta_Cell_2_pPS | 4159 | 0.0060557 | [-0.02 0.04] | 0.696642922 | 0.930772879 |
| FGF_23 | Total_GRS | 4159 | 0.041135371 | [0.01 0.07] | 0.008065258 | 0.197592038 |
| HSP_27 | Cholesterol_pPS | 4159 | -0.006794057 | [-0.04 0.02] | 0.661827487 | 0.924111434 |
| HSP_27 | Lipodystrophy_1_pPS | 4159 | -0.012674234 | [-0.04 0.02] | 0.414516171 | 0.817764515 |
| HSP_27 | Lipodystrophy_2_pPS | 4159 | 0.014784481 | [-0.02 0.05] | 0.341175978 | 0.787547587 |
| HSP_27 | Liver-Lipid_pPS | 4159 | 0.002333332 | [-0.03 0.03] | 0.880595007 | 0.979035738 |
| HSP_27 | ASAT_Pos_pPS | 4159 | -0.023227729 | [-0.05 0.01] | 0.134770537 | 0.629316669 |
| HSP_27 | ALP_Neg_pPS | 4159 | 0.010737182 | [-0.02 0.04] | 0.489404672 | 0.858689586 |
| HSP_27 | SHBG-LpA_pPS | 4159 | 0.024107513 | [-0.01 0.05] | 0.120611027 | 0.615837655 |
| HSP_27 | Obesity_pPS | 4159 | -0.020300229 | [-0.05 0.01] | 0.19120641 | 0.693468715 |
| HSP_27 | VAT_Neg_pPS | 4159 | 0.005408007 | [-0.03 0.04] | 0.727721476 | 0.942134248 |
| HSP_27 | Bilirubin_pPS | 4159 | -0.002767419 | [-0.03 0.03] | 0.858596818 | 0.977006629 |
| HSP_27 | Proinsulin_pPS | 4159 | 0.00708595 | [-0.02 0.04] | 0.648257981 | 0.9184764 |
| HSP_27 | Hyper_Insulin_pPS | 4159 | 0.004032216 | [-0.03 0.03] | 0.795182985 | 0.958074488 |
| HSP_27 | Beta_Cell_1_pPS | 4159 | 0.0130243 | [-0.02 0.04] | 0.401743629 | 0.81230319 |
| HSP_27 | Beta_Cell_2_pPS | 4159 | -0.024516538 | [-0.05 0.01] | 0.114437514 | 0.607547265 |
| HSP_27 | Total_GRS | 4159 | 0.002661903 | [-0.03 0.03] | 0.863934605 | 0.977006629 |
| IGFBP_2 | Cholesterol_pPS | 4225 | 0.00677365 | [-0.02 0.04] | 0.660273123 | 0.924111434 |
| IGFBP_2 | Lipodystrophy_1_pPS | 4225 | -0.04669463 | [-0.08 -0.02] | 0.002432679 | 0.090660952 |
| IGFBP_2 | Lipodystrophy_2_pPS | 4225 | -0.038107693 | [-0.07 -0.01] | 0.013374274 | 0.242414912 |
| IGFBP_2 | Liver-Lipid_pPS | 4225 | 0.01459538 | [-0.02 0.04] | 0.34357721 | 0.790391668 |
| IGFBP_2 | ASAT_Pos_pPS | 4225 | -0.037266357 | [-0.07 -0.01] | 0.015563531 | 0.258767146 |
| IGFBP_2 | ALP_Neg_pPS | 4225 | -0.004279122 | [-0.03 0.03] | 0.781268365 | 0.955139045 |
| IGFBP_2 | SHBG-LpA_pPS | 4225 | -0.027631672 | [-0.06 0. ] | 0.072922337 | 0.502472104 |
| IGFBP_2 | Obesity_pPS | 4225 | -0.046797084 | [-0.08 -0.02] | 0.002379558 | 0.089966861 |
| IGFBP_2 | VAT_Neg_pPS | 4225 | -0.054679868 | [-0.08 -0.02] | 0.000384144 | 0.025859464 |
| IGFBP_2 | Bilirubin_pPS | 4225 | 0.010736599 | [-0.02 0.04] | 0.485990293 | 0.854965648 |
| IGFBP_2 | Proinsulin_pPS | 4225 | 0.01795047 | [-0.01 0.05] | 0.244071415 | 0.732214246 |
| IGFBP_2 | Hyper_Insulin_pPS | 4225 | -0.033253094 | [-0.06 -0. ] | 0.030901735 | 0.369452038 |
| IGFBP_2 | Beta_Cell_1_pPS | 4225 | -0.014139289 | [-0.04 0.02] | 0.358869572 | 0.7979754 |
| IGFBP_2 | Beta_Cell_2_pPS | 4225 | 0.011040875 | [-0.02 0.04] | 0.473715897 | 0.846453261 |
| IGFBP_2 | Total_GRS | 4225 | -0.05608413 | [-0.09 -0.03] | 0.00027045 | 0.019630339 |
| ITGB1BP2 | Cholesterol_pPS | 4159 | -0.002649083 | [-0.03 0.03] | 0.864583588 | 0.977006629 |
| ITGB1BP2 | Lipodystrophy_1_pPS | 4159 | -0.007222745 | [-0.04 0.02] | 0.641938283 | 0.916400472 |
| ITGB1BP2 | Lipodystrophy_2_pPS | 4159 | 0.018561846 | [-0.01 0.05] | 0.232058272 | 0.726631332 |
| ITGB1BP2 | Liver-Lipid_pPS | 4159 | -0.028025495 | [-0.06 0. ] | 0.071141386 | 0.495833904 |
| ITGB1BP2 | ASAT_Pos_pPS | 4159 | 0.013583483 | [-0.02 0.04] | 0.381837874 | 0.806027092 |
| ITGB1BP2 | ALP_Neg_pPS | 4159 | -0.01280358 | [-0.04 0.02] | 0.409769131 | 0.814814699 |
| ITGB1BP2 | SHBG-LpA_pPS | 4159 | 0.009947719 | [-0.02 0.04] | 0.521894362 | 0.872980599 |
| ITGB1BP2 | Obesity_pPS | 4159 | -0.014085896 | [-0.04 0.02] | 0.36447878 | 0.798887077 |
| ITGB1BP2 | VAT_Neg_pPS | 4159 | 0.012317296 | [-0.02 0.04] | 0.427783439 | 0.82738773 |
| ITGB1BP2 | Bilirubin_pPS | 4159 | 0.006693367 | [-0.02 0.04] | 0.66653455 | 0.924640793 |
| ITGB1BP2 | Proinsulin_pPS | 4159 | 0.026825524 | [-0. 0.06] | 0.084117609 | 0.534941474 |
| ITGB1BP2 | Hyper_Insulin_pPS | 4159 | 0.002385506 | [-0.03 0.03] | 0.877945788 | 0.979031336 |
| ITGB1BP2 | Beta_Cell_1_pPS | 4159 | -0.00082051 | [-0.03 0.03] | 0.957873256 | 0.99612031 |
| ITGB1BP2 | Beta_Cell_2_pPS | 4159 | 0.004531244 | [-0.03 0.03] | 0.770506107 | 0.95301975 |
| ITGB1BP2 | Total_GRS | 4159 | 0.00495356 | [-0.03 0.04] | 0.749801446 | 0.948757065 |
| SCF | Cholesterol_pPS | 4159 | -0.010729835 | [-0.04 0.02] | 0.489701959 | 0.858689586 |
| SCF | Lipodystrophy_1_pPS | 4159 | -0.019903983 | [-0.05 0.01] | 0.200017616 | 0.700947446 |
| SCF | Lipodystrophy_2_pPS | 4159 | -0.006423946 | [-0.04 0.02] | 0.679193669 | 0.925579558 |
| SCF | Liver-Lipid_pPS | 4159 | -0.031678556 | [-0.06 -0. ] | 0.041359546 | 0.407686951 |
| SCF | ASAT_Pos_pPS | 4159 | -0.029155696 | [-0.06 0. ] | 0.060466247 | 0.463868622 |
| SCF | ALP_Neg_pPS | 4159 | -0.011498216 | [-0.04 0.02] | 0.459145752 | 0.837016034 |
| SCF | SHBG-LpA_pPS | 4159 | 0.007478483 | [-0.02 0.04] | 0.630193195 | 0.914460114 |
| SCF | Obesity_pPS | 4159 | -0.03646175 | [-0.07 -0.01] | 0.018869065 | 0.29257652 |
| SCF | VAT_Neg_pPS | 4159 | -0.004184416 | [-0.03 0.03] | 0.787633707 | 0.957247257 |
| SCF | Bilirubin_pPS | 4159 | 0.004602195 | [-0.03 0.04] | 0.767015702 | 0.952160028 |
| SCF | Proinsulin_pPS | 4159 | -0.014694073 | [-0.05 0.02] | 0.344136824 | 0.790391668 |
| SCF | Hyper_Insulin_pPS | 4159 | -0.013252853 | [-0.04 0.02] | 0.393533514 | 0.812191725 |
| SCF | Beta_Cell_1_pPS | 4159 | 0.004607671 | [-0.03 0.04] | 0.766746501 | 0.952160028 |
| SCF | Beta_Cell_2_pPS | 4159 | 0.017883516 | [-0.01 0.05] | 0.24957045 | 0.738661262 |
| SCF | Total_GRS | 4159 | -0.018769895 | [-0.05 0.01] | 0.226866209 | 0.724711501 |
| Notch_3 | Cholesterol_pPS | 4225 | 0.017073525 | [-0.01 0.05] | 0.267881731 | 0.746520328 |
| Notch_3 | Lipodystrophy_1_pPS | 4225 | -0.009476487 | [-0.04 0.02] | 0.538602569 | 0.88195499 |
| Notch_3 | Lipodystrophy_2_pPS | 4225 | 0.006442413 | [-0.02 0.04] | 0.675916412 | 0.925361755 |
| Notch_3 | Liver-Lipid_pPS | 4225 | 0.03384906 | [0. 0.06] | 0.028017607 | 0.356352975 |

**b) Partial Correlation**

| Proteomics | PPS | Sample_Size | r | CI95% | p | fdr_p_adjusted |
|---|---|---|---|---|---|---|
| PTX3 | ASAT_Pos_pPS | 4159 | 0.001265475 | [-0.03 0.03] | 0.935171046 | 0.994163908 |
| PTX3 | ALP_Neg_pPS | 4159 | -0.042475952 | [-0.07 -0.01] | 0.006305657 | 0.193373473 |
| PTX3 | SHBG-LpA_pPS | 4159 | 0.004153113 | [-0.03 0.03] | 0.789507872 | 0.967638637 |
| PTX3 | Obesity_pPS | 4159 | -0.013738661 | [-0.04 0.02] | 0.377172468 | 0.849453763 |
| PTX3 | VAT_Neg_pPS | 4159 | -0.024524735 | [-0.05 0.01] | 0.114885913 | 0.681046037 |
| PTX3 | Bilirubin_pPS | 4159 | 0.024229686 | [-0.01 0.05] | 0.119320025 | 0.681046037 |
| PTX3 | Proinsulin_pPS | 4159 | 0.003731585 | [-0.03 0.03] | 0.810442709 | 0.967638637 |
| PTX3 | Hyper_Insulin_pPS | 4159 | 0.001188384 | [-0.03 0.03] | 0.939112433 | 0.99430893 |
| PTX3 | Beta_Cell_1_pPS | 4159 | 0.020633864 | [-0.01 0.05] | 0.184700997 | 0.738879686 |
| PTX3 | Beta_Cell_2_pPS | 4159 | -0.008677066 | [-0.04 0.02] | 0.577019171 | 0.909113996 |
| SPON1 | Cholesterol_pPS | 4225 | 0.005807706 | [-0.02 0.04] | 0.70671356 | 0.945970482 |
| SPON1 | Lipodystrophy_1_pPS | 4225 | 0.011097371 | [-0.02 0.04] | 0.472140983 | 0.883720482 |
| SPON1 | Lipodystrophy_2_pPS | 4225 | -0.003479673 | [-0.03 0.03] | 0.821635339 | 0.968668398 |
| SPON1 | Liver-Lipid_pPS | 4225 | -0.011513077 | [-0.04 0.02] | 0.455706516 | 0.874087852 |
| SPON1 | ASAT_Pos_pPS | 4225 | 0.01213911 | [-0.02 0.04] | 0.431575173 | 0.864060889 |
| SPON1 | ALP_Neg_pPS | 4225 | 0.011470005 | [-0.02 0.04] | 0.457394225 | 0.874172623 |
| SPON1 | SHBG-LpA_pPS | 4225 | 0.016974488 | [-0.01 0.05] | 0.271408671 | 0.808820203 |
| SPON1 | Obesity_pPS | 4225 | -0.009193359 | [-0.04 0.02] | 0.551420426 | 0.899241939 |
| SPON1 | VAT_Neg_pPS | 4225 | -0.005259813 | [-0.04 0.02] | 0.733272395 | 0.948247837 |
| SPON1 | Bilirubin_pPS | 4225 | 0.032056063 | [0. 0.06] | 0.037765965 | 0.461066941 |
| SPON1 | Proinsulin_pPS | 4225 | 0.028715445 | [-0. 0.06] | 0.062771957 | 0.558815902 |
| SPON1 | Hyper_Insulin_pPS | 4225 | 0.007709118 | [-0.02 0.04] | 0.617450658 | 0.921525431 |
| SPON1 | Beta_Cell_1_pPS | 4225 | -0.034754499 | [-0.06 -0. ] | 0.024299601 | 0.384023142 |
| SPON1 | Beta_Cell_2_pPS | 4225 | 0.006869349 | [-0.02 0.04] | 0.656278227 | 0.936605382 |
| STK4 | Cholesterol_pPS | 4159 | 0.000413355 | [-0.03 0.03] | 0.978803422 | 0.998803911 |
| STK4 | Lipodystrophy_1_pPS | 4159 | 0.014642079 | [-0.02 0.05] | 0.346604627 | 0.847284666 |
| STK4 | Lipodystrophy_2_pPS | 4159 | -0.001277759 | [-0.03 0.03] | 0.934543171 | 0.994163908 |
| STK4 | Liver-Lipid_pPS | 4159 | -0.017879945 | [-0.05 0.01] | 0.250408983 | 0.799790128 |
| STK4 | ASAT_Pos_pPS | 4159 | -0.006295327 | [-0.04 0.02] | 0.685736919 | 0.944421056 |
| STK4 | ALP_Neg_pPS | 4159 | -0.026056471 | [-0.06 0. ] | 0.09391274 | 0.639997931 |
| STK4 | SHBG-LpA_pPS | 4159 | 0.026015716 | [-0. 0.06] | 0.094428008 | 0.64003016 |
| STK4 | Obesity_pPS | 4159 | -0.012551163 | [-0.04 0.02] | 0.419793431 | 0.859608807 |
| STK4 | VAT_Neg_pPS | 4159 | 0.009572641 | [-0.02 0.04] | 0.53834851 | 0.899241939 |
| STK4 | Bilirubin_pPS | 4159 | 0.006011063 | [-0.02 0.04] | 0.699218915 | 0.945970482 |
| STK4 | Proinsulin_pPS | 4159 | 0.02006966 | [-0.01 0.05] | 0.197001222 | 0.760832307 |
| STK4 | Hyper_Insulin_pPS | 4159 | -0.008487396 | [-0.04 0.02] | 0.585373913 | 0.911131843 |
| STK4 | Beta_Cell_1_pPS | 4159 | -0.012328792 | [-0.04 0.02] | 0.428078091 | 0.864060889 |
| STK4 | Beta_Cell_2_pPS | 4159 | -0.004027367 | [-0.03 0.03] | 0.795737811 | 0.967638637 |
| THBS2 | Cholesterol_pPS | 4159 | 0.013300491 | [-0.02 0.04] | 0.392578784 | 0.85037326 |
| THBS2 | Lipodystrophy_1_pPS | 4159 | 0.021308062 | [-0.01 0.05] | 0.170759214 | 0.726214013 |
| THBS2 | Lipodystrophy_2_pPS | 4159 | 0.004006376 | [-0.03 0.03] | 0.796779023 | 0.967638637 |
| THBS2 | Liver-Lipid_pPS | 4159 | -0.015218661 | [-0.05 0.02] | 0.327944426 | 0.84456682 |
| THBS2 | ASAT_Pos_pPS | 4159 | 0.028220832 | [-0. 0.06] | 0.069630847 | 0.573238056 |
| THBS2 | ALP_Neg_pPS | 4159 | -0.01079412 | [-0.04 0.02] | 0.487788115 | 0.895803896 |
| THBS2 | SHBG-LpA_pPS | 4159 | 0.017116054 | [-0.01 0.05] | 0.27122547 | 0.808820203 |
| THBS2 | Obesity_pPS | 4159 | 0.027361264 | [-0. 0.06] | 0.07857372 | 0.602398521 |
| THBS2 | VAT_Neg_pPS | 4159 | 0.021965066 | [-0.01 0.05] | 0.157946049 | 0.712555204 |
| THBS2 | Bilirubin_pPS | 4159 | -0.005680927 | [-0.04 0.02] | 0.714996129 | 0.945970482 |
| THBS2 | Proinsulin_pPS | 4159 | 0.008230844 | [-0.02 0.04] | 0.596763273 | 0.918301271 |
| THBS2 | Hyper_Insulin_pPS | 4159 | 0.02479561 | [-0.01 0.06] | 0.110930167 | 0.680721977 |
| THBS2 | Beta_Cell_1_pPS | 4159 | -0.015367956 | [-0.05 0.02] | 0.323220968 | 0.841027489 |
| THBS2 | Beta_Cell_2_pPS | 4159 | -0.009971718 | [-0.04 0.02] | 0.521545086 | 0.895803896 |
| CDH5 | Cholesterol_pPS | 4225 | 0.020131036 | [-0.01 0.05] | 0.192102653 | 0.750920235 |
| CDH5 | Lipodystrophy_1_pPS | 4225 | -0.007724375 | [-0.04 0.02] | 0.616754634 | 0.921525431 |
| CDH5 | Lipodystrophy_2_pPS | 4225 | 0.013782072 | [-0.02 0.04] | 0.371880265 | 0.847284666 |
| CDH5 | Liver-Lipid_pPS | 4225 | 0.010779961 | [-0.02 0.04] | 0.484906028 | 0.895803896 |
| CDH5 | ASAT_Pos_pPS | 4225 | -0.006658904 | [-0.04 0.02] | 0.666161096 | 0.937489141 |
| CDH5 | ALP_Neg_pPS | 4225 | 0.074978515 | [0.04 0.1 ] | 1.15E-06 | 0.000184681 |
| CDH5 | SHBG-LpA_pPS | 4225 | -0.003761934 | [-0.03 0.03] | 0.807440324 | 0.967638637 |
| CDH5 | Obesity_pPS | 4225 | 0.00549743 | [-0.02 0.04] | 0.721712663 | 0.946314812 |
| CDH5 | VAT_Neg_pPS | 4225 | -0.008609217 | [-0.04 0.02] | 0.57699101 | 0.909113996 |
| CDH5 | Bilirubin_pPS | 4225 | 0.002697534 | [-0.03 0.03] | 0.861261589 | 0.97556306 |
| CDH5 | Proinsulin_pPS | 4225 | 0.025448472 | [-0. 0.06] | 0.099142393 | 0.649849377 |
| CDH5 | Hyper_Insulin_pPS | 4225 | -0.001663409 | [-0.03 0.03] | 0.914178761 | 0.989461462 |
| CDH5 | Beta_Cell_1_pPS | 4225 | -0.012586209 | [-0.04 0.02] | 0.414803869 | 0.854508456 |
| CDH5 | Beta_Cell_2_pPS | 4225 | 0.029184534 | [-0. 0.06] | 0.058595808 | 0.558815902 |
| COL1A1 | Cholesterol_pPS | 4225 | 0.004506107 | [-0.03 0.03] | 0.770329688 | 0.960953645 |
| COL1A1 | Lipodystrophy_1_pPS | 4225 | -0.00907368 | [-0.04 0.02] | 0.556613703 | 0.900085938 |
| COL1A1 | Lipodystrophy_2_pPS | 4225 | -0.008991846 | [-0.04 0.02] | 0.560178387 | 0.902076574 |
| COL1A1 | Liver-Lipid_pPS | 4225 | 0.018461236 | [-0.01 0.05] | 0.231629983 | 0.786988264 |
| COL1A1 | ASAT_Pos_pPS | 4225 | -0.013054011 | [-0.04 0.02] | 0.397675028 | 0.85037326 |
| COL1A1 | ALP_Neg_pPS | 4225 | -0.038749815 | [-0.07 -0.01] | 0.012022925 | 0.284138119 |
| COL1A1 | SHBG-LpA_pPS | 4225 | -0.00060691 | [-0.03 0.03] | 0.96863479 | 0.996924797 |
| COL1A1 | Obesity_pPS | 4225 | -0.032173562 | [-0.06 -0. ] | 0.037068915 | 0.457947978 |
| COL1A1 | VAT_Neg_pPS | 4225 | -0.019192046 | [-0.05 0.01] | 0.213672219 | 0.767305935 |
| COL1A1 | Bilirubin_pPS | 4225 | -0.000373056 | [-0.03 0.03] | 0.980717311 | 0.998803911 |
| COL1A1 | Proinsulin_pPS | 4225 | -0.005239857 | [-0.04 0.03] | 0.734246051 | 0.948434512 |
| COL1A1 | Hyper_Insulin_pPS | 4225 | 0.004461413 | [-0.03 0.03] | 0.772544676 | 0.962056893 |
| COL1A1 | Beta_Cell_1_pPS | 4225 | -0.002108634 | [-0.03 0.03] | 0.891335448 | 0.981230818 |
| COL1A1 | Beta_Cell_2_pPS | 4225 | 0.028182255 | [-0. 0.06] | 0.067814432 | 0.568910429 |
| EPHB4 | Cholesterol_pPS | 4225 | 0.011881967 | [-0.02 0.04] | 0.441396314 | 0.866107938 |
| EPHB4 | Lipodystrophy_1_pPS | 4225 | 0.010593223 | [-0.02 0.04] | 0.492502384 | 0.895803896 |
| EPHB4 | Lipodystrophy_2_pPS | 4225 | -0.003021054 | [-0.03 0.03] | 0.844822385 | 0.972178273 |
| EPHB4 | Liver-Lipid_pPS | 4225 | 0.012876039 | [-0.02 0.04] | 0.404140756 | 0.85241904 |
| EPHB4 | ASAT_Pos_pPS | 4225 | -0.007719203 | [-0.04 0.02] | 0.616990549 | 0.921525431 |
| EPHB4 | ALP_Neg_pPS | 4225 | -0.10534785 | [-0.14 -0.08] | 7.68E-12 | 1.98E-09 |
| EPHB4 | SHBG-LpA_pPS | 4225 | 0.003640977 | [-0.03 0.03] | 0.813515833 | 0.967790883 |
| EPHB4 | Obesity_pPS | 4225 | 0.009950254 | [-0.02 0.04] | 0.519138363 | 0.895803896 |
| EPHB4 | VAT_Neg_pPS | 4225 | -0.027926927 | [-0.06 0. ] | 0.070344508 | 0.573666874 |
| EPHB4 | Bilirubin_pPS | 4225 | -0.000204709 | [-0.03 0.03] | 0.989418184 | 0.998803911 |
| EPHB4 | Proinsulin_pPS | 4225 | 0.018444232 | [-0.01 0.05] | 0.232060199 | 0.786988264 |
| EPHB4 | Hyper_Insulin_pPS | 4225 | 0.010031449 | [-0.02 0.04] | 0.515734196 | 0.895803896 |
| EPHB4 | Beta_Cell_1_pPS | 4225 | 0.004063876 | [-0.03 0.03] | 0.792325548 | 0.967638637 |
| EPHB4 | Beta_Cell_2_pPS | 4225 | 0.020470154 | [-0.01 0.05] | 0.184719921 | 0.738879686 |
| JAM_A | Cholesterol_pPS | 4225 | 0.003270554 | [-0.03 0.03] | 0.832190065 | 0.971458352 |
| JAM_A | Lipodystrophy_1_pPS | 4225 | 0.017770684 | [-0.01 0.05] | 0.249559469 | 0.799583871 |
| JAM_A | Lipodystrophy_2_pPS | 4225 | 0.026059793 | [-0. 0.06] | 0.091286204 | 0.63465347 |
| JAM_A | Liver-Lipid_pPS | 4225 | -0.029007215 | [-0.06 0. ] | 0.060146476 | 0.558815902 |
| JAM_A | ASAT_Pos_pPS | 4225 | 0.003818218 | [-0.03 0.03] | 0.804617206 | 0.967638637 |
| JAM_A | ALP_Neg_pPS | 4225 | -0.013390624 | [-0.04 0.02] | 0.385617621 | 0.85037326 |
| JAM_A | SHBG-LpA_pPS | 4225 | -0.001033191 | [-0.03 0.03] | 0.946630638 | 0.996425456 |
| JAM_A | Obesity_pPS | 4225 | -0.011276503 | [-0.04 0.02] | 0.465019474 | 0.879924083 |
| JAM_A | VAT_Neg_pPS | 4225 | -8.07E-06 | [-0.03 0.03] | 0.999583089 | 0.999583089 |
| JAM_A | Bilirubin_pPS | 4225 | 0.002383069 | [-0.03 0.03] | 0.87729846 | 0.97781424 |
| JAM_A | Proinsulin_pPS | 4225 | 0.017131066 | [-0.01 0.05] | 0.267011553 | 0.808113719 |
| JAM_A | Hyper_Insulin_pPS | 4225 | -0.002708837 | [-0.03 0.03] | 0.860686179 | 0.97556306 |
| JAM_A | Beta_Cell_1_pPS | 4225 | -0.002366187 | [-0.03 0.03] | 0.878160925 | 0.97781424 |

**Table S7: Associations between pPSs and plasma proteins in SCAPIS**

**a) Spearman Correlation**

| Proteomics | PPS | Sample_Size | r | CI95% | p | fdr_p_adjusted |
|---|---|---|---|---|---|---|
| Notch_3 | ASAT_Pos_pPS | 4225 | 0.004384327 | [-0.03 0.03] | 0.776032445 | 0.953637204 |
| Notch_3 | ALP_Neg_pPS | 4225 | -0.033079667 | [-0.06 -0. ] | 0.031787462 | 0.373333595 |
| Notch_3 | SHBG-LpA_pPS | 4225 | 0.007308853 | [-0.02 0.04] | 0.635310333 | 0.914460114 |
| Notch_3 | Obesity_pPS | 4225 | 0.011495037 | [-0.02 0.04] | 0.455716448 | 0.835860883 |
| Notch_3 | VAT_Neg_pPS | 4225 | 0.010482657 | [-0.02 0.04] | 0.496364412 | 0.863780476 |
| Notch_3 | Bilirubin_pPS | 4225 | -0.014901358 | [-0.05 0.02] | 0.333554796 | 0.779518406 |
| Notch_3 | Proinsulin_pPS | 4225 | 0.019371899 | [-0.01 0.05] | 0.20870698 | 0.709689105 |
| Notch_3 | Hyper_Insulin_pPS | 4225 | 0.029373301 | [-0. 0.06] | 0.05659874 | 0.451781658 |
| Notch_3 | Beta_Cell_1_pPS | 4225 | -0.000741468 | [-0.03 0.03] | 0.961626444 | 0.99612031 |
| Notch_3 | Beta_Cell_2_pPS | 4225 | 0.030333657 | [0. 0.06] | 0.048981297 | 0.438587647 |
| Notch_3 | Total_GRS | 4225 | 0.02814138 | [-0. 0.06] | 0.06778855 | 0.480967608 |
| PTX3 | Cholesterol_pPS | 4159 | 0.005730245 | [-0.02 0.04] | 0.712199619 | 0.940386535 |
| PTX3 | Lipodystrophy_1_pPS | 4159 | -0.023926223 | [-0.05 0.01] | 0.123429284 | 0.617146422 |
| PTX3 | Lipodystrophy_2_pPS | 4159 | 0.000984315 | [-0.03 0.03] | 0.949473434 | 0.993154165 |
| PTX3 | Liver-Lipid_pPS | 4159 | -0.011066045 | [-0.04 0.02] | 0.476198933 | 0.84751951 |
| PTX3 | ASAT_Pos_pPS | 4159 | -0.001706658 | [-0.03 0.03] | 0.912511711 | 0.990056084 |
| PTX3 | ALP_Neg_pPS | 4159 | -0.043532931 | [-0.07 -0.01] | 0.00504912 | 0.140763349 |
| PTX3 | SHBG-LpA_pPS | 4159 | 0.001926858 | [-0.03 0.03] | 0.901278058 | 0.987003157 |
| PTX3 | Obesity_pPS | 4159 | -0.014080768 | [-0.04 0.02] | 0.364653432 | 0.798887077 |
| PTX3 | VAT_Neg_pPS | 4159 | -0.022953312 | [-0.05 0.01] | 0.139440376 | 0.634028726 |
| PTX3 | Bilirubin_pPS | 4159 | 0.024176304 | [-0.01 0.05] | 0.119554899 | 0.615837655 |
| PTX3 | Proinsulin_pPS | 4159 | -0.000397117 | [-0.03 0.03] | 0.979603878 | 0.996862516 |
| PTX3 | Hyper_Insulin_pPS | 4159 | -0.005550064 | [-0.04 0.02] | 0.720864513 | 0.941593022 |
| PTX3 | Beta_Cell_1_pPS | 4159 | 0.017446992 | [-0.01 0.05] | 0.261316762 | 0.745847221 |
| PTX3 | Beta_Cell_2_pPS | 4159 | -0.004850046 | [-0.04 0.03] | 0.754860362 | 0.950403646 |
| PTX3 | Total_GRS | 4159 | -0.011894607 | [-0.04 0.02] | 0.443809828 | 0.831350198 |
| SPON1 | Cholesterol_pPS | 4225 | 0.005626994 | [-0.02 0.04] | 0.715014855 | 0.94141015 |
| SPON1 | Lipodystrophy_1_pPS | 4225 | 0.015038146 | [-0.02 0.05] | 0.329135896 | 0.775125841 |
| SPON1 | Lipodystrophy_2_pPS | 4225 | -0.000325181 | [-0.03 0.03] | 0.983165515 | 0.99799074 |
| SPON1 | Liver-Lipid_pPS | 4225 | -0.01125765 | [-0.04 0.02] | 0.465076278 | 0.841554994 |
| SPON1 | ASAT_Pos_pPS | 4225 | 0.013869214 | [-0.02 0.04] | 0.368123479 | 0.799230784 |
| SPON1 | ALP_Neg_pPS | 4225 | 0.016026508 | [-0.01 0.05] | 0.298339568 | 0.759754073 |
| SPON1 | SHBG-LpA_pPS | 4225 | 0.018205166 | [-0.01 0.05] | 0.237443313 | 0.726631332 |
| SPON1 | Obesity_pPS | 4225 | -0.003848194 | [-0.03 0.03] | 0.802816312 | 0.958794037 |
| SPON1 | VAT_Neg_pPS | 4225 | -0.006546003 | [-0.04 0.02] | 0.671008714 | 0.925160508 |
| SPON1 | Bilirubin_pPS | 4225 | 0.031675668 | [0. 0.06] | 0.03979233 | 0.406766041 |
| SPON1 | Proinsulin_pPS | 4225 | 0.023151419 | [-0.01 0.05] | 0.13297858 | 0.629316669 |
| SPON1 | Hyper_Insulin_pPS | 4225 | 0.008964596 | [-0.02 0.04] | 0.560762238 | 0.89774001 |
| SPON1 | Beta_Cell_1_pPS | 4225 | -0.02553832 | [-0.06 0. ] | 0.09743658 | 0.572901389 |
| SPON1 | Beta_Cell_2_pPS | 4225 | 0.002849998 | [-0.03 0.03] | 0.853282308 | 0.976392691 |
| SPON1 | Total_GRS | 4225 | 0.016729919 | [-0.01 0.05] | 0.277632508 | 0.747422497 |
| STK4 | Cholesterol_pPS | 4159 | -0.001781824 | [-0.03 0.03] | 0.908674995 | 0.990056084 |
| STK4 | Lipodystrophy_1_pPS | 4159 | 0.013704852 | [-0.02 0.04] | 0.377598635 | 0.803170077 |
| STK4 | Lipodystrophy_2_pPS | 4159 | -0.000152629 | [-0.03 0.03] | 0.992160186 | 0.999402232 |
| STK4 | Liver-Lipid_pPS | 4159 | -0.019266595 | [-0.05 0.01] | 0.214806795 | 0.715184753 |
| STK4 | ASAT_Pos_pPS | 4159 | -0.005126487 | [-0.04 0.03] | 0.74137423 | 0.945527195 |
| STK4 | ALP_Neg_pPS | 4159 | -0.02445125 | [-0.05 0.01] | 0.115405904 | 0.610816194 |
| STK4 | SHBG-LpA_pPS | 4159 | 0.029159822 | [-0. 0.06] | 0.060429859 | 0.463868622 |
| STK4 | Obesity_pPS | 4159 | -0.011412806 | [-0.04 0.02] | 0.462488631 | 0.83934244 |
| STK4 | VAT_Neg_pPS | 4159 | 0.009510343 | [-0.02 0.04] | 0.540359997 | 0.883003903 |
| STK4 | Bilirubin_pPS | 4159 | 0.005836896 | [-0.02 0.04] | 0.707088115 | 0.937738119 |
| STK4 | Proinsulin_pPS | 4159 | 0.011888413 | [-0.02 0.04] | 0.444047187 | 0.831350198 |
| STK4 | Hyper_Insulin_pPS | 4159 | -0.008350542 | [-0.04 0.02] | 0.59085404 | 0.904970671 |
| STK4 | Beta_Cell_1_pPS | 4159 | -0.007437615 | [-0.04 0.02] | 0.632063971 | 0.914460114 |
| STK4 | Beta_Cell_2_pPS | 4159 | -0.003865058 | [-0.03 0.03] | 0.803496295 | 0.959191079 |
| STK4 | Total_GRS | 4159 | -0.007930623 | [-0.04 0.02] | 0.609656362 | 0.909483337 |
| THBS2 | Cholesterol_pPS | 4159 | 0.015178229 | [-0.02 0.05] | 0.328471547 | 0.775125841 |
| THBS2 | Lipodystrophy_1_pPS | 4159 | 0.032754169 | [0. 0.06] | 0.034926723 | 0.386023413 |
| THBS2 | Lipodystrophy_2_pPS | 4159 | 0.013202601 | [-0.02 0.04] | 0.395329916 | 0.812191725 |
| THBS2 | Liver-Lipid_pPS | 4159 | -0.013157387 | [-0.04 0.02] | 0.396950421 | 0.812191725 |
| THBS2 | ASAT_Pos_pPS | 4159 | 0.035749605 | [0.01 0.07] | 0.021322968 | 0.313142997 |
| THBS2 | ALP_Neg_pPS | 4159 | -0.009187588 | [-0.04 0.02] | 0.554192734 | 0.896004422 |
| THBS2 | SHBG-LpA_pPS | 4159 | 0.020884485 | [-0.01 0.05] | 0.17874034 | 0.679661798 |
| THBS2 | Obesity_pPS | 4159 | 0.032491744 | [0. 0.06] | 0.036411767 | 0.386599178 |
| THBS2 | VAT_Neg_pPS | 4159 | 0.023987549 | [-0.01 0.05] | 0.122470245 | 0.615837655 |
| THBS2 | Bilirubin_pPS | 4159 | -0.004828822 | [-0.04 0.03] | 0.755898928 | 0.950403646 |
| THBS2 | Proinsulin_pPS | 4159 | 0.005317764 | [-0.03 0.04] | 0.732088757 | 0.942134248 |
| THBS2 | Hyper_Insulin_pPS | 4159 | 0.033599474 | [0. 0.06] | 0.030489626 | 0.369452038 |
| THBS2 | Beta_Cell_1_pPS | 4159 | -0.012839587 | [-0.04 0.02] | 0.408543443 | 0.813957763 |
| THBS2 | Beta_Cell_2_pPS | 4159 | -0.011139887 | [-0.04 0.02] | 0.473260832 | 0.846453261 |
| THBS2 | Total_GRS | 4159 | 0.037581727 | [0.01 0.07] | 0.015508408 | 0.258767146 |
| CDH5 | Cholesterol_pPS | 4225 | 0.023004043 | [-0.01 0.05] | 0.135465388 | 0.629316669 |
| CDH5 | Lipodystrophy_1_pPS | 4225 | -0.007390285 | [-0.04 0.02] | 0.631547378 | 0.914460114 |
| CDH5 | Lipodystrophy_2_pPS | 4225 | 0.014050642 | [-0.02 0.04] | 0.361890755 | 0.7979754 |
| CDH5 | Liver-Lipid_pPS | 4225 | 0.01586937 | [-0.01 0.05] | 0.303102793 | 0.76319014 |
| CDH5 | ASAT_Pos_pPS | 4225 | -0.007251965 | [-0.04 0.02] | 0.637944716 | 0.915577927 |
| CDH5 | ALP_Neg_pPS | 4225 | 0.081579449 | [0.05 0.11] | 1.14E-07 | 2.10E-05 |
| CDH5 | SHBG-LpA_pPS | 4225 | -0.005680084 | [-0.04 0.02] | 0.712445016 | 0.940386535 |
| CDH5 | Obesity_pPS | 4225 | 0.012159603 | [-0.02 0.04] | 0.430085148 | 0.827992564 |
| CDH5 | VAT_Neg_pPS | 4225 | -0.006466377 | [-0.04 0.02] | 0.674779841 | 0.925361755 |
| CDH5 | Bilirubin_pPS | 4225 | 0.002726528 | [-0.03 0.03] | 0.859571019 | 0.977006629 |
| CDH5 | Proinsulin_pPS | 4225 | 0.042457134 | [0.01 0.07] | 0.005847423 | 0.152834705 |
| CDH5 | Hyper_Insulin_pPS | 4225 | 0.000553011 | [-0.03 0.03] | 0.971374849 | 0.996862516 |
| CDH5 | Beta_Cell_1_pPS | 4225 | 0.001163591 | [-0.03 0.03] | 0.939814073 | 0.992305601 |
| CDH5 | Beta_Cell_2_pPS | 4225 | 0.03284501 | [0. 0.06] | 0.033020483 | 0.378159894 |
| CDH5 | Total_GRS | 4225 | 0.0305137 | [0. 0.06] | 0.047653518 | 0.436063541 |
| COL1A1 | Cholesterol_pPS | 4225 | 0.002613352 | [-0.03 0.03] | 0.865343266 | 0.977006629 |
| COL1A1 | Lipodystrophy_1_pPS | 4225 | -0.014302372 | [-0.04 0.02] | 0.353353096 | 0.796092727 |
| COL1A1 | Lipodystrophy_2_pPS | 4225 | -0.012158905 | [-0.04 0.02] | 0.43011164 | 0.827992564 |
| COL1A1 | Liver-Lipid_pPS | 4225 | 0.01659873 | [-0.01 0.05] | 0.281418329 | 0.748127084 |
| COL1A1 | ASAT_Pos_pPS | 4225 | -0.01737295 | [-0.05 0.01] | 0.259578726 | 0.743022111 |
| COL1A1 | ALP_Neg_pPS | 4225 | -0.039934869 | [-0.07 -0.01] | 0.009532483 | 0.215652903 |
| COL1A1 | SHBG-LpA_pPS | 4225 | -0.001339624 | [-0.03 0.03] | 0.930730295 | 0.990056084 |
| COL1A1 | Obesity_pPS | 4225 | -0.033396715 | [-0.06 -0. ] | 0.030184344 | 0.369452038 |
| COL1A1 | VAT_Neg_pPS | 4225 | -0.019071065 | [-0.05 0.01] | 0.215863372 | 0.715705689 |
| COL1A1 | Bilirubin_pPS | 4225 | -0.001444196 | [-0.03 0.03] | 0.925338276 | 0.990056084 |
| COL1A1 | Proinsulin_pPS | 4225 | -0.011932813 | [-0.04 0.02] | 0.438736658 | 0.830394412 |
| COL1A1 | Hyper_Insulin_pPS | 4225 | -0.001652988 | [-0.03 0.03] | 0.91458291 | 0.990056084 |
| COL1A1 | Beta_Cell_1_pPS | 4225 | 0.000583479 | [-0.03 0.03] | 0.969798509 | 0.996862516 |
| COL1A1 | Beta_Cell_2_pPS | 4225 | 0.025633296 | [-0. 0.06] | 0.096197184 | 0.568531537 |
| COL1A1 | Total_GRS | 4225 | -0.026077489 | [-0.06 0. ] | 0.090567166 | 0.55302075 |
| EPHB4 | Cholesterol_pPS | 4225 | 0.014294246 | [-0.02 0.04] | 0.353626697 | 0.796092727 |
| EPHB4 | Lipodystrophy_1_pPS | 4225 | 0.008398219 | [-0.02 0.04] | 0.58578355 | 0.903925569 |
| EPHB4 | Lipodystrophy_2_pPS | 4225 | -0.005339916 | [-0.04 0.02] | 0.728966291 | 0.942134248 |
| EPHB4 | Liver-Lipid_pPS | 4225 | 0.010124852 | [-0.02 0.04] | 0.511179653 | 0.870690038 |
| EPHB4 | ASAT_Pos_pPS | 4225 | -0.005448225 | [-0.04 0.02] | 0.72369184 | 0.942134248 |
| EPHB4 | ALP_Neg_pPS | 4225 | -0.102422943 | [-0.13 -0.07] | 2.67E-11 | 7.38E-09 |

**b) Partial Correlation**

| Proteomics | PPS | Sample_Size | r | CI95% | p | fdr_p_adjusted |
|---|---|---|---|---|---|---|
| JAM_A | Beta_Cell_2_pPS | 4225 | 0.001650968 | [-0.03 0.03] | 0.914818192 | 0.989461462 |
| FS | Cholesterol_pPS | 4159 | 0.013206001 | [-0.02 0.04] | 0.395950428 | 0.85037326 |
| FS | Lipodystrophy_1_pPS | 4159 | -0.008087914 | [-0.04 0.02] | 0.603151911 | 0.918301271 |
| FS | Lipodystrophy_2_pPS | 4159 | 0.007223205 | [-0.02 0.04] | 0.642439849 | 0.933976497 |
| FS | Liver-Lipid_pPS | 4159 | -0.093561058 | [-0.12 -0.06] | 1.67E-09 | 3.91E-07 |
| FS | ASAT_Pos_pPS | 4159 | -0.033594009 | [-0.06 -0. ] | 0.030778614 | 0.419027441 |
| FS | ALP_Neg_pPS | 4159 | 0.01167073 | [-0.02 0.04] | 0.45314506 | 0.871771227 |
| FS | SHBG-LpA_pPS | 4159 | 0.018271064 | [-0.01 0.05] | 0.240193792 | 0.790085251 |
| FS | Obesity_pPS | 4159 | 0.004619162 | [-0.03 0.04] | 0.76653806 | 0.960245786 |
| FS | VAT_Neg_pPS | 4159 | -0.015615621 | [-0.05 0.01] | 0.315483469 | 0.837820016 |
| FS | Bilirubin_pPS | 4159 | 0.002563801 | [-0.03 0.03] | 0.869106855 | 0.977511154 |
| FS | Proinsulin_pPS | 4159 | 0.003837164 | [-0.03 0.03] | 0.805185803 | 0.967638637 |
| FS | Hyper_Insulin_pPS | 4159 | 0.00654949 | [-0.02 0.04] | 0.673766466 | 0.939698114 |
| FS | Beta_Cell_1_pPS | 4159 | 0.016424708 | [-0.01 0.05] | 0.291060502 | 0.81655641 |
| FS | Beta_Cell_2_pPS | 4159 | -0.004588531 | [-0.04 0.03] | 0.768041703 | 0.960245786 |
| PLC | Cholesterol_pPS | 4223 | 0.006362476 | [-0.02 0.04] | 0.680252362 | 0.943176659 |
| PLC | Lipodystrophy_1_pPS | 4223 | 0.019863726 | [-0.01 0.05] | 0.198180158 | 0.763097292 |
| PLC | Lipodystrophy_2_pPS | 4223 | -0.005796343 | [-0.04 0.02] | 0.707327436 | 0.945970482 |
| PLC | Liver-Lipid_pPS | 4223 | 0.018779413 | [-0.01 0.05] | 0.223794439 | 0.780100778 |
| PLC | ASAT_Pos_pPS | 4223 | -0.010355516 | [-0.04 0.02] | 0.502365448 | 0.895803896 |
| PLC | ALP_Neg_pPS | 4223 | 0.005687372 | [-0.02 0.04] | 0.712582964 | 0.945970482 |
| PLC | SHBG-LpA_pPS | 4223 | 0.031861263 | [0. 0.06] | 0.038992746 | 0.467187506 |
| PLC | Obesity_pPS | 4223 | 0.013618887 | [-0.02 0.04] | 0.377683415 | 0.84970522 |
| PLC | VAT_Neg_pPS | 4223 | -0.010949456 | [-0.04 0.02] | 0.478171309 | 0.88808168 |
| PLC | Bilirubin_pPS | 4223 | -0.013379458 | [-0.04 0.02] | 0.386127169 | 0.85037326 |
| PLC | Proinsulin_pPS | 4223 | 0.003159284 | [-0.03 0.03] | 0.837856663 | 0.972099644 |
| PLC | Hyper_Insulin_pPS | 4223 | 0.007684758 | [-0.02 0.04] | 0.618646373 | 0.92152711 |
| PLC | Beta_Cell_1_pPS | 4223 | 0.005959393 | [-0.02 0.04] | 0.699489804 | 0.945970482 |
| PLC | Beta_Cell_2_pPS | 4223 | -0.00739213 | [-0.04 0.02] | 0.632070519 | 0.930939769 |
| IGFBP_1 | Cholesterol_pPS | 4225 | 0.008848941 | [-0.02 0.04] | 0.566429798 | 0.90516325 |
| IGFBP_1 | Lipodystrophy_1_pPS | 4225 | -0.01634587 | [-0.05 0.01] | 0.289559319 | 0.81655641 |
| IGFBP_1 | Lipodystrophy_2_pPS | 4225 | -0.007115193 | [-0.04 0.02] | 0.644808809 | 0.934865668 |
| IGFBP_1 | Liver-Lipid_pPS | 4225 | -0.090079654 | [-0.12 -0.06] | 4.98E-09 | 1.07E-06 |
| IGFBP_1 | ASAT_Pos_pPS | 4225 | -0.001372842 | [-0.03 0.03] | 0.929126513 | 0.991797911 |
| IGFBP_1 | ALP_Neg_pPS | 4225 | 0.009556995 | [-0.02 0.04] | 0.535788676 | 0.898959051 |
| IGFBP_1 | SHBG-LpA_pPS | 4225 | 0.009711631 | [-0.02 0.04] | 0.529209539 | 0.898959051 |
| IGFBP_1 | Obesity_pPS | 4225 | -0.052942075 | [-0.08 -0.02] | 0.00059821 | 0.042010707 |
| IGFBP_1 | VAT_Neg_pPS | 4225 | -0.029711711 | [-0.06 0. ] | 0.054180483 | 0.549483952 |
| IGFBP_1 | Bilirubin_pPS | 4225 | -0.00376834 | [-0.03 0.03] | 0.807118878 | 0.967638637 |
| IGFBP_1 | Proinsulin_pPS | 4225 | 0.015140434 | [-0.02 0.05] | 0.326604363 | 0.84456682 |
| IGFBP_1 | Hyper_Insulin_pPS | 4225 | -0.020678608 | [-0.05 0.01] | 0.180287182 | 0.738879686 |
| IGFBP_1 | Beta_Cell_1_pPS | 4225 | -0.012391101 | [-0.04 0.02] | 0.4220748 | 0.860518922 |
| IGFBP_1 | Beta_Cell_2_pPS | 4225 | 0.04280325 | [0.01 0.07] | 0.005530135 | 0.177399096 |
| IGFBP_7 | Cholesterol_pPS | 4225 | 0.024417314 | [-0.01 0.05] | 0.11360674 | 0.681046037 |
| IGFBP_7 | Lipodystrophy_1_pPS | 4225 | 0.01065859 | [-0.02 0.04] | 0.489836083 | 0.895803896 |
| IGFBP_7 | Lipodystrophy_2_pPS | 4225 | 0.010035586 | [-0.02 0.04] | 0.515561037 | 0.895803896 |
| IGFBP_7 | Liver-Lipid_pPS | 4225 | -0.009062905 | [-0.04 0.02] | 0.55708242 | 0.900278741 |
| IGFBP_7 | ASAT_Pos_pPS | 4225 | 0.020391191 | [-0.01 0.05] | 0.186419954 | 0.742222258 |
| IGFBP_7 | ALP_Neg_pPS | 4225 | -0.006146818 | [-0.04 0.02] | 0.690449968 | 0.945319264 |
| IGFBP_7 | SHBG-LpA_pPS | 4225 | 0.017808878 | [-0.01 0.05] | 0.248543149 | 0.797743002 |
| IGFBP_7 | Obesity_pPS | 4225 | -0.015703063 | [-0.05 0.01] | 0.308946738 | 0.830335001 |
| IGFBP_7 | VAT_Neg_pPS | 4225 | -0.010336924 | [-0.04 0.02] | 0.503031375 | 0.895803896 |
| IGFBP_7 | Bilirubin_pPS | 4225 | -0.006595792 | [-0.04 0.02] | 0.669136391 | 0.937489141 |
| IGFBP_7 | Proinsulin_pPS | 4225 | 0.01506283 | [-0.02 0.05] | 0.329090353 | 0.84456682 |
| IGFBP_7 | Hyper_Insulin_pPS | 4225 | 0.009007695 | [-0.02 0.04] | 0.559487152 | 0.902076574 |
| IGFBP_7 | Beta_Cell_1_pPS | 4225 | -0.003410815 | [-0.03 0.03] | 0.825107298 | 0.970677755 |
| IGFBP_7 | Beta_Cell_2_pPS | 4225 | 0.005676687 | [-0.02 0.04] | 0.713033678 | 0.945970482 |
| MB | Cholesterol_pPS | 4225 | 0.013820702 | [-0.02 0.04] | 0.370541237 | 0.847284666 |
| MB | Lipodystrophy_1_pPS | 4225 | 0.032765833 | [0. 0.06] | 0.033719785 | 0.436493301 |
| MB | Lipodystrophy_2_pPS | 4225 | 0.001483396 | [-0.03 0.03] | 0.923435981 | 0.990503296 |
| MB | Liver-Lipid_pPS | 4225 | 0.018430923 | [-0.01 0.05] | 0.2323973 | 0.786988264 |
| MB | ASAT_Pos_pPS | 4225 | -0.003330044 | [-0.03 0.03] | 0.829184296 | 0.971458352 |
| MB | ALP_Neg_pPS | 4225 | -0.01887852 | [-0.05 0.01] | 0.221249346 | 0.778604255 |
| MB | SHBG-LpA_pPS | 4225 | 0.017467625 | [-0.01 0.05] | 0.257726563 | 0.80473167 |
| MB | Obesity_pPS | 4225 | 0.023826052 | [-0.01 0.05] | 0.122621244 | 0.684037897 |
| MB | VAT_Neg_pPS | 4225 | -0.026708708 | [-0.06 0. ] | 0.083502503 | 0.614725404 |
| MB | Bilirubin_pPS | 4225 | -0.000284313 | [-0.03 0.03] | 0.985303731 | 0.998803911 |
| MB | Proinsulin_pPS | 4225 | -0.002441273 | [-0.03 0.03] | 0.874326212 | 0.97781424 |
| MB | Hyper_Insulin_pPS | 4225 | -0.000240207 | [-0.03 0.03] | 0.987583353 | 0.998803911 |
| MB | Beta_Cell_1_pPS | 4225 | 0.022591308 | [-0.01 0.05] | 0.143239016 | 0.697681852 |
| MB | Beta_Cell_2_pPS | 4225 | -0.02228553 | [-0.05 0.01] | 0.148734249 | 0.703126624 |
| MMP_2 | Cholesterol_pPS | 4225 | 0.000889491 | [-0.03 0.03] | 0.954044565 | 0.996425456 |
| MMP_2 | Lipodystrophy_1_pPS | 4225 | -0.030016618 | [-0.06 0. ] | 0.051755914 | 0.540728413 |
| MMP_2 | Lipodystrophy_2_pPS | 4225 | 0.018128858 | [-0.01 0.05] | 0.240142351 | 0.790085251 |
| MMP_2 | Liver-Lipid_pPS | 4225 | 0.023068171 | [-0.01 0.05] | 0.134981726 | 0.694065456 |
| MMP_2 | ASAT_Pos_pPS | 4225 | -0.003982224 | [-0.03 0.03] | 0.796405529 | 0.967638637 |
| MMP_2 | ALP_Neg_pPS | 4225 | -0.003792813 | [-0.03 0.03] | 0.805891185 | 0.967638637 |
| MMP_2 | SHBG-LpA_pPS | 4225 | 0.00753668 | [-0.02 0.04] | 0.625341435 | 0.927058714 |
| MMP_2 | Obesity_pPS | 4225 | -0.006651918 | [-0.04 0.02] | 0.666490173 | 0.937489141 |
| MMP_2 | VAT_Neg_pPS | 4225 | -0.020082145 | [-0.05 0.01] | 0.193184675 | 0.752352981 |
| MMP_2 | Bilirubin_pPS | 4225 | -0.020747516 | [-0.05 0.01] | 0.178839374 | 0.738879686 |
| MMP_2 | Proinsulin_pPS | 4225 | 0.020892428 | [-0.01 0.05] | 0.175822922 | 0.732083915 |
| MMP_2 | Hyper_Insulin_pPS | 4225 | 0.018532293 | [-0.01 0.05] | 0.229838307 | 0.786988264 |
| MMP_2 | Beta_Cell_1_pPS | 4225 | -0.005583396 | [-0.04 0.02] | 0.717545955 | 0.945970482 |
| MMP_2 | Beta_Cell_2_pPS | 4225 | 0.026927451 | [-0. 0.06] | 0.081003111 | 0.610643339 |
| BNP | Cholesterol_pPS | 4159 | 0.010766222 | [-0.02 0.04] | 0.488913622 | 0.895803896 |
| BNP | Lipodystrophy_1_pPS | 4159 | -0.024011736 | [-0.05 0.01] | 0.12268071 | 0.684037897 |
| BNP | Lipodystrophy_2_pPS | 4159 | 0.00173806 | [-0.03 0.03] | 0.911047858 | 0.989461462 |
| BNP | Liver-Lipid_pPS | 4159 | 0.017822038 | [-0.01 0.05] | 0.251946797 | 0.799790128 |
| BNP | ASAT_Pos_pPS | 4159 | -0.00567797 | [-0.04 0.02] | 0.715137984 | 0.945970482 |
| BNP | ALP_Neg_pPS | 4159 | 0.019279009 | [-0.01 0.05] | 0.215231772 | 0.767947366 |
| BNP | SHBG-LpA_pPS | 4159 | 0.025226079 | [-0.01 0.06] | 0.104865823 | 0.663720785 |
| BNP | Obesity_pPS | 4159 | 0.004356485 | [-0.03 0.03] | 0.779460631 | 0.964905159 |
| BNP | VAT_Neg_pPS | 4159 | -0.01597856 | [-0.05 0.01] | 0.304366125 | 0.826182442 |
| BNP | Bilirubin_pPS | 4159 | -0.006639297 | [-0.04 0.02] | 0.669556251 | 0.937489141 |
| BNP | Proinsulin_pPS | 4159 | 0.021743401 | [-0.01 0.05] | 0.162185059 | 0.718997225 |
| BNP | Hyper_Insulin_pPS | 4159 | 0.008043905 | [-0.02 0.04] | 0.605125156 | 0.918301271 |
| BNP | Beta_Cell_1_pPS | 4159 | -0.01135334 | [-0.04 0.02] | 0.465525134 | 0.879924083 |
| BNP | Beta_Cell_2_pPS | 4159 | 0.008186363 | [-0.02 0.04] | 0.598748163 | 0.918301271 |
| PAPPA | Cholesterol_pPS | 4159 | -0.005417883 | [-0.04 0.03] | 0.727655114 | 0.946981479 |
| PAPPA | Lipodystrophy_1_pPS | 4159 | -0.018951569 | [-0.05 0.01] | 0.223127253 | 0.780100778 |
| PAPPA | Lipodystrophy_2_pPS | 4159 | -0.014385475 | [-0.04 0.02] | 0.355122297 | 0.847284666 |
| PAPPA | Liver-Lipid_pPS | 4159 | 0.003295349 | [-0.03 0.03] | 0.83225147 | 0.971458352 |
| PAPPA | ASAT_Pos_pPS | 4159 | 0.006718124 | [-0.02 0.04] | 0.665869324 | 0.937489141 |
| PAPPA | ALP_Neg_pPS | 4159 | -0.026603724 | [-0.06 0. ] | 0.087209771 | 0.616770438 |
| PAPPA | SHBG-LpA_pPS | 4159 | -0.021709658 | [-0.05 0.01] | 0.162837807 | 0.718997225 |
| PAPPA | Obesity_pPS | 4159 | -0.034624163 | [-0.07 -0. ] | 0.026001057 | 0.396323801 |

**Table S7: Associations between pPSs and plasma proteins in SCAPIS**

**a) Spearman Correlation**

| Proteomics | PPS | Sample_Size | r | CI95% | p | fdr_p_adjusted |
|---|---|---|---|---|---|---|
| EPHB4 | SHBG-LpA_pPS | 4225 | 0.006604941 | [-0.02 0.04] | 0.668222728 | 0.924640793 |
| EPHB4 | Obesity_pPS | 4225 | 0.012898205 | [-0.02 0.04] | 0.402604797 | 0.81230319 |
| EPHB4 | VAT_Neg_pPS | 4225 | -0.02494199 | [-0.06 0.01] | 0.105512527 | 0.595530825 |
| EPHB4 | Bilirubin_pPS | 4225 | 0.000270692 | [-0.03 0.03] | 0.985986064 | 0.998703927 |
| EPHB4 | Proinsulin_pPS | 4225 | 0.008016937 | [-0.02 0.04] | 0.602914235 | 0.908795882 |
| EPHB4 | Hyper_Insulin_pPS | 4225 | 0.007390004 | [-0.02 0.04] | 0.63156033 | 0.914460114 |
| EPHB4 | Beta_Cell_1_pPS | 4225 | 0.012307395 | [-0.02 0.04] | 0.424500918 | 0.826250025 |
| EPHB4 | Beta_Cell_2_pPS | 4225 | 0.023294701 | [-0.01 0.05] | 0.130594871 | 0.626855379 |
| EPHB4 | Total_GRS | 4225 | 0.006076726 | [-0.02 0.04] | 0.693350357 | 0.930013012 |
| JAM_A | Cholesterol_pPS | 4225 | 0.002650099 | [-0.03 0.03] | 0.863468293 | 0.977006629 |
| JAM_A | Lipodystrophy_1_pPS | 4225 | 0.019194099 | [-0.01 0.05] | 0.212915511 | 0.712633026 |
| JAM_A | Lipodystrophy_2_pPS | 4225 | 0.027198502 | [-0. 0.06] | 0.077531351 | 0.514390694 |
| JAM_A | Liver-Lipid_pPS | 4225 | -0.028444993 | [-0.06 0. ] | 0.064874403 | 0.475856102 |
| JAM_A | ASAT_Pos_pPS | 4225 | 0.008820487 | [-0.02 0.04] | 0.567079382 | 0.899863296 |
| JAM_A | ALP_Neg_pPS | 4225 | -0.011458537 | [-0.04 0.02] | 0.45714867 | 0.835860883 |
| JAM_A | SHBG-LpA_pPS | 4225 | 0.001631399 | [-0.03 0.03] | 0.915694306 | 0.990056084 |
| JAM_A | Obesity_pPS | 4225 | -0.008839531 | [-0.04 0.02] | 0.566242643 | 0.899863296 |
| JAM_A | VAT_Neg_pPS | 4225 | 0.003477468 | [-0.03 0.03] | 0.821475307 | 0.963461584 |
| JAM_A | Bilirubin_pPS | 4225 | 0.003667328 | [-0.03 0.03] | 0.81190613 | 0.962155826 |
| JAM_A | Proinsulin_pPS | 4225 | 0.01392134 | [-0.02 0.04] | 0.366325963 | 0.798890546 |
| JAM_A | Hyper_Insulin_pPS | 4225 | -0.000368707 | [-0.03 0.03] | 0.980912583 | 0.996862516 |
| JAM_A | Beta_Cell_1_pPS | 4225 | 0.00217142 | [-0.03 0.03] | 0.887948901 | 0.979291846 |
| JAM_A | Beta_Cell_2_pPS | 4225 | 0.00340759 | [-0.03 0.03] | 0.825003968 | 0.964449615 |
| JAM_A | Total_GRS | 4225 | 0.017431099 | [-0.01 0.05] | 0.257987152 | 0.740941247 |
| FS | Cholesterol_pPS | 4159 | 0.011635387 | [-0.02 0.04] | 0.453805451 | 0.834640464 |
| FS | Lipodystrophy_1_pPS | 4159 | -0.011842842 | [-0.04 0.02] | 0.445795794 | 0.831350198 |
| FS | Lipodystrophy_2_pPS | 4159 | -0.000351866 | [-0.03 0.03] | 0.98192759 | 0.99746785 |
| FS | Liver-Lipid_pPS | 4159 | -0.092548655 | [-0.12 -0.06] | 2.36E-09 | 5.92E-07 |
| FS | ASAT_Pos_pPS | 4159 | -0.034907491 | [-0.07 -0. ] | 0.024579235 | 0.333340061 |
| FS | ALP_Neg_pPS | 4159 | 0.009014867 | [-0.02 0.04] | 0.561665799 | 0.897748415 |
| FS | SHBG-LpA_pPS | 4159 | 0.016061601 | [-0.01 0.05] | 0.301100116 | 0.76319014 |
| FS | Obesity_pPS | 4159 | 0.004200918 | [-0.03 0.03] | 0.786816386 | 0.957247257 |
| FS | VAT_Neg_pPS | 4159 | -0.014159916 | [-0.04 0.02] | 0.361963632 | 0.7979754 |
| FS | Bilirubin_pPS | 4159 | 0.003220053 | [-0.03 0.03] | 0.835775218 | 0.970026164 |
| FS | Proinsulin_pPS | 4159 | 0.008848973 | [-0.02 0.04] | 0.568889001 | 0.900387238 |
| FS | Hyper_Insulin_pPS | 4159 | -0.001244848 | [-0.03 0.03] | 0.936125457 | 0.99138381 |
| FS | Beta_Cell_1_pPS | 4159 | 0.011312875 | [-0.02 0.04] | 0.466416999 | 0.841780762 |
| FS | Beta_Cell_2_pPS | 4159 | -0.002225006 | [-0.03 0.03] | 0.886099732 | 0.979291846 |
| FS | Total_GRS | 4159 | 0.017337387 | [-0.01 0.05] | 0.264325154 | 0.746520328 |
| PLC | Cholesterol_pPS | 4223 | 0.008250235 | [-0.02 0.04] | 0.592493461 | 0.905925461 |
| PLC | Lipodystrophy_1_pPS | 4223 | 0.021651515 | [-0.01 0.05] | 0.160091956 | 0.663913634 |
| PLC | Lipodystrophy_2_pPS | 4223 | -0.005671823 | [-0.04 0.02] | 0.712909883 | 0.940550324 |
| PLC | Liver-Lipid_pPS | 4223 | 0.0197544 | [-0.01 0.05] | 0.199964984 | 0.700947446 |
| PLC | ASAT_Pos_pPS | 4223 | -0.006370399 | [-0.04 0.02] | 0.679408208 | 0.925579558 |
| PLC | ALP_Neg_pPS | 4223 | 0.006938391 | [-0.02 0.04] | 0.652624394 | 0.919948884 |
| PLC | SHBG-LpA_pPS | 4223 | 0.032756698 | [0. 0.06] | 0.03353707 | 0.381862422 |
| PLC | Obesity_pPS | 4223 | 0.015745788 | [-0.01 0.05] | 0.306999103 | 0.76365449 |
| PLC | VAT_Neg_pPS | 4223 | -0.008333138 | [-0.04 0.02] | 0.588780122 | 0.904476019 |
| PLC | Bilirubin_pPS | 4223 | -0.013600314 | [-0.04 0.02] | 0.377596573 | 0.803170077 |
| PLC | Proinsulin_pPS | 4223 | 0.007850336 | [-0.02 0.04] | 0.610554343 | 0.909483337 |
| PLC | Hyper_Insulin_pPS | 4223 | 0.010433937 | [-0.02 0.04] | 0.498470196 | 0.863780476 |
| PLC | Beta_Cell_1_pPS | 4223 | 0.004761851 | [-0.03 0.03] | 0.757384579 | 0.950403646 |
| PLC | Beta_Cell_2_pPS | 4223 | -0.004974225 | [-0.04 0.03] | 0.74692649 | 0.947292085 |
| PLC | Total_GRS | 4223 | 0.016851639 | [-0.01 0.05] | 0.274265077 | 0.746520328 |
| IGFBP_1 | Cholesterol_pPS | 4225 | 0.002498244 | [-0.03 0.03] | 0.871221396 | 0.977006629 |
| IGFBP_1 | Lipodystrophy_1_pPS | 4225 | -0.023142595 | [-0.05 0.01] | 0.133126472 | 0.629316669 |
| IGFBP_1 | Lipodystrophy_2_pPS | 4225 | -0.014585183 | [-0.04 0.02] | 0.343914513 | 0.790391668 |
| IGFBP_1 | Liver-Lipid_pPS | 4225 | -0.090399593 | [-0.12 -0.06] | 4.15E-09 | 9.56E-07 |
| IGFBP_1 | ASAT_Pos_pPS | 4225 | -0.010752657 | [-0.04 0.02] | 0.485338222 | 0.854965648 |
| IGFBP_1 | ALP_Neg_pPS | 4225 | 0.006919224 | [-0.02 0.04] | 0.653444254 | 0.919948884 |
| IGFBP_1 | SHBG-LpA_pPS | 4225 | 0.006970959 | [-0.02 0.04] | 0.651024322 | 0.919948884 |
| IGFBP_1 | Obesity_pPS | 4225 | -0.053074616 | [-0.08 -0.02] | 0.000568205 | 0.035220685 |
| IGFBP_1 | VAT_Neg_pPS | 4225 | -0.032323731 | [-0.06 -0. ] | 0.035906711 | 0.386599178 |
| IGFBP_1 | Bilirubin_pPS | 4225 | -0.003045772 | [-0.03 0.03] | 0.843330137 | 0.973480208 |
| IGFBP_1 | Proinsulin_pPS | 4225 | 0.012037943 | [-0.02 0.04] | 0.43471382 | 0.828998134 |
| IGFBP_1 | Hyper_Insulin_pPS | 4225 | -0.031629982 | [-0.06 -0. ] | 0.040079226 | 0.407062052 |
| IGFBP_1 | Beta_Cell_1_pPS | 4225 | -0.005138521 | [-0.04 0.03] | 0.738807873 | 0.943595433 |
| IGFBP_1 | Beta_Cell_2_pPS | 4225 | 0.03727978 | [0.01 0.07] | 0.015526265 | 0.258767146 |
| IGFBP_1 | Total_GRS | 4225 | -0.017902074 | [-0.05 0.01] | 0.245345403 | 0.735237038 |
| IGFBP_7 | Cholesterol_pPS | 4225 | 0.023645464 | [-0.01 0.05] | 0.124899304 | 0.621120863 |
| IGFBP_7 | Lipodystrophy_1_pPS | 4225 | 0.01632359 | [-0.01 0.05] | 0.289471526 | 0.753421877 |
| IGFBP_7 | Lipodystrophy_2_pPS | 4225 | 0.013241629 | [-0.02 0.04] | 0.390194155 | 0.81106215 |
| IGFBP_7 | Liver-Lipid_pPS | 4225 | -0.007487817 | [-0.04 0.02] | 0.627052942 | 0.914245177 |
| IGFBP_7 | ASAT_Pos_pPS | 4225 | 0.023218581 | [-0.01 0.05] | 0.13185709 | 0.629316669 |
| IGFBP_7 | ALP_Neg_pPS | 4225 | -0.004047396 | [-0.03 0.03] | 0.792835779 | 0.958013595 |
| IGFBP_7 | SHBG-LpA_pPS | 4225 | 0.019084149 | [-0.01 0.05] | 0.215548494 | 0.715705689 |
| IGFBP_7 | Obesity_pPS | 4225 | -0.01022648 | [-0.04 0.02] | 0.506948334 | 0.867976055 |
| IGFBP_7 | VAT_Neg_pPS | 4225 | -0.007167371 | [-0.04 0.02] | 0.641870628 | 0.916400472 |
| IGFBP_7 | Bilirubin_pPS | 4225 | -0.005867643 | [-0.04 0.02] | 0.703392431 | 0.936951308 |
| IGFBP_7 | Proinsulin_pPS | 4225 | 0.014078371 | [-0.02 0.04] | 0.360944015 | 0.7979754 |
| IGFBP_7 | Hyper_Insulin_pPS | 4225 | 0.011463149 | [-0.02 0.04] | 0.456967551 | 0.835860883 |
| IGFBP_7 | Beta_Cell_1_pPS | 4225 | 0.001117702 | [-0.03 0.03] | 0.942183393 | 0.992903797 |
| IGFBP_7 | Beta_Cell_2_pPS | 4225 | 0.006801742 | [-0.02 0.04] | 0.658953095 | 0.923203321 |
| IGFBP_7 | Total_GRS | 4225 | 0.021037589 | [-0.01 0.05] | 0.172175798 | 0.674627844 |
| MB | Cholesterol_pPS | 4225 | 0.016877893 | [-0.01 0.05] | 0.273404115 | 0.746520328 |
| MB | Lipodystrophy_1_pPS | 4225 | 0.03485546 | [0. 0.06] | 0.023672622 | 0.327897946 |
| MB | Lipodystrophy_2_pPS | 4225 | -0.000115073 | [-0.03 0.03] | 0.994042356 | 0.999464614 |
| MB | Liver-Lipid_pPS | 4225 | 0.018264613 | [-0.01 0.05] | 0.235914769 | 0.726631332 |
| MB | ASAT_Pos_pPS | 4225 | 0.004241468 | [-0.03 0.03] | 0.783144804 | 0.955139045 |
| MB | ALP_Neg_pPS | 4225 | -0.018370181 | [-0.05 0.01] | 0.23321749 | 0.726631332 |
| MB | SHBG-LpA_pPS | 4225 | 0.018133091 | [-0.01 0.05] | 0.239305934 | 0.728373408 |
| MB | Obesity_pPS | 4225 | 0.025918042 | [-0. 0.06] | 0.092556833 | 0.559254737 |
| MB | VAT_Neg_pPS | 4225 | -0.024022079 | [-0.05 0.01] | 0.119001489 | 0.615837655 |
| MB | Bilirubin_pPS | 4225 | 0.000497466 | [-0.03 0.03] | 0.974248973 | 0.996862516 |
| MB | Proinsulin_pPS | 4225 | 0.000837 | [-0.03 0.03] | 0.95668692 | 0.99612031 |
| MB | Hyper_Insulin_pPS | 4225 | 0.003312445 | [-0.03 0.03] | 0.829814305 | 0.966693035 |
| MB | Beta_Cell_1_pPS | 4225 | 0.015782654 | [-0.01 0.05] | 0.305752895 | 0.76365449 |
| MB | Beta_Cell_2_pPS | 4225 | -0.018261902 | [-0.05 0.01] | 0.235984309 | 0.726631332 |
| MB | Total_GRS | 4225 | 0.008436301 | [-0.02 0.04] | 0.584085036 | 0.903823705 |
| MMP_2 | Cholesterol_pPS | 4225 | 0.00307213 | [-0.03 0.03] | 0.841992143 | 0.972342391 |
| MMP_2 | Lipodystrophy_1_pPS | 4225 | -0.029077337 | [-0.06 0. ] | 0.059136135 | 0.459762627 |
| MMP_2 | Lipodystrophy_2_pPS | 4225 | 0.016496801 | [-0.01 0.05] | 0.284383838 | 0.749111537 |
| MMP_2 | Liver-Lipid_pPS | 4225 | 0.025209399 | [-0. 0.06] | 0.101827723 | 0.586731767 |
| MMP_2 | ASAT_Pos_pPS | 4225 | -0.007329943 | [-0.04 0.02] | 0.634334871 | 0.914460114 |
| MMP_2 | ALP_Neg_pPS | 4225 | 0.000745138 | [-0.03 0.03] | 0.961436655 | 0.99612031 |
| MMP_2 | SHBG-LpA_pPS | 4225 | 0.00630312 | [-0.02 0.04] | 0.682537309 | 0.926969309 |
| MMP_2 | Obesity_pPS | 4225 | -0.003587239 | [-0.03 0.03] | 0.81593935 | 0.963461584 |

**b) Partial Correlation**

| Proteomics | PPS | Sample_Size | r | CI95% | p | fdr_p_adjusted |
|---|---|---|---|---|---|---|
| PAPPA | VAT_Neg_pPS | 4159 | -0.016430927 | [-0.05 0.01] | 0.290877842 | 0.81655641 |
| PAPPA | Bilirubin_pPS | 4159 | 0.021436317 | [-0.01 0.05] | 0.168198656 | 0.722132896 |
| PAPPA | Proinsulin_pPS | 4159 | 0.048047328 | [0.02 0.08] | 0.00200082 | 0.099117562 |
| PAPPA | Hyper_Insulin_pPS | 4159 | 0.032664068 | [0. 0.06] | 0.035720016 | 0.449078407 |
| PAPPA | Beta_Cell_1_pPS | 4159 | -0.020294563 | [-0.05 0.01] | 0.192028125 | 0.750920235 |
| PAPPA | Beta_Cell_2_pPS | 4159 | -0.022046047 | [-0.05 0.01] | 0.156418499 | 0.708464493 |
| PDGF_subunit_A | Cholesterol_pPS | 4225 | -0.006273405 | [-0.04 0.02] | 0.684414872 | 0.944323894 |
| PDGF_subunit_A | Lipodystrophy_1_pPS | 4225 | 0.023323243 | [-0.01 0.05] | 0.130718519 | 0.694065456 |
| PDGF_subunit_A | Lipodystrophy_2_pPS | 4225 | 0.005366142 | [-0.02 0.04] | 0.728091968 | 0.946981479 |
| PDGF_subunit_A | Liver-Lipid_pPS | 4225 | -0.013499898 | [-0.04 0.02] | 0.381751955 | 0.85037326 |
| PDGF_subunit_A | ASAT_Pos_pPS | 4225 | -0.002245562 | [-0.03 0.03] | 0.884327255 | 0.979375326 |
| PDGF_subunit_A | ALP_Neg_pPS | 4225 | 0.003947918 | [-0.03 0.03] | 0.798121354 | 0.967638637 |
| PDGF_subunit_A | SHBG-LpA_pPS | 4225 | -0.012904288 | [-0.04 0.02] | 0.403110267 | 0.85241904 |
| PDGF_subunit_A | Obesity_pPS | 4225 | -0.004781319 | [-0.04 0.03] | 0.756732509 | 0.95462436 |
| PDGF_subunit_A | VAT_Neg_pPS | 4225 | -0.016706403 | [-0.05 0.01] | 0.279051766 | 0.808820203 |
| PDGF_subunit_A | Bilirubin_pPS | 4225 | 0.003680027 | [-0.03 0.03] | 0.811553162 | 0.967638637 |
| PDGF_subunit_A | Proinsulin_pPS | 4225 | 0.000581502 | [-0.03 0.03] | 0.969947273 | 0.996924797 |
| PDGF_subunit_A | Hyper_Insulin_pPS | 4225 | -0.023163805 | [-0.05 0.01] | 0.133370912 | 0.694065456 |
| PDGF_subunit_A | Beta_Cell_1_pPS | 4225 | -0.012563561 | [-0.04 0.02] | 0.415644059 | 0.854508456 |
| PDGF_subunit_A | Beta_Cell_2_pPS | 4225 | 0.00471995 | [-0.03 0.03] | 0.759758172 | 0.956884516 |
| PDGF_subunit_B | Cholesterol_pPS | 4159 | -0.008371061 | [-0.04 0.02] | 0.590525959 | 0.914729327 |
| PDGF_subunit_B | Lipodystrophy_1_pPS | 4159 | 0.014832352 | [-0.02 0.05] | 0.34037339 | 0.847042484 |
| PDGF_subunit_B | Lipodystrophy_2_pPS | 4159 | -0.00269501 | [-0.03 0.03] | 0.862473153 | 0.97556306 |
| PDGF_subunit_B | Liver-Lipid_pPS | 4159 | -0.017330146 | [-0.05 0.01] | 0.265275302 | 0.80722329 |
| PDGF_subunit_B | ASAT_Pos_pPS | 4159 | -0.001688548 | [-0.03 0.03] | 0.91357176 | 0.989461462 |
| PDGF_subunit_B | ALP_Neg_pPS | 4159 | 0.008069288 | [-0.02 0.04] | 0.603986703 | 0.918301271 |
| PDGF_subunit_B | SHBG-LpA_pPS | 4159 | -0.004399361 | [-0.03 0.03] | 0.777347031 | 0.964867039 |
| PDGF_subunit_B | Obesity_pPS | 4159 | -0.01543385 | [-0.05 0.02] | 0.321150344 | 0.83927892 |
| PDGF_subunit_B | VAT_Neg_pPS | 4159 | -0.018087502 | [-0.05 0.01] | 0.244950896 | 0.79432283 |
| PDGF_subunit_B | Bilirubin_pPS | 4159 | -0.003218927 | [-0.03 0.03] | 0.83608586 | 0.972099644 |
| PDGF_subunit_B | Proinsulin_pPS | 4159 | 0.005801084 | [-0.02 0.04] | 0.709239302 | 0.945970482 |
| PDGF_subunit_B | Hyper_Insulin_pPS | 4159 | -0.028218697 | [-0.06 0. ] | 0.069651984 | 0.573238056 |
| PDGF_subunit_B | Beta_Cell_1_pPS | 4159 | -0.009969369 | [-0.04 0.02] | 0.521643195 | 0.895803896 |
| PDGF_subunit_B | Beta_Cell_2_pPS | 4159 | -0.007936972 | [-0.04 0.02] | 0.609931848 | 0.919490897 |
| PECAM_1 | Cholesterol_pPS | 4225 | -0.002783746 | [-0.03 0.03] | 0.856874664 | 0.97556306 |
| PECAM_1 | Lipodystrophy_1_pPS | 4225 | 0.018525024 | [-0.01 0.05] | 0.230021148 | 0.786988264 |
| PECAM_1 | Lipodystrophy_2_pPS | 4225 | 0.029195814 | [-0. 0.06] | 0.058498301 | 0.558815902 |
| PECAM_1 | Liver-Lipid_pPS | 4225 | -0.010253105 | [-0.04 0.02] | 0.506500342 | 0.895803896 |
| PECAM_1 | ASAT_Pos_pPS | 4225 | 0.001450357 | [-0.03 0.03] | 0.925136231 | 0.990503296 |
| PECAM_1 | ALP_Neg_pPS | 4225 | -0.114691946 | [-0.14 -0.08] | 8.97E-14 | 3.30E-11 |
| PECAM_1 | SHBG-LpA_pPS | 4225 | -0.006662117 | [-0.04 0.02] | 0.666009772 | 0.937489141 |
| PECAM_1 | Obesity_pPS | 4225 | -0.006029927 | [-0.04 0.02] | 0.696040326 | 0.945970482 |
| PECAM_1 | VAT_Neg_pPS | 4225 | -0.001159109 | [-0.03 0.03] | 0.940137936 | 0.994985753 |
| PECAM_1 | Bilirubin_pPS | 4225 | 0.003039529 | [-0.03 0.03] | 0.843885593 | 0.972178273 |
| PECAM_1 | Proinsulin_pPS | 4225 | 0.021428984 | [-0.01 0.05] | 0.16498395 | 0.720336704 |
| PECAM_1 | Hyper_Insulin_pPS | 4225 | 0.004893269 | [-0.03 0.04] | 0.751222665 | 0.954135344 |
| PECAM_1 | Beta_Cell_1_pPS | 4225 | -0.008990183 | [-0.04 0.02] | 0.560250955 | 0.902076574 |
| PECAM_1 | Beta_Cell_2_pPS | 4225 | 0.024952461 | [-0.01 0.06] | 0.105905331 | 0.665922758 |
| PGF | Cholesterol_pPS | 4159 | 0.02049627 | [-0.01 0.05] | 0.187646988 | 0.745954693 |
| PGF | Lipodystrophy_1_pPS | 4159 | 0.021147978 | [-0.01 0.05] | 0.173996043 | 0.730702407 |
| PGF | Lipodystrophy_2_pPS | 4159 | 0.003862292 | [-0.03 0.03] | 0.803935978 | 0.967638637 |
| PGF | Liver-Lipid_pPS | 4159 | 0.014129555 | [-0.02 0.04] | 0.363747661 | 0.847284666 |
| PGF | ASAT_Pos_pPS | 4159 | -0.008493662 | [-0.04 0.02] | 0.585097007 | 0.911131843 |
| PGF | ALP_Neg_pPS | 4159 | 0.007241478 | [-0.02 0.04] | 0.641598688 | 0.933976497 |
| PGF | SHBG-LpA_pPS | 4159 | 0.016241313 | [-0.01 0.05] | 0.296481862 | 0.817900851 |
| PGF | Obesity_pPS | 4159 | 0.010208841 | [-0.02 0.04] | 0.511690182 | 0.895803896 |
| PGF | VAT_Neg_pPS | 4159 | -0.004568679 | [-0.04 0.03] | 0.769016714 | 0.960245786 |
| PGF | Bilirubin_pPS | 4159 | 0.008932732 | [-0.02 0.04] | 0.56584708 | 0.90516325 |
| PGF | Proinsulin_pPS | 4159 | 0.001406112 | [-0.03 0.03] | 0.927984995 | 0.991797911 |
| PGF | Hyper_Insulin_pPS | 4159 | 0.011920518 | [-0.02 0.04] | 0.443533921 | 0.867534836 |
| PGF | Beta_Cell_1_pPS | 4159 | -0.00392336 | [-0.03 0.03] | 0.800900546 | 0.967638637 |
| PGF | Beta_Cell_2_pPS | 4159 | -0.006230535 | [-0.04 0.02] | 0.688801255 | 0.944989936 |
| t_PA | Cholesterol_pPS | 4225 | 0.009498011 | [-0.02 0.04] | 0.538309004 | 0.899241939 |
| t_PA | Lipodystrophy_1_pPS | 4225 | 0.046222051 | [0.02 0.08] | 0.002733303 | 0.119687959 |
| t_PA | Lipodystrophy_2_pPS | 4225 | 0.015681865 | [-0.01 0.05] | 0.309600368 | 0.830335001 |
| t_PA | Liver-Lipid_pPS | 4225 | 0.006941403 | [-0.02 0.04] | 0.652908144 | 0.936605382 |
| t_PA | ASAT_Pos_pPS | 4225 | -0.004023387 | [-0.03 0.03] | 0.794347969 | 0.967638637 |
| t_PA | ALP_Neg_pPS | 4225 | 0.004670945 | [-0.03 0.03] | 0.762176873 | 0.958675598 |
| t_PA | SHBG-LpA_pPS | 4225 | -0.000264784 | [-0.03 0.03] | 0.986313078 | 0.998803911 |
| t_PA | Obesity_pPS | 4225 | 0.036830932 | [0.01 0.07] | 0.016985726 | 0.33440804 |
| t_PA | VAT_Neg_pPS | 4225 | 0.024264843 | [-0.01 0.05] | 0.115879839 | 0.681046037 |
| t_PA | Bilirubin_pPS | 4225 | 0.00431233 | [-0.03 0.03] | 0.779946261 | 0.964905159 |
| t_PA | Proinsulin_pPS | 4225 | 0.002808546 | [-0.03 0.03] | 0.855613511 | 0.97556306 |
| t_PA | Hyper_Insulin_pPS | 4225 | -0.005194421 | [-0.04 0.03] | 0.736464438 | 0.949205935 |
| t_PA | Beta_Cell_1_pPS | 4225 | 0.015852861 | [-0.01 0.05] | 0.304353831 | 0.826182442 |
| t_PA | Beta_Cell_2_pPS | 4225 | 0.007674595 | [-0.02 0.04] | 0.619026963 | 0.92152711 |
| PRTN3 | Cholesterol_pPS | 3944 | 0.000271173 | [-0.03 0.03] | 0.986460144 | 0.998803911 |
| PRTN3 | Lipodystrophy_1_pPS | 3944 | 0.000844544 | [-0.03 0.03] | 0.957848861 | 0.996425456 |
| PRTN3 | Lipodystrophy_2_pPS | 3944 | 0.000619128 | [-0.03 0.03] | 0.969092683 | 0.996924797 |
| PRTN3 | Liver-Lipid_pPS | 3944 | 0.014845475 | [-0.02 0.05] | 0.352832116 | 0.847284666 |
| PRTN3 | ASAT_Pos_pPS | 3944 | 0.026604915 | [-0. 0.06] | 0.095856484 | 0.643037244 |
| PRTN3 | ALP_Neg_pPS | 3944 | 0.003352678 | [-0.03 0.03] | 0.833809944 | 0.971458352 |
| PRTN3 | SHBG-LpA_pPS | 3944 | -0.008788934 | [-0.04 0.02] | 0.58229246 | 0.909633339 |
| PRTN3 | Obesity_pPS | 3944 | 0.010583678 | [-0.02 0.04] | 0.507736879 | 0.895803896 |
| PRTN3 | VAT_Neg_pPS | 3944 | -0.009928017 | [-0.04 0.02] | 0.534382551 | 0.898959051 |
| PRTN3 | Bilirubin_pPS | 3944 | 0.020199564 | [-0.01 0.05] | 0.206137795 | 0.765145476 |
| PRTN3 | Proinsulin_pPS | 3944 | 0.010266207 | [-0.02 0.04] | 0.520551071 | 0.895803896 |
| PRTN3 | Hyper_Insulin_pPS | 3944 | -0.021761997 | [-0.05 0.01] | 0.17317597 | 0.72956083 |
| PRTN3 | Beta_Cell_1_pPS | 3944 | 0.018847347 | [-0.01 0.05] | 0.238155998 | 0.790085251 |
| PRTN3 | Beta_Cell_2_pPS | 3944 | -0.026298259 | [-0.06 0.01] | 0.099747108 | 0.651687563 |
| REN | Cholesterol_pPS | 4158 | 0.012503485 | [-0.02 0.04] | 0.421617925 | 0.860518922 |
| REN | Lipodystrophy_1_pPS | 4158 | 0.005335408 | [-0.03 0.04] | 0.731670997 | 0.947604067 |
| REN | Lipodystrophy_2_pPS | 4158 | 0.000378821 | [-0.03 0.03] | 0.980576296 | 0.998803911 |
| REN | Liver-Lipid_pPS | 4158 | -0.021792544 | [-0.05 0.01] | 0.161288667 | 0.718997225 |
| REN | ASAT_Pos_pPS | 4158 | 0.015315243 | [-0.02 0.05] | 0.324942199 | 0.842951766 |
| REN | ALP_Neg_pPS | 4158 | 0.009608217 | [-0.02 0.04] | 0.536888867 | 0.898959051 |
| REN | SHBG-LpA_pPS | 4158 | -0.002368201 | [-0.03 0.03] | 0.879027571 | 0.97781424 |
| REN | Obesity_pPS | 4158 | 0.007843652 | [-0.02 0.04] | 0.614183251 | 0.921048876 |
| REN | VAT_Neg_pPS | 4158 | 0.019700698 | [-0.01 0.05] | 0.205417739 | 0.765145476 |
| REN | Bilirubin_pPS | 4158 | 0.022158515 | [-0.01 0.05] | 0.154365511 | 0.708464493 |
| REN | Proinsulin_pPS | 4158 | 0.000114258 | [-0.03 0.03] | 0.994141005 | 0.998803911 |
| REN | Hyper_Insulin_pPS | 4158 | 0.004574937 | [-0.03 0.04] | 0.768736501 | 0.960245786 |
| REN | Beta_Cell_1_pPS | 4158 | -0.013177973 | [-0.04 0.02] | 0.39701101 | 0.85037326 |
| REN | Beta_Cell_2_pPS | 4158 | 0.006588168 | [-0.02 0.04] | 0.671989329 | 0.939253669 |
| PSGL_1 | Cholesterol_pPS | 4159 | -0.000107187 | [-0.03 0.03] | 0.994502913 | 0.998803911 |
| PSGL_1 | Lipodystrophy_1_pPS | 4159 | 0.033054243 | [0. 0.06] | 0.03356994 | 0.436493301 |
| PSGL_1 | Lipodystrophy_2_pPS | 4159 | 0.01950024 | [-0.01 0.05] | 0.21001246 | 0.767305935 |

**Table S7: Associations between pPSs and plasma proteins in SCAPIS**

**a) Spearman Correlation**

| Proteomics | PPS | Sample_Size | r | CI95% | p | fdr_p_adjusted |
|---|---|---|---|---|---|---|
| MMP_2 | VAT_Neg_pPS | 4225 | -0.01478379 | [-0.04 0.02] | 0.337383263 | 0.782502359 |
| MMP_2 | Bilirubin_pPS | 4225 | -0.021747585 | [-0.05 0.01] | 0.158146706 | 0.659342762 |
| MMP_2 | Proinsulin_pPS | 4225 | 0.02438988 | [-0.01 0.05] | 0.113454547 | 0.604978148 |
| MMP_2 | Hyper_Insulin_pPS | 4225 | 0.015535205 | [-0.01 0.05] | 0.313399299 | 0.76365449 |
| MMP_2 | Beta_Cell_1_pPS | 4225 | 0.003877189 | [-0.03 0.03] | 0.801361541 | 0.958074488 |
| MMP_2 | Beta_Cell_2_pPS | 4225 | 0.029826246 | [-0. 0.06] | 0.052891345 | 0.441027526 |
| MMP_2 | Total_GRS | 4225 | 0.016597157 | [-0.01 0.05] | 0.281463914 | 0.748127084 |
| BNP | Cholesterol_pPS | 4159 | 0.01289831 | [-0.02 0.04] | 0.406313105 | 0.813210382 |
| BNP | Lipodystrophy_1_pPS | 4159 | -0.023082832 | [-0.05 0.01] | 0.137221012 | 0.629316669 |
| BNP | Lipodystrophy_2_pPS | 4159 | 0.000102907 | [-0.03 0.03] | 0.994714132 | 0.999464614 |
| BNP | Liver-Lipid_pPS | 4159 | 0.019531394 | [-0.01 0.05] | 0.208569753 | 0.709689105 |
| BNP | ASAT_Pos_pPS | 4159 | -0.009731813 | [-0.04 0.02] | 0.530969039 | 0.877412567 |
| BNP | ALP_Neg_pPS | 4159 | 0.023270822 | [-0.01 0.05] | 0.134048317 | 0.629316669 |
| BNP | SHBG-LpA_pPS | 4159 | 0.023158899 | [-0.01 0.05] | 0.135930315 | 0.629316669 |
| BNP | Obesity_pPS | 4159 | 0.008011377 | [-0.02 0.04] | 0.606019927 | 0.909483337 |
| BNP | VAT_Neg_pPS | 4159 | -0.014840657 | [-0.05 0.02] | 0.339344448 | 0.785730433 |
| BNP | Bilirubin_pPS | 4159 | -0.008050395 | [-0.04 0.02] | 0.604266367 | 0.908869305 |
| BNP | Proinsulin_pPS | 4159 | 0.026237122 | [-0. 0.06] | 0.091146871 | 0.55533193 |
| BNP | Hyper_Insulin_pPS | 4159 | 0.005773439 | [-0.02 0.04] | 0.710127879 | 0.938946716 |
| BNP | Beta_Cell_1_pPS | 4159 | -0.005393379 | [-0.04 0.03] | 0.72842879 | 0.942134248 |
| BNP | Beta_Cell_2_pPS | 4159 | 0.00985606 | [-0.02 0.04] | 0.52573711 | 0.873942847 |
| BNP | Total_GRS | 4159 | 0.006471155 | [-0.02 0.04] | 0.676968814 | 0.925579558 |
| PAPPA | Cholesterol_pPS | 4159 | -0.006791025 | [-0.04 0.02] | 0.661969047 | 0.924111434 |
| PAPPA | Lipodystrophy_1_pPS | 4159 | -0.018857883 | [-0.05 0.01] | 0.224695485 | 0.724402982 |
| PAPPA | Lipodystrophy_2_pPS | 4159 | -0.014620794 | [-0.05 0.02] | 0.346548725 | 0.790391668 |
| PAPPA | Liver-Lipid_pPS | 4159 | 0.002082872 | [-0.03 0.03] | 0.893330738 | 0.981916701 |
| PAPPA | ASAT_Pos_pPS | 4159 | 0.003697666 | [-0.03 0.03] | 0.811843573 | 0.962155826 |
| PAPPA | ALP_Neg_pPS | 4159 | -0.01831757 | [-0.05 0.01] | 0.238261361 | 0.726631332 |
| PAPPA | SHBG-LpA_pPS | 4159 | -0.021313538 | [-0.05 0.01] | 0.169978048 | 0.674050881 |
| PAPPA | Obesity_pPS | 4159 | -0.028548497 | [-0.06 0. ] | 0.066023533 | 0.476590869 |
| PAPPA | VAT_Neg_pPS | 4159 | -0.019905693 | [-0.05 0.01] | 0.199978947 | 0.700947446 |
| PAPPA | Bilirubin_pPS | 4159 | 0.020425594 | [-0.01 0.05] | 0.188479049 | 0.690839542 |
| PAPPA | Proinsulin_pPS | 4159 | 0.03126722 | [0. 0.06] | 0.044071849 | 0.424281318 |
| PAPPA | Hyper_Insulin_pPS | 4159 | 0.025329578 | [-0.01 0.06] | 0.102906372 | 0.586842904 |
| PAPPA | Beta_Cell_1_pPS | 4159 | -0.014216576 | [-0.04 0.02] | 0.36004571 | 0.7979754 |
| PAPPA | Beta_Cell_2_pPS | 4159 | -0.021257198 | [-0.05 0.01] | 0.171109921 | 0.674627844 |
| PAPPA | Total_GRS | 4159 | -0.022730193 | [-0.05 0.01] | 0.143328241 | 0.641415816 |
| PDGF_subunit_A | Cholesterol_pPS | 4225 | -0.006229327 | [-0.04 0.02] | 0.68605479 | 0.926969309 |
| PDGF_subunit_A | Lipodystrophy_1_pPS | 4225 | 0.018967017 | [-0.01 0.05] | 0.218379171 | 0.718175854 |
| PDGF_subunit_A | Lipodystrophy_2_pPS | 4225 | 0.002480642 | [-0.03 0.03] | 0.872120918 | 0.977006629 |
| PDGF_subunit_A | Liver-Lipid_pPS | 4225 | -0.014843858 | [-0.05 0.02] | 0.335423696 | 0.781240001 |
| PDGF_subunit_A | ASAT_Pos_pPS | 4225 | -0.000416047 | [-0.03 0.03] | 0.978462423 | 0.996862516 |
| PDGF_subunit_A | ALP_Neg_pPS | 4225 | 0.003673125 | [-0.03 0.03] | 0.811614369 | 0.962155826 |
| PDGF_subunit_A | SHBG-LpA_pPS | 4225 | -0.012831121 | [-0.04 0.02] | 0.405056412 | 0.812467802 |
| PDGF_subunit_A | Obesity_pPS | 4225 | -0.006152402 | [-0.04 0.02] | 0.689728835 | 0.927705451 |
| PDGF_subunit_A | VAT_Neg_pPS | 4225 | -0.019832405 | [-0.05 0.01] | 0.198087694 | 0.700925686 |
| PDGF_subunit_A | Bilirubin_pPS | 4225 | 0.004103283 | [-0.03 0.03] | 0.790041717 | 0.957247257 |
| PDGF_subunit_A | Proinsulin_pPS | 4225 | -0.003962359 | [-0.03 0.03] | 0.797092308 | 0.958074488 |
| PDGF_subunit_A | Hyper_Insulin_pPS | 4225 | -0.02308744 | [-0.05 0.01] | 0.134053775 | 0.629316669 |
| PDGF_subunit_A | Beta_Cell_1_pPS | 4225 | -0.014268848 | [-0.04 0.02] | 0.354482685 | 0.797369366 |
| PDGF_subunit_A | Beta_Cell_2_pPS | 4225 | -0.000878149 | [-0.03 0.03] | 0.954559805 | 0.99612031 |
| PDGF_subunit_A | Total_GRS | 4225 | -0.010227199 | [-0.04 0.02] | 0.506918459 | 0.867976055 |
| PDGF_subunit_B | Cholesterol_pPS | 4159 | -0.010096028 | [-0.04 0.02] | 0.515707366 | 0.870853148 |
| PDGF_subunit_B | Lipodystrophy_1_pPS | 4159 | 0.009316942 | [-0.02 0.04] | 0.548628087 | 0.891638418 |
| PDGF_subunit_B | Lipodystrophy_2_pPS | 4159 | -0.006762417 | [-0.04 0.02] | 0.663305182 | 0.924111434 |
| PDGF_subunit_B | Liver-Lipid_pPS | 4159 | -0.019622722 | [-0.05 0.01] | 0.206449359 | 0.709139138 |
| PDGF_subunit_B | ASAT_Pos_pPS | 4159 | -0.002958897 | [-0.03 0.03] | 0.848926991 | 0.97528582 |
| PDGF_subunit_B | ALP_Neg_pPS | 4159 | 0.007892107 | [-0.02 0.04] | 0.611394207 | 0.909483337 |
| PDGF_subunit_B | SHBG-LpA_pPS | 4159 | -0.004953983 | [-0.04 0.03] | 0.749780793 | 0.948757065 |
| PDGF_subunit_B | Obesity_pPS | 4159 | -0.017355706 | [-0.05 0.01] | 0.263820707 | 0.746520328 |
| PDGF_subunit_B | VAT_Neg_pPS | 4159 | -0.022776603 | [-0.05 0.01] | 0.142512771 | 0.640622936 |
| PDGF_subunit_B | Bilirubin_pPS | 4159 | -0.003219866 | [-0.03 0.03] | 0.835784636 | 0.970026164 |
| PDGF_subunit_B | Proinsulin_pPS | 4159 | -0.000593922 | [-0.03 0.03] | 0.969500028 | 0.996862516 |
| PDGF_subunit_B | Hyper_Insulin_pPS | 4159 | -0.030748627 | [-0.06 -0. ] | 0.047703881 | 0.436063541 |
| PDGF_subunit_B | Beta_Cell_1_pPS | 4159 | -0.013909859 | [-0.04 0.02] | 0.370504191 | 0.799802236 |
| PDGF_subunit_B | Beta_Cell_2_pPS | 4159 | -0.013064188 | [-0.04 0.02] | 0.40030343 | 0.81230319 |
| PDGF_subunit_B | Total_GRS | 4159 | -0.022479443 | [-0.05 0.01] | 0.14779619 | 0.645439059 |
| PECAM_1 | Cholesterol_pPS | 4225 | -0.002035539 | [-0.03 0.03] | 0.894918711 | 0.982879285 |
| PECAM_1 | Lipodystrophy_1_pPS | 4225 | 0.018194932 | [-0.01 0.05] | 0.237707156 | 0.726631332 |
| PECAM_1 | Lipodystrophy_2_pPS | 4225 | 0.030694919 | [0. 0.06] | 0.046347741 | 0.434802524 |
| PECAM_1 | Liver-Lipid_pPS | 4225 | -0.012952603 | [-0.04 0.02] | 0.400623323 | 0.81230319 |
| PECAM_1 | ASAT_Pos_pPS | 4225 | 0.007220506 | [-0.02 0.04] | 0.639403522 | 0.91627919 |
| PECAM_1 | ALP_Neg_pPS | 4225 | -0.112150734 | [-0.14 -0.08] | 2.87E-13 | 9.90E-11 |
| PECAM_1 | SHBG-LpA_pPS | 4225 | -0.000462258 | [-0.03 0.03] | 0.976070929 | 0.996862516 |
| PECAM_1 | Obesity_pPS | 4225 | -0.004333154 | [-0.03 0.03] | 0.778577995 | 0.953637204 |
| PECAM_1 | VAT_Neg_pPS | 4225 | 0.004493692 | [-0.03 0.03] | 0.770600286 | 0.95301975 |
| PECAM_1 | Bilirubin_pPS | 4225 | 0.004202379 | [-0.03 0.03] | 0.785094043 | 0.956248702 |
| PECAM_1 | Proinsulin_pPS | 4225 | 0.004368539 | [-0.03 0.03] | 0.776817586 | 0.953637204 |
| PECAM_1 | Hyper_Insulin_pPS | 4225 | 0.006073507 | [-0.02 0.04] | 0.693504554 | 0.930013012 |
| PECAM_1 | Beta_Cell_1_pPS | 4225 | 0.00273578 | [-0.03 0.03] | 0.859099473 | 0.977006629 |
| PECAM_1 | Beta_Cell_2_pPS | 4225 | 0.025890698 | [-0. 0.06] | 0.092901537 | 0.559254737 |
| PECAM_1 | Total_GRS | 4225 | 0.021003644 | [-0.01 0.05] | 0.172869113 | 0.674627844 |
| PGF | Cholesterol_pPS | 4159 | 0.022595626 | [-0.01 0.05] | 0.145712941 | 0.641415816 |
| PGF | Lipodystrophy_1_pPS | 4159 | 0.023756137 | [-0.01 0.05] | 0.126119853 | 0.623533826 |
| PGF | Lipodystrophy_2_pPS | 4159 | 0.006254343 | [-0.02 0.04] | 0.687209617 | 0.926969309 |
| PGF | Liver-Lipid_pPS | 4159 | 0.015431867 | [-0.02 0.05] | 0.320452381 | 0.770109076 |
| PGF | ASAT_Pos_pPS | 4159 | -0.002960966 | [-0.03 0.03] | 0.848822647 | 0.97528582 |
| PGF | ALP_Neg_pPS | 4159 | 0.008512986 | [-0.02 0.04] | 0.583652643 | 0.903823705 |
| PGF | SHBG-LpA_pPS | 4159 | 0.017046994 | [-0.01 0.05] | 0.272410828 | 0.746520328 |
| PGF | Obesity_pPS | 4159 | 0.013884069 | [-0.02 0.04] | 0.371392119 | 0.799802236 |
| PGF | VAT_Neg_pPS | 4159 | -0.002818026 | [-0.03 0.03] | 0.856038989 | 0.977006629 |
| PGF | Bilirubin_pPS | 4159 | 0.008511094 | [-0.02 0.04] | 0.583736279 | 0.903823705 |
| PGF | Proinsulin_pPS | 4159 | 0.003121395 | [-0.03 0.03] | 0.840738464 | 0.972342391 |
| PGF | Hyper_Insulin_pPS | 4159 | 0.015951454 | [-0.01 0.05] | 0.304427673 | 0.76365449 |
| PGF | Beta_Cell_1_pPS | 4159 | -0.005020612 | [-0.04 0.03] | 0.746530196 | 0.947292085 |
| PGF | Beta_Cell_2_pPS | 4159 | -0.006238402 | [-0.04 0.02] | 0.687964852 | 0.926969309 |
| PGF | Total_GRS | 4159 | 0.027103781 | [-0. 0.06] | 0.080949693 | 0.5244628 |
| t_PA | Cholesterol_pPS | 4225 | 0.012983264 | [-0.02 0.04] | 0.399509093 | 0.81230319 |
| t_PA | Lipodystrophy_1_pPS | 4225 | 0.04995811 | [0.02 0.08] | 0.001179773 | 0.052518946 |
| t_PA | Lipodystrophy_2_pPS | 4225 | 0.021353232 | [-0.01 0.05] | 0.165828044 | 0.664911429 |
| t_PA | Liver-Lipid_pPS | 4225 | 0.009687727 | [-0.02 0.04] | 0.529587694 | 0.876821856 |
| t_PA | ASAT_Pos_pPS | 4225 | 0.007849033 | [-0.02 0.04] | 0.610528821 | 0.909483337 |
| t_PA | ALP_Neg_pPS | 4225 | 0.007406887 | [-0.02 0.04] | 0.630781363 | 0.914460114 |
| t_PA | SHBG-LpA_pPS | 4225 | 0.004636057 | [-0.03 0.03] | 0.76354582 | 0.951648335 |
| t_PA | Obesity_pPS | 4225 | 0.040714597 | [0.01 0.07] | 0.008217514 | 0.197592038 |
| t_PA | VAT_Neg_pPS | 4225 | 0.030525772 | [0. 0.06] | 0.047565581 | 0.436063541 |
| t_PA | Bilirubin_pPS | 4225 | 0.006881448 | [-0.02 0.04] | 0.65521357 | 0.92076856 |

**b) Partial Correlation**

| Proteomics | PPS | Sample_Size | r | CI95% | p | fdr_p_adjusted |
|---|---|---|---|---|---|---|
| PSGL_1 | Liver-Lipid_pPS | 4159 | -0.035907026 | [-0.07 -0.01] | 0.020958501 | 0.354719791 |
| PSGL_1 | ASAT_Pos_pPS | 4159 | -0.02488946 | [-0.06 0.01] | 0.10958498 | 0.676681438 |
| PSGL_1 | ALP_Neg_pPS | 4159 | -0.018567754 | [-0.05 0.01] | 0.232643199 | 0.786988264 |
| PSGL_1 | SHBG-LpA_pPS | 4159 | 0.022769924 | [-0.01 0.05] | 0.143257801 | 0.697681852 |
| PSGL_1 | Obesity_pPS | 4159 | 0.031085796 | [0. 0.06] | 0.045654169 | 0.512674866 |
| PSGL_1 | VAT_Neg_pPS | 4159 | 0.011503054 | [-0.02 0.04] | 0.459662128 | 0.874172623 |
| PSGL_1 | Bilirubin_pPS | 4159 | 0.009602729 | [-0.02 0.04] | 0.537072197 | 0.898959051 |
| PSGL_1 | Proinsulin_pPS | 4159 | -0.001565578 | [-0.03 0.03] | 0.919843919 | 0.99041009 |
| PSGL_1 | Hyper_Insulin_pPS | 4159 | -0.021492174 | [-0.05 0.01] | 0.167092543 | 0.722132896 |
| PSGL_1 | Beta_Cell_1_pPS | 4159 | -0.008924205 | [-0.04 0.02] | 0.566218028 | 0.90516325 |
| PSGL_1 | Beta_Cell_2_pPS | 4159 | 0.00750045 | [-0.02 0.04] | 0.629726873 | 0.929613998 |
| TIE2 | Cholesterol_pPS | 4159 | 0.01630712 | [-0.01 0.05] | 0.294528832 | 0.817571414 |
| TIE2 | Lipodystrophy_1_pPS | 4159 | -0.012161272 | [-0.04 0.02] | 0.434381571 | 0.864182741 |
| TIE2 | Lipodystrophy_2_pPS | 4159 | 0.033985202 | [0. 0.06] | 0.028882295 | 0.414433192 |
| TIE2 | Liver-Lipid_pPS | 4159 | -0.023211987 | [-0.05 0.01] | 0.135648807 | 0.694065456 |
| TIE2 | ASAT_Pos_pPS | 4159 | -0.029113988 | [-0.06 0. ] | 0.061241322 | 0.558815902 |
| TIE2 | ALP_Neg_pPS | 4159 | 0.079966746 | [0.05 0.11] | 2.63E-07 | 4.52E-05 |
| TIE2 | SHBG-LpA_pPS | 4159 | -0.017508584 | [-0.05 0.01] | 0.260385251 | 0.807171938 |
| TIE2 | Obesity_pPS | 4159 | -0.005883973 | [-0.04 0.02] | 0.705277617 | 0.945970482 |
| TIE2 | VAT_Neg_pPS | 4159 | 0.018226412 | [-0.01 0.05] | 0.24134493 | 0.790972698 |
| TIE2 | Bilirubin_pPS | 4159 | 0.000573227 | [-0.03 0.03] | 0.970608459 | 0.996924797 |
| TIE2 | Proinsulin_pPS | 4159 | 0.036961556 | [0.01 0.07] | 0.017473946 | 0.33440804 |
| TIE2 | Hyper_Insulin_pPS | 4159 | -0.022461397 | [-0.05 0.01] | 0.148759321 | 0.703126624 |
| TIE2 | Beta_Cell_1_pPS | 4159 | -0.0135441 | [-0.04 0.02] | 0.383966792 | 0.85037326 |
| TIE2 | Beta_Cell_2_pPS | 4159 | -0.014488081 | [-0.04 0.02] | 0.351700699 | 0.847284666 |
| TFPI | Cholesterol_pPS | 4225 | -0.069046891 | [-0.1 -0.04] | 7.51E-06 | 0.00107443 |
| TFPI | Lipodystrophy_1_pPS | 4225 | 0.022200853 | [-0.01 0.05] | 0.150284085 | 0.706166997 |
| TFPI | Lipodystrophy_2_pPS | 4225 | -0.011837736 | [-0.04 0.02] | 0.443098475 | 0.867534836 |
| TFPI | Liver-Lipid_pPS | 4225 | -0.033505342 | [-0.06 -0. ] | 0.029904149 | 0.416452728 |
| TFPI | ASAT_Pos_pPS | 4225 | 0.024564146 | [-0.01 0.05] | 0.111451033 | 0.680721977 |
| TFPI | ALP_Neg_pPS | 4225 | 0.016265271 | [-0.01 0.05] | 0.291944275 | 0.81655641 |
| TFPI | SHBG-LpA_pPS | 4225 | -0.008450246 | [-0.04 0.02] | 0.584045374 | 0.91071482 |
| TFPI | Obesity_pPS | 4225 | -0.00572128 | [-0.04 0.02] | 0.710880348 | 0.945970482 |
| TFPI | VAT_Neg_pPS | 4225 | -0.00267618 | [-0.03 0.03] | 0.862348856 | 0.97556306 |
| TFPI | Bilirubin_pPS | 4225 | -0.015737819 | [-0.05 0.01] | 0.30787702 | 0.830335001 |
| TFPI | Proinsulin_pPS | 4225 | 0.016784342 | [-0.01 0.05] | 0.276814779 | 0.808820203 |
| TFPI | Hyper_Insulin_pPS | 4225 | -0.015684093 | [-0.05 0.01] | 0.309531606 | 0.830335001 |
| TFPI | Beta_Cell_1_pPS | 4225 | 0.008501532 | [-0.02 0.04] | 0.581765159 | 0.909583681 |
| TFPI | Beta_Cell_2_pPS | 4225 | 0.002405101 | [-0.03 0.03] | 0.876173201 | 0.97781424 |
| TM | Cholesterol_pPS | 4159 | 0.003947244 | [-0.03 0.03] | 0.799714229 | 0.967638637 |
| TM | Lipodystrophy_1_pPS | 4159 | -0.004889949 | [-0.04 0.03] | 0.753284152 | 0.95462436 |
| TM | Lipodystrophy_2_pPS | 4159 | 0.010290833 | [-0.02 0.04] | 0.508305315 | 0.895803896 |
| TM | Liver-Lipid_pPS | 4159 | 0.033370104 | [0. 0.06] | 0.031911358 | 0.428023414 |
| TM | ASAT_Pos_pPS | 4159 | -0.003669652 | [-0.03 0.03] | 0.813530377 | 0.967790883 |
| TM | ALP_Neg_pPS | 4159 | 0.106430892 | [0.08 0.14] | 6.86E-12 | 1.96E-09 |
| TM | SHBG-LpA_pPS | 4159 | 0.019558288 | [-0.01 0.05] | 0.208658278 | 0.767305935 |
| TM | Obesity_pPS | 4159 | 0.010800038 | [-0.02 0.04] | 0.487549572 | 0.895803896 |
| TM | VAT_Neg_pPS | 4159 | -0.01330942 | [-0.04 0.02] | 0.392261063 | 0.85037326 |
| TM | Bilirubin_pPS | 4159 | -0.00578529 | [-0.04 0.02] | 0.709995068 | 0.945970482 |
| TM | Proinsulin_pPS | 4159 | 0.020301479 | [-0.01 0.05] | 0.191876678 | 0.750920235 |
| TM | Hyper_Insulin_pPS | 4159 | 0.009255269 | [-0.02 0.04] | 0.551902759 | 0.899241939 |
| TM | Beta_Cell_1_pPS | 4159 | 0.017294688 | [-0.01 0.05] | 0.266254501 | 0.807858179 |
| TM | Beta_Cell_2_pPS | 4159 | -0.012997643 | [-0.04 0.02] | 0.403446678 | 0.85241904 |
| VEGFD | Cholesterol_pPS | 4159 | -0.008877189 | [-0.04 0.02] | 0.568265354 | 0.905613993 |
| VEGFD | Lipodystrophy_1_pPS | 4159 | 0.027145941 | [-0. 0.06] | 0.080954527 | 0.610643339 |
| VEGFD | Lipodystrophy_2_pPS | 4159 | -0.005419596 | [-0.04 0.03] | 0.727572421 | 0.946981479 |
| VEGFD | Liver-Lipid_pPS | 4159 | -0.00508911 | [-0.04 0.03] | 0.743581988 | 0.952599446 |
| VEGFD | ASAT_Pos_pPS | 4159 | -0.034776821 | [-0.07 -0. ] | 0.025350643 | 0.392967754 |
| VEGFD | ALP_Neg_pPS | 4159 | 0.007292702 | [-0.02 0.04] | 0.639243073 | 0.933760412 |
| VEGFD | SHBG-LpA_pPS | 4159 | 0.024609338 | [-0.01 0.06] | 0.113638677 | 0.681046037 |
| VEGFD | Obesity_pPS | 4159 | -0.001681739 | [-0.03 0.03] | 0.913918911 | 0.989461462 |
| VEGFD | VAT_Neg_pPS | 4159 | 0.002700616 | [-0.03 0.03] | 0.862189976 | 0.97556306 |
| VEGFD | Bilirubin_pPS | 4159 | 0.005386098 | [-0.03 0.04] | 0.729189864 | 0.946981479 |
| VEGFD | Proinsulin_pPS | 4159 | 0.020668535 | [-0.01 0.05] | 0.183964092 | 0.738879686 |
| VEGFD | Hyper_Insulin_pPS | 4159 | -0.01411013 | [-0.04 0.02] | 0.364407657 | 0.847284666 |
| VEGFD | Beta_Cell_1_pPS | 4159 | 0.00261451 | [-0.03 0.03] | 0.866542006 | 0.976043816 |
| VEGFD | Beta_Cell_2_pPS | 4159 | 0.003614119 | [-0.03 0.03] | 0.816301456 | 0.968668398 |
| vWF | Cholesterol_pPS | 4225 | -0.00900189 | [-0.04 0.02] | 0.559740289 | 0.902076574 |
| vWF | Lipodystrophy_1_pPS | 4225 | -0.012460198 | [-0.04 0.02] | 0.419491316 | 0.859608807 |
| vWF | Lipodystrophy_2_pPS | 4225 | 0.00304025 | [-0.03 0.03] | 0.843849023 | 0.972178273 |
| vWF | Liver-Lipid_pPS | 4225 | -0.028414805 | [-0.06 0. ] | 0.065575636 | 0.563327578 |
| vWF | ASAT_Pos_pPS | 4225 | -0.001214802 | [-0.03 0.03] | 0.937267452 | 0.994163908 |
| vWF | ALP_Neg_pPS | 4225 | 0.194436877 | [0.17 0.22] | 4.65E-37 | 3.00E-34 |
| vWF | SHBG-LpA_pPS | 4225 | -0.001677488 | [-0.03 0.03] | 0.913455217 | 0.989461462 |
| vWF | Obesity_pPS | 4225 | -0.018329883 | [-0.05 0.01] | 0.234968035 | 0.788664502 |
| vWF | VAT_Neg_pPS | 4225 | -0.013337468 | [-0.04 0.02] | 0.387506678 | 0.85037326 |
| vWF | Bilirubin_pPS | 4225 | 0.018110234 | [-0.01 0.05] | 0.240625737 | 0.790085251 |
| vWF | Proinsulin_pPS | 4225 | 0.036041687 | [0.01 0.07] | 0.019500005 | 0.341714381 |
| vWF | Hyper_Insulin_pPS | 4225 | 0.016217256 | [-0.01 0.05] | 0.293371326 | 0.816999498 |
| vWF | Beta_Cell_1_pPS | 4225 | -0.025591211 | [-0.06 0. ] | 0.097261381 | 0.64755296 |
| vWF | Beta_Cell_2_pPS | 4225 | -0.00933983 | [-0.04 0.02] | 0.545097156 | 0.899241939 |
| AMBP | Cholesterol_pPS | 4159 | 0.010359298 | [-0.02 0.04] | 0.505487895 | 0.895803896 |
| AMBP | Lipodystrophy_1_pPS | 4159 | 0.016696614 | [-0.01 0.05] | 0.283146039 | 0.81132836 |
| AMBP | Lipodystrophy_2_pPS | 4159 | 0.002309923 | [-0.03 0.03] | 0.881968267 | 0.978445416 |
| AMBP | Liver-Lipid_pPS | 4159 | -0.024372654 | [-0.05 0.01] | 0.117154997 | 0.681046037 |
| AMBP | ASAT_Pos_pPS | 4159 | -0.021722558 | [-0.05 0.01] | 0.162588023 | 0.718997225 |
| AMBP | ALP_Neg_pPS | 4159 | 0.000506101 | [-0.03 0.03] | 0.974048992 | 0.997444743 |
| AMBP | SHBG-LpA_pPS | 4159 | 0.014041382 | [-0.02 0.04] | 0.366749477 | 0.847284666 |
| AMBP | Obesity_pPS | 4159 | 0.012283073 | [-0.02 0.04] | 0.429793107 | 0.864060889 |
| AMBP | VAT_Neg_pPS | 4159 | -0.017945526 | [-0.05 0.01] | 0.248675323 | 0.797743002 |
| AMBP | Bilirubin_pPS | 4159 | 0.009398562 | [-0.02 0.04] | 0.545762299 | 0.899241939 |
| AMBP | Proinsulin_pPS | 4159 | 0.003517781 | [-0.03 0.03] | 0.821114151 | 0.968668398 |
| AMBP | Hyper_Insulin_pPS | 4159 | 0.009880312 | [-0.02 0.04] | 0.52536984 | 0.898044266 |
| AMBP | Beta_Cell_1_pPS | 4159 | 0.016776787 | [-0.01 0.05] | 0.280840505 | 0.808820203 |
| AMBP | Beta_Cell_2_pPS | 4159 | -0.024032311 | [-0.05 0.01] | 0.122360328 | 0.684037897 |
| AZU1 | Cholesterol_pPS | 3944 | -0.01165266 | [-0.04 0.02] | 0.465835967 | 0.879924083 |
| AZU1 | Lipodystrophy_1_pPS | 3944 | 0.016979641 | [-0.01 0.05] | 0.287920011 | 0.81609738 |
| AZU1 | Lipodystrophy_2_pPS | 3944 | 0.023826457 | [-0.01 0.06] | 0.135878085 | 0.694065456 |
| AZU1 | Liver-Lipid_pPS | 3944 | -0.021754338 | [-0.05 0.01] | 0.17332734 | 0.72956083 |
| AZU1 | ASAT_Pos_pPS | 3944 | 0.006992754 | [-0.02 0.04] | 0.661657782 | 0.937489141 |
| AZU1 | ALP_Neg_pPS | 3944 | 0.014578997 | [-0.02 0.05] | 0.361542129 | 0.847284666 |
| AZU1 | SHBG-LpA_pPS | 3944 | -0.015296599 | [-0.05 0.02] | 0.338392152 | 0.844649909 |
| AZU1 | Obesity_pPS | 3944 | 9.63E-05 | [-0.03 0.03] | 0.995193765 | 0.998803911 |
| AZU1 | VAT_Neg_pPS | 3944 | -0.027380529 | [-0.06 0. ] | 0.086556632 | 0.615938909 |
| AZU1 | Bilirubin_pPS | 3944 | 0.023821613 | [-0.01 0.06] | 0.135957688 | 0.694065456 |
| AZU1 | Proinsulin_pPS | 3944 | 0.021406529 | [-0.01 0.05] | 0.18030525 | 0.738879686 |
| AZU1 | Hyper_Insulin_pPS | 3944 | -0.011692312 | [-0.04 0.02] | 0.464319536 | 0.879924083 |

**Table S7: Associations between pPSs and plasma proteins in SCAPIS**

**a) Spearman Correlation**

| Proteomics | PPS | Sample_Size | r | CI95% | p | fdr_p_adjusted |
|---|---|---|---|---|---|---|
| t_PA | Proinsulin_pPS | 4225 | 0.016275619 | [-0.01 0.05] | 0.290891342 | 0.753421877 |
| t_PA | Hyper_Insulin_pPS | 4225 | 0.006662548 | [-0.02 0.04] | 0.66550404 | 0.924111434 |
| t_PA | Beta_Cell_1_pPS | 4225 | 0.024835622 | [-0.01 0.05] | 0.107007319 | 0.597786172 |
| t_PA | Beta_Cell_2_pPS | 4225 | 0.016386848 | [-0.01 0.05] | 0.287606396 | 0.751557788 |
| t_PA | Total_GRS | 4225 | 0.059891841 | [0.03 0.09] | 0.000100337 | 0.010651153 |
| PRTN3 | Cholesterol_pPS | 3944 | 0.00111462 | [-0.03 0.03] | 0.944296523 | 0.993154165 |
| PRTN3 | Lipodystrophy_1_pPS | 3944 | 0.003919533 | [-0.03 0.04] | 0.805914693 | 0.960193229 |
| PRTN3 | Lipodystrophy_2_pPS | 3944 | 0.000460338 | [-0.03 0.03] | 0.976978954 | 0.996862516 |
| PRTN3 | Liver-Lipid_pPS | 3944 | 0.014607543 | [-0.02 0.05] | 0.359806814 | 0.7979754 |
| PRTN3 | ASAT_Pos_pPS | 3944 | 0.027939534 | [-0. 0.06] | 0.079816481 | 0.521702218 |
| PRTN3 | ALP_Neg_pPS | 3944 | 0.005726668 | [-0.03 0.04] | 0.71960759 | 0.941449667 |
| PRTN3 | SHBG-LpA_pPS | 3944 | -0.010165734 | [-0.04 0.02] | 0.523953642 | 0.872980599 |
| PRTN3 | Obesity_pPS | 3944 | 0.012345896 | [-0.02 0.04] | 0.438965742 | 0.830394412 |
| PRTN3 | VAT_Neg_pPS | 3944 | -0.011112841 | [-0.04 0.02] | 0.486029385 | 0.854965648 |
| PRTN3 | Bilirubin_pPS | 3944 | 0.021656971 | [-0.01 0.05] | 0.174545513 | 0.676609012 |
| PRTN3 | Proinsulin_pPS | 3944 | 0.014389407 | [-0.02 0.05] | 0.367026526 | 0.798890546 |
| PRTN3 | Hyper_Insulin_pPS | 3944 | -0.019852874 | [-0.05 0.01] | 0.213273507 | 0.712633026 |
| PRTN3 | Beta_Cell_1_pPS | 3944 | 0.017588667 | [-0.01 0.05] | 0.270181968 | 0.746520328 |
| PRTN3 | Beta_Cell_2_pPS | 3944 | -0.020808306 | [-0.05 0.01] | 0.192054653 | 0.694207068 |
| PRTN3 | Total_GRS | 3944 | 0.003581023 | [-0.03 0.03] | 0.822384035 | 0.963461584 |
| REN | Cholesterol_pPS | 4158 | 0.01220752 | [-0.02 0.04] | 0.431968472 | 0.827992564 |
| REN | Lipodystrophy_1_pPS | 4158 | 0.009888608 | [-0.02 0.04] | 0.524420954 | 0.872980599 |
| REN | Lipodystrophy_2_pPS | 4158 | 0.005389229 | [-0.03 0.04] | 0.728660925 | 0.942134248 |
| REN | Liver-Lipid_pPS | 4158 | -0.020805998 | [-0.05 0.01] | 0.180431509 | 0.680315527 |
| REN | ASAT_Pos_pPS | 4158 | 0.017131824 | [-0.01 0.05] | 0.27008934 | 0.746520328 |
| REN | ALP_Neg_pPS | 4158 | 0.009541589 | [-0.02 0.04] | 0.53907901 | 0.88195499 |
| REN | SHBG-LpA_pPS | 4158 | -0.001427645 | [-0.03 0.03] | 0.926779393 | 0.990056084 |
| REN | Obesity_pPS | 4158 | 0.010164879 | [-0.02 0.04] | 0.512898949 | 0.870690038 |
| REN | VAT_Neg_pPS | 4158 | 0.019118995 | [-0.01 0.05] | 0.218396681 | 0.718175854 |
| REN | Bilirubin_pPS | 4158 | 0.022848207 | [-0.01 0.05] | 0.141309612 | 0.638321653 |
| REN | Proinsulin_pPS | 4158 | 0.000460253 | [-0.03 0.03] | 0.976364936 | 0.996862516 |
| REN | Hyper_Insulin_pPS | 4158 | 0.008258867 | [-0.02 0.04] | 0.594980518 | 0.907233991 |
| REN | Beta_Cell_1_pPS | 4158 | -0.009242737 | [-0.04 0.02] | 0.55186494 | 0.893341486 |
| REN | Beta_Cell_2_pPS | 4158 | 0.004569936 | [-0.03 0.04] | 0.768629202 | 0.952961063 |
| REN | Total_GRS | 4158 | 0.00800938 | [-0.02 0.04] | 0.606153179 | 0.909483337 |
| PSGL_1 | Cholesterol_pPS | 4159 | 0.001728789 | [-0.03 0.03] | 0.911381885 | 0.990056084 |
| PSGL_1 | Lipodystrophy_1_pPS | 4159 | 0.030542722 | [0. 0.06] | 0.049214143 | 0.438587647 |
| PSGL_1 | Lipodystrophy_2_pPS | 4159 | 0.019092563 | [-0.01 0.05] | 0.218978435 | 0.718175854 |
| PSGL_1 | Liver-Lipid_pPS | 4159 | -0.036684931 | [-0.07 -0.01] | 0.018152631 | 0.290017524 |
| PSGL_1 | ASAT_Pos_pPS | 4159 | -0.018495144 | [-0.05 0.01] | 0.233740596 | 0.726631332 |
| PSGL_1 | ALP_Neg_pPS | 4159 | -0.020078163 | [-0.05 0.01] | 0.196108582 | 0.697416791 |
| PSGL_1 | SHBG-LpA_pPS | 4159 | 0.025876725 | [-0. 0.06] | 0.095680476 | 0.567652214 |
| PSGL_1 | Obesity_pPS | 4159 | 0.029274055 | [-0. 0.06] | 0.059429437 | 0.46074507 |
| PSGL_1 | VAT_Neg_pPS | 4159 | 0.013452314 | [-0.02 0.04] | 0.386452087 | 0.807658934 |
| PSGL_1 | Bilirubin_pPS | 4159 | 0.01017398 | [-0.02 0.04] | 0.512470778 | 0.870690038 |
| PSGL_1 | Proinsulin_pPS | 4159 | -0.001957985 | [-0.03 0.03] | 0.899691675 | 0.985767774 |
| PSGL_1 | Hyper_Insulin_pPS | 4159 | -0.017004971 | [-0.05 0.01] | 0.273594787 | 0.746520328 |
| PSGL_1 | Beta_Cell_1_pPS | 4159 | -0.007681791 | [-0.04 0.02] | 0.620921997 | 0.910907479 |
| PSGL_1 | Beta_Cell_2_pPS | 4159 | 0.005585057 | [-0.02 0.04] | 0.719178847 | 0.941449667 |
| PSGL_1 | Total_GRS | 4159 | 0.042536729 | [0.01 0.07] | 0.006150266 | 0.156589778 |
| TIE2 | Cholesterol_pPS | 4159 | 0.016641214 | [-0.01 0.05] | 0.283990146 | 0.749111537 |
| TIE2 | Lipodystrophy_1_pPS | 4159 | -0.017763261 | [-0.05 0.01] | 0.252768918 | 0.739680398 |
| TIE2 | Lipodystrophy_2_pPS | 4159 | 0.031721704 | [0. 0.06] | 0.041083407 | 0.407062052 |
| TIE2 | Liver-Lipid_pPS | 4159 | -0.020057959 | [-0.05 0.01] | 0.196559106 | 0.697416791 |
| TIE2 | ASAT_Pos_pPS | 4159 | -0.031170056 | [-0.06 -0. ] | 0.044734009 | 0.424281318 |
| TIE2 | ALP_Neg_pPS | 4159 | 0.086608135 | [0.06 0.12] | 2.32E-08 | 4.58E-06 |
| TIE2 | SHBG-LpA_pPS | 4159 | -0.018500403 | [-0.05 0.01] | 0.23360764 | 0.726631332 |
| TIE2 | Obesity_pPS | 4159 | -0.001662821 | [-0.03 0.03] | 0.914750256 | 0.990056084 |
| TIE2 | VAT_Neg_pPS | 4159 | 0.016195645 | [-0.01 0.05] | 0.297083431 | 0.759609171 |
| TIE2 | Bilirubin_pPS | 4159 | 0.000672976 | [-0.03 0.03] | 0.965442691 | 0.99612031 |
| TIE2 | Proinsulin_pPS | 4159 | 0.04496648 | [0.01 0.08] | 0.003775821 | 0.117810613 |
| TIE2 | Hyper_Insulin_pPS | 4159 | -0.022406817 | [-0.05 0.01] | 0.149109926 | 0.648099837 |
| TIE2 | Beta_Cell_1_pPS | 4159 | -0.007756573 | [-0.04 0.02] | 0.61752682 | 0.909483337 |
| TIE2 | Beta_Cell_2_pPS | 4159 | -0.009686095 | [-0.04 0.02] | 0.532900796 | 0.878093252 |
| TIE2 | Total_GRS | 4159 | 0.001416206 | [-0.03 0.03] | 0.927355693 | 0.990056084 |
| TFPI | Cholesterol_pPS | 4225 | -0.071186396 | [-0.1 -0.04] | 3.75E-06 | 0.000544056 |
| TFPI | Lipodystrophy_1_pPS | 4225 | 0.02315569 | [-0.01 0.05] | 0.132907046 | 0.629316669 |
| TFPI | Lipodystrophy_2_pPS | 4225 | -0.013139557 | [-0.04 0.02] | 0.393858299 | 0.812191725 |
| TFPI | Liver-Lipid_pPS | 4225 | -0.034404658 | [-0.06 -0. ] | 0.025540826 | 0.338445886 |
| TFPI | ASAT_Pos_pPS | 4225 | 0.026974456 | [-0. 0.06] | 0.080006754 | 0.521702218 |
| TFPI | ALP_Neg_pPS | 4225 | 0.018049194 | [-0.01 0.05] | 0.241487003 | 0.729918816 |
| TFPI | SHBG-LpA_pPS | 4225 | -0.005999195 | [-0.04 0.02] | 0.69706795 | 0.930772879 |
| TFPI | Obesity_pPS | 4225 | -0.009077676 | [-0.04 0.02] | 0.555829282 | 0.896216632 |
| TFPI | VAT_Neg_pPS | 4225 | -0.001590016 | [-0.03 0.03] | 0.917825171 | 0.990056084 |
| TFPI | Bilirubin_pPS | 4225 | -0.012926597 | [-0.04 0.02] | 0.401569873 | 0.81230319 |
| TFPI | Proinsulin_pPS | 4225 | 0.020457795 | [-0.01 0.05] | 0.184306371 | 0.686251968 |
| TFPI | Hyper_Insulin_pPS | 4225 | -0.014501661 | [-0.04 0.02] | 0.346685113 | 0.790391668 |
| TFPI | Beta_Cell_1_pPS | 4225 | 0.01566903 | [-0.01 0.05] | 0.309248514 | 0.76365449 |
| TFPI | Beta_Cell_2_pPS | 4225 | 0.005286262 | [-0.02 0.04] | 0.731583943 | 0.942134248 |
| TFPI | Total_GRS | 4225 | -0.001316601 | [-0.03 0.03] | 0.931917846 | 0.990113904 |
| TM | Cholesterol_pPS | 4159 | 0.00714932 | [-0.02 0.04] | 0.645327228 | 0.916568313 |
| TM | Lipodystrophy_1_pPS | 4159 | 0.000270112 | [-0.03 0.03] | 0.986126111 | 0.998703927 |
| TM | Lipodystrophy_2_pPS | 4159 | 0.009933605 | [-0.02 0.04] | 0.522485152 | 0.872980599 |
| TM | Liver-Lipid_pPS | 4159 | 0.038574186 | [0.01 0.07] | 0.012982628 | 0.242414912 |
| TM | ASAT_Pos_pPS | 4159 | -0.001559421 | [-0.03 0.03] | 0.920033014 | 0.990056084 |
| TM | ALP_Neg_pPS | 4159 | 0.112571332 | [0.08 0.14] | 3.59E-13 | 1.10E-10 |
| TM | SHBG-LpA_pPS | 4159 | 0.016335578 | [-0.01 0.05] | 0.292928668 | 0.753479147 |
| TM | Obesity_pPS | 4159 | 0.016616132 | [-0.01 0.05] | 0.284716668 | 0.749111537 |
| TM | VAT_Neg_pPS | 4159 | -0.008105503 | [-0.04 0.02] | 0.601793624 | 0.907623171 |
| TM | Bilirubin_pPS | 4159 | -0.005762053 | [-0.04 0.02] | 0.710673801 | 0.938946716 |
| TM | Proinsulin_pPS | 4159 | 0.045691489 | [0.02 0.08] | 0.003249915 | 0.106782928 |
| TM | Hyper_Insulin_pPS | 4159 | 0.013045236 | [-0.02 0.04] | 0.400987331 | 0.81230319 |
| TM | Beta_Cell_1_pPS | 4159 | 0.021176505 | [-0.01 0.05] | 0.172740889 | 0.674627844 |
| TM | Beta_Cell_2_pPS | 4159 | -0.002579676 | [-0.03 0.03] | 0.868098639 | 0.977006629 |
| TM | Total_GRS | 4159 | 0.043103974 | [0.01 0.07] | 0.005499348 | 0.150279217 |
| VEGFD | Cholesterol_pPS | 4159 | -0.009344045 | [-0.04 0.02] | 0.547465666 | 0.8909229 |
| VEGFD | Lipodystrophy_1_pPS | 4159 | 0.021463467 | [-0.01 0.05] | 0.166993269 | 0.666299341 |
| VEGFD | Lipodystrophy_2_pPS | 4159 | -0.007611366 | [-0.04 0.02] | 0.624126781 | 0.91287224 |
| VEGFD | Liver-Lipid_pPS | 4159 | -0.00538112 | [-0.04 0.03] | 0.729021738 | 0.942134248 |
| VEGFD | ASAT_Pos_pPS | 4159 | -0.031868065 | [-0.06 -0. ] | 0.040158312 | 0.407062052 |
| VEGFD | ALP_Neg_pPS | 4159 | 0.010538023 | [-0.02 0.04] | 0.497496875 | 0.863780476 |
| VEGFD | SHBG-LpA_pPS | 4159 | 0.027283587 | [-0. 0.06] | 0.078954623 | 0.520082958 |
| VEGFD | Obesity_pPS | 4159 | -0.000919369 | [-0.03 0.03] | 0.952803186 | 0.995735249 |
| VEGFD | VAT_Neg_pPS | 4159 | 0.004657248 | [-0.03 0.04] | 0.764310683 | 0.951939298 |
| VEGFD | Bilirubin_pPS | 4159 | 0.005408252 | [-0.03 0.04] | 0.727709606 | 0.942134248 |
| VEGFD | Proinsulin_pPS | 4159 | 0.025422477 | [-0.01 0.06] | 0.101649809 | 0.586731767 |
| VEGFD | Hyper_Insulin_pPS | 4159 | -0.01369293 | [-0.04 0.02] | 0.378013743 | 0.803170077 |

**b) Partial Correlation**

| Proteomics | PPS | Sample_Size | r | CI95% | p | fdr_p_adjusted |
|---|---|---|---|---|---|---|
| AZU1 | Beta_Cell_1_pPS | 3944 | -0.003824637 | [-0.04 0.03] | 0.810831035 | 0.967638637 |
| AZU1 | Beta_Cell_2_pPS | 3944 | -0.021632852 | [-0.05 0.01] | 0.175741343 | 0.732083915 |
| BLM_hydrolase | Cholesterol_pPS | 4074 | 0.00388308 | [-0.03 0.03] | 0.804899081 | 0.967638637 |
| BLM_hydrolase | Lipodystrophy_1_pPS | 4074 | 0.01310848 | [-0.02 0.04] | 0.404340805 | 0.85241904 |
| BLM_hydrolase | Lipodystrophy_2_pPS | 4074 | 0.020036749 | [-0.01 0.05] | 0.202412705 | 0.765145476 |
| BLM_hydrolase | Liver-Lipid_pPS | 4074 | -0.021661914 | [-0.05 0.01] | 0.168165738 | 0.722132896 |
| BLM_hydrolase | ASAT_Pos_pPS | 4074 | -0.00558057 | [-0.04 0.03] | 0.722593647 | 0.946314812 |
| BLM_hydrolase | ALP_Neg_pPS | 4074 | -0.00817936 | [-0.04 0.02] | 0.602844052 | 0.918301271 |
| BLM_hydrolase | SHBG-LpA_pPS | 4074 | 0.012403304 | [-0.02 0.04] | 0.430094118 | 0.864060889 |
| BLM_hydrolase | Obesity_pPS | 4074 | 0.000748544 | [-0.03 0.03] | 0.962021896 | 0.996924797 |
| BLM_hydrolase | VAT_Neg_pPS | 4074 | -0.015998076 | [-0.05 0.01] | 0.308803214 | 0.830335001 |
| BLM_hydrolase | Bilirubin_pPS | 4074 | 2.90E-05 | [-0.03 0.03] | 0.998529942 | 0.999305801 |
| BLM_hydrolase | Proinsulin_pPS | 4074 | 0.011855567 | [-0.02 0.04] | 0.45073869 | 0.870351501 |
| BLM_hydrolase | Hyper_Insulin_pPS | 4074 | -0.019958448 | [-0.05 0.01] | 0.204182471 | 0.765145476 |
| BLM_hydrolase | Beta_Cell_1_pPS | 4074 | -0.002555607 | [-0.03 0.03] | 0.870858719 | 0.977511154 |
| BLM_hydrolase | Beta_Cell_2_pPS | 4074 | 0.010231532 | [-0.02 0.04] | 0.515132792 | 0.895803896 |
| CSTB | Cholesterol_pPS | 4087 | 0.006246356 | [-0.02 0.04] | 0.69064086 | 0.945319264 |
| CSTB | Lipodystrophy_1_pPS | 4087 | 0.008102887 | [-0.02 0.04] | 0.605660628 | 0.918301271 |
| CSTB | Lipodystrophy_2_pPS | 4087 | 0.005060669 | [-0.03 0.04] | 0.74712048 | 0.954135344 |
| CSTB | Liver-Lipid_pPS | 4087 | 0.023108702 | [-0.01 0.05] | 0.140872619 | 0.697681852 |
| CSTB | ASAT_Pos_pPS | 4087 | -0.006528085 | [-0.04 0.02] | 0.677456723 | 0.942803089 |
| CSTB | ALP_Neg_pPS | 4087 | -0.008880765 | [-0.04 0.02] | 0.571501154 | 0.907308519 |
| CSTB | SHBG-LpA_pPS | 4087 | 0.001624963 | [-0.03 0.03] | 0.917539724 | 0.99041009 |
| CSTB | Obesity_pPS | 4087 | 0.023142517 | [-0.01 0.05] | 0.140291989 | 0.697681852 |
| CSTB | VAT_Neg_pPS | 4087 | -0.022447933 | [-0.05 0.01] | 0.152592939 | 0.706977356 |
| CSTB | Bilirubin_pPS | 4087 | -0.000895824 | [-0.03 0.03] | 0.954484083 | 0.996425456 |
| CSTB | Proinsulin_pPS | 4087 | 0.01577947 | [-0.01 0.05] | 0.314685517 | 0.836563355 |
| CSTB | Hyper_Insulin_pPS | 4087 | -0.009775348 | [-0.04 0.02] | 0.533387438 | 0.898959051 |
| CSTB | Beta_Cell_1_pPS | 4087 | -0.024955115 | [-0.06 0.01] | 0.111780045 | 0.680721977 |
| CSTB | Beta_Cell_2_pPS | 4087 | 0.000194397 | [-0.03 0.03] | 0.990117766 | 0.998803911 |
| CTRC | Cholesterol_pPS | 4157 | 0.00375161 | [-0.03 0.03] | 0.809490224 | 0.967638637 |
| CTRC | Lipodystrophy_1_pPS | 4157 | 0.004598738 | [-0.03 0.04] | 0.767595196 | 0.960245786 |
| CTRC | Lipodystrophy_2_pPS | 4157 | 0.031456072 | [0. 0.06] | 0.043186186 | 0.494433848 |
| CTRC | Liver-Lipid_pPS | 4157 | -0.053339089 | [-0.08 -0.02] | 0.000603415 | 0.042010707 |
| CTRC | ASAT_Pos_pPS | 4157 | -0.01124666 | [-0.04 0.02] | 0.469835582 | 0.882783706 |
| CTRC | ALP_Neg_pPS | 4157 | 0.109195697 | [0.08 0.14] | 1.95E-12 | 6.28E-10 |
| CTRC | SHBG-LpA_pPS | 4157 | -0.03547238 | [-0.07 -0. ] | 0.022594953 | 0.36838354 |
| CTRC | Obesity_pPS | 4157 | -0.007307734 | [-0.04 0.02] | 0.638633745 | 0.933760412 |
| CTRC | VAT_Neg_pPS | 4157 | -0.007149792 | [-0.04 0.02] | 0.645903861 | 0.935271696 |
| CTRC | Bilirubin_pPS | 4157 | -0.013881657 | [-0.04 0.02] | 0.372342175 | 0.847284666 |
| CTRC | Proinsulin_pPS | 4157 | 0.021059162 | [-0.01 0.05] | 0.175916127 | 0.732083915 |
| CTRC | Hyper_Insulin_pPS | 4157 | 0.019809346 | [-0.01 0.05] | 0.20298364 | 0.765145476 |
| CTRC | Beta_Cell_1_pPS | 4157 | -0.006030954 | [-0.04 0.02] | 0.698341856 | 0.945970482 |
| CTRC | Beta_Cell_2_pPS | 4157 | 0.004671793 | [-0.03 0.04] | 0.764011949 | 0.959095176 |
| KLK6 | Cholesterol_pPS | 4225 | 0.018688027 | [-0.01 0.05] | 0.225946071 | 0.784416549 |
| KLK6 | Lipodystrophy_1_pPS | 4225 | -0.009186282 | [-0.04 0.02] | 0.551726849 | 0.899241939 |
| KLK6 | Lipodystrophy_2_pPS | 4225 | 0.03363381 | [0. 0.06] | 0.029280606 | 0.414433192 |
| KLK6 | Liver-Lipid_pPS | 4225 | -0.001907889 | [-0.03 0.03] | 0.901625135 | 0.985399384 |
| KLK6 | ASAT_Pos_pPS | 4225 | 0.009361875 | [-0.02 0.04] | 0.544148577 | 0.899241939 |
| KLK6 | ALP_Neg_pPS | 4225 | 0.006221931 | [-0.02 0.04] | 0.686866493 | 0.944571269 |
| KLK6 | SHBG-LpA_pPS | 4225 | -0.00311496 | [-0.03 0.03] | 0.840063009 | 0.972178273 |
| KLK6 | Obesity_pPS | 4225 | -0.035050078 | [-0.07 -0. ] | 0.023115128 | 0.369842052 |
| KLK6 | VAT_Neg_pPS | 4225 | -0.026631592 | [-0.06 0. ] | 0.084398386 | 0.614725404 |
| KLK6 | Bilirubin_pPS | 4225 | -0.006677679 | [-0.04 0.02] | 0.665277002 | 0.937489141 |
| KLK6 | Proinsulin_pPS | 4225 | 0.005949372 | [-0.02 0.04] | 0.699902558 | 0.945970482 |
| KLK6 | Hyper_Insulin_pPS | 4225 | -0.002697025 | [-0.03 0.03] | 0.861287483 | 0.97556306 |
| KLK6 | Beta_Cell_1_pPS | 4225 | -0.010204082 | [-0.04 0.02] | 0.50853508 | 0.895803896 |
| KLK6 | Beta_Cell_2_pPS | 4225 | 0.040762254 | [0.01 0.07] | 0.008241619 | 0.228283991 |
| MERTK | Cholesterol_pPS | 4159 | 0.013395283 | [-0.02 0.04] | 0.389213921 | 0.85037326 |
| MERTK | Lipodystrophy_1_pPS | 4159 | 0.003732698 | [-0.03 0.03] | 0.810387205 | 0.967638637 |
| MERTK | Lipodystrophy_2_pPS | 4159 | 0.01986429 | [-0.01 0.05] | 0.201624172 | 0.765145476 |
| MERTK | Liver-Lipid_pPS | 4159 | -0.013978958 | [-0.04 0.02] | 0.368883978 | 0.847284666 |
| MERTK | ASAT_Pos_pPS | 4159 | -0.028942613 | [-0.06 0. ] | 0.062782779 | 0.558815902 |
| MERTK | ALP_Neg_pPS | 4159 | -0.062437892 | [-0.09 -0.03] | 5.89E-05 | 0.006926058 |
| MERTK | SHBG-LpA_pPS | 4159 | 0.013510846 | [-0.02 0.04] | 0.385135521 | 0.85037326 |
| MERTK | Obesity_pPS | 4159 | 0.020649555 | [-0.01 0.05] | 0.184367236 | 0.738879686 |
| MERTK | VAT_Neg_pPS | 4159 | -0.006115407 | [-0.04 0.02] | 0.694258817 | 0.945970482 |
| MERTK | Bilirubin_pPS | 4159 | 0.028637666 | [-0. 0.06] | 0.06560492 | 0.563327578 |
| MERTK | Proinsulin_pPS | 4159 | 0.024290888 | [-0.01 0.05] | 0.118389407 | 0.681046037 |
| MERTK | Hyper_Insulin_pPS | 4159 | -0.014198906 | [-0.04 0.02] | 0.361397465 | 0.847284666 |
| MERTK | Beta_Cell_1_pPS | 4159 | -0.004925419 | [-0.04 0.03] | 0.751553328 | 0.954135344 |
| MERTK | Beta_Cell_2_pPS | 4159 | 0.003296695 | [-0.03 0.03] | 0.832183971 | 0.971458352 |
| PI3 | Cholesterol_pPS | 3943 | 0.012179891 | [-0.02 0.04] | 0.445957228 | 0.869647841 |
| PI3 | Lipodystrophy_1_pPS | 3943 | 0.026795198 | [-0. 0.06] | 0.093545882 | 0.639188839 |
| PI3 | Lipodystrophy_2_pPS | 3943 | 0.015293052 | [-0.02 0.05] | 0.338565888 | 0.844649909 |
| PI3 | Liver-Lipid_pPS | 3943 | -0.006272461 | [-0.04 0.03] | 0.694691354 | 0.945970482 |
| PI3 | ASAT_Pos_pPS | 3943 | 0.001701226 | [-0.03 0.03] | 0.915223333 | 0.989461462 |
| PI3 | ALP_Neg_pPS | 3943 | -0.008132989 | [-0.04 0.02] | 0.610807119 | 0.919490897 |
| PI3 | SHBG-LpA_pPS | 3943 | 0.003686194 | [-0.03 0.03] | 0.817578533 | 0.968668398 |
| PI3 | Obesity_pPS | 3943 | 3.51E-05 | [-0.03 0.03] | 0.998245557 | 0.999305801 |
| PI3 | VAT_Neg_pPS | 3943 | 0.0025649 | [-0.03 0.03] | 0.872490228 | 0.97781424 |
| PI3 | Bilirubin_pPS | 3943 | 0.033549179 | [0. 0.06] | 0.035737994 | 0.449078407 |
| PI3 | Proinsulin_pPS | 3943 | -0.012250276 | [-0.04 0.02] | 0.443333051 | 0.867534836 |
| PI3 | Hyper_Insulin_pPS | 3943 | 0.001538817 | [-0.03 0.03] | 0.923290333 | 0.990503296 |
| PI3 | Beta_Cell_1_pPS | 3943 | -0.003848857 | [-0.04 0.03] | 0.809679815 | 0.967638637 |
| PI3 | Beta_Cell_2_pPS | 3943 | -0.001733861 | [-0.03 0.03] | 0.913603363 | 0.989461462 |
| PRSS27 | Cholesterol_pPS | 4158 | -0.006946193 | [-0.04 0.02] | 0.655286629 | 0.936605382 |
| PRSS27 | Lipodystrophy_1_pPS | 4158 | -0.00725694 | [-0.04 0.02] | 0.640927645 | 0.933976497 |
| PRSS27 | Lipodystrophy_2_pPS | 4158 | 0.005226131 | [-0.03 0.04] | 0.73696113 | 0.949205935 |
| PRSS27 | Liver-Lipid_pPS | 4158 | 0.014585969 | [-0.02 0.05] | 0.348514278 | 0.847284666 |
| PRSS27 | ASAT_Pos_pPS | 4158 | -0.008873576 | [-0.04 0.02] | 0.568469653 | 0.905613993 |
| PRSS27 | ALP_Neg_pPS | 4158 | -0.00977286 | [-0.04 0.02] | 0.529934116 | 0.898959051 |
| PRSS27 | SHBG-LpA_pPS | 4158 | 0.006749667 | [-0.02 0.04] | 0.664434361 | 0.937489141 |
| PRSS27 | Obesity_pPS | 4158 | -0.018011053 | [-0.05 0.01] | 0.247008673 | 0.796363381 |
| PRSS27 | VAT_Neg_pPS | 4158 | -0.005798136 | [-0.04 0.02] | 0.709413931 | 0.945970482 |
| PRSS27 | Bilirubin_pPS | 4158 | -0.026333087 | [-0.06 0. ] | 0.090513771 | 0.634479633 |
| PRSS27 | Proinsulin_pPS | 4158 | -0.027412149 | [-0.06 0. ] | 0.07805552 | 0.602398521 |
| PRSS27 | Hyper_Insulin_pPS | 4158 | 0.007187964 | [-0.02 0.04] | 0.644103577 | 0.934766655 |
| PRSS27 | Beta_Cell_1_pPS | 4158 | 0.021664003 | [-0.01 0.05] | 0.163775122 | 0.718997225 |
| PRSS27 | Beta_Cell_2_pPS | 4158 | 0.020833766 | [-0.01 0.05] | 0.180534885 | 0.738879686 |
| PRSS8 | Cholesterol_pPS | 4159 | -0.003214706 | [-0.03 0.03] | 0.836297743 | 0.972099644 |
| PRSS8 | Lipodystrophy_1_pPS | 4159 | 0.035358168 | [0. 0.07] | 0.023000954 | 0.369842052 |
| PRSS8 | Lipodystrophy_2_pPS | 4159 | 0.048915499 | [0.02 0.08] | 0.00165513 | 0.093356425 |
| PRSS8 | Liver-Lipid_pPS | 4159 | -0.038296236 | [-0.07 -0.01] | 0.013798729 | 0.309091537 |
| PRSS8 | ASAT_Pos_pPS | 4159 | -0.006312468 | [-0.04 0.02] | 0.684927071 | 0.944421056 |
| PRSS8 | ALP_Neg_pPS | 4159 | -0.003191075 | [-0.03 0.03] | 0.837484255 | 0.972099644 |
| PRSS8 | SHBG-LpA_pPS | 4159 | 0.011536894 | [-0.02 0.04] | 0.458342658 | 0.874172623 |

**Table S7: Associations between pPSs and plasma proteins in SCAPIS**

**a) Spearman Correlation**

| Proteomics | PPS | Sample_Size | r | CI95% | p | fdr_p_adjusted |
|---|---|---|---|---|---|---|
| VEGFD | Beta_Cell_1_pPS | 4159 | 0.010493427 | [-0.02 0.04] | 0.499318605 | 0.863780476 |
| VEGFD | Beta_Cell_2_pPS | 4159 | 0.008884221 | [-0.02 0.04] | 0.567350574 | 0.899863296 |
| VEGFD | Total_GRS | 4159 | 0.01064711 | [-0.02 0.04] | 0.493055665 | 0.860742337 |
| vWF | Cholesterol_pPS | 4225 | -0.008871309 | [-0.04 0.02] | 0.56484767 | 0.899863296 |
| vWF | Lipodystrophy_1_pPS | 4225 | -0.008357257 | [-0.04 0.02] | 0.587613083 | 0.904020128 |
| vWF | Lipodystrophy_2_pPS | 4225 | 0.002485995 | [-0.03 0.03] | 0.871847342 | 0.977006629 |
| vWF | Liver-Lipid_pPS | 4225 | -0.020123733 | [-0.05 0.01] | 0.191576912 | 0.693900626 |
| vWF | ASAT_Pos_pPS | 4225 | -0.002546518 | [-0.03 0.03] | 0.868755347 | 0.977006629 |
| vWF | ALP_Neg_pPS | 4225 | 0.201854487 | [0.17 0.23] | 5.55E-40 | 3.83E-37 |
| vWF | SHBG-LpA_pPS | 4225 | -0.007123023 | [-0.04 0.02] | 0.643932757 | 0.916568313 |
| vWF | Obesity_pPS | 4225 | -0.011153172 | [-0.04 0.02] | 0.469229308 | 0.844602693 |
| vWF | VAT_Neg_pPS | 4225 | -0.014548576 | [-0.04 0.02] | 0.345127098 | 0.790391668 |
| vWF | Bilirubin_pPS | 4225 | 0.017067279 | [-0.01 0.05] | 0.268056865 | 0.746520328 |
| vWF | Proinsulin_pPS | 4225 | 0.058005416 | [0.03 0.09] | 0.000165189 | 0.015197379 |
| vWF | Hyper_Insulin_pPS | 4225 | 0.015667108 | [-0.01 0.05] | 0.309307845 | 0.76365449 |
| vWF | Beta_Cell_1_pPS | 4225 | -0.019720397 | [-0.05 0.01] | 0.200633508 | 0.700947446 |
| vWF | Beta_Cell_2_pPS | 4225 | -0.009012803 | [-0.04 0.02] | 0.55865669 | 0.897093962 |
| vWF | Total_GRS | 4225 | -0.000392388 | [-0.03 0.03] | 0.979686933 | 0.996862516 |
| AMBP | Cholesterol_pPS | 4159 | 0.011528942 | [-0.02 0.04] | 0.457946485 | 0.83593406 |
| AMBP | Lipodystrophy_1_pPS | 4159 | 0.016168321 | [-0.01 0.05] | 0.297899285 | 0.759754073 |
| AMBP | Lipodystrophy_2_pPS | 4159 | -0.001062423 | [-0.03 0.03] | 0.94547002 | 0.993154165 |
| AMBP | Liver-Lipid_pPS | 4159 | -0.024030756 | [-0.05 0.01] | 0.121798055 | 0.615837655 |
| AMBP | ASAT_Pos_pPS | 4159 | -0.017016094 | [-0.05 0.01] | 0.273281055 | 0.746520328 |
| AMBP | ALP_Neg_pPS | 4159 | 0.000571699 | [-0.03 0.03] | 0.970640716 | 0.996862516 |
| AMBP | SHBG-LpA_pPS | 4159 | 0.013647886 | [-0.02 0.04] | 0.379584742 | 0.805487278 |
| AMBP | Obesity_pPS | 4159 | 0.013884908 | [-0.02 0.04] | 0.371363201 | 0.799802236 |
| AMBP | VAT_Neg_pPS | 4159 | -0.016393641 | [-0.05 0.01] | 0.291216221 | 0.753421877 |
| AMBP | Bilirubin_pPS | 4159 | 0.009641842 | [-0.02 0.04] | 0.534774067 | 0.880129054 |
| AMBP | Proinsulin_pPS | 4159 | 0.005900043 | [-0.02 0.04] | 0.704067881 | 0.937028372 |
| AMBP | Hyper_Insulin_pPS | 4159 | 0.008627552 | [-0.02 0.04] | 0.578598453 | 0.903823705 |
| AMBP | Beta_Cell_1_pPS | 4159 | 0.00989474 | [-0.02 0.04] | 0.524113725 | 0.872980599 |
| AMBP | Beta_Cell_2_pPS | 4159 | -0.020857519 | [-0.05 0.01] | 0.179302059 | 0.679661798 |
| AMBP | Total_GRS | 4159 | 0.007224672 | [-0.02 0.04] | 0.641849457 | 0.916400472 |
| AZU1 | Cholesterol_pPS | 3944 | -0.010216896 | [-0.04 0.02] | 0.521866928 | 0.872980599 |
| AZU1 | Lipodystrophy_1_pPS | 3944 | 0.017208145 | [-0.01 0.05] | 0.280683635 | 0.748127084 |
| AZU1 | Lipodystrophy_2_pPS | 3944 | 0.020473912 | [-0.01 0.05] | 0.199297157 | 0.700947446 |
| AZU1 | Liver-Lipid_pPS | 3944 | -0.021696941 | [-0.05 0.01] | 0.173751271 | 0.674627844 |
| AZU1 | ASAT_Pos_pPS | 3944 | 0.012398488 | [-0.02 0.04] | 0.437018338 | 0.829429813 |
| AZU1 | ALP_Neg_pPS | 3944 | 0.017657236 | [-0.01 0.05] | 0.268318614 | 0.746520328 |
| AZU1 | SHBG-LpA_pPS | 3944 | -0.016078402 | [-0.05 0.02] | 0.313477781 | 0.76365449 |
| AZU1 | Obesity_pPS | 3944 | 0.00210854 | [-0.03 0.03] | 0.894845501 | 0.982879285 |
| AZU1 | VAT_Neg_pPS | 3944 | -0.027305485 | [-0.06 0. ] | 0.08689975 | 0.54140702 |
| AZU1 | Bilirubin_pPS | 3944 | 0.02470408 | [-0.01 0.06] | 0.121422714 | 0.615837655 |
| AZU1 | Proinsulin_pPS | 3944 | 0.018681575 | [-0.01 0.05] | 0.24153034 | 0.729918816 |
| AZU1 | Hyper_Insulin_pPS | 3944 | -0.011498891 | [-0.04 0.02] | 0.471007883 | 0.846342289 |
| AZU1 | Beta_Cell_1_pPS | 3944 | -0.006002403 | [-0.04 0.03] | 0.706717392 | 0.937738119 |
| AZU1 | Beta_Cell_2_pPS | 3944 | -0.021915884 | [-0.05 0.01] | 0.169448486 | 0.672917731 |
| AZU1 | Total_GRS | 3944 | 0.006670661 | [-0.02 0.04] | 0.675830252 | 0.925361755 |
| BLM_hydrolase | Cholesterol_pPS | 4074 | 0.00465241 | [-0.03 0.04] | 0.766904594 | 0.952160028 |
| BLM_hydrolase | Lipodystrophy_1_pPS | 4074 | 0.011287253 | [-0.02 0.04] | 0.472029221 | 0.846453261 |
| BLM_hydrolase | Lipodystrophy_2_pPS | 4074 | 0.017069079 | [-0.01 0.05] | 0.276761996 | 0.747422497 |
| BLM_hydrolase | Liver-Lipid_pPS | 4074 | -0.021801049 | [-0.05 0.01] | 0.164771454 | 0.664372291 |
| BLM_hydrolase | ASAT_Pos_pPS | 4074 | -0.003523694 | [-0.03 0.03] | 0.822360443 | 0.963461584 |
| BLM_hydrolase | ALP_Neg_pPS | 4074 | -0.007485239 | [-0.04 0.02] | 0.633417128 | 0.914460114 |
| BLM_hydrolase | SHBG-LpA_pPS | 4074 | 0.012878946 | [-0.02 0.04] | 0.411870815 | 0.8164748 |
| BLM_hydrolase | Obesity_pPS | 4074 | 0.001316262 | [-0.03 0.03] | 0.933164072 | 0.990208704 |
| BLM_hydrolase | VAT_Neg_pPS | 4074 | -0.013959062 | [-0.04 0.02] | 0.373769942 | 0.80093559 |
| BLM_hydrolase | Bilirubin_pPS | 4074 | 0.000707601 | [-0.03 0.03] | 0.964040173 | 0.99612031 |
| BLM_hydrolase | Proinsulin_pPS | 4074 | 0.012042372 | [-0.02 0.04] | 0.442904377 | 0.831350198 |
| BLM_hydrolase | Hyper_Insulin_pPS | 4074 | -0.019549523 | [-0.05 0.01] | 0.212874059 | 0.712633026 |
| BLM_hydrolase | Beta_Cell_1_pPS | 4074 | 0.000522208 | [-0.03 0.03] | 0.973457636 | 0.996862516 |
| BLM_hydrolase | Beta_Cell_2_pPS | 4074 | 0.010268258 | [-0.02 0.04] | 0.512950001 | 0.870690038 |
| BLM_hydrolase | Total_GRS | 4074 | 0.004717655 | [-0.03 0.04] | 0.763732269 | 0.951648335 |
| CSTB | Cholesterol_pPS | 4087 | 0.010462372 | [-0.02 0.04] | 0.504333718 | 0.866725443 |
| CSTB | Lipodystrophy_1_pPS | 4087 | 0.005817784 | [-0.02 0.04] | 0.710435183 | 0.938946716 |
| CSTB | Lipodystrophy_2_pPS | 4087 | 0.003050809 | [-0.03 0.03] | 0.845634607 | 0.974332356 |
| CSTB | Liver-Lipid_pPS | 4087 | 0.022298447 | [-0.01 0.05] | 0.154684623 | 0.650979658 |
| CSTB | ASAT_Pos_pPS | 4087 | -0.005398701 | [-0.04 0.03] | 0.730450163 | 0.942134248 |
| CSTB | ALP_Neg_pPS | 4087 | -0.005276118 | [-0.04 0.03] | 0.736340185 | 0.942470055 |
| CSTB | SHBG-LpA_pPS | 4087 | 0.001196629 | [-0.03 0.03] | 0.939129308 | 0.992211394 |
| CSTB | Obesity_pPS | 4087 | 0.025262147 | [-0.01 0.06] | 0.106877082 | 0.597786172 |
| CSTB | VAT_Neg_pPS | 4087 | -0.025381364 | [-0.06 0.01] | 0.105231907 | 0.595164067 |
| CSTB | Bilirubin_pPS | 4087 | -0.002039273 | [-0.03 0.03] | 0.896456736 | 0.984176847 |
| CSTB | Proinsulin_pPS | 4087 | 0.010483078 | [-0.02 0.04] | 0.503490391 | 0.865815252 |
| CSTB | Hyper_Insulin_pPS | 4087 | -0.008913272 | [-0.04 0.02] | 0.569476991 | 0.900720055 |
| CSTB | Beta_Cell_1_pPS | 4087 | -0.023940255 | [-0.05 0.01] | 0.12651411 | 0.623533826 |
| CSTB | Beta_Cell_2_pPS | 4087 | -0.00431779 | [-0.04 0.03] | 0.7828968 | 0.955139045 |
| CSTB | Total_GRS | 4087 | 0.005875808 | [-0.02 0.04] | 0.707679332 | 0.937738119 |
| CTRC | Cholesterol_pPS | 4157 | 0.004719405 | [-0.03 0.04] | 0.761315876 | 0.950403646 |
| CTRC | Lipodystrophy_1_pPS | 4157 | 0.005505557 | [-0.02 0.04] | 0.723074554 | 0.942134248 |
| CTRC | Lipodystrophy_2_pPS | 4157 | 0.030398458 | [-0. 0.06] | 0.050351517 | 0.440215663 |
| CTRC | Liver-Lipid_pPS | 4157 | -0.047016699 | [-0.08 -0.02] | 0.002463613 | 0.090660952 |
| CTRC | ASAT_Pos_pPS | 4157 | -0.007776848 | [-0.04 0.02] | 0.61669276 | 0.909483337 |
| CTRC | ALP_Neg_pPS | 4157 | 0.114200489 | [0.08 0.14] | 1.65E-13 | 6.51E-11 |
| CTRC | SHBG-LpA_pPS | 4157 | -0.036948315 | [-0.07 -0.01] | 0.017364897 | 0.283592393 |
| CTRC | Obesity_pPS | 4157 | -0.00181853 | [-0.03 0.03] | 0.906824578 | 0.990045821 |
| CTRC | VAT_Neg_pPS | 4157 | -0.004561222 | [-0.04 0.03] | 0.769084936 | 0.952961063 |
| CTRC | Bilirubin_pPS | 4157 | -0.012357019 | [-0.04 0.02] | 0.426406444 | 0.82738773 |
| CTRC | Proinsulin_pPS | 4157 | 0.036133756 | [0.01 0.07] | 0.019996703 | 0.306616116 |
| CTRC | Hyper_Insulin_pPS | 4157 | 0.020556546 | [-0.01 0.05] | 0.185767057 | 0.68727393 |
| CTRC | Beta_Cell_1_pPS | 4157 | -0.002330303 | [-0.03 0.03] | 0.880777441 | 0.979035738 |
| CTRC | Beta_Cell_2_pPS | 4157 | 0.006563064 | [-0.02 0.04] | 0.672719843 | 0.925361755 |
| CTRC | Total_GRS | 4157 | 0.022086278 | [-0.01 0.05] | 0.15511449 | 0.650979658 |
| KLK6 | Cholesterol_pPS | 4225 | 0.018040283 | [-0.01 0.05] | 0.241719492 | 0.729918816 |
| KLK6 | Lipodystrophy_1_pPS | 4225 | -0.008818397 | [-0.04 0.02] | 0.567171238 | 0.899863296 |
| KLK6 | Lipodystrophy_2_pPS | 4225 | 0.0324179 | [0. 0.06] | 0.035369939 | 0.386023413 |
| KLK6 | Liver-Lipid_pPS | 4225 | 4.67E-05 | [-0.03 0.03] | 0.997583867 | 0.999464614 |
| KLK6 | ASAT_Pos_pPS | 4225 | 0.008076308 | [-0.02 0.04] | 0.60023194 | 0.907623171 |
| KLK6 | ALP_Neg_pPS | 4225 | 0.007164447 | [-0.02 0.04] | 0.642006513 | 0.916400472 |
| KLK6 | SHBG-LpA_pPS | 4225 | -0.004679408 | [-0.03 0.03] | 0.761401547 | 0.950403646 |
| KLK6 | Obesity_pPS | 4225 | -0.032449808 | [-0.06 -0. ] | 0.035189622 | 0.386023413 |
| KLK6 | VAT_Neg_pPS | 4225 | -0.022099424 | [-0.05 0.01] | 0.151523564 | 0.649125224 |
| KLK6 | Bilirubin_pPS | 4225 | -0.006694434 | [-0.04 0.02] | 0.664001087 | 0.924111434 |
| KLK6 | Proinsulin_pPS | 4225 | 0.007015647 | [-0.02 0.04] | 0.648936957 | 0.91896665 |
| KLK6 | Hyper_Insulin_pPS | 4225 | -0.004074412 | [-0.03 0.03] | 0.791484761 | 0.957693091 |
| KLK6 | Beta_Cell_1_pPS | 4225 | -0.003245976 | [-0.03 0.03] | 0.833178647 | 0.969057339 |
| KLK6 | Beta_Cell_2_pPS | 4225 | 0.038236987 | [0.01 0.07] | 0.013063142 | 0.242414912 |

**b) Partial Correlation**

| Proteomics | PPS | Sample_Size | r | CI95% | p | fdr_p_adjusted |
|---|---|---|---|---|---|---|
| PRSS8 | Obesity_pPS | 4159 | 0.048668198 | [0.02 0.08] | 0.001747539 | 0.093784569 |
| PRSS8 | VAT_Neg_pPS | 4159 | 0.008927563 | [-0.02 0.04] | 0.566071936 | 0.90516325 |
| PRSS8 | Bilirubin_pPS | 4159 | -0.024207305 | [-0.05 0.01] | 0.119661775 | 0.681046037 |
| PRSS8 | Proinsulin_pPS | 4159 | 0.008007189 | [-0.02 0.04] | 0.606773645 | 0.918301271 |
| PRSS8 | Hyper_Insulin_pPS | 4159 | 0.000773409 | [-0.03 0.03] | 0.960351716 | 0.996425456 |
| PRSS8 | Beta_Cell_1_pPS | 4159 | 0.0138934 | [-0.02 0.04] | 0.371822022 | 0.847284666 |
| PRSS8 | Beta_Cell_2_pPS | 4159 | -0.005924044 | [-0.04 0.02] | 0.703365329 | 0.945970482 |
| SERPINA12 | Cholesterol_pPS | 4159 | 0.016409292 | [-0.01 0.05] | 0.291513636 | 0.81655641 |
| SERPINA12 | Lipodystrophy_1_pPS | 4159 | 0.021665844 | [-0.01 0.05] | 0.163688323 | 0.718997225 |
| SERPINA12 | Lipodystrophy_2_pPS | 4159 | 0.040250359 | [0.01 0.07] | 0.00964759 | 0.248521918 |
| SERPINA12 | Liver-Lipid_pPS | 4159 | -0.002192924 | [-0.03 0.03] | 0.887906155 | 0.98038845 |
| SERPINA12 | ASAT_Pos_pPS | 4159 | 0.005765642 | [-0.02 0.04] | 0.710935619 | 0.945970482 |
| SERPINA12 | ALP_Neg_pPS | 4159 | 0.007515058 | [-0.02 0.04] | 0.629060028 | 0.929162059 |
| SERPINA12 | SHBG-LpA_pPS | 4159 | 0.021320552 | [-0.01 0.05] | 0.170508578 | 0.726214013 |
| SERPINA12 | Obesity_pPS | 4159 | -0.013682261 | [-0.04 0.02] | 0.379134364 | 0.85037326 |
| SERPINA12 | VAT_Neg_pPS | 4159 | 0.026006614 | [-0. 0.06] | 0.094543404 | 0.64003016 |
| SERPINA12 | Bilirubin_pPS | 4159 | -0.002555019 | [-0.03 0.03] | 0.869551219 | 0.977511154 |
| SERPINA12 | Proinsulin_pPS | 4159 | -0.011479532 | [-0.04 0.02] | 0.460580518 | 0.874967119 |
| SERPINA12 | Hyper_Insulin_pPS | 4159 | -0.017135824 | [-0.05 0.01] | 0.27067219 | 0.808820203 |
| SERPINA12 | Beta_Cell_1_pPS | 4159 | -0.018437784 | [-0.05 0.01] | 0.235929882 | 0.789292696 |
| SERPINA12 | Beta_Cell_2_pPS | 4159 | 0.003020862 | [-0.03 0.03] | 0.846041509 | 0.972947736 |
| TIMP4 | Cholesterol_pPS | 4224 | -0.008542761 | [-0.04 0.02] | 0.579980261 | 0.909583681 |
| TIMP4 | Lipodystrophy_1_pPS | 4224 | -0.03742703 | [-0.07 -0.01] | 0.015292049 | 0.317679992 |
| TIMP4 | Lipodystrophy_2_pPS | 4224 | 0.012682377 | [-0.02 0.04] | 0.411303187 | 0.854426619 |
| TIMP4 | Liver-Lipid_pPS | 4224 | 0.021898859 | [-0.01 0.05] | 0.155961135 | 0.708464493 |
| TIMP4 | ASAT_Pos_pPS | 4224 | -0.007067108 | [-0.04 0.02] | 0.647084771 | 0.935848959 |
| TIMP4 | ALP_Neg_pPS | 4224 | 0.01672922 | [-0.01 0.05] | 0.278452874 | 0.808820203 |
| TIMP4 | SHBG-LpA_pPS | 4224 | 0.013742607 | [-0.02 0.04] | 0.373308244 | 0.847284666 |
| TIMP4 | Obesity_pPS | 4224 | -0.011114043 | [-0.04 0.02] | 0.471528463 | 0.883720482 |
| TIMP4 | VAT_Neg_pPS | 4224 | -0.023554969 | [-0.05 0.01] | 0.126981972 | 0.694065456 |
| TIMP4 | Bilirubin_pPS | 4224 | 0.010247888 | [-0.02 0.04] | 0.506767296 | 0.895803896 |
| TIMP4 | Proinsulin_pPS | 4224 | 0.019303866 | [-0.01 0.05] | 0.211069988 | 0.767305935 |
| TIMP4 | Hyper_Insulin_pPS | 4224 | -0.000613799 | [-0.03 0.03] | 0.968282734 | 0.996924797 |
| TIMP4 | Beta_Cell_1_pPS | 4224 | -0.005490324 | [-0.04 0.02] | 0.722089172 | 0.946314812 |
| TIMP4 | Beta_Cell_2_pPS | 4224 | -0.008174675 | [-0.04 0.02] | 0.596411798 | 0.918301271 |
| ADAM_TS13 | Cholesterol_pPS | 4159 | 0.016981846 | [-0.01 0.05] | 0.275001708 | 0.808820203 |
| ADAM_TS13 | Lipodystrophy_1_pPS | 4159 | -0.01089517 | [-0.04 0.02] | 0.483723217 | 0.895803896 |
| ADAM_TS13 | Lipodystrophy_2_pPS | 4159 | -0.001963512 | [-0.03 0.03] | 0.899567057 | 0.985399384 |
| ADAM_TS13 | Liver-Lipid_pPS | 4159 | -0.000724027 | [-0.03 0.03] | 0.962881366 | 0.996924797 |
| ADAM_TS13 | ASAT_Pos_pPS | 4159 | -0.014576088 | [-0.05 0.02] | 0.348782585 | 0.847284666 |
| ADAM_TS13 | ALP_Neg_pPS | 4159 | -0.005530244 | [-0.04 0.02] | 0.722238435 | 0.946314812 |
| ADAM_TS13 | SHBG-LpA_pPS | 4159 | 0.030357896 | [-0. 0.06] | 0.050967113 | 0.537121778 |
| ADAM_TS13 | Obesity_pPS | 4159 | -0.001225593 | [-0.03 0.03] | 0.937209896 | 0.994163908 |
| ADAM_TS13 | VAT_Neg_pPS | 4159 | 0.011499024 | [-0.02 0.04] | 0.459819413 | 0.874172623 |
| ADAM_TS13 | Bilirubin_pPS | 4159 | 0.003870092 | [-0.03 0.03] | 0.803548108 | 0.967638637 |
| ADAM_TS13 | Proinsulin_pPS | 4159 | 0.00486963 | [-0.03 0.04] | 0.754276223 | 0.95462436 |
| ADAM_TS13 | Hyper_Insulin_pPS | 4159 | 0.000242819 | [-0.03 0.03] | 0.987547413 | 0.998803911 |
| ADAM_TS13 | Beta_Cell_1_pPS | 4159 | 0.012476618 | [-0.02 0.04] | 0.422560113 | 0.860518922 |
| ADAM_TS13 | Beta_Cell_2_pPS | 4159 | -0.02757275 | [-0.06 0. ] | 0.076291088 | 0.597342987 |
| PAR_1 | Cholesterol_pPS | 4159 | 0.005000957 | [-0.03 0.04] | 0.74787148 | 0.954135344 |
| PAR_1 | Lipodystrophy_1_pPS | 4159 | -0.012086987 | [-0.04 0.02] | 0.437193829 | 0.865276785 |
| PAR_1 | Lipodystrophy_2_pPS | 4159 | 0.025126282 | [-0.01 0.06] | 0.106247769 | 0.665922758 |
| PAR_1 | Liver-Lipid_pPS | 4159 | 0.01408394 | [-0.02 0.04] | 0.365298661 | 0.847284666 |
| PAR_1 | ASAT_Pos_pPS | 4159 | 0.002941915 | [-0.03 0.03] | 0.850016681 | 0.974254359 |
| PAR_1 | ALP_Neg_pPS | 4159 | -0.023171301 | [-0.05 0.01] | 0.136335808 | 0.694065456 |
| PAR_1 | SHBG-LpA_pPS | 4159 | 0.018056567 | [-0.01 0.05] | 0.245759043 | 0.79432283 |
| PAR_1 | Obesity_pPS | 4159 | 0.009264168 | [-0.02 0.04] | 0.55152042 | 0.899241939 |
| PAR_1 | VAT_Neg_pPS | 4159 | -0.009191523 | [-0.04 0.02] | 0.554645266 | 0.899898936 |
| PAR_1 | Bilirubin_pPS | 4159 | 0.012203252 | [-0.02 0.04] | 0.432796903 | 0.864060889 |
| PAR_1 | Proinsulin_pPS | 4159 | 0.01934664 | [-0.01 0.05] | 0.213626391 | 0.767305935 |
| PAR_1 | Hyper_Insulin_pPS | 4159 | -0.028334643 | [-0.06 0. ] | 0.068512004 | 0.571035672 |
| PAR_1 | Beta_Cell_1_pPS | 4159 | -0.007308732 | [-0.04 0.02] | 0.638506644 | 0.933760412 |
| PAR_1 | Beta_Cell_2_pPS | 4159 | 0.007918134 | [-0.02 0.04] | 0.610780368 | 0.919490897 |
| GP6 | Cholesterol_pPS | 4225 | 0.000116118 | [-0.03 0.03] | 0.993997501 | 0.998803911 |
| GP6 | Lipodystrophy_1_pPS | 4225 | 0.005063258 | [-0.03 0.04] | 0.742880619 | 0.95254379 |
| GP6 | Lipodystrophy_2_pPS | 4225 | 0.017997372 | [-0.01 0.05] | 0.243569876 | 0.793218712 |
| GP6 | Liver-Lipid_pPS | 4225 | 0.002130588 | [-0.03 0.03] | 0.890211234 | 0.981230818 |
| GP6 | ASAT_Pos_pPS | 4225 | 0.028964716 | [-0. 0.06] | 0.060523138 | 0.558815902 |
| GP6 | ALP_Neg_pPS | 4225 | -0.025921377 | [-0.06 0. ] | 0.093019705 | 0.638855661 |
| GP6 | SHBG-LpA_pPS | 4225 | -0.005468727 | [-0.04 0.02] | 0.723105712 | 0.946504225 |
| GP6 | Obesity_pPS | 4225 | -0.011498841 | [-0.04 0.02] | 0.456263944 | 0.874172623 |
| GP6 | VAT_Neg_pPS | 4225 | -0.011794217 | [-0.04 0.02] | 0.444776847 | 0.868646822 |
| GP6 | Bilirubin_pPS | 4225 | 0.004968453 | [-0.03 0.04] | 0.747529428 | 0.954135344 |
| GP6 | Proinsulin_pPS | 4225 | 0.021458733 | [-0.01 0.05] | 0.164398046 | 0.718997225 |
| GP6 | Hyper_Insulin_pPS | 4225 | -0.012208101 | [-0.04 0.02] | 0.428961901 | 0.864060889 |
| GP6 | Beta_Cell_1_pPS | 4225 | -0.011993332 | [-0.04 0.02] | 0.437127296 | 0.865276785 |
| GP6 | Beta_Cell_2_pPS | 4225 | 0.008339571 | [-0.02 0.04] | 0.588980147 | 0.913889104 |
| MMP12 | Cholesterol_pPS | 4159 | -0.019679609 | [-0.05 0.01] | 0.205848483 | 0.765145476 |
| MMP12 | Lipodystrophy_1_pPS | 4159 | 0.022661685 | [-0.01 0.05] | 0.145169826 | 0.697681852 |
| MMP12 | Lipodystrophy_2_pPS | 4159 | 0.012053864 | [-0.02 0.04] | 0.438451166 | 0.865276785 |
| MMP12 | Liver-Lipid_pPS | 4159 | -0.016506736 | [-0.05 0.01] | 0.288657372 | 0.81655641 |
| MMP12 | ASAT_Pos_pPS | 4159 | -0.021233333 | [-0.05 0.01] | 0.172264547 | 0.727559563 |
| MMP12 | ALP_Neg_pPS | 4159 | 0.023329364 | [-0.01 0.05] | 0.133681838 | 0.694065456 |
| MMP12 | SHBG-LpA_pPS | 4159 | 0.013419579 | [-0.02 0.04] | 0.388354279 | 0.85037326 |
| MMP12 | Obesity_pPS | 4159 | 0.008627762 | [-0.02 0.04] | 0.579185535 | 0.909583681 |
| MMP12 | VAT_Neg_pPS | 4159 | -0.001768764 | [-0.03 0.03] | 0.909483136 | 0.988841471 |
| MMP12 | Bilirubin_pPS | 4159 | -0.012189116 | [-0.04 0.02] | 0.433330158 | 0.864060889 |
| MMP12 | Proinsulin_pPS | 4159 | -0.011758276 | [-0.04 0.02] | 0.449763198 | 0.870351501 |
| MMP12 | Hyper_Insulin_pPS | 4159 | 0.016365548 | [-0.01 0.05] | 0.292802011 | 0.816672929 |
| MMP12 | Beta_Cell_1_pPS | 4159 | 0.014724992 | [-0.02 0.05] | 0.343880422 | 0.847284666 |
| MMP12 | Beta_Cell_2_pPS | 4159 | 0.00241009 | [-0.03 0.03] | 0.876889917 | 0.97781424 |
| MMP7 | Cholesterol_pPS | 4159 | -0.002894282 | [-0.03 0.03] | 0.852417009 | 0.974979087 |
| MMP7 | Lipodystrophy_1_pPS | 4159 | 0.005846032 | [-0.02 0.04] | 0.707090034 | 0.945970482 |
| MMP7 | Lipodystrophy_2_pPS | 4159 | 0.045273611 | [0.01 0.08] | 0.003596596 | 0.137844373 |
| MMP7 | Liver-Lipid_pPS | 4159 | -0.036268818 | [-0.07 -0.01] | 0.019700228 | 0.342890447 |
| MMP7 | ASAT_Pos_pPS | 4159 | 0.010327491 | [-0.02 0.04] | 0.506795795 | 0.895803896 |
| MMP7 | ALP_Neg_pPS | 4159 | -0.020737145 | [-0.05 0.01] | 0.182512259 | 0.738879686 |
| MMP7 | SHBG-LpA_pPS | 4159 | 0.01248698 | [-0.02 0.04] | 0.422174914 | 0.860518922 |
| MMP7 | Obesity_pPS | 4159 | -0.001245325 | [-0.03 0.03] | 0.936201073 | 0.994163908 |
| MMP7 | VAT_Neg_pPS | 4159 | 0.040562338 | [0.01 0.07] | 0.009099624 | 0.248521918 |
| MMP7 | Bilirubin_pPS | 4159 | 0.007261283 | [-0.02 0.04] | 0.640687487 | 0.933976497 |
| MMP7 | Proinsulin_pPS | 4159 | -0.013023871 | [-0.04 0.02] | 0.402498404 | 0.85241904 |
| MMP7 | Hyper_Insulin_pPS | 4159 | 0.000278358 | [-0.03 0.03] | 0.985725071 | 0.998803911 |
| MMP7 | Beta_Cell_1_pPS | 4159 | 0.04138138 | [0.01 0.07] | 0.007790891 | 0.21814494 |
| MMP7 | Beta_Cell_2_pPS | 4159 | -0.016966579 | [-0.05 0.01] | 0.275433543 | 0.808820203 |
| uPA | Cholesterol_pPS | 4225 | -0.013283734 | [-0.04 0.02] | 0.389421973 | 0.85037326 |
| uPA | Lipodystrophy_1_pPS | 4225 | 0.016196754 | [-0.01 0.05] | 0.293982089 | 0.817312323 |

**Table S7: Associations between pPSs and plasma proteins in SCAPIS**

**a) Spearman Correlation**

| Proteomics | PPS | Sample_Size | r | CI95% | p | fdr_p_adjusted |
|---|---|---|---|---|---|---|
| KLK6 | Total_GRS | 4225 | 0.011066836 | [-0.02 0.04] | 0.472676602 | 0.846453261 |
| MERTK | Cholesterol_pPS | 4159 | 0.016019011 | [-0.01 0.05] | 0.302383894 | 0.76319014 |
| MERTK | Lipodystrophy_1_pPS | 4159 | -0.001056174 | [-0.03 0.03] | 0.945790261 | 0.993154165 |
| MERTK | Lipodystrophy_2_pPS | 4159 | 0.016373683 | [-0.01 0.05] | 0.291804093 | 0.753421877 |
| MERTK | Liver-Lipid_pPS | 4159 | -0.015705817 | [-0.05 0.01] | 0.311935933 | 0.76365449 |
| MERTK | ASAT_Pos_pPS | 4159 | -0.026903499 | [-0.06 0. ] | 0.083219915 | 0.534925551 |
| MERTK | ALP_Neg_pPS | 4159 | -0.059503521 | [-0.09 -0.03] | 0.000126024 | 0.012882403 |
| MERTK | SHBG-LpA_pPS | 4159 | 0.015427063 | [-0.02 0.05] | 0.320603057 | 0.770109076 |
| MERTK | Obesity_pPS | 4159 | 0.022281424 | [-0.01 0.05] | 0.151399141 | 0.649125224 |
| MERTK | VAT_Neg_pPS | 4159 | -0.004466183 | [-0.03 0.03] | 0.773710798 | 0.953322233 |
| MERTK | Bilirubin_pPS | 4159 | 0.028427353 | [-0. 0.06] | 0.067181166 | 0.477886642 |
| MERTK | Proinsulin_pPS | 4159 | 0.01761189 | [-0.01 0.05] | 0.25683545 | 0.740941247 |
| MERTK | Hyper_Insulin_pPS | 4159 | -0.015687884 | [-0.05 0.01] | 0.31248882 | 0.76365449 |
| MERTK | Beta_Cell_1_pPS | 4159 | 0.001410284 | [-0.03 0.03] | 0.927658658 | 0.990056084 |
| MERTK | Beta_Cell_2_pPS | 4159 | 0.006419226 | [-0.02 0.04] | 0.679416226 | 0.925579558 |
| MERTK | Total_GRS | 4159 | -0.005246684 | [-0.04 0.03] | 0.735534831 | 0.942470055 |
| PI3 | Cholesterol_pPS | 3943 | 0.012301099 | [-0.02 0.04] | 0.440686564 | 0.831350198 |
| PI3 | Lipodystrophy_1_pPS | 3943 | 0.029612967 | [-0. 0.06] | 0.063385002 | 0.473548288 |
| PI3 | Lipodystrophy_2_pPS | 3943 | 0.017652606 | [-0.01 0.05] | 0.268505076 | 0.746520328 |
| PI3 | Liver-Lipid_pPS | 3943 | -0.00659571 | [-0.04 0.02] | 0.679307178 | 0.925579558 |
| PI3 | ASAT_Pos_pPS | 3943 | 0.008641751 | [-0.02 0.04] | 0.588055764 | 0.904197163 |
| PI3 | ALP_Neg_pPS | 3943 | -0.010360607 | [-0.04 0.02] | 0.516081848 | 0.870853148 |
| PI3 | SHBG-LpA_pPS | 3943 | 0.004863032 | [-0.03 0.04] | 0.760514162 | 0.950403646 |
| PI3 | Obesity_pPS | 3943 | 0.000834414 | [-0.03 0.03] | 0.958290269 | 0.99612031 |
| PI3 | VAT_Neg_pPS | 3943 | 0.004495383 | [-0.03 0.04] | 0.778126103 | 0.953637204 |
| PI3 | Bilirubin_pPS | 3943 | 0.033737219 | [0. 0.06] | 0.034415039 | 0.383006082 |
| PI3 | Proinsulin_pPS | 3943 | -0.014917553 | [-0.05 0.02] | 0.349762205 | 0.793215848 |
| PI3 | Hyper_Insulin_pPS | 3943 | 0.005667535 | [-0.03 0.04] | 0.722416392 | 0.942134248 |
| PI3 | Beta_Cell_1_pPS | 3943 | -0.006417729 | [-0.04 0.02] | 0.6874979 | 0.926969309 |
| PI3 | Beta_Cell_2_pPS | 3943 | -0.004409414 | [-0.04 0.03] | 0.78226122 | 0.955139045 |
| PI3 | Total_GRS | 3943 | 0.009691623 | [-0.02 0.04] | 0.54354346 | 0.886924725 |
| PRSS27 | Cholesterol_pPS | 4158 | -0.008471842 | [-0.04 0.02] | 0.585517992 | 0.903925569 |
| PRSS27 | Lipodystrophy_1_pPS | 4158 | -0.008506227 | [-0.04 0.02] | 0.583996836 | 0.903823705 |
| PRSS27 | Lipodystrophy_2_pPS | 4158 | 0.003342854 | [-0.03 0.03] | 0.829626836 | 0.966693035 |
| PRSS27 | Liver-Lipid_pPS | 4158 | 0.015045011 | [-0.02 0.05] | 0.332793457 | 0.779058474 |
| PRSS27 | ASAT_Pos_pPS | 4158 | -0.010003465 | [-0.04 0.02] | 0.519614713 | 0.872596104 |
| PRSS27 | ALP_Neg_pPS | 4158 | -0.014530434 | [-0.04 0.02] | 0.349595706 | 0.793215848 |
| PRSS27 | SHBG-LpA_pPS | 4158 | 0.006181967 | [-0.02 0.04] | 0.690676525 | 0.928527622 |
| PRSS27 | Obesity_pPS | 4158 | -0.022178693 | [-0.05 0.01] | 0.153344075 | 0.649125224 |
| PRSS27 | VAT_Neg_pPS | 4158 | -0.000831384 | [-0.03 0.03] | 0.957320596 | 0.99612031 |
| PRSS27 | Bilirubin_pPS | 4158 | -0.026593413 | [-0.06 0. ] | 0.0868743 | 0.54140702 |
| PRSS27 | Proinsulin_pPS | 4158 | -0.022713005 | [-0.05 0.01] | 0.143679489 | 0.641415816 |
| PRSS27 | Hyper_Insulin_pPS | 4158 | 0.005287886 | [-0.03 0.04] | 0.733567572 | 0.942134248 |
| PRSS27 | Beta_Cell_1_pPS | 4158 | 0.018379854 | [-0.01 0.05] | 0.236725295 | 0.726631332 |
| PRSS27 | Beta_Cell_2_pPS | 4158 | 0.022232179 | [-0.01 0.05] | 0.152354916 | 0.649125224 |
| PRSS27 | Total_GRS | 4158 | -0.017068334 | [-0.05 0.01] | 0.271868786 | 0.746520328 |
| PRSS8 | Cholesterol_pPS | 4159 | 0.001572832 | [-0.03 0.03] | 0.919347628 | 0.990056084 |
| PRSS8 | Lipodystrophy_1_pPS | 4159 | 0.041113893 | [0.01 0.07] | 0.008098361 | 0.197592038 |
| PRSS8 | Lipodystrophy_2_pPS | 4159 | 0.050882491 | [0.02 0.08] | 0.0010461 | 0.050992277 |
| PRSS8 | Liver-Lipid_pPS | 4159 | -0.035109613 | [-0.07 -0. ] | 0.023760721 | 0.327897946 |
| PRSS8 | ASAT_Pos_pPS | 4159 | 0.00646573 | [-0.02 0.04] | 0.677224301 | 0.925579558 |
| PRSS8 | ALP_Neg_pPS | 4159 | -0.001621504 | [-0.03 0.03] | 0.916860698 | 0.990056084 |
| PRSS8 | SHBG-LpA_pPS | 4159 | 0.015728109 | [-0.01 0.05] | 0.311249574 | 0.76365449 |
| PRSS8 | Obesity_pPS | 4159 | 0.050487218 | [0.02 0.08] | 0.001144632 | 0.051789895 |
| PRSS8 | VAT_Neg_pPS | 4159 | 0.017750929 | [-0.01 0.05] | 0.253098523 | 0.739680398 |
| PRSS8 | Bilirubin_pPS | 4159 | -0.021745639 | [-0.05 0.01] | 0.16148284 | 0.664061293 |
| PRSS8 | Proinsulin_pPS | 4159 | 0.017204451 | [-0.01 0.05] | 0.268005862 | 0.746520328 |
| PRSS8 | Hyper_Insulin_pPS | 4159 | 0.010539359 | [-0.02 0.04] | 0.497442347 | 0.863780476 |
| PRSS8 | Beta_Cell_1_pPS | 4159 | 0.015196065 | [-0.02 0.05] | 0.327903438 | 0.775125841 |
| PRSS8 | Beta_Cell_2_pPS | 4159 | 0.000173623 | [-0.03 0.03] | 0.991081859 | 0.999402232 |
| PRSS8 | Total_GRS | 4159 | 0.060421363 | [0.03 0.09] | 9.89E-05 | 0.010651153 |
| SERPINA12 | Cholesterol_pPS | 4159 | 0.014975147 | [-0.02 0.05] | 0.334985301 | 0.780877897 |
| SERPINA12 | Lipodystrophy_1_pPS | 4159 | 0.025316459 | [-0.01 0.06] | 0.103084809 | 0.586842904 |
| SERPINA12 | Lipodystrophy_2_pPS | 4159 | 0.046037898 | [0.02 0.08] | 0.003022989 | 0.105623173 |
| SERPINA12 | Liver-Lipid_pPS | 4159 | -0.001421048 | [-0.03 0.03] | 0.927108049 | 0.990056084 |
| SERPINA12 | ASAT_Pos_pPS | 4159 | 0.011052767 | [-0.02 0.04] | 0.47672832 | 0.847790054 |
| SERPINA12 | ALP_Neg_pPS | 4159 | 0.00478867 | [-0.03 0.04] | 0.757864887 | 0.950403646 |
| SERPINA12 | SHBG-LpA_pPS | 4159 | 0.024035333 | [-0.01 0.05] | 0.121727012 | 0.615837655 |
| SERPINA12 | Obesity_pPS | 4159 | -0.014142907 | [-0.04 0.02] | 0.362540609 | 0.7979754 |
| SERPINA12 | VAT_Neg_pPS | 4159 | 0.028976002 | [-0. 0.06] | 0.062068963 | 0.468061031 |
| SERPINA12 | Bilirubin_pPS | 4159 | -0.002643158 | [-0.03 0.03] | 0.864883553 | 0.977006629 |
| SERPINA12 | Proinsulin_pPS | 4159 | -0.015836312 | [-0.05 0.01] | 0.307932107 | 0.76365449 |
| SERPINA12 | Hyper_Insulin_pPS | 4159 | -0.009152477 | [-0.04 0.02] | 0.555707943 | 0.896216632 |
| SERPINA12 | Beta_Cell_1_pPS | 4159 | -0.020556622 | [-0.05 0.01] | 0.185659254 | 0.68727393 |
| SERPINA12 | Beta_Cell_2_pPS | 4159 | -0.001808794 | [-0.03 0.03] | 0.907298873 | 0.990056084 |
| SERPINA12 | Total_GRS | 4159 | 0.008170863 | [-0.02 0.04] | 0.598866731 | 0.907623171 |
| TIMP4 | Cholesterol_pPS | 4224 | -0.007625483 | [-0.04 0.02] | 0.620774079 | 0.910907479 |
| TIMP4 | Lipodystrophy_1_pPS | 4224 | -0.038567666 | [-0.07 -0.01] | 0.012306661 | 0.237527156 |
| TIMP4 | Lipodystrophy_2_pPS | 4224 | 0.007681071 | [-0.02 0.04] | 0.618230128 | 0.909483337 |
| TIMP4 | Liver-Lipid_pPS | 4224 | 0.021858732 | [-0.01 0.05] | 0.156080314 | 0.653689932 |
| TIMP4 | ASAT_Pos_pPS | 4224 | -0.012910388 | [-0.04 0.02] | 0.402216393 | 0.81230319 |
| TIMP4 | ALP_Neg_pPS | 4224 | 0.01980912 | [-0.01 0.05] | 0.198668287 | 0.700947446 |
| TIMP4 | SHBG-LpA_pPS | 4224 | 0.010041853 | [-0.02 0.04] | 0.514698939 | 0.870853148 |
| TIMP4 | Obesity_pPS | 4224 | -0.010980744 | [-0.04 0.02] | 0.47618036 | 0.84751951 |
| TIMP4 | VAT_Neg_pPS | 4224 | -0.022962331 | [-0.05 0.01] | 0.136222226 | 0.629316669 |
| TIMP4 | Bilirubin_pPS | 4224 | 0.008228853 | [-0.02 0.04] | 0.59340901 | 0.905925461 |
| TIMP4 | Proinsulin_pPS | 4224 | 0.01943399 | [-0.01 0.05] | 0.207305545 | 0.709689105 |
| TIMP4 | Hyper_Insulin_pPS | 4224 | -0.006581955 | [-0.04 0.02] | 0.669345648 | 0.925084621 |
| TIMP4 | Beta_Cell_1_pPS | 4224 | -0.005528772 | [-0.04 0.02] | 0.719809691 | 0.941449667 |
| TIMP4 | Beta_Cell_2_pPS | 4224 | -0.007106598 | [-0.04 0.02] | 0.644736474 | 0.916568313 |
| TIMP4 | Total_GRS | 4224 | -0.027693123 | [-0.06 0. ] | 0.072320896 | 0.501521789 |
| ADAM_TS13 | Cholesterol_pPS | 4159 | 0.015787539 | [-0.01 0.05] | 0.309424563 | 0.76365449 |
| ADAM_TS13 | Lipodystrophy_1_pPS | 4159 | -0.010798052 | [-0.04 0.02] | 0.486945623 | 0.855691721 |
| ADAM_TS13 | Lipodystrophy_2_pPS | 4159 | -0.002606713 | [-0.03 0.03] | 0.866729043 | 0.977006629 |
| ADAM_TS13 | Liver-Lipid_pPS | 4159 | -0.000793313 | [-0.03 0.03] | 0.959268344 | 0.99612031 |
| ADAM_TS13 | ASAT_Pos_pPS | 4159 | -0.016043459 | [-0.05 0.01] | 0.301646516 | 0.76319014 |
| ADAM_TS13 | ALP_Neg_pPS | 4159 | -0.006018954 | [-0.04 0.02] | 0.69839313 | 0.932091412 |
| ADAM_TS13 | SHBG-LpA_pPS | 4159 | 0.030091172 | [-0. 0.06] | 0.052666925 | 0.441027526 |
| ADAM_TS13 | Obesity_pPS | 4159 | -0.000476827 | [-0.03 0.03] | 0.975511144 | 0.996862516 |
| ADAM_TS13 | VAT_Neg_pPS | 4159 | 0.011717748 | [-0.02 0.04] | 0.450615904 | 0.831350198 |
| ADAM_TS13 | Bilirubin_pPS | 4159 | 0.00310342 | [-0.03 0.03] | 0.841643423 | 0.972342391 |
| ADAM_TS13 | Proinsulin_pPS | 4159 | 0.004825542 | [-0.03 0.04] | 0.756059476 | 0.950403646 |
| ADAM_TS13 | Hyper_Insulin_pPS | 4159 | -0.0013407 | [-0.03 0.03] | 0.931218912 | 0.990056084 |
| ADAM_TS13 | Beta_Cell_1_pPS | 4159 | 0.008063364 | [-0.02 0.04] | 0.603684017 | 0.908869305 |
| ADAM_TS13 | Beta_Cell_2_pPS | 4159 | -0.022609895 | [-0.05 0.01] | 0.145458646 | 0.641415816 |
| ADAM_TS13 | Total_GRS | 4159 | 0.004063427 | [-0.03 0.03] | 0.793633284 | 0.958013595 |
| PAR_1 | Cholesterol_pPS | 4159 | 0.007014898 | [-0.02 0.04] | 0.651550526 | 0.919948884 |

**b) Partial Correlation**

| Proteomics | PPS | Sample_Size | r | CI95% | p | fdr_p_adjusted |
|---|---|---|---|---|---|---|
| uPA | Lipodystrophy_2_pPS | 4225 | 0.02146555 | [-0.01 0.05] | 0.164263996 | 0.718997225 |
| uPA | Liver-Lipid_pPS | 4225 | 0.026543476 | [-0. 0.06] | 0.085431559 | 0.614725404 |
| uPA | ASAT_Pos_pPS | 4225 | 0.014055565 | [-0.02 0.04] | 0.36246476 | 0.847284666 |
| uPA | ALP_Neg_pPS | 4225 | -0.015711727 | [-0.05 0.01] | 0.308679853 | 0.830335001 |
| uPA | SHBG-LpA_pPS | 4225 | 0.011065843 | [-0.02 0.04] | 0.473400582 | 0.88406027 |
| uPA | Obesity_pPS | 4225 | 0.003153052 | [-0.03 0.03] | 0.838134048 | 0.972099644 |
| uPA | VAT_Neg_pPS | 4225 | -0.013570425 | [-0.04 0.02] | 0.379269657 | 0.85037326 |
| uPA | Bilirubin_pPS | 4225 | -0.014849694 | [-0.05 0.02] | 0.335980918 | 0.844649909 |
| uPA | Proinsulin_pPS | 4225 | 0.018340741 | [-0.01 0.05] | 0.234690827 | 0.788664502 |
| uPA | Hyper_Insulin_pPS | 4225 | -0.012911297 | [-0.04 0.02] | 0.402854847 | 0.85241904 |
| uPA | Beta_Cell_1_pPS | 4225 | -0.01431769 | [-0.04 0.02] | 0.353582046 | 0.847284666 |
| uPA | Beta_Cell_2_pPS | 4225 | 0.045127525 | [0.01 0.08] | 0.003442155 | 0.134348358 |
| CD93 | Cholesterol_pPS | 4225 | 0.009551665 | [-0.02 0.04] | 0.53601615 | 0.898959051 |
| CD93 | Lipodystrophy_1_pPS | 4225 | 0.006360207 | [-0.02 0.04] | 0.680288134 | 0.943176659 |
| CD93 | Lipodystrophy_2_pPS | 4225 | 0.012442493 | [-0.02 0.04] | 0.420152406 | 0.859660522 |
| CD93 | Liver-Lipid_pPS | 4225 | 0.02426949 | [-0.01 0.05] | 0.115810035 | 0.681046037 |
| CD93 | ASAT_Pos_pPS | 4225 | -0.012264835 | [-0.04 0.02] | 0.426819774 | 0.864060889 |
| CD93 | ALP_Neg_pPS | 4225 | 0.014135021 | [-0.02 0.04] | 0.359757558 | 0.847284666 |
| CD93 | SHBG-LpA_pPS | 4225 | 0.015007931 | [-0.02 0.05] | 0.330856386 | 0.844649909 |
| CD93 | Obesity_pPS | 4225 | 0.013424952 | [-0.02 0.04] | 0.38440066 | 0.85037326 |
| CD93 | VAT_Neg_pPS | 4225 | -0.029998355 | [-0.06 0. ] | 0.051898542 | 0.540728413 |
| CD93 | Bilirubin_pPS | 4225 | -0.015914821 | [-0.05 0.01] | 0.302467397 | 0.826182442 |
| CD93 | Proinsulin_pPS | 4225 | 0.017327043 | [-0.01 0.05] | 0.261577353 | 0.807171938 |
| CD93 | Hyper_Insulin_pPS | 4225 | 0.017322822 | [-0.01 0.05] | 0.261693588 | 0.807171938 |
| CD93 | Beta_Cell_1_pPS | 4225 | 0.00624635 | [-0.02 0.04] | 0.685703027 | 0.944421056 |
| CD93 | Beta_Cell_2_pPS | 4225 | 0.032267078 | [0. 0.06] | 0.036521998 | 0.454495981 |
| CEACAM8 | Cholesterol_pPS | 4159 | -0.016907604 | [-0.05 0.01] | 0.277106003 | 0.808820203 |
| CEACAM8 | Lipodystrophy_1_pPS | 4159 | -0.000225729 | [-0.03 0.03] | 0.98842381 | 0.998803911 |
| CEACAM8 | Lipodystrophy_2_pPS | 4159 | 0.008610243 | [-0.02 0.04] | 0.579956212 | 0.909583681 |
| CEACAM8 | Liver-Lipid_pPS | 4159 | 0.0183836 | [-0.01 0.05] | 0.23730974 | 0.78980606 |
| CEACAM8 | ASAT_Pos_pPS | 4159 | -0.010053188 | [-0.04 0.02] | 0.518148202 | 0.895803896 |
| CEACAM8 | ALP_Neg_pPS | 4159 | 0.006840149 | [-0.02 0.04] | 0.660177835 | 0.937489141 |
| CEACAM8 | SHBG-LpA_pPS | 4159 | 0.0059377 | [-0.02 0.04] | 0.702714041 | 0.945970482 |
| CEACAM8 | Obesity_pPS | 4159 | 0.015754394 | [-0.01 0.05] | 0.311201557 | 0.833007217 |
| CEACAM8 | VAT_Neg_pPS | 4159 | -0.009714247 | [-0.04 0.02] | 0.532355122 | 0.898959051 |
| CEACAM8 | Bilirubin_pPS | 4159 | 0.023298588 | [-0.01 0.05] | 0.134195434 | 0.694065456 |
| CEACAM8 | Proinsulin_pPS | 4159 | 0.036872423 | [0.01 0.07] | 0.017747447 | 0.33440804 |
| CEACAM8 | Hyper_Insulin_pPS | 4159 | -0.031789182 | [-0.06 -0. ] | 0.040971165 | 0.4797351 |
| CEACAM8 | Beta_Cell_1_pPS | 4159 | -0.003415607 | [-0.03 0.03] | 0.826225746 | 0.970677755 |
| CEACAM8 | Beta_Cell_2_pPS | 4159 | -0.048882818 | [-0.08 -0.02] | 0.001667079 | 0.093356425 |
| DLK_1 | Cholesterol_pPS | 4225 | 0.012972621 | [-0.02 0.04] | 0.400624167 | 0.85241904 |
| DLK_1 | Lipodystrophy_1_pPS | 4225 | 0.019583876 | [-0.01 0.05] | 0.204468128 | 0.765145476 |
| DLK_1 | Lipodystrophy_2_pPS | 4225 | 0.01764843 | [-0.01 0.05] | 0.25283205 | 0.799790128 |
| DLK_1 | Liver-Lipid_pPS | 4225 | 0.044021515 | [0.01 0.07] | 0.004324675 | 0.154727258 |
| DLK_1 | ASAT_Pos_pPS | 4225 | 0.010100779 | [-0.02 0.04] | 0.512836674 | 0.895803896 |
| DLK_1 | ALP_Neg_pPS | 4225 | -0.010218406 | [-0.04 0.02] | 0.507940118 | 0.895803896 |
| DLK_1 | SHBG-LpA_pPS | 4225 | -0.002895275 | [-0.03 0.03] | 0.851206075 | 0.974635716 |
| DLK_1 | Obesity_pPS | 4225 | 0.015054309 | [-0.02 0.05] | 0.329364061 | 0.84456682 |
| DLK_1 | VAT_Neg_pPS | 4225 | 0.000649195 | [-0.03 0.03] | 0.96645077 | 0.996924797 |
| DLK_1 | Bilirubin_pPS | 4225 | -0.008806704 | [-0.04 0.02] | 0.568283815 | 0.905613993 |
| DLK_1 | Proinsulin_pPS | 4225 | -0.022133319 | [-0.05 0.01] | 0.151528974 | 0.706166997 |
| DLK_1 | Hyper_Insulin_pPS | 4225 | 0.011707502 | [-0.02 0.04] | 0.448131961 | 0.870351501 |
| DLK_1 | Beta_Cell_1_pPS | 4225 | 0.022353074 | [-0.01 0.05] | 0.147506748 | 0.703126624 |
| DLK_1 | Beta_Cell_2_pPS | 4225 | 0.013203935 | [-0.02 0.04] | 0.392276956 | 0.85037326 |
| Ep_CAM | Cholesterol_pPS | 4225 | -0.005350507 | [-0.04 0.02] | 0.728852976 | 0.946981479 |
| Ep_CAM | Lipodystrophy_1_pPS | 4225 | 0.002529422 | [-0.03 0.03] | 0.869828187 | 0.977511154 |
| Ep_CAM | Lipodystrophy_2_pPS | 4225 | 0.005865246 | [-0.02 0.04] | 0.703944303 | 0.945970482 |
| Ep_CAM | Liver-Lipid_pPS | 4225 | 0.002134076 | [-0.03 0.03] | 0.890032616 | 0.981230818 |
| Ep_CAM | ASAT_Pos_pPS | 4225 | 0.009534903 | [-0.02 0.04] | 0.536731935 | 0.898959051 |
| Ep_CAM | ALP_Neg_pPS | 4225 | -0.004881672 | [-0.04 0.03] | 0.751792848 | 0.954135344 |
| Ep_CAM | SHBG-LpA_pPS | 4225 | -0.010234503 | [-0.04 0.02] | 0.507271914 | 0.895803896 |
| Ep_CAM | Obesity_pPS | 4225 | -0.004420636 | [-0.03 0.03] | 0.774567128 | 0.962510816 |
| Ep_CAM | VAT_Neg_pPS | 4225 | -0.018542791 | [-0.05 0.01] | 0.229574453 | 0.786988264 |
| Ep_CAM | Bilirubin_pPS | 4225 | -0.00959533 | [-0.04 0.02] | 0.534153859 | 0.898959051 |
| Ep_CAM | Proinsulin_pPS | 4225 | 0.008910409 | [-0.02 0.04] | 0.563736745 | 0.904956448 |
| Ep_CAM | Hyper_Insulin_pPS | 4225 | 0.009312356 | [-0.02 0.04] | 0.546280462 | 0.899241939 |
| Ep_CAM | Beta_Cell_1_pPS | 4225 | -0.014853085 | [-0.05 0.02] | 0.335870568 | 0.844649909 |
| Ep_CAM | Beta_Cell_2_pPS | 4225 | -0.000543988 | [-0.03 0.03] | 0.971885216 | 0.997444743 |
| KIM1 | Cholesterol_pPS | 4159 | -0.014911883 | [-0.05 0.02] | 0.337790215 | 0.844649909 |
| KIM1 | Lipodystrophy_1_pPS | 4159 | 0.005579148 | [-0.02 0.04] | 0.719885164 | 0.946314812 |
| KIM1 | Lipodystrophy_2_pPS | 4159 | 0.01861139 | [-0.01 0.05] | 0.231547061 | 0.786988264 |
| KIM1 | Liver-Lipid_pPS | 4159 | -0.012210348 | [-0.04 0.02] | 0.432529403 | 0.864060889 |
| KIM1 | ASAT_Pos_pPS | 4159 | 0.009905185 | [-0.02 0.04] | 0.524327647 | 0.897453833 |
| KIM1 | ALP_Neg_pPS | 4159 | -0.022280082 | [-0.05 0.01] | 0.15206686 | 0.70648128 |
| KIM1 | SHBG-LpA_pPS | 4159 | -0.008080667 | [-0.04 0.02] | 0.603476644 | 0.918301271 |
| KIM1 | Obesity_pPS | 4159 | 0.01281951 | [-0.02 0.04] | 0.409922377 | 0.854336605 |
| KIM1 | VAT_Neg_pPS | 4159 | -0.017875965 | [-0.05 0.01] | 0.250514479 | 0.799790128 |
| KIM1 | Bilirubin_pPS | 4159 | -0.008190646 | [-0.04 0.02] | 0.598556918 | 0.918301271 |
| KIM1 | Proinsulin_pPS | 4159 | 0.019356537 | [-0.01 0.05] | 0.213392203 | 0.767305935 |
| KIM1 | Hyper_Insulin_pPS | 4159 | 0.000686789 | [-0.03 0.03] | 0.964789186 | 0.996924797 |
| KIM1 | Beta_Cell_1_pPS | 4159 | 0.00080363 | [-0.03 0.03] | 0.958803825 | 0.996425456 |
| KIM1 | Beta_Cell_2_pPS | 4159 | 0.000638004 | [-0.03 0.03] | 0.967288862 | 0.996924797 |
| IL_18BP | Cholesterol_pPS | 4225 | 0.01365398 | [-0.02 0.04] | 0.376341717 | 0.849453763 |
| IL_18BP | Lipodystrophy_1_pPS | 4225 | 0.02509599 | [-0.01 0.06] | 0.103911795 | 0.662566294 |
| IL_18BP | Lipodystrophy_2_pPS | 4225 | 0.018394308 | [-0.01 0.05] | 0.233326583 | 0.786988264 |
| IL_18BP | Liver-Lipid_pPS | 4225 | 0.013822656 | [-0.02 0.04] | 0.370473589 | 0.847284666 |
| IL_18BP | ASAT_Pos_pPS | 4225 | -0.009083255 | [-0.04 0.02] | 0.556197316 | 0.899977567 |
| IL_18BP | ALP_Neg_pPS | 4225 | -0.019395924 | [-0.05 0.01] | 0.208846505 | 0.767305935 |
| IL_18BP | SHBG-LpA_pPS | 4225 | 0.006378378 | [-0.02 0.04] | 0.679425441 | 0.943176659 |
| IL_18BP | Obesity_pPS | 4225 | 0.01984231 | [-0.01 0.05] | 0.198557397 | 0.763408738 |
| IL_18BP | VAT_Neg_pPS | 4225 | -0.008349899 | [-0.04 0.02] | 0.588518832 | 0.913818271 |
| IL_18BP | Bilirubin_pPS | 4225 | -0.002335506 | [-0.03 0.03] | 0.879728605 | 0.97781424 |
| IL_18BP | Proinsulin_pPS | 4225 | 0.03143515 | [0. 0.06] | 0.041637257 | 0.481247748 |
| IL_18BP | Hyper_Insulin_pPS | 4225 | -0.005419761 | [-0.04 0.02] | 0.72548432 | 0.946981479 |
| IL_18BP | Beta_Cell_1_pPS | 4225 | -0.003789576 | [-0.03 0.03] | 0.806053522 | 0.967638637 |
| IL_18BP | Beta_Cell_2_pPS | 4225 | 0.013684657 | [-0.02 0.04] | 0.375270236 | 0.849453763 |
| Gal_4 | Cholesterol_pPS | 4225 | 0.00898915 | [-0.02 0.04] | 0.560296009 | 0.902076574 |
| Gal_4 | Lipodystrophy_1_pPS | 4225 | 0.035783052 | [0.01 0.07] | 0.020391829 | 0.350195677 |
| Gal_4 | Lipodystrophy_2_pPS | 4225 | 0.032608118 | [0. 0.06] | 0.034585324 | 0.443820658 |
| Gal_4 | Liver-Lipid_pPS | 4225 | -0.009545299 | [-0.04 0.02] | 0.536287954 | 0.898959051 |
| Gal_4 | ASAT_Pos_pPS | 4225 | 0.021689552 | [-0.01 0.05] | 0.159905188 | 0.717834051 |
| Gal_4 | ALP_Neg_pPS | 4225 | -0.236386806 | [-0.26 -0.21] | 1.99E-54 | 1.71E-51 |
| Gal_4 | SHBG-LpA_pPS | 4225 | 0.009577056 | [-0.02 0.04] | 0.534932818 | 0.898959051 |
| Gal_4 | Obesity_pPS | 4225 | 0.041471568 | [0.01 0.07] | 0.007187684 | 0.208039037 |
| Gal_4 | VAT_Neg_pPS | 4225 | 0.000941376 | [-0.03 0.03] | 0.951367181 | 0.996425456 |
| Gal_4 | Bilirubin_pPS | 4225 | 0.005566238 | [-0.02 0.04] | 0.7183769 | 0.94608328 |
| Gal_4 | Proinsulin_pPS | 4225 | 0.001926196 | [-0.03 0.03] | 0.900686055 | 0.985399384 |

**Table S7: Associations between pPSs and plasma proteins in SCAPIS**

**a) Spearman Correlation**

| Proteomics | PPS | Sample_Size | r | CI95% | p | fdr_p_adjusted |
|---|---|---|---|---|---|---|
| PAR_1 | Lipodystrophy_1_pPS | 4159 | -0.012737594 | [-0.04 0.02] | 0.412186771 | 0.8164748 |
| PAR_1 | Lipodystrophy_2_pPS | 4159 | 0.023099117 | [-0.01 0.05] | 0.136943894 | 0.629316669 |
| PAR_1 | Liver-Lipid_pPS | 4159 | 0.01322947 | [-0.02 0.04] | 0.394368771 | 0.812191725 |
| PAR_1 | ASAT_Pos_pPS | 4159 | 0.001363232 | [-0.03 0.03] | 0.930065902 | 0.990056084 |
| PAR_1 | ALP_Neg_pPS | 4159 | -0.020599608 | [-0.05 0.01] | 0.184741019 | 0.686251968 |
| PAR_1 | SHBG-LpA_pPS | 4159 | 0.01786812 | [-0.01 0.05] | 0.249978352 | 0.738661262 |
| PAR_1 | Obesity_pPS | 4159 | 0.010081181 | [-0.02 0.04] | 0.516325037 | 0.870853148 |
| PAR_1 | VAT_Neg_pPS | 4159 | -0.007517231 | [-0.04 0.02] | 0.62842168 | 0.914460114 |
| PAR_1 | Bilirubin_pPS | 4159 | 0.011902091 | [-0.02 0.04] | 0.443523135 | 0.831350198 |
| PAR_1 | Proinsulin_pPS | 4159 | 0.018325724 | [-0.01 0.05] | 0.238052432 | 0.726631332 |
| PAR_1 | Hyper_Insulin_pPS | 4159 | -0.027795384 | [-0.06 0. ] | 0.073493916 | 0.502472104 |
| PAR_1 | Beta_Cell_1_pPS | 4159 | -0.000861134 | [-0.03 0.03] | 0.955789611 | 0.99612031 |
| PAR_1 | Beta_Cell_2_pPS | 4159 | 0.00984065 | [-0.02 0.04] | 0.526384579 | 0.873942847 |
| PAR_1 | Total_GRS | 4159 | 0.017307236 | [-0.01 0.05] | 0.265156894 | 0.746520328 |
| GP6 | Cholesterol_pPS | 4225 | 0.00021845 | [-0.03 0.03] | 0.988690473 | 0.999402232 |
| GP6 | Lipodystrophy_1_pPS | 4225 | 0.008058326 | [-0.02 0.04] | 0.601043772 | 0.907623171 |
| GP6 | Lipodystrophy_2_pPS | 4225 | 0.019536163 | [-0.01 0.05] | 0.204872714 | 0.70858232 |
| GP6 | Liver-Lipid_pPS | 4225 | 0.001423483 | [-0.03 0.03] | 0.926406011 | 0.990056084 |
| GP6 | ASAT_Pos_pPS | 4225 | 0.030406054 | [0. 0.06] | 0.048443699 | 0.438114317 |
| GP6 | ALP_Neg_pPS | 4225 | -0.022492006 | [-0.05 0.01] | 0.14438514 | 0.641415816 |
| GP6 | SHBG-LpA_pPS | 4225 | -0.00409981 | [-0.03 0.03] | 0.790215269 | 0.957247257 |
| GP6 | Obesity_pPS | 4225 | -0.008674643 | [-0.04 0.02] | 0.573507074 | 0.903411176 |
| GP6 | VAT_Neg_pPS | 4225 | -0.010786523 | [-0.04 0.02] | 0.483964637 | 0.854586077 |
| GP6 | Bilirubin_pPS | 4225 | 0.005852344 | [-0.02 0.04] | 0.704129291 | 0.937028372 |
| GP6 | Proinsulin_pPS | 4225 | 0.015055195 | [-0.02 0.05] | 0.32858778 | 0.775125841 |
| GP6 | Hyper_Insulin_pPS | 4225 | -0.010361186 | [-0.04 0.02] | 0.501368233 | 0.863780476 |
| GP6 | Beta_Cell_1_pPS | 4225 | -0.004769659 | [-0.03 0.03] | 0.756943393 | 0.950403646 |
| GP6 | Beta_Cell_2_pPS | 4225 | 0.007778138 | [-0.02 0.04] | 0.613756686 | 0.909483337 |
| GP6 | Total_GRS | 4225 | 0.005163492 | [-0.03 0.04] | 0.73758526 | 0.942470055 |
| MMP12 | Cholesterol_pPS | 4159 | -0.018846007 | [-0.05 0.01] | 0.224987606 | 0.724402982 |
| MMP12 | Lipodystrophy_1_pPS | 4159 | 0.023901921 | [-0.01 0.05] | 0.123810959 | 0.617935349 |
| MMP12 | Lipodystrophy_2_pPS | 4159 | 0.011275689 | [-0.02 0.04] | 0.467883513 | 0.84347387 |
| MMP12 | Liver-Lipid_pPS | 4159 | -0.014772369 | [-0.05 0.02] | 0.341571695 | 0.787583858 |
| MMP12 | ASAT_Pos_pPS | 4159 | -0.014539382 | [-0.04 0.02] | 0.349240858 | 0.793215848 |
| MMP12 | ALP_Neg_pPS | 4159 | 0.021720717 | [-0.01 0.05] | 0.161963938 | 0.664061293 |
| MMP12 | SHBG-LpA_pPS | 4159 | 0.014644223 | [-0.02 0.05] | 0.345776431 | 0.790391668 |
| MMP12 | Obesity_pPS | 4159 | 0.006716376 | [-0.02 0.04] | 0.665457761 | 0.924111434 |
| MMP12 | VAT_Neg_pPS | 4159 | 0.004519873 | [-0.03 0.03] | 0.771065902 | 0.95301975 |
| MMP12 | Bilirubin_pPS | 4159 | -0.011660909 | [-0.04 0.02] | 0.452815712 | 0.834174785 |
| MMP12 | Proinsulin_pPS | 4159 | -0.002804722 | [-0.03 0.03] | 0.856711263 | 0.977006629 |
| MMP12 | Hyper_Insulin_pPS | 4159 | 0.018701083 | [-0.01 0.05] | 0.228574243 | 0.726631332 |
| MMP12 | Beta_Cell_1_pPS | 4159 | 0.011951908 | [-0.02 0.04] | 0.44161738 | 0.831350198 |
| MMP12 | Beta_Cell_2_pPS | 4159 | 0.004778155 | [-0.03 0.04] | 0.758380006 | 0.950403646 |
| MMP12 | Total_GRS | 4159 | 0.019189044 | [-0.01 0.05] | 0.216658598 | 0.716140995 |
| MMP7 | Cholesterol_pPS | 4159 | -0.005449243 | [-0.04 0.02] | 0.72572879 | 0.942134248 |
| MMP7 | Lipodystrophy_1_pPS | 4159 | 0.014033613 | [-0.02 0.04] | 0.366261922 | 0.798890546 |
| MMP7 | Lipodystrophy_2_pPS | 4159 | 0.049592702 | [0.02 0.08] | 0.001400081 | 0.061336891 |
| MMP7 | Liver-Lipid_pPS | 4159 | -0.034460817 | [-0.06 -0. ] | 0.026475134 | 0.344676278 |
| MMP7 | ASAT_Pos_pPS | 4159 | 0.017992761 | [-0.01 0.05] | 0.246689431 | 0.736582988 |
| MMP7 | ALP_Neg_pPS | 4159 | -0.024587039 | [-0.05 0.01] | 0.113398997 | 0.604978148 |
| MMP7 | SHBG-LpA_pPS | 4159 | 0.015993001 | [-0.01 0.05] | 0.303169657 | 0.76319014 |
| MMP7 | Obesity_pPS | 4159 | -0.003054736 | [-0.03 0.03] | 0.844095459 | 0.973549297 |
| MMP7 | VAT_Neg_pPS | 4159 | 0.050853291 | [0.02 0.08] | 0.001053101 | 0.050992277 |
| MMP7 | Bilirubin_pPS | 4159 | 0.009820887 | [-0.02 0.04] | 0.527215522 | 0.873942847 |
| MMP7 | Proinsulin_pPS | 4159 | -0.006250226 | [-0.04 0.02] | 0.68740464 | 0.926969309 |
| MMP7 | Hyper_Insulin_pPS | 4159 | 0.006531702 | [-0.02 0.04] | 0.674119477 | 0.925361755 |
| MMP7 | Beta_Cell_1_pPS | 4159 | 0.039137539 | [0.01 0.07] | 0.011717026 | 0.23392385 |
| MMP7 | Beta_Cell_2_pPS | 4159 | -0.005599829 | [-0.04 0.02] | 0.718467708 | 0.941449667 |
| MMP7 | Total_GRS | 4159 | 0.03838325 | [0.01 0.07] | 0.013438218 | 0.242414912 |
| uPA | Cholesterol_pPS | 4225 | -0.010607875 | [-0.04 0.02] | 0.49123427 | 0.859085266 |
| uPA | Lipodystrophy_1_pPS | 4225 | 0.018254229 | [-0.01 0.05] | 0.236181264 | 0.726631332 |
| uPA | Lipodystrophy_2_pPS | 4225 | 0.023035941 | [-0.01 0.05] | 0.134924122 | 0.629316669 |
| uPA | Liver-Lipid_pPS | 4225 | 0.0274392 | [-0. 0.06] | 0.074941842 | 0.507976478 |
| uPA | ASAT_Pos_pPS | 4225 | 0.017455491 | [-0.01 0.05] | 0.257321538 | 0.740941247 |
| uPA | ALP_Neg_pPS | 4225 | -0.011218267 | [-0.04 0.02] | 0.466639336 | 0.841780762 |
| uPA | SHBG-LpA_pPS | 4225 | 0.013587086 | [-0.02 0.04] | 0.377947454 | 0.803170077 |
| uPA | Obesity_pPS | 4225 | 0.005490815 | [-0.02 0.04] | 0.721621364 | 0.942134248 |
| uPA | VAT_Neg_pPS | 4225 | -0.007671615 | [-0.04 0.02] | 0.618620869 | 0.909483337 |
| uPA | Bilirubin_pPS | 4225 | -0.013955996 | [-0.04 0.02] | 0.365133893 | 0.798890546 |
| uPA | Proinsulin_pPS | 4225 | 0.022004684 | [-0.01 0.05] | 0.153285844 | 0.649125224 |
| uPA | Hyper_Insulin_pPS | 4225 | -0.007752439 | [-0.04 0.02] | 0.61492867 | 0.909483337 |
| uPA | Beta_Cell_1_pPS | 4225 | 0.001724998 | [-0.03 0.03] | 0.910877016 | 0.990056084 |
| uPA | Beta_Cell_2_pPS | 4225 | 0.045807481 | [0.02 0.08] | 0.002940087 | 0.105384924 |
| uPA | Total_GRS | 4225 | 0.02656823 | [-0. 0.06] | 0.084658503 | 0.535911625 |
| CD93 | Cholesterol_pPS | 4225 | 0.013837787 | [-0.02 0.04] | 0.369209873 | 0.799230784 |
| CD93 | Lipodystrophy_1_pPS | 4225 | 0.008098888 | [-0.02 0.04] | 0.599213274 | 0.907623171 |
| CD93 | Lipodystrophy_2_pPS | 4225 | 0.011022655 | [-0.02 0.04] | 0.474446047 | 0.847005879 |
| CD93 | Liver-Lipid_pPS | 4225 | 0.027929245 | [-0. 0.06] | 0.069887882 | 0.491184667 |
| CD93 | ASAT_Pos_pPS | 4225 | -0.00872019 | [-0.04 0.02] | 0.571496013 | 0.902878647 |
| CD93 | ALP_Neg_pPS | 4225 | 0.019340499 | [-0.01 0.05] | 0.209445821 | 0.709689105 |
| CD93 | SHBG-LpA_pPS | 4225 | 0.014870891 | [-0.02 0.05] | 0.33454421 | 0.780508892 |
| CD93 | Obesity_pPS | 4225 | 0.01881808 | [-0.01 0.05] | 0.222016894 | 0.722207328 |
| CD93 | VAT_Neg_pPS | 4225 | -0.022423797 | [-0.05 0.01] | 0.145606566 | 0.641415816 |
| CD93 | Bilirubin_pPS | 4225 | -0.01560157 | [-0.05 0.01] | 0.311336318 | 0.76365449 |
| CD93 | Proinsulin_pPS | 4225 | 0.031030793 | [0. 0.06] | 0.044007182 | 0.424281318 |
| CD93 | Hyper_Insulin_pPS | 4225 | 0.019059926 | [-0.01 0.05] | 0.216131704 | 0.715705689 |
| CD93 | Beta_Cell_1_pPS | 4225 | 0.017216857 | [-0.01 0.05] | 0.263884661 | 0.746520328 |
| CD93 | Beta_Cell_2_pPS | 4225 | 0.038156386 | [0.01 0.07] | 0.01325634 | 0.242414912 |
| CD93 | Total_GRS | 4225 | 0.04419291 | [0.01 0.07] | 0.004117622 | 0.124886119 |
| CEACAM8 | Cholesterol_pPS | 4159 | -0.014514 | [-0.04 0.02] | 0.350082903 | 0.793291306 |
| CEACAM8 | Lipodystrophy_1_pPS | 4159 | -0.003820081 | [-0.03 0.03] | 0.805736994 | 0.960193229 |
| CEACAM8 | Lipodystrophy_2_pPS | 4159 | 0.004006649 | [-0.03 0.03] | 0.79645303 | 0.958074488 |
| CEACAM8 | Liver-Lipid_pPS | 4159 | 0.015858578 | [-0.01 0.05] | 0.30725233 | 0.76365449 |
| CEACAM8 | ASAT_Pos_pPS | 4159 | -0.009923651 | [-0.04 0.02] | 0.522901986 | 0.872980599 |
| CEACAM8 | ALP_Neg_pPS | 4159 | 0.013012252 | [-0.02 0.04] | 0.402179251 | 0.81230319 |
| CEACAM8 | SHBG-LpA_pPS | 4159 | 0.005346753 | [-0.03 0.04] | 0.73068492 | 0.942134248 |
| CEACAM8 | Obesity_pPS | 4159 | 0.016494279 | [-0.01 0.05] | 0.288264076 | 0.751993242 |
| CEACAM8 | VAT_Neg_pPS | 4159 | -0.013500786 | [-0.04 0.02] | 0.384743004 | 0.806422804 |
| CEACAM8 | Bilirubin_pPS | 4159 | 0.022709349 | [-0.01 0.05] | 0.143695656 | 0.641415816 |
| CEACAM8 | Proinsulin_pPS | 4159 | 0.032935951 | [0. 0.06] | 0.033928606 | 0.382561193 |
| CEACAM8 | Hyper_Insulin_pPS | 4159 | -0.032562439 | [-0.06 -0. ] | 0.036006506 | 0.386599178 |
| CEACAM8 | Beta_Cell_1_pPS | 4159 | -0.005308676 | [-0.04 0.03] | 0.732529057 | 0.942134248 |
| CEACAM8 | Beta_Cell_2_pPS | 4159 | -0.044816553 | [-0.08 -0.01] | 0.003893782 | 0.119409314 |
| CEACAM8 | Total_GRS | 4159 | -0.003752141 | [-0.03 0.03] | 0.809124669 | 0.96133624 |
| DLK_1 | Cholesterol_pPS | 4225 | 0.015864584 | [-0.01 0.05] | 0.303248665 | 0.76319014 |
| DLK_1 | Lipodystrophy_1_pPS | 4225 | 0.025357245 | [-0. 0.06] | 0.099834859 | 0.585019558 |
| DLK_1 | Lipodystrophy_2_pPS | 4225 | 0.022017606 | [-0.01 0.05] | 0.153044566 | 0.649125224 |

**b) Partial Correlation**

| Proteomics | PPS | Sample_Size | r | CI95% | p | fdr_p_adjusted |
|---|---|---|---|---|---|---|
| Gal_4 | Hyper_Insulin_pPS | 4225 | -0.006191934 | [-0.04 0.02] | 0.68829673 | 0.944989936 |
| Gal_4 | Beta_Cell_1_pPS | 4225 | -0.020693621 | [-0.05 0.01] | 0.179971004 | 0.738879686 |
| Gal_4 | Beta_Cell_2_pPS | 4225 | 0.014956387 | [-0.02 0.05] | 0.332520076 | 0.844649909 |
| MEPE | Cholesterol_pPS | 4225 | 0.013416636 | [-0.02 0.04] | 0.384695248 | 0.85037326 |
| MEPE | Lipodystrophy_1_pPS | 4225 | -0.004491857 | [-0.03 0.03] | 0.771035687 | 0.961368794 |
| MEPE | Lipodystrophy_2_pPS | 4225 | 0.003871166 | [-0.03 0.03] | 0.801963729 | 0.967638637 |
| MEPE | Liver-Lipid_pPS | 4225 | 0.018010352 | [-0.01 0.05] | 0.243229987 | 0.793114488 |
| MEPE | ASAT_Pos_pPS | 4225 | 0.00090598 | [-0.03 0.03] | 0.953193638 | 0.996425456 |
| MEPE | ALP_Neg_pPS | 4225 | -0.009373102 | [-0.04 0.02] | 0.543665783 | 0.899241939 |
| MEPE | SHBG-LpA_pPS | 4225 | 0.019314473 | [-0.01 0.05] | 0.210764855 | 0.767305935 |
| MEPE | Obesity_pPS | 4225 | -0.022366485 | [-0.05 0.01] | 0.147263942 | 0.703126624 |
| MEPE | VAT_Neg_pPS | 4225 | -0.01912653 | [-0.05 0.01] | 0.215239906 | 0.767947366 |
| MEPE | Bilirubin_pPS | 4225 | -0.01639954 | [-0.05 0.01] | 0.287978462 | 0.81609738 |
| MEPE | Proinsulin_pPS | 4225 | 0.037779566 | [0.01 0.07] | 0.014343611 | 0.312663737 |
| MEPE | Hyper_Insulin_pPS | 4225 | 0.001359557 | [-0.03 0.03] | 0.929810542 | 0.991797911 |
| MEPE | Beta_Cell_1_pPS | 4225 | -0.017319688 | [-0.05 0.01] | 0.26177991 | 0.807171938 |
| MEPE | Beta_Cell_2_pPS | 4225 | 0.000101167 | [-0.03 0.03] | 0.994770347 | 0.998803911 |
| hOSCAR | Cholesterol_pPS | 4159 | 0.001463723 | [-0.03 0.03] | 0.925042892 | 0.990503296 |
| hOSCAR | Lipodystrophy_1_pPS | 4159 | 0.011911548 | [-0.02 0.04] | 0.443877018 | 0.867547191 |
| hOSCAR | Lipodystrophy_2_pPS | 4159 | 0.012289772 | [-0.02 0.04] | 0.429541558 | 0.864060889 |
| hOSCAR | Liver-Lipid_pPS | 4159 | 0.020653782 | [-0.01 0.05] | 0.1842774 | 0.738879686 |
| hOSCAR | ASAT_Pos_pPS | 4159 | -0.03306072 | [-0.06 -0. ] | 0.033535205 | 0.436493301 |
| hOSCAR | ALP_Neg_pPS | 4159 | 0.009489388 | [-0.02 0.04] | 0.541887836 | 0.899241939 |
| hOSCAR | SHBG-LpA_pPS | 4159 | 0.001063097 | [-0.03 0.03] | 0.945521009 | 0.996180826 |
| hOSCAR | Obesity_pPS | 4159 | 0.023422653 | [-0.01 0.05] | 0.13213432 | 0.694065456 |
| hOSCAR | VAT_Neg_pPS | 4159 | 0.002979942 | [-0.03 0.03] | 0.84810147 | 0.97401221 |
| hOSCAR | Bilirubin_pPS | 4159 | 0.003627342 | [-0.03 0.03] | 0.815641436 | 0.968668398 |
| hOSCAR | Proinsulin_pPS | 4159 | 0.021835625 | [-0.01 0.05] | 0.160411104 | 0.717834051 |
| hOSCAR | Hyper_Insulin_pPS | 4159 | -0.007710863 | [-0.04 0.02] | 0.620150852 | 0.922349073 |
| hOSCAR | Beta_Cell_1_pPS | 4159 | -0.003050454 | [-0.03 0.03] | 0.844552471 | 0.972178273 |
| hOSCAR | Beta_Cell_2_pPS | 4159 | -0.02438247 | [-0.05 0.01] | 0.117007485 | 0.681046037 |
| PRELP | Cholesterol_pPS | 4158 | -0.008396293 | [-0.04 0.02] | 0.58945182 | 0.913889104 |
| PRELP | Lipodystrophy_1_pPS | 4158 | -0.030702538 | [-0.06 -0. ] | 0.048417498 | 0.520248282 |
| PRELP | Lipodystrophy_2_pPS | 4158 | 0.00800965 | [-0.02 0.04] | 0.606706644 | 0.918301271 |
| PRELP | Liver-Lipid_pPS | 4158 | 0.003732192 | [-0.03 0.03] | 0.810434955 | 0.967638637 |
| PRELP | ASAT_Pos_pPS | 4158 | -0.027536918 | [-0.06 0. ] | 0.07670967 | 0.598800331 |
| PRELP | ALP_Neg_pPS | 4158 | -0.012253861 | [-0.04 0.02] | 0.430946762 | 0.864060889 |
| PRELP | SHBG-LpA_pPS | 4158 | 0.046143008 | [0.02 0.08] | 0.003005803 | 0.124886277 |
| PRELP | Obesity_pPS | 4158 | 0.008576622 | [-0.02 0.04] | 0.581482362 | 0.909583681 |
| PRELP | VAT_Neg_pPS | 4158 | -0.009190093 | [-0.04 0.02] | 0.554754777 | 0.899898936 |
| PRELP | Bilirubin_pPS | 4158 | -0.000261298 | [-0.03 0.03] | 0.986601465 | 0.998803911 |
| PRELP | Proinsulin_pPS | 4158 | 0.030402422 | [-0. 0.06] | 0.050655827 | 0.537121778 |
| PRELP | Hyper_Insulin_pPS | 4158 | -0.001720914 | [-0.03 0.03] | 0.9119324 | 0.989461462 |
| PRELP | Beta_Cell_1_pPS | 4158 | -0.025738194 | [-0.06 0. ] | 0.098037909 | 0.64755296 |
| PRELP | Beta_Cell_2_pPS | 4158 | -0.003220054 | [-0.03 0.03] | 0.836048856 | 0.972099644 |
| SCGB3A2 | Cholesterol_pPS | 4225 | 0.005307285 | [-0.02 0.04] | 0.730958052 | 0.947604067 |
| SCGB3A2 | Lipodystrophy_1_pPS | 4225 | -0.043848718 | [-0.07 -0.01] | 0.004479735 | 0.15594322 |
| SCGB3A2 | Lipodystrophy_2_pPS | 4225 | -0.010528701 | [-0.04 0.02] | 0.495141831 | 0.895803896 |
| SCGB3A2 | Liver-Lipid_pPS | 4225 | -0.010107111 | [-0.04 0.02] | 0.512572439 | 0.895803896 |
| SCGB3A2 | ASAT_Pos_pPS | 4225 | -0.023421039 | [-0.05 0.01] | 0.129111945 | 0.694065456 |
| SCGB3A2 | ALP_Neg_pPS | 4225 | -0.031121361 | [-0.06 -0. ] | 0.04371818 | 0.49830987 |
| SCGB3A2 | SHBG-LpA_pPS | 4225 | 0.004032191 | [-0.03 0.03] | 0.79390812 | 0.967638637 |
| SCGB3A2 | Obesity_pPS | 4225 | -0.018139989 | [-0.05 0.01] | 0.239853752 | 0.790085251 |
| SCGB3A2 | VAT_Neg_pPS | 4225 | -0.012146244 | [-0.04 0.02] | 0.431304541 | 0.864060889 |
| SCGB3A2 | Bilirubin_pPS | 4225 | -0.010979947 | [-0.04 0.02] | 0.476841654 | 0.886891048 |
| SCGB3A2 | Proinsulin_pPS | 4225 | 0.032928327 | [0. 0.06] | 0.032847456 | 0.433923314 |
| SCGB3A2 | Hyper_Insulin_pPS | 4225 | -0.014796285 | [-0.05 0.02] | 0.337722049 | 0.844649909 |
| SCGB3A2 | Beta_Cell_1_pPS | 4225 | 0.009387483 | [-0.02 0.04] | 0.543047699 | 0.899241939 |
| SCGB3A2 | Beta_Cell_2_pPS | 4225 | 0.007370902 | [-0.02 0.04] | 0.632968141 | 0.931124222 |
| SHPS_1 | Cholesterol_pPS | 4225 | 0.014470294 | [-0.02 0.04] | 0.348474588 | 0.847284666 |
| SHPS_1 | Lipodystrophy_1_pPS | 4225 | 0.010151138 | [-0.02 0.04] | 0.510737307 | 0.895803896 |
| SHPS_1 | Lipodystrophy_2_pPS | 4225 | 0.03688692 | [0.01 0.07] | 0.016818688 | 0.33440804 |
| SHPS_1 | Liver-Lipid_pPS | 4225 | 0.03343977 | [0. 0.06] | 0.030226791 | 0.416452728 |
| SHPS_1 | ASAT_Pos_pPS | 4225 | -0.014020254 | [-0.04 0.02] | 0.363671951 | 0.847284666 |
| SHPS_1 | ALP_Neg_pPS | 4225 | -0.036763712 | [-0.07 -0.01] | 0.017188199 | 0.33440804 |
| SHPS_1 | SHBG-LpA_pPS | 4225 | 0.008131482 | [-0.02 0.04] | 0.598310196 | 0.918301271 |
| SHPS_1 | Obesity_pPS | 4225 | -0.002831792 | [-0.03 0.03] | 0.854431739 | 0.975477892 |
| SHPS_1 | VAT_Neg_pPS | 4225 | -0.004811773 | [-0.04 0.03] | 0.755232411 | 0.95462436 |
| SHPS_1 | Bilirubin_pPS | 4225 | 0.008704601 | [-0.02 0.04] | 0.572777676 | 0.907786603 |
| SHPS_1 | Proinsulin_pPS | 4225 | 0.009228202 | [-0.02 0.04] | 0.549912943 | 0.899241939 |
| SHPS_1 | Hyper_Insulin_pPS | 4225 | 0.017031941 | [-0.01 0.05] | 0.269789518 | 0.808113719 |
| SHPS_1 | Beta_Cell_1_pPS | 4225 | 0.006281439 | [-0.02 0.04] | 0.684032513 | 0.944323894 |
| SHPS_1 | Beta_Cell_2_pPS | 4225 | 0.003810505 | [-0.03 0.03] | 0.805003916 | 0.967638637 |
| TFF3 | Cholesterol_pPS | 4225 | 0.01921673 | [-0.01 0.05] | 0.213083707 | 0.767305935 |
| TFF3 | Lipodystrophy_1_pPS | 4225 | 0.029398057 | [-0. 0.06] | 0.056772736 | 0.558815902 |
| TFF3 | Lipodystrophy_2_pPS | 4225 | -0.001774209 | [-0.03 0.03] | 0.908486518 | 0.988841471 |
| TFF3 | Liver-Lipid_pPS | 4225 | 0.00508543 | [-0.03 0.04] | 0.741794761 | 0.951867558 |
| TFF3 | ASAT_Pos_pPS | 4225 | -0.012369434 | [-0.04 0.02] | 0.422886804 | 0.860518922 |
| TFF3 | ALP_Neg_pPS | 4225 | -0.005738106 | [-0.04 0.02] | 0.710068487 | 0.945970482 |
| TFF3 | SHBG-LpA_pPS | 4225 | 0.006927704 | [-0.02 0.04] | 0.653548321 | 0.936605382 |
| TFF3 | Obesity_pPS | 4225 | 0.010312112 | [-0.02 0.04] | 0.504056934 | 0.895803896 |
| TFF3 | VAT_Neg_pPS | 4225 | -0.025930271 | [-0.06 0. ] | 0.092907533 | 0.638855661 |
| TFF3 | Bilirubin_pPS | 4225 | 0.016791377 | [-0.01 0.05] | 0.276613493 | 0.808820203 |
| TFF3 | Proinsulin_pPS | 4225 | 0.012796868 | [-0.02 0.04] | 0.407037115 | 0.853674654 |
| TFF3 | Hyper_Insulin_pPS | 4225 | -0.001582122 | [-0.03 0.03] | 0.918357592 | 0.99041009 |
| TFF3 | Beta_Cell_1_pPS | 4225 | 0.012671189 | [-0.02 0.04] | 0.41166026 | 0.854426619 |
| TFF3 | Beta_Cell_2_pPS | 4225 | 0.005754161 | [-0.02 0.04] | 0.709294063 | 0.945970482 |
| TLT_2 | Cholesterol_pPS | 4225 | -0.003025393 | [-0.03 0.03] | 0.844602366 | 0.972178273 |
| TLT_2 | Lipodystrophy_1_pPS | 4225 | 0.015331068 | [-0.01 0.05] | 0.320549447 | 0.83927892 |
| TLT_2 | Lipodystrophy_2_pPS | 4225 | 0.007198343 | [-0.02 0.04] | 0.640948423 | 0.933976497 |
| TLT_2 | Liver-Lipid_pPS | 4225 | 0.028888244 | [-0. 0.06] | 0.061205835 | 0.558815902 |
| TLT_2 | ASAT_Pos_pPS | 4225 | -0.00937872 | [-0.04 0.02] | 0.543424309 | 0.899241939 |
| TLT_2 | ALP_Neg_pPS | 4225 | 0.024115337 | [-0.01 0.05] | 0.1181433 | 0.681046037 |
| TLT_2 | SHBG-LpA_pPS | 4225 | 0.004641544 | [-0.03 0.03] | 0.763629151 | 0.959095176 |
| TLT_2 | Obesity_pPS | 4225 | 0.001590828 | [-0.03 0.03] | 0.917909939 | 0.99041009 |
| TLT_2 | VAT_Neg_pPS | 4225 | -0.000934809 | [-0.03 0.03] | 0.951705999 | 0.996425456 |
| TLT_2 | Bilirubin_pPS | 4225 | -0.00091532 | [-0.03 0.03] | 0.952711683 | 0.996425456 |
| TLT_2 | Proinsulin_pPS | 4225 | 0.022129715 | [-0.01 0.05] | 0.151595633 | 0.706166997 |
| TLT_2 | Hyper_Insulin_pPS | 4225 | -0.01411917 | [-0.04 0.02] | 0.360296614 | 0.847284666 |
| TLT_2 | Beta_Cell_1_pPS | 4225 | 0.001192264 | [-0.03 0.03] | 0.938428972 | 0.994163908 |
| TLT_2 | Beta_Cell_2_pPS | 4225 | 0.011091072 | [-0.02 0.04] | 0.472392509 | 0.883720482 |
| NT_proBNP | Cholesterol_pPS | 4225 | -0.000533496 | [-0.03 0.03] | 0.972427244 | 0.997444743 |
| NT_proBNP | Lipodystrophy_1_pPS | 4225 | -0.018474283 | [-0.05 0.01] | 0.231300254 | 0.786988264 |
| NT_proBNP | Lipodystrophy_2_pPS | 4225 | 0.005814493 | [-0.02 0.04] | 0.706386746 | 0.945970482 |
| NT_proBNP | Liver-Lipid_pPS | 4225 | 0.019448785 | [-0.01 0.05] | 0.207608273 | 0.767305935 |
| NT_proBNP | ASAT_Pos_pPS | 4225 | -0.001310968 | [-0.03 0.03] | 0.932312898 | 0.993233261 |
| NT_proBNP | ALP_Neg_pPS | 4225 | 0.01328897 | [-0.02 0.04] | 0.389235093 | 0.85037326 |

**Table S7: Associations between pPSs and plasma proteins in SCAPIS**

**a) Spearman Correlation**

| Proteomics | PPS | Sample_Size | r | CI95% | p | fdr_p_adjusted |
|---|---|---|---|---|---|---|
| DLK_1 | Liver-Lipid_pPS | 4225 | 0.046134176 | [0.02 0.08] | 0.002742955 | 0.099612573 |
| DLK_1 | ASAT_Pos_pPS | 4225 | 0.017607621 | [-0.01 0.05] | 0.25319709 | 0.739680398 |
| DLK_1 | ALP_Neg_pPS | 4225 | -0.010983604 | [-0.04 0.02] | 0.47601308 | 0.84751951 |
| DLK_1 | SHBG-LpA_pPS | 4225 | -0.001547981 | [-0.03 0.03] | 0.919990235 | 0.990056084 |
| DLK_1 | Obesity_pPS | 4225 | 0.016512241 | [-0.01 0.05] | 0.283933274 | 0.749111537 |
| DLK_1 | VAT_Neg_pPS | 4225 | 0.008052334 | [-0.02 0.04] | 0.601314427 | 0.907623171 |
| DLK_1 | Bilirubin_pPS | 4225 | -0.007781133 | [-0.04 0.02] | 0.613620193 | 0.909483337 |
| DLK_1 | Proinsulin_pPS | 4225 | -0.012525482 | [-0.04 0.02] | 0.416338511 | 0.817860704 |
| DLK_1 | Hyper_Insulin_pPS | 4225 | 0.019536672 | [-0.01 0.05] | 0.204860922 | 0.70858232 |
| DLK_1 | Beta_Cell_1_pPS | 4225 | 0.022584156 | [-0.01 0.05] | 0.142747502 | 0.640622936 |
| DLK_1 | Beta_Cell_2_pPS | 4225 | 0.017753251 | [-0.01 0.05] | 0.249292221 | 0.738661262 |
| DLK_1 | Total_GRS | 4225 | 0.033208885 | [0. 0.06] | 0.031125485 | 0.369452038 |
| Ep_CAM | Cholesterol_pPS | 4225 | -0.004341878 | [-0.03 0.03] | 0.778143881 | 0.953637204 |
| Ep_CAM | Lipodystrophy_1_pPS | 4225 | 0.003517489 | [-0.03 0.03] | 0.819455939 | 0.963461584 |
| Ep_CAM | Lipodystrophy_2_pPS | 4225 | 0.005515363 | [-0.02 0.04] | 0.720428919 | 0.94146961 |
| Ep_CAM | Liver-Lipid_pPS | 4225 | 0.001971899 | [-0.03 0.03] | 0.898185832 | 0.984642786 |
| Ep_CAM | ASAT_Pos_pPS | 4225 | 0.010371008 | [-0.02 0.04] | 0.500962648 | 0.863780476 |
| Ep_CAM | ALP_Neg_pPS | 4225 | -0.002893094 | [-0.03 0.03] | 0.85108948 | 0.97528582 |
| Ep_CAM | SHBG-LpA_pPS | 4225 | -0.010374266 | [-0.04 0.02] | 0.500828142 | 0.863780476 |
| Ep_CAM | Obesity_pPS | 4225 | -0.003229732 | [-0.03 0.03] | 0.834001262 | 0.969605511 |
| Ep_CAM | VAT_Neg_pPS | 4225 | -0.019419841 | [-0.05 0.01] | 0.207582576 | 0.709689105 |
| Ep_CAM | Bilirubin_pPS | 4225 | -0.009939226 | [-0.04 0.02] | 0.518955647 | 0.872596104 |
| Ep_CAM | Proinsulin_pPS | 4225 | 0.002379569 | [-0.03 0.03] | 0.877289165 | 0.978705778 |
| Ep_CAM | Hyper_Insulin_pPS | 4225 | 0.008636418 | [-0.02 0.04] | 0.575197403 | 0.903554258 |
| Ep_CAM | Beta_Cell_1_pPS | 4225 | -0.015731898 | [-0.05 0.01] | 0.307311148 | 0.76365449 |
| Ep_CAM | Beta_Cell_2_pPS | 4225 | -0.004805391 | [-0.03 0.03] | 0.75518053 | 0.950403646 |
| Ep_CAM | Total_GRS | 4225 | 0.009044872 | [-0.02 0.04] | 0.557258131 | 0.896793725 |
| KIM1 | Cholesterol_pPS | 4159 | -0.012536168 | [-0.04 0.02] | 0.41961893 | 0.820585997 |
| KIM1 | Lipodystrophy_1_pPS | 4159 | 0.007480677 | [-0.02 0.04] | 0.630092829 | 0.914460114 |
| KIM1 | Lipodystrophy_2_pPS | 4159 | 0.017190566 | [-0.01 0.05] | 0.268392331 | 0.746520328 |
| KIM1 | Liver-Lipid_pPS | 4159 | -0.012151111 | [-0.04 0.02] | 0.434043839 | 0.828998134 |
| KIM1 | ASAT_Pos_pPS | 4159 | 0.01326065 | [-0.02 0.04] | 0.393255208 | 0.812191725 |
| KIM1 | ALP_Neg_pPS | 4159 | -0.018983747 | [-0.05 0.01] | 0.221616185 | 0.722207328 |
| KIM1 | SHBG-LpA_pPS | 4159 | -0.006819071 | [-0.04 0.02] | 0.660660224 | 0.924111434 |
| KIM1 | Obesity_pPS | 4159 | 0.014222217 | [-0.02 0.04] | 0.359855085 | 0.7979754 |
| KIM1 | VAT_Neg_pPS | 4159 | -0.015076733 | [-0.05 0.02] | 0.331716656 | 0.778452202 |
| KIM1 | Bilirubin_pPS | 4159 | -0.007150552 | [-0.04 0.02] | 0.645270321 | 0.916568313 |
| KIM1 | Proinsulin_pPS | 4159 | 0.017630802 | [-0.01 0.05] | 0.256324917 | 0.740941247 |
| KIM1 | Hyper_Insulin_pPS | 4159 | 0.001481824 | [-0.03 0.03] | 0.923999821 | 0.990056084 |
| KIM1 | Beta_Cell_1_pPS | 4159 | 0.004629856 | [-0.03 0.04] | 0.76565624 | 0.952160028 |
| KIM1 | Beta_Cell_2_pPS | 4159 | 0.002549083 | [-0.03 0.03] | 0.869648786 | 0.977006629 |
| KIM1 | Total_GRS | 4159 | 0.025216586 | [-0.01 0.06] | 0.104451326 | 0.593032011 |
| IL_18BP | Cholesterol_pPS | 4225 | 0.017595982 | [-0.01 0.05] | 0.253511007 | 0.739680398 |
| IL_18BP | Lipodystrophy_1_pPS | 4225 | 0.025147007 | [-0.01 0.06] | 0.102678174 | 0.586842904 |
| IL_18BP | Lipodystrophy_2_pPS | 4225 | 0.019349499 | [-0.01 0.05] | 0.209233861 | 0.709689105 |
| IL_18BP | Liver-Lipid_pPS | 4225 | 0.015304868 | [-0.01 0.05] | 0.320629039 | 0.770109076 |
| IL_18BP | ASAT_Pos_pPS | 4225 | -0.002755572 | [-0.03 0.03] | 0.858090901 | 0.977006629 |
| IL_18BP | ALP_Neg_pPS | 4225 | -0.012625073 | [-0.04 0.02] | 0.412642087 | 0.8164748 |
| IL_18BP | SHBG-LpA_pPS | 4225 | 0.00973373 | [-0.02 0.04] | 0.527634688 | 0.874112688 |
| IL_18BP | Obesity_pPS | 4225 | 0.025869819 | [-0. 0.06] | 0.093165449 | 0.559254737 |
| IL_18BP | VAT_Neg_pPS | 4225 | -0.004099582 | [-0.03 0.03] | 0.790226668 | 0.957247257 |
| IL_18BP | Bilirubin_pPS | 4225 | -0.001296782 | [-0.03 0.03] | 0.932940251 | 0.990208704 |
| IL_18BP | Proinsulin_pPS | 4225 | 0.035101252 | [0. 0.07] | 0.022704845 | 0.320646716 |
| IL_18BP | Hyper_Insulin_pPS | 4225 | -0.00044454 | [-0.03 0.03] | 0.976987826 | 0.996862516 |
| IL_18BP | Beta_Cell_1_pPS | 4225 | 0.008816466 | [-0.02 0.04] | 0.567256154 | 0.899863296 |
| IL_18BP | Beta_Cell_2_pPS | 4225 | 0.019584631 | [-0.01 0.05] | 0.20375121 | 0.708253575 |
| IL_18BP | Total_GRS | 4225 | 0.035785825 | [0.01 0.07] | 0.020188414 | 0.307845423 |
| Gal_4 | Cholesterol_pPS | 4225 | 0.012566652 | [-0.02 0.04] | 0.414808088 | 0.817764515 |
| Gal_4 | Lipodystrophy_1_pPS | 4225 | 0.037587029 | [0.01 0.07] | 0.01469446 | 0.252355767 |
| Gal_4 | Lipodystrophy_2_pPS | 4225 | 0.035528532 | [0.01 0.07] | 0.021104129 | 0.313142997 |
| Gal_4 | Liver-Lipid_pPS | 4225 | -0.017450146 | [-0.05 0.01] | 0.257467271 | 0.740941247 |
| Gal_4 | ASAT_Pos_pPS | 4225 | 0.030505381 | [0. 0.06] | 0.047714199 | 0.436063541 |
| Gal_4 | ALP_Neg_pPS | 4225 | -0.238239763 | [-0.27 -0.21] | 1.90E-55 | 1.75E-52 |
| Gal_4 | SHBG-LpA_pPS | 4225 | 0.018713765 | [-0.01 0.05] | 0.224590441 | 0.724402982 |
| Gal_4 | Obesity_pPS | 4225 | 0.038954308 | [0.01 0.07] | 0.011450506 | 0.232377917 |
| Gal_4 | VAT_Neg_pPS | 4225 | 0.004441576 | [-0.03 0.03] | 0.773187509 | 0.953322233 |
| Gal_4 | Bilirubin_pPS | 4225 | 0.0065832 | [-0.02 0.04] | 0.669249875 | 0.925084621 |
| Gal_4 | Proinsulin_pPS | 4225 | -0.035466558 | [-0.07 -0.01] | 0.02133003 | 0.313142997 |
| Gal_4 | Hyper_Insulin_pPS | 4225 | -0.00111225 | [-0.03 0.03] | 0.942464934 | 0.992903797 |
| Gal_4 | Beta_Cell_1_pPS | 4225 | -0.016718675 | [-0.05 0.01] | 0.277955624 | 0.747422497 |
| Gal_4 | Beta_Cell_2_pPS | 4225 | 0.007907371 | [-0.02 0.04] | 0.60787828 | 0.909483337 |
| Gal_4 | Total_GRS | 4225 | 0.043535784 | [0.01 0.07] | 0.004708828 | 0.136074023 |
| MEPE | Cholesterol_pPS | 4225 | 0.013334404 | [-0.02 0.04] | 0.386881739 | 0.80771074 |
| MEPE | Lipodystrophy_1_pPS | 4225 | -0.005041423 | [-0.04 0.03] | 0.743568206 | 0.94573652 |
| MEPE | Lipodystrophy_2_pPS | 4225 | 0.003409486 | [-0.03 0.03] | 0.824908203 | 0.964449615 |
| MEPE | Liver-Lipid_pPS | 4225 | 0.01836919 | [-0.01 0.05] | 0.233242692 | 0.726631332 |
| MEPE | ASAT_Pos_pPS | 4225 | -0.001213393 | [-0.03 0.03] | 0.937243333 | 0.991487772 |
| MEPE | ALP_Neg_pPS | 4225 | -0.003080105 | [-0.03 0.03] | 0.841587388 | 0.972342391 |
| MEPE | SHBG-LpA_pPS | 4225 | 0.019984963 | [-0.01 0.05] | 0.194658341 | 0.696574877 |
| MEPE | Obesity_pPS | 4225 | -0.016970749 | [-0.05 0.01] | 0.27077329 | 0.746520328 |
| MEPE | VAT_Neg_pPS | 4225 | -0.019204321 | [-0.05 0.01] | 0.21267191 | 0.712633026 |
| MEPE | Bilirubin_pPS | 4225 | -0.017370015 | [-0.05 0.01] | 0.259659244 | 0.743022111 |
| MEPE | Proinsulin_pPS | 4225 | 0.031506515 | [0. 0.06] | 0.040863348 | 0.407062052 |
| MEPE | Hyper_Insulin_pPS | 4225 | -0.00068077 | [-0.03 0.03] | 0.964765662 | 0.99612031 |
| MEPE | Beta_Cell_1_pPS | 4225 | -0.01005735 | [-0.04 0.02] | 0.514000334 | 0.870853148 |
| MEPE | Beta_Cell_2_pPS | 4225 | 0.000988534 | [-0.03 0.03] | 0.948855251 | 0.993154165 |
| MEPE | Total_GRS | 4225 | -0.009070674 | [-0.04 0.02] | 0.556134123 | 0.896216632 |
| hOSCAR | Cholesterol_pPS | 4159 | 0.005218867 | [-0.03 0.04] | 0.736884882 | 0.942470055 |
| hOSCAR | Lipodystrophy_1_pPS | 4159 | 0.007680475 | [-0.02 0.04] | 0.620981815 | 0.910907479 |
| hOSCAR | Lipodystrophy_2_pPS | 4159 | 0.010450633 | [-0.02 0.04] | 0.501070069 | 0.863780476 |
| hOSCAR | Liver-Lipid_pPS | 4159 | 0.020817924 | [-0.01 0.05] | 0.180129245 | 0.680104948 |
| hOSCAR | ASAT_Pos_pPS | 4159 | -0.029481471 | [-0.06 0. ] | 0.057648071 | 0.45329986 |
| hOSCAR | ALP_Neg_pPS | 4159 | 0.014576828 | [-0.02 0.04] | 0.348000936 | 0.79247738 |
| hOSCAR | SHBG-LpA_pPS | 4159 | 0.002135387 | [-0.03 0.03] | 0.890658007 | 0.981178631 |
| hOSCAR | Obesity_pPS | 4159 | 0.02553779 | [-0. 0.06] | 0.100107085 | 0.585371936 |
| hOSCAR | VAT_Neg_pPS | 4159 | 0.002577772 | [-0.03 0.03] | 0.868195102 | 0.977006629 |
| hOSCAR | Bilirubin_pPS | 4159 | 0.003023035 | [-0.03 0.03] | 0.845692977 | 0.974332356 |
| hOSCAR | Proinsulin_pPS | 4159 | 0.023179191 | [-0.01 0.05] | 0.135587603 | 0.629316669 |
| hOSCAR | Hyper_Insulin_pPS | 4159 | -0.005697874 | [-0.04 0.02] | 0.713753661 | 0.940764137 |
| hOSCAR | Beta_Cell_1_pPS | 4159 | -0.002670423 | [-0.03 0.03] | 0.863503373 | 0.977006629 |
| hOSCAR | Beta_Cell_2_pPS | 4159 | -0.020236854 | [-0.05 0.01] | 0.192596142 | 0.694856668 |
| hOSCAR | Total_GRS | 4159 | 0.015609746 | [-0.01 0.05] | 0.314905429 | 0.764236758 |
| PRELP | Cholesterol_pPS | 4158 | -0.006550993 | [-0.04 0.02] | 0.673249749 | 0.925361755 |
| PRELP | Lipodystrophy_1_pPS | 4158 | -0.033683247 | [-0.06 -0. ] | 0.030097357 | 0.369452038 |
| PRELP | Lipodystrophy_2_pPS | 4158 | 0.003393412 | [-0.03 0.03] | 0.827090452 | 0.966003345 |
| PRELP | Liver-Lipid_pPS | 4158 | 0.002691864 | [-0.03 0.03] | 0.862434729 | 0.977006629 |
| PRELP | ASAT_Pos_pPS | 4158 | -0.03318761 | [-0.06 -0. ] | 0.032608089 | 0.376562033 |

**b) Partial Correlation**

| Proteomics | PPS | Sample_Size | r | CI95% | p | fdr_p_adjusted |
|---|---|---|---|---|---|---|
| NT_proBNP | SHBG-LpA_pPS | 4225 | 0.016760299 | [-0.01 0.05] | 0.277503548 | 0.808820203 |
| NT_proBNP | Obesity_pPS | 4225 | -0.011863523 | [-0.04 0.02] | 0.442105657 | 0.866715504 |
| NT_proBNP | VAT_Neg_pPS | 4225 | -0.026568842 | [-0.06 0. ] | 0.085133099 | 0.614725404 |
| NT_proBNP | Bilirubin_pPS | 4225 | -0.014515957 | [-0.04 0.02] | 0.346955465 | 0.847284666 |
| NT_proBNP | Proinsulin_pPS | 4225 | 0.03215896 | [0. 0.06] | 0.037154941 | 0.457947978 |
| NT_proBNP | Hyper_Insulin_pPS | 4225 | 0.011679404 | [-0.02 0.04] | 0.4492222 | 0.870351501 |
| NT_proBNP | Beta_Cell_1_pPS | 4225 | -0.020398053 | [-0.05 0.01] | 0.186271754 | 0.742222258 |
| NT_proBNP | Beta_Cell_2_pPS | 4225 | 0.012840728 | [-0.02 0.04] | 0.405431042 | 0.852958996 |

**Table S7: Associations between pPSs and plasma proteins in SCAPIS**

**a) Spearman Correlation**

| Proteomics | PPS | Sample_Size | r | CI95% | p | fdr_p_adjusted |
|---|---|---|---|---|---|---|
| PRELP | ALP_Neg_pPS | 4158 | -0.008747326 | [-0.04 0.02] | 0.573382909 | 0.903411176 |
| PRELP | SHBG-LpA_pPS | 4158 | 0.045104344 | [0.01 0.08] | 0.003674338 | 0.116565202 |
| PRELP | Obesity_pPS | 4158 | 0.008501391 | [-0.02 0.04] | 0.584210685 | 0.903823705 |
| PRELP | VAT_Neg_pPS | 4158 | -0.009181945 | [-0.04 0.02] | 0.554483896 | 0.896004422 |
| PRELP | Bilirubin_pPS | 4158 | -0.002927023 | [-0.03 0.03] | 0.850552947 | 0.97528582 |
| PRELP | Proinsulin_pPS | 4158 | 0.024371563 | [-0.01 0.05] | 0.116640678 | 0.611872879 |
| PRELP | Hyper_Insulin_pPS | 4158 | -0.007329495 | [-0.04 0.02] | 0.637065194 | 0.915577927 |
| PRELP | Beta_Cell_1_pPS | 4158 | -0.021627795 | [-0.05 0.01] | 0.163818087 | 0.664061293 |
| PRELP | Beta_Cell_2_pPS | 4158 | -0.004115309 | [-0.03 0.03] | 0.791083673 | 0.957627604 |
| PRELP | Total_GRS | 4158 | -0.001010736 | [-0.03 0.03] | 0.948125348 | 0.993154165 |
| SCGB3A2 | Cholesterol_pPS | 4225 | 0.003528535 | [-0.03 0.03] | 0.818898785 | 0.963461584 |
| SCGB3A2 | Lipodystrophy_1_pPS | 4225 | -0.053030624 | [-0.08 -0.02] | 0.00057425 | 0.035220685 |
| SCGB3A2 | Lipodystrophy_2_pPS | 4225 | -0.018206408 | [-0.05 0.01] | 0.237411316 | 0.726631332 |
| SCGB3A2 | Liver-Lipid_pPS | 4225 | -0.011461096 | [-0.04 0.02] | 0.457048178 | 0.835860883 |
| SCGB3A2 | ASAT_Pos_pPS | 4225 | -0.034667819 | [-0.06 -0. ] | 0.024435333 | 0.333340061 |
| SCGB3A2 | ALP_Neg_pPS | 4225 | -0.027764732 | [-0.06 0. ] | 0.071552417 | 0.497442495 |
| SCGB3A2 | SHBG-LpA_pPS | 4225 | 0.002221067 | [-0.03 0.03] | 0.885404462 | 0.979291846 |
| SCGB3A2 | Obesity_pPS | 4225 | -0.017571101 | [-0.05 0.01] | 0.254182972 | 0.739680398 |
| SCGB3A2 | VAT_Neg_pPS | 4225 | -0.014217397 | [-0.04 0.02] | 0.356220774 | 0.7979754 |
| SCGB3A2 | Bilirubin_pPS | 4225 | -0.011648837 | [-0.04 0.02] | 0.44970936 | 0.831350198 |
| SCGB3A2 | Proinsulin_pPS | 4225 | 0.031001201 | [0. 0.06] | 0.044209309 | 0.424281318 |
| SCGB3A2 | Hyper_Insulin_pPS | 4225 | -0.025896674 | [-0.06 0. ] | 0.092826113 | 0.559254737 |
| SCGB3A2 | Beta_Cell_1_pPS | 4225 | 0.017602077 | [-0.01 0.05] | 0.253346597 | 0.739680398 |
| SCGB3A2 | Beta_Cell_2_pPS | 4225 | 0.014230546 | [-0.02 0.04] | 0.355776067 | 0.7979754 |
| SCGB3A2 | Total_GRS | 4225 | -0.031842013 | [-0.06 -0. ] | 0.038762408 | 0.39919495 |
| SHPS_1 | Cholesterol_pPS | 4225 | 0.01701052 | [-0.01 0.05] | 0.269651828 | 0.746520328 |
| SHPS_1 | Lipodystrophy_1_pPS | 4225 | 0.012538696 | [-0.02 0.04] | 0.415846945 | 0.817860704 |
| SHPS_1 | Lipodystrophy_2_pPS | 4225 | 0.038709625 | [0.01 0.07] | 0.011979554 | 0.234493393 |
| SHPS_1 | Liver-Lipid_pPS | 4225 | 0.034467228 | [0. 0.06] | 0.025274137 | 0.336988491 |
| SHPS_1 | ASAT_Pos_pPS | 4225 | -0.007087578 | [-0.04 0.02] | 0.645582899 | 0.916568313 |
| SHPS_1 | ALP_Neg_pPS | 4225 | -0.033619479 | [-0.06 -0. ] | 0.029099947 | 0.365072067 |
| SHPS_1 | SHBG-LpA_pPS | 4225 | 0.010603406 | [-0.02 0.04] | 0.491416887 | 0.859085266 |
| SHPS_1 | Obesity_pPS | 4225 | 0.00058149 | [-0.03 0.03] | 0.969901409 | 0.996862516 |
| SHPS_1 | VAT_Neg_pPS | 4225 | 0.003486822 | [-0.03 0.03] | 0.821003215 | 0.963461584 |
| SHPS_1 | Bilirubin_pPS | 4225 | 0.008396657 | [-0.02 0.04] | 0.585853263 | 0.903925569 |
| SHPS_1 | Proinsulin_pPS | 4225 | 0.007953464 | [-0.02 0.04] | 0.605787753 | 0.909483337 |
| SHPS_1 | Hyper_Insulin_pPS | 4225 | 0.020470815 | [-0.01 0.05] | 0.184027207 | 0.686251968 |
| SHPS_1 | Beta_Cell_1_pPS | 4225 | 0.010171474 | [-0.02 0.04] | 0.509236237 | 0.868660083 |
| SHPS_1 | Beta_Cell_2_pPS | 4225 | 0.008988686 | [-0.02 0.04] | 0.559709566 | 0.897095472 |
| SHPS_1 | Total_GRS | 4225 | 0.022493613 | [-0.01 0.05] | 0.144356466 | 0.641415816 |
| TFF3 | Cholesterol_pPS | 4225 | 0.021441315 | [-0.01 0.05] | 0.164088446 | 0.664061293 |
| TFF3 | Lipodystrophy_1_pPS | 4225 | 0.028542303 | [-0. 0.06] | 0.063962556 | 0.47584004 |
| TFF3 | Lipodystrophy_2_pPS | 4225 | -0.003867119 | [-0.03 0.03] | 0.801866691 | 0.958074488 |
| TFF3 | Liver-Lipid_pPS | 4225 | 0.005897541 | [-0.02 0.04] | 0.70195326 | 0.93593768 |
| TFF3 | ASAT_Pos_pPS | 4225 | -0.006702578 | [-0.04 0.02] | 0.663617428 | 0.924111434 |
| TFF3 | ALP_Neg_pPS | 4225 | -0.002606571 | [-0.03 0.03] | 0.865689327 | 0.977006629 |
| TFF3 | SHBG-LpA_pPS | 4225 | 0.007679349 | [-0.02 0.04] | 0.618267161 | 0.909483337 |
| TFF3 | Obesity_pPS | 4225 | 0.014891548 | [-0.02 0.05] | 0.333873182 | 0.779602355 |
| TFF3 | VAT_Neg_pPS | 4225 | -0.023432727 | [-0.05 0.01] | 0.128330081 | 0.623575746 |
| TFF3 | Bilirubin_pPS | 4225 | 0.017888466 | [-0.01 0.05] | 0.245704457 | 0.735514428 |
| TFF3 | Proinsulin_pPS | 4225 | 0.019346167 | [-0.01 0.05] | 0.209312315 | 0.709689105 |
| TFF3 | Hyper_Insulin_pPS | 4225 | -3.59E-05 | [-0.03 0.03] | 0.998141173 | 0.999464614 |
| TFF3 | Beta_Cell_1_pPS | 4225 | 0.017257396 | [-0.01 0.05] | 0.262761645 | 0.746520328 |
| TFF3 | Beta_Cell_2_pPS | 4225 | 0.010859083 | [-0.02 0.04] | 0.481028737 | 0.852690632 |
| TFF3 | Total_GRS | 4225 | 0.018788009 | [-0.01 0.05] | 0.222756594 | 0.722453819 |
| TLT_2 | Cholesterol_pPS | 4225 | -0.001343772 | [-0.03 0.03] | 0.930516369 | 0.990056084 |
| TLT_2 | Lipodystrophy_1_pPS | 4225 | 0.013800368 | [-0.02 0.04] | 0.370505986 | 0.799802236 |
| TLT_2 | Lipodystrophy_2_pPS | 4225 | 0.007503966 | [-0.02 0.04] | 0.626310124 | 0.913767992 |
| TLT_2 | Liver-Lipid_pPS | 4225 | 0.030630657 | [0. 0.06] | 0.046807296 | 0.434976894 |
| TLT_2 | ASAT_Pos_pPS | 4225 | -0.00667749 | [-0.04 0.02] | 0.66479958 | 0.924111434 |
| TLT_2 | ALP_Neg_pPS | 4225 | 0.029691193 | [-0. 0.06] | 0.053974918 | 0.446020284 |
| TLT_2 | SHBG-LpA_pPS | 4225 | 0.006076977 | [-0.02 0.04] | 0.693338321 | 0.930013012 |
| TLT_2 | Obesity_pPS | 4225 | 0.004822442 | [-0.03 0.04] | 0.754339763 | 0.950403646 |
| TLT_2 | VAT_Neg_pPS | 4225 | 0.001746959 | [-0.03 0.03] | 0.909747224 | 0.990056084 |
| TLT_2 | Bilirubin_pPS | 4225 | -0.000378388 | [-0.03 0.03] | 0.980411529 | 0.996862516 |
| TLT_2 | Proinsulin_pPS | 4225 | 0.031398392 | [0. 0.06] | 0.04156066 | 0.408211461 |
| TLT_2 | Hyper_Insulin_pPS | 4225 | -0.01044232 | [-0.04 0.02] | 0.4980231 | 0.863780476 |
| TLT_2 | Beta_Cell_1_pPS | 4225 | 0.012020967 | [-0.02 0.04] | 0.435361982 | 0.828998134 |
| TLT_2 | Beta_Cell_2_pPS | 4225 | 0.017217511 | [-0.01 0.05] | 0.263866515 | 0.746520328 |
| TLT_2 | Total_GRS | 4225 | 0.017100308 | [-0.01 0.05] | 0.267131698 | 0.746520328 |
| NT_proBNP | Cholesterol_pPS | 4225 | 0.000973855 | [-0.03 0.03] | 0.94961371 | 0.993154165 |
| NT_proBNP | Lipodystrophy_1_pPS | 4225 | -0.018051501 | [-0.05 0.01] | 0.241426842 | 0.729918816 |
| NT_proBNP | Lipodystrophy_2_pPS | 4225 | 0.003965458 | [-0.03 0.03] | 0.796937104 | 0.958074488 |
| NT_proBNP | Liver-Lipid_pPS | 4225 | 0.020865555 | [-0.01 0.05] | 0.175711119 | 0.678995855 |
| NT_proBNP | ASAT_Pos_pPS | 4225 | -0.004719419 | [-0.03 0.03] | 0.759424146 | 0.950403646 |
| NT_proBNP | ALP_Neg_pPS | 4225 | 0.019319068 | [-0.01 0.05] | 0.209951152 | 0.709689105 |
| NT_proBNP | SHBG-LpA_pPS | 4225 | 0.015490804 | [-0.01 0.05] | 0.314784531 | 0.764236758 |
| NT_proBNP | Obesity_pPS | 4225 | -0.00768742 | [-0.04 0.02] | 0.61789808 | 0.909483337 |
| NT_proBNP | VAT_Neg_pPS | 4225 | -0.025249001 | [-0.06 0. ] | 0.101290839 | 0.586731767 |
| NT_proBNP | Bilirubin_pPS | 4225 | -0.015944551 | [-0.05 0.01] | 0.300817631 | 0.76319014 |
| NT_proBNP | Proinsulin_pPS | 4225 | 0.031522657 | [0. 0.06] | 0.040760099 | 0.407062052 |
| NT_proBNP | Hyper_Insulin_pPS | 4225 | 0.00847197 | [-0.02 0.04] | 0.582496245 | 0.903823705 |
| NT_proBNP | Beta_Cell_1_pPS | 4225 | -0.012408908 | [-0.04 0.02] | 0.420689996 | 0.821148791 |
| NT_proBNP | Beta_Cell_2_pPS | 4225 | 0.012432504 | [-0.02 0.04] | 0.419807039 | 0.820585997 |
| NT_proBNP | Total_GRS | 4225 | -0.004229063 | [-0.03 0.03] | 0.783763251 | 0.955343908 |

**b) Partial Correlation**

| Proteomics | PPS | Sample_Size | r | CI95% | p | fdr_p_adjusted |
|---|---|---|---|---|---|---|

**Table S8: Protein functional and pathway annotations**

| Variable_Name | Uniprot | Gene | Extracted_Pathway_Annotation | Extracted_Disease_Annotation | Uniport_Annotation | Elsevier_Annotation | KEGG_2021_Human_Annotation | MSigDB_Hallmark_2020_Annotation |
|---|---|---|---|---|---|---|---|---|
| ACE2 | Q9BYF1 | ACE2 | Energy Homeostasis | Cardiovascular Diseases | Enzymes, Metabolic proteins, Transporters | Proteins Involved in Myocardial Ischemia\|I | Coronavirus disease\|Renin-angiotensin system\|Protein digestion and absorption | |
| ADAM TS13 | Q76LX8 | ADAMTS13 | Complement and Coagulation Cascades | | Disease related genes, Enzymes, Human di | Proteins Involved in Thrombophilia\|Proteins Involved in Antiphospholipid Syndrome | | |
| ADM | P35318 | ADM | Immune Response and Inflammation | Cardiovascular Diseases | Cancer-related genes, Plasma proteins, Ho | Proteins Involved in Atherosclerosis\|Protei | Vascular smooth muscle contraction\|Neur | Inflammatory Response\|Hypoxia |
| AGRP | O00253 | AGRP | Immune Response and Inflammation | Metabolic Disorders | Disease related genes, Human disease rela | Proteins Involved in Obesity\|Proteins Invol | Adipocytokine signaling pathway | |
| ALCAM | Q13740 | ALCAM | Immune Response and Inflammation | | Cancer-related genes, CD markers, Plasma | Proteins Involved in Melanoma\|Hematopc | Cell adhesion molecules | IL-2/STAT5 Signaling\|Cholesterol Homeostasis |
| AMBP | P02760 | AMBP | Enzymatic and Detoxification | | Plasma proteins, Host-virus interaction, Ox | Proteins Involved in Female Infertility | | |
| ANG 1 | Q15389 | ANGPT1 | Lipid Metabolism | Cardiovascular Diseases | Cancer-related genes, Disease related gene | Proteins Involved in Myocardial Ischemia\|I | Rheumatoid arthritis\|PI3K-Akt signaling pa | Adipogenesis |
| AP N | P15144 | ANPEP | Energy Homeostasis | Cardiovascular Diseases | CD markers, Enzymes, Metabolic proteins, | Proteins Involved in Arterial Hypertension\| | Hematopoietic cell lineage\|Renin-angioter | Epithelial Mesenchymal Transition |
| AXL | P30530 | AXL | Immune Response and Inflammation | Cancer-Related Diseases | Disease related genes, Enzymes, FDA appro | Proteins Involved in Glomerulonephritis\|Proteins Involved in Glioma\|Proteins with Alt | | Inflammatory Response |
| AZU1 | P20160 | AZU1 | Enzymatic and Detoxification | | Enzymes, Plasma proteins, Chemotaxis, Antibiotic, Antimicrobial, Heparin-binding, Ser | | Neutrophil extracellular trap formation | |
| BLM hydrolase | Q13867 | BLMH | Enzymatic and Detoxification | | Cancer-related genes, Enzymes, Plasma proteins, Hydrolase, Protease, Thiol protease | | | |
| BMP 6 | P22004 | BMP6 | Immune Response and Inflammation | Cardiovascular Diseases | Chondrogenesis, Differentiation, Osteogen | Proteins Involved in Cholesteatoma\|BMP S | Cytokine-cytokine receptor interaction\|TG | Bile Acid Metabolism |
| BNP | P16860 | NPPB | Cardiovascular Function | Cardiovascular Diseases | Candidate cardiovascular disease genes, Pl | Proteins Involved in Myocardial Ischemia\|I | Vascular smooth muscle contraction\|Thermogenesis\|cGMP-PKG signaling pathway | |
| BOC | Q9BWV1 | BOC | Growth and Cell Signaling | | Cell adhesion | Hedgehog Family Signaling | Axon guidance\|Hedgehog signaling pathway | |
| CA5A | P35218 | CA5A | Energy Homeostasis | | Disease related genes, Enzymes, FDA approved drug targets, Human disease related ge | | Nitrogen metabolism | |
| CASP 3 | P42574 | CASP3 | Immune Response and Inflammation | Cardiovascular Diseases | Cancer-related genes, Enzymes, FDA appro | Proteins Involved in Myocardial Ischemia\|I | Lipid and atherosclerosis\|TNF signaling pa | Complement\|Apoptosis\|IL-2/STAT5 Signaling\|Interferon Gamma Response\|UV Response Up |
| CCL15 | Q16663 | CCL15 | Immune Response and Inflammation | Cardiovascular Diseases | Plasma proteins, Chemotaxis, Cytokine, He | Proteins Involved in Atherosclerosis\|CCR1 | Cytokine-cytokine receptor interaction\|Viral protein interaction with cytokine and cytokine receptor\|Chemokine signaling pathway | |
| CCL16 | O15467 | CCL16 | Immune Response and Inflammation | | Cancer-related genes, Plasma proteins, Ch | CCR1 Expression Targets\|Nociception Expr | Cytokine-cytokine receptor interaction\|Viral protein interaction with cytokine and cytokine receptor\|Chemokine signaling pathway | |
| CCL17 | Q92583 | CCL17 | Immune Response and Inflammation | Cardiovascular Diseases | Chemotaxis, Inflammatory response, Cytok | Proteins Involved in Systemic Scleroderma | Cytokine-cytokine receptor interaction\|Vir | Inflammatory Response |
| CCL24 | O00175 | CCL24 | Immune Response and Inflammation | Inflammatory and Immune-Related Diseases | Chemotaxis, Inflammatory response, Cytok | anti-Inflammatory Function of Macrophag | Cytokine-cytokine receptor interaction\|Vir | Inflammatory Response |
| CCL3 | P10147 | CCL3 | Immune Response and Inflammation | Cardiovascular Diseases | Cancer-related genes, Chemotaxis, Inflamr | Proteins Involved in Atherosclerosis\|Protei | Cytokine-cytokine receptor interaction\|Viral protein interaction with cytokine and cytokine receptor\|Lipid and atherosclerosis\|Rheumatoid arthritis\|Chemokine signaling pathway\|Human cytomegalovirus infection\|Chagas disease\|Toll-like receptor signaling pathway | |
| CD163 | Q86VB7 | CD163 | Immune Response and Inflammation | Inflammatory and Immune-Related Diseases | CD markers, Plasma proteins, Acute phase | Proteins Involved in Endometriosis\|anti-Inflammatory Function of Macrophage M2 Lineage\|Macrophage Related Iron Uptake and Release\|Hemoglobin Reduction and Leukocyte Adhesion Initiate Vasospasm | | |
| CD4 | P01730 | CD4 | Immune Response and Inflammation | Cardiovascular Diseases | CD markers, Disease related genes, FDA ap | Proteins Involved in Atherosclerosis\|Prote | Cytokine-cytokine receptor interaction\|Cel | Allograft Rejection |
| CD40 L | P29965 | CD40LG | Immune Response and Inflammation | Cardiovascular Diseases | Cancer-related genes, Candidate cardiovas | Proteins Involved in Myocardial Ischemia\|I | Cytokine-cytokine receptor interaction\|Lip | Complement\|Allograft Rejection\|KRAS Signaling Dn |
| CD84 | Q9UIB8 | CD84 | Immune Response and Inflammation | | CD markers, Adaptive immunity, Autophagy, Cell adhesion, Immunity, Innate immunity, Receptor | | | |
| CD93 | Q9NPY3 | CD93 | Other Functions | | CD markers, Plasma proteins, Cell adhesior | Hemapoietic Stem Cell -> Lymphoid Multipotent Progenitor Surface Expression Markers\|Complement System Defects in Systemic Lupus Erythematosis | | |
| CDH5 | P33151 | CDH5 | Cardiovascular Function | Cardiovascular Diseases | Cancer-related genes, CD markers, Plasma | Proteins Involved in Melanoma\|Proteins Ir | Fluid shear stress and atherosclerosis\|Cell adhesion molecules\|Leukocyte transendothelial migration | |
| CEACAM8 | P31997 | CEACAM8 | Other Functions | | CD markers, Plasma proteins, Cell adhesior | Myoblast -> Neutrophil Surface Expression Markers | | |
| CHI3L1 | P36222 | CHI3L1 | Immune Response and Inflammation | | Cancer-related genes, Disease related gene | Proteins Involved in Chronic Obstructive Pulmonary Disease\|Proteins Involved in Glioma\|Proteins Involved in Age-Related Macular Degeneration\|Proteins Involved in Psoriatic Arthritis\|Glioblastoma, Mesenchymal Subtype | | |
| CHIT1 | Q13231 | CHIT1 | Energy Homeostasis | Metabolic Disorders | Candidate cardiovascular disease genes, Er | Proteins Involved in non-Alcoholic Fatty Liv | Amino sugar and nucleotide sugar metabolism | |
| CNTN1 | Q12860 | CNTN1 | Growth and Cell Signaling | | Disease related genes, Human disease related genes, Plasma proteins, Cell adhesion, N | | Cell adhesion molecules | Apical Junction |
| COL1A1 | P02452 | COL1A1 | Cardiovascular Function | Metabolic Disorders | Cancer-related genes, Disease related gene | Proteins Involved in Osteoporosis\|Hepatic | PI3K-Akt signaling pathway\|AGE-RAGE sig | Epithelial Mesenchymal Transition\|UV Response Dn\|Myogenesis |
| CPA1 | P15085 | CPA1 | Energy Homeostasis | | Enzymes, Metabolic proteins, Carboxypeptidase, Hydrolase, Metalloprotease, Proteas | | Protein digestion and absorption\|Pancreatic secretion | |
| CPB1 | P15086 | CPB1 | Growth and Cell Signaling | | Enzymes, Plasma proteins, Carboxypeptidase, Hydrolase, Metalloprotease, Protease | | Protein digestion and absorption\|Pancreat | KRAS Signaling Dn |
| CSTB | P04080 | CSTB | Enzymatic and Detoxification | | Cancer-related genes, Disease related gene | Proteins Involved in Congenital Deafness | | |
| CTRC | Q99895 | CTRC | Enzymatic and Detoxification | | Disease related genes, Enzymes, Human disease related genes, Potential drug targets, Hydrolase, Protease, Serine protease | | | |
| CTSD | P07339 | CTSD | Lipid Metabolism | Cardiovascular Diseases | Cancer-related genes, Disease related gene | Proteins Involved in Myocardial Ischemia\|I | Apoptosis\|Tuberculosis\|Lysosome\|Estrog | Complement\|p53 Pathway |
| CTSL1 | P07711 | CTSL | Energy Homeostasis | | Cancer-related genes, Enzymes, Metabolic | Proteins Involved in Melanoma\|Alveolar M | Fluid shear stress and atherosclerosis\|Rhe | Complement |
| CTSZ | Q9UBR2 | CTSZ | Energy Homeostasis | | Enzymes, Metabolic proteins, Plasma proteins, Hydrolase, Protease, Thiol protease | | Apoptosis\|Lysosome | IL-2/STAT5 Signaling |
| CXCL1 | P09341 | CXCL1 | Immune Response and Inflammation | Cardiovascular Diseases | Cancer-related genes, Plasma proteins, Inf | Proteins Involved in Atherosclerosis\|Prote | Cytokine-cytokine receptor interaction\|Vir | Epithelial Mesenchymal Transition\|IL-6/JAK/STAT3 Signaling\|Complement\|TNF-alpha Signaling via NF-kB |
| CXCL16 | Q9H2A7 | CXCL16 | Immune Response and Inflammation | Cardiovascular Diseases | Transporters, Chemotaxis, Cytokine | Proteins Involved in Glomerulonephritis\|P | Cytokine-cytokine receptor interaction\|Ch | Cholesterol Homeostasis |
| DCN | P07585 | DCN | Growth and Cell Signaling | Cardiovascular Diseases | Cancer-related genes, Disease related gene | Proteins Involved in Atherosclerosis\|Prote | Proteoglycans in cancer\|TGF-beta signalin | Epithelial Mesenchymal Transition\|Apoptosis\|Hypoxia\|Glycolysis |
| DECR1 | Q16698 | DECR1 | Lipid Metabolism | | Disease related genes, Enzymes, Metabolic proteins, Plasma proteins, Potential drug targets, Fatty acid metabolism, Lipid metabo | | | Adipogenesis\|Fatty Acid Metabolism\|Oxidative Phosphorylation |
| Dkk 1 | O94907 | DKK1 | Growth and Cell Signaling | Metabolic Disorders | Wnt signaling pathway, Developmental pro | Proteins Involved in Diabetic Nephropathy | Alzheimer disease\|Pathways of neurodege | Epithelial Mesenchymal Transition\|Wnt-beta Catenin Signaling |
| DLK 1 | P80370 | DLK1 | Other Functions | | | Proteins Involved in Glioma\|Proteins Involved in Glioblastoma\|PHOX2B Effects in Neuroblastoma\|Neuroblastoma | | |
| EGFR | P00533 | EGFR | Energy Homeostasis | Cardiovascular Diseases | Cancer-related genes, Disease related gene | Proteins Involved in Atherosclerosis\|Prote | PI3K-Akt signaling pathway\|MAPK signalin | Allograft Rejection\|Hypoxia\|Apical Junction\|Glycolysis\|PI3K/AKT/mTOR Signaling\|Protein Secretion |
| Ep CAM | P16422 | EPCAM | Other Functions | Cancer-Related Diseases | Cancer-related genes, CD markers, Disease | Proteins with Altered Expression in Cancer Metastases\|Proteins Involved in Hepatocel | | UV Response Up |
| EPHB4 | P54760 | EPHB4 | Cardiovascular Function | Cancer-Related Diseases | Cancer-related genes, Disease related gene | Proteins with Altered Expression in Cancer | Axon guidance | Apical Surface |
| FABP2 | P12104 | FABP2 | Lipid Metabolism | Cardiovascular Diseases | Cancer-related genes, Metabolic proteins, | Proteins Involved in Myocardial Ischemia | PPAR signaling pathway\|Fat digestion and | Fatty Acid Metabolism |
| FABP4 | P15090 | FABP4 | Lipid Metabolism | Metabolic Disorders | Cancer-related genes, Metabolic proteins, | Proteins Involved in non-Alcoholic Fatty Liv | PPAR signaling pathway\|Regulation of lipo | Adipogenesis |
| FAS | P25445 | FAS | Immune Response and Inflammation | Cardiovascular Diseases | Cancer-related genes, Candidate cardiovas | Proteins Involved in Myocardial Ischemia\|I | Cytokine-cytokine receptor interaction\|Lip | Epithelial Mesenchymal Transition\|IL-6/JAK/STAT3 Signaling\|Allograft Rejection\|Apoptosis\|Interferon Gamma Response\|Xenobiotic Metabolism\|p53 Pathway |
| FGF 21 | Q9NSA1 | FGF21 | Growth and Cell Signaling | Metabolic Disorders | RAS pathway related proteins, Growth fact | Proteins Involved in non-Alcoholic Fatty Liv | PI3K-Akt signaling pathway\|MAPK signaling pathway\|Pathways in cancer\|Rap1 signaling pathway\|Ras signaling pathway\|Melanoma\|Regulation of actin cytoskeleton\|Calcium signaling pathway\|Breast cancer\|Gastric cancer\|Thermogenesis | |
| FGF 23 | Q9GZV9 | FGF23 | Growth and Cell Signaling | | Cancer-related genes, Disease related gene | Vitamin D Deficite and Dentin Formation\| | PI3K-Akt signaling pathway\|MAPK signaling pathway\|Pathways in cancer\|Rap1 signaling pathway\|Ras signaling pathway\|Melanoma\|Regulation of actin cytoskeleton\|Calcium signaling pathway\|Breast cancer\|Parathyroid hormone synthesis, secretion and action\|Gastric cancer | |
| FS | P19883 | FST | Cardiovascular Function | | Cancer-related genes, Candidate cardiovas | Proteins Involved in Polycystic Ovary Syndr | TGF-beta signaling pathway | Myogenesis |
| Gal 3 | P17931 | LGALS3 | Immune Response and Inflammation | Metabolic Disorders | Cancer-related genes, Plasma proteins, Dif | Proteins Involved in Endometriosis\|Proteins Involved in Chronic Obstructive Pulmonar | | Complement\|Apoptosis\|Cholesterol Homeostasis |
| Gal 4 | P56470 | LGALS4 | Other Functions | | Cancer-related genes, Plasma proteins | | | |
| Gal 9 | O00182 | LGALS9 | Immune Response and Inflammation | Inflammatory and Immune-Related Diseases | Chemotaxis, Immunity | Proteins Involved in Myocarditis\|Proteins Involved in Melanoma\|Proteins Involved in Ulcerative Colitis\|Uric Acid Excretion Impairment in Kidney in Gout | | |
| GDF 15 | Q99988 | GDF15 | Immune Response and Inflammation | Cardiovascular Diseases | Cancer-related genes, Disease related gene | Proteins Involved in Myocardial Ischemia\|I | Cytokine-cytokine receptor interaction | |
| GDF 2 | Q9UK05 | GDF2 | Immune Response and Inflammation | | Disease related genes, Human disease rela | Proteins Involved in Osteoporosis\|Heredit | Cytokine-cytokine receptor interaction | |
| GH | P01241 | GH1 | Immune Response and Inflammation | Cardiovascular Diseases | Cancer-related genes, Candidate cardiovas | Proteins Involved in Atherosclerosis\|Prote | Cytokine-cytokine receptor interaction\|PI3K-Akt signaling pathway\|JAK-STAT signaling pathway\|Neuroactive ligand-receptor interaction\|Growth hormone synthesis, secretion and action | |
| GIF | P27352 | CBLIF | Energy Homeostasis | | Disease related genes, Human disease rela | P-cell Activation | Vitamin digestion and absorption | |
| GLO1 | Q04760 | GLO1 | Energy Homeostasis | | Cancer-related genes, Enzymes, Metabolic proteins, Plasma proteins, Lyase | | Pyruvate metabolism | Myc Targets V1 |
| GP6 | Q9HCN6 | GP6 | Complement and Coagulation Cascades | Inflammatory and Immune-Related Diseases | Disease related genes, Human disease rela | Megakaryocyte -> Platelet Precursor Cell S | ECM-receptor interaction\|Platelet activation | |
| GRN | P28799 | GRN | Immune Response and Inflammation | | Disease related genes, Human disease rela | Proteins Involved in Amyotrophic Lateral Sclerosis\|Proteins Involved in Polycystic Kidney Disease\|Genes with Mutations Associated with Frontotemporal Dementia\|Cortisol in Resolving Inflammation\|Proteins Involved in Frontotemporal Dementia | | |
| GT | P51161 | FABP6 | Lipid Metabolism | | Metabolic proteins, Lipid transport, Transport | | PPAR signaling pathway | Peroxisome |
| HAOX1 | Q9UJM8 | HAO1 | Energy Homeostasis | | Enzymes, FDA approved drug targets, Metabolic proteins, Amino-acid biosynthesis, Ox | | Peroxisome\|Glyoxylate and dicarboxylate | Bile Acid Metabolism |
| HB EGF | Q99075 | HBEGF | Immune Response and Inflammation | Cardiovascular Diseases | Growth factor, Heparin-binding, Receptor | Proteins Involved in Atherosclerosis\|Prote | Proteoglycans in cancer\|Epithelial cell sign | Inflammatory Response\|KRAS Signaling Up\|TNF-alpha Signaling via NF-kB\|p53 Pathway\|Myogenesis |
| HO 1 | P09601 | HMOX1 | Energy Homeostasis | Cardiovascular Diseases | Cancer-related genes, Disease related gene | Proteins Involved in Myocardial Ischemia\|I | Fluid shear stress and atherosclerosis\|Path | IL-6/JAK/STAT3 Signaling\|Apoptosis\|Hypoxia\|UV Response Up\|Xenobiotic Metabolism\|p53 Pathway |
| hOSCAR | Q8IYS5 | OSCAR | Other Functions | | Receptor | Osteoclast Activation in Multiple Myeloma | Osteoclast differentiation | |
| HSP 27 | P04792 | HSPB1 | Growth and Cell Signaling | Cardiovascular Diseases | Cancer-related genes, Disease related gene | Proteins Involved in Myocardial Ischemia\|I | MAPK signaling pathway\|Amoebiasis\|VEG | Apoptosis\|Apical Surface |
| ICAM 2 | P13598 | ICAM2 | Immune Response and Inflammation | Metabolic Disorders | CD markers, Plasma proteins, Cell adhesior | Megakaryocyte -> Platelet Precursor Cell S | Cell adhesion molecules\|Natural killer cell | Apical Junction |
| IDUA | P35475 | IDUA | Energy Homeostasis | | Disease related genes, Enzymes, Human disease related genes, Metabolic proteins, Pla | | Lysosome\|Glycosaminoglycan degradation | Glycolysis\|KRAS Signaling Dn |
| IGFBP 1 | P08833 | IGFBP1 | Cardiovascular Function | Cardiovascular Diseases | Candidate cardiovascular disease genes, Pl | Proteins Involved in Arterial Hypertension\|Proteins Involved in Polycystic Ovary Syndr | | Hypoxia\|Xenobiotic Metabolism\|Unfolded Protein Response |
| IGFBP 2 | P18065 | IGFBP2 | Growth and Cell Signaling | Cancer-Related Diseases | Cancer-related genes, Plasma proteins, Gr | Proteins Involved in Glioma\|Proteins Involved in Glioblastoma\|Proteins Involved in As | | Epithelial Mesenchymal Transition\|UV Response Up\|KRAS Signaling Dn |
| IGFBP 7 | Q16270 | IGFBP7 | Cardiovascular Function | | Disease related genes, Plasma proteins, Ce | Proteins Involved in Glioma\|Proteins Involved in Colorectal Neoplasms | | Myogenesis |
| IgG Fc receptor II b | P31994 | FCGR2B | Growth and Cell Signaling | Inflammatory and Immune-Related Diseases | Cancer-related genes, CD markers, Disease | Proteins Involved in Glomerulonephritis\|P | Osteoclast differentiation\|Phagosome\|Tul | Allograft Rejection |
| IL 17D | Q8TAD2 | IL17D | Immune Response and Inflammation | | Cytokine | | Cytokine-cytokine receptor interaction\|IL-17 signaling pathway\|JAK-STAT signaling pathway\|Th17 cell differentiation\|C-type lectin receptor signaling pathway | |
| IL 17RA | Q96F46 | IL17RA | Immune Response and Inflammation | Cancer-Related Diseases | CD markers, Disease related genes, Human | Proteins Involved in Endometriosis\|Protein | Cytokine-cytokine receptor interaction\|IL- | IL-6/JAK/STAT3 Signaling |
| IL 18BP | O95998 | IL18BP | Other Functions | | Disease related genes, Human disease related genes | | | Interferon Gamma Response |
| IL 1ra | P18510 | IL1RN | Immune Response and Inflammation | Cardiovascular Diseases | Cancer-related genes, Candidate cardiovas | Proteins Involved in Endometriosis\|Protein | Cytokine-cytokine receptor interaction | |
| IL 1RT1 | P14778 | IL1R1 | Immune Response and Inflammation | Metabolic Disorders | Cancer-related genes, CD markers, Enzyme | Proteins Involved in Endometriosis\|Protein | Cytokine-cytokine receptor interaction\|Flu | Inflammatory Response\|IL-6/JAK/STAT3 Signaling\|Xenobiotic Metabolism |
| IL 1RT2 | P27930 | IL1R2 | Immune Response and Inflammation | Inflammatory and Immune-Related Diseases | Cancer-related genes, CD markers, Plasma | Proteins Involved in Endometriosis\|Protein | Cytokine-cytokine receptor interaction\|Flu | IL-6/JAK/STAT3 Signaling\|IL-2/STAT5 Signaling |
| IL 27 | Q8NEV9 | IL27 | Immune Response and Inflammation | Cardiovascular Diseases | Immunity, Inflammatory response, Innate | Proteins Involved in Glomerulonephritis\|P | Cytokine-cytokine receptor interaction | |
| IL 4RA | P24394 | IL4R | Immune Response and Inflammation | Metabolic Disorders | Cancer-related genes, CD markers, FDA ap | Proteins Involved in Glioma\|anti-Inflamma | Cytokine-cytokine receptor interaction\|PI3 | Inflammatory Response\|IL-6/JAK/STAT3 Signaling\|Allograft Rejection\|IL-2/STAT5 Signaling\|Interferon Gamma Response\|Interferon Alpha Response |
| IL 6RA | P08887 | IL6R | Immune Response and Inflammation | Cardiovascular Diseases | Cancer-related genes, Candidate cardiovas | Proteins Involved in Endometriosis\|Protein | Cytokine-cytokine receptor interaction\|Viral protein interaction with cytokine and cytokine receptor\|PI3K-Akt signaling pathway\|Hematopoietic cell lineage\|Pathways in cancer\|JAK-STAT signaling pathway\|HIF-1 signaling pathway\|Th17 cell differentiation\|Human cytomegalovirus infection\|Coronavirus disease\|Non-alcoholic fatty liver disease | |
| IL16 | Q14005 | IL16 | Immune Response and Inflammation | Inflammatory and Immune-Related Diseases | Cancer-related genes, Human disease relat | Proteins Involved in Endometriosis\|Protein | Cytokine-cytokine receptor interaction | Allograft Rejection |
| IL18 | Q14116 | IL18 | Immune Response and Inflammation | Cardiovascular Diseases | Cancer-related genes, Candidate cardiovas | Proteins Involved in Atherosclerosis\|Prote | Cytokine-cytokine receptor interaction\|Vir | Inflammatory Response\|Allograft Rejection\|Apoptosis\|TNF-alpha Signaling via NF-kB |
| IL1RL2 | Q9HB29 | IL1RL2 | Immune Response and Inflammation | | Enzymes, FDA approved drug targets, Imm | IL36 Receptor Antagonist (IL1F5) Deficienc | Cytokine-cytokine receptor interaction | KRAS Signaling Up |
| IL2 RA | P01589 | IL2RA | Immune Response and Inflammation | Cardiovascular Diseases | Cancer-related genes, CD markers, Disease | Proteins Involved in Multiple Sclerosis\|Pro | Cytokine-cytokine receptor interaction\|Vir | IL-6/JAK/STAT3 Signaling\|Allograft Rejection\|IL-2/STAT5 Signaling |
| IL6 | P05231 | IL6 | Immune Response and Inflammation | Cardiovascular Diseases | Cancer-related genes, Candidate cardiovas | Proteins Involved in Atherosclerosis\|Prote | Cytokine-cytokine receptor interaction\|Vir | Inflammatory Response\|Epithelial Mesenchymal Transition\|IL-6/JAK/STAT3 Signaling\|Complement\|Allograft Rejection\|Apoptosis\|TNF-alpha Signaling via NF-kB\|Hypoxia\|Interferon Gamma Response\|UV Response Up |
| ITGB1BP2 | Q9UKP3 | ITGB1BP2 | Growth and Cell Signaling | Cardiovascular Diseases | | Proteins Involved in Dilated Cardiomyopathy | | KRAS Signaling Dn |
| ITGB2 | P05107 | ITGB2 | Immune Response and Inflammation | Cardiovascular Diseases | CD markers, Disease related genes, Human | Proteins Involved in Atherosclerosis\|Prote | Rheumatoid arthritis\|Malaria\|Cell adhesic | KRAS Signaling Up\|Allograft Rejection\|mTORC1 Signaling |
| JAM A | Q9Y624 | F11R | Cardiovascular Function | Cardiovascular Diseases | CD markers, Plasma proteins, Host-virus in | Proteins Involved in Atherosclerosis\|Leuko | Cell adhesion molecules\|Epithelial cell signaling in Helicobacter pylori infection\|Leukocyte transendothelial migration\|Tight junction | |
| KIM1 | Q96D42 | HAVCR1 | Other Functions | | CD markers, Host-virus interaction, Host ce | T-lymphoid precursor cell -> CD8+ Naive T-cell Surface Expression Markers\|Eat me Signal: Apoptotic Cell Initiates Phagocytosis\|Proteins Involved in Polycystic Kidney Disease | | |
| KLK6 | Q92876 | KLK6 | Enzymatic and Detoxification | | Cancer-related genes, Enzymes, Plasma pr | Proteins Involved in Alzheimer's Disease\|Proteins Involved in Multiple Sclerosis\|Proteins Involved in Parkinson's Disease\|Proteins Involved in Vascular Dementia | | |
| LDL receptor | P01130 | LDLR | Immune Response and Inflammation | Cardiovascular Diseases | Disease related genes, Human disease rela | Proteins Involved in Atherosclerosis\|Prote | Lipid and atherosclerosis\|Cholesterol met | Inflammatory Response\|TNF-alpha Signaling via NF-kB\|Cholesterol Homeostasis\|UV Response Dn\|mTORC1 Signaling |
| LEP | P41159 | LEP | Immune Response and Inflammation | Cardiovascular Diseases | Cancer-related genes, Candidate cardiovas | Proteins Involved in Atherosclerosis\|Prote | Cytokine-cytokine receptor interaction\|JA | Adipogenesis |
| LOX 1 | P78380 | OLR1 | Immune Response and Inflammation | Cardiovascular Diseases | Disease related genes, Human disease rela | Proteins Involved in Atherosclerosis\|Prote | Lipid and atherosclerosis\|Phagosome\|PPA | Inflammatory Response\|Complement\|Coagulation\|TNF-alpha Signaling via NF-kB\|Angiogenesis |
| LPL | P06858 | LPL | Lipid Metabolism | Cardiovascular Diseases | Candidate cardiovascular disease genes, Di | Proteins Involved in Atherosclerosis\|Prote | PPAR signaling pathway\|Cholesterol metal | Angiogenesis\|Cholesterol Homeostasis\|Adipogenesis |
| LTBR | P36941 | LTBR | Immune Response and Inflammation | Cancer-Related Diseases | Cancer-related genes, Plasma proteins, Ap | Multiple Myeloma\|NFKB Signaling Activati | Cytokine-cytokine receptor interaction\|Vir | IL-6/JAK/STAT3 Signaling |
| MARCO | Q9UEW3 | MARCO | Immune Response and Inflammation | Cardiovascular Diseases | Plasma proteins, Immunity, Innate immuni | Proteins Involved in Atherosclerosis | Phagosome | Inflammatory Response |
| MB | P02144 | MB | Cardiovascular Function | Cardiovascular Diseases | Candidate cardiovascular disease genes, Fl | Proteins Involved in Myocardial Ischemia\|Proteins Involved in Myocarditis | | Myogenesis |
| MCP 1 | P13500 | CCL2 | Immune Response and Inflammation | Cardiovascular Diseases | Cancer-related genes, Candidate cardiovas | Proteins Involved in Atherosclerosis\|Prote | Cytokine-cytokine receptor interaction\|Vir | Inflammatory Response\|Allograft Rejection\|TNF-alpha Signaling via NF-kB\|Interferon Gamma Response\|Unfolded Protein Response |
| MEPE | Q9NQ76 | MEPE | Other Functions | | Biomineralization | | | |
| MERTK | Q12866 | MERTK | Enzymatic and Detoxification | Cardiovascular Diseases | Disease related genes, Enzymes, Human di | Proteins Involved in Atherosclerosis\|Proteins with Altered Expression in Cancer-Associ | | Glycolysis\|Androgen Response |
| MMP 2 | P08253 | MMP2 | Cardiovascular Function | Cardiovascular Diseases | Cancer-related genes, Candidate cardiovas | Proteins Involved in Atherosclerosis\|Prote | Fluid shear stress and atherosclerosis\|AGE | Epithelial Mesenchymal Transition\|Coagulation\|Apoptosis\|Apical Junction |
| MMP 3 | P08254 | MMP3 | Immune Response and Inflammation | Cardiovascular Diseases | Cancer-related genes, Candidate cardiovas | Proteins Involved in Atherosclerosis\|Prote | Lipid and atherosclerosis\|Rheumatoid arth | Epithelial Mesenchymal Transition\|Coagulation |
| MMP 9 | P14780 | MMP9 | Lipid Metabolism | Cardiovascular Diseases | Cancer-related genes, Candidate cardiovas | Proteins Involved in Atherosclerosis\|Prote | Lipid and atherosclerosis\|Fluid shear stres | Coagulation\|KRAS Signaling Up\|Allograft Rejection\|Apical Junction |
| MMP12 | P39900 | MMP12 | Complement and Coagulation Cascades | Cardiovascular Diseases | Cancer-related genes, Enzymes, Hydrolase, | Proteins Involved in Atherosclerosis\|Proteins Involved in Chronic Obstructive Pulmona | | Complement |
| MMP7 | P09237 | MMP7 | Complement and Coagulation Cascades | Metabolic Disorders | Cancer-related genes, Enzymes, Plasma pr | Proteins Involved in Endometriosis\|Protein | Human T-cell leukemia virus 1 infection\|W | Coagulation |
| MPO | P05164 | MPO | Energy Homeostasis | Cardiovascular Diseases | Cancer-related genes, Candidate cardiovas | Proteins Involved in Myocardial Ischemia\|I | Phagosome\|Neutrophil extracellular trap f | Reactive Oxygen Species Pathway |
| NEMO | Q9Y6K9 | IKBKG | Immune Response and Inflammation | Metabolic Disorders | Disease related genes, Human disease rela | Proteins Involved in non-Alcoholic Fatty Liv | Lipid and atherosclerosis\|Fluid shear stres | Apical Junction |
| Notch 3 | Q9UM47 | NOTCH3 | Growth and Cell Signaling | Cardiovascular Diseases | Cancer-related genes, Disease related gene | Proteins Involved in Glioma\|Proteins Invol | Pathways in cancer\|Human papillomavirus | Notch Signaling |
| NT proBNP | | | Other Functions | | | | | |
| OPG | O00300 | TNFRSF11B | Immune Response and Inflammation | Cardiovascular Diseases | Cancer-related genes, Candidate cardiovas | Proteins Involved in Endometriosis\|Protein | Cytokine-cytokine receptor interaction\|Os | Epithelial Mesenchymal Transition\|Apical Junction |
| OPN | P10451 | SPP1 | Immune Response and Inflammation | Cardiovascular Diseases | Cancer-related genes, Plasma proteins, Bic | Proteins Involved in Myocardial Ischemia\|I | PI3K-Akt signaling pathway\|Focal adhesior | Epithelial Mesenchymal Transition\|KRAS Signaling Up\|IL-2/STAT5 Signaling\|Angiogenesis |
| PAI | P05121 | SERPINE1 | Immune Response and Inflammation | Cardiovascular Diseases | Cancer-related genes, Candidate cardiovas | Proteins Involved in Atherosclerosis\|Prote | AGE-RAGE signaling pathway in diabetic cc | Inflammatory Response\|Epithelial Mesenchymal Transition\|Complement\|Coagulation\|TNF-alpha Signaling via NF-kB\|Hypoxia\|Xenobiotic Metabolism\|UV Response Dn\|TGF-beta Signaling |
| PAPPA | Q13219 | PAPPA | Cardiovascular Function | | Cancer-related genes, Candidate cardiovas | Proteins Involved in Endometriosis\|Proteins Involved in Osteoporosis | | |
| PAR 1 | P25116 | F2R | Complement and Coagulation Cascades | Cardiovascular Diseases | FDA approved drug targets, G-protein cou | Proteins Involved in Glomerulonephritis\|P | PI3K-Akt signaling pathway\|Complement a | Allograft Rejection\|Apoptosis\|p53 Pathway |
| PARP 1 | P09874 | PARP1 | Immune Response and Inflammation | Cardiovascular Diseases | Cancer-related genes, Enzymes, FDA appro | Proteins Involved in Myocardial Ischemia\|I | NF-kappa B signaling pathway\|Apoptosis\|Diabetic cardiomyopathy\|Necroptosis\|Base excision repair | |
| PCSK9 | Q8NBP7 | PCSK9 | Lipid Metabolism | | Disease related genes, Enzymes, FDA appro | Proteins Involved in Male Infertility | Cholesterol metabolism | Complement\|Apical Surface |
| PD L2 | Q9BQ51 | PDCD1LG2 | Immune Response and Inflammation | Metabolic Disorders | Cancer-related genes, CD markers, Adaptiv | Proteins Involved in Myocarditis\|Periphera | Cell adhesion molecules | KRAS Signaling Up |
| PDGF subunit A | P04085 | PDGFA | Cardiovascular Function | Cardiovascular Diseases | Cancer-related genes, Candidate cardiovas | Proteins Involved in Pulmonary Hypertensi | Fluid shear stress and atherosclerosis\|PI3K | Angiogenesis\|p53 Pathway |
| PDGF subunit B | P01127 | PDGFB | Cardiovascular Function | Cardiovascular Diseases | Cancer-related genes, Candidate cardiovas | Proteins Involved in Pulmonary Hypertensi | Fluid shear stress and atherosclerosis\|PI3K | Complement\|Coagulation\|Hypoxia |
| PECAM 1 | P16284 | PECAM1 | Cardiovascular Function | Cardiovascular Diseases | Cancer-related genes, Candidate cardiovas | Proteins Involved in Atherosclerosis\|Prote | Fluid shear stress and atherosclerosis\|Mal | Coagulation\|KRAS Signaling Up\|Apical Junction |
| PGF | P49763 | PGF | Cardiovascular Function | Cardiovascular Diseases | Cancer-related genes, Candidate cardiovas | Proteins Involved in Myocardial Ischemia\|I | PI3K-Akt signaling pathway\|MAPK signalin | Hypoxia |
| PGLYRP1 | O75594 | PGLYRP1 | Immune Response and Inflammation | | Metabolic proteins, Plasma proteins, Immunity, Innate immunity, Antibiotic, Antimicrobial | | | Angiogenesis |
| PI3 | P19957 | PI3 | Enzymatic and Detoxification | | Plasma proteins, Protease inhibitor, Serine protease inhibitor | | | |
| PIgR | P01833 | PIGR | Immune Response and Inflammation | | Cancer-related genes, Plasma proteins, Tra | Proteins Involved in Chronic Obstructive Pu | Intestinal immune network for IgA product | KRAS Signaling Up |
| PLC | P98160 | HSPG2 | Cardiovascular Function | Cardiovascular Diseases | Disease related genes, Human disease rela | Proteins Involved in Myocardial Ischemia\|I | Proteoglycans in cancer\|ECM-receptor interaction\|Hepatitis B | |
| PON3 | Q15166 | PON3 | Energy Homeostasis | | Candidate cardiovascular disease genes, Enzymes, Metabolic proteins, Plasma proteins, Hydrolase | | | |
| PRELP | P51888 | PRELP | Other Functions | | Plasma proteins, Heparin-binding | Proteins Involved in Hutchinson-Gilford Progeria Sindrome | | |
| PRSS27 | Q9BQR3 | PRSS27 | Enzymatic and Detoxification | | Enzymes, Hydrolase, Protease, Serine protease | | | |
| PRSS8 | Q16651 | PRSS8 | Enzymatic and Detoxification | | Enzymes, Hydrolase, Protease, Serine prot | Proteins Involved in Ichthyosis | | |
| PRTN3 | P24158 | PRTN3 | Cardiovascular Function | | Candidate cardiovascular disease genes, Di | Proteins Involved in Glomerulonephritis\|Proteins Involved in Thrombophilia\|Neutrophil Degranulation via FPR1/IL8\|Neutrophil Associated Lung Damage in Chronic Obstructive Pulmonary Disease\|Cathepsins Role in Periodontitis\|Salivary Proteins Role in Periodontitis | | |
| PSGL 1 | Q14242 | SELPLG | Cardiovascular Function | Cardiovascular Diseases | Candidate cardiovascular disease genes, Cl | Proteins Involved in Atherosclerosis\|Leuko | Cell adhesion molecules\|Neutrophil extracellular trap formation\|Staphylococcus aureus infection | |

**Table S8: Protein functional and pathway annotations**

| Variable_Name | Uniprot | Gene | Extracted_Pathway_Annotation | Extracted_Disease_Annotation | Uniport_Annotation | Elsevier_Annotation | KEGG_2021_Human_Annotation | MSigDB_Hallmark_2020_Annotation |
|---|---|---|---|---|---|---|---|---|
| PSP D | P35247 | SFTPD | Immune Response and Inflammation | | Gaseous exchange, Immunity, Innate immu | Proteins Involved in Chronic Obstructive Pu | Phagosome | |
| PTX3 | P26022 | PTX3 | Growth and Cell Signaling | Cardiovascular Diseases | Plasma proteins | Proteins Involved in Myocardial Ischemia\|Proteins Involved in Chronic Obstructive Pul | | Epithelial Mesenchymal Transition\|TNF-alpha Signaling via NF-kB |
| RAGE | Q15109 | AGER | Immune Response and Inflammation | Cardiovascular Diseases | Cancer-related genes, Plasma proteins, Tra | Proteins Involved in Atherosclerosis\|Protei | Lipid and atherosclerosis\|AGE-RAGE signal | IL-2/STAT5 Signaling |
| RARRES2 | Q99969 | RARRES2 | Immune Response and Inflammation | Metabolic Disorders | Plasma proteins, Chemotaxis, Differentiati | Proteins Involved in Endometriosis\|Proteins Involved in non-Alcoholic Fatty Liver Disease\|Adipokines Production by Adipocyte\|Adipokines Production by Adipocyte Impaired in Obesity | | |
| REN | P00797 | REN | Cardiovascular Function | Cardiovascular Diseases | Candidate cardiovascular disease genes, Di | Proteins Involved in Atherosclerosis\|Protei | Renin-angiotensin system\|Diabetic cardiomyopathy\|Renin secretion | |
| RETN | Q9HD89 | RETN | Lipid Metabolism | Cardiovascular Diseases | Candidate cardiovascular disease genes, Pl | Proteins Involved in Atherosclerosis\|Proteins Involved in non-Alcoholic Fatty Liver Dise | | KRAS Signaling Up\|Adipogenesis |
| SCF | P21583 | KITLG | Growth and Cell Signaling | Cancer-Related Diseases | Cancer-related genes, Disease related gene | Proteins Involved in Endometriosis\|Protei | PI3K-Akt signaling pathway\|Hematopoietic cell lineage\|MAPK signaling pathway\|Pathways in cancer\|Rap1 signaling pathway\|Ras signaling pathway\|Phospholipase D signaling pathway\|Melanogenesis | |
| SCGB3A2 | Q96PL1 | SCGB3A2 | Other Functions | | Human disease related genes | Proteins Involved in Graves Disease | | |
| SELE | P16581 | SELE | Immune Response and Inflammation | Cardiovascular Diseases | Cancer-related genes, Candidate cardiovas | Proteins Involved in Atherosclerosis\|Protei | Lipid and atherosclerosis\|Fluid shear stres | Inflammatory Response |
| SELP | P16109 | SELP | Lipid Metabolism | Cardiovascular Diseases | Cancer-related genes, Candidate cardiovas | Proteins Involved in Atherosclerosis\|Protei | Lipid and atherosclerosis\|Malaria\|Cell adh | IL-2/STAT5 Signaling\|Interferon Gamma Response |
| SERPINA12 | Q8IW75 | SERPINA12 | Enzymatic and Detoxification | Metabolic Disorders | Protease inhibitor, Serine protease inhibitc | Adipokines Production by Adipocyte\|Adipokines Production by Adipocyte Impaired in Obesity | | |
| SHPS 1 | P78324 | SIRPA | Other Functions | | CD markers, Plasma proteins | Natural Killer Cell Precursor -> Natural Kille | Osteoclast differentiation | Apical Junction |
| SLAMF7 | Q9NQ25 | SLAMF7 | Immune Response and Inflammation | | CD markers, FDA approved drug targets, A | Natural Killer Cell Activation through ITSM-Containing Receptors | | Interferon Gamma Response |
| SOD2 | P04179 | SOD2 | Lipid Metabolism | Cardiovascular Diseases | Cancer-related genes, Disease related gene | Proteins Involved in Myocardial Ischemia\|I | Lipid and atherosclerosis\|FoxO signaling p | Apoptosis\|TNF-alpha Signaling via NF-kB\|Interferon Gamma Response\|UV Response Up\|Reactive Oxygen Species Pathway\|Pperoxisome |
| SORT1 | Q99523 | SORT1 | Lipid Metabolism | | Disease related genes, Potential drug targe | Proteins Involved in Amyotrophic Lateral S | Cholesterol metabolism\|Lysosome\|Neurotrophin signaling pathway | |
| SPON1 | Q9HCB6 | SPON1 | Growth and Cell Signaling | | Plasma proteins, Cell adhesion | Proteins Involved in Neuroblastoma\|Amyloid beta and APP Intracellular Transport in A | | KRAS Signaling Up |
| SPON2 | Q9BUD6 | SPON2 | Immune Response and Inflammation | | Plasma proteins, Cell adhesion, Immunity, | MYOC in Cell Adhesion in Glaucoma | | |
| SRC | P12931 | SRC | Immune Response and Inflammation | Cardiovascular Diseases | Cancer-related genes, Disease related gene | Proteins Involved in Glomerulonephritis\|Pr | Lipid and atherosclerosis\|Fluid shear stres | Complement\|Apical Junction |
| ST2 | Q01638 | IL1RL1 | Immune Response and Inflammation | | Candidate cardiovascular disease genes, Er | Proteins Involved in Osteoporosis\|Myelobl | Cytokine-cytokine receptor interaction | IL-2/STAT5 Signaling |
| STK4 | Q13043 | STK4 | Growth and Cell Signaling | Cardiovascular Diseases | Disease related genes, Enzymes, Potential | Proteins Involved in Myocardial Ischemia\|I | MAPK signaling pathway\|Pathways in cancer\|Ras signaling pathway\|FoxO signaling pathway\|Non-small cell lung cancer | |
| t PA | P00750 | PLAT | Cardiovascular Function | Cardiovascular Diseases | Cancer-related genes, Candidate cardiovas | Proteins Involved in Atherosclerosis\|Protei | Fluid shear stress and atherosclerosis\|Com | Complement\|Coagulation\|KRAS Signaling Up\|Apoptosis |
| TF | P13726 | F3 | Immune Response and Inflammation | Cardiovascular Diseases | Cancer-related genes, Candidate cardiovas | Proteins Involved in Atherosclerosis\|Protei | AGE-RAGE signaling pathway in diabetic cc | Inflammatory Response\|Complement\|Coagulation\|TNF-alpha Signaling via NF-kB\|Hypoxia\|UV Response Dn |
| TFF3 | Q07654 | TFF3 | Other Functions | | Cancer-related genes, Plasma proteins | Proteins Involved in Ulcerative Colitis\|Proteins Involved in Hearing Loss\|Proteins Invol | | Glycolysis\|Estrogen Response Early\|Estrogen Response Late |
| TFPI | P10646 | TFPI | Cardiovascular Function | Cardiovascular Diseases | Candidate cardiovascular disease genes, FI | Proteins Involved in Myocardial Ischemia\|I | Complement and coagulation cascades | KRAS Signaling Up\|UV Response Dn |
| TGM2 | P21980 | TGM2 | Energy Homeostasis | Cardiovascular Diseases | Disease related genes, Enzymes, Metabolic | Proteins Involved in Atherosclerosis\|Protei | Huntington disease | Epithelial Mesenchymal Transition\|IL-2/STAT5 Signaling\|Hypoxia\|Estrogen Response Early |
| THBS2 | P35442 | THBS2 | Growth and Cell Signaling | Cardiovascular Diseases | Cancer-related genes, Disease related gene | Proteins Involved in Glomerulonephritis\|Pr | PI3K-Akt signaling pathway\|Malaria\|Phag | Epithelial Mesenchymal Transition |
| THPO | P40225 | THPO | Immune Response and Inflammation | Inflammatory and Immune-Related Diseases | Cancer-related genes, Disease related gene | Megakaryocyte -> Platelet Precursor Cell S | Cytokine-cytokine receptor interaction\|Hematopoietic cell lineage\|JAK-STAT signaling pathway | |
| TIE2 | Q02763 | TEK | Cardiovascular Function | Cardiovascular Diseases | Cancer-related genes, CD markers, Disease | Proteins Involved in Pulmonary Hypertensi | Rheumatoid arthritis\|PI3K-Akt signaling pathway\|MAPK signaling pathway\|Rap1 signaling pathway\|HIF-1 signaling pathway\|Ras signaling pathway | |
| TIMP4 | Q99727 | TIMP4 | Enzymatic and Detoxification | | Plasma proteins, Metalloenzyme inhibitor, | Proteins Involved in Periodontitis\|Extracellular Matrix Turnover\|ADAM33 Role in Asthma | | |
| TLT 2 | Q5T2D2 | TREML2 | Other Functions | | Plasma proteins, Receptor | | | |
| TM | P07204 | THBD | Cardiovascular Function | Cardiovascular Diseases | Candidate cardiovascular disease genes, CI | Proteins Involved in Pulmonary Hypertensi | Fluid shear stress and atherosclerosis\|AGE | Coagulation\|Angiogenesis |
| TNF R1 | P19438 | TNFRSF1A | Immune Response and Inflammation | Cardiovascular Diseases | Cancer-related genes, Candidate cardiovas | Proteins Involved in Atherosclerosis\|Protei | Cytokine-cytokine receptor interaction\|Vir | IL-6/JAK/STAT3 Signaling\|Xenobiotic Metabolism\|PI3K/AKT/mTOR Signaling |
| TNF R2 | P20333 | TNFRSF1B | Immune Response and Inflammation | Cardiovascular Diseases | Cancer-related genes, Candidate cardiovas | Proteins Involved in Atherosclerosis\|Protei | Cytokine-cytokine receptor interaction\|Vir | Inflammatory Response\|IL-6/JAK/STAT3 Signaling\|KRAS Signaling Up\|IL-2/STAT5 Signaling |
| TNFRSF10A | O00220 | TNFRSF10A | Immune Response and Inflammation | Inflammatory and Immune-Related Diseases | Cancer-related genes, CD markers, Apopto | TNFR -> NF-kB Signaling\|Proteins with Alte | Cytokine-cytokine receptor interaction\|Viral protein interaction with cytokine and cytokine receptor\|Lipid and atherosclerosis\|Apoptosis\|Pathogenic Escherichia coli infection\|Influenza A\|p53 signaling pathway\|Natural killer cell mediated cytotoxicity\|Salmonella infection\|Necroptosis | |
| TNFRSF10C | O14798 | TNFRSF10C | Immune Response and Inflammation | Inflammatory and Immune-Related Diseases | Cancer-related genes, CD markers, Apopto | anti-Inflammatory Function of Macrophag | Cytokine-cytokine receptor interaction\|Viral protein interaction with cytokine and cytokine receptor | |
| TNFRSF11A | Q9Y6Q6 | TNFRSF11A | Immune Response and Inflammation | Inflammatory and Immune-Related Diseases | CD markers, Disease related genes, Human | Proteins Involved in Osteoporosis\|Multiple | Cytokine-cytokine receptor interaction\|Rheumatoid arthritis\|NF-kappa B signaling pathway\|Osteoclast differentiation\|Prolactin signaling pathway | |
| TNFRSF13B | O14836 | TNFRSF13B | Immune Response and Inflammation | Cancer-Related Diseases | CD markers, Disease related genes, Human | Proteins Involved in Glomerulonephritis\|Pr | Cytokine-cytokine receptor interaction\|Intestinal immune network for IgA production\|Primary immunodeficiency | |
| TNFRSF14 | Q92956 | TNFRSF14 | Immune Response and Inflammation | Cancer-Related Diseases | Cancer-related genes, CD markers, Human | Proteins with Altered Expression in Cancer- | Cytokine-cytokine receptor interaction\|Viral protein interaction with cytokine and cytokine receptor\|Herpes simplex virus 1 infection | |
| TNFSF13B | Q9Y275 | TNFSF13B | Immune Response and Inflammation | Inflammatory and Immune-Related Diseases | Cancer-related genes, CD markers, FDA ap | Proteins Involved in Chronic Obstructive Pu | Cytokine-cytokine receptor interaction\|Rheumatoid arthritis\|NF-kappa B signaling pathway\|Intestinal immune network for IgA production | |
| TR | P02786 | TFRC | Energy Homeostasis | Inflammatory and Immune-Related Diseases | Cancer-related genes, CD markers, Disease | Proteins Involved in Glioma\|Megakaryocyt | Hematopoietic cell lineage\|HIF-1 signaling | UV Response Up\|mTORC1 Signaling\|heme Metabolism\|E2F Targets |
| TR AP | P13686 | ACP5 | Energy Homeostasis | | Disease related genes, Enzymes, Human di | Osteoclast Activation in Psoriatic Arthritis\| | Rheumatoid arthritis\|Osteoclast differenti | heme Metabolism |
| TRAIL R2 | O14763 | TNFRSF10B | Immune Response and Inflammation | Inflammatory and Immune-Related Diseases | Cancer-related genes, CD markers, Disease | TNFR -> NF-kB Signaling\|TNFR -> AP-1/ATF | Cytokine-cytokine receptor interaction\|Viral protein interaction with cytokine and cytokine receptor\|Lipid and atherosclerosis\|Apoptosis\|Pathogenic Escherichia coli infection\|Influenza A\|p53 signaling pathway\|Natural killer cell mediated cytotoxicity\|Salmonella infection\|Necroptosis | |
| U PAR | Q03405 | PLAUR | Immune Response and Inflammation | Cardiovascular Diseases | Cancer-related genes, CD markers, FDA ap | Proteins Involved in Atherosclerosis\|Protei | Complement and coagulation cascades\|Pr | Inflammatory Response\|Epithelial Mesenchymal Transition\|Complement\|KRAS Signaling Up\|TNF-alpha Signaling via NF-kB\|Hypoxia\|Cholesterol Homeostasis\|Apical Surface |
| uPA | P00749 | PLAU | Complement and Coagulation Cascades | Cardiovascular Diseases | Cancer-related genes, Disease related gene | Proteins Involved in Atherosclerosis\|Protei | Complement and coagulation cascades\|NF | Coagulation\|KRAS Signaling Up\|TNF-alpha Signaling via NF-kB |
| VEGFD | O43915 | VEGFD | Cardiovascular Function | Cancer-Related Diseases | Cancer-related genes, Plasma proteins, RA | Proteins Involved in Systemic Scleroderma | PI3K-Akt signaling pathway\|TNF signaling pathway\|MAPK signaling pathway\|AGE-RAGE signaling pathway in diabetic complications\|Pathways in cancer\|Rap1 signaling pathway\|Ras signaling pathway\|Focal adhesion\|Relaxin signaling pathway\|Calcium signaling pathway | |
| VSIG2 | Q96IQ7 | VSIG2 | Lipid Metabolism | | Lipid metabolism | | | |
| vWF | P04275 | VWF | Cardiovascular Function | Cardiovascular Diseases | Cancer-related genes, Candidate cardiovas | Proteins Involved in Atherosclerosis\|Protei | PI3K-Akt signaling pathway\|Complement a | Coagulation\|Apical Junction |
| XCL1 | P47992 | XCL1 | Immune Response and Inflammation | | Chemotaxis, Cytokine | | Cytokine-cytokine receptor interaction\|Vir | Interferon Gamma Response |

**Table S9: Associations between pPSs and global metabolomic profile in SCAPIS**

**a) Spearman Correlation**

| Metabolomics | PPS | Samp | r | CI95% | p | fdr_p_adjusted |
|---|---|---|---|---|---|---|
| AcAce | Cholesterol_pPS | 4255 | -0.013781894 | [-0.04 0.02] | 0.369448212 | 0.643918869 |
| AcAce | Lipodystrophy_1_pPS | 4255 | -0.0133617 | [-0.04 0.02] | 0.384223354 | 0.654405057 |
| AcAce | Lipodystrophy_2_pPS | 4255 | -0.028429502 | [-0.06 0. ] | 0.064072009 | 0.210789049 |
| AcAce | Liver-Lipid_pPS | 4255 | -0.00905682 | [-0.04 0.02] | 0.555335291 | 0.76163312 |
| AcAce | ASAT_Pos_pPS | 4255 | -0.021060349 | [-0.05 0.01] | 0.170193122 | 0.43221059 |
| AcAce | ALP_Neg_pPS | 4255 | -0.029361127 | [-0.06 0. ] | 0.05582708 | 0.189479812 |
| AcAce | SHBG-LpA_pPS | 4255 | 0.016206562 | [-0.01 0.05] | 0.291230346 | 0.577755652 |
| AcAce | Obesity_pPS | 4255 | 0.020542654 | [-0.01 0.05] | 0.18094183 | 0.45159215 |
| AcAce | VAT_Neg_pPS | 4255 | -0.039876649 | [-0.07 -0.01] | 0.009383253 | 0.045127976 |
| AcAce | Bilirubin_pPS | 4255 | 0.028205041 | [-0. 0.06] | 0.066202075 | 0.215777718 |
| AcAce | Proinsulin_pPS | 4255 | -0.001973511 | [-0.03 0.03] | 0.897742667 | 0.949937874 |
| AcAce | Hyper_Insulin_pPS | 4255 | -0.020648477 | [-0.05 0.01] | 0.178704676 | 0.447805525 |
| AcAce | Beta_Cell_1_pPS | 4255 | 0.00783664 | [-0.02 0.04] | 0.609825207 | 0.794699313 |
| AcAce | Beta_Cell_2_pPS | 4255 | -0.02789202 | [-0.06 0. ] | 0.069269614 | 0.224107575 |
| AcAce | Total_GRS | 4255 | -0.019366789 | [-0.05 0.01] | 0.207212822 | 0.486292673 |
| Ace | Cholesterol_pPS | 4254 | -0.02014185 | [-0.05 0.01] | 0.189655817 | 0.462286054 |
| Ace | Lipodystrophy_1_pPS | 4254 | 0.005289954 | [-0.02 0.04] | 0.730516187 | 0.871985098 |
| Ace | Lipodystrophy_2_pPS | 4254 | -0.026676053 | [-0.06 0. ] | 0.082347383 | 0.25748562 |
| Ace | Liver-Lipid_pPS | 4254 | 0.02959559 | [-0. 0.06] | 0.0539256 | 0.184482317 |
| Ace | ASAT_Pos_pPS | 4254 | -0.008736413 | [-0.04 0.02] | 0.56945705 | 0.770653232 |
| Ace | ALP_Neg_pPS | 4254 | -0.045766083 | [-0.08 -0.02] | 0.002868795 | 0.018368853 |
| Ace | SHBG-LpA_pPS | 4254 | 0.008905967 | [-0.02 0.04] | 0.561987535 | 0.767334778 |
| Ace | Obesity_pPS | 4254 | 0.008313312 | [-0.02 0.04] | 0.588300047 | 0.780398021 |
| Ace | VAT_Neg_pPS | 4254 | -0.022504411 | [-0.05 0.01] | 0.142789944 | 0.390668915 |
| Ace | Bilirubin_pPS | 4254 | 0.01308537 | [-0.02 0.04] | 0.394189791 | 0.661609625 |
| Ace | Proinsulin_pPS | 4254 | -0.017516425 | [-0.05 0.01] | 0.254033427 | 0.536320571 |
| Ace | Hyper_Insulin_pPS | 4254 | 0.006152882 | [-0.02 0.04] | 0.688694882 | 0.844625798 |
| Ace | Beta_Cell_1_pPS | 4254 | 0.011540501 | [-0.02 0.04] | 0.452385655 | 0.695176638 |
| Ace | Beta_Cell_2_pPS | 4254 | 0.017863859 | [-0.01 0.05] | 0.244734388 | 0.525480742 |
| Ace | Total_GRS | 4254 | -0.006964427 | [-0.04 0.02] | 0.650210791 | 0.818487176 |
| Ala | Cholesterol_pPS | 4255 | 0.003933203 | [-0.03 0.03] | 0.797852224 | 0.905020106 |
| Ala | Lipodystrophy_1_pPS | 4255 | 0.043489304 | [0.01 0.07] | 0.004606642 | 0.025363691 |
| Ala | Lipodystrophy_2_pPS | 4255 | -0.001403719 | [-0.03 0.03] | 0.927167609 | 0.964488129 |
| Ala | Liver-Lipid_pPS | 4255 | -0.069899712 | [-0.1 -0.04] | 5.18E-06 | 0.000102871 |
| Ala | ASAT_Pos_pPS | 4255 | 0.007641238 | [-0.02 0.04] | 0.618767423 | 0.800033829 |
| Ala | ALP_Neg_pPS | 4255 | 0.02983923 | [-0. 0.06] | 0.051951286 | 0.179445264 |
| Ala | SHBG-LpA_pPS | 4255 | 0.015063129 | [-0.02 0.05] | 0.326615727 | 0.609784284 |
| Ala | Obesity_pPS | 4255 | 0.01747083 | [-0.01 0.05] | 0.255215729 | 0.537758092 |
| Ala | VAT_Neg_pPS | 4255 | 0.023022918 | [-0.01 0.05] | 0.13376188 | 0.373106933 |
| Ala | Bilirubin_pPS | 4255 | -0.022238481 | [-0.05 0.01] | 0.147524928 | 0.397040113 |
| Ala | Proinsulin_pPS | 4255 | 0.003798888 | [-0.03 0.03] | 0.804613268 | 0.909247074 |
| Ala | Hyper_Insulin_pPS | 4255 | 0.009838755 | [-0.02 0.04] | 0.521713312 | 0.738176158 |
| Ala | Beta_Cell_1_pPS | 4255 | -0.00302257 | [-0.03 0.03] | 0.843962467 | 0.92810697 |
| Ala | Beta_Cell_2_pPS | 4255 | 0.023110144 | [-0.01 0.05] | 0.132294951 | 0.370838552 |
| Ala | Total_GRS | 4255 | 0.031737726 | [0. 0.06] | 0.038710265 | 0.144657696 |
| Alb | Cholesterol_pPS | 4254 | 0.01036772 | [-0.02 0.04] | 0.499627912 | 0.726085278 |
| Alb | Lipodystrophy_1_pPS | 4254 | 0.008688861 | [-0.02 0.04] | 0.57156036 | 0.771552531 |
| Alb | Lipodystrophy_2_pPS | 4254 | -0.003620454 | [-0.03 0.03] | 0.81363925 | 0.913746697 |
| Alb | Liver-Lipid_pPS | 4254 | -0.047844363 | [-0.08 -0.02] | 0.001827012 | 0.013239667 |
| Alb | ASAT_Pos_pPS | 4254 | -0.002121739 | [-0.03 0.03] | 0.890122152 | 0.948487102 |
| Alb | ALP_Neg_pPS | 4254 | -0.015109028 | [-0.05 0.01] | 0.325200689 | 0.608687153 |
| Alb | SHBG-LpA_pPS | 4254 | -0.019611775 | [-0.05 0.01] | 0.201576204 | 0.477769015 |
| Alb | Obesity_pPS | 4254 | -0.007397832 | [-0.04 0.02] | 0.630026116 | 0.803929814 |
| Alb | VAT_Neg_pPS | 4254 | -0.003405232 | [-0.03 0.03] | 0.824531695 | 0.921344342 |
| Alb | Bilirubin_pPS | 4254 | 0.009120891 | [-0.02 0.04] | 0.552587869 | 0.760607725 |
| Alb | Proinsulin_pPS | 4254 | 0.013164556 | [-0.02 0.04] | 0.391334168 | 0.659396536 |
| Alb | Hyper_Insulin_pPS | 4254 | 0.000220873 | [-0.03 0.03] | 0.988525732 | 0.992226344 |
| Alb | Beta_Cell_1_pPS | 4254 | 0.020446225 | [-0.01 0.05] | 0.183050069 | 0.453397688 |
| Alb | Beta_Cell_2_pPS | 4254 | 0.015041446 | [-0.02 0.05] | 0.327369706 | 0.61008516 |
| Alb | Total_GRS | 4254 | 0.010286227 | [-0.02 0.04] | 0.503005186 | 0.727744052 |
| bOHBut | Cholesterol_pPS | 4249 | -0.009002079 | [-0.04 0.02] | 0.558006899 | 0.763832034 |
| bOHBut | Lipodystrophy_1_pPS | 4249 | -0.042293582 | [-0.07 -0.01] | 0.005897911 | 0.031083583 |
| bOHBut | Lipodystrophy_2_pPS | 4249 | -0.018974987 | [-0.05 0.01] | 0.216878451 | 0.497544682 |
| bOHBut | Liver-Lipid_pPS | 4249 | 0.015429846 | [-0.01 0.05] | 0.315317555 | 0.60298573 |
| bOHBut | ASAT_Pos_pPS | 4249 | -0.020198371 | [-0.05 0.01] | 0.188676859 | 0.460423052 |
| bOHBut | ALP_Neg_pPS | 4249 | -0.010589668 | [-0.04 0.02] | 0.490747022 | 0.720501275 |
| bOHBut | SHBG-LpA_pPS | 4249 | -0.007308911 | [-0.04 0.02] | 0.634345551 | 0.807999529 |
| bOHBut | Obesity_pPS | 4249 | 0.005105735 | [-0.03 0.04] | 0.739702102 | 0.874675307 |
| bOHBut | VAT_Neg_pPS | 4249 | -0.023462713 | [-0.05 0.01] | 0.126760966 | 0.359462978 |
| bOHBut | Bilirubin_pPS | 4249 | 0.026829965 | [-0. 0.06] | 0.080772455 | 0.254539888 |
| bOHBut | Proinsulin_pPS | 4249 | 0.021202746 | [-0.01 0.05] | 0.167622922 | 0.429571287 |
| bOHBut | Hyper_Insulin_pPS | 4249 | -0.010604082 | [-0.04 0.02] | 0.490156933 | 0.720501275 |
| bOHBut | Beta_Cell_1_pPS | 4249 | 0.03228206 | [0. 0.06] | 0.035619966 | 0.134990859 |
| bOHBut | Beta_Cell_2_pPS | 4249 | -0.008442699 | [-0.04 0.02] | 0.58272969 | 0.778303354 |
| bOHBut | Total_GRS | 4249 | -0.010596913 | [-0.04 0.02] | 0.490450391 | 0.720501275 |
| Cit | Cholesterol_pPS | 4255 | -0.008821623 | [-0.04 0.02] | 0.565651467 | 0.768674587 |
| Cit | Lipodystrophy_1_pPS | 4255 | -0.011204173 | [-0.04 0.02] | 0.465617112 | 0.706538163 |
| Cit | Lipodystrophy_2_pPS | 4255 | -0.029372194 | [-0.06 0. ] | 0.05573471 | 0.189479812 |
| Cit | Liver-Lipid_pPS | 4255 | -0.019119072 | [-0.05 0.01] | 0.213083606 | 0.493880952 |
| Cit | ASAT_Pos_pPS | 4255 | 0.002819697 | [-0.03 0.03] | 0.85431428 | 0.932572076 |
| Cit | ALP_Neg_pPS | 4255 | 0.0112345 | [-0.02 0.04] | 0.464410388 | 0.705495951 |
| Cit | SHBG-LpA_pPS | 4255 | 0.00429337 | [-0.03 0.03] | 0.779798397 | 0.894802239 |
| Cit | Obesity_pPS | 4255 | -0.013954209 | [-0.04 0.02] | 0.363492616 | 0.639224774 |
| Cit | VAT_Neg_pPS | 4255 | -0.014545222 | [-0.04 0.02] | 0.343525565 | 0.62658362 |
| Cit | Bilirubin_pPS | 4255 | -0.008875852 | [-0.04 0.02] | 0.563264755 | 0.767793567 |
| Cit | Proinsulin_pPS | 4255 | 0.012717464 | [-0.02 0.04] | 0.407566546 | 0.671010017 |
| Cit | Hyper_Insulin_pPS | 4255 | 0.00909839 | [-0.02 0.04] | 0.553521556 | 0.760607725 |
| Cit | Beta_Cell_1_pPS | 4255 | -0.002336649 | [-0.03 0.03] | 0.879060148 | 0.944681371 |
| Cit | Beta_Cell_2_pPS | 4255 | 0.018285441 | [-0.01 0.05] | 0.233719778 | 0.516301672 |
| Cit | Total_GRS | 4255 | -0.012831273 | [-0.04 0.02] | 0.40338242 | 0.668142396 |
| Crea | Cholesterol_pPS | 4249 | 0.015153383 | [-0.01 0.05] | 0.324067735 | 0.608687153 |
| Crea | Lipodystrophy_1_pPS | 4249 | 0.013728578 | [-0.02 0.04] | 0.371641753 | 0.643918869 |
| Crea | Lipodystrophy_2_pPS | 4249 | -0.002045055 | [-0.03 0.03] | 0.89413164 | 0.948580863 |
| Crea | Liver-Lipid_pPS | 4249 | 0.051871201 | [0.02 0.08] | 0.000730973 | 0.006846889 |

**b) Partial Correlation**

| Metabolomics | PPS | Samp | r | CI95% | p | fdr_p_adjusted |
|---|---|---|---|---|---|---|
| AcAce | Cholesterol_pPS | 4255 | -0.015454018 | [-0.05 0.01] | 0.314960254 | 0.616791721 |
| AcAce | Lipodystrophy_1_pPS | 4255 | -0.008776892 | [-0.04 0.02] | 0.568218099 | 0.797755947 |
| AcAce | Lipodystrophy_2_pPS | 4255 | -0.018819631 | [-0.05 0.01] | 0.221047972 | 0.514579116 |
| AcAce | Liver-Lipid_pPS | 4255 | -0.005073479 | [-0.04 0.03] | 0.741493796 | 0.873045882 |
| AcAce | ASAT_Pos_pPS | 4255 | -0.018599629 | [-0.05 0.01] | 0.226494556 | 0.521665942 |
| AcAce | ALP_Neg_pPS | 4255 | -0.027748069 | [-0.06 0. ] | 0.0711521 | 0.245174706 |
| AcAce | SHBG-LpA_pPS | 4255 | 0.019644463 | [-0.01 0.05] | 0.201464221 | 0.486527589 |
| AcAce | Obesity_pPS | 4255 | 0.024375435 | [-0.01 0.05] | 0.112941921 | 0.350015057 |
| AcAce | VAT_Neg_pPS | 4255 | -0.036868381 | [-0.07 -0.01] | 0.016486266 | 0.07895582 |
| AcAce | Bilirubin_pPS | 4255 | 0.029183137 | [-0. 0.06] | 0.057713308 | 0.210843145 |
| AcAce | Proinsulin_pPS | 4255 | 0.000233607 | [-0.03 0.03] | 0.987881461 | 0.993363084 |
| AcAce | Hyper_Insulin_pPS | 4255 | -0.013227947 | [-0.04 0.02] | 0.389729229 | 0.668708134 |
| AcAce | Beta_Cell_1_pPS | 4255 | 0.017727752 | [-0.01 0.05] | 0.249019034 | 0.545779139 |
| AcAce | Beta_Cell_2_pPS | 4255 | -0.029125449 | [-0.06 0. ] | 0.058209611 | 0.211498441 |
| Ace | Cholesterol_pPS | 4254 | -0.021559607 | [-0.05 0.01] | 0.160979126 | 0.435513797 |
| Ace | Lipodystrophy_1_pPS | 4254 | 0.006964478 | [-0.02 0.04] | 0.650709378 | 0.844400309 |
| Ace | Lipodystrophy_2_pPS | 4254 | -0.023260097 | [-0.05 0.01] | 0.130437413 | 0.3879647 |
| Ace | Liver-Lipid_pPS | 4254 | 0.032335661 | [0. 0.06] | 0.035488425 | 0.139309465 |
| Ace | ASAT_Pos_pPS | 4254 | -0.009550722 | [-0.04 0.02] | 0.534651399 | 0.781586915 |
| Ace | ALP_Neg_pPS | 4254 | -0.043794155 | [-0.07 -0.01] | 0.004392697 | 0.028368318 |
| Ace | SHBG-LpA_pPS | 4254 | 0.009029495 | [-0.02 0.04] | 0.557181333 | 0.796929123 |
| Ace | Obesity_pPS | 4254 | 0.011783733 | [-0.02 0.04] | 0.443613727 | 0.706833249 |
| Ace | VAT_Neg_pPS | 4254 | -0.02423251 | [-0.05 0.01] | 0.115112456 | 0.354001747 |
| Ace | Bilirubin_pPS | 4254 | 0.013657363 | [-0.02 0.04] | 0.374579723 | 0.655035906 |
| Ace | Proinsulin_pPS | 4254 | -0.017459106 | [-0.05 0.01] | 0.256320919 | 0.55296819 |
| Ace | Hyper_Insulin_pPS | 4254 | 0.008667945 | [-0.02 0.04] | 0.573076089 | 0.800878787 |
| Ace | Beta_Cell_1_pPS | 4254 | 0.011676109 | [-0.02 0.04] | 0.447788141 | 0.709019056 |
| Ace | Beta_Cell_2_pPS | 4254 | 0.018261077 | [-0.01 0.05] | 0.235117308 | 0.530670632 |
| Ala | Cholesterol_pPS | 4255 | 0.004738791 | [-0.03 0.03] | 0.757995412 | 0.881758753 |
| Ala | Lipodystrophy_1_pPS | 4255 | 0.040777643 | [0.01 0.07] | 0.007991374 | 0.045450937 |
| Ala | Lipodystrophy_2_pPS | 4255 | -0.00558583 | [-0.04 0.02] | 0.71646354 | 0.866099998 |
| Ala | Liver-Lipid_pPS | 4255 | -0.073063941 | [-0.1 -0.04] | 1.97E-06 | 4.23E-05 |
| Ala | ASAT_Pos_pPS | 4255 | 0.001851865 | [-0.03 0.03] | 0.904160789 | 0.96224905 |
| Ala | ALP_Neg_pPS | 4255 | 0.032007633 | [0. 0.06] | 0.037374916 | 0.145791207 |
| Ala | SHBG-LpA_pPS | 4255 | 0.011376147 | [-0.02 0.04] | 0.459486775 | 0.72261785 |
| Ala | Obesity_pPS | 4255 | 0.015380035 | [-0.01 0.05] | 0.31728286 | 0.618500765 |
| Ala | VAT_Neg_pPS | 4255 | 0.02157517 | [-0.01 0.05] | 0.160627527 | 0.43515062 |
| Ala | Bilirubin_pPS | 4255 | -0.024537387 | [-0.05 0.01] | 0.110568701 | 0.344222613 |
| Ala | Proinsulin_pPS | 4255 | -0.004000646 | [-0.03 0.03] | 0.794771244 | 0.901491235 |
| Ala | Hyper_Insulin_pPS | 4255 | 0.003501403 | [-0.03 0.03] | 0.819911203 | 0.915483674 |
| Ala | Beta_Cell_1_pPS | 4255 | -0.009090272 | [-0.04 0.02] | 0.554483775 | 0.795753776 |
| Ala | Beta_Cell_2_pPS | 4255 | 0.02541056 | [-0. 0.06] | 0.098444521 | 0.321510491 |
| Alb | Cholesterol_pPS | 4254 | 0.012010996 | [-0.02 0.04] | 0.434872206 | 0.699850608 |
| Alb | Lipodystrophy_1_pPS | 4254 | 0.011115756 | [-0.02 0.04] | 0.469879744 | 0.732061671 |
| Alb | Lipodystrophy_2_pPS | 4254 | -0.002192231 | [-0.03 0.03] | 0.886668889 | 0.952011446 |
| Alb | Liver-Lipid_pPS | 4254 | -0.048216839 | [-0.08 -0.02] | 0.00170992 | 0.013424546 |
| Alb | ASAT_Pos_pPS | 4254 | -0.003658713 | [-0.03 0.03] | 0.811990181 | 0.911348446 |
| Alb | ALP_Neg_pPS | 4254 | -0.015816411 | [-0.05 0.01] | 0.303802365 | 0.602663767 |
| Alb | SHBG-LpA_pPS | 4254 | -0.020884059 | [-0.05 0.01] | 0.174510373 | 0.453447086 |
| Alb | Obesity_pPS | 4254 | -0.010137219 | [-0.04 0.02] | 0.509861642 | 0.766145193 |
| Alb | VAT_Neg_pPS | 4254 | -0.004348158 | [-0.03 0.03] | 0.777420796 | 0.891327566 |
| Alb | Bilirubin_pPS | 4254 | 0.006962457 | [-0.02 0.04] | 0.650804035 | 0.844400309 |
| Alb | Proinsulin_pPS | 4254 | 0.011583868 | [-0.02 0.04] | 0.45138362 | 0.713235997 |
| Alb | Hyper_Insulin_pPS | 4254 | 0.002452379 | [-0.03 0.03] | 0.873327606 | 0.946048963 |
| Alb | Beta_Cell_1_pPS | 4254 | 0.014750448 | [-0.02 0.04] | 0.337557995 | 0.631792226 |
| Alb | Beta_Cell_2_pPS | 4254 | 0.010469593 | [-0.02 0.04] | 0.496094593 | 0.753425828 |
| bOHBut | Cholesterol_pPS | 4249 | -0.009779486 | [-0.04 0.02] | 0.525155559 | 0.775912494 |
| bOHBut | Lipodystrophy_1_pPS | 4249 | -0.038655073 | [-0.07 -0.01] | 0.011988666 | 0.062018887 |
| bOHBut | Lipodystrophy_2_pPS | 4249 | -0.011279767 | [-0.04 0.02] | 0.463616718 | 0.726829028 |
| bOHBut | Liver-Lipid_pPS | 4249 | 0.016995316 | [-0.01 0.05] | 0.269455367 | 0.569377194 |
| bOHBut | ASAT_Pos_pPS | 4249 | -0.014240289 | [-0.04 0.02] | 0.354819137 | 0.640530128 |
| bOHBut | ALP_Neg_pPS | 4249 | -0.01290632 | [-0.04 0.02] | 0.401694828 | 0.677078992 |
| bOHBut | SHBG-LpA_pPS | 4249 | -0.003325699 | [-0.03 0.03] | 0.828924257 | 0.920924729 |
| bOHBut | Obesity_pPS | 4249 | 0.005889545 | [-0.02 0.04] | 0.701968039 | 0.859880159 |
| bOHBut | VAT_Neg_pPS | 4249 | -0.023652983 | [-0.05 0.01] | 0.124287933 | 0.375867736 |
| bOHBut | Bilirubin_pPS | 4249 | 0.02725104 | [-0. 0.06] | 0.076575227 | 0.261610248 |
| bOHBut | Proinsulin_pPS | 4249 | 0.015199839 | [-0.01 0.05] | 0.323330329 | 0.620844623 |
| bOHBut | Hyper_Insulin_pPS | 4249 | -0.002035228 | [-0.03 0.03] | 0.894798324 | 0.955915818 |
| bOHBut | Beta_Cell_1_pPS | 4249 | 0.032543045 | [0. 0.06] | 0.034431501 | 0.136498743 |
| bOHBut | Beta_Cell_2_pPS | 4249 | -0.018690008 | [-0.05 0.01] | 0.224574663 | 0.519767023 |
| Cit | Cholesterol_pPS | 4255 | -0.006779976 | [-0.04 0.02] | 0.659333253 | 0.845602289 |
| Cit | Lipodystrophy_1_pPS | 4255 | -0.011137988 | [-0.04 0.02] | 0.468939354 | 0.731735454 |
| Cit | Lipodystrophy_2_pPS | 4255 | -0.028184838 | [-0.06 0. ] | 0.066814395 | 0.234401596 |
| Cit | Liver-Lipid_pPS | 4255 | -0.019485361 | [-0.05 0.01] | 0.205139762 | 0.490083299 |
| Cit | ASAT_Pos_pPS | 4255 | 0.007713747 | [-0.02 0.04] | 0.615984562 | 0.828794354 |
| Cit | ALP_Neg_pPS | 4255 | 0.010077949 | [-0.02 0.04] | 0.512289369 | 0.767666412 |
| Cit | SHBG-LpA_pPS | 4255 | 0.005246023 | [-0.02 0.04] | 0.733032266 | 0.869905511 |
| Cit | Obesity_pPS | 4255 | -0.015685474 | [-0.05 0.01] | 0.307766339 | 0.607642507 |
| Cit | VAT_Neg_pPS | 4255 | -0.012783797 | [-0.04 0.02] | 0.405845637 | 0.678602222 |
| Cit | Bilirubin_pPS | 4255 | -0.008907299 | [-0.04 0.02] | 0.562483095 | 0.797755947 |
| Cit | Proinsulin_pPS | 4255 | 0.012262191 | [-0.02 0.04] | 0.42527207 | 0.693893617 |
| Cit | Hyper_Insulin_pPS | 4255 | 0.013797353 | [-0.02 0.04] | 0.369646382 | 0.653351391 |
| Cit | Beta_Cell_1_pPS | 4255 | -0.008856753 | [-0.04 0.02] | 0.564702708 | 0.797755947 |
| Cit | Beta_Cell_2_pPS | 4255 | 0.019685445 | [-0.01 0.05] | 0.200525269 | 0.486527589 |
| Crea | Cholesterol_pPS | 4249 | 0.012126952 | [-0.02 0.04] | 0.430723457 | 0.696591166 |
| Crea | Lipodystrophy_1_pPS | 4249 | 0.01688892 | [-0.01 0.05] | 0.272465158 | 0.572772495 |
| Crea | Lipodystrophy_2_pPS | 4249 | 0.004124185 | [-0.03 0.03] | 0.788728497 | 0.898710558 |
| Crea | Liver-Lipid_pPS | 4249 | 0.053114831 | [0.02 0.08] | 0.000553421 | 0.005893343 |
| Crea | ASAT_Pos_pPS | 4249 | -0.024093254 | [-0.05 0.01] | 0.117432928 | 0.360031733 |
| Crea | ALP_Neg_pPS | 4249 | -0.004610554 | [-0.03 0.03] | 0.764510181 | 0.885222315 |
| Crea | SHBG-LpA_pPS | 4249 | 0.019275635 | [-0.01 0.05] | 0.210382784 | 0.4972684 |
| Crea | Obesity_pPS | 4249 | 0.013054288 | [-0.02 0.04] | 0.396319168 | 0.673455453 |
| Crea | VAT_Neg_pPS | 4249 | -0.042175891 | [-0.07 -0.01] | 0.00611562 | 0.036116434 |
| Crea | Bilirubin_pPS | 4249 | 0.018090445 | [-0.01 0.05] | 0.23979955 | 0.53580212 |

**Table S9: Associations between pPSs and global metabolomic profile in SCAPIS**

**a) Spearman Correlation**

| Metabolomics | PPS | Samp | r | CI95% | p | fdr_p_adjusted |
|---|---|---|---|---|---|---|
| Crea | ASAT_Pos_pPS | 4249 | -0.020481265 | [-0.05 0.01] | 0.182558754 | 0.452703501 |
| Crea | ALP_Neg_pPS | 4249 | -0.006422126 | [-0.04 0.02] | 0.676011789 | 0.834318347 |
| Crea | SHBG-LpA_pPS | 4249 | 0.016327961 | [-0.01 0.05] | 0.287972098 | 0.57504794 |
| Crea | Obesity_pPS | 4249 | 0.011363616 | [-0.02 0.04] | 0.459610189 | 0.700511576 |
| Crea | VAT_Neg_pPS | 4249 | -0.040351038 | [-0.07 -0.01] | 0.008618328 | 0.042302775 |
| Crea | Bilirubin_pPS | 4249 | 0.016787737 | [-0.01 0.05] | 0.27461076 | 0.559392289 |
| Crea | Proinsulin_pPS | 4249 | -0.015852004 | [-0.05 0.01] | 0.302258198 | 0.587800395 |
| Crea | Hyper_Insulin_pPS | 4249 | -0.01370375 | [-0.04 0.02] | 0.3725074 | 0.64437772 |
| Crea | Beta_Cell_1_pPS | 4249 | 0.012435212 | [-0.02 0.04] | 0.418383863 | 0.679359111 |
| Crea | Beta_Cell_2_pPS | 4249 | -0.011808458 | [-0.04 0.02] | 0.442226418 | 0.692820992 |
| Crea | Total_GRS | 4249 | -0.00682649 | [-0.04 0.02] | 0.656879467 | 0.82301779 |
| DHA | Cholesterol_pPS | 4223 | -0.0532224 | [-0.08 -0.02] | 0.000550001 | 0.00559124 |
| DHA | Lipodystrophy_1_pPS | 4223 | 0.026929655 | [-0. 0.06] | 0.080581308 | 0.254539888 |
| DHA | Lipodystrophy_2_pPS | 4223 | -0.025971092 | [-0.06 0. ] | 0.091968191 | 0.283436451 |
| DHA | Liver-Lipid_pPS | 4223 | 0.028746924 | [-0. 0.06] | 0.062141699 | 0.206018461 |
| DHA | ASAT_Pos_pPS | 4223 | -0.011005992 | [-0.04 0.02] | 0.475219232 | 0.712330715 |
| DHA | ALP_Neg_pPS | 4223 | 0.069067218 | [0.04 0.1 ] | 7.26E-06 | 0.000134227 |
| DHA | SHBG-LpA_pPS | 4223 | 0.007882613 | [-0.02 0.04] | 0.609087551 | 0.794220546 |
| DHA | Obesity_pPS | 4223 | 0.002033051 | [-0.03 0.03] | 0.895071174 | 0.948580863 |
| DHA | VAT_Neg_pPS | 4223 | 0.002442391 | [-0.03 0.03] | 0.874105866 | 0.942282701 |
| DHA | Bilirubin_pPS | 4223 | -0.036066801 | [-0.07 -0.01] | 0.019256838 | 0.082777391 |
| DHA | Proinsulin_pPS | 4223 | -0.008938497 | [-0.04 0.02] | 0.561996707 | 0.767334778 |
| DHA | Hyper_Insulin_pPS | 4223 | -0.007952051 | [-0.04 0.02] | 0.605937395 | 0.792521775 |
| DHA | Beta_Cell_1_pPS | 4223 | 0.036261749 | [0.01 0.07] | 0.018613589 | 0.080334304 |
| DHA | Beta_Cell_2_pPS | 4223 | 0.021289214 | [-0.01 0.05] | 0.167201895 | 0.429518642 |
| DHA | Total_GRS | 4223 | 0.006079782 | [-0.02 0.04] | 0.693273155 | 0.847816943 |
| EstC | Cholesterol_pPS | 4223 | -0.134191345 | [-0.16 -0.1 ] | 2.23E-18 | 1.45E-16 |
| EstC | Lipodystrophy_1_pPS | 4223 | 0.023536916 | [-0.01 0.05] | 0.12673098 | 0.359462978 |
| EstC | Lipodystrophy_2_pPS | 4223 | -0.059791063 | [-0.09 -0.03] | 0.000103475 | 0.00147325 |
| EstC | Liver-Lipid_pPS | 4223 | -0.00536419 | [-0.04 0.02] | 0.727845177 | 0.870249668 |
| EstC | ASAT_Pos_pPS | 4223 | -0.028420994 | [-0.06 0. ] | 0.065164731 | 0.213076751 |
| EstC | ALP_Neg_pPS | 4223 | 0.053639719 | [0.02 0.08] | 0.000497175 | 0.005151884 |
| EstC | SHBG-LpA_pPS | 4223 | 0.018753342 | [-0.01 0.05] | 0.223721553 | 0.505139718 |
| EstC | Obesity_pPS | 4223 | -0.009574004 | [-0.04 0.02] | 0.53452845 | 0.749387925 |
| EstC | VAT_Neg_pPS | 4223 | 0.001293838 | [-0.03 0.03] | 0.933108034 | 0.96878835 |
| EstC | Bilirubin_pPS | 4223 | -0.012955549 | [-0.04 0.02] | 0.400628114 | 0.666677506 |
| EstC | Proinsulin_pPS | 4223 | -0.002322998 | [-0.03 0.03] | 0.880212397 | 0.944972769 |
| EstC | Hyper_Insulin_pPS | 4223 | -0.022225291 | [-0.05 0.01] | 0.149302833 | 0.398714883 |
| EstC | Beta_Cell_1_pPS | 4223 | 0.021372397 | [-0.01 0.05] | 0.165548854 | 0.427012507 |
| EstC | Beta_Cell_2_pPS | 4223 | 0.025589456 | [-0. 0.06] | 0.096846989 | 0.296343497 |
| EstC | Total_GRS | 4223 | -0.013175038 | [-0.04 0.02] | 0.392694704 | 0.660507155 |
| FAw3 | Cholesterol_pPS | 4223 | -0.068454541 | [-0.1 -0.04] | 8.75E-06 | 0.000157645 |
| FAw3 | Lipodystrophy_1_pPS | 4223 | 0.03047078 | [0. 0.06] | 0.04802032 | 0.169170784 |
| FAw3 | Lipodystrophy_2_pPS | 4223 | -0.028031196 | [-0.06 0. ] | 0.068938309 | 0.223594589 |
| FAw3 | Liver-Lipid_pPS | 4223 | 0.013372779 | [-0.02 0.04] | 0.385629527 | 0.654929006 |
| FAw3 | ASAT_Pos_pPS | 4223 | -0.008534655 | [-0.04 0.02] | 0.579798976 | 0.776336407 |
| FAw3 | ALP_Neg_pPS | 4223 | 0.070880054 | [0.04 0.1 ] | 4.14E-06 | 8.39E-05 |
| FAw3 | SHBG-LpA_pPS | 4223 | 0.007087625 | [-0.02 0.04] | 0.645659126 | 0.817402268 |
| FAw3 | Obesity_pPS | 4223 | 1.23E-05 | [-0.03 0.03] | 0.999364206 | 0.999364206 |
| FAw3 | VAT_Neg_pPS | 4223 | 0.00216862 | [-0.03 0.03] | 0.888118811 | 0.946826466 |
| FAw3 | Bilirubin_pPS | 4223 | -0.037228446 | [-0.07 -0.01] | 0.015693929 | 0.069266417 |
| FAw3 | Proinsulin_pPS | 4223 | -0.004878067 | [-0.04 0.03] | 0.75165604 | 0.881192614 |
| FAw3 | Hyper_Insulin_pPS | 4223 | -0.011080604 | [-0.04 0.02] | 0.472231137 | 0.710023653 |
| FAw3 | Beta_Cell_1_pPS | 4223 | 0.043916408 | [0.01 0.07] | 0.004366947 | 0.024735931 |
| FAw3 | Beta_Cell_2_pPS | 4223 | 0.027923962 | [-0. 0.06] | 0.070007338 | 0.226153223 |
| FAw3 | Total_GRS | 4223 | 0.007908533 | [-0.02 0.04] | 0.607910804 | 0.793168294 |
| FAw6 | Cholesterol_pPS | 4223 | -0.142515845 | [-0.17 -0.11] | 1.50E-20 | 1.19E-18 |
| FAw6 | Lipodystrophy_1_pPS | 4223 | 0.02871194 | [-0. 0.06] | 0.062460522 | 0.20643732 |
| FAw6 | Lipodystrophy_2_pPS | 4223 | -0.058428807 | [-0.09 -0.03] | 0.000148425 | 0.001872771 |
| FAw6 | Liver-Lipid_pPS | 4223 | -0.034533171 | [-0.06 -0. ] | 0.025030149 | 0.102461202 |
| FAw6 | ASAT_Pos_pPS | 4223 | -0.025444467 | [-0.06 0. ] | 0.098753894 | 0.300038388 |
| FAw6 | ALP_Neg_pPS | 4223 | 0.05114848 | [0.02 0.08] | 0.000899123 | 0.007776689 |
| FAw6 | SHBG-LpA_pPS | 4223 | 0.016698071 | [-0.01 0.05] | 0.278662557 | 0.564963312 |
| FAw6 | Obesity_pPS | 4223 | 0.005366113 | [-0.02 0.04] | 0.727751482 | 0.870249668 |
| FAw6 | VAT_Neg_pPS | 4223 | -0.002545287 | [-0.03 0.03] | 0.868849098 | 0.940757973 |
| FAw6 | Bilirubin_pPS | 4223 | -0.018765804 | [-0.05 0.01] | 0.223413907 | 0.504976639 |
| FAw6 | Proinsulin_pPS | 4223 | 0.01032231 | [-0.02 0.04] | 0.503076745 | 0.727744052 |
| FAw6 | Hyper_Insulin_pPS | 4223 | -0.023325928 | [-0.05 0.01] | 0.130171038 | 0.366907855 |
| FAw6 | Beta_Cell_1_pPS | 4223 | 0.029834142 | [-0. 0.06] | 0.052884879 | 0.18179177 |
| FAw6 | Beta_Cell_2_pPS | 4223 | 0.02527414 | [-0. 0.06] | 0.101032192 | 0.305231058 |
| FAw6 | Total_GRS | 4223 | -0.0057984 | [-0.04 0.02] | 0.706796087 | 0.858243888 |
| FreeC | Cholesterol_pPS | 4223 | -0.136320549 | [-0.17 -0.11] | 6.39E-19 | 4.42E-17 |
| FreeC | Lipodystrophy_1_pPS | 4223 | 0.029002949 | [-0. 0.06] | 0.059849213 | 0.199652507 |
| FreeC | Lipodystrophy_2_pPS | 4223 | -0.057025438 | [-0.09 -0.03] | 0.000213551 | 0.002543104 |
| FreeC | Liver-Lipid_pPS | 4223 | -0.001034917 | [-0.03 0.03] | 0.946471727 | 0.974500144 |
| FreeC | ASAT_Pos_pPS | 4223 | -0.026206488 | [-0.06 0. ] | 0.089058665 | 0.275260571 |
| FreeC | ALP_Neg_pPS | 4223 | 0.055674817 | [0.03 0.09] | 0.000300819 | 0.003360716 |
| FreeC | SHBG-LpA_pPS | 4223 | 0.017635701 | [-0.01 0.05] | 0.252553565 | 0.534775319 |
| FreeC | Obesity_pPS | 4223 | -0.003199039 | [-0.03 0.03] | 0.835594671 | 0.925322958 |
| FreeC | VAT_Neg_pPS | 4223 | 0.003128113 | [-0.03 0.03] | 0.83918943 | 0.926178923 |
| FreeC | Bilirubin_pPS | 4223 | -0.012022203 | [-0.04 0.02] | 0.435423859 | 0.690341841 |
| FreeC | Proinsulin_pPS | 4223 | 0.000181602 | [-0.03 0.03] | 0.990600317 | 0.993843629 |
| FreeC | Hyper_Insulin_pPS | 4223 | -0.021364596 | [-0.05 0.01] | 0.165703365 | 0.427012507 |
| FreeC | Beta_Cell_1_pPS | 4223 | 0.019866857 | [-0.01 0.05] | 0.197415852 | 0.47103115 |
| FreeC | Beta_Cell_2_pPS | 4223 | 0.026855528 | [-0. 0.06] | 0.081418949 | 0.255700798 |
| FreeC | Total_GRS | 4223 | -0.006769421 | [-0.04 0.02] | 0.660547518 | 0.824723182 |
| Gln | Cholesterol_pPS | 4252 | -0.006496349 | [-0.04 0.02] | 0.672374553 | 0.832193459 |
| Gln | Lipodystrophy_1_pPS | 4252 | -0.006804838 | [-0.04 0.02] | 0.657784938 | 0.823091532 |
| Gln | Lipodystrophy_2_pPS | 4252 | 0.000862198 | [-0.03 0.03] | 0.95524165 | 0.977573158 |
| Gln | Liver-Lipid_pPS | 4252 | 0.051754218 | [0.02 0.08] | 0.000748262 | 0.006948151 |
| Gln | ASAT_Pos_pPS | 4252 | -0.000392694 | [-0.03 0.03] | 0.979606017 | 0.987896053 |
| Gln | ALP_Neg_pPS | 4252 | 0.011866669 | [-0.02 0.04] | 0.439817839 | 0.691142319 |
| Gln | SHBG-LpA_pPS | 4252 | 0.005082806 | [-0.03 0.04] | 0.740740496 | 0.875420586 |
| Gln | Obesity_pPS | 4252 | -0.017836697 | [-0.05 0.01] | 0.245564043 | 0.526734873 |

**b) Partial Correlation**

| Metabolomics | PPS | Samp | r | CI95% | p | fdr_p_adjusted |
|---|---|---|---|---|---|---|
| Crea | Proinsulin_pPS | 4249 | -0.021308368 | [-0.05 0.01] | 0.166166461 | 0.441605751 |
| Crea | Hyper_Insulin_pPS | 4249 | -0.013225984 | [-0.04 0.02] | 0.390136213 | 0.668709503 |
| Crea | Beta_Cell_1_pPS | 4249 | 0.023947193 | [-0.01 0.05] | 0.119673415 | 0.364405465 |
| Crea | Beta_Cell_2_pPS | 4249 | -0.01301313 | [-0.04 0.02] | 0.39781007 | 0.673455453 |
| DHA | Cholesterol_pPS | 4223 | -0.053227871 | [-0.08 -0.02] | 0.000560259 | 0.005903363 |
| DHA | Lipodystrophy_1_pPS | 4223 | 0.02871946 | [-0. 0.06] | 0.062797836 | 0.224855592 |
| DHA | Lipodystrophy_2_pPS | 4223 | -0.023564268 | [-0.05 0.01] | 0.126877249 | 0.379116795 |
| DHA | Liver-Lipid_pPS | 4223 | 0.026528692 | [-0. 0.06] | 0.085680502 | 0.287323895 |
| DHA | ASAT_Pos_pPS | 4223 | -0.013599121 | [-0.04 0.02] | 0.378376138 | 0.658703502 |
| DHA | ALP_Neg_pPS | 4223 | 0.07275525 | [0.04 0.1 ] | 2.37E-06 | 4.95E-05 |
| DHA | SHBG-LpA_pPS | 4223 | 0.008381088 | [-0.02 0.04] | 0.587215848 | 0.814131667 |
| DHA | Obesity_pPS | 4223 | 0.008797759 | [-0.02 0.04] | 0.568768943 | 0.797755947 |
| DHA | VAT_Neg_pPS | 4223 | -0.001583622 | [-0.03 0.03] | 0.918299867 | 0.967598071 |
| DHA | Bilirubin_pPS | 4223 | -0.037494952 | [-0.07 -0.01] | 0.01511955 | 0.074008166 |
| DHA | Proinsulin_pPS | 4223 | -0.03405182 | [-0.06 -0. ] | 0.027365284 | 0.115580797 |
| DHA | Hyper_Insulin_pPS | 4223 | -0.01174731 | [-0.04 0.02] | 0.446698289 | 0.709019056 |
| DHA | Beta_Cell_1_pPS | 4223 | 0.038418373 | [0.01 0.07] | 0.012796613 | 0.065354132 |
| DHA | Beta_Cell_2_pPS | 4223 | 0.016264948 | [-0.01 0.05] | 0.292068867 | 0.588843777 |
| EstC | Cholesterol_pPS | 4223 | -0.132220636 | [-0.16 -0.1 ] | 7.83E-18 | 4.48E-16 |
| EstC | Lipodystrophy_1_pPS | 4223 | 0.03146899 | [0. 0.06] | 0.041466425 | 0.159953339 |
| EstC | Lipodystrophy_2_pPS | 4223 | -0.053015865 | [-0.08 -0.02] | 0.000589508 | 0.006115 |
| EstC | Liver-Lipid_pPS | 4223 | -0.003242134 | [-0.03 0.03] | 0.833665902 | 0.923118991 |
| EstC | ASAT_Pos_pPS | 4223 | -0.027926487 | [-0.06 0. ] | 0.070415859 | 0.243475906 |
| EstC | ALP_Neg_pPS | 4223 | 0.057576497 | [0.03 0.09] | 0.000189603 | 0.002497277 |
| EstC | SHBG-LpA_pPS | 4223 | 0.018018774 | [-0.01 0.05] | 0.243121917 | 0.540756944 |
| EstC | Obesity_pPS | 4223 | 0.004334861 | [-0.03 0.03] | 0.778877698 | 0.891327566 |
| EstC | VAT_Neg_pPS | 4223 | 0.000406794 | [-0.03 0.03] | 0.978978861 | 0.99236237 |
| EstC | Bilirubin_pPS | 4223 | -0.013838862 | [-0.04 0.02] | 0.370026847 | 0.653351391 |
| EstC | Proinsulin_pPS | 4223 | -0.018711934 | [-0.05 0.01] | 0.225463299 | 0.521221161 |
| EstC | Hyper_Insulin_pPS | 4223 | -0.020160994 | [-0.05 0.01] | 0.191547682 | 0.474464643 |
| EstC | Beta_Cell_1_pPS | 4223 | 0.01672805 | [-0.01 0.05] | 0.278543761 | 0.576656518 |
| EstC | Beta_Cell_2_pPS | 4223 | 0.025123291 | [-0.01 0.06] | 0.103618266 | 0.332441936 |
| FAw3 | Cholesterol_pPS | 4223 | -0.067956281 | [-0.1 -0.04] | 1.05E-05 | 0.000212144 |
| FAw3 | Lipodystrophy_1_pPS | 4223 | 0.03250308 | [0. 0.06] | 0.035215825 | 0.138783627 |
| FAw3 | Lipodystrophy_2_pPS | 4223 | -0.024690148 | [-0.05 0.01] | 0.109711574 | 0.343926372 |
| FAw3 | Liver-Lipid_pPS | 4223 | 0.011100272 | [-0.02 0.04] | 0.472130742 | 0.734922314 |
| FAw3 | ASAT_Pos_pPS | 4223 | -0.011489762 | [-0.04 0.02] | 0.456726912 | 0.72054159 |
| FAw3 | ALP_Neg_pPS | 4223 | 0.074954872 | [0.04 0.1 ] | 1.16E-06 | 2.53E-05 |
| FAw3 | SHBG-LpA_pPS | 4223 | 0.007232579 | [-0.02 0.04] | 0.63944161 | 0.836160747 |
| FAw3 | Obesity_pPS | 4223 | 0.007809002 | [-0.02 0.04] | 0.612984647 | 0.828794354 |
| FAw3 | VAT_Neg_pPS | 4223 | -0.002937844 | [-0.03 0.03] | 0.849079968 | 0.932962731 |
| FAw3 | Bilirubin_pPS | 4223 | -0.039458604 | [-0.07 -0.01] | 0.010562924 | 0.05639193 |
| FAw3 | Proinsulin_pPS | 4223 | -0.032383321 | [-0.06 -0. ] | 0.035895898 | 0.140633243 |
| FAw3 | Hyper_Insulin_pPS | 4223 | -0.014545041 | [-0.04 0.02] | 0.346105033 | 0.637861136 |
| FAw3 | Beta_Cell_1_pPS | 4223 | 0.04405351 | [0.01 0.07] | 0.004305718 | 0.028142462 |
| FAw3 | Beta_Cell_2_pPS | 4223 | 0.021696431 | [-0.01 0.05] | 0.159872238 | 0.433691357 |
| FAw6 | Cholesterol_pPS | 4223 | -0.141547554 | [-0.17 -0.11] | 3.12E-20 | 2.31E-18 |
| FAw6 | Lipodystrophy_1_pPS | 4223 | 0.036446126 | [0.01 0.07] | 0.018201282 | 0.084362666 |
| FAw6 | Lipodystrophy_2_pPS | 4223 | -0.050121946 | [-0.08 -0.02] | 0.001159848 | 0.009756369 |
| FAw6 | Liver-Lipid_pPS | 4223 | -0.033130718 | [-0.06 -0. ] | 0.031828757 | 0.12795416 |
| FAw6 | ASAT_Pos_pPS | 4223 | -0.026544 | [-0.06 0. ] | 0.0854999 | 0.287199329 |
| FAw6 | ALP_Neg_pPS | 4223 | 0.054241401 | [0.02 0.08] | 0.000438185 | 0.004846662 |
| FAw6 | SHBG-LpA_pPS | 4223 | 0.015339871 | [-0.01 0.05] | 0.320386955 | 0.620672596 |
| FAw6 | Obesity_pPS | 4223 | 0.019034656 | [-0.01 0.05] | 0.217561824 | 0.510619896 |
| FAw6 | VAT_Neg_pPS | 4223 | -0.004417206 | [-0.03 0.03] | 0.77478954 | 0.89042977 |
| FAw6 | Bilirubin_pPS | 4223 | -0.020891388 | [-0.05 0.01] | 0.175947403 | 0.453447086 |
| FAw6 | Proinsulin_pPS | 4223 | -0.008266085 | [-0.04 0.02] | 0.592355555 | 0.817111563 |
| FAw6 | Hyper_Insulin_pPS | 4223 | -0.021183138 | [-0.05 0.01] | 0.169987928 | 0.447195575 |
| FAw6 | Beta_Cell_1_pPS | 4223 | 0.023610913 | [-0.01 0.05] | 0.126126833 | 0.378852721 |
| FAw6 | Beta_Cell_2_pPS | 4223 | 0.022270591 | [-0.01 0.05] | 0.149103648 | 0.424616648 |
| FreeC | Cholesterol_pPS | 4223 | -0.135182763 | [-0.16 -0.11] | 1.41E-18 | 9.11E-17 |
| FreeC | Lipodystrophy_1_pPS | 4223 | 0.036473344 | [0.01 0.07] | 0.018114832 | 0.084339289 |
| FreeC | Lipodystrophy_2_pPS | 4223 | -0.050906986 | [-0.08 -0.02] | 0.000968499 | 0.008545465 |
| FreeC | Liver-Lipid_pPS | 4223 | 0.000969759 | [-0.03 0.03] | 0.949914694 | 0.978747744 |
| FreeC | ASAT_Pos_pPS | 4223 | -0.027000834 | [-0.06 0. ] | 0.08025036 | 0.271387198 |
| FreeC | ALP_Neg_pPS | 4223 | 0.058821246 | [0.03 0.09] | 0.000137117 | 0.001893161 |
| FreeC | SHBG-LpA_pPS | 4223 | 0.016347122 | [-0.01 0.05] | 0.289637287 | 0.5869802 |
| FreeC | Obesity_pPS | 4223 | 0.010450356 | [-0.02 0.04] | 0.498459228 | 0.754822494 |
| FreeC | VAT_Neg_pPS | 4223 | 0.001976222 | [-0.03 0.03] | 0.898144762 | 0.958467917 |
| FreeC | Bilirubin_pPS | 4223 | -0.013008003 | [-0.04 0.02] | 0.399452831 | 0.674856176 |
| FreeC | Proinsulin_pPS | 4223 | -0.016718339 | [-0.05 0.01] | 0.278822932 | 0.576656518 |
| FreeC | Hyper_Insulin_pPS | 4223 | -0.020780581 | [-0.05 0.01] | 0.178251167 | 0.455834788 |
| FreeC | Beta_Cell_1_pPS | 4223 | 0.01383732 | [-0.02 0.04] | 0.370080159 | 0.653351391 |
| FreeC | Beta_Cell_2_pPS | 4223 | 0.026399915 | [-0. 0.06] | 0.087212019 | 0.291839812 |
| Gln | Cholesterol_pPS | 4252 | -0.006196267 | [-0.04 0.02] | 0.687136649 | 0.858568103 |
| Gln | Lipodystrophy_1_pPS | 4252 | -0.005466873 | [-0.04 0.02] | 0.722343616 | 0.866319132 |
| Gln | Lipodystrophy_2_pPS | 4252 | -0.000275463 | [-0.03 0.03] | 0.985715411 | 0.993363084 |
| Gln | Liver-Lipid_pPS | 4252 | 0.050382581 | [0.02 0.08] | 0.001050074 | 0.009004786 |
| Gln | ASAT_Pos_pPS | 4252 | 0.00214547 | [-0.03 0.03] | 0.889096578 | 0.953849648 |
| Gln | ALP_Neg_pPS | 4252 | 0.007365326 | [-0.02 0.04] | 0.632134723 | 0.835456904 |
| Gln | SHBG-LpA_pPS | 4252 | 0.004295326 | [-0.03 0.03] | 0.780105946 | 0.891560895 |
| Gln | Obesity_pPS | 4252 | -0.019971022 | [-0.05 0.01] | 0.19422923 | 0.478398712 |
| Gln | VAT_Neg_pPS | 4252 | -0.00057086 | [-0.03 0.03] | 0.970402309 | 0.988674515 |
| Gln | Bilirubin_pPS | 4252 | -0.005995199 | [-0.04 0.02] | 0.696781804 | 0.858963776 |
| Gln | Proinsulin_pPS | 4252 | 0.015005135 | [-0.02 0.05] | 0.329396122 | 0.627451033 |
| Gln | Hyper_Insulin_pPS | 4252 | -0.012256654 | [-0.04 0.02] | 0.425645438 | 0.693893617 |
| Gln | Beta_Cell_1_pPS | 4252 | -0.00102407 | [-0.03 0.03] | 0.946931485 | 0.978747744 |
| Gln | Beta_Cell_2_pPS | 4252 | 0.045860633 | [0.02 0.08] | 0.002860599 | 0.019954421 |
| Gp | Cholesterol_pPS | 4254 | -0.053271441 | [-0.08 -0.02] | 0.000528815 | 0.005661435 |
| Gp | Lipodystrophy_1_pPS | 4254 | 0.039332738 | [0.01 0.07] | 0.010525441 | 0.056342065 |
| Gp | Lipodystrophy_2_pPS | 4254 | 0.010346485 | [-0.02 0.04] | 0.501164312 | 0.756659844 |
| Gp | Liver-Lipid_pPS | 4254 | -0.063620651 | [-0.09 -0.03] | 3.47E-05 | 0.000631351 |
| Gp | ASAT_Pos_pPS | 4254 | 0.031341075 | [0. 0.06] | 0.04154672 | 0.159954873 |
| Gp | ALP_Neg_pPS | 4254 | 0.021277023 | [-0.01 0.05] | 0.166539631 | 0.441605751 |

**Table S9: Associations between pPSs and global metabolomic profile in SCAPIS**

**a) Spearman Correlation**

| Metabolomics | PPS | Samp | r | CI95% | p | fdr_p_adjusted |
|---|---|---|---|---|---|---|
| Gln | VAT_Neg_pPS | 4252 | 0.003489328 | [-0.03 0.03] | 0.820313099 | 0.918836343 |
| Gln | Bilirubin_pPS | 4252 | -0.005856674 | [-0.04 0.02] | 0.703017904 | 0.855830536 |
| Gln | Proinsulin_pPS | 4252 | 0.024349287 | [-0.01 0.05] | 0.112903135 | 0.330843202 |
| Gln | Hyper_Insulin_pPS | 4252 | -0.010752145 | [-0.04 0.02] | 0.483963066 | 0.717164725 |
| Gln | Beta_Cell_1_pPS | 4252 | 0.016961107 | [-0.01 0.05] | 0.269514621 | 0.554275035 |
| Gln | Beta_Cell_2_pPS | 4252 | 0.051142036 | [0.02 0.08] | 0.000864333 | 0.007629602 |
| Gln | Total_GRS | 4252 | 0.014061671 | [-0.02 0.04] | 0.35997908 | 0.637617776 |
| Gp | Cholesterol_pPS | 4254 | -0.05152465 | [-0.08 -0.02] | 0.00078772 | 0.007220765 |
| Gp | Lipodystrophy_1_pPS | 4254 | 0.048967147 | [0.02 0.08] | 0.00142157 | 0.01104807 |
| Gp | Lipodystrophy_2_pPS | 4254 | 0.012005608 | [-0.02 0.04] | 0.434372884 | 0.69017025 |
| Gp | Liver-Lipid_pPS | 4254 | -0.061215919 | [-0.09 -0.03] | 6.62E-05 | 0.000999769 |
| Gp | ASAT_Pos_pPS | 4254 | 0.042914624 | [0.01 0.07] | 0.005181614 | 0.027786407 |
| Gp | ALP_Neg_pPS | 4254 | 0.020685412 | [-0.01 0.05] | 0.177979865 | 0.447033736 |
| Gp | SHBG-LpA_pPS | 4254 | -0.001775613 | [-0.03 0.03] | 0.907959415 | 0.952872131 |
| Gp | Obesity_pPS | 4254 | 0.037727087 | [0.01 0.07] | 0.013997019 | 0.062549179 |
| Gp | VAT_Neg_pPS | 4254 | 0.015213871 | [-0.01 0.05] | 0.321854339 | 0.608511855 |
| Gp | Bilirubin_pPS | 4254 | 0.008340735 | [-0.02 0.04] | 0.587070094 | 0.779730867 |
| Gp | Proinsulin_pPS | 4254 | 0.017524017 | [-0.01 0.05] | 0.253827629 | 0.536320571 |
| Gp | Hyper_Insulin_pPS | 4254 | 0.010341538 | [-0.02 0.04] | 0.500711635 | 0.726675547 |
| Gp | Beta_Cell_1_pPS | 4254 | 0.039802451 | [0.01 0.07] | 0.009524729 | 0.045603891 |
| Gp | Beta_Cell_2_pPS | 4254 | 0.001535534 | [-0.03 0.03] | 0.92035951 | 0.960199976 |
| Gp | Total_GRS | 4254 | 0.039352446 | [0.01 0.07] | 0.010368617 | 0.048666705 |
| HDL2_C | Cholesterol_pPS | 4254 | 0.039746988 | [0.01 0.07] | 0.00962532 | 0.045880691 |
| HDL2_C | Lipodystrophy_1_pPS | 4254 | -0.075313252 | [-0.11 -0.05] | 9.07E-07 | 2.09E-05 |
| HDL2_C | Lipodystrophy_2_pPS | 4254 | -0.042331433 | [-0.07 -0.01] | 0.005824581 | 0.030772723 |
| HDL2_C | Liver-Lipid_pPS | 4254 | -0.005189502 | [-0.04 0.02] | 0.73543986 | 0.873878512 |
| HDL2_C | ASAT_Pos_pPS | 4254 | -0.059263446 | [-0.09 -0.03] | 0.000112406 | 0.001535736 |
| HDL2_C | ALP_Neg_pPS | 4254 | 0.015467919 | [-0.01 0.05] | 0.313839259 | 0.602672525 |
| HDL2_C | SHBG-LpA_pPS | 4254 | 0.007047949 | [-0.02 0.04] | 0.646300348 | 0.817402268 |
| HDL2_C | Obesity_pPS | 4254 | -0.029961643 | [-0.06 0. ] | 0.051023285 | 0.177332222 |
| HDL2_C | VAT_Neg_pPS | 4254 | -0.031378392 | [-0.06 -0. ] | 0.04099352 | 0.151084365 |
| HDL2_C | Bilirubin_pPS | 4254 | -0.004925897 | [-0.04 0.03] | 0.748411824 | 0.879158468 |
| HDL2_C | Proinsulin_pPS | 4254 | -0.006643314 | [-0.04 0.02] | 0.665334082 | 0.826370357 |
| HDL2_C | Hyper_Insulin_pPS | 4254 | -0.018618389 | [-0.05 0.01] | 0.225368765 | 0.506725367 |
| HDL2_C | Beta_Cell_1_pPS | 4254 | 0.001925459 | [-0.03 0.03] | 0.900231005 | 0.951229313 |
| HDL2_C | Beta_Cell_2_pPS | 4254 | 0.008818274 | [-0.02 0.04] | 0.565844836 | 0.768674587 |
| HDL2_C | Total_GRS | 4254 | -0.0469076 | [-0.08 -0.02] | 0.002243834 | 0.015183038 |
| HDL3_C | Cholesterol_pPS | 4254 | -0.06021271 | [-0.09 -0.03] | 8.71E-05 | 0.001270302 |
| HDL3_C | Lipodystrophy_1_pPS | 4254 | -0.029473092 | [-0.06 0. ] | 0.054927091 | 0.187310986 |
| HDL3_C | Lipodystrophy_2_pPS | 4254 | -0.051455682 | [-0.08 -0.02] | 0.000800637 | 0.007255522 |
| HDL3_C | Liver-Lipid_pPS | 4254 | -0.000235975 | [-0.03 0.03] | 0.987741249 | 0.991903081 |
| HDL3_C | ASAT_Pos_pPS | 4254 | -0.048337242 | [-0.08 -0.02] | 0.001637471 | 0.012324121 |
| HDL3_C | ALP_Neg_pPS | 4254 | 0.033447699 | [0. 0.06] | 0.029373484 | 0.117768453 |
| HDL3_C | SHBG-LpA_pPS | 4254 | 0.008503164 | [-0.02 0.04] | 0.579809289 | 0.776336407 |
| HDL3_C | Obesity_pPS | 4254 | -0.023874758 | [-0.05 0.01] | 0.120007642 | 0.347390543 |
| HDL3_C | VAT_Neg_pPS | 4254 | -0.017115164 | [-0.05 0.01] | 0.265075601 | 0.551490946 |
| HDL3_C | Bilirubin_pPS | 4254 | -0.015822232 | [-0.05 0.01] | 0.302881937 | 0.588479851 |
| HDL3_C | Proinsulin_pPS | 4254 | -0.00472926 | [-0.03 0.03] | 0.758135114 | 0.886212435 |
| HDL3_C | Hyper_Insulin_pPS | 4254 | -0.030378072 | [-0.06 -0. ] | 0.047881724 | 0.169170784 |
| HDL3_C | Beta_Cell_1_pPS | 4254 | 0.012542673 | [-0.02 0.04] | 0.414097798 | 0.677528433 |
| HDL3_C | Beta_Cell_2_pPS | 4254 | 0.019061221 | [-0.01 0.05] | 0.214525829 | 0.495859809 |
| HDL3_C | Total_GRS | 4254 | -0.035643661 | [-0.07 -0.01] | 0.020257295 | 0.086385483 |
| HDL_C | Cholesterol_pPS | 4254 | 0.032489773 | [0. 0.06] | 0.034343563 | 0.131105874 |
| HDL_C | Lipodystrophy_1_pPS | 4254 | -0.073283652 | [-0.1 -0.04] | 1.77E-06 | 4.04E-05 |
| HDL_C | Lipodystrophy_2_pPS | 4254 | -0.043489176 | [-0.07 -0.01] | 0.00461158 | 0.025363691 |
| HDL_C | Liver-Lipid_pPS | 4254 | -0.004578847 | [-0.03 0.03] | 0.765598688 | 0.889697961 |
| HDL_C | ASAT_Pos_pPS | 4254 | -0.059480862 | [-0.09 -0.03] | 0.00010605 | 0.001477118 |
| HDL_C | ALP_Neg_pPS | 4254 | 0.017080917 | [-0.01 0.05] | 0.266033131 | 0.552249619 |
| HDL_C | SHBG-LpA_pPS | 4254 | 0.007468073 | [-0.02 0.04] | 0.626780141 | 0.803929814 |
| HDL_C | Obesity_pPS | 4254 | -0.030603514 | [-0.06 -0. ] | 0.046249866 | 0.165896258 |
| HDL_C | VAT_Neg_pPS | 4254 | -0.030581072 | [-0.06 -0. ] | 0.046410186 | 0.166134098 |
| HDL_C | Bilirubin_pPS | 4254 | -0.006419203 | [-0.04 0.02] | 0.675970619 | 0.834318347 |
| HDL_C | Proinsulin_pPS | 4254 | -0.007007829 | [-0.04 0.02] | 0.64817755 | 0.818328925 |
| HDL_C | Hyper_Insulin_pPS | 4254 | -0.020002818 | [-0.05 0.01] | 0.192730943 | 0.464503229 |
| HDL_C | Beta_Cell_1_pPS | 4254 | 0.002638794 | [-0.03 0.03] | 0.863582199 | 0.937915857 |
| HDL_C | Beta_Cell_2_pPS | 4254 | 0.009938247 | [-0.02 0.04] | 0.517561024 | 0.736667707 |
| HDL_C | Total_GRS | 4254 | -0.047228797 | [-0.08 -0.02] | 0.002091991 | 0.014475227 |
| HDL_D | Cholesterol_pPS | 4254 | 0.029898792 | [-0. 0.06] | 0.051512105 | 0.178215267 |
| HDL_D | Lipodystrophy_1_pPS | 4254 | -0.065451105 | [-0.1 -0.04] | 1.99E-05 | 0.000330627 |
| HDL_D | Lipodystrophy_2_pPS | 4254 | -0.04077578 | [-0.07 -0.01] | 0.007905616 | 0.039344654 |
| HDL_D | Liver-Lipid_pPS | 4254 | 0.020380755 | [-0.01 0.05] | 0.184456344 | 0.455303634 |
| HDL_D | ASAT_Pos_pPS | 4254 | -0.052067164 | [-0.08 -0.02] | 0.000692657 | 0.006603329 |
| HDL_D | ALP_Neg_pPS | 4254 | 0.018207367 | [-0.01 0.05] | 0.235777841 | 0.518179785 |
| HDL_D | SHBG-LpA_pPS | 4254 | 0.009095043 | [-0.02 0.04] | 0.55371424 | 0.760607725 |
| HDL_D | Obesity_pPS | 4254 | -0.03429266 | [-0.06 -0. ] | 0.025515893 | 0.104052452 |
| HDL_D | VAT_Neg_pPS | 4254 | -0.016169035 | [-0.05 0.01] | 0.292406079 | 0.57860797 |
| HDL_D | Bilirubin_pPS | 4254 | -0.008134624 | [-0.04 0.02] | 0.596343595 | 0.787173546 |
| HDL_D | Proinsulin_pPS | 4254 | -0.007405235 | [-0.04 0.02] | 0.629683641 | 0.803929814 |
| HDL_D | Hyper_Insulin_pPS | 4254 | -0.012829041 | [-0.04 0.02] | 0.403519687 | 0.668142396 |
| HDL_D | Beta_Cell_1_pPS | 4254 | -0.00958714 | [-0.04 0.02] | 0.532465484 | 0.746983952 |
| HDL_D | Beta_Cell_2_pPS | 4254 | 0.011466139 | [-0.02 0.04] | 0.455304185 | 0.696320363 |
| HDL_D | Total_GRS | 4254 | -0.045966767 | [-0.08 -0.02] | 0.002748542 | 0.01786552 |
| HDL_TG | Cholesterol_pPS | 4254 | -0.058609622 | [-0.09 -0.03] | 0.000133755 | 0.001717987 |
| HDL_TG | Lipodystrophy_1_pPS | 4254 | 0.041711756 | [0.01 0.07] | 0.006585768 | 0.034087051 |
| HDL_TG | Lipodystrophy_2_pPS | 4254 | -0.028421297 | [-0.06 0. ] | 0.064180268 | 0.21082186 |
| HDL_TG | Liver-Lipid_pPS | 4254 | -0.045823094 | [-0.08 -0.02] | 0.002834154 | 0.018311022 |
| HDL_TG | ASAT_Pos_pPS | 4254 | 0.016003192 | [-0.01 0.05] | 0.297384953 | 0.583614568 |
| HDL_TG | ALP_Neg_pPS | 4254 | 0.046433198 | [0.02 0.08] | 0.002486627 | 0.016513356 |
| HDL_TG | SHBG-LpA_pPS | 4254 | -0.007752846 | [-0.04 0.02] | 0.613694708 | 0.796469827 |
| HDL_TG | Obesity_pPS | 4254 | 0.016133659 | [-0.01 0.05] | 0.2934634 | 0.57909751 |
| HDL_TG | VAT_Neg_pPS | 4254 | 0.004993564 | [-0.03 0.04] | 0.745074996 | 0.87716019 |
| HDL_TG | Bilirubin_pPS | 4254 | -0.010511347 | [-0.04 0.02] | 0.493705037 | 0.721879553 |
| HDL_TG | Proinsulin_pPS | 4254 | 0.026373947 | [-0. 0.06] | 0.085879733 | 0.266973952 |
| HDL_TG | Hyper_Insulin_pPS | 4254 | 0.006487163 | [-0.02 0.04] | 0.672738209 | 0.832193459 |

**b) Partial Correlation**

| Metabolomics | PPS | Samp | r | CI95% | p | fdr_p_adjusted |
|---|---|---|---|---|---|---|
| Gp | SHBG-LpA_pPS | 4254 | -0.005119048 | [-0.04 0.03] | 0.739285729 | 0.873045882 |
| Gp | Obesity_pPS | 4254 | 0.039049491 | [0.01 0.07] | 0.011096787 | 0.058516788 |
| Gp | VAT_Neg_pPS | 4254 | 0.007454437 | [-0.02 0.04] | 0.627938008 | 0.835009289 |
| Gp | Bilirubin_pPS | 4254 | 0.003858067 | [-0.03 0.03] | 0.801953456 | 0.906328459 |
| Gp | Proinsulin_pPS | 4254 | 0.000993396 | [-0.03 0.03] | 0.948506645 | 0.978747744 |
| Gp | Hyper_Insulin_pPS | 4254 | 0.002218874 | [-0.03 0.03] | 0.885301003 | 0.951558497 |
| Gp | Beta_Cell_1_pPS | 4254 | 0.036203648 | [0.01 0.07] | 0.018551306 | 0.08518283 |
| Gp | Beta_Cell_2_pPS | 4254 | -0.007838555 | [-0.04 0.02] | 0.610328666 | 0.828224707 |
| HDL2_C | Cholesterol_pPS | 4254 | 0.043262005 | [0.01 0.07] | 0.004895255 | 0.030247843 |
| HDL2_C | Lipodystrophy_1_pPS | 4254 | -0.064007706 | [-0.09 -0.03] | 3.11E-05 | 0.000575864 |
| HDL2_C | Lipodystrophy_2_pPS | 4254 | -0.032405232 | [-0.06 -0. ] | 0.035094411 | 0.138783627 |
| HDL2_C | Liver-Lipid_pPS | 4254 | -0.004551904 | [-0.03 0.03] | 0.767285222 | 0.88638489 |
| HDL2_C | ASAT_Pos_pPS | 4254 | -0.042107105 | [-0.07 -0.01] | 0.006168972 | 0.03632436 |
| HDL2_C | ALP_Neg_pPS | 4254 | 0.019535987 | [-0.01 0.05] | 0.204018461 | 0.489742157 |
| HDL2_C | SHBG-LpA_pPS | 4254 | 0.015177454 | [-0.01 0.05] | 0.323757136 | 0.620844623 |
| HDL2_C | Obesity_pPS | 4254 | -0.028731744 | [-0.06 0. ] | 0.061722975 | 0.222247116 |
| HDL2_C | VAT_Neg_pPS | 4254 | -0.025121181 | [-0.06 0.01] | 0.102379773 | 0.330444899 |
| HDL2_C | Bilirubin_pPS | 4254 | -0.002108035 | [-0.03 0.03] | 0.890993864 | 0.954053428 |
| HDL2_C | Proinsulin_pPS | 4254 | -0.009647388 | [-0.04 0.02] | 0.530524125 | 0.779815932 |
| HDL2_C | Hyper_Insulin_pPS | 4254 | -0.002481993 | [-0.03 0.03] | 0.871811097 | 0.946048963 |
| HDL2_C | Beta_Cell_1_pPS | 4254 | 0.008620766 | [-0.02 0.04] | 0.575165949 | 0.802986213 |
| HDL2_C | Beta_Cell_2_pPS | 4254 | 0.010005487 | [-0.02 0.04] | 0.515376809 | 0.769988337 |
| HDL3_C | Cholesterol_pPS | 4254 | -0.057205399 | [-0.09 -0.03] | 0.000197605 | 0.002568863 |
| HDL3_C | Lipodystrophy_1_pPS | 4254 | -0.017715561 | [-0.05 0.01] | 0.249400695 | 0.545779139 |
| HDL3_C | Lipodystrophy_2_pPS | 4254 | -0.041157738 | [-0.07 -0.01] | 0.007431523 | 0.043124372 |
| HDL3_C | Liver-Lipid_pPS | 4254 | 0.001797074 | [-0.03 0.03] | 0.906994282 | 0.96224905 |
| HDL3_C | ASAT_Pos_pPS | 4254 | -0.038497696 | [-0.07 -0.01] | 0.012289439 | 0.063085785 |
| HDL3_C | ALP_Neg_pPS | 4254 | 0.037023572 | [0.01 0.07] | 0.016049639 | 0.077826401 |
| HDL3_C | SHBG-LpA_pPS | 4254 | 0.012094193 | [-0.02 0.04] | 0.431697137 | 0.69698199 |
| HDL3_C | Obesity_pPS | 4254 | -0.014558799 | [-0.04 0.02] | 0.343873205 | 0.637439033 |
| HDL3_C | VAT_Neg_pPS | 4254 | -0.013994458 | [-0.04 0.02] | 0.362903835 | 0.650367164 |
| HDL3_C | Bilirubin_pPS | 4254 | -0.014686593 | [-0.04 0.02] | 0.339653823 | 0.634316189 |
| HDL3_C | Proinsulin_pPS | 4254 | -0.013900438 | [-0.04 0.02] | 0.366137251 | 0.650982927 |
| HDL3_C | Hyper_Insulin_pPS | 4254 | -0.020325115 | [-0.05 0.01] | 0.186333485 | 0.468642761 |
| HDL3_C | Beta_Cell_1_pPS | 4254 | 0.013426261 | [-0.02 0.04] | 0.382716917 | 0.661657399 |
| HDL3_C | Beta_Cell_2_pPS | 4254 | 0.019305608 | [-0.01 0.05] | 0.209404636 | 0.496715736 |
| HDL_C | Cholesterol_pPS | 4254 | 0.036066221 | [0.01 0.07] | 0.019002418 | 0.086657954 |
| HDL_C | Lipodystrophy_1_pPS | 4254 | -0.061716783 | [-0.09 -0.03] | 5.92E-05 | 0.000960172 |
| HDL_C | Lipodystrophy_2_pPS | 4254 | -0.033371881 | [-0.06 -0. ] | 0.029994596 | 0.122549349 |
| HDL_C | Liver-Lipid_pPS | 4254 | -0.003817207 | [-0.03 0.03] | 0.804007958 | 0.907699133 |
| HDL_C | ASAT_Pos_pPS | 4254 | -0.042524213 | [-0.07 -0.01] | 0.005678064 | 0.034034385 |
| HDL_C | ALP_Neg_pPS | 4254 | 0.021246141 | [-0.01 0.05] | 0.167155946 | 0.442294174 |
| HDL_C | SHBG-LpA_pPS | 4254 | 0.015382301 | [-0.01 0.05] | 0.31726878 | 0.618500765 |
| HDL_C | Obesity_pPS | 4254 | -0.028640007 | [-0.06 0. ] | 0.062558997 | 0.22445002 |
| HDL_C | VAT_Neg_pPS | 4254 | -0.024481395 | [-0.05 0.01] | 0.111427029 | 0.345855679 |
| HDL_C | Bilirubin_pPS | 4254 | -0.003667157 | [-0.03 0.03] | 0.811564419 | 0.911348446 |
| HDL_C | Proinsulin_pPS | 4254 | -0.01048268 | [-0.04 0.02] | 0.495546381 | 0.753425828 |
| HDL_C | Hyper_Insulin_pPS | 4254 | -0.004059319 | [-0.03 0.03] | 0.791854136 | 0.899824971 |
| HDL_C | Beta_Cell_1_pPS | 4254 | 0.009048977 | [-0.02 0.04] | 0.556330983 | 0.796929123 |
| HDL_C | Beta_Cell_2_pPS | 4254 | 0.011146317 | [-0.02 0.04] | 0.468659598 | 0.731735454 |
| HDL_D | Cholesterol_pPS | 4254 | 0.033434261 | [0. 0.06] | 0.029688578 | 0.122035016 |
| HDL_D | Lipodystrophy_1_pPS | 4254 | -0.055494544 | [-0.09 -0.03] | 0.000305527 | 0.003662667 |
| HDL_D | Lipodystrophy_2_pPS | 4254 | -0.034843302 | [-0.06 -0. ] | 0.023457223 | 0.10276009 |
| HDL_D | Liver-Lipid_pPS | 4254 | 0.021166494 | [-0.01 0.05] | 0.168753373 | 0.444593986 |
| HDL_D | ASAT_Pos_pPS | 4254 | -0.036172083 | [-0.07 -0.01] | 0.018654082 | 0.085458747 |
| HDL_D | ALP_Neg_pPS | 4254 | 0.020589834 | [-0.01 0.05] | 0.180662266 | 0.459575421 |
| HDL_D | SHBG-LpA_pPS | 4254 | 0.015186187 | [-0.01 0.05] | 0.323478769 | 0.620844623 |
| HDL_D | Obesity_pPS | 4254 | -0.033259232 | [-0.06 -0. ] | 0.030554094 | 0.124252113 |
| HDL_D | VAT_Neg_pPS | 4254 | -0.009564529 | [-0.04 0.02] | 0.534060909 | 0.781586915 |
| HDL_D | Bilirubin_pPS | 4254 | -0.004875055 | [-0.04 0.03] | 0.75129164 | 0.876790721 |
| HDL_D | Proinsulin_pPS | 4254 | -0.007242274 | [-0.04 0.02] | 0.637756751 | 0.835456904 |
| HDL_D | Hyper_Insulin_pPS | 4254 | -0.00015229 | [-0.03 0.03] | 0.992100573 | 0.994934804 |
| HDL_D | Beta_Cell_1_pPS | 4254 | -0.006878906 | [-0.04 0.02] | 0.654720865 | 0.844628798 |
| HDL_D | Beta_Cell_2_pPS | 4254 | 0.01458622 | [-0.02 0.04] | 0.342965016 | 0.636982428 |
| HDL_TG | Cholesterol_pPS | 4254 | -0.05837337 | [-0.09 -0.03] | 0.000145762 | 0.001985135 |
| HDL_TG | Lipodystrophy_1_pPS | 4254 | 0.040204625 | [0.01 0.07] | 0.008927406 | 0.049576757 |
| HDL_TG | Lipodystrophy_2_pPS | 4254 | -0.029044954 | [-0.06 0. ] | 0.058937997 | 0.21375701 |
| HDL_TG | Liver-Lipid_pPS | 4254 | -0.047354225 | [-0.08 -0.02] | 0.002067929 | 0.015801504 |
| HDL_TG | ASAT_Pos_pPS | 4254 | 0.010736316 | [-0.02 0.04] | 0.485174359 | 0.744305799 |
| HDL_TG | ALP_Neg_pPS | 4254 | 0.043566415 | [0.01 0.07] | 0.004601755 | 0.029166809 |
| HDL_TG | SHBG-LpA_pPS | 4254 | -0.011033209 | [-0.04 0.02] | 0.473184247 | 0.734922314 |
| HDL_TG | Obesity_pPS | 4254 | 0.016337256 | [-0.01 0.05] | 0.288153502 | 0.586008484 |
| HDL_TG | VAT_Neg_pPS | 4254 | 0.005099837 | [-0.03 0.04] | 0.740228767 | 0.873045882 |
| HDL_TG | Bilirubin_pPS | 4254 | -0.013319243 | [-0.04 0.02] | 0.386521499 | 0.664790413 |
| HDL_TG | Proinsulin_pPS | 4254 | 0.016453769 | [-0.01 0.05] | 0.284728611 | 0.583442529 |
| HDL_TG | Hyper_Insulin_pPS | 4254 | 0.003022159 | [-0.03 0.03] | 0.84423789 | 0.930194967 |
| HDL_TG | Beta_Cell_1_pPS | 4254 | 0.001601219 | [-0.03 0.03] | 0.917091744 | 0.966833951 |
| HDL_TG | Beta_Cell_2_pPS | 4254 | 0.008445906 | [-0.02 0.04] | 0.582942894 | 0.810452551 |
| His | Cholesterol_pPS | 4249 | -0.019084076 | [-0.05 0.01] | 0.214951735 | 0.505086119 |
| His | Lipodystrophy_1_pPS | 4249 | -0.001971362 | [-0.03 0.03] | 0.898081232 | 0.958467917 |
| His | Lipodystrophy_2_pPS | 4249 | 0.002432838 | [-0.03 0.03] | 0.874402265 | 0.946048963 |
| His | Liver-Lipid_pPS | 4249 | 0.021298171 | [-0.01 0.05] | 0.166369301 | 0.441605751 |
| His | ASAT_Pos_pPS | 4249 | -0.004621317 | [-0.03 0.03] | 0.763976722 | 0.885222315 |
| His | ALP_Neg_pPS | 4249 | -0.014293061 | [-0.04 0.02] | 0.353038597 | 0.639407383 |
| His | SHBG-LpA_pPS | 4249 | 0.004404961 | [-0.03 0.03] | 0.774720945 | 0.89042977 |
| His | Obesity_pPS | 4249 | 0.014477756 | [-0.02 0.04] | 0.34685167 | 0.637861136 |
| His | VAT_Neg_pPS | 4249 | -0.028022943 | [-0.06 0. ] | 0.068592528 | 0.239237354 |
| His | Bilirubin_pPS | 4249 | 0.006821382 | [-0.02 0.04] | 0.65761289 | 0.844628798 |
| His | Proinsulin_pPS | 4249 | -0.008098822 | [-0.04 0.02] | 0.598739571 | 0.820871565 |
| His | Hyper_Insulin_pPS | 4249 | 0.009318607 | [-0.02 0.04] | 0.544865642 | 0.789070018 |
| His | Beta_Cell_1_pPS | 4249 | 0.012165357 | [-0.02 0.04] | 0.429265089 | 0.696591166 |
| His | Beta_Cell_2_pPS | 4249 | 0.017888314 | [-0.01 0.05] | 0.245092636 | 0.542782585 |
| IDL_C | Cholesterol_pPS | 4255 | -0.14605175 | [-0.18 -0.12] | 1.33E-21 | 1.16E-19 |
| IDL_C | Lipodystrophy_1_pPS | 4255 | 0.040933908 | [0.01 0.07] | 0.007753791 | 0.044351684 |

**Table S9: Associations between pPSs and global metabolomic profile in SCAPIS**

**a) Spearman Correlation**

| Metabolomics | PPS | Samp | r | CI95% | p | fdr_p_adjusted |
|---|---|---|---|---|---|---|
| HDL_TG | Beta_Cell_1_pPS | 4254 | 0.011177225 | [-0.02 0.04] | 0.466743393 | 0.706538163 |
| HDL_TG | Beta_Cell_2_pPS | 4254 | 0.011028661 | [-0.02 0.04] | 0.472687075 | 0.710023653 |
| HDL_TG | Total_GRS | 4254 | 0.020612342 | [-0.01 0.05] | 0.179517569 | 0.449317602 |
| His | Cholesterol_pPS | 4249 | -0.016346278 | [-0.05 0.01] | 0.287431556 | 0.57504794 |
| His | Lipodystrophy_1_pPS | 4249 | -0.001243123 | [-0.03 0.03] | 0.935526388 | 0.969715263 |
| His | Lipodystrophy_2_pPS | 4249 | -0.00123766 | [-0.03 0.03] | 0.935809135 | 0.969715263 |
| His | Liver-Lipid_pPS | 4249 | 0.021071727 | [-0.01 0.05] | 0.170264778 | 0.43221059 |
| His | ASAT_Pos_pPS | 4249 | -0.003061452 | [-0.03 0.03] | 0.84209185 | 0.927489034 |
| His | ALP_Neg_pPS | 4249 | -0.014495327 | [-0.04 0.02] | 0.345525181 | 0.62662761 |
| His | SHBG-LpA_pPS | 4249 | 0.003157938 | [-0.03 0.03] | 0.83718373 | 0.926178923 |
| His | Obesity_pPS | 4249 | 0.012785168 | [-0.02 0.04] | 0.405406858 | 0.669952012 |
| His | VAT_Neg_pPS | 4249 | -0.023575575 | [-0.05 0.01] | 0.124944183 | 0.357416198 |
| His | Bilirubin_pPS | 4249 | 0.00661729 | [-0.02 0.04] | 0.666750599 | 0.827171796 |
| His | Proinsulin_pPS | 4249 | -0.002543476 | [-0.03 0.03] | 0.868541017 | 0.940757973 |
| His | Hyper_Insulin_pPS | 4249 | 0.007991535 | [-0.02 0.04] | 0.60303446 | 0.791256366 |
| His | Beta_Cell_1_pPS | 4249 | 0.013253845 | [-0.02 0.04] | 0.388409248 | 0.657347349 |
| His | Beta_Cell_2_pPS | 4249 | 0.019355058 | [-0.01 0.05] | 0.207809962 | 0.486940844 |
| His | Total_GRS | 4249 | 0.012659201 | [-0.02 0.04] | 0.41004999 | 0.672444364 |
| IDL_C | Cholesterol_pPS | 4255 | -0.148062327 | [-0.18 -0.12] | 3.16E-22 | 2.95E-20 |
| IDL_C | Lipodystrophy_1_pPS | 4255 | 0.035092257 | [0.01 0.07] | 0.022261223 | 0.09344486 |
| IDL_C | Lipodystrophy_2_pPS | 4255 | -0.050995063 | [-0.08 -0.02] | 0.000890818 | 0.007736053 |
| IDL_C | Liver-Lipid_pPS | 4255 | 0.01021588 | [-0.02 0.04] | 0.505880061 | 0.729504373 |
| IDL_C | ASAT_Pos_pPS | 4255 | -0.015906045 | [-0.05 0.01] | 0.300270617 | 0.586594238 |
| IDL_C | ALP_Neg_pPS | 4255 | 0.05029172 | [0.02 0.08] | 0.001049066 | 0.008755823 |
| IDL_C | SHBG-LpA_pPS | 4255 | 0.017754681 | [-0.01 0.05] | 0.247574679 | 0.529459308 |
| IDL_C | Obesity_pPS | 4255 | -0.01042012 | [-0.04 0.02] | 0.497411933 | 0.724295386 |
| IDL_C | VAT_Neg_pPS | 4255 | 0.011674188 | [-0.02 0.04] | 0.447111861 | 0.693159645 |
| IDL_C | Bilirubin_pPS | 4255 | -0.012814422 | [-0.04 0.02] | 0.40400032 | 0.668142396 |
| IDL_C | Proinsulin_pPS | 4255 | -0.005484084 | [-0.04 0.02] | 0.721000911 | 0.866412859 |
| IDL_C | Hyper_Insulin_pPS | 4255 | -0.020578184 | [-0.05 0.01] | 0.180188393 | 0.450470983 |
| IDL_C | Beta_Cell_1_pPS | 4255 | 0.017042976 | [-0.01 0.05] | 0.267040286 | 0.552363948 |
| IDL_C | Beta_Cell_2_pPS | 4255 | 0.024873359 | [-0.01 0.05] | 0.105236165 | 0.314239045 |
| IDL_C | Total_GRS | 4255 | -0.006283219 | [-0.04 0.02] | 0.682420964 | 0.838849838 |
| IDL_CE | Cholesterol_pPS | 4255 | -0.149788801 | [-0.18 -0.12] | 1.02E-22 | 1.16E-20 |
| IDL_CE | Lipodystrophy_1_pPS | 4255 | 0.039166659 | [0.01 0.07] | 0.010726727 | 0.049802661 |
| IDL_CE | Lipodystrophy_2_pPS | 4255 | -0.051025729 | [-0.08 -0.02] | 0.00088445 | 0.007711973 |
| IDL_CE | Liver-Lipid_pPS | 4255 | 0.005104006 | [-0.02 0.04] | 0.739609453 | 0.874675307 |
| IDL_CE | ASAT_Pos_pPS | 4255 | -0.011992319 | [-0.04 0.02] | 0.434827557 | 0.690341841 |
| IDL_CE | ALP_Neg_pPS | 4255 | 0.049736967 | [0.02 0.08] | 0.001191808 | 0.009574639 |
| IDL_CE | SHBG-LpA_pPS | 4255 | 0.018373355 | [-0.01 0.05] | 0.231478984 | 0.515064752 |
| IDL_CE | Obesity_pPS | 4255 | -0.008341297 | [-0.04 0.02] | 0.587000812 | 0.779730867 |
| IDL_CE | VAT_Neg_pPS | 4255 | 0.011793002 | [-0.02 0.04] | 0.442501053 | 0.692820992 |
| IDL_CE | Bilirubin_pPS | 4255 | -0.014264395 | [-0.04 0.02] | 0.35292425 | 0.633491645 |
| IDL_CE | Proinsulin_pPS | 4255 | -0.004784433 | [-0.03 0.03] | 0.755375024 | 0.883949497 |
| IDL_CE | Hyper_Insulin_pPS | 4255 | -0.019652923 | [-0.05 0.01] | 0.200578658 | 0.475930554 |
| IDL_CE | Beta_Cell_1_pPS | 4255 | 0.018150946 | [-0.01 0.05] | 0.237177515 | 0.5196586 |
| IDL_CE | Beta_Cell_2_pPS | 4255 | 0.023440037 | [-0.01 0.05] | 0.126859429 | 0.359462978 |
| IDL_CE | Total_GRS | 4255 | -0.004032909 | [-0.03 0.03] | 0.792843048 | 0.90169386 |
| IDL_FC | Cholesterol_pPS | 4255 | -0.140306698 | [-0.17 -0.11] | 4.23E-20 | 3.13E-18 |
| IDL_FC | Lipodystrophy_1_pPS | 4255 | 0.02411092 | [-0.01 0.05] | 0.116342968 | 0.339070199 |
| IDL_FC | Lipodystrophy_2_pPS | 4255 | -0.050102785 | [-0.08 -0.02] | 0.001095802 | 0.009017049 |
| IDL_FC | Liver-Lipid_pPS | 4255 | 0.023537094 | [-0.01 0.05] | 0.125293791 | 0.357416198 |
| IDL_FC | ASAT_Pos_pPS | 4255 | -0.024427933 | [-0.05 0.01] | 0.111617924 | 0.327524552 |
| IDL_FC | ALP_Neg_pPS | 4255 | 0.051076766 | [0.02 0.08] | 0.000873945 | 0.007682834 |
| IDL_FC | SHBG-LpA_pPS | 4255 | 0.015768488 | [-0.01 0.05] | 0.304470417 | 0.590496424 |
| IDL_FC | Obesity_pPS | 4255 | -0.015964987 | [-0.05 0.01] | 0.298482922 | 0.584165937 |
| IDL_FC | VAT_Neg_pPS | 4255 | 0.009647316 | [-0.02 0.04] | 0.529847156 | 0.744284315 |
| IDL_FC | Bilirubin_pPS | 4255 | -0.010888667 | [-0.04 0.02] | 0.478273951 | 0.714911237 |
| IDL_FC | Proinsulin_pPS | 4255 | -0.006125618 | [-0.04 0.02] | 0.689967929 | 0.845220564 |
| IDL_FC | Hyper_Insulin_pPS | 4255 | -0.021485833 | [-0.05 0.01] | 0.161723003 | 0.420479807 |
| IDL_FC | Beta_Cell_1_pPS | 4255 | 0.014342082 | [-0.02 0.04] | 0.350308118 | 0.629849885 |
| IDL_FC | Beta_Cell_2_pPS | 4255 | 0.027190494 | [-0. 0.06] | 0.076569262 | 0.24404319 |
| IDL_FC | Total_GRS | 4255 | -0.011709492 | [-0.04 0.02] | 0.445738983 | 0.693159645 |
| IDL_L | Cholesterol_pPS | 4254 | -0.149104789 | [-0.18 -0.12] | 1.62E-22 | 1.66E-20 |
| IDL_L | Lipodystrophy_1_pPS | 4254 | 0.040641188 | [0.01 0.07] | 0.008113732 | 0.04019389 |
| IDL_L | Lipodystrophy_2_pPS | 4254 | -0.047341052 | [-0.08 -0.02] | 0.002041186 | 0.0142154 |
| IDL_L | Liver-Lipid_pPS | 4254 | 0.008270671 | [-0.02 0.04] | 0.590214967 | 0.781971034 |
| IDL_L | ASAT_Pos_pPS | 4254 | -0.017159812 | [-0.05 0.01] | 0.263830852 | 0.550503091 |
| IDL_L | ALP_Neg_pPS | 4254 | 0.05220726 | [0.02 0.08] | 0.000669902 | 0.006472703 |
| IDL_L | SHBG-LpA_pPS | 4254 | 0.018177167 | [-0.01 0.05] | 0.236555861 | 0.519357545 |
| IDL_L | Obesity_pPS | 4254 | -0.00797603 | [-0.04 0.02] | 0.603524284 | 0.791295592 |
| IDL_L | VAT_Neg_pPS | 4254 | 0.011977583 | [-0.02 0.04] | 0.435446392 | 0.690341841 |
| IDL_L | Bilirubin_pPS | 4254 | -0.012081448 | [-0.04 0.02] | 0.431475521 | 0.688963257 |
| IDL_L | Proinsulin_pPS | 4254 | -0.003055421 | [-0.03 0.03] | 0.842307057 | 0.927489034 |
| IDL_L | Hyper_Insulin_pPS | 4254 | -0.019034587 | [-0.05 0.01] | 0.215167156 | 0.496093758 |
| IDL_L | Beta_Cell_1_pPS | 4254 | 0.015249073 | [-0.01 0.05] | 0.320735833 | 0.608204326 |
| IDL_L | Beta_Cell_2_pPS | 4254 | 0.024964057 | [-0.01 0.06] | 0.104013672 | 0.312041015 |
| IDL_L | Total_GRS | 4254 | -0.002400769 | [-0.03 0.03] | 0.875782565 | 0.943048997 |
| IDL_P | Cholesterol_pPS | 4254 | -0.148350427 | [-0.18 -0.12] | 2.65E-22 | 2.59E-20 |
| IDL_P | Lipodystrophy_1_pPS | 4254 | 0.043673381 | [0.01 0.07] | 0.004441222 | 0.024873164 |
| IDL_P | Lipodystrophy_2_pPS | 4254 | -0.046455832 | [-0.08 -0.02] | 0.002474517 | 0.016483975 |
| IDL_P | Liver-Lipid_pPS | 4254 | 0.006979655 | [-0.02 0.04] | 0.649497107 | 0.818487176 |
| IDL_P | ASAT_Pos_pPS | 4254 | -0.015557313 | [-0.05 0.01] | 0.311050403 | 0.598925597 |
| IDL_P | ALP_Neg_pPS | 4254 | 0.052648352 | [0.02 0.08] | 0.000602716 | 0.005958484 |
| IDL_P | SHBG-LpA_pPS | 4254 | 0.018017468 | [-0.01 0.05] | 0.240700158 | 0.523632697 |
| IDL_P | Obesity_pPS | 4254 | -0.005650822 | [-0.04 0.02] | 0.712920876 | 0.86291356 |
| IDL_P | VAT_Neg_pPS | 4254 | 0.012717752 | [-0.02 0.04] | 0.407611213 | 0.671010017 |
| IDL_P | Bilirubin_pPS | 4254 | -0.011517082 | [-0.04 0.02] | 0.453303664 | 0.696017437 |
| IDL_P | Proinsulin_pPS | 4254 | -0.002253036 | [-0.03 0.03] | 0.883369901 | 0.945095197 |
| IDL_P | Hyper_Insulin_pPS | 4254 | -0.018228132 | [-0.05 0.01] | 0.235243967 | 0.518179785 |
| IDL_P | Beta_Cell_1_pPS | 4254 | 0.015136407 | [-0.01 0.05] | 0.324324633 | 0.608687153 |
| IDL_P | Beta_Cell_2_pPS | 4254 | 0.023685433 | [-0.01 0.05] | 0.122974208 | 0.354065765 |
| IDL_P | Total_GRS | 4254 | -0.000673186 | [-0.03 0.03] | 0.965038246 | 0.982019059 |
| IDL_PL | Cholesterol_pPS | 4255 | -0.149625475 | [-0.18 -0.12] | 1.14E-22 | 1.22E-20 |

**b) Partial Correlation**

| Metabolomics | PPS | Samp | r | CI95% | p | fdr_p_adjusted |
|---|---|---|---|---|---|---|
| IDL_C | Lipodystrophy_2_pPS | 4255 | -0.046638845 | [-0.08 -0.02] | 0.002412763 | 0.017438093 |
| IDL_C | Liver-Lipid_pPS | 4255 | 0.012782419 | [-0.02 0.04] | 0.405896242 | 0.678602222 |
| IDL_C | ASAT_Pos_pPS | 4255 | -0.018057558 | [-0.05 0.01] | 0.240321252 | 0.53636917 |
| IDL_C | ALP_Neg_pPS | 4255 | 0.053324784 | [0.02 0.08] | 0.000521225 | 0.005640495 |
| IDL_C | SHBG-LpA_pPS | 4255 | 0.015180771 | [-0.01 0.05] | 0.323594167 | 0.620844623 |
| IDL_C | Obesity_pPS | 4255 | 0.004070716 | [-0.03 0.03] | 0.791259161 | 0.899824971 |
| IDL_C | VAT_Neg_pPS | 4255 | 0.009867591 | [-0.02 0.04] | 0.521133714 | 0.773759954 |
| IDL_C | Bilirubin_pPS | 4255 | -0.013654212 | [-0.04 0.02] | 0.374633423 | 0.655035906 |
| IDL_C | Proinsulin_pPS | 4255 | -0.019656275 | [-0.05 0.01] | 0.20119325 | 0.486527589 |
| IDL_C | Hyper_Insulin_pPS | 4255 | -0.021901711 | [-0.05 0.01] | 0.154387673 | 0.428688101 |
| IDL_C | Beta_Cell_1_pPS | 4255 | 0.011119271 | [-0.02 0.04] | 0.469686783 | 0.732061671 |
| IDL_C | Beta_Cell_2_pPS | 4255 | 0.024893932 | [-0.01 0.05] | 0.105482749 | 0.336804566 |
| IDL_CE | Cholesterol_pPS | 4255 | -0.147793314 | [-0.18 -0.12] | 4.35E-22 | 4.35E-20 |
| IDL_CE | Lipodystrophy_1_pPS | 4255 | 0.044342055 | [0.01 0.07] | 0.003920296 | 0.02598819 |
| IDL_CE | Lipodystrophy_2_pPS | 4255 | -0.046937517 | [-0.08 -0.02] | 0.002261706 | 0.016876323 |
| IDL_CE | Liver-Lipid_pPS | 4255 | 0.007539486 | [-0.02 0.04] | 0.62397928 | 0.83335992 |
| IDL_CE | ASAT_Pos_pPS | 4255 | -0.014834456 | [-0.04 0.02] | 0.334756252 | 0.630462857 |
| IDL_CE | ALP_Neg_pPS | 4255 | 0.052823156 | [0.02 0.08] | 0.000588369 | 0.006115 |
| IDL_CE | SHBG-LpA_pPS | 4255 | 0.015498129 | [-0.01 0.05] | 0.313580785 | 0.61536839 |
| IDL_CE | Obesity_pPS | 4255 | 0.006047782 | [-0.02 0.04] | 0.694152781 | 0.858808524 |
| IDL_CE | VAT_Neg_pPS | 4255 | 0.009812773 | [-0.02 0.04] | 0.523451356 | 0.774477051 |
| IDL_CE | Bilirubin_pPS | 4255 | -0.015407558 | [-0.05 0.01] | 0.316417497 | 0.618017395 |
| IDL_CE | Proinsulin_pPS | 4255 | -0.019090088 | [-0.05 0.01] | 0.214481682 | 0.504573828 |
| IDL_CE | Hyper_Insulin_pPS | 4255 | -0.02149689 | [-0.05 0.01] | 0.162151296 | 0.43691372 |
| IDL_CE | Beta_Cell_1_pPS | 4255 | 0.012365856 | [-0.02 0.04] | 0.421368597 | 0.692597028 |
| IDL_CE | Beta_Cell_2_pPS | 4255 | 0.023246705 | [-0.01 0.05] | 0.13061349 | 0.3879647 |
| IDL_FC | Cholesterol_pPS | 4255 | -0.13830503 | [-0.17 -0.11] | 1.63E-19 | 1.12E-17 |
| IDL_FC | Lipodystrophy_1_pPS | 4255 | 0.031304294 | [0. 0.06] | 0.04176241 | 0.160169244 |
| IDL_FC | Lipodystrophy_2_pPS | 4255 | -0.045125786 | [-0.08 -0.02] | 0.003329686 | 0.022673575 |
| IDL_FC | Liver-Lipid_pPS | 4255 | 0.026275491 | [-0. 0.06] | 0.087505209 | 0.291975714 |
| IDL_FC | ASAT_Pos_pPS | 4255 | -0.024764265 | [-0.05 0.01] | 0.107310479 | 0.340519287 |
| IDL_FC | ALP_Neg_pPS | 4255 | 0.05369043 | [0.02 0.08] | 0.000476858 | 0.005245437 |
| IDL_FC | SHBG-LpA_pPS | 4255 | 0.014034621 | [-0.02 0.04] | 0.361471284 | 0.648445797 |
| IDL_FC | Obesity_pPS | 4255 | -0.001769253 | [-0.03 0.03] | 0.908416935 | 0.96224905 |
| IDL_FC | VAT_Neg_pPS | 4255 | 0.008242408 | [-0.02 0.04] | 0.592011014 | 0.817111563 |
| IDL_FC | Bilirubin_pPS | 4255 | -0.010988152 | [-0.04 0.02] | 0.474941103 | 0.735233474 |
| IDL_FC | Proinsulin_pPS | 4255 | -0.019610694 | [-0.05 0.01] | 0.202240298 | 0.487833345 |
| IDL_FC | Hyper_Insulin_pPS | 4255 | -0.021363181 | [-0.05 0.01] | 0.164779218 | 0.441248115 |
| IDL_FC | Beta_Cell_1_pPS | 4255 | 0.008306386 | [-0.02 0.04] | 0.589139034 | 0.815104592 |
| IDL_FC | Beta_Cell_2_pPS | 4255 | 0.027550622 | [-0. 0.06] | 0.073187407 | 0.250892446 |
| IDL_L | Cholesterol_pPS | 4254 | -0.14757808 | [-0.18 -0.12] | 5.05E-22 | 4.82E-20 |
| IDL_L | Lipodystrophy_1_pPS | 4254 | 0.046652941 | [0.02 0.08] | 0.002408296 | 0.017438093 |
| IDL_L | Lipodystrophy_2_pPS | 4254 | -0.043095844 | [-0.07 -0.01] | 0.005062507 | 0.030805894 |
| IDL_L | Liver-Lipid_pPS | 4254 | 0.010584365 | [-0.02 0.04] | 0.491373988 | 0.751475077 |
| IDL_L | ASAT_Pos_pPS | 4254 | -0.020821377 | [-0.05 0.01] | 0.175807689 | 0.453447086 |
| IDL_L | ALP_Neg_pPS | 4254 | 0.054765188 | [0.02 0.08] | 0.000366581 | 0.004169853 |
| IDL_L | SHBG-LpA_pPS | 4254 | 0.015195552 | [-0.01 0.05] | 0.323180463 | 0.620844623 |
| IDL_L | Obesity_pPS | 4254 | 0.006176955 | [-0.02 0.04] | 0.687995252 | 0.858568103 |
| IDL_L | VAT_Neg_pPS | 4254 | 0.009805764 | [-0.02 0.04] | 0.523797166 | 0.774477051 |
| IDL_L | Bilirubin_pPS | 4254 | -0.012996229 | [-0.04 0.02] | 0.398144049 | 0.673455453 |
| IDL_L | Proinsulin_pPS | 4254 | -0.017214442 | [-0.05 0.01] | 0.263046188 | 0.559637055 |
| IDL_L | Hyper_Insulin_pPS | 4254 | -0.021081695 | [-0.05 0.01] | 0.170466714 | 0.447866617 |
| IDL_L | Beta_Cell_1_pPS | 4254 | 0.008554213 | [-0.02 0.04] | 0.578120135 | 0.804865443 |
| IDL_L | Beta_Cell_2_pPS | 4254 | 0.025232072 | [-0. 0.06] | 0.100872796 | 0.327836589 |
| IDL_P | Cholesterol_pPS | 4254 | -0.147002256 | [-0.18 -0.12] | 7.32E-22 | 6.66E-20 |
| IDL_P | Lipodystrophy_1_pPS | 4254 | 0.049327257 | [0.02 0.08] | 0.001332929 | 0.011026957 |
| IDL_P | Lipodystrophy_2_pPS | 4254 | -0.042528202 | [-0.07 -0.01] | 0.005673545 | 0.034034385 |
| IDL_P | Liver-Lipid_pPS | 4254 | 0.009183598 | [-0.02 0.04] | 0.55047238 | 0.792838637 |
| IDL_P | ASAT_Pos_pPS | 4254 | -0.019863832 | [-0.05 0.01] | 0.196528387 | 0.482168909 |
| IDL_P | ALP_Neg_pPS | 4254 | 0.055019958 | [0.02 0.09] | 0.000344063 | 0.003970654 |
| IDL_P | SHBG-LpA_pPS | 4254 | 0.01479852 | [-0.02 0.04] | 0.335985702 | 0.63058718 |
| IDL_P | Obesity_pPS | 4254 | 0.008229603 | [-0.02 0.04] | 0.592630364 | 0.817111563 |
| IDL_P | VAT_Neg_pPS | 4254 | 0.01051331 | [-0.02 0.04] | 0.494287597 | 0.753425828 |
| IDL_P | Bilirubin_pPS | 4254 | -0.012542019 | [-0.04 0.02] | 0.414838817 | 0.68636968 |
| IDL_P | Proinsulin_pPS | 4254 | -0.016459063 | [-0.05 0.01] | 0.284573644 | 0.583442529 |
| IDL_P | Hyper_Insulin_pPS | 4254 | -0.020831561 | [-0.05 0.01] | 0.175596425 | 0.453447086 |
| IDL_P | Beta_Cell_1_pPS | 4254 | 0.008405039 | [-0.02 0.04] | 0.584767536 | 0.811861724 |
| IDL_P | Beta_Cell_2_pPS | 4254 | 0.023816435 | [-0.01 0.05] | 0.121486882 | 0.368590185 |
| IDL_PL | Cholesterol_pPS | 4255 | -0.147936875 | [-0.18 -0.12] | 3.97E-22 | 4.18E-20 |
| IDL_PL | Lipodystrophy_1_pPS | 4255 | 0.041808424 | [0.01 0.07] | 0.00653727 | 0.038267876 |
| IDL_PL | Lipodystrophy_2_pPS | 4255 | -0.043818095 | [-0.07 -0.01] | 0.004366597 | 0.028368318 |
| IDL_PL | Liver-Lipid_pPS | 4255 | 0.015037137 | [-0.02 0.05] | 0.328193879 | 0.626352856 |
| IDL_PL | ASAT_Pos_pPS | 4255 | -0.019928909 | [-0.05 0.01] | 0.19501305 | 0.47962669 |
| IDL_PL | ALP_Neg_pPS | 4255 | 0.054657737 | [0.02 0.08] | 0.000375882 | 0.004251504 |
| IDL_PL | SHBG-LpA_pPS | 4255 | 0.014751699 | [-0.02 0.04] | 0.337459877 | 0.631792226 |
| IDL_PL | Obesity_pPS | 4255 | 0.004836153 | [-0.03 0.03] | 0.753183233 | 0.878099599 |
| IDL_PL | VAT_Neg_pPS | 4255 | 0.008824423 | [-0.02 0.04] | 0.566124581 | 0.797755947 |
| IDL_PL | Bilirubin_pPS | 4255 | -0.011737424 | [-0.04 0.02] | 0.445353341 | 0.708330803 |
| IDL_PL | Proinsulin_pPS | 4255 | -0.018567524 | [-0.05 0.01] | 0.227297326 | 0.522130731 |
| IDL_PL | Hyper_Insulin_pPS | 4255 | -0.02170015 | [-0.05 0.01] | 0.158217159 | 0.431435756 |
| IDL_PL | Beta_Cell_1_pPS | 4255 | 0.009514662 | [-0.02 0.04] | 0.536146108 | 0.781765847 |
| IDL_PL | Beta_Cell_2_pPS | 4255 | 0.024328119 | [-0.01 0.05] | 0.113642797 | 0.351642782 |
| IDL_TG | Cholesterol_pPS | 4255 | -0.101425728 | [-0.13 -0.07] | 3.80E-11 | 1.46E-09 |
| IDL_TG | Lipodystrophy_1_pPS | 4255 | 0.072598864 | [0.04 0.1 ] | 2.28E-06 | 4.81E-05 |
| IDL_TG | Lipodystrophy_2_pPS | 4255 | 0.001624959 | [-0.03 0.03] | 0.915857146 | 0.966041099 |
| IDL_TG | Liver-Lipid_pPS | 4255 | -0.016714773 | [-0.05 0.01] | 0.277099752 | 0.57509971 |
| IDL_TG | ASAT_Pos_pPS | 4255 | -0.01001825 | [-0.04 0.02] | 0.514791394 | 0.769688104 |
| IDL_TG | ALP_Neg_pPS | 4255 | 0.042288212 | [0.01 0.07] | 0.005945404 | 0.035319581 |
| IDL_TG | SHBG-LpA_pPS | 4255 | 0.001096745 | [-0.03 0.03] | 0.943151417 | 0.978747744 |
| IDL_TG | Obesity_pPS | 4255 | 0.017236588 | [-0.01 0.05] | 0.262376027 | 0.559637055 |
| IDL_TG | VAT_Neg_pPS | 4255 | 0.000672234 | [-0.03 0.03] | 0.965137031 | 0.985818539 |
| IDL_TG | Bilirubin_pPS | 4255 | -0.006256572 | [-0.04 0.02] | 0.68415381 | 0.858568103 |
| IDL_TG | Proinsulin_pPS | 4255 | 0.006245054 | [-0.02 0.04] | 0.684703988 | 0.858568103 |
| IDL_TG | Hyper_Insulin_pPS | 4255 | -0.006905156 | [-0.04 0.02] | 0.65345095 | 0.844628798 |

**Table S9: Associations between pPSs and global metabolomic profile in SCAPIS**

**a) Spearman Correlation**

| Metabolomics | PPS | Samp | r | CI95% | p | fdr_p_adjusted |
|---|---|---|---|---|---|---|
| IDL_PL | Lipodystrophy_1_pPS | 4255 | 0.035842322 | [0.01 0.07] | 0.019555404 | 0.083725234 |
| IDL_PL | Lipodystrophy_2_pPS | 4255 | -0.048359062 | [-0.08 -0.02] | 0.001627442 | 0.012291769 |
| IDL_PL | Liver-Lipid_pPS | 4255 | 0.012457536 | [-0.02 0.04] | 0.4172191 | 0.678719122 |
| IDL_PL | ASAT_Pos_pPS | 4255 | -0.017294327 | [-0.05 0.01] | 0.260048864 | 0.546250184 |
| IDL_PL | ALP_Neg_pPS | 4255 | 0.051961717 | [0.02 0.08] | 0.000709223 | 0.006701686 |
| IDL_PL | SHBG-LpA_pPS | 4255 | 0.017333347 | [-0.01 0.05] | 0.258974973 | 0.544609134 |
| IDL_PL | Obesity_pPS | 4255 | -0.009697828 | [-0.04 0.02] | 0.527694773 | 0.743176087 |
| IDL_PL | VAT_Neg_pPS | 4255 | 0.010870659 | [-0.02 0.04] | 0.479001954 | 0.71550083 |
| IDL_PL | Bilirubin_pPS | 4255 | -0.011038099 | [-0.04 0.02] | 0.472256027 | 0.710023653 |
| IDL_PL | Proinsulin_pPS | 4255 | -0.004592643 | [-0.03 0.03] | 0.764886285 | 0.889697961 |
| IDL_PL | Hyper_Insulin_pPS | 4255 | -0.020227322 | [-0.05 0.01] | 0.187731305 | 0.459363377 |
| IDL_PL | Beta_Cell_1_pPS | 4255 | 0.015476113 | [-0.01 0.05] | 0.313525879 | 0.602610224 |
| IDL_PL | Beta_Cell_2_pPS | 4255 | 0.024133781 | [-0.01 0.05] | 0.115997042 | 0.33852198 |
| IDL_PL | Total_GRS | 4255 | -0.00535174 | [-0.04 0.02] | 0.727462361 | 0.870249668 |
| IDL_TG | Cholesterol_pPS | 4255 | -0.099760962 | [-0.13 -0.07] | 7.39E-11 | 2.99E-09 |
| IDL_TG | Lipodystrophy_1_pPS | 4255 | 0.072372473 | [0.04 0.1 ] | 2.37E-06 | 5.35E-05 |
| IDL_TG | Lipodystrophy_2_pPS | 4255 | 0.000878882 | [-0.03 0.03] | 0.954360379 | 0.977573158 |
| IDL_TG | Liver-Lipid_pPS | 4255 | -0.016426111 | [-0.05 0.01] | 0.284742631 | 0.573495723 |
| IDL_TG | ASAT_Pos_pPS | 4255 | 0.003137167 | [-0.03 0.03] | 0.838126815 | 0.926178923 |
| IDL_TG | ALP_Neg_pPS | 4255 | 0.044649429 | [0.01 0.07] | 0.003626074 | 0.021420079 |
| IDL_TG | SHBG-LpA_pPS | 4255 | 0.006615024 | [-0.02 0.04] | 0.66663613 | 0.827171796 |
| IDL_TG | Obesity_pPS | 4255 | 0.011610772 | [-0.02 0.04] | 0.449584004 | 0.694281993 |
| IDL_TG | VAT_Neg_pPS | 4255 | 0.005441961 | [-0.02 0.04] | 0.723055371 | 0.867909218 |
| IDL_TG | Bilirubin_pPS | 4255 | -0.004525063 | [-0.03 0.03] | 0.768246242 | 0.889703209 |
| IDL_TG | Proinsulin_pPS | 4255 | 0.012907662 | [-0.02 0.04] | 0.400588526 | 0.666677506 |
| IDL_TG | Hyper_Insulin_pPS | 4255 | 0.000968746 | [-0.03 0.03] | 0.949699713 | 0.97525391 |
| IDL_TG | Beta_Cell_1_pPS | 4255 | -0.000961846 | [-0.03 0.03] | 0.950057498 | 0.97525391 |
| IDL_TG | Beta_Cell_2_pPS | 4255 | 0.006643997 | [-0.02 0.04] | 0.665264691 | 0.826370357 |
| IDL_TG | Total_GRS | 4255 | 0.027059414 | [-0. 0.06] | 0.078000397 | 0.247501258 |
| Ile | Cholesterol_pPS | 4255 | -0.024743237 | [-0.05 0.01] | 0.107069792 | 0.317216442 |
| Ile | Lipodystrophy_1_pPS | 4255 | 0.051229695 | [0.02 0.08] | 0.000843155 | 0.0075357 |
| Ile | Lipodystrophy_2_pPS | 4255 | 0.011942989 | [-0.02 0.04] | 0.436719551 | 0.690884659 |
| Ile | Liver-Lipid_pPS | 4255 | -0.030985929 | [-0.06 -0. ] | 0.043564081 | 0.158113289 |
| Ile | ASAT_Pos_pPS | 4255 | 0.016917703 | [-0.01 0.05] | 0.270572644 | 0.554396235 |
| Ile | ALP_Neg_pPS | 4255 | 0.004402637 | [-0.03 0.03] | 0.774344147 | 0.891684366 |
| Ile | SHBG-LpA_pPS | 4255 | -0.008407358 | [-0.04 0.02] | 0.584042505 | 0.778602346 |
| Ile | Obesity_pPS | 4255 | 0.035519781 | [0.01 0.07] | 0.020681605 | 0.087672021 |
| Ile | VAT_Neg_pPS | 4255 | 0.017706667 | [-0.01 0.05] | 0.248855798 | 0.53033113 |
| Ile | Bilirubin_pPS | 4255 | -0.0058016 | [-0.04 0.02] | 0.705580431 | 0.857976205 |
| Ile | Proinsulin_pPS | 4255 | -0.004095973 | [-0.03 0.03] | 0.789679152 | 0.899856387 |
| Ile | Hyper_Insulin_pPS | 4255 | 0.01860347 | [-0.01 0.05] | 0.225686034 | 0.506907374 |
| Ile | Beta_Cell_1_pPS | 4255 | 0.030364456 | [0. 0.06] | 0.047955481 | 0.169170784 |
| Ile | Beta_Cell_2_pPS | 4255 | -0.001830335 | [-0.03 0.03] | 0.905124888 | 0.952872131 |
| Ile | Total_GRS | 4255 | 0.044171519 | [0.01 0.07] | 0.004004398 | 0.023089876 |
| L_HDL_C | Cholesterol_pPS | 4255 | 0.055524478 | [0.03 0.09] | 0.000296319 | 0.003327774 |
| L_HDL_C | Lipodystrophy_1_pPS | 4255 | -0.071622361 | [-0.1 -0.04] | 3.01E-06 | 6.53E-05 |
| L_HDL_C | Lipodystrophy_2_pPS | 4255 | -0.035381259 | [-0.07 -0.01] | 0.021182367 | 0.089617707 |
| L_HDL_C | Liver-Lipid_pPS | 4255 | 0.008606588 | [-0.02 0.04] | 0.575163097 | 0.77308819 |
| L_HDL_C | ASAT_Pos_pPS | 4255 | -0.056000325 | [-0.09 -0.03] | 0.000262694 | 0.003013255 |
| L_HDL_C | ALP_Neg_pPS | 4255 | 0.012069568 | [-0.02 0.04] | 0.431874336 | 0.688963257 |
| L_HDL_C | SHBG-LpA_pPS | 4255 | 0.005129822 | [-0.02 0.04] | 0.738340529 | 0.874278301 |
| L_HDL_C | Obesity_pPS | 4255 | -0.032365682 | [-0.06 -0. ] | 0.035015678 | 0.133171327 |
| L_HDL_C | VAT_Neg_pPS | 4255 | -0.023773298 | [-0.05 0.01] | 0.121546454 | 0.350897905 |
| L_HDL_C | Bilirubin_pPS | 4255 | -0.002821185 | [-0.03 0.03] | 0.854238254 | 0.932572076 |
| L_HDL_C | Proinsulin_pPS | 4255 | -0.00470839 | [-0.03 0.03] | 0.759141844 | 0.886905913 |
| L_HDL_C | Hyper_Insulin_pPS | 4255 | -0.013993236 | [-0.04 0.02] | 0.362152146 | 0.639224774 |
| L_HDL_C | Beta_Cell_1_pPS | 4255 | -0.00701894 | [-0.04 0.02] | 0.647618675 | 0.818104863 |
| L_HDL_C | Beta_Cell_2_pPS | 4255 | 0.006856314 | [-0.02 0.04] | 0.655248829 | 0.821454552 |
| L_HDL_C | Total_GRS | 4255 | -0.045534243 | [-0.08 -0.02] | 0.003010257 | 0.019047198 |
| L_HDL_CE | Cholesterol_pPS | 4255 | 0.056135496 | [0.03 0.09] | 0.000253816 | 0.002927066 |
| L_HDL_CE | Lipodystrophy_1_pPS | 4255 | -0.071322845 | [-0.1 -0.04] | 3.31E-06 | 6.97E-05 |
| L_HDL_CE | Lipodystrophy_2_pPS | 4255 | -0.035343775 | [-0.07 -0.01] | 0.021319678 | 0.09002108 |
| L_HDL_CE | Liver-Lipid_pPS | 4255 | 0.007316955 | [-0.02 0.04] | 0.633732424 | 0.807999529 |
| L_HDL_CE | ASAT_Pos_pPS | 4255 | -0.055562027 | [-0.09 -0.03] | 0.000293526 | 0.003313754 |
| L_HDL_CE | ALP_Neg_pPS | 4255 | 0.01205767 | [-0.02 0.04] | 0.432328459 | 0.688963257 |
| L_HDL_CE | SHBG-LpA_pPS | 4255 | 0.005138501 | [-0.02 0.04] | 0.737914116 | 0.874278301 |
| L_HDL_CE | Obesity_pPS | 4255 | -0.032127265 | [-0.06 -0. ] | 0.036381356 | 0.137183756 |
| L_HDL_CE | VAT_Neg_pPS | 4255 | -0.023957447 | [-0.05 0.01] | 0.118686201 | 0.344961924 |
| L_HDL_CE | Bilirubin_pPS | 4255 | -0.002695545 | [-0.03 0.03] | 0.860661751 | 0.93707304 |
| L_HDL_CE | Proinsulin_pPS | 4255 | -0.004400395 | [-0.03 0.03] | 0.774455932 | 0.891684366 |
| L_HDL_CE | Hyper_Insulin_pPS | 4255 | -0.013745052 | [-0.04 0.02] | 0.370729375 | 0.643918869 |
| L_HDL_CE | Beta_Cell_1_pPS | 4255 | -0.006933431 | [-0.04 0.02] | 0.651626092 | 0.819307132 |
| L_HDL_CE | Beta_Cell_2_pPS | 4255 | 0.006671083 | [-0.02 0.04] | 0.663983571 | 0.825654931 |
| L_HDL_CE | Total_GRS | 4255 | -0.045147465 | [-0.08 -0.02] | 0.003266658 | 0.019849806 |
| L_HDL_FC | Cholesterol_pPS | 4255 | 0.053383294 | [0.02 0.08] | 0.000503751 | 0.005194934 |
| L_HDL_FC | Lipodystrophy_1_pPS | 4255 | -0.072257488 | [-0.1 -0.04] | 2.46E-06 | 5.49E-05 |
| L_HDL_FC | Lipodystrophy_2_pPS | 4255 | -0.035556172 | [-0.07 -0.01] | 0.020551771 | 0.087294156 |
| L_HDL_FC | Liver-Lipid_pPS | 4255 | 0.012530023 | [-0.02 0.04] | 0.41451379 | 0.677692134 |
| L_HDL_FC | ASAT_Pos_pPS | 4255 | -0.057193184 | [-0.09 -0.03] | 0.000193449 | 0.002357661 |
| L_HDL_FC | ALP_Neg_pPS | 4255 | 0.011891369 | [-0.02 0.04] | 0.438704392 | 0.691142319 |
| L_HDL_FC | SHBG-LpA_pPS | 4255 | 0.005323147 | [-0.02 0.04] | 0.728860975 | 0.870493759 |
| L_HDL_FC | Obesity_pPS | 4255 | -0.03331296 | [-0.06 -0. ] | 0.030013444 | 0.118780144 |
| L_HDL_FC | VAT_Neg_pPS | 4255 | -0.023509597 | [-0.05 0.01] | 0.125735802 | 0.358171707 |
| L_HDL_FC | Bilirubin_pPS | 4255 | -0.004003639 | [-0.03 0.03] | 0.794312682 | 0.901959081 |
| L_HDL_FC | Proinsulin_pPS | 4255 | -0.005784016 | [-0.04 0.02] | 0.706431361 | 0.858243888 |
| L_HDL_FC | Hyper_Insulin_pPS | 4255 | -0.014652486 | [-0.04 0.02] | 0.33997824 | 0.622761166 |
| L_HDL_FC | Beta_Cell_1_pPS | 4255 | -0.007138534 | [-0.04 0.02] | 0.64203102 | 0.81440363 |
| L_HDL_FC | Beta_Cell_2_pPS | 4255 | 0.007704058 | [-0.02 0.04] | 0.615886341 | 0.798233354 |
| L_HDL_FC | Total_GRS | 4255 | -0.046578465 | [-0.08 -0.02] | 0.002406985 | 0.016084061 |
| L_HDL_L | Cholesterol_pPS | 4255 | 0.050883413 | [0.02 0.08] | 0.000914365 | 0.007854666 |
| L_HDL_L | Lipodystrophy_1_pPS | 4255 | -0.070207671 | [-0.1 -0.04] | 4.71E-06 | 9.43E-05 |
| L_HDL_L | Lipodystrophy_2_pPS | 4255 | -0.038226556 | [-0.07 -0.01] | 0.012767487 | 0.058021734 |
| L_HDL_L | Liver-Lipid_pPS | 4255 | 0.005619349 | [-0.02 0.04] | 0.714417321 | 0.86291356 |
| L_HDL_L | ASAT_Pos_pPS | 4255 | -0.056456333 | [-0.09 -0.03] | 0.000233855 | 0.002735646 |

**b) Partial Correlation**

| Metabolomics | PPS | Samp | r | CI95% | p | fdr_p_adjusted |
|---|---|---|---|---|---|---|
| IDL_TG | Beta_Cell_1_pPS | 4255 | -0.00723896 | [-0.04 0.02] | 0.637870893 | 0.835456904 |
| IDL_TG | Beta_Cell_2_pPS | 4255 | 0.007641389 | [-0.02 0.04] | 0.619298757 | 0.830432761 |
| Ile | Cholesterol_pPS | 4255 | -0.027048126 | [-0.06 0. ] | 0.078582589 | 0.267100753 |
| Ile | Lipodystrophy_1_pPS | 4255 | 0.045914017 | [0.02 0.08] | 0.002818504 | 0.01988601 |
| Ile | Lipodystrophy_2_pPS | 4255 | 0.009677362 | [-0.02 0.04] | 0.529198914 | 0.778529119 |
| Ile | Liver-Lipid_pPS | 4255 | -0.032940049 | [-0.06 -0. ] | 0.03216791 | 0.12896057 |
| Ile | ASAT_Pos_pPS | 4255 | 0.004530685 | [-0.03 0.03] | 0.768312345 | 0.886548308 |
| Ile | ALP_Neg_pPS | 4255 | 0.006231811 | [-0.02 0.04] | 0.685336766 | 0.858568103 |
| Ile | SHBG-LpA_pPS | 4255 | -0.012261471 | [-0.04 0.02] | 0.425299265 | 0.693893617 |
| Ile | Obesity_pPS | 4255 | 0.036884887 | [0.01 0.07] | 0.016437975 | 0.07895582 |
| Ile | VAT_Neg_pPS | 4255 | 0.011317401 | [-0.02 0.04] | 0.46180845 | 0.725129817 |
| Ile | Bilirubin_pPS | 4255 | -0.008929028 | [-0.04 0.02] | 0.561530229 | 0.797755947 |
| Ile | Proinsulin_pPS | 4255 | -0.016033056 | [-0.05 0.01] | 0.297168849 | 0.594932035 |
| Ile | Hyper_Insulin_pPS | 4255 | 0.010402079 | [-0.02 0.04] | 0.498816264 | 0.754822494 |
| Ile | Beta_Cell_1_pPS | 4255 | 0.032661454 | [0. 0.06] | 0.033654322 | 0.133682445 |
| Ile | Beta_Cell_2_pPS | 4255 | -0.008197413 | [-0.04 0.02] | 0.594034653 | 0.817359021 |
| L_HDL_C | Cholesterol_pPS | 4255 | 0.058894161 | [0.03 0.09] | 0.000126807 | 0.001775291 |
| L_HDL_C | Lipodystrophy_1_pPS | 4255 | -0.061084319 | [-0.09 -0.03] | 7.03E-05 | 0.001099076 |
| L_HDL_C | Lipodystrophy_2_pPS | 4255 | -0.028388262 | [-0.06 0. ] | 0.064869717 | 0.229450836 |
| L_HDL_C | Liver-Lipid_pPS | 4255 | 0.009034566 | [-0.02 0.04] | 0.556913276 | 0.796929123 |
| L_HDL_C | ASAT_Pos_pPS | 4255 | -0.039713423 | [-0.07 -0.01] | 0.00979013 | 0.052972541 |
| L_HDL_C | ALP_Neg_pPS | 4255 | 0.014461238 | [-0.02 0.04] | 0.347059755 | 0.637861136 |
| L_HDL_C | SHBG-LpA_pPS | 4255 | 0.012243671 | [-0.02 0.04] | 0.425971656 | 0.693893617 |
| L_HDL_C | Obesity_pPS | 4255 | -0.033538561 | [-0.06 -0. ] | 0.029163824 | 0.120881936 |
| L_HDL_C | VAT_Neg_pPS | 4255 | -0.016795577 | [-0.05 0.01] | 0.274783672 | 0.573178999 |
| L_HDL_C | Bilirubin_pPS | 4255 | 0.000339304 | [-0.03 0.03] | 0.98239905 | 0.993363084 |
| L_HDL_C | Proinsulin_pPS | 4255 | -0.003963393 | [-0.03 0.03] | 0.796640185 | 0.90258837 |
| L_HDL_C | Hyper_Insulin_pPS | 4255 | 0.000202234 | [-0.03 0.03] | 0.989508864 | 0.993977294 |
| L_HDL_C | Beta_Cell_1_pPS | 4255 | -0.002323203 | [-0.03 0.03] | 0.879933875 | 0.94864169 |
| L_HDL_C | Beta_Cell_2_pPS | 4255 | 0.008969449 | [-0.02 0.04] | 0.559759747 | 0.797755947 |
| L_HDL_CE | Cholesterol_pPS | 4255 | 0.059510912 | [0.03 0.09] | 0.000107598 | 0.001538655 |
| L_HDL_CE | Lipodystrophy_1_pPS | 4255 | -0.060880527 | [-0.09 -0.03] | 7.43E-05 | 0.001135479 |
| L_HDL_CE | Lipodystrophy_2_pPS | 4255 | -0.028373112 | [-0.06 0. ] | 0.065012911 | 0.229551761 |
| L_HDL_CE | Liver-Lipid_pPS | 4255 | 0.007706643 | [-0.02 0.04] | 0.616309579 | 0.828794354 |
| L_HDL_CE | ASAT_Pos_pPS | 4255 | -0.039332205 | [-0.07 -0.01] | 0.010517335 | 0.056342065 |
| L_HDL_CE | ALP_Neg_pPS | 4255 | 0.014438961 | [-0.02 0.04] | 0.347803135 | 0.637861136 |
| L_HDL_CE | SHBG-LpA_pPS | 4255 | 0.012245738 | [-0.02 0.04] | 0.425893537 | 0.693893617 |
| L_HDL_CE | Obesity_pPS | 4255 | -0.0334184 | [-0.06 -0. ] | 0.029746797 | 0.122035016 |
| L_HDL_CE | VAT_Neg_pPS | 4255 | -0.016977588 | [-0.05 0.01] | 0.269615175 | 0.569377194 |
| L_HDL_CE | Bilirubin_pPS | 4255 | 0.00042684 | [-0.03 0.03] | 0.977859283 | 0.991729628 |
| L_HDL_CE | Proinsulin_pPS | 4255 | -0.003651928 | [-0.03 0.03] | 0.812310529 | 0.911348446 |
| L_HDL_CE | Hyper_Insulin_pPS | 4255 | 0.000445298 | [-0.03 0.03] | 0.976902137 | 0.991343605 |
| L_HDL_CE | Beta_Cell_1_pPS | 4255 | -0.002239097 | [-0.03 0.03] | 0.884249392 | 0.951245182 |
| L_HDL_CE | Beta_Cell_2_pPS | 4255 | 0.008752942 | [-0.02 0.04] | 0.569274422 | 0.797755947 |
| L_HDL_FC | Cholesterol_pPS | 4255 | 0.056749561 | [0.03 0.09] | 0.000221808 | 0.00282841 |
| L_HDL_FC | Lipodystrophy_1_pPS | 4255 | -0.061438917 | [-0.09 -0.03] | 6.37E-05 | 0.001020992 |
| L_HDL_FC | Lipodystrophy_2_pPS | 4255 | -0.02847279 | [-0.06 0. ] | 0.064075496 | 0.227445291 |
| L_HDL_FC | Liver-Lipid_pPS | 4255 | 0.013067225 | [-0.02 0.04] | 0.39551571 | 0.673455453 |
| L_HDL_FC | ASAT_Pos_pPS | 4255 | -0.040757395 | [-0.07 -0.01] | 0.00802263 | 0.045499449 |
| L_HDL_FC | ALP_Neg_pPS | 4255 | 0.014344525 | [-0.02 0.04] | 0.350965642 | 0.637861136 |
| L_HDL_FC | SHBG-LpA_pPS | 4255 | 0.012437732 | [-0.02 0.04] | 0.418674472 | 0.691002715 |
| L_HDL_FC | Obesity_pPS | 4255 | -0.034100379 | [-0.06 -0. ] | 0.026566748 | 0.113056469 |
| L_HDL_FC | VAT_Neg_pPS | 4255 | -0.016599164 | [-0.05 0.01] | 0.280436554 | 0.578797919 |
| L_HDL_FC | Bilirubin_pPS | 4255 | -0.000740417 | [-0.03 0.03] | 0.961603577 | 0.984217976 |
| L_HDL_FC | Proinsulin_pPS | 4255 | -0.005067578 | [-0.04 0.03] | 0.741783739 | 0.873045882 |
| L_HDL_FC | Hyper_Insulin_pPS | 4255 | -0.000443729 | [-0.03 0.03] | 0.976983483 | 0.991343605 |
| L_HDL_FC | Beta_Cell_1_pPS | 4255 | -0.00242296 | [-0.03 0.03] | 0.874819885 | 0.946048963 |
| L_HDL_FC | Beta_Cell_2_pPS | 4255 | 0.00993589 | [-0.02 0.04] | 0.518253541 | 0.772556657 |
| L_HDL_L | Cholesterol_pPS | 4255 | 0.054435026 | [0.02 0.08] | 0.000397181 | 0.00441754 |
| L_HDL_L | Lipodystrophy_1_pPS | 4255 | -0.059466502 | [-0.09 -0.03] | 0.000108884 | 0.001545999 |
| L_HDL_L | Lipodystrophy_2_pPS | 4255 | -0.03082718 | [-0.06 -0. ] | 0.044980594 | 0.169587853 |
| L_HDL_L | Liver-Lipid_pPS | 4255 | 0.005994717 | [-0.02 0.04] | 0.696702635 | 0.858963776 |
| L_HDL_L | ASAT_Pos_pPS | 4255 | -0.040192672 | [-0.07 -0.01] | 0.008939665 | 0.049576757 |
| L_HDL_L | ALP_Neg_pPS | 4255 | 0.018521462 | [-0.01 0.05] | 0.228452611 | 0.522699573 |
| L_HDL_L | SHBG-LpA_pPS | 4255 | 0.012282927 | [-0.02 0.04] | 0.424489579 | 0.693893617 |
| L_HDL_L | Obesity_pPS | 4255 | -0.034205797 | [-0.06 -0. ] | 0.026102357 | 0.111899184 |
| L_HDL_L | VAT_Neg_pPS | 4255 | -0.018170375 | [-0.05 0.01] | 0.2373957 | 0.533407623 |
| L_HDL_L | Bilirubin_pPS | 4255 | -0.000307686 | [-0.03 0.03] | 0.984038994 | 0.993363084 |
| L_HDL_L | Proinsulin_pPS | 4255 | -0.003662352 | [-0.03 0.03] | 0.811784843 | 0.911348446 |
| L_HDL_L | Hyper_Insulin_pPS | 4255 | 0.000237462 | [-0.03 0.03] | 0.987681449 | 0.993363084 |
| L_HDL_L | Beta_Cell_1_pPS | 4255 | -0.001211486 | [-0.03 0.03] | 0.937215624 | 0.978170683 |
| L_HDL_L | Beta_Cell_2_pPS | 4255 | 0.009370395 | [-0.02 0.04] | 0.542345984 | 0.787936618 |
| L_HDL_P | Cholesterol_pPS | 4255 | 0.053500782 | [0.02 0.08] | 0.00049941 | 0.00546349 |
| L_HDL_P | Lipodystrophy_1_pPS | 4255 | -0.058965296 | [-0.09 -0.03] | 0.000124437 | 0.00175438 |
| L_HDL_P | Lipodystrophy_2_pPS | 4255 | -0.031310482 | [-0.06 -0. ] | 0.041721984 | 0.160169244 |
| L_HDL_P | Liver-Lipid_pPS | 4255 | 0.005429523 | [-0.02 0.04] | 0.724068927 | 0.866459051 |
| L_HDL_P | ASAT_Pos_pPS | 4255 | -0.04010902 | [-0.07 -0.01] | 0.009083184 | 0.050095139 |
| L_HDL_P | ALP_Neg_pPS | 4255 | 0.019275149 | [-0.01 0.05] | 0.210070447 | 0.4972684 |
| L_HDL_P | SHBG-LpA_pPS | 4255 | 0.012145233 | [-0.02 0.04] | 0.429701342 | 0.696591166 |
| L_HDL_P | Obesity_pPS | 4255 | -0.034309248 | [-0.06 -0. ] | 0.02565347 | 0.110210831 |
| L_HDL_P | VAT_Neg_pPS | 4255 | -0.018203557 | [-0.05 0.01] | 0.236540036 | 0.532559144 |
| L_HDL_P | Bilirubin_pPS | 4255 | -0.000377878 | [-0.03 0.03] | 0.980398495 | 0.992796048 |
| L_HDL_P | Proinsulin_pPS | 4255 | -0.003459462 | [-0.03 0.03] | 0.822032007 | 0.916829013 |
| L_HDL_P | Hyper_Insulin_pPS | 4255 | 0.000260077 | [-0.03 0.03] | 0.986508393 | 0.993363084 |
| L_HDL_P | Beta_Cell_1_pPS | 4255 | -0.001068371 | [-0.03 0.03] | 0.944619758 | 0.978747744 |
| L_HDL_P | Beta_Cell_2_pPS | 4255 | 0.009317548 | [-0.02 0.04] | 0.544625712 | 0.789070018 |
| L_HDL_PL | Cholesterol_pPS | 4255 | 0.052153867 | [0.02 0.08] | 0.000690525 | 0.006776628 |
| L_HDL_PL | Lipodystrophy_1_pPS | 4255 | -0.059812725 | [-0.09 -0.03] | 9.92E-05 | 0.001439582 |
| L_HDL_PL | Lipodystrophy_2_pPS | 4255 | -0.031431275 | [-0.06 -0. ] | 0.040939493 | 0.15824097 |
| L_HDL_PL | Liver-Lipid_pPS | 4255 | 0.002762219 | [-0.03 0.03] | 0.857468429 | 0.937541939 |
| L_HDL_PL | ASAT_Pos_pPS | 4255 | -0.041053288 | [-0.07 -0.01] | 0.007576572 | 0.04364299 |
| L_HDL_PL | ALP_Neg_pPS | 4255 | 0.02071881 | [-0.01 0.05] | 0.177894699 | 0.455834788 |
| L_HDL_PL | SHBG-LpA_pPS | 4255 | 0.012830007 | [-0.02 0.04] | 0.404150547 | 0.677078992 |
| L_HDL_PL | Obesity_pPS | 4255 | -0.033432131 | [-0.06 -0. ] | 0.029679675 | 0.122035016 |

**Table S9: Associations between pPSs and global metabolomic profile in SCAPIS**

**a) Spearman Correlation**

| Metabolomics | PPS | Samp | r | CI95% | p | fdr_p_adjusted |
|---|---|---|---|---|---|---|
| L_HDL_L | ALP_Neg_pPS | 4255 | 0.016105963 | [-0.01 0.05] | 0.294236016 | 0.580088468 |
| L_HDL_L | SHBG-LpA_pPS | 4255 | 0.005131451 | [-0.02 0.04] | 0.738260485 | 0.874278301 |
| L_HDL_L | Obesity_pPS | 4255 | -0.033289497 | [-0.06 -0. ] | 0.030129502 | 0.119019856 |
| L_HDL_L | VAT_Neg_pPS | 4255 | -0.025151704 | [-0.06 0. ] | 0.101397496 | 0.305903838 |
| L_HDL_L | Bilirubin_pPS | 4255 | -0.003357337 | [-0.03 0.03] | 0.826940259 | 0.921447717 |
| L_HDL_L | Proinsulin_pPS | 4255 | -0.003595446 | [-0.03 0.03] | 0.814881714 | 0.914663148 |
| L_HDL_L | Hyper_Insulin_pPS | 4255 | -0.014222889 | [-0.04 0.02] | 0.354327032 | 0.634686582 |
| L_HDL_L | Beta_Cell_1_pPS | 4255 | -0.005765607 | [-0.04 0.02] | 0.707322579 | 0.858243888 |
| L_HDL_L | Beta_Cell_2_pPS | 4255 | 0.007406515 | [-0.02 0.04] | 0.629584046 | 0.803929814 |
| L_HDL_L | Total_GRS | 4255 | -0.045543794 | [-0.08 -0.02] | 0.003004164 | 0.019047198 |
| L_HDL_P | Cholesterol_pPS | 4255 | 0.049914522 | [0.02 0.08] | 0.00114428 | 0.009343882 |
| L_HDL_P | Lipodystrophy_1_pPS | 4255 | -0.069713499 | [-0.1 -0.04] | 5.49E-06 | 0.000106052 |
| L_HDL_P | Lipodystrophy_2_pPS | 4255 | -0.038733428 | [-0.07 -0.01] | 0.011628112 | 0.053524249 |
| L_HDL_P | Liver-Lipid_pPS | 4255 | 0.005065699 | [-0.03 0.04] | 0.741493663 | 0.875828143 |
| L_HDL_P | ASAT_Pos_pPS | 4255 | -0.056326919 | [-0.09 -0.03] | 0.000241723 | 0.002802675 |
| L_HDL_P | ALP_Neg_pPS | 4255 | 0.016889612 | [-0.01 0.05] | 0.271369103 | 0.554396235 |
| L_HDL_P | SHBG-LpA_pPS | 4255 | 0.005021683 | [-0.03 0.04] | 0.743660599 | 0.876709962 |
| L_HDL_P | Obesity_pPS | 4255 | -0.033418905 | [-0.06 -0. ] | 0.029494168 | 0.1180317 |
| L_HDL_P | VAT_Neg_pPS | 4255 | -0.025195048 | [-0.06 0. ] | 0.100809899 | 0.305231058 |
| L_HDL_P | Bilirubin_pPS | 4255 | -0.003395501 | [-0.03 0.03] | 0.825004673 | 0.921344342 |
| L_HDL_P | Proinsulin_pPS | 4255 | -0.003269587 | [-0.03 0.03] | 0.831394648 | 0.922752159 |
| L_HDL_P | Hyper_Insulin_pPS | 4255 | -0.014198119 | [-0.04 0.02] | 0.355165864 | 0.634858982 |
| L_HDL_P | Beta_Cell_1_pPS | 4255 | -0.00554647 | [-0.04 0.02] | 0.717961855 | 0.865184371 |
| L_HDL_P | Beta_Cell_2_pPS | 4255 | 0.007402006 | [-0.02 0.04] | 0.629792632 | 0.803929814 |
| L_HDL_P | Total_GRS | 4255 | -0.045478207 | [-0.08 -0.02] | 0.00304623 | 0.01910574 |
| L_HDL_PL | Cholesterol_pPS | 4255 | 0.048595901 | [0.02 0.08] | 0.00154342 | 0.011854253 |
| L_HDL_PL | Lipodystrophy_1_pPS | 4255 | -0.070730679 | [-0.1 -0.04] | 3.99E-06 | 8.24E-05 |
| L_HDL_PL | Lipodystrophy_2_pPS | 4255 | -0.039525741 | [-0.07 -0.01] | 0.010027264 | 0.047244496 |
| L_HDL_PL | Liver-Lipid_pPS | 4255 | 0.002385788 | [-0.03 0.03] | 0.876537017 | 0.943108963 |
| L_HDL_PL | ASAT_Pos_pPS | 4255 | -0.05741023 | [-0.09 -0.03] | 0.00018286 | 0.00225422 |
| L_HDL_PL | ALP_Neg_pPS | 4255 | 0.018071768 | [-0.01 0.05] | 0.239229922 | 0.521492055 |
| L_HDL_PL | SHBG-LpA_pPS | 4255 | 0.005382141 | [-0.02 0.04] | 0.725976369 | 0.870169158 |
| L_HDL_PL | Obesity_pPS | 4255 | -0.032824138 | [-0.06 -0. ] | 0.032512271 | 0.126338445 |
| L_HDL_PL | VAT_Neg_pPS | 4255 | -0.027623498 | [-0.06 0. ] | 0.071993181 | 0.231869931 |
| L_HDL_PL | Bilirubin_pPS | 4255 | -0.002659032 | [-0.03 0.03] | 0.862530285 | 0.937517799 |
| L_HDL_PL | Proinsulin_pPS | 4255 | -0.003266481 | [-0.03 0.03] | 0.831552412 | 0.922752159 |
| L_HDL_PL | Hyper_Insulin_pPS | 4255 | -0.015164762 | [-0.05 0.01] | 0.323361937 | 0.608687153 |
| L_HDL_PL | Beta_Cell_1_pPS | 4255 | -0.00411226 | [-0.03 0.03] | 0.788862586 | 0.899856387 |
| L_HDL_PL | Beta_Cell_2_pPS | 4255 | 0.00702602 | [-0.02 0.04] | 0.647287329 | 0.818104863 |
| L_HDL_PL | Total_GRS | 4255 | -0.045647754 | [-0.08 -0.02] | 0.002938571 | 0.018703958 |
| L_HDL_TG | Cholesterol_pPS | 4255 | -0.010995885 | [-0.04 0.02] | 0.473951803 | 0.711425205 |
| L_HDL_TG | Lipodystrophy_1_pPS | 4255 | -0.013043859 | [-0.04 0.02] | 0.395636074 | 0.662999515 |
| L_HDL_TG | Lipodystrophy_2_pPS | 4255 | -0.04878574 | [-0.08 -0.02] | 0.001478977 | 0.011411529 |
| L_HDL_TG | Liver-Lipid_pPS | 4255 | -0.003659071 | [-0.03 0.03] | 0.811666806 | 0.912487054 |
| L_HDL_TG | ASAT_Pos_pPS | 4255 | -0.028587213 | [-0.06 0. ] | 0.062609473 | 0.206611259 |
| L_HDL_TG | ALP_Neg_pPS | 4255 | 0.043493436 | [0.01 0.07] | 0.00460276 | 0.025363691 |
| L_HDL_TG | SHBG-LpA_pPS | 4255 | 0.000645847 | [-0.03 0.03] | 0.966453255 | 0.982019059 |
| L_HDL_TG | Obesity_pPS | 4255 | -0.028325078 | [-0.06 0. ] | 0.065055822 | 0.213045402 |
| L_HDL_TG | VAT_Neg_pPS | 4255 | -0.012484988 | [-0.04 0.02] | 0.416193316 | 0.677854717 |
| L_HDL_TG | Bilirubin_pPS | 4255 | -0.011049891 | [-0.04 0.02] | 0.47178292 | 0.710023653 |
| L_HDL_TG | Proinsulin_pPS | 4255 | 0.011186494 | [-0.02 0.04] | 0.466321346 | 0.706538163 |
| L_HDL_TG | Hyper_Insulin_pPS | 4255 | -0.006372305 | [-0.04 0.02] | 0.678168908 | 0.836400502 |
| L_HDL_TG | Beta_Cell_1_pPS | 4255 | -0.008363564 | [-0.04 0.02] | 0.586002859 | 0.779730867 |
| L_HDL_TG | Beta_Cell_2_pPS | 4255 | 0.016255225 | [-0.01 0.05] | 0.289783859 | 0.576610739 |
| L_HDL_TG | Total_GRS | 4255 | -0.026201456 | [-0.06 0. ] | 0.087913399 | 0.272112901 |
| L_LDL_C | Cholesterol_pPS | 4255 | -0.153142605 | [-0.18 -0.12] | 1.11E-23 | 1.74E-21 |
| L_LDL_C | Lipodystrophy_1_pPS | 4255 | 0.037899962 | [0.01 0.07] | 0.013552939 | 0.061073641 |
| L_LDL_C | Lipodystrophy_2_pPS | 4255 | -0.045789019 | [-0.08 -0.02] | 0.002851533 | 0.018367983 |
| L_LDL_C | Liver-Lipid_pPS | 4255 | 0.008209289 | [-0.02 0.04] | 0.592932923 | 0.785087111 |
| L_LDL_C | ASAT_Pos_pPS | 4255 | -0.014176529 | [-0.04 0.02] | 0.355898042 | 0.635109235 |
| L_LDL_C | ALP_Neg_pPS | 4255 | 0.04822373 | [0.02 0.08] | 0.001677321 | 0.01249255 |
| L_LDL_C | SHBG-LpA_pPS | 4255 | 0.017882707 | [-0.01 0.05] | 0.244181227 | 0.525480742 |
| L_LDL_C | Obesity_pPS | 4255 | -0.008524643 | [-0.04 0.02] | 0.578807543 | 0.776133311 |
| L_LDL_C | VAT_Neg_pPS | 4255 | 0.011692995 | [-0.02 0.04] | 0.446380227 | 0.693159645 |
| L_LDL_C | Bilirubin_pPS | 4255 | -0.012953506 | [-0.04 0.02] | 0.398917381 | 0.666036824 |
| L_LDL_C | Proinsulin_pPS | 4255 | -0.006448341 | [-0.04 0.02] | 0.674547793 | 0.833951018 |
| L_LDL_C | Hyper_Insulin_pPS | 4255 | -0.020507503 | [-0.05 0.01] | 0.181689522 | 0.45159215 |
| L_LDL_C | Beta_Cell_1_pPS | 4255 | 0.019413077 | [-0.01 0.05] | 0.206128949 | 0.484809862 |
| L_LDL_C | Beta_Cell_2_pPS | 4255 | 0.023071966 | [-0.01 0.05] | 0.132935466 | 0.371768677 |
| L_LDL_C | Total_GRS | 4255 | -0.003921819 | [-0.03 0.03] | 0.798424689 | 0.905190781 |
| L_LDL_CE | Cholesterol_pPS | 4255 | -0.15445325 | [-0.18 -0.12] | 4.57E-24 | 1.36E-21 |
| L_LDL_CE | Lipodystrophy_1_pPS | 4255 | 0.041058293 | [0.01 0.07] | 0.00747721 | 0.037534473 |
| L_LDL_CE | Lipodystrophy_2_pPS | 4255 | -0.044533866 | [-0.07 -0.01] | 0.003714452 | 0.021709807 |
| L_LDL_CE | Liver-Lipid_pPS | 4255 | 0.005031011 | [-0.03 0.04] | 0.743201187 | 0.876709962 |
| L_LDL_CE | ASAT_Pos_pPS | 4255 | -0.012096316 | [-0.04 0.02] | 0.430854475 | 0.688963257 |
| L_LDL_CE | ALP_Neg_pPS | 4255 | 0.047989975 | [0.02 0.08] | 0.001766801 | 0.01292441 |
| L_LDL_CE | SHBG-LpA_pPS | 4255 | 0.018163199 | [-0.01 0.05] | 0.236861006 | 0.519495765 |
| L_LDL_CE | Obesity_pPS | 4255 | -0.006857444 | [-0.04 0.02] | 0.655195685 | 0.821454552 |
| L_LDL_CE | VAT_Neg_pPS | 4255 | 0.011732458 | [-0.02 0.04] | 0.444847182 | 0.692953671 |
| L_LDL_CE | Bilirubin_pPS | 4255 | -0.013701904 | [-0.04 0.02] | 0.372233305 | 0.64437772 |
| L_LDL_CE | Proinsulin_pPS | 4255 | -0.005836565 | [-0.04 0.02] | 0.703889563 | 0.856405622 |
| L_LDL_CE | Hyper_Insulin_pPS | 4255 | -0.020026012 | [-0.05 0.01] | 0.192162991 | 0.464503229 |
| L_LDL_CE | Beta_Cell_1_pPS | 4255 | 0.020332983 | [-0.01 0.05] | 0.185435669 | 0.456669931 |
| L_LDL_CE | Beta_Cell_2_pPS | 4255 | 0.022305783 | [-0.01 0.05] | 0.146303253 | 0.396530158 |
| L_LDL_CE | Total_GRS | 4255 | -0.001888986 | [-0.03 0.03] | 0.902099773 | 0.95179735 |
| L_LDL_FC | Cholesterol_pPS | 4255 | -0.146777948 | [-0.18 -0.12] | 7.24E-22 | 6.48E-20 |
| L_LDL_FC | Lipodystrophy_1_pPS | 4255 | 0.02747599 | [-0. 0.06] | 0.073526257 | 0.235745623 |
| L_LDL_FC | Lipodystrophy_2_pPS | 4255 | -0.048103379 | [-0.08 -0.02] | 0.001722857 | 0.0127432 |
| L_LDL_FC | Liver-Lipid_pPS | 4255 | 0.019113826 | [-0.01 0.05] | 0.213209213 | 0.493880952 |
| L_LDL_FC | ASAT_Pos_pPS | 4255 | -0.02001541 | [-0.05 0.01] | 0.192398483 | 0.464503229 |
| L_LDL_FC | ALP_Neg_pPS | 4255 | 0.048416942 | [0.02 0.08] | 0.001606528 | 0.012219869 |
| L_LDL_FC | SHBG-LpA_pPS | 4255 | 0.016708507 | [-0.01 0.05] | 0.276542468 | 0.561195453 |
| L_LDL_FC | Obesity_pPS | 4255 | -0.013757948 | [-0.04 0.02] | 0.370280612 | 0.643918869 |
| L_LDL_FC | VAT_Neg_pPS | 4255 | 0.010513239 | [-0.02 0.04] | 0.493576304 | 0.721879553 |

**b) Partial Correlation**

| Metabolomics | PPS | Samp | r | CI95% | p | fdr_p_adjusted |
|---|---|---|---|---|---|---|
| L_HDL_PL | VAT_Neg_pPS | 4255 | -0.020734524 | [-0.05 0.01] | 0.177565864 | 0.455752383 |
| L_HDL_PL | Bilirubin_pPS | 4255 | 0.000333521 | [-0.03 0.03] | 0.982698984 | 0.993363084 |
| L_HDL_PL | Proinsulin_pPS | 4255 | -0.004142047 | [-0.03 0.03] | 0.787688184 | 0.898036301 |
| L_HDL_PL | Hyper_Insulin_pPS | 4255 | -0.000238183 | [-0.03 0.03] | 0.987644091 | 0.993363084 |
| L_HDL_PL | Beta_Cell_1_pPS | 4255 | 0.000956855 | [-0.03 0.03] | 0.950392415 | 0.978747744 |
| L_HDL_PL | Beta_Cell_2_pPS | 4255 | 0.008763214 | [-0.02 0.04] | 0.568821272 | 0.797755947 |
| L_HDL_TG | Cholesterol_pPS | 4255 | -0.00744931 | [-0.04 0.02] | 0.628133861 | 0.835009289 |
| L_HDL_TG | Lipodystrophy_1_pPS | 4255 | -0.005526017 | [-0.04 0.02] | 0.719370596 | 0.866319132 |
| L_HDL_TG | Lipodystrophy_2_pPS | 4255 | -0.045552483 | [-0.08 -0.02] | 0.003043346 | 0.021082278 |
| L_HDL_TG | Liver-Lipid_pPS | 4255 | -0.003731934 | [-0.03 0.03] | 0.808277827 | 0.910620264 |
| L_HDL_TG | ASAT_Pos_pPS | 4255 | -0.020810222 | [-0.05 0.01] | 0.175988205 | 0.453447086 |
| L_HDL_TG | ALP_Neg_pPS | 4255 | 0.042205079 | [0.01 0.07] | 0.006044365 | 0.035801237 |
| L_HDL_TG | SHBG-LpA_pPS | 4255 | 0.00236782 | [-0.03 0.03] | 0.877645971 | 0.946684933 |
| L_HDL_TG | Obesity_pPS | 4255 | -0.028543016 | [-0.06 0. ] | 0.063421774 | 0.226094412 |
| L_HDL_TG | VAT_Neg_pPS | 4255 | -0.00656965 | [-0.04 0.02] | 0.669264131 | 0.850060323 |
| L_HDL_TG | Bilirubin_pPS | 4255 | -0.010068802 | [-0.04 0.02] | 0.512672324 | 0.767666412 |
| L_HDL_TG | Proinsulin_pPS | 4255 | 0.008899618 | [-0.02 0.04] | 0.562820142 | 0.797755947 |
| L_HDL_TG | Hyper_Insulin_pPS | 4255 | 0.00036445 | [-0.03 0.03] | 0.981094914 | 0.992998998 |
| L_HDL_TG | Beta_Cell_1_pPS | 4255 | -0.013658647 | [-0.04 0.02] | 0.374478285 | 0.655035906 |
| L_HDL_TG | Beta_Cell_2_pPS | 4255 | 0.018950789 | [-0.01 0.05] | 0.217845891 | 0.510687908 |
| L_LDL_C | Cholesterol_pPS | 4255 | -0.15140774 | [-0.18 -0.12] | 4.10E-23 | 5.47E-21 |
| L_LDL_C | Lipodystrophy_1_pPS | 4255 | 0.043274262 | [0.01 0.07] | 0.004878061 | 0.030234918 |
| L_LDL_C | Lipodystrophy_2_pPS | 4255 | -0.041118959 | [-0.07 -0.01] | 0.007480639 | 0.043283928 |
| L_LDL_C | Liver-Lipid_pPS | 4255 | 0.010770446 | [-0.02 0.04] | 0.483735975 | 0.743238236 |
| L_LDL_C | ASAT_Pos_pPS | 4255 | -0.017467997 | [-0.05 0.01] | 0.256022555 | 0.55296819 |
| L_LDL_C | ALP_Neg_pPS | 4255 | 0.051543628 | [0.02 0.08] | 0.000797802 | 0.00742884 |
| L_LDL_C | SHBG-LpA_pPS | 4255 | 0.015128314 | [-0.02 0.05] | 0.325269149 | 0.623147212 |
| L_LDL_C | Obesity_pPS | 4255 | 0.006637384 | [-0.02 0.04] | 0.666059544 | 0.849453588 |
| L_LDL_C | VAT_Neg_pPS | 4255 | 0.008846862 | [-0.02 0.04] | 0.565137498 | 0.797755947 |
| L_LDL_C | Bilirubin_pPS | 4255 | -0.013983179 | [-0.04 0.02] | 0.363234046 | 0.650367164 |
| L_LDL_C | Proinsulin_pPS | 4255 | -0.021039169 | [-0.05 0.01] | 0.171280169 | 0.448811801 |
| L_LDL_C | Hyper_Insulin_pPS | 4255 | -0.022355309 | [-0.05 0.01] | 0.146027671 | 0.420039364 |
| L_LDL_C | Beta_Cell_1_pPS | 4255 | 0.014434542 | [-0.02 0.04] | 0.347950703 | 0.637861136 |
| L_LDL_C | Beta_Cell_2_pPS | 4255 | 0.022612497 | [-0.01 0.05] | 0.141443943 | 0.410988061 |
| L_LDL_CE | Cholesterol_pPS | 4255 | -0.152823572 | [-0.18 -0.12] | 1.60E-23 | 4.03E-21 |
| L_LDL_CE | Lipodystrophy_1_pPS | 4255 | 0.046054551 | [0.02 0.08] | 0.002735276 | 0.019557221 |
| L_LDL_CE | Lipodystrophy_2_pPS | 4255 | -0.039954316 | [-0.07 -0.01] | 0.009354032 | 0.051306227 |
| L_LDL_CE | Liver-Lipid_pPS | 4255 | 0.007472818 | [-0.02 0.04] | 0.62704965 | 0.834676462 |
| L_LDL_CE | ASAT_Pos_pPS | 4255 | -0.016065327 | [-0.05 0.01] | 0.296197451 | 0.594033661 |
| L_LDL_CE | ALP_Neg_pPS | 4255 | 0.05130912 | [0.02 0.08] | 0.000842993 | 0.007741614 |
| L_LDL_CE | SHBG-LpA_pPS | 4255 | 0.01517945 | [-0.01 0.05] | 0.323636281 | 0.620844623 |
| L_LDL_CE | Obesity_pPS | 4255 | 0.00827363 | [-0.02 0.04] | 0.590608624 | 0.816573526 |
| L_LDL_CE | VAT_Neg_pPS | 4255 | 0.008579886 | [-0.02 0.04] | 0.576934658 | 0.803773963 |
| L_LDL_CE | Bilirubin_pPS | 4255 | -0.014936155 | [-0.05 0.02] | 0.331452952 | 0.62795236 |
| L_LDL_CE | Proinsulin_pPS | 4255 | -0.020646256 | [-0.05 0.01] | 0.179418798 | 0.457575075 |
| L_LDL_CE | Hyper_Insulin_pPS | 4255 | -0.022317258 | [-0.05 0.01] | 0.146715366 | 0.420206242 |
| L_LDL_CE | Beta_Cell_1_pPS | 4255 | 0.015447201 | [-0.01 0.05] | 0.315173792 | 0.616791721 |
| L_LDL_CE | Beta_Cell_2_pPS | 4255 | 0.021632937 | [-0.01 0.05] | 0.159509999 | 0.433462108 |
| L_LDL_FC | Cholesterol_pPS | 4255 | -0.144813289 | [-0.17 -0.12] | 2.92E-21 | 2.44E-19 |
| L_LDL_FC | Lipodystrophy_1_pPS | 4255 | 0.033795007 | [0. 0.06] | 0.02795245 | 0.117318251 |
| L_LDL_FC | Lipodystrophy_2_pPS | 4255 | -0.04313335 | [-0.07 -0.01] | 0.005019138 | 0.03072879 |
| L_LDL_FC | Liver-Lipid_pPS | 4255 | 0.021923157 | [-0.01 0.05] | 0.153984388 | 0.428688101 |
| L_LDL_FC | ASAT_Pos_pPS | 4255 | -0.021239369 | [-0.05 0.01] | 0.167241104 | 0.442294174 |
| L_LDL_FC | ALP_Neg_pPS | 4255 | 0.051628222 | [0.02 0.08] | 0.00078206 | 0.007350631 |
| L_LDL_FC | SHBG-LpA_pPS | 4255 | 0.014745249 | [-0.02 0.04] | 0.33767117 | 0.631792226 |
| L_LDL_FC | Obesity_pPS | 4255 | 0.001234922 | [-0.03 0.03] | 0.936003688 | 0.978170683 |
| L_LDL_FC | VAT_Neg_pPS | 4255 | 0.008416243 | [-0.02 0.04] | 0.584222578 | 0.81166801 |
| L_LDL_FC | Bilirubin_pPS | 4255 | -0.012325205 | [-0.04 0.02] | 0.422896779 | 0.693893617 |
| L_LDL_FC | Proinsulin_pPS | 4255 | -0.021949618 | [-0.05 0.01] | 0.153487919 | 0.428688101 |
| L_LDL_FC | Hyper_Insulin_pPS | 4255 | -0.021542431 | [-0.05 0.01] | 0.16126349 | 0.435694341 |
| L_LDL_FC | Beta_Cell_1_pPS | 4255 | 0.01192442 | [-0.02 0.04] | 0.438136324 | 0.702280961 |
| L_LDL_FC | Beta_Cell_2_pPS | 4255 | 0.024907256 | [-0.01 0.06] | 0.105296343 | 0.336746451 |
| L_LDL_L | Cholesterol_pPS | 4255 | -0.151549036 | [-0.18 -0.12] | 3.73E-23 | 5.34E-21 |
| L_LDL_L | Lipodystrophy_1_pPS | 4255 | 0.045852884 | [0.02 0.08] | 0.002855425 | 0.019954421 |
| L_LDL_L | Lipodystrophy_2_pPS | 4255 | -0.040432213 | [-0.07 -0.01] | 0.008539827 | 0.047890008 |
| L_LDL_L | Liver-Lipid_pPS | 4255 | 0.008372459 | [-0.02 0.04] | 0.586179735 | 0.813258371 |
| L_LDL_L | ASAT_Pos_pPS | 4255 | -0.019265674 | [-0.05 0.01] | 0.210294689 | 0.4972684 |
| L_LDL_L | ALP_Neg_pPS | 4255 | 0.052505119 | [0.02 0.08] | 0.000635019 | 0.006420748 |
| L_LDL_L | SHBG-LpA_pPS | 4255 | 0.014915329 | [-0.02 0.05] | 0.332127693 | 0.62795236 |
| L_LDL_L | Obesity_pPS | 4255 | 0.007930193 | [-0.02 0.04] | 0.606117633 | 0.824474694 |
| L_LDL_L | VAT_Neg_pPS | 4255 | 0.008205705 | [-0.02 0.04] | 0.59366152 | 0.817359021 |
| L_LDL_L | Bilirubin_pPS | 4255 | -0.013485615 | [-0.04 0.02] | 0.380560441 | 0.661235752 |
| L_LDL_L | Proinsulin_pPS | 4255 | -0.018807965 | [-0.05 0.01] | 0.221334407 | 0.514647483 |
| L_LDL_L | Hyper_Insulin_pPS | 4255 | -0.02202168 | [-0.05 0.01] | 0.152142016 | 0.427792579 |
| L_LDL_L | Beta_Cell_1_pPS | 4255 | 0.013397344 | [-0.02 0.04] | 0.383686789 | 0.66173638 |
| L_LDL_L | Beta_Cell_2_pPS | 4255 | 0.022469386 | [-0.01 0.05] | 0.14398071 | 0.4165382 |
| L_LDL_P | Cholesterol_pPS | 4255 | -0.151583951 | [-0.18 -0.12] | 3.65E-23 | 5.34E-21 |
| L_LDL_P | Lipodystrophy_1_pPS | 4255 | 0.04766463 | [0.02 0.08] | 0.001928701 | 0.014908341 |
| L_LDL_P | Lipodystrophy_2_pPS | 4255 | -0.039753185 | [-0.07 -0.01] | 0.009716923 | 0.052718915 |
| L_LDL_P | Liver-Lipid_pPS | 4255 | 0.006911654 | [-0.02 0.04] | 0.65314619 | 0.844628798 |
| L_LDL_P | ASAT_Pos_pPS | 4255 | -0.019499581 | [-0.05 0.01] | 0.204809291 | 0.489878376 |
| L_LDL_P | ALP_Neg_pPS | 4255 | 0.052860581 | [0.02 0.08] | 0.000583094 | 0.0061118 |
| L_LDL_P | SHBG-LpA_pPS | 4255 | 0.014879331 | [-0.02 0.04] | 0.333296047 | 0.628957504 |
| L_LDL_P | Obesity_pPS | 4255 | 0.008732617 | [-0.02 0.04] | 0.570171583 | 0.798240216 |
| L_LDL_P | VAT_Neg_pPS | 4255 | 0.007964698 | [-0.02 0.04] | 0.604551241 | 0.824462932 |
| L_LDL_P | Bilirubin_pPS | 4255 | -0.013295437 | [-0.04 0.02] | 0.387314796 | 0.665583022 |
| L_LDL_P | Proinsulin_pPS | 4255 | -0.017813727 | [-0.05 0.01] | 0.246730747 | 0.544603039 |
| L_LDL_P | Hyper_Insulin_pPS | 4255 | -0.021730367 | [-0.05 0.01] | 0.157638517 | 0.431435756 |
| L_LDL_P | Beta_Cell_1_pPS | 4255 | 0.013015315 | [-0.02 0.04] | 0.397395687 | 0.673455453 |
| L_LDL_P | Beta_Cell_2_pPS | 4255 | 0.02219772 | [-0.01 0.05] | 0.148891928 | 0.424616648 |
| L_LDL_PL | Cholesterol_pPS | 4255 | -0.151841163 | [-0.18 -0.12] | 3.08E-23 | 5.13E-21 |
| L_LDL_PL | Lipodystrophy_1_pPS | 4255 | 0.044954895 | [0.01 0.07] | 0.003451088 | 0.023262887 |
| L_LDL_PL | Lipodystrophy_2_pPS | 4255 | -0.043296122 | [-0.07 -0.01] | 0.0048565 | 0.03019476 |
| L_LDL_PL | Liver-Lipid_pPS | 4255 | 0.005167156 | [-0.02 0.04] | 0.73689593 | 0.87275313 |

**Table S9: Associations between pPSs and global metabolomic profile in SCAPIS**

**a) Spearman Correlation**

| Metabolomics | PPS | Samp | r | CI95% | p | fdr_p_adjusted |
|---|---|---|---|---|---|---|
| L_LDL_FC | Bilirubin_pPS | 4255 | -0.011925297 | [-0.04 0.02] | 0.437399239 | 0.690884659 |
| L_LDL_FC | Proinsulin_pPS | 4255 | -0.008098823 | [-0.04 0.02] | 0.59791801 | 0.787890007 |
| L_LDL_FC | Hyper_Insulin_pPS | 4255 | -0.021096143 | [-0.05 0.01] | 0.169468012 | 0.431822279 |
| L_LDL_FC | Beta_Cell_1_pPS | 4255 | 0.01688913 | [-0.01 0.05] | 0.271382772 | 0.554396235 |
| L_LDL_FC | Beta_Cell_2_pPS | 4255 | 0.024864522 | [-0.01 0.05] | 0.105359901 | 0.314239045 |
| L_LDL_FC | Total_GRS | 4255 | -0.009907613 | [-0.04 0.02] | 0.518803479 | 0.736667707 |
| L_LDL_L | Cholesterol_pPS | 4255 | -0.153075605 | [-0.18 -0.12] | 1.16E-23 | 1.74E-21 |
| L_LDL_L | Lipodystrophy_1_pPS | 4255 | 0.040275722 | [0.01 0.07] | 0.008695886 | 0.04248901 |
| L_LDL_L | Lipodystrophy_2_pPS | 4255 | -0.045206179 | [-0.08 -0.02] | 0.003226496 | 0.019717477 |
| L_LDL_L | Liver-Lipid_pPS | 4255 | 0.005941842 | [-0.02 0.04] | 0.69880731 | 0.852954013 |
| L_LDL_L | ASAT_Pos_pPS | 4255 | -0.015426146 | [-0.05 0.01] | 0.315091018 | 0.60298573 |
| L_LDL_L | ALP_Neg_pPS | 4255 | 0.049578986 | [0.02 0.08] | 0.001235624 | 0.009852836 |
| L_LDL_L | SHBG-LpA_pPS | 4255 | 0.017861864 | [-0.01 0.05] | 0.244731449 | 0.525480742 |
| L_LDL_L | Obesity_pPS | 4255 | -0.006924288 | [-0.04 0.02] | 0.652055144 | 0.819366306 |
| L_LDL_L | VAT_Neg_pPS | 4255 | 0.011118794 | [-0.02 0.04] | 0.46902368 | 0.707973539 |
| L_LDL_L | Bilirubin_pPS | 4255 | -0.01241187 | [-0.04 0.02] | 0.418928701 | 0.679729247 |
| L_LDL_L | Proinsulin_pPS | 4255 | -0.004199626 | [-0.03 0.03] | 0.784486452 | 0.897605875 |
| L_LDL_L | Hyper_Insulin_pPS | 4255 | -0.0200215 | [-0.05 0.01] | 0.192263179 | 0.464503229 |
| L_LDL_L | Beta_Cell_1_pPS | 4255 | 0.018853805 | [-0.01 0.05] | 0.219502405 | 0.500648222 |
| L_LDL_L | Beta_Cell_2_pPS | 4255 | 0.023012331 | [-0.01 0.05] | 0.133940766 | 0.373120705 |
| L_LDL_L | Total_GRS | 4255 | -0.002326059 | [-0.03 0.03] | 0.879604087 | 0.944792572 |
| L_LDL_P | Cholesterol_pPS | 4255 | -0.152996702 | [-0.18 -0.12] | 1.22E-23 | 1.74E-21 |
| L_LDL_P | Lipodystrophy_1_pPS | 4255 | 0.042158806 | [0.01 0.07] | 0.006022286 | 0.03158387 |
| L_LDL_P | Lipodystrophy_2_pPS | 4255 | -0.044479853 | [-0.07 -0.01] | 0.003756426 | 0.021836135 |
| L_LDL_P | Liver-Lipid_pPS | 4255 | 0.004572762 | [-0.03 0.03] | 0.765874284 | 0.889697961 |
| L_LDL_P | ASAT_Pos_pPS | 4255 | -0.015238993 | [-0.05 0.01] | 0.320998787 | 0.608204326 |
| L_LDL_P | ALP_Neg_pPS | 4255 | 0.050112913 | [0.02 0.08] | 0.001093248 | 0.009017049 |
| L_LDL_P | SHBG-LpA_pPS | 4255 | 0.017968887 | [-0.01 0.05] | 0.24191542 | 0.525211109 |
| L_LDL_P | Obesity_pPS | 4255 | -0.005935123 | [-0.04 0.02] | 0.699131294 | 0.852954013 |
| L_LDL_P | VAT_Neg_pPS | 4255 | 0.010987018 | [-0.02 0.04] | 0.474308433 | 0.711462649 |
| L_LDL_P | Bilirubin_pPS | 4255 | -0.012165607 | [-0.04 0.02] | 0.428219037 | 0.687522331 |
| L_LDL_P | Proinsulin_pPS | 4255 | -0.003190316 | [-0.03 0.03] | 0.835423336 | 0.925322958 |
| L_LDL_P | Hyper_Insulin_pPS | 4255 | -0.0195098 | [-0.05 0.01] | 0.203877402 | 0.482157693 |
| L_LDL_P | Beta_Cell_1_pPS | 4255 | 0.018664028 | [-0.01 0.05] | 0.224178884 | 0.505575551 |
| L_LDL_P | Beta_Cell_2_pPS | 4255 | 0.022793469 | [-0.01 0.05] | 0.13768069 | 0.380083759 |
| L_LDL_P | Total_GRS | 4255 | -0.000985675 | [-0.03 0.03] | 0.94882187 | 0.975190662 |
| L_LDL_PL | Cholesterol_pPS | 4255 | -0.15352636 | [-0.18 -0.12] | 8.54E-24 | 1.63E-21 |
| L_LDL_PL | Lipodystrophy_1_pPS | 4255 | 0.039517112 | [0.01 0.07] | 0.010043585 | 0.047244496 |
| L_LDL_PL | Lipodystrophy_2_pPS | 4255 | -0.048238121 | [-0.08 -0.02] | 0.001671951 | 0.01249255 |
| L_LDL_PL | Liver-Lipid_pPS | 4255 | 0.002752845 | [-0.03 0.03] | 0.857731068 | 0.934399767 |
| L_LDL_PL | ASAT_Pos_pPS | 4255 | -0.014614771 | [-0.04 0.02] | 0.341222825 | 0.623975243 |
| L_LDL_PL | ALP_Neg_pPS | 4255 | 0.049797057 | [0.02 0.08] | 0.001175523 | 0.00951508 |
| L_LDL_PL | SHBG-LpA_pPS | 4255 | 0.018285999 | [-0.01 0.05] | 0.233705498 | 0.516301672 |
| L_LDL_PL | Obesity_pPS | 4255 | -0.006692287 | [-0.04 0.02] | 0.662981323 | 0.825654931 |
| L_LDL_PL | VAT_Neg_pPS | 4255 | 0.010008298 | [-0.02 0.04] | 0.514563825 | 0.733381664 |
| L_LDL_PL | Bilirubin_pPS | 4255 | -0.012942681 | [-0.04 0.02] | 0.399311588 | 0.666036824 |
| L_LDL_PL | Proinsulin_pPS | 4255 | -0.004546803 | [-0.03 0.03] | 0.767164878 | 0.889703209 |
| L_LDL_PL | Hyper_Insulin_pPS | 4255 | -0.019732119 | [-0.05 0.01] | 0.198770135 | 0.473735488 |
| L_LDL_PL | Beta_Cell_1_pPS | 4255 | 0.020099679 | [-0.01 0.05] | 0.190532439 | 0.463895666 |
| L_LDL_PL | Beta_Cell_2_pPS | 4255 | 0.022443153 | [-0.01 0.05] | 0.143833712 | 0.391527046 |
| L_LDL_PL | Total_GRS | 4255 | -0.003009512 | [-0.03 0.03] | 0.844628015 | 0.92810697 |
| L_LDL_TG | Cholesterol_pPS | 4255 | -0.107396572 | [-0.14 -0.08] | 2.32E-12 | 1.04E-10 |
| L_LDL_TG | Lipodystrophy_1_pPS | 4255 | 0.058955563 | [0.03 0.09] | 0.000121799 | 0.001604501 |
| L_LDL_TG | Lipodystrophy_2_pPS | 4255 | -0.017096496 | [-0.05 0.01] | 0.265540896 | 0.551923664 |
| L_LDL_TG | Liver-Lipid_pPS | 4255 | -0.012452581 | [-0.04 0.02] | 0.417404406 | 0.678719122 |
| L_LDL_TG | ASAT_Pos_pPS | 4255 | -0.009903788 | [-0.04 0.02] | 0.518964912 | 0.736667707 |
| L_LDL_TG | ALP_Neg_pPS | 4255 | 0.053083327 | [0.02 0.08] | 0.000541826 | 0.005560841 |
| L_LDL_TG | SHBG-LpA_pPS | 4255 | 0.010181 | [-0.02 0.04] | 0.507333775 | 0.73035634 |
| L_LDL_TG | Obesity_pPS | 4255 | 0.007603283 | [-0.02 0.04] | 0.620510997 | 0.801322148 |
| L_LDL_TG | VAT_Neg_pPS | 4255 | 0.001836395 | [-0.03 0.03] | 0.904812277 | 0.952872131 |
| L_LDL_TG | Bilirubin_pPS | 4255 | -0.005606188 | [-0.04 0.02] | 0.715056991 | 0.863138574 |
| L_LDL_TG | Proinsulin_pPS | 4255 | 0.010478015 | [-0.02 0.04] | 0.49502532 | 0.722332865 |
| L_LDL_TG | Hyper_Insulin_pPS | 4255 | -0.007479751 | [-0.04 0.02] | 0.626200486 | 0.803929814 |
| L_LDL_TG | Beta_Cell_1_pPS | 4255 | 0.000103432 | [-0.03 0.03] | 0.994625986 | 0.996787043 |
| L_LDL_TG | Beta_Cell_2_pPS | 4255 | 0.011350192 | [-0.02 0.04] | 0.459822983 | 0.700511576 |
| L_LDL_TG | Total_GRS | 4255 | 0.016370253 | [-0.01 0.05] | 0.286383894 | 0.57504794 |
| L_VLDL_C | Cholesterol_pPS | 4255 | -0.083462674 | [-0.11 -0.05] | 5.19E-08 | 1.39E-06 |
| L_VLDL_C | Lipodystrophy_1_pPS | 4255 | 0.065167182 | [0.04 0.1 ] | 2.16E-05 | 0.000353437 |
| L_VLDL_C | Lipodystrophy_2_pPS | 4255 | 0.003367633 | [-0.03 0.03] | 0.826417955 | 0.921344342 |
| L_VLDL_C | Liver-Lipid_pPS | 4255 | -0.045683165 | [-0.08 -0.02] | 0.002916528 | 0.018618907 |
| L_VLDL_C | ASAT_Pos_pPS | 4255 | 0.046405564 | [0.02 0.08] | 0.002498538 | 0.01654125 |
| L_VLDL_C | ALP_Neg_pPS | 4255 | 0.007470946 | [-0.02 0.04] | 0.62660689 | 0.803929814 |
| L_VLDL_C | SHBG-LpA_pPS | 4255 | -0.002980175 | [-0.03 0.03] | 0.846123565 | 0.92810697 |
| L_VLDL_C | Obesity_pPS | 4255 | 0.031774119 | [0. 0.06] | 0.038487453 | 0.144076068 |
| L_VLDL_C | VAT_Neg_pPS | 4255 | 0.02095744 | [-0.01 0.05] | 0.172290778 | 0.436320801 |
| L_VLDL_C | Bilirubin_pPS | 4255 | -0.010511972 | [-0.04 0.02] | 0.493628371 | 0.721879553 |
| L_VLDL_C | Proinsulin_pPS | 4255 | 0.011408535 | [-0.02 0.04] | 0.457519225 | 0.697993412 |
| L_VLDL_C | Hyper_Insulin_pPS | 4255 | 0.015965362 | [-0.01 0.05] | 0.298471577 | 0.584165937 |
| L_VLDL_C | Beta_Cell_1_pPS | 4255 | 0.012923018 | [-0.02 0.04] | 0.400028289 | 0.666677506 |
| L_VLDL_C | Beta_Cell_2_pPS | 4255 | -0.000299549 | [-0.03 0.03] | 0.984437176 | 0.990439842 |
| L_VLDL_C | Total_GRS | 4255 | 0.03800181 | [0.01 0.07] | 0.013303536 | 0.060075966 |
| L_VLDL_CE | Cholesterol_pPS | 4255 | -0.08438446 | [-0.11 -0.05] | 3.69E-08 | 1.02E-06 |
| L_VLDL_CE | Lipodystrophy_1_pPS | 4255 | 0.065001149 | [0.03 0.09] | 2.26E-05 | 0.000364851 |
| L_VLDL_CE | Lipodystrophy_2_pPS | 4255 | 0.000544052 | [-0.03 0.03] | 0.971738321 | 0.983305352 |
| L_VLDL_CE | Liver-Lipid_pPS | 4255 | -0.039595921 | [-0.07 -0.01] | 0.009895402 | 0.046752503 |
| L_VLDL_CE | ASAT_Pos_pPS | 4255 | 0.047546934 | [0.02 0.08] | 0.001948533 | 0.013858578 |
| L_VLDL_CE | ALP_Neg_pPS | 4255 | 0.010912949 | [-0.02 0.04] | 0.477293212 | 0.714440991 |
| L_VLDL_CE | SHBG-LpA_pPS | 4255 | -0.003894426 | [-0.03 0.03] | 0.799802618 | 0.905317475 |
| L_VLDL_CE | Obesity_pPS | 4255 | 0.030208324 | [0. 0.06] | 0.049115402 | 0.171863846 |
| L_VLDL_CE | VAT_Neg_pPS | 4255 | 0.021264452 | [-0.01 0.05] | 0.166089465 | 0.427172545 |
| L_VLDL_CE | Bilirubin_pPS | 4255 | -0.012041075 | [-0.04 0.02] | 0.432962291 | 0.68946111 |
| L_VLDL_CE | Proinsulin_pPS | 4255 | 0.012203365 | [-0.02 0.04] | 0.426786874 | 0.687522331 |
| L_VLDL_CE | Hyper_Insulin_pPS | 4255 | 0.016037631 | [-0.01 0.05] | 0.296289447 | 0.58253058 |
| L_VLDL_CE | Beta_Cell_1_pPS | 4255 | 0.013734685 | [-0.02 0.04] | 0.371090369 | 0.643918869 |

**b) Partial Correlation**

| Metabolomics | PPS | Samp | r | CI95% | p | fdr_p_adjusted |
|---|---|---|---|---|---|---|
| L_LDL_PL | ASAT_Pos_pPS | 4255 | -0.018108893 | [-0.05 0.01] | 0.238986907 | 0.535744184 |
| L_LDL_PL | ALP_Neg_pPS | 4255 | 0.053027211 | [0.02 0.08] | 0.000560139 | 0.005903363 |
| L_LDL_PL | SHBG-LpA_pPS | 4255 | 0.015530311 | [-0.01 0.05] | 0.312576886 | 0.614714072 |
| L_LDL_PL | Obesity_pPS | 4255 | 0.008245303 | [-0.02 0.04] | 0.59188092 | 0.817111563 |
| L_LDL_PL | VAT_Neg_pPS | 4255 | 0.00728683 | [-0.02 0.04] | 0.635649407 | 0.835456904 |
| L_LDL_PL | Bilirubin_pPS | 4255 | -0.01410438 | [-0.04 0.02] | 0.359089441 | 0.64484135 |
| L_LDL_PL | Proinsulin_pPS | 4255 | -0.01951902 | [-0.05 0.01] | 0.20435814 | 0.489878376 |
| L_LDL_PL | Hyper_Insulin_pPS | 4255 | -0.021347862 | [-0.05 0.01] | 0.165082337 | 0.441248115 |
| L_LDL_PL | Beta_Cell_1_pPS | 4255 | 0.015077692 | [-0.02 0.05] | 0.326890863 | 0.625128976 |
| L_LDL_PL | Beta_Cell_2_pPS | 4255 | 0.021782317 | [-0.01 0.05] | 0.156647461 | 0.431435756 |
| L_LDL_TG | Cholesterol_pPS | 4255 | -0.108255282 | [-0.14 -0.08] | 1.67E-12 | 6.97E-11 |
| L_LDL_TG | Lipodystrophy_1_pPS | 4255 | 0.062209717 | [0.03 0.09] | 5.15E-05 | 0.000852296 |
| L_LDL_TG | Lipodystrophy_2_pPS | 4255 | -0.014141831 | [-0.04 0.02] | 0.357814817 | 0.64419538 |
| L_LDL_TG | Liver-Lipid_pPS | 4255 | -0.012093739 | [-0.04 0.02] | 0.431659926 | 0.69698199 |
| L_LDL_TG | ASAT_Pos_pPS | 4255 | -0.019102151 | [-0.05 0.01] | 0.214192126 | 0.504485455 |
| L_LDL_TG | ALP_Neg_pPS | 4255 | 0.051852123 | [0.02 0.08] | 0.000741774 | 0.007105415 |
| L_LDL_TG | SHBG-LpA_pPS | 4255 | 0.006156775 | [-0.02 0.04] | 0.688926323 | 0.858568103 |
| L_LDL_TG | Obesity_pPS | 4255 | 0.015207379 | [-0.01 0.05] | 0.322746696 | 0.620844623 |
| L_LDL_TG | VAT_Neg_pPS | 4255 | -0.001019265 | [-0.03 0.03] | 0.947161413 | 0.978747744 |
| L_LDL_TG | Bilirubin_pPS | 4255 | -0.006753928 | [-0.04 0.02] | 0.660559969 | 0.8466332 |
| L_LDL_TG | Proinsulin_pPS | 4255 | 0.001544565 | [-0.03 0.03] | 0.920005724 | 0.968376161 |
| L_LDL_TG | Hyper_Insulin_pPS | 4255 | -0.012385536 | [-0.04 0.02] | 0.420629937 | 0.692597028 |
| L_LDL_TG | Beta_Cell_1_pPS | 4255 | -0.005872237 | [-0.04 0.02] | 0.702601128 | 0.859880159 |
| L_LDL_TG | Beta_Cell_2_pPS | 4255 | 0.012831648 | [-0.02 0.04] | 0.404090418 | 0.677078992 |
| L_VLDL_C | Cholesterol_pPS | 4255 | -0.085140626 | [-0.11 -0.06] | 2.93E-08 | 7.51E-07 |
| L_VLDL_C | Lipodystrophy_1_pPS | 4255 | 0.056854476 | [0.03 0.09] | 0.000215917 | 0.002788809 |
| L_VLDL_C | Lipodystrophy_2_pPS | 4255 | -0.000802373 | [-0.03 0.03] | 0.958393491 | 0.981434153 |
| L_VLDL_C | Liver-Lipid_pPS | 4255 | -0.046392992 | [-0.08 -0.02] | 0.002543972 | 0.018254598 |
| L_VLDL_C | ASAT_Pos_pPS | 4255 | 0.033701553 | [0. 0.06] | 0.028388781 | 0.118900294 |
| L_VLDL_C | ALP_Neg_pPS | 4255 | 0.006196119 | [-0.02 0.04] | 0.687043334 | 0.858568103 |
| L_VLDL_C | SHBG-LpA_pPS | 4255 | -0.009541157 | [-0.04 0.02] | 0.535012388 | 0.781586915 |
| L_VLDL_C | Obesity_pPS | 4255 | 0.035014109 | [0. 0.07] | 0.022768999 | 0.100625906 |
| L_VLDL_C | VAT_Neg_pPS | 4255 | 0.015926739 | [-0.01 0.05] | 0.300384152 | 0.597661337 |
| L_VLDL_C | Bilirubin_pPS | 4255 | -0.014276971 | [-0.04 0.02] | 0.353239044 | 0.639407383 |
| L_VLDL_C | Proinsulin_pPS | 4255 | 0.006276993 | [-0.02 0.04] | 0.683178802 | 0.858568103 |
| L_VLDL_C | Hyper_Insulin_pPS | 4255 | 0.006031392 | [-0.02 0.04] | 0.694939964 | 0.858808524 |
| L_VLDL_C | Beta_Cell_1_pPS | 4255 | 0.006585763 | [-0.02 0.04] | 0.66850122 | 0.849884836 |
| L_VLDL_C | Beta_Cell_2_pPS | 4255 | -0.002793661 | [-0.03 0.03] | 0.855863677 | 0.93707811 |
| L_VLDL_CE | Cholesterol_pPS | 4255 | -0.085919888 | [-0.12 -0.06] | 2.18E-08 | 5.83E-07 |
| L_VLDL_CE | Lipodystrophy_1_pPS | 4255 | 0.05665152 | [0.03 0.09] | 0.00022745 | 0.002863866 |
| L_VLDL_CE | Lipodystrophy_2_pPS | 4255 | -0.003975418 | [-0.03 0.03] | 0.796036766 | 0.902415406 |
| L_VLDL_CE | Liver-Lipid_pPS | 4255 | -0.040392437 | [-0.07 -0.01] | 0.008605094 | 0.048121224 |
| L_VLDL_CE | ASAT_Pos_pPS | 4255 | 0.035210592 | [0.01 0.07] | 0.022016937 | 0.098388186 |
| L_VLDL_CE | ALP_Neg_pPS | 4255 | 0.009355944 | [-0.02 0.04] | 0.542968924 | 0.788269605 |
| L_VLDL_CE | SHBG-LpA_pPS | 4255 | -0.010441616 | [-0.04 0.02] | 0.497185835 | 0.753597539 |
| L_VLDL_CE | Obesity_pPS | 4255 | 0.033291177 | [0. 0.06] | 0.03037492 | 0.123850487 |
| L_VLDL_CE | VAT_Neg_pPS | 4255 | 0.016402122 | [-0.01 0.05] | 0.28618637 | 0.583962408 |
| L_VLDL_CE | Bilirubin_pPS | 4255 | -0.015806734 | [-0.05 0.01] | 0.304041161 | 0.602663767 |
| L_VLDL_CE | Proinsulin_pPS | 4255 | 0.006276436 | [-0.02 0.04] | 0.683205376 | 0.858568103 |
| L_VLDL_CE | Hyper_Insulin_pPS | 4255 | 0.005928968 | [-0.02 0.04] | 0.699866782 | 0.85959098 |
| L_VLDL_CE | Beta_Cell_1_pPS | 4255 | 0.006518956 | [-0.02 0.04] | 0.671666482 | 0.851013914 |
| L_VLDL_CE | Beta_Cell_2_pPS | 4255 | -0.000571398 | [-0.03 0.03] | 0.970363901 | 0.988674515 |
| L_VLDL_FC | Cholesterol_pPS | 4255 | -0.083648379 | [-0.11 -0.05] | 5.09E-08 | 1.27E-06 |
| L_VLDL_FC | Lipodystrophy_1_pPS | 4255 | 0.056584168 | [0.03 0.09] | 0.000231403 | 0.002895433 |
| L_VLDL_FC | Lipodystrophy_2_pPS | 4255 | 0.002699447 | [-0.03 0.03] | 0.860673985 | 0.940026906 |
| L_VLDL_FC | Liver-Lipid_pPS | 4255 | -0.053342 | [-0.08 -0.02] | 0.000519052 | 0.005640495 |
| L_VLDL_FC | ASAT_Pos_pPS | 4255 | 0.031225415 | [0. 0.06] | 0.04228061 | 0.161230059 |
| L_VLDL_FC | ALP_Neg_pPS | 4255 | 0.00311192 | [-0.03 0.03] | 0.839654718 | 0.925654596 |
| L_VLDL_FC | SHBG-LpA_pPS | 4255 | -0.00883502 | [-0.04 0.02] | 0.565658326 | 0.797755947 |
| L_VLDL_FC | Obesity_pPS | 4255 | 0.03647116 | [0.01 0.07] | 0.017686546 | 0.083314038 |
| L_VLDL_FC | VAT_Neg_pPS | 4255 | 0.014914067 | [-0.02 0.05] | 0.332168606 | 0.62795236 |
| L_VLDL_FC | Bilirubin_pPS | 4255 | -0.012840418 | [-0.04 0.02] | 0.403769227 | 0.677078992 |
| L_VLDL_FC | Proinsulin_pPS | 4255 | 0.006530065 | [-0.02 0.04] | 0.671139742 | 0.850932085 |
| L_VLDL_FC | Hyper_Insulin_pPS | 4255 | 0.005430902 | [-0.02 0.04] | 0.724001731 | 0.866459051 |
| L_VLDL_FC | Beta_Cell_1_pPS | 4255 | 0.006459609 | [-0.02 0.04] | 0.674483141 | 0.853549462 |
| L_VLDL_FC | Beta_Cell_2_pPS | 4255 | -0.004492267 | [-0.03 0.03] | 0.770221473 | 0.888239279 |
| L_VLDL_L | Cholesterol_pPS | 4255 | -0.08137271 | [-0.11 -0.05] | 1.16E-07 | 2.67E-06 |
| L_VLDL_L | Lipodystrophy_1_pPS | 4255 | 0.056543712 | [0.03 0.09] | 0.000233809 | 0.002907361 |
| L_VLDL_L | Lipodystrophy_2_pPS | 4255 | 0.005160875 | [-0.02 0.04] | 0.737203935 | 0.87275313 |
| L_VLDL_L | Liver-Lipid_pPS | 4255 | -0.052064824 | [-0.08 -0.02] | 0.000705296 | 0.00685438 |
| L_VLDL_L | ASAT_Pos_pPS | 4255 | 0.033824752 | [0. 0.06] | 0.027814791 | 0.116985741 |
| L_VLDL_L | ALP_Neg_pPS | 4255 | -0.000960791 | [-0.03 0.03] | 0.950188624 | 0.978747744 |
| L_VLDL_L | SHBG-LpA_pPS | 4255 | -0.010743181 | [-0.04 0.02] | 0.484843617 | 0.744305799 |
| L_VLDL_L | Obesity_pPS | 4255 | 0.036855919 | [0.01 0.07] | 0.016522805 | 0.07895582 |
| L_VLDL_L | VAT_Neg_pPS | 4255 | 0.013642893 | [-0.02 0.04] | 0.375029572 | 0.655156372 |
| L_VLDL_L | Bilirubin_pPS | 4255 | -0.011251835 | [-0.04 0.02] | 0.46440737 | 0.727498869 |
| L_VLDL_L | Proinsulin_pPS | 4255 | 0.007146513 | [-0.02 0.04] | 0.642170152 | 0.83808647 |
| L_VLDL_L | Hyper_Insulin_pPS | 4255 | 0.007382145 | [-0.02 0.04] | 0.631236012 | 0.835456904 |
| L_VLDL_L | Beta_Cell_1_pPS | 4255 | 0.00613435 | [-0.02 0.04] | 0.690000446 | 0.858568103 |
| L_VLDL_L | Beta_Cell_2_pPS | 4255 | -0.004880948 | [-0.04 0.03] | 0.750972481 | 0.876790721 |
| L_VLDL_P | Cholesterol_pPS | 4255 | -0.080787438 | [-0.11 -0.05] | 1.43E-07 | 3.25E-06 |
| L_VLDL_P | Lipodystrophy_1_pPS | 4255 | 0.056380611 | [0.03 0.09] | 0.000243747 | 0.002993749 |
| L_VLDL_P | Lipodystrophy_2_pPS | 4255 | 0.005790366 | [-0.02 0.04] | 0.706553922 | 0.862512775 |
| L_VLDL_P | Liver-Lipid_pPS | 4255 | -0.052530052 | [-0.08 -0.02] | 0.000631241 | 0.006420748 |
| L_VLDL_P | ASAT_Pos_pPS | 4255 | 0.034026119 | [0. 0.06] | 0.026898142 | 0.113847952 |
| L_VLDL_P | ALP_Neg_pPS | 4255 | -0.001775882 | [-0.03 0.03] | 0.908075332 | 0.96224905 |
| L_VLDL_P | SHBG-LpA_pPS | 4255 | -0.010926207 | [-0.04 0.02] | 0.47743457 | 0.737518525 |
| L_VLDL_P | Obesity_pPS | 4255 | 0.036947857 | [0.01 0.07] | 0.016254891 | 0.078604569 |
| L_VLDL_P | VAT_Neg_pPS | 4255 | 0.013403692 | [-0.02 0.04] | 0.383461087 | 0.66173638 |
| L_VLDL_P | Bilirubin_pPS | 4255 | -0.01100611 | [-0.04 0.02] | 0.47421955 | 0.735233474 |
| L_VLDL_P | Proinsulin_pPS | 4255 | 0.007302677 | [-0.02 0.04] | 0.634914717 | 0.835456904 |
| L_VLDL_P | Hyper_Insulin_pPS | 4255 | 0.007617773 | [-0.02 0.04] | 0.620382107 | 0.831328634 |
| L_VLDL_P | Beta_Cell_1_pPS | 4255 | 0.006012114 | [-0.02 0.04] | 0.695866327 | 0.858892964 |
| L_VLDL_P | Beta_Cell_2_pPS | 4255 | -0.005035488 | [-0.04 0.03] | 0.743361134 | 0.873512116 |

**Table S9: Associations between pPSs and global metabolomic profile in SCAPIS**

**a) Spearman Correlation**

| Metabolomics | PPS | Samp | r | CI95% | p | fdr_p_adjusted |
|---|---|---|---|---|---|---|
| L_VLDL_CE | Beta_Cell_2_pPS | 4255 | 0.002103644 | [-0.03 0.03] | 0.891040612 | 0.948487102 |
| L_VLDL_CE | Total_GRS | 4255 | 0.037040958 | [0.01 0.07] | 0.015826046 | 0.069706094 |
| L_VLDL_FC | Cholesterol_pPS | 4255 | -0.081838677 | [-0.11 -0.05] | 9.39E-08 | 2.43E-06 |
| L_VLDL_FC | Lipodystrophy_1_pPS | 4255 | 0.064558504 | [0.03 0.09] | 2.57E-05 | 0.000411719 |
| L_VLDL_FC | Lipodystrophy_2_pPS | 4255 | 0.006296189 | [-0.02 0.04] | 0.681801299 | 0.838849838 |
| L_VLDL_FC | Liver-Lipid_pPS | 4255 | -0.052732124 | [-0.08 -0.02] | 0.000589804 | 0.005911819 |
| L_VLDL_FC | ASAT_Pos_pPS | 4255 | 0.044125829 | [0.01 0.07] | 0.004042381 | 0.023184243 |
| L_VLDL_FC | ALP_Neg_pPS | 4255 | 0.00409443 | [-0.03 0.03] | 0.78975651 | 0.899856387 |
| L_VLDL_FC | SHBG-LpA_pPS | 4255 | -0.002340466 | [-0.03 0.03] | 0.878864113 | 0.944681371 |
| L_VLDL_FC | Obesity_pPS | 4255 | 0.033071367 | [0. 0.06] | 0.03122703 | 0.122453346 |
| L_VLDL_FC | VAT_Neg_pPS | 4255 | 0.020031622 | [-0.01 0.05] | 0.192038452 | 0.464503229 |
| L_VLDL_FC | Bilirubin_pPS | 4255 | -0.009091778 | [-0.04 0.02] | 0.553809864 | 0.760607725 |
| L_VLDL_FC | Proinsulin_pPS | 4255 | 0.010846238 | [-0.02 0.04] | 0.479990229 | 0.716353047 |
| L_VLDL_FC | Hyper_Insulin_pPS | 4255 | 0.014985057 | [-0.02 0.05] | 0.329129625 | 0.610965216 |
| L_VLDL_FC | Beta_Cell_1_pPS | 4255 | 0.011948989 | [-0.02 0.04] | 0.43648917 | 0.690884659 |
| L_VLDL_FC | Beta_Cell_2_pPS | 4255 | -0.002243631 | [-0.03 0.03] | 0.883839702 | 0.945095197 |
| L_VLDL_FC | Total_GRS | 4255 | 0.038497688 | [0.01 0.07] | 0.012146297 | 0.055670529 |
| L_VLDL_L | Cholesterol_pPS | 4255 | -0.079314052 | [-0.11 -0.05] | 2.30E-07 | 5.68E-06 |
| L_VLDL_L | Lipodystrophy_1_pPS | 4255 | 0.065162334 | [0.04 0.1 ] | 2.16E-05 | 0.000353437 |
| L_VLDL_L | Lipodystrophy_2_pPS | 4255 | 0.009090205 | [-0.02 0.04] | 0.553878446 | 0.760607725 |
| L_VLDL_L | Liver-Lipid_pPS | 4255 | -0.051444208 | [-0.08 -0.02] | 0.000801659 | 0.007255522 |
| L_VLDL_L | ASAT_Pos_pPS | 4255 | 0.047190662 | [0.02 0.08] | 0.00210695 | 0.014531858 |
| L_VLDL_L | ALP_Neg_pPS | 4255 | 0.000269486 | [-0.03 0.03] | 0.98599889 | 0.991081359 |
| L_VLDL_L | SHBG-LpA_pPS | 4255 | -0.004168219 | [-0.03 0.03] | 0.786038831 | 0.898293123 |
| L_VLDL_L | Obesity_pPS | 4255 | 0.033902265 | [0. 0.06] | 0.027221941 | 0.109757637 |
| L_VLDL_L | VAT_Neg_pPS | 4255 | 0.019046438 | [-0.01 0.05] | 0.214827557 | 0.496022723 |
| L_VLDL_L | Bilirubin_pPS | 4255 | -0.007463505 | [-0.04 0.02] | 0.626950407 | 0.803929814 |
| L_VLDL_L | Proinsulin_pPS | 4255 | 0.010744235 | [-0.02 0.04] | 0.484130082 | 0.717164725 |
| L_VLDL_L | Hyper_Insulin_pPS | 4255 | 0.017192076 | [-0.01 0.05] | 0.262877593 | 0.550119451 |
| L_VLDL_L | Beta_Cell_1_pPS | 4255 | 0.011654521 | [-0.02 0.04] | 0.447877726 | 0.693644564 |
| L_VLDL_L | Beta_Cell_2_pPS | 4255 | -0.002657361 | [-0.03 0.03] | 0.862615823 | 0.937517799 |
| L_VLDL_L | Total_GRS | 4255 | 0.039896561 | [0.01 0.07] | 0.009347841 | 0.045058692 |
| L_VLDL_P | Cholesterol_pPS | 4255 | -0.078692029 | [-0.11 -0.05] | 2.86E-07 | 6.98E-06 |
| L_VLDL_P | Lipodystrophy_1_pPS | 4255 | 0.06506271 | [0.04 0.09] | 2.22E-05 | 0.000361089 |
| L_VLDL_P | Lipodystrophy_2_pPS | 4255 | 0.00972457 | [-0.02 0.04] | 0.526557071 | 0.742580484 |
| L_VLDL_P | Liver-Lipid_pPS | 4255 | -0.051911999 | [-0.08 -0.02] | 0.000717653 | 0.006751607 |
| L_VLDL_P | ASAT_Pos_pPS | 4255 | 0.047441374 | [0.02 0.08] | 0.001994292 | 0.013934058 |
| L_VLDL_P | ALP_Neg_pPS | 4255 | -0.000533983 | [-0.03 0.03] | 0.972261149 | 0.983305352 |
| L_VLDL_P | SHBG-LpA_pPS | 4255 | -0.004351894 | [-0.03 0.03] | 0.776875689 | 0.89275053 |
| L_VLDL_P | Obesity_pPS | 4255 | 0.034057936 | [0. 0.06] | 0.026523044 | 0.107343263 |
| L_VLDL_P | VAT_Neg_pPS | 4255 | 0.018843144 | [-0.01 0.05] | 0.219763247 | 0.500648222 |
| L_VLDL_P | Bilirubin_pPS | 4255 | -0.007215761 | [-0.04 0.02] | 0.638433521 | 0.811859149 |
| L_VLDL_P | Proinsulin_pPS | 4255 | 0.010714649 | [-0.02 0.04] | 0.485334444 | 0.71746546 |
| L_VLDL_P | Hyper_Insulin_pPS | 4255 | 0.017417568 | [-0.01 0.05] | 0.256667555 | 0.540286463 |
| L_VLDL_P | Beta_Cell_1_pPS | 4255 | 0.011479119 | [-0.02 0.04] | 0.454740831 | 0.696320363 |
| L_VLDL_P | Beta_Cell_2_pPS | 4255 | -0.002845904 | [-0.03 0.03] | 0.852975605 | 0.932534492 |
| L_VLDL_P | Total_GRS | 4255 | 0.040097464 | [0.01 0.07] | 0.008997158 | 0.043662678 |
| L_VLDL_PL | Cholesterol_pPS | 4255 | -0.083672044 | [-0.11 -0.05] | 4.81E-08 | 1.31E-06 |
| L_VLDL_PL | Lipodystrophy_1_pPS | 4255 | 0.066385585 | [0.04 0.1 ] | 1.51E-05 | 0.000258504 |
| L_VLDL_PL | Lipodystrophy_2_pPS | 4255 | 0.008063759 | [-0.02 0.04] | 0.599504324 | 0.788918266 |
| L_VLDL_PL | Liver-Lipid_pPS | 4255 | -0.05009733 | [-0.08 -0.02] | 0.001097179 | 0.009017049 |
| L_VLDL_PL | ASAT_Pos_pPS | 4255 | 0.046159009 | [0.02 0.08] | 0.002634606 | 0.017229362 |
| L_VLDL_PL | ALP_Neg_pPS | 4255 | 0.002435056 | [-0.03 0.03] | 0.874008517 | 0.942282701 |
| L_VLDL_PL | SHBG-LpA_pPS | 4255 | -0.003112345 | [-0.03 0.03] | 0.839390129 | 0.926178923 |
| L_VLDL_PL | Obesity_pPS | 4255 | 0.032879936 | [0. 0.06] | 0.032218306 | 0.125423352 |
| L_VLDL_PL | VAT_Neg_pPS | 4255 | 0.019016383 | [-0.01 0.05] | 0.215552159 | 0.496093758 |
| L_VLDL_PL | Bilirubin_pPS | 4255 | -0.007892014 | [-0.04 0.02] | 0.607301637 | 0.792855759 |
| L_VLDL_PL | Proinsulin_pPS | 4255 | 0.010918022 | [-0.02 0.04] | 0.477088445 | 0.714440991 |
| L_VLDL_PL | Hyper_Insulin_pPS | 4255 | 0.016464576 | [-0.01 0.05] | 0.283616096 | 0.571763651 |
| L_VLDL_PL | Beta_Cell_1_pPS | 4255 | 0.01226279 | [-0.02 0.04] | 0.424538555 | 0.68520331 |
| L_VLDL_PL | Beta_Cell_2_pPS | 4255 | -0.002243406 | [-0.03 0.03] | 0.883851266 | 0.945095197 |
| L_VLDL_PL | Total_GRS | 4255 | 0.039623709 | [0.01 0.07] | 0.00984362 | 0.046610518 |
| L_VLDL_TG | Cholesterol_pPS | 4255 | -0.075823781 | [-0.11 -0.05] | 7.63E-07 | 1.78E-05 |
| L_VLDL_TG | Lipodystrophy_1_pPS | 4255 | 0.064468542 | [0.03 0.09] | 2.64E-05 | 0.000419427 |
| L_VLDL_TG | Lipodystrophy_2_pPS | 4255 | 0.011932274 | [-0.02 0.04] | 0.437131106 | 0.690884659 |
| L_VLDL_TG | Liver-Lipid_pPS | 4255 | -0.054435945 | [-0.08 -0.02] | 0.000388978 | 0.00419275 |
| L_VLDL_TG | ASAT_Pos_pPS | 4255 | 0.04770058 | [0.02 0.08] | 0.001883651 | 0.013595948 |
| L_VLDL_TG | ALP_Neg_pPS | 4255 | -0.00332651 | [-0.03 0.03] | 0.828504505 | 0.92223257 |
| L_VLDL_TG | SHBG-LpA_pPS | 4255 | -0.00498245 | [-0.03 0.03] | 0.745593742 | 0.8772894 |
| L_VLDL_TG | Obesity_pPS | 4255 | 0.035024706 | [0. 0.07] | 0.022520172 | 0.094347206 |
| L_VLDL_TG | VAT_Neg_pPS | 4255 | 0.018090606 | [-0.01 0.05] | 0.238740499 | 0.520954599 |
| L_VLDL_TG | Bilirubin_pPS | 4255 | -0.006283699 | [-0.04 0.02] | 0.682398045 | 0.838849838 |
| L_VLDL_TG | Proinsulin_pPS | 4255 | 0.010686457 | [-0.02 0.04] | 0.486483601 | 0.717611897 |
| L_VLDL_TG | Hyper_Insulin_pPS | 4255 | 0.017868235 | [-0.01 0.05] | 0.244563176 | 0.525480742 |
| L_VLDL_TG | Beta_Cell_1_pPS | 4255 | 0.010727855 | [-0.02 0.04] | 0.484796667 | 0.717164725 |
| L_VLDL_TG | Beta_Cell_2_pPS | 4255 | -0.003912592 | [-0.03 0.03] | 0.79888877 | 0.905238463 |
| L_VLDL_TG | Total_GRS | 4255 | 0.040693786 | [0.01 0.07] | 0.008024393 | 0.039843342 |
| LA | Cholesterol_pPS | 4223 | -0.155714122 | [-0.19 -0.13] | 2.88E-24 | 1.25E-21 |
| LA | Lipodystrophy_1_pPS | 4223 | 0.029865008 | [-0. 0.06] | 0.052639877 | 0.181240026 |
| LA | Lipodystrophy_2_pPS | 4223 | -0.052759793 | [-0.08 -0.02] | 0.000614643 | 0.00604775 |
| LA | Liver-Lipid_pPS | 4223 | -0.046007405 | [-0.08 -0.02] | 0.002824517 | 0.018303894 |
| LA | ASAT_Pos_pPS | 4223 | -0.025546431 | [-0.06 0. ] | 0.097409767 | 0.297217569 |
| LA | ALP_Neg_pPS | 4223 | 0.036699584 | [0.01 0.07] | 0.017237018 | 0.074844948 |
| LA | SHBG-LpA_pPS | 4223 | 0.016255747 | [-0.01 0.05] | 0.29159551 | 0.577755652 |
| LA | Obesity_pPS | 4223 | 0.003790068 | [-0.03 0.03] | 0.805779926 | 0.909683127 |
| LA | VAT_Neg_pPS | 4223 | -0.001603699 | [-0.03 0.03] | 0.917140155 | 0.957786011 |
| LA | Bilirubin_pPS | 4223 | -0.020138668 | [-0.05 0.01] | 0.191352899 | 0.464503229 |
| LA | Proinsulin_pPS | 4223 | 0.013294061 | [-0.02 0.04] | 0.388432716 | 0.657347349 |
| LA | Hyper_Insulin_pPS | 4223 | -0.027164147 | [-0.06 0. ] | 0.077977492 | 0.247501258 |
| LA | Beta_Cell_1_pPS | 4223 | 0.022813681 | [-0.01 0.05] | 0.138824291 | 0.3817668 |
| LA | Beta_Cell_2_pPS | 4223 | 0.023751579 | [-0.01 0.05] | 0.123304022 | 0.354065765 |
| LA | Total_GRS | 4223 | -0.007657087 | [-0.04 0.02] | 0.619368797 | 0.800328957 |
| Lac | Cholesterol_pPS | 4255 | -0.00434013 | [-0.03 0.03] | 0.777462933 | 0.89275053 |
| Lac | Lipodystrophy_1_pPS | 4255 | 0.034641484 | [0. 0.06] | 0.024039321 | 0.099162199 |

**b) Partial Correlation**

| Metabolomics | PPS | Samp | r | CI95% | p | fdr_p_adjusted |
|---|---|---|---|---|---|---|
| L_VLDL_PL | Cholesterol_pPS | 4255 | -0.085655945 | [-0.12 -0.06] | 2.41E-08 | 6.35E-07 |
| L_VLDL_PL | Lipodystrophy_1_pPS | 4255 | 0.058006669 | [0.03 0.09] | 0.000160183 | 0.002152255 |
| L_VLDL_PL | Lipodystrophy_2_pPS | 4255 | 0.004344714 | [-0.03 0.03] | 0.777566849 | 0.891327566 |
| L_VLDL_PL | Liver-Lipid_pPS | 4255 | -0.050680896 | [-0.08 -0.02] | 0.000976012 | 0.00857007 |
| L_VLDL_PL | ASAT_Pos_pPS | 4255 | 0.032675455 | [0. 0.06] | 0.033578243 | 0.133645413 |
| L_VLDL_PL | ALP_Neg_pPS | 4255 | 0.001213855 | [-0.03 0.03] | 0.937093094 | 0.978170683 |
| L_VLDL_PL | SHBG-LpA_pPS | 4255 | -0.009751917 | [-0.04 0.02] | 0.526030429 | 0.776631946 |
| L_VLDL_PL | Obesity_pPS | 4255 | 0.036241353 | [0.01 0.07] | 0.018415354 | 0.084752963 |
| L_VLDL_PL | VAT_Neg_pPS | 4255 | 0.013458482 | [-0.02 0.04] | 0.38151965 | 0.661657399 |
| L_VLDL_PL | Bilirubin_pPS | 4255 | -0.011669114 | [-0.04 0.02] | 0.448006547 | 0.709019056 |
| L_VLDL_PL | Proinsulin_pPS | 4255 | 0.006868129 | [-0.02 0.04] | 0.655188664 | 0.844628798 |
| L_VLDL_PL | Hyper_Insulin_pPS | 4255 | 0.00663087 | [-0.02 0.04] | 0.666367448 | 0.849453588 |
| L_VLDL_PL | Beta_Cell_1_pPS | 4255 | 0.006637028 | [-0.02 0.04] | 0.666076368 | 0.849453588 |
| L_VLDL_PL | Beta_Cell_2_pPS | 4255 | -0.004564286 | [-0.03 0.03] | 0.766643725 | 0.886155161 |
| L_VLDL_TG | Cholesterol_pPS | 4255 | -0.078066483 | [-0.11 -0.05] | 3.70E-07 | 8.15E-06 |
| L_VLDL_TG | Lipodystrophy_1_pPS | 4255 | 0.055679361 | [0.03 0.09] | 0.000291161 | 0.003511473 |
| L_VLDL_TG | Lipodystrophy_2_pPS | 4255 | 0.008061765 | [-0.02 0.04] | 0.600154553 | 0.821264125 |
| L_VLDL_TG | Liver-Lipid_pPS | 4255 | -0.055040288 | [-0.09 -0.02] | 0.000341769 | 0.003970654 |
| L_VLDL_TG | ASAT_Pos_pPS | 4255 | 0.034153932 | [0. 0.06] | 0.026329952 | 0.112393527 |
| L_VLDL_TG | ALP_Neg_pPS | 4255 | -0.004585772 | [-0.03 0.03] | 0.76557728 | 0.885433688 |
| L_VLDL_TG | SHBG-LpA_pPS | 4255 | -0.011523953 | [-0.04 0.02] | 0.453674343 | 0.716290249 |
| L_VLDL_TG | Obesity_pPS | 4255 | 0.03767479 | [0.01 0.07] | 0.014267553 | 0.071081259 |
| L_VLDL_TG | VAT_Neg_pPS | 4255 | 0.012614704 | [-0.02 0.04] | 0.41208445 | 0.6835071 |
| L_VLDL_TG | Bilirubin_pPS | 4255 | -0.010074348 | [-0.04 0.02] | 0.512440108 | 0.767666412 |
| L_VLDL_TG | Proinsulin_pPS | 4255 | 0.007953464 | [-0.02 0.04] | 0.605061022 | 0.824474694 |
| L_VLDL_TG | Hyper_Insulin_pPS | 4255 | 0.008168354 | [-0.02 0.04] | 0.595343275 | 0.818035166 |
| L_VLDL_TG | Beta_Cell_1_pPS | 4255 | 0.005573424 | [-0.02 0.04] | 0.717066161 | 0.866099998 |
| L_VLDL_TG | Beta_Cell_2_pPS | 4255 | -0.005979568 | [-0.04 0.02] | 0.697431205 | 0.859078268 |
| LA | Cholesterol_pPS | 4223 | -0.154650574 | [-0.18 -0.12] | 6.92E-24 | 3.20E-21 |
| LA | Lipodystrophy_1_pPS | 4223 | 0.038690503 | [0.01 0.07] | 0.012175167 | 0.062821352 |
| LA | Lipodystrophy_2_pPS | 4223 | -0.043933137 | [-0.07 -0.01] | 0.00441276 | 0.028406254 |
| LA | Liver-Lipid_pPS | 4223 | -0.043783121 | [-0.07 -0.01] | 0.004549533 | 0.029006894 |
| LA | ASAT_Pos_pPS | 4223 | -0.027046727 | [-0.06 0. ] | 0.079737793 | 0.270110088 |
| LA | ALP_Neg_pPS | 4223 | 0.038998883 | [0.01 0.07] | 0.011503383 | 0.059973366 |
| LA | SHBG-LpA_pPS | 4223 | 0.013801957 | [-0.02 0.04] | 0.371304586 | 0.653802291 |
| LA | Obesity_pPS | 4223 | 0.018412201 | [-0.01 0.05] | 0.232983489 | 0.52883554 |
| LA | VAT_Neg_pPS | 4223 | -0.003321949 | [-0.03 0.03] | 0.829633099 | 0.921201034 |
| LA | Bilirubin_pPS | 4223 | -0.02231169 | [-0.05 0.01] | 0.148354587 | 0.42368885 |
| LA | Proinsulin_pPS | 4223 | 8.27E-05 | [-0.03 0.03] | 0.995725712 | 0.996223326 |
| LA | Hyper_Insulin_pPS | 4223 | -0.024669293 | [-0.05 0.01] | 0.110011958 | 0.343926372 |
| LA | Beta_Cell_1_pPS | 4223 | 0.014899817 | [-0.02 0.05] | 0.334467361 | 0.630462857 |
| LA | Beta_Cell_2_pPS | 4223 | 0.022197469 | [-0.01 0.05] | 0.150443495 | 0.426611725 |
| Lac | Cholesterol_pPS | 4255 | -0.002942904 | [-0.03 0.03] | 0.848254734 | 0.932567807 |
| Lac | Lipodystrophy_1_pPS | 4255 | 0.035752966 | [0.01 0.07] | 0.020051989 | 0.090211421 |
| Lac | Lipodystrophy_2_pPS | 4255 | -0.009526661 | [-0.04 0.02] | 0.535632991 | 0.781586915 |
| Lac | Liver-Lipid_pPS | 4255 | -0.039025459 | [-0.07 -0.01] | 0.011137051 | 0.058520675 |
| Lac | ASAT_Pos_pPS | 4255 | 0.000444139 | [-0.03 0.03] | 0.976962252 | 0.991343605 |
| Lac | ALP_Neg_pPS | 4255 | 0.020700375 | [-0.01 0.05] | 0.178281039 | 0.455834788 |
| Lac | SHBG-LpA_pPS | 4255 | -0.022050879 | [-0.05 0.01] | 0.151599237 | 0.427467143 |
| Lac | Obesity_pPS | 4255 | -0.005269175 | [-0.04 0.02] | 0.731899333 | 0.869076195 |
| Lac | VAT_Neg_pPS | 4255 | 0.007427364 | [-0.02 0.04] | 0.629146747 | 0.835246543 |
| Lac | Bilirubin_pPS | 4255 | -0.024616567 | [-0.05 0.01] | 0.109422833 | 0.343900332 |
| Lac | Proinsulin_pPS | 4255 | 0.007360789 | [-0.02 0.04] | 0.632223696 | 0.835456904 |
| Lac | Hyper_Insulin_pPS | 4255 | 0.006061636 | [-0.02 0.04] | 0.693487638 | 0.858603742 |
| Lac | Beta_Cell_1_pPS | 4255 | -0.006809375 | [-0.04 0.02] | 0.657949851 | 0.844628798 |
| Lac | Beta_Cell_2_pPS | 4255 | -0.018591486 | [-0.05 0.01] | 0.226697987 | 0.521665942 |
| LDL_C | Cholesterol_pPS | 4254 | -0.152364411 | [-0.18 -0.12] | 2.20E-23 | 4.73E-21 |
| LDL_C | Lipodystrophy_1_pPS | 4254 | 0.043108049 | [0.01 0.07] | 0.005050049 | 0.030805894 |
| LDL_C | Lipodystrophy_2_pPS | 4254 | -0.03921673 | [-0.07 -0.01] | 0.010756204 | 0.057247273 |
| LDL_C | Liver-Lipid_pPS | 4254 | 0.011641531 | [-0.02 0.04] | 0.449134054 | 0.710242004 |
| LDL_C | ASAT_Pos_pPS | 4254 | -0.018597629 | [-0.05 0.01] | 0.226599445 | 0.521665942 |
| LDL_C | ALP_Neg_pPS | 4254 | 0.050769667 | [0.02 0.08] | 0.000957437 | 0.008545465 |
| LDL_C | SHBG-LpA_pPS | 4254 | 0.014955501 | [-0.02 0.05] | 0.330884162 | 0.62795236 |
| LDL_C | Obesity_pPS | 4254 | 0.006812328 | [-0.02 0.04] | 0.65784889 | 0.844628798 |
| LDL_C | VAT_Neg_pPS | 4254 | 0.008979624 | [-0.02 0.04] | 0.559360966 | 0.797755947 |
| LDL_C | Bilirubin_pPS | 4254 | -0.014585449 | [-0.04 0.02] | 0.342990538 | 0.636982428 |
| LDL_C | Proinsulin_pPS | 4254 | -0.022621809 | [-0.05 0.01] | 0.141327151 | 0.410988061 |
| LDL_C | Hyper_Insulin_pPS | 4254 | -0.022154209 | [-0.05 0.01] | 0.14973843 | 0.425214661 |
| LDL_C | Beta_Cell_1_pPS | 4254 | 0.015986794 | [-0.01 0.05] | 0.298622252 | 0.596053587 |
| LDL_C | Beta_Cell_2_pPS | 4254 | 0.023066486 | [-0.01 0.05] | 0.133669528 | 0.39645392 |
| LDL_D | Cholesterol_pPS | 4254 | 0.059906279 | [0.03 0.09] | 9.70E-05 | 0.001426225 |
| LDL_D | Lipodystrophy_1_pPS | 4254 | -0.002682646 | [-0.03 0.03] | 0.861548532 | 0.940272439 |
| LDL_D | Lipodystrophy_2_pPS | 4254 | -0.012566217 | [-0.04 0.02] | 0.413939094 | 0.685447531 |
| LDL_D | Liver-Lipid_pPS | 4254 | 0.023618952 | [-0.01 0.05] | 0.124607407 | 0.376265503 |
| LDL_D | ASAT_Pos_pPS | 4254 | 0.017466527 | [-0.01 0.05] | 0.256118806 | 0.55296819 |
| LDL_D | ALP_Neg_pPS | 4254 | 0.002611912 | [-0.03 0.03] | 0.865163742 | 0.94287306 |
| LDL_D | SHBG-LpA_pPS | 4254 | -0.003408748 | [-0.03 0.03] | 0.824618571 | 0.91920177 |
| LDL_D | Obesity_pPS | 4254 | -0.020169783 | [-0.05 0.01] | 0.189721974 | 0.473428767 |
| LDL_D | VAT_Neg_pPS | 4254 | 0.010551426 | [-0.02 0.04] | 0.49272351 | 0.752427511 |
| LDL_D | Bilirubin_pPS | 4254 | -1.64E-05 | [-0.03 0.03] | 0.999150638 | 0.999150638 |
| LDL_D | Proinsulin_pPS | 4254 | 0.030750357 | [0. 0.06] | 0.045543384 | 0.171386945 |
| LDL_D | Hyper_Insulin_pPS | 4254 | 0.019683498 | [-0.01 0.05] | 0.200623065 | 0.486527589 |
| LDL_D | Beta_Cell_1_pPS | 4254 | -0.04002165 | [-0.07 -0.01] | 0.009243573 | 0.050839653 |
| LDL_D | Beta_Cell_2_pPS | 4254 | -0.005974004 | [-0.04 0.02] | 0.697732899 | 0.859078268 |
| LDL_TG | Cholesterol_pPS | 4254 | -0.113546745 | [-0.14 -0.08] | 1.30E-13 | 5.80E-12 |
| LDL_TG | Lipodystrophy_1_pPS | 4254 | 0.062865118 | [0.03 0.09] | 4.30E-05 | 0.000757137 |
| LDL_TG | Lipodystrophy_2_pPS | 4254 | -0.014932492 | [-0.05 0.02] | 0.331628756 | 0.62795236 |
| LDL_TG | Liver-Lipid_pPS | 4254 | -0.018858793 | [-0.05 0.01] | 0.220142915 | 0.513666802 |
| LDL_TG | ASAT_Pos_pPS | 4254 | -0.018690622 | [-0.05 0.01] | 0.224285151 | 0.519697769 |
| LDL_TG | ALP_Neg_pPS | 4254 | 0.051490134 | [0.02 0.08] | 0.000809065 | 0.007498835 |
| LDL_TG | SHBG-LpA_pPS | 4254 | 0.005478573 | [-0.02 0.04] | 0.721710935 | 0.866319132 |
| LDL_TG | Obesity_pPS | 4254 | 0.017982058 | [-0.01 0.05] | 0.242348932 | 0.540176565 |
| LDL_TG | VAT_Neg_pPS | 4254 | -0.001862465 | [-0.03 0.03] | 0.903626242 | 0.96224905 |
| LDL_TG | Bilirubin_pPS | 4254 | -0.00663657 | [-0.04 0.02] | 0.666135134 | 0.849453588 |

**Table S9: Associations between pPSs and global metabolomic profile in SCAPIS**

**a) Spearman Correlation**

| Metabolomics | PPS | Samp | r | CI95% | p | fdr_p_adjusted |
|---|---|---|---|---|---|---|
| Lac | Lipodystrophy_2_pPS | 4255 | -0.007536646 | [-0.04 0.02] | 0.623577281 | 0.802865107 |
| Lac | Liver-Lipid_pPS | 4255 | -0.038192099 | [-0.07 -0.01] | 0.012848411 | 0.058249914 |
| Lac | ASAT_Pos_pPS | 4255 | 0.005135073 | [-0.02 0.04] | 0.738082518 | 0.874278301 |
| Lac | ALP_Neg_pPS | 4255 | 0.021844333 | [-0.01 0.05] | 0.154836616 | 0.407130084 |
| Lac | SHBG-LpA_pPS | 4255 | -0.01927433 | [-0.05 0.01] | 0.20939019 | 0.488466046 |
| Lac | Obesity_pPS | 4255 | -0.003378116 | [-0.03 0.03] | 0.825886285 | 0.921344342 |
| Lac | VAT_Neg_pPS | 4255 | 0.003815204 | [-0.03 0.03] | 0.803791163 | 0.908872981 |
| Lac | Bilirubin_pPS | 4255 | -0.023289065 | [-0.05 0.01] | 0.129324975 | 0.36526936 |
| Lac | Proinsulin_pPS | 4255 | 0.002984258 | [-0.03 0.03] | 0.84591537 | 0.92810697 |
| Lac | Hyper_Insulin_pPS | 4255 | 0.008416238 | [-0.02 0.04] | 0.583645414 | 0.778556849 |
| Lac | Beta_Cell_1_pPS | 4255 | -0.010166129 | [-0.04 0.02] | 0.507954258 | 0.730759144 |
| Lac | Beta_Cell_2_pPS | 4255 | -0.021084226 | [-0.05 0.01] | 0.169709175 | 0.431822279 |
| Lac | Total_GRS | 4255 | 0.004461793 | [-0.03 0.03] | 0.771395903 | 0.891027884 |
| LDL_C | Cholesterol_pPS | 4254 | -0.153993054 | [-0.18 -0.12] | 6.31E-24 | 1.50E-21 |
| LDL_C | Lipodystrophy_1_pPS | 4254 | 0.037590185 | [0.01 0.07] | 0.01434857 | 0.063721909 |
| LDL_C | Lipodystrophy_2_pPS | 4254 | -0.044029959 | [-0.07 -0.01] | 0.004127556 | 0.02354683 |
| LDL_C | Liver-Lipid_pPS | 4254 | 0.009121113 | [-0.02 0.04] | 0.552578205 | 0.760607725 |
| LDL_C | ASAT_Pos_pPS | 4254 | -0.014990235 | [-0.05 0.02] | 0.329019547 | 0.610965216 |
| LDL_C | ALP_Neg_pPS | 4254 | 0.04716263 | [0.02 0.08] | 0.002122478 | 0.014592035 |
| LDL_C | SHBG-LpA_pPS | 4254 | 0.017635798 | [-0.01 0.05] | 0.250811075 | 0.532136256 |
| LDL_C | Obesity_pPS | 4254 | -0.008640992 | [-0.04 0.02] | 0.573681438 | 0.772791937 |
| LDL_C | VAT_Neg_pPS | 4254 | 0.012155094 | [-0.02 0.04] | 0.428672736 | 0.68773599 |
| LDL_C | Bilirubin_pPS | 4254 | -0.013513749 | [-0.04 0.02] | 0.378891896 | 0.650178493 |
| LDL_C | Proinsulin_pPS | 4254 | -0.007553087 | [-0.04 0.02] | 0.622861161 | 0.80242474 |
| LDL_C | Hyper_Insulin_pPS | 4254 | -0.020285061 | [-0.05 0.01] | 0.186526198 | 0.458380551 |
| LDL_C | Beta_Cell_1_pPS | 4254 | 0.020725251 | [-0.01 0.05] | 0.17714563 | 0.445459996 |
| LDL_C | Beta_Cell_2_pPS | 4254 | 0.023636462 | [-0.01 0.05] | 0.123750805 | 0.354873631 |
| LDL_C | Total_GRS | 4254 | -0.003763164 | [-0.03 0.03] | 0.806436363 | 0.909945291 |
| LDL_D | Cholesterol_pPS | 4254 | 0.059395725 | [0.03 0.09] | 0.000108497 | 0.001491835 |
| LDL_D | Lipodystrophy_1_pPS | 4254 | 0.000674526 | [-0.03 0.03] | 0.964968697 | 0.982019059 |
| LDL_D | Lipodystrophy_2_pPS | 4254 | -0.004445692 | [-0.03 0.03] | 0.772224166 | 0.891027884 |
| LDL_D | Liver-Lipid_pPS | 4254 | 0.025101157 | [-0. 0.06] | 0.102126612 | 0.307239246 |
| LDL_D | ASAT_Pos_pPS | 4254 | 0.015700624 | [-0.01 0.05] | 0.306613713 | 0.592510283 |
| LDL_D | ALP_Neg_pPS | 4254 | 0.009220964 | [-0.02 0.04] | 0.548237654 | 0.760607725 |
| LDL_D | SHBG-LpA_pPS | 4254 | -0.002204309 | [-0.03 0.03] | 0.885874869 | 0.946315535 |
| LDL_D | Obesity_pPS | 4254 | -0.009422077 | [-0.04 0.02] | 0.539546692 | 0.753959384 |
| LDL_D | VAT_Neg_pPS | 4254 | 0.004850233 | [-0.03 0.03] | 0.752148591 | 0.881192614 |
| LDL_D | Bilirubin_pPS | 4254 | -0.001204048 | [-0.03 0.03] | 0.937512016 | 0.970084172 |
| LDL_D | Proinsulin_pPS | 4254 | 0.019262108 | [-0.01 0.05] | 0.209732974 | 0.488466046 |
| LDL_D | Hyper_Insulin_pPS | 4254 | 0.021862226 | [-0.01 0.05] | 0.154547451 | 0.407130084 |
| LDL_D | Beta_Cell_1_pPS | 4254 | -0.033340827 | [-0.06 -0. ] | 0.029895436 | 0.118780144 |
| LDL_D | Beta_Cell_2_pPS | 4254 | -0.009899451 | [-0.04 0.02] | 0.519197248 | 0.736667707 |
| LDL_D | Total_GRS | 4254 | 0.001676911 | [-0.03 0.03] | 0.913054808 | 0.955833364 |
| LDL_TG | Cholesterol_pPS | 4254 | -0.112611992 | [-0.14 -0.08] | 1.90E-13 | 9.05E-12 |
| LDL_TG | Lipodystrophy_1_pPS | 4254 | 0.059559519 | [0.03 0.09] | 0.000103835 | 0.00147325 |
| LDL_TG | Lipodystrophy_2_pPS | 4254 | -0.01843372 | [-0.05 0.01] | 0.230004168 | 0.513581257 |
| LDL_TG | Liver-Lipid_pPS | 4254 | -0.019210734 | [-0.05 0.01] | 0.21095125 | 0.490238819 |
| LDL_TG | ASAT_Pos_pPS | 4254 | -0.009176973 | [-0.04 0.02] | 0.550147898 | 0.760607725 |
| LDL_TG | ALP_Neg_pPS | 4254 | 0.052287601 | [0.02 0.08] | 0.000657168 | 0.0063784 |
| LDL_TG | SHBG-LpA_pPS | 4254 | 0.009432665 | [-0.02 0.04] | 0.539091028 | 0.753813726 |
| LDL_TG | Obesity_pPS | 4254 | 0.009813691 | [-0.02 0.04] | 0.522823622 | 0.739259504 |
| LDL_TG | VAT_Neg_pPS | 4254 | 0.001198389 | [-0.03 0.03] | 0.937805075 | 0.970084172 |
| LDL_TG | Bilirubin_pPS | 4254 | -0.005193092 | [-0.04 0.02] | 0.735263733 | 0.873878512 |
| LDL_TG | Proinsulin_pPS | 4254 | 0.010816613 | [-0.02 0.04] | 0.481242303 | 0.716353047 |
| LDL_TG | Hyper_Insulin_pPS | 4254 | -0.007993132 | [-0.04 0.02] | 0.602748123 | 0.791256366 |
| LDL_TG | Beta_Cell_1_pPS | 4254 | 0.003373793 | [-0.03 0.03] | 0.826125735 | 0.921344342 |
| LDL_TG | Beta_Cell_2_pPS | 4254 | 0.011573986 | [-0.02 0.04] | 0.451074941 | 0.694584169 |
| LDL_TG | Total_GRS | 4254 | 0.017012523 | [-0.01 0.05] | 0.267952484 | 0.553183906 |
| Leu | Cholesterol_pPS | 4254 | -0.027543979 | [-0.06 0. ] | 0.072850118 | 0.2339274 |
| Leu | Lipodystrophy_1_pPS | 4254 | 0.035304266 | [0.01 0.07] | 0.021480618 | 0.090344954 |
| Leu | Lipodystrophy_2_pPS | 4254 | 0.012495321 | [-0.02 0.04] | 0.415862593 | 0.677830746 |
| Leu | Liver-Lipid_pPS | 4254 | -0.029184804 | [-0.06 0. ] | 0.05734531 | 0.193379811 |
| Leu | ASAT_Pos_pPS | 4254 | 0.002520903 | [-0.03 0.03] | 0.869621014 | 0.940757973 |
| Leu | ALP_Neg_pPS | 4254 | 0.002181492 | [-0.03 0.03] | 0.887048185 | 0.946626048 |
| Leu | SHBG-LpA_pPS | 4254 | -0.002560929 | [-0.03 0.03] | 0.86756987 | 0.940757973 |
| Leu | Obesity_pPS | 4254 | 0.031179781 | [0. 0.06] | 0.042290086 | 0.154790985 |
| Leu | VAT_Neg_pPS | 4254 | 0.012240951 | [-0.02 0.04] | 0.425418607 | 0.686107453 |
| Leu | Bilirubin_pPS | 4254 | -0.0070495 | [-0.04 0.02] | 0.646227816 | 0.817402268 |
| Leu | Proinsulin_pPS | 4254 | 0.005523677 | [-0.02 0.04] | 0.719103418 | 0.865374699 |
| Leu | Hyper_Insulin_pPS | 4254 | 0.012521904 | [-0.02 0.04] | 0.414871323 | 0.677760082 |
| Leu | Beta_Cell_1_pPS | 4254 | 0.039151019 | [0.01 0.07] | 0.010767441 | 0.049883717 |
| Leu | Beta_Cell_2_pPS | 4254 | 0.008348206 | [-0.02 0.04] | 0.586735178 | 0.779730867 |
| Leu | Total_GRS | 4254 | 0.048013854 | [0.02 0.08] | 0.00175967 | 0.01292441 |
| M_HDL_C | Cholesterol_pPS | 4255 | 0.049909269 | [0.02 0.08] | 0.00114566 | 0.009343882 |
| M_HDL_C | Lipodystrophy_1_pPS | 4255 | -0.061725171 | [-0.09 -0.03] | 5.74E-05 | 0.000882187 |
| M_HDL_C | Lipodystrophy_2_pPS | 4255 | -0.027241051 | [-0.06 0. ] | 0.076023036 | 0.242662815 |
| M_HDL_C | Liver-Lipid_pPS | 4255 | -0.050867238 | [-0.08 -0.02] | 0.000917823 | 0.007854666 |
| M_HDL_C | ASAT_Pos_pPS | 4255 | -0.037875871 | [-0.07 -0.01] | 0.013612542 | 0.061213586 |
| M_HDL_C | ALP_Neg_pPS | 4255 | 0.00370868 | [-0.03 0.03] | 0.80916233 | 0.911104041 |
| M_HDL_C | SHBG-LpA_pPS | 4255 | -0.002504556 | [-0.03 0.03] | 0.870443856 | 0.940757973 |
| M_HDL_C | Obesity_pPS | 4255 | -0.020652056 | [-0.05 0.01] | 0.178629379 | 0.447805525 |
| M_HDL_C | VAT_Neg_pPS | 4255 | -0.05086117 | [-0.08 -0.02] | 0.000919124 | 0.007854666 |
| M_HDL_C | Bilirubin_pPS | 4255 | 0.014202855 | [-0.02 0.04] | 0.355005392 | 0.634858982 |
| M_HDL_C | Proinsulin_pPS | 4255 | 0.000439819 | [-0.03 0.03] | 0.977151188 | 0.985883959 |
| M_HDL_C | Hyper_Insulin_pPS | 4255 | -0.008363709 | [-0.04 0.02] | 0.585996381 | 0.779730867 |
| M_HDL_C | Beta_Cell_1_pPS | 4255 | 0.007983874 | [-0.02 0.04] | 0.603125482 | 0.791256366 |
| M_HDL_C | Beta_Cell_2_pPS | 4255 | -0.001714594 | [-0.03 0.03] | 0.911098607 | 0.954725626 |
| M_HDL_C | Total_GRS | 4255 | -0.032653086 | [-0.06 -0. ] | 0.033427777 | 0.128499249 |
| M_HDL_CE | Cholesterol_pPS | 4255 | 0.051810711 | [0.02 0.08] | 0.000735117 | 0.006855765 |
| M_HDL_CE | Lipodystrophy_1_pPS | 4255 | -0.061712845 | [-0.09 -0.03] | 5.76E-05 | 0.000882187 |
| M_HDL_CE | Lipodystrophy_2_pPS | 4255 | -0.025715868 | [-0.06 0. ] | 0.093959413 | 0.288743469 |
| M_HDL_CE | Liver-Lipid_pPS | 4255 | -0.052372802 | [-0.08 -0.02] | 0.00064296 | 0.006268864 |
| M_HDL_CE | ASAT_Pos_pPS | 4255 | -0.036791373 | [-0.07 -0.01] | 0.016546475 | 0.072285516 |
| M_HDL_CE | ALP_Neg_pPS | 4255 | 0.000766755 | [-0.03 0.03] | 0.960177828 | 0.981183397 |

**b) Partial Correlation**

| Metabolomics | PPS | Samp | r | CI95% | p | fdr_p_adjusted |
|---|---|---|---|---|---|---|
| LDL_TG | Proinsulin_pPS | 4254 | 0.000851713 | [-0.03 0.03] | 0.95584273 | 0.981227298 |
| LDL_TG | Hyper_Insulin_pPS | 4254 | -0.012642653 | [-0.04 0.02] | 0.411104706 | 0.682580842 |
| LDL_TG | Beta_Cell_1_pPS | 4254 | -0.00249246 | [-0.03 0.03] | 0.871275209 | 0.946048963 |
| LDL_TG | Beta_Cell_2_pPS | 4254 | 0.012606189 | [-0.02 0.04] | 0.412455427 | 0.683556097 |
| Leu | Cholesterol_pPS | 4254 | -0.029823909 | [-0.06 0. ] | 0.052461226 | 0.194135629 |
| Leu | Lipodystrophy_1_pPS | 4254 | 0.032366353 | [0. 0.06] | 0.03531414 | 0.138897657 |
| Leu | Lipodystrophy_2_pPS | 4254 | 0.013176275 | [-0.02 0.04] | 0.391639954 | 0.670712736 |
| Leu | Liver-Lipid_pPS | 4254 | -0.031123176 | [-0.06 -0. ] | 0.042984991 | 0.163294026 |
| Leu | ASAT_Pos_pPS | 4254 | -0.007349835 | [-0.04 0.02] | 0.632770834 | 0.835456904 |
| Leu | ALP_Neg_pPS | 4254 | 0.003256108 | [-0.03 0.03] | 0.832352673 | 0.922174904 |
| Leu | SHBG-LpA_pPS | 4254 | -0.005488658 | [-0.04 0.02] | 0.721219994 | 0.866319132 |
| Leu | Obesity_pPS | 4254 | 0.032936298 | [0. 0.06] | 0.032207935 | 0.12896057 |
| Leu | VAT_Neg_pPS | 4254 | 0.004282956 | [-0.03 0.03] | 0.780672452 | 0.891560895 |
| Leu | Bilirubin_pPS | 4254 | -0.009857359 | [-0.04 0.02] | 0.521615192 | 0.773759954 |
| Leu | Proinsulin_pPS | 4254 | -0.009034744 | [-0.04 0.02] | 0.556952171 | 0.796929123 |
| Leu | Hyper_Insulin_pPS | 4254 | 0.007325887 | [-0.02 0.04] | 0.633879515 | 0.835456904 |
| Leu | Beta_Cell_1_pPS | 4254 | 0.038528705 | [0.01 0.07] | 0.012219519 | 0.062888115 |
| Leu | Beta_Cell_2_pPS | 4254 | -0.000825932 | [-0.03 0.03] | 0.957178049 | 0.981227298 |
| M_HDL_C | Cholesterol_pPS | 4255 | 0.052517463 | [0.02 0.08] | 0.000633146 | 0.006420748 |
| M_HDL_C | Lipodystrophy_1_pPS | 4255 | -0.054771198 | [-0.08 -0.02] | 0.000365446 | 0.004169853 |
| M_HDL_C | Lipodystrophy_2_pPS | 4255 | -0.016150517 | [-0.05 0.01] | 0.293643337 | 0.590234901 |
| M_HDL_C | Liver-Lipid_pPS | 4255 | -0.050891436 | [-0.08 -0.02] | 0.000929411 | 0.008496257 |
| M_HDL_C | ASAT_Pos_pPS | 4255 | -0.025466595 | [-0.06 0. ] | 0.097704214 | 0.319614112 |
| M_HDL_C | ALP_Neg_pPS | 4255 | 0.00757862 | [-0.02 0.04] | 0.622179981 | 0.833180148 |
| M_HDL_C | SHBG-LpA_pPS | 4255 | 0.005946303 | [-0.02 0.04] | 0.699032032 | 0.859092773 |
| M_HDL_C | Obesity_pPS | 4255 | -0.022336304 | [-0.05 0.01] | 0.146370836 | 0.420206242 |
| M_HDL_C | VAT_Neg_pPS | 4255 | -0.046777612 | [-0.08 -0.02] | 0.002341467 | 0.017233888 |
| M_HDL_C | Bilirubin_pPS | 4255 | 0.015505955 | [-0.01 0.05] | 0.313336454 | 0.61536839 |
| M_HDL_C | Proinsulin_pPS | 4255 | -0.001142436 | [-0.03 0.03] | 0.940787308 | 0.978430443 |
| M_HDL_C | Hyper_Insulin_pPS | 4255 | 0.008025032 | [-0.02 0.04] | 0.601816707 | 0.822414367 |
| M_HDL_C | Beta_Cell_1_pPS | 4255 | 0.017264246 | [-0.01 0.05] | 0.261610972 | 0.558959622 |
| M_HDL_C | Beta_Cell_2_pPS | 4255 | -0.001517834 | [-0.03 0.03] | 0.921385633 | 0.96931899 |
| M_HDL_CE | Cholesterol_pPS | 4255 | 0.054441296 | [0.02 0.08] | 0.000396567 | 0.00441754 |
| M_HDL_CE | Lipodystrophy_1_pPS | 4255 | -0.055002235 | [-0.08 -0.02] | 0.000345027 | 0.003970654 |
| M_HDL_CE | Lipodystrophy_2_pPS | 4255 | -0.014794489 | [-0.04 0.02] | 0.336060212 | 0.63058718 |
| M_HDL_CE | Liver-Lipid_pPS | 4255 | -0.052368714 | [-0.08 -0.02] | 0.000656062 | 0.006567183 |
| M_HDL_CE | ASAT_Pos_pPS | 4255 | -0.024526365 | [-0.05 0.01] | 0.110728952 | 0.344222613 |
| M_HDL_CE | ALP_Neg_pPS | 4255 | 0.004797227 | [-0.03 0.03] | 0.755106018 | 0.878908284 |
| M_HDL_CE | SHBG-LpA_pPS | 4255 | 0.005415478 | [-0.02 0.04] | 0.724753675 | 0.866514653 |
| M_HDL_CE | Obesity_pPS | 4255 | -0.022642499 | [-0.05 0.01] | 0.140916527 | 0.410647579 |
| M_HDL_CE | VAT_Neg_pPS | 4255 | -0.046651252 | [-0.08 -0.02] | 0.002406308 | 0.017438093 |
| M_HDL_CE | Bilirubin_pPS | 4255 | 0.016440732 | [-0.01 0.05] | 0.285053484 | 0.583442529 |
| M_HDL_CE | Proinsulin_pPS | 4255 | -0.001696586 | [-0.03 0.03] | 0.912162902 | 0.965195628 |
| M_HDL_CE | Hyper_Insulin_pPS | 4255 | 0.008804814 | [-0.02 0.04] | 0.566987813 | 0.797755947 |
| M_HDL_CE | Beta_Cell_1_pPS | 4255 | 0.017114979 | [-0.01 0.05] | 0.265758144 | 0.563609961 |
| M_HDL_CE | Beta_Cell_2_pPS | 4255 | -0.001672132 | [-0.03 0.03] | 0.913423951 | 0.966019414 |
| M_HDL_FC | Cholesterol_pPS | 4255 | 0.043971713 | [0.01 0.07] | 0.004231199 | 0.027864672 |
| M_HDL_FC | Lipodystrophy_1_pPS | 4255 | -0.051671616 | [-0.08 -0.02] | 0.000774097 | 0.007318078 |
| M_HDL_FC | Lipodystrophy_2_pPS | 4255 | -0.020523163 | [-0.05 0.01] | 0.1820267 | 0.462437397 |
| M_HDL_FC | Liver-Lipid_pPS | 4255 | -0.043899167 | [-0.07 -0.01] | 0.004294659 | 0.028142462 |
| M_HDL_FC | ASAT_Pos_pPS | 4255 | -0.027538502 | [-0.06 0. ] | 0.073313874 | 0.250896368 |
| M_HDL_FC | ALP_Neg_pPS | 4255 | 0.01838284 | [-0.01 0.05] | 0.231954583 | 0.527097702 |
| M_HDL_FC | SHBG-LpA_pPS | 4255 | 0.007925782 | [-0.02 0.04] | 0.606318018 | 0.824474694 |
| M_HDL_FC | Obesity_pPS | 4255 | -0.021700175 | [-0.05 0.01] | 0.158216678 | 0.431435756 |
| M_HDL_FC | VAT_Neg_pPS | 4255 | -0.04483358 | [-0.07 -0.01] | 0.003539699 | 0.023700596 |
| M_HDL_FC | Bilirubin_pPS | 4255 | 0.011986657 | [-0.02 0.04] | 0.435749324 | 0.700698913 |
| M_HDL_FC | Proinsulin_pPS | 4255 | 0.00130153 | [-0.03 0.03] | 0.932559908 | 0.977479024 |
| M_HDL_FC | Hyper_Insulin_pPS | 4255 | 0.006161396 | [-0.02 0.04] | 0.688705075 | 0.858568103 |
| M_HDL_FC | Beta_Cell_1_pPS | 4255 | 0.016870308 | [-0.01 0.05] | 0.27265344 | 0.572772495 |
| M_HDL_FC | Beta_Cell_2_pPS | 4255 | -0.000872119 | [-0.03 0.03] | 0.95478057 | 0.980852662 |
| M_HDL_L | Cholesterol_pPS | 4255 | 0.039063486 | [0.01 0.07] | 0.011058414 | 0.058516788 |
| M_HDL_L | Lipodystrophy_1_pPS | 4255 | -0.047218067 | [-0.08 -0.02] | 0.002127754 | 0.016074577 |
| M_HDL_L | Lipodystrophy_2_pPS | 4255 | -0.019651968 | [-0.05 0.01] | 0.20129202 | 0.486527589 |
| M_HDL_L | Liver-Lipid_pPS | 4255 | -0.054523014 | [-0.08 -0.02] | 0.000388635 | 0.004371056 |
| M_HDL_L | ASAT_Pos_pPS | 4255 | -0.019799043 | [-0.05 0.01] | 0.197933433 | 0.483126879 |
| M_HDL_L | ALP_Neg_pPS | 4255 | 0.015784688 | [-0.01 0.05] | 0.304716195 | 0.603404375 |
| M_HDL_L | SHBG-LpA_pPS | 4255 | 0.003198104 | [-0.03 0.03] | 0.835276784 | 0.923370581 |
| M_HDL_L | Obesity_pPS | 4255 | -0.017355008 | [-0.05 0.01] | 0.259111267 | 0.555700813 |
| M_HDL_L | VAT_Neg_pPS | 4255 | -0.047309012 | [-0.08 -0.02] | 0.002085917 | 0.01587835 |
| M_HDL_L | Bilirubin_pPS | 4255 | 0.014330063 | [-0.02 0.04] | 0.351451543 | 0.637902074 |
| M_HDL_L | Proinsulin_pPS | 4255 | 0.004410657 | [-0.03 0.03] | 0.774281623 | 0.89042977 |
| M_HDL_L | Hyper_Insulin_pPS | 4255 | 0.009675932 | [-0.02 0.04] | 0.529259806 | 0.778529119 |
| M_HDL_L | Beta_Cell_1_pPS | 4255 | 0.0176052 | [-0.01 0.05] | 0.252306409 | 0.54979503 |
| M_HDL_L | Beta_Cell_2_pPS | 4255 | -0.004419007 | [-0.03 0.03] | 0.773865928 | 0.89042977 |
| M_HDL_P | Cholesterol_pPS | 4255 | 0.035802048 | [0.01 0.07] | 0.019881962 | 0.089647945 |
| M_HDL_P | Lipodystrophy_1_pPS | 4255 | -0.044916726 | [-0.07 -0.01] | 0.003478747 | 0.023370643 |
| M_HDL_P | Lipodystrophy_2_pPS | 4255 | -0.019790831 | [-0.05 0.01] | 0.198125458 | 0.483126879 |
| M_HDL_P | Liver-Lipid_pPS | 4255 | -0.055988217 | [-0.09 -0.03] | 0.000269303 | 0.003287461 |
| M_HDL_P | ASAT_Pos_pPS | 4255 | -0.018268068 | [-0.05 0.01] | 0.234882749 | 0.530670632 |
| M_HDL_P | ALP_Neg_pPS | 4255 | 0.016845388 | [-0.01 0.05] | 0.27336252 | 0.573178999 |
| M_HDL_P | SHBG-LpA_pPS | 4255 | 0.002080155 | [-0.03 0.03] | 0.892414043 | 0.954690071 |
| M_HDL_P | Obesity_pPS | 4255 | -0.015993008 | [-0.05 0.01] | 0.298377294 | 0.596053587 |
| M_HDL_P | VAT_Neg_pPS | 4255 | -0.047168661 | [-0.08 -0.02] | 0.002150803 | 0.016187619 |
| M_HDL_P | Bilirubin_pPS | 4255 | 0.014291534 | [-0.02 0.04] | 0.352748158 | 0.639407383 |
| M_HDL_P | Proinsulin_pPS | 4255 | 0.005503167 | [-0.02 0.04] | 0.720482231 | 0.866319132 |
| M_HDL_P | Hyper_Insulin_pPS | 4255 | 0.010225549 | [-0.02 0.04] | 0.50613063 | 0.76185979 |
| M_HDL_P | Beta_Cell_1_pPS | 4255 | 0.017334112 | [-0.01 0.05] | 0.259685287 | 0.555700813 |
| M_HDL_P | Beta_Cell_2_pPS | 4255 | -0.005297578 | [-0.04 0.02] | 0.730510245 | 0.869031685 |
| M_HDL_PL | Cholesterol_pPS | 4255 | 0.035321271 | [0.01 0.07] | 0.021602883 | 0.096970787 |
| M_HDL_PL | Lipodystrophy_1_pPS | 4255 | -0.044969386 | [-0.08 -0.01] | 0.003440639 | 0.023262887 |
| M_HDL_PL | Lipodystrophy_2_pPS | 4255 | -0.021028423 | [-0.05 0.01] | 0.171499015 | 0.448811801 |
| M_HDL_PL | Liver-Lipid_pPS | 4255 | -0.048898468 | [-0.08 -0.02] | 0.001466442 | 0.011837974 |
| M_HDL_PL | ASAT_Pos_pPS | 4255 | -0.019419434 | [-0.05 0.01] | 0.206676997 | 0.49296928 |
| M_HDL_PL | ALP_Neg_pPS | 4255 | 0.021690919 | [-0.01 0.05] | 0.158394246 | 0.431435756 |

**Table S9: Associations between pPSs and global metabolomic profile in SCAPIS**

**a) Spearman Correlation**

| Metabolomics | PPS | Samp | r | CI95% | p | fdr_p_adjusted |
|---|---|---|---|---|---|---|
| M_HDL_CE | SHBG-LpA_pPS | 4255 | -0.00299543 | [-0.03 0.03] | 0.845345789 | 0.92810697 |
| M_HDL_CE | Obesity_pPS | 4255 | -0.020857206 | [-0.05 0.01] | 0.17435243 | 0.439983485 |
| M_HDL_CE | VAT_Neg_pPS | 4255 | -0.050640345 | [-0.08 -0.02] | 0.000967634 | 0.008236412 |
| M_HDL_CE | Bilirubin_pPS | 4255 | 0.015156948 | [-0.01 0.05] | 0.323611357 | 0.608687153 |
| M_HDL_CE | Proinsulin_pPS | 4255 | -0.000715269 | [-0.03 0.03] | 0.962849828 | 0.982019059 |
| M_HDL_CE | Hyper_Insulin_pPS | 4255 | -0.007553526 | [-0.04 0.02] | 0.622799947 | 0.80242474 |
| M_HDL_CE | Beta_Cell_1_pPS | 4255 | 0.007579277 | [-0.02 0.04] | 0.621614854 | 0.802264658 |
| M_HDL_CE | Beta_Cell_2_pPS | 4255 | -0.002050524 | [-0.03 0.03] | 0.893775455 | 0.948580863 |
| M_HDL_CE | Total_GRS | 4255 | -0.032803883 | [-0.06 -0. ] | 0.032619546 | 0.126526086 |
| M_HDL_FC | Cholesterol_pPS | 4255 | 0.041433789 | [0.01 0.07] | 0.006948738 | 0.035488197 |
| M_HDL_FC | Lipodystrophy_1_pPS | 4255 | -0.05902283 | [-0.09 -0.03] | 0.000119637 | 0.001603884 |
| M_HDL_FC | Lipodystrophy_2_pPS | 4255 | -0.031577003 | [-0.06 -0. ] | 0.039707435 | 0.147612563 |
| M_HDL_FC | Liver-Lipid_pPS | 4255 | -0.043679681 | [-0.07 -0.01] | 0.004430827 | 0.024873164 |
| M_HDL_FC | ASAT_Pos_pPS | 4255 | -0.039919345 | [-0.07 -0.01] | 0.009307467 | 0.044965127 |
| M_HDL_FC | ALP_Neg_pPS | 4255 | 0.0153222 | [-0.01 0.05] | 0.318363373 | 0.60701283 |
| M_HDL_FC | SHBG-LpA_pPS | 4255 | -0.000288747 | [-0.03 0.03] | 0.984998281 | 0.990539763 |
| M_HDL_FC | Obesity_pPS | 4255 | -0.020259585 | [-0.05 0.01] | 0.187028135 | 0.459010697 |
| M_HDL_FC | VAT_Neg_pPS | 4255 | -0.048998061 | [-0.08 -0.02] | 0.001409843 | 0.010996774 |
| M_HDL_FC | Bilirubin_pPS | 4255 | 0.010666125 | [-0.02 0.04] | 0.487313235 | 0.717916819 |
| M_HDL_FC | Proinsulin_pPS | 4255 | 0.005009031 | [-0.03 0.04] | 0.744283842 | 0.876709962 |
| M_HDL_FC | Hyper_Insulin_pPS | 4255 | -0.009653577 | [-0.04 0.02] | 0.529580128 | 0.744284315 |
| M_HDL_FC | Beta_Cell_1_pPS | 4255 | 0.009073115 | [-0.02 0.04] | 0.554623973 | 0.761144225 |
| M_HDL_FC | Beta_Cell_2_pPS | 4255 | -0.000342993 | [-0.03 0.03] | 0.982180398 | 0.990005874 |
| M_HDL_FC | Total_GRS | 4255 | -0.031027609 | [-0.06 -0. ] | 0.043280778 | 0.157351303 |
| M_HDL_L | Cholesterol_pPS | 4255 | 0.036866719 | [0.01 0.07] | 0.016326012 | 0.071467952 |
| M_HDL_L | Lipodystrophy_1_pPS | 4255 | -0.052958284 | [-0.08 -0.02] | 0.000558475 | 0.005650608 |
| M_HDL_L | Lipodystrophy_2_pPS | 4255 | -0.030376786 | [-0.06 -0. ] | 0.047864869 | 0.169170784 |
| M_HDL_L | Liver-Lipid_pPS | 4255 | -0.054279182 | [-0.08 -0.02] | 0.000404362 | 0.004315208 |
| M_HDL_L | ASAT_Pos_pPS | 4255 | -0.030680784 | [-0.06 -0. ] | 0.045675897 | 0.164111891 |
| M_HDL_L | ALP_Neg_pPS | 4255 | 0.01314183 | [-0.02 0.04] | 0.392096576 | 0.660162603 |
| M_HDL_L | SHBG-LpA_pPS | 4255 | -0.004615621 | [-0.03 0.03] | 0.763744854 | 0.889697961 |
| M_HDL_L | Obesity_pPS | 4255 | -0.015442934 | [-0.05 0.01] | 0.314564595 | 0.60298573 |
| M_HDL_L | VAT_Neg_pPS | 4255 | -0.05123797 | [-0.08 -0.02] | 0.000841518 | 0.0075357 |
| M_HDL_L | Bilirubin_pPS | 4255 | 0.013549364 | [-0.02 0.04] | 0.377580425 | 0.648966355 |
| M_HDL_L | Proinsulin_pPS | 4255 | 0.007554341 | [-0.02 0.04] | 0.622762418 | 0.80242474 |
| M_HDL_L | Hyper_Insulin_pPS | 4255 | -0.005557377 | [-0.04 0.02] | 0.717431021 | 0.865030658 |
| M_HDL_L | Beta_Cell_1_pPS | 4255 | 0.009888345 | [-0.02 0.04] | 0.519616896 | 0.736667707 |
| M_HDL_L | Beta_Cell_2_pPS | 4255 | -0.003721182 | [-0.03 0.03] | 0.808531498 | 0.911007872 |
| M_HDL_L | Total_GRS | 4255 | -0.026907705 | [-0.06 0. ] | 0.07968385 | 0.252097136 |
| M_HDL_P | Cholesterol_pPS | 4255 | 0.033727608 | [0. 0.06] | 0.028024929 | 0.112783253 |
| M_HDL_P | Lipodystrophy_1_pPS | 4255 | -0.050310556 | [-0.08 -0.02] | 0.001044509 | 0.008751843 |
| M_HDL_P | Lipodystrophy_2_pPS | 4255 | -0.030354289 | [-0.06 -0. ] | 0.048030306 | 0.169170784 |
| M_HDL_P | Liver-Lipid_pPS | 4255 | -0.055722114 | [-0.09 -0.03] | 0.000281892 | 0.003199246 |
| M_HDL_P | ASAT_Pos_pPS | 4255 | -0.028617379 | [-0.06 0. ] | 0.062332892 | 0.206333417 |
| M_HDL_P | ALP_Neg_pPS | 4255 | 0.014451755 | [-0.02 0.04] | 0.346635804 | 0.62662761 |
| M_HDL_P | SHBG-LpA_pPS | 4255 | -0.00551009 | [-0.04 0.02] | 0.71973355 | 0.865374699 |
| M_HDL_P | Obesity_pPS | 4255 | -0.014057501 | [-0.04 0.02] | 0.359951586 | 0.637617776 |
| M_HDL_P | VAT_Neg_pPS | 4255 | -0.051037699 | [-0.08 -0.02] | 0.000881976 | 0.007711973 |
| M_HDL_P | Bilirubin_pPS | 4255 | 0.013646343 | [-0.02 0.04] | 0.374175486 | 0.645644896 |
| M_HDL_P | Proinsulin_pPS | 4255 | 0.008717664 | [-0.02 0.04] | 0.570240438 | 0.771226822 |
| M_HDL_P | Hyper_Insulin_pPS | 4255 | -0.00463663 | [-0.03 0.03] | 0.762701706 | 0.889654204 |
| M_HDL_P | Beta_Cell_1_pPS | 4255 | 0.009870908 | [-0.02 0.04] | 0.520353533 | 0.737224788 |
| M_HDL_P | Beta_Cell_2_pPS | 4255 | -0.004533776 | [-0.03 0.03] | 0.767812829 | 0.889703209 |
| M_HDL_P | Total_GRS | 4255 | -0.025366991 | [-0.06 0. ] | 0.09850561 | 0.299708557 |
| M_HDL_PL | Cholesterol_pPS | 4255 | 0.033335851 | [0. 0.06] | 0.029900586 | 0.118780144 |
| M_HDL_PL | Lipodystrophy_1_pPS | 4255 | -0.050554576 | [-0.08 -0.02] | 0.000987107 | 0.008368949 |
| M_HDL_PL | Lipodystrophy_2_pPS | 4255 | -0.031332891 | [-0.06 -0. ] | 0.04126361 | 0.151818943 |
| M_HDL_PL | Liver-Lipid_pPS | 4255 | -0.048464145 | [-0.08 -0.02] | 0.001589656 | 0.012134564 |
| M_HDL_PL | ASAT_Pos_pPS | 4255 | -0.030012265 | [-0.06 0. ] | 0.05060519 | 0.176501028 |
| M_HDL_PL | ALP_Neg_pPS | 4255 | 0.019979836 | [-0.01 0.05] | 0.193190256 | 0.465087653 |
| M_HDL_PL | SHBG-LpA_pPS | 4255 | -0.003663774 | [-0.03 0.03] | 0.811429325 | 0.912487054 |
| M_HDL_PL | Obesity_pPS | 4255 | -0.014033659 | [-0.04 0.02] | 0.360766998 | 0.638486148 |
| M_HDL_PL | VAT_Neg_pPS | 4255 | -0.049436154 | [-0.08 -0.02] | 0.001276512 | 0.010103761 |
| M_HDL_PL | Bilirubin_pPS | 4255 | 0.011481133 | [-0.02 0.04] | 0.454661667 | 0.696320363 |
| M_HDL_PL | Proinsulin_pPS | 4255 | 0.011870716 | [-0.02 0.04] | 0.439499976 | 0.691142319 |
| M_HDL_PL | Hyper_Insulin_pPS | 4255 | -0.005700425 | [-0.04 0.02] | 0.710481347 | 0.861007056 |
| M_HDL_PL | Beta_Cell_1_pPS | 4255 | 0.009775363 | [-0.02 0.04] | 0.524399604 | 0.74051162 |
| M_HDL_PL | Beta_Cell_2_pPS | 4255 | -0.004287654 | [-0.03 0.03] | 0.780084003 | 0.894802239 |
| M_HDL_PL | Total_GRS | 4255 | -0.024530345 | [-0.05 0.01] | 0.110124247 | 0.324473227 |
| M_HDL_TG | Cholesterol_pPS | 4255 | -0.055818351 | [-0.09 -0.03] | 0.000275107 | 0.003138853 |
| M_HDL_TG | Lipodystrophy_1_pPS | 4255 | 0.053584568 | [0.02 0.08] | 0.000479618 | 0.004994081 |
| M_HDL_TG | Lipodystrophy_2_pPS | 4255 | -0.009021912 | [-0.04 0.02] | 0.556860611 | 0.763199634 |
| M_HDL_TG | Liver-Lipid_pPS | 4255 | -0.04202187 | [-0.07 -0.01] | 0.006188244 | 0.032296308 |
| M_HDL_TG | ASAT_Pos_pPS | 4255 | 0.04058597 | [0.01 0.07] | 0.008192973 | 0.040492918 |
| M_HDL_TG | ALP_Neg_pPS | 4255 | 0.026379138 | [-0. 0.06] | 0.085781069 | 0.266973952 |
| M_HDL_TG | SHBG-LpA_pPS | 4255 | -0.008785479 | [-0.04 0.02] | 0.567244956 | 0.769278765 |
| M_HDL_TG | Obesity_pPS | 4255 | 0.020516354 | [-0.01 0.05] | 0.18150105 | 0.45159215 |
| M_HDL_TG | VAT_Neg_pPS | 4255 | 0.004931624 | [-0.03 0.04] | 0.748100528 | 0.879158468 |
| M_HDL_TG | Bilirubin_pPS | 4255 | -0.001700171 | [-0.03 0.03] | 0.911843342 | 0.955031235 |
| M_HDL_TG | Proinsulin_pPS | 4255 | 0.021087491 | [-0.01 0.05] | 0.169643076 | 0.431822279 |
| M_HDL_TG | Hyper_Insulin_pPS | 4255 | 0.015927289 | [-0.01 0.05] | 0.29962548 | 0.585867506 |
| M_HDL_TG | Beta_Cell_1_pPS | 4255 | 0.011454432 | [-0.02 0.04] | 0.455711517 | 0.696320363 |
| M_HDL_TG | Beta_Cell_2_pPS | 4255 | -0.001276317 | [-0.03 0.03] | 0.933761972 | 0.968998273 |
| M_HDL_TG | Total_GRS | 4255 | 0.031314485 | [0. 0.06] | 0.041383013 | 0.151852919 |
| M_LDL_C | Cholesterol_pPS | 4255 | -0.153789367 | [-0.18 -0.12] | 7.15E-24 | 1.53E-21 |
| M_LDL_C | Lipodystrophy_1_pPS | 4255 | 0.037626213 | [0.01 0.07] | 0.014243865 | 0.063519939 |
| M_LDL_C | Lipodystrophy_2_pPS | 4255 | -0.041251044 | [-0.07 -0.01] | 0.007201592 | 0.036432584 |
| M_LDL_C | Liver-Lipid_pPS | 4255 | 0.009504325 | [-0.02 0.04] | 0.535964258 | 0.75041993 |
| M_LDL_C | ASAT_Pos_pPS | 4255 | -0.013348006 | [-0.04 0.02] | 0.384710851 | 0.654405057 |
| M_LDL_C | ALP_Neg_pPS | 4255 | 0.045283276 | [0.02 0.08] | 0.003174444 | 0.019679715 |
| M_LDL_C | SHBG-LpA_pPS | 4255 | 0.0167487 | [-0.01 0.05] | 0.275388569 | 0.560079455 |
| M_LDL_C | Obesity_pPS | 4255 | -0.007913028 | [-0.04 0.02] | 0.60634514 | 0.792571801 |
| M_LDL_C | VAT_Neg_pPS | 4255 | 0.011851859 | [-0.02 0.04] | 0.440227128 | 0.691279056 |
| M_LDL_C | Bilirubin_pPS | 4255 | -0.014156277 | [-0.04 0.02] | 0.356585686 | 0.635279317 |

**b) Partial Correlation**

| Metabolomics | PPS | Samp | r | CI95% | p | fdr_p_adjusted |
|---|---|---|---|---|---|---|
| M_HDL_PL | SHBG-LpA_pPS | 4255 | 0.003853316 | [-0.03 0.03] | 0.80216933 | 0.906328459 |
| M_HDL_PL | Obesity_pPS | 4255 | -0.016239167 | [-0.05 0.01] | 0.291001231 | 0.588469156 |
| M_HDL_PL | VAT_Neg_pPS | 4255 | -0.045430579 | [-0.08 -0.02] | 0.00312277 | 0.02155788 |
| M_HDL_PL | Bilirubin_pPS | 4255 | 0.012254228 | [-0.02 0.04] | 0.425572791 | 0.693893617 |
| M_HDL_PL | Proinsulin_pPS | 4255 | 0.007831252 | [-0.02 0.04] | 0.610619215 | 0.828224707 |
| M_HDL_PL | Hyper_Insulin_pPS | 4255 | 0.008771644 | [-0.02 0.04] | 0.56844948 | 0.797755947 |
| M_HDL_PL | Beta_Cell_1_pPS | 4255 | 0.016313942 | [-0.01 0.05] | 0.288785123 | 0.586356812 |
| M_HDL_PL | Beta_Cell_2_pPS | 4255 | -0.005560128 | [-0.04 0.02] | 0.717712236 | 0.866099998 |
| M_HDL_TG | Cholesterol_pPS | 4255 | -0.056466575 | [-0.09 -0.03] | 0.000238461 | 0.002946901 |
| M_HDL_TG | Lipodystrophy_1_pPS | 4255 | 0.046687753 | [0.02 0.08] | 0.002387411 | 0.017438093 |
| M_HDL_TG | Lipodystrophy_2_pPS | 4255 | -0.012117404 | [-0.04 0.02] | 0.430759172 | 0.696591166 |
| M_HDL_TG | Liver-Lipid_pPS | 4255 | -0.04333498 | [-0.07 -0.01] | 0.004818386 | 0.030051115 |
| M_HDL_TG | ASAT_Pos_pPS | 4255 | 0.031129961 | [0. 0.06] | 0.042914965 | 0.163294026 |
| M_HDL_TG | ALP_Neg_pPS | 4255 | 0.023500887 | [-0.01 0.05] | 0.126457781 | 0.378994725 |
| M_HDL_TG | SHBG-LpA_pPS | 4255 | -0.01300997 | [-0.04 0.02] | 0.397589565 | 0.673455453 |
| M_HDL_TG | Obesity_pPS | 4255 | 0.019839992 | [-0.01 0.05] | 0.197013175 | 0.482766677 |
| M_HDL_TG | VAT_Neg_pPS | 4255 | 0.00257935 | [-0.03 0.03] | 0.866813298 | 0.943130556 |
| M_HDL_TG | Bilirubin_pPS | 4255 | -0.004964024 | [-0.04 0.03] | 0.746877759 | 0.874926432 |
| M_HDL_TG | Proinsulin_pPS | 4255 | 0.015049439 | [-0.02 0.05] | 0.327798264 | 0.626194775 |
| M_HDL_TG | Hyper_Insulin_pPS | 4255 | 0.00930664 | [-0.02 0.04] | 0.545096821 | 0.789070018 |
| M_HDL_TG | Beta_Cell_1_pPS | 4255 | 0.004715874 | [-0.03 0.03] | 0.759129434 | 0.882565114 |
| M_HDL_TG | Beta_Cell_2_pPS | 4255 | -0.004056849 | [-0.03 0.03] | 0.791953845 | 0.899824971 |
| M_LDL_C | Cholesterol_pPS | 4255 | -0.15223714 | [-0.18 -0.12] | 2.36E-23 | 4.73E-21 |
| M_LDL_C | Lipodystrophy_1_pPS | 4255 | 0.042654816 | [0.01 0.07] | 0.005526182 | 0.033323545 |
| M_LDL_C | Lipodystrophy_2_pPS | 4255 | -0.036401946 | [-0.07 -0.01] | 0.017903346 | 0.084078549 |
| M_LDL_C | Liver-Lipid_pPS | 4255 | 0.012019169 | [-0.02 0.04] | 0.434505355 | 0.699850608 |
| M_LDL_C | ASAT_Pos_pPS | 4255 | -0.017149196 | [-0.05 0.01] | 0.264803512 | 0.562180946 |
| M_LDL_C | ALP_Neg_pPS | 4255 | 0.048787809 | [0.02 0.08] | 0.001503352 | 0.012087189 |
| M_LDL_C | SHBG-LpA_pPS | 4255 | 0.01396548 | [-0.02 0.04] | 0.36384177 | 0.650367164 |
| M_LDL_C | Obesity_pPS | 4255 | 0.007561155 | [-0.02 0.04] | 0.622984553 | 0.833339789 |
| M_LDL_C | VAT_Neg_pPS | 4255 | 0.008179807 | [-0.02 0.04] | 0.594827357 | 0.817887615 |
| M_LDL_C | Bilirubin_pPS | 4255 | -0.015278937 | [-0.05 0.01] | 0.320474779 | 0.620672596 |
| M_LDL_C | Proinsulin_pPS | 4255 | -0.022686645 | [-0.05 0.01] | 0.140143197 | 0.409586394 |
| M_LDL_C | Hyper_Insulin_pPS | 4255 | -0.021849808 | [-0.05 0.01] | 0.155366978 | 0.42902716 |
| M_LDL_C | Beta_Cell_1_pPS | 4255 | 0.01772284 | [-0.01 0.05] | 0.249150215 | 0.545779139 |
| M_LDL_C | Beta_Cell_2_pPS | 4255 | 0.021351688 | [-0.01 0.05] | 0.165006585 | 0.441248115 |
| M_LDL_CE | Cholesterol_pPS | 4255 | -0.152812588 | [-0.18 -0.12] | 1.61E-23 | 4.03E-21 |
| M_LDL_CE | Lipodystrophy_1_pPS | 4255 | 0.043662749 | [0.01 0.07] | 0.004507498 | 0.028878276 |
| M_LDL_CE | Lipodystrophy_2_pPS | 4255 | -0.035071117 | [-0.07 -0. ] | 0.02254853 | 0.099894945 |
| M_LDL_CE | Liver-Lipid_pPS | 4255 | 0.012777508 | [-0.02 0.04] | 0.406076654 | 0.678602222 |
| M_LDL_CE | ASAT_Pos_pPS | 4255 | -0.016171069 | [-0.05 0.01] | 0.293029388 | 0.589592798 |
| M_LDL_CE | ALP_Neg_pPS | 4255 | 0.048352826 | [0.02 0.08] | 0.00165688 | 0.013110961 |
| M_LDL_CE | SHBG-LpA_pPS | 4255 | 0.013788581 | [-0.02 0.04] | 0.369950808 | 0.653351391 |
| M_LDL_CE | Obesity_pPS | 4255 | 0.007654079 | [-0.02 0.04] | 0.618716934 | 0.830208648 |
| M_LDL_CE | VAT_Neg_pPS | 4255 | 0.008477261 | [-0.02 0.04] | 0.581500112 | 0.809008495 |
| M_LDL_CE | Bilirubin_pPS | 4255 | -0.015256462 | [-0.05 0.01] | 0.321187218 | 0.620672596 |
| M_LDL_CE | Proinsulin_pPS | 4255 | -0.02245495 | [-0.05 0.01] | 0.14423851 | 0.4165382 |
| M_LDL_CE | Hyper_Insulin_pPS | 4255 | -0.021851045 | [-0.05 0.01] | 0.15534358 | 0.42902716 |
| M_LDL_CE | Beta_Cell_1_pPS | 4255 | 0.017035124 | [-0.01 0.05] | 0.267995302 | 0.567753011 |
| M_LDL_CE | Beta_Cell_2_pPS | 4255 | 0.02131052 | [-0.01 0.05] | 0.165822972 | 0.441605751 |
| M_LDL_FC | Cholesterol_pPS | 4255 | -0.148565972 | [-0.18 -0.12] | 2.64E-22 | 3.11E-20 |
| M_LDL_FC | Lipodystrophy_1_pPS | 4255 | 0.038073478 | [0.01 0.07] | 0.013271256 | 0.067434148 |
| M_LDL_FC | Lipodystrophy_2_pPS | 4255 | -0.041265394 | [-0.07 -0.01] | 0.007270635 | 0.042436766 |
| M_LDL_FC | Liver-Lipid_pPS | 4255 | 0.00912286 | [-0.02 0.04] | 0.55306495 | 0.795753776 |
| M_LDL_FC | ASAT_Pos_pPS | 4255 | -0.020615082 | [-0.05 0.01] | 0.180076608 | 0.458668408 |
| M_LDL_FC | ALP_Neg_pPS | 4255 | 0.049066312 | [0.02 0.08] | 0.001412051 | 0.01154043 |
| M_LDL_FC | SHBG-LpA_pPS | 4255 | 0.014361914 | [-0.02 0.04] | 0.350381932 | 0.637861136 |
| M_LDL_FC | Obesity_pPS | 4255 | 0.00718028 | [-0.02 0.04] | 0.6405984 | 0.836753752 |
| M_LDL_FC | VAT_Neg_pPS | 4255 | 0.006198837 | [-0.02 0.04] | 0.686913281 | 0.858568103 |
| M_LDL_FC | Bilirubin_pPS | 4255 | -0.015918909 | [-0.05 0.01] | 0.300621862 | 0.597661337 |
| M_LDL_FC | Proinsulin_pPS | 4255 | -0.022936519 | [-0.05 0.01] | 0.135827393 | 0.400480768 |
| M_LDL_FC | Hyper_Insulin_pPS | 4255 | -0.02173678 | [-0.05 0.01] | 0.157515912 | 0.431435756 |
| M_LDL_FC | Beta_Cell_1_pPS | 4255 | 0.020412433 | [-0.01 0.05] | 0.184396591 | 0.466062214 |
| M_LDL_FC | Beta_Cell_2_pPS | 4255 | 0.020989956 | [-0.01 0.05] | 0.172284136 | 0.45027786 |
| M_LDL_L | Cholesterol_pPS | 4255 | -0.153905873 | [-0.18 -0.12] | 7.74E-24 | 3.20E-21 |
| M_LDL_L | Lipodystrophy_1_pPS | 4255 | 0.046848067 | [0.02 0.08] | 0.002306012 | 0.017035555 |
| M_LDL_L | Lipodystrophy_2_pPS | 4255 | -0.037017288 | [-0.07 -0.01] | 0.016055097 | 0.077826401 |
| M_LDL_L | Liver-Lipid_pPS | 4255 | 0.006583345 | [-0.02 0.04] | 0.668615692 | 0.849884836 |
| M_LDL_L | ASAT_Pos_pPS | 4255 | -0.018832489 | [-0.05 0.01] | 0.220732571 | 0.514443081 |
| M_LDL_L | ALP_Neg_pPS | 4255 | 0.0502769 | [0.02 0.08] | 0.001071551 | 0.009128699 |
| M_LDL_L | SHBG-LpA_pPS | 4255 | 0.013928478 | [-0.02 0.04] | 0.365114366 | 0.650898451 |
| M_LDL_L | Obesity_pPS | 4255 | 0.009913204 | [-0.02 0.04] | 0.519209303 | 0.772830501 |
| M_LDL_L | VAT_Neg_pPS | 4255 | 0.007005668 | [-0.02 0.04] | 0.648743287 | 0.843394061 |
| M_LDL_L | Bilirubin_pPS | 4255 | -0.014933748 | [-0.05 0.02] | 0.331530909 | 0.62795236 |
| M_LDL_L | Proinsulin_pPS | 4255 | -0.020264627 | [-0.05 0.01] | 0.187594516 | 0.470631855 |
| M_LDL_L | Hyper_Insulin_pPS | 4255 | -0.021905832 | [-0.05 0.01] | 0.154310117 | 0.428688101 |
| M_LDL_L | Beta_Cell_1_pPS | 4255 | 0.017863019 | [-0.01 0.05] | 0.24542546 | 0.542919083 |
| M_LDL_L | Beta_Cell_2_pPS | 4255 | 0.020825685 | [-0.01 0.05] | 0.175667207 | 0.453447086 |
| M_LDL_P | Cholesterol_pPS | 4255 | -0.154264293 | [-0.18 -0.12] | 6.08E-24 | 3.20E-21 |
| M_LDL_P | Lipodystrophy_1_pPS | 4255 | 0.047935042 | [0.02 0.08] | 0.00181779 | 0.014105489 |
| M_LDL_P | Lipodystrophy_2_pPS | 4255 | -0.036585484 | [-0.07 -0.01] | 0.017333486 | 0.082231372 |
| M_LDL_P | Liver-Lipid_pPS | 4255 | 0.005708928 | [-0.02 0.04] | 0.710493677 | 0.864160597 |
| M_LDL_P | ASAT_Pos_pPS | 4255 | -0.019183953 | [-0.05 0.01] | 0.212235993 | 0.501057144 |
| M_LDL_P | ALP_Neg_pPS | 4255 | 0.050514442 | [0.02 0.08] | 0.001014378 | 0.008829501 |
| M_LDL_P | SHBG-LpA_pPS | 4255 | 0.013859327 | [-0.02 0.04] | 0.367500048 | 0.652247425 |
| M_LDL_P | Obesity_pPS | 4255 | 0.010491002 | [-0.02 0.04] | 0.495153184 | 0.753425828 |
| M_LDL_P | VAT_Neg_pPS | 4255 | 0.006891371 | [-0.02 0.04] | 0.654097684 | 0.844628798 |
| M_LDL_P | Bilirubin_pPS | 4255 | -0.014643788 | [-0.04 0.02] | 0.341006294 | 0.635656053 |
| M_LDL_P | Proinsulin_pPS | 4255 | -0.019412453 | [-0.05 0.01] | 0.206840257 | 0.49296928 |
| M_LDL_P | Hyper_Insulin_pPS | 4255 | -0.022027037 | [-0.05 0.01] | 0.152042328 | 0.427792579 |
| M_LDL_P | Beta_Cell_1_pPS | 4255 | 0.017414364 | [-0.01 0.05] | 0.257485492 | 0.553690606 |
| M_LDL_P | Beta_Cell_2_pPS | 4255 | 0.020758828 | [-0.01 0.05] | 0.177058196 | 0.455032746 |
| M_LDL_PL | Cholesterol_pPS | 4255 | -0.154122013 | [-0.18 -0.12] | 6.69E-24 | 3.20E-21 |
| M_LDL_PL | Lipodystrophy_1_pPS | 4255 | 0.050659714 | [0.02 0.08] | 0.000980819 | 0.008574667 |

**Table S9: Associations between pPSs and global metabolomic profile in SCAPIS**

**a) Spearman Correlation**

| Metabolomics | PPS | Samp | r | CI95% | p | fdr_p_adjusted |
|---|---|---|---|---|---|---|
| M_LDL_C | Proinsulin_pPS | 4255 | -0.00778136 | [-0.04 0.02] | 0.612349197 | 0.795619093 |
| M_LDL_C | Hyper_Insulin_pPS | 4255 | -0.019795446 | [-0.05 0.01] | 0.197332616 | 0.47103115 |
| M_LDL_C | Beta_Cell_1_pPS | 4255 | 0.021892158 | [-0.01 0.05] | 0.153935041 | 0.406638747 |
| M_LDL_C | Beta_Cell_2_pPS | 4255 | 0.022371719 | [-0.01 0.05] | 0.145113892 | 0.394011771 |
| M_LDL_C | Total_GRS | 4255 | -0.003081524 | [-0.03 0.03] | 0.840959289 | 0.926956667 |
| M_LDL_CE | Cholesterol_pPS | 4255 | -0.154297426 | [-0.18 -0.12] | 5.08E-24 | 1.36E-21 |
| M_LDL_CE | Lipodystrophy_1_pPS | 4255 | 0.038872336 | [0.01 0.07] | 0.01133204 | 0.052273605 |
| M_LDL_CE | Lipodystrophy_2_pPS | 4255 | -0.039712142 | [-0.07 -0.01] | 0.009680427 | 0.046041058 |
| M_LDL_CE | Liver-Lipid_pPS | 4255 | 0.010268601 | [-0.02 0.04] | 0.503686915 | 0.727744052 |
| M_LDL_CE | ASAT_Pos_pPS | 4255 | -0.012062746 | [-0.04 0.02] | 0.432134674 | 0.688963257 |
| M_LDL_CE | ALP_Neg_pPS | 4255 | 0.044948083 | [0.01 0.07] | 0.00340646 | 0.020524878 |
| M_LDL_CE | SHBG-LpA_pPS | 4255 | 0.016710772 | [-0.01 0.05] | 0.276477356 | 0.561195453 |
| M_LDL_CE | Obesity_pPS | 4255 | -0.00778118 | [-0.04 0.02] | 0.612357425 | 0.795619093 |
| M_LDL_CE | VAT_Neg_pPS | 4255 | 0.012299057 | [-0.02 0.04] | 0.423169818 | 0.684539412 |
| M_LDL_CE | Bilirubin_pPS | 4255 | -0.014114642 | [-0.04 0.02] | 0.358002007 | 0.636744863 |
| M_LDL_CE | Proinsulin_pPS | 4255 | -0.007780563 | [-0.04 0.02] | 0.612385605 | 0.795619093 |
| M_LDL_CE | Hyper_Insulin_pPS | 4255 | -0.019466356 | [-0.05 0.01] | 0.204886488 | 0.482946722 |
| M_LDL_CE | Beta_Cell_1_pPS | 4255 | 0.021324358 | [-0.01 0.05] | 0.16489924 | 0.426155265 |
| M_LDL_CE | Beta_Cell_2_pPS | 4255 | 0.022226062 | [-0.01 0.05] | 0.147751225 | 0.397150849 |
| M_LDL_CE | Total_GRS | 4255 | -0.002082413 | [-0.03 0.03] | 0.892133499 | 0.948580863 |
| M_LDL_FC | Cholesterol_pPS | 4255 | -0.150351777 | [-0.18 -0.12] | 7.08E-23 | 8.93E-21 |
| M_LDL_FC | Lipodystrophy_1_pPS | 4255 | 0.032125699 | [0. 0.06] | 0.03639047 | 0.137183756 |
| M_LDL_FC | Lipodystrophy_2_pPS | 4255 | -0.046997933 | [-0.08 -0.02] | 0.002197488 | 0.014916492 |
| M_LDL_FC | Liver-Lipid_pPS | 4255 | 0.006569275 | [-0.02 0.04] | 0.668803989 | 0.829239629 |
| M_LDL_FC | ASAT_Pos_pPS | 4255 | -0.018114229 | [-0.05 0.01] | 0.238127736 | 0.520146634 |
| M_LDL_FC | ALP_Neg_pPS | 4255 | 0.045291152 | [0.02 0.08] | 0.00316917 | 0.019679715 |
| M_LDL_FC | SHBG-LpA_pPS | 4255 | 0.016569578 | [-0.01 0.05] | 0.28055633 | 0.567194465 |
| M_LDL_FC | Obesity_pPS | 4255 | -0.008276324 | [-0.04 0.02] | 0.589917078 | 0.781971034 |
| M_LDL_FC | VAT_Neg_pPS | 4255 | 0.009227052 | [-0.02 0.04] | 0.547926361 | 0.760617725 |
| M_LDL_FC | Bilirubin_pPS | 4255 | -0.014874748 | [-0.04 0.02] | 0.332702837 | 0.613626471 |
| M_LDL_FC | Proinsulin_pPS | 4255 | -0.00744274 | [-0.04 0.02] | 0.627909475 | 0.803929814 |
| M_LDL_FC | Hyper_Insulin_pPS | 4255 | -0.021093203 | [-0.05 0.01] | 0.169527476 | 0.431822279 |
| M_LDL_FC | Beta_Cell_1_pPS | 4255 | 0.024029147 | [-0.01 0.05] | 0.117586915 | 0.342230574 |
| M_LDL_FC | Beta_Cell_2_pPS | 4255 | 0.022454735 | [-0.01 0.05] | 0.143626979 | 0.391461081 |
| M_LDL_FC | Total_GRS | 4255 | -0.006965487 | [-0.04 0.02] | 0.65012257 | 0.818487176 |
| M_LDL_L | Cholesterol_pPS | 4255 | -0.155256646 | [-0.18 -0.13] | 2.65E-24 | 1.25E-21 |
| M_LDL_L | Lipodystrophy_1_pPS | 4255 | 0.041566665 | [0.01 0.07] | 0.00676992 | 0.034823688 |
| M_LDL_L | Lipodystrophy_2_pPS | 4255 | -0.042121183 | [-0.07 -0.01] | 0.006067479 | 0.031743272 |
| M_LDL_L | Liver-Lipid_pPS | 4255 | 0.004220431 | [-0.03 0.03] | 0.783445316 | 0.897605875 |
| M_LDL_L | ASAT_Pos_pPS | 4255 | -0.014287165 | [-0.04 0.02] | 0.352156202 | 0.632642423 |
| M_LDL_L | ALP_Neg_pPS | 4255 | 0.047086946 | [0.02 0.08] | 0.002155239 | 0.014722887 |
| M_LDL_L | SHBG-LpA_pPS | 4255 | 0.016958653 | [-0.01 0.05] | 0.269414448 | 0.554275035 |
| M_LDL_L | Obesity_pPS | 4255 | -0.005334153 | [-0.04 0.02] | 0.72832249 | 0.870335232 |
| M_LDL_L | VAT_Neg_pPS | 4255 | 0.010732772 | [-0.02 0.04] | 0.484596523 | 0.717164725 |
| M_LDL_L | Bilirubin_pPS | 4255 | -0.013630086 | [-0.04 0.02] | 0.374744939 | 0.645644896 |
| M_LDL_L | Proinsulin_pPS | 4255 | -0.004940854 | [-0.04 0.03] | 0.747645079 | 0.879158468 |
| M_LDL_L | Hyper_Insulin_pPS | 4255 | -0.019692622 | [-0.05 0.01] | 0.199670604 | 0.474826436 |
| M_LDL_L | Beta_Cell_1_pPS | 4255 | 0.022490054 | [-0.01 0.05] | 0.142997919 | 0.390739537 |
| M_LDL_L | Beta_Cell_2_pPS | 4255 | 0.022133377 | [-0.01 0.05] | 0.149448376 | 0.398714883 |
| M_LDL_L | Total_GRS | 4255 | -0.00065432 | [-0.03 0.03] | 0.966013436 | 0.982019059 |
| M_LDL_P | Cholesterol_pPS | 4255 | -0.155532432 | [-0.18 -0.13] | 2.19E-24 | 1.25E-21 |
| M_LDL_P | Lipodystrophy_1_pPS | 4255 | 0.042619577 | [0.01 0.07] | 0.005492887 | 0.029236336 |
| M_LDL_P | Lipodystrophy_2_pPS | 4255 | -0.041699784 | [-0.07 -0.01] | 0.006594931 | 0.034087051 |
| M_LDL_P | Liver-Lipid_pPS | 4255 | 0.003381258 | [-0.03 0.03] | 0.825726926 | 0.921334342 |
| M_LDL_P | ASAT_Pos_pPS | 4255 | -0.01440424 | [-0.04 0.02] | 0.348223782 | 0.628208589 |
| M_LDL_P | ALP_Neg_pPS | 4255 | 0.047470678 | [0.02 0.08] | 0.001981491 | 0.013934058 |
| M_LDL_P | SHBG-LpA_pPS | 4255 | 0.016981472 | [-0.01 0.05] | 0.268770558 | 0.553806769 |
| M_LDL_P | Obesity_pPS | 4255 | -0.004669086 | [-0.03 0.03] | 0.76109097 | 0.888699036 |
| M_LDL_P | VAT_Neg_pPS | 4255 | 0.010655427 | [-0.02 0.04] | 0.487750112 | 0.718067255 |
| M_LDL_P | Bilirubin_pPS | 4255 | -0.013313433 | [-0.04 0.02] | 0.385943401 | 0.65494351 |
| M_LDL_P | Proinsulin_pPS | 4255 | -0.004105852 | [-0.03 0.03] | 0.789183824 | 0.899856387 |
| M_LDL_P | Hyper_Insulin_pPS | 4255 | -0.019694514 | [-0.05 0.01] | 0.199627386 | 0.474826436 |
| M_LDL_P | Beta_Cell_1_pPS | 4255 | 0.02225204 | [-0.01 0.05] | 0.147278178 | 0.396873985 |
| M_LDL_P | Beta_Cell_2_pPS | 4255 | 0.022088091 | [-0.01 0.05] | 0.150283006 | 0.400443538 |
| M_LDL_P | Total_GRS | 4255 | 8.96E-05 | [-0.03 0.03] | 0.99534458 | 0.996787043 |
| M_LDL_PL | Cholesterol_pPS | 4255 | -0.155385417 | [-0.18 -0.13] | 2.42E-24 | 1.25E-21 |
| M_LDL_PL | Lipodystrophy_1_pPS | 4255 | 0.04619174 | [0.02 0.08] | 0.002616161 | 0.017161058 |
| M_LDL_PL | Lipodystrophy_2_pPS | 4255 | -0.046274626 | [-0.08 -0.02] | 0.002569978 | 0.016909825 |
| M_LDL_PL | Liver-Lipid_pPS | 4255 | -0.009715994 | [-0.04 0.02] | 0.526921764 | 0.742606559 |
| M_LDL_PL | ASAT_Pos_pPS | 4255 | -0.009432396 | [-0.04 0.02] | 0.539054754 | 0.753813726 |
| M_LDL_PL | ALP_Neg_pPS | 4255 | 0.047578759 | [0.02 0.08] | 0.001934928 | 0.013834735 |
| M_LDL_PL | SHBG-LpA_pPS | 4255 | 0.017919314 | [-0.01 0.05] | 0.243216966 | 0.525480742 |
| M_LDL_PL | Obesity_pPS | 4255 | -8.87E-05 | [-0.03 0.03] | 0.995392935 | 0.996787043 |
| M_LDL_PL | VAT_Neg_pPS | 4255 | 0.007063574 | [-0.02 0.04] | 0.64553095 | 0.817402268 |
| M_LDL_PL | Bilirubin_pPS | 4255 | -0.016147098 | [-0.05 0.01] | 0.293004488 | 0.578724334 |
| M_LDL_PL | Proinsulin_pPS | 4255 | -0.00177773 | [-0.03 0.03] | 0.907839349 | 0.952872131 |
| M_LDL_PL | Hyper_Insulin_pPS | 4255 | -0.01787647 | [-0.05 0.01] | 0.244345792 | 0.525480742 |
| M_LDL_PL | Beta_Cell_1_pPS | 4255 | 0.024560185 | [-0.01 0.05] | 0.109692022 | 0.323644273 |
| M_LDL_PL | Beta_Cell_2_pPS | 4255 | 0.019070923 | [-0.01 0.05] | 0.214238517 | 0.495729902 |
| M_LDL_PL | Total_GRS | 4255 | 0.002036772 | [-0.03 0.03] | 0.894483686 | 0.948580863 |
| M_LDL_TG | Cholesterol_pPS | 4255 | -0.112725833 | [-0.14 -0.08] | 1.78E-13 | 8.70E-12 |
| M_LDL_TG | Lipodystrophy_1_pPS | 4255 | 0.053803381 | [0.02 0.08] | 0.000454607 | 0.004780062 |
| M_LDL_TG | Lipodystrophy_2_pPS | 4255 | -0.022145606 | [-0.05 0.01] | 0.149223606 | 0.398714883 |
| M_LDL_TG | Liver-Lipid_pPS | 4255 | -0.015207925 | [-0.05 0.01] | 0.321986459 | 0.608511855 |
| M_LDL_TG | ASAT_Pos_pPS | 4255 | -0.014445361 | [-0.04 0.02] | 0.346849225 | 0.62662761 |
| M_LDL_TG | ALP_Neg_pPS | 4255 | 0.052523894 | [0.02 0.08] | 0.000620088 | 0.006073462 |
| M_LDL_TG | SHBG-LpA_pPS | 4255 | 0.008915243 | [-0.02 0.04] | 0.56153413 | 0.767334778 |
| M_LDL_TG | Obesity_pPS | 4255 | 0.008148572 | [-0.02 0.04] | 0.595670585 | 0.786769338 |
| M_LDL_TG | VAT_Neg_pPS | 4255 | -0.002833374 | [-0.03 0.03] | 0.85361559 | 0.932572076 |
| M_LDL_TG | Bilirubin_pPS | 4255 | -0.002426227 | [-0.03 0.03] | 0.874461523 | 0.942282701 |
| M_LDL_TG | Proinsulin_pPS | 4255 | 0.010016087 | [-0.02 0.04] | 0.5142366 | 0.733381664 |
| M_LDL_TG | Hyper_Insulin_pPS | 4255 | -0.01077041 | [-0.04 0.02] | 0.483065927 | 0.717164725 |
| M_LDL_TG | Beta_Cell_1_pPS | 4255 | 0.002899523 | [-0.03 0.03] | 0.850237973 | 0.930490026 |
| M_LDL_TG | Beta_Cell_2_pPS | 4255 | 0.011303183 | [-0.02 0.04] | 0.461683917 | 0.702847411 |

**b) Partial Correlation**

| Metabolomics | PPS | Samp | r | CI95% | p | fdr_p_adjusted |
|---|---|---|---|---|---|---|
| M_LDL_PL | Lipodystrophy_2_pPS | 4255 | -0.040986887 | [-0.07 -0.01] | 0.007674691 | 0.044025017 |
| M_LDL_PL | Liver-Lipid_pPS | 4255 | -0.007707137 | [-0.04 0.02] | 0.616286981 | 0.828794354 |
| M_LDL_PL | ASAT_Pos_pPS | 4255 | -0.01452442 | [-0.04 0.02] | 0.344956909 | 0.637861136 |
| M_LDL_PL | ALP_Neg_pPS | 4255 | 0.050784644 | [0.02 0.08] | 0.000952783 | 0.008545465 |
| M_LDL_PL | SHBG-LpA_pPS | 4255 | 0.014793816 | [-0.02 0.04] | 0.336082178 | 0.63058718 |
| M_LDL_PL | Obesity_pPS | 4255 | 0.014599512 | [-0.02 0.04] | 0.342468256 | 0.636982428 |
| M_LDL_PL | VAT_Neg_pPS | 4255 | 0.003560052 | [-0.03 0.03] | 0.816947718 | 0.913698059 |
| M_LDL_PL | Bilirubin_pPS | 4255 | -0.017971618 | [-0.05 0.01] | 0.242566799 | 0.540176565 |
| M_LDL_PL | Proinsulin_pPS | 4255 | -0.017592278 | [-0.05 0.01] | 0.252654798 | 0.549798809 |
| M_LDL_PL | Hyper_Insulin_pPS | 4255 | -0.020040595 | [-0.05 0.01] | 0.192521891 | 0.475838056 |
| M_LDL_PL | Beta_Cell_1_pPS | 4255 | 0.020393189 | [-0.01 0.05] | 0.184810789 | 0.466062214 |
| M_LDL_PL | Beta_Cell_2_pPS | 4255 | 0.017335577 | [-0.01 0.05] | 0.259645022 | 0.555700813 |
| M_LDL_TG | Cholesterol_pPS | 4255 | -0.113571308 | [-0.14 -0.08] | 1.28E-13 | 5.80E-12 |
| M_LDL_TG | Lipodystrophy_1_pPS | 4255 | 0.058230159 | [0.03 0.09] | 0.000152149 | 0.002058124 |
| M_LDL_TG | Lipodystrophy_2_pPS | 4255 | -0.017556771 | [-0.05 0.01] | 0.253613801 | 0.550091907 |
| M_LDL_TG | Liver-Lipid_pPS | 4255 | -0.014449194 | [-0.04 0.02] | 0.347461526 | 0.637861136 |
| M_LDL_TG | ASAT_Pos_pPS | 4255 | -0.02265822 | [-0.05 0.01] | 0.140640746 | 0.41044136 |
| M_LDL_TG | ALP_Neg_pPS | 4255 | 0.051752377 | [0.02 0.08] | 0.000759477 | 0.007240349 |
| M_LDL_TG | SHBG-LpA_pPS | 4255 | 0.005598443 | [-0.02 0.04] | 0.715851064 | 0.866099998 |
| M_LDL_TG | Obesity_pPS | 4255 | 0.016770393 | [-0.01 0.05] | 0.275504102 | 0.573942989 |
| M_LDL_TG | VAT_Neg_pPS | 4255 | -0.005379339 | [-0.04 0.02] | 0.726516563 | 0.866797473 |
| M_LDL_TG | Bilirubin_pPS | 4255 | -0.003491512 | [-0.03 0.03] | 0.820411197 | 0.915531336 |
| M_LDL_TG | Proinsulin_pPS | 4255 | 0.000280936 | [-0.03 0.03] | 0.985426444 | 0.993363084 |
| M_LDL_TG | Hyper_Insulin_pPS | 4255 | -0.014118406 | [-0.04 0.02] | 0.358611749 | 0.64484135 |
| M_LDL_TG | Beta_Cell_1_pPS | 4255 | -0.002275887 | [-0.03 0.03] | 0.882361237 | 0.949724299 |
| M_LDL_TG | Beta_Cell_2_pPS | 4255 | 0.012462582 | [-0.02 0.04] | 0.417745416 | 0.690608028 |
| M_VLDL_C | Cholesterol_pPS | 4255 | -0.101472943 | [-0.13 -0.07] | 3.72E-11 | 1.46E-09 |
| M_VLDL_C | Lipodystrophy_1_pPS | 4255 | 0.061031941 | [0.03 0.09] | 7.13E-05 | 0.001106313 |
| M_VLDL_C | Lipodystrophy_2_pPS | 4255 | -0.009983884 | [-0.04 0.02] | 0.51623455 | 0.770120395 |
| M_VLDL_C | Liver-Lipid_pPS | 4255 | -0.036379966 | [-0.07 -0.01] | 0.017972677 | 0.084078549 |
| M_VLDL_C | ASAT_Pos_pPS | 4255 | 0.027202212 | [-0. 0.06] | 0.076894849 | 0.262254664 |
| M_VLDL_C | ALP_Neg_pPS | 4255 | 0.018528064 | [-0.01 0.05] | 0.228286759 | 0.522699573 |
| M_VLDL_C | SHBG-LpA_pPS | 4255 | -0.005450336 | [-0.04 0.02] | 0.723054652 | 0.866319132 |
| M_VLDL_C | Obesity_pPS | 4255 | 0.030409789 | [0. 0.06] | 0.047965027 | 0.17948782 |
| M_VLDL_C | VAT_Neg_pPS | 4255 | 0.014570813 | [-0.02 0.04] | 0.343418017 | 0.637185237 |
| M_VLDL_C | Bilirubin_pPS | 4255 | -0.020856659 | [-0.05 0.01] | 0.175025563 | 0.453447086 |
| M_VLDL_C | Proinsulin_pPS | 4255 | 0.004030888 | [-0.03 0.03] | 0.793254935 | 0.900281394 |
| M_VLDL_C | Hyper_Insulin_pPS | 4255 | 0.001415362 | [-0.03 0.03] | 0.926677564 | 0.974374203 |
| M_VLDL_C | Beta_Cell_1_pPS | 4255 | 0.010826641 | [-0.02 0.04] | 0.481457382 | 0.740874464 |
| M_VLDL_C | Beta_Cell_2_pPS | 4255 | 0.00252283 | [-0.03 0.03] | 0.869705384 | 0.945763269 |
| M_VLDL_CE | Cholesterol_pPS | 4255 | -0.107253222 | [-0.14 -0.08] | 2.68E-12 | 1.09E-10 |
| M_VLDL_CE | Lipodystrophy_1_pPS | 4255 | 0.058432711 | [0.03 0.09] | 0.000143244 | 0.001964208 |
| M_VLDL_CE | Lipodystrophy_2_pPS | 4255 | -0.020847821 | [-0.05 0.01] | 0.175208471 | 0.453447086 |
| M_VLDL_CE | Liver-Lipid_pPS | 4255 | -0.026725917 | [-0.06 0. ] | 0.082209246 | 0.277174517 |
| M_VLDL_CE | ASAT_Pos_pPS | 4255 | 0.02475116 | [-0.01 0.05] | 0.107496598 | 0.340519287 |
| M_VLDL_CE | ALP_Neg_pPS | 4255 | 0.028218767 | [-0. 0.06] | 0.066486741 | 0.234342351 |
| M_VLDL_CE | SHBG-LpA_pPS | 4255 | -0.003271643 | [-0.03 0.03] | 0.831545151 | 0.92191249 |
| M_VLDL_CE | Obesity_pPS | 4255 | 0.026039657 | [-0. 0.06] | 0.090385915 | 0.300585719 |
| M_VLDL_CE | VAT_Neg_pPS | 4255 | 0.014428747 | [-0.02 0.04] | 0.348144299 | 0.637861136 |
| M_VLDL_CE | Bilirubin_pPS | 4255 | -0.025541886 | [-0.06 0. ] | 0.09671652 | 0.317420447 |
| M_VLDL_CE | Proinsulin_pPS | 4255 | 0.001132773 | [-0.03 0.03] | 0.94128723 | 0.978430443 |
| M_VLDL_CE | Hyper_Insulin_pPS | 4255 | -0.000627843 | [-0.03 0.03] | 0.967437882 | 0.987161386 |
| M_VLDL_CE | Beta_Cell_1_pPS | 4255 | 0.012018114 | [-0.02 0.04] | 0.434545698 | 0.699850608 |
| M_VLDL_CE | Beta_Cell_2_pPS | 4255 | 0.006617418 | [-0.02 0.04] | 0.667003516 | 0.849453588 |
| M_VLDL_FC | Cholesterol_pPS | 4255 | -0.089327043 | [-0.12 -0.06] | 5.90E-09 | 1.69E-07 |
| M_VLDL_FC | Lipodystrophy_1_pPS | 4255 | 0.062208298 | [0.03 0.09] | 5.15E-05 | 0.000852296 |
| M_VLDL_FC | Lipodystrophy_2_pPS | 4255 | 0.005104916 | [-0.03 0.04] | 0.739949756 | 0.873045882 |
| M_VLDL_FC | Liver-Lipid_pPS | 4255 | -0.047595143 | [-0.08 -0.02] | 0.001959162 | 0.015085545 |
| M_VLDL_FC | ASAT_Pos_pPS | 4255 | 0.02883893 | [-0. 0.06] | 0.060727401 | 0.219451729 |
| M_VLDL_FC | ALP_Neg_pPS | 4255 | 0.006249124 | [-0.02 0.04] | 0.684509567 | 0.858568103 |
| M_VLDL_FC | SHBG-LpA_pPS | 4255 | -0.009115507 | [-0.04 0.02] | 0.553384925 | 0.795753776 |
| M_VLDL_FC | Obesity_pPS | 4255 | 0.034473102 | [0. 0.06] | 0.024956146 | 0.107486249 |
| M_VLDL_FC | VAT_Neg_pPS | 4255 | 0.012851693 | [-0.02 0.04] | 0.403356529 | 0.677078992 |
| M_VLDL_FC | Bilirubin_pPS | 4255 | -0.013904377 | [-0.04 0.02] | 0.365944733 | 0.650982927 |
| M_VLDL_FC | Proinsulin_pPS | 4255 | 0.006885809 | [-0.02 0.04] | 0.654358677 | 0.844628798 |
| M_VLDL_FC | Hyper_Insulin_pPS | 4255 | 0.003696235 | [-0.03 0.03] | 0.810076628 | 0.911108657 |
| M_VLDL_FC | Beta_Cell_1_pPS | 4255 | 0.00806745 | [-0.02 0.04] | 0.599897491 | 0.821264125 |
| M_VLDL_FC | Beta_Cell_2_pPS | 4255 | -0.001010407 | [-0.03 0.03] | 0.947619939 | 0.978747744 |
| M_VLDL_L | Cholesterol_pPS | 4255 | -0.090890747 | [-0.12 -0.06] | 3.18E-09 | 9.65E-08 |
| M_VLDL_L | Lipodystrophy_1_pPS | 4255 | 0.061690599 | [0.03 0.09] | 5.95E-05 | 0.000960172 |
| M_VLDL_L | Lipodystrophy_2_pPS | 4255 | 0.004063477 | [-0.03 0.03] | 0.791621797 | 0.899824971 |
| M_VLDL_L | Liver-Lipid_pPS | 4255 | -0.045251596 | [-0.08 -0.02] | 0.003242809 | 0.022233233 |
| M_VLDL_L | ASAT_Pos_pPS | 4255 | 0.031430657 | [0. 0.06] | 0.040943468 | 0.15824097 |
| M_VLDL_L | ALP_Neg_pPS | 4255 | 0.004320312 | [-0.03 0.03] | 0.778783516 | 0.891327566 |
| M_VLDL_L | SHBG-LpA_pPS | 4255 | -0.009183624 | [-0.04 0.02] | 0.550424087 | 0.792838637 |
| M_VLDL_L | Obesity_pPS | 4255 | 0.034725153 | [0. 0.06] | 0.023915519 | 0.104084498 |
| M_VLDL_L | VAT_Neg_pPS | 4255 | 0.013102465 | [-0.02 0.04] | 0.394242562 | 0.672569903 |
| M_VLDL_L | Bilirubin_pPS | 4255 | -0.01416246 | [-0.04 0.02] | 0.357113929 | 0.643512228 |
| M_VLDL_L | Proinsulin_pPS | 4255 | 0.006071171 | [-0.02 0.04] | 0.693029998 | 0.858568103 |
| M_VLDL_L | Hyper_Insulin_pPS | 4255 | 0.004594462 | [-0.03 0.03] | 0.765146112 | 0.885433688 |
| M_VLDL_L | Beta_Cell_1_pPS | 4255 | 0.009025868 | [-0.02 0.04] | 0.557293093 | 0.796929123 |
| M_VLDL_L | Beta_Cell_2_pPS | 4255 | -0.001377094 | [-0.03 0.03] | 0.928654631 | 0.974930195 |
| M_VLDL_P | Cholesterol_pPS | 4255 | -0.089586819 | [-0.12 -0.06] | 5.33E-09 | 1.55E-07 |
| M_VLDL_P | Lipodystrophy_1_pPS | 4255 | 0.061347011 | [0.03 0.09] | 6.54E-05 | 0.001038839 |
| M_VLDL_P | Lipodystrophy_2_pPS | 4255 | 0.005152564 | [-0.02 0.04] | 0.737611536 | 0.87275313 |
| M_VLDL_P | Liver-Lipid_pPS | 4255 | -0.046029662 | [-0.08 -0.02] | 0.00274985 | 0.019591458 |
| M_VLDL_P | ASAT_Pos_pPS | 4255 | 0.031998225 | [0. 0.06] | 0.037430909 | 0.145791207 |
| M_VLDL_P | ALP_Neg_pPS | 4255 | 0.002991789 | [-0.03 0.03] | 0.845765387 | 0.930853384 |
| M_VLDL_P | SHBG-LpA_pPS | 4255 | -0.009550622 | [-0.04 0.02] | 0.534607382 | 0.781586915 |
| M_VLDL_P | Obesity_pPS | 4255 | 0.034944564 | [0. 0.07] | 0.023040486 | 0.101378139 |
| M_VLDL_P | VAT_Neg_pPS | 4255 | 0.013001833 | [-0.02 0.04] | 0.397884833 | 0.673455453 |
| M_VLDL_P | Bilirubin_pPS | 4255 | -0.013425509 | [-0.04 0.02] | 0.382687299 | 0.661657399 |
| M_VLDL_P | Proinsulin_pPS | 4255 | 0.006261449 | [-0.02 0.04] | 0.683920923 | 0.858568103 |
| M_VLDL_P | Hyper_Insulin_pPS | 4255 | 0.004886459 | [-0.03 0.04] | 0.750700589 | 0.876790721 |

**Table S9: Associations between pPSs and global metabolomic profile in SCAPIS**

**a) Spearman Correlation**

| Metabolomics | PPS | Samp | r | CI95% | p | fdr_p_adjusted |
|---|---|---|---|---|---|---|
| M_LDL_TG | Total_GRS | 4255 | 0.012494314 | [-0.02 0.04] | 0.415845178 | 0.677830746 |
| M_VLDL_C | Cholesterol_pPS | 4255 | -0.10043409 | [-0.13 -0.07] | 5.50E-11 | 2.27E-09 |
| M_VLDL_C | Lipodystrophy_1_pPS | 4255 | 0.067436983 | [0.04 0.1 ] | 1.10E-05 | 0.000193471 |
| M_VLDL_C | Lipodystrophy_2_pPS | 4255 | -0.006981309 | [-0.04 0.02] | 0.649381036 | 0.818487176 |
| M_VLDL_C | Liver-Lipid_pPS | 4255 | -0.035756788 | [-0.07 -0.01] | 0.019848716 | 0.084811744 |
| M_VLDL_C | ASAT_Pos_pPS | 4255 | 0.039225516 | [0.01 0.07] | 0.010609187 | 0.049363789 |
| M_VLDL_C | ALP_Neg_pPS | 4255 | 0.019681556 | [-0.01 0.05] | 0.199923425 | 0.474901159 |
| M_VLDL_C | SHBG-LpA_pPS | 4255 | 0.00110488 | [-0.03 0.03] | 0.942642648 | 0.973139992 |
| M_VLDL_C | Obesity_pPS | 4255 | 0.025466866 | [-0. 0.06] | 0.097186559 | 0.29695893 |
| M_VLDL_C | VAT_Neg_pPS | 4255 | 0.019818217 | [-0.01 0.05] | 0.196817572 | 0.47103115 |
| M_VLDL_C | Bilirubin_pPS | 4255 | -0.017117724 | [-0.05 0.01] | 0.264947773 | 0.551490946 |
| M_VLDL_C | Proinsulin_pPS | 4255 | 0.012621658 | [-0.02 0.04] | 0.411108765 | 0.673665623 |
| M_VLDL_C | Hyper_Insulin_pPS | 4255 | 0.010893754 | [-0.02 0.04] | 0.478068401 | 0.714911237 |
| M_VLDL_C | Beta_Cell_1_pPS | 4255 | 0.018300752 | [-0.01 0.05] | 0.233328431 | 0.516301672 |
| M_VLDL_C | Beta_Cell_2_pPS | 4255 | 0.005861054 | [-0.02 0.04] | 0.702706164 | 0.855830536 |
| M_VLDL_C | Total_GRS | 4255 | 0.033272323 | [0. 0.06] | 0.030214694 | 0.119136982 |
| M_VLDL_CE | Cholesterol_pPS | 4255 | -0.106885716 | [-0.14 -0.08] | 2.95E-12 | 1.29E-10 |
| M_VLDL_CE | Lipodystrophy_1_pPS | 4255 | 0.063604001 | [0.03 0.09] | 3.38E-05 | 0.000533541 |
| M_VLDL_CE | Lipodystrophy_2_pPS | 4255 | -0.018315353 | [-0.05 0.01] | 0.232955645 | 0.516301672 |
| M_VLDL_CE | Liver-Lipid_pPS | 4255 | -0.026228719 | [-0.06 0. ] | 0.087583471 | 0.271608134 |
| M_VLDL_CE | ASAT_Pos_pPS | 4255 | 0.0349085 | [0. 0.06] | 0.022971767 | 0.095493101 |
| M_VLDL_CE | ALP_Neg_pPS | 4255 | 0.029068664 | [-0. 0.06] | 0.058314538 | 0.195952676 |
| M_VLDL_CE | SHBG-LpA_pPS | 4255 | 0.002761908 | [-0.03 0.03] | 0.857267704 | 0.934369525 |
| M_VLDL_CE | Obesity_pPS | 4255 | 0.020239722 | [-0.01 0.05] | 0.187460833 | 0.459363377 |
| M_VLDL_CE | VAT_Neg_pPS | 4255 | 0.018816851 | [-0.01 0.05] | 0.220407485 | 0.501351065 |
| M_VLDL_CE | Bilirubin_pPS | 4255 | -0.022022715 | [-0.05 0.01] | 0.151494149 | 0.401178951 |
| M_VLDL_CE | Proinsulin_pPS | 4255 | 0.011751845 | [-0.02 0.04] | 0.444095159 | 0.692953671 |
| M_VLDL_CE | Hyper_Insulin_pPS | 4255 | 0.007954313 | [-0.02 0.04] | 0.604467981 | 0.791565213 |
| M_VLDL_CE | Beta_Cell_1_pPS | 4255 | 0.019945244 | [-0.01 0.05] | 0.193962461 | 0.466423181 |
| M_VLDL_CE | Beta_Cell_2_pPS | 4255 | 0.009921238 | [-0.02 0.04] | 0.518228701 | 0.736667707 |
| M_VLDL_CE | Total_GRS | 4255 | 0.026807493 | [-0. 0.06] | 0.080811965 | 0.254539888 |
| M_VLDL_FC | Cholesterol_pPS | 4255 | -0.08739117 | [-0.12 -0.06] | 1.19E-08 | 3.48E-07 |
| M_VLDL_FC | Lipodystrophy_1_pPS | 4255 | 0.06974816 | [0.04 0.1 ] | 5.43E-06 | 0.000106052 |
| M_VLDL_FC | Lipodystrophy_2_pPS | 4255 | 0.008443525 | [-0.02 0.04] | 0.582425865 | 0.778303354 |
| M_VLDL_FC | Liver-Lipid_pPS | 4255 | -0.046818987 | [-0.08 -0.02] | 0.002284728 | 0.015362825 |
| M_VLDL_FC | ASAT_Pos_pPS | 4255 | 0.042779678 | [0.01 0.07] | 0.005319012 | 0.028381297 |
| M_VLDL_FC | ALP_Neg_pPS | 4255 | 0.007665481 | [-0.02 0.04] | 0.617654904 | 0.799138801 |
| M_VLDL_FC | SHBG-LpA_pPS | 4255 | -0.002266126 | [-0.03 0.03] | 0.882683452 | 0.945095197 |
| M_VLDL_FC | Obesity_pPS | 4255 | 0.030834235 | [0. 0.06] | 0.044603372 | 0.160975011 |
| M_VLDL_FC | VAT_Neg_pPS | 4255 | 0.018976678 | [-0.01 0.05] | 0.21651214 | 0.497544682 |
| M_VLDL_FC | Bilirubin_pPS | 4255 | -0.010041498 | [-0.04 0.02] | 0.513169771 | 0.733343877 |
| M_VLDL_FC | Proinsulin_pPS | 4255 | 0.012604713 | [-0.02 0.04] | 0.411737161 | 0.674180313 |
| M_VLDL_FC | Hyper_Insulin_pPS | 4255 | 0.013884357 | [-0.02 0.04] | 0.365899584 | 0.64109339 |
| M_VLDL_FC | Beta_Cell_1_pPS | 4255 | 0.014668476 | [-0.02 0.04] | 0.339451454 | 0.622327666 |
| M_VLDL_FC | Beta_Cell_2_pPS | 4255 | 0.001828902 | [-0.03 0.03] | 0.905198851 | 0.952872131 |
| M_VLDL_FC | Total_GRS | 4255 | 0.040300215 | [0.01 0.07] | 0.008655203 | 0.04238678 |
| M_VLDL_L | Cholesterol_pPS | 4255 | -0.088968457 | [-0.12 -0.06] | 6.43E-09 | 2.03E-07 |
| M_VLDL_L | Lipodystrophy_1_pPS | 4255 | 0.069724407 | [0.04 0.1 ] | 5.47E-06 | 0.000106052 |
| M_VLDL_L | Lipodystrophy_2_pPS | 4255 | 0.007669384 | [-0.02 0.04] | 0.617475847 | 0.799138801 |
| M_VLDL_L | Liver-Lipid_pPS | 4255 | -0.044559203 | [-0.07 -0.01] | 0.003694909 | 0.021654589 |
| M_VLDL_L | ASAT_Pos_pPS | 4255 | 0.045369888 | [0.02 0.08] | 0.003116881 | 0.019491862 |
| M_VLDL_L | ALP_Neg_pPS | 4255 | 0.0056757 | [-0.02 0.04] | 0.71168085 | 0.8619737 |
| M_VLDL_L | SHBG-LpA_pPS | 4255 | -0.002283388 | [-0.03 0.03] | 0.881796342 | 0.945095197 |
| M_VLDL_L | Obesity_pPS | 4255 | 0.030962739 | [0. 0.06] | 0.043721624 | 0.158291421 |
| M_VLDL_L | VAT_Neg_pPS | 4255 | 0.019335703 | [-0.01 0.05] | 0.207943036 | 0.486940844 |
| M_VLDL_L | Bilirubin_pPS | 4255 | -0.01026102 | [-0.04 0.02] | 0.504001973 | 0.727744052 |
| M_VLDL_L | Proinsulin_pPS | 4255 | 0.011734228 | [-0.02 0.04] | 0.444778479 | 0.692953671 |
| M_VLDL_L | Hyper_Insulin_pPS | 4255 | 0.014926892 | [-0.02 0.04] | 0.331010635 | 0.612969968 |
| M_VLDL_L | Beta_Cell_1_pPS | 4255 | 0.015564602 | [-0.01 0.05] | 0.310766692 | 0.598916941 |
| M_VLDL_L | Beta_Cell_2_pPS | 4255 | 0.00160002 | [-0.03 0.03] | 0.917017033 | 0.957786011 |
| M_VLDL_L | Total_GRS | 4255 | 0.040168177 | [0.01 0.07] | 0.008876549 | 0.043175051 |
| M_VLDL_P | Cholesterol_pPS | 4255 | -0.087610351 | [-0.12 -0.06] | 1.09E-08 | 3.25E-07 |
| M_VLDL_P | Lipodystrophy_1_pPS | 4255 | 0.069536731 | [0.04 0.1 ] | 5.80E-06 | 0.000111024 |
| M_VLDL_P | Lipodystrophy_2_pPS | 4255 | 0.008823927 | [-0.02 0.04] | 0.565549989 | 0.768674587 |
| M_VLDL_P | Liver-Lipid_pPS | 4255 | -0.045342451 | [-0.08 -0.02] | 0.003135011 | 0.019548254 |
| M_VLDL_P | ASAT_Pos_pPS | 4255 | 0.046019932 | [0.02 0.08] | 0.002714314 | 0.01769667 |
| M_VLDL_P | ALP_Neg_pPS | 4255 | 0.004353513 | [-0.03 0.03] | 0.776794883 | 0.89275053 |
| M_VLDL_P | SHBG-LpA_pPS | 4255 | -0.00265418 | [-0.03 0.03] | 0.862778617 | 0.937517799 |
| M_VLDL_P | Obesity_pPS | 4255 | 0.031309649 | [0. 0.06] | 0.041414432 | 0.151852919 |
| M_VLDL_P | VAT_Neg_pPS | 4255 | 0.019264661 | [-0.01 0.05] | 0.209618827 | 0.488466046 |
| M_VLDL_P | Bilirubin_pPS | 4255 | -0.009519538 | [-0.04 0.02] | 0.535311756 | 0.749995896 |
| M_VLDL_P | Proinsulin_pPS | 4255 | 0.011602328 | [-0.02 0.04] | 0.449913735 | 0.694291339 |
| M_VLDL_P | Hyper_Insulin_pPS | 4255 | 0.015244996 | [-0.01 0.05] | 0.320808185 | 0.608204326 |
| M_VLDL_P | Beta_Cell_1_pPS | 4255 | 0.015230876 | [-0.01 0.05] | 0.321256644 | 0.608204326 |
| M_VLDL_P | Beta_Cell_2_pPS | 4255 | 0.001084728 | [-0.03 0.03] | 0.943687014 | 0.973645332 |
| M_VLDL_P | Total_GRS | 4255 | 0.040547875 | [0.01 0.07] | 0.008253291 | 0.040697263 |
| M_VLDL_PL | Cholesterol_pPS | 4255 | -0.09237011 | [-0.12 -0.06] | 1.66E-09 | 5.74E-08 |
| M_VLDL_PL | Lipodystrophy_1_pPS | 4255 | 0.070652682 | [0.04 0.1 ] | 4.09E-06 | 8.36E-05 |
| M_VLDL_PL | Lipodystrophy_2_pPS | 4255 | 0.007174914 | [-0.02 0.04] | 0.640335258 | 0.813214404 |
| M_VLDL_PL | Liver-Lipid_pPS | 4255 | -0.043135927 | [-0.07 -0.01] | 0.004949816 | 0.026743968 |
| M_VLDL_PL | ASAT_Pos_pPS | 4255 | 0.044180179 | [0.01 0.07] | 0.003997235 | 0.023089876 |
| M_VLDL_PL | ALP_Neg_pPS | 4255 | 0.007724099 | [-0.02 0.04] | 0.614968458 | 0.7975256 |
| M_VLDL_PL | SHBG-LpA_pPS | 4255 | -0.001179163 | [-0.03 0.03] | 0.938793744 | 0.970463894 |
| M_VLDL_PL | Obesity_pPS | 4255 | 0.029788411 | [-0. 0.06] | 0.05235227 | 0.18053958 |
| M_VLDL_PL | VAT_Neg_pPS | 4255 | 0.019122414 | [-0.01 0.05] | 0.213003615 | 0.493880952 |
| M_VLDL_PL | Bilirubin_pPS | 4255 | -0.010299018 | [-0.04 0.02] | 0.502423882 | 0.727744052 |
| M_VLDL_PL | Proinsulin_pPS | 4255 | 0.011795511 | [-0.02 0.04] | 0.442404006 | 0.692820992 |
| M_VLDL_PL | Hyper_Insulin_pPS | 4255 | 0.014179682 | [-0.02 0.04] | 0.355791052 | 0.635109235 |
| M_VLDL_PL | Beta_Cell_1_pPS | 4255 | 0.016350862 | [-0.01 0.05] | 0.286955147 | 0.57504794 |
| M_VLDL_PL | Beta_Cell_2_pPS | 4255 | 0.001787106 | [-0.03 0.03] | 0.907355457 | 0.952872131 |
| M_VLDL_PL | Total_GRS | 4255 | 0.039836967 | [0.01 0.07] | 0.009454182 | 0.045367382 |
| M_VLDL_TG | Cholesterol_pPS | 4255 | -0.079432745 | [-0.11 -0.05] | 2.21E-07 | 5.51E-06 |
| M_VLDL_TG | Lipodystrophy_1_pPS | 4255 | 0.068616324 | [0.04 0.1 ] | 7.69E-06 | 0.000141069 |
| M_VLDL_TG | Lipodystrophy_2_pPS | 4255 | 0.015257944 | [-0.01 0.05] | 0.320397324 | 0.608204326 |

**b) Partial Correlation**

| Metabolomics | PPS | Samp | r | CI95% | p | fdr_p_adjusted |
|---|---|---|---|---|---|---|
| M_VLDL_P | Beta_Cell_1_pPS | 4255 | 0.008808076 | [-0.02 0.04] | 0.566844176 | 0.797755947 |
| M_VLDL_P | Beta_Cell_2_pPS | 4255 | -0.001841068 | [-0.03 0.03] | 0.904716892 | 0.96224905 |
| M_VLDL_PL | Cholesterol_pPS | 4255 | -0.094218627 | [-0.12 -0.06] | 8.27E-10 | 2.67E-08 |
| M_VLDL_PL | Lipodystrophy_1_pPS | 4255 | 0.062844237 | [0.03 0.09] | 4.31E-05 | 0.000757137 |
| M_VLDL_PL | Lipodystrophy_2_pPS | 4255 | 0.003810458 | [-0.03 0.03] | 0.804324806 | 0.907699133 |
| M_VLDL_PL | Liver-Lipid_pPS | 4255 | -0.043803912 | [-0.07 -0.01] | 0.004379293 | 0.028368318 |
| M_VLDL_PL | ASAT_Pos_pPS | 4255 | 0.030116796 | [-0. 0.06] | 0.050157881 | 0.186646984 |
| M_VLDL_PL | ALP_Neg_pPS | 4255 | 0.00641375 | [-0.02 0.04] | 0.676662759 | 0.855435982 |
| M_VLDL_PL | SHBG-LpA_pPS | 4255 | -0.008086254 | [-0.04 0.02] | 0.599047631 | 0.820871565 |
| M_VLDL_PL | Obesity_pPS | 4255 | 0.033906192 | [0. 0.06] | 0.027440875 | 0.115656067 |
| M_VLDL_PL | VAT_Neg_pPS | 4255 | 0.012681796 | [-0.02 0.04] | 0.409602278 | 0.682216107 |
| M_VLDL_PL | Bilirubin_pPS | 4255 | -0.014176771 | [-0.04 0.02] | 0.356628194 | 0.643215897 |
| M_VLDL_PL | Proinsulin_pPS | 4255 | 0.005654173 | [-0.02 0.04] | 0.71314696 | 0.86476088 |
| M_VLDL_PL | Hyper_Insulin_pPS | 4255 | 0.003854138 | [-0.03 0.03] | 0.802128016 | 0.906328459 |
| M_VLDL_PL | Beta_Cell_1_pPS | 4255 | 0.009856361 | [-0.02 0.04] | 0.521608072 | 0.773759954 |
| M_VLDL_PL | Beta_Cell_2_pPS | 4255 | -0.001342902 | [-0.03 0.03] | 0.930421536 | 0.976259914 |
| M_VLDL_TG | Cholesterol_pPS | 4255 | -0.081819313 | [-0.11 -0.05] | 9.89E-08 | 2.32E-06 |
| M_VLDL_TG | Lipodystrophy_1_pPS | 4255 | 0.059874431 | [0.03 0.09] | 9.76E-05 | 0.001426225 |
| M_VLDL_TG | Lipodystrophy_2_pPS | 4255 | 0.011377383 | [-0.02 0.04] | 0.459437995 | 0.72261785 |
| M_VLDL_TG | Liver-Lipid_pPS | 4255 | -0.050405987 | [-0.08 -0.02] | 0.001040122 | 0.008975536 |
| M_VLDL_TG | ASAT_Pos_pPS | 4255 | 0.033202597 | [0. 0.06] | 0.030818948 | 0.124645523 |
| M_VLDL_TG | ALP_Neg_pPS | 4255 | -0.003211123 | [-0.03 0.03] | 0.834615846 | 0.923149681 |
| M_VLDL_TG | SHBG-LpA_pPS | 4255 | -0.010981194 | [-0.04 0.02] | 0.475220837 | 0.735233474 |
| M_VLDL_TG | Obesity_pPS | 4255 | 0.036904764 | [0.01 0.07] | 0.016379989 | 0.07895582 |
| M_VLDL_TG | VAT_Neg_pPS | 4255 | 0.012281009 | [-0.02 0.04] | 0.424561927 | 0.693893617 |
| M_VLDL_TG | Bilirubin_pPS | 4255 | -0.010592385 | [-0.04 0.02] | 0.49099446 | 0.751475077 |
| M_VLDL_TG | Proinsulin_pPS | 4255 | 0.007763385 | [-0.02 0.04] | 0.613715544 | 0.828794354 |
| M_VLDL_TG | Hyper_Insulin_pPS | 4255 | 0.006095456 | [-0.02 0.04] | 0.691864891 | 0.858568103 |
| M_VLDL_TG | Beta_Cell_1_pPS | 4255 | 0.007375546 | [-0.02 0.04] | 0.63154115 | 0.835456904 |
| M_VLDL_TG | Beta_Cell_2_pPS | 4255 | 0.003269471 | [-0.03 0.03] | 0.831655323 | 0.92191249 |
| MUFA | Cholesterol_pPS | 4223 | -0.110242044 | [-0.14 -0.08] | 7.92E-13 | 3.45E-11 |
| MUFA | Lipodystrophy_1_pPS | 4223 | 0.05093741 | [0.02 0.08] | 0.000961708 | 0.008545465 |
| MUFA | Lipodystrophy_2_pPS | 4223 | -0.02357142 | [-0.05 0.01] | 0.126761963 | 0.379116795 |
| MUFA | Liver-Lipid_pPS | 4223 | -0.058151881 | [-0.09 -0.03] | 0.00016335 | 0.002180178 |
| MUFA | ASAT_Pos_pPS | 4223 | 0.00958941 | [-0.02 0.04] | 0.534503538 | 0.781586915 |
| MUFA | ALP_Neg_pPS | 4223 | 0.034748463 | [0. 0.06] | 0.024358256 | 0.105781405 |
| MUFA | SHBG-LpA_pPS | 4223 | 0.001963945 | [-0.03 0.03] | 0.898774092 | 0.95862852 |
| MUFA | Obesity_pPS | 4223 | 0.03996398 | [0.01 0.07] | 0.009608512 | 0.052335396 |
| MUFA | VAT_Neg_pPS | 4223 | -0.005135934 | [-0.04 0.03] | 0.739383072 | 0.873045882 |
| MUFA | Bilirubin_pPS | 4223 | -0.017278148 | [-0.05 0.01] | 0.263039699 | 0.559637055 |
| MUFA | Proinsulin_pPS | 4223 | -0.001223865 | [-0.03 0.03] | 0.936815436 | 0.978170683 |
| MUFA | Hyper_Insulin_pPS | 4223 | -0.005657956 | [-0.04 0.02] | 0.714004007 | 0.865276043 |
| MUFA | Beta_Cell_1_pPS | 4223 | 0.020037864 | [-0.01 0.05] | 0.194274802 | 0.478398712 |
| MUFA | Beta_Cell_2_pPS | 4223 | 0.001127322 | [-0.03 0.03] | 0.941790454 | 0.978445505 |
| PC | Cholesterol_pPS | 4223 | -0.087429901 | [-0.12 -0.06] | 1.40E-08 | 3.83E-07 |
| PC | Lipodystrophy_1_pPS | 4223 | 0.022243858 | [-0.01 0.05] | 0.149592431 | 0.425214661 |
| PC | Lipodystrophy_2_pPS | 4223 | -0.04046696 | [-0.07 -0.01] | 0.00873571 | 0.048715573 |
| PC | Liver-Lipid_pPS | 4223 | -0.0410261 | [-0.07 -0.01] | 0.007849176 | 0.044769376 |
| PC | ASAT_Pos_pPS | 4223 | -0.030406677 | [-0.06 -0. ] | 0.048840962 | 0.182424637 |
| PC | ALP_Neg_pPS | 4223 | 0.061184963 | [0.03 0.09] | 7.28E-05 | 0.001121654 |
| PC | SHBG-LpA_pPS | 4223 | 0.017982823 | [-0.01 0.05] | 0.244063603 | 0.541702144 |
| PC | Obesity_pPS | 4223 | 0.007267232 | [-0.02 0.04] | 0.637837619 | 0.835456904 |
| PC | VAT_Neg_pPS | 4223 | -0.015991732 | [-0.05 0.01] | 0.300251787 | 0.597661337 |
| PC | Bilirubin_pPS | 4223 | -0.011729034 | [-0.04 0.02] | 0.447405754 | 0.709019056 |
| PC | Proinsulin_pPS | 4223 | -0.009377959 | [-0.04 0.02] | 0.543553049 | 0.788545801 |
| PC | Hyper_Insulin_pPS | 4223 | -0.016737636 | [-0.05 0.01] | 0.278268378 | 0.576656518 |
| PC | Beta_Cell_1_pPS | 4223 | 0.018161344 | [-0.01 0.05] | 0.239412743 | 0.535744184 |
| PC | Beta_Cell_2_pPS | 4223 | 0.014458923 | [-0.02 0.04] | 0.348968385 | 0.637861136 |
| Phe | Cholesterol_pPS | 4254 | -0.006092362 | [-0.04 0.02] | 0.692047844 | 0.858568103 |
| Phe | Lipodystrophy_1_pPS | 4254 | 0.005088741 | [-0.03 0.04] | 0.740773645 | 0.873045882 |
| Phe | Lipodystrophy_2_pPS | 4254 | -0.00023318 | [-0.03 0.03] | 0.987905045 | 0.993363084 |
| Phe | Liver-Lipid_pPS | 4254 | 0.024313706 | [-0.01 0.05] | 0.113899736 | 0.351893938 |
| Phe | ASAT_Pos_pPS | 4254 | 0.021858567 | [-0.01 0.05] | 0.155250237 | 0.42902716 |
| Phe | ALP_Neg_pPS | 4254 | 0.001777641 | [-0.03 0.03] | 0.907995533 | 0.96224905 |
| Phe | SHBG-LpA_pPS | 4254 | -0.006839194 | [-0.04 0.02] | 0.656585918 | 0.844628798 |
| Phe | Obesity_pPS | 4254 | 0.004457401 | [-0.03 0.03] | 0.771981522 | 0.889756481 |
| Phe | VAT_Neg_pPS | 4254 | -0.014824144 | [-0.04 0.02] | 0.33514955 | 0.63058718 |
| Phe | Bilirubin_pPS | 4254 | 0.000995536 | [-0.03 0.03] | 0.948395846 | 0.978747744 |
| Phe | Proinsulin_pPS | 4254 | -0.006224055 | [-0.04 0.02] | 0.685742652 | 0.858568103 |
| Phe | Hyper_Insulin_pPS | 4254 | 0.010785481 | [-0.02 0.04] | 0.483177536 | 0.742950405 |
| Phe | Beta_Cell_1_pPS | 4254 | 0.00953734 | [-0.02 0.04] | 0.535224062 | 0.781586915 |
| Phe | Beta_Cell_2_pPS | 4254 | -0.009830065 | [-0.04 0.02] | 0.52276887 | 0.774477051 |
| PUFA | Cholesterol_pPS | 4223 | -0.133346883 | [-0.16 -0.1 ] | 4.10E-18 | 2.56E-16 |
| PUFA | Lipodystrophy_1_pPS | 4223 | 0.037815949 | [0.01 0.07] | 0.01427306 | 0.071081259 |
| PUFA | Lipodystrophy_2_pPS | 4223 | -0.047305483 | [-0.08 -0.02] | 0.002170745 | 0.016276523 |
| PUFA | Liver-Lipid_pPS | 4223 | -0.028201577 | [-0.06 0. ] | 0.067691484 | 0.236920195 |
| PUFA | ASAT_Pos_pPS | 4223 | -0.024661921 | [-0.05 0.01] | 0.110118284 | 0.343926372 |
| PUFA | ALP_Neg_pPS | 4223 | 0.060575376 | [0.03 0.09] | 8.59E-05 | 0.001293444 |
| PUFA | SHBG-LpA_pPS | 4223 | 0.015133912 | [-0.02 0.05] | 0.326928091 | 0.625128976 |
| PUFA | Obesity_pPS | 4223 | 0.018386086 | [-0.01 0.05] | 0.233647008 | 0.529741009 |
| PUFA | VAT_Neg_pPS | 4223 | -0.004060989 | [-0.03 0.03] | 0.79251802 | 0.899955233 |
| PUFA | Bilirubin_pPS | 4223 | -0.024621659 | [-0.05 0.01] | 0.110700471 | 0.344222613 |
| PUFA | Proinsulin_pPS | 4223 | -0.012785843 | [-0.04 0.02] | 0.407553204 | 0.68050168 |
| PUFA | Hyper_Insulin_pPS | 4223 | -0.020469349 | [-0.05 0.01] | 0.184841857 | 0.466062214 |
| PUFA | Beta_Cell_1_pPS | 4223 | 0.027075136 | [-0. 0.06] | 0.079421833 | 0.269495778 |
| PUFA | Beta_Cell_2_pPS | 4223 | 0.024355522 | [-0.01 0.05] | 0.11461008 | 0.352999047 |
| Remnant_C | Cholesterol_pPS | 4254 | -0.140604617 | [-0.17 -0.11] | 4.05E-20 | 2.90E-18 |
| Remnant_C | Lipodystrophy_1_pPS | 4254 | 0.059752792 | [0.03 0.09] | 0.000101034 | 0.001455181 |
| Remnant_C | Lipodystrophy_2_pPS | 4254 | -0.035124784 | [-0.07 -0. ] | 0.022358545 | 0.099692223 |
| Remnant_C | Liver-Lipid_pPS | 4254 | -0.009532757 | [-0.04 0.02] | 0.53542022 | 0.781586915 |
| Remnant_C | ASAT_Pos_pPS | 4254 | -0.000533459 | [-0.03 0.03] | 0.972334109 | 0.989702967 |
| Remnant_C | ALP_Neg_pPS | 4254 | 0.048446358 | [0.02 0.08] | 0.001624772 | 0.012959334 |
| Remnant_C | SHBG-LpA_pPS | 4254 | 0.011791393 | [-0.02 0.04] | 0.44331746 | 0.706833249 |
| Remnant_C | Obesity_pPS | 4254 | 0.018419584 | [-0.01 0.05] | 0.231077803 | 0.526899501 |

**Table S9: Associations between pPSs and global metabolomic profile in SCAPIS**

**a) Spearman Correlation**

| Metabolomics | PPS | Samp | r | CI95% | p | fdr_p_adjusted |
|---|---|---|---|---|---|---|
| M_VLDL_TG | Liver-Lipid_pPS | 4255 | -0.049646654 | [-0.08 -0.02] | 0.001216677 | 0.009737958 |
| M_VLDL_TG | ASAT_Pos_pPS | 4255 | 0.04768808 | [0.02 0.08] | 0.001888854 | 0.013595948 |
| M_VLDL_TG | ALP_Neg_pPS | 4255 | -0.001714442 | [-0.03 0.03] | 0.91110646 | 0.954725626 |
| M_VLDL_TG | SHBG-LpA_pPS | 4255 | -0.004090694 | [-0.03 0.03] | 0.789943859 | 0.899856387 |
| M_VLDL_TG | Obesity_pPS | 4255 | 0.034045074 | [0. 0.06] | 0.026580196 | 0.10737198 |
| M_VLDL_TG | VAT_Neg_pPS | 4255 | 0.018769229 | [-0.01 0.05] | 0.2215778 | 0.502934916 |
| M_VLDL_TG | Bilirubin_pPS | 4255 | -0.00667348 | [-0.04 0.02] | 0.663870225 | 0.825654931 |
| M_VLDL_TG | Proinsulin_pPS | 4255 | 0.011758101 | [-0.02 0.04] | 0.443852633 | 0.692953671 |
| M_VLDL_TG | Hyper_Insulin_pPS | 4255 | 0.016607011 | [-0.01 0.05] | 0.279470948 | 0.566067218 |
| M_VLDL_TG | Beta_Cell_1_pPS | 4255 | 0.013459171 | [-0.02 0.04] | 0.380764192 | 0.652142798 |
| M_VLDL_TG | Beta_Cell_2_pPS | 4255 | -0.000549009 | [-0.03 0.03] | 0.971480911 | 0.983305352 |
| M_VLDL_TG | Total_GRS | 4255 | 0.043377218 | [0.01 0.07] | 0.004713084 | 0.025724087 |
| MUFA | Cholesterol_pPS | 4223 | -0.108802485 | [-0.14 -0.08] | 1.45E-12 | 6.61E-11 |
| MUFA | Lipodystrophy_1_pPS | 4223 | 0.052841097 | [0.02 0.08] | 0.000602793 | 0.005958484 |
| MUFA | Lipodystrophy_2_pPS | 4223 | -0.026802541 | [-0.06 0. ] | 0.082022027 | 0.257218197 |
| MUFA | Liver-Lipid_pPS | 4223 | -0.058020568 | [-0.09 -0.03] | 0.000165129 | 0.002059312 |
| MUFA | ASAT_Pos_pPS | 4223 | 0.017935716 | [-0.01 0.05] | 0.244571378 | 0.525480742 |
| MUFA | ALP_Neg_pPS | 4223 | 0.033353114 | [0. 0.06] | 0.030440019 | 0.119585788 |
| MUFA | SHBG-LpA_pPS | 4223 | 0.005144985 | [-0.03 0.04] | 0.73855118 | 0.874278301 |
| MUFA | Obesity_pPS | 4223 | 0.032089207 | [0. 0.06] | 0.037318926 | 0.140191061 |
| MUFA | VAT_Neg_pPS | 4223 | -0.001786549 | [-0.03 0.03] | 0.907732764 | 0.952872131 |
| MUFA | Bilirubin_pPS | 4223 | -0.013813745 | [-0.04 0.02] | 0.370156017 | 0.643918869 |
| MUFA | Proinsulin_pPS | 4223 | 0.012162959 | [-0.02 0.04] | 0.430067457 | 0.688963257 |
| MUFA | Hyper_Insulin_pPS | 4223 | -0.001613534 | [-0.03 0.03] | 0.916633878 | 0.957786011 |
| MUFA | Beta_Cell_1_pPS | 4223 | 0.022984628 | [-0.01 0.05] | 0.135888605 | 0.377077696 |
| MUFA | Beta_Cell_2_pPS | 4223 | 0.003866372 | [-0.03 0.03] | 0.801950281 | 0.907269701 |
| MUFA | Total_GRS | 4223 | 0.019004387 | [-0.01 0.05] | 0.217582466 | 0.498626484 |
| PC | Cholesterol_pPS | 4223 | -0.088979956 | [-0.12 -0.06] | 7.30E-09 | 2.27E-07 |
| PC | Lipodystrophy_1_pPS | 4223 | 0.014570546 | [-0.02 0.04] | 0.344513628 | 0.62662761 |
| PC | Lipodystrophy_2_pPS | 4223 | -0.049634924 | [-0.08 -0.02] | 0.001273214 | 0.010103761 |
| PC | Liver-Lipid_pPS | 4223 | -0.041548162 | [-0.07 -0.01] | 0.007006897 | 0.035700223 |
| PC | ASAT_Pos_pPS | 4223 | -0.031771244 | [-0.06 -0. ] | 0.039244422 | 0.146144592 |
| PC | ALP_Neg_pPS | 4223 | 0.057705146 | [0.03 0.09] | 0.00017923 | 0.002222239 |
| PC | SHBG-LpA_pPS | 4223 | 0.016641032 | [-0.01 0.05] | 0.280308017 | 0.567194465 |
| PC | Obesity_pPS | 4223 | -0.002063212 | [-0.03 0.03] | 0.893523739 | 0.948580863 |
| PC | VAT_Neg_pPS | 4223 | -0.016127991 | [-0.05 0.01] | 0.295404793 | 0.58132411 |
| PC | Bilirubin_pPS | 4223 | -0.010719916 | [-0.04 0.02] | 0.486771707 | 0.717611897 |
| PC | Proinsulin_pPS | 4223 | 0.006351954 | [-0.02 0.04] | 0.680285044 | 0.83766442 |
| PC | Hyper_Insulin_pPS | 4223 | -0.021641594 | [-0.05 0.01] | 0.16028355 | 0.417242979 |
| PC | Beta_Cell_1_pPS | 4223 | 0.018812001 | [-0.01 0.05] | 0.222276131 | 0.502934916 |
| PC | Beta_Cell_2_pPS | 4223 | 0.015241589 | [-0.01 0.05] | 0.322749112 | 0.608687153 |
| PC | Total_GRS | 4223 | -0.011880959 | [-0.04 0.02] | 0.440837283 | 0.691730776 |
| Phe | Cholesterol_pPS | 4254 | -0.004840478 | [-0.03 0.03] | 0.752630812 | 0.881218937 |
| Phe | Lipodystrophy_1_pPS | 4254 | 0.010204557 | [-0.02 0.04] | 0.50640187 | 0.729504373 |
| Phe | Lipodystrophy_2_pPS | 4254 | 0.00105295 | [-0.03 0.03] | 0.94534058 | 0.974414005 |
| Phe | Liver-Lipid_pPS | 4254 | 0.024684556 | [-0.01 0.05] | 0.107946482 | 0.318932788 |
| Phe | ASAT_Pos_pPS | 4254 | 0.024825987 | [-0.01 0.05] | 0.105941979 | 0.314744523 |
| Phe | ALP_Neg_pPS | 4254 | 0.002264452 | [-0.03 0.03] | 0.882783218 | 0.945095197 |
| Phe | SHBG-LpA_pPS | 4254 | -0.007467944 | [-0.04 0.02] | 0.626786107 | 0.803929814 |
| Phe | Obesity_pPS | 4254 | 0.005182794 | [-0.02 0.04] | 0.735769041 | 0.873878512 |
| Phe | VAT_Neg_pPS | 4254 | -0.01340424 | [-0.04 0.02] | 0.382767415 | 0.653782443 |
| Phe | Bilirubin_pPS | 4254 | 0.001354636 | [-0.03 0.03] | 0.929715864 | 0.966201806 |
| Phe | Proinsulin_pPS | 4254 | -0.004553636 | [-0.03 0.03] | 0.766851802 | 0.889703209 |
| Phe | Hyper_Insulin_pPS | 4254 | 0.012982321 | [-0.02 0.04] | 0.397924783 | 0.665794586 |
| Phe | Beta_Cell_1_pPS | 4254 | 0.005863565 | [-0.02 0.04] | 0.702618302 | 0.855830536 |
| Phe | Beta_Cell_2_pPS | 4254 | -0.009178561 | [-0.04 0.02] | 0.550078855 | 0.760607725 |
| Phe | Total_GRS | 4254 | 0.009158315 | [-0.02 0.04] | 0.550959074 | 0.760607725 |
| PUFA | Cholesterol_pPS | 4223 | -0.134194468 | [-0.16 -0.1 ] | 2.23E-18 | 1.45E-16 |
| PUFA | Lipodystrophy_1_pPS | 4223 | 0.030944025 | [0. 0.06] | 0.044652742 | 0.160975011 |
| PUFA | Lipodystrophy_2_pPS | 4223 | -0.054921794 | [-0.08 -0.02] | 0.000362983 | 0.00397244 |
| PUFA | Liver-Lipid_pPS | 4223 | -0.028934643 | [-0.06 0. ] | 0.06045387 | 0.201356445 |
| PUFA | ASAT_Pos_pPS | 4223 | -0.023067246 | [-0.05 0.01] | 0.134487097 | 0.37415671 |
| PUFA | ALP_Neg_pPS | 4223 | 0.057131461 | [0.03 0.09] | 0.00020782 | 0.002496149 |
| PUFA | SHBG-LpA_pPS | 4223 | 0.016327495 | [-0.01 0.05] | 0.289470717 | 0.576522459 |
| PUFA | Obesity_pPS | 4223 | 0.005374346 | [-0.02 0.04] | 0.727350402 | 0.870249668 |
| PUFA | VAT_Neg_pPS | 4223 | -0.001484223 | [-0.03 0.03] | 0.92329342 | 0.962324775 |
| PUFA | Bilirubin_pPS | 4223 | -0.022347472 | [-0.05 0.01] | 0.147078713 | 0.396835019 |
| PUFA | Proinsulin_pPS | 4223 | 0.008220187 | [-0.02 0.04] | 0.593841967 | 0.785805687 |
| PUFA | Hyper_Insulin_pPS | 4223 | -0.021536457 | [-0.05 0.01] | 0.162324692 | 0.421533251 |
| PUFA | Beta_Cell_1_pPS | 4223 | 0.032716988 | [0. 0.06] | 0.033752517 | 0.129284194 |
| PUFA | Beta_Cell_2_pPS | 4223 | 0.027816687 | [-0. 0.06] | 0.071090359 | 0.229306497 |
| PUFA | Total_GRS | 4223 | -0.002515852 | [-0.03 0.03] | 0.870352299 | 0.940757973 |
| Remnant_C | Cholesterol_pPS | 4254 | -0.141120786 | [-0.17 -0.11] | 2.59E-20 | 1.98E-18 |
| Remnant_C | Lipodystrophy_1_pPS | 4254 | 0.058640364 | [0.03 0.09] | 0.000132671 | 0.00171433 |
| Remnant_C | Lipodystrophy_2_pPS | 4254 | -0.036950833 | [-0.07 -0.01] | 0.016095451 | 0.070747425 |
| Remnant_C | Liver-Lipid_pPS | 4254 | -0.010414249 | [-0.04 0.02] | 0.497705032 | 0.724295386 |
| Remnant_C | ASAT_Pos_pPS | 4254 | 0.007074708 | [-0.02 0.04] | 0.645049587 | 0.817402268 |
| Remnant_C | ALP_Neg_pPS | 4254 | 0.047451255 | [0.02 0.08] | 0.001992412 | 0.013934058 |
| Remnant_C | SHBG-LpA_pPS | 4254 | 0.016476342 | [-0.01 0.05] | 0.283328896 | 0.571763651 |
| Remnant_C | Obesity_pPS | 4254 | 0.007307023 | [-0.02 0.04] | 0.634233137 | 0.807999529 |
| Remnant_C | VAT_Neg_pPS | 4254 | 0.012688259 | [-0.02 0.04] | 0.408699644 | 0.671770679 |
| Remnant_C | Bilirubin_pPS | 4254 | -0.021570819 | [-0.05 0.01] | 0.160119261 | 0.417242979 |
| Remnant_C | Proinsulin_pPS | 4254 | 0.006746485 | [-0.02 0.04] | 0.660459866 | 0.824723182 |
| Remnant_C | Hyper_Insulin_pPS | 4254 | -0.009321564 | [-0.04 0.02] | 0.54388187 | 0.758531414 |
| Remnant_C | Beta_Cell_1_pPS | 4254 | 0.020283612 | [-0.01 0.05] | 0.186557679 | 0.458380551 |
| Remnant_C | Beta_Cell_2_pPS | 4254 | 0.018657889 | [-0.01 0.05] | 0.224385979 | 0.505575551 |
| Remnant_C | Total_GRS | 4254 | 0.012504364 | [-0.02 0.04] | 0.415525221 | 0.677830746 |
| S_HDL_C | Cholesterol_pPS | 4255 | -0.067189248 | [-0.1 -0.04] | 1.19E-05 | 0.000206723 |
| S_HDL_C | Lipodystrophy_1_pPS | 4255 | -0.004575611 | [-0.03 0.03] | 0.765732677 | 0.889697961 |
| S_HDL_C | Lipodystrophy_2_pPS | 4255 | -0.029626704 | [-0.06 0. ] | 0.053645443 | 0.183817052 |
| S_HDL_C | Liver-Lipid_pPS | 4255 | -0.004468166 | [-0.03 0.03] | 0.771078496 | 0.891027884 |
| S_HDL_C | ASAT_Pos_pPS | 4255 | -0.012856924 | [-0.04 0.02] | 0.402442939 | 0.668142396 |
| S_HDL_C | ALP_Neg_pPS | 4255 | 0.017485338 | [-0.01 0.05] | 0.254821259 | 0.537454867 |
| S_HDL_C | SHBG-LpA_pPS | 4255 | -0.006426195 | [-0.04 0.02] | 0.67560171 | 0.834318347 |

**b) Partial Correlation**

| Metabolomics | PPS | Samp | r | CI95% | p | fdr_p_adjusted |
|---|---|---|---|---|---|---|
| Remnant_C | VAT_Neg_pPS | 4254 | 0.008950055 | [-0.02 0.04] | 0.560655267 | 0.797755947 |
| Remnant_C | Bilirubin_pPS | 4254 | -0.02404644 | [-0.05 0.01] | 0.117929848 | 0.361002379 |
| Remnant_C | Proinsulin_pPS | 4254 | -0.007752029 | [-0.04 0.02] | 0.614276204 | 0.828794354 |
| Remnant_C | Hyper_Insulin_pPS | 4254 | -0.014940261 | [-0.05 0.02] | 0.331377217 | 0.62795236 |
| Remnant_C | Beta_Cell_1_pPS | 4254 | 0.012688752 | [-0.02 0.04] | 0.40940082 | 0.682216107 |
| Remnant_C | Beta_Cell_2_pPS | 4254 | 0.016795463 | [-0.01 0.05] | 0.274843711 | 0.573178999 |
| S_HDL_C | Cholesterol_pPS | 4255 | -0.065277143 | [-0.1 -0.04] | 2.15E-05 | 0.000409776 |
| S_HDL_C | Lipodystrophy_1_pPS | 4255 | 0.000189897 | [-0.03 0.03] | 0.990148817 | 0.99412133 |
| S_HDL_C | Lipodystrophy_2_pPS | 4255 | -0.021388097 | [-0.05 0.01] | 0.164287106 | 0.441248115 |
| S_HDL_C | Liver-Lipid_pPS | 4255 | -0.002442005 | [-0.03 0.03] | 0.873844175 | 0.946048963 |
| S_HDL_C | ASAT_Pos_pPS | 4255 | -0.010669656 | [-0.04 0.02] | 0.487837403 | 0.747818133 |
| S_HDL_C | ALP_Neg_pPS | 4255 | 0.022318323 | [-0.01 0.05] | 0.146696085 | 0.420206242 |
| S_HDL_C | SHBG-LpA_pPS | 4255 | -0.003749051 | [-0.03 0.03] | 0.807415712 | 0.91016118 |
| S_HDL_C | Obesity_pPS | 4255 | -0.003707655 | [-0.03 0.03] | 0.809501124 | 0.91103332 |
| S_HDL_C | VAT_Neg_pPS | 4255 | -0.019804917 | [-0.05 0.01] | 0.197806284 | 0.483126879 |
| S_HDL_C | Bilirubin_pPS | 4255 | 0.002854706 | [-0.03 0.03] | 0.852749708 | 0.935454749 |
| S_HDL_C | Proinsulin_pPS | 4255 | -0.025203013 | [-0.06 0. ] | 0.101225593 | 0.328449979 |
| S_HDL_C | Hyper_Insulin_pPS | 4255 | -0.014671877 | [-0.04 0.02] | 0.340080884 | 0.634521837 |
| S_HDL_C | Beta_Cell_1_pPS | 4255 | 0.035069773 | [0. 0.07] | 0.022553704 | 0.099894945 |
| S_HDL_C | Beta_Cell_2_pPS | 4255 | -0.000602052 | [-0.03 0.03] | 0.968774814 | 0.988021996 |
| S_HDL_CE | Cholesterol_pPS | 4255 | -0.082512492 | [-0.11 -0.05] | 7.70E-08 | 1.83E-06 |
| S_HDL_CE | Lipodystrophy_1_pPS | 4255 | 0.003604713 | [-0.03 0.03] | 0.814692786 | 0.912503804 |
| S_HDL_CE | Lipodystrophy_2_pPS | 4255 | -0.024849186 | [-0.05 0.01] | 0.106110658 | 0.338269962 |
| S_HDL_CE | Liver-Lipid_pPS | 4255 | 0.014394513 | [-0.02 0.04] | 0.349289358 | 0.637861136 |
| S_HDL_CE | ASAT_Pos_pPS | 4255 | -0.010129698 | [-0.04 0.02] | 0.510125646 | 0.766145193 |
| S_HDL_CE | ALP_Neg_pPS | 4255 | 0.024184807 | [-0.01 0.05] | 0.115786561 | 0.355528672 |
| S_HDL_CE | SHBG-LpA_pPS | 4255 | -0.001173536 | [-0.03 0.03] | 0.93917851 | 0.978430443 |
| S_HDL_CE | Obesity_pPS | 4255 | -0.008893257 | [-0.04 0.02] | 0.563099296 | 0.797755947 |
| S_HDL_CE | VAT_Neg_pPS | 4255 | -0.015336641 | [-0.05 0.01] | 0.318650351 | 0.61940234 |
| S_HDL_CE | Bilirubin_pPS | 4255 | -0.001027691 | [-0.03 0.03] | 0.94672522 | 0.978747744 |
| S_HDL_CE | Proinsulin_pPS | 4255 | -0.02849019 | [-0.06 0. ] | 0.063913008 | 0.227271478 |
| S_HDL_CE | Hyper_Insulin_pPS | 4255 | -0.014986218 | [-0.05 0.02] | 0.329834614 | 0.627689066 |
| S_HDL_CE | Beta_Cell_1_pPS | 4255 | 0.032864398 | [0. 0.06] | 0.03256587 | 0.130133476 |
| S_HDL_CE | Beta_Cell_2_pPS | 4255 | 0.001781434 | [-0.03 0.03] | 0.907789208 | 0.96224905 |
| S_HDL_FC | Cholesterol_pPS | 4255 | 0.045909873 | [0.02 0.08] | 0.002820992 | 0.01988601 |
| S_HDL_FC | Lipodystrophy_1_pPS | 4255 | -0.015895492 | [-0.05 0.01] | 0.301333519 | 0.59848185 |
| S_HDL_FC | Lipodystrophy_2_pPS | 4255 | 0.010502356 | [-0.02 0.04] | 0.494686539 | 0.753425828 |
| S_HDL_FC | Liver-Lipid_pPS | 4255 | -0.072709439 | [-0.1 -0.04] | 2.20E-06 | 4.69E-05 |
| S_HDL_FC | ASAT_Pos_pPS | 4255 | -0.0072347 | [-0.04 0.02] | 0.638068735 | 0.835456904 |
| S_HDL_FC | ALP_Neg_pPS | 4255 | -0.002788994 | [-0.03 0.03] | 0.85610183 | 0.93707811 |
| S_HDL_FC | SHBG-LpA_pPS | 4255 | -0.005287691 | [-0.04 0.02] | 0.730993685 | 0.869031685 |
| S_HDL_FC | Obesity_pPS | 4255 | 0.007695164 | [-0.02 0.04] | 0.616834959 | 0.828794354 |
| S_HDL_FC | VAT_Neg_pPS | 4255 | -0.039039892 | [-0.07 -0.01] | 0.011107083 | 0.058516788 |
| S_HDL_FC | Bilirubin_pPS | 4255 | 0.015014751 | [-0.02 0.05] | 0.328914568 | 0.627130443 |
| S_HDL_FC | Proinsulin_pPS | 4255 | 0.007996676 | [-0.02 0.04] | 0.603101195 | 0.823046075 |
| S_HDL_FC | Hyper_Insulin_pPS | 4255 | 0.004216617 | [-0.03 0.03] | 0.783959838 | 0.894804787 |
| S_HDL_FC | Beta_Cell_1_pPS | 4255 | 0.025490632 | [-0. 0.06] | 0.097388017 | 0.319101162 |
| S_HDL_FC | Beta_Cell_2_pPS | 4255 | -0.017015641 | [-0.05 0.01] | 0.26854308 | 0.5683121 |
| S_HDL_L | Cholesterol_pPS | 4255 | -0.011275995 | [-0.04 0.02] | 0.463448771 | 0.726829028 |
| S_HDL_L | Lipodystrophy_1_pPS | 4255 | 0.005713847 | [-0.02 0.04] | 0.710255521 | 0.864160597 |
| S_HDL_L | Lipodystrophy_2_pPS | 4255 | -0.005749127 | [-0.04 0.02] | 0.708548013 | 0.863367695 |
| S_HDL_L | Liver-Lipid_pPS | 4255 | -0.056686182 | [-0.09 -0.03] | 0.00022544 | 0.002856526 |
| S_HDL_L | ASAT_Pos_pPS | 4255 | 0.003852611 | [-0.03 0.03] | 0.802204809 | 0.906328459 |
| S_HDL_L | ALP_Neg_pPS | 4255 | 0.008595817 | [-0.02 0.04] | 0.576227443 | 0.803773963 |
| S_HDL_L | SHBG-LpA_pPS | 4255 | -0.006931825 | [-0.04 0.02] | 0.652200498 | 0.844628798 |
| S_HDL_L | Obesity_pPS | 4255 | 0.013755391 | [-0.02 0.04] | 0.371104052 | 0.653802291 |
| S_HDL_L | VAT_Neg_pPS | 4255 | -0.029403588 | [-0.06 0. ] | 0.055849009 | 0.20440533 |
| S_HDL_L | Bilirubin_pPS | 4255 | 0.01628375 | [-0.01 0.05] | 0.28967854 | 0.5869802 |
| S_HDL_L | Proinsulin_pPS | 4255 | -0.000837674 | [-0.03 0.03] | 0.956568377 | 0.981227298 |
| S_HDL_L | Hyper_Insulin_pPS | 4255 | 0.000686479 | [-0.03 0.03] | 0.964398796 | 0.985567325 |
| S_HDL_L | Beta_Cell_1_pPS | 4255 | 0.029153661 | [-0. 0.06] | 0.057966455 | 0.211382229 |
| S_HDL_L | Beta_Cell_2_pPS | 4255 | -0.018088898 | [-0.05 0.01] | 0.239506016 | 0.535744184 |
| S_HDL_P | Cholesterol_pPS | 4255 | -0.012125368 | [-0.04 0.02] | 0.430456315 | 0.696591166 |
| S_HDL_P | Lipodystrophy_1_pPS | 4255 | 0.009266126 | [-0.02 0.04] | 0.546848502 | 0.791033743 |
| S_HDL_P | Lipodystrophy_2_pPS | 4255 | -0.004432524 | [-0.03 0.03] | 0.773193125 | 0.89042977 |
| S_HDL_P | Liver-Lipid_pPS | 4255 | -0.058796168 | [-0.09 -0.03] | 0.000130141 | 0.001809321 |
| S_HDL_P | ASAT_Pos_pPS | 4255 | 0.005662179 | [-0.02 0.04] | 0.712758824 | 0.86476088 |
| S_HDL_P | ALP_Neg_pPS | 4255 | 0.008802571 | [-0.02 0.04] | 0.567086609 | 0.797755947 |
| S_HDL_P | SHBG-LpA_pPS | 4255 | -0.007259388 | [-0.04 0.02] | 0.636922482 | 0.835456904 |
| S_HDL_P | Obesity_pPS | 4255 | 0.01556508 | [-0.01 0.05] | 0.31149463 | 0.613188052 |
| S_HDL_P | VAT_Neg_pPS | 4255 | -0.02859986 | [-0.06 0. ] | 0.062896669 | 0.224855592 |
| S_HDL_P | Bilirubin_pPS | 4255 | 0.016644366 | [-0.01 0.05] | 0.279128672 | 0.576693087 |
| S_HDL_P | Proinsulin_pPS | 4255 | 0.001253501 | [-0.03 0.03] | 0.935042969 | 0.978170683 |
| S_HDL_P | Hyper_Insulin_pPS | 4255 | 0.001401599 | [-0.03 0.03] | 0.927388552 | 0.974609911 |
| S_HDL_P | Beta_Cell_1_pPS | 4255 | 0.027907127 | [-0. 0.06] | 0.069546458 | 0.241818772 |
| S_HDL_P | Beta_Cell_2_pPS | 4255 | -0.018883815 | [-0.05 0.01] | 0.219476787 | 0.513309027 |
| S_HDL_PL | Cholesterol_pPS | 4255 | 0.040475722 | [0.01 0.07] | 0.008468942 | 0.047625902 |
| S_HDL_PL | Lipodystrophy_1_pPS | 4255 | -0.007213081 | [-0.04 0.02] | 0.639073165 | 0.836160747 |
| S_HDL_PL | Lipodystrophy_2_pPS | 4255 | 0.01192972 | [-0.02 0.04] | 0.437932772 | 0.702280961 |
| S_HDL_PL | Liver-Lipid_pPS | 4255 | -0.066887065 | [-0.1 -0.04] | 1.34E-05 | 0.000268636 |
| S_HDL_PL | ASAT_Pos_pPS | 4255 | 0.005817094 | [-0.02 0.04] | 0.705262577 | 0.861987595 |
| S_HDL_PL | ALP_Neg_pPS | 4255 | -0.007548863 | [-0.04 0.02] | 0.623547954 | 0.833339789 |
| S_HDL_PL | SHBG-LpA_pPS | 4255 | -0.007521477 | [-0.04 0.02] | 0.624808046 | 0.833566935 |
| S_HDL_PL | Obesity_pPS | 4255 | 0.011013868 | [-0.02 0.04] | 0.473906823 | 0.735233474 |
| S_HDL_PL | VAT_Neg_pPS | 4255 | -0.034946339 | [-0.07 -0. ] | 0.023033522 | 0.101378139 |
| S_HDL_PL | Bilirubin_pPS | 4255 | 0.021219532 | [-0.01 0.05] | 0.167638081 | 0.442759153 |
| S_HDL_PL | Proinsulin_pPS | 4255 | 0.011187388 | [-0.02 0.04] | 0.466969869 | 0.73037006 |
| S_HDL_PL | Hyper_Insulin_pPS | 4255 | 0.008028549 | [-0.02 0.04] | 0.601657473 | 0.822414367 |
| S_HDL_PL | Beta_Cell_1_pPS | 4255 | 0.013591286 | [-0.02 0.04] | 0.376838898 | 0.657170274 |
| S_HDL_PL | Beta_Cell_2_pPS | 4255 | -0.020076355 | [-0.05 0.01] | 0.191729219 | 0.474464643 |
| S_HDL_TG | Cholesterol_pPS | 4255 | -0.061885012 | [-0.09 -0.03] | 5.64E-05 | 0.000924773 |
| S_HDL_TG | Lipodystrophy_1_pPS | 4255 | 0.062492498 | [0.03 0.09] | 4.76E-05 | 0.000814207 |
| S_HDL_TG | Lipodystrophy_2_pPS | 4255 | 0.015490095 | [-0.01 0.05] | 0.313831731 | 0.61536839 |
| S_HDL_TG | Liver-Lipid_pPS | 4255 | -0.035919536 | [-0.07 -0.01] | 0.019480072 | 0.088360646 |

**Table S9: Associations between pPSs and global metabolomic profile in SCAPIS**

**a) Spearman Correlation**

| Metabolomics | PPS | Samp | r | CI95% | p | fdr_p_adjusted |
|---|---|---|---|---|---|---|
| S_HDL_C | Obesity_pPS | 4255 | -0.013329494 | [-0.04 0.02] | 0.385370524 | 0.654929006 |
| S_HDL_C | VAT_Neg_pPS | 4255 | -0.018966573 | [-0.05 0.01] | 0.21675694 | 0.497544682 |
| S_HDL_C | Bilirubin_pPS | 4255 | 0.003434797 | [-0.03 0.03] | 0.823012836 | 0.920418422 |
| S_HDL_C | Proinsulin_pPS | 4255 | -0.01511885 | [-0.05 0.01] | 0.324829176 | 0.608687153 |
| S_HDL_C | Hyper_Insulin_pPS | 4255 | -0.020036324 | [-0.05 0.01] | 0.191934124 | 0.464503229 |
| S_HDL_C | Beta_Cell_1_pPS | 4255 | 0.02819564 | [-0. 0.06] | 0.066292548 | 0.215777718 |
| S_HDL_C | Beta_Cell_2_pPS | 4255 | 0.001944755 | [-0.03 0.03] | 0.899224677 | 0.95063427 |
| S_HDL_C | Total_GRS | 4255 | -0.022022733 | [-0.05 0.01] | 0.15149381 | 0.401178951 |
| S_HDL_CE | Cholesterol_pPS | 4255 | -0.084862045 | [-0.11 -0.05] | 3.09E-08 | 8.61E-07 |
| S_HDL_CE | Lipodystrophy_1_pPS | 4255 | -0.001146114 | [-0.03 0.03] | 0.940505953 | 0.971765544 |
| S_HDL_CE | Lipodystrophy_2_pPS | 4255 | -0.032317107 | [-0.06 -0. ] | 0.035290314 | 0.133978272 |
| S_HDL_CE | Liver-Lipid_pPS | 4255 | 0.011889586 | [-0.02 0.04] | 0.438773056 | 0.691142319 |
| S_HDL_CE | ASAT_Pos_pPS | 4255 | -0.012082625 | [-0.04 0.02] | 0.431376332 | 0.688963257 |
| S_HDL_CE | ALP_Neg_pPS | 4255 | 0.019319459 | [-0.01 0.05] | 0.208325356 | 0.487304132 |
| S_HDL_CE | SHBG-LpA_pPS | 4255 | -0.003143058 | [-0.03 0.03] | 0.837827071 | 0.926178923 |
| S_HDL_CE | Obesity_pPS | 4255 | -0.02022352 | [-0.05 0.01] | 0.187814304 | 0.459363377 |
| S_HDL_CE | VAT_Neg_pPS | 4255 | -0.014003235 | [-0.04 0.02] | 0.361809198 | 0.639224774 |
| S_HDL_CE | Bilirubin_pPS | 4255 | -0.000758637 | [-0.03 0.03] | 0.96059913 | 0.981183397 |
| S_HDL_CE | Proinsulin_pPS | 4255 | -0.018889583 | [-0.05 0.01] | 0.218628645 | 0.499423263 |
| S_HDL_CE | Hyper_Insulin_pPS | 4255 | -0.019492192 | [-0.05 0.01] | 0.204285947 | 0.482591802 |
| S_HDL_CE | Beta_Cell_1_pPS | 4255 | 0.026647448 | [-0. 0.06] | 0.082640498 | 0.258026008 |
| S_HDL_CE | Beta_Cell_2_pPS | 4255 | 0.003718789 | [-0.03 0.03] | 0.808652209 | 0.911007872 |
| S_HDL_CE | Total_GRS | 4255 | -0.024933817 | [-0.05 0.01] | 0.104392728 | 0.312304604 |
| S_HDL_FC | Cholesterol_pPS | 4255 | 0.046354986 | [0.02 0.08] | 0.002525916 | 0.016671044 |
| S_HDL_FC | Lipodystrophy_1_pPS | 4255 | -0.017165937 | [-0.05 0.01] | 0.263604117 | 0.550503091 |
| S_HDL_FC | Lipodystrophy_2_pPS | 4255 | 0.003015031 | [-0.03 0.03] | 0.844346719 | 0.92810697 |
| S_HDL_FC | Liver-Lipid_pPS | 4255 | -0.071685174 | [-0.1 -0.04] | 2.95E-06 | 6.47E-05 |
| S_HDL_FC | ASAT_Pos_pPS | 4255 | -0.008647736 | [-0.04 0.02] | 0.573337187 | 0.772791937 |
| S_HDL_FC | ALP_Neg_pPS | 4255 | -0.004425927 | [-0.03 0.03] | 0.773183023 | 0.891654615 |
| S_HDL_FC | SHBG-LpA_pPS | 4255 | -0.009690217 | [-0.04 0.02] | 0.528018815 | 0.743176087 |
| S_HDL_FC | Obesity_pPS | 4255 | 0.011639231 | [-0.02 0.04] | 0.448473617 | 0.694066312 |
| S_HDL_FC | VAT_Neg_pPS | 4255 | -0.039268759 | [-0.07 -0.01] | 0.010523561 | 0.049286108 |
| S_HDL_FC | Bilirubin_pPS | 4255 | 0.016190596 | [-0.01 0.05] | 0.291706001 | 0.577755652 |
| S_HDL_FC | Proinsulin_pPS | 4255 | 0.011537959 | [-0.02 0.04] | 0.452431975 | 0.695176638 |
| S_HDL_FC | Hyper_Insulin_pPS | 4255 | -0.003224641 | [-0.03 0.03] | 0.833678319 | 0.924155036 |
| S_HDL_FC | Beta_Cell_1_pPS | 4255 | 0.0175328 | [-0.01 0.05] | 0.253533717 | 0.536320571 |
| S_HDL_FC | Beta_Cell_2_pPS | 4255 | -0.013362145 | [-0.04 0.02] | 0.384207515 | 0.654405057 |
| S_HDL_FC | Total_GRS | 4255 | -0.000319864 | [-0.03 0.03] | 0.983381837 | 0.990093216 |
| S_HDL_L | Cholesterol_pPS | 4255 | -0.011136833 | [-0.04 0.02] | 0.468302796 | 0.707899576 |
| S_HDL_L | Lipodystrophy_1_pPS | 4255 | 0.006233552 | [-0.02 0.04] | 0.684795978 | 0.841287155 |
| S_HDL_L | Lipodystrophy_2_pPS | 4255 | -0.011893133 | [-0.04 0.02] | 0.4386365 | 0.691142319 |
| S_HDL_L | Liver-Lipid_pPS | 4255 | -0.05690682 | [-0.09 -0.03] | 0.000208303 | 0.002496149 |
| S_HDL_L | ASAT_Pos_pPS | 4255 | 0.005735783 | [-0.02 0.04] | 0.708767264 | 0.859415365 |
| S_HDL_L | ALP_Neg_pPS | 4255 | 0.006540843 | [-0.02 0.04] | 0.670152641 | 0.829952319 |
| S_HDL_L | SHBG-LpA_pPS | 4255 | -0.009312336 | [-0.04 0.02] | 0.544233029 | 0.758531414 |
| S_HDL_L | Obesity_pPS | 4255 | 0.012949041 | [-0.02 0.04] | 0.399079968 | 0.666036824 |
| S_HDL_L | VAT_Neg_pPS | 4255 | -0.028987967 | [-0.06 0. ] | 0.059016867 | 0.197182522 |
| S_HDL_L | Bilirubin_pPS | 4255 | 0.018128514 | [-0.01 0.05] | 0.237757729 | 0.519979223 |
| S_HDL_L | Proinsulin_pPS | 4255 | 0.005469283 | [-0.02 0.04] | 0.72172256 | 0.866794452 |
| S_HDL_L | Hyper_Insulin_pPS | 4255 | -0.00328592 | [-0.03 0.03] | 0.830565167 | 0.922611229 |
| S_HDL_L | Beta_Cell_1_pPS | 4255 | 0.022458691 | [-0.01 0.05] | 0.143556414 | 0.391461081 |
| S_HDL_L | Beta_Cell_2_pPS | 4255 | -0.01443919 | [-0.04 0.02] | 0.347055292 | 0.62662761 |
| S_HDL_L | Total_GRS | 4255 | -0.000583874 | [-0.03 0.03] | 0.969670664 | 0.983305352 |
| S_HDL_P | Cholesterol_pPS | 4255 | -0.011776153 | [-0.04 0.02] | 0.443153283 | 0.692953671 |
| S_HDL_P | Lipodystrophy_1_pPS | 4255 | 0.010390995 | [-0.02 0.04] | 0.498614892 | 0.725104368 |
| S_HDL_P | Lipodystrophy_2_pPS | 4255 | -0.010084878 | [-0.04 0.02] | 0.511351282 | 0.732208611 |
| S_HDL_P | Liver-Lipid_pPS | 4255 | -0.058880081 | [-0.09 -0.03] | 0.000124269 | 0.001615492 |
| S_HDL_P | ASAT_Pos_pPS | 4255 | 0.008450719 | [-0.02 0.04] | 0.582104559 | 0.778303354 |
| S_HDL_P | ALP_Neg_pPS | 4255 | 0.007165988 | [-0.02 0.04] | 0.640751159 | 0.813261086 |
| S_HDL_P | SHBG-LpA_pPS | 4255 | -0.009233858 | [-0.04 0.02] | 0.547631163 | 0.760607725 |
| S_HDL_P | Obesity_pPS | 4255 | 0.01517742 | [-0.01 0.05] | 0.322958152 | 0.608687153 |
| S_HDL_P | VAT_Neg_pPS | 4255 | -0.027964258 | [-0.06 0. ] | 0.068551531 | 0.222792474 |
| S_HDL_P | Bilirubin_pPS | 4255 | 0.018643731 | [-0.01 0.05] | 0.224683234 | 0.505714099 |
| S_HDL_P | Proinsulin_pPS | 4255 | 0.007428366 | [-0.02 0.04] | 0.628573723 | 0.803929814 |
| S_HDL_P | Hyper_Insulin_pPS | 4255 | -0.001814382 | [-0.03 0.03] | 0.905947955 | 0.952872131 |
| S_HDL_P | Beta_Cell_1_pPS | 4255 | 0.021788901 | [-0.01 0.05] | 0.155886619 | 0.408773591 |
| S_HDL_P | Beta_Cell_2_pPS | 4255 | -0.015158089 | [-0.05 0.01] | 0.323574904 | 0.608687153 |
| S_HDL_P | Total_GRS | 4255 | 0.00292072 | [-0.03 0.03] | 0.849156247 | 0.929780576 |
| S_HDL_PL | Cholesterol_pPS | 4255 | 0.041537172 | [0.01 0.07] | 0.006809248 | 0.034942196 |
| S_HDL_PL | Lipodystrophy_1_pPS | 4255 | -0.0056183 | [-0.04 0.02] | 0.71446829 | 0.86291356 |
| S_HDL_PL | Lipodystrophy_2_pPS | 4255 | 0.007197321 | [-0.02 0.04] | 0.639291725 | 0.812370112 |
| S_HDL_PL | Liver-Lipid_pPS | 4255 | -0.066163758 | [-0.1 -0.04] | 1.61E-05 | 0.000271706 |
| S_HDL_PL | ASAT_Pos_pPS | 4255 | 0.006681312 | [-0.02 0.04] | 0.663499971 | 0.825654931 |
| S_HDL_PL | ALP_Neg_pPS | 4255 | -0.008097268 | [-0.04 0.02] | 0.597988313 | 0.787890007 |
| S_HDL_PL | SHBG-LpA_pPS | 4255 | -0.010735601 | [-0.04 0.02] | 0.484481362 | 0.717164725 |
| S_HDL_PL | Obesity_pPS | 4255 | 0.015748488 | [-0.01 0.05] | 0.30508428 | 0.591152466 |
| S_HDL_PL | VAT_Neg_pPS | 4255 | -0.035622237 | [-0.07 -0.01] | 0.020317887 | 0.086471961 |
| S_HDL_PL | Bilirubin_pPS | 4255 | 0.022414746 | [-0.01 0.05] | 0.144341763 | 0.392412017 |
| S_HDL_PL | Proinsulin_pPS | 4255 | 0.010696401 | [-0.02 0.04] | 0.486078081 | 0.717575695 |
| S_HDL_PL | Hyper_Insulin_pPS | 4255 | 0.003289665 | [-0.03 0.03] | 0.83037498 | 0.922611229 |
| S_HDL_PL | Beta_Cell_1_pPS | 4255 | 0.006670984 | [-0.02 0.04] | 0.663988231 | 0.825654931 |
| S_HDL_PL | Beta_Cell_2_pPS | 4255 | -0.018742372 | [-0.05 0.01] | 0.222239782 | 0.502934916 |
| S_HDL_PL | Total_GRS | 4255 | 0.006091347 | [-0.02 0.04] | 0.691613128 | 0.846635599 |
| S_HDL_TG | Cholesterol_pPS | 4255 | -0.059204123 | [-0.09 -0.03] | 0.000113989 | 0.001547513 |
| S_HDL_TG | Lipodystrophy_1_pPS | 4255 | 0.071325722 | [0.04 0.1 ] | 3.31E-06 | 6.97E-05 |
| S_HDL_TG | Lipodystrophy_2_pPS | 4255 | 0.020058231 | [-0.01 0.05] | 0.191448579 | 0.464503229 |
| S_HDL_TG | Liver-Lipid_pPS | 4255 | -0.034686085 | [-0.06 -0. ] | 0.023858067 | 0.098604148 |
| S_HDL_TG | ASAT_Pos_pPS | 4255 | 0.047069457 | [0.02 0.08] | 0.00216348 | 0.014732269 |
| S_HDL_TG | ALP_Neg_pPS | 4255 | 0.00882635 | [-0.02 0.04] | 0.565443225 | 0.768674587 |
| S_HDL_TG | SHBG-LpA_pPS | 4255 | -0.005999571 | [-0.04 0.02] | 0.696026145 | 0.850214169 |
| S_HDL_TG | Obesity_pPS | 4255 | 0.030355229 | [0. 0.06] | 0.048023388 | 0.169170784 |
| S_HDL_TG | VAT_Neg_pPS | 4255 | 0.012186726 | [-0.02 0.04] | 0.427417637 | 0.687522331 |
| S_HDL_TG | Bilirubin_pPS | 4255 | 0.002520518 | [-0.03 0.03] | 0.869625495 | 0.940757973 |
| S_HDL_TG | Proinsulin_pPS | 4255 | 0.020473728 | [-0.01 0.05] | 0.182410104 | 0.452703501 |

**b) Partial Correlation**

| Metabolomics | PPS | Samp | r | CI95% | p | fdr_p_adjusted |
|---|---|---|---|---|---|---|
| S_HDL_TG | ASAT_Pos_pPS | 4255 | 0.031814381 | [0. 0.06] | 0.0385395 | 0.149817629 |
| S_HDL_TG | ALP_Neg_pPS | 4255 | 0.004972358 | [-0.03 0.04] | 0.746467375 | 0.874926432 |
| S_HDL_TG | SHBG-LpA_pPS | 4255 | -0.012425892 | [-0.04 0.02] | 0.419117569 | 0.691164228 |
| S_HDL_TG | Obesity_pPS | 4255 | 0.029860163 | [-0. 0.06] | 0.052147063 | 0.193330408 |
| S_HDL_TG | VAT_Neg_pPS | 4255 | 0.006352777 | [-0.02 0.04] | 0.679564967 | 0.857328072 |
| S_HDL_TG | Bilirubin_pPS | 4255 | -0.000978583 | [-0.03 0.03] | 0.949267435 | 0.978747744 |
| S_HDL_TG | Proinsulin_pPS | 4255 | 0.017231977 | [-0.01 0.05] | 0.26250371 | 0.559637055 |
| S_HDL_TG | Hyper_Insulin_pPS | 4255 | 0.005950777 | [-0.02 0.04] | 0.69881664 | 0.859092773 |
| S_HDL_TG | Beta_Cell_1_pPS | 4255 | 0.002676662 | [-0.03 0.03] | 0.861838125 | 0.940272439 |
| S_HDL_TG | Beta_Cell_2_pPS | 4255 | -0.009454691 | [-0.04 0.02] | 0.538719547 | 0.784375661 |
| S_LDL_C | Cholesterol_pPS | 4255 | -0.151021273 | [-0.18 -0.12] | 5.29E-23 | 6.62E-21 |
| S_LDL_C | Lipodystrophy_1_pPS | 4255 | 0.038929357 | [0.01 0.07] | 0.01133763 | 0.059263537 |
| S_LDL_C | Lipodystrophy_2_pPS | 4255 | -0.037874404 | [-0.07 -0.01] | 0.013760773 | 0.069393116 |
| S_LDL_C | Liver-Lipid_pPS | 4255 | 0.014362622 | [-0.02 0.04] | 0.350358179 | 0.637861136 |
| S_LDL_C | ASAT_Pos_pPS | 4255 | -0.019723374 | [-0.05 0.01] | 0.199659141 | 0.486274453 |
| S_LDL_C | ALP_Neg_pPS | 4255 | 0.048745662 | [0.02 0.08] | 0.001517633 | 0.012153205 |
| S_LDL_C | SHBG-LpA_pPS | 4255 | 0.013437016 | [-0.02 0.04] | 0.382279546 | 0.661657399 |
| S_LDL_C | Obesity_pPS | 4255 | 0.006812456 | [-0.02 0.04] | 0.657804977 | 0.844628798 |
| S_LDL_C | VAT_Neg_pPS | 4255 | 0.007065646 | [-0.02 0.04] | 0.645940777 | 0.841909788 |
| S_LDL_C | Bilirubin_pPS | 4255 | -0.014605956 | [-0.04 0.02] | 0.342255221 | 0.636982428 |
| S_LDL_C | Proinsulin_pPS | 4255 | -0.023011672 | [-0.05 0.01] | 0.134549643 | 0.397885355 |
| S_LDL_C | Hyper_Insulin_pPS | 4255 | -0.02206201 | [-0.05 0.01] | 0.151392711 | 0.427467143 |
| S_LDL_C | Beta_Cell_1_pPS | 4255 | 0.018610449 | [-0.01 0.05] | 0.226224481 | 0.521665942 |
| S_LDL_C | Beta_Cell_2_pPS | 4255 | 0.022506405 | [-0.01 0.05] | 0.143321198 | 0.415237394 |
| S_LDL_CE | Cholesterol_pPS | 4255 | -0.151937341 | [-0.18 -0.12] | 2.89E-23 | 5.13E-21 |
| S_LDL_CE | Lipodystrophy_1_pPS | 4255 | 0.039806189 | [0.01 0.07] | 0.009620093 | 0.052335396 |
| S_LDL_CE | Lipodystrophy_2_pPS | 4255 | -0.036544689 | [-0.07 -0.01] | 0.017458755 | 0.08243497 |
| S_LDL_CE | Liver-Lipid_pPS | 4255 | 0.016393522 | [-0.01 0.05] | 0.286439103 | 0.583962408 |
| S_LDL_CE | ASAT_Pos_pPS | 4255 | -0.018396916 | [-0.05 0.01] | 0.231597262 | 0.527097702 |
| S_LDL_CE | ALP_Neg_pPS | 4255 | 0.048385657 | [0.02 0.08] | 0.001644796 | 0.013066989 |
| S_LDL_CE | SHBG-LpA_pPS | 4255 | 0.013478223 | [-0.02 0.04] | 0.38082159 | 0.661235752 |
| S_LDL_CE | Obesity_pPS | 4255 | 0.006948703 | [-0.02 0.04] | 0.651409638 | 0.844628798 |
| S_LDL_CE | VAT_Neg_pPS | 4255 | 0.008138311 | [-0.02 0.04] | 0.596697606 | 0.818772177 |
| S_LDL_CE | Bilirubin_pPS | 4255 | -0.0144041 | [-0.04 0.02] | 0.348968445 | 0.637861136 |
| S_LDL_CE | Proinsulin_pPS | 4255 | -0.022716205 | [-0.05 0.01] | 0.139627197 | 0.408674924 |
| S_LDL_CE | Hyper_Insulin_pPS | 4255 | -0.02151613 | [-0.05 0.01] | 0.161775771 | 0.436489344 |
| S_LDL_CE | Beta_Cell_1_pPS | 4255 | 0.017458019 | [-0.01 0.05] | 0.25629427 | 0.55296819 |
| S_LDL_CE | Beta_Cell_2_pPS | 4255 | 0.02207677 | [-0.01 0.05] | 0.15111919 | 0.427467143 |
| S_LDL_FC | Cholesterol_pPS | 4255 | -0.142858386 | [-0.17 -0.11] | 9.96E-21 | 7.67E-19 |
| S_LDL_FC | Lipodystrophy_1_pPS | 4255 | 0.034865848 | [0. 0.06] | 0.02335116 | 0.102519784 |
| S_LDL_FC | Lipodystrophy_2_pPS | 4255 | -0.041837592 | [-0.07 -0.01] | 0.006499829 | 0.038160289 |
| S_LDL_FC | Liver-Lipid_pPS | 4255 | 0.005529137 | [-0.02 0.04] | 0.719218853 | 0.866319132 |
| S_LDL_FC | ASAT_Pos_pPS | 4255 | -0.023427605 | [-0.05 0.01] | 0.127645201 | 0.380843059 |
| S_LDL_FC | ALP_Neg_pPS | 4255 | 0.048088702 | [0.02 0.08] | 0.001757009 | 0.013740358 |
| S_LDL_FC | SHBG-LpA_pPS | 4255 | 0.013329526 | [-0.02 0.04] | 0.386098727 | 0.664634265 |
| S_LDL_FC | Obesity_pPS | 4255 | 0.006864352 | [-0.02 0.04] | 0.655366023 | 0.844628798 |
| S_LDL_FC | VAT_Neg_pPS | 4255 | 0.002593255 | [-0.03 0.03] | 0.866102044 | 0.943130556 |
| S_LDL_FC | Bilirubin_pPS | 4255 | -0.016057814 | [-0.05 0.01] | 0.29642339 | 0.594033661 |
| S_LDL_FC | Proinsulin_pPS | 4255 | -0.022897138 | [-0.05 0.01] | 0.136500683 | 0.40187407 |
| S_LDL_FC | Hyper_Insulin_pPS | 4255 | -0.023793268 | [-0.05 0.01] | 0.12180564 | 0.368918143 |
| S_LDL_FC | Beta_Cell_1_pPS | 4255 | 0.02169523 | [-0.01 0.05] | 0.158311515 | 0.431435756 |
| S_LDL_FC | Beta_Cell_2_pPS | 4255 | 0.021932776 | [-0.01 0.05] | 0.153803771 | 0.428688101 |
| S_LDL_L | Cholesterol_pPS | 4255 | -0.153105687 | [-0.18 -0.12] | 1.32E-23 | 4.03E-21 |
| S_LDL_L | Lipodystrophy_1_pPS | 4255 | 0.04353402 | [0.01 0.07] | 0.004627363 | 0.029166809 |
| S_LDL_L | Lipodystrophy_2_pPS | 4255 | -0.039552668 | [-0.07 -0.01] | 0.010091137 | 0.054454059 |
| S_LDL_L | Liver-Lipid_pPS | 4255 | 0.00538249 | [-0.02 0.04] | 0.726362804 | 0.866797473 |
| S_LDL_L | ASAT_Pos_pPS | 4255 | -0.021629765 | [-0.05 0.01] | 0.159571216 | 0.433462108 |
| S_LDL_L | ALP_Neg_pPS | 4255 | 0.05045923 | [0.02 0.08] | 0.001027409 | 0.008904213 |
| S_LDL_L | SHBG-LpA_pPS | 4255 | 0.013721363 | [-0.02 0.04] | 0.372288734 | 0.654940286 |
| S_LDL_L | Obesity_pPS | 4255 | 0.010838528 | [-0.02 0.04] | 0.480976136 | 0.740874464 |
| S_LDL_L | VAT_Neg_pPS | 4255 | 0.004965754 | [-0.03 0.04] | 0.746792579 | 0.874926432 |
| S_LDL_L | Bilirubin_pPS | 4255 | -0.014504576 | [-0.04 0.02] | 0.345616489 | 0.637861136 |
| S_LDL_L | Proinsulin_pPS | 4255 | -0.020512717 | [-0.05 0.01] | 0.182249304 | 0.462437397 |
| S_LDL_L | Hyper_Insulin_pPS | 4255 | -0.022540451 | [-0.05 0.01] | 0.142716695 | 0.414085252 |
| S_LDL_L | Beta_Cell_1_pPS | 4255 | 0.020328925 | [-0.01 0.05] | 0.186198929 | 0.468642761 |
| S_LDL_L | Beta_Cell_2_pPS | 4255 | 0.021366125 | [-0.01 0.05] | 0.164721019 | 0.441248115 |
| S_LDL_P | Cholesterol_pPS | 4255 | -0.153859057 | [-0.18 -0.12] | 7.99E-24 | 3.20E-21 |
| S_LDL_P | Lipodystrophy_1_pPS | 4255 | 0.045185757 | [0.02 0.08] | 0.003288013 | 0.022466219 |
| S_LDL_P | Lipodystrophy_2_pPS | 4255 | -0.039200095 | [-0.07 -0.01] | 0.010780331 | 0.057247273 |
| S_LDL_P | Liver-Lipid_pPS | 4255 | 0.00354938 | [-0.03 0.03] | 0.817486761 | 0.913698059 |
| S_LDL_P | ASAT_Pos_pPS | 4255 | -0.021756379 | [-0.05 0.01] | 0.157141687 | 0.431435756 |
| S_LDL_P | ALP_Neg_pPS | 4255 | 0.050739433 | [0.02 0.08] | 0.000962842 | 0.008545465 |
| S_LDL_P | SHBG-LpA_pPS | 4255 | 0.013659375 | [-0.02 0.04] | 0.374452843 | 0.655035906 |
| S_LDL_P | Obesity_pPS | 4255 | 0.011777502 | [-0.02 0.04] | 0.443800896 | 0.706833249 |
| S_LDL_P | VAT_Neg_pPS | 4255 | 0.004950461 | [-0.03 0.04] | 0.747545803 | 0.8751969 |
| S_LDL_P | Bilirubin_pPS | 4255 | -0.014531935 | [-0.04 0.02] | 0.344707336 | 0.637861136 |
| S_LDL_P | Proinsulin_pPS | 4255 | -0.019556707 | [-0.05 0.01] | 0.203485566 | 0.489049344 |
| S_LDL_P | Hyper_Insulin_pPS | 4255 | -0.022441726 | [-0.05 0.01] | 0.144475005 | 0.4165382 |
| S_LDL_P | Beta_Cell_1_pPS | 4255 | 0.020229414 | [-0.01 0.05] | 0.188363591 | 0.471379886 |
| S_LDL_P | Beta_Cell_2_pPS | 4255 | 0.020955511 | [-0.01 0.05] | 0.172989457 | 0.451531803 |
| S_LDL_PL | Cholesterol_pPS | 4255 | -0.148365007 | [-0.18 -0.12] | 3.01E-22 | 3.34E-20 |
| S_LDL_PL | Lipodystrophy_1_pPS | 4255 | 0.04120259 | [0.01 0.07] | 0.007360047 | 0.042833763 |
| S_LDL_PL | Lipodystrophy_2_pPS | 4255 | -0.046915669 | [-0.08 -0.02] | 0.002272455 | 0.016876323 |
| S_LDL_PL | Liver-Lipid_pPS | 4255 | 0.013487449 | [-0.04 0.02] | 0.380495646 | 0.661235752 |
| S_LDL_PL | ASAT_Pos_pPS | 4255 | -0.01982752 | [-0.05 0.01] | 0.197294938 | 0.482866095 |
| S_LDL_PL | ALP_Neg_pPS | 4255 | 0.051455899 | [0.02 0.08] | 0.000814438 | 0.00751385 |
| S_LDL_PL | SHBG-LpA_pPS | 4255 | 0.013901862 | [-0.02 0.04] | 0.36603145 | 0.650982927 |
| S_LDL_PL | Obesity_pPS | 4255 | 0.01519344 | [-0.01 0.05] | 0.323190487 | 0.620844623 |
| S_LDL_PL | VAT_Neg_pPS | 4255 | -0.001625037 | [-0.03 0.03] | 0.91585312 | 0.966041099 |
| S_LDL_PL | Bilirubin_pPS | 4255 | -0.016758018 | [-0.05 0.01] | 0.275858579 | 0.574084069 |
| S_LDL_PL | Proinsulin_pPS | 4255 | -0.017701633 | [-0.05 0.01] | 0.249717128 | 0.545779139 |
| S_LDL_PL | Hyper_Insulin_pPS | 4255 | -0.020115336 | [-0.05 0.01] | 0.190867895 | 0.474162704 |
| S_LDL_PL | Beta_Cell_1_pPS | 4255 | 0.025947093 | [-0. 0.06] | 0.091537221 | 0.303406485 |
| S_LDL_PL | Beta_Cell_2_pPS | 4255 | 0.016217954 | [-0.01 0.05] | 0.291632001 | 0.588843777 |

**Table S9: Associations between pPSs and global metabolomic profile in SCAPIS**

**a) Spearman Correlation**

| Metabolomics | PPS | Samp | r | CI95% | p | fdr_p_adjusted |
|---|---|---|---|---|---|---|
| S_HDL_TG | Hyper_Insulin_pPS | 4255 | 0.017023857 | [-0.01 0.05] | 0.267577332 | 0.552941597 |
| S_HDL_TG | Beta_Cell_1_pPS | 4255 | 0.009402707 | [-0.02 0.04] | 0.540332967 | 0.754566546 |
| S_HDL_TG | Beta_Cell_2_pPS | 4255 | -0.006205432 | [-0.04 0.02] | 0.686141984 | 0.842205031 |
| S_HDL_TG | Total_GRS | 4255 | 0.043637793 | [0.01 0.07] | 0.004468981 | 0.024963447 |
| S_LDL_C | Cholesterol_pPS | 4255 | -0.1526013 | [-0.18 -0.12] | 1.59E-23 | 2.13E-21 |
| S_LDL_C | Lipodystrophy_1_pPS | 4255 | 0.033256032 | [0. 0.06] | 0.0302957 | 0.119237203 |
| S_LDL_C | Lipodystrophy_2_pPS | 4255 | -0.043248575 | [-0.07 -0.01] | 0.00483799 | 0.026243573 |
| S_LDL_C | Liver-Lipid_pPS | 4255 | 0.011747917 | [-0.02 0.04] | 0.444247472 | 0.692953671 |
| S_LDL_C | ASAT_Pos_pPS | 4255 | -0.016745849 | [-0.05 0.01] | 0.275470315 | 0.560079455 |
| S_LDL_C | ALP_Neg_pPS | 4255 | 0.045220151 | [0.02 0.08] | 0.003217006 | 0.019717477 |
| S_LDL_C | SHBG-LpA_pPS | 4255 | 0.015846811 | [-0.01 0.05] | 0.302074372 | 0.587800395 |
| S_LDL_C | Obesity_pPS | 4255 | -0.00877604 | [-0.04 0.02] | 0.567661438 | 0.769278765 |
| S_LDL_C | VAT_Neg_pPS | 4255 | 0.010534115 | [-0.02 0.04] | 0.492718546 | 0.721879553 |
| S_LDL_C | Bilirubin_pPS | 4255 | -0.013636528 | [-0.04 0.02] | 0.374519236 | 0.645644896 |
| S_LDL_C | Proinsulin_pPS | 4255 | -0.007750001 | [-0.04 0.02] | 0.613783042 | 0.796469827 |
| S_LDL_C | Hyper_Insulin_pPS | 4255 | -0.020794999 | [-0.05 0.01] | 0.175641161 | 0.442714795 |
| S_LDL_C | Beta_Cell_1_pPS | 4255 | 0.022807967 | [-0.01 0.05] | 0.13743049 | 0.379881959 |
| S_LDL_C | Beta_Cell_2_pPS | 4255 | 0.023726317 | [-0.01 0.05] | 0.122284727 | 0.352554757 |
| S_LDL_C | Total_GRS | 4255 | -0.00526328 | [-0.04 0.02] | 0.731792158 | 0.872537948 |
| S_LDL_CE | Cholesterol_pPS | 4255 | -0.153433483 | [-0.18 -0.12] | 9.09E-24 | 1.63E-21 |
| S_LDL_CE | Lipodystrophy_1_pPS | 4255 | 0.034542021 | [0. 0.06] | 0.024447837 | 0.100460938 |
| S_LDL_CE | Lipodystrophy_2_pPS | 4255 | -0.041516304 | [-0.07 -0.01] | 0.006837201 | 0.035001898 |
| S_LDL_CE | Liver-Lipid_pPS | 4255 | 0.013774562 | [-0.02 0.04] | 0.369702946 | 0.643918869 |
| S_LDL_CE | ASAT_Pos_pPS | 4255 | -0.015066408 | [-0.05 0.02] | 0.326510407 | 0.609784284 |
| S_LDL_CE | ALP_Neg_pPS | 4255 | 0.0450211 | [0.01 0.08] | 0.003354642 | 0.020269596 |
| S_LDL_CE | SHBG-LpA_pPS | 4255 | 0.016076702 | [-0.01 0.05] | 0.295114179 | 0.581285504 |
| S_LDL_CE | Obesity_pPS | 4255 | -0.008585316 | [-0.04 0.02] | 0.576108108 | 0.773311572 |
| S_LDL_CE | VAT_Neg_pPS | 4255 | 0.011768629 | [-0.02 0.04] | 0.443444665 | 0.692953671 |
| S_LDL_CE | Bilirubin_pPS | 4255 | -0.013453959 | [-0.04 0.02] | 0.380948683 | 0.652142798 |
| S_LDL_CE | Proinsulin_pPS | 4255 | -0.007807506 | [-0.04 0.02] | 0.611154846 | 0.795465501 |
| S_LDL_CE | Hyper_Insulin_pPS | 4255 | -0.019792004 | [-0.05 0.01] | 0.197410551 | 0.47103115 |
| S_LDL_CE | Beta_Cell_1_pPS | 4255 | 0.021818833 | [-0.01 0.05] | 0.155318967 | 0.407783579 |
| S_LDL_CE | Beta_Cell_2_pPS | 4255 | 0.02317658 | [-0.01 0.05] | 0.131186055 | 0.36928358 |
| S_LDL_CE | Total_GRS | 4255 | -0.00406899 | [-0.03 0.03] | 0.79103244 | 0.900618145 |
| S_LDL_FC | Cholesterol_pPS | 4255 | -0.144692858 | [-0.17 -0.12] | 2.74E-21 | 2.35E-19 |
| S_LDL_FC | Lipodystrophy_1_pPS | 4255 | 0.027918385 | [-0. 0.06] | 0.069006815 | 0.223594589 |
| S_LDL_FC | Lipodystrophy_2_pPS | 4255 | -0.048584395 | [-0.08 -0.02] | 0.001547408 | 0.011854253 |
| S_LDL_FC | Liver-Lipid_pPS | 4255 | 0.002966845 | [-0.03 0.03] | 0.846803304 | 0.92810697 |
| S_LDL_FC | ASAT_Pos_pPS | 4255 | -0.021868495 | [-0.05 0.01] | 0.154380631 | 0.407130084 |
| S_LDL_FC | ALP_Neg_pPS | 4255 | 0.044138976 | [0.01 0.07] | 0.004031418 | 0.023183357 |
| S_LDL_FC | SHBG-LpA_pPS | 4255 | 0.014998261 | [-0.02 0.05] | 0.32870356 | 0.610965216 |
| S_LDL_FC | Obesity_pPS | 4255 | -0.008425771 | [-0.04 0.02] | 0.58321919 | 0.77847241 |
| S_LDL_FC | VAT_Neg_pPS | 4255 | 0.005134507 | [-0.02 0.04] | 0.738110324 | 0.874278301 |
| S_LDL_FC | Bilirubin_pPS | 4255 | -0.015054919 | [-0.05 0.02] | 0.326879487 | 0.609784284 |
| S_LDL_FC | Proinsulin_pPS | 4255 | -0.007251411 | [-0.04 0.02] | 0.63677569 | 0.810613564 |
| S_LDL_FC | Hyper_Insulin_pPS | 4255 | -0.024239547 | [-0.05 0.01] | 0.114407187 | 0.334337081 |
| S_LDL_FC | Beta_Cell_1_pPS | 4255 | 0.024841165 | [-0.01 0.05] | 0.105687496 | 0.314423966 |
| S_LDL_FC | Beta_Cell_2_pPS | 4255 | 0.023398447 | [-0.01 0.05] | 0.127534961 | 0.360900385 |
| S_LDL_FC | Total_GRS | 4255 | -0.009663603 | [-0.04 0.02] | 0.529152668 | 0.744283589 |
| S_LDL_L | Cholesterol_pPS | 4255 | -0.154449606 | [-0.18 -0.12] | 4.58E-24 | 1.36E-21 |
| S_LDL_L | Lipodystrophy_1_pPS | 4255 | 0.037530096 | [0.01 0.07] | 0.014493725 | 0.064101114 |
| S_LDL_L | Lipodystrophy_2_pPS | 4255 | -0.045484542 | [-0.08 -0.02] | 0.003042143 | 0.01910574 |
| S_LDL_L | Liver-Lipid_pPS | 4255 | 0.002959163 | [-0.03 0.03] | 0.847195081 | 0.92810697 |
| S_LDL_L | ASAT_Pos_pPS | 4255 | -0.017890458 | [-0.05 0.01] | 0.24397683 | 0.525480742 |
| S_LDL_L | ALP_Neg_pPS | 4255 | 0.047120998 | [0.02 0.08] | 0.002139274 | 0.014660517 |
| S_LDL_L | SHBG-LpA_pPS | 4255 | 0.016308893 | [-0.01 0.05] | 0.288194189 | 0.57504794 |
| S_LDL_L | Obesity_pPS | 4255 | -0.004592136 | [-0.03 0.03] | 0.764911473 | 0.889697961 |
| S_LDL_L | VAT_Neg_pPS | 4255 | 0.008453093 | [-0.02 0.04] | 0.581998541 | 0.778303354 |
| S_LDL_L | Bilirubin_pPS | 4255 | -0.013239539 | [-0.04 0.02] | 0.388585752 | 0.657347349 |
| S_LDL_L | Proinsulin_pPS | 4255 | -0.004340382 | [-0.03 0.03] | 0.77745037 | 0.89275053 |
| S_LDL_L | Hyper_Insulin_pPS | 4255 | -0.021277576 | [-0.05 0.01] | 0.165828167 | 0.427012507 |
| S_LDL_L | Beta_Cell_1_pPS | 4255 | 0.024774084 | [-0.01 0.05] | 0.106632834 | 0.316358823 |
| S_LDL_L | Beta_Cell_2_pPS | 4255 | 0.023144061 | [-0.01 0.05] | 0.131727931 | 0.369964431 |
| S_LDL_L | Total_GRS | 4255 | -0.002525292 | [-0.03 0.03] | 0.869380804 | 0.940757973 |
| S_LDL_P | Cholesterol_pPS | 4255 | -0.155115049 | [-0.18 -0.13] | 2.91E-24 | 1.25E-21 |
| S_LDL_P | Lipodystrophy_1_pPS | 4255 | 0.039226231 | [0.01 0.07] | 0.010607767 | 0.049363789 |
| S_LDL_P | Lipodystrophy_2_pPS | 4255 | -0.045112304 | [-0.08 -0.02] | 0.003290926 | 0.019940781 |
| S_LDL_P | Liver-Lipid_pPS | 4255 | 0.001194984 | [-0.03 0.03] | 0.93797416 | 0.970084172 |
| S_LDL_P | ASAT_Pos_pPS | 4255 | -0.017651891 | [-0.05 0.01] | 0.250323004 | 0.531626579 |
| S_LDL_P | ALP_Neg_pPS | 4255 | 0.047540757 | [0.02 0.08] | 0.001951184 | 0.013858578 |
| S_LDL_P | SHBG-LpA_pPS | 4255 | 0.016395343 | [-0.01 0.05] | 0.285645885 | 0.574775256 |
| S_LDL_P | Obesity_pPS | 4255 | -0.003558937 | [-0.03 0.03] | 0.816727832 | 0.915817998 |
| S_LDL_P | VAT_Neg_pPS | 4255 | 0.008521806 | [-0.02 0.04] | 0.578933938 | 0.776133311 |
| S_LDL_P | Bilirubin_pPS | 4255 | -0.013189369 | [-0.04 0.02] | 0.390386037 | 0.658788398 |
| S_LDL_P | Proinsulin_pPS | 4255 | -0.003327889 | [-0.03 0.03] | 0.828434497 | 0.92223257 |
| S_LDL_P | Hyper_Insulin_pPS | 4255 | -0.020992201 | [-0.05 0.01] | 0.171580066 | 0.435034565 |
| S_LDL_P | Beta_Cell_1_pPS | 4255 | 0.024856035 | [-0.01 0.05] | 0.10547884 | 0.314239045 |
| S_LDL_P | Beta_Cell_2_pPS | 4255 | 0.022841076 | [-0.01 0.05] | 0.136860405 | 0.378794282 |
| S_LDL_P | Total_GRS | 4255 | -0.00133419 | [-0.03 0.03] | 0.930765824 | 0.966824549 |
| S_LDL_PL | Cholesterol_pPS | 4255 | -0.149811184 | [-0.18 -0.12] | 1.01E-22 | 1.16E-20 |
| S_LDL_PL | Lipodystrophy_1_pPS | 4255 | 0.035321161 | [0.01 0.07] | 0.02140289 | 0.090194888 |
| S_LDL_PL | Lipodystrophy_2_pPS | 4255 | -0.054079189 | [-0.08 -0.02] | 0.000424814 | 0.004488803 |
| S_LDL_PL | Liver-Lipid_pPS | 4255 | -0.015542168 | [-0.05 0.01] | 0.311464677 | 0.599185411 |
| S_LDL_PL | ASAT_Pos_pPS | 4255 | -0.01691511 | [-0.05 0.01] | 0.270646082 | 0.554396235 |
| S_LDL_PL | ALP_Neg_pPS | 4255 | 0.047845497 | [0.02 0.08] | 0.001824278 | 0.013239667 |
| S_LDL_PL | SHBG-LpA_pPS | 4255 | 0.015807908 | [-0.01 0.05] | 0.303262921 | 0.588686846 |
| S_LDL_PL | Obesity_pPS | 4255 | 0.000484755 | [-0.03 0.03] | 0.974817485 | 0.984455511 |
| S_LDL_PL | VAT_Neg_pPS | 4255 | 0.001007395 | [-0.03 0.03] | 0.947695711 | 0.974500144 |
| S_LDL_PL | Bilirubin_pPS | 4255 | -0.015053535 | [-0.05 0.02] | 0.326923975 | 0.609784284 |
| S_LDL_PL | Proinsulin_pPS | 4255 | -0.000550097 | [-0.03 0.03] | 0.971424446 | 0.983305352 |
| S_LDL_PL | Hyper_Insulin_pPS | 4255 | -0.020521806 | [-0.05 0.01] | 0.181385012 | 0.45159215 |
| S_LDL_PL | Beta_Cell_1_pPS | 4255 | 0.028994701 | [-0. 0.06] | 0.058957987 | 0.197182522 |
| S_LDL_PL | Beta_Cell_2_pPS | 4255 | 0.018921193 | [-0.01 0.05] | 0.217858773 | 0.498643401 |
| S_LDL_PL | Total_GRS | 4255 | -0.00369782 | [-0.03 0.03] | 0.809710415 | 0.911242833 |

**b) Partial Correlation**

| Metabolomics | PPS | Samp | r | CI95% | p | fdr_p_adjusted |
|---|---|---|---|---|---|---|
| S_LDL_TG | Cholesterol_pPS | 4255 | -0.11869865 | [-0.15 -0.09] | 9.55E-15 | 4.66E-13 |
| S_LDL_TG | Lipodystrophy_1_pPS | 4255 | 0.062625953 | [0.03 0.09] | 4.58E-05 | 0.000797984 |
| S_LDL_TG | Lipodystrophy_2_pPS | 4255 | -0.014387283 | [-0.04 0.02] | 0.349531496 | 0.637861136 |
| S_LDL_TG | Liver-Lipid_pPS | 4255 | -0.037463126 | [-0.07 -0.01] | 0.014822837 | 0.073091921 |
| S_LDL_TG | ASAT_Pos_pPS | 4255 | -0.0081532 | [-0.04 0.02] | 0.59602625 | 0.818411902 |
| S_LDL_TG | ALP_Neg_pPS | 4255 | 0.039516515 | [0.01 0.07] | 0.010159956 | 0.054678045 |
| S_LDL_TG | SHBG-LpA_pPS | 4255 | 0.002870941 | [-0.03 0.03] | 0.851921921 | 0.935059038 |
| S_LDL_TG | Obesity_pPS | 4255 | 0.027854977 | [-0. 0.06] | 0.070069587 | 0.243118395 |
| S_LDL_TG | VAT_Neg_pPS | 4255 | -0.004666561 | [-0.03 0.03] | 0.761571449 | 0.884377054 |
| S_LDL_TG | Bilirubin_pPS | 4255 | -0.010826652 | [-0.04 0.02] | 0.481456937 | 0.740874464 |
| S_LDL_TG | Proinsulin_pPS | 4255 | 0.003001013 | [-0.03 0.03] | 0.845295868 | 0.930848365 |
| S_LDL_TG | Hyper_Insulin_pPS | 4255 | -0.013433844 | [-0.04 0.02] | 0.382391924 | 0.661657399 |
| S_LDL_TG | Beta_Cell_1_pPS | 4255 | 0.008204995 | [-0.02 0.04] | 0.593693433 | 0.817359021 |
| S_LDL_TG | Beta_Cell_2_pPS | 4255 | 0.009008187 | [-0.02 0.04] | 0.558069037 | 0.797469102 |
| S_VLDL_C | Cholesterol_pPS | 4255 | -0.115959447 | [-0.15 -0.09] | 3.87E-14 | 1.80E-12 |
| S_VLDL_C | Lipodystrophy_1_pPS | 4255 | 0.06381446 | [0.03 0.09] | 3.28E-05 | 0.000601648 |
| S_VLDL_C | Lipodystrophy_2_pPS | 4255 | -0.020827878 | [-0.05 0.01] | 0.175621731 | 0.453447086 |
| S_VLDL_C | Liver-Lipid_pPS | 4255 | -0.011682784 | [-0.04 0.02] | 0.44747489 | 0.709019056 |
| S_VLDL_C | ASAT_Pos_pPS | 4255 | 0.012672383 | [-0.02 0.04] | 0.409949982 | 0.68222765 |
| S_VLDL_C | ALP_Neg_pPS | 4255 | 0.037434378 | [0.01 0.07] | 0.014899704 | 0.073290438 |
| S_VLDL_C | SHBG-LpA_pPS | 4255 | 0.001194276 | [-0.03 0.03] | 0.938105749 | 0.978170683 |
| S_VLDL_C | Obesity_pPS | 4255 | 0.018562562 | [-0.01 0.05] | 0.227421577 | 0.522130731 |
| S_VLDL_C | VAT_Neg_pPS | 4255 | 0.007242877 | [-0.02 0.04] | 0.637688969 | 0.835456904 |
| S_VLDL_C | Bilirubin_pPS | 4255 | -0.020787884 | [-0.05 0.01] | 0.176452678 | 0.454059461 |
| S_VLDL_C | Proinsulin_pPS | 4255 | -0.001112279 | [-0.03 0.03] | 0.942347626 | 0.97851657 |
| S_VLDL_C | Hyper_Insulin_pPS | 4255 | -0.007261668 | [-0.04 0.02] | 0.63681666 | 0.835456904 |
| S_VLDL_C | Beta_Cell_1_pPS | 4255 | 0.013097989 | [-0.02 0.04] | 0.394404129 | 0.672569903 |
| S_VLDL_C | Beta_Cell_2_pPS | 4255 | 0.010490582 | [-0.02 0.04] | 0.495170446 | 0.753425828 |
| S_VLDL_CE | Cholesterol_pPS | 4255 | -0.12272099 | [-0.15 -0.09] | 1.15E-15 | 5.91E-14 |
| S_VLDL_CE | Lipodystrophy_1_pPS | 4255 | 0.060152597 | [0.03 0.09] | 9.05E-05 | 0.001352793 |
| S_VLDL_CE | Lipodystrophy_2_pPS | 4255 | -0.027622022 | [-0.06 0. ] | 0.072445982 | 0.248842895 |
| S_VLDL_CE | Liver-Lipid_pPS | 4255 | -0.00027538 | [-0.03 0.03] | 0.985714645 | 0.993363084 |
| S_VLDL_CE | ASAT_Pos_pPS | 4255 | 0.009233019 | [-0.02 0.04] | 0.548281949 | 0.791962815 |
| S_VLDL_CE | ALP_Neg_pPS | 4255 | 0.043185382 | [0.01 0.07] | 0.004966622 | 0.030500541 |
| S_VLDL_CE | SHBG-LpA_pPS | 4255 | 0.005449682 | [-0.02 0.04] | 0.723086548 | 0.866319132 |
| S_VLDL_CE | Obesity_pPS | 4255 | 0.015313152 | [-0.01 0.05] | 0.319392197 | 0.620197069 |
| S_VLDL_CE | VAT_Neg_pPS | 4255 | 0.008585144 | [-0.02 0.04] | 0.576701201 | 0.803773963 |
| S_VLDL_CE | Bilirubin_pPS | 4255 | -0.022434607 | [-0.05 0.01] | 0.144602422 | 0.4165382 |
| S_VLDL_CE | Proinsulin_pPS | 4255 | -0.005339252 | [-0.04 0.02] | 0.728473731 | 0.868097863 |
| S_VLDL_CE | Hyper_Insulin_pPS | 4255 | -0.009697102 | [-0.04 0.02] | 0.528359046 | 0.77834791 |
| S_VLDL_CE | Beta_Cell_1_pPS | 4255 | 0.01217076 | [-0.02 0.04] | 0.428732347 | 0.696591166 |
| S_VLDL_CE | Beta_Cell_2_pPS | 4255 | 0.01571961 | [-0.01 0.05] | 0.30671462 | 0.60676153 |
| S_VLDL_FC | Cholesterol_pPS | 4255 | -0.097794728 | [-0.13 -0.07] | 1.84E-10 | 6.71E-09 |
| S_VLDL_FC | Lipodystrophy_1_pPS | 4255 | 0.064529348 | [0.03 0.09] | 2.67E-05 | 0.000504059 |
| S_VLDL_FC | Lipodystrophy_2_pPS | 4255 | -0.004828277 | [-0.03 0.03] | 0.753572196 | 0.878099599 |
| S_VLDL_FC | Liver-Lipid_pPS | 4255 | -0.031269872 | [-0.06 -0. ] | 0.041987884 | 0.160489904 |
| S_VLDL_FC | ASAT_Pos_pPS | 4255 | 0.017763928 | [-0.01 0.05] | 0.248054371 | 0.545779139 |
| S_VLDL_FC | ALP_Neg_pPS | 4255 | 0.024685347 | [-0.01 0.05] | 0.108435108 | 0.342480177 |
| S_VLDL_FC | SHBG-LpA_pPS | 4255 | -0.005290853 | [-0.04 0.02] | 0.730839094 | 0.869031685 |
| S_VLDL_FC | Obesity_pPS | 4255 | 0.024571886 | [-0.01 0.05] | 0.110068285 | 0.343926372 |
| S_VLDL_FC | VAT_Neg_pPS | 4255 | 0.004365442 | [-0.03 0.03] | 0.776533743 | 0.891327566 |
| S_VLDL_FC | Bilirubin_pPS | 4255 | -0.012181377 | [-0.04 0.02] | 0.428329702 | 0.696591166 |
| S_VLDL_FC | Proinsulin_pPS | 4255 | 0.005078314 | [-0.03 0.04] | 0.741256261 | 0.873045882 |
| S_VLDL_FC | Hyper_Insulin_pPS | 4255 | -0.002476074 | [-0.03 0.03] | 0.872099153 | 0.946048963 |
| S_VLDL_FC | Beta_Cell_1_pPS | 4255 | 0.013020046 | [-0.02 0.04] | 0.397224125 | 0.673455453 |
| S_VLDL_FC | Beta_Cell_2_pPS | 4255 | 0.001222684 | [-0.03 0.03] | 0.936636501 | 0.978170683 |
| S_VLDL_L | Cholesterol_pPS | 4255 | -0.099561456 | [-0.13 -0.07] | 8.61E-11 | 3.19E-09 |
| S_VLDL_L | Lipodystrophy_1_pPS | 4255 | 0.06551957 | [0.04 0.1 ] | 2.00E-05 | 0.000389219 |
| S_VLDL_L | Lipodystrophy_2_pPS | 4255 | -0.00137685 | [-0.03 0.03] | 0.928667274 | 0.974930195 |
| S_VLDL_L | Liver-Lipid_pPS | 4255 | -0.031068981 | [-0.06 -0. ] | 0.043324408 | 0.164271714 |
| S_VLDL_L | ASAT_Pos_pPS | 4255 | 0.022735302 | [-0.01 0.05] | 0.13929463 | 0.408298462 |
| S_VLDL_L | ALP_Neg_pPS | 4255 | 0.018912019 | [-0.01 0.05] | 0.218788912 | 0.512298715 |
| S_VLDL_L | SHBG-LpA_pPS | 4255 | -0.005410816 | [-0.04 0.02] | 0.724981041 | 0.866514653 |
| S_VLDL_L | Obesity_pPS | 4255 | 0.027774533 | [-0. 0.06] | 0.070882872 | 0.244668121 |
| S_VLDL_L | VAT_Neg_pPS | 4255 | 0.007439444 | [-0.02 0.04] | 0.628589162 | 0.835060055 |
| S_VLDL_L | Bilirubin_pPS | 4255 | -0.013671779 | [-0.04 0.02] | 0.374019177 | 0.655035906 |
| S_VLDL_L | Proinsulin_pPS | 4255 | 0.005041652 | [-0.03 0.04] | 0.743058055 | 0.873512116 |
| S_VLDL_L | Hyper_Insulin_pPS | 4255 | -0.000870083 | [-0.03 0.03] | 0.954886034 | 0.980852662 |
| S_VLDL_L | Beta_Cell_1_pPS | 4255 | 0.011972218 | [-0.02 0.04] | 0.436302452 | 0.700747966 |
| S_VLDL_L | Beta_Cell_2_pPS | 4255 | 0.001060466 | [-0.03 0.03] | 0.94502885 | 0.978747744 |
| S_VLDL_P | Cholesterol_pPS | 4255 | -0.097028413 | [-0.13 -0.07] | 2.55E-10 | 9.13E-09 |
| S_VLDL_P | Lipodystrophy_1_pPS | 4255 | 0.065356222 | [0.04 0.1 ] | 2.10E-05 | 0.000404295 |
| S_VLDL_P | Lipodystrophy_2_pPS | 4255 | 0.001152712 | [-0.03 0.03] | 0.940255714 | 0.978430443 |
| S_VLDL_P | Liver-Lipid_pPS | 4255 | -0.033402813 | [-0.06 -0. ] | 0.029823146 | 0.122098033 |
| S_VLDL_P | ASAT_Pos_pPS | 4255 | 0.023932352 | [-0.01 0.05] | 0.119640115 | 0.364405465 |
| S_VLDL_P | ALP_Neg_pPS | 4255 | 0.016245955 | [-0.01 0.05] | 0.290799582 | 0.588469156 |
| S_VLDL_P | SHBG-LpA_pPS | 4255 | -0.006401284 | [-0.04 0.02] | 0.67725576 | 0.855435982 |
| S_VLDL_P | Obesity_pPS | 4255 | 0.029014159 | [-0. 0.06] | 0.05917708 | 0.214236011 |
| S_VLDL_P | VAT_Neg_pPS | 4255 | 0.007773667 | [-0.02 0.04] | 0.613246007 | 0.828794354 |
| S_VLDL_P | Bilirubin_pPS | 4255 | -0.012878934 | [-0.04 0.02] | 0.40236042 | 0.677078992 |
| S_VLDL_P | Proinsulin_pPS | 4255 | 0.005802317 | [-0.02 0.04] | 0.705976397 | 0.862333586 |
| S_VLDL_P | Hyper_Insulin_pPS | 4255 | 0.000145537 | [-0.03 0.03] | 0.992449952 | 0.994934804 |
| S_VLDL_P | Beta_Cell_1_pPS | 4255 | 0.011426489 | [-0.02 0.04] | 0.45750242 | 0.721196728 |
| S_VLDL_P | Beta_Cell_2_pPS | 4255 | 0.000448048 | [-0.03 0.03] | 0.976759549 | 0.991343605 |
| S_VLDL_PL | Cholesterol_pPS | 4255 | -0.092463205 | [-0.12 -0.06] | 1.69E-09 | 5.22E-08 |
| S_VLDL_PL | Lipodystrophy_1_pPS | 4255 | 0.062428643 | [0.03 0.09] | 4.84E-05 | 0.000814944 |
| S_VLDL_PL | Lipodystrophy_2_pPS | 4255 | -0.001559263 | [-0.03 0.03] | 0.919247127 | 0.968086664 |
| S_VLDL_PL | Liver-Lipid_pPS | 4255 | -0.033234256 | [-0.06 -0. ] | 0.030659612 | 0.124252113 |
| S_VLDL_PL | ASAT_Pos_pPS | 4255 | 0.021924332 | [-0.01 0.05] | 0.153962324 | 0.428688101 |
| S_VLDL_PL | ALP_Neg_pPS | 4255 | 0.021162821 | [-0.01 0.05] | 0.168776938 | 0.444593986 |
| S_VLDL_PL | SHBG-LpA_pPS | 4255 | -0.006104483 | [-0.04 0.02] | 0.691431986 | 0.858568103 |
| S_VLDL_PL | Obesity_pPS | 4255 | 0.026723357 | [-0. 0.06] | 0.082238593 | 0.277174517 |
| S_VLDL_PL | VAT_Neg_pPS | 4255 | 0.003364028 | [-0.03 0.03] | 0.826862595 | 0.920677928 |
| S_VLDL_PL | Bilirubin_pPS | 4255 | -0.007850147 | [-0.04 0.02] | 0.609758397 | 0.828179316 |

**Table S9: Associations between pPSs and global metabolomic profile in SCAPIS**

**a) Spearman Correlation**

| Metabolomics | PPS | Samp | r | CI95% | p | fdr_p_adjusted |
|---|---|---|---|---|---|---|
| S_LDL_TG | Cholesterol_pPS | 4255 | -0.117485781 | [-0.15 -0.09] | 1.63E-14 | 8.51E-13 |
| S_LDL_TG | Lipodystrophy_1_pPS | 4255 | 0.061026431 | [0.03 0.09] | 6.96E-05 | 0.001043798 |
| S_LDL_TG | Lipodystrophy_2_pPS | 4255 | -0.018424891 | [-0.05 0.01] | 0.230172525 | 0.513581257 |
| S_LDL_TG | Liver-Lipid_pPS | 4255 | -0.037809369 | [-0.07 -0.01] | 0.013778241 | 0.061829137 |
| S_LDL_TG | ASAT_Pos_pPS | 4255 | 0.00189556 | [-0.03 0.03] | 0.901760829 | 0.95179735 |
| S_LDL_TG | ALP_Neg_pPS | 4255 | 0.03999974 | [0.01 0.07] | 0.009166243 | 0.044382826 |
| S_LDL_TG | SHBG-LpA_pPS | 4255 | 0.007214093 | [-0.02 0.04] | 0.638511135 | 0.811859149 |
| S_LDL_TG | Obesity_pPS | 4255 | 0.019471469 | [-0.01 0.05] | 0.204767538 | 0.482946722 |
| S_LDL_TG | VAT_Neg_pPS | 4255 | -0.000622628 | [-0.03 0.03] | 0.967658663 | 0.98241424 |
| S_LDL_TG | Bilirubin_pPS | 4255 | -0.008322327 | [-0.04 0.02] | 0.587851571 | 0.780285656 |
| S_LDL_TG | Proinsulin_pPS | 4255 | 0.014140988 | [-0.02 0.04] | 0.357105361 | 0.635677178 |
| S_LDL_TG | Hyper_Insulin_pPS | 4255 | -0.008689345 | [-0.04 0.02] | 0.571493582 | 0.771552531 |
| S_LDL_TG | Beta_Cell_1_pPS | 4255 | 0.01393838 | [-0.02 0.04] | 0.364037193 | 0.639224774 |
| S_LDL_TG | Beta_Cell_2_pPS | 4255 | 0.010355815 | [-0.02 0.04] | 0.500069977 | 0.72623568 |
| S_LDL_TG | Total_GRS | 4255 | 0.02174173 | [-0.01 0.05] | 0.15678436 | 0.410124941 |
| S_VLDL_C | Cholesterol_pPS | 4255 | -0.115795246 | [-0.15 -0.09] | 3.85E-14 | 1.92E-12 |
| S_VLDL_C | Lipodystrophy_1_pPS | 4255 | 0.066332703 | [0.04 0.1 ] | 1.53E-05 | 0.000260505 |
| S_VLDL_C | Lipodystrophy_2_pPS | 4255 | -0.020208651 | [-0.05 0.01] | 0.18813914 | 0.459633775 |
| S_VLDL_C | Liver-Lipid_pPS | 4255 | -0.011617783 | [-0.04 0.02] | 0.449310313 | 0.694281993 |
| S_VLDL_C | ASAT_Pos_pPS | 4255 | 0.02302697 | [-0.01 0.05] | 0.133693451 | 0.373106933 |
| S_VLDL_C | ALP_Neg_pPS | 4255 | 0.038307055 | [0.01 0.07] | 0.012580185 | 0.057291924 |
| S_VLDL_C | SHBG-LpA_pPS | 4255 | 0.006750877 | [-0.02 0.04] | 0.660215102 | 0.824723182 |
| S_VLDL_C | Obesity_pPS | 4255 | 0.011455598 | [-0.02 0.04] | 0.455665637 | 0.696320363 |
| S_VLDL_C | VAT_Neg_pPS | 4255 | 0.012188724 | [-0.02 0.04] | 0.427341863 | 0.687522331 |
| S_VLDL_C | Bilirubin_pPS | 4255 | -0.017793137 | [-0.05 0.01] | 0.246551901 | 0.528325503 |
| S_VLDL_C | Proinsulin_pPS | 4255 | 0.011144114 | [-0.02 0.04] | 0.468012009 | 0.707899576 |
| S_VLDL_C | Hyper_Insulin_pPS | 4255 | 0.000652471 | [-0.03 0.03] | 0.966109384 | 0.982019059 |
| S_VLDL_C | Beta_Cell_1_pPS | 4255 | 0.020912078 | [-0.01 0.05] | 0.173221524 | 0.437644487 |
| S_VLDL_C | Beta_Cell_2_pPS | 4255 | 0.013654997 | [-0.02 0.04] | 0.373872571 | 0.645644896 |
| S_VLDL_C | Total_GRS | 4255 | 0.021621665 | [-0.01 0.05] | 0.159087126 | 0.415641761 |
| S_VLDL_CE | Cholesterol_pPS | 4255 | -0.122970047 | [-0.15 -0.09] | 9.11E-16 | 5.01E-14 |
| S_VLDL_CE | Lipodystrophy_1_pPS | 4255 | 0.061577079 | [0.03 0.09] | 5.98E-05 | 0.000909509 |
| S_VLDL_CE | Lipodystrophy_2_pPS | 4255 | -0.027351525 | [-0.06 0. ] | 0.074840512 | 0.23924426 |
| S_VLDL_CE | Liver-Lipid_pPS | 4255 | -0.000568635 | [-0.03 0.03] | 0.97046187 | 0.983305352 |
| S_VLDL_CE | ASAT_Pos_pPS | 4255 | 0.017949102 | [-0.01 0.05] | 0.242434289 | 0.525274293 |
| S_VLDL_CE | ALP_Neg_pPS | 4255 | 0.043488404 | [0.01 0.07] | 0.004607488 | 0.025363691 |
| S_VLDL_CE | SHBG-LpA_pPS | 4255 | 0.010548429 | [-0.02 0.04] | 0.492130902 | 0.721879553 |
| S_VLDL_CE | Obesity_pPS | 4255 | 0.00690906 | [-0.02 0.04] | 0.652770059 | 0.819784413 |
| S_VLDL_CE | VAT_Neg_pPS | 4255 | 0.013110538 | [-0.02 0.04] | 0.393225006 | 0.660507155 |
| S_VLDL_CE | Bilirubin_pPS | 4255 | -0.019852782 | [-0.05 0.01] | 0.196037632 | 0.469833208 |
| S_VLDL_CE | Proinsulin_pPS | 4255 | 0.008118671 | [-0.02 0.04] | 0.597020892 | 0.787582911 |
| S_VLDL_CE | Hyper_Insulin_pPS | 4255 | -0.002426965 | [-0.03 0.03] | 0.874423672 | 0.942282701 |
| S_VLDL_CE | Beta_Cell_1_pPS | 4255 | 0.020396872 | [-0.01 0.05] | 0.184057706 | 0.455303634 |
| S_VLDL_CE | Beta_Cell_2_pPS | 4255 | 0.018297962 | [-0.01 0.05] | 0.233399703 | 0.516301672 |
| S_VLDL_CE | Total_GRS | 4255 | 0.016207198 | [-0.01 0.05] | 0.291211423 | 0.577755652 |
| S_VLDL_FC | Cholesterol_pPS | 4255 | -0.096684256 | [-0.13 -0.07] | 2.78E-10 | 1.08E-08 |
| S_VLDL_FC | Lipodystrophy_1_pPS | 4255 | 0.069134847 | [0.04 0.1 ] | 6.56E-06 | 0.000123486 |
| S_VLDL_FC | Lipodystrophy_2_pPS | 4255 | -0.003734062 | [-0.03 0.03] | 0.807881716 | 0.911007872 |
| S_VLDL_FC | Liver-Lipid_pPS | 4255 | -0.030713843 | [-0.06 -0. ] | 0.045443018 | 0.163549118 |
| S_VLDL_FC | ASAT_Pos_pPS | 4255 | 0.030457928 | [0. 0.06] | 0.047272142 | 0.168157123 |
| S_VLDL_FC | ALP_Neg_pPS | 4255 | 0.026225391 | [-0. 0.06] | 0.087623697 | 0.271608134 |
| S_VLDL_FC | SHBG-LpA_pPS | 4255 | 0.000488403 | [-0.03 0.03] | 0.974628036 | 0.984455511 |
| S_VLDL_FC | Obesity_pPS | 4255 | 0.01988732 | [-0.01 0.05] | 0.195260593 | 0.468494377 |
| S_VLDL_FC | VAT_Neg_pPS | 4255 | 0.010056284 | [-0.02 0.04] | 0.512549578 | 0.732945897 |
| S_VLDL_FC | Bilirubin_pPS | 4255 | -0.008774619 | [-0.04 0.02] | 0.567724142 | 0.769278765 |
| S_VLDL_FC | Proinsulin_pPS | 4255 | 0.01446593 | [-0.02 0.04] | 0.346162964 | 0.62662761 |
| S_VLDL_FC | Hyper_Insulin_pPS | 4255 | 0.006085116 | [-0.02 0.04] | 0.69191245 | 0.846635599 |
| S_VLDL_FC | Beta_Cell_1_pPS | 4255 | 0.0194013 | [-0.01 0.05] | 0.206404323 | 0.484925819 |
| S_VLDL_FC | Beta_Cell_2_pPS | 4255 | 0.004860779 | [-0.03 0.03] | 0.751599073 | 0.881192614 |
| S_VLDL_FC | Total_GRS | 4255 | 0.031133615 | [0. 0.06] | 0.042571963 | 0.155300782 |
| S_VLDL_L | Cholesterol_pPS | 4255 | -0.098111662 | [-0.13 -0.07] | 1.51E-10 | 6.00E-09 |
| S_VLDL_L | Lipodystrophy_1_pPS | 4255 | 0.071429627 | [0.04 0.1 ] | 3.20E-06 | 6.87E-05 |
| S_VLDL_L | Lipodystrophy_2_pPS | 4255 | 0.000694578 | [-0.03 0.03] | 0.963923737 | 0.982019059 |
| S_VLDL_L | Liver-Lipid_pPS | 4255 | -0.030488853 | [-0.06 -0. ] | 0.047047877 | 0.1679163 |
| S_VLDL_L | ASAT_Pos_pPS | 4255 | 0.036234718 | [0.01 0.07] | 0.018257796 | 0.078957605 |
| S_VLDL_L | ALP_Neg_pPS | 4255 | 0.020518681 | [-0.01 0.05] | 0.181451504 | 0.45159215 |
| S_VLDL_L | SHBG-LpA_pPS | 4255 | 0.000931526 | [-0.03 0.03] | 0.951629864 | 0.976205671 |
| S_VLDL_L | Obesity_pPS | 4255 | 0.023102083 | [-0.01 0.05] | 0.132429991 | 0.370838552 |
| S_VLDL_L | VAT_Neg_pPS | 4255 | 0.013725984 | [-0.02 0.04] | 0.37139353 | 0.643918869 |
| S_VLDL_L | Bilirubin_pPS | 4255 | -0.01010719 | [-0.04 0.02] | 0.51041726 | 0.7313594 |
| S_VLDL_L | Proinsulin_pPS | 4255 | 0.013631183 | [-0.02 0.04] | 0.374706506 | 0.645644896 |
| S_VLDL_L | Hyper_Insulin_pPS | 4255 | 0.008779694 | [-0.02 0.04] | 0.567500163 | 0.769278765 |
| S_VLDL_L | Beta_Cell_1_pPS | 4255 | 0.018749989 | [-0.01 0.05] | 0.222051893 | 0.502934916 |
| S_VLDL_L | Beta_Cell_2_pPS | 4255 | 0.004635889 | [-0.03 0.03] | 0.7627385 | 0.889654204 |
| S_VLDL_L | Total_GRS | 4255 | 0.034942081 | [0. 0.06] | 0.022840467 | 0.095131655 |
| S_VLDL_P | Cholesterol_pPS | 4255 | -0.095390858 | [-0.13 -0.07] | 4.79E-10 | 1.80E-08 |
| S_VLDL_P | Lipodystrophy_1_pPS | 4255 | 0.071682672 | [0.04 0.1 ] | 2.95E-06 | 6.47E-05 |
| S_VLDL_P | Lipodystrophy_2_pPS | 4255 | 0.003468904 | [-0.03 0.03] | 0.821284883 | 0.919444716 |
| S_VLDL_P | Liver-Lipid_pPS | 4255 | -0.03274816 | [-0.06 -0. ] | 0.032916236 | 0.127023499 |
| S_VLDL_P | ASAT_Pos_pPS | 4255 | 0.037766839 | [0.01 0.07] | 0.013885146 | 0.062178787 |
| S_VLDL_P | ALP_Neg_pPS | 4255 | 0.017951743 | [-0.01 0.05] | 0.242364977 | 0.525274293 |
| S_VLDL_P | SHBG-LpA_pPS | 4255 | 4.79E-05 | [-0.03 0.03] | 0.997511807 | 0.998442756 |
| S_VLDL_P | Obesity_pPS | 4255 | 0.024686143 | [-0.01 0.05] | 0.107882283 | 0.318932788 |
| S_VLDL_P | VAT_Neg_pPS | 4255 | 0.014218439 | [-0.02 0.04] | 0.354477634 | 0.634686582 |
| S_VLDL_P | Bilirubin_pPS | 4255 | -0.009253644 | [-0.04 0.02] | 0.54677345 | 0.760607725 |
| S_VLDL_P | Proinsulin_pPS | 4255 | 0.013876075 | [-0.02 0.04] | 0.366185636 | 0.64109339 |
| S_VLDL_P | Hyper_Insulin_pPS | 4255 | 0.010013502 | [-0.02 0.04] | 0.514345173 | 0.733381664 |
| S_VLDL_P | Beta_Cell_1_pPS | 4255 | 0.018209572 | [-0.01 0.05] | 0.235665891 | 0.518179785 |
| S_VLDL_P | Beta_Cell_2_pPS | 4255 | 0.004008623 | [-0.03 0.03] | 0.794062404 | 0.901959081 |
| S_VLDL_P | Total_GRS | 4255 | 0.03670631 | [0.01 0.07] | 0.016798507 | 0.07308884 |
| S_VLDL_PL | Cholesterol_pPS | 4255 | -0.091037657 | [-0.12 -0.06] | 2.84E-09 | 9.52E-08 |
| S_VLDL_PL | Lipodystrophy_1_pPS | 4255 | 0.068297711 | [0.04 0.1 ] | 8.48E-06 | 0.000154154 |
| S_VLDL_PL | Lipodystrophy_2_pPS | 4255 | -3.55E-05 | [-0.03 0.03] | 0.998153569 | 0.998619126 |
| S_VLDL_PL | Liver-Lipid_pPS | 4255 | -0.032632506 | [-0.06 -0. ] | 0.033539399 | 0.128697693 |

**b) Partial Correlation**

| Metabolomics | PPS | Samp | r | CI95% | p | fdr_p_adjusted |
|---|---|---|---|---|---|---|
| S_VLDL_PL | Proinsulin_pPS | 4255 | 0.005486377 | [-0.02 0.04] | 0.721299442 | 0.866319132 |
| S_VLDL_PL | Hyper_Insulin_pPS | 4255 | 0.000256924 | [-0.03 0.03] | 0.986671947 | 0.993363084 |
| S_VLDL_PL | Beta_Cell_1_pPS | 4255 | 0.012714399 | [-0.02 0.04] | 0.408399273 | 0.681346121 |
| S_VLDL_PL | Beta_Cell_2_pPS | 4255 | -0.00243256 | [-0.03 0.03] | 0.874328059 | 0.946048963 |
| S_VLDL_TG | Cholesterol_pPS | 4255 | -0.080027187 | [-0.11 -0.05] | 1.87E-07 | 4.21E-06 |
| S_VLDL_TG | Lipodystrophy_1_pPS | 4255 | 0.063078553 | [0.03 0.09] | 4.04E-05 | 0.00072794 |
| S_VLDL_TG | Lipodystrophy_2_pPS | 4255 | 0.014382973 | [-0.02 0.04] | 0.349675876 | 0.637861136 |
| S_VLDL_TG | Liver-Lipid_pPS | 4255 | -0.045858982 | [-0.08 -0.02] | 0.002851722 | 0.019954421 |
| S_VLDL_TG | ASAT_Pos_pPS | 4255 | 0.028180663 | [-0. 0.06] | 0.066854801 | 0.234401596 |
| S_VLDL_TG | ALP_Neg_pPS | 4255 | 0.00356863 | [-0.03 0.03] | 0.816514479 | 0.913698059 |
| S_VLDL_TG | SHBG-LpA_pPS | 4255 | -0.011734963 | [-0.04 0.02] | 0.445448791 | 0.708330803 |
| S_VLDL_TG | Obesity_pPS | 4255 | 0.034156337 | [0. 0.06] | 0.026319358 | 0.112393527 |
| S_VLDL_TG | VAT_Neg_pPS | 4255 | 0.008794191 | [-0.02 0.04] | 0.567455741 | 0.797755947 |
| S_VLDL_TG | Bilirubin_pPS | 4255 | -0.008854095 | [-0.04 0.02] | 0.56481954 | 0.797755947 |
| S_VLDL_TG | Proinsulin_pPS | 4255 | 0.00920041 | [-0.02 0.04] | 0.549695675 | 0.792838637 |
| S_VLDL_TG | Hyper_Insulin_pPS | 4255 | 0.004283024 | [-0.03 0.03] | 0.780643772 | 0.891560895 |
| S_VLDL_TG | Beta_Cell_1_pPS | 4255 | 0.007556576 | [-0.02 0.04] | 0.623193218 | 0.833339789 |
| S_VLDL_TG | Beta_Cell_2_pPS | 4255 | -0.002135145 | [-0.03 0.03] | 0.889587947 | 0.953849648 |
| Serum_C | Cholesterol_pPS | 4254 | -0.135714741 | [-0.17 -0.11] | 7.72E-19 | 5.15E-17 |
| Serum_C | Lipodystrophy_1_pPS | 4254 | 0.033245155 | [0. 0.06] | 0.030624637 | 0.124252113 |
| Serum_C | Lipodystrophy_2_pPS | 4254 | -0.051976309 | [-0.08 -0.02] | 0.000721318 | 0.006976228 |
| Serum_C | Liver-Lipid_pPS | 4254 | -0.001049496 | [-0.03 0.03] | 0.94560304 | 0.978747744 |
| Serum_C | ASAT_Pos_pPS | 4254 | -0.029495285 | [-0.06 0. ] | 0.055117213 | 0.202096449 |
| Serum_C | ALP_Neg_pPS | 4254 | 0.057727076 | [0.03 0.09] | 0.000172611 | 0.002288518 |
| Serum_C | SHBG-LpA_pPS | 4254 | 0.015418761 | [-0.01 0.05] | 0.316122948 | 0.618017395 |
| Serum_C | Obesity_pPS | 4254 | 0.00640931 | [-0.02 0.04] | 0.676910033 | 0.855435982 |
| Serum_C | VAT_Neg_pPS | 4254 | 0.000726391 | [-0.03 0.03] | 0.962334869 | 0.984463162 |
| Serum_C | Bilirubin_pPS | 4254 | -0.012843961 | [-0.04 0.02] | 0.403695133 | 0.677078992 |
| Serum_C | Proinsulin_pPS | 4254 | -0.018197479 | [-0.05 0.01] | 0.236752067 | 0.532559144 |
| Serum_C | Hyper_Insulin_pPS | 4254 | -0.019507043 | [-0.05 0.01] | 0.204689583 | 0.489878376 |
| Serum_C | Beta_Cell_1_pPS | 4254 | 0.017339485 | [-0.01 0.05] | 0.259593977 | 0.555700813 |
| Serum_C | Beta_Cell_2_pPS | 4254 | 0.024652357 | [-0.01 0.05] | 0.108949844 | 0.343027199 |
| Serum_TG | Cholesterol_pPS | 4254 | -0.090381545 | [-0.12 -0.06] | 3.91E-09 | 1.17E-07 |
| Serum_TG | Lipodystrophy_1_pPS | 4254 | 0.062438868 | [0.03 0.09] | 4.84E-05 | 0.000814944 |
| Serum_TG | Lipodystrophy_2_pPS | 4254 | 0.005915915 | [-0.02 0.04] | 0.700529269 | 0.859877129 |
| Serum_TG | Liver-Lipid_pPS | 4254 | -0.0464781 | [-0.08 -0.02] | 0.002500784 | 0.018009245 |
| Serum_TG | ASAT_Pos_pPS | 4254 | 0.024650697 | [-0.01 0.05] | 0.108973676 | 0.343027199 |
| Serum_TG | ALP_Neg_pPS | 4254 | 0.009527788 | [-0.02 0.04] | 0.535632985 | 0.781586915 |
| Serum_TG | SHBG-LpA_pPS | 4254 | -0.007920094 | [-0.04 0.02] | 0.606618993 | 0.824474694 |
| Serum_TG | Obesity_pPS | 4254 | 0.034750197 | [0. 0.06] | 0.023830797 | 0.103941735 |
| Serum_TG | VAT_Neg_pPS | 4254 | 0.0103812 | [-0.02 0.04] | 0.499729143 | 0.755062449 |
| Serum_TG | Bilirubin_pPS | 4254 | -0.010462226 | [-0.04 0.02] | 0.496387946 | 0.753425828 |
| Serum_TG | Proinsulin_pPS | 4254 | 0.007338008 | [-0.02 0.04] | 0.633318236 | 0.835456904 |
| Serum_TG | Hyper_Insulin_pPS | 4254 | 0.003337198 | [-0.03 0.03] | 0.828241893 | 0.920813717 |
| Serum_TG | Beta_Cell_1_pPS | 4254 | 0.006072417 | [-0.02 0.04] | 0.693004656 | 0.858568103 |
| Serum_TG | Beta_Cell_2_pPS | 4254 | -0.001632983 | [-0.03 0.03] | 0.915453198 | 0.966041099 |
| SFA | Cholesterol_pPS | 4223 | -0.109111067 | [-0.14 -0.08] | 1.35E-12 | 5.77E-11 |
| SFA | Lipodystrophy_1_pPS | 4223 | 0.048213995 | [0.02 0.08] | 0.00177946 | 0.013861787 |
| SFA | Lipodystrophy_2_pPS | 4223 | -0.036517664 | [-0.07 -0.01] | 0.017974835 | 0.084078549 |
| SFA | Liver-Lipid_pPS | 4223 | -0.04120278 | [-0.07 -0.01] | 0.007586294 | 0.04364299 |
| SFA | ASAT_Pos_pPS | 4223 | -0.005368804 | [-0.04 0.02] | 0.728024703 | 0.868079485 |
| SFA | ALP_Neg_pPS | 4223 | 0.052535468 | [0.02 0.08] | 0.000661121 | 0.006584896 |
| SFA | SHBG-LpA_pPS | 4223 | 0.012985019 | [-0.02 0.04] | 0.40028634 | 0.675694143 |
| SFA | Obesity_pPS | 4223 | 0.027725252 | [-0. 0.06] | 0.072465239 | 0.248842895 |
| SFA | VAT_Neg_pPS | 4223 | -0.006587474 | [-0.04 0.02] | 0.669602962 | 0.850060323 |
| SFA | Bilirubin_pPS | 4223 | -0.0165661 | [-0.05 0.01] | 0.283224314 | 0.583348844 |
| SFA | Proinsulin_pPS | 4223 | -0.005746633 | [-0.04 0.02] | 0.709723201 | 0.864160597 |
| SFA | Hyper_Insulin_pPS | 4223 | -0.011906825 | [-0.04 0.02] | 0.440550301 | 0.705021345 |
| SFA | Beta_Cell_1_pPS | 4223 | 0.022083258 | [-0.01 0.05] | 0.152554566 | 0.428350969 |
| SFA | Beta_Cell_2_pPS | 4223 | 0.007078904 | [-0.02 0.04] | 0.646575072 | 0.842188221 |
| SM | Cholesterol_pPS | 4223 | -0.104466646 | [-0.13 -0.07] | 1.16E-11 | 4.64E-10 |
| SM | Lipodystrophy_1_pPS | 4223 | 0.007374673 | [-0.02 0.04] | 0.63287525 | 0.835456904 |
| SM | Lipodystrophy_2_pPS | 4223 | -0.053492056 | [-0.08 -0.02] | 0.000525703 | 0.005658368 |
| SM | Liver-Lipid_pPS | 4223 | 0.013720837 | [-0.02 0.04] | 0.374122825 | 0.655035906 |
| SM | ASAT_Pos_pPS | 4223 | -0.030231952 | [-0.06 0. ] | 0.050153529 | 0.186646984 |
| SM | ALP_Neg_pPS | 4223 | 0.062737544 | [0.03 0.09] | 4.75E-05 | 0.000814207 |
| SM | SHBG-LpA_pPS | 4223 | 0.012909405 | [-0.02 0.04] | 0.403035861 | 0.677078992 |
| SM | Obesity_pPS | 4223 | -0.002196505 | [-0.03 0.03] | 0.88686381 | 0.952011446 |
| SM | VAT_Neg_pPS | 4223 | -0.003867432 | [-0.03 0.03] | 0.802196957 | 0.906328459 |
| SM | Bilirubin_pPS | 4223 | -0.006217969 | [-0.04 0.02] | 0.687125962 | 0.858568103 |
| SM | Proinsulin_pPS | 4223 | -0.021385885 | [-0.05 0.01] | 0.165936455 | 0.441605751 |
| SM | Hyper_Insulin_pPS | 4223 | -0.011236543 | [-0.04 0.02] | 0.466709089 | 0.73037006 |
| SM | Beta_Cell_1_pPS | 4223 | 0.025204919 | [-0.01 0.06] | 0.102500641 | 0.330444899 |
| SM | Beta_Cell_2_pPS | 4223 | 0.024110849 | [-0.01 0.05] | 0.118299458 | 0.361580937 |
| TotCho | Cholesterol_pPS | 4223 | -0.088426852 | [-0.12 -0.06] | 9.53E-09 | 2.65E-07 |
| TotCho | Lipodystrophy_1_pPS | 4223 | 0.01574515 | [-0.01 0.05] | 0.307766984 | 0.607642507 |
| TotCho | Lipodystrophy_2_pPS | 4223 | -0.043402382 | [-0.07 -0.01] | 0.004914062 | 0.030270619 |
| TotCho | Liver-Lipid_pPS | 4223 | -0.02285503 | [-0.05 0.01] | 0.138719885 | 0.407209986 |
| TotCho | ASAT_Pos_pPS | 4223 | -0.03107031 | [-0.06 -0. ] | 0.044115346 | 0.166639477 |
| TotCho | ALP_Neg_pPS | 4223 | 0.06650025 | [0.04 0.1 ] | 1.62E-05 | 0.000320435 |
| TotCho | SHBG-LpA_pPS | 4223 | 0.017669519 | [-0.01 0.05] | 0.252378438 | 0.54979503 |
| TotCho | Obesity_pPS | 4223 | 0.006621028 | [-0.02 0.04] | 0.668020427 | 0.849884836 |
| TotCho | VAT_Neg_pPS | 4223 | -0.013725628 | [-0.04 0.02] | 0.373956028 | 0.655035906 |
| TotCho | Bilirubin_pPS | 4223 | -0.005420293 | [-0.04 0.02] | 0.725521202 | 0.866642867 |
| TotCho | Proinsulin_pPS | 4223 | -0.012412878 | [-0.04 0.02] | 0.42137022 | 0.692597028 |
| TotCho | Hyper_Insulin_pPS | 4223 | -0.017769765 | [-0.05 0.01] | 0.249696787 | 0.545779139 |
| TotCho | Beta_Cell_1_pPS | 4223 | 0.016858242 | [-0.01 0.05] | 0.274819433 | 0.573178999 |
| TotCho | Beta_Cell_2_pPS | 4223 | 0.016968462 | [-0.01 0.05] | 0.271693095 | 0.572772495 |
| TotFA | Cholesterol_pPS | 4223 | -0.121906317 | [-0.15 -0.09] | 2.27E-15 | 1.13E-13 |
| TotFA | Lipodystrophy_1_pPS | 4223 | 0.04762159 | [0.02 0.08] | 0.002026445 | 0.015543841 |
| TotFA | Lipodystrophy_2_pPS | 4223 | -0.037666434 | [-0.07 -0.01] | 0.014661998 | 0.072477332 |
| TotFA | Liver-Lipid_pPS | 4223 | -0.044612692 | [-0.07 -0.01] | 0.003838678 | 0.025531672 |
| TotFA | ASAT_Pos_pPS | 4223 | -0.006143452 | [-0.04 0.02] | 0.690680615 | 0.858568103 |
| TotFA | ALP_Neg_pPS | 4223 | 0.052625662 | [0.02 0.08] | 0.000647088 | 0.006509896 |

**Table S9: Associations between pPSs and global metabolomic profile in SCAPIS**

**a) Spearman Correlation**

| Metabolomics | PPS | Samp | r | CI95% | p | fdr_p_adjusted |
|---|---|---|---|---|---|---|
| S_VLDL_PL | ASAT_Pos_pPS | 4255 | 0.035000592 | [0. 0.07] | 0.022613246 | 0.09455246 |
| S_VLDL_PL | ALP_Neg_pPS | 4255 | 0.022735823 | [-0.01 0.05] | 0.138679008 | 0.3817668 |
| S_VLDL_PL | SHBG-LpA_pPS | 4255 | -0.000524342 | [-0.03 0.03] | 0.972761751 | 0.983305352 |
| S_VLDL_PL | Obesity_pPS | 4255 | 0.022691736 | [-0.01 0.05] | 0.139446277 | 0.382986254 |
| S_VLDL_PL | VAT_Neg_pPS | 4255 | 0.009153144 | [-0.02 0.04] | 0.551137046 | 0.760607725 |
| S_VLDL_PL | Bilirubin_pPS | 4255 | -0.004449905 | [-0.03 0.03] | 0.771988095 | 0.891027884 |
| S_VLDL_PL | Proinsulin_pPS | 4255 | 0.013953566 | [-0.02 0.04] | 0.363514745 | 0.639224774 |
| S_VLDL_PL | Hyper_Insulin_pPS | 4255 | 0.0090144 | [-0.02 0.04] | 0.557189104 | 0.763199634 |
| S_VLDL_PL | Beta_Cell_1_pPS | 4255 | 0.018214609 | [-0.01 0.05] | 0.235536324 | 0.518179785 |
| S_VLDL_PL | Beta_Cell_2_pPS | 4255 | 0.001102933 | [-0.03 0.03] | 0.942743545 | 0.973139992 |
| S_VLDL_PL | Total_GRS | 4255 | 0.033321483 | [0. 0.06] | 0.029971381 | 0.118780144 |
| S_VLDL_TG | Cholesterol_pPS | 4255 | -0.077499851 | [-0.11 -0.05] | 4.32E-07 | 1.03E-05 |
| S_VLDL_TG | Lipodystrophy_1_pPS | 4255 | 0.070964981 | [0.04 0.1 ] | 3.71E-06 | 7.73E-05 |
| S_VLDL_TG | Lipodystrophy_2_pPS | 4255 | 0.017710544 | [-0.01 0.05] | 0.248752173 | 0.53033113 |
| S_VLDL_TG | Liver-Lipid_pPS | 4255 | -0.044833398 | [-0.07 -0.01] | 0.00348932 | 0.020906679 |
| S_VLDL_TG | ASAT_Pos_pPS | 4255 | 0.043241478 | [0.01 0.07] | 0.004844967 | 0.026243573 |
| S_VLDL_TG | ALP_Neg_pPS | 4255 | 0.005618839 | [-0.02 0.04] | 0.714442114 | 0.86291356 |
| S_VLDL_TG | SHBG-LpA_pPS | 4255 | -0.005016953 | [-0.04 0.03] | 0.743893581 | 0.876709962 |
| S_VLDL_TG | Obesity_pPS | 4255 | 0.031713058 | [0. 0.06] | 0.038861915 | 0.14497184 |
| S_VLDL_TG | VAT_Neg_pPS | 4255 | 0.015648688 | [-0.01 0.05] | 0.30815966 | 0.594443297 |
| S_VLDL_TG | Bilirubin_pPS | 4255 | -0.005035641 | [-0.04 0.03] | 0.742973225 | 0.876709962 |
| S_VLDL_TG | Proinsulin_pPS | 4255 | 0.014371115 | [-0.02 0.04] | 0.349333592 | 0.629258643 |
| S_VLDL_TG | Hyper_Insulin_pPS | 4255 | 0.014882495 | [-0.02 0.04] | 0.332451066 | 0.613626471 |
| S_VLDL_TG | Beta_Cell_1_pPS | 4255 | 0.013977197 | [-0.02 0.04] | 0.362702662 | 0.639224774 |
| S_VLDL_TG | Beta_Cell_2_pPS | 4255 | 0.000907914 | [-0.03 0.03] | 0.952854455 | 0.976994649 |
| S_VLDL_TG | Total_GRS | 4255 | 0.04434888 | [0.01 0.07] | 0.003860002 | 0.022377579 |
| Serum_C | Cholesterol_pPS | 4254 | -0.137575844 | [-0.17 -0.11] | 2.25E-19 | 1.61E-17 |
| Serum_C | Lipodystrophy_1_pPS | 4254 | 0.025241838 | [-0. 0.06] | 0.100218683 | 0.304058098 |
| Serum_C | Lipodystrophy_2_pPS | 4254 | -0.059085404 | [-0.09 -0.03] | 0.000117877 | 0.001590227 |
| Serum_C | Liver-Lipid_pPS | 4254 | -0.003128088 | [-0.03 0.03] | 0.838607614 | 0.926178923 |
| Serum_C | ASAT_Pos_pPS | 4254 | -0.029023404 | [-0.06 0. ] | 0.058737385 | 0.196862018 |
| Serum_C | ALP_Neg_pPS | 4254 | 0.05435364 | [0.02 0.08] | 0.000397615 | 0.004264416 |
| Serum_C | SHBG-LpA_pPS | 4254 | 0.016473679 | [-0.01 0.05] | 0.283406736 | 0.571763651 |
| Serum_C | Obesity_pPS | 4254 | -0.007461043 | [-0.04 0.02] | 0.627104701 | 0.803929814 |
| Serum_C | VAT_Neg_pPS | 4254 | 0.001811271 | [-0.03 0.03] | 0.906119498 | 0.952872131 |
| Serum_C | Bilirubin_pPS | 4254 | -0.012068435 | [-0.04 0.02] | 0.431971868 | 0.688963257 |
| Serum_C | Proinsulin_pPS | 4254 | -0.001537248 | [-0.03 0.03] | 0.920270914 | 0.960199976 |
| Serum_C | Hyper_Insulin_pPS | 4254 | -0.021392914 | [-0.05 0.01] | 0.163594712 | 0.423805142 |
| Serum_C | Beta_Cell_1_pPS | 4254 | 0.022072621 | [-0.01 0.05] | 0.150617055 | 0.40083571 |
| Serum_C | Beta_Cell_2_pPS | 4254 | 0.025732914 | [-0. 0.06] | 0.093780383 | 0.288606775 |
| Serum_C | Total_GRS | 4254 | -0.009956738 | [-0.04 0.02] | 0.516782109 | 0.736054199 |
| Serum_TG | Cholesterol_pPS | 4254 | -0.088232746 | [-0.12 -0.06] | 8.60E-09 | 2.64E-07 |
| Serum_TG | Lipodystrophy_1_pPS | 4254 | 0.069359198 | [0.04 0.1 ] | 6.14E-06 | 0.000116544 |
| Serum_TG | Lipodystrophy_2_pPS | 4254 | 0.008606302 | [-0.02 0.04] | 0.575220863 | 0.77308819 |
| Serum_TG | Liver-Lipid_pPS | 4254 | -0.04577601 | [-0.08 -0.02] | 0.002862735 | 0.018368853 |
| Serum_TG | ASAT_Pos_pPS | 4254 | 0.038712226 | [0.01 0.07] | 0.011683773 | 0.065665296 |
| Serum_TG | ALP_Neg_pPS | 4254 | 0.011034591 | [-0.02 0.04] | 0.472449034 | 0.710023653 |
| Serum_TG | SHBG-LpA_pPS | 4254 | -0.001413148 | [-0.03 0.03] | 0.92668839 | 0.964457349 |
| Serum_TG | Obesity_pPS | 4254 | 0.030960639 | [0. 0.06] | 0.043760752 | 0.158291421 |
| Serum_TG | VAT_Neg_pPS | 4254 | 0.016329217 | [-0.01 0.05] | 0.287650556 | 0.57504794 |
| Serum_TG | Bilirubin_pPS | 4254 | -0.006859188 | [-0.04 0.02] | 0.655151697 | 0.821454552 |
| Serum_TG | Proinsulin_pPS | 4254 | 0.012998246 | [-0.02 0.04] | 0.397346203 | 0.665345516 |
| Serum_TG | Hyper_Insulin_pPS | 4254 | 0.013119858 | [-0.02 0.04] | 0.392944506 | 0.660507155 |
| Serum_TG | Beta_Cell_1_pPS | 4254 | 0.012279811 | [-0.02 0.04] | 0.423950495 | 0.684769436 |
| Serum_TG | Beta_Cell_2_pPS | 4254 | 0.000666744 | [-0.03 0.03] | 0.965372577 | 0.982019059 |
| Serum_TG | Total_GRS | 4254 | 0.03964044 | [0.01 0.07] | 0.009821219 | 0.046607336 |
| SFA | Cholesterol_pPS | 4223 | -0.108923355 | [-0.14 -0.08] | 1.37E-12 | 6.38E-11 |
| SFA | Lipodystrophy_1_pPS | 4223 | 0.046769427 | [0.02 0.08] | 0.002399525 | 0.016084061 |
| SFA | Lipodystrophy_2_pPS | 4223 | -0.041244319 | [-0.07 -0.01] | 0.007433349 | 0.037516548 |
| SFA | Liver-Lipid_pPS | 4223 | -0.041205555 | [-0.07 -0.01] | 0.007489396 | 0.037534473 |
| SFA | ASAT_Pos_pPS | 4223 | 0.000255955 | [-0.03 0.03] | 0.98675208 | 0.991373869 |
| SFA | ALP_Neg_pPS | 4223 | 0.050521665 | [0.02 0.08] | 0.001039603 | 0.008744895 |
| SFA | SHBG-LpA_pPS | 4223 | 0.014963179 | [-0.02 0.05] | 0.331667827 | 0.61329956 |
| SFA | Obesity_pPS | 4223 | 0.018984893 | [-0.01 0.05] | 0.218054783 | 0.498643401 |
| SFA | VAT_Neg_pPS | 4223 | -0.004169396 | [-0.03 0.03] | 0.786789281 | 0.898649099 |
| SFA | Bilirubin_pPS | 4223 | -0.013895919 | [-0.04 0.02] | 0.36731571 | 0.641606025 |
| SFA | Proinsulin_pPS | 4223 | 0.011152995 | [-0.02 0.04] | 0.469341899 | 0.707973539 |
| SFA | Hyper_Insulin_pPS | 4223 | -0.009772628 | [-0.04 0.02] | 0.526084481 | 0.742402112 |
| SFA | Beta_Cell_1_pPS | 4223 | 0.025278835 | [-0. 0.06] | 0.10096883 | 0.305231058 |
| SFA | Beta_Cell_2_pPS | 4223 | 0.010050552 | [-0.02 0.04] | 0.51438475 | 0.733381664 |
| SFA | Total_GRS | 4223 | 0.010502191 | [-0.02 0.04] | 0.495664631 | 0.722774054 |
| SM | Cholesterol_pPS | 4223 | -0.106498665 | [-0.14 -0.08] | 4.25E-12 | 1.82E-10 |
| SM | Lipodystrophy_1_pPS | 4223 | -9.88E-05 | [-0.03 0.03] | 0.994888247 | 0.996787043 |
| SM | Lipodystrophy_2_pPS | 4223 | -0.060793553 | [-0.09 -0.03] | 7.90E-05 | 0.001168191 |
| SM | Liver-Lipid_pPS | 4223 | 0.012366937 | [-0.02 0.04] | 0.422373427 | 0.683766793 |
| SM | ASAT_Pos_pPS | 4223 | -0.034236782 | [-0.06 -0. ] | 0.026304097 | 0.106658389 |
| SM | ALP_Neg_pPS | 4223 | 0.059174918 | [0.03 0.09] | 0.000121927 | 0.001604501 |
| SM | SHBG-LpA_pPS | 4223 | 0.011333103 | [-0.02 0.04] | 0.462196122 | 0.703128142 |
| SM | Obesity_pPS | 4223 | -0.013919796 | [-0.04 0.02] | 0.366492995 | 0.64109339 |
| SM | VAT_Neg_pPS | 4223 | -0.003042661 | [-0.03 0.03] | 0.843524822 | 0.92810697 |
| SM | Bilirubin_pPS | 4223 | -0.006049821 | [-0.04 0.02] | 0.694708483 | 0.849088146 |
| SM | Proinsulin_pPS | 4223 | -0.001986282 | [-0.03 0.03] | 0.897471489 | 0.949937874 |
| SM | Hyper_Insulin_pPS | 4223 | -0.01607923 | [-0.05 0.01] | 0.296867449 | 0.583132489 |
| SM | Beta_Cell_1_pPS | 4223 | 0.028506886 | [-0. 0.06] | 0.064356545 | 0.211077659 |
| SM | Beta_Cell_2_pPS | 4223 | 0.027101044 | [-0. 0.06] | 0.078671365 | 0.249261563 |
| SM | Total_GRS | 4223 | -0.015953767 | [-0.05 0.01] | 0.30065311 | 0.586807026 |
| TotCho | Cholesterol_pPS | 4223 | -0.089781723 | [-0.12 -0.06] | 5.34E-09 | 1.71E-07 |
| TotCho | Lipodystrophy_1_pPS | 4223 | 0.007984959 | [-0.02 0.04] | 0.604447024 | 0.791565213 |
| TotCho | Lipodystrophy_2_pPS | 4223 | -0.052279112 | [-0.08 -0.02] | 0.000689247 | 0.006600154 |
| TotCho | Liver-Lipid_pPS | 4223 | -0.023581037 | [-0.05 0.01] | 0.126020625 | 0.358506951 |
| TotCho | ASAT_Pos_pPS | 4223 | -0.033711214 | [-0.06 -0. ] | 0.028701105 | 0.115288145 |
| TotCho | ALP_Neg_pPS | 4223 | 0.063194405 | [0.03 0.09] | 4.07E-05 | 0.000636576 |
| TotCho | SHBG-LpA_pPS | 4223 | 0.015707782 | [-0.01 0.05] | 0.308168273 | 0.594443297 |
| TotCho | Obesity_pPS | 4223 | -0.003149294 | [-0.03 0.03] | 0.838115546 | 0.926178923 |

**b) Partial Correlation**

| Metabolomics | PPS | Samp | r | CI95% | p | fdr_p_adjusted |
|---|---|---|---|---|---|---|
| TotFA | SHBG-LpA_pPS | 4223 | 0.010614804 | [-0.02 0.04] | 0.491724451 | 0.751475077 |
| TotFA | Obesity_pPS | 4223 | 0.030540767 | [0. 0.06] | 0.047853305 | 0.179405089 |
| TotFA | VAT_Neg_pPS | 4223 | -0.005291084 | [-0.04 0.02] | 0.731809072 | 0.869076195 |
| TotFA | Bilirubin_pPS | 4223 | -0.019424502 | [-0.05 0.01] | 0.208284806 | 0.495232994 |
| TotFA | Proinsulin_pPS | 4223 | -0.006036899 | [-0.04 0.02] | 0.695775274 | 0.858892964 |
| TotFA | Hyper_Insulin_pPS | 4223 | -0.013040189 | [-0.04 0.02] | 0.398287341 | 0.673455453 |
| TotFA | Beta_Cell_1_pPS | 4223 | 0.025922593 | [-0. 0.06] | 0.093082294 | 0.307509494 |
| TotFA | Beta_Cell_2_pPS | 4223 | 0.009651958 | [-0.02 0.04] | 0.531841274 | 0.781178452 |
| TotPG | Cholesterol_pPS | 4223 | -0.085577603 | [-0.12 -0.06] | 2.80E-08 | 7.29E-07 |
| TotPG | Lipodystrophy_1_pPS | 4223 | 0.019778391 | [-0.01 0.05] | 0.200114744 | 0.486527589 |
| TotPG | Lipodystrophy_2_pPS | 4223 | -0.043702463 | [-0.07 -0.01] | 0.004624647 | 0.029166809 |
| TotPG | Liver-Lipid_pPS | 4223 | -0.043125486 | [-0.07 -0.01] | 0.005195538 | 0.031519599 |
| TotPG | ASAT_Pos_pPS | 4223 | -0.026705994 | [-0.06 0. ] | 0.083607519 | 0.281314712 |
| TotPG | ALP_Neg_pPS | 4223 | 0.060343641 | [0.03 0.09] | 9.15E-05 | 0.001356391 |
| TotPG | SHBG-LpA_pPS | 4223 | 0.016501763 | [-0.01 0.05] | 0.285098436 | 0.583442529 |
| TotPG | Obesity_pPS | 4223 | 0.009235 | [-0.02 0.04] | 0.549714177 | 0.792838637 |
| TotPG | VAT_Neg_pPS | 4223 | -0.017395596 | [-0.05 0.01] | 0.259808172 | 0.555700813 |
| TotPG | Bilirubin_pPS | 4223 | -0.012418577 | [-0.04 0.02] | 0.421157049 | 0.692597028 |
| TotPG | Proinsulin_pPS | 4223 | -0.007508596 | [-0.04 0.02] | 0.626712855 | 0.834676462 |
| TotPG | Hyper_Insulin_pPS | 4223 | -0.014021011 | [-0.04 0.02] | 0.363760351 | 0.650367164 |
| TotPG | Beta_Cell_1_pPS | 4223 | 0.021946007 | [-0.01 0.05] | 0.155121226 | 0.42902716 |
| TotPG | Beta_Cell_2_pPS | 4223 | 0.009850911 | [-0.02 0.04] | 0.523418094 | 0.774477051 |
| Tyr | Cholesterol_pPS | 4250 | 0.025067402 | [-0.01 0.06] | 0.103280221 | 0.33188925 |
| Tyr | Lipodystrophy_1_pPS | 4250 | 0.00925816 | [-0.02 0.04] | 0.547430299 | 0.79130358 |
| Tyr | Lipodystrophy_2_pPS | 4250 | 0.016807306 | [-0.01 0.05] | 0.274732571 | 0.573178999 |
| Tyr | Liver-Lipid_pPS | 4250 | 0.03370008 | [0. 0.06] | 0.028489503 | 0.119017391 |
| Tyr | ASAT_Pos_pPS | 4250 | 0.002051982 | [-0.03 0.03] | 0.89392489 | 0.955492594 |
| Tyr | ALP_Neg_pPS | 4250 | -0.011844157 | [-0.04 0.02] | 0.441496047 | 0.705970516 |
| Tyr | SHBG-LpA_pPS | 4250 | 0.004937599 | [-0.03 0.04] | 0.748323347 | 0.875595172 |
| Tyr | Obesity_pPS | 4250 | 0.029152662 | [-0. 0.06] | 0.058123542 | 0.211498441 |
| Tyr | VAT_Neg_pPS | 4250 | 0.014354876 | [-0.02 0.04] | 0.35090314 | 0.637861136 |
| Tyr | Bilirubin_pPS | 4250 | -0.015357941 | [-0.05 0.01] | 0.318264977 | 0.61940234 |
| Tyr | Proinsulin_pPS | 4250 | 0.007742568 | [-0.02 0.04] | 0.614876012 | 0.828794354 |
| Tyr | Hyper_Insulin_pPS | 4250 | 0.013403362 | [-0.02 0.04] | 0.383754214 | 0.66173638 |
| Tyr | Beta_Cell_1_pPS | 4250 | -0.002398461 | [-0.03 0.03] | 0.87614807 | 0.946048963 |
| Tyr | Beta_Cell_2_pPS | 4250 | 0.008008675 | [-0.02 0.04] | 0.602772152 | 0.823046075 |
| UnSat | Cholesterol_pPS | 4223 | 0.04549899 | [0.02 0.08] | 0.003191978 | 0.021959928 |
| UnSat | Lipodystrophy_1_pPS | 4223 | -0.02477769 | [-0.05 0.01] | 0.108457758 | 0.342480177 |
| UnSat | Lipodystrophy_2_pPS | 4223 | -0.013854479 | [-0.04 0.02] | 0.369486963 | 0.653351391 |
| UnSat | Liver-Lipid_pPS | 4223 | 0.08375595 | [0.05 0.11] | 5.49E-08 | 1.34E-06 |
| UnSat | ASAT_Pos_pPS | 4223 | -0.028162385 | [-0.06 0. ] | 0.068074253 | 0.237844075 |
| UnSat | ALP_Neg_pPS | 4223 | 0.045783999 | [0.02 0.08] | 0.003006152 | 0.02089693 |
| UnSat | SHBG-LpA_pPS | 4223 | -0.002082544 | [-0.03 0.03] | 0.89269721 | 0.954690071 |
| UnSat | Obesity_pPS | 4223 | -0.034674559 | [-0.06 -0. ] | 0.024663067 | 0.106642464 |
| UnSat | VAT_Neg_pPS | 4223 | -0.001749797 | [-0.03 0.03] | 0.909761684 | 0.963163877 |
| UnSat | Bilirubin_pPS | 4223 | -0.028646497 | [-0.06 0. ] | 0.063469061 | 0.226094412 |
| UnSat | Proinsulin_pPS | 4223 | -0.025281128 | [-0.06 0. ] | 0.101465888 | 0.328696939 |
| UnSat | Hyper_Insulin_pPS | 4223 | -0.00281935 | [-0.03 0.03] | 0.855098357 | 0.93700433 |
| UnSat | Beta_Cell_1_pPS | 4223 | 0.011822626 | [-0.02 0.04] | 0.443789388 | 0.706833249 |
| UnSat | Beta_Cell_2_pPS | 4223 | 0.033227342 | [0. 0.06] | 0.031332955 | 0.126468902 |
| Val | Cholesterol_pPS | 4254 | -0.000811961 | [-0.03 0.03] | 0.957901742 | 0.981432594 |
| Val | Lipodystrophy_1_pPS | 4254 | 0.002901362 | [-0.03 0.03] | 0.850388767 | 0.933888267 |
| Val | Lipodystrophy_2_pPS | 4254 | 0.016712971 | [-0.01 0.05] | 0.277208402 | 0.57509971 |
| Val | Liver-Lipid_pPS | 4254 | 0.001200973 | [-0.03 0.03] | 0.937766698 | 0.978170683 |
| Val | ASAT_Pos_pPS | 4254 | -0.009879618 | [-0.04 0.02] | 0.52067526 | 0.773759954 |
| Val | ALP_Neg_pPS | 4254 | 0.000117305 | [-0.03 0.03] | 0.993915271 | 0.995905092 |
| Val | SHBG-LpA_pPS | 4254 | 0.004692622 | [-0.03 0.03] | 0.760308087 | 0.883422397 |
| Val | Obesity_pPS | 4254 | 0.020457364 | [-0.01 0.05] | 0.183484053 | 0.464981107 |
| Val | VAT_Neg_pPS | 4254 | -0.002461976 | [-0.03 0.03] | 0.872836089 | 0.946048963 |
| Val | Bilirubin_pPS | 4254 | -0.009464318 | [-0.04 0.02] | 0.538354191 | 0.784375661 |
| Val | Proinsulin_pPS | 4254 | -0.007921954 | [-0.04 0.02] | 0.606534507 | 0.824474694 |
| Val | Hyper_Insulin_pPS | 4254 | 0.003165973 | [-0.03 0.03] | 0.836927382 | 0.92461824 |
| Val | Beta_Cell_1_pPS | 4254 | 0.027184568 | [-0. 0.06] | 0.077121568 | 0.262580577 |
| Val | Beta_Cell_2_pPS | 4254 | 8.84E-05 | [-0.03 0.03] | 0.995415361 | 0.996223326 |
| VLDL_C | Cholesterol_pPS | 4254 | -0.117253308 | [-0.15 -0.09] | 2.02E-14 | 9.64E-13 |
| VLDL_C | Lipodystrophy_1_pPS | 4254 | 0.063024583 | [0.03 0.09] | 4.11E-05 | 0.000734043 |
| VLDL_C | Lipodystrophy_2_pPS | 4254 | -0.019590303 | [-0.05 0.01] | 0.202763398 | 0.487899426 |
| VLDL_C | Liver-Lipid_pPS | 4254 | -0.021330004 | [-0.05 0.01] | 0.16548625 | 0.441605751 |
| VLDL_C | ASAT_Pos_pPS | 4254 | 0.016886764 | [-0.01 0.05] | 0.272211294 | 0.572772495 |
| VLDL_C | ALP_Neg_pPS | 4254 | 0.033515821 | [0. 0.06] | 0.029292515 | 0.121164494 |
| VLDL_C | SHBG-LpA_pPS | 4254 | 0.001794184 | [-0.03 0.03] | 0.907143131 | 0.96224905 |
| VLDL_C | Obesity_pPS | 4254 | 0.024291455 | [-0.01 0.05] | 0.114231072 | 0.352373815 |
| VLDL_C | VAT_Neg_pPS | 4254 | 0.011036961 | [-0.02 0.04] | 0.473033768 | 0.734922314 |
| VLDL_C | Bilirubin_pPS | 4254 | -0.022283305 | [-0.05 0.01] | 0.147378973 | 0.421503863 |
| VLDL_C | Proinsulin_pPS | 4254 | 0.000532179 | [-0.03 0.03] | 0.972400468 | 0.989702967 |
| VLDL_C | Hyper_Insulin_pPS | 4254 | -0.00587131 | [-0.04 0.02] | 0.702679291 | 0.859880159 |
| VLDL_C | Beta_Cell_1_pPS | 4254 | 0.012126465 | [-0.02 0.04] | 0.43046912 | 0.696591166 |
| VLDL_C | Beta_Cell_2_pPS | 4254 | 0.007696768 | [-0.02 0.04] | 0.616805977 | 0.828794354 |
| VLDL_D | Cholesterol_pPS | 4254 | -0.045951039 | [-0.08 -0.02] | 0.002799595 | 0.019875137 |
| VLDL_D | Lipodystrophy_1_pPS | 4254 | 0.049059513 | [0.02 0.08] | 0.001416067 | 0.01154043 |
| VLDL_D | Lipodystrophy_2_pPS | 4254 | 0.016872524 | [-0.01 0.05] | 0.272647162 | 0.572772495 |
| VLDL_D | Liver-Lipid_pPS | 4254 | -0.068553061 | [-0.1 -0.04] | 8.12E-06 | 0.000165885 |
| VLDL_D | ASAT_Pos_pPS | 4254 | 0.035095262 | [0. 0.07] | 0.022471638 | 0.099894945 |
| VLDL_D | ALP_Neg_pPS | 4254 | -0.016795205 | [-0.05 0.01] | 0.274851068 | 0.573178999 |
| VLDL_D | SHBG-LpA_pPS | 4254 | -0.016181353 | [-0.05 0.01] | 0.292779594 | 0.589592798 |
| VLDL_D | Obesity_pPS | 4254 | 0.038973152 | [0.01 0.07] | 0.011255412 | 0.058987788 |
| VLDL_D | VAT_Neg_pPS | 4254 | 0.017477559 | [-0.01 0.05] | 0.255818576 | 0.55296819 |
| VLDL_D | Bilirubin_pPS | 4254 | -0.006088655 | [-0.04 0.02] | 0.692225634 | 0.858568103 |
| VLDL_D | Proinsulin_pPS | 4254 | 0.009923457 | [-0.02 0.04] | 0.518826699 | 0.772830501 |
| VLDL_D | Hyper_Insulin_pPS | 4254 | 0.011039156 | [-0.02 0.04] | 0.47294575 | 0.734922314 |
| VLDL_D | Beta_Cell_1_pPS | 4254 | 0.003599776 | [-0.03 0.03] | 0.814963437 | 0.912503804 |
| VLDL_D | Beta_Cell_2_pPS | 4254 | -0.008750515 | [-0.04 0.02] | 0.569427197 | 0.797755947 |
| VLDL_TG | Cholesterol_pPS | 4254 | -0.083039824 | [-0.11 -0.05] | 6.38E-08 | 1.54E-06 |
| VLDL_TG | Lipodystrophy_1_pPS | 4254 | 0.060670928 | [0.03 0.09] | 7.88E-05 | 0.001195499 |

**Table S9: Associations between pPSs and global metabolomic profile in SCAPIS**

**a) Spearman Correlation**

| Metabolomics | PPS | Samp | r | CI95% | p | fdr_p_adjusted |
|---|---|---|---|---|---|---|
| TotCho | VAT_Neg_pPS | 4223 | -0.014218674 | [-0.04 0.02] | 0.356291799 | 0.635279317 |
| TotCho | Bilirubin_pPS | 4223 | -0.00491853 | [-0.04 0.03] | 0.749664695 | 0.880148205 |
| TotCho | Proinsulin_pPS | 4223 | 0.004585672 | [-0.03 0.03] | 0.766094235 | 0.889697961 |
| TotCho | Hyper_Insulin_pPS | 4223 | -0.022715594 | [-0.05 0.01] | 0.140530648 | 0.385470894 |
| TotCho | Beta_Cell_1_pPS | 4223 | 0.018103296 | [-0.01 0.05] | 0.240190606 | 0.523054671 |
| TotCho | Beta_Cell_2_pPS | 4223 | 0.017729772 | [-0.01 0.05] | 0.250031421 | 0.531533596 |
| TotCho | Total_GRS | 4223 | -0.015498589 | [-0.05 0.01] | 0.314656331 | 0.60298573 |
| TotFA | Cholesterol_pPS | 4223 | -0.121626714 | [-0.15 -0.09] | 2.38E-15 | 1.28E-13 |
| TotFA | Lipodystrophy_1_pPS | 4223 | 0.045680895 | [0.02 0.08] | 0.003026807 | 0.019095589 |
| TotFA | Lipodystrophy_2_pPS | 4223 | -0.042985931 | [-0.07 -0.01] | 0.005272208 | 0.028201712 |
| TotFA | Liver-Lipid_pPS | 4223 | -0.044660569 | [-0.07 -0.01] | 0.003747022 | 0.021836135 |
| TotFA | ASAT_Pos_pPS | 4223 | -0.000623066 | [-0.03 0.03] | 0.967758176 | 0.98241424 |
| TotFA | ALP_Neg_pPS | 4223 | 0.050376734 | [0.02 0.08] | 0.001074852 | 0.00893627 |
| TotFA | SHBG-LpA_pPS | 4223 | 0.012800351 | [-0.02 0.04] | 0.406295383 | 0.67066604 |
| TotFA | Obesity_pPS | 4223 | 0.020457011 | [-0.01 0.05] | 0.184427459 | 0.455303634 |
| TotFA | VAT_Neg_pPS | 4223 | -0.002274715 | [-0.03 0.03] | 0.882683985 | 0.945095197 |
| TotFA | Bilirubin_pPS | 4223 | -0.016410416 | [-0.05 0.01] | 0.287028079 | 0.57504794 |
| TotFA | Proinsulin_pPS | 4223 | 0.012433991 | [-0.02 0.04] | 0.419861887 | 0.680728456 |
| TotFA | Hyper_Insulin_pPS | 4223 | -0.011143847 | [-0.04 0.02] | 0.46970648 | 0.708025579 |
| TotFA | Beta_Cell_1_pPS | 4223 | 0.03005529 | [-0. 0.06] | 0.051150383 | 0.177332222 |
| TotFA | Beta_Cell_2_pPS | 4223 | 0.013527031 | [-0.02 0.04] | 0.380172456 | 0.651854451 |
| TotFA | Total_GRS | 4223 | 0.010156691 | [-0.02 0.04] | 0.5099526 | 0.7313594 |
| TotPG | Cholesterol_pPS | 4223 | -0.087104679 | [-0.12 -0.06] | 1.50E-08 | 4.35E-07 |
| TotPG | Lipodystrophy_1_pPS | 4223 | 0.012871484 | [-0.02 0.04] | 0.403691938 | 0.668142396 |
| TotPG | Lipodystrophy_2_pPS | 4223 | -0.052853876 | [-0.08 -0.02] | 0.00060095 | 0.005958484 |
| TotPG | Liver-Lipid_pPS | 4223 | -0.043558815 | [-0.07 -0.01] | 0.004696714 | 0.025700132 |
| TotPG | ASAT_Pos_pPS | 4223 | -0.028307747 | [-0.06 0. ] | 0.066243085 | 0.215777718 |
| TotPG | ALP_Neg_pPS | 4223 | 0.0571393 | [0.03 0.09] | 0.000207402 | 0.002496149 |
| TotPG | SHBG-LpA_pPS | 4223 | 0.014977543 | [-0.02 0.05] | 0.33120382 | 0.612969968 |
| TotPG | Obesity_pPS | 4223 | 0.000463774 | [-0.03 0.03] | 0.97599813 | 0.985183995 |
| TotPG | VAT_Neg_pPS | 4223 | -0.017772554 | [-0.05 0.01] | 0.248890223 | 0.53033113 |
| TotPG | Bilirubin_pPS | 4223 | -0.011218574 | [-0.04 0.02] | 0.466733015 | 0.706538163 |
| TotPG | Proinsulin_pPS | 4223 | 0.008659217 | [-0.02 0.04] | 0.574279882 | 0.772791937 |
| TotPG | Hyper_Insulin_pPS | 4223 | -0.019282294 | [-0.05 0.01] | 0.210928758 | 0.490238819 |
| TotPG | Beta_Cell_1_pPS | 4223 | 0.022106517 | [-0.01 0.05] | 0.151489429 | 0.401178951 |
| TotPG | Beta_Cell_2_pPS | 4223 | 0.011715142 | [-0.02 0.04] | 0.447241468 | 0.693159645 |
| TotPG | Total_GRS | 4223 | -0.011503437 | [-0.04 0.02] | 0.455494305 | 0.696320363 |
| Tyr | Cholesterol_pPS | 4250 | 0.029348973 | [-0. 0.06] | 0.056073521 | 0.189712465 |
| Tyr | Lipodystrophy_1_pPS | 4250 | 0.014370132 | [-0.02 0.04] | 0.349650819 | 0.629258643 |
| Tyr | Lipodystrophy_2_pPS | 4250 | 0.022551489 | [-0.01 0.05] | 0.142143943 | 0.389398158 |
| Tyr | Liver-Lipid_pPS | 4250 | 0.036282538 | [0.01 0.07] | 0.018173144 | 0.078750289 |
| Tyr | ASAT_Pos_pPS | 4250 | 0.007516789 | [-0.02 0.04] | 0.624696595 | 0.803823753 |
| Tyr | ALP_Neg_pPS | 4250 | -0.007929536 | [-0.04 0.02] | 0.605806914 | 0.792521775 |
| Tyr | SHBG-LpA_pPS | 4250 | 0.00790612 | [-0.02 0.04] | 0.606871704 | 0.792776982 |
| Tyr | Obesity_pPS | 4250 | 0.03432726 | [0. 0.06] | 0.025436965 | 0.103928172 |
| Tyr | VAT_Neg_pPS | 4250 | 0.018789602 | [-0.01 0.05] | 0.221349042 | 0.502934916 |
| Tyr | Bilirubin_pPS | 4250 | -0.015124549 | [-0.05 0.01] | 0.324932221 | 0.608687153 |
| Tyr | Proinsulin_pPS | 4250 | 0.012683072 | [-0.02 0.04] | 0.409112379 | 0.67193419 |
| Tyr | Hyper_Insulin_pPS | 4250 | 0.021150448 | [-0.01 0.05] | 0.168623472 | 0.431105302 |
| Tyr | Beta_Cell_1_pPS | 4250 | 0.004463647 | [-0.03 0.03] | 0.77143469 | 0.891027884 |
| Tyr | Beta_Cell_2_pPS | 4250 | 0.012369584 | [-0.02 0.04] | 0.420789552 | 0.681717212 |
| Tyr | Total_GRS | 4250 | 0.05150544 | [0.02 0.08] | 0.00079584 | 0.007255522 |
| UnSat | Cholesterol_pPS | 4223 | 0.043842127 | [0.01 0.07] | 0.004433669 | 0.024873164 |
| UnSat | Lipodystrophy_1_pPS | 4223 | -0.031165716 | [-0.06 -0. ] | 0.043145031 | 0.157124096 |
| UnSat | Lipodystrophy_2_pPS | 4223 | -0.014131702 | [-0.04 0.02] | 0.359241685 | 0.63736428 |
| UnSat | Liver-Lipid_pPS | 4223 | 0.086301326 | [0.06 0.12] | 2.03E-08 | 5.82E-07 |
| UnSat | ASAT_Pos_pPS | 4223 | -0.036215327 | [-0.07 -0.01] | 0.018765036 | 0.080825307 |
| UnSat | ALP_Neg_pPS | 4223 | 0.044806958 | [0.01 0.07] | 0.003634923 | 0.021420079 |
| UnSat | SHBG-LpA_pPS | 4223 | -0.005785859 | [-0.04 0.02] | 0.707401023 | 0.858243888 |
| UnSat | Obesity_pPS | 4223 | -0.033460976 | [-0.06 -0. ] | 0.029906966 | 0.118780144 |
| UnSat | VAT_Neg_pPS | 4223 | -0.002038715 | [-0.03 0.03] | 0.894780568 | 0.948580863 |
| UnSat | Bilirubin_pPS | 4223 | -0.030454465 | [-0.06 -0. ] | 0.048140124 | 0.169279617 |
| UnSat | Proinsulin_pPS | 4223 | -0.011907866 | [-0.04 0.02] | 0.439803065 | 0.691142319 |
| UnSat | Hyper_Insulin_pPS | 4223 | -0.005203137 | [-0.04 0.03] | 0.735705979 | 0.873878512 |
| UnSat | Beta_Cell_1_pPS | 4223 | 0.012492615 | [-0.02 0.04] | 0.417673306 | 0.678719122 |
| UnSat | Beta_Cell_2_pPS | 4223 | 0.034875951 | [0. 0.07] | 0.023623707 | 0.097824039 |
| UnSat | Total_GRS | 4223 | -0.021516957 | [-0.05 0.01] | 0.162705416 | 0.422011025 |
| Val | Cholesterol_pPS | 4254 | 0.001423823 | [-0.03 0.03] | 0.926136149 | 0.964350504 |
| Val | Lipodystrophy_1_pPS | 4254 | 0.004368787 | [-0.03 0.03] | 0.776058338 | 0.89275053 |
| Val | Lipodystrophy_2_pPS | 4254 | 0.015388721 | [-0.01 0.05] | 0.31632371 | 0.604198004 |
| Val | Liver-Lipid_pPS | 4254 | 0.0028026 | [-0.03 0.03] | 0.855204716 | 0.933069234 |
| Val | ASAT_Pos_pPS | 4254 | -0.006202271 | [-0.04 0.02] | 0.686328389 | 0.842205031 |
| Val | ALP_Neg_pPS | 4254 | -0.000650085 | [-0.03 0.03] | 0.966237268 | 0.982019059 |
| Val | SHBG-LpA_pPS | 4254 | 0.004849243 | [-0.03 0.03] | 0.752197518 | 0.881192614 |
| Val | Obesity_pPS | 4254 | 0.019932813 | [-0.01 0.05] | 0.194293117 | 0.466695113 |
| Val | VAT_Neg_pPS | 4254 | 0.003294144 | [-0.03 0.03] | 0.830167271 | 0.922611229 |
| Val | Bilirubin_pPS | 4254 | -0.008633271 | [-0.04 0.02] | 0.574023898 | 0.772791937 |
| Val | Proinsulin_pPS | 4254 | 0.002688103 | [-0.03 0.03] | 0.861058723 | 0.93707304 |
| Val | Hyper_Insulin_pPS | 4254 | 0.005260985 | [-0.02 0.04] | 0.731934972 | 0.872537948 |
| Val | Beta_Cell_1_pPS | 4254 | 0.026076493 | [-0. 0.06] | 0.08947628 | 0.276153412 |
| Val | Beta_Cell_2_pPS | 4254 | 0.006812612 | [-0.02 0.04] | 0.657343251 | 0.823091532 |
| Val | Total_GRS | 4254 | 0.03116921 | [0. 0.06] | 0.04236005 | 0.154790985 |
| VLDL_C | Cholesterol_pPS | 4254 | -0.116828527 | [-0.15 -0.09] | 2.29E-14 | 1.17E-12 |
| VLDL_C | Lipodystrophy_1_pPS | 4254 | 0.06661252 | [0.04 0.1 ] | 1.41E-05 | 0.000244171 |
| VLDL_C | Lipodystrophy_2_pPS | 4254 | -0.018406302 | [-0.05 0.01] | 0.230698133 | 0.513860327 |
| VLDL_C | Liver-Lipid_pPS | 4254 | -0.021158407 | [-0.05 0.01] | 0.168262375 | 0.430695458 |
| VLDL_C | ASAT_Pos_pPS | 4254 | 0.027613227 | [-0. 0.06] | 0.072132661 | 0.23197085 |
| VLDL_C | ALP_Neg_pPS | 4254 | 0.034253185 | [0. 0.06] | 0.025685836 | 0.104348709 |
| VLDL_C | SHBG-LpA_pPS | 4254 | 0.007665344 | [-0.02 0.04] | 0.617702625 | 0.799138801 |
| VLDL_C | Obesity_pPS | 4254 | 0.017227993 | [-0.01 0.05] | 0.261937829 | 0.548688129 |
| VLDL_C | VAT_Neg_pPS | 4254 | 0.015879415 | [-0.01 0.05] | 0.301137665 | 0.587218446 |
| VLDL_C | Bilirubin_pPS | 4254 | -0.019020356 | [-0.05 0.01] | 0.215510392 | 0.496093758 |
| VLDL_C | Proinsulin_pPS | 4254 | 0.012118038 | [-0.02 0.04] | 0.43008165 | 0.688963257 |
| VLDL_C | Hyper_Insulin_pPS | 4254 | 0.0025021 | [-0.03 0.03] | 0.870584884 | 0.940757973 |

**b) Partial Correlation**

| Metabolomics | PPS | Samp | r | CI95% | p | fdr_p_adjusted |
|---|---|---|---|---|---|---|
| VLDL_TG | Lipodystrophy_2_pPS | 4254 | 0.010283686 | [-0.02 0.04] | 0.50376603 | 0.760014764 |
| VLDL_TG | Liver-Lipid_pPS | 4254 | -0.050022491 | [-0.08 -0.02] | 0.001137617 | 0.009609748 |
| VLDL_TG | ASAT_Pos_pPS | 4254 | 0.030712004 | [0. 0.06] | 0.045813703 | 0.172080737 |
| VLDL_TG | ALP_Neg_pPS | 4254 | 0.000381369 | [-0.03 0.03] | 0.980219747 | 0.992796048 |
| VLDL_TG | SHBG-LpA_pPS | 4254 | -0.009853784 | [-0.04 0.02] | 0.521766203 | 0.773759954 |
| VLDL_TG | Obesity_pPS | 4254 | 0.035915492 | [0.01 0.07] | 0.019508194 | 0.088360646 |
| VLDL_TG | VAT_Neg_pPS | 4254 | 0.012077877 | [-0.02 0.04] | 0.432318724 | 0.697423114 |
| VLDL_TG | Bilirubin_pPS | 4254 | -0.009741675 | [-0.04 0.02] | 0.526514061 | 0.776773139 |
| VLDL_TG | Proinsulin_pPS | 4254 | 0.007246267 | [-0.02 0.04] | 0.63757135 | 0.835456904 |
| VLDL_TG | Hyper_Insulin_pPS | 4254 | 0.006040891 | [-0.02 0.04] | 0.69451804 | 0.858808524 |
| VLDL_TG | Beta_Cell_1_pPS | 4254 | 0.00680795 | [-0.02 0.04] | 0.65805477 | 0.844628798 |
| VLDL_TG | Beta_Cell_2_pPS | 4254 | -0.003212188 | [-0.03 0.03] | 0.834581097 | 0.923149681 |
| XL_HDL_C | Cholesterol_pPS | 4255 | -0.014500707 | [-0.04 0.02] | 0.345745168 | 0.637861136 |
| XL_HDL_C | Lipodystrophy_1_pPS | 4255 | -0.044999641 | [-0.08 -0.01] | 0.003418916 | 0.023202274 |
| XL_HDL_C | Lipodystrophy_2_pPS | 4255 | -0.036787729 | [-0.07 -0.01] | 0.016724016 | 0.07971781 |
| XL_HDL_C | Liver-Lipid_pPS | 4255 | 0.020156763 | [-0.01 0.05] | 0.189955653 | 0.473428767 |
| XL_HDL_C | ASAT_Pos_pPS | 4255 | -0.034544549 | [-0.06 -0. ] | 0.024657258 | 0.106642464 |
| XL_HDL_C | ALP_Neg_pPS | 4255 | 0.025647518 | [-0. 0.06] | 0.095344281 | 0.313946136 |
| XL_HDL_C | SHBG-LpA_pPS | 4255 | 0.017884121 | [-0.01 0.05] | 0.244868148 | 0.542782585 |
| XL_HDL_C | Obesity_pPS | 4255 | -0.020649811 | [-0.05 0.01] | 0.179343885 | 0.457575075 |
| XL_HDL_C | VAT_Neg_pPS | 4255 | -0.007673788 | [-0.04 0.02] | 0.617813812 | 0.829552818 |
| XL_HDL_C | Bilirubin_pPS | 4255 | -0.014340216 | [-0.04 0.02] | 0.351110376 | 0.637861136 |
| XL_HDL_C | Proinsulin_pPS | 4255 | -0.016210667 | [-0.05 0.01] | 0.291848897 | 0.588843777 |
| XL_HDL_C | Hyper_Insulin_pPS | 4255 | -0.011967588 | [-0.04 0.02] | 0.436479877 | 0.700747966 |
| XL_HDL_C | Beta_Cell_1_pPS | 4255 | 0.003384391 | [-0.03 0.03] | 0.82583132 | 0.920041348 |
| XL_HDL_C | Beta_Cell_2_pPS | 4255 | 0.020299434 | [-0.01 0.05] | 0.186838509 | 0.46932333 |
| XL_HDL_CE | Cholesterol_pPS | 4255 | -0.01989469 | [-0.05 0.01] | 0.195781023 | 0.480924673 |
| XL_HDL_CE | Lipodystrophy_1_pPS | 4255 | -0.043437776 | [-0.07 -0.01] | 0.004718858 | 0.029614901 |
| XL_HDL_CE | Lipodystrophy_2_pPS | 4255 | -0.036696073 | [-0.07 -0.01] | 0.016997852 | 0.080830642 |
| XL_HDL_CE | Liver-Lipid_pPS | 4255 | 0.020114032 | [-0.01 0.05] | 0.190896673 | 0.474162704 |
| XL_HDL_CE | ASAT_Pos_pPS | 4255 | -0.033657747 | [-0.06 -0. ] | 0.028595319 | 0.119018357 |
| XL_HDL_CE | ALP_Neg_pPS | 4255 | 0.026176336 | [-0. 0.06] | 0.088707235 | 0.295493983 |
| XL_HDL_CE | SHBG-LpA_pPS | 4255 | 0.017824594 | [-0.01 0.05] | 0.246442558 | 0.544567329 |
| XL_HDL_CE | Obesity_pPS | 4255 | -0.018470934 | [-0.05 0.01] | 0.229724701 | 0.525010105 |
| XL_HDL_CE | VAT_Neg_pPS | 4255 | -0.00725011 | [-0.04 0.02] | 0.637353158 | 0.835456904 |
| XL_HDL_CE | Bilirubin_pPS | 4255 | -0.014689099 | [-0.04 0.02] | 0.339514281 | 0.634316189 |
| XL_HDL_CE | Proinsulin_pPS | 4255 | -0.016422051 | [-0.05 0.01] | 0.285601238 | 0.583442529 |
| XL_HDL_CE | Hyper_Insulin_pPS | 4255 | -0.014102899 | [-0.04 0.02] | 0.359139913 | 0.64484135 |
| XL_HDL_CE | Beta_Cell_1_pPS | 4255 | 0.004530843 | [-0.03 0.03] | 0.768304481 | 0.886548308 |
| XL_HDL_CE | Beta_Cell_2_pPS | 4255 | 0.02009137 | [-0.01 0.05] | 0.191397109 | 0.474464643 |
| XL_HDL_FC | Cholesterol_pPS | 4255 | -0.001198076 | [-0.03 0.03] | 0.937909207 | 0.978170683 |
| XL_HDL_FC | Lipodystrophy_1_pPS | 4255 | -0.048223856 | [-0.08 -0.02] | 0.001705097 | 0.013424546 |
| XL_HDL_FC | Lipodystrophy_2_pPS | 4255 | -0.037402204 | [-0.07 -0.01] | 0.014986149 | 0.073534975 |
| XL_HDL_FC | Liver-Lipid_pPS | 4255 | 0.020148914 | [-0.01 0.05] | 0.190128236 | 0.473428767 |
| XL_HDL_FC | ASAT_Pos_pPS | 4255 | -0.036260641 | [-0.07 -0.01] | 0.018353191 | 0.084752963 |
| XL_HDL_FC | ALP_Neg_pPS | 4255 | 0.023923979 | [-0.01 0.05] | 0.119769628 | 0.364405465 |
| XL_HDL_FC | SHBG-LpA_pPS | 4255 | 0.017575989 | [-0.01 0.05] | 0.25309444 | 0.549908188 |
| XL_HDL_FC | Obesity_pPS | 4255 | -0.025322531 | [-0.06 0. ] | 0.099616541 | 0.324808329 |
| XL_HDL_FC | VAT_Neg_pPS | 4255 | -0.008810787 | [-0.04 0.02] | 0.566724811 | 0.797755947 |
| XL_HDL_FC | Bilirubin_pPS | 4255 | -0.013373423 | [-0.04 0.02] | 0.38453625 | 0.662514262 |
| XL_HDL_FC | Proinsulin_pPS | 4255 | -0.014233798 | [-0.04 0.02] | 0.354696824 | 0.640530128 |
| XL_HDL_FC | Hyper_Insulin_pPS | 4255 | -0.006662614 | [-0.04 0.02] | 0.664867448 | 0.849453588 |
| XL_HDL_FC | Beta_Cell_1_pPS | 4255 | 0.000176715 | [-0.03 0.03] | 0.990832629 | 0.994309234 |
| XL_HDL_FC | Beta_Cell_2_pPS | 4255 | 0.020235197 | [-0.01 0.05] | 0.188237277 | 0.471379886 |
| XL_HDL_L | Cholesterol_pPS | 4255 | 0.001941118 | [-0.03 0.03] | 0.899565603 | 0.958961841 |
| XL_HDL_L | Lipodystrophy_1_pPS | 4255 | -0.049047494 | [-0.08 -0.02] | 0.001418055 | 0.01154043 |
| XL_HDL_L | Lipodystrophy_2_pPS | 4255 | -0.037537678 | [-0.07 -0.01] | 0.014625122 | 0.072473995 |
| XL_HDL_L | Liver-Lipid_pPS | 4255 | 0.023515554 | [-0.01 0.05] | 0.126221161 | 0.378852721 |
| XL_HDL_L | ASAT_Pos_pPS | 4255 | -0.037780489 | [-0.07 -0.01] | 0.013997191 | 0.070407981 |
| XL_HDL_L | ALP_Neg_pPS | 4255 | 0.024757422 | [-0.01 0.05] | 0.107407636 | 0.340519287 |
| XL_HDL_L | SHBG-LpA_pPS | 4255 | 0.016737396 | [-0.01 0.05] | 0.276449993 | 0.574717432 |
| XL_HDL_L | Obesity_pPS | 4255 | -0.028442095 | [-0.06 0. ] | 0.064362965 | 0.228061337 |
| XL_HDL_L | VAT_Neg_pPS | 4255 | -0.009107538 | [-0.04 0.02] | 0.55373182 | 0.795753776 |
| XL_HDL_L | Bilirubin_pPS | 4255 | -0.010439963 | [-0.04 0.02] | 0.497253921 | 0.753597539 |
| XL_HDL_L | Proinsulin_pPS | 4255 | -0.011175455 | [-0.04 0.02] | 0.467445213 | 0.730542792 |
| XL_HDL_L | Hyper_Insulin_pPS | 4255 | -0.005951492 | [-0.04 0.02] | 0.698782256 | 0.859092773 |
| XL_HDL_L | Beta_Cell_1_pPS | 4255 | -0.00210477 | [-0.03 0.03] | 0.891148806 | 0.954053428 |
| XL_HDL_L | Beta_Cell_2_pPS | 4255 | 0.018783831 | [-0.01 0.05] | 0.221927823 | 0.514831403 |
| XL_HDL_P | Cholesterol_pPS | 4255 | 0.00238854 | [-0.03 0.03] | 0.87658383 | 0.946048963 |
| XL_HDL_P | Lipodystrophy_1_pPS | 4255 | -0.048914727 | [-0.08 -0.02] | 0.00146109 | 0.011837974 |
| XL_HDL_P | Lipodystrophy_2_pPS | 4255 | -0.037714724 | [-0.07 -0.01] | 0.014164872 | 0.070895183 |
| XL_HDL_P | Liver-Lipid_pPS | 4255 | 0.023575574 | [-0.01 0.05] | 0.125256475 | 0.377655817 |
| XL_HDL_P | ASAT_Pos_pPS | 4255 | -0.037977911 | [-0.07 -0.01] | 0.013504298 | 0.068271729 |
| XL_HDL_P | ALP_Neg_pPS | 4255 | 0.024971084 | [-0.01 0.06] | 0.10440701 | 0.334436534 |
| XL_HDL_P | SHBG-LpA_pPS | 4255 | 0.016578552 | [-0.01 0.05] | 0.281034332 | 0.579434329 |
| XL_HDL_P | Obesity_pPS | 4255 | -0.028824868 | [-0.06 0. ] | 0.060853261 | 0.219510322 |
| XL_HDL_P | VAT_Neg_pPS | 4255 | -0.00909802 | [-0.04 0.02] | 0.554146275 | 0.795753776 |
| XL_HDL_P | Bilirubin_pPS | 4255 | -0.010087568 | [-0.04 0.02] | 0.511886824 | 0.767666412 |
| XL_HDL_P | Proinsulin_pPS | 4255 | -0.010647121 | [-0.04 0.02] | 0.488756963 | 0.748654506 |
| XL_HDL_P | Hyper_Insulin_pPS | 4255 | -0.005576591 | [-0.04 0.02] | 0.71691235 | 0.866099998 |
| XL_HDL_P | Beta_Cell_1_pPS | 4255 | -0.002401616 | [-0.03 0.03] | 0.875913647 | 0.946048963 |
| XL_HDL_P | Beta_Cell_2_pPS | 4255 | 0.018524562 | [-0.01 0.05] | 0.228374725 | 0.522699573 |
| XL_HDL_PL | Cholesterol_pPS | 4255 | 0.021429937 | [-0.01 0.05] | 0.163463207 | 0.43985664 |
| XL_HDL_PL | Lipodystrophy_1_pPS | 4255 | -0.055043218 | [-0.09 -0.02] | 0.000341519 | 0.003970654 |
| XL_HDL_PL | Lipodystrophy_2_pPS | 4255 | -0.038164647 | [-0.07 -0.01] | 0.013052259 | 0.066490132 |
| XL_HDL_PL | Liver-Lipid_pPS | 4255 | 0.02790435 | [-0. 0.06] | 0.069574232 | 0.241818772 |
| XL_HDL_PL | ASAT_Pos_pPS | 4255 | -0.038695317 | [-0.07 -0.01] | 0.01183984 | 0.06140767 |
| XL_HDL_PL | ALP_Neg_pPS | 4255 | 0.021742781 | [-0.01 0.05] | 0.157401269 | 0.431435756 |
| XL_HDL_PL | SHBG-LpA_pPS | 4255 | 0.014878329 | [-0.02 0.04] | 0.333328627 | 0.628957504 |
| XL_HDL_PL | Obesity_pPS | 4255 | -0.032790895 | [-0.06 -0. ] | 0.032956559 | 0.131432331 |
| XL_HDL_PL | VAT_Neg_pPS | 4255 | -0.01046585 | [-0.04 0.02] | 0.496187831 | 0.753425828 |
| XL_HDL_PL | Bilirubin_pPS | 4255 | -0.006546979 | [-0.04 0.02] | 0.670338085 | 0.850454275 |
| XL_HDL_PL | Proinsulin_pPS | 4255 | -0.007518024 | [-0.04 0.02] | 0.624967018 | 0.833566935 |
| XL_HDL_PL | Hyper_Insulin_pPS | 4255 | -0.001085543 | [-0.03 0.03] | 0.943731108 | 0.978747744 |

**Table S9: Associations between pPSs and global metabolomic profile in SCAPIS**

**a) Spearman Correlation**

| Metabolomics | PPS | Samp | r | CI95% | p | fdr_p_adjusted |
|---|---|---|---|---|---|---|
| VLDL_C | Beta_Cell_1_pPS | 4254 | 0.02000835 | [-0.01 0.05] | 0.192607888 | 0.464503229 |
| VLDL_C | Beta_Cell_2_pPS | 4254 | 0.010828694 | [-0.02 0.04] | 0.480752625 | 0.716353047 |
| VLDL_C | Total_GRS | 4254 | 0.025588837 | [-0. 0.06] | 0.095633974 | 0.293049822 |
| VLDL_D | Cholesterol_pPS | 4254 | -0.043575647 | [-0.07 -0.01] | 0.004530886 | 0.025235002 |
| VLDL_D | Lipodystrophy_1_pPS | 4254 | 0.058458148 | [0.03 0.09] | 0.000139219 | 0.001767006 |
| VLDL_D | Lipodystrophy_2_pPS | 4254 | 0.022046504 | [-0.01 0.05] | 0.151100767 | 0.401178951 |
| VLDL_D | Liver-Lipid_pPS | 4254 | -0.067502375 | [-0.1 -0.04] | 1.08E-05 | 0.000191726 |
| VLDL_D | ASAT_Pos_pPS | 4254 | 0.048401954 | [0.02 0.08] | 0.001613975 | 0.012233133 |
| VLDL_D | ALP_Neg_pPS | 4254 | -0.015716317 | [-0.05 0.01] | 0.306130443 | 0.592255382 |
| VLDL_D | SHBG-LpA_pPS | 4254 | -0.009783731 | [-0.04 0.02] | 0.524093503 | 0.74051162 |
| VLDL_D | Obesity_pPS | 4254 | 0.039065794 | [0.01 0.07] | 0.010940251 | 0.050575084 |
| VLDL_D | VAT_Neg_pPS | 4254 | 0.022302452 | [-0.01 0.05] | 0.146411135 | 0.396530158 |
| VLDL_D | Bilirubin_pPS | 4254 | -0.002191545 | [-0.03 0.03] | 0.886531202 | 0.946545261 |
| VLDL_D | Proinsulin_pPS | 4254 | 0.010113358 | [-0.02 0.04] | 0.510209088 | 0.7313594 |
| VLDL_D | Hyper_Insulin_pPS | 4254 | 0.020740186 | [-0.01 0.05] | 0.176833623 | 0.445197325 |
| VLDL_D | Beta_Cell_1_pPS | 4254 | 0.008033185 | [-0.02 0.04] | 0.600931995 | 0.790312158 |
| VLDL_D | Beta_Cell_2_pPS | 4254 | -0.006712604 | [-0.04 0.02] | 0.662058966 | 0.825654931 |
| VLDL_D | Total_GRS | 4254 | 0.043117866 | [0.01 0.07] | 0.004973074 | 0.026802121 |
| VLDL_TG | Cholesterol_pPS | 4254 | -0.080753704 | [-0.11 -0.05] | 1.39E-07 | 3.51E-06 |
| VLDL_TG | Lipodystrophy_1_pPS | 4254 | 0.069031052 | [0.04 0.1 ] | 6.79E-06 | 0.0001267 |
| VLDL_TG | Lipodystrophy_2_pPS | 4254 | 0.013945075 | [-0.02 0.04] | 0.36386337 | 0.639224774 |
| VLDL_TG | Liver-Lipid_pPS | 4254 | -0.049245087 | [-0.08 -0.02] | 0.001334919 | 0.010527207 |
| VLDL_TG | ASAT_Pos_pPS | 4254 | 0.045223422 | [0.02 0.08] | 0.003218401 | 0.019717477 |
| VLDL_TG | ALP_Neg_pPS | 4254 | 0.001766122 | [-0.03 0.03] | 0.908449187 | 0.952872131 |
| VLDL_TG | SHBG-LpA_pPS | 4254 | -0.003099383 | [-0.03 0.03] | 0.840068547 | 0.926450917 |
| VLDL_TG | Obesity_pPS | 4254 | 0.032883855 | [0. 0.06] | 0.032218078 | 0.125423352 |
| VLDL_TG | VAT_Neg_pPS | 4254 | 0.018427155 | [-0.01 0.05] | 0.230170195 | 0.513581257 |
| VLDL_TG | Bilirubin_pPS | 4254 | -0.005928963 | [-0.04 0.02] | 0.699462055 | 0.852954013 |
| VLDL_TG | Proinsulin_pPS | 4254 | 0.011577591 | [-0.02 0.04] | 0.450933958 | 0.694584169 |
| VLDL_TG | Hyper_Insulin_pPS | 4254 | 0.016370905 | [-0.01 0.05] | 0.28642152 | 0.57504794 |
| VLDL_TG | Beta_Cell_1_pPS | 4254 | 0.012724894 | [-0.02 0.04] | 0.407347893 | 0.671010017 |
| VLDL_TG | Beta_Cell_2_pPS | 4254 | -0.000733228 | [-0.03 0.03] | 0.961922205 | 0.982019059 |
| VLDL_TG | Total_GRS | 4254 | 0.042519548 | [0.01 0.07] | 0.005609732 | 0.029710801 |
| XL_HDL_C | Cholesterol_pPS | 4255 | -0.0175779 | [-0.05 0.01] | 0.252314455 | 0.534775319 |
| XL_HDL_C | Lipodystrophy_1_pPS | 4255 | -0.055210615 | [-0.09 -0.03] | 0.00032066 | 0.00356381 |
| XL_HDL_C | Lipodystrophy_2_pPS | 4255 | -0.044601393 | [-0.07 -0.01] | 0.003662575 | 0.021523898 |
| XL_HDL_C | Liver-Lipid_pPS | 4255 | 0.018584192 | [-0.01 0.05] | 0.22616734 | 0.507457055 |
| XL_HDL_C | ASAT_Pos_pPS | 4255 | -0.048811421 | [-0.08 -0.02] | 0.001470452 | 0.011386712 |
| XL_HDL_C | ALP_Neg_pPS | 4255 | 0.021760258 | [-0.01 0.05] | 0.156431285 | 0.409700984 |
| XL_HDL_C | SHBG-LpA_pPS | 4255 | 0.012747378 | [-0.02 0.04] | 0.406464267 | 0.67066604 |
| XL_HDL_C | Obesity_pPS | 4255 | -0.027087146 | [-0.06 0. ] | 0.077695814 | 0.247266351 |
| XL_HDL_C | VAT_Neg_pPS | 4255 | -0.012169299 | [-0.04 0.02] | 0.428078877 | 0.687522331 |
| XL_HDL_C | Bilirubin_pPS | 4255 | -0.016842013 | [-0.05 0.01] | 0.27272232 | 0.556602641 |
| XL_HDL_C | Proinsulin_pPS | 4255 | -0.011197361 | [-0.04 0.02] | 0.465888403 | 0.706538163 |
| XL_HDL_C | Hyper_Insulin_pPS | 4255 | -0.023141368 | [-0.05 0.01] | 0.131772879 | 0.369964431 |
| XL_HDL_C | Beta_Cell_1_pPS | 4255 | 0.001224592 | [-0.03 0.03] | 0.936440535 | 0.969900989 |
| XL_HDL_C | Beta_Cell_2_pPS | 4255 | 0.018269601 | [-0.01 0.05] | 0.234125148 | 0.516665065 |
| XL_HDL_C | Total_GRS | 4255 | -0.041798848 | [-0.07 -0.01] | 0.00646736 | 0.033589556 |
| XL_HDL_CE | Cholesterol_pPS | 4255 | -0.022851136 | [-0.05 0.01] | 0.13668755 | 0.378794282 |
| XL_HDL_CE | Lipodystrophy_1_pPS | 4255 | -0.053592878 | [-0.08 -0.02] | 0.000478645 | 0.004994081 |
| XL_HDL_CE | Lipodystrophy_2_pPS | 4255 | -0.044659957 | [-0.07 -0.01] | 0.003618119 | 0.021420079 |
| XL_HDL_CE | Liver-Lipid_pPS | 4255 | 0.018433816 | [-0.01 0.05] | 0.229946807 | 0.513581257 |
| XL_HDL_CE | ASAT_Pos_pPS | 4255 | -0.047630482 | [-0.08 -0.02] | 0.001913002 | 0.01372371 |
| XL_HDL_CE | ALP_Neg_pPS | 4255 | 0.022271276 | [-0.01 0.05] | 0.146928667 | 0.396835019 |
| XL_HDL_CE | SHBG-LpA_pPS | 4255 | 0.012833605 | [-0.02 0.04] | 0.403296982 | 0.668142396 |
| XL_HDL_CE | Obesity_pPS | 4255 | -0.02542025 | [-0.06 0. ] | 0.097800444 | 0.297985728 |
| XL_HDL_CE | VAT_Neg_pPS | 4255 | -0.01157507 | [-0.04 0.02] | 0.450979158 | 0.694584169 |
| XL_HDL_CE | Bilirubin_pPS | 4255 | -0.017063268 | [-0.05 0.01] | 0.26647112 | 0.552249619 |
| XL_HDL_CE | Proinsulin_pPS | 4255 | -0.010827516 | [-0.04 0.02] | 0.480748638 | 0.716353047 |
| XL_HDL_CE | Hyper_Insulin_pPS | 4255 | -0.024946219 | [-0.05 0.01] | 0.104220383 | 0.312224471 |
| XL_HDL_CE | Beta_Cell_1_pPS | 4255 | 0.00254409 | [-0.03 0.03] | 0.868417299 | 0.940757973 |
| XL_HDL_CE | Beta_Cell_2_pPS | 4255 | 0.018325634 | [-0.01 0.05] | 0.232693418 | 0.516301672 |
| XL_HDL_CE | Total_GRS | 4255 | -0.040962856 | [-0.07 -0.01] | 0.007617145 | 0.038085724 |
| XL_HDL_FC | Cholesterol_pPS | 4255 | -0.004495559 | [-0.03 0.03] | 0.769714544 | 0.890527344 |
| XL_HDL_FC | Lipodystrophy_1_pPS | 4255 | -0.058512275 | [-0.09 -0.03] | 0.000136993 | 0.001749106 |
| XL_HDL_FC | Lipodystrophy_2_pPS | 4255 | -0.044771661 | [-0.07 -0.01] | 0.00353468 | 0.021002463 |
| XL_HDL_FC | Liver-Lipid_pPS | 4255 | 0.018838983 | [-0.01 0.05] | 0.219865093 | 0.500648222 |
| XL_HDL_FC | ASAT_Pos_pPS | 4255 | -0.051128521 | [-0.08 -0.02] | 0.00086341 | 0.007629602 |
| XL_HDL_FC | ALP_Neg_pPS | 4255 | 0.020361892 | [-0.01 0.05] | 0.184811209 | 0.455655222 |
| XL_HDL_FC | SHBG-LpA_pPS | 4255 | 0.012176105 | [-0.02 0.04] | 0.427820537 | 0.687522331 |
| XL_HDL_FC | Obesity_pPS | 4255 | -0.030274681 | [-0.06 -0. ] | 0.048619583 | 0.170685771 |
| XL_HDL_FC | VAT_Neg_pPS | 4255 | -0.013822723 | [-0.04 0.02] | 0.368031617 | 0.642333457 |
| XL_HDL_FC | Bilirubin_pPS | 4255 | -0.016148206 | [-0.05 0.01] | 0.292971372 | 0.578724334 |
| XL_HDL_FC | Proinsulin_pPS | 4255 | -0.010775846 | [-0.04 0.02] | 0.482845059 | 0.717164725 |
| XL_HDL_FC | Hyper_Insulin_pPS | 4255 | -0.018555349 | [-0.05 0.01] | 0.226888813 | 0.50854389 |
| XL_HDL_FC | Beta_Cell_1_pPS | 4255 | -0.002095783 | [-0.03 0.03] | 0.89144522 | 0.948487102 |
| XL_HDL_FC | Beta_Cell_2_pPS | 4255 | 0.017689835 | [-0.01 0.05] | 0.249306016 | 0.530517267 |
| XL_HDL_FC | Total_GRS | 4255 | -0.043461102 | [-0.07 -0.01] | 0.004633217 | 0.02541752 |
| XL_HDL_L | Cholesterol_pPS | 4255 | -0.001442924 | [-0.03 0.03] | 0.925139338 | 0.963780418 |
| XL_HDL_L | Lipodystrophy_1_pPS | 4255 | -0.059669854 | [-0.09 -0.03] | 0.000100611 | 0.001458178 |
| XL_HDL_L | Lipodystrophy_2_pPS | 4255 | -0.044795781 | [-0.07 -0.01] | 0.003516895 | 0.021002463 |
| XL_HDL_L | Liver-Lipid_pPS | 4255 | 0.022189199 | [-0.01 0.05] | 0.148424442 | 0.398461113 |
| XL_HDL_L | ASAT_Pos_pPS | 4255 | -0.052851859 | [-0.08 -0.02] | 0.000573019 | 0.005770548 |
| XL_HDL_L | ALP_Neg_pPS | 4255 | 0.021842024 | [-0.01 0.05] | 0.154880256 | 0.407130084 |
| XL_HDL_L | SHBG-LpA_pPS | 4255 | 0.011269512 | [-0.02 0.04] | 0.463019416 | 0.703881394 |
| XL_HDL_L | Obesity_pPS | 4255 | -0.032771463 | [-0.06 -0. ] | 0.032791883 | 0.126964963 |
| XL_HDL_L | VAT_Neg_pPS | 4255 | -0.014496451 | [-0.04 0.02] | 0.345146271 | 0.62662761 |
| XL_HDL_L | Bilirubin_pPS | 4255 | -0.013401154 | [-0.04 0.02] | 0.382820862 | 0.653782443 |
| XL_HDL_L | Proinsulin_pPS | 4255 | -0.008589819 | [-0.04 0.02] | 0.57590801 | 0.773311572 |
| XL_HDL_L | Hyper_Insulin_pPS | 4255 | -0.018126546 | [-0.05 0.01] | 0.23780868 | 0.519979223 |
| XL_HDL_L | Beta_Cell_1_pPS | 4255 | -0.003972073 | [-0.03 0.03] | 0.7958984 | 0.903281518 |
| XL_HDL_L | Beta_Cell_2_pPS | 4255 | 0.016230669 | [-0.01 0.05] | 0.29051318 | 0.577526201 |
| XL_HDL_L | Total_GRS | 4255 | -0.045164519 | [-0.08 -0.02] | 0.003254946 | 0.019834827 |
| XL_HDL_P | Cholesterol_pPS | 4255 | -0.001012238 | [-0.03 0.03] | 0.947444637 | 0.974500144 |

**b) Partial Correlation**

| Metabolomics | PPS | Samp | r | CI95% | p | fdr_p_adjusted |
|---|---|---|---|---|---|---|
| XL_HDL_PL | Beta_Cell_1_pPS | 4255 | -0.005684067 | [-0.04 0.02] | 0.711697958 | 0.86476088 |
| XL_HDL_PL | Beta_Cell_2_pPS | 4255 | 0.016021628 | [-0.01 0.05] | 0.29751334 | 0.59502668 |
| XL_HDL_TG | Cholesterol_pPS | 4255 | -0.065992989 | [-0.1 -0.04] | 1.74E-05 | 0.000342113 |
| XL_HDL_TG | Lipodystrophy_1_pPS | 4255 | 0.025609059 | [-0. 0.06] | 0.095842077 | 0.315067058 |
| XL_HDL_TG | Lipodystrophy_2_pPS | 4255 | -0.038718255 | [-0.07 -0.01] | 0.011789761 | 0.061306756 |
| XL_HDL_TG | Liver-Lipid_pPS | 4255 | -0.02989145 | [-0.06 0. ] | 0.051901075 | 0.192775422 |
| XL_HDL_TG | ASAT_Pos_pPS | 4255 | -0.013137026 | [-0.04 0.02] | 0.392996327 | 0.671954495 |
| XL_HDL_TG | ALP_Neg_pPS | 4255 | 0.040177587 | [0.01 0.07] | 0.008965395 | 0.049582102 |
| XL_HDL_TG | SHBG-LpA_pPS | 4255 | 0.002214906 | [-0.03 0.03] | 0.885491249 | 0.951558497 |
| XL_HDL_TG | Obesity_pPS | 4255 | 0.003542078 | [-0.03 0.03] | 0.817855605 | 0.913698059 |
| XL_HDL_TG | VAT_Neg_pPS | 4255 | 0.011798956 | [-0.02 0.04] | 0.442971092 | 0.706833249 |
| XL_HDL_TG | Bilirubin_pPS | 4255 | -0.025065656 | [-0.06 0.01] | 0.103100291 | 0.331843701 |
| XL_HDL_TG | Proinsulin_pPS | 4255 | 0.002945928 | [-0.03 0.03] | 0.8481007 | 0.932567807 |
| XL_HDL_TG | Hyper_Insulin_pPS | 4255 | -0.005672676 | [-0.04 0.02] | 0.712249967 | 0.86476088 |
| XL_HDL_TG | Beta_Cell_1_pPS | 4255 | 0.001054542 | [-0.03 0.03] | 0.945335473 | 0.978747744 |
| XL_HDL_TG | Beta_Cell_2_pPS | 4255 | 0.012254583 | [-0.02 0.04] | 0.425559401 | 0.693893617 |
| XL_VLDL_C | Cholesterol_pPS | 4255 | -0.090325297 | [-0.12 -0.06] | 3.98E-09 | 1.17E-07 |
| XL_VLDL_C | Lipodystrophy_1_pPS | 4255 | 0.05208189 | [0.02 0.08] | 0.000702442 | 0.00685438 |
| XL_VLDL_C | Lipodystrophy_2_pPS | 4255 | -0.010018275 | [-0.04 0.02] | 0.514790335 | 0.769688104 |
| XL_VLDL_C | Liver-Lipid_pPS | 4255 | -0.044325779 | [-0.07 -0.01] | 0.003933512 | 0.025989737 |
| XL_VLDL_C | ASAT_Pos_pPS | 4255 | 0.034470858 | [0. 0.06] | 0.024965587 | 0.107486249 |
| XL_VLDL_C | ALP_Neg_pPS | 4255 | 0.016430407 | [-0.01 0.05] | 0.285356128 | 0.583442529 |
| XL_VLDL_C | SHBG-LpA_pPS | 4255 | -0.005453731 | [-0.04 0.02] | 0.722889244 | 0.866319132 |
| XL_VLDL_C | Obesity_pPS | 4255 | 0.033670377 | [0. 0.06] | 0.028535638 | 0.119017391 |
| XL_VLDL_C | VAT_Neg_pPS | 4255 | 0.025276114 | [-0. 0.06] | 0.100239017 | 0.326306524 |
| XL_VLDL_C | Bilirubin_pPS | 4255 | -0.018232196 | [-0.05 0.01] | 0.235803285 | 0.531619568 |
| XL_VLDL_C | Proinsulin_pPS | 4255 | 0.003706594 | [-0.03 0.03] | 0.809554583 | 0.91103332 |
| XL_VLDL_C | Hyper_Insulin_pPS | 4255 | 0.003344758 | [-0.03 0.03] | 0.827838822 | 0.920813717 |
| XL_VLDL_C | Beta_Cell_1_pPS | 4255 | 0.007031048 | [-0.02 0.04] | 0.647556778 | 0.842918511 |
| XL_VLDL_C | Beta_Cell_2_pPS | 4255 | -0.002127017 | [-0.03 0.03] | 0.890005566 | 0.953849648 |
| XL_VLDL_CE | Cholesterol_pPS | 4255 | -0.085962206 | [-0.12 -0.06] | 2.15E-08 | 5.81E-07 |
| XL_VLDL_CE | Lipodystrophy_1_pPS | 4255 | 0.052217289 | [0.02 0.08] | 0.000680181 | 0.0067412 |
| XL_VLDL_CE | Lipodystrophy_2_pPS | 4255 | -0.007700937 | [-0.04 0.02] | 0.616570727 | 0.828794354 |
| XL_VLDL_CE | Liver-Lipid_pPS | 4255 | -0.042300425 | [-0.07 -0.01] | 0.005930989 | 0.035319581 |
| XL_VLDL_CE | ASAT_Pos_pPS | 4255 | 0.036363307 | [0.01 0.07] | 0.018025382 | 0.084118448 |
| XL_VLDL_CE | ALP_Neg_pPS | 4255 | 0.015922581 | [-0.01 0.05] | 0.300510356 | 0.597661337 |
| XL_VLDL_CE | SHBG-LpA_pPS | 4255 | -0.007348762 | [-0.04 0.02] | 0.632780276 | 0.835456904 |
| XL_VLDL_CE | Obesity_pPS | 4255 | 0.032129044 | [0. 0.06] | 0.036658687 | 0.143341196 |
| XL_VLDL_CE | VAT_Neg_pPS | 4255 | 0.026290974 | [-0. 0.06] | 0.087318705 | 0.291839812 |
| XL_VLDL_CE | Bilirubin_pPS | 4255 | -0.01744422 | [-0.05 0.01] | 0.256670394 | 0.55312608 |
| XL_VLDL_CE | Proinsulin_pPS | 4255 | 0.004820169 | [-0.03 0.03] | 0.753972633 | 0.878099599 |
| XL_VLDL_CE | Hyper_Insulin_pPS | 4255 | 0.004644156 | [-0.03 0.03] | 0.762681743 | 0.885152957 |
| XL_VLDL_CE | Beta_Cell_1_pPS | 4255 | 0.005772588 | [-0.02 0.04] | 0.707413307 | 0.863035613 |
| XL_VLDL_CE | Beta_Cell_2_pPS | 4255 | -0.000930875 | [-0.03 0.03] | 0.951737675 | 0.979629216 |
| XL_VLDL_FC | Cholesterol_pPS | 4255 | -0.094450255 | [-0.12 -0.06] | 7.52E-10 | 2.47E-08 |
| XL_VLDL_FC | Lipodystrophy_1_pPS | 4255 | 0.050765967 | [0.02 0.08] | 0.00095692 | 0.008545465 |
| XL_VLDL_FC | Lipodystrophy_2_pPS | 4255 | -0.012339131 | [-0.04 0.02] | 0.422372899 | 0.69367559 |
| XL_VLDL_FC | Liver-Lipid_pPS | 4255 | -0.04728927 | [-0.08 -0.02] | 0.002094934 | 0.015886586 |
| XL_VLDL_FC | ASAT_Pos_pPS | 4255 | 0.031770596 | [0. 0.06] | 0.038807602 | 0.150567478 |
| XL_VLDL_FC | ALP_Neg_pPS | 4255 | 0.016821208 | [-0.01 0.05] | 0.274051754 | 0.573178999 |
| XL_VLDL_FC | SHBG-LpA_pPS | 4255 | -0.003601286 | [-0.03 0.03] | 0.814865719 | 0.912503804 |
| XL_VLDL_FC | Obesity_pPS | 4255 | 0.035218762 | [0.01 0.07] | 0.021986138 | 0.098388186 |
| XL_VLDL_FC | VAT_Neg_pPS | 4255 | 0.022809969 | [-0.01 0.05] | 0.138000151 | 0.405692073 |
| XL_VLDL_FC | Bilirubin_pPS | 4255 | -0.018384845 | [-0.05 0.01] | 0.231903673 | 0.527097702 |
| XL_VLDL_FC | Proinsulin_pPS | 4255 | 0.002393547 | [-0.03 0.03] | 0.876327186 | 0.946048963 |
| XL_VLDL_FC | Hyper_Insulin_pPS | 4255 | 0.001158462 | [-0.03 0.03] | 0.939958242 | 0.978430443 |
| XL_VLDL_FC | Beta_Cell_1_pPS | 4255 | 0.00883342 | [-0.02 0.04] | 0.565728724 | 0.797755947 |
| XL_VLDL_FC | Beta_Cell_2_pPS | 4255 | -0.003794695 | [-0.03 0.03] | 0.805117938 | 0.908082317 |
| XL_VLDL_L | Cholesterol_pPS | 4255 | -0.084730091 | [-0.11 -0.05] | 3.41E-08 | 8.64E-07 |
| XL_VLDL_L | Lipodystrophy_1_pPS | 4255 | 0.050868968 | [0.02 0.08] | 0.000934281 | 0.008501961 |
| XL_VLDL_L | Lipodystrophy_2_pPS | 4255 | -0.003334385 | [-0.03 0.03] | 0.828364388 | 0.920813717 |
| XL_VLDL_L | Liver-Lipid_pPS | 4255 | -0.051878783 | [-0.08 -0.02] | 0.000737108 | 0.007094663 |
| XL_VLDL_L | ASAT_Pos_pPS | 4255 | 0.03588542 | [0.01 0.07] | 0.019596037 | 0.088558161 |
| XL_VLDL_L | ALP_Neg_pPS | 4255 | 0.006107901 | [-0.02 0.04] | 0.691268113 | 0.858568103 |
| XL_VLDL_L | SHBG-LpA_pPS | 4255 | -0.006727109 | [-0.04 0.02] | 0.661823895 | 0.847168439 |
| XL_VLDL_L | Obesity_pPS | 4255 | 0.036855267 | [0.01 0.07] | 0.01652472 | 0.07895582 |
| XL_VLDL_L | VAT_Neg_pPS | 4255 | 0.019588248 | [-0.01 0.05] | 0.20275736 | 0.487899426 |
| XL_VLDL_L | Bilirubin_pPS | 4255 | -0.015636414 | [-0.05 0.01] | 0.309282035 | 0.610032152 |
| XL_VLDL_L | Proinsulin_pPS | 4255 | 0.004313315 | [-0.03 0.03] | 0.779132488 | 0.891327566 |
| XL_VLDL_L | Hyper_Insulin_pPS | 4255 | 0.00529242 | [-0.02 0.04] | 0.73076246 | 0.869031685 |
| XL_VLDL_L | Beta_Cell_1_pPS | 4255 | 0.006731151 | [-0.02 0.04] | 0.661633331 | 0.847168439 |
| XL_VLDL_L | Beta_Cell_2_pPS | 4255 | -0.005168239 | [-0.04 0.02] | 0.736842858 | 0.87275313 |
| XL_VLDL_P | Cholesterol_pPS | 4255 | -0.083537765 | [-0.11 -0.05] | 5.30E-08 | 1.31E-06 |
| XL_VLDL_P | Lipodystrophy_1_pPS | 4255 | 0.050712232 | [0.02 0.08] | 0.000968941 | 0.008545465 |
| XL_VLDL_P | Lipodystrophy_2_pPS | 4255 | -0.002304521 | [-0.03 0.03] | 0.88089214 | 0.949163651 |
| XL_VLDL_P | Liver-Lipid_pPS | 4255 | -0.052656984 | [-0.08 -0.02] | 0.00061233 | 0.006286587 |
| XL_VLDL_P | ASAT_Pos_pPS | 4255 | 0.03599798 | [0.01 0.07] | 0.019215707 | 0.087431465 |
| XL_VLDL_P | ALP_Neg_pPS | 4255 | 0.005101375 | [-0.03 0.04] | 0.740123646 | 0.873045882 |
| XL_VLDL_P | SHBG-LpA_pPS | 4255 | -0.00699116 | [-0.04 0.02] | 0.649421942 | 0.843700667 |
| XL_VLDL_P | Obesity_pPS | 4255 | 0.037047309 | [0.01 0.07] | 0.01596938 | 0.077787588 |
| XL_VLDL_P | VAT_Neg_pPS | 4255 | 0.019166102 | [-0.01 0.05] | 0.212661752 | 0.501470939 |
| XL_VLDL_P | Bilirubin_pPS | 4255 | -0.015261836 | [-0.05 0.01] | 0.321016775 | 0.620672596 |
| XL_VLDL_P | Proinsulin_pPS | 4255 | 0.004314995 | [-0.03 0.03] | 0.779048724 | 0.891327566 |
| XL_VLDL_P | Hyper_Insulin_pPS | 4255 | 0.005481639 | [-0.02 0.04] | 0.721530068 | 0.866319132 |
| XL_VLDL_P | Beta_Cell_1_pPS | 4255 | 0.006621664 | [-0.02 0.04] | 0.66680273 | 0.849453588 |
| XL_VLDL_P | Beta_Cell_2_pPS | 4255 | -0.005570062 | [-0.04 0.02] | 0.717229513 | 0.866099998 |
| XL_VLDL_PL | Cholesterol_pPS | 4255 | -0.092511695 | [-0.12 -0.06] | 1.66E-09 | 5.19E-08 |
| XL_VLDL_PL | Lipodystrophy_1_pPS | 4255 | 0.051666996 | [0.02 0.08] | 0.000774941 | 0.007318078 |
| XL_VLDL_PL | Lipodystrophy_2_pPS | 4255 | -0.008322676 | [-0.04 0.02] | 0.588408798 | 0.815104592 |
| XL_VLDL_PL | Liver-Lipid_pPS | 4255 | -0.049573604 | [-0.08 -0.02] | 0.001258757 | 0.010544065 |
| XL_VLDL_PL | ASAT_Pos_pPS | 4255 | 0.033057799 | [0. 0.06] | 0.031556782 | 0.127116051 |
| XL_VLDL_PL | ALP_Neg_pPS | 4255 | 0.011688992 | [-0.02 0.04] | 0.447233552 | 0.709019056 |
| XL_VLDL_PL | SHBG-LpA_pPS | 4255 | -0.005026488 | [-0.04 0.03] | 0.743803683 | 0.873512116 |
| XL_VLDL_PL | Obesity_pPS | 4255 | 0.036246962 | [0.01 0.07] | 0.018397257 | 0.084752963 |

**Table S9: Associations between pPSs and global metabolomic profile in SCAPIS**

**a) Spearman Correlation**

| Metabolomics | PPS | Samp | r | CI95% | p | fdr_p_adjusted |
|---|---|---|---|---|---|---|
| XL_HDL_P | Lipodystrophy_1_pPS | 4255 | -0.059552722 | [-0.09 -0.03] | 0.000103829 | 0.00147325 |
| XL_HDL_P | Lipodystrophy_2_pPS | 4255 | -0.044931168 | [-0.07 -0.01] | 0.003418568 | 0.020540133 |
| XL_HDL_P | Liver-Lipid_pPS | 4255 | 0.022265691 | [-0.01 0.05] | 0.147030082 | 0.396835019 |
| XL_HDL_P | ASAT_Pos_pPS | 4255 | -0.053027406 | [-0.08 -0.02] | 0.000549213 | 0.00559124 |
| XL_HDL_P | ALP_Neg_pPS | 4255 | 0.022161332 | [-0.01 0.05] | 0.148934933 | 0.398714883 |
| XL_HDL_P | SHBG-LpA_pPS | 4255 | 0.0111271 | [-0.02 0.04] | 0.468691681 | 0.707973539 |
| XL_HDL_P | Obesity_pPS | 4255 | -0.033029033 | [-0.06 -0. ] | 0.031443965 | 0.122854838 |
| XL_HDL_P | VAT_Neg_pPS | 4255 | -0.014533319 | [-0.04 0.02] | 0.343920657 | 0.62662761 |
| XL_HDL_P | Bilirubin_pPS | 4255 | -0.013060729 | [-0.04 0.02] | 0.395025224 | 0.662493437 |
| XL_HDL_P | Proinsulin_pPS | 4255 | -0.0081675 | [-0.04 0.02] | 0.594816523 | 0.786125349 |
| XL_HDL_P | Hyper_Insulin_pPS | 4255 | -0.017774519 | [-0.05 0.01] | 0.247046689 | 0.528857434 |
| XL_HDL_P | Beta_Cell_1_pPS | 4255 | -0.004188588 | [-0.03 0.03] | 0.785038985 | 0.897605875 |
| XL_HDL_P | Beta_Cell_2_pPS | 4255 | 0.015990182 | [-0.01 0.05] | 0.297720933 | 0.583739854 |
| XL_HDL_P | Total_GRS | 4255 | -0.045226099 | [-0.08 -0.02] | 0.003212973 | 0.019717477 |
| XL_HDL_PL | Cholesterol_pPS | 4255 | 0.017972935 | [-0.01 0.05] | 0.24180936 | 0.525211109 |
| XL_HDL_PL | Lipodystrophy_1_pPS | 4255 | -0.065699938 | [-0.1 -0.04] | 1.84E-05 | 0.000309067 |
| XL_HDL_PL | Lipodystrophy_2_pPS | 4255 | -0.044773485 | [-0.07 -0.01] | 0.003533332 | 0.021002463 |
| XL_HDL_PL | Liver-Lipid_pPS | 4255 | 0.02678537 | [-0. 0.06] | 0.081062751 | 0.254955428 |
| XL_HDL_PL | ASAT_Pos_pPS | 4255 | -0.054745771 | [-0.08 -0.02] | 0.000360167 | 0.003961835 |
| XL_HDL_PL | ALP_Neg_pPS | 4255 | 0.01958158 | [-0.01 0.05] | 0.2022181 | 0.478761397 |
| XL_HDL_PL | SHBG-LpA_pPS | 4255 | 0.008872627 | [-0.02 0.04] | 0.563406561 | 0.767793567 |
| XL_HDL_PL | Obesity_pPS | 4255 | -0.03494256 | [-0.06 -0. ] | 0.022838598 | 0.095131655 |
| XL_HDL_PL | VAT_Neg_pPS | 4255 | -0.016901022 | [-0.05 0.01] | 0.2710454 | 0.554396235 |
| XL_HDL_PL | Bilirubin_pPS | 4255 | -0.00989307 | [-0.04 0.02] | 0.519417335 | 0.736667707 |
| XL_HDL_PL | Proinsulin_pPS | 4255 | -0.006953458 | [-0.04 0.02] | 0.650686598 | 0.81860572 |
| XL_HDL_PL | Hyper_Insulin_pPS | 4255 | -0.014083439 | [-0.04 0.02] | 0.359065809 | 0.63736428 |
| XL_HDL_PL | Beta_Cell_1_pPS | 4255 | -0.007809945 | [-0.04 0.02] | 0.611043483 | 0.795465501 |
| XL_HDL_PL | Beta_Cell_2_pPS | 4255 | 0.013225115 | [-0.02 0.04] | 0.389102843 | 0.657703386 |
| XL_HDL_PL | Total_GRS | 4255 | -0.048151686 | [-0.08 -0.02] | 0.001704445 | 0.012650635 |
| XL_HDL_TG | Cholesterol_pPS | 4255 | -0.06810972 | [-0.1 -0.04] | 8.98E-06 | 0.000160501 |
| XL_HDL_TG | Lipodystrophy_1_pPS | 4255 | 0.021338853 | [-0.01 0.05] | 0.164612218 | 0.425926667 |
| XL_HDL_TG | Lipodystrophy_2_pPS | 4255 | -0.040803359 | [-0.07 -0.01] | 0.007856255 | 0.039189922 |
| XL_HDL_TG | Liver-Lipid_pPS | 4255 | -0.029946326 | [-0.06 0. ] | 0.051114662 | 0.177332222 |
| XL_HDL_TG | ASAT_Pos_pPS | 4255 | -0.013181576 | [-0.04 0.02] | 0.390666127 | 0.658788398 |
| XL_HDL_TG | ALP_Neg_pPS | 4255 | 0.040174581 | [0.01 0.07] | 0.008865699 | 0.043175051 |
| XL_HDL_TG | SHBG-LpA_pPS | 4255 | 0.004534091 | [-0.03 0.03] | 0.767797136 | 0.889703209 |
| XL_HDL_TG | Obesity_pPS | 4255 | -0.001258574 | [-0.03 0.03] | 0.934680713 | 0.969482654 |
| XL_HDL_TG | VAT_Neg_pPS | 4255 | 0.010070511 | [-0.02 0.04] | 0.511953178 | 0.732581432 |
| XL_HDL_TG | Bilirubin_pPS | 4255 | -0.023900435 | [-0.05 0.01] | 0.119566032 | 0.346579918 |
| XL_HDL_TG | Proinsulin_pPS | 4255 | 0.010632109 | [-0.02 0.04] | 0.488703001 | 0.718976637 |
| XL_HDL_TG | Hyper_Insulin_pPS | 4255 | -0.006126669 | [-0.04 0.02] | 0.689917472 | 0.845220564 |
| XL_HDL_TG | Beta_Cell_1_pPS | 4255 | 0.006982615 | [-0.02 0.04] | 0.649319827 | 0.818487176 |
| XL_HDL_TG | Beta_Cell_2_pPS | 4255 | 0.012825021 | [-0.02 0.04] | 0.403611617 | 0.668142396 |
| XL_HDL_TG | Total_GRS | 4255 | -0.001526496 | [-0.03 0.03] | 0.920817378 | 0.960210635 |
| XL_VLDL_C | Cholesterol_pPS | 4255 | -0.089519075 | [-0.12 -0.06] | 5.18E-09 | 1.69E-07 |
| XL_VLDL_C | Lipodystrophy_1_pPS | 4255 | 0.060223024 | [0.03 0.09] | 8.66E-05 | 0.001270302 |
| XL_VLDL_C | Lipodystrophy_2_pPS | 4255 | -0.005210518 | [-0.04 0.02] | 0.734378733 | 0.873878512 |
| XL_VLDL_C | Liver-Lipid_pPS | 4255 | -0.043559467 | [-0.07 -0.01] | 0.004541124 | 0.025235002 |
| XL_VLDL_C | ASAT_Pos_pPS | 4255 | 0.045243723 | [0.02 0.08] | 0.003201052 | 0.019717477 |
| XL_VLDL_C | ALP_Neg_pPS | 4255 | 0.017054122 | [-0.01 0.05] | 0.266727555 | 0.552249619 |
| XL_VLDL_C | SHBG-LpA_pPS | 4255 | 0.000958184 | [-0.03 0.03] | 0.9502474 | 0.97525391 |
| XL_VLDL_C | Obesity_pPS | 4255 | 0.029489075 | [-0. 0.06] | 0.054766973 | 0.187062353 |
| XL_VLDL_C | VAT_Neg_pPS | 4255 | 0.029335001 | [-0. 0.06] | 0.056045629 | 0.189712465 |
| XL_VLDL_C | Bilirubin_pPS | 4255 | -0.014545481 | [-0.04 0.02] | 0.343516994 | 0.62658362 |
| XL_VLDL_C | Proinsulin_pPS | 4255 | 0.010732286 | [-0.02 0.04] | 0.484616289 | 0.717164725 |
| XL_VLDL_C | Hyper_Insulin_pPS | 4255 | 0.013193484 | [-0.02 0.04] | 0.390238193 | 0.658788398 |
| XL_VLDL_C | Beta_Cell_1_pPS | 4255 | 0.013957578 | [-0.02 0.04] | 0.363376774 | 0.639224774 |
| XL_VLDL_C | Beta_Cell_2_pPS | 4255 | 0.000704796 | [-0.03 0.03] | 0.963393354 | 0.982019059 |
| XL_VLDL_C | Total_GRS | 4255 | 0.034871688 | [0. 0.06] | 0.023116456 | 0.095908699 |
| XL_VLDL_CE | Cholesterol_pPS | 4255 | -0.085110971 | [-0.11 -0.06] | 2.82E-08 | 7.95E-07 |
| XL_VLDL_CE | Lipodystrophy_1_pPS | 4255 | 0.060858309 | [0.03 0.09] | 7.29E-05 | 0.001085453 |
| XL_VLDL_CE | Lipodystrophy_2_pPS | 4255 | -0.002209262 | [-0.03 0.03] | 0.885606799 | 0.946315535 |
| XL_VLDL_CE | Liver-Lipid_pPS | 4255 | -0.041314452 | [-0.07 -0.01] | 0.007112937 | 0.036069149 |
| XL_VLDL_CE | ASAT_Pos_pPS | 4255 | 0.047443267 | [0.02 0.08] | 0.001993463 | 0.013934058 |
| XL_VLDL_CE | ALP_Neg_pPS | 4255 | 0.016928344 | [-0.01 0.05] | 0.270271349 | 0.554396235 |
| XL_VLDL_CE | SHBG-LpA_pPS | 4255 | -0.00078037 | [-0.03 0.03] | 0.959471334 | 0.980965687 |
| XL_VLDL_CE | Obesity_pPS | 4255 | 0.028687336 | [-0. 0.06] | 0.061695388 | 0.204855428 |
| XL_VLDL_CE | VAT_Neg_pPS | 4255 | 0.030568962 | [0. 0.06] | 0.046471076 | 0.166134098 |
| XL_VLDL_CE | Bilirubin_pPS | 4255 | -0.013722582 | [-0.04 0.02] | 0.371512088 | 0.643918869 |
| XL_VLDL_CE | Proinsulin_pPS | 4255 | 0.011626716 | [-0.02 0.04] | 0.448961718 | 0.694281993 |
| XL_VLDL_CE | Hyper_Insulin_pPS | 4255 | 0.014980829 | [-0.02 0.05] | 0.329266102 | 0.610965216 |
| XL_VLDL_CE | Beta_Cell_1_pPS | 4255 | 0.0134394 | [-0.02 0.04] | 0.381464312 | 0.652504744 |
| XL_VLDL_CE | Beta_Cell_2_pPS | 4255 | 0.001966129 | [-0.03 0.03] | 0.898123081 | 0.949937874 |
| XL_VLDL_CE | Total_GRS | 4255 | 0.035916034 | [0.01 0.07] | 0.019305676 | 0.082821349 |
| XL_VLDL_FC | Cholesterol_pPS | 4255 | -0.093703175 | [-0.12 -0.06] | 9.64E-10 | 3.39E-08 |
| XL_VLDL_FC | Lipodystrophy_1_pPS | 4255 | 0.058104611 | [0.03 0.09] | 0.000152526 | 0.001913261 |
| XL_VLDL_FC | Lipodystrophy_2_pPS | 4255 | -0.008645327 | [-0.04 0.02] | 0.573443994 | 0.772791937 |
| XL_VLDL_FC | Liver-Lipid_pPS | 4255 | -0.046835551 | [-0.08 -0.02] | 0.002276521 | 0.015355779 |
| XL_VLDL_FC | ASAT_Pos_pPS | 4255 | 0.041954222 | [0.01 0.07] | 0.006271738 | 0.032652617 |
| XL_VLDL_FC | ALP_Neg_pPS | 4255 | 0.016937954 | [-0.01 0.05] | 0.269999442 | 0.554396235 |
| XL_VLDL_FC | SHBG-LpA_pPS | 4255 | 0.002422912 | [-0.03 0.03] | 0.874631635 | 0.942282701 |
| XL_VLDL_FC | Obesity_pPS | 4255 | 0.030150966 | [0. 0.06] | 0.049547385 | 0.173093062 |
| XL_VLDL_FC | VAT_Neg_pPS | 4255 | 0.026507158 | [-0. 0.06] | 0.084270773 | 0.262733732 |
| XL_VLDL_FC | Bilirubin_pPS | 4255 | -0.014772263 | [-0.04 0.02] | 0.336044957 | 0.618195912 |
| XL_VLDL_FC | Proinsulin_pPS | 4255 | 0.009615019 | [-0.02 0.04] | 0.531225717 | 0.745732436 |
| XL_VLDL_FC | Hyper_Insulin_pPS | 4255 | 0.010119414 | [-0.02 0.04] | 0.50990592 | 0.7313594 |
| XL_VLDL_FC | Beta_Cell_1_pPS | 4255 | 0.014616045 | [-0.02 0.04] | 0.341180749 | 0.623975243 |
| XL_VLDL_FC | Beta_Cell_2_pPS | 4255 | -0.001074368 | [-0.03 0.03] | 0.944223982 | 0.973730981 |
| XL_VLDL_FC | Total_GRS | 4255 | 0.03248472 | [0. 0.06] | 0.03435035 | 0.131105874 |
| XL_VLDL_L | Cholesterol_pPS | 4255 | -0.08336361 | [-0.11 -0.05] | 5.39E-08 | 1.43E-06 |
| XL_VLDL_L | Lipodystrophy_1_pPS | 4255 | 0.059507917 | [0.03 0.09] | 0.000105085 | 0.00147325 |
| XL_VLDL_L | Lipodystrophy_2_pPS | 4255 | 0.000889265 | [-0.03 0.03] | 0.953821784 | 0.97751922 |
| XL_VLDL_L | Liver-Lipid_pPS | 4255 | -0.051311162 | [-0.08 -0.02] | 0.000827167 | 0.007454928 |
| XL_VLDL_L | ASAT_Pos_pPS | 4255 | 0.047287773 | [0.02 0.08] | 0.002062638 | 0.014318313 |

**b) Partial Correlation**

| Metabolomics | PPS | Samp | r | CI95% | p | fdr_p_adjusted |
|---|---|---|---|---|---|---|
| XL_VLDL_PL | VAT_Neg_pPS | 4255 | 0.019330321 | [-0.01 0.05] | 0.208768124 | 0.495793339 |
| XL_VLDL_PL | Bilirubin_pPS | 4255 | -0.016368592 | [-0.05 0.01] | 0.287172669 | 0.584862344 |
| XL_VLDL_PL | Proinsulin_pPS | 4255 | 0.002810532 | [-0.03 0.03] | 0.855002824 | 0.93700433 |
| XL_VLDL_PL | Hyper_Insulin_pPS | 4255 | 0.003296877 | [-0.03 0.03] | 0.830265583 | 0.921392293 |
| XL_VLDL_PL | Beta_Cell_1_pPS | 4255 | 0.00937283 | [-0.02 0.04] | 0.542241085 | 0.787936618 |
| XL_VLDL_PL | Beta_Cell_2_pPS | 4255 | -0.005015148 | [-0.04 0.03] | 0.744361473 | 0.873512116 |
| XL_VLDL_TG | Cholesterol_pPS | 4255 | -0.07952837 | [-0.11 -0.05] | 2.23E-07 | 4.96E-06 |
| XL_VLDL_TG | Lipodystrophy_1_pPS | 4255 | 0.05013035 | [0.02 0.08] | 0.001108294 | 0.009401714 |
| XL_VLDL_TG | Lipodystrophy_2_pPS | 4255 | 0.000453784 | [-0.03 0.03] | 0.976462089 | 0.991343605 |
| XL_VLDL_TG | Liver-Lipid_pPS | 4255 | -0.055001372 | [-0.08 -0.02] | 0.000345102 | 0.003970654 |
| XL_VLDL_TG | ASAT_Pos_pPS | 4255 | 0.036307124 | [0.01 0.07] | 0.018204132 | 0.084362666 |
| XL_VLDL_TG | ALP_Neg_pPS | 4255 | 0.001770994 | [-0.03 0.03] | 0.908327227 | 0.96224905 |
| XL_VLDL_TG | SHBG-LpA_pPS | 4255 | -0.007728871 | [-0.04 0.02] | 0.615292814 | 0.828794354 |
| XL_VLDL_TG | Obesity_pPS | 4255 | 0.037640571 | [0.01 0.07] | 0.01435606 | 0.071317202 |
| XL_VLDL_TG | VAT_Neg_pPS | 4255 | 0.017570066 | [-0.01 0.05] | 0.25325442 | 0.549908188 |
| XL_VLDL_TG | Bilirubin_pPS | 4255 | -0.013945671 | [-0.04 0.02] | 0.3645227 | 0.650898451 |
| XL_VLDL_TG | Proinsulin_pPS | 4255 | 0.004615995 | [-0.03 0.03] | 0.764077962 | 0.885222315 |
| XL_VLDL_TG | Hyper_Insulin_pPS | 4255 | 0.006120629 | [-0.02 0.04] | 0.690657957 | 0.858568103 |
| XL_VLDL_TG | Beta_Cell_1_pPS | 4255 | 0.005892403 | [-0.02 0.04] | 0.701628704 | 0.859880159 |
| XL_VLDL_TG | Beta_Cell_2_pPS | 4255 | -0.006319563 | [-0.04 0.02] | 0.681147902 | 0.858274475 |
| XS_VLDL_C | Cholesterol_pPS | 4255 | -0.126419714 | [-0.16 -0.1 ] | 1.55E-16 | 8.60E-15 |
| XS_VLDL_C | Lipodystrophy_1_pPS | 4255 | 0.046908428 | [0.02 0.08] | 0.002276028 | 0.016876323 |
| XS_VLDL_C | Lipodystrophy_2_pPS | 4255 | -0.038024803 | [-0.07 -0.01] | 0.013389503 | 0.067862747 |
| XS_VLDL_C | Liver-Lipid_pPS | 4255 | 0.019675091 | [-0.01 0.05] | 0.200762202 | 0.486527589 |
| XS_VLDL_C | ASAT_Pos_pPS | 4255 | -0.009702179 | [-0.04 0.02] | 0.52814315 | 0.77834791 |
| XS_VLDL_C | ALP_Neg_pPS | 4255 | 0.056808759 | [0.03 0.09] | 0.000218466 | 0.002803644 |
| XS_VLDL_C | SHBG-LpA_pPS | 4255 | 0.011898387 | [-0.02 0.04] | 0.439137018 | 0.703321847 |
| XS_VLDL_C | Obesity_pPS | 4255 | 0.006156761 | [-0.02 0.04] | 0.688926982 | 0.858568103 |
| XS_VLDL_C | VAT_Neg_pPS | 4255 | 0.010931584 | [-0.02 0.04] | 0.477217844 | 0.737518525 |
| XS_VLDL_C | Bilirubin_pPS | 4255 | -0.013106372 | [-0.04 0.02] | 0.394101559 | 0.672569903 |
| XS_VLDL_C | Proinsulin_pPS | 4255 | -0.008905701 | [-0.04 0.02] | 0.562553231 | 0.797755947 |
| XS_VLDL_C | Hyper_Insulin_pPS | 4255 | -0.01774425 | [-0.05 0.01] | 0.248578772 | 0.545779139 |
| XS_VLDL_C | Beta_Cell_1_pPS | 4255 | 0.005475586 | [-0.02 0.04] | 0.721824823 | 0.866319132 |
| XS_VLDL_C | Beta_Cell_2_pPS | 4255 | 0.023415826 | [-0.01 0.05] | 0.127836866 | 0.380847331 |
| XS_VLDL_CE | Cholesterol_pPS | 4255 | -0.126155637 | [-0.16 -0.1 ] | 1.79E-16 | 9.68E-15 |
| XS_VLDL_CE | Lipodystrophy_1_pPS | 4255 | 0.043528158 | [0.01 0.07] | 0.00463289 | 0.029166809 |
| XS_VLDL_CE | Lipodystrophy_2_pPS | 4255 | -0.040502418 | [-0.07 -0.01] | 0.00842571 | 0.047625902 |
| XS_VLDL_CE | Liver-Lipid_pPS | 4255 | 0.022960065 | [-0.01 0.05] | 0.13542606 | 0.399886389 |
| XS_VLDL_CE | ASAT_Pos_pPS | 4255 | -0.004627227 | [-0.03 0.03] | 0.763520993 | 0.885222315 |
| XS_VLDL_CE | ALP_Neg_pPS | 4255 | 0.05527634 | [0.03 0.09] | 0.000322187 | 0.003826979 |
| XS_VLDL_CE | SHBG-LpA_pPS | 4255 | 0.011350817 | [-0.02 0.04] | 0.460487023 | 0.723622464 |
| XS_VLDL_CE | Obesity_pPS | 4255 | 0.006301508 | [-0.02 0.04] | 0.68200897 | 0.858568103 |
| XS_VLDL_CE | VAT_Neg_pPS | 4255 | 0.013135916 | [-0.02 0.04] | 0.39303632 | 0.671954495 |
| XS_VLDL_CE | Bilirubin_pPS | 4255 | -0.014370947 | [-0.04 0.02] | 0.350078965 | 0.637861136 |
| XS_VLDL_CE | Proinsulin_pPS | 4255 | -0.008824348 | [-0.04 0.02] | 0.566127874 | 0.797755947 |
| XS_VLDL_CE | Hyper_Insulin_pPS | 4255 | -0.017941105 | [-0.05 0.01] | 0.243367636 | 0.540756944 |
| XS_VLDL_CE | Beta_Cell_1_pPS | 4255 | 0.005835136 | [-0.02 0.04] | 0.704391373 | 0.861448704 |
| XS_VLDL_CE | Beta_Cell_2_pPS | 4255 | 0.021955858 | [-0.01 0.05] | 0.153371011 | 0.428688101 |
| XS_VLDL_FC | Cholesterol_pPS | 4255 | -0.123364177 | [-0.15 -0.09] | 8.16E-16 | 4.30E-14 |
| XS_VLDL_FC | Lipodystrophy_1_pPS | 4255 | 0.052734283 | [0.02 0.08] | 0.000601073 | 0.006202825 |
| XS_VLDL_FC | Lipodystrophy_2_pPS | 4255 | -0.029712994 | [-0.06 0. ] | 0.053317236 | 0.196576625 |
| XS_VLDL_FC | Liver-Lipid_pPS | 4255 | 0.011389889 | [-0.02 0.04] | 0.458944624 | 0.72261785 |
| XS_VLDL_FC | ASAT_Pos_pPS | 4255 | -0.020423605 | [-0.05 0.01] | 0.18415645 | 0.466062214 |
| XS_VLDL_FC | ALP_Neg_pPS | 4255 | 0.057332786 | [0.03 0.09] | 0.000190871 | 0.00249754 |
| XS_VLDL_FC | SHBG-LpA_pPS | 4255 | 0.01218628 | [-0.02 0.04] | 0.428143817 | 0.696591166 |
| XS_VLDL_FC | Obesity_pPS | 4255 | 0.004869548 | [-0.03 0.04] | 0.751534904 | 0.876790721 |
| XS_VLDL_FC | VAT_Neg_pPS | 4255 | 0.005761694 | [-0.02 0.04] | 0.707940135 | 0.863152345 |
| XS_VLDL_FC | Bilirubin_pPS | 4255 | -0.010845544 | [-0.04 0.02] | 0.480692214 | 0.740874464 |
| XS_VLDL_FC | Proinsulin_pPS | 4255 | -0.007727036 | [-0.04 0.02] | 0.615376739 | 0.828794354 |
| XS_VLDL_FC | Hyper_Insulin_pPS | 4255 | -0.016471066 | [-0.05 0.01] | 0.284165537 | 0.583442529 |
| XS_VLDL_FC | Beta_Cell_1_pPS | 4255 | 0.004889961 | [-0.03 0.04] | 0.750527874 | 0.876790721 |
| XS_VLDL_FC | Beta_Cell_2_pPS | 4255 | 0.024835216 | [-0.01 0.05] | 0.106307305 | 0.338358068 |
| XS_VLDL_L | Cholesterol_pPS | 4255 | -0.132279604 | [-0.16 -0.1 ] | 5.69E-18 | 3.45E-16 |
| XS_VLDL_L | Lipodystrophy_1_pPS | 4255 | 0.061147344 | [0.03 0.09] | 6.91E-05 | 0.001088757 |
| XS_VLDL_L | Lipodystrophy_2_pPS | 4255 | -0.027842724 | [-0.06 0. ] | 0.070192965 | 0.243125115 |
| XS_VLDL_L | Liver-Lipid_pPS | 4255 | 0.003646865 | [-0.03 0.03] | 0.812565922 | 0.911348446 |
| XS_VLDL_L | ASAT_Pos_pPS | 4255 | -0.008662846 | [-0.04 0.02] | 0.573256395 | 0.800878787 |
| XS_VLDL_L | ALP_Neg_pPS | 4255 | 0.051595975 | [0.02 0.08] | 0.000788027 | 0.007372102 |
| XS_VLDL_L | SHBG-LpA_pPS | 4255 | 0.010141081 | [-0.02 0.04] | 0.50965035 | 0.766145193 |
| XS_VLDL_L | Obesity_pPS | 4255 | 0.013563394 | [-0.02 0.04] | 0.377819003 | 0.658306043 |
| XS_VLDL_L | VAT_Neg_pPS | 4255 | 0.007789108 | [-0.02 0.04] | 0.612541131 | 0.828794354 |
| XS_VLDL_L | Bilirubin_pPS | 4255 | -0.015335907 | [-0.05 0.01] | 0.318673532 | 0.61940234 |
| XS_VLDL_L | Proinsulin_pPS | 4255 | -0.004161317 | [-0.03 0.03] | 0.786724228 | 0.897448379 |
| XS_VLDL_L | Hyper_Insulin_pPS | 4255 | -0.016470483 | [-0.05 0.01] | 0.284182601 | 0.583442529 |
| XS_VLDL_L | Beta_Cell_1_pPS | 4255 | 0.005217342 | [-0.02 0.04] | 0.73443656 | 0.871055684 |
| XS_VLDL_L | Beta_Cell_2_pPS | 4255 | 0.019980022 | [-0.01 0.05] | 0.193870049 | 0.478398712 |
| XS_VLDL_P | Cholesterol_pPS | 4255 | -0.131917651 | [-0.16 -0.1 ] | 7.00E-18 | 4.12E-16 |
| XS_VLDL_P | Lipodystrophy_1_pPS | 4255 | 0.064029106 | [0.03 0.09] | 3.08E-05 | 0.000575864 |
| XS_VLDL_P | Lipodystrophy_2_pPS | 4255 | -0.02514884 | [-0.06 0. ] | 0.101961708 | 0.329769532 |
| XS_VLDL_P | Liver-Lipid_pPS | 4255 | -0.000102772 | [-0.03 0.03] | 0.994668443 | 0.996161192 |
| XS_VLDL_P | ASAT_Pos_pPS | 4255 | -0.006630523 | [-0.04 0.02] | 0.666383862 | 0.849453588 |
| XS_VLDL_P | ALP_Neg_pPS | 4255 | 0.04912644 | [0.02 0.08] | 0.001393022 | 0.011476667 |
| XS_VLDL_P | SHBG-LpA_pPS | 4255 | 0.008903218 | [-0.02 0.04] | 0.562662142 | 0.797755947 |
| XS_VLDL_P | Obesity_pPS | 4255 | 0.015264544 | [-0.01 0.05] | 0.320930901 | 0.620672596 |
| XS_VLDL_P | VAT_Neg_pPS | 4255 | 0.007504565 | [-0.02 0.04] | 0.625586745 | 0.833837991 |
| XS_VLDL_P | Bilirubin_pPS | 4255 | -0.016329656 | [-0.05 0.01] | 0.288320857 | 0.586008484 |
| XS_VLDL_P | Proinsulin_pPS | 4255 | -0.002582093 | [-0.03 0.03] | 0.866672976 | 0.943130556 |
| XS_VLDL_P | Hyper_Insulin_pPS | 4255 | -0.015585336 | [-0.05 0.01] | 0.3108653 | 0.612551507 |
| XS_VLDL_P | Beta_Cell_1_pPS | 4255 | 0.005016161 | [-0.03 0.04] | 0.744311668 | 0.873512116 |
| XS_VLDL_P | Beta_Cell_2_pPS | 4255 | 0.018134224 | [-0.01 0.05] | 0.238330421 | 0.534907515 |
| XS_VLDL_PL | Cholesterol_pPS | 4255 | -0.143576364 | [-0.17 -0.11] | 6.36E-21 | 5.09E-19 |
| XS_VLDL_PL | Lipodystrophy_1_pPS | 4255 | 0.055796096 | [0.03 0.09] | 0.000282711 | 0.003430229 |
| XS_VLDL_PL | Lipodystrophy_2_pPS | 4255 | -0.034093297 | [-0.06 -0. ] | 0.0265982 | 0.113056469 |
| XS_VLDL_PL | Liver-Lipid_pPS | 4255 | 0.010902558 | [-0.02 0.04] | 0.478388398 | 0.738422184 |

**Table S9: Associations between pPSs and global metabolomic profile in SCAPIS**

**a) Spearman Correlation**

| Metabolomics | PPS | Samp | r | CI95% | p | fdr_p_adjusted |
|---|---|---|---|---|---|---|
| XL_VLDL_L | ALP_Neg_pPS | 4255 | 0.006551036 | [-0.02 0.04] | 0.66966904 | 0.82983252 |
| XL_VLDL_L | SHBG-LpA_pPS | 4255 | -0.000525658 | [-0.03 0.03] | 0.97269342 | 0.983305352 |
| XL_VLDL_L | Obesity_pPS | 4255 | 0.03305583 | [0. 0.06] | 0.031306501 | 0.122540959 |
| XL_VLDL_L | VAT_Neg_pPS | 4255 | 0.023942063 | [-0.01 0.05] | 0.118923115 | 0.345182791 |
| XL_VLDL_L | Bilirubin_pPS | 4255 | -0.011934139 | [-0.04 0.02] | 0.437059475 | 0.690884659 |
| XL_VLDL_L | Proinsulin_pPS | 4255 | 0.00917721 | [-0.02 0.04] | 0.550090557 | 0.760607725 |
| XL_VLDL_L | Hyper_Insulin_pPS | 4255 | 0.014703303 | [-0.02 0.04] | 0.338305915 | 0.620758073 |
| XL_VLDL_L | Beta_Cell_1_pPS | 4255 | 0.012031443 | [-0.02 0.04] | 0.4333304 | 0.689535391 |
| XL_VLDL_L | Beta_Cell_2_pPS | 4255 | -0.002971067 | [-0.03 0.03] | 0.846588026 | 0.92810697 |
| XL_VLDL_L | Total_GRS | 4255 | 0.036708704 | [0.01 0.07] | 0.016791368 | 0.07308884 |
| XL_VLDL_P | Cholesterol_pPS | 4255 | -0.082115763 | [-0.11 -0.05] | 8.49E-08 | 2.22E-06 |
| XL_VLDL_P | Lipodystrophy_1_pPS | 4255 | 0.059411187 | [0.03 0.09] | 0.000107846 | 0.001491835 |
| XL_VLDL_P | Lipodystrophy_2_pPS | 4255 | 0.001899248 | [-0.03 0.03] | 0.901570629 | 0.95179735 |
| XL_VLDL_P | Liver-Lipid_pPS | 4255 | -0.052093976 | [-0.08 -0.02] | 0.000687242 | 0.006600154 |
| XL_VLDL_P | ASAT_Pos_pPS | 4255 | 0.047483035 | [0.02 0.08] | 0.001976116 | 0.013934058 |
| XL_VLDL_P | ALP_Neg_pPS | 4255 | 0.005518254 | [-0.02 0.04] | 0.719335809 | 0.865374699 |
| XL_VLDL_P | SHBG-LpA_pPS | 4255 | -0.000814506 | [-0.03 0.03] | 0.957700084 | 0.979621688 |
| XL_VLDL_P | Obesity_pPS | 4255 | 0.03333178 | [0. 0.06] | 0.029920632 | 0.118780144 |
| XL_VLDL_P | VAT_Neg_pPS | 4255 | 0.02354775 | [-0.01 0.05] | 0.125122816 | 0.357416198 |
| XL_VLDL_P | Bilirubin_pPS | 4255 | -0.011562599 | [-0.04 0.02] | 0.451467079 | 0.694689301 |
| XL_VLDL_P | Proinsulin_pPS | 4255 | 0.008918653 | [-0.02 0.04] | 0.561384413 | 0.767334778 |
| XL_VLDL_P | Hyper_Insulin_pPS | 4255 | 0.014878102 | [-0.02 0.04] | 0.332593815 | 0.613626471 |
| XL_VLDL_P | Beta_Cell_1_pPS | 4255 | 0.011709502 | [-0.02 0.04] | 0.445738596 | 0.693159645 |
| XL_VLDL_P | Beta_Cell_2_pPS | 4255 | -0.003455627 | [-0.03 0.03] | 0.821957435 | 0.919717631 |
| XL_VLDL_P | Total_GRS | 4255 | 0.036925548 | [0.01 0.07] | 0.016155679 | 0.070866934 |
| XL_VLDL_PL | Cholesterol_pPS | 4255 | -0.091428603 | [-0.12 -0.06] | 2.43E-09 | 8.27E-08 |
| XL_VLDL_PL | Lipodystrophy_1_pPS | 4255 | 0.059516619 | [0.03 0.09] | 0.00010484 | 0.00147325 |
| XL_VLDL_PL | Lipodystrophy_2_pPS | 4255 | -0.004774206 | [-0.03 0.03] | 0.75588129 | 0.884059633 |
| XL_VLDL_PL | Liver-Lipid_pPS | 4255 | -0.049141197 | [-0.08 -0.02] | 0.001364931 | 0.010685316 |
| XL_VLDL_PL | ASAT_Pos_pPS | 4255 | 0.04402548 | [0.01 0.07] | 0.004126954 | 0.02354683 |
| XL_VLDL_PL | ALP_Neg_pPS | 4255 | 0.0118807 | [-0.02 0.04] | 0.439115275 | 0.691142319 |
| XL_VLDL_PL | SHBG-LpA_pPS | 4255 | 0.001015161 | [-0.03 0.03] | 0.947293124 | 0.974500144 |
| XL_VLDL_PL | Obesity_pPS | 4255 | 0.031498858 | [0. 0.06] | 0.040200084 | 0.149185433 |
| XL_VLDL_PL | VAT_Neg_pPS | 4255 | 0.02353646 | [-0.01 0.05] | 0.125303954 | 0.357416198 |
| XL_VLDL_PL | Bilirubin_pPS | 4255 | -0.012690585 | [-0.04 0.02] | 0.408558485 | 0.671770679 |
| XL_VLDL_PL | Proinsulin_pPS | 4255 | 0.009175695 | [-0.02 0.04] | 0.550156388 | 0.760607725 |
| XL_VLDL_PL | Hyper_Insulin_pPS | 4255 | 0.012278513 | [-0.02 0.04] | 0.423944843 | 0.684769436 |
| XL_VLDL_PL | Beta_Cell_1_pPS | 4255 | 0.014567367 | [-0.02 0.04] | 0.342791273 | 0.626309439 |
| XL_VLDL_PL | Beta_Cell_2_pPS | 4255 | -0.00238227 | [-0.03 0.03] | 0.876717609 | 0.943108963 |
| XL_VLDL_PL | Total_GRS | 4255 | 0.034582079 | [0. 0.06] | 0.024282591 | 0.099973429 |
| XL_VLDL_TG | Cholesterol_pPS | 4255 | -0.077925896 | [-0.11 -0.05] | 3.73E-07 | 8.99E-06 |
| XL_VLDL_TG | Lipodystrophy_1_pPS | 4255 | 0.058972739 | [0.03 0.09] | 0.000121243 | 0.001604501 |
| XL_VLDL_TG | Lipodystrophy_2_pPS | 4255 | 0.004582576 | [-0.03 0.03] | 0.765386549 | 0.889697961 |
| XL_VLDL_TG | Liver-Lipid_pPS | 4255 | -0.054464242 | [-0.08 -0.02] | 0.00038626 | 0.004184479 |
| XL_VLDL_TG | ASAT_Pos_pPS | 4255 | 0.04797811 | [0.02 0.08] | 0.001771458 | 0.01292441 |
| XL_VLDL_TG | ALP_Neg_pPS | 4255 | 0.002160438 | [-0.03 0.03] | 0.888118039 | 0.946826466 |
| XL_VLDL_TG | SHBG-LpA_pPS | 4255 | -0.001641406 | [-0.03 0.03] | 0.914878617 | 0.957275431 |
| XL_VLDL_TG | Obesity_pPS | 4255 | 0.0342553 | [0. 0.06] | 0.025659287 | 0.104348709 |
| XL_VLDL_TG | VAT_Neg_pPS | 4255 | 0.021979283 | [-0.01 0.05] | 0.152302861 | 0.402823226 |
| XL_VLDL_TG | Bilirubin_pPS | 4255 | -0.010267817 | [-0.04 0.02] | 0.503719495 | 0.727744052 |
| XL_VLDL_TG | Proinsulin_pPS | 4255 | 0.008367253 | [-0.02 0.04] | 0.585837638 | 0.779730867 |
| XL_VLDL_TG | Hyper_Insulin_pPS | 4255 | 0.015416049 | [-0.01 0.05] | 0.31540792 | 0.60298573 |
| XL_VLDL_TG | Beta_Cell_1_pPS | 4255 | 0.010493714 | [-0.02 0.04] | 0.494379224 | 0.722332865 |
| XL_VLDL_TG | Beta_Cell_2_pPS | 4255 | -0.004518206 | [-0.03 0.03] | 0.768587435 | 0.889703209 |
| XL_VLDL_TG | Total_GRS | 4255 | 0.037547425 | [0.01 0.07] | 0.014448395 | 0.064032658 |
| XS_VLDL_C | Cholesterol_pPS | 4255 | -0.127434826 | [-0.16 -0.1 ] | 7.92E-17 | 4.72E-15 |
| XS_VLDL_C | Lipodystrophy_1_pPS | 4255 | 0.04374437 | [0.01 0.07] | 0.004372485 | 0.024735931 |
| XS_VLDL_C | Lipodystrophy_2_pPS | 4255 | -0.039785482 | [-0.07 -0.01] | 0.009546921 | 0.045608344 |
| XS_VLDL_C | Liver-Lipid_pPS | 4255 | 0.018418533 | [-0.01 0.05] | 0.230333412 | 0.513581257 |
| XS_VLDL_C | ASAT_Pos_pPS | 4255 | -0.005378482 | [-0.04 0.02] | 0.726155148 | 0.870169158 |
| XS_VLDL_C | ALP_Neg_pPS | 4255 | 0.056598245 | [0.03 0.09] | 0.000225505 | 0.002657736 |
| XS_VLDL_C | SHBG-LpA_pPS | 4255 | 0.015307566 | [-0.01 0.05] | 0.318825829 | 0.60735471 |
| XS_VLDL_C | Obesity_pPS | 4255 | -0.004231004 | [-0.03 0.03] | 0.782916359 | 0.897511134 |
| XS_VLDL_C | VAT_Neg_pPS | 4255 | 0.013350516 | [-0.02 0.04] | 0.384621468 | 0.654405057 |
| XS_VLDL_C | Bilirubin_pPS | 4255 | -0.012014652 | [-0.04 0.02] | 0.433972603 | 0.690045392 |
| XS_VLDL_C | Proinsulin_pPS | 4255 | 0.005392938 | [-0.02 0.04] | 0.725448857 | 0.870169158 |
| XS_VLDL_C | Hyper_Insulin_pPS | 4255 | -0.013537092 | [-0.04 0.02] | 0.37801267 | 0.649189092 |
| XS_VLDL_C | Beta_Cell_1_pPS | 4255 | 0.014467854 | [-0.02 0.04] | 0.346098816 | 0.62662761 |
| XS_VLDL_C | Beta_Cell_2_pPS | 4255 | 0.024518368 | [-0.01 0.05] | 0.110298122 | 0.324539741 |
| XS_VLDL_C | Total_GRS | 4255 | 0.004198252 | [-0.03 0.03] | 0.784555238 | 0.897605875 |
| XS_VLDL_CE | Cholesterol_pPS | 4255 | -0.127322007 | [-0.16 -0.1 ] | 8.43E-17 | 4.89E-15 |
| XS_VLDL_CE | Lipodystrophy_1_pPS | 4255 | 0.041053618 | [0.01 0.07] | 0.00748401 | 0.037534473 |
| XS_VLDL_CE | Lipodystrophy_2_pPS | 4255 | -0.041675074 | [-0.07 -0.01] | 0.006627103 | 0.034170999 |
| XS_VLDL_CE | Liver-Lipid_pPS | 4255 | 0.021579933 | [-0.01 0.05] | 0.159893476 | 0.417240274 |
| XS_VLDL_CE | ASAT_Pos_pPS | 4255 | -0.001035058 | [-0.03 0.03] | 0.946261614 | 0.974500144 |
| XS_VLDL_CE | ALP_Neg_pPS | 4255 | 0.055114299 | [0.03 0.09] | 0.000328497 | 0.00363209 |
| XS_VLDL_CE | SHBG-LpA_pPS | 4255 | 0.014709753 | [-0.02 0.04] | 0.338094011 | 0.620758073 |
| XS_VLDL_CE | Obesity_pPS | 4255 | -0.004025034 | [-0.03 0.03] | 0.793238375 | 0.90169386 |
| XS_VLDL_CE | VAT_Neg_pPS | 4255 | 0.015123133 | [-0.01 0.05] | 0.324692142 | 0.608687153 |
| XS_VLDL_CE | Bilirubin_pPS | 4255 | -0.013240423 | [-0.04 0.02] | 0.38855409 | 0.657347349 |
| XS_VLDL_CE | Proinsulin_pPS | 4255 | 0.005225418 | [-0.02 0.04] | 0.733647986 | 0.873778417 |
| XS_VLDL_CE | Hyper_Insulin_pPS | 4255 | -0.013615606 | [-0.04 0.02] | 0.375252622 | 0.646000701 |
| XS_VLDL_CE | Beta_Cell_1_pPS | 4255 | 0.014930915 | [-0.02 0.05] | 0.330880311 | 0.612969968 |
| XS_VLDL_CE | Beta_Cell_2_pPS | 4255 | 0.023283325 | [-0.01 0.05] | 0.129419447 | 0.36526936 |
| XS_VLDL_CE | Total_GRS | 4255 | 0.003791814 | [-0.03 0.03] | 0.804969787 | 0.909247074 |
| XS_VLDL_FC | Cholesterol_pPS | 4255 | -0.124055571 | [-0.15 -0.09] | 5.07E-16 | 2.86E-14 |
| XS_VLDL_FC | Lipodystrophy_1_pPS | 4255 | 0.048233394 | [0.02 0.08] | 0.001673713 | 0.01249255 |
| XS_VLDL_FC | Lipodystrophy_2_pPS | 4255 | -0.032746438 | [-0.06 -0. ] | 0.032925439 | 0.127023499 |
| XS_VLDL_FC | Liver-Lipid_pPS | 4255 | 0.010482305 | [-0.02 0.04] | 0.494848731 | 0.722332865 |
| XS_VLDL_FC | ASAT_Pos_pPS | 4255 | -0.014499422 | [-0.04 0.02] | 0.3450474 | 0.62662761 |
| XS_VLDL_FC | ALP_Neg_pPS | 4255 | 0.057213619 | [0.03 0.09] | 0.000192428 | 0.002357661 |
| XS_VLDL_FC | SHBG-LpA_pPS | 4255 | 0.01571202 | [-0.01 0.05] | 0.306205696 | 0.592255382 |
| XS_VLDL_FC | Obesity_pPS | 4255 | -0.005254963 | [-0.04 0.02] | 0.732199677 | 0.872537948 |
| XS_VLDL_FC | VAT_Neg_pPS | 4255 | 0.009249195 | [-0.02 0.04] | 0.546966257 | 0.760607725 |

**b) Partial Correlation**

| Metabolomics | PPS | Samp | r | CI95% | p | fdr_p_adjusted |
|---|---|---|---|---|---|---|
| XS_VLDL_PL | ASAT_Pos_pPS | 4255 | -0.019362915 | [-0.05 0.01] | 0.208001499 | 0.495147445 |
| XS_VLDL_PL | ALP_Neg_pPS | 4255 | 0.055265575 | [0.03 0.09] | 0.000323057 | 0.003826979 |
| XS_VLDL_PL | SHBG-LpA_pPS | 4255 | 0.013837446 | [-0.02 0.04] | 0.368256951 | 0.653011884 |
| XS_VLDL_PL | Obesity_pPS | 4255 | 0.010990854 | [-0.02 0.04] | 0.474832482 | 0.735233474 |
| XS_VLDL_PL | VAT_Neg_pPS | 4255 | 0.00717743 | [-0.02 0.04] | 0.64073102 | 0.836753752 |
| XS_VLDL_PL | Bilirubin_pPS | 4255 | -0.010254297 | [-0.04 0.02] | 0.504935684 | 0.761205753 |
| XS_VLDL_PL | Proinsulin_pPS | 4255 | -0.009994226 | [-0.04 0.02] | 0.515800035 | 0.770045988 |
| XS_VLDL_PL | Hyper_Insulin_pPS | 4255 | -0.018862492 | [-0.05 0.01] | 0.219997861 | 0.513666802 |
| XS_VLDL_PL | Beta_Cell_1_pPS | 4255 | 0.006351827 | [-0.02 0.04] | 0.679610215 | 0.857328072 |
| XS_VLDL_PL | Beta_Cell_2_pPS | 4255 | 0.023025334 | [-0.01 0.05] | 0.134318353 | 0.397788969 |
| XS_VLDL_TG | Cholesterol_pPS | 4255 | -0.093042156 | [-0.12 -0.06] | 1.34E-09 | 4.25E-08 |
| XS_VLDL_TG | Lipodystrophy_1_pPS | 4255 | 0.071797464 | [0.04 0.1 ] | 2.95E-06 | 6.09E-05 |
| XS_VLDL_TG | Lipodystrophy_2_pPS | 4255 | 0.013881452 | [-0.02 0.04] | 0.366735695 | 0.651468378 |
| XS_VLDL_TG | Liver-Lipid_pPS | 4255 | -0.033423216 | [-0.06 -0. ] | 0.029723241 | 0.122035016 |
| XS_VLDL_TG | ASAT_Pos_pPS | 4255 | 0.013225933 | [-0.02 0.04] | 0.389801395 | 0.668708134 |
| XS_VLDL_TG | ALP_Neg_pPS | 4255 | 0.020217481 | [-0.01 0.05] | 0.1886244 | 0.471443258 |
| XS_VLDL_TG | SHBG-LpA_pPS | 4255 | -0.006809554 | [-0.04 0.02] | 0.657941458 | 0.844628798 |
| XS_VLDL_TG | Obesity_pPS | 4255 | 0.025964426 | [-0. 0.06] | 0.091320741 | 0.303190917 |
| XS_VLDL_TG | VAT_Neg_pPS | 4255 | 0.002644273 | [-0.03 0.03] | 0.86349344 | 0.941565287 |
| XS_VLDL_TG | Bilirubin_pPS | 4255 | -0.007119927 | [-0.04 0.02] | 0.643408809 | 0.839155984 |
| XS_VLDL_TG | Proinsulin_pPS | 4255 | 0.009437331 | [-0.02 0.04] | 0.539465379 | 0.784890762 |
| XS_VLDL_TG | Hyper_Insulin_pPS | 4255 | -0.001309398 | [-0.03 0.03] | 0.932153194 | 0.977479024 |
| XS_VLDL_TG | Beta_Cell_1_pPS | 4255 | 0.001812453 | [-0.03 0.03] | 0.906190972 | 0.96224905 |
| XS_VLDL_TG | Beta_Cell_2_pPS | 4255 | 0.000298461 | [-0.03 0.03] | 0.984517438 | 0.993363084 |
| XXL_VLDL_C | Cholesterol_pPS | 4255 | -0.09607745 | [-0.13 -0.07] | 3.82E-10 | 1.34E-08 |
| XXL_VLDL_C | Lipodystrophy_1_pPS | 4255 | 0.049397681 | [0.02 0.08] | 0.001310087 | 0.01088296 |
| XXL_VLDL_C | Lipodystrophy_2_pPS | 4255 | -0.018263203 | [-0.05 0.01] | 0.23500744 | 0.530670632 |
| XXL_VLDL_C | Liver-Lipid_pPS | 4255 | -0.039901867 | [-0.07 -0.01] | 0.00944748 | 0.051677198 |
| XXL_VLDL_C | ASAT_Pos_pPS | 4255 | 0.030968418 | [0. 0.06] | 0.044006837 | 0.166543834 |
| XXL_VLDL_C | ALP_Neg_pPS | 4255 | 0.023811934 | [-0.01 0.05] | 0.121513248 | 0.368590185 |
| XXL_VLDL_C | SHBG-LpA_pPS | 4255 | -0.004384529 | [-0.03 0.03] | 0.775582815 | 0.89083006 |
| XXL_VLDL_C | Obesity_pPS | 4255 | 0.032386511 | [0. 0.06] | 0.035178434 | 0.138783627 |
| XXL_VLDL_C | VAT_Neg_pPS | 4255 | 0.025767706 | [-0. 0.06] | 0.093801956 | 0.309376468 |
| XXL_VLDL_C | Bilirubin_pPS | 4255 | -0.018287188 | [-0.05 0.01] | 0.23439312 | 0.530670632 |
| XXL_VLDL_C | Proinsulin_pPS | 4255 | 0.002449964 | [-0.03 0.03] | 0.873436462 | 0.946048963 |
| XXL_VLDL_C | Hyper_Insulin_pPS | 4255 | 0.001827976 | [-0.03 0.03] | 0.905391285 | 0.96224905 |
| XXL_VLDL_C | Beta_Cell_1_pPS | 4255 | 0.008685569 | [-0.02 0.04] | 0.57225087 | 0.800591364 |
| XXL_VLDL_C | Beta_Cell_2_pPS | 4255 | 0.00064194 | [-0.03 0.03] | 0.966707184 | 0.986918808 |
| XXL_VLDL_CE | Cholesterol_pPS | 4255 | -0.10077493 | [-0.13 -0.07] | 5.06E-11 | 1.91E-09 |
| XXL_VLDL_CE | Lipodystrophy_1_pPS | 4255 | 0.050354691 | [0.02 0.08] | 0.001052507 | 0.009004786 |
| XXL_VLDL_CE | Lipodystrophy_2_pPS | 4255 | -0.023545217 | [-0.05 0.01] | 0.125743675 | 0.378554642 |
| XXL_VLDL_CE | Liver-Lipid_pPS | 4255 | -0.029746596 | [-0.06 0. ] | 0.053048148 | 0.195945372 |
| XXL_VLDL_CE | ASAT_Pos_pPS | 4255 | 0.029496908 | [-0. 0.06] | 0.055075064 | 0.202096449 |
| XXL_VLDL_CE | ALP_Neg_pPS | 4255 | 0.029596668 | [-0. 0.06] | 0.054257621 | 0.199676023 |
| XXL_VLDL_CE | SHBG-LpA_pPS | 4255 | -0.003148598 | [-0.03 0.03] | 0.837790952 | 0.92461824 |
| XXL_VLDL_CE | Obesity_pPS | 4255 | 0.028204705 | [-0. 0.06] | 0.066622378 | 0.234401596 |
| XXL_VLDL_CE | VAT_Neg_pPS | 4255 | 0.025932426 | [-0. 0.06] | 0.091720725 | 0.303512218 |
| XXL_VLDL_CE | Bilirubin_pPS | 4255 | -0.021753736 | [-0.05 0.01] | 0.157192106 | 0.431435756 |
| XXL_VLDL_CE | Proinsulin_pPS | 4255 | 0.001136084 | [-0.03 0.03] | 0.941115954 | 0.978430443 |
| XXL_VLDL_CE | Hyper_Insulin_pPS | 4255 | 0.001263752 | [-0.03 0.03] | 0.934512967 | 0.978170683 |
| XXL_VLDL_CE | Beta_Cell_1_pPS | 4255 | 0.008307012 | [-0.02 0.04] | 0.589110971 | 0.815104592 |
| XXL_VLDL_CE | Beta_Cell_2_pPS | 4255 | 0.005666241 | [-0.02 0.04] | 0.712561875 | 0.86476088 |
| XXL_VLDL_FC | Cholesterol_pPS | 4255 | -0.081794388 | [-0.11 -0.05] | 9.98E-08 | 2.32E-06 |
| XXL_VLDL_FC | Lipodystrophy_1_pPS | 4255 | 0.046679409 | [0.02 0.08] | 0.002391719 | 0.017438093 |
| XXL_VLDL_FC | Lipodystrophy_2_pPS | 4255 | -0.006805071 | [-0.04 0.02] | 0.65815231 | 0.844628798 |
| XXL_VLDL_FC | Liver-Lipid_pPS | 4255 | -0.05219073 | [-0.08 -0.02] | 0.000684495 | 0.006750541 |
| XXL_VLDL_FC | ASAT_Pos_pPS | 4255 | 0.03126706 | [0. 0.06] | 0.042006349 | 0.160489904 |
| XXL_VLDL_FC | ALP_Neg_pPS | 4255 | 0.012647813 | [-0.02 0.04] | 0.410858422 | 0.682580842 |
| XXL_VLDL_FC | SHBG-LpA_pPS | 4255 | -0.005883778 | [-0.04 0.02] | 0.702044534 | 0.859880159 |
| XXL_VLDL_FC | Obesity_pPS | 4255 | 0.036131292 | [0.01 0.07] | 0.018773613 | 0.085809983 |
| XXL_VLDL_FC | VAT_Neg_pPS | 4255 | 0.020866147 | [-0.01 0.05] | 0.17482936 | 0.453447086 |
| XXL_VLDL_FC | Bilirubin_pPS | 4255 | -0.012638986 | [-0.04 0.02] | 0.411185063 | 0.682580842 |
| XXL_VLDL_FC | Proinsulin_pPS | 4255 | 0.00407593 | [-0.03 0.03] | 0.790997956 | 0.899824971 |
| XXL_VLDL_FC | Hyper_Insulin_pPS | 4255 | 0.001642137 | [-0.03 0.03] | 0.914970999 | 0.966041099 |
| XXL_VLDL_FC | Beta_Cell_1_pPS | 4255 | 0.008852563 | [-0.02 0.04] | 0.564886852 | 0.797755947 |
| XXL_VLDL_FC | Beta_Cell_2_pPS | 4255 | -0.006510776 | [-0.04 0.02] | 0.672054445 | 0.851013914 |
| XXL_VLDL_L | Cholesterol_pPS | 4255 | -0.095828887 | [-0.13 -0.07] | 4.24E-10 | 1.46E-08 |
| XXL_VLDL_L | Lipodystrophy_1_pPS | 4255 | 0.042886642 | [0.01 0.07] | 0.005275041 | 0.031905232 |
| XXL_VLDL_L | Lipodystrophy_2_pPS | 4255 | -0.017637834 | [-0.05 0.01] | 0.251428106 | 0.548919377 |
| XXL_VLDL_L | Liver-Lipid_pPS | 4255 | -0.043875494 | [-0.07 -0.01] | 0.004315552 | 0.028142462 |
| XXL_VLDL_L | ASAT_Pos_pPS | 4255 | 0.033577725 | [0. 0.06] | 0.028975955 | 0.120352411 |
| XXL_VLDL_L | ALP_Neg_pPS | 4255 | 0.017794519 | [-0.01 0.05] | 0.247240711 | 0.545127647 |
| XXL_VLDL_L | SHBG-LpA_pPS | 4255 | -0.003133079 | [-0.03 0.03] | 0.838579447 | 0.924978542 |
| XXL_VLDL_L | Obesity_pPS | 4255 | 0.03717754 | [0.01 0.07] | 0.015602179 | 0.076184301 |
| XXL_VLDL_L | VAT_Neg_pPS | 4255 | 0.021986959 | [-0.01 0.05] | 0.152789381 | 0.428409441 |
| XXL_VLDL_L | Bilirubin_pPS | 4255 | -0.014265764 | [-0.04 0.02] | 0.35361708 | 0.639513454 |
| XXL_VLDL_L | Proinsulin_pPS | 4255 | -0.000894257 | [-0.03 0.03] | 0.953633989 | 0.980572802 |
| XXL_VLDL_L | Hyper_Insulin_pPS | 4255 | 0.000914962 | [-0.03 0.03] | 0.952561698 | 0.979973545 |
| XXL_VLDL_L | Beta_Cell_1_pPS | 4255 | 0.012440513 | [-0.02 0.04] | 0.418570448 | 0.691002715 |
| XXL_VLDL_L | Beta_Cell_2_pPS | 4255 | -0.005567587 | [-0.04 0.02] | 0.717349776 | 0.866099998 |
| XXL_VLDL_P | Cholesterol_pPS | 4255 | -0.095637248 | [-0.13 -0.07] | 4.59E-10 | 1.56E-08 |
| XXL_VLDL_P | Lipodystrophy_1_pPS | 4255 | 0.042326701 | [0.01 0.07] | 0.005900082 | 0.035259597 |
| XXL_VLDL_P | Lipodystrophy_2_pPS | 4255 | -0.018435372 | [-0.05 0.01] | 0.230623019 | 0.526462126 |
| XXL_VLDL_P | Liver-Lipid_pPS | 4255 | -0.043347774 | [-0.07 -0.01] | 0.004805895 | 0.030051115 |
| XXL_VLDL_P | ASAT_Pos_pPS | 4255 | 0.034043554 | [0. 0.06] | 0.026820019 | 0.113757794 |
| XXL_VLDL_P | ALP_Neg_pPS | 4255 | 0.017703406 | [-0.01 0.05] | 0.249669718 | 0.545779139 |
| XXL_VLDL_P | SHBG-LpA_pPS | 4255 | -0.00315104 | [-0.03 0.03] | 0.837666909 | 0.92461824 |
| XXL_VLDL_P | Obesity_pPS | 4255 | 0.03772141 | [0.01 0.07] | 0.014147742 | 0.070895183 |
| XXL_VLDL_P | VAT_Neg_pPS | 4255 | 0.022055286 | [-0.01 0.05] | 0.151517448 | 0.427467143 |
| XXL_VLDL_P | Bilirubin_pPS | 4255 | -0.013749319 | [-0.04 0.02] | 0.371315287 | 0.653802291 |
| XXL_VLDL_P | Proinsulin_pPS | 4255 | -0.001639422 | [-0.03 0.03] | 0.915111074 | 0.966041099 |
| XXL_VLDL_P | Hyper_Insulin_pPS | 4255 | 0.000825194 | [-0.03 0.03] | 0.957211245 | 0.981227298 |
| XXL_VLDL_P | Beta_Cell_1_pPS | 4255 | 0.012855359 | [-0.02 0.04] | 0.403222379 | 0.677078992 |
| XXL_VLDL_P | Beta_Cell_2_pPS | 4255 | -0.006318096 | [-0.04 0.02] | 0.681217852 | 0.858274475 |

**Table S9: Associations between pPSs and global metabolomic profile in SCAPIS**

**a) Spearman Correlation**

| Metabolomics | PPS | Samp | r | CI95% | p | fdr_p_adjusted |
|---|---|---|---|---|---|---|
| XS_VLDL_FC | Bilirubin_pPS | 4255 | -0.00984237 | [-0.04 0.02] | 0.521560378 | 0.738176158 |
| XS_VLDL_FC | Proinsulin_pPS | 4255 | 0.00635323 | [-0.02 0.04] | 0.679078476 | 0.836659007 |
| XS_VLDL_FC | Hyper_Insulin_pPS | 4255 | -0.012666066 | [-0.04 0.02] | 0.409464613 | 0.671998159 |
| XS_VLDL_FC | Beta_Cell_1_pPS | 4255 | 0.013363417 | [-0.02 0.04] | 0.384162247 | 0.654405057 |
| XS_VLDL_FC | Beta_Cell_2_pPS | 4255 | 0.025658767 | [-0. 0.06] | 0.094691753 | 0.290577698 |
| XS_VLDL_FC | Total_GRS | 4255 | 0.005533669 | [-0.02 0.04] | 0.718585108 | 0.865374699 |
| XS_VLDL_L | Cholesterol_pPS | 4255 | -0.132411328 | [-0.16 -0.1 ] | 4.69E-18 | 2.96E-16 |
| XS_VLDL_L | Lipodystrophy_1_pPS | 4255 | 0.058884748 | [0.03 0.09] | 0.000124115 | 0.001615492 |
| XS_VLDL_L | Lipodystrophy_2_pPS | 4255 | -0.029644544 | [-0.06 0. ] | 0.053501485 | 0.183617095 |
| XS_VLDL_L | Liver-Lipid_pPS | 4255 | 0.002765438 | [-0.03 0.03] | 0.857087219 | 0.934369525 |
| XS_VLDL_L | ASAT_Pos_pPS | 4255 | -0.000611779 | [-0.03 0.03] | 0.968221877 | 0.982420021 |
| XS_VLDL_L | ALP_Neg_pPS | 4255 | 0.051995838 | [0.02 0.08] | 0.00070349 | 0.006676932 |
| XS_VLDL_L | SHBG-LpA_pPS | 4255 | 0.014856435 | [-0.02 0.04] | 0.333298434 | 0.614196857 |
| XS_VLDL_L | Obesity_pPS | 4255 | 0.003549772 | [-0.03 0.03] | 0.817191444 | 0.915817998 |
| XS_VLDL_L | VAT_Neg_pPS | 4255 | 0.011702003 | [-0.02 0.04] | 0.446030001 | 0.693159645 |
| XS_VLDL_L | Bilirubin_pPS | 4255 | -0.013564579 | [-0.04 0.02] | 0.377044975 | 0.648565735 |
| XS_VLDL_L | Proinsulin_pPS | 4255 | 0.00916553 | [-0.02 0.04] | 0.550598323 | 0.760607725 |
| XS_VLDL_L | Hyper_Insulin_pPS | 4255 | -0.010520268 | [-0.04 0.02] | 0.493287384 | 0.721879553 |
| XS_VLDL_L | Beta_Cell_1_pPS | 4255 | 0.014219146 | [-0.02 0.04] | 0.35445373 | 0.634686582 |
| XS_VLDL_L | Beta_Cell_2_pPS | 4255 | 0.021195928 | [-0.01 0.05] | 0.167458811 | 0.429571287 |
| XS_VLDL_L | Total_GRS | 4255 | 0.013898136 | [-0.02 0.04] | 0.365423988 | 0.640911247 |
| XS_VLDL_P | Cholesterol_pPS | 4255 | -0.131845795 | [-0.16 -0.1 ] | 6.50E-18 | 3.98E-16 |
| XS_VLDL_P | Lipodystrophy_1_pPS | 4255 | 0.062311074 | [0.03 0.09] | 4.87E-05 | 0.000757585 |
| XS_VLDL_P | Lipodystrophy_2_pPS | 4255 | -0.026685151 | [-0.06 0. ] | 0.082206749 | 0.257421135 |
| XS_VLDL_P | Liver-Lipid_pPS | 4255 | -0.000866498 | [-0.03 0.03] | 0.955002758 | 0.977573158 |
| XS_VLDL_P | ASAT_Pos_pPS | 4255 | 0.002293877 | [-0.03 0.03] | 0.881257393 | 0.945095197 |
| XS_VLDL_P | ALP_Neg_pPS | 4255 | 0.049774744 | [0.02 0.08] | 0.001181546 | 0.009527879 |
| XS_VLDL_P | SHBG-LpA_pPS | 4255 | 0.013979449 | [-0.02 0.04] | 0.362625352 | 0.639224774 |
| XS_VLDL_P | Obesity_pPS | 4255 | 0.005597558 | [-0.02 0.04] | 0.715476519 | 0.863159242 |
| XS_VLDL_P | VAT_Neg_pPS | 4255 | 0.01167328 | [-0.02 0.04] | 0.447147206 | 0.693159645 |
| XS_VLDL_P | Bilirubin_pPS | 4255 | -0.014360612 | [-0.04 0.02] | 0.349685922 | 0.629258643 |
| XS_VLDL_P | Proinsulin_pPS | 4255 | 0.01012674 | [-0.02 0.04] | 0.509599565 | 0.7313594 |
| XS_VLDL_P | Hyper_Insulin_pPS | 4255 | -0.009103033 | [-0.04 0.02] | 0.55331914 | 0.760607725 |
| XS_VLDL_P | Beta_Cell_1_pPS | 4255 | 0.013934684 | [-0.02 0.04] | 0.364164416 | 0.639224774 |
| XS_VLDL_P | Beta_Cell_2_pPS | 4255 | 0.019413983 | [-0.01 0.05] | 0.20610777 | 0.484809862 |
| XS_VLDL_P | Total_GRS | 4255 | 0.016298865 | [-0.01 0.05] | 0.28849078 | 0.575104761 |
| XS_VLDL_PL | Cholesterol_pPS | 4255 | -0.144066778 | [-0.17 -0.11] | 4.07E-21 | 3.36E-19 |
| XS_VLDL_PL | Lipodystrophy_1_pPS | 4255 | 0.051172437 | [0.02 0.08] | 0.000854563 | 0.007605968 |
| XS_VLDL_PL | Lipodystrophy_2_pPS | 4255 | -0.037605997 | [-0.07 -0.01] | 0.014296101 | 0.063620614 |
| XS_VLDL_PL | Liver-Lipid_pPS | 4255 | 0.009202543 | [-0.02 0.04] | 0.548990058 | 0.760607725 |
| XS_VLDL_PL | ASAT_Pos_pPS | 4255 | -0.012965443 | [-0.04 0.02] | 0.398482922 | 0.666036824 |
| XS_VLDL_PL | ALP_Neg_pPS | 4255 | 0.054522715 | [0.02 0.08] | 0.000380699 | 0.004145175 |
| XS_VLDL_PL | SHBG-LpA_pPS | 4255 | 0.017702347 | [-0.01 0.05] | 0.248971304 | 0.53033113 |
| XS_VLDL_PL | Obesity_pPS | 4255 | -0.001366572 | [-0.03 0.03] | 0.929089848 | 0.966019256 |
| XS_VLDL_PL | VAT_Neg_pPS | 4255 | 0.01070514 | [-0.02 0.04] | 0.485721906 | 0.717543724 |
| XS_VLDL_PL | Bilirubin_pPS | 4255 | -0.00913203 | [-0.04 0.02] | 0.552055936 | 0.760607725 |
| XS_VLDL_PL | Proinsulin_pPS | 4255 | 0.004190707 | [-0.03 0.03] | 0.78493289 | 0.897605875 |
| XS_VLDL_PL | Hyper_Insulin_pPS | 4255 | -0.0148368 | [-0.04 0.02] | 0.333937831 | 0.614846907 |
| XS_VLDL_PL | Beta_Cell_1_pPS | 4255 | 0.014458208 | [-0.02 0.04] | 0.346420494 | 0.62662761 |
| XS_VLDL_PL | Beta_Cell_2_pPS | 4255 | 0.023671006 | [-0.01 0.05] | 0.123158402 | 0.354065765 |
| XS_VLDL_PL | Total_GRS | 4255 | 0.006365807 | [-0.02 0.04] | 0.678478728 | 0.836400502 |
| XS_VLDL_TG | Cholesterol_pPS | 4255 | -0.090724515 | [-0.12 -0.06] | 3.22E-09 | 1.06E-07 |
| XS_VLDL_TG | Lipodystrophy_1_pPS | 4255 | 0.076613155 | [0.05 0.11] | 5.84E-07 | 1.38E-05 |
| XS_VLDL_TG | Lipodystrophy_2_pPS | 4255 | 0.0153537 | [-0.01 0.05] | 0.317369354 | 0.605655929 |
| XS_VLDL_TG | Liver-Lipid_pPS | 4255 | -0.032662873 | [-0.06 -0. ] | 0.033374808 | 0.128499249 |
| XS_VLDL_TG | ASAT_Pos_pPS | 4255 | 0.028729902 | [-0. 0.06] | 0.061310129 | 0.203891825 |
| XS_VLDL_TG | ALP_Neg_pPS | 4255 | 0.022733066 | [-0.01 0.05] | 0.1387269 | 0.3817668 |
| XS_VLDL_TG | SHBG-LpA_pPS | 4255 | -0.000315066 | [-0.03 0.03] | 0.983631069 | 0.990093216 |
| XS_VLDL_TG | Obesity_pPS | 4255 | 0.022144812 | [-0.01 0.05] | 0.149238198 | 0.398714883 |
| XS_VLDL_TG | VAT_Neg_pPS | 4255 | 0.00936475 | [-0.02 0.04] | 0.541969332 | 0.756359283 |
| XS_VLDL_TG | Bilirubin_pPS | 4255 | -0.004048907 | [-0.03 0.03] | 0.792040125 | 0.901287039 |
| XS_VLDL_TG | Proinsulin_pPS | 4255 | 0.015000509 | [-0.02 0.05] | 0.328631063 | 0.610965216 |
| XS_VLDL_TG | Hyper_Insulin_pPS | 4255 | 0.008737843 | [-0.02 0.04] | 0.569348313 | 0.770653232 |
| XS_VLDL_TG | Beta_Cell_1_pPS | 4255 | 0.008020144 | [-0.02 0.04] | 0.601480159 | 0.790548371 |
| XS_VLDL_TG | Beta_Cell_2_pPS | 4255 | 0.001773441 | [-0.03 0.03] | 0.908060707 | 0.952872131 |
| XS_VLDL_TG | Total_GRS | 4255 | 0.038318367 | [0.01 0.07] | 0.012554062 | 0.057291924 |
| XXL_VLDL_C | Cholesterol_pPS | 4255 | -0.095665988 | [-0.13 -0.07] | 4.26E-10 | 1.63E-08 |
| XXL_VLDL_C | Lipodystrophy_1_pPS | 4255 | 0.056442809 | [0.03 0.09] | 0.000234666 | 0.002735646 |
| XXL_VLDL_C | Lipodystrophy_2_pPS | 4255 | -0.014100768 | [-0.04 0.02] | 0.358474775 | 0.637057491 |
| XXL_VLDL_C | Liver-Lipid_pPS | 4255 | -0.039241648 | [-0.07 -0.01] | 0.010577173 | 0.049363789 |
| XXL_VLDL_C | ASAT_Pos_pPS | 4255 | 0.040370238 | [0.01 0.07] | 0.00853983 | 0.042013614 |
| XXL_VLDL_C | ALP_Neg_pPS | 4255 | 0.02429917 | [-0.01 0.05] | 0.113518538 | 0.33219272 |
| XXL_VLDL_C | SHBG-LpA_pPS | 4255 | 0.001597395 | [-0.03 0.03] | 0.917152666 | 0.957786011 |
| XXL_VLDL_C | Obesity_pPS | 4255 | 0.027462827 | [-0. 0.06] | 0.073664344 | 0.235835846 |
| XXL_VLDL_C | VAT_Neg_pPS | 4255 | 0.029234521 | [-0. 0.06] | 0.056892825 | 0.192181275 |
| XXL_VLDL_C | Bilirubin_pPS | 4255 | -0.014739689 | [-0.04 0.02] | 0.337111718 | 0.619626938 |
| XXL_VLDL_C | Proinsulin_pPS | 4255 | 0.011452861 | [-0.02 0.04] | 0.455773328 | 0.696320363 |
| XXL_VLDL_C | Hyper_Insulin_pPS | 4255 | 0.010773624 | [-0.02 0.04] | 0.482935336 | 0.717164725 |
| XXL_VLDL_C | Beta_Cell_1_pPS | 4255 | 0.016309153 | [-0.01 0.05] | 0.288186509 | 0.57504794 |
| XXL_VLDL_C | Beta_Cell_2_pPS | 4255 | 0.003896079 | [-0.03 0.03] | 0.799719437 | 0.905317475 |
| XXL_VLDL_C | Total_GRS | 4255 | 0.031484914 | [0. 0.06] | 0.040288539 | 0.14925547 |
| XXL_VLDL_CE | Cholesterol_pPS | 4255 | -0.10070518 | [-0.13 -0.07] | 4.88E-11 | 2.05E-09 |
| XXL_VLDL_CE | Lipodystrophy_1_pPS | 4255 | 0.05679131 | [0.03 0.09] | 0.000214593 | 0.002543104 |
| XXL_VLDL_CE | Lipodystrophy_2_pPS | 4255 | -0.019278909 | [-0.05 0.01] | 0.209281958 | 0.488466046 |
| XXL_VLDL_CE | Liver-Lipid_pPS | 4255 | -0.029061718 | [-0.06 0. ] | 0.058374713 | 0.195952676 |
| XXL_VLDL_CE | ASAT_Pos_pPS | 4255 | 0.038317542 | [0.01 0.07] | 0.012555966 | 0.057291924 |
| XXL_VLDL_CE | ALP_Neg_pPS | 4255 | 0.030240574 | [0. 0.06] | 0.048873903 | 0.171298237 |
| XXL_VLDL_CE | SHBG-LpA_pPS | 4255 | 0.00284649 | [-0.03 0.03] | 0.852945673 | 0.932534492 |
| XXL_VLDL_CE | Obesity_pPS | 4255 | 0.022923616 | [-0.01 0.05] | 0.135447152 | 0.376339562 |
| XXL_VLDL_CE | VAT_Neg_pPS | 4255 | 0.029361006 | [-0. 0.06] | 0.055828084 | 0.189479812 |
| XXL_VLDL_CE | Bilirubin_pPS | 4255 | -0.018340817 | [-0.05 0.01] | 0.232306548 | 0.516301672 |
| XXL_VLDL_CE | Proinsulin_pPS | 4255 | 0.011427664 | [-0.02 0.04] | 0.456765327 | 0.697339236 |
| XXL_VLDL_CE | Hyper_Insulin_pPS | 4255 | 0.010210574 | [-0.02 0.04] | 0.50610106 | 0.729504373 |
| XXL_VLDL_CE | Beta_Cell_1_pPS | 4255 | 0.017130063 | [-0.01 0.05] | 0.264603443 | 0.551490946 |

**b) Partial Correlation**

| Metabolomics | PPS | Samp | r | CI95% | p | fdr_p_adjusted |
|---|---|---|---|---|---|---|
| XXL_VLDL_PL | Cholesterol_pPS | 4255 | -0.089016847 | [-0.12 -0.06] | 6.66E-09 | 1.88E-07 |
| XXL_VLDL_PL | Lipodystrophy_1_pPS | 4255 | 0.044681778 | [0.01 0.07] | 0.003653493 | 0.024380977 |
| XXL_VLDL_PL | Lipodystrophy_2_pPS | 4255 | -0.010383891 | [-0.04 0.02] | 0.499567275 | 0.755062449 |
| XXL_VLDL_PL | Liver-Lipid_pPS | 4255 | -0.049501575 | [-0.08 -0.02] | 0.001279545 | 0.010673534 |
| XXL_VLDL_PL | ASAT_Pos_pPS | 4255 | 0.028689291 | [-0. 0.06] | 0.062077807 | 0.223123466 |
| XXL_VLDL_PL | ALP_Neg_pPS | 4255 | 0.017414422 | [-0.01 0.05] | 0.257483892 | 0.553690606 |
| XXL_VLDL_PL | SHBG-LpA_pPS | 4255 | -0.002753168 | [-0.03 0.03] | 0.857930485 | 0.937541939 |
| XXL_VLDL_PL | Obesity_pPS | 4255 | 0.036552994 | [0.01 0.07] | 0.017433187 | 0.08243497 |
| XXL_VLDL_PL | VAT_Neg_pPS | 4255 | 0.017167694 | [-0.01 0.05] | 0.264288408 | 0.561683006 |
| XXL_VLDL_PL | Bilirubin_pPS | 4255 | -0.010233491 | [-0.04 0.02] | 0.505800387 | 0.76185979 |
| XXL_VLDL_PL | Proinsulin_pPS | 4255 | 0.000709316 | [-0.03 0.03] | 0.963215247 | 0.984860534 |
| XXL_VLDL_PL | Hyper_Insulin_pPS | 4255 | -0.000205571 | [-0.03 0.03] | 0.98933572 | 0.993977294 |
| XXL_VLDL_PL | Beta_Cell_1_pPS | 4255 | 0.013935457 | [-0.02 0.04] | 0.364874115 | 0.650898451 |
| XXL_VLDL_PL | Beta_Cell_2_pPS | 4255 | -0.007005211 | [-0.04 0.02] | 0.648764662 | 0.843394061 |
| XXL_VLDL_TG | Cholesterol_pPS | 4255 | -0.095407012 | [-0.13 -0.07] | 5.05E-10 | 1.69E-08 |
| XXL_VLDL_TG | Lipodystrophy_1_pPS | 4255 | 0.040488247 | [0.01 0.07] | 0.008448634 | 0.047625902 |
| XXL_VLDL_TG | Lipodystrophy_2_pPS | 4255 | -0.018786407 | [-0.05 0.01] | 0.221864431 | 0.514831403 |
| XXL_VLDL_TG | Liver-Lipid_pPS | 4255 | -0.043654672 | [-0.07 -0.01] | 0.004514935 | 0.028878276 |
| XXL_VLDL_TG | ASAT_Pos_pPS | 4255 | 0.034761834 | [0. 0.06] | 0.023767254 | 0.103890924 |
| XXL_VLDL_TG | ALP_Neg_pPS | 4255 | 0.016092428 | [-0.01 0.05] | 0.295383302 | 0.593136781 |
| XXL_VLDL_TG | SHBG-LpA_pPS | 4255 | -0.00275667 | [-0.03 0.03] | 0.857751712 | 0.937541939 |
| XXL_VLDL_TG | Obesity_pPS | 4255 | 0.038398321 | [0.01 0.07] | 0.012505474 | 0.064030585 |
| XXL_VLDL_TG | VAT_Neg_pPS | 4255 | 0.021050725 | [-0.01 0.05] | 0.171045057 | 0.448797122 |
| XXL_VLDL_TG | Bilirubin_pPS | 4255 | -0.013621218 | [-0.04 0.02] | 0.37578883 | 0.655910408 |
| XXL_VLDL_TG | Proinsulin_pPS | 4255 | -0.002294266 | [-0.03 0.03] | 0.881418217 | 0.949219618 |
| XXL_VLDL_TG | Hyper_Insulin_pPS | 4255 | 0.000518458 | [-0.03 0.03] | 0.973108601 | 0.989920436 |
| XXL_VLDL_TG | Beta_Cell_1_pPS | 4255 | 0.012993951 | [-0.02 0.04] | 0.398170954 | 0.673455453 |
| XXL_VLDL_TG | Beta_Cell_2_pPS | 4255 | -0.007279692 | [-0.04 0.02] | 0.635980446 | 0.835456904 |

**Table S9: Associations between pPSs and global metabolomic profile in SCAPIS**

**a) Spearman Correlation**

| Metabolomics | PPS | Samp | r | CI95% | p | fdr_p_adjusted |
|---|---|---|---|---|---|---|
| XXL_VLDL_CE | Beta_Cell_2_pPS | 4255 | 0.008888107 | [-0.02 0.04] | 0.562726032 | 0.767793567 |
| XXL_VLDL_CE | Total_GRS | 4255 | 0.028508163 | [-0. 0.06] | 0.063339063 | 0.208697833 |
| XXL_VLDL_FC | Cholesterol_pPS | 4255 | -0.080779156 | [-0.11 -0.05] | 1.37E-07 | 3.51E-06 |
| XXL_VLDL_FC | Lipodystrophy_1_pPS | 4255 | 0.054157553 | [0.02 0.08] | 0.000416688 | 0.004424734 |
| XXL_VLDL_FC | Lipodystrophy_2_pPS | 4255 | -0.003315771 | [-0.03 0.03] | 0.829049571 | 0.922360648 |
| XXL_VLDL_FC | Liver-Lipid_pPS | 4255 | -0.051663927 | [-0.08 -0.02] | 0.000761126 | 0.007037137 |
| XXL_VLDL_FC | ASAT_Pos_pPS | 4255 | 0.041370048 | [0.01 0.07] | 0.007036012 | 0.035763616 |
| XXL_VLDL_FC | ALP_Neg_pPS | 4255 | 0.012875922 | [-0.02 0.04] | 0.401747979 | 0.668022802 |
| XXL_VLDL_FC | SHBG-LpA_pPS | 4255 | -0.000334507 | [-0.03 0.03] | 0.982621214 | 0.990005874 |
| XXL_VLDL_FC | Obesity_pPS | 4255 | 0.032179686 | [0. 0.06] | 0.036077248 | 0.13648271 |
| XXL_VLDL_FC | VAT_Neg_pPS | 4255 | 0.024433417 | [-0.01 0.05] | 0.111537538 | 0.327524552 |
| XXL_VLDL_FC | Bilirubin_pPS | 4255 | -0.009092893 | [-0.04 0.02] | 0.553761229 | 0.760607725 |
| XXL_VLDL_FC | Proinsulin_pPS | 4255 | 0.010412636 | [-0.02 0.04] | 0.497720932 | 0.724295386 |
| XXL_VLDL_FC | Hyper_Insulin_pPS | 4255 | 0.01014172 | [-0.02 0.04] | 0.508973514 | 0.7313594 |
| XXL_VLDL_FC | Beta_Cell_1_pPS | 4255 | 0.013860516 | [-0.02 0.04] | 0.366723352 | 0.64109339 |
| XXL_VLDL_FC | Beta_Cell_2_pPS | 4255 | -0.00364886 | [-0.03 0.03] | 0.812182532 | 0.912588544 |
| XXL_VLDL_FC | Total_GRS | 4255 | 0.034463134 | [0. 0.06] | 0.024776107 | 0.101615199 |
| XXL_VLDL_L | Cholesterol_pPS | 4255 | -0.095201005 | [-0.12 -0.07] | 5.18E-10 | 1.92E-08 |
| XXL_VLDL_L | Lipodystrophy_1_pPS | 4255 | 0.050387546 | [0.02 0.08] | 0.001026073 | 0.008665065 |
| XXL_VLDL_L | Lipodystrophy_2_pPS | 4255 | -0.014489803 | [-0.04 0.02] | 0.345367565 | 0.62662761 |
| XXL_VLDL_L | Liver-Lipid_pPS | 4255 | -0.043740872 | [-0.07 -0.01] | 0.004375622 | 0.024735931 |
| XXL_VLDL_L | ASAT_Pos_pPS | 4255 | 0.042223887 | [0.01 0.07] | 0.005944826 | 0.031254047 |
| XXL_VLDL_L | ALP_Neg_pPS | 4255 | 0.0172865 | [-0.01 0.05] | 0.260264656 | 0.546250184 |
| XXL_VLDL_L | SHBG-LpA_pPS | 4255 | 0.001982401 | [-0.03 0.03] | 0.897284588 | 0.949937874 |
| XXL_VLDL_L | Obesity_pPS | 4255 | 0.03141993 | [0. 0.06] | 0.040702926 | 0.150530649 |
| XXL_VLDL_L | VAT_Neg_pPS | 4255 | 0.025118587 | [-0. 0.06] | 0.101848284 | 0.306832261 |
| XXL_VLDL_L | Bilirubin_pPS | 4255 | -0.010823279 | [-0.04 0.02] | 0.480920368 | 0.716353047 |
| XXL_VLDL_L | Proinsulin_pPS | 4255 | 0.007407506 | [-0.02 0.04] | 0.629538205 | 0.803929814 |
| XXL_VLDL_L | Hyper_Insulin_pPS | 4255 | 0.008828398 | [-0.02 0.04] | 0.565353044 | 0.768674587 |
| XXL_VLDL_L | Beta_Cell_1_pPS | 4255 | 0.017158462 | [-0.01 0.05] | 0.263812123 | 0.550503091 |
| XXL_VLDL_L | Beta_Cell_2_pPS | 4255 | -0.002451522 | [-0.03 0.03] | 0.87316372 | 0.942282701 |
| XXL_VLDL_L | Total_GRS | 4255 | 0.030475943 | [0. 0.06] | 0.047141388 | 0.167970561 |
| XXL_VLDL_P | Cholesterol_pPS | 4255 | -0.095009241 | [-0.12 -0.07] | 5.61E-10 | 2.04E-08 |
| XXL_VLDL_P | Lipodystrophy_1_pPS | 4255 | 0.049892001 | [0.02 0.08] | 0.001150209 | 0.009345451 |
| XXL_VLDL_P | Lipodystrophy_2_pPS | 4255 | -0.015285763 | [-0.05 0.01] | 0.31951568 | 0.608128779 |
| XXL_VLDL_P | Liver-Lipid_pPS | 4255 | -0.043273664 | [-0.07 -0.01] | 0.004813397 | 0.026204915 |
| XXL_VLDL_P | ASAT_Pos_pPS | 4255 | 0.042569979 | [0.01 0.07] | 0.005547784 | 0.029455436 |
| XXL_VLDL_P | ALP_Neg_pPS | 4255 | 0.017070534 | [-0.01 0.05] | 0.266267514 | 0.552249619 |
| XXL_VLDL_P | SHBG-LpA_pPS | 4255 | 0.001871135 | [-0.03 0.03] | 0.903020362 | 0.952300234 |
| XXL_VLDL_P | Obesity_pPS | 4255 | 0.031878651 | [0. 0.06] | 0.037853506 | 0.141950648 |
| XXL_VLDL_P | VAT_Neg_pPS | 4255 | 0.025039121 | [-0.01 0.06] | 0.102936487 | 0.309241967 |
| XXL_VLDL_P | Bilirubin_pPS | 4255 | -0.010325928 | [-0.04 0.02] | 0.501307902 | 0.727048985 |
| XXL_VLDL_P | Proinsulin_pPS | 4255 | 0.006679873 | [-0.02 0.04] | 0.663568025 | 0.825654931 |
| XXL_VLDL_P | Hyper_Insulin_pPS | 4255 | 0.008695092 | [-0.02 0.04] | 0.57123917 | 0.771552531 |
| XXL_VLDL_P | Beta_Cell_1_pPS | 4255 | 0.017237492 | [-0.01 0.05] | 0.261618561 | 0.548555046 |
| XXL_VLDL_P | Beta_Cell_2_pPS | 4255 | -0.003227714 | [-0.03 0.03] | 0.833522134 | 0.924155036 |
| XXL_VLDL_P | Total_GRS | 4255 | 0.029938657 | [-0. 0.06] | 0.051174194 | 0.177332222 |
| XXL_VLDL_PL | Cholesterol_pPS | 4255 | -0.088132937 | [-0.12 -0.06] | 8.91E-09 | 2.69E-07 |
| XXL_VLDL_PL | Lipodystrophy_1_pPS | 4255 | 0.051632978 | [0.02 0.08] | 0.000766718 | 0.007058413 |
| XXL_VLDL_PL | Lipodystrophy_2_pPS | 4255 | -0.008168536 | [-0.04 0.02] | 0.594769813 | 0.786125349 |
| XXL_VLDL_PL | Liver-Lipid_pPS | 4255 | -0.049189787 | [-0.08 -0.02] | 0.001349987 | 0.01060704 |
| XXL_VLDL_PL | ASAT_Pos_pPS | 4255 | 0.038182087 | [0.01 0.07] | 0.012872009 | 0.058249914 |
| XXL_VLDL_PL | ALP_Neg_pPS | 4255 | 0.016997313 | [-0.01 0.05] | 0.268324165 | 0.55341859 |
| XXL_VLDL_PL | SHBG-LpA_pPS | 4255 | 0.002106788 | [-0.03 0.03] | 0.890878782 | 0.948487102 |
| XXL_VLDL_PL | Obesity_pPS | 4255 | 0.031403814 | [0. 0.06] | 0.040806253 | 0.150653034 |
| XXL_VLDL_PL | VAT_Neg_pPS | 4255 | 0.020945996 | [-0.01 0.05] | 0.172525231 | 0.436399316 |
| XXL_VLDL_PL | Bilirubin_pPS | 4255 | -0.006795966 | [-0.04 0.02] | 0.6580895 | 0.823091532 |
| XXL_VLDL_PL | Proinsulin_pPS | 4255 | 0.009090972 | [-0.02 0.04] | 0.553845014 | 0.760607725 |
| XXL_VLDL_PL | Hyper_Insulin_pPS | 4255 | 0.007401486 | [-0.02 0.04] | 0.629816708 | 0.803929814 |
| XXL_VLDL_PL | Beta_Cell_1_pPS | 4255 | 0.017901758 | [-0.01 0.05] | 0.243679077 | 0.525480742 |
| XXL_VLDL_PL | Beta_Cell_2_pPS | 4255 | -0.003556368 | [-0.03 0.03] | 0.816857804 | 0.915817998 |
| XXL_VLDL_PL | Total_GRS | 4255 | 0.03210504 | [0. 0.06] | 0.036510941 | 0.137396436 |
| XXL_VLDL_TG | Cholesterol_pPS | 4255 | -0.094761644 | [-0.12 -0.06] | 6.22E-10 | 2.22E-08 |
| XXL_VLDL_TG | Lipodystrophy_1_pPS | 4255 | 0.048079437 | [0.02 0.08] | 0.00173205 | 0.012767171 |
| XXL_VLDL_TG | Lipodystrophy_2_pPS | 4255 | -0.015859062 | [-0.05 0.01] | 0.301700721 | 0.587782058 |
| XXL_VLDL_TG | Liver-Lipid_pPS | 4255 | -0.043733629 | [-0.07 -0.01] | 0.004382123 | 0.024735931 |
| XXL_VLDL_TG | ASAT_Pos_pPS | 4255 | 0.042939468 | [0.01 0.07] | 0.005150445 | 0.027688481 |
| XXL_VLDL_TG | ALP_Neg_pPS | 4255 | 0.015254755 | [-0.01 0.05] | 0.320498487 | 0.608204326 |
| XXL_VLDL_TG | SHBG-LpA_pPS | 4255 | 0.00205446 | [-0.03 0.03] | 0.89357276 | 0.948580863 |
| XXL_VLDL_TG | Obesity_pPS | 4255 | 0.032374907 | [0. 0.06] | 0.034963725 | 0.133171327 |
| XXL_VLDL_TG | VAT_Neg_pPS | 4255 | 0.023827548 | [-0.01 0.05] | 0.120698283 | 0.348918892 |
| XXL_VLDL_TG | Bilirubin_pPS | 4255 | -0.010257168 | [-0.04 0.02] | 0.50416208 | 0.727744052 |
| XXL_VLDL_TG | Proinsulin_pPS | 4255 | 0.005639117 | [-0.02 0.04] | 0.713456976 | 0.86291356 |
| XXL_VLDL_TG | Hyper_Insulin_pPS | 4255 | 0.008080558 | [-0.02 0.04] | 0.598744101 | 0.788401533 |
| XXL_VLDL_TG | Beta_Cell_1_pPS | 4255 | 0.016781609 | [-0.01 0.05] | 0.274446223 | 0.559392289 |
| XXL_VLDL_TG | Beta_Cell_2_pPS | 4255 | -0.004411629 | [-0.03 0.03] | 0.773895767 | 0.891684366 |
| XXL_VLDL_TG | Total_GRS | 4255 | 0.029171694 | [-0. 0.06] | 0.057427944 | 0.193379811 |

**b) Partial Correlation**

| Metabolomics | PPS | Samp | r | CI95% | p | fdr_p_adjusted |
|---|---|---|---|---|---|---|

**Table S10: Elastic Net Regression Summary Statistics (generated from SCAPIS)**

| PPS | Omic_Layer | Sample_Size | Selected_Variables_Number | r2_train | r2_test | lambda | l1_ratio (alpha) | p_value | fdr_p_adjusted |
|---|---|---|---|---|---|---|---|---|---|
| Cholesterol_pPS | Anthropometry | 24255 | 0 | 0 | -0.000227009 | 115.1682353 | 0.0001 | 0.719 | 0.823913043 |
| Cholesterol_pPS | Biochemistry | 21936 | 21 | 0.05626623 | 0.050809317 | 0.001492067 | 0.10009 | 0 | 0 |
| Cholesterol_pPS | Proteomics | 3547 | 14 | 0.026729384 | 0.009074736 | 0.030162901 | 1 | 0 | 0 |
| Cholesterol_pPS | Metabolomics | 4214 | 62 | 0.070148216 | 0.037073649 | 0.002905653 | 0.40006 | 0 | 0 |
| Cholesterol_pPS | Merged_Omics | 2815 | 63 | 0.129716453 | 0.046168595 | 0.018867165 | 1 | 0 | 0 |
| Lipodystrophy_1_pPS | Anthropometry | 24255 | 8 | 0.004194524 | 0.003324586 | 0.009724837 | 0.10009 | 0 | 0 |
| Lipodystrophy_1_pPS | Biochemistry | 21936 | 12 | 0.00736391 | 0.006415542 | 0.004630211 | 1 | 0 | 0 |
| Lipodystrophy_1_pPS | Proteomics | 3547 | 183 | 0.018278847 | 0.00354214 | 4.75077728 | 0.0001 | 0.017 | 0.028977273 |
| Lipodystrophy_1_pPS | Metabolomics | 4214 | 136 | 0.005516298 | 0.004167842 | 9.856956676 | 0.0001 | 0.015 | 0.026162791 |
| Lipodystrophy_1_pPS | Merged_Omics | 2815 | 311 | 0.008842379 | 0.00282957 | 30.63644076 | 0.0001 | 0.009 | 0.016875 |
| Lipodystrophy_2_pPS | Anthropometry | 24255 | 10 | 0.001617291 | 0.001274231 | 0.373877471 | 0.0001 | 0 | 0 |
| Lipodystrophy_2_pPS | Biochemistry | 21936 | 15 | 0.011339682 | 0.011599761 | 0.029699311 | 0.10009 | 0 | 0 |
| Lipodystrophy_2_pPS | Proteomics | 3547 | 26 | 0.024411119 | 0.003764646 | 0.251979647 | 0.10009 | 0.004 | 0.008108108 |
| Lipodystrophy_2_pPS | Metabolomics | 4214 | 142 | 0.014382282 | -0.00297949 | 1.109493241 | 0.0001 | 0.932 | 0.944594595 |
| Lipodystrophy_2_pPS | Merged_Omics | 2815 | 333 | 0.057332251 | 0.017342218 | 2.258197227 | 0.0001 | 0 | 0 |
| Liver-Lipid_pPS | Anthropometry | 24255 | 6 | 0.001928931 | 0.001470696 | 0.001805613 | 1 | 0 | 0 |
| Liver-Lipid_pPS | Biochemistry | 21936 | 21 | 0.026665047 | 0.027228056 | 0.001313715 | 0.10009 | 0 | 0 |
| Liver-Lipid_pPS | Proteomics | 3547 | 46 | 0.069788548 | 0.037138074 | 0.018713061 | 1 | 0 | 0 |
| Liver-Lipid_pPS | Metabolomics | 4214 | 50 | 0.05738676 | 0.030190654 | 0.003173754 | 0.40006 | 0 | 0 |
| Liver-Lipid_pPS | Merged_Omics | 2815 | 57 | 0.098384209 | 0.050844068 | 0.021718306 | 1 | 0 | 0 |
| ASAT_Pos_pPS | Anthropometry | 24255 | 10 | 0.001035854 | 0.00133845 | 0.426388405 | 0.0001 | 0.001 | 0.002142857 |
| ASAT_Pos_pPS | Biochemistry | 21936 | 11 | 0.004112157 | 0.002589973 | 0.004901279 | 1 | 0 | 0 |
| ASAT_Pos_pPS | Proteomics | 3547 | 180 | 0.03100955 | 0.001271044 | 2.353192318 | 0.0001 | 0.007 | 0.013815789 |
| ASAT_Pos_pPS | Metabolomics | 4214 | 0 | 0 | -2.29E-05 | 359.3053831 | 0.0001 | 0.69 | 0.823913043 |
| ASAT_Pos_pPS | Merged_Omics | 2815 | 13 | 0.012270452 | -0.004757539 | 0.396812927 | 0.10009 | 0.01 | 0.018292683 |
| ALP_Neg_pPS | Anthropometry | 24255 | 10 | 0.001101543 | 0.001126805 | 0.16387208 | 0.0001 | 0 | 0 |
| ALP_Neg_pPS | Biochemistry | 21936 | 18 | 0.012922248 | 0.011788276 | 0.001054523 | 1 | 0 | 0 |
| ALP_Neg_pPS | Proteomics | 3547 | 104 | 0.396883421 | 0.345880066 | 0.008186242 | 1 | 0 | 0 |
| ALP_Neg_pPS | Metabolomics | 4214 | 43 | 0.031153354 | 0.002252129 | 0.001590719 | 1 | 0.026 | 0.043333333 |
| ALP_Neg_pPS | Merged_Omics | 2815 | 133 | 0.409688351 | 0.339205946 | 0.046421024 | 0.20008 | 0 | 0 |
| SHBG-LpA_pPS | Anthropometry | 24255 | 0 | 0 | -0.000697168 | 114.5695247 | 0.0001 | 0.693 | 0.823913043 |
| SHBG-LpA_pPS | Biochemistry | 21936 | 17 | 0.00720147 | 0.006408074 | 0.016631797 | 0.10009 | 0 | 0 |
| SHBG-LpA_pPS | Proteomics | 3547 | 161 | 0.003486839 | -0.000923523 | 30.97633085 | 0.0001 | 0.769 | 0.823928571 |
| SHBG-LpA_pPS | Metabolomics | 4214 | 0 | 0 | -0.000313855 | 338.0480906 | 0.0001 | 0.626 | 0.823913043 |
| SHBG-LpA_pPS | Merged_Omics | 2815 | 5 | 0.006638611 | -0.000696643 | 0.04558656 | 1 | 0.036 | 0.057446809 |
| Obesity_pPS | Anthropometry | 24255 | 6 | 0.013561623 | 0.009237362 | 0.001822454 | 1 | 0 | 0 |
| Obesity_pPS | Biochemistry | 21936 | 18 | 0.007921659 | 0.004025853 | 0.010165803 | 0.10009 | 0 | 0 |
| Obesity_pPS | Proteomics | 3547 | 180 | 0.0264863 | -0.007645233 | 3.777631983 | 0.0001 | 0.892 | 0.929166667 |
| Obesity_pPS | Metabolomics | 4214 | 0 | 0 | -0.000183298 | 293.24857 | 0.0001 | 0.691 | 0.823913043 |
| Obesity_pPS | Merged_Omics | 2815 | 9 | 0.010716112 | -0.001061749 | 0.047053115 | 1 | 0.785 | 0.829225352 |
| VAT_Neg_pPS | Anthropometry | 24255 | 9 | 0.004060591 | 0.000520522 | 0.003328961 | 0.10009 | 0 | 0 |
| VAT_Neg_pPS | Biochemistry | 21936 | 21 | 0.009092439 | 0.005213715 | 0.001688285 | 0.10009 | 0 | 0 |
| VAT_Neg_pPS | Proteomics | 3547 | 161 | 0.004947674 | -0.002541346 | 24.03671956 | 0.0001 | 0.049 | 0.075 |
| VAT_Neg_pPS | Metabolomics | 4214 | 7 | 0.004060398 | -0.002584987 | 0.278341607 | 0.10009 | 0.907 | 0.931849315 |
| VAT_Neg_pPS | Merged_Omics | 2815 | 0 | 0 | -0.003614518 | 586.4690051 | 0.0001 | 0.728 | 0.823913043 |
| Bilirubin_pPS | Anthropometry | 24255 | 0 | 0 | -1.96E-05 | 62.17377375 | 0.0001 | 0.758 | 0.823913043 |
| Bilirubin_pPS | Biochemistry | 21936 | 1 | 0.00066193 | -0.000216072 | 0.009019223 | 1 | 0.009 | 0.016875 |
| Bilirubin_pPS | Proteomics | 3547 | 0 | 0 | -0.000144979 | 449.7393419 | 0.0001 | 0.574 | 0.812264151 |
| Bilirubin_pPS | Metabolomics | 4214 | 140 | 0.007408877 | -0.005543612 | 1.67196987 | 0.0001 | 0.972 | 0.972 |
| Bilirubin_pPS | Merged_Omics | 2815 | 0 | 0 | -0.001047447 | 442.0792795 | 0.0001 | 0.691 | 0.823913043 |
| Proinsulin_pPS | Anthropometry | 24255 | 1 | 0.000469913 | 0.000110683 | 0.011236557 | 1 | 0.056 | 0.084 |
| Proinsulin_pPS | Biochemistry | 21936 | 19 | 0.009543491 | 0.005095793 | 0.001238729 | 1 | 0 | 0 |
| Proinsulin_pPS | Proteomics | 3547 | 184 | 0.064066156 | 0.003911542 | 0.941016347 | 0.0001 | 0.001 | 0.002142857 |
| Proinsulin_pPS | Metabolomics | 4214 | 0 | 0 | -0.000132754 | 0.040226992 | 1 | 0.705 | 0.823913043 |
| Proinsulin_pPS | Merged_Omics | 2815 | 343 | 0.059585089 | 0.005160384 | 1.928673234 | 0.0001 | 0.001 | 0.002142857 |
| Hyper_Insulin_pPS | Anthropometry | 24255 | 0 | 0 | -0.000299039 | 101.4320837 | 0.0001 | 0.704 | 0.823913043 |
| Hyper_Insulin_pPS | Biochemistry | 21936 | 18 | 0.005444241 | 0.001702722 | 0.000959467 | 1 | 0 | 0 |
| Hyper_Insulin_pPS | Proteomics | 3547 | 1 | 0 | -0.00087401 | 498.9769964 | 0.0001 | 0.614 | 0.823913043 |
| Hyper_Insulin_pPS | Metabolomics | 4214 | 17 | 9.47E-05 | -0.00179025 | 189.237374 | 0.0001 | 0.739 | 0.823913043 |
| Hyper_Insulin_pPS | Merged_Omics | 2815 | 0 | 0 | -0.000183487 | 627.8462649 | 0.0001 | 0.721 | 0.823913043 |
| Beta_Cell_1_pPS | Anthropometry | 24255 | 8 | 8.14E-05 | -0.000995769 | 10.01611405 | 0.0001 | 0.126 | 0.181730769 |
| Beta_Cell_1_pPS | Biochemistry | 21936 | 5 | 0.011160379 | 0.008098588 | 0.007854458 | 1 | 0 | 0 |
| Beta_Cell_1_pPS | Proteomics | 3547 | 143 | 0.002283894 | -0.000726967 | 44.31595873 | 0.0001 | 0.689 | 0.823913043 |
| Beta_Cell_1_pPS | Metabolomics | 4214 | 123 | 0.001134979 | 0.000197933 | 51.03186725 | 0.0001 | 0.1 | 0.147058824 |
| Beta_Cell_1_pPS | Merged_Omics | 2815 | 1 | 0 | -0.000200278 | 573.6385649 | 0.0001 | 0.719 | 0.823913043 |
| Beta_Cell_2_pPS | Anthropometry | 24255 | 7 | 0.001087764 | 0.001876355 | 0.0008336 | 0.80002 | 0 | 0 |
| Beta_Cell_2_pPS | Biochemistry | 21936 | 21 | 0.044475094 | 0.04094696 | 0.000506957 | 1 | 0 | 0 |
| Beta_Cell_2_pPS | Proteomics | 3547 | 167 | 0.006247045 | 0.003906889 | 15.90363831 | 0.0001 | 0.014 | 0.025 |
| Beta_Cell_2_pPS | Metabolomics | 4214 | 1 | 0.001362767 | 0.000336969 | 0.035566179 | 1 | 0.034 | 0.055434783 |
| Beta_Cell_2_pPS | Merged_Omics | 2815 | 32 | 0.027443101 | 0.006141835 | 0.31333516 | 0.10009 | 0 | 0 |
| Total_GRS | Anthropometry | 24255 | 5 | 0.003124881 | 0.001727443 | 0.003181595 | 1 | 0 | 0 |
| Total_GRS | Biochemistry | 21936 | 21 | 0.033400467 | 0.024040088 | 0.002534447 | 0.10009 | 0 | 0 |
| Total_GRS | Proteomics | 3547 | 181 | 0.034696523 | 0.001020194 | 2.564965296 | 0.0001 | 0.038 | 0.059375 |
| Total_GRS | Metabolomics | 4214 | 1 | 0 | -0.00014177 | 368.6289098 | 0.0001 | 0.748 | 0.823913043 |

**Table S10: Elastic Net Regression Summary Statistics (generated from SCAPIS)**

| PPS | Omic_Layer | Sample_Size | Selected_Variables_Number | r2_train | r2_test | lambda | l1_ratio (alpha) | p_value | fdr_p_adjusted |
|---|---|---|---|---|---|---|---|---|---|
| Total_GRS | Merged_Omics | 2815 | 12 | 0.035588239 | 0.005566847 | 0.044333399 | 1 | 0.002 | 0.004166667 |

**Table S11: Elastic Net Model Coefficients (generated from SCAPIS)**

**a) Anthropometry Models**

| Anthropometry | Cholesterol_pMMS | Lipodystrophy_1_pMMS | Lipodystrophy_2_pMMS | Liver-Lipid_pMMS | ASAT_Pos_pMMS | ALP_Neg_pMMS | SHBG-LpA_pMMS | Obesity_pMMS | VAT_Neg_pMMS | Bilirubin_pMMS | Proinsulin_pMMS | Hyper_Insulin_pMMS | Beta_Cell_1_pMMS | Beta_Cell_2_pMMS | Total_GRS |
|---|---|---|---|---|---|---|---|---|---|---|---|---|---|---|---|
| Waist/hip_Ratio | 0 | 0.05072994 | 0.004775644 | 0.028256559 | 0.009743092 | 0.014394009 | 0 | 0.00623528 | -0.034997685 | 0 | 0 | 0 | 0.000474506 | 0.007736932 | 0.027351913 |
| BMI | 0 | -0.008625366 | 0.010147522 | 0 | -0.007408494 | 0.007454051 | 0 | 0.03266898 | 0.072068841 | 0 | 0 | 0 | 0.000806015 | 0.050505579 | 0.02381983 |
| Waist | 0 | 0 | -0.002275338 | 0 | 0.001627279 | 0.006812882 | 0 | 0 | -0.031755848 | 0 | 0 | 0 | 0.000738624 | 0 | 0 |
| Hip | 0 | -0.023752175 | -0.00890038 | -0.003015972 | -0.007500196 | -0.007026152 | 0 | 0 | -0.020554403 | 0 | 0 | 0 | 0.00049398 | 0 | 0 |
| Weight | 0 | -0.031498078 | -0.004929182 | 0 | -0.008512294 | -0.001066454 | 0 | 0.096722812 | 0 | 0 | 0.013244192 | 0 | 0.000625982 | -0.048869044 | 0 |
| Height | 0 | 0 | -0.025763052 | 0.038437239 | -0.004302385 | -0.008063659 | 0 | 0 | -0.040289167 | 0 | 0 | 0 | 0 | 0 | -0.035947724 |
| DBP_Mean | 0 | 0.009193166 | 0.007847099 | 0.004837055 | 0.005406364 | -0.019840041 | 0 | -0.004467178 | -0.0386711 | 0 | 0 | 0 | 0.000921557 | -0.02672034 | 0 |
| SBP_Mean | 0 | 0.018445611 | -0.002850088 | 0 | 0.012387302 | 0.016514549 | 0 | 0 | 0.034854098 | 0 | 0 | 0 | 0.000416402 | 0.009188133 | 0.001809817 |
| Pulse | 0 | -0.004465425 | 0.002046413 | -0.015868799 | -0.004564344 | -0.015566253 | 0 | -0.003836978 | 0.02119921 | 0 | 0 | 0 | -3.97E-05 | -4.03E-05 | 0.004589562 |
| Sex | 0 | -0.057304397 | 0.002983156 | -0.090104791 | -0.006224097 | -0.008435153 | 0 | -0.119023278 | 0.111157332 | 0 | 0 | 0 | 0 | 0.025697796 | 0 |

**b) Biochemistry Models**

| Biochemistry | Cholesterol_pMMS | Lipodystrophy_1_pMMS | Lipodystrophy_2_pMMS | Liver-Lipid_pMMS | ASAT_Pos_pMMS | ALP_Neg_pMMS | SHBG-LpA_pMMS | Obesity_pMMS | VAT_Neg_pMMS | Bilirubin_pMMS | Proinsulin_pMMS | Hyper_Insulin_pMMS | Beta_Cell_1_pMMS | Beta_Cell_2_pMMS | Total_GRS |
|---|---|---|---|---|---|---|---|---|---|---|---|---|---|---|---|
| Hba1c | 0.017970919 | 0.010134482 | 0.000399622 | 0.02125132 | 0.024897243 | 0.060201129 | 0.014614758 | 0.033732833 | 0.021216221 | 0 | 0.071432452 | 0.022197805 | 0.050810457 | 0.111480026 | 0.101911328 |
| Glucose | -0.028551213 | 0.021270849 | 0.021552626 | 0.029181195 | 0 | 0.031347511 | 0.004222757 | -0.015976693 | 0.038256454 | 0 | 0.037304035 | 0 | 0.084580132 | 0.17304364 | 0.109791872 |
| PGluc_Cap | 0.009215314 | 0 | 0.007444528 | 0.046830658 | 0 | 0.00478625 | 0.00632834 | 0 | 0.010094583 | 0 | 0.023886136 | 0.013504918 | 0 | 0.023438045 | 0.025825129 |
| Insulin | 0.027424104 | 0 | 0.010364826 | 0.062150459 | 0.025182119 | -0.022177328 | -0.018811634 | 0.050823088 | -0.015350062 | 0 | -0.058258385 | 0.030443213 | -0.044627691 | -0.12119727 | -0.059705457 |
| Ldl | -0.018378361 | 0 | 0 | -0.10378593 | 0 | 0 | -0.018945718 | 0 | 0.077173679 | 0 | 0.042460704 | -0.054751652 | 0.004903438 | 0.124684356 | 0.048573869 |
| ApoB | -0.266208027 | 0.020817509 | 0 | -0.011993529 | 0 | -0.06141492 | 0.02412925 | -0.032259314 | 0.022699056 | 0 | -0.032215795 | 0.004768265 | 0 | -0.017868912 | -0.040161439 |
| Cholesterol | 0.067047755 | 0 | -0.017397433 | 0.147531696 | 0 | 0.11038112 | 0 | 0 | -0.129698004 | -0.017192764 | 0 | 0.048144613 | 0 | -0.128857042 | -0.041659318 |
| Lpa | 0.033029473 | 0.00802927 | -0.011866093 | -0.005696333 | -0.00446654 | 0 | 0.064412168 | 0.005267826 | 0.045894636 | 0 | -0.009601009 | 0 | 0 | 0.007180086 | 0.005664405 |
| Tg | -0.015323177 | 0.034209906 | 0 | -0.119726416 | 0.002266989 | -0.028655997 | 0.013357262 | 0.015307981 | 0.053729398 | 0 | 0.003690865 | 0.002215558 | 0 | 0.039634402 | 0.03631492 |
| ApoA1 | 0.176625626 | -0.000643662 | -0.024898287 | -0.10154577 | 0 | 0.024420163 | -0.004363179 | 0.020558531 | -0.017390912 | 0 | 0.034832033 | 0.002799802 | 0 | 0.015692693 | 0.026311029 |
| Hdl | -0.126026769 | -0.030989206 | -0.011166312 | 0.012121672 | 0 | -0.066421494 | 0 | -0.00875317 | 0.047850123 | 0 | -0.041528786 | -0.041444266 | 0 | 0.036963235 | -0.046401936 |
| ALAT | 0.080667184 | 0.002421917 | 0.06734705 | 0.082370888 | 0 | 0.064311967 | 0.004373486 | 0.026840483 | 0.031502725 | 0 | 0.008801273 | 0.00616354 | 0 | -0.016611056 | 0.058641979 |
| ASAT | -0.026979666 | 0.015142968 | 0.017316119 | -0.046710952 | 0.017432482 | -0.061525093 | -0.018169162 | -0.01012156 | -0.009007989 | 0 | -0.008137919 | 0.017792865 | 0 | -0.007821327 | -0.003826404 |
| GGT2 | -0.018835661 | 0 | 0 | -0.068621566 | 0.011215828 | 0.017757419 | 0 | -0.00899732 | -0.003315886 | 0 | 0.003513612 | 0 | 0 | -0.003981245 | -0.012424754 |
| Crp | 0.145423658 | -0.028165886 | -0.025549187 | -0.087411557 | -0.011723298 | -0.00209715 | -0.018430198 | 0.038547348 | -0.011408143 | 0 | 0 | -0.003747313 | 0 | -0.001968499 | -0.006224114 |
| Alfa1Antitrypsin | -0.033470174 | 0 | 0.000201309 | 0.053536832 | -0.015424481 | 0.00382723 | -0.008904263 | -0.015589717 | -0.020645244 | 0 | 0.002476594 | -0.017485027 | 0 | 0.007114065 | -0.012338414 |
| TroponinI | -0.019782836 | 0 | -0.016152663 | 0.01196495 | 0 | 0.008030775 | 0.003385711 | 0.012767116 | -0.002012734 | 0 | 0.000230996 | -0.000221542 | 0 | -0.003858633 | -0.007219992 |
| NTproBNP | -0.014940876 | 0 | 0 | 0.026754802 | -0.003195651 | 0.021556196 | 0.012669179 | 0.006418523 | -0.007873092 | 0 | 0.004817864 | -0.007332727 | 0 | -0.037598075 | -0.017995669 |
| HB | 0.028922404 | -0.014884435 | -0.021747723 | 0.016892496 | 0 | -0.051742241 | -0.02406151 | 0.014594934 | -0.008536558 | 0 | 0.002489745 | -0.021157064 | 0 | -0.02531456 | -0.021689543 |
| Creatinine | 0.024184642 | 0 | -0.027242837 | 0.012473851 | -0.022213305 | 0.01689312 | 0.0204445 | -0.014610609 | -0.019040967 | 0 | 0.006743735 | 0.016469219 | -0.003107577 | 0.000372412 | -0.028691125 |
| CystatinC | 0.005422444 | -0.031515464 | 0 | 0.044437599 | -0.013841796 | 0 | 0 | 0.003302622 | -0.018468508 | 0 | -0.000683585 | -0.036733642 | 0 | 0.005283979 | -0.006919778 |

**Table S11: Elastic Net Model Coefficients (generated from SCAPIS)**

**c) Proteomics Models**

| Proteomics | Cholesterol_pMMS | Lipodystrophy_1_pMMS | Lipodystrophy_2_pMMS | Liver-Lipid_pMMS | ASAT_Pos_pMMS | ALP_Neg_pMMS | SHBG-LpA_pMMS | Obesity_pMMS | VAT_Neg_pMMS | Bilirubin_pMMS | Proinsulin_pMMS | Hyper_Insulin_pMMS | Beta_Cell_1_pMMS | Beta_Cell_2_pMMS | Total_GRS |
|---|---|---|---|---|---|---|---|---|---|---|---|---|---|---|---|
| FABP4 | 0 | -0.001779735 | 0 | 0 | 0.002844678 | 0 | 0.00086148 | 0.005774895 | -0.000991309 | 0 | 0.007457886 | 0 | -0.000673073 | -0.000952983 | -0.000881493 |
| SELP | 0 | 0.002344463 | 0 | 0 | 0.005179644 | -0.104264748 | 0 | -0.001265149 | -0.000821531 | 0 | 0.009805334 | 0 | 4.48E-05 | 0 | 0.000244262 |
| GT | 0 | -0.002707982 | 0.001369547 | 0.024059814 | 0.002546188 | 0.046155822 | 0.000790913 | -0.001641539 | 0.0006611 | 0 | 0.000980492 | 0 | -0.00023997 | -0.000296792 | -0.00378188 |
| PCSK9 | -0.017001656 | 0.005203877 | 0 | -0.057086494 | 0.003534041 | 0 | -0.000518332 | -0.004459756 | -0.000546735 | 0 | 0.010376282 | 0 | -0.000417165 | 0 | -4.92E-05 |
| CTSD | 0 | 0.006388805 | 0.019884307 | 0 | -0.002078614 | 0 | -0.000148801 | 0.002563351 | 0.00054152 | 0 | -0.001611614 | 0 | 0.000528418 | 0.000577462 | 0.008325598 |
| CTSZ | 0.060391648 | 0.002133907 | 0 | 0 | -0.002058194 | 0 | -0.000984046 | 0.001755817 | 0.000561512 | 0 | -0.01574683 | 0 | 6.66E-05 | -0.001409899 | -0.001292341 |
| CA5A | 0.020259027 | -0.000490372 | 0.038017249 | 0 | 0.000680596 | 0.049284066 | 1.01E-05 | 0 | 0.000615653 | 0 | -0.017689981 | 0 | 0 | -0.000873326 | 0.00625432 |
| ACE2 | 0 | -0.003998133 | 0.001387345 | -0.00036554 | -0.003875642 | 0 | -0.000306049 | 0.00514297 | 0.000579348 | 0 | -0.006427443 | 0 | -0.000331734 | -0.001584339 | 0.002531254 |
| IDUA | 0.002915898 | 0.000829705 | 0.016476796 | 0 | -0.011037909 | 0 | 0.00028913 | -0.004297071 | 0.001046623 | 0 | -0.004329335 | 0 | 2.10E-05 | 0.000731357 | 0.001910499 |
| HAOX1 | 0 | -0.003145057 | 0.01516049 | -0.017666418 | 0.000389568 | 0 | -0.000131602 | 0.001551655 | 0.00107726 | 0 | -0.015289438 | 0 | 0 | -0.000702723 | 0.004884039 |
| CPA1 | 0 | 0.004272546 | 0 | -0.003031889 | 0.000107696 | 0 | -0.000966582 | -0.004509605 | -0.00143341 | 0 | 0.002614855 | 0 | -0.000591512 | -0.000313424 | -0.008825723 |
| PON3 | 0 | -0.003084197 | 0 | -0.009007848 | -0.004137653 | 0 | -3.18E-06 | -0.00909565 | -0.001751635 | 0 | -0.012712459 | 0 | -0.000263949 | 0.000614241 | -0.013499088 |
| AP_N | 0 | 0.000874217 | 0 | 0 | -0.005167477 | 0 | -3.51E-06 | 0.001695059 | 0.000723195 | 0 | 0.033382508 | 0 | 0.000923587 | 0.000854986 | 0.00891919 |
| GIF | 0 | 0.002106172 | 0 | 0 | 0.00418607 | -0.080391391 | -0.000319766 | -0.002491771 | -0.000108552 | 0 | -0.032900317 | 0 | -0.000742608 | 2.01E-05 | -0.005993524 |
| ICAM_2 | 0 | 0.000584883 | 0 | 0 | 0.001998948 | -0.160531144 | -0.000355387 | 0.006113332 | -0.00035974 | 0 | -0.037300921 | 0 | 0 | 0 | -0.000968037 |
| PD_L2 | 0 | -0.00087166 | 0 | 0 | 0.002607005 | 0 | 0.000737188 | 0 | 0.000569506 | 0 | -0.005527369 | 0 | -4.02E-06 | -0.000258337 | -0.004636232 |
| Gal_9 | 0 | 0.001779085 | 0 | 0 | 0.001237688 | -0.018416837 | 0.000572878 | 0.005491977 | 0.000359094 | 0 | -0.005790218 | 0 | -2.40E-05 | -0.000701907 | -1.03E-05 |
| SELE | 0 | 0.002674924 | 0 | 0 | 0.010273926 | -0.363407802 | -0.000475214 | 0.006930586 | -0.00048827 | 0 | -0.031572956 | 0 | 0 | 0.000756338 | 0.000413027 |
| TF | -0.013394256 | 0.003433162 | 0.012405003 | 0.002421623 | 0.005926846 | -0.010937673 | 0.000623963 | -0.004615318 | -0.001186033 | 0 | -0.000525435 | 0 | -0.000867321 | 0.000721263 | 0.005177299 |
| ADM | 0 | -0.003250126 | -0.012008778 | 0.003946009 | -0.00074038 | 0 | 0.00086614 | 0.009053802 | -0.000769596 | 0 | 0.008434607 | 0 | -0.000139687 | -0.00081013 | -0.000944543 |
| IL_1ra | 0 | -9.08E-05 | 0 | 0 | 0.00784913 | 0 | 0.000486619 | 0.00484462 | -0.000956109 | 0 | -0.027402057 | 0 | -0.000591924 | -0.00139157 | -0.000157379 |
| IL6 | 0 | -0.00449262 | 0 | 0.014182777 | 0.001521198 | 0 | 0.000502933 | 0.006430918 | 0.000345874 | 0 | 0.004228303 | 0 | 0 | -0.000130435 | 0.004947293 |
| LEP | 0 | -0.004365282 | 0 | 0.043251546 | 0.009060709 | 0 | 0.000325666 | 0.00658214 | -0.000523532 | 0 | -0.001662992 | 0 | -0.000590008 | -0.001035049 | 4.11E-05 |
| IL_1RT2 | 0 | -0.002728154 | 0.004199811 | -0.000984084 | 0.002816309 | 0 | -0.000140217 | -0.002153662 | 9.37E-05 | 0 | 0.005525552 | 0 | 0 | 0.000997196 | 0.001482754 |
| PARP_1 | 0 | 0.00602536 | 0.00521232 | 0 | 0 | 0.002930272 | 0.000966944 | 0.003199564 | 0.000385214 | 0 | 0.010098082 | 0 | -0.000447646 | 0.00092899 | 0.00989405 |
| U_PAR | 0 | 0.002374666 | 0 | 0 | 0.001082906 | 0.053648126 | 0.000714741 | 0.003788284 | -6.21E-05 | 0 | 0.02403047 | 0 | -0.000256276 | 0.000870314 | 0.009245831 |
| LDL_receptor | 0 | 0.00282696 | 0 | -0.026312631 | 0.007020735 | 0 | 0 | 0.002126352 | -0.000246973 | 0 | 0.008745529 | 0 | 0.000484726 | 0.000402534 | 0.004212413 |
| FGF_21 | 0 | 0.00716444 | 0 | -0.068058109 | 0.009007781 | 0 | 0.000217117 | -0.000847895 | 0.00198282 | 0 | -0.012067588 | 0 | 0 | 0 | 0.003338323 |
| IGFBP_2 | 0 | -0.002612601 | 0 | 0.040559593 | -0.00598709 | 0 | -9.37E-05 | -0.004723274 | -0.001841186 | 0 | -0.004524835 | 0 | 0 | 0 | -0.010182028 |
| CPB1 | 0 | 0.003334436 | 0 | 0 | -0.003129521 | -0.101606338 | -0.000988243 | -0.00446661 | -0.001500065 | 0 | -0.016998517 | 0 | -0.000539014 | -0.001054389 | -0.010859615 |
| FGF_23 | 0 | -0.005220118 | 0 | -0.027407983 | 0.008978394 | 0 | -0.000299649 | 0.005828194 | 0.000450825 | 0 | -0.002066854 | 0 | 0.000378689 | 0.001085046 | 0.000999163 |
| vWF | 0 | -0.003412986 | 0 | -0.002997321 | -0.008896949 | 0.144797008 | 7.79E-05 | -0.009076892 | -0.000940652 | 0 | 0.015870598 | 0 | -0.000610262 | -0.000658709 | -0.008423401 |
| TIE2 | 0 | -0.008478504 | 0 | 0 | -0.010498039 | 0.080108215 | -1.29E-05 | -0.002147283 | -7.35E-05 | 0 | 0.020127558 | 0 | -0.000345739 | -0.00050983 | -0.003704237 |
| CDH5 | 0 | -0.006988076 | 0 | 0 | -0.009348889 | 0.176361898 | -5.68E-05 | 0.002199371 | -0.000398117 | 0 | 0.013767829 | 0 | -0.000103636 | 0.001190068 | 0.001965003 |
| TM | 0 | -0.005184948 | 0 | 0.032846598 | 0.002328178 | 0.10238379 | 0.000656121 | 0.001463677 | -0.000137382 | 0 | 0.025017928 | 0 | 0.000332008 | 2.06E-05 | 0.004957693 |
| PECAM_1 | 0 | -0.000190224 | 0 | 0 | 0.000389193 | 0 | 1.31E-05 | 0.001700537 | 0.000189735 | 0 | -0.00438969 | 0 | 0 | 0.000472235 | 1.59E-05 |
| EPHB4 | 0 | -0.002847292 | 0 | 0 | -0.001917758 | -0.125198603 | 0 | 0.000431401 | -9.20E-05 | 0 | 0.000333358 | 0 | 0 | 0.000659937 | -0.003124005 |
| IGFBP_1 | 0 | 0.001075563 | 0 | -0.106690487 | 0.011385353 | -0.03037638 | 0.000631813 | -0.006470553 | -0.000547058 | 0 | -0.005660875 | 0 | -9.92E-05 | 0.001978476 | 0.002042114 |
| FS | 0 | 0.000139175 | 0 | -0.070071972 | -0.007140239 | -0.007715585 | 0.000684218 | -0.003161983 | -6.18E-05 | 0 | -0.000734778 | 0 | 0.000682807 | 0.000927534 | 0.005905141 |
| TFPI | -0.077635102 | -0.000814174 | 0 | 0 | 0.006493953 | 0.049756288 | -1.79E-05 | -0.002394218 | 0 | 0 | -0.000989355 | 0 | 0.000270758 | 0.000152097 | -0.00221557 |
| t_PA | 0 | 0.001720388 | 0 | 0 | -0.006615222 | 0 | -0.000334277 | -0.001240585 | 0.000419331 | 0 | 0.003999578 | 0 | 0.000206192 | 0.000387505 | 0.002068378 |
| CTRC | 0 | -0.000310808 | 0 | -0.044492381 | 0.003656607 | 0.136053058 | -0.001050151 | -0.00050857 | -0.000856451 | 0 | 0.014896312 | 0 | -0.000550108 | 0.000542978 | 0.005718899 |
| MERTK | 0 | -0.003233314 | 0 | -0.000432309 | -0.013501037 | -0.083794071 | 0 | 0.00702127 | -0.000271897 | 0 | 0.002961396 | 0 | 0.000212234 | 0.000117758 | -0.000842433 |
| PRSS8 | 0 | 0.001222975 | 0 | 0 | -0.000383675 | 0.030150381 | 0 | 0.004180525 | 0.000642309 | 0 | -0.003434898 | 0 | -1.34E-05 | 1.23E-05 | 0.002270468 |
| MMP7 | 0 | 0.002627298 | 0.027136648 | 0 | 0.007110766 | -0.010927504 | 0.00046224 | -0.005288572 | 0.001898598 | 0 | -0.012101704 | 0 | 0.00024567 | -0.000462556 | 0.007269906 |
| Gal_4 | 0.004204534 | 0.001515314 | 0.008869639 | 0 | 0.00739029 | -0.206085401 | 3.21E-05 | 0.003530296 | -0.000179383 | 0 | -0.013565711 | 0 | -0.00033425 | 0.001221143 | 0.010158709 |
| SCGB3A2 | 0 | -0.004529833 | 0 | 0 | -0.005284267 | 0 | -0.00011684 | -0.004177358 | -0.000326584 | 0 | 0.01374386 | 0 | 0 | 0.000387442 | -0.006220303 |
| CHIT1 | 0 | -0.003060469 | -0.011867599 | 0 | 0.003790793 | -0.027573389 | 0 | 0.000200607 | -0.000810653 | 0 | -0.014150103 | 0 | -0.000305132 | -0.000941209 | -0.014489365 |
| PTX3 | 0 | -0.001354805 | 0 | 0 | 0.002437143 | -0.010580876 | 0.000156149 | 0.002351801 | -0.001368543 | 0 | -0.00256637 | 0 | 0 | 0 | 0.000535058 |
| MMP_3 | 0 | 0.000747082 | 0 | 0 | 0.00112368 | 0.046850823 | 0 | 0.000611863 | -0.000123909 | 0 | 8.62E-06 | 0 | 0.000215557 | 3.25E-06 | 0.002089922 |
| MMP_2 | 0 | -0.002488023 | 0 | 0 | -0.002708054 | 0.024970874 | 0 | -0.001279668 | -0.000851475 | 0 | 0.005504391 | 0 | -9.05E-05 | 0.000173724 | 0.00067752 |
| BNP | 0 | -0.002707418 | 0 | 0 | -0.003894784 | 0 | 0.00047888 | 0.003523459 | -0.000383466 | 0 | -0.001971051 | 0 | 0.000268262 | 0.000759005 | 0.003326116 |
| ST2 | 0 | -0.003167201 | 0 | 0 | -0.005566935 | 0 | 0 | -0.00352585 | 0 | 0 | -0.005842664 | 0 | -3.27E-05 | -0.00151836 | -0.007641676 |
| XCL1 | 0 | 0.004755884 | 0 | 0.02134244 | 0.00532484 | -0.003492738 | 0.000453096 | -0.00068232 | 0.0009692 | 0 | -0.012227709 | 0 | -1.27E-05 | -3.94E-05 | 0.003521514 |
| KLK6 | 0 | -0.005418984 | 0 | 0 | 0.001049728 | 0.009409316 | -0.000256504 | -0.005218367 | -0.000267994 | 0 | 0.002350043 | 0 | 0 | 0.001674428 | 6.64E-05 |
| CD40_L | 0 | -0.000752369 | 0 | 0 | 0.003470032 | 0 | 6.30E-06 | -0.000879529 | 0.000197496 | 0 | -0.006700244 | 0 | -3.40E-05 | 0 | -0.002774453 |
| TNFRSF14 | 0 | -0.001723284 | 0 | 0 | 0.002665207 | 0 | 0.00039434 | -0.000245844 | 0.000282195 | 0 | 0.005286319 | 0 | -8.90E-05 | 0.000720079 | -5.24E-05 |
| CD4 | 0 | -0.00047089 | 0.002967066 | 0 | 0.002980817 | -0.003261685 | 0.001489604 | 0.000986568 | 0.000322191 | 0 | 0.022468954 | 0 | 0.00041295 | -0.000931201 | 0.007842068 |
| Gal_3 | 0 | 0.002217396 | 0 | 0 | 0.002410917 | 0.011102381 | 0.000461649 | -0.001354993 | 0 | 0 | 0.00730759 | 0 | 0.000163092 | 0.002169547 | 0.004960665 |
| HB_EGF | 0 | 0.001298276 | 0 | 0 | 0.003913572 | 0 | -0.000146086 | -0.002997389 | -0.000347039 | 0 | 0.000639182 | 0 | -6.65E-05 | -0.001162415 | -0.006685188 |
| DLK_1 | 0 | 0.002342082 | 0 | 0.023180507 | 0.009624318 | 0 | 0.000333845 | 0.001862045 | 0.000479166 | 0 | -0.006672053 | 0 | 0.000431141 | 0.001712233 | 0.011546706 |
| REN | 0 | -0.001030704 | 0 | 0 | -0.002173482 | 0 | 0 | -0.003435481 | 0.000438744 | 0 | 0.001907982 | 0 | -0.000655278 | 0.00072619 | -0.00327439 |
| TIMP4 | 0 | -0.004544198 | 0 | 0.018756943 | -0.00557292 | 0.009991015 | 0 | -0.001233135 | -0.000778257 | 0 | -0.003403213 | 0 | -0.00020332 | 0.000631311 | -0.004067935 |
| BMP_6 | 0 | 0.00085169 | 0 | 0.005605439 | 0.01104881 | 0.02089574 | 0.000218136 | -0.000949746 | 8.45E-05 | 0 | 0.01967454 | 0 | -0.000161184 | 0.00167848 | 0.010055678 |
| PSP_D | 0 | -0.000344435 | 0 | 0.010602952 | 7.53E-05 | 0.017024069 | -0.000578807 | 0.007052064 | -5.14E-05 | 0 | 0.011473331 | 0 | 0 | 0.000647958 | 0.006405949 |
| CD163 | 0 | 0.001716598 | 0 | 0 | 0.003011555 | 0.012560269 | 0.00062222 | 0.007907174 | -4.38E-05 | 0 | 0.0141064 | 0 | -0.000285196 | 0.00084298 | 0.00989699 |
| PRELP | 0 | -0.003233703 | 0 | 0 | -0.00096258 | 0 | 0.001409711 | 0.001575275 | -0.00139725 | 0 | -0.001686058 | 0 | -0.000626792 | -0.000362859 | -0.005418338 |
| PGLYRP1 | 0 | -0.00105347 | 0 | 0 | 0.006484084 | 0 | -0.000263401 | -0.002830572 | -0.000898463 | 0 | 0.008424208 | 0 | 0 | -0.000820652 | 2.44E-05 |
| AMBP | 0 | -0.003201057 | 0 | 0 | -0.00503912 | -0.033245726 | 9.33E-05 | -0.000886321 | -3.50E-05 | 0 | 0.001736024 | 0 | 0 | -0.001217198 | -0.006719516 |
| FABP2 | 0 | -0.003968578 | 0 | 0 | -0.001592838 | 0.083649424 | 0.000202925 | 0.00298539 | -0.001610557 | 0 | -0.017515176 | 0 | -0.000692825 | -0.000323417 | -0.003360122 |
| HO_1 | -0.006896782 | -0.003976838 | 0 | 0 | 0.008858107 | 0 | 0.000203543 | -0.000973272 | 0.000325227 | 0 | 0.004584256 | 0 | -0.00052849 | 0.000956167 | 0.000943606 |
| COL1A1 | 0 | -0.000849523 | 0 | 0 | -0.007147836 | -0.014799295 | -0.000358786 | -0.008610897 | -0.001392367 | 0 | -0.02072248 | 0 | -0.000164801 | 0.000883573 | -0.011340777 |
| DCN | 0 | -0.002780534 | 0 | 0.01310322 | -0.007622644 | -0.013275131 | 0.00057381 | 0.002737055 | -5.73E-05 | 0 | -0.000539458 | 0 | -0.000562117 | 0 | 0.000744736 |
| CXCL16 | 0 | 0.005764551 | 0 | 0 | 0.004138855 | 0 | 3.83E-05 | -0.001889667 | -0.001173378 | 0 | -0.009667432 | 0 | 0 | 0.000211755 | 0.002057209 |
| GLO1 | -0.005681934 | 0.003129802 | 0 | 0 | -0.004040218 | -0.00040351 | -0.000695939 | 0.00183794 | 0.000590342 | 0 | 0.006082189 | 0 | -0.000297044 | -0.001326798 | 0.001188658 |
| MMP_9 | 0 | -0.005192287 | 0 | 0 | -0.002775725 | 0.029519061 | 0 | 0.000736674 | -0.000667865 | 0 | 0.008901451 | 0 | -0.00076807 | -0.000181723 | -0.003616468 |

**Table S11: Elastic Net Model Coefficients (generated from SCAPIS)**

**c) Proteomics Models**

| Proteomics | Cholesterol_pMMS | Lipodystrophy_1_pMMS | Lipodystrophy_2_pMMS | Liver-Lipid_pMMS | ASAT_Pos_pMMS | ALP_Neg_pMMS | SHBG-LpA_pMMS | Obesity_pMMS | VAT_Neg_pMMS | Bilirubin_pMMS | Proinsulin_pMMS | Hyper_Insulin_pMMS | Beta_Cell_1_pMMS | Beta_Cell_2_pMMS | Total_GRS |
|---|---|---|---|---|---|---|---|---|---|---|---|---|---|---|---|
| Notch_3 | 0 | 0.000893078 | 0 | 0 | 0.001624713 | -0.02697347 | 0 | 0.004921088 | 7.33E-05 | 0 | -0.004764171 | 0 | -3.88E-05 | 0.000755755 | 0.005714457 |
| IL_4RA | 0 | -0.00320204 | 0 | 0 | -0.001801061 | 0.003415115 | 0 | -0.00727891 | -0.000103643 | 0 | 0.00692568 | 0 | 0.000211331 | -0.000739974 | -0.005406533 |
| SERPINA12 | 0 | 0.001359163 | 0.020401323 | 0 | -0.002612151 | 0.005747683 | 0.000835868 | 0.000983149 | 0.000677954 | 0 | -0.008985387 | 0 | -0.000529211 | 0.001044448 | 0.005338445 |
| Ep_CAM | 0 | 0.002829279 | 0 | 0 | 0.003810337 | 0.009226379 | 0.000313328 | -0.000922241 | -0.001225558 | 0 | 0.004332979 | 0 | -0.000399451 | 0.000334328 | 0.0049266 |
| LPL | 0 | -0.001856934 | 0 | 0 | -0.001561192 | -0.046663236 | 0.000452128 | 0.006159175 | -0.001338845 | 0 | 0.000949647 | 0 | -0.000295938 | -0.000673664 | 0.001618125 |
| HSP_27 | 0 | -0.000662734 | 0 | 0 | -0.012064806 | 0.01851671 | 0.000222436 | -0.002068265 | -0.000522443 | 0 | 0.014611565 | 0 | 0 | -0.001646539 | -0.001461235 |
| RAGE | 0 | 0.00343115 | 0 | 0 | -0.000124717 | -0.028945033 | 0.000956395 | -0.006406016 | 0 | 0 | -0.015560347 | 0 | -0.000604856 | 0.000396907 | -0.002983963 |
| IL_27 | 0 | 0.000561302 | 0 | 0 | -0.005778188 | 0 | 0.000390081 | -0.001200116 | 0 | 0 | -0.002946188 | 0 | 0 | -0.000385069 | -0.004095978 |
| DECR1 | 0 | 0 | 0 | 0 | -0.000968867 | -0.010585386 | 0.000378919 | -0.000805944 | 0.000376085 | 0 | -0.006508664 | 0 | 0 | 3.50E-05 | 0 |
| MEPE | 0 | -0.005405638 | 0 | 0 | 0.001762238 | 0 | 0 | -0.005284042 | -0.001723595 | 0 | 0.017961815 | 0 | 0.000329332 | 0.001257564 | -0.005690976 |
| TNFRSF10A | 0 | 0.003586639 | 0 | 0 | 0.007419763 | 0.021287533 | 0.00067616 | -0.006118952 | 7.94E-05 | 0 | -0.01948214 | 0 | -0.000378544 | -0.001303436 | -0.001968309 |
| OPN | 0 | -0.00131155 | 0 | 0.010936528 | -0.004682439 | 0 | 0.000479511 | -0.003595229 | -0.001676767 | 0 | -0.003390408 | 0 | -0.000280044 | 0.000952539 | -0.004633391 |
| RETN | 0 | -0.001516853 | 0 | 0 | -0.000816237 | 0 | -3.06E-05 | 0.000384508 | -0.000768056 | 0 | -0.015651208 | 0 | -0.000273684 | -0.001459866 | -0.003001334 |
| Dkk_1 | 0 | 0.00143933 | 0 | 0 | 0.002996529 | 0 | -0.000238656 | -0.001473965 | -0.000442057 | 0 | 0.004820504 | 0 | -6.09E-05 | 0 | -0.000750736 |
| CCL3 | 0 | -0.002126094 | 0 | 0 | -0.00042339 | -0.010610301 | 2.36E-05 | 0.000107722 | 0 | 0 | -0.0097665 | 0 | -8.06E-05 | -0.00217022 | -0.012045387 |
| SOD2 | 0 | -0.002391297 | 0 | 0 | -0.005522308 | 0.000951533 | 0.000795377 | -0.005077057 | 0.000547909 | 0 | -0.004461915 | 0 | 0.000226438 | -0.001035029 | -0.007942504 |
| IL1RL2 | 0.005779244 | 0.000460763 | 0 | 0 | 0.003663077 | 0 | 0.001095941 | -0.003813684 | 0 | 0 | 0.01021155 | 0 | 0.000870915 | -0.000259547 | -0.001612284 |
| MCP_1 | 0 | 1.49E-05 | -0.003930896 | 0 | -0.004159759 | 0.008843914 | -0.000941588 | 0.002766774 | -0.001228277 | 0 | 0.010107305 | 0 | -6.93E-05 | 0 | -0.004134438 |
| CCL24 | 0 | 0.000654274 | 0 | 0 | 0.000730497 | 0 | 0.000490651 | -0.001427781 | -0.000284479 | 0 | -0.004880097 | 0 | -0.000144676 | 0 | -0.00165548 |
| SHPS_1 | 0 | -0.004711001 | 0 | 0.004943689 | 0 | 0 | 0.000194006 | -0.004675135 | 9.48E-05 | 0 | -0.007992319 | 0 | 0.000175599 | 0.000268699 | -0.003739699 |
| ITGB2 | 0 | 0.000733803 | 0 | 0 | 0.00598521 | 0 | 0.000591407 | 0.001355316 | -0.000378327 | 0 | 0.022132737 | 0 | 0.000372139 | 0.001992935 | 0.009801159 |
| TLT_2 | 0 | -0.002164679 | 0 | 0 | -0.006880334 | 0.042948692 | -1.17E-05 | 0.002250327 | -3.04E-05 | 0 | 0.010911233 | 0 | -7.79E-06 | 0.000182751 | 0.002684994 |
| JAM_A | 0 | 0.000988367 | 0 | 0 | -0.001083785 | 0 | 0.0002204 | -0.000159366 | 0.000120742 | 0 | -0.006774444 | 0 | 0 | -0.00010491 | -0.000329539 |
| BLM_hydrolase | 0 | -0.000253413 | 0 | 0 | -0.003476754 | 0 | -1.00E-05 | -0.001169663 | -0.000583678 | 0 | -0.000280975 | 0 | 0.000105796 | 0.000623314 | -0.001961028 |
| PI3 | 0 | 0.003544662 | 0 | 0 | 0.003397435 | 0 | -0.000161934 | -0.000922513 | -9.23E-05 | 0 | -0.012425678 | 0 | -0.000226895 | -0.001107023 | -0.003776565 |
| ANG_1 | 0 | 0.00222149 | 0 | 0 | 0 | -0.035447285 | -0.000291233 | -0.003848178 | 0 | 0 | -0.00349086 | 0 | -0.000407623 | -2.68E-05 | -0.00454279 |
| TNFRSF10C | -0.002211227 | 0.002792498 | 0 | 0 | -0.005569637 | 0 | 4.90E-05 | -0.006893373 | -0.000595591 | 0 | 0.00848191 | 0 | -0.000406509 | -0.000549413 | -0.001997515 |
| hOSCAR | 0 | -0.001570306 | 0 | 0 | -0.00762097 | -0.014419628 | 0.000427661 | 0.005556221 | -0.000407065 | 0 | -0.004395225 | 0 | -0.000430337 | -0.001856096 | -0.003640301 |
| GH | 0 | -2.32E-05 | 0 | 0 | 0.000419181 | -0.031847466 | 0.000525093 | -0.000725117 | 0.000150204 | 0 | -0.001558916 | 0 | -0.000116436 | -0.001046591 | -0.001510182 |
| RARRES2 | 0 | -0.001118695 | 0 | 0 | 0.004689231 | 0.073574675 | -0.00012042 | 0.005932067 | -0.000558799 | 0 | -0.004020449 | 0 | 4.03E-05 | -0.001307511 | 0.003598154 |
| CD93 | 0 | -0.003095351 | 0 | 0 | -0.005514533 | -0.021168368 | 0.00027025 | 0.003080592 | -0.000166277 | 0 | 0.007451765 | 0 | 0.000171755 | 0.001644974 | 0.003746512 |
| KIM1 | 0 | -0.001341135 | 0 | 0 | 0.000406479 | 0 | -6.86E-05 | -0.001202624 | -0.000644501 | 0 | 0.010655818 | 0 | 0 | 0.000340036 | 0.002765466 |
| FAS | 0 | -0.001047628 | 0 | 0 | 0.001012806 | -0.013478914 | 0.000462549 | 0.001076054 | 0 | 0 | 0.000932006 | 0 | -0.000163642 | -0.000547831 | 0.004840338 |
| PIgR | 0 | -0.001486329 | 0 | 0 | 0.000456952 | -0.021731842 | 0.000442026 | 0.002294455 | -0.000246979 | 0 | -0.008216482 | 0 | -0.000579381 | -0.001386623 | -0.003430679 |
| CCL15 | 0 | 0.006204738 | 0 | 0 | 0.001649748 | -0.002773306 | -0.000117999 | 0.003619116 | -0.000389732 | 0 | -0.006356649 | 0 | -0.000109745 | -0.000514747 | 0.000137779 |
| CTSL1 | 0 | -0.002230968 | 0.0023302 | 0 | 0.003163603 | 0 | -0.000237137 | 0.002223269 | 0 | 0 | 0.00262321 | 0 | -0.000414847 | 0.00031408 | 0.004692051 |
| TNF_R1 | 0 | -0.002127283 | 0 | 0 | 0.003633918 | 0.018198723 | 0.00046788 | 0.001276692 | -0.00066999 | 0 | -0.003636089 | 0 | -0.000225322 | -1.19E-05 | -0.001712774 |
| PAI | 0 | 0.00316668 | 0 | 0 | 0.007906067 | 0.051011585 | -0.000528541 | -0.001611081 | 0 | 0 | -0.008251783 | 0 | -0.000586771 | -0.000655573 | -0.003388151 |
| VEGFD | 0 | 0.004276224 | 0 | 0 | -0.008867824 | -3.97E-06 | 0.000946034 | 0.004726581 | 0.000461136 | 0 | 0.012638655 | 0 | -8.98E-05 | -0.000376748 | 0.002701232 |
| CNTN1 | 0 | 0.003654211 | 0 | 0 | 0.012287758 | 0 | 0.000347756 | -0.001051902 | 0.000324766 | 0 | -0.013674983 | 0 | 0 | 0.001918477 | 0.005601812 |
| PDGF_subunit_A | 0 | 0.003830906 | 0 | 0 | -0.000531402 | 0 | -0.000483726 | -0.00225698 | -0.000488909 | 0 | -0.002528446 | 0 | -0.000226857 | 0 | 0.000184533 |
| CCL16 | 0 | 0.001732606 | 0 | 0 | 0.006520803 | 0 | 5.49E-06 | -0.000440588 | -0.000486233 | 0 | -0.006275246 | 0 | 7.51E-05 | -0.001966674 | -0.000139905 |
| IGFBP_7 | 0 | -5.34E-05 | 0 | 0 | 0.012535579 | 0.030544846 | -7.11E-07 | -0.005035464 | -1.53E-05 | 0 | -0.012122404 | 0 | -0.000318083 | 0.000254697 | -0.001588891 |
| IL_6RA | 0 | -0.00321306 | -0.016194286 | -0.03010391 | -0.001675049 | -0.00184245 | 0 | 0.000838748 | -0.001021669 | 0 | 0.005160283 | 0 | 0.000444709 | 0.000860877 | -0.008276458 |
| GRN | 0 | -0.001642578 | 0 | 0 | 0.003665776 | 0.072847182 | -9.27E-05 | 0.003289244 | -0.000468029 | 0 | -0.010887174 | 0 | 0.000185705 | 0.001181975 | -0.001235852 |
| LOX_1 | 0 | -0.001481893 | 0 | 0 | 0.002453913 | 0 | 0.000219464 | 0.001126092 | -0.000100342 | 0 | 0.006044243 | 0 | -0.00045698 | -5.02E-05 | -0.001972214 |
| TNFRSF13B | 0 | 0.006461166 | 0 | 0 | -0.00185466 | 0 | 0.001446248 | 0.001436416 | 0.000310547 | 0 | 0.006752153 | 0 | 0.000761812 | 0.001106145 | 0.004367915 |
| GP6 | 0 | -0.001322123 | 0 | 0 | 0.007303636 | 0 | 3.32E-05 | 0.003266278 | 0 | 0 | -0.00225701 | 0 | 0 | 0.000390189 | 0.004092461 |
| CASP_3 | 0 | 0.000365374 | 0 | 0 | 0.002359347 | 0 | 0.000212416 | 0.001403424 | 1.29E-05 | 0 | 0.009951986 | 0 | 0 | 9.18E-05 | 0.003087002 |
| SPON1 | 0.010998544 | 0.002214766 | 0 | 0 | -0.001204644 | 0.006338062 | 0.00038019 | 0.001013346 | -0.000193046 | 0 | 0.001973085 | 0 | -0.000652565 | -0.000947887 | 0.00306066 |
| IL18 | 0 | 0.00278474 | 0.000514389 | 0 | -0.000295 | 0 | -0.000198226 | 0.002531863 | 0.00115464 | 0 | 0.010972977 | 0 | 0 | 0.000772463 | 0.008480553 |
| PRSS27 | 0 | -0.000972953 | 0 | 0.005577972 | 0.001882714 | 0 | 2.98E-05 | -0.003065222 | 0.000904015 | 0 | -0.017481977 | 0 | 0.000385083 | 0.00122612 | -0.00500808 |
| AZU1 | 0 | 0.001380216 | 0 | -0.014027059 | 0.000263537 | 0.019728968 | -0.000187515 | 0.000242597 | -0.000950394 | 0 | -0.007338174 | 0 | -0.000471014 | -0.000618638 | -0.001265126 |
| CEACAM8 | 0 | 9.83E-05 | 0 | 0.009456485 | -0.002755203 | 0 | 4.44E-05 | 0.002169963 | 0 | 0 | -0.00539299 | 0 | -0.000353845 | -0.001538005 | -0.006460925 |
| BOC | 0 | 0.001540807 | 0 | 0.001389306 | 0.000480478 | 0 | 0.000328338 | -0.001292436 | 0.000608627 | 0 | -0.010229076 | 0 | -0.000438697 | 0.001361063 | 0.004857617 |
| MB | 0 | 0.000976144 | -0.001964009 | 0 | -0.004912829 | -0.018380376 | -9.49E-05 | 0.001828354 | -0.000483174 | 0 | -0.002795621 | 0 | 0.000534017 | -0.00068581 | -0.006008034 |
| PAPPA | 0 | -0.001147661 | 0 | 0 | 0.005567382 | -0.020388954 | -0.000930473 | -0.011291247 | -0.00142472 | 0 | 0.014162334 | 0 | -0.000381138 | -0.000572241 | -0.008317624 |
| TNF_R2 | 0 | -0.002718917 | 0 | 0 | -0.004757956 | 0.002798076 | 0.000474165 | 0.001857321 | -6.89E-06 | 0 | -0.002766118 | 0 | -0.000260387 | 0.000115678 | -0.000747377 |
| TR | 0 | 0.003559401 | 0 | 0 | -0.000905434 | -0.017628481 | 0.000120401 | 0.005160146 | 0 | 0 | 0.007532706 | 0 | 0 | 0.001213383 | 0.003981101 |
| GDF_15 | 0 | -0.000259739 | 0 | 0.022751638 | -0.006089282 | 0.010152665 | 0.000112219 | -0.001496347 | -3.78E-07 | 0 | -0.002270211 | 0 | -4.06E-05 | -0.001136966 | -0.002500228 |
| PLC | 0 | -0.00065572 | -0.01347314 | 0 | -0.01075274 | 0 | 0.000719147 | -0.004108269 | -0.000930289 | 0 | -0.007643309 | 0 | 0 | -0.000380139 | -0.011686563 |
| PAR_1 | 0 | -0.00277347 | 0 | 0.027456501 | 0.002218187 | 0.040981741 | 0.00069166 | 0.004950323 | 0.000151539 | 0 | -0.006888241 | 0 | -8.41E-05 | 0.000652829 | 0.001987092 |
| CSTB | 0 | -0.003864607 | 0 | 0 | -0.007714873 | 0 | -0.000301141 | 0.003278378 | -0.000333138 | 0 | 0.001348016 | 0 | 0 | 7.12E-06 | -0.002944253 |
| NEMO | 0 | -0.00020733 | 0 | 0 | 0.002904311 | -0.025465331 | 0.000341209 | -0.00299498 | 0.0005944 | 0 | 0.001300075 | 0 | 0.000112203 | 0.000339763 | -0.000817792 |
| uPA | 0 | 0.001390048 | 0 | 0.021577798 | 0.00518024 | 0 | 0 | 0.00141042 | -0.000484339 | 0 | -0.00181653 | 0 | 0 | 0.001398511 | 0.004123306 |
| CHI3L1 | 0 | 0.004805743 | 0 | 0 | 0.006975052 | 0 | 0 | 0.002738783 | 8.00E-05 | 0 | 0.005948685 | 0 | -8.77E-05 | -0.001087781 | 0.0026803 |
| TNFSF13B | 0 | 0.000805427 | 0 | 0 | -0.008086241 | 0.028102792 | -0.00011333 | 0.006061946 | 0.000228544 | 0 | 0.006202973 | 0 | 0 | 0.000269964 | 0.008298457 |
| CXCL1 | 0 | -0.000690669 | 0 | -0.012477963 | -0.005103746 | 0 | -0.000584422 | -0.003972284 | 0.000427713 | 0 | 0.003590055 | 0 | 0 | 0.000718842 | -0.000337426 |
| IL_17RA | 0 | -0.004903462 | 0 | 0 | -0.014338084 | 0.037694536 | -0.001370272 | -0.001473074 | 0.00035483 | 0 | -0.000981101 | 0 | 0 | 0.002230261 | 0.002624954 |
| NT_proBNP | 0 | 0.000633642 | 0 | 0 | -0.002220172 | 0 | 0.000585913 | 0.000402699 | -0.000638161 | 0 | 0.015529988 | 0 | 0.000223648 | 0.001349936 | 0.005459417 |
| MARCO | 0 | 0.00035459 | 0 | 0 | -0.005657396 | -0.048554277 | 0.000457281 | -0.001010178 | -0.000187658 | 0 | -0.017878652 | 0 | 0 | 0.002004361 | 0.002214953 |
| TNFRSF11A | 0 | -0.002137517 | 0 | 0 | -0.000662659 | 0 | 0.001109101 | 0.004672379 | -0.001273045 | 0 | 0.014048232 | 0 | -0.000171178 | -0.000802706 | 0.000622671 |
| ALCAM | 0 | 0.003171138 | 0 | 0 | 0 | 0 | 0.000684354 | 0.001803767 | -0.000621149 | 0 | 0.004742759 | 0 | 0 | 0.001200727 | 0.005245732 |
| IgG_Fc_receptor_II | 0 | 0.000958259 | -0.001886958 | 0.00871273 | 0.003404159 | -0.017086834 | -0.000245934 | -0.00137158 | 0.000405181 | 0 | 0.002372534 | 0 | -8.25E-05 | -0.001639046 | -0.000181006 |
| IL2_RA | 0 | 0.004788462 | 0.016158396 | 0 | 0.006920585 | 0 | 0 | 0.004292558 | -0.000782814 | 0 | 0.005166731 | 0 | 0 | 0.001463627 | 0.010476151 |
| SCF | 0 | -0.00377394 | 0 | -0.03606453 | -0.004780087 | -0.08705834 | 0.000367156 | -0.007791474 | 0 | 0 | -0.033455389 | 0 | -0.000389719 | 0 | -0.011090949 |

**Table S11: Elastic Net Model Coefficients (generated from SCAPIS)**

**c) Proteomics Models**

| Proteomics | Cholesterol_pMMS | Lipodystrophy_1_pMMS | Lipodystrophy_2_pMMS | Liver-Lipid_pMMS | ASAT_Pos_pMMS | ALP_Neg_pMMS | SHBG-LpA_pMMS | Obesity_pMMS | VAT_Neg_pMMS | Bilirubin_pMMS | Proinsulin_pMMS | Hyper_Insulin_pMMS | Beta_Cell_1_pMMS | Beta_Cell_2_pMMS | Total_GRS |
|---|---|---|---|---|---|---|---|---|---|---|---|---|---|---|---|
| THPO | 0 | -0.002755839 | 0 | 0 | -0.005520737 | -0.007679221 | 0 | 0.003710403 | -0.000503103 | 0 | -0.01536251 | 0 | -0.000432464 | -7.96E-05 | -0.004281032 |
| STK4 | 0 | 0.001518236 | 0 | 0 | -0.003999529 | -0.000551392 | 0.000926086 | -0.000994727 | 0.000320913 | 0 | -0.006449458 | 0 | 0 | -0.000286842 | -0.004541107 |
| CD84 | 0 | 0.002045798 | 0 | 0 | 0.005997011 | 0.00946041 | 0.000177706 | 0.00087038 | 0.000101669 | 0 | 0.008626904 | 0 | -0.000352406 | -0.000508592 | 0.004648023 |
| PRTN3 | 0 | 0.000629964 | 0 | 0 | 0.00942508 | -0.013158474 | -0.000239803 | 0.00396934 | -0.001253254 | 0 | -0.010168779 | 0 | 0 | -0.001056878 | -0.000988814 |
| TRAIL_R2 | 0 | 0.00213625 | 0 | 0 | 6.59E-06 | -0.006165633 | 0.000125409 | 0 | 0 | 0 | 0.012563232 | 0 | -6.69E-05 | 0 | 0.007905708 |
| TR_AP | 0 | 0.00438124 | 0 | 0.014129063 | -0.008318988 | 0 | -0.00081768 | -0.008999254 | -0.000373172 | 0 | 0.006834038 | 0 | 0.000164272 | 0.002798347 | 0.004646515 |
| EGFR | 0 | 0.000790502 | 0 | -0.000533156 | 0.002396059 | -0.021354475 | 0 | -0.001575698 | -0.0007661 | 0 | -0.004505887 | 0 | 0.000610021 | 0.000935749 | 0.00035041 |
| THBS2 | 0 | 0.005229938 | 0 | 0 | 0.004344006 | 0 | 0.000889351 | 0.008564573 | 0 | 0 | 0.000924963 | 1.15E-19 | -0.000887343 | -0.000598863 | 0.00604193 |
| ITGB1BP2 | 0 | -0.002248156 | 0 | 0 | 0.003364664 | 0 | 0.00048146 | -0.000407676 | 0.000649083 | 0 | 0.017999223 | 0 | 5.73E-05 | 0.000512608 | 0.005068608 |
| IL_1RT1 | 0 | -0.002847589 | 0 | 0 | -0.004366269 | 0.053228455 | -0.000550051 | -0.002943606 | -0.000605712 | 0 | 0.003749666 | 0 | 8.23E-05 | 0.000428905 | -0.004265604 |
| GDF_2 | 0 | 0.000690573 | -0.01800294 | -0.004094789 | 0.002232063 | 3.97E-05 | 0.000571755 | -0.005484064 | -0.000231076 | 0 | -0.000123189 | 0 | 0.000160974 | 0 | -0.00739133 |
| IL_18BP | 0 | -0.00096716 | 0 | 0 | -0.003202988 | 0 | 0.000123813 | 0.00201558 | 0.000320548 | 0 | 0.011457435 | 0 | 0 | 0.000601239 | 0.002757883 |
| MMP12 | 0 | 0.001880422 | 0 | 0 | -0.002980496 | 0.017795387 | 0.000604235 | 0.000124223 | -0.000139908 | 0 | -0.012564077 | 0 | 0.000120598 | 0 | 0.002638591 |
| SORT1 | 0 | -0.001967328 | 0 | 0 | -0.003679742 | 0.015716435 | 0.000312904 | 0.000727619 | 0 | 0 | 0.016034004 | 0 | -2.13E-05 | 0.000257794 | 0.001696387 |
| AXL | 0 | -0.003816995 | 0 | 0 | -0.011303613 | 0.01594402 | 4.93E-05 | -0.002179808 | 0 | 0 | 0.005909581 | 0 | 0.000203016 | 0.001663855 | 0.002625653 |
| AGRP | -0.033675164 | 0.006114054 | -0.010325099 | 0 | 0.00142235 | 0 | 0.000426317 | 0.000236039 | -0.000734027 | 0 | 0.019660823 | 0 | 0.000415374 | -0.001531795 | -0.00167092 |
| PSGL_1 | 0 | 0.001740341 | 0 | -0.015757431 | -0.004680553 | 0 | 0.000968112 | 0.001512116 | 0.000737334 | 0 | 0.010554377 | 0 | -0.000155045 | 0.001424054 | 0.009055385 |
| VSIG2 | 0 | -0.000819561 | 0 | 0 | 0.009917125 | 0.023677169 | 0.000350732 | -0.006087426 | 0.00143748 | 0 | 0.012214257 | 0 | -0.000119077 | 0.001406004 | 0.008118313 |
| TGM2 | -0.012350685 | 0.001094196 | 0 | 0 | -0.010493057 | -0.019120083 | 7.88E-05 | -0.001133813 | 0 | 0 | -0.010308456 | 0 | 0 | -0.001503557 | -0.007814625 |
| PGF | 0 | 0.000107172 | 0 | 0 | -0.00440306 | -0.017911323 | 0.000378762 | 0.000435896 | -2.76E-05 | 0 | 0.004747903 | 0 | -0.000300659 | -0.000634627 | -0.00187209 |
| SRC | 0 | -0.000307151 | 0 | 0 | -0.001535392 | 0 | 0 | 0.00193541 | 0 | 0 | 0.008844452 | 0 | 0.000122551 | 0.000317618 | 0.001490591 |
| ADAM_TS13 | 0 | -0.002539163 | 0 | 0 | -0.005365044 | 0 | 0.00051007 | 0.000691377 | -0.000236135 | 0 | -0.007115509 | 0 | 0 | -0.00162939 | -0.002614009 |
| IL16 | 0 | 0.000220404 | 0 | 0.001778123 | 0.002856771 | -0.018729948 | 0.000181729 | -0.00022588 | -0.000195997 | 0 | -0.015694761 | 0 | -0.0001417 | 0 | -0.000787401 |
| IL_17D | 0 | -0.000957531 | 0 | 0 | 0.000413796 | 0 | 0.000133798 | -0.010723006 | -0.000965506 | 0 | -0.012981307 | 0 | -0.000786902 | -0.001800783 | -0.009333013 |
| MPO | 0 | 0.005744778 | 0 | 0 | -0.003085951 | 0.001298746 | 0 | -0.001102645 | 0 | 0 | 0.005587007 | 0 | -6.93E-05 | 0.000330299 | 0.003730468 |
| OPG | 0 | 0.001030106 | 0 | 0 | 0.007496644 | -0.030433328 | -3.28E-05 | 0 | -0.000548269 | 0 | 0.010047737 | 0 | -0.000578223 | -0.000654533 | 0 |
| SLAMF7 | 0 | 0.002660199 | -0.001547389 | 0 | 0.00526876 | 0.041609139 | -0.000496396 | -0.00020341 | 0.001388403 | 0 | -0.012262446 | 0 | -0.000116926 | 0.000320136 | -0.003637336 |
| TFF3 | 0 | 0.002189103 | 0 | 0 | -0.003965281 | 0 | 0.000175497 | 0.004054936 | -0.000230545 | 0 | -0.002529093 | 0 | 8.60E-05 | 0.000194302 | 0.00211692 |
| PDGF_subunit_B | 0 | 0.003005257 | 0 | 0 | 0.000154066 | 0 | -0.000182635 | -0.002713402 | -0.000132506 | 0 | -0.004683836 | 0 | -0.000277033 | -5.89E-05 | -0.001593624 |
| CCL17 | 0 | 0.00368838 | 0 | -0.005084286 | 0.010057782 | 0.002851849 | 0.000259159 | 0.002258698 | 0 | 0 | 0.014525288 | 0 | -0.000581687 | -0.000190411 | 0.009875663 |
| LTBR | 0 | 0.002024255 | 0 | 0 | -0.002682873 | 0.002771967 | -0.000437067 | -0.000421684 | -0.001075149 | 0 | 0.003322501 | 0 | -0.000423129 | -0.00026862 | -0.00432502 |
| SPON2 | 0 | 0.003906768 | 0 | 0 | 0.001092084 | 0.020967831 | 0.000666831 | 0.00376702 | 0.000518425 | 0 | 0.000573026 | 0 | -0.000611626 | -0.001508662 | 0 |

Table S11: Elastic Net Model Coefficients (generated from SCAPIS)

d) Metabolomics Models

| Metabolomics | Cholesterol_pMMS | Lipodystrophy_1_pMMS | Lipodystrophy_2_pMMS | Liver-Lipid_pMMS | ASAT_Pos_pMMS | ALP_Neg_pMMS | SHBG-LpA_pMMS | Obesity_pMMS | VAT_Neg_pMMS | Bilirubin_pMMS | Proinsulin_pMMS | Hyper_Insulin_pMMS | Beta_Cell_1_pMMS | Beta_Cell_2_pMMS | Total_GRS |
|---|---|---|---|---|---|---|---|---|---|---|---|---|---|---|---|
| HDL_C | 0 | -0.001837658 | -0.004623258 | 0 | 0 | 0 | 0 | 0 | 0 | 0 | 0 | -1.79E-05 | 0 | 0 | 0 |
| HDL2_C | 0 | -0.001913899 | -0.004602986 | 0 | 0 | 0 | 0 | 0 | 0 | 0.000160385 | 0 | -1.42E-05 | 0 | 0 | 0 |
| HDL3_C | -0.066151327 | -0.00073536 | -0.003100863 | -0.077949019 | 0 | -0.13654928 | 0 | 0 | 0 | -0.002533113 | 0 | -3.40E-05 | 0.000145757 | 0 | 0 |
| HDL_D | -0.02477527 | -0.001930342 | -0.00364991 | 0 | 0 | 0.083801132 | 0 | 0 | 0 | 0.001278177 | 0 | 0 | -6.07E-05 | 0 | 0 |
| HDL_TG | 0.117902805 | -0.000581634 | -0.006595177 | -0.088127686 | 0 | 0.146902336 | 0 | 0 | 0 | 0.000455981 | 0 | 0 | 6.39E-05 | 0 | 0 |
| S_HDL_C | 0.104433246 | -9.76E-05 | -0.006017503 | 0.059921044 | 0 | 0.119398386 | 0 | 0 | 0 | -0.001220632 | 0 | 0 | 0.000659367 | 0 | 0 |
| S_HDL_CE | 0 | -0.000115229 | -0.008086919 | 0.027636022 | 0 | 0 | 0 | 0 | 0 | -0.001682747 | 0 | 0 | 0.00062159 | 0 | 0 |
| S_HDL_FC | 0.003387843 | -0.000555156 | 0.006412333 | -0.010353542 | 0 | 0 | 0 | 0 | -0.011652586 | 0.000161292 | 0 | -0.000126625 | 0.000256691 | 0 | 0 |
| S_HDL_L | 0 | -0.000180588 | -0.00289028 | 0 | 0 | 0 | 0 | 0 | 0 | 0.000680226 | 0 | -7.62E-05 | 0.000454372 | 0 | 0 |
| S_HDL_P | 0 | -0.000150834 | -0.002654839 | 0 | 0 | 0 | 0 | 0 | 0 | 0.000914393 | 0 | -7.52E-05 | 0.000428226 | 0 | 0 |
| S_HDL_PL | -0.044257023 | -0.000785537 | 0.001638123 | 0 | 0 | 0 | 0 | 0 | -0.01359023 | 0.001461076 | 0 | -0.000116542 | 3.13E-05 | 0 | 0 |
| S_HDL_TG | -0.059770281 | 0.000983171 | -0.000194192 | 0.42125025 | 0 | 0.006641542 | 0 | 0 | 0 | 0.000313837 | 0 | 0 | 5.47E-05 | 0 | 0 |
| M_HDL_C | 0 | -0.001623157 | -0.002802425 | 0 | 0 | 0 | 0 | 0 | -0.002353797 | 0.001604734 | 0 | -9.82E-05 | 1.39E-05 | 0 | 0 |
| M_HDL_CE | 0 | -0.001666653 | -0.002655721 | -0.05378106 | 0 | -0.082565168 | 0 | 0 | -0.003288312 | 0.001517628 | 0 | -9.93E-05 | 7.02E-06 | 0 | 0 |
| M_HDL_FC | 0 | -0.001409616 | -0.003264746 | 0 | 0 | 0 | 0 | 0 | 0 | 0.001929963 | 0 | -8.53E-05 | 2.75E-05 | 0 | 0 |
| M_HDL_L | 0 | -0.001698994 | -0.004561679 | 0 | 0 | 0 | 0 | 0 | -0.003941613 | 0.002025276 | 0 | -0.00010472 | 4.27E-05 | 0 | 0 |
| M_HDL_P | 0 | -0.001714987 | -0.004575859 | 0 | 0 | 0 | 0 | 0 | -0.004732367 | 0.002051396 | 0 | -0.000106636 | 4.55E-05 | 0 | 0 |
| M_HDL_PL | 0 | -0.001657728 | -0.004311105 | 0 | 0 | 0 | 0 | 0 | 0 | 0.002246686 | 0 | -0.000101758 | 2.85E-05 | 0 | 0 |
| M_HDL_TG | -0.184112558 | 0 | -0.007359517 | 0 | 0 | 0 | 0 | 0 | 0 | 0.000108514 | 0 | 0 | 1.41E-05 | 0 | 0 |
| L_HDL_C | -0.00906019 | -0.001585544 | -0.000945229 | 0 | 0 | 0 | 0 | 0 | 0 | 0.002534726 | 0 | 0 | 0 | 0 | 0 |
| L_HDL_CE | -0.03150439 | -0.00160094 | -0.00088206 | 0 | 0 | 0 | 0 | 0 | 0 | 0.002550314 | 0 | 0 | 0 | 0 | 0 |
| L_HDL_FC | 0 | -0.00154209 | -0.001049311 | 0 | 0 | 0 | 0 | 0 | 0 | 0.002087127 | 0 | 0 | -2.60E-06 | 0 | 0 |
| L_HDL_L | 0 | -0.001593051 | -0.002079821 | 0 | 0 | 0 | 0 | 0 | 0 | 0.002677585 | 0 | 0 | 0 | 0 | 0 |
| L_HDL_P | 0 | -0.001604131 | -0.002300118 | 0 | 0 | 0 | 0 | 0 | 0 | 0.002743776 | 0 | 0 | 0 | 0 | 0 |
| L_HDL_PL | 0 | -0.00157236 | -0.002614844 | 0 | 0 | 0 | 0 | 0 | 0 | 0.00296036 | 0 | 0 | 0 | 0 | 0 |
| L_HDL_TG | -0.048755572 | -0.000794224 | -0.012237119 | 0.09861363 | 0 | 0 | 0 | 0 | 0 | 0.001428934 | 0 | 0 | 0 | 0 | 0 |
| XL_HDL_C | 0 | -0.0019628 | -0.000689741 | 0 | 0 | 0 | 0 | 0 | 0 | -0.004625914 | 0 | 0 | 0 | 0 | 0 |
| XL_HDL_CE | -0.071427708 | -0.001939403 | -0.00092664 | 0 | 0 | 0 | 0 | 0 | 0 | -0.0045023 | 0 | 0 | 0 | 0 | -2.61E-20 |
| XL_HDL_FC | 0 | -0.001973264 | -9.35E-05 | 0 | 0 | 0.015467691 | 0 | 0 | 0 | -0.00480544 | 0 | 0 | -1.44E-05 | 0 | 0 |
| XL_HDL_L | 0 | -0.001990883 | -0.001871006 | 0.003731005 | 0 | 0 | 0 | 0 | 0 | -0.002499939 | 0 | 0 | -1.29E-05 | 0 | 0 |
| XL_HDL_P | 0 | -0.00199888 | -0.002082043 | 0.082191903 | 0 | 0 | 0 | 0 | 0 | -0.002299018 | 0 | 0 | -1.17E-05 | 0 | 0 |
| XL_HDL_PL | 0 | -0.001902931 | -0.003305667 | 0 | 0 | 0 | 0 | 0 | 0 | -0.000249675 | 0 | 0 | -5.74E-05 | 0 | 0 |
| XL_HDL_TG | 0.003048079 | -0.001046254 | -0.003823536 | 0 | 0 | -0.018357811 | 0 | 0 | 0 | -0.005250439 | 0 | 0 | 1.04E-05 | 0 | 0 |
| IDL_C | 0 | 0.00034938 | -0.005018001 | 0 | 0 | 0 | 0 | 0 | 0 | -0.001452219 | 0 | 0 | 0.000125029 | 0 | 0 |
| IDL_CE | 0 | 0.000337936 | -0.005264818 | 0 | 0 | 0 | 0 | 0 | 0 | -0.002170948 | 0 | 0 | 0.000129682 | 0 | 0 |
| IDL_FC | 0 | 0.000475528 | -0.00424124 | 0 | 0 | 0 | 0 | 0 | 0 | -0.00013907 | 0 | 0 | 0.000110267 | 0 | 0 |
| IDL_L | 0 | 0.00054457 | -0.003318815 | 0 | 0 | 0 | 0 | 0 | 0 | -0.000640624 | 0 | 0 | 0.00011129 | 0 | 0 |
| IDL_P | 0 | 0.000647251 | -0.003246486 | 0 | 0 | 0 | 0 | 0 | 0 | -0.000183438 | 0 | 0 | 0.000111584 | 0 | 0 |
| IDL_PL | 0 | 0.000504582 | -0.004384954 | -0.014959603 | 0 | 0 | 0 | 0 | 0 | -0.000329637 | 0 | 0 | 9.78E-05 | 0 | 0 |
| IDL_TG | 0 | 0.002085394 | 0.014186794 | 0 | 0 | 0 | 0 | 0 | 0 | 0.004581838 | 0 | 0 | 0 | 0 | 0 |
| LDL_C | 0 | 0.000747585 | -0.00188817 | 0 | 0 | 0 | 0 | 0 | 0 | -0.000468777 | 0 | 0 | 0.000230617 | 0 | 0 |
| LDL_D | 0.02725333 | -0.000603053 | -0.008293198 | 0.000723127 | 0 | -0.000102729 | 0 | 0 | 0 | -0.003666031 | 0 | 0 | -0.000744203 | 0 | 0 |
| LDL_TG | -0.029304737 | 0.001284523 | 0.009763635 | 0 | 0 | 0 | 0 | 0 | 0 | 0.004007228 | 0 | 0 | 0 | 0 | 0 |
| S_LDL_C | -0.012139803 | 0.000792013 | -0.001197584 | 0 | 0 | 0 | 0 | 0 | 0 | -0.000433822 | 0 | 0 | 0.000295879 | 0 | 0 |
| S_LDL_CE | -0.19852475 | 0.000841241 | -0.001016726 | 0 | 0 | 0 | 0 | 0 | 0 | 2.40E-05 | 0 | 0 | 0.000292491 | 0 | 0 |
| S_LDL_FC | 0 | 0.00063628 | -0.002216354 | 0 | 0 | 0 | 0 | 0 | 0 | -0.003564275 | 0 | 0 | 0.000294264 | 0 | 0 |
| S_LDL_L | -0.061226709 | 0.000703329 | -0.001370257 | 0 | 0 | 0 | 0 | 0 | 0 | -0.000879292 | 0 | 0 | 0.000304323 | 0 | 0 |
| S_LDL_P | -0.038116045 | 0.000712405 | -0.001112631 | 0 | 0 | 0 | 0 | 0 | 0 | -0.000775695 | 0 | 0 | 0.000306062 | 0 | 0 |
| S_LDL_PL | 0 | 7.76E-05 | -0.002804907 | 0 | 0 | 0 | 0 | 0 | 0 | -0.003794783 | 0 | 0 | 0.000308534 | 0 | 0 |
| S_LDL_TG | -0.01344188 | 0.000672561 | 0.005079603 | 0 | 0 | 0 | 0 | 0 | 0 | 0.00425282 | 0 | 0 | 0.000115232 | 0 | 0 |
| M_LDL_C | -0.019541352 | 0.000838165 | -0.000652289 | 0 | 0 | 0 | 0 | 0 | 0 | -0.000469285 | 0 | 0 | 0.000274534 | 0 | 0 |
| M_LDL_CE | -0.080366572 | 0.000926048 | -0.000779457 | 0 | 0 | 0 | 0 | 0 | 0 | 0 | 0 | 0 | 0.000273872 | 0 | 0 |
| M_LDL_FC | 0 | 0.000522232 | -0.000766675 | 0 | 0 | 0 | 0 | 0 | 0 | -0.002667158 | 0 | 0 | 0.000276865 | 0 | 0 |
| M_LDL_L | 0 | 0.000844201 | -0.000604324 | 0 | 0 | 0 | 0 | 0 | 0 | -0.000605475 | 0 | 0 | 0.000267141 | 0 | 0 |
| M_LDL_P | 0 | 0.000854454 | -0.000419804 | 0 | 0 | 0 | 0 | 0 | 0 | -0.000385768 | 0 | 0 | 0.000263095 | 0 | 0 |
| M_LDL_PL | -0.014820243 | 0.000456213 | -0.001942712 | 0 | 0 | 0 | 0 | 0 | 0 | -0.002885643 | 0 | 0 | 0.000249613 | 0 | 0 |
| M_LDL_TG | -0.003124807 | 0.001276975 | 0.008888005 | 0 | 0 | 0 | 0 | 0 | 0 | 0.00479226 | 0 | 0 | 0 | 0 | 0 |
| L_LDL_C | -0.019211372 | 0.000654361 | -0.002829663 | 0 | 0 | 0 | 0 | 0 | 0 | -0.000694426 | 0 | 0 | 0.000180199 | 0 | 0 |
| L_LDL_CE | -0.043552822 | 0.000674078 | -0.002563781 | 0 | 0 | 0 | 0 | 0 | 0 | -0.000775279 | 0 | 0 | 0.000194408 | 0 | 0 |
| L_LDL_FC | 0 | 0.000519294 | -0.002836829 | 0 | 0 | 0 | 0 | 0 | 0 | -0.000734089 | 0 | 0 | 0.000145782 | 0 | 0 |
| L_LDL_L | 0 | 0.000652028 | -0.002225524 | 0 | 0 | 0 | 0 | 0 | 0 | -0.000572098 | 0 | 0 | 0.000168136 | 0 | 0 |
| L_LDL_P | 0 | 0.000701904 | -0.001842394 | 0 | 0 | 0 | 0 | 0 | 0 | -0.000408096 | 0 | 0 | 0.000166249 | 0 | 0 |
| L_LDL_PL | -0.055882068 | 0.000470389 | -0.003426559 | -0.054768216 | 0 | 0 | 0 | 0 | 0 | -0.001300556 | 0 | 0 | 0.000169265 | 0 | 0 |
| L_LDL_TG | -0.037316003 | 0.001568011 | 0.011688615 | 0 | 0 | 0 | 0 | 0 | 0 | 0.003262806 | 0 | 0 | 0 | 0 | 0 |
| VLDL_C | 0 | 0.000438991 | -0.002215156 | 0 | 0 | -0.147992153 | 0 | 0 | 0 | 0.000431889 | 0 | 0 | 0.000188398 | 0 | 0 |
| VLDL_D | -0.023851043 | 3.35E-05 | -0.002954657 | 0 | 0 | 0.204770965 | 0 | 0 | 0 | -0.001988306 | 0 | 0 | 0 | 0 | 0 |
| VLDL_TG | 0 | 0.000530076 | 0.000261627 | 0 | 0 | 0 | 0 | 0 | 0 | -0.000908745 | 0 | 0 | 2.61E-05 | 0 | 0 |
| Serum_TG | 0 | 0.000365379 | 0.002736384 | 0 | 0 | 0 | 0 | 0 | 0 | 0.001493765 | 0 | 0 | 2.50E-05 | 0 | 0 |
| XS_VLDL_C | 0 | 0.000415265 | -0.003209221 | 0.010695314 | 0 | 0 | 0 | 0 | 0 | 0.000304553 | 0 | 0 | 7.98E-05 | 0 | 0 |
| XS_VLDL_CE | 0 | 3.51E-05 | -0.00537086 | 0.02017799 | 0 | 0 | 0 | 0 | 0 | -0.000437721 | 0 | 0 | 5.85E-05 | 0 | 0 |
| XS_VLDL_FC | 0.129283446 | 0.00124845 | 0.00211864 | 0.149906425 | 0 | 0 | 0 | 0 | 0 | 0.002506081 | 0 | 0 | 0.000107216 | 0 | 0 |
| XS_VLDL_L | 0 | 0.001008857 | 0.00157593 | 0 | 0 | 0 | 0 | 0 | 0 | 0.00164201 | 0 | 0 | 9.32E-05 | 0 | 0 |
| XS_VLDL_P | 0 | 0.001063133 | 0.002356095 | 0 | 0 | 0 | 0 | 0 | 0 | 0.001809858 | 0 | 0 | 9.32E-05 | 0 | 0 |
| XS_VLDL_PL | 0 | 0.001159945 | 0.000156296 | 0 | 0 | 0 | 0 | 0 | 0 | 0.001358237 | 0 | 0 | 0.000105209 | 0 | 0 |
| XS_VLDL_TG | 0 | 0.001523348 | 0.008695478 | -0.041936437 | 0 | 0 | 0 | 0 | 0 | 0.003230713 | 0 | 0 | 2.70E-05 | 0 | 0 |

**Table S11: Elastic Net Model Coefficients (generated from SCAPIS)**

**d) Metabolomics Models**

| Metabolomics | Cholesterol_pMMS | Lipodystrophy_1_pMMS | Lipodystrophy_2_pMMS | Liver-Lipid_pMMS | ASAT_Pos_pMMS | ALP_Neg_pMMS | SHBG-LpA_pMMS | Obesity_pMMS | VAT_Neg_pMMS | Bilirubin_pMMS | Proinsulin_pMMS | Hyper_Insulin_pMMS | Beta_Cell_1_pMMS | Beta_Cell_2_pMMS | Total_GRS |
|---|---|---|---|---|---|---|---|---|---|---|---|---|---|---|---|
| S_VLDL_C | 0 | 0.000739865 | -0.003329126 | 0 | 0 | 0 | 0 | 0 | 0 | 0.001186686 | 0 | 0 | 0.000251649 | 0 | 0 |
| S_VLDL_CE | 0 | 0.00064992 | -0.005285472 | 0 | 0 | 0 | 0 | 0 | 0 | 8.49E-05 | 0 | 0 | 0.000246949 | 0 | 0 |
| S_VLDL_FC | 0 | 0.000772471 | 4.79E-05 | -0.049769494 | 0 | 0 | 0 | 0 | 0 | 0.002583676 | 0 | 0 | 0.000210674 | 0 | 0 |
| S_VLDL_L | 0 | 0.000837606 | 2.23E-05 | 0 | 0 | 0 | 0 | 0 | 0 | 0.00120954 | 0 | 0 | 0.000172128 | 0 | 0 |
| S_VLDL_P | 0 | 0.000840267 | 0.00045886 | 0 | 0 | 0 | 0 | 0 | 0 | 0.001061564 | 0 | 0 | 0.000156139 | 0 | 0 |
| S_VLDL_PL | 0 | 0.000710903 | -0.001351719 | 0 | 0 | 0.178668724 | 0 | 0 | 0 | 0.002147432 | 0 | 0 | 0.000163419 | 0 | 0 |
| S_VLDL_TG | 0.122310896 | 0.000901637 | 0.002399568 | -0.156141118 | 0 | 0 | 0 | 0 | 0 | -0.000171289 | 0 | 0 | 5.54E-05 | 0 | 0 |
| M_VLDL_C | 0.040701933 | 0.000440245 | -0.001648584 | -0.088580104 | 0 | 0 | 0 | 0 | 0 | -0.000558743 | 0 | 0 | 0.000120661 | 0 | 0 |
| M_VLDL_CE | 0.315748578 | 0.000313099 | -0.00495201 | 0 | 0 | -0.017513396 | 0 | 0 | 0 | -0.001872687 | 0 | 0 | 0.000149896 | 0 | 0 |
| M_VLDL_FC | 0 | 0.000534703 | 0.001753411 | -0.211778554 | 0 | 0 | 0 | 0 | 0 | -8.18E-05 | 0 | 0 | 6.09E-05 | 0 | 0 |
| M_VLDL_L | 0 | 0.000599096 | 0 | 0 | 0 | 0 | 0 | 0 | 0 | -0.000996384 | 0 | 0 | 7.59E-05 | 0 | 0 |
| M_VLDL_P | 0 | 0.000604718 | -0.000271768 | 0 | 0 | 0 | 0 | 0 | 0 | -0.001117929 | 0 | 0 | 6.64E-05 | 0 | 0 |
| M_VLDL_PL | 0 | 0.000679964 | 0.000852233 | 0 | 0 | 0 | 0 | 0 | 0 | -0.000523065 | 0 | 0 | 0.000100183 | 0 | 0 |
| M_VLDL_TG | 0 | 0.000668669 | -6.56E-05 | 0 | 0 | 0 | 0 | 0 | 0 | -0.001708861 | 0 | 0 | 3.62E-05 | 0 | 0 |
| L_VLDL_C | 0 | 0.00022808 | -0.00217081 | 0 | 0 | 0 | 0 | 0 | 0 | -0.000549279 | 0 | 0 | 1.47E-05 | 0 | 0 |
| L_VLDL_CE | -0.103838376 | 0.000330553 | -0.004121275 | 0 | 0 | 0 | 0 | 0 | 0 | -0.001274576 | 0 | 0 | 8.61E-06 | 0 | 0 |
| L_VLDL_FC | -0.106623516 | 0.000153363 | -0.00026766 | 0 | 0 | 0 | 0 | 0 | 0 | -3.15E-05 | 0 | 0 | 2.63E-05 | 0 | 0 |
| L_VLDL_L | 0 | 0.000249752 | -0.000535757 | 0 | 0 | -0.105869632 | 0 | 0 | 0 | 0.000182519 | 0 | 0 | 0 | 0 | 0 |
| L_VLDL_P | 0 | 0.000248264 | -0.000462753 | 0 | 0 | -0.040649046 | 0 | 0 | 0 | 0.00022586 | 0 | 0 | 0 | 0 | 0 |
| L_VLDL_PL | -0.020111366 | 0.000315451 | 1.66E-05 | 0 | 0 | -0.35478332 | 0 | 0 | 0 | 0.000242142 | 0 | 0 | 1.83E-05 | 0 | 0 |
| L_VLDL_TG | 0 | 0.000230925 | -3.27E-05 | 0 | 0 | 0 | 0 | 0 | 0 | 0.000356377 | 0 | 0 | 0 | 0 | 0 |
| XL_VLDL_C | 0 | -3.42E-05 | -0.00137713 | 0 | 0 | 0 | 0 | 0 | 0 | -0.00120714 | 0 | 0 | 4.52E-05 | 0 | 0 |
| XL_VLDL_CE | -0.085278196 | 0 | -0.001547026 | -0.099524838 | 0 | 0.316662984 | 0 | 0 | 0 | -0.001072124 | 0 | 0 | 1.98E-05 | 0 | 0 |
| XL_VLDL_FC | 0 | -0.000314312 | -0.001398991 | 0 | 0 | 0 | 0 | 0 | 0 | -0.001181366 | 0 | 0 | 6.98E-05 | 0 | 0 |
| XL_VLDL_L | 0 | -0.000278161 | -0.00045045 | 0 | 0 | 0 | 0 | 0 | 0 | -0.001203797 | 0 | 0 | 2.11E-05 | 0 | 0 |
| XL_VLDL_P | 0 | -0.00027873 | -0.000186051 | 0 | 0 | 0 | 0 | 0 | 0 | -0.001116765 | 0 | 0 | 1.46E-05 | 0 | 0 |
| XL_VLDL_PL | 0 | -0.00030127 | -0.000864063 | 0.243511936 | 0 | -0.105484458 | 0 | 0 | 0 | -0.001148671 | 0 | 0 | 8.04E-05 | 0 | 0 |
| XL_VLDL_TG | 0.030441697 | -0.000305235 | 0.000422311 | 0 | 0 | 0 | 0 | 0 | 0 | -0.000778449 | 0 | 0 | 0 | 0 | 0 |
| XXL_VLDL_C | 0 | -0.000179779 | -0.003614725 | 0 | 0 | 0 | 0 | 0 | 0 | 0.000506803 | 0 | 0 | 3.72E-05 | 0 | 0 |
| XXL_VLDL_CE | -0.006891799 | 0 | -0.004796974 | 0 | 0 | 0 | 0 | 0 | 0 | -0.000420464 | 0 | 0 | 8.93E-06 | 0 | 0 |
| XXL_VLDL_FC | 0 | -0.000512195 | -0.000783386 | 0 | 0 | 0 | 0 | 0 | 0 | 0.001699177 | 0 | 0 | 2.54E-05 | 0 | 0 |
| XXL_VLDL_L | 0 | -0.000744762 | -0.006593384 | 0 | 0 | 0 | 0 | 0 | 0 | 0.001374409 | 0 | 0 | 4.41E-05 | 0 | 0 |
| XXL_VLDL_P | 0 | -0.00078196 | -0.007297559 | 0.007456648 | 0 | 0 | 0 | 0 | 0 | 0.00158665 | 0 | 0 | 4.26E-05 | 0 | 0 |
| XXL_VLDL_PL | 0.045952549 | -0.000608685 | -0.002840808 | 0 | 0 | 0.006038884 | 0 | 0 | 0 | 0.003103804 | 0 | 0 | 8.10E-05 | 0 | 0 |
| XXL_VLDL_TG | 0 | -0.000919461 | -0.008052566 | 0.122488395 | 0 | 0.012831363 | 0 | 0 | 0 | 0.001264723 | 0 | 0 | 2.49E-05 | 0 | 0 |
| Ala | 0.016519818 | 0.001832573 | 0.004413496 | -0.130906665 | 0 | 0.003707244 | 0 | 0 | 0 | -0.006974523 | 0 | 0 | -6.54E-05 | 0 | 0 |
| Cit | -0.004831944 | -0.000818083 | -0.008626892 | -0.003515456 | 0 | 0.006870106 | 0 | 0 | 0 | 0.00380812 | 0 | 0 | 0.000150465 | 0 | 0 |
| Gln | -0.036680878 | -0.001375037 | 0.011846195 | 0.043271985 | 0 | 0.002542187 | 0 | 0 | 0 | 0.006129255 | 0 | 0 | 0.000139683 | 0.015536108 | 0 |
| His | -0.00979898 | -0.000639614 | -0.00913373 | 0.020015942 | 0 | 0.004677688 | 0 | 0 | 0 | 0.000241389 | 0 | 0 | 0.000242644 | 0 | 0 |
| Ile | 0.051967733 | 8.60E-05 | 0.002060379 | 0.000728947 | 0 | 0.022282643 | 0 | 0 | 0 | 0.002399648 | 0 | 0 | 0.000446523 | 0 | 0 |
| Leu | -0.080943936 | -3.14E-05 | 0.001046549 | -0.066072948 | 0 | 0.007592983 | 0 | 0 | 0 | -0.000321194 | 0 | 0 | 0.000721539 | 0 | 0 |
| Phe | -0.026934883 | -0.001362926 | 0.00400565 | 0.020092292 | 0 | -0.012806446 | 0 | 0 | 0 | 0.003131796 | 0 | 0 | -0.000193844 | 0 | 0 |
| Tyr | 0.045781581 | 0.000262546 | 0.004927724 | 0.08561493 | 0 | -0.005193198 | 0 | 0 | 0 | -0.006481651 | 0 | 0 | -5.32E-06 | 0 | 0 |
| Val | 0.034301211 | 0.000241351 | 0.006572041 | 0.008700091 | 0 | 0 | 0 | 0 | 0 | 0.000307334 | 0 | 0 | 0.000354413 | 0 | 0 |
| Ace | 0.007307393 | 0.001228371 | -0.012670779 | 0.001232682 | 0 | -0.032151279 | 0 | 0 | 0 | 0.014636498 | 0 | 0 | -6.94E-05 | 0 | 0 |
| AcAce | -0.059362351 | -0.000254323 | -0.013019515 | -0.086417134 | 0 | -0.00402086 | 0 | 0 | -0.004290779 | 0.001348181 | 0 | 0 | 9.43E-05 | 0 | 0 |
| bOHBut | 0.056122411 | -0.002460578 | -0.007129448 | 0.054014963 | 0 | 0.026093033 | 0 | 0 | 0 | 0.002763335 | 0 | 0 | 0.000777363 | 0 | 0 |
| LA | -0.201720678 | -0.000561302 | 0.002926341 | -0.096396126 | 0 | -0.118924703 | 0 | 0 | 0 | -0.000944969 | 0 | 0 | 0.000325793 | 0 | 0 |
| DHA | 0 | 0.001382877 | 0.011943152 | -0.037502211 | 0 | -0.044711595 | 0 | 0 | 0 | -0.006829663 | 0 | 0 | 0.000486756 | 0 | 0 |
| MUFA | -0.2941474 | -0.000411735 | -0.0013965 | -0.063987807 | 0 | -0.043270405 | 0 | 0 | 0 | -0.000589744 | 0 | 0 | 0.000230889 | 0 | 0 |
| PUFA | 0 | -0.000242354 | 0.002022354 | 0 | 0 | 0 | 0 | 0 | 0 | -0.00328161 | 0 | 0 | 0.000476274 | 0 | 0 |
| FAw3 | -0.136491802 | 0.000934604 | 0.012334602 | 0 | 0 | 0 | 0 | 0 | 0 | -0.006970548 | 0 | 0 | 0.000647146 | 0 | 0 |
| FAw6 | 0 | -0.000531104 | -0.000143181 | 0 | 0 | 0 | 0 | 0 | 0 | -0.002081978 | 0 | 0 | 0.000428286 | 0 | 0 |
| SFA | 0.562668471 | 0 | 0.000814411 | 0.232568105 | 0 | 0.119421696 | 0 | 0 | 0 | -0.001755533 | 0 | -2.13E-05 | 0.000171515 | 0 | 0 |
| UnSat | 0.148991144 | 0.000665367 | 0.010044287 | 0.130493206 | 0 | 0.106510735 | 0 | 0 | 0 | -0.006045884 | 0 | 0 | 0.000336518 | 0 | 0 |
| TotFA | 0 | -0.000124363 | 0.000352177 | 0 | 0 | 0 | 0 | 0 | 0 | -0.001627606 | 0 | 0 | 0.000319947 | 0 | 0 |
| EstC | 0 | 0 | -0.004206972 | -0.1141144 | 0 | 0 | 0 | 0 | 0 | -0.001829721 | 0 | 0 | 0.000243127 | 0 | 0 |
| FreeC | 0 | 4.16E-06 | -0.004727068 | 0 | 0 | 0 | 0 | 0 | 0 | -0.001729026 | 0 | 0 | 0.000203561 | 0 | 0 |
| PC | 0.172843479 | -0.000597144 | 0.003982823 | -0.158862799 | 0 | 0 | 0 | 0 | 0 | 0.000278361 | 0 | -6.69E-05 | 0.000206289 | 0 | 0 |
| Remnant_C | 0 | 0.000494573 | -0.00317513 | 0 | 0 | -0.040698871 | 0 | 0 | 0 | -0.001508458 | 0 | 0 | 0.000196614 | 0 | 0 |
| Serum_C | 0 | 0 | -0.004335544 | 0 | 0 | 0 | 0 | 0 | 0 | -0.001802575 | 0 | 0 | 0.000234569 | 0 | 0 |
| SM | 0.042003762 | -0.001382426 | -0.004549991 | 0.055386078 | 0 | 0.13627835 | 0 | 0 | 0 | 0 | 0 | 0 | 0.000264548 | 0 | 0 |
| TotCho | 0 | -0.000678947 | 0.002534972 | 0.086182706 | 0 | 0.095623643 | 0 | 0 | 0 | 0.001932881 | 0 | -4.84E-05 | 0.000175614 | 0 | 0 |
| TotPG | 0 | -0.000527447 | 0.000242256 | 0 | 0 | 0 | 0 | 0 | 0 | -0.000140576 | 0 | -6.82E-05 | 0.000233478 | 0 | 0 |
| Alb | 0.050741019 | 0.000760949 | 0.012545794 | -0.036023001 | 0 | -0.045891725 | 0 | 0 | 0 | -0.001097538 | 0 | 0 | 0.000457212 | 0 | 0 |
| Crea | 0.023327983 | 0 | -0.008290361 | 0.057554653 | 0 | -0.001053036 | 0 | 0 | 0 | 0.009329483 | 0 | 0 | 0 | 0 | 0 |
| Lac | 0 | 0.001730569 | 0.000895426 | 0.011023013 | 0 | 0.038304672 | 0 | 0 | 0 | -0.00833503 | 0 | 0 | -8.81E-05 | 0 | 0 |
| Gp | -0.032056143 | -0.000433833 | 0.011230824 | -0.048259407 | 0 | 0.039896295 | 0 | 0 | 0 | 0.00383088 | 0 | 0 | 0.000560245 | 0 | 0 |

**Table S11: Elastic Net Model Coefficients (generated from SCAPIS)**

**e) Merged_Omics Models**

| Omics_Variables | Cholesterol_pMMS | Lipodystrophy_1_pMMS | Lipodystrophy_2_pMMS | Liver-Lipid_pMMS | ASAT_Pos_pMMS | ALP_Neg_pMMS | SHBG-LpA_pMMS | Obesity_pMMS | VAT_Neg_pMMS | Bilirubin_pMMS | Proinsulin_pMMS | Hyper_Insulin_pMMS | Beta_Cell_1_pMMS | Beta_Cell_2_pMMS | Total_GRS |
|---|---|---|---|---|---|---|---|---|---|---|---|---|---|---|---|
| Hba1c | 0.02179822 | 0 | 0.013071397 | 0 | 0.017871169 | 0.067263369 | 0.005707732 | 0.004680535 | 0 | 0 | 0.014386736 | 0 | 0 | 0.025453497 | 0.079197172 |
| Glucose | 0 | 0.000642971 | 0.014475814 | 0 | 0 | 0 | 0 | 0 | 0 | 0 | 0.006728712 | 0 | 0 | 0.034313618 | 0.050470495 |
| PGluc_Cap | 0 | 0.000164317 | 0.002479244 | 0.049128071 | 0 | 0 | 0 | 0 | 0 | 0 | 0.001695298 | 0 | 0 | 0.028302132 | 0 |
| Insulin | 0.054135047 | 0.000688497 | 0.002504271 | 0 | 0.01173871 | 0 | 0 | 0.001900186 | 0 | 0 | -0.011325042 | 0 | 0 | -0.02463227 | 0 |
| Ldl | 0 | 0.000355928 | -0.000283171 | 0 | 0 | 0 | 0 | 0 | 0 | 0 | -0.001550303 | 0 | 0 | 0 | 0 |
| ApoB | -0.197847728 | 0.000698086 | -0.000489343 | 0 | 0 | 0 | 0 | 0 | 0 | 0 | -0.001457095 | 0 | 0 | 0 | 0 |
| Cholesterol | 0 | 2.00E-05 | -0.003031453 | 0 | 0 | 0 | 0 | 0 | 0 | 0 | -0.004205378 | 0 | 0 | 0 | 0 |
| Lpa | 0.034195757 | 0 | 0.014608299 | 0 | 0 | 0.004378343 | 0.00993295 | 0 | 0 | 0 | -0.00569871 | 0 | 0 | 0 | 0 |
| Tg | 0 | 0.000991878 | 0.000141419 | 0 | 0 | 0 | 0 | 0 | 0 | 0 | -0.004469595 | 0 | 0 | 0 | 0 |
| ApoA1 | 0.081793501 | -0.000973174 | -0.008546037 | 0 | -0.010276594 | 0 | 0 | 0 | 0 | 0 | 0.00291909 | 0 | 0 | 0 | 0 |
| Hdl | 0 | -0.000982865 | -0.002957212 | 0 | -0.006509999 | 0 | 0 | 0 | 0 | 0 | -0.007213231 | 0 | 0 | 0 | 0 |
| ALAT | 0 | -6.31E-05 | 0.017280859 | 0 | 0 | 0.011371614 | 0 | 0 | 0 | 0 | 0.007563363 | 0 | 0 | 0 | 0 |
| ASAT | 0 | -5.17E-05 | 0.003325512 | 0 | 0 | 0 | 0 | 0 | 0 | 0 | -0.010316411 | 0 | 0 | -0.000552499 | 0 |
| GGT2 | 0 | 2.09E-05 | 0.00218316 | -0.026934052 | 0 | 0.006002451 | 0 | 0 | 0 | 0 | -0.003319756 | 0 | 0 | 0 | 0 |
| Crp | 0.118680381 | -0.000247322 | -0.003995871 | -0.006409667 | 0 | 0 | 0 | 0.025260227 | 0 | 0 | 0.005521989 | 0 | 0 | -0.00251915 | 0 |
| Alfa1Antitrypsin | 0 | -0.001083535 | -0.007388412 | 0 | 0 | 0 | 0 | 0 | 0 | 0 | -0.000442926 | 0 | 0 | -0.010625665 | 0 |
| TroponinI | 0 | -0.000670125 | -0.000365727 | 0.027509521 | 0 | 0.003849916 | 0 | 0 | 0 | 0 | 0.000849599 | 0 | 0 | 0 | 0 |
| NTproBNP | 0 | 1.35E-05 | 0.001166591 | 0 | 0 | 0.0320083 | 0 | 0 | 0 | 0 | 0.004910836 | 0 | 0 | 0 | 0 |
| HB | 0 | 5.69E-05 | -0.005167975 | 0 | 0 | -0.022820604 | 0 | 0 | 0 | 0 | 0.00378538 | 0 | 0 | 0 | 0 |
| Creatinine | 0.021803786 | -5.60E-05 | -0.003538811 | 0 | 0 | -0.014589603 | 0 | 0 | 0 | 0 | -0.007895706 | 0 | 0 | 0 | 0 |
| CystatinC | 0 | -0.000110266 | -0.002787057 | 0 | 0 | 0.029723308 | 0 | 0 | 0 | 0 | 0.004594647 | 0 | 0 | -0.009583432 | 0 |
| FABP4 | 0 | -0.000138289 | -0.003212116 | 0 | 0 | 0 | 0 | 0 | 0 | 0 | 0.006788818 | 0 | 0 | 0 | 0 |
| SELP | 0 | 0.001114655 | 0.005134312 | 0 | 0 | -0.092504184 | 0 | 0 | 0 | 0 | 0.005838771 | 0 | 0 | 0 | 0 |
| GT | 0 | 0.000325041 | 0.001223793 | 0.004824863 | 0 | 0.052536814 | 0 | 0 | 0 | 0 | 0.00857595 | 0 | 0 | 0 | 0 |
| PCSK9 | -0.040017201 | 0.001440545 | 1.41E-05 | 0 | 0 | 0.00511546 | 0 | 0 | 0 | 0 | 0.003077545 | 0 | 0 | 0 | 0 |
| CTSD | 0 | 0.001505831 | 0.007227996 | 0 | 0 | 0 | 0 | 0 | 0 | 0 | 0.002965144 | 0 | 0 | 0 | 0 |
| CTSZ | 0.080198713 | 0.000560786 | 0.005374924 | 0 | 0 | 0.03290333 | 0 | 0 | 0 | 0 | -0.007927954 | 0 | 0 | 0 | 0 |
| CA5A | 0.047566261 | 0.000577878 | 0.018699962 | 0 | 0 | 0.048709244 | 0 | 0 | 0 | 0 | -0.008279203 | 0 | 0 | 0 | 0.004242775 |
| ACE2 | 0 | -0.000471799 | 0.008273852 | 0 | 0 | -0.020477705 | 0 | 0 | 0 | 0 | -0.012908397 | 0 | 0 | 0 | 0 |
| IDUA | 0 | 0.000866862 | 0.008472977 | 0.013034917 | 0 | 0.001418794 | 0 | 0 | 0 | 0 | -0.004127022 | 0 | 0 | 0 | 0 |
| HAOX1 | 0 | 0.000654017 | 0.010526211 | -0.004813198 | 0 | 0.014485133 | 0 | 0 | 0 | 0 | -0.000358573 | 0 | 0 | 0 | 0.00751433 |
| CPA1 | -0.001864623 | 0.001227071 | -0.00282519 | 0 | 0 | -0.030173598 | 0 | 0 | 0 | 0 | -0.000817025 | 0 | 0 | 0 | 0 |
| PON3 | 0 | 0 | 0.002338015 | 0 | 0 | 0 | 0 | 0 | 0 | 0 | 0.000685724 | 0 | 0 | 0 | 0 |
| AP_N | 0 | 0.000411094 | 0.01080174 | 0 | 0 | 0 | 0 | 0 | 0 | 0 | 0.015384783 | 0 | 0 | 0 | 0.020818108 |
| GIF | 0.009311099 | 0.000337961 | -0.006671929 | 0.000164843 | 0 | -0.078724179 | 0 | 0 | 0 | 0 | -0.007183766 | 0 | 0 | 0 | 0 |
| ICAM_2 | 0 | 0.000666016 | -0.001377796 | 0.017852163 | 0 | -0.198483181 | 0 | 0 | 0 | 0 | -0.017415519 | 0 | 0 | 0 | 0 |
| PD_L2 | 0 | 0.000610941 | 0.003613852 | 0 | 0 | 0 | 0 | 0 | 0 | 0 | -0.012279426 | 0 | 0 | 0 | 0 |
| Gal_9 | 0 | 0.000496878 | -0.009018489 | -0.03727268 | 0 | -0.043812262 | 0 | 0 | 0 | 0 | -0.01008059 | 0 | 0 | 0 | 0 |
| SELE | 0 | 0.001395134 | -0.007394101 | 0 | 0.003048336 | -0.311055479 | 0 | 0 | 0 | 0 | -0.019410723 | 0 | 0 | 0 | 0 |
| TF | -0.044349934 | 0.000271431 | 0.013225941 | 0.007733926 | 0 | 0 | 0 | 0 | 0 | 0 | 0.004092338 | 0 | 0 | 0 | 0 |
| ADM | 0 | -0.000211883 | 0.001063605 | 0.03025806 | 0 | 0 | 0 | 0.015380374 | 0 | 0 | -0.00012698 | 0 | 0 | 0 | 0 |
| IL_1ra | 0 | 0.000253313 | 0.0011034 | 0 | 0 | 0.006667732 | 0 | 0 | 0 | 0 | -0.009471638 | 0 | 0 | 0 | 0 |
| IL6 | -0.007519843 | -0.000730864 | -0.001224465 | 0 | 0 | -0.002720043 | 0 | 0 | 0 | 0 | -0.004158533 | 0 | 0 | 0 | 0 |
| LEP | 0 | -0.00023714 | 0.011662972 | 0.048118267 | 0.010010398 | 0 | 0 | 0 | 0 | 0 | -0.006312128 | 0 | 0 | 0 | 0 |
| IL_1RT2 | 0 | 0.000145429 | 0.013517749 | 0 | 0 | 0 | 0 | 0 | 0 | 0 | 0.006179815 | 0 | 0 | 0 | 0 |
| PARP_1 | 0 | 0.001417045 | 0.015790511 | 0 | 0 | 0 | 0 | 0 | 0 | 0 | 0.019257 | 0 | 0 | 0 | 0.032141578 |
| U_PAR | -0.003482468 | 0.000210815 | 0.001553351 | 0 | 0 | 0.101592435 | 0 | 0 | 0 | 0 | 0.016436134 | 0 | 0 | 0 | 0 |
| LDL_receptor | 0.040528328 | 0.000692683 | -0.004843111 | 0 | 0 | -0.024204193 | 0 | 0 | 0 | 0 | -0.003728842 | 0 | 0 | 0 | 0 |
| FGF_21 | 0 | 0.001489999 | -0.001813054 | -0.040906937 | 0.017094732 | 0.001757443 | 0 | 0 | 0 | 0 | -0.005826777 | 0 | 0 | 0 | 0 |
| IGFBP_2 | 0 | -0.001301092 | -0.009612068 | 0.052570073 | -0.005322195 | 0 | 0 | 0 | 0 | 0 | -0.001619114 | 0 | 0 | 0 | -0.002572799 |
| CPB1 | 0 | 0.001420487 | 0.000127986 | 0 | 0 | -0.058636796 | 0 | 0 | 0 | 0 | -0.008227056 | 0 | 0 | 0 | 0 |
| FGF_23 | 0 | -0.000528834 | 0.008023288 | -0.037413986 | 0 | 0 | 0 | 0.003916109 | 0 | 0 | 0.009409222 | 0 | 0 | 0.002152005 | 0 |
| vWF | 0 | 0.000258967 | -0.004427103 | -0.003291673 | 0 | 0.127983385 | 0 | 0 | 0 | 0 | 0.015206426 | 0 | 0 | 0 | 0 |
| TIE2 | 0.014491483 | -0.000474196 | 0.004564605 | 0 | 0 | 0.125914132 | 0 | 0 | 0 | 0 | 0.013766106 | 0 | 0 | 0 | 0 |
| CDH5 | 0 | -0.00038392 | 0.000879007 | 0 | 0 | 0.153329212 | 0 | 0 | 0 | 0 | 0.013602899 | 0 | 0 | 0 | 0.007721691 |
| TM | 0 | -0.000470846 | -0.000521434 | 0.025787016 | 0 | 0.046102987 | 0 | 0 | 0 | 0 | 0.006920272 | 0 | 0 | 0 | 0 |
| PECAM_1 | -0.010133874 | 0.000858426 | 0.000326696 | 0 | 0 | -0.024020565 | 0 | 0 | 0 | 0 | -0.005305729 | 0 | 0 | 0 | 0 |
| EPHB4 | 0 | 3.68E-05 | -0.011264696 | 0 | 0 | -0.10322964 | 0 | 0 | 0 | 0 | -0.004204254 | 0 | 0 | 0 | 0 |
| IGFBP_1 | -0.012144313 | -0.000885698 | -0.003206678 | -0.10086446 | 0 | -0.006473853 | 0 | 0 | 0 | 0 | -0.00269466 | 0 | 0 | 0.014286095 | 0 |
| FS | 0 | -0.000468631 | -0.004761714 | -0.07457598 | 0 | -0.027977634 | 0 | 0 | 0 | 0 | -0.0053298 | 0 | 0 | 0.001316223 | 0 |
| TFPI | 0 | -0.000101402 | -0.004575264 | 0 | 0 | 0.012825796 | 0 | 0 | 0 | 0 | 0.00495568 | 0 | 0 | 0 | 0 |
| t_PA | 0 | 0.001702253 | -0.003670835 | 0.008791136 | 0 | 0.013192681 | 0 | 0.007862803 | 0 | 0 | 0.010067457 | 0 | 0 | 0 | 0.012544423 |
| CTRC | 0 | 8.19E-06 | -0.00117681 | -0.036478969 | 0 | 0.132016428 | 0 | 0 | 0 | 0 | 0.015090776 | 0 | 0 | 0 | 0 |
| MERTK | 0 | 7.43E-05 | -0.003872608 | 0 | -0.001239757 | -0.035878427 | 0 | 0 | 0 | 0 | 0.006899515 | 0 | 0 | 0 | 0 |
| PRSS8 | 0 | 0.000962953 | 0.011984065 | 0 | 0 | 0.035331325 | 0 | 0 | 0 | 0 | 0.007742666 | 0 | 0 | 0 | 0 |
| MMP7 | -0.016411047 | 0 | 0.005315144 | -0.00482869 | 0 | 0 | 0 | 0 | 0 | 0 | -0.00983691 | 0 | 0 | 0 | 0 |
| Gal_4 | 0 | 0.001085171 | -0.001227121 | 0 | 0 | -0.185416685 | 0 | 0 | 0 | 0 | -0.01341858 | 0 | 0 | 0 | 0 |
| SCGB3A2 | -0.006860492 | -0.000628155 | -0.009464849 | 0 | -0.002769364 | 0 | 0 | 0 | 0 | 0 | 0.010056556 | 0 | 0 | 0 | 0 |
| THPO | 0 | -0.00015591 | -0.000745519 | 0 | 0 | -0.012464022 | 0 | 0 | 0 | 0 | -0.009258451 | 0 | 0 | 0.000830775 | 0 |
| PLC | 0 | 0 | -0.002161314 | 0 | 0 | -0.025355145 | 0 | 0 | 0 | 0 | 0.001024519 | 0 | 0 | 0 | 0 |
| STK4 | 0 | 0.000192143 | -0.00355437 | 0 | 0 | 0 | 0.018332216 | 0 | 0 | 0 | 0.000949583 | 0 | 0 | 0 | 0 |
| DCN | 0 | -9.06E-05 | -0.006000515 | 0.030284735 | 0 | -0.004007661 | 0 | 0 | 0 | 0 | 0.003290971 | 0 | 0 | -0.000987219 | 0 |
| CD163 | 0 | 0.000258337 | -0.001829515 | 0 | 0 | 0.028675471 | 0 | 0 | 0 | 0 | 3.82E-05 | 0 | 0 | 0 | 0 |
| IL_6RA | 0 | -0.00028656 | -0.003742015 | 0 | 0 | -0.000724452 | 0 | 0 | 0 | 0 | -0.007547268 | 0 | 0 | 0 | 0 |
| PGF | 0 | 0.000215846 | -0.001304912 | 0.026775646 | 0 | -0.003355872 | 0 | 0 | 0 | 0 | -0.004229283 | 0 | 0 | 0 | 0 |
| CD4 | 0 | -0.000321852 | 0.009878131 | 0 | 0 | -0.006360915 | 0.022982551 | 0 | 0 | 0 | 0 | 0 | 0 | 0 | 0 |

**Table S11: Elastic Net Model Coefficients (generated from SCAPIS)**

**e) Merged_Omics Models**

| Omics_Variables | Cholesterol_pMMS | Lipodystrophy_1_pMMS | Lipodystrophy_2_pMMS | Liver-Lipid_pMMS | ASAT_Pos_pMMS | ALP_Neg_pMMS | SHBG-LpA_pMMS | Obesity_pMMS | VAT_Neg_pMMS | Bilirubin_pMMS | Proinsulin_pMMS | Hyper_Insulin_pMMS | Beta_Cell_1_pMMS | Beta_Cell_2_pMMS | Total_GRS |
|---|---|---|---|---|---|---|---|---|---|---|---|---|---|---|---|
| CD40_L | 0 | 0.000101241 | 0.00106656 | 0 | 0 | 0 | 0 | 0 | 0 | 0 | -0.007107324 | 0 | 0 | 0 | 0 |
| REN | 0 | -0.000109453 | -0.002300081 | 0 | 0 | 0 | 0 | 0 | 0 | 0 | -0.00244567 | 0 | 0 | 0 | 0 |
| CCL17 | -0.024685366 | 0.000318984 | 0.000849055 | -0.009340377 | 0 | 0 | 0 | 0 | 0 | 0 | -0.004742392 | 0 | 0 | 0 | 0 |
| IL18 | 0 | 0.00016143 | 0.009469592 | 0 | 0 | 0 | 0 | 0 | 0 | 0 | -0.000193411 | 0 | 0 | 0 | 0 |
| CTSL1 | 0 | -0.000117005 | 0.006068884 | 0 | 0 | -0.008638813 | 0 | 0 | 0 | 0 | -0.000226493 | 0 | 0 | 0 | 0 |
| SOD2 | 0 | -0.000231639 | -0.001607113 | 0 | 0 | 0 | 0 | 0 | 0 | 0 | 0.007388315 | 0 | 0 | 0 | 0 |
| IL1RL2 | 0.029773139 | -0.000116084 | -0.003733489 | 0 | 0 | -0.00361128 | 0 | 0 | 0 | 0 | 0.000195063 | 0 | 0 | 0 | 0 |
| ANG_1 | 0 | 0 | -0.003741861 | 0 | 0 | -0.012573669 | 0 | 0 | 0 | 0 | -0.002829284 | 0 | 0 | 0 | 0 |
| IL_27 | -0.011908475 | 0.000150942 | 0.004926477 | 0 | 0 | 0.012412083 | 0 | 0 | 0 | 0 | 0.002417904 | 0 | 0 | 0 | 0 |
| BOC | 0 | 0.00020937 | 0.000131977 | 0 | 0 | 0 | 0 | 0 | 0 | 0 | -0.000811019 | 0 | 0 | 0 | 0 |
| CCL3 | 0 | -0.000214214 | 0.001717575 | 0 | 0 | 0 | 0 | 0 | 0 | 0 | 0.005108397 | 0 | 0 | 0 | 0 |
| Ep_CAM | 0 | 0.000376026 | -0.007804621 | 0 | 0 | 0 | 0 | 0 | 0 | 0 | -0.006429909 | 0 | 0 | 0 | 0 |
| NT_proBNP | 0 | 9.42E-05 | 0.007561327 | 0 | 0 | 0 | 0 | 0 | 0 | 0 | 1.75E-05 | 0 | 0 | 0 | 0 |
| GP6 | 0 | 8.64E-05 | 0.003764719 | 0 | 0.01434014 | 0.015848444 | 0 | 0 | 0 | 0 | -0.009771451 | 0 | 0 | 0 | 0 |
| OPN | -0.045711648 | -0.000227816 | 0.003472169 | 0.007440706 | 0 | 0 | 0 | 0 | 0 | 0 | 0.001685816 | 0 | 0 | 0 | 0 |
| AXL | 0 | 2.11E-05 | 6.29E-05 | 0.000154336 | 0 | 0.003507494 | 0 | 0 | 0 | 0 | 0.005736551 | 0 | 0 | 0 | 0 |
| CHIT1 | -0.006399045 | -0.000723856 | -0.006365237 | 0 | 0 | -0.033926347 | 0 | 0 | 0 | 0 | 0.003792918 | 0 | 0 | 0 | 0 |
| BMP_6 | -0.011188639 | -8.02E-05 | -0.001251674 | 0 | 0 | 0.024457215 | 0 | 0 | 0 | 0 | 0.003088282 | 0 | 0 | 0 | 0 |
| XCL1 | 0 | 0.00045919 | -0.011042579 | 0.009999662 | 0 | 0 | 0 | 0 | 0 | 0 | 0.005179809 | 0 | 0 | 0 | 0 |
| HSP_27 | 0 | -0.000532631 | 0.008055684 | 0 | 0 | 0 | 0 | 0 | 0 | 0 | -0.000454039 | 0 | 0 | 0 | 0 |
| PIgR | 0.003199996 | -0.000323765 | 3.95E-05 | 0 | 0 | -0.040127094 | 0 | 0 | 0 | 0 | 0.001556147 | 0 | 0 | 0 | 0 |
| TNF_R2 | 0 | 0 | 0 | 0 | 0 | 0 | 0 | 0 | 0 | 0 | 0.007313654 | 0 | 0 | -0.013396676 | 0 |
| MPO | 0 | 0 | 0.0028966 | 0 | 0 | 0 | 0 | 0 | 0 | 0 | -0.002900038 | 0 | 0 | 0 | 0 |
| PSP_D | 0.000173698 | -0.000149215 | 0.00526757 | 0 | 0 | 0.010686562 | 0 | 0 | 0 | 0 | 0.007561952 | 0 | 0 | 0.001412557 | 0 |
| CD84 | 0.026519816 | 0.000863053 | 0.009303372 | 0.01755662 | 0 | 0.000858411 | 0.00043093 | 0 | 0 | 0 | -0.0024954 | 0 | 0 | 0 | 0 |
| SPON2 | 0 | 0.000304485 | -0.001717077 | 0 | 0 | 0 | 0 | 0 | 0 | 0 | 0.008251348 | 0 | 0 | 0 | 0 |
| IGFBP_7 | 0 | 2.82E-05 | 0.000377558 | 0 | 0 | 0.026118326 | 0 | 0 | 0 | 0 | 0.000835394 | 0 | 0 | 0 | 0 |
| SPON1 | 0 | -0.000643145 | -0.00592444 | 0 | 0 | 0.002006982 | 0 | 0 | 0 | 0 | 0.010549331 | 0 | 0 | 0.008156083 | 0 |
| DLK_1 | 0 | 0.000273164 | 0.005727594 | 0.028508645 | 0 | -0.008247367 | 0 | 0 | 0 | 0 | 0.001507109 | 0 | 0 | 0 | 0 |
| MMP12 | 0 | 3.15E-05 | 0.000661061 | 0 | 0 | 0.030101455 | 0 | 0 | 0 | 0 | -0.00847271 | 0 | 0 | 0 | 0 |
| PRSS27 | 0 | -0.000290073 | -0.006620766 | 0 | 0 | 0 | 0 | 0 | 0 | 0 | 0.007613636 | 0 | 0 | 0 | 0 |
| FAS | 0.008324635 | 0.000762766 | 0.005995994 | 0 | 0 | 0.002983094 | 0 | 0 | 0 | 0 | 0.002760695 | 0 | 0 | 0 | 0 |
| RAGE | 0 | -0.000253439 | -9.24E-05 | -0.000479521 | 0 | -0.026133679 | 0 | 0 | 0 | 0 | 0.00480678 | 0 | 0 | 0 | 0 |
| MARCO | 0 | 0 | -0.002678317 | 0.003766936 | 0 | -0.038646187 | 0 | 0 | 0 | 0 | -0.008510725 | 0 | 0 | -0.015748967 | 0 |
| ITGB2 | 0 | 0.000425469 | -0.001325506 | 0 | 0 | 0 | 0 | 0 | 0 | 0 | -0.01690496 | 0 | 0 | 0 | 0 |
| PDGF_subunit_B | 0 | 0.000214093 | -0.004565752 | 0 | 0 | 0 | 0 | 0 | 0 | 0 | 0.006466115 | 0 | 0 | 0 | 0 |
| SHPS_1 | 0.041446141 | 0.000767762 | 0.004135264 | 0 | 0 | 0 | 0 | 0 | 0 | 0 | -0.008384791 | 0 | 0 | 0 | 0 |
| TNFSF13B | 0 | 0.000769097 | 0.007978873 | 0 | 0 | 0.015336785 | 0 | 0 | 0 | 0 | -0.01557294 | 0 | 0 | 0 | 0 |
| GLO1 | 0 | 0.00113714 | 0 | 0 | 0 | 0 | 0 | 0 | 0 | 0 | -0.00537663 | 0 | 0 | 0 | 0 |
| CSTB | 0 | -0.000318722 | -0.009066828 | 0 | 0 | 0 | 0 | 0 | 0 | 0 | -0.012465937 | 0 | 0 | 0 | 0 |
| Gal_3 | 0 | 0.0003731 | -0.000406594 | 0 | 0 | 0 | 0 | 0 | 0 | 0 | -0.003648147 | 0 | 0 | 0 | 0 |
| CD93 | 0 | 0 | 0.001876985 | 0 | 0 | -0.011397652 | 0 | 0 | 0 | 0 | 0.009049681 | 0 | 0 | 0 | 0 |
| GDF_15 | 0 | 0 | 0.008331235 | 0 | 0 | 0 | 0 | 0 | 0 | 0 | 0.004569293 | 0 | 0 | -0.003655359 | 0 |
| GH | -0.001382391 | 0 | 0.005808555 | 0 | 0 | -0.016335467 | 0 | 0 | 0 | 0 | -0.012311781 | 0 | 0 | 0 | 0 |
| AGRP | 0 | 4.95E-05 | -0.007925878 | 0 | 0 | 0 | 0 | 0 | 0 | 0 | 0 | 0 | 0 | 0 | 0 |
| TNFRSF10A | -0.037784134 | 0.000666512 | -0.004239769 | 0 | 0 | 0 | 0 | 0 | 0 | 0 | -0.01099208 | 0 | 0 | 0 | 0 |
| TNFRSF10C | 0 | -0.00039751 | 0.005687957 | 0 | 0 | 0.011525501 | 0 | 0 | 0 | 0 | 0.000974612 | 0 | 0 | 0.027672472 | 0 |
| THBS2 | 0 | 0.000543832 | -0.00676054 | 0 | 0 | -0.017061655 | 0 | 0 | 0 | 0 | -0.004956647 | 0 | 0 | 0 | 0 |
| CXCL16 | 0 | 0.000317217 | -0.002929116 | 0 | 0 | 0 | 0 | 0 | 0 | 0 | 0.001585783 | 0 | 0 | 0 | 0 |
| CEACAM8 | 0 | 6.41E-05 | 0.001301079 | 0 | 0 | 0 | 0 | 0 | 0 | 0 | 0.00138444 | 0 | 0 | 0 | 0 |
| IL_18BP | 0 | 0.000411826 | 0.001604239 | 0 | 0 | 0 | 0 | 0 | 0 | 0 | -0.001083019 | 0 | 0 | 0 | 0 |
| COL1A1 | 0 | -0.00011505 | -0.002394947 | 0 | 0 | -0.018735712 | 0 | 0 | 0 | 0 | 0.007233035 | 0 | 0 | -0.000946274 | 0 |
| PSGL_1 | 0.007075229 | -0.000161156 | 0.00241621 | 0 | 0 | 0 | 0 | 0 | 0 | 0 | 0.001816712 | 0 | 0 | 0 | 0 |
| TR | -0.010075805 | 0.000679935 | 0 | 0 | 0 | -0.027204368 | 0 | 0 | 0 | 0 | 0.010938636 | 0 | 0 | 0 | 0 |
| AZU1 | 0 | 0.000729543 | 0.009066905 | -0.02866811 | 0 | -0.00308986 | 0 | 0 | 0 | 0 | -0.001763229 | 0 | 0 | 0 | 0 |
| CXCL1 | -0.008156846 | 0.00037389 | 0.001995803 | 0 | 0 | 0.000651331 | 0 | 0 | 0 | 0 | -0.00506192 | 0 | 0 | 0 | 0 |
| PI3 | 0 | 0.000680553 | 0.001697267 | 0 | 0 | 0 | 0 | 0 | 0 | 0 | 0.001658886 | 0 | 0 | 0 | 0 |
| TR_AP | 0 | 0.00087598 | 0.000699277 | 0.019270037 | 0 | 0.005054378 | 0 | 0 | 0 | 0 | 0.00661857 | 0 | 0 | 0.00202016 | 0 |
| PRTN3 | -0.01197524 | 0.00032752 | -0.00905592 | 0 | 0 | -0.00875797 | 0 | 0 | 0 | 0 | -0.004105769 | 0 | 0 | 0 | 0 |
| PGLYRP1 | 0 | 0 | -0.00011842 | 0 | 0 | 0 | 0 | 0 | 0 | 0 | -0.005939304 | 0 | 0 | 0 | 0 |
| PTX3 | 0 | -0.000317667 | -0.003967892 | 0 | 0 | -0.008775712 | 0 | 0 | 0 | 0 | 0.005694249 | 0 | 0 | 0 | 0 |
| DECR1 | 0 | 9.01E-05 | -0.002040162 | 0 | 0 | 0 | 0 | 0 | 0 | 0 | -0.006694341 | 0 | 0 | 0 | 0 |
| CHI3L1 | 0 | 0.001186056 | 0.00201717 | 0 | 0 | 0.002475477 | 0 | 0 | 0 | 0 | 0.003832938 | 0 | 0 | 0 | 0 |
| LOX_1 | -0.015655406 | 0 | -0.007369762 | -0.001304437 | 0 | -0.019625412 | 0 | 0 | 0 | 0 | 0.004095051 | 0 | 0 | 0 | 0 |
| CCL15 | 0 | 0.000244806 | -0.000544736 | 0 | 0 | 0 | 0 | 0 | 0 | 0 | 0.001578122 | 0 | 0 | 0 | 0 |
| TGM2 | -0.007542188 | 0.001147366 | -0.006744872 | 0 | 0 | -0.026907341 | 0 | 0 | 0 | 0 | 0.005918877 | 0 | 0 | 0 | 0 |
| MB | 0 | 0.000260867 | -0.01154151 | 0 | 0 | 0 | 0 | 0.012104818 | 0 | 0 | 0.004951287 | 0 | 0 | 0 | 0 |
| CNTN1 | 0 | 0.000855892 | 0.010046086 | 0 | 0 | 0 | 0 | 0 | 0 | 0 | -0.001858995 | 0 | 0 | 0 | 0 |
| SLAMF7 | 0 | 0.000317282 | -0.002085814 | -0.015729881 | 0 | 0 | 0 | 0 | 0 | 0 | 0.004955865 | 0 | 0 | 0 | 0 |
| MMP_2 | 0 | -0.000472335 | 0 | 0 | 0 | 0.045397272 | 0 | 0 | 0 | 0 | 0.000561589 | 0 | 0 | 0 | 0 |
| hOSCAR | -0.008492392 | -0.000245821 | 0.003853961 | 0 | 0 | 0 | 0 | 0 | 0 | 0 | -0.007007454 | 0 | 0 | 0 | 0 |
| TNFRSF14 | 0 | 0.000303695 | 0.002205526 | 0 | 0 | 0.030423887 | 0 | 0 | 0 | 0 | 0.01084389 | 0 | 0 | 0 | 0 |
| MCP_1 | 0 | 0 | -0.006120683 | 0 | 0 | 0 | 0 | 0 | 0 | 0 | 0.003871411 | 0 | 0 | 0 | 0 |
| CCL16 | 0 | 0.000210583 | 0.002387806 | 0 | 0 | 0 | 0 | 0 | 0 | 0 | -0.002472867 | 0 | 0 | 0 | 0 |
| IgG_Fc_receptor_I | -0.017221851 | -0.000421598 | -0.013045318 | 0 | 0 | 0.003379114 | 0 | 0 | 0 | 0 | -0.006144441 | 0 | 0 | 0 | 0 |
| LTBR | 0 | 0.000443966 | -0.008782406 | 0 | 0 | 0 | 0 | 0 | 0 | 0 | -0.004689168 | 0 | 0 | 0 | 0 |

**Table S11: Elastic Net Model Coefficients (generated from SCAPIS)**

**e) Merged_Omics Models**

| Omics_Variables | Cholesterol_pMMS | Lipodystrophy_1_pMMS | Lipodystrophy_2_pMMS | Liver-Lipid_pMMS | ASAT_Pos_pMMS | ALP_Neg_pMMS | SHBG-LpA_pMMS | Obesity_pMMS | VAT_Neg_pMMS | Bilirubin_pMMS | Proinsulin_pMMS | Hyper_Insulin_pMMS | Beta_Cell_1_pMMS | Beta_Cell_2_pMMS | Total_GRS |
|---|---|---|---|---|---|---|---|---|---|---|---|---|---|---|---|
| Dkk_1 | 0 | 0.000255623 | -0.00147193 | 0 | 0 | 0 | 0 | 0 | 0 | 0 | -0.000524142 | 0 | 0 | 0 | 0 |
| MMP_9 | 0 | 0 | 0.000985424 | 0 | 0 | 0.032382514 | 0 | 0 | 0 | 0 | 0.007072342 | 0 | 0 | 0 | 0 |
| SORT1 | 0 | 0.000255561 | -0.002877314 | 0 | 0 | 0.03760989 | 0 | 0 | 0 | 0 | -0.014591216 | 0 | 0 | 0 | -0.024739833 |
| TNFRSF11A | 0.014653974 | 0.000219136 | -0.00305416 | 0 | 0 | 0 | 0 | 0 | 0 | 0 | -0.005896018 | 0 | 0 | 0 | 0 |
| SCF | -0.024176729 | -0.00031591 | 0.004093721 | -0.075184956 | 0 | -0.027787628 | 0 | 0 | 0 | 0 | 0.011623437 | 0 | 0 | 0.007080718 | 0.003899847 |
| PRELP | 0 | -0.000666083 | -0.007728875 | 0 | 0 | -0.003389225 | 0 | 0 | 0 | 0 | -0.008666293 | 0 | 0 | -0.001939397 | 0 |
| PAPPA | 0 | 1.67E-05 | -0.005023042 | 0 | 0 | 0 | 0 | 0 | 0 | 0 | -0.000760255 | 0 | 0 | 0 | 0 |
| TRAIL_R2 | 0 | 0.000175599 | 0.006389333 | 0 | 0 | -0.012530567 | 0 | 0 | 0 | 0 | 0.001974696 | 0 | 0 | -0.000639466 | 0 |
| ADAM_TS13 | 0.00530706 | -0.000371422 | 0.00390555 | 0 | 0 | 0 | 0 | 0 | 0 | 0 | -0.005483379 | 0 | 0 | -0.007897475 | 0 |
| BNP | 0 | -0.000494193 | -0.003593071 | 0 | 0 | 0.000916508 | 0 | 0 | 0 | 0 | -0.007412949 | 0 | 0 | 0 | 0 |
| IL16 | 0.005878467 | 0.000275323 | 0.000494977 | 0.051378933 | 0 | -0.023658083 | 0 | 0 | 0 | 0 | -0.001950287 | 0 | 0 | 0 | 0 |
| uPA | 0 | 0.000339251 | 0.003697294 | 0 | 0 | 0 | 0 | 0 | 0 | 0 | 0.00156413 | 0 | 0 | 0 | 0 |
| ALCAM | 0 | 0.000826351 | -0.001846199 | 0 | 0 | -0.014717971 | 0 | 0 | 0 | 0 | 0.003515188 | 0 | 0 | 0 | 0 |
| AMBP | 0 | -0.000121242 | -0.005665534 | 0 | 0 | 0 | 0 | 0 | 0 | 0 | -0.005928677 | 0 | 0 | 0 | 0 |
| LPL | -0.019859146 | -0.000387601 | 0.008857546 | 0 | 0 | -0.040055028 | 0 | 0 | 0 | 0 | -0.004024465 | 0 | 0 | 0 | 0 |
| GDF_2 | 0.009481738 | 0.000681738 | -0.005003671 | 0 | 0 | -0.014638643 | 0 | 0 | 0 | 0 | -0.004809293 | 0 | 0 | 0 | 0 |
| Notch_3 | 0 | 0 | 0.002786983 | 0 | 0 | -0.007919316 | 0 | 0 | 0 | 0 | 0.002916742 | 0 | 0 | -0.010947333 | 0 |
| TFF3 | 0 | 0.000891379 | -0.004189545 | 0.004348063 | 0 | 0 | 0 | 0 | 0 | 0 | 0.008658508 | 0 | 0 | 0 | 0.004972963 |
| NEMO | 0 | 6.20E-05 | 0.000464397 | 0 | 0 | 0 | 0 | 0 | 0 | 0 | 0.015556274 | 0 | 0 | 0 | 0 |
| RARRES2 | 0 | -0.000262216 | 0.006164645 | 0 | 0 | 0.045148205 | 0 | 0 | 0 | 0 | -0.002605023 | 0 | 0 | 0 | 0 |
| IL2_RA | 0 | 0.000872453 | 0.009934249 | 0 | 0 | 0 | 0 | 0 | 0 | 0 | 0.007960482 | 0 | 0 | 0 | 0 |
| TIMP4 | -0.00213347 | -0.001137848 | 0.008463439 | 0 | 0 | 0.001192433 | 0 | 0 | 0 | 0 | -0.003504174 | 0 | 0 | 0 | 0 |
| OPG | 0 | 3.86E-05 | 0.003595613 | 0 | 0 | -0.027863718 | 0 | 0 | 0 | 0 | -0.007724936 | 0 | 0 | 0 | 0 |
| IL_4RA | 0 | 0.000175004 | -0.007529826 | 0 | 0 | 0.000966878 | 0 | 0 | 0 | 0 | -0.000471116 | 0 | 0 | 0.015591816 | 0 |
| KLK6 | 0 | 0 | 0.00996484 | 0.007835628 | 0 | 0.018916993 | 0 | 0 | 0 | 0 | -0.000420091 | 0 | 0 | 0 | 0 |
| CASP_3 | 0 | 0.000879325 | 0.005470515 | 0 | 0 | 0 | 0 | 0 | 0 | 0 | 0.001804572 | 0 | 0 | 0 | 0 |
| BLM_hydrolase | 0 | 0.000770605 | 0.003654777 | 0 | 0 | 0.004793746 | 0 | 0 | 0 | 0 | -0.002222998 | 0 | 0 | 0 | 0 |
| SERPINA12 | 0 | 0.001043695 | 0.020698869 | 0 | 0 | 0 | 0 | 0 | 0 | 0 | 0.011596936 | 0 | 0 | 0 | 0 |
| IL_1RT1 | 0 | 0 | 0 | 0 | 0 | 0.03053799 | 0 | 0 | 0 | 0 | 0.01359983 | 0 | 0 | 0 | 0 |
| JAM_A | 0 | 0.000999558 | 0.006400354 | 0 | 0 | 0 | 0 | 0 | 0 | 0 | -0.00346837 | 0 | 0 | 0 | 0 |
| GRN | 0 | 0.000117954 | -0.017724192 | 0 | 0 | 0.050999873 | 0 | 0 | 0 | 0 | 0.011961211 | 0 | 0 | 0 | 0 |
| PAR_1 | 0 | 0 | -0.000538864 | 0 | 0 | 0 | 0 | 0 | 0 | 0 | -0.015685109 | 0 | 0 | 0 | 0 |
| HB_EGF | 0 | 0 | 0 | 0 | 0 | 0 | 0 | 0 | 0 | 0 | 0.009441233 | 0 | 0 | 0 | 0 |
| VSIG2 | 0 | -0.000241309 | 0.00794556 | 0.010000808 | 0 | 0.027318605 | 0 | 0 | 0 | 0 | -0.00654151 | 0 | 0 | -0.005471468 | 0 |
| EGFR | 0 | 0.000613636 | 0.004403317 | -0.020389953 | 0 | -0.001503219 | 0 | 0 | 0 | 0 | -0.009167021 | 0 | 0 | 0 | 0 |
| IL_17RA | 0 | 0.000112058 | 0.000869461 | 0 | 0 | 0.04503266 | 0 | 0 | 0 | 0 | 0.003496334 | 0 | 0 | 0 | 0 |
| TNFRSF13B | 0.000982872 | 0.000813676 | -0.002069959 | -0.021100586 | 0 | -0.010872933 | 0 | 0 | 0 | 0 | 0.006425128 | 0 | 0 | 0 | 0 |
| MMP_3 | 0 | 0.000703534 | 0.00316669 | 0 | 0 | 0.027612 | 0 | 0 | 0 | 0 | 0.002816057 | 0 | 0 | 0 | 0 |
| FABP2 | 0 | 0 | -0.006771192 | 0 | 0 | 0.071260666 | 0 | 0 | 0 | 0 | 0.006222129 | 0 | 0 | 0 | 0 |
| CCL24 | 0 | 0 | 6.71E-05 | 0 | 0 | 0 | 0 | 0 | 0 | 0 | 0.004863239 | 0 | 0 | 0 | 0 |
| ST2 | -0.036951727 | -0.000266133 | -0.014591418 | 0 | 0 | 0 | 0 | 0 | 0 | 0 | 0.00340125 | 0 | 0 | 0 | 0 |
| PAI | 0 | 0.000611914 | 0.002453758 | 0 | 0 | 0.005558504 | 0 | 0 | 0 | 0 | 0.005460862 | 0 | 0 | 0 | 0 |
| SRC | 0 | 0.00027583 | -0.000922946 | -0.0088003 | 0 | 0 | 0 | 0 | 0 | 0 | -0.008228284 | 0 | 0 | 0 | 0 |
| ITGB1BP2 | 0 | 0.000114266 | 0.008807875 | 0 | 0 | 0.002630514 | 0 | 0 | 0 | 0 | -0.010042184 | 0 | 0 | 0 | 0 |
| HO_1 | -0.03195694 | -0.000240084 | 0.005825181 | 0 | 0 | 0 | 0 | 0 | 0 | 0 | 0.014914004 | 0 | 0 | 0 | 0 |
| VEGFD | 0 | 0.000183442 | -0.004059026 | 0 | 0 | -0.019181833 | 0 | 0 | 0 | 0 | -0.005793494 | 0 | 0 | 0 | 0 |
| MEPE | 0 | -0.000330285 | -0.001169294 | 0 | 0 | 0 | 0 | 0 | 0 | 0 | -0.010188526 | 0 | 0 | 0 | 0 |
| KIM1 | 0 | 0.001043095 | 0.003363056 | 0 | 0 | -0.007524035 | 0 | 0.005205769 | 0 | 0 | -0.005408279 | 0 | 0 | 0 | 0 |
| PDGF_subunit_A | 0 | 0.0005476 | 0.000751469 | 0 | 0 | 0 | 0 | 0 | 0 | 0 | -0.006306108 | 0 | 0 | 0 | 0 |
| IL_17D | 0.021976234 | -0.000863493 | 0.013135354 | 0 | 0 | -0.008915111 | 0 | -0.023068535 | 0 | 0 | 0.003000064 | 0 | 0 | -0.011842431 | 0 |
| TNF_R1 | 0 | -0.000135501 | 0.002416418 | 0 | 0 | 0.027740488 | 0 | 0 | 0 | 0 | 0.002605546 | 0 | 0 | 0 | 0 |
| TLT_2 | 0 | 8.52E-05 | 0.004391402 | 0.000633017 | 0 | 0.040132841 | 0 | 0 | 0 | 0 | -0.00572944 | 0 | 0 | 0 | 0 |
| RETN | -0.023628147 | 1.51E-05 | 0.014146539 | 0 | 0 | 0.00827206 | 0 | 0 | 0 | 0 | 0.002330008 | 0 | 0 | 0 | 0 |
| HDL2_C | 0 | -0.000705913 | -0.003991385 | 0 | 0 | 0 | 0 | 0 | 0 | 0 | -0.001383363 | 0 | 0 | 0 | 0 |
| HDL3_C | 0 | 0 | 0.00045535 | 0 | 0 | 0 | 0 | 0 | 0 | 0 | -0.002496218 | 0 | 0 | 0 | 0 |
| HDL_C | 0 | -0.000652515 | -0.003617868 | 0 | 0 | 0 | 0 | 0 | 0 | 0 | -0.001584931 | 0 | 0 | 0 | 0 |
| HDL_D | 0 | -0.000737281 | -0.001362442 | 0 | 0 | 0 | 0 | 0 | 0 | 0 | 0.000372278 | 0 | 0 | 0 | 0 |
| HDL_TG | 0 | -3.28E-05 | -0.002634544 | 0 | 0 | 0 | 0 | 0 | 0 | 0 | 0.003974779 | 0 | 0 | 0 | 0 |
| L_HDL_C | 0 | -0.000578724 | -0.001327551 | 0 | 0 | 0 | 0 | 0 | 0 | 0 | 0.000309622 | 0 | 0 | 0 | 0 |
| L_HDL_CE | 0 | -0.000572007 | -0.001371753 | 0 | 0 | 0 | 0 | 0 | 0 | 0 | 0.000347531 | 0 | 0 | 0 | 0 |
| L_HDL_FC | 0 | -0.000594689 | -0.001399793 | 0 | 0 | 0 | 0 | 0 | 0 | 0 | 0.000185558 | 0 | 0 | 0 | 0 |
| L_HDL_L | 0 | -0.000583021 | -0.001647588 | 0 | 0 | 0 | 0 | 0 | 0 | 0 | 0.000755357 | 0 | 0 | 0 | 0 |
| L_HDL_P | 0 | -0.000585765 | -0.001697979 | 0 | 0 | 0 | 0 | 0 | 0 | 0 | 0.000813209 | 0 | 0 | 0 | 0 |
| L_HDL_PL | 0 | -0.000575723 | -0.001906296 | 0 | 0 | 0 | 0 | 0 | 0 | 0 | 0.000446138 | 0 | 0 | 0 | 0 |
| L_HDL_TG | 0 | -0.000554618 | -0.003780351 | 0.0076305 | 0 | 0.001884777 | 0 | 0 | 0 | 0 | 0.005007765 | 0 | 0 | 0 | 0 |
| M_HDL_C | 0 | -0.000402127 | -0.004068166 | 0 | 0 | 0 | 0 | 0 | 0 | 0 | -0.00120941 | 0 | 0 | 0 | 0 |
| M_HDL_CE | 0 | -0.000414072 | -0.00425246 | 0 | 0 | 0 | 0 | 0 | 0 | 0 | -0.001479795 | 0 | 0 | 0 | 0 |
| M_HDL_FC | 0 | -0.000354964 | -0.003462943 | 0 | 0 | 0 | 0 | 0 | 0 | 0 | -0.000422127 | 0 | 0 | 0 | 0 |
| M_HDL_L | 0 | -0.000336929 | -0.004616792 | 0 | 0 | 0 | 0 | 0 | 0 | 0 | 0 | 0 | 0 | 0 | 0 |
| M_HDL_P | 0 | -0.000326262 | -0.004659184 | 0 | 0 | 0 | 0 | 0 | 0 | 0 | 0.00014663 | 0 | 0 | 0 | 0 |
| M_HDL_PL | 0 | -0.000237247 | -0.003685896 | 0 | 0 | 0.000729418 | 0 | 0 | 0 | 0 | 0.001255684 | 0 | 0 | 0 | 0 |
| M_HDL_TG | 0 | 0.00013639 | -0.003423072 | 0 | 0 | 0 | 0 | 0 | 0 | 0 | -7.08E-06 | 0 | 0 | 0 | 0 |
| S_HDL_C | 0 | 0 | -0.005433249 | 0 | 0 | 0 | 0 | 0 | 0 | 0 | -0.000278712 | 0 | 0 | 0 | 0 |
| S_HDL_CE | 0 | 0 | -0.005871937 | 0 | 0 | 0 | 0 | 0 | 0 | 0 | -0.000915492 | 0 | 0 | 0 | 0 |
| S_HDL_FC | 0.007159136 | 7.98E-05 | 0.000118983 | -0.009051345 | 0 | 0 | 0 | 0 | 0 | 0 | 0.002179447 | 0 | 0 | 0 | 0 |
| S_HDL_L | 0 | 0.000334541 | -0.003556203 | 0 | 0 | 0 | 0 | 0 | 0 | 0 | 0.001168725 | 0 | 0 | 0 | 0 |

**Table S11: Elastic Net Model Coefficients (generated from SCAPIS)**

**e) Merged_Omics Models**

| Omics_Variables | Cholesterol_pMMS | Lipodystrophy_1_pMMS | Lipodystrophy_2_pMMS | Liver-Lipid_pMMS | ASAT_Pos_pMMS | ALP_Neg_pMMS | SHBG-LpA_pMMS | Obesity_pMMS | VAT_Neg_pMMS | Bilirubin_pMMS | Proinsulin_pMMS | Hyper_Insulin_pMMS | Beta_Cell_1_pMMS | Beta_Cell_2_pMMS | Total_GRS |
|---|---|---|---|---|---|---|---|---|---|---|---|---|---|---|---|
| S_HDL_P | 0 | 0.000383364 | -0.003361344 | 0 | 0 | 0 | 0 | 0 | 0 | 0 | 0.001345361 | 0 | 0 | 0 | 0 |
| S_HDL_PL | 0 | 0.000282705 | -0.000992138 | 0 | 0 | 0 | 0 | 0 | 0 | 0 | 0.000198504 | 0 | 0 | 0 | 0 |
| S_HDL_TG | 0 | 0.000865952 | 0.001974202 | 0 | 0 | 0 | 0 | 0 | 0 | 0 | 0.002856846 | 0 | 0 | 0 | 0 |
| XL_HDL_C | 0 | -0.000425016 | -6.25E-05 | 0 | 0 | 0 | 0 | 0 | 0 | 0 | -0.002060848 | 0 | 0 | 0 | 0 |
| XL_HDL_CE | 0 | -0.000389282 | 0 | 0.001091148 | 0 | 0 | 0 | 0 | 0 | 0 | -0.001638624 | 0 | 0 | 0 | 0 |
| XL_HDL_FC | 0 | -0.000529311 | -0.000387239 | 0 | 0 | 0 | 0 | 0 | 0 | 0 | -0.002790958 | 0 | 0 | 0 | 0 |
| XL_HDL_L | 0 | -0.000527904 | -0.001157987 | 0 | 0 | 0 | 0 | 0 | 0 | 0 | -0.002191745 | 0 | 0 | 0 | 0 |
| XL_HDL_P | 0 | -0.000531375 | -0.001290379 | 0 | 0 | 0 | 0 | 0 | 0 | 0 | -0.002101609 | 0 | 0 | 0 | 0 |
| XL_HDL_PL | 0 | -0.000611088 | -0.002134486 | 0 | 0 | 0 | 0 | 0 | 0 | 0 | -0.002812706 | 0 | 0 | 0 | 0 |
| XL_HDL_TG | 0 | -0.000223545 | -0.001120412 | 0 | 0 | 0 | 0 | 0 | 0 | 0 | 0.003977137 | 0 | 0 | 0.000713397 | 0 |
| IDL_C | 0 | 0 | -0.00096405 | 0 | 0 | 0 | 0 | 0 | 0 | 0 | -0.000512616 | 0 | 0 | 0 | 0 |
| IDL_CE | 0 | 6.45E-06 | -0.00134332 | 0 | 0 | 0 | 0 | 0 | 0 | 0 | -0.000368226 | 0 | 0 | 0 | 0 |
| IDL_FC | 0 | 0 | -0.000379082 | 0 | 0 | 0 | 0 | 0 | 0 | 0 | -0.000247786 | 0 | 0 | 0 | 0 |
| IDL_L | 0 | 7.80E-05 | 0 | 0 | 0 | 0 | 0 | 0 | 0 | 0 | -0.000893754 | 0 | 0 | 0 | 0 |
| IDL_P | 0 | 0.000128619 | -0.000175205 | 0 | 0 | 0 | 0 | 0 | 0 | 0 | -0.001519768 | 0 | 0 | 0 | 0 |
| IDL_PL | 0 | 5.08E-05 | -0.000256346 | 0 | 0 | 0 | 0 | 0 | 0 | 0 | -0.00095982 | 0 | 0 | 0 | 0 |
| IDL_TG | 0 | 0.00085088 | 0.007595466 | 0 | 0 | 0 | 0 | 0 | 0 | 0 | -0.001234224 | 0 | 0 | 0 | 0 |
| L_LDL_C | 0 | 8.95E-05 | -0.000262668 | 0 | 0 | 0 | 0 | 0 | 0 | 0 | 0 | 0 | 0 | 0 | 0 |
| L_LDL_CE | 0 | 0.000120104 | -0.000177575 | 0 | 0 | 0 | 0 | 0 | 0 | 0 | 5.43E-05 | 0 | 0 | 0 | 0 |
| L_LDL_FC | 0 | 0 | -0.00015215 | 0 | 0 | 0 | 0 | 0 | 0 | 0 | -0.000343337 | 0 | 0 | 0 | 0 |
| L_LDL_L | 0 | 0.000126897 | 3.08E-05 | 0 | 0 | 0 | 0 | 0 | 0 | 0 | -0.000235025 | 0 | 0 | 0 | 0 |
| L_LDL_P | 0 | 0.000154563 | 0.000246907 | 0 | 0 | 0 | 0 | 0 | 0 | 0 | -0.000356895 | 0 | 0 | 0 | 0 |
| L_LDL_PL | 0 | 9.44E-05 | -0.000441523 | 0 | 0 | 0 | 0 | 0 | 0 | 0 | -0.000599359 | 0 | 0 | 0 | 0 |
| L_LDL_TG | 0 | 0.000686677 | 0.007061615 | 0 | 0 | 0 | 0 | 0 | 0 | 0 | -0.00140027 | 0 | 0 | 0 | 0 |
| LDL_C | 0 | 9.88E-05 | 0 | 0 | 0 | 0 | 0 | 0 | 0 | 0 | 0.000211588 | 0 | 0 | 0 | 0 |
| LDL_D | 0 | -0.000114059 | 0.001090029 | 0 | 0.007500371 | 0.03136201 | 0 | 0 | 0 | 0 | -0.005380893 | 0 | 0 | 0 | 0 |
| LDL_TG | 0 | 0.000701063 | 0.006126617 | 0 | 0 | 0 | 0 | 0 | 0 | 0 | -0.000871079 | 0 | 0 | 0 | 0 |
| M_LDL_C | 0 | 0.000104507 | 0.000190044 | 0 | 0 | 0 | 0 | 0 | 0 | 0 | 0.000642418 | 0 | 0 | 0 | 0 |
| M_LDL_CE | 0 | 0.000117836 | 0.000204382 | 0 | 0 | 0 | 0 | 0 | 0 | 0 | 0.000736119 | 0 | 0 | 0 | 0 |
| M_LDL_FC | 0 | 5.81E-05 | -0.000120132 | 0 | 0 | 0 | 0 | 0 | 0 | 0 | 0.000135005 | 0 | 0 | 0 | 0 |
| M_LDL_L | 0 | 0.000179992 | 0.000380805 | 0 | 0 | 0 | 0 | 0 | 0 | 0 | 0.000308193 | 0 | 0 | 0 | 0 |
| M_LDL_P | 0 | 0.000196537 | 0.000528501 | 0 | 0 | 0 | 0 | 0 | 0 | 0 | 0.000255874 | 0 | 0 | 0 | 0 |
| M_LDL_PL | 0 | 0.000219551 | -0.000706347 | 0 | 0 | 0 | 0 | 0 | 0 | 0 | -0.000329533 | 0 | 0 | 0 | 0 |
| M_LDL_TG | 0 | 0.000719325 | 0.006101564 | 0 | 0 | 0 | 0 | 0 | 0 | 0 | -0.000993423 | 0 | 0 | 0 | 0 |
| S_LDL_C | 0 | 7.25E-05 | 0 | 0 | 0 | 0 | 0 | 0 | 0 | 0 | 0.000847509 | 0 | 0 | 0 | 0 |
| S_LDL_CE | 0 | 7.28E-05 | 0 | 0 | 0 | 0 | 0 | 0 | 0 | 0 | 0.000962492 | 0 | 0 | 0 | 0 |
| S_LDL_FC | 0 | 0.000106649 | -0.000870291 | 0 | 0 | 0 | 0 | 0 | 0 | 0 | 0.000302437 | 0 | 0 | 0 | 0 |
| S_LDL_L | 0 | 0.000148417 | -8.55E-05 | 0 | 0 | 0 | 0 | 0 | 0 | 0 | 0.000729755 | 0 | 0 | 0 | 0 |
| S_LDL_P | 0 | 0.000174285 | 0 | 0 | 0 | 0 | 0 | 0 | 0 | 0 | 0.000741538 | 0 | 0 | 0 | 0 |
| S_LDL_PL | 0 | 0.00016165 | -0.001581512 | 0 | 0 | 0 | 0 | 0 | 0 | 0 | 0.000115492 | 0 | 0 | 0 | 0 |
| S_LDL_TG | 0 | 0.000681929 | 0.003944675 | 0 | 0 | 0 | 0 | 0 | 0 | 0 | 0.000324836 | 0 | 0 | 0 | 0 |
| L_VLDL_C | 0 | 0.000400913 | -0.000668987 | 0 | 0 | 0 | 0 | 0 | 0 | 0 | 0.000409157 | 0 | 0 | 0 | 0 |
| L_VLDL_CE | 0 | 0.000301006 | -0.001263581 | 0 | 0 | 0 | 0 | 0 | 0 | 0 | 0.000210806 | 0 | 0 | 0 | 0 |
| L_VLDL_FC | 0 | 0.000510025 | 0 | 0 | 0 | 0 | 0 | 0 | 0 | 0 | 0.000338489 | 0 | 0 | 0 | 0 |
| L_VLDL_L | 0 | 0.000573375 | -0.000214816 | 0 | 0 | 0 | 0 | 0 | 0 | 0 | 0.000541993 | 0 | 0 | 0 | 0 |
| L_VLDL_P | 0 | 0.000590544 | -0.000207823 | 0 | 0 | 0 | 0 | 0 | 0 | 0 | 0.000459481 | 0 | 0 | 0 | 0 |
| L_VLDL_PL | 0 | 0.000568729 | 8.63E-05 | 0 | 0 | 0 | 0 | 0 | 0 | 0 | 0.00085109 | 0 | 0 | 0 | 0 |
| L_VLDL_TG | 0 | 0.000645729 | -0.000191746 | 0 | 0 | 0 | 0 | 0 | 0 | 0 | 0.000573841 | 0 | 0 | 0 | 0 |
| M_VLDL_C | 0 | 0.000215469 | -0.001307741 | 0 | 0 | 0 | 0 | 0 | 0 | 0 | 0.000902373 | 0 | 0 | 0 | 0 |
| M_VLDL_CE | 0 | 0 | -0.003014439 | 0 | 0 | 0 | 0 | 0 | 0 | 0 | 0.000116116 | 0 | 0 | 0 | 0 |
| M_VLDL_FC | 0 | 0.000627692 | 0.000790692 | 0 | 0 | 0 | 0 | 0 | 0 | 0 | 0.001902666 | 0 | 0 | 0 | 0 |
| M_VLDL_L | 0 | 0.000611593 | 0 | 0 | 0 | 0 | 0 | 0 | 0 | 0 | 0.001861079 | 0 | 0 | 0 | 0 |
| M_VLDL_P | 0 | 0.000645467 | 0 | 0 | 0 | 0 | 0 | 0 | 0 | 0 | 0.001813823 | 0 | 0 | 0 | 0 |
| M_VLDL_PL | 0 | 0.000609248 | 0.000272684 | 0 | 0 | 0 | 0 | 0 | 0 | 0 | 0.001838816 | 0 | 0 | 0 | 0 |
| M_VLDL_TG | 0 | 0.000807507 | 0.000517491 | 0 | 0 | 0 | 0 | 0 | 0 | 0 | 0.002220918 | 0 | 0 | 0 | 0 |
| S_VLDL_C | 0 | 3.50E-05 | -0.002977513 | 0 | 0 | 0 | 0 | 0 | 0 | 0 | 9.83E-05 | 0 | 0 | 0 | 0 |
| S_VLDL_CE | 0 | -3.87E-06 | -0.003892087 | 0 | 0 | 0 | 0 | 0 | 0 | 0 | -0.000294948 | 0 | 0 | 0 | 0 |
| S_VLDL_FC | 0 | 0.000449248 | -0.000286123 | 0 | 0 | 0 | 0 | 0 | 0 | 0 | 0.000554951 | 0 | 0 | 0 | 0 |
| S_VLDL_L | 0 | 0.000498956 | -5.06E-05 | 0 | 0 | 0 | 0 | 0 | 0 | 0 | 0.000984065 | 0 | 0 | 0 | 0 |
| S_VLDL_P | 0 | 0.000547283 | 0.000172741 | 0 | 0 | 0 | 0 | 0 | 0 | 0 | 0.001165368 | 0 | 0 | 0 | 0 |
| S_VLDL_PL | 0 | 0.000527849 | -0.000181744 | 0 | 0 | 0 | 0 | 0 | 0 | 0 | -0.000284986 | 0 | 0 | 0 | 0 |
| S_VLDL_TG | 0 | 0.000821233 | 0.002533855 | 0 | 0 | 0 | 0 | 0 | 0 | 0 | 0.001633164 | 0 | 0 | 0 | 0 |
| Serum_TG | 0 | 0.00066993 | 0.001691205 | 0 | 0 | 0 | 0 | 0 | 0 | 0 | 0.001820722 | 0 | 0 | 0 | 0 |
| VLDL_C | 0 | 7.78E-05 | -0.002143595 | 0 | 0 | 0 | 0 | 0 | 0 | 0 | 0.001064084 | 0 | 0 | 0 | 0 |
| VLDL_D | 0 | 0.000676869 | -0.00069571 | 0 | 0 | 0 | 0 | 0 | 0 | 0 | 0.001624558 | 0 | 0 | 0 | 0 |
| VLDL_TG | 0 | 0.000769905 | 0.001083917 | 0 | 0 | 0 | 0 | 0 | 0 | 0 | 0.001829599 | 0 | 0 | 0 | 0 |
| XL_VLDL_C | 0 | 2.44E-05 | -0.001327707 | 0 | 0 | 0 | 0 | 0 | 0 | 0 | 0.00088523 | 0 | 0 | 0 | 0 |
| XL_VLDL_CE | 0 | 0 | -0.001246399 | 0 | 0 | 0 | 0 | 0 | 0 | 0 | 0.000908349 | 0 | 0 | 0 | 0 |
| XL_VLDL_FC | 0 | 4.52E-05 | -0.001454405 | 0 | 0 | 0 | 0 | 0 | 0 | 0 | 0.000419808 | 0 | 0 | 0 | 0 |
| XL_VLDL_L | 0 | 0.000188321 | -0.001237488 | 0 | 0 | 0 | 0 | 0 | 0 | 0 | -0.000670085 | 0 | 0 | 0 | 0 |
| XL_VLDL_P | 0 | 0.000194691 | -0.001112356 | 0 | 0 | 0 | 0 | 0 | 0 | 0 | -0.000907709 | 0 | 0 | 0 | 0 |
| XL_VLDL_PL | 0 | 0.000132246 | -0.001259419 | 0 | 0 | 0 | 0 | 0 | 0 | 0 | -0.000171812 | 0 | 0 | 0 | 0 |
| XL_VLDL_TG | 0 | 0.000246834 | -0.001181887 | 0 | 0 | 0 | 0 | 0 | 0 | 0 | -0.001479398 | 0 | 0 | 0 | 0 |
| XS_VLDL_C | 0 | -2.27E-05 | -0.001183816 | 0 | 0 | 0 | 0 | 0 | 0 | 0 | -0.001904332 | 0 | 0 | 0 | 0 |
| XS_VLDL_CE | 0 | -6.72E-05 | -0.00187941 | 0 | 0 | 0 | 0 | 0 | 0 | 0 | -0.001955919 | 0 | 0 | 0 | 0 |
| XS_VLDL_FC | 0 | 0 | 0.0001139 | 0 | 0 | 0 | 0 | 0 | 0 | 0 | -0.002220353 | 0 | 0 | 0 | 0 |

**Table S11: Elastic Net Model Coefficients (generated from SCAPIS)**

**e) Merged_Omics Models**

| Omics_Variables | Cholesterol_pMMS | Lipodystrophy_1_pMMS | Lipodystrophy_2_pMMS | Liver-Lipid_pMMS | ASAT_Pos_pMMS | ALP_Neg_pMMS | SHBG-LpA_pMMS | Obesity_pMMS | VAT_Neg_pMMS | Bilirubin_pMMS | Proinsulin_pMMS | Hyper_Insulin_pMMS | Beta_Cell_1_pMMS | Beta_Cell_2_pMMS | Total_GRS |
|---|---|---|---|---|---|---|---|---|---|---|---|---|---|---|---|
| XS_VLDL_L | 0 | 0.000122999 | 0.000312945 | 0 | 0 | 0 | 0 | 0 | 0 | 0 | -0.001559558 | 0 | 0 | 0 | 0 |
| XS_VLDL_P | 0 | 0.000197378 | 0.000751958 | 0 | 0 | 0 | 0 | 0 | 0 | 0 | -0.001342867 | 0 | 0 | 0 | 0 |
| XS_VLDL_PL | 0 | 0.00015098 | 0.000643956 | 0 | 0 | 0 | 0 | 0 | 0 | 0 | -0.001946137 | 0 | 0 | 0 | 0 |
| XS_VLDL_TG | 0 | 0.000839414 | 0.004886734 | 0 | 0 | 0 | 0 | 0 | 0 | 0 | 0.000357482 | 0 | 0 | 0 | 0 |
| XXL_VLDL_C | 0 | 0 | -0.002689897 | 0 | 0 | 0 | 0 | 0 | 0 | 0 | 0.001041559 | 0 | 0 | 0 | 0 |
| XXL_VLDL_CE | 0 | -4.25E-05 | -0.003022491 | 0 | 0 | 0 | 0 | 0 | 0 | 0 | 0.001456713 | 0 | 0 | 0 | 0 |
| XXL_VLDL_FC | 0 | 7.53E-05 | -0.001800106 | 0 | 0 | 0 | 0 | 0 | 0 | 0 | -0.000352025 | 0 | 0 | 0 | 0 |
| XXL_VLDL_L | 0 | 0 | -0.004175457 | 0 | 0 | 0 | 0 | 0 | 0 | 0 | 6.37E-05 | 0 | 0 | 0 | 0 |
| XXL_VLDL_P | 0 | 1.18E-06 | -0.004634782 | 0 | 0 | 0 | 0 | 0 | 0 | 0 | -0.000142509 | 0 | 0 | 0 | 0 |
| XXL_VLDL_PL | 0 | 9.98E-05 | -0.001968895 | 0 | 0 | 0 | 0 | 0 | 0 | 0 | -0.000303596 | 0 | 0 | 0 | 0 |
| XXL_VLDL_TG | 0 | 2.43E-05 | -0.005049411 | 0 | 0 | 0 | 0 | 0 | 0 | 0 | -0.000409109 | 0 | 0 | 0 | 0 |
| AcAce | -0.010452809 | -0.000651894 | -0.010522925 | 0 | 0 | 0 | 0 | 0 | 0 | 0 | -0.003175962 | 0 | 0 | 0 | 0 |
| Ace | 0 | 0.000607077 | -0.009366727 | 0.007906501 | 0 | -0.020614714 | 0 | 0 | 0 | 0 | -0.013236428 | 0 | 0 | 0 | 0 |
| Ala | 0 | 0.001150909 | 0.002777975 | -0.08185671 | 0 | 0 | 0 | 0 | 0 | 0 | 0.00077512 | 0 | 0 | 0.002740293 | 0 |
| Alb | 0.050253617 | -0.000760861 | 0.007400474 | -0.009219233 | 0 | 0 | 0 | 0 | 0 | 0 | 0.006137316 | 0 | 0 | 0 | 0 |
| bOHBut | 0 | -0.001427785 | -0.004264655 | 0 | 0 | 0 | 0 | 0 | 0 | 0 | 0.007196434 | 0 | 0 | 0 | 0 |
| Cit | 0 | -0.000397834 | -0.012902461 | 0 | 0 | 0 | 0 | 0 | 0 | 0 | -0.00128519 | 0 | 0 | 0 | 0 |
| Crea | 0 | 0.000540416 | -0.006691277 | 0.032497294 | 0 | 0 | 0 | 0 | 0 | 0 | -0.006192979 | 0 | 0 | 0 | 0 |
| DHA | 0.021763792 | -0.000289377 | 0.0013651 | 0 | 0 | 0 | 0 | 0 | 0 | 0 | -0.001208479 | 0 | 0 | 0 | 0 |
| EstC | 0 | 0 | -0.001909237 | 0 | 0 | 0 | 0 | 0 | 0 | 0 | 1.23E-05 | 0 | 0 | 0 | 0 |
| FAw3 | 0 | -0.000404867 | -0.000620916 | 0 | 0 | 0.053048009 | 0 | 0 | 0 | 0 | 0.001136149 | 0 | 0 | 0 | 0 |
| FAw6 | 0 | -0.000114653 | -0.002260112 | 0 | 0 | 0 | 0 | 0 | 0 | 0 | 0.001087553 | 0 | 0 | 0 | 0 |
| FreeC | 0 | 0 | -0.000859957 | 0 | 0 | 0 | 0 | 0 | 0 | 0 | 0 | 0 | 0 | 0 | 0 |
| Gln | 0 | -0.000520437 | 0.00470152 | 0.01681301 | 0 | -0.001771929 | 0 | 0 | 0 | 0 | 0.008792331 | 0 | 0 | 0.009331552 | 0 |
| Gp | -0.041041368 | -0.000386605 | 0.005178423 | -0.003616577 | 0 | 0.033277676 | 0 | 0 | 0 | 0 | 0.006977765 | 0 | 0 | 0 | 0 |
| His | 0.006385281 | -0.000673936 | -0.003549594 | 0 | 0 | -0.011823941 | 0 | 0 | 0 | 0 | -0.002761042 | 0 | 0 | 0 | 0 |
| Ile | 0 | 0.000608028 | -0.000261767 | 0 | 0 | 0 | 0 | 0 | 0 | 0 | -0.001440087 | 0 | 0 | 0 | 0 |
| LA | -0.050404178 | -5.35E-05 | -0.001134187 | 0 | 0 | -0.012897318 | 0 | 0 | 0 | 0 | 0.002892725 | 0 | 0 | 0 | 0 |
| Lac | 0 | 0.00160769 | -0.000552738 | 0 | 0 | 0.001258653 | 0 | 0 | 0 | 0 | -0.000565084 | 0 | 0 | 0 | 0 |
| Leu | 0 | 0.000281142 | -0.002442586 | 0 | 0 | 0.056746549 | 0 | 0 | 0 | 0 | 0.003920329 | 0 | 1.26E-19 | 0 | 0 |
| MUFA | 0 | 0.000124123 | -0.000381761 | 0 | 0 | 0 | 0 | 0 | 0 | 0 | -0.00020083 | 0 | 0 | 0 | 0 |
| PC | 0 | -0.000151274 | -0.000733119 | 0 | -0.006053318 | 0 | 0 | 0 | 0 | 0 | 0.000443403 | 0 | 0 | 0 | 0 |
| Phe | -0.021781756 | 0.000571402 | -0.009612741 | 0 | 0 | 0 | 0 | 0 | 0 | 0 | -0.019140654 | 0 | 0 | 0 | 0 |
| PUFA | 0 | -0.000177705 | -0.00202834 | 0 | 0 | 0 | 0 | 0 | 0 | 0 | 0.000924172 | 0 | 0 | 0 | 0 |
| Remnant_C | 0 | 2.25E-05 | -0.001664918 | 0 | 0 | 0 | 0 | 0 | 0 | 0 | 0.000238963 | 0 | 0 | 0 | 0 |
| Serum_C | 0 | 0 | -0.001618422 | 0 | 0 | 0 | 0 | 0 | 0 | 0 | 0.000123703 | 0 | 0 | 0 | 0 |
| SFA | 0 | 0.000124999 | -0.002171051 | 0 | 0 | 0 | 0 | 0 | 0 | 0 | -0.002197784 | 0 | 0 | 0 | 0 |
| SM | 0 | -0.000525662 | -0.002031434 | 0 | 0 | 0.004355152 | 0 | 0 | 0 | 0 | -0.000151868 | 0 | 0 | 0 | 0 |
| TotCho | 0 | -0.000289082 | -0.001218289 | 0 | 0 | 0.017755828 | 0 | 0 | 0 | 0 | 0.000591045 | 0 | 0 | 0 | 0 |
| TotFA | 0 | 5.14E-06 | -0.002091606 | 0 | 0 | 0 | 0 | 0 | 0 | 0 | -0.000502952 | 0 | 0 | 0 | 0 |
| TotPG | 0 | -0.000198761 | -0.001359895 | 0 | 0 | 0 | 0 | 0 | 0 | 0 | 0.002274994 | 0 | 0 | 0 | 0 |
| Tyr | 0 | 0.000469833 | 0.004651207 | 0.008645121 | 0 | 0.014655731 | 0 | 0 | 0 | 0 | 0.002422277 | 0 | 0 | 0 | 0 |
| UnSat | 0.005376415 | -0.000919454 | 0.000826823 | 0.042109936 | 0 | 0.031516119 | 0 | 0 | 0 | 0 | 0.001129085 | 0 | 0 | 0 | 0 |
| Val | 0 | 0.000191953 | 0.001341436 | 0 | 0 | 0 | 0 | 0 | 0 | 0 | -0.000807863 | 0 | 0 | 0 | 0 |

**Table S12: Associations between pPSs, pMMSs and imaging risk scores in SCAPIS**

**a) Spearman Correlation with Genetic PPSs**

| Riskscore | PPS | Sample_Size | r | CI95% | p | fdr_p_adjusted |
|---|---|---|---|---|---|---|
| CACS_Tot | Cholesterol_PPS | 26125 | -0.036954262 | [-0.05 -0.02] | 2.32E-09 | 3.00E-08 |
| CACS_Tot | Lipodystrophy_1_PPS | 26125 | 0.017426192 | [0.01 0.03] | 0.004861545 | 0.010417597 |
| CACS_Tot | Lipodystrophy_2_PPS | 26125 | -0.000573477 | [-0.01 0.01] | 0.926168302 | 0.963586303 |
| CACS_Tot | Liver-Lipid_PPS | 26125 | -0.018975986 | [-0.03 -0.01] | 0.002165395 | 0.005128566 |
| CACS_Tot | ASAT_Pos_PPS | 26125 | 0.006428931 | [-0.01 0.02] | 0.298876756 | 0.373595945 |
| CACS_Tot | ALP_Neg_PPS | 26125 | 0.018120211 | [0.01 0.03] | 0.003408881 | 0.007669981 |
| CACS_Tot | SHBG-LpA_PPS | 26125 | 0.022416845 | [0.01 0.03] | 0.000291504 | 0.001190685 |
| CACS_Tot | Obesity_PPS | 26125 | 0.019016722 | [0.01 0.03] | 0.002118182 | 0.005128566 |
| CACS_Tot | VAT_Neg_PPS | 26125 | 0.007602489 | [-0. 0.02] | 0.219265156 | 0.281912344 |
| CACS_Tot | Bilirubin_PPS | 26125 | 0.007994725 | [-0. 0.02] | 0.196402916 | 0.259945035 |
| CACS_Tot | Proinsulin_PPS | 26125 | 0.016926711 | [0. 0.03] | 0.00623159 | 0.012746434 |
| CACS_Tot | Hyper_Insulin_PPS | 26125 | 0.002499006 | [-0.01 0.01] | 0.686353426 | 0.73537867 |
| CACS_Tot | Beta_Cell_1_PPS | 26125 | 0.035809752 | [0.02 0.05] | 7.11E-09 | 6.40E-08 |
| CACS_Tot | Beta_Cell_2_PPS | 26125 | -0.004926085 | [-0.02 0.01] | 0.426033054 | 0.504512827 |
| CACS_Tot | Total_GRS | 26125 | 0.021964607 | [0.01 0.03] | 0.000385769 | 0.001239971 |
| DukeIndex | Cholesterol_PPS | 24938 | -0.035533116 | [-0.05 -0.02] | 2.01E-08 | 1.50E-07 |
| DukeIndex | Lipodystrophy_1_PPS | 24938 | 0.020225707 | [0.01 0.03] | 0.001406104 | 0.003954668 |
| DukeIndex | Lipodystrophy_2_PPS | 24938 | -0.004559792 | [-0.02 0.01] | 0.471607366 | 0.530558287 |
| DukeIndex | Liver-Lipid_PPS | 24938 | -0.024448224 | [-0.04 -0.01] | 0.000113266 | 0.00058413 |
| DukeIndex | ASAT_Pos_PPS | 24938 | 0.011159817 | [-0. 0.02] | 0.078090404 | 0.114486417 |
| DukeIndex | ALP_Neg_PPS | 24938 | 0.015160113 | [0. 0.03] | 0.016688863 | 0.030039953 |
| DukeIndex | SHBG-LpA_PPS | 24938 | 0.022745211 | [0.01 0.04] | 0.000329043 | 0.001190685 |
| DukeIndex | Obesity_PPS | 24938 | 0.011076332 | [-0. 0.02] | 0.080343943 | 0.114486417 |
| DukeIndex | VAT_Neg_PPS | 24938 | 0.011385768 | [-0. 0.02] | 0.072248391 | 0.112109572 |
| DukeIndex | Bilirubin_PPS | 24938 | 0.011393919 | [-0. 0.02] | 0.072044528 | 0.112109572 |
| DukeIndex | Proinsulin_PPS | 24938 | 0.017210896 | [0. 0.03] | 0.006581373 | 0.012876599 |
| DukeIndex | Hyper_Insulin_PPS | 24938 | 0.005393951 | [-0.01 0.02] | 0.394458986 | 0.479747416 |
| DukeIndex | Beta_Cell_1_PPS | 24938 | 0.043283349 | [0.03 0.06] | 8.12E-12 | 1.83E-10 |
| DukeIndex | Beta_Cell_2_PPS | 24938 | -0.000456612 | [-0.01 0.01] | 0.942533395 | 0.963586303 |
| DukeIndex | Total_GRS | 24938 | 0.022671878 | [0.01 0.04] | 0.000343976 | 0.001190685 |
| SIS | Cholesterol_PPS | 24938 | -0.037680909 | [-0.05 -0.03] | 2.67E-09 | 3.00E-08 |
| SIS | Lipodystrophy_1_PPS | 24938 | 0.021368357 | [0.01 0.03] | 0.000741259 | 0.002223777 |
| SIS | Lipodystrophy_2_PPS | 24938 | -0.004183448 | [-0.02 0.01] | 0.508964288 | 0.55861934 |
| SIS | Liver-Lipid_PPS | 24938 | -0.025390704 | [-0.04 -0.01] | 6.09E-05 | 0.000391789 |
| SIS | ASAT_Pos_PPS | 24938 | 0.010946377 | [-0. 0.02] | 0.083956706 | 0.114486417 |
| SIS | ALP_Neg_PPS | 24938 | 0.016153924 | [0. 0.03] | 0.010759471 | 0.020174008 |
| SIS | SHBG-LpA_PPS | 24938 | 0.02284637 | [0.01 0.04] | 0.000309438 | 0.001190685 |
| SIS | Obesity_PPS | 24938 | 0.013161282 | [0. 0.03] | 0.037719948 | 0.065284525 |
| SIS | VAT_Neg_PPS | 24938 | 0.01166464 | [-0. 0.02] | 0.065536594 | 0.109227657 |
| SIS | Bilirubin_PPS | 24938 | 0.011023598 | [-0. 0.02] | 0.081794418 | 0.114486417 |
| SIS | Proinsulin_PPS | 24938 | 0.019834833 | [0.01 0.03] | 0.00173816 | 0.004601012 |
| SIS | Hyper_Insulin_PPS | 24938 | 0.004780391 | [-0.01 0.02] | 0.450433122 | 0.519730525 |
| SIS | Beta_Cell_1_PPS | 24938 | 0.046315534 | [0.03 0.06] | 2.56E-13 | 1.15E-11 |
| SIS | Beta_Cell_2_PPS | 24938 | 0.000289182 | [-0.01 0.01] | 0.963586303 | 0.963586303 |
| SIS | Total_GRS | 24938 | 0.024400302 | [0.01 0.04] | 0.000116826 | 0.00058413 |

**b) Partial Correlation with Genetic PPSs**

| Riskscore | PPS | Sample_Size | r | CI95% | p | fdr_p_adjusted |
|---|---|---|---|---|---|---|
| CACS_Tot | Cholesterol_pPS | 26125 | -0.038971341 | [-0.05 -0.03] | 3.01E-10 | 4.22E-09 |
| CACS_Tot | Lipodystrophy_1_pPS | 26125 | 0.013548738 | [0. 0.03] | 0.028606933 | 0.060074559 |
| CACS_Tot | Lipodystrophy_2_pPS | 26125 | 0.000696637 | [-0.01 0.01] | 0.910395374 | 0.93260014 |
| CACS_Tot | Liver-Lipid_pPS | 26125 | -0.021547724 | [-0.03 -0.01] | 0.00049884 | 0.002095126 |
| CACS_Tot | ASAT_Pos_pPS | 26125 | 0.002410531 | [-0.01 0.01] | 0.696969049 | 0.791154056 |
| CACS_Tot | ALP_Neg_pPS | 26125 | 0.018165906 | [0.01 0.03] | 0.003336621 | 0.010009864 |
| CACS_Tot | SHBG-LpA_pPS | 26125 | 0.021885572 | [0.01 0.03] | 0.000406271 | 0.001895931 |
| CACS_Tot | Obesity_pPS | 26125 | 0.020336707 | [0.01 0.03] | 0.001017373 | 0.003286896 |
| CACS_Tot | VAT_Neg_pPS | 26125 | 0.00287132 | [-0.01 0.02] | 0.642751878 | 0.771302254 |
| CACS_Tot | Bilirubin_pPS | 26125 | 0.00657419 | [-0.01 0.02] | 0.288211359 | 0.432317038 |
| CACS_Tot | Proinsulin_pPS | 26125 | 0.001924033 | [-0.01 0.01] | 0.755935555 | 0.826720244 |
| CACS_Tot | Hyper_Insulin_pPS | 26125 | -0.000251728 | [-0.01 0.01] | 0.967562175 | 0.967562175 |
| CACS_Tot | Beta_Cell_1_pPS | 26125 | 0.035017644 | [0.02 0.05] | 1.52E-08 | 1.07E-07 |
| CACS_Tot | Beta_Cell_2_pPS | 26125 | -0.01318765 | [-0.03 -0. ] | 0.033128658 | 0.066257316 |
| DukeIndex | Cholesterol_pPS | 24938 | -0.03658805 | [-0.05 -0.02] | 7.62E-09 | 6.40E-08 |
| DukeIndex | Lipodystrophy_1_pPS | 24938 | 0.015577497 | [0. 0.03] | 0.013941974 | 0.032531273 |
| DukeIndex | Lipodystrophy_2_pPS | 24938 | -0.00417272 | [-0.02 0.01] | 0.510161791 | 0.669587351 |
| DukeIndex | Liver-Lipid_pPS | 24938 | -0.027193387 | [-0.04 -0.01] | 1.76E-05 | 9.27E-05 |
| DukeIndex | ASAT_Pos_pPS | 24938 | 0.006869908 | [-0.01 0.02] | 0.278234212 | 0.432317038 |
| DukeIndex | ALP_Neg_pPS | 24938 | 0.01559604 | [0. 0.03] | 0.013828716 | 0.032531273 |
| DukeIndex | SHBG-LpA_pPS | 24938 | 0.021411393 | [0.01 0.03] | 0.00072549 | 0.002539215 |
| DukeIndex | Obesity_pPS | 24938 | 0.011866678 | [-0. 0.02] | 0.061069648 | 0.116587509 |
| DukeIndex | VAT_Neg_pPS | 24938 | 0.005839879 | [-0.01 0.02] | 0.35667491 | 0.489241011 |
| DukeIndex | Bilirubin_pPS | 24938 | 0.009822005 | [-0. 0.02] | 0.121081417 | 0.203416781 |
| DukeIndex | Proinsulin_pPS | 24938 | 0.001622699 | [-0.01 0.01] | 0.797864603 | 0.837757833 |
| DukeIndex | Hyper_Insulin_pPS | 24938 | 0.002690743 | [-0.01 0.02] | 0.671068212 | 0.782912914 |
| DukeIndex | Beta_Cell_1_pPS | 24938 | 0.041293946 | [0.03 0.05] | 7.02E-11 | 1.47E-09 |
| DukeIndex | Beta_Cell_2_pPS | 24938 | -0.01044498 | [-0.02 0. ] | 0.0992332 | 0.173658101 |
| SIS | Cholesterol_pPS | 24938 | -0.038917671 | [-0.05 -0.03] | 8.01E-10 | 8.41E-09 |
| SIS | Lipodystrophy_1_pPS | 24938 | 0.016757455 | [0. 0.03] | 0.00816849 | 0.022871772 |
| SIS | Lipodystrophy_2_pPS | 24938 | -0.003603736 | [-0.02 0.01] | 0.569504337 | 0.724823702 |
| SIS | Liver-Lipid_pPS | 24938 | -0.028366616 | [-0.04 -0.02] | 7.53E-06 | 4.52E-05 |
| SIS | ASAT_Pos_pPS | 24938 | 0.006310837 | [-0.01 0.02] | 0.31922432 | 0.462324877 |
| SIS | ALP_Neg_pPS | 24938 | 0.016296476 | [0. 0.03] | 0.010103889 | 0.026522708 |
| SIS | SHBG-LpA_pPS | 24938 | 0.021447083 | [0.01 0.03] | 0.000710758 | 0.002539215 |
| SIS | Obesity_pPS | 24938 | 0.013883491 | [0. 0.03] | 0.028426813 | 0.060074559 |
| SIS | VAT_Neg_pPS | 24938 | 0.005786274 | [-0.01 0.02] | 0.36110646 | 0.489241011 |
| SIS | Bilirubin_pPS | 24938 | 0.009380242 | [-0. 0.02] | 0.138734019 | 0.224108801 |
| SIS | Proinsulin_pPS | 24938 | 0.003018895 | [-0.01 0.02] | 0.63373593 | 0.771302254 |
| SIS | Hyper_Insulin_pPS | 24938 | 0.001871787 | [-0.01 0.01] | 0.767668798 | 0.826720244 |
| SIS | Beta_Cell_1_pPS | 24938 | 0.043748335 | [0.03 0.06] | 4.91E-12 | 2.06E-10 |
| SIS | Beta_Cell_2_pPS | 24938 | -0.010523669 | [-0.02 0. ] | 0.096712841 | 0.173658101 |

**Table S12: Associations between pPSs, pMMSs and imaging risk scores in SCAPIS**

**c) Spearman Correlation with pMMSs Derived from Biochemistry Elastic Net Models**

| Riskscore | pMMS | Sample_Si | r | CI95% | p | fdr_p_adjusted |
|---|---|---|---|---|---|---|
| CACS_Tot | Cholesterol_pM | 23479 | -0.08152 | [-0.09 -0.0 | 6.70E-36 | 4.69E-35 |
| CACS_Tot | Lipodystrophy_1 | 23479 | 0.109528 | [0.1 0.12] | 1.51E-63 | 2.12E-62 |
| CACS_Tot | Lipodystrophy_2 | 23479 | 0.026448 | [0.01 0.04] | 5.08E-05 | 5.47E-05 |
| CACS_Tot | Liver-Lipid_pMN | 23479 | -0.00069 | [-0.01 0.0 | 0.916263 | 0.916263268 |
| CACS_Tot | ASAT_Pos_pMN | 23479 | 0.061691 | [0.05 0.07] | 3.11E-21 | 7.69E-21 |
| CACS_Tot | ALP_Neg_pMM | 23479 | 0.05586 | [0.04 0.07] | 1.10E-17 | 1.84E-17 |
| CACS_Tot | SHBG-LpA_pMN | 23479 | 0.053851 | [0.04 0.07] | 1.52E-16 | 2.36E-16 |
| CACS_Tot | Obesity_pMMS_ | 23479 | 0.055458 | [0.04 0.07] | 1.87E-17 | 3.02E-17 |
| CACS_Tot | VAT_Neg_pMM | 23479 | 0.073544 | [0.06 0.09] | 1.63E-29 | 7.62E-29 |
| CACS_Tot | Proinsulin_pMN | 23479 | 0.047184 | [0.03 0.06] | 4.77E-13 | 6.26E-13 |
| CACS_Tot | Hyper_Insulin_p | 23479 | 0.038857 | [0.03 0.05] | 2.61E-09 | 2.88E-09 |
| CACS_Tot | Beta_Cell_1_pN | 23479 | 0.055974 | [0.04 0.07] | 9.41E-18 | 1.65E-17 |
| CACS_Tot | Beta_Cell_2_pN | 23479 | 0.039609 | [0.03 0.05] | 1.28E-09 | 1.45E-09 |
| CACS_Tot | Total_GRS_biocl | 23479 | 0.056143 | [0.04 0.07] | 7.51E-18 | 1.37E-17 |
| DukeIndex | Cholesterol_pM | 22493 | -0.08294 | [-0.1 -0.07 | 1.28E-35 | 7.69E-35 |
| DukeIndex | Lipodystrophy_1 | 22493 | 0.126241 | [0.11 0.14] | 1.61E-80 | 3.38E-79 |
| DukeIndex | Lipodystrophy_2 | 22493 | 0.043687 | [0.03 0.06] | 5.63E-11 | 6.76E-11 |
| DukeIndex | Liver-Lipid_pMN | 22493 | -0.00982 | [-0.02 0. ] | 0.140949 | 0.147996327 |
| DukeIndex | ASAT_Pos_pMN | 22493 | 0.072895 | [0.06 0.09] | 7.14E-28 | 2.72E-27 |
| DukeIndex | ALP_Neg_pMM | 22493 | 0.062679 | [0.05 0.08] | 5.12E-21 | 1.19E-20 |
| DukeIndex | SHBG-LpA_pMN | 22493 | 0.061376 | [0.05 0.07] | 3.24E-20 | 6.81E-20 |
| DukeIndex | Obesity_pMMS_ | 22493 | 0.064168 | [0.05 0.08] | 5.94E-22 | 1.56E-21 |
| DukeIndex | VAT_Neg_pMM | 22493 | 0.095561 | [0.08 0.11] | 9.19E-47 | 7.72E-46 |
| DukeIndex | Proinsulin_pMN | 22493 | 0.049955 | [0.04 0.06] | 6.67E-14 | 9.65E-14 |
| DukeIndex | Hyper_Insulin_p | 22493 | 0.048357 | [0.04 0.06] | 4.04E-13 | 5.47E-13 |
| DukeIndex | Beta_Cell_1_pN | 22493 | 0.058052 | [0.05 0.07] | 3.01E-18 | 5.75E-18 |
| DukeIndex | Beta_Cell_2_pN | 22493 | 0.042002 | [0.03 0.06] | 2.98E-10 | 3.47E-10 |
| DukeIndex | Total_GRS_biocl | 22493 | 0.06865 | [0.06 0.08] | 6.70E-25 | 2.17E-24 |
| SIS | Cholesterol_pM | 22493 | -0.08276 | [-0.1 -0.07 | 1.80E-35 | 9.47E-35 |
| SIS | Lipodystrophy_1 | 22493 | 0.126633 | [0.11 0.14] | 5.15E-81 | 2.16E-79 |
| SIS | Lipodystrophy_2 | 22493 | 0.043889 | [0.03 0.06] | 4.60E-11 | 5.68E-11 |
| SIS | Liver-Lipid_pMN | 22493 | -0.00691 | [-0.02 0.0 | 0.30021 | 0.307532382 |
| SIS | ASAT_Pos_pMN | 22493 | 0.07461 | [0.06 0.09] | 4.00E-29 | 1.68E-28 |
| SIS | ALP_Neg_pMM | 22493 | 0.064345 | [0.05 0.08] | 4.59E-22 | 1.28E-21 |
| SIS | SHBG-LpA_pMN | 22493 | 0.061643 | [0.05 0.07] | 2.23E-20 | 4.93E-20 |
| SIS | Obesity_pMMS_ | 22493 | 0.066861 | [0.05 0.08] | 1.06E-23 | 3.19E-23 |
| SIS | VAT_Neg_pMM | 22493 | 0.096137 | [0.08 0.11] | 2.61E-47 | 2.74E-46 |
| SIS | Proinsulin_pMN | 22493 | 0.051438 | [0.04 0.06] | 1.19E-14 | 1.79E-14 |
| SIS | Hyper_Insulin_p | 22493 | 0.048975 | [0.04 0.06] | 2.02E-13 | 2.83E-13 |
| SIS | Beta_Cell_1_pN | 22493 | 0.060366 | [0.05 0.07] | 1.32E-19 | 2.64E-19 |
| SIS | Beta_Cell_2_pN | 22493 | 0.043893 | [0.03 0.06] | 4.58E-11 | 5.68E-11 |
| SIS | Total_GRS_biocl | 22493 | 0.070104 | [0.06 0.08] | 6.73E-26 | 2.36E-25 |

**d) Spearman Correlation with pMMSs Derived from Proteomics Elastic Net Models**

| Riskscore | pMMS | Sample_Si | r | CI95% | p | fdr_p_adjusted |
|---|---|---|---|---|---|---|
| CACS_Tot | Cholesterol_pMMS_ | 3919 | -0.01435 | [-0.05 0.0 | 0.36981 | 0.403429392 |
| CACS_Tot | Lipodystrophy_1_pN | 3919 | 0.10456 | [0.07 0.14] | 5.70E-11 | 4.56E-10 |
| CACS_Tot | Lipodystrophy_2_pN | 3919 | 0.062904 | [0.03 0.09] | 8.34E-05 | 0.000333484 |
| CACS_Tot | Liver-Lipid_pMMS_p | 3919 | -0.03857 | [-0.07 -0.0 | 0.015902 | 0.023852505 |
| CACS_Tot | ASAT_Pos_pMMS_p | 3919 | 0.059781 | [0.03 0.09] | 0.000185 | 0.00055463 |
| CACS_Tot | ALP_Neg_pMMS_pr | 3919 | 0.023755 | [-0.01 0.0 | 0.137664 | 0.157330622 |
| CACS_Tot | Proinsulin_pMMS_p | 3919 | 0.04068 | [0.01 0.07] | 0.010992 | 0.018843451 |
| CACS_Tot | Beta_Cell_2_pMMS_ | 3919 | -0.06165 | [-0.09 -0.0 | 0.000115 | 0.000395659 |
| DukeIndex | Cholesterol_pMMS_ | 3956 | -0.00941 | [-0.04 0.0 | 0.554738 | 0.578856825 |
| DukeIndex | Lipodystrophy_1_pN | 3956 | 0.112093 | [0.08 0.14] | 1.67E-12 | 2.00E-11 |
| DukeIndex | Lipodystrophy_2_pN | 3956 | 0.068945 | [0.04 0.1 ] | 1.47E-05 | 7.04E-05 |
| DukeIndex | Liver-Lipid_pMMS_p | 3956 | -0.04265 | [-0.07 -0.0 | 0.007382 | 0.013629207 |
| DukeIndex | ASAT_Pos_pMMS_p | 3956 | 0.049889 | [0.02 0.08] | 0.001724 | 0.00352889 |
| DukeIndex | ALP_Neg_pMMS_pr | 3956 | 0.029135 | [-0. 0.06] | 0.067325 | 0.080790287 |
| DukeIndex | Proinsulin_pMMS_p | 3956 | 0.032819 | [0. 0.06] | 0.039304 | 0.052405261 |
| DukeIndex | Beta_Cell_2_pMMS_ | 3956 | -0.05653 | [-0.09 -0.0 | 0.000383 | 0.000935935 |
| SIS | Cholesterol_pMMS_ | 3956 | -0.00749 | [-0.04 0.0 | 0.638364 | 0.63836395 |
| SIS | Lipodystrophy_1_pN | 3956 | 0.114924 | [0.08 0.15] | 4.52E-13 | 1.08E-11 |
| SIS | Lipodystrophy_2_pN | 3956 | 0.073843 | [0.04 0.1 ] | 3.44E-06 | 2.07E-05 |
| SIS | Liver-Lipid_pMMS_p | 3956 | -0.03911 | [-0.07 -0.0 | 0.014027 | 0.022443291 |
| SIS | ASAT_Pos_pMMS_p | 3956 | 0.049781 | [0.02 0.08] | 0.001764 | 0.00352889 |
| SIS | ALP_Neg_pMMS_pr | 3956 | 0.030466 | [-0. 0.06] | 0.055731 | 0.070397653 |
| SIS | Proinsulin_pMMS_p | 3956 | 0.037566 | [0.01 0.07] | 0.01831 | 0.025850105 |
| SIS | Beta_Cell_2_pMMS_ | 3956 | -0.05645 | [-0.09 -0.0 | 0.00039 | 0.000935935 |

**Table S12: Associations between pPSs, pMMSs and imaging risk scores in SCAPIS**

**e) Spearman Correlation with pMMSs Derived from Metabolomics Elastic Net Models**

| Riskscore | pMMS | Sample_Size | r | CI95% | p | fdr_p_adjusted |
|---|---|---|---|---|---|---|
| CACS_Tot | Cholesterol_pMMS_metabolomics | 4645 | -0.030794113 | [-0.06 -0. ] | 0.036085031 | 0.079934091 |
| CACS_Tot | Lipodystrophy_1_pMMS_metabolomics | 4645 | 0.089022763 | [0.06 0.12] | 1.28E-09 | 5.12E-09 |
| CACS_Tot | Liver-Lipid_pMMS_metabolomics | 4645 | -0.004542946 | [-0.03 0.02] | 0.757215789 | 0.826053588 |
| CACS_Tot | ALP_Neg_pMMS_metabolomics | 4645 | -0.001105346 | [-0.03 0.03] | 0.940042579 | 0.940042579 |
| DukeIndex | Cholesterol_pMMS_metabolomics | 4694 | -0.033091576 | [-0.06 -0. ] | 0.023554975 | 0.070664926 |
| DukeIndex | Lipodystrophy_1_pMMS_metabolomics | 4694 | 0.105610586 | [0.08 0.13] | 4.33E-13 | 4.31E-12 |
| DukeIndex | Liver-Lipid_pMMS_metabolomics | 4694 | -0.017041191 | [-0.05 0.01] | 0.243687551 | 0.365531326 |
| DukeIndex | ALP_Neg_pMMS_metabolomics | 4694 | 0.008645146 | [-0.02 0.04] | 0.554253466 | 0.665104159 |
| SIS | Cholesterol_pMMS_metabolomics | 4694 | -0.030020762 | [-0.06 -0. ] | 0.039967046 | 0.079934091 |
| SIS | Lipodystrophy_1_pMMS_metabolomics | 4694 | 0.104613626 | [0.08 0.13] | 7.18E-13 | 4.31E-12 |
| SIS | Liver-Lipid_pMMS_metabolomics | 4694 | -0.019671633 | [-0.05 0.01] | 0.178365384 | 0.305769229 |
| SIS | ALP_Neg_pMMS_metabolomics | 4694 | 0.009213318 | [-0.02 0.04] | 0.528519497 | 0.665104159 |

**f) Spearman Correlation with pMMSs Derived from Merged-omics Elastic Net Models**

| Riskscore | pMMS | Sample_Size | r | CI95% | p | fdr_p_adjusted |
|---|---|---|---|---|---|---|
| CACS_Tot | Cholesterol_pMMS_merged_omics | 3076 | -0.058394026 | [-0.09 -0.02] | 0.001221768 | 0.002199182 |
| CACS_Tot | Lipodystrophy_1_pMMS_merged_omics | 3076 | 0.101165935 | [0.07 0.14] | 2.00E-08 | 9.00E-08 |
| CACS_Tot | Lipodystrophy_2_pMMS_merged_omics | 3076 | 0.06992866 | [0.03 0.11] | 0.000107084 | 0.000240939 |
| CACS_Tot | Liver-Lipid_pMMS_merged_omics | 3076 | -0.01603235 | [-0.05 0.02] | 0.375003455 | 0.405003732 |
| CACS_Tot | ASAT_Pos_pMMS_merged_omics | 3076 | 0.124582064 | [0.09 0.16] | 4.51E-12 | 1.22E-10 |
| CACS_Tot | ALP_Neg_pMMS_merged_omics | 3076 | 0.031423142 | [-0. 0.07] | 0.082017564 | 0.105451153 |
| CACS_Tot | Proinsulin_pMMS_merged_omics | 3076 | -0.041943806 | [-0.08 -0.01] | 0.020243328 | 0.032151168 |
| CACS_Tot | Beta_Cell_2_pMMS_merged_omics | 3076 | 0.019202467 | [-0.02 0.05] | 0.287969603 | 0.323965803 |
| CACS_Tot | Total_GRS_merged_omics | 3076 | 0.090018306 | [0.05 0.13] | 6.01E-07 | 2.32E-06 |
| DukeIndex | Cholesterol_pMMS_merged_omics | 3106 | -0.062874268 | [-0.1 -0.03] | 0.000466392 | 0.000968661 |
| DukeIndex | Lipodystrophy_1_pMMS_merged_omics | 3106 | 0.10808727 | [0.07 0.14] | 1.67E-09 | 1.12E-08 |
| DukeIndex | Lipodystrophy_2_pMMS_merged_omics | 3106 | 0.075623631 | [0.04 0.11] | 2.54E-05 | 6.25E-05 |
| DukeIndex | Liver-Lipid_pMMS_merged_omics | 3106 | -0.01272761 | [-0.05 0.02] | 0.47913169 | 0.497559832 |
| DukeIndex | ASAT_Pos_pMMS_merged_omics | 3106 | 0.119863494 | [0.08 0.15] | 2.25E-11 | 2.03E-10 |
| DukeIndex | ALP_Neg_pMMS_merged_omics | 3106 | 0.037823016 | [0. 0.07] | 0.035399384 | 0.047789169 |
| DukeIndex | Proinsulin_pMMS_merged_omics | 3106 | -0.041997292 | [-0.08 -0.01] | 0.019484313 | 0.032151168 |
| DukeIndex | Beta_Cell_2_pMMS_merged_omics | 3106 | 0.024692097 | [-0.01 0.06] | 0.169714724 | 0.202967626 |
| DukeIndex | Total_GRS_merged_omics | 3106 | 0.082336514 | [0.05 0.12] | 4.52E-06 | 1.36E-05 |
| SIS | Cholesterol_pMMS_merged_omics | 3106 | -0.059880782 | [-0.09 -0.02] | 0.000860888 | 0.001660285 |
| SIS | Lipodystrophy_1_pMMS_merged_omics | 3106 | 0.106993194 | [0.07 0.14] | 2.43E-09 | 1.31E-08 |
| SIS | Lipodystrophy_2_pMMS_merged_omics | 3106 | 0.08162804 | [0.05 0.12] | 5.46E-06 | 1.47E-05 |
| SIS | Liver-Lipid_pMMS_merged_omics | 3106 | -0.010992475 | [-0.05 0.02] | 0.541057978 | 0.541057978 |
| SIS | ASAT_Pos_pMMS_merged_omics | 3106 | 0.121310397 | [0.09 0.16] | 1.29E-11 | 1.74E-10 |
| SIS | ALP_Neg_pMMS_merged_omics | 3106 | 0.040456038 | [0.01 0.08] | 0.024428139 | 0.036642208 |
| SIS | Proinsulin_pMMS_merged_omics | 3106 | -0.039541524 | [-0.07 -0. ] | 0.027848166 | 0.039573709 |
| SIS | Beta_Cell_2_pMMS_merged_omics | 3106 | 0.024509248 | [-0.01 0.06] | 0.172898348 | 0.202967626 |
| SIS | Total_GRS_merged_omics | 3106 | 0.086374539 | [0.05 0.12] | 1.50E-06 | 5.06E-06 |

**Table S13: pPS associations with Categorical Coronary Artery Calcium Scores (CACS) in SCAPIS**

| CACS | PPS | Sample_Size | coefficient | CI 2.5% | CI 97.5% | p | fdr_p_adjusted |
|---|---|---|---|---|---|---|---|
| CACS_1 | Cholesterol_pPS | 26125 | -0.081763587 | -0.110363511 | -0.053163663 | 2.12E-08 | 6.43E-07 |
| CACS_2 | Cholesterol_pPS | 26125 | -0.095146075 | -0.145380581 | -0.044911569 | 0.000205715 | 0.001322451 |
| CACS_3 | Cholesterol_pPS | 26125 | -0.098104502 | -0.159020968 | -0.037188036 | 0.0015979 | 0.006536864 |
| CACS_1 | Lipodystrophy_1_pPS | 26125 | 0.025264711 | -0.003104603 | 0.053634025 | 0.080899938 | 0.145619889 |
| CACS_2 | Lipodystrophy_1_pPS | 26125 | 0.068061947 | 0.018232497 | 0.117891397 | 0.007427894 | 0.025711939 |
| CACS_3 | Lipodystrophy_1_pPS | 26125 | 0.074567074 | 0.014141797 | 0.134992352 | 0.015579375 | 0.041239523 |
| CACS_1 | Lipodystrophy_2_pPS | 26125 | 0.013652046 | -0.014799446 | 0.042103538 | 0.346967503 | 0.487923051 |
| CACS_2 | Lipodystrophy_2_pPS | 26125 | -0.029615838 | -0.07958963 | 0.020357953 | 0.24541583 | 0.368123746 |
| CACS_3 | Lipodystrophy_2_pPS | 26125 | 0.017357966 | -0.043242347 | 0.077958279 | 0.574512267 | 0.698731136 |
| CACS_1 | Liver-Lipid_pPS | 26125 | -0.029187087 | -0.057628454 | -0.000745721 | 0.044288554 | 0.094904045 |
| CACS_2 | Liver-Lipid_pPS | 26125 | -0.104281281 | -0.154237288 | -0.054325275 | 4.30E-05 | 0.000483528 |
| CACS_3 | Liver-Lipid_pPS | 26125 | -0.002535739 | -0.063114485 | 0.058043007 | 0.934611508 | 0.985562063 |
| CACS_1 | ASAT_Pos_pPS | 26125 | 0.023848419 | -0.004731072 | 0.052427909 | 0.101938359 | 0.176431775 |
| CACS_2 | ASAT_Pos_pPS | 26125 | -0.017297797 | -0.067496412 | 0.032900819 | 0.499421506 | 0.624276882 |
| CACS_3 | ASAT_Pos_pPS | 26125 | 0.000543193 | -0.060329751 | 0.061416136 | 0.986045585 | 0.986045585 |
| CACS_1 | ALP_Neg_pPS | 26125 | 0.031923035 | 0.003353395 | 0.060492675 | 0.028524452 | 0.065411713 |
| CACS_2 | ALP_Neg_pPS | 26125 | 0.067482903 | 0.01730159 | 0.117664217 | 0.008397754 | 0.026992781 |
| CACS_3 | ALP_Neg_pPS | 26125 | 0.039650396 | -0.021201566 | 0.100502359 | 0.201561421 | 0.323937998 |
| CACS_1 | SHBG-LpA_pPS | 26125 | 0.057039345 | 0.028495408 | 0.085583282 | 9.00E-05 | 0.000809719 |
| CACS_2 | SHBG-LpA_pPS | 26125 | 0.055829434 | 0.005693267 | 0.1059656 | 0.029071873 | 0.065411713 |
| CACS_3 | SHBG-LpA_pPS | 26125 | 0.061330821 | 0.000533605 | 0.122128036 | 0.048022992 | 0.098228847 |
| CACS_1 | Obesity_pPS | 26125 | 0.018254737 | -0.010295608 | 0.046805081 | 0.210131492 | 0.326066109 |
| CACS_2 | Obesity_pPS | 26125 | 0.086466543 | 0.036319122 | 0.136613965 | 0.000726926 | 0.003634631 |
| CACS_3 | Obesity_pPS | 26125 | 0.116254417 | 0.055443554 | 0.17706528 | 0.000179268 | 0.001322451 |
| CACS_1 | VAT_Neg_pPS | 26125 | 0.01157459 | -0.01690953 | 0.04005871 | 0.425764443 | 0.577955707 |
| CACS_2 | VAT_Neg_pPS | 26125 | 0.064217603 | 0.014186501 | 0.114248704 | 0.011881007 | 0.033415331 |
| CACS_3 | VAT_Neg_pPS | 26125 | 0.016030553 | -0.044639255 | 0.076700362 | 0.604533929 | 0.715895442 |
| CACS_1 | Bilirubin_pPS | 26125 | 0.000613009 | -0.02575906 | 0.026985078 | 0.963660683 | 0.985562063 |
| CACS_2 | Bilirubin_pPS | 26125 | 0.041509065 | -0.004812312 | 0.087830442 | 0.079027515 | 0.145619889 |
| CACS_3 | Bilirubin_pPS | 26125 | 0.005174855 | -0.050996386 | 0.061346097 | 0.856704281 | 0.963792316 |
| CACS_1 | Proinsulin_pPS | 26125 | 0.026861212 | -0.001760889 | 0.055483313 | 0.065857677 | 0.128851976 |
| CACS_2 | Proinsulin_pPS | 26125 | 0.040439046 | -0.009834413 | 0.090712505 | 0.11489232 | 0.191487199 |
| CACS_3 | Proinsulin_pPS | 26125 | 0.092374228 | 0.031410526 | 0.15333793 | 0.002981285 | 0.011179819 |
| CACS_1 | Hyper_Insulin_pPS | 26125 | -0.010354627 | -0.03883216 | 0.018122907 | 0.47604435 | 0.612057021 |
| CACS_2 | Hyper_Insulin_pPS | 26125 | 0.019849739 | -0.030169793 | 0.069869272 | 0.436677645 | 0.577955707 |
| CACS_3 | Hyper_Insulin_pPS | 26125 | 0.030360225 | -0.030295555 | 0.091016006 | 0.326566171 | 0.474047668 |
| CACS_1 | Beta_Cell_1_pPS | 26125 | 0.065025299 | 0.03659888 | 0.093451718 | 7.37E-06 | 0.000110551 |
| CACS_2 | Beta_Cell_1_pPS | 26125 | 0.084413107 | 0.034483355 | 0.134342859 | 0.000921816 | 0.004148171 |
| CACS_3 | Beta_Cell_1_pPS | 26125 | 0.17149075 | 0.110943841 | 0.232037658 | 2.86E-08 | 6.43E-07 |
| CACS_1 | Beta_Cell_2_pPS | 26125 | 0.002070761 | -0.026277481 | 0.030419003 | 0.88615182 | 0.972605656 |
| CACS_2 | Beta_Cell_2_pPS | 26125 | -0.010272408 | -0.060064845 | 0.039520029 | 0.685945223 | 0.791475257 |
| CACS_3 | Beta_Cell_2_pPS | 26125 | 0.002085276 | -0.058295119 | 0.06246567 | 0.946031535 | 0.985562063 |
| CACS_1 | Total_GRS | 26125 | 0.037601549 | 0.009183867 | 0.066019232 | 0.009505911 | 0.028517733 |
| CACS_2 | Total_GRS | 26125 | 0.05751255 | 0.007598144 | 0.107426957 | 0.02392763 | 0.059819074 |
| CACS_3 | Total_GRS | 26125 | 0.105448024 | 0.044919725 | 0.165976324 | 0.000639574 | 0.003597604 |

**Table S14: Associations between pPSs, pMMSs and imaging-derived phenotypes in SCAPIS**

**a) Logistic Regression with Genetic pPSs**

| PPS | Imaging_Variables | Sample_Size | Cases (Positive | coefficient | CI 2.5% | CI 97.5% | p | fdr_p_adjusted |
|---|---|---|---|---|---|---|---|---|
| Cholesterol_pPS | Above50_Any | 24938 | 1353 | -0.048675288 | -0.105146938 | 0.007796362 | 0.091147526 | 0.2474132 |
| Lipodystrophy_1_pPS | Above50_Any | 24938 | 1353 | 0.073379561 | 0.016620432 | 0.13013869 | 0.011280467 | 0.065802722 |
| Lipodystrophy_2_pPS | Above50_Any | 24938 | 1353 | 0.006509756 | -0.049557674 | 0.062577187 | 0.819985772 | 0.946137429 |
| Liver-Lipid_pPS | Above50_Any | 24938 | 1353 | -0.028190392 | -0.0843388 | 0.027958016 | 0.325096952 | 0.566322466 |
| ASAT_Pos_pPS | Above50_Any | 24938 | 1353 | -0.012221804 | -0.068206055 | 0.043762447 | 0.668741685 | 0.861544335 |
| ALP_Neg_pPS | Above50_Any | 24938 | 1353 | 0.075862701 | 0.019711309 | 0.132014093 | 0.008097262 | 0.056680831 |
| SHBG-LpA_pPS | Above50_Any | 24938 | 1353 | 0.037453791 | -0.018890006 | 0.093797587 | 0.192622551 | 0.417128379 |
| Obesity_pPS | Above50_Any | 24938 | 1353 | 0.066721678 | 0.010532848 | 0.122910507 | 0.019945686 | 0.085443271 |
| VAT_Neg_pPS | Above50_Any | 24938 | 1353 | 0.028036917 | -0.028396902 | 0.084470735 | 0.33019022 | 0.566322466 |
| Bilirubin_pPS | Above50_Any | 24938 | 1353 | 0.02641125 | -0.034242818 | 0.087065318 | 0.393410858 | 0.607472648 |
| Proinsulin_pPS | Above50_Any | 24938 | 1353 | 0.066306567 | 0.010457456 | 0.122155677 | 0.01996755 | 0.085443271 |
| Hyper_Insulin_pPS | Above50_Any | 24938 | 1353 | 0.021062351 | -0.035345973 | 0.077470675 | 0.464270758 | 0.686597599 |
| Beta_Cell_1_pPS | Above50_Any | 24938 | 1353 | 0.121791177 | 0.065192963 | 0.178389392 | 2.47E-05 | 0.000410702 |
| Beta_Cell_2_pPS | Above50_Any | 24938 | 1353 | -0.003541433 | -0.059821575 | 0.052738708 | 0.901845055 | 0.977366923 |
| Total_GRS | Above50_Any | 24938 | 1353 | 0.067065684 | 0.01043688 | 0.123694489 | 0.020276486 | 0.085443271 |
| Cholesterol_pPS | AortaGt4cm | 26533 | 945 | 0.042919651 | -0.022648208 | 0.108487509 | 0.199505931 | 0.418962454 |
| Lipodystrophy_1_pPS | AortaGt4cm | 26533 | 945 | -0.041931588 | -0.108152629 | 0.024289453 | 0.214582933 | 0.425117132 |
| Lipodystrophy_2_pPS | AortaGt4cm | 26533 | 945 | -0.032076801 | -0.097993445 | 0.033839843 | 0.340199636 | 0.566999393 |
| Liver-Lipid_pPS | AortaGt4cm | 26533 | 945 | -0.030517786 | -0.096285697 | 0.035250126 | 0.363103543 | 0.586551877 |
| ASAT_Pos_pPS | AortaGt4cm | 26533 | 945 | -0.076735414 | -0.142399668 | -0.011071159 | 0.021997048 | 0.088834231 |
| ALP_Neg_pPS | AortaGt4cm | 26533 | 945 | 0.025318382 | -0.04038345 | 0.091020215 | 0.45008248 | 0.675123719 |
| SHBG-LpA_pPS | AortaGt4cm | 26533 | 945 | -0.044006354 | -0.109906711 | 0.021894003 | 0.190599629 | 0.417128379 |
| Obesity_pPS | AortaGt4cm | 26533 | 945 | 0.010845775 | -0.054822094 | 0.076513644 | 0.746157797 | 0.903131698 |
| VAT_Neg_pPS | AortaGt4cm | 26533 | 945 | -0.078233691 | -0.14432791 | -0.012139473 | 0.020343636 | 0.085443271 |
| Bilirubin_pPS | AortaGt4cm | 26533 | 945 | 0.009864887 | -0.061107758 | 0.080837532 | 0.785294164 | 0.936998718 |
| Proinsulin_pPS | AortaGt4cm | 26533 | 945 | 0.023206374 | -0.042197116 | 0.088609863 | 0.486784828 | 0.691668079 |
| Hyper_Insulin_pPS | AortaGt4cm | 26533 | 945 | 0.014520134 | -0.051520866 | 0.080561134 | 0.666520455 | 0.861544335 |
| Beta_Cell_1_pPS | AortaGt4cm | 26533 | 945 | -0.054442202 | -0.120563379 | 0.011678975 | 0.106576069 | 0.266440172 |
| Beta_Cell_2_pPS | AortaGt4cm | 26533 | 945 | -0.006557896 | -0.072563476 | 0.059447684 | 0.84560464 | 0.965092252 |
| Total_GRS | AortaGt4cm | 26533 | 945 | -0.044475516 | -0.110644323 | 0.021693291 | 0.187706487 | 0.417128379 |
| Cholesterol_pPS | AorticValveCalc | 26468 | 1817 | 0.014988676 | -0.033185848 | 0.063163199 | 0.541988251 | 0.739074888 |
| Lipodystrophy_1_pPS | AorticValveCalc | 26468 | 1817 | 0.024069355 | -0.024802076 | 0.072940786 | 0.334399932 | 0.566322466 |
| Lipodystrophy_2_pPS | AorticValveCalc | 26468 | 1817 | 0.009991211 | -0.038510773 | 0.058493195 | 0.68640054 | 0.868338032 |
| Liver-Lipid_pPS | AorticValveCalc | 26468 | 1817 | 0.032683407 | -0.015850243 | 0.081217056 | 0.186877101 | 0.417128379 |
| ASAT_Pos_pPS | AorticValveCalc | 26468 | 1817 | 0.007916318 | -0.040438815 | 0.056271451 | 0.748309122 | 0.903131698 |
| ALP_Neg_pPS | AorticValveCalc | 26468 | 1817 | 0.024303771 | -0.024112707 | 0.072720249 | 0.325189636 | 0.566322466 |
| SHBG-LpA_pPS | AorticValveCalc | 26468 | 1817 | 0.025400918 | -0.023114284 | 0.073916121 | 0.304811075 | 0.551813153 |
| Obesity_pPS | AorticValveCalc | 26468 | 1817 | 0.032006863 | -0.016363481 | 0.080377207 | 0.19465991 | 0.417128379 |
| VAT_Neg_pPS | AorticValveCalc | 26468 | 1817 | 0.040282961 | -0.008295754 | 0.088861676 | 0.104106687 | 0.266440172 |
| Bilirubin_pPS | AorticValveCalc | 26468 | 1817 | -0.009605586 | -0.062062465 | 0.042851293 | 0.719672343 | 0.895443306 |
| Proinsulin_pPS | AorticValveCalc | 26468 | 1817 | 0.005745884 | -0.042433984 | 0.053925752 | 0.815184192 | 0.946137429 |
| Hyper_Insulin_pPS | AorticValveCalc | 26468 | 1817 | -0.003415997 | -0.052089913 | 0.045257918 | 0.890593886 | 0.977366923 |
| Beta_Cell_1_pPS | AorticValveCalc | 26468 | 1817 | 0.050788306 | 0.002111348 | 0.099465263 | 0.040857144 | 0.147931037 |
| Beta_Cell_2_pPS | AorticValveCalc | 26468 | 1817 | -0.002442124 | -0.051024664 | 0.046140417 | 0.921517384 | 0.977366923 |
| Total_GRS | AorticValveCalc | 26468 | 1817 | 0.020486278 | -0.028185435 | 0.06915799 | 0.409392552 | 0.622988666 |
| Cholesterol_pPS | Atherosclerosis_Any | 24938 | 10388 | -0.082800952 | -0.10978651 | -0.055815394 | 1.81E-09 | 6.34E-08 |
| Lipodystrophy_1_pPS | Atherosclerosis_Any | 24938 | 10388 | 0.033398308 | 0.006218445 | 0.06057817 | 0.016023424 | 0.084122976 |
| Lipodystrophy_2_pPS | Atherosclerosis_Any | 24938 | 10388 | -0.001897147 | -0.028960319 | 0.025166025 | 0.890718715 | 0.977366923 |
| Liver-Lipid_pPS | Atherosclerosis_Any | 24938 | 10388 | -0.059639114 | -0.086752493 | -0.032525735 | 1.62E-05 | 0.000341018 |
| ASAT_Pos_pPS | Atherosclerosis_Any | 24938 | 10388 | 0.021867963 | -0.005054972 | 0.048790957 | 0.111392059 | 0.272003864 |
| ALP_Neg_pPS | Atherosclerosis_Any | 24938 | 10388 | 0.032360654 | 0.005447336 | 0.059273972 | 0.018439794 | 0.085443271 |
| SHBG-LpA_pPS | Atherosclerosis_Any | 24938 | 10388 | 0.050894399 | 0.023892408 | 0.07789639 | 0.000220566 | 0.002315945 |
| Obesity_pPS | Atherosclerosis_Any | 24938 | 10388 | 0.029037146 | 0.002063828 | 0.056010465 | 0.034864555 | 0.130742082 |
| VAT_Neg_pPS | Atherosclerosis_Any | 24938 | 10388 | 0.02480988 | -0.002215788 | 0.051835547 | 0.07197599 | 0.218573345 |
| Bilirubin_pPS | Atherosclerosis_Any | 24938 | 10388 | 0.018711688 | -0.010476379 | 0.047899755 | 0.208941849 | 0.425117132 |
| Proinsulin_pPS | Atherosclerosis_Any | 24938 | 10388 | 0.037431163 | 0.010465087 | 0.064397239 | 0.006516483 | 0.048873623 |
| Hyper_Insulin_pPS | Atherosclerosis_Any | 24938 | 10388 | 0.001700351 | -0.025334746 | 0.028735447 | 0.901893013 | 0.977366923 |
| Beta_Cell_1_pPS | Atherosclerosis_Any | 24938 | 10388 | 0.092510066 | 0.065405517 | 0.119614614 | 2.24E-11 | 1.18E-09 |
| Beta_Cell_2_pPS | Atherosclerosis_Any | 24938 | 10388 | 0.007073905 | -0.020010124 | 0.034157934 | 0.608713638 | 0.809049772 |
| Total_GRS | Atherosclerosis_Any | 24938 | 10388 | 0.045903539 | 0.018755112 | 0.073051966 | 0.000919788 | 0.008048148 |
| Cholesterol_pPS | Bicuspid | 24839 | 130 | 0.037161933 | -0.135323387 | 0.209647253 | 0.6728251 | 0.861544335 |
| Lipodystrophy_1_pPS | Bicuspid | 24839 | 130 | -0.003587745 | -0.177794661 | 0.170619172 | 0.967802185 | 0.996472143 |
| Lipodystrophy_2_pPS | Bicuspid | 24839 | 130 | 0.002616402 | -0.171085351 | 0.176318155 | 0.976448133 | 0.996472143 |
| Liver-Lipid_pPS | Bicuspid | 24839 | 130 | -0.079275765 | -0.252795967 | 0.094244436 | 0.370549721 | 0.58951092 |
| ASAT_Pos_pPS | Bicuspid | 24839 | 130 | 0.00038956 | -0.172293457 | 0.173072576 | 0.996472143 | 0.996472143 |
| ALP_Neg_pPS | Bicuspid | 24839 | 130 | 0.158793669 | -0.01407493 | 0.331662268 | 0.071800014 | 0.218573345 |
| SHBG-LpA_pPS | Bicuspid | 24839 | 130 | 0.07577956 | -0.097550831 | 0.249109952 | 0.391505265 | 0.607472648 |
| Obesity_pPS | Bicuspid | 24839 | 130 | 0.096326109 | -0.076838836 | 0.269491054 | 0.275596362 | 0.526138509 |
| VAT_Neg_pPS | Bicuspid | 24839 | 130 | 0.08257628 | -0.091342277 | 0.256494838 | 0.352066686 | 0.577609407 |
| Bilirubin_pPS | Bicuspid | 24839 | 130 | -0.170792097 | -0.364062906 | 0.022478712 | 0.083272349 | 0.236313422 |
| Proinsulin_pPS | Bicuspid | 24839 | 130 | 0.05478674 | -0.117906951 | 0.22748043 | 0.534076813 | 0.737869282 |
| Hyper_Insulin_pPS | Bicuspid | 24839 | 130 | 0.14956603 | -0.024357322 | 0.323489381 | 0.091896332 | 0.2474132 |
| Beta_Cell_1_pPS | Bicuspid | 24839 | 130 | -0.062971804 | -0.236612506 | 0.110668898 | 0.477212972 | 0.691668079 |
| Beta_Cell_2_pPS | Bicuspid | 24839 | 130 | -0.002818954 | -0.176531624 | 0.170893715 | 0.974627021 | 0.996472143 |
| Total_GRS | Bicuspid | 24839 | 130 | 0.159332776 | -0.014767086 | 0.333432638 | 0.072857782 | 0.218573345 |
| Cholesterol_pPS | Carotidplaque_Any | 26903 | 14665 | -0.043668653 | -0.068314829 | -0.019022477 | 0.000515235 | 0.004918148 |

**b) Logistic Regression with pMMSs Derived from Biochemistry Elastic Net Models**

| pMMS | Imaging_Variables | Sample_Size | Cases (Positive | coefficient | CI 2.5% | CI 97.5% | p | fdr_p_adjusted |
|---|---|---|---|---|---|---|---|---|
| Cholesterol_pMMS_biochemistry | Above50_Any | 22493 | 1181 | -0.687586553 | -0.944148465 | -0.43102464 | 1.50E-07 | 3.19E-07 |
| Lipodystrophy_1_pMMS_biochemistr | Above50_Any | 22493 | 1181 | 4.227020735 | 3.396002657 | 5.058038812 | 2.07E-23 | 1.35E-22 |
| Lipodystrophy_2_pMMS_biochemistr | Above50_Any | 22493 | 1181 | 1.03436217 | 0.409899073 | 1.658825266 | 0.001168379 | 0.002008793 |
| Liver-Lipid_pMMS_biochemistry | Above50_Any | 22493 | 1181 | 0.222504107 | -0.147908863 | 0.592917076 | 0.239061949 | 0.292850888 |
| ASAT_Pos_pMMS_biochemistry | Above50_Any | 22493 | 1181 | 2.859253172 | 1.707656895 | 4.010849449 | 1.14E-06 | 2.37E-06 |
| ALP_Neg_pMMS_biochemistry | Above50_Any | 22493 | 1181 | 1.646935809 | 1.081697217 | 2.212174402 | 1.12E-08 | 2.56E-08 |
| SHBG-LpA_pMMS_biochemistry | Above50_Any | 22493 | 1181 | 3.392703943 | 2.635301637 | 4.150106248 | 1.64E-18 | 7.32E-18 |
| Obesity_pMMS_biochemistry | Above50_Any | 22493 | 1181 | 2.808410141 | 2.101860783 | 3.514959499 | 6.67E-15 | 2.11E-14 |
| VAT_Neg_pMMS_biochemistry | Above50_Any | 22493 | 1181 | 2.781851625 | 2.130784293 | 3.432918957 | 5.55E-17 | 2.17E-16 |
| Proinsulin_pMMS_biochemistry | Above50_Any | 22493 | 1181 | 1.120156724 | 0.469553913 | 1.770759536 | 0.000739468 | 0.001341997 |
| Hyper_Insulin_pMMS_biochemistry | Above50_Any | 22493 | 1181 | 1.980070026 | 1.134636916 | 2.825503136 | 4.42E-06 | 9.03E-06 |
| Beta_Cell_1_pMMS_biochemistry | Above50_Any | 22493 | 1181 | 1.512889252 | 0.837167511 | 2.188610994 | 1.14E-05 | 2.29E-05 |
| Beta_Cell_2_pMMS_biochemistry | Above50_Any | 22493 | 1181 | 0.343344593 | 0.052772957 | 0.63391623 | 0.020562191 | 0.031485855 |
| Total_GRS_biochemistry | Above50_Any | 22493 | 1181 | 0.938032932 | 0.605360948 | 1.270704915 | 3.27E-08 | 7.11E-08 |
| Cholesterol_pMMS_biochemistry | AortaGt4cm | 23934 | 868 | 0.184614987 | -0.10406743 | 0.473297405 | 0.2100549 | 0.26391513 |
| Lipodystrophy_1_pMMS_biochemistr | AortaGt4cm | 23934 | 868 | -0.813183689 | -1.754561448 | 0.128194071 | 0.090443701 | 0.123103926 |
| Lipodystrophy_2_pMMS_biochemistr | AortaGt4cm | 23934 | 868 | -0.600234068 | -1.322117579 | 0.121649443 | 0.103169722 | 0.138501818 |
| Liver-Lipid_pMMS_biochemistry | AortaGt4cm | 23934 | 868 | 0.250674268 | -0.175023972 | 0.676372509 | 0.248445501 | 0.300588384 |
| ASAT_Pos_pMMS_biochemistry | AortaGt4cm | 23934 | 868 | -0.978450042 | -2.298417403 | 0.34151732 | 0.146262826 | 0.19111676 |
| ALP_Neg_pMMS_biochemistry | AortaGt4cm | 23934 | 868 | 0.352929013 | -0.292239409 | 0.998097435 | 0.283645124 | 0.338990515 |
| SHBG-LpA_pMMS_biochemistry | AortaGt4cm | 23934 | 868 | 0.106742331 | -0.755927339 | 0.969412001 | 0.808380427 | 0.85184174 |
| Obesity_pMMS_biochemistry | AortaGt4cm | 23934 | 868 | 1.480147966 | 0.674847041 | 2.285448891 | 0.000315262 | 0.000582937 |
| VAT_Neg_pMMS_biochemistry | AortaGt4cm | 23934 | 868 | -0.158481369 | -0.897595411 | 0.580632673 | 0.674297792 | 0.726166853 |
| Proinsulin_pMMS_biochemistry | AortaGt4cm | 23934 | 868 | -0.083104938 | -0.829965219 | 0.663755344 | 0.827359066 | 0.857735126 |
| Hyper_Insulin_pMMS_biochemistry | AortaGt4cm | 23934 | 868 | -1.098328746 | -2.087785646 | -0.108871846 | 0.029583493 | 0.044602804 |
| Beta_Cell_1_pMMS_biochemistry | AortaGt4cm | 23934 | 868 | -0.005080395 | -0.780875272 | 0.770714482 | 0.989759366 | 0.997770323 |
| Beta_Cell_2_pMMS_biochemistry | AortaGt4cm | 23934 | 868 | -0.072086105 | -0.405228825 | 0.261056615 | 0.671492136 | 0.726166853 |
| Total_GRS_biochemistry | AortaGt4cm | 23934 | 868 | -0.130500485 | -0.515899038 | 0.254898068 | 0.506903551 | 0.584429976 |
| Cholesterol_pMMS_biochemistry | AorticValveCalc | 23796 | 1651 | -0.261974931 | -0.476932917 | -0.047016945 | 0.016909968 | 0.026728658 |
| Lipodystrophy_1_pMMS_biochemistr | AorticValveCalc | 23796 | 1651 | 2.241644837 | 1.53798277 | 2.945306904 | 4.27E-10 | 1.05E-09 |
| Lipodystrophy_2_pMMS_biochemistr | AorticValveCalc | 23796 | 1651 | 0.080954857 | -0.447460603 | 0.609370318 | 0.763969346 | 0.813793434 |
| Liver-Lipid_pMMS_biochemistry | AorticValveCalc | 23796 | 1651 | 0.034246995 | -0.281170489 | 0.349664479 | 0.831477928 | 0.857735126 |
| ASAT_Pos_pMMS_biochemistry | AorticValveCalc | 23796 | 1651 | 1.362497472 | 0.383057013 | 2.341937932 | 0.006400964 | 0.010454908 |
| ALP_Neg_pMMS_biochemistry | AorticValveCalc | 23796 | 1651 | 0.826765098 | 0.345532604 | 1.307997592 | 0.00075922 | 0.001352792 |
| SHBG-LpA_pMMS_biochemistry | AorticValveCalc | 23796 | 1651 | 4.073258585 | 3.432154654 | 4.714362516 | 1.35E-35 | 1.66E-34 |
| Obesity_pMMS_biochemistry | AorticValveCalc | 23796 | 1651 | 1.704308126 | 1.104632407 | 2.303983844 | 2.54E-08 | 5.66E-08 |
| VAT_Neg_pMMS_biochemistry | AorticValveCalc | 23796 | 1651 | 2.36069403 | 1.818603442 | 2.902784617 | 1.40E-17 | 5.71E-17 |
| Proinsulin_pMMS_biochemistry | AorticValveCalc | 23796 | 1651 | 0.146442105 | -0.409556134 | 0.702440344 | 0.60569594 | 0.682278185 |
| Hyper_Insulin_pMMS_biochemistry | AorticValveCalc | 23796 | 1651 | 0.883437029 | 0.151541405 | 1.615332652 | 0.017992084 | 0.027987686 |
| Beta_Cell_1_pMMS_biochemistry | AorticValveCalc | 23796 | 1651 | 0.765363781 | 0.193655324 | 1.337072238 | 0.00869384 | 0.013967153 |
| Beta_Cell_2_pMMS_biochemistry | AorticValveCalc | 23796 | 1651 | 0.150420126 | -0.095990716 | 0.396830969 | 0.231521351 | 0.287203701 |
| Total_GRS_biochemistry | AorticValveCalc | 23796 | 1651 | 0.411815427 | 0.130018823 | 0.693612032 | 0.004179629 | 0.006942435 |
| Cholesterol_pMMS_biochemistry | Atherosclerosis_Any | 22493 | 9253 | -0.775531331 | -0.896803894 | -0.654258769 | 4.87E-36 | 6.82E-35 |
| Lipodystrophy_1_pMMS_biochemistr | Atherosclerosis_Any | 22493 | 9253 | 3.741093141 | 3.338474789 | 4.143711494 | 4.16E-74 | 4.08E-72 |
| Lipodystrophy_2_pMMS_biochemistr | Atherosclerosis_Any | 22493 | 9253 | 0.997798367 | 0.699569628 | 1.296027106 | 5.47E-11 | 1.38E-10 |
| Liver-Lipid_pMMS_biochemistry | Atherosclerosis_Any | 22493 | 9253 | -0.189487781 | -0.365888582 | -0.01308698 | 0.035258979 | 0.052354242 |
| ASAT_Pos_pMMS_biochemistry | Atherosclerosis_Any | 22493 | 9253 | 3.196211113 | 2.635626125 | 3.756796102 | 5.41E-29 | 4.42E-28 |
| ALP_Neg_pMMS_biochemistry | Atherosclerosis_Any | 22493 | 9253 | 1.373792549 | 1.100776889 | 1.64680821 | 6.06E-23 | 3.71E-22 |
| SHBG-LpA_pMMS_biochemistry | Atherosclerosis_Any | 22493 | 9253 | 1.646713809 | 1.282243304 | 2.011184315 | 8.35E-19 | 3.89E-18 |
| Obesity_pMMS_biochemistry | Atherosclerosis_Any | 22493 | 9253 | 1.546115026 | 1.20443133 | 1.887798722 | 7.39E-19 | 3.62E-18 |
| VAT_Neg_pMMS_biochemistry | Atherosclerosis_Any | 22493 | 9253 | 2.213906776 | 1.905221181 | 2.522592371 | 6.98E-45 | 1.37E-43 |
| Proinsulin_pMMS_biochemistry | Atherosclerosis_Any | 22493 | 9253 | 1.342116468 | 1.026076584 | 1.658156351 | 8.55E-17 | 3.10E-16 |
| Hyper_Insulin_pMMS_biochemistry | Atherosclerosis_Any | 22493 | 9253 | 1.622388841 | 1.204109718 | 2.040667965 | 2.91E-14 | 8.92E-14 |
| Beta_Cell_1_pMMS_biochemistry | Atherosclerosis_Any | 22493 | 9253 | 1.559301269 | 1.23458785 | 1.884014688 | 4.87E-21 | 2.51E-20 |
| Beta_Cell_2_pMMS_biochemistry | Atherosclerosis_Any | 22493 | 9253 | 0.494497771 | 0.354930183 | 0.634065358 | 3.78E-12 | 1.13E-11 |
| Total_GRS_biochemistry | Atherosclerosis_Any | 22493 | 9253 | 0.898053638 | 0.737160166 | 1.05894711 | 7.43E-28 | 5.60E-27 |
| Cholesterol_pMMS_biochemistry | Bicuspid | 22400 | 115 | -0.010436408 | -0.788052295 | 0.76717948 | 0.979014275 | 0.997770323 |
| Lipodystrophy_1_pMMS_biochemistr | Bicuspid | 22400 | 115 | -2.322029935 | -4.873534929 | 0.229475059 | 0.074473982 | 0.102795073 |
| Lipodystrophy_2_pMMS_biochemistr | Bicuspid | 22400 | 115 | -3.344338642 | -5.354539359 | -1.334137924 | 0.001111138 | 0.001944492 |
| Liver-Lipid_pMMS_biochemistry | Bicuspid | 22400 | 115 | 0.290772137 | -0.856307509 | 1.437851782 | 0.619309346 | 0.689685409 |
| ASAT_Pos_pMMS_biochemistry | Bicuspid | 22400 | 115 | -3.860068923 | -7.512852807 | -0.20728504 | 0.038341386 | 0.056021809 |
| ALP_Neg_pMMS_biochemistry | Bicuspid | 22400 | 115 | 1.129795541 | -0.620064727 | 2.87965581 | 0.205710492 | 0.26391513 |
| SHBG-LpA_pMMS_biochemistry | Bicuspid | 22400 | 115 | 0.65422017 | -1.679752671 | 2.988193011 | 0.582741765 | 0.664054569 |
| Obesity_pMMS_biochemistry | Bicuspid | 22400 | 115 | -0.91180493 | -3.134552321 | 1.310942461 | 0.421392838 | 0.49754817 |
| VAT_Neg_pMMS_biochemistry | Bicuspid | 22400 | 115 | -3.123090231 | -5.106946518 | -1.139233945 | 0.002032254 | 0.003433809 |
| Proinsulin_pMMS_biochemistry | Bicuspid | 22400 | 115 | -0.002902517 | -2.038633965 | 2.032828931 | 0.997770323 | 0.997770323 |
| Hyper_Insulin_pMMS_biochemistry | Bicuspid | 22400 | 115 | -2.162410391 | -4.898238618 | 0.573417836 | 0.121342311 | 0.160696574 |
| Beta_Cell_1_pMMS_biochemistry | Bicuspid | 22400 | 115 | -0.459371201 | -2.558592376 | 1.639849975 | 0.667997717 | 0.726166853 |
| Beta_Cell_2_pMMS_biochemistry | Bicuspid | 22400 | 115 | -0.350865753 | -1.251821714 | 0.550090208 | 0.445294724 | 0.519510511 |
| Total_GRS_biochemistry | Bicuspid | 22400 | 115 | -1.113720835 | -2.170517862 | -0.056923807 | 0.038872275 | 0.056021809 |
| Cholesterol_pMMS_biochemistry | Carotidplaque_Any | 24186 | 13003 | -0.763747061 | -0.874402244 | -0.653091878 | 1.07E-41 | 1.75E-40 |
| Lipodystrophy_1_pMMS_biochemistr | Carotidplaque_Any | 24186 | 13003 | 2.779398188 | 2.413257707 | 3.14553867 | 4.56E-50 | 1.49E-48 |
| Lipodystrophy_2_pMMS_biochemistr | Carotidplaque_Any | 24186 | 13003 | 0.270886699 | 0.000771473 | 0.541001924 | 0.049349249 | 0.069088948 |
| Liver-Lipid_pMMS_biochemistry | Carotidplaque_Any | 24186 | 13003 | -0.103244012 | -0.264103876 | 0.057615851 | 0.20840786 | 0.26391513 |
| ASAT_Pos_pMMS_biochemistry | Carotidplaque_Any | 24186 | 13003 | 1.693805827 | 1.187530739 | 2.200080916 | 5.48E-11 | 1.38E-10 |
| ALP_Neg_pMMS_biochemistry | Carotidplaque_Any | 24186 | 13003 | 1.061427532 | 0.812756533 | 1.310098531 | 5.97E-17 | 2.25E-16 |

**Table S14: Associations between pPSs, pMMSs and imaging-derived phenotypes in SCAPIS**

**a) Logistic Regression with Genetic pPSs**

| PPS | Imaging_Variables | Sample_Size | Cases (Positive | coefficient | CI 2.5% | CI 97.5% | p | fdr_p_adjusted |
|---|---|---|---|---|---|---|---|---|
| Lipodystrophy_1_pPS | Carotidplaque_Any | 26903 | 14665 | 0.022417061 | -0.002380865 | 0.047214987 | 0.076429987 | 0.222920795 |
| Lipodystrophy_2_pPS | Carotidplaque_Any | 26903 | 14665 | 0.000232525 | -0.024564424 | 0.025029473 | 0.98533661 | 0.996472143 |
| Liver-Lipid_pPS | Carotidplaque_Any | 26903 | 14665 | -0.020887547 | -0.045660159 | 0.003885066 | 0.09841543 | 0.258340503 |
| ASAT_Pos_pPS | Carotidplaque_Any | 26903 | 14665 | 0.004419534 | -0.020192923 | 0.029031991 | 0.724882676 | 0.895443306 |
| ALP_Neg_pPS | Carotidplaque_Any | 26903 | 14665 | -0.000198058 | -0.024852437 | 0.024456321 | 0.987437703 | 0.996472143 |
| SHBG-LpA_pPS | Carotidplaque_Any | 26903 | 14665 | -0.003208244 | -0.027890406 | 0.021473918 | 0.798907999 | 0.942531909 |
| Obesity_pPS | Carotidplaque_Any | 26903 | 14665 | -0.013258226 | -0.037943837 | 0.011427386 | 0.292495187 | 0.538806923 |
| VAT_Neg_pPS | Carotidplaque_Any | 26903 | 14665 | 0.049691965 | 0.024957902 | 0.074426027 | 8.23E-05 | 0.001079902 |
| Bilirubin_pPS | Carotidplaque_Any | 26903 | 14665 | -0.00923446 | -0.035941511 | 0.017472592 | 0.497965736 | 0.69715203 |
| Proinsulin_pPS | Carotidplaque_Any | 26903 | 14665 | 0.008719385 | -0.015893021 | 0.033331791 | 0.487461313 | 0.691668079 |
| Hyper_Insulin_pPS | Carotidplaque_Any | 26903 | 14665 | 0.018418889 | -0.006328569 | 0.043166346 | 0.144633886 | 0.345149045 |
| Beta_Cell_1_pPS | Carotidplaque_Any | 26903 | 14665 | 0.033002997 | 0.008216182 | 0.057789811 | 0.009063732 | 0.059480743 |
| Beta_Cell_2_pPS | Carotidplaque_Any | 26903 | 14665 | 0.013376515 | -0.01144456 | 0.038197591 | 0.290849822 | 0.538806923 |
| Total_GRS | Carotidplaque_Any | 26903 | 14665 | 0.02389239 | -0.000896373 | 0.048681153 | 0.058879667 | 0.199431129 |
| Cholesterol_pPS | SCCTStenosis_Any | 23025 | 10267 | -0.080434784 | -0.108278191 | -0.052591378 | 1.50E-08 | 3.93E-07 |
| Lipodystrophy_1_pPS | SCCTStenosis_Any | 23025 | 10267 | 0.027358921 | -0.000738899 | 0.055456741 | 0.05633669 | 0.197178415 |
| Lipodystrophy_2_pPS | SCCTStenosis_Any | 23025 | 10267 | 0.001658891 | -0.026257428 | 0.02957521 | 0.907281384 | 0.977366923 |
| Liver-Lipid_pPS | SCCTStenosis_Any | 23025 | 10267 | -0.060086653 | -0.088165156 | -0.03200815 | 2.74E-05 | 0.000410702 |
| ASAT_Pos_pPS | SCCTStenosis_Any | 23025 | 10267 | 0.017610754 | -0.010168044 | 0.045389552 | 0.214035051 | 0.425117132 |
| ALP_Neg_pPS | SCCTStenosis_Any | 23025 | 10267 | 0.035378794 | 0.007551769 | 0.063205819 | 0.012707468 | 0.070225479 |
| SHBG-LpA_pPS | SCCTStenosis_Any | 23025 | 10267 | 0.053131919 | 0.025277193 | 0.080986645 | 0.000185075 | 0.00215921 |
| Obesity_pPS | SCCTStenosis_Any | 23025 | 10267 | 0.032209234 | 0.004339973 | 0.060078495 | 0.023501536 | 0.091394861 |
| VAT_Neg_pPS | SCCTStenosis_Any | 23025 | 10267 | 0.026108767 | -0.001777349 | 0.053994883 | 0.066499211 | 0.218200537 |
| Bilirubin_pPS | SCCTStenosis_Any | 23025 | 10267 | 0.018816382 | -0.011316833 | 0.048949598 | 0.220997601 | 0.429717558 |
| Proinsulin_pPS | SCCTStenosis_Any | 23025 | 10267 | 0.036200428 | 0.008338884 | 0.064061971 | 0.010878555 | 0.065802722 |
| Hyper_Insulin_pPS | SCCTStenosis_Any | 23025 | 10267 | 0.00156332 | -0.02637854 | 0.02950518 | 0.912680548 | 0.977366923 |
| Beta_Cell_1_pPS | SCCTStenosis_Any | 23025 | 10267 | 0.09557869 | 0.067586934 | 0.123570446 | 2.20E-11 | 1.18E-09 |
| Beta_Cell_2_pPS | SCCTStenosis_Any | 23025 | 10267 | 0.007547863 | -0.020447346 | 0.035543072 | 0.597200125 | 0.803923245 |
| Total_GRS | SCCTStenosis_Any | 23025 | 10267 | 0.045696827 | 0.017653915 | 0.073739739 | 0.001404015 | 0.011340123 |

**b) Logistic Regression with pMMSs Derived from Biochemistry Elastic Net Models**

| pMMS | Imaging_Variables | Sample_Size | Cases (Positive | coefficient | CI 2.5% | CI 97.5% | p | fdr_p_adjusted |
|---|---|---|---|---|---|---|---|---|
| SHBG-LpA_pMMS_biochemistry | Carotidplaque_Any | 24186 | 13003 | 1.129131222 | 0.797241918 | 1.461020527 | 2.59E-11 | 6.87E-11 |
| Obesity_pMMS_biochemistry | Carotidplaque_Any | 24186 | 13003 | 0.631383355 | 0.320954866 | 0.941811843 | 6.71E-05 | 0.000128911 |
| VAT_Neg_pMMS_biochemistry | Carotidplaque_Any | 24186 | 13003 | 1.148792117 | 0.871196635 | 1.426387599 | 5.02E-16 | 1.70E-15 |
| Proinsulin_pMMS_biochemistry | Carotidplaque_Any | 24186 | 13003 | 0.876567369 | 0.589093123 | 1.164041614 | 2.28E-09 | 5.46E-09 |
| Hyper_Insulin_pMMS_biochemistry | Carotidplaque_Any | 24186 | 13003 | 0.768991046 | 0.38693663 | 1.151045462 | 7.98E-05 | 0.000150409 |
| Beta_Cell_1_pMMS_biochemistry | Carotidplaque_Any | 24186 | 13003 | 1.03248162 | 0.7381988 | 1.326764441 | 6.14E-12 | 1.69E-11 |
| Beta_Cell_2_pMMS_biochemistry | Carotidplaque_Any | 24186 | 13003 | 0.259703392 | 0.132972333 | 0.386434451 | 5.91E-05 | 0.000115798 |
| Total_GRS_biochemistry | Carotidplaque_Any | 24186 | 13003 | 0.441032868 | 0.295589162 | 0.586476575 | 2.79E-09 | 6.52E-09 |
| Cholesterol_pMMS_biochemistry | SCCTStenosis_Any | 20746 | 9147 | -0.753032276 | -0.878454024 | -0.627610528 | 5.73E-32 | 6.24E-31 |
| Lipodystrophy_1_pMMS_biochemistry | SCCTStenosis_Any | 20746 | 9147 | 3.803681848 | 3.387486547 | 4.21987715 | 9.43E-72 | 4.62E-70 |
| Lipodystrophy_2_pMMS_biochemistry | SCCTStenosis_Any | 20746 | 9147 | 1.083463731 | 0.774562949 | 1.392364514 | 6.22E-12 | 1.69E-11 |
| Liver-Lipid_pMMS_biochemistry | SCCTStenosis_Any | 20746 | 9147 | -0.187009234 | -0.369285942 | -0.004732527 | 0.04434116 | 0.062977299 |
| ASAT_Pos_pMMS_biochemistry | SCCTStenosis_Any | 20746 | 9147 | 3.334005353 | 2.753085494 | 3.914925213 | 2.35E-29 | 2.31E-28 |
| ALP_Neg_pMMS_biochemistry | SCCTStenosis_Any | 20746 | 9147 | 1.397541512 | 1.115672422 | 1.679410603 | 2.53E-22 | 1.46E-21 |
| SHBG-LpA_pMMS_biochemistry | SCCTStenosis_Any | 20746 | 9147 | 1.649663259 | 1.272618389 | 2.026708129 | 9.88E-18 | 4.21E-17 |
| Obesity_pMMS_biochemistry | SCCTStenosis_Any | 20746 | 9147 | 1.821009031 | 1.465799119 | 2.176218943 | 9.39E-24 | 6.57E-23 |
| VAT_Neg_pMMS_biochemistry | SCCTStenosis_Any | 20746 | 9147 | 2.300327913 | 1.982311614 | 2.618344213 | 1.27E-45 | 3.11E-44 |
| Proinsulin_pMMS_biochemistry | SCCTStenosis_Any | 20746 | 9147 | 1.377570275 | 1.051272471 | 1.703868079 | 1.29E-16 | 4.51E-16 |
| Hyper_Insulin_pMMS_biochemistry | SCCTStenosis_Any | 20746 | 9147 | 1.763732897 | 1.330844905 | 2.196620889 | 1.40E-15 | 4.57E-15 |
| Beta_Cell_1_pMMS_biochemistry | SCCTStenosis_Any | 20746 | 9147 | 1.611197402 | 1.275993787 | 1.946401017 | 4.48E-21 | 2.44E-20 |
| Beta_Cell_2_pMMS_biochemistry | SCCTStenosis_Any | 20746 | 9147 | 0.507871136 | 0.36382806 | 0.651914212 | 4.83E-12 | 1.39E-11 |
| Total_GRS_biochemistry | SCCTStenosis_Any | 20746 | 9147 | 0.950299431 | 0.783992913 | 1.116605949 | 4.10E-29 | 3.65E-28 |

**Table S14: Associations between pPSs, pMMSs and imaging-derived phenotypes in SCAPIS**

**c) Logistic Regression with pMMSs Derived from Proteomics Elastic Net Models**

| pMMS | Imaging_Variables | Sample_Size | Cases (Positive | coefficient | CI 2.5% | CI 97.5% | p | fdr_p_adjusted |
|---|---|---|---|---|---|---|---|---|
| Cholesterol_pMMS_proteomics | Above50_Any | 3956 | 251 | -0.347073066 | -1.665484951 | 0.971338818 | 0.605880803 | 0.709323867 |
| Lipodystrophy_1_pMMS_proteomics | Above50_Any | 3956 | 251 | 6.97868587 | 4.122732268 | 9.834639472 | 1.67E-06 | 2.00E-05 |
| Lipodystrophy_2_pMMS_proteomics | Above50_Any | 3956 | 251 | 1.950884348 | 0.435037283 | 3.466731414 | 0.011653732 | 0.036258288 |
| Liver-Lipid_pMMS_proteomics | Above50_Any | 3956 | 251 | -0.684545292 | -1.406624672 | 0.037534088 | 0.063157092 | 0.126314183 |
| ASAT_Pos_pMMS_proteomics | Above50_Any | 3956 | 251 | 2.299948627 | 0.52138399 | 4.078513264 | 0.01125999 | 0.036258288 |
| ALP_Neg_pMMS_proteomics | Above50_Any | 3956 | 251 | -0.026490105 | -0.255232991 | 0.202252781 | 0.820440681 | 0.875136726 |
| Proinsulin_pMMS_proteomics | Above50_Any | 3956 | 251 | 0.204505826 | -0.752724579 | 1.161736232 | 0.675411203 | 0.771898518 |
| Beta_Cell_2_pMMS_proteomics | Above50_Any | 3956 | 251 | -2.332250324 | -8.644088026 | 3.979587378 | 0.468933839 | 0.640317918 |
| Cholesterol_pMMS_proteomics | AortaGt4cm | 3884 | 116 | 1.416035951 | -0.455628932 | 3.287700835 | 0.138116939 | 0.236771895 |
| Lipodystrophy_1_pMMS_proteomics | AortaGt4cm | 3884 | 116 | 1.796179495 | -2.232441451 | 5.824800441 | 0.382194984 | 0.555919977 |
| Lipodystrophy_2_pMMS_proteomics | AortaGt4cm | 3884 | 116 | 0.790584893 | -1.404451169 | 2.985620955 | 0.480238439 | 0.640317918 |
| Liver-Lipid_pMMS_proteomics | AortaGt4cm | 3884 | 116 | -0.173770029 | -1.220648651 | 0.873108592 | 0.744929899 | 0.831549655 |
| ASAT_Pos_pMMS_proteomics | AortaGt4cm | 3884 | 116 | 0.013645935 | -2.495077447 | 2.522369317 | 0.991493901 | 0.991493901 |
| ALP_Neg_pMMS_proteomics | AortaGt4cm | 3884 | 116 | 0.124741047 | -0.202578801 | 0.452060896 | 0.455100383 | 0.640317918 |
| Proinsulin_pMMS_proteomics | AortaGt4cm | 3884 | 116 | -0.379361327 | -1.738649483 | 0.97992683 | 0.584376753 | 0.709323867 |
| Beta_Cell_2_pMMS_proteomics | AortaGt4cm | 3884 | 116 | -4.0903847 | -13.03409634 | 4.853326937 | 0.370047311 | 0.555070966 |
| Cholesterol_pMMS_proteomics | AorticValveCalc | 3936 | 268 | -1.372243697 | -2.647161033 | -0.097326362 | 0.034894 | 0.076220025 |
| Lipodystrophy_1_pMMS_proteomics | AorticValveCalc | 3936 | 268 | 2.00658979 | -0.736729265 | 4.749908846 | 0.151684203 | 0.251063509 |
| Lipodystrophy_2_pMMS_proteomics | AorticValveCalc | 3936 | 268 | 2.023233459 | 0.550452114 | 3.496014804 | 0.00709184 | 0.02836736 |
| Liver-Lipid_pMMS_proteomics | AorticValveCalc | 3936 | 268 | -0.398836486 | -1.099606384 | 0.301933413 | 0.264638464 | 0.418573113 |
| ASAT_Pos_pMMS_proteomics | AorticValveCalc | 3936 | 268 | -0.451789821 | -2.165040415 | 1.261460773 | 0.605261603 | 0.709323867 |
| ALP_Neg_pMMS_proteomics | AorticValveCalc | 3936 | 268 | 0.075961123 | -0.143830916 | 0.295753163 | 0.498169692 | 0.646274194 |
| Proinsulin_pMMS_proteomics | AorticValveCalc | 3936 | 268 | 0.901320762 | -0.022021967 | 1.824663492 | 0.055720108 | 0.116285443 |
| Beta_Cell_2_pMMS_proteomics | AorticValveCalc | 3936 | 268 | -6.704783378 | -12.71357933 | -0.695987423 | 0.028743773 | 0.072615848 |
| Cholesterol_pMMS_proteomics | Atherosclerosis_Any | 3956 | 1649 | -0.206091234 | -0.878946896 | 0.466764428 | 0.548290333 | 0.692577262 |
| Lipodystrophy_1_pMMS_proteomics | Atherosclerosis_Any | 3956 | 1649 | 4.352599828 | 2.873694212 | 5.831505444 | 8.00E-09 | 3.84E-07 |
| Lipodystrophy_2_pMMS_proteomics | Atherosclerosis_Any | 3956 | 1649 | 1.959100056 | 1.149928511 | 2.768271601 | 2.08E-06 | 2.00E-05 |
| Liver-Lipid_pMMS_proteomics | Atherosclerosis_Any | 3956 | 1649 | -0.505454251 | -0.880355691 | -0.13055281 | 0.008229966 | 0.030387567 |
| ASAT_Pos_pMMS_proteomics | Atherosclerosis_Any | 3956 | 1649 | 1.458223562 | 0.535469769 | 2.380977355 | 0.001952776 | 0.009373325 |
| ALP_Neg_pMMS_proteomics | Atherosclerosis_Any | 3956 | 1649 | 0.096182027 | -0.019587243 | 0.211951298 | 0.103449929 | 0.183910985 |
| Proinsulin_pMMS_proteomics | Atherosclerosis_Any | 3956 | 1649 | 0.439173567 | -0.052219405 | 0.930566539 | 0.079828451 | 0.147375602 |
| Beta_Cell_2_pMMS_proteomics | Atherosclerosis_Any | 3956 | 1649 | -5.257374086 | -8.489089293 | -2.025658878 | 0.001430222 | 0.008581332 |
| Cholesterol_pMMS_proteomics | Carotidplaque_Any | 3985 | 2290 | -0.81953074 | -1.467788688 | -0.171272793 | 0.013219683 | 0.037326163 |
| Lipodystrophy_1_pMMS_proteomics | Carotidplaque_Any | 3985 | 2290 | 3.155707595 | 1.736345258 | 4.575069933 | 1.31E-05 | 0.000105177 |
| Lipodystrophy_2_pMMS_proteomics | Carotidplaque_Any | 3985 | 2290 | 0.926824662 | 0.145376183 | 1.708273141 | 0.020094193 | 0.053584514 |
| Liver-Lipid_pMMS_proteomics | Carotidplaque_Any | 3985 | 2290 | 0.051858845 | -0.308575497 | 0.412293187 | 0.777945672 | 0.848668005 |
| ASAT_Pos_pMMS_proteomics | Carotidplaque_Any | 3985 | 2290 | 1.406586647 | 0.51752912 | 2.295644173 | 0.001929476 | 0.009373325 |
| ALP_Neg_pMMS_proteomics | Carotidplaque_Any | 3985 | 2290 | 0.002577429 | -0.10867034 | 0.113825198 | 0.96378118 | 0.984287162 |
| Proinsulin_pMMS_proteomics | Carotidplaque_Any | 3985 | 2290 | 0.26604169 | -0.20699567 | 0.739079049 | 0.270328469 | 0.418573113 |
| Beta_Cell_2_pMMS_proteomics | Carotidplaque_Any | 3985 | 2290 | -3.41743918 | -6.508912532 | -0.325965827 | 0.030263601 | 0.072632643 |
| Cholesterol_pMMS_proteomics | SCCTStenosis_Any | 3669 | 1627 | -0.037670123 | -0.730839703 | 0.655499457 | 0.915174754 | 0.954964961 |
| Lipodystrophy_1_pMMS_proteomics | SCCTStenosis_Any | 3669 | 1627 | 4.220232143 | 2.702003949 | 5.738460337 | 5.09E-08 | 1.22E-06 |
| Lipodystrophy_2_pMMS_proteomics | SCCTStenosis_Any | 3669 | 1627 | 2.201164072 | 1.36213944 | 3.040188704 | 2.72E-07 | 4.35E-06 |
| Liver-Lipid_pMMS_proteomics | SCCTStenosis_Any | 3669 | 1627 | -0.495808429 | -0.883025002 | -0.108591856 | 0.012086096 | 0.036258288 |
| ASAT_Pos_pMMS_proteomics | SCCTStenosis_Any | 3669 | 1627 | 1.407601936 | 0.456376811 | 2.358827061 | 0.003727921 | 0.01626729 |
| ALP_Neg_pMMS_proteomics | SCCTStenosis_Any | 3669 | 1627 | 0.128530428 | 0.009089645 | 0.247971211 | 0.034934178 | 0.076220025 |
| Proinsulin_pMMS_proteomics | SCCTStenosis_Any | 3669 | 1627 | 0.477134317 | -0.032511203 | 0.986779837 | 0.066515416 | 0.127709598 |
| Beta_Cell_2_pMMS_proteomics | SCCTStenosis_Any | 3669 | 1627 | -5.962832525 | -9.292100603 | -2.633564446 | 0.000447497 | 0.003068548 |

**d) Logistic Regression with pMMSs Derived from Metabolomics Elastic Net Models**

| pMMS | Imaging_Variables | Sample_Size | Cases (Positive | coefficient | CI 2.5% | CI 97.5% | p | fdr_p_adjusted |
|---|---|---|---|---|---|---|---|---|
| Cholesterol_pMMS_metabolomics | Above50_Any | 4694 | 285 | -0.15257642 | -0.654394959 | 0.349242119 | 0.551227894 | 0.661473472 |
| Lipodystrophy_1_pMMS_metabolomics | Above50_Any | 4694 | 285 | 3.583004566 | 0.900825812 | 6.26518332 | 0.00883873 | 0.035354921 |
| Liver-Lipid_pMMS_metabolomics | Above50_Any | 4694 | 285 | -0.053541705 | -0.62907008 | 0.521986669 | 0.855318677 | 0.962671445 |
| ALP_Neg_pMMS_metabolomics | Above50_Any | 4694 | 285 | 0.497008894 | -0.328972039 | 1.322989826 | 0.238259357 | 0.381214972 |
| Cholesterol_pMMS_metabolomics | AortaGt4cm | 4620 | 130 | -0.025714109 | -0.746213868 | 0.694785651 | 0.944233712 | 0.985287352 |
| Lipodystrophy_1_pMMS_metabolomics | AortaGt4cm | 4620 | 130 | 2.924022916 | -0.910321035 | 6.758366868 | 0.135007347 | 0.249244334 |
| Liver-Lipid_pMMS_metabolomics | AortaGt4cm | 4620 | 130 | -1.147460395 | -1.972675906 | -0.322244883 | 0.006423808 | 0.03083428 |
| ALP_Neg_pMMS_metabolomics | AortaGt4cm | 4620 | 130 | -0.473651456 | -1.671931866 | 0.724628953 | 0.438501186 | 0.613130903 |
| Cholesterol_pMMS_metabolomics | AorticValveCalc | 4673 | 312 | 0.550240525 | 0.079976 | 1.02050505 | 0.021831476 | 0.05821727 |
| Lipodystrophy_1_pMMS_metabolomics | AorticValveCalc | 4673 | 312 | 2.855887038 | 0.325517442 | 5.386256633 | 0.026959615 | 0.064703077 |
| Liver-Lipid_pMMS_metabolomics | AorticValveCalc | 4673 | 312 | -0.223700443 | -0.771418155 | 0.324017269 | 0.423424565 | 0.613130903 |
| ALP_Neg_pMMS_metabolomics | AorticValveCalc | 4673 | 312 | -0.059251362 | -0.844630273 | 0.726127549 | 0.882448825 | 0.962671445 |
| Cholesterol_pMMS_metabolomics | Atherosclerosis_Any | 4694 | 1954 | -0.318551236 | -0.570741367 | -0.066361106 | 0.013297254 | 0.045590583 |
| Lipodystrophy_1_pMMS_metabolomics | Atherosclerosis_Any | 4694 | 1954 | 4.6927766 | 3.345498612 | 6.040054588 | 8.68E-12 | 1.04E-10 |
| Liver-Lipid_pMMS_metabolomics | Atherosclerosis_Any | 4694 | 1954 | -0.230271795 | -0.514192066 | 0.053648476 | 0.111921214 | 0.223842427 |
| ALP_Neg_pMMS_metabolomics | Atherosclerosis_Any | 4694 | 1954 | 0.127517256 | -0.281953067 | 0.536987578 | 0.541615397 | 0.661473472 |
| Cholesterol_pMMS_metabolomics | Carotidplaque_Any | 4722 | 2716 | -0.644858073 | -0.890612663 | -0.399103482 | 2.70E-07 | 1.62E-06 |
| Lipodystrophy_1_pMMS_metabolomics | Carotidplaque_Any | 4722 | 2716 | 3.82223572 | 2.521021534 | 5.123449906 | 8.55E-09 | 6.84E-08 |
| Liver-Lipid_pMMS_metabolomics | Carotidplaque_Any | 4722 | 2716 | 0.000695152 | -0.272015803 | 0.273406107 | 0.996013759 | 0.996013759 |
| ALP_Neg_pMMS_metabolomics | Carotidplaque_Any | 4722 | 2716 | 0.489277231 | 0.094065535 | 0.884488927 | 0.015246891 | 0.045740672 |
| Cholesterol_pMMS_metabolomics | SCCTStenosis_Any | 4345 | 1925 | -0.281807518 | -0.542067297 | -0.021547738 | 0.033817953 | 0.073784625 |
| Lipodystrophy_1_pMMS_metabolomics | SCCTStenosis_Any | 4345 | 1925 | 5.017963966 | 3.616791271 | 6.41913666 | 2.23E-12 | 5.36E-11 |
| Liver-Lipid_pMMS_metabolomics | SCCTStenosis_Any | 4345 | 1925 | -0.205925256 | -0.497958545 | 0.086108032 | 0.166954794 | 0.286208218 |
| ALP_Neg_pMMS_metabolomics | SCCTStenosis_Any | 4345 | 1925 | 0.159199222 | -0.262971069 | 0.581369513 | 0.459848177 | 0.613130903 |

**Table S14: Associations between pPSs, pMMSs and imaging-derived phenotypes in SCAPIS**

**e) Logistic Regression with pMMSs Derived from Merged-omics Elastic Net Models**

| pMMS | Imaging_Variables | Sample_Size | Cases (Positive | coefficient | CI 2.5% | CI 97.5% | p | fdr_p_adjusted |
|---|---|---|---|---|---|---|---|---|
| Cholesterol_pMMS_merged_omics | Above50_Any | 3106 | 182 | -0.550652401 | -1.122169617 | 0.020864815 | 0.058970919 | 0.098284865 |
| Lipodystrophy_1_pMMS_merged_omics | Above50_Any | 3106 | 182 | 5.417101019 | 1.491118266 | 9.343083772 | 0.006843336 | 0.016743762 |
| Lipodystrophy_2_pMMS_merged_omics | Above50_Any | 3106 | 182 | 1.753139489 | 0.606783493 | 2.899495485 | 0.00272291 | 0.00816873 |
| Liver-Lipid_pMMS_merged_omics | Above50_Any | 3106 | 182 | -0.375430794 | -1.129029918 | 0.378168331 | 0.328855338 | 0.448439097 |
| ASAT_Pos_pMMS_merged_omics | Above50_Any | 3106 | 182 | 6.286277765 | 3.131763608 | 9.440791921 | 9.39E-05 | 0.000352189 |
| ALP_Neg_pMMS_merged_omics | Above50_Any | 3106 | 182 | 0.247089668 | -0.032656564 | 0.526835901 | 0.083422363 | 0.129448494 |
| Proinsulin_pMMS_merged_omics | Above50_Any | 3106 | 182 | -0.269518453 | -1.724376288 | 1.185339382 | 0.716536561 | 0.806103631 |
| Beta_Cell_2_pMMS_merged_omics | Above50_Any | 3106 | 182 | 2.537038032 | 0.418781984 | 4.655294081 | 0.018902338 | 0.038663873 |
| Total_GRS_merged_omics | Above50_Any | 3106 | 182 | 2.730101284 | 1.37812436 | 4.082078209 | 7.56E-05 | 0.000309406 |
| Cholesterol_pMMS_merged_omics | AorticValveCalc | 3079 | 219 | -0.265298055 | -0.782632385 | 0.252036276 | 0.314846935 | 0.442753502 |
| Lipodystrophy_1_pMMS_merged_omics | AorticValveCalc | 3079 | 219 | 4.920402305 | 1.36753523 | 8.47326938 | 0.006640108 | 0.016743762 |
| Lipodystrophy_2_pMMS_merged_omics | AorticValveCalc | 3079 | 219 | 0.783812489 | -0.269575905 | 1.837200884 | 0.144734867 | 0.217102301 |
| Liver-Lipid_pMMS_merged_omics | AorticValveCalc | 3079 | 219 | -0.041506051 | -0.732666726 | 0.649654625 | 0.906304504 | 0.924868794 |
| ASAT_Pos_pMMS_merged_omics | AorticValveCalc | 3079 | 219 | 2.846266563 | -0.017219435 | 5.709752561 | 0.051393688 | 0.088950614 |
| ALP_Neg_pMMS_merged_omics | AorticValveCalc | 3079 | 219 | 0.090906305 | -0.162116184 | 0.343928794 | 0.481321406 | 0.617161538 |
| Proinsulin_pMMS_merged_omics | AorticValveCalc | 3079 | 219 | 0.103145448 | -1.213057113 | 1.41934801 | 0.87792954 | 0.924868794 |
| Beta_Cell_2_pMMS_merged_omics | AorticValveCalc | 3079 | 219 | -0.092045326 | -2.005094873 | 1.821004221 | 0.924868794 | 0.924868794 |
| Total_GRS_merged_omics | AorticValveCalc | 3079 | 219 | 1.617630682 | 0.37731676 | 2.857944604 | 0.010582045 | 0.023731907 |
| Cholesterol_pMMS_merged_omics | Atherosclerosis_Any | 3106 | 1248 | -0.490470369 | -0.774809687 | -0.20613105 | 0.00072266 | 0.002322835 |
| Lipodystrophy_1_pMMS_merged_omics | Atherosclerosis_Any | 3106 | 1248 | 5.666256753 | 3.743526349 | 7.588987157 | 7.65E-09 | 8.61E-08 |
| Lipodystrophy_2_pMMS_merged_omics | Atherosclerosis_Any | 3106 | 1248 | 1.259179421 | 0.676863548 | 1.841495295 | 2.25E-05 | 0.000112681 |
| Liver-Lipid_pMMS_merged_omics | Atherosclerosis_Any | 3106 | 1248 | -0.154518404 | -0.52830964 | 0.219272831 | 0.417817784 | 0.552994125 |
| ASAT_Pos_pMMS_merged_omics | Atherosclerosis_Any | 3106 | 1248 | 5.210994401 | 3.626821193 | 6.795167608 | 1.14E-10 | 2.56E-09 |
| ALP_Neg_pMMS_merged_omics | Atherosclerosis_Any | 3106 | 1248 | 0.121460696 | -0.014989513 | 0.257910905 | 0.081044876 | 0.129448494 |
| Proinsulin_pMMS_merged_omics | Atherosclerosis_Any | 3106 | 1248 | -0.764670852 | -1.485000128 | -0.044341575 | 0.037469232 | 0.07025481 |
| Beta_Cell_2_pMMS_merged_omics | Atherosclerosis_Any | 3106 | 1248 | 0.36214541 | -0.674971521 | 1.399262342 | 0.49372923 | 0.617161538 |
| Total_GRS_merged_omics | Atherosclerosis_Any | 3106 | 1248 | 1.523244152 | 0.840102507 | 2.206385798 | 1.24E-05 | 6.98E-05 |
| Cholesterol_pMMS_merged_omics | Carotidplaque_Any | 3129 | 1740 | -0.757717222 | -1.031768757 | -0.483665687 | 5.99E-08 | 5.39E-07 |
| Lipodystrophy_1_pMMS_merged_omics | Carotidplaque_Any | 3129 | 1740 | 3.81654341 | 1.996289906 | 5.636796914 | 3.97E-05 | 0.000178451 |
| Lipodystrophy_2_pMMS_merged_omics | Carotidplaque_Any | 3129 | 1740 | 0.358145872 | -0.195559393 | 0.911851137 | 0.204891656 | 0.297423371 |
| Liver-Lipid_pMMS_merged_omics | Carotidplaque_Any | 3129 | 1740 | -0.024778609 | -0.381343118 | 0.331785899 | 0.891660745 | 0.924868794 |
| ASAT_Pos_pMMS_merged_omics | Carotidplaque_Any | 3129 | 1740 | 2.116155012 | 0.62170624 | 3.610603783 | 0.005514582 | 0.015509763 |
| ALP_Neg_pMMS_merged_omics | Carotidplaque_Any | 3129 | 1740 | 0.009063799 | -0.120517445 | 0.138645043 | 0.890957052 | 0.924868794 |
| Proinsulin_pMMS_merged_omics | Carotidplaque_Any | 3129 | 1740 | -0.888798919 | -1.574542895 | -0.203054944 | 0.01107489 | 0.023731907 |
| Beta_Cell_2_pMMS_merged_omics | Carotidplaque_Any | 3129 | 1740 | 0.214252557 | -0.778447428 | 1.206952542 | 0.672284088 | 0.77571241 |
| Total_GRS_merged_omics | Carotidplaque_Any | 3129 | 1740 | 1.167232606 | 0.515489063 | 1.818976149 | 0.000447812 | 0.001550118 |
| Cholesterol_pMMS_merged_omics | SCCTStenosis_Any | 2861 | 1229 | -0.40289207 | -0.696057008 | -0.109727132 | 0.007069588 | 0.016743762 |
| Lipodystrophy_1_pMMS_merged_omics | SCCTStenosis_Any | 2861 | 1229 | 6.063787093 | 4.066413407 | 8.061160779 | 2.68E-09 | 4.02E-08 |
| Lipodystrophy_2_pMMS_merged_omics | SCCTStenosis_Any | 2861 | 1229 | 1.500970739 | 0.894200613 | 2.107740865 | 1.24E-06 | 9.34E-06 |
| Liver-Lipid_pMMS_merged_omics | SCCTStenosis_Any | 2861 | 1229 | -0.119075738 | -0.50549169 | 0.267340215 | 0.545862702 | 0.663887071 |
| ASAT_Pos_pMMS_merged_omics | SCCTStenosis_Any | 2861 | 1229 | 5.91463168 | 4.25923722 | 7.57002614 | 2.51E-12 | 1.13E-10 |
| ALP_Neg_pMMS_merged_omics | SCCTStenosis_Any | 2861 | 1229 | 0.157838662 | 0.016968434 | 0.29870889 | 0.02808835 | 0.054955468 |
| Proinsulin_pMMS_merged_omics | SCCTStenosis_Any | 2861 | 1229 | -0.75112395 | -1.495850344 | -0.006397555 | 0.048064127 | 0.086515429 |
| Beta_Cell_2_pMMS_merged_omics | SCCTStenosis_Any | 2861 | 1229 | 0.284806223 | -0.793037295 | 1.362649742 | 0.60453142 | 0.715892471 |
| Total_GRS_merged_omics | SCCTStenosis_Any | 2861 | 1229 | 1.740481425 | 1.02612014 | 2.45484271 | 1.79E-06 | 1.15E-05 |

**Table S15: Associations between pPSs and health history in SCAPIS**

| PPS | Health_History | Health_History_Heading | Sample_Size | Cases (Positive I | coefficient | CI 2.5% | CI 97.5% | p | fdr_p_adjusted |
|---|---|---|---|---|---|---|---|---|---|
| Cholesterol_pPS | cqhe_cad | Coronary artery disease | 28239 | 702 | -0.025113216 | -0.10191 | 0.051688 | 0.521596906 | 0.806593154 |
| Lipodystrophy_1_pPS | cqhe_cad | Coronary artery disease | 28239 | 702 | -0.041728848 | -0.11777 | 0.034314 | 0.28213486 | 0.587780958 |
| Lipodystrophy_2_pPS | cqhe_cad | Coronary artery disease | 28239 | 702 | -0.027704537 | -0.10385 | 0.048439 | 0.475769816 | 0.779031707 |
| Liver-Lipid_pPS | cqhe_cad | Coronary artery disease | 28239 | 702 | 0.03284374 | -0.04304 | 0.108732 | 0.396299249 | 0.733887498 |
| ASAT_Pos_pPS | cqhe_cad | Coronary artery disease | 28239 | 702 | 0.045577385 | -0.03012 | 0.121276 | 0.237971765 | 0.517329924 |
| ALP_Neg_pPS | cqhe_cad | Coronary artery disease | 28239 | 702 | 0.103820011 | 0.027645 | 0.179995 | 0.007556424 | 0.036915048 |
| SHBG-LpA_pPS | cqhe_cad | Coronary artery disease | 28239 | 702 | -0.006349368 | -0.08267 | 0.069972 | 0.870476172 | 0.932653041 |
| Obesity_pPS | cqhe_cad | Coronary artery disease | 28239 | 702 | 0.057346645 | -0.01879 | 0.133479 | 0.139852482 | 0.393867342 |
| VAT_Neg_pPS | cqhe_cad | Coronary artery disease | 28239 | 702 | 0.050813997 | -0.0257 | 0.127323 | 0.19301307 | 0.455998238 |
| Bilirubin_pPS | cqhe_cad | Coronary artery disease | 28239 | 702 | -0.017353487 | -0.09986 | 0.065157 | 0.680181826 | 0.872027982 |
| Proinsulin_pPS | cqhe_cad | Coronary artery disease | 28239 | 702 | -0.005368223 | -0.08082 | 0.070086 | 0.889100157 | 0.943804768 |
| Hyper_Insulin_pPS | cqhe_cad | Coronary artery disease | 28239 | 702 | 0.056227778 | -0.02023 | 0.132682 | 0.149458583 | 0.393867342 |
| Beta_Cell_1_pPS | cqhe_cad | Coronary artery disease | 28239 | 702 | 0.064560082 | -0.01183 | 0.140946 | 0.097615064 | 0.325383546 |
| Beta_Cell_2_pPS | cqhe_cad | Coronary artery disease | 28239 | 702 | 0.056209983 | -0.02 | 0.132419 | 0.148284156 | 0.393867342 |
| Total_GRS | cqhe_cad | Coronary artery disease | 28239 | 702 | 0.017864337 | -0.05825 | 0.09398 | 0.645513568 | 0.872027982 |
| Cholesterol_pPS | cqhe009 | Atrial fibrillation | 28236 | 519 | -0.029522106 | -0.11659 | 0.057548 | 0.506338994 | 0.791154679 |
| Lipodystrophy_1_pPS | cqhe009 | Atrial fibrillation | 28236 | 519 | -0.015645135 | -0.1032 | 0.071906 | 0.726159504 | 0.90769938 |
| Lipodystrophy_2_pPS | cqhe009 | Atrial fibrillation | 28236 | 519 | 0.038612231 | -0.04932 | 0.126547 | 0.389444927 | 0.730209237 |
| Liver-Lipid_pPS | cqhe009 | Atrial fibrillation | 28236 | 519 | -0.009822825 | -0.09747 | 0.077821 | 0.826131517 | 0.928523428 |
| ASAT_Pos_pPS | cqhe009 | Atrial fibrillation | 28236 | 519 | 0.010763562 | -0.07659 | 0.098119 | 0.809168649 | 0.928523428 |
| ALP_Neg_pPS | cqhe009 | Atrial fibrillation | 28236 | 519 | 0.072437149 | -0.01532 | 0.160197 | 0.105714967 | 0.337453309 |
| SHBG-LpA_pPS | cqhe009 | Atrial fibrillation | 28236 | 519 | 0.001027437 | -0.08682 | 0.088874 | 0.981711358 | 0.981711358 |
| Obesity_pPS | cqhe009 | Atrial fibrillation | 28236 | 519 | 0.057464665 | -0.02995 | 0.144881 | 0.197599236 | 0.455998238 |
| VAT_Neg_pPS | cqhe009 | Atrial fibrillation | 28236 | 519 | -0.01419695 | -0.10208 | 0.073685 | 0.751528316 | 0.924010225 |
| Bilirubin_pPS | cqhe009 | Atrial fibrillation | 28236 | 519 | 0.020000239 | -0.07461 | 0.11461 | 0.678632105 | 0.872027982 |
| Proinsulin_pPS | cqhe009 | Atrial fibrillation | 28236 | 519 | 0.022607687 | -0.06475 | 0.109966 | 0.611995737 | 0.86738823 |
| Hyper_Insulin_pPS | cqhe009 | Atrial fibrillation | 28236 | 519 | 0.009265027 | -0.07875 | 0.097277 | 0.836536267 | 0.929484741 |
| Beta_Cell_1_pPS | cqhe009 | Atrial fibrillation | 28236 | 519 | -0.024313591 | -0.11226 | 0.063636 | 0.587935497 | 0.864611025 |
| Beta_Cell_2_pPS | cqhe009 | Atrial fibrillation | 28236 | 519 | 0.022914468 | -0.06497 | 0.110796 | 0.609317374 | 0.86738823 |
| Total_GRS | cqhe009 | Atrial fibrillation | 28236 | 519 | 0.020081831 | -0.06731 | 0.107472 | 0.652430993 | 0.872027982 |
| Cholesterol_pPS | cqhe011 | Heart failure | 28236 | 144 | -0.155818483 | -0.32326 | 0.011626 | 0.068170802 | 0.249404284 |
| Lipodystrophy_1_pPS | cqhe011 | Heart failure | 28236 | 144 | -0.107744366 | -0.27283 | 0.057345 | 0.2008411 | 0.456457046 |
| Lipodystrophy_2_pPS | cqhe011 | Heart failure | 28236 | 144 | 0.101836607 | -0.0639 | 0.267576 | 0.228482052 | 0.504004526 |
| Liver-Lipid_pPS | cqhe011 | Heart failure | 28236 | 144 | -0.06890168 | -0.23367 | 0.095869 | 0.41244796 | 0.736514214 |
| ASAT_Pos_pPS | cqhe011 | Heart failure | 28236 | 144 | 0.11027532 | -0.05412 | 0.274673 | 0.188605909 | 0.455998238 |
| ALP_Neg_pPS | cqhe011 | Heart failure | 28236 | 144 | 0.063499737 | -0.1014 | 0.228396 | 0.450392264 | 0.767714087 |
| SHBG-LpA_pPS | cqhe011 | Heart failure | 28236 | 144 | 0.190629855 | 0.024842 | 0.356418 | 0.024218128 | 0.110082399 |
| Obesity_pPS | cqhe011 | Heart failure | 28236 | 144 | 0.109609714 | -0.05607 | 0.275293 | 0.194755893 | 0.455998238 |
| VAT_Neg_pPS | cqhe011 | Heart failure | 28236 | 144 | 0.008284633 | -0.15787 | 0.174435 | 0.92214787 | 0.953946072 |
| Bilirubin_pPS | cqhe011 | Heart failure | 28236 | 144 | 0.064232605 | -0.11341 | 0.241873 | 0.478511708 | 0.779031707 |
| Proinsulin_pPS | cqhe011 | Heart failure | 28236 | 144 | 0.065599076 | -0.09791 | 0.22911 | 0.431680205 | 0.744276215 |
| Hyper_Insulin_pPS | cqhe011 | Heart failure | 28236 | 144 | 0.021210553 | -0.14484 | 0.187258 | 0.802308147 | 0.928523428 |
| Beta_Cell_1_pPS | cqhe011 | Heart failure | 28236 | 144 | 0.0183412 | -0.14772 | 0.1844 | 0.828614617 | 0.928523428 |
| Beta_Cell_2_pPS | cqhe011 | Heart failure | 28236 | 144 | -0.033285387 | -0.19886 | 0.132288 | 0.69357073 | 0.881657708 |
| Total_GRS | cqhe011 | Heart failure | 28236 | 144 | 0.037209533 | -0.12827 | 0.202691 | 0.659423751 | 0.872027982 |
| Cholesterol_pPS | cqhe029 | Stroke | 28236 | 407 | 0.013861049 | -0.08411 | 0.111837 | 0.78156145 | 0.928523428 |
| Lipodystrophy_1_pPS | cqhe029 | Stroke | 28236 | 407 | -0.040547446 | -0.13907 | 0.057973 | 0.419870165 | 0.738505913 |
| Lipodystrophy_2_pPS | cqhe029 | Stroke | 28236 | 407 | 0.13459669 | 0.035722 | 0.233472 | 0.00762911 | 0.036915048 |
| Liver-Lipid_pPS | cqhe029 | Stroke | 28236 | 407 | -0.094480775 | -0.19322 | 0.00426 | 0.060738118 | 0.227767942 |
| ASAT_Pos_pPS | cqhe029 | Stroke | 28236 | 407 | 0.023102411 | -0.07504 | 0.121249 | 0.644549224 | 0.872027982 |
| ALP_Neg_pPS | cqhe029 | Stroke | 28236 | 407 | -0.001349622 | -0.09992 | 0.097216 | 0.978589717 | 0.981711358 |
| SHBG-LpA_pPS | cqhe029 | Stroke | 28236 | 407 | 0.044757726 | -0.05403 | 0.143541 | 0.374518099 | 0.711110314 |
| Obesity_pPS | cqhe029 | Stroke | 28236 | 407 | 0.03109025 | -0.06725 | 0.129432 | 0.535499398 | 0.819611492 |
| VAT_Neg_pPS | cqhe029 | Stroke | 28236 | 407 | 0.046702272 | -0.05202 | 0.145427 | 0.353835017 | 0.689288995 |
| Bilirubin_pPS | cqhe029 | Stroke | 28236 | 407 | 0.105605668 | 0.000353 | 0.210858 | 0.04923606 | 0.19435287 |
| Proinsulin_pPS | cqhe029 | Stroke | 28236 | 407 | 0.071971851 | -0.02591 | 0.169851 | 0.149532912 | 0.393867342 |
| Hyper_Insulin_pPS | cqhe029 | Stroke | 28236 | 407 | 0.072000784 | -0.02681 | 0.17081 | 0.153234361 | 0.39629576 |
| Beta_Cell_1_pPS | cqhe029 | Stroke | 28236 | 407 | 0.057616223 | -0.04119 | 0.156419 | 0.253061354 | 0.542274329 |
| Beta_Cell_2_pPS | cqhe029 | Stroke | 28236 | 407 | -0.011142414 | -0.10978 | 0.087498 | 0.824782958 | 0.928523428 |
| Total_GRS | cqhe029 | Stroke | 28236 | 407 | 0.081158228 | -0.01718 | 0.179497 | 0.105760858 | 0.337453309 |
| Cholesterol_pPS | cqhe034 | Hypertension | 28311 | 6460 | -0.00193087 | -0.03019 | 0.026329 | 0.893468513 | 0.943804768 |
| Lipodystrophy_1_pPS | cqhe034 | Hypertension | 28311 | 6460 | 0.028514487 | 0.000132 | 0.056897 | 0.04894623 | 0.19435287 |
| Lipodystrophy_2_pPS | cqhe034 | Hypertension | 28311 | 6460 | 0.029444228 | 0.000955 | 0.057934 | 0.042800749 | 0.178336456 |
| Liver-Lipid_pPS | cqhe034 | Hypertension | 28311 | 6460 | -0.010196284 | -0.03868 | 0.018292 | 0.482999659 | 0.779031707 |
| ASAT_Pos_pPS | cqhe034 | Hypertension | 28311 | 6460 | 0.048578084 | 0.020282 | 0.076874 | 0.000765975 | 0.005222557 |
| ALP_Neg_pPS | cqhe034 | Hypertension | 28311 | 6460 | 0.020853572 | -0.00752 | 0.049223 | 0.14966959 | 0.393867342 |
| SHBG-LpA_pPS | cqhe034 | Hypertension | 28311 | 6460 | -0.01413405 | -0.04254 | 0.014276 | 0.329510531 | 0.667926753 |
| Obesity_pPS | cqhe034 | Hypertension | 28311 | 6460 | 0.099998385 | 0.071565 | 0.128432 | 5.46E-12 | 1.64E-10 |
| VAT_Neg_pPS | cqhe034 | Hypertension | 28311 | 6460 | 0.050157241 | 0.021708 | 0.078606 | 0.000549274 | 0.003923388 |
| Bilirubin_pPS | cqhe034 | Hypertension | 28311 | 6460 | -0.003828355 | -0.03451 | 0.026857 | 0.806822662 | 0.928523428 |
| Proinsulin_pPS | cqhe034 | Hypertension | 28311 | 6460 | 0.025660411 | -0.00259 | 0.053914 | 0.075062374 | 0.261845491 |
| Hyper_Insulin_pPS | cqhe034 | Hypertension | 28311 | 6460 | 0.05823499 | 0.029804 | 0.086666 | 5.96E-05 | 0.000496361 |

**Table S15: Associations between pPSs and health history in SCAPIS**

| PPS | Health_History | Health_History_Heading | Sample_Size | Cases (Positive I | coefficient | CI 2.5% | CI 97.5% | p | fdr_p_adjusted |
|---|---|---|---|---|---|---|---|---|---|
| Beta_Cell_1_pPS | cqhe034 | Hypertension | 28311 | 6460 | 0.051481196 | 0.023001 | 0.079962 | 0.000395871 | 0.002969033 |
| Beta_Cell_2_pPS | cqhe034 | Hypertension | 28311 | 6460 | 0.007552398 | -0.02092 | 0.036026 | 0.603160371 | 0.86738823 |
| Total_GRS | cqhe034 | Hypertension | 28311 | 6460 | 0.097754711 | 0.069312 | 0.126197 | 1.63E-11 | 4.06E-10 |
| Cholesterol_pPS | cqhe036 | Hyperlipidemia | 28311 | 3350 | -0.196372092 | -0.23335 | -0.1594 | 2.24E-25 | 1.68E-23 |
| Lipodystrophy_1_pPS | cqhe036 | Hyperlipidemia | 28311 | 3350 | 0.06936422 | 0.032628 | 0.1061 | 0.000214957 | 0.00169703 |
| Lipodystrophy_2_pPS | cqhe036 | Hyperlipidemia | 28311 | 3350 | 0.028008224 | -0.00881 | 0.064827 | 0.135975063 | 0.393867342 |
| Liver-Lipid_pPS | cqhe036 | Hyperlipidemia | 28311 | 3350 | -0.013465731 | -0.05026 | 0.023332 | 0.473237434 | 0.779031707 |
| ASAT_Pos_pPS | cqhe036 | Hyperlipidemia | 28311 | 3350 | 0.05372096 | 0.017134 | 0.090308 | 0.004004286 | 0.022246031 |
| ALP_Neg_pPS | cqhe036 | Hyperlipidemia | 28311 | 3350 | 0.07550751 | 0.038774 | 0.112241 | 5.61E-05 | 0.000494665 |
| SHBG-LpA_pPS | cqhe036 | Hyperlipidemia | 28311 | 3350 | 0.038863517 | 0.002095 | 0.075632 | 0.038296565 | 0.164128134 |
| Obesity_pPS | cqhe036 | Hyperlipidemia | 28311 | 3350 | 0.060305385 | 0.0236 | 0.097011 | 0.001281425 | 0.008357119 |
| VAT_Neg_pPS | cqhe036 | Hyperlipidemia | 28311 | 3350 | 0.050170189 | 0.013348 | 0.086992 | 0.00757498 | 0.036915048 |
| Bilirubin_pPS | cqhe036 | Hyperlipidemia | 28311 | 3350 | -0.011622222 | -0.05134 | 0.028098 | 0.56631388 | 0.841060219 |
| Proinsulin_pPS | cqhe036 | Hyperlipidemia | 28311 | 3350 | 0.003386859 | -0.03312 | 0.039898 | 0.855732512 | 0.932653041 |
| Hyper_Insulin_pPS | cqhe036 | Hyperlipidemia | 28311 | 3350 | 0.051133674 | 0.014353 | 0.087915 | 0.006434488 | 0.03447047 |
| Beta_Cell_1_pPS | cqhe036 | Hyperlipidemia | 28311 | 3350 | 0.082761712 | 0.045906 | 0.119617 | 1.08E-05 | 0.00012024 |
| Beta_Cell_2_pPS | cqhe036 | Hyperlipidemia | 28311 | 3350 | 0.059315855 | 0.022484 | 0.096148 | 0.001597389 | 0.00998368 |
| Total_GRS | cqhe036 | Hyperlipidemia | 28311 | 3350 | 0.119629861 | 0.082842 | 0.156418 | 1.85E-10 | 3.46E-09 |
| Cholesterol_pPS | cqhe038 | Diabetes | 28311 | 1256 | 0.086869728 | 0.028915 | 0.144825 | 0.003305201 | 0.019068465 |
| Lipodystrophy_1_pPS | cqhe038 | Diabetes | 28311 | 1256 | 0.128179738 | 0.070785 | 0.185575 | 1.20E-05 | 0.00012024 |
| Lipodystrophy_2_pPS | cqhe038 | Diabetes | 28311 | 1256 | 0.088317052 | 0.030749 | 0.145885 | 0.002639836 | 0.015839017 |
| Liver-Lipid_pPS | cqhe038 | Diabetes | 28311 | 1256 | 0.132426465 | 0.075056 | 0.189797 | 6.06E-06 | 8.27E-05 |
| ASAT_Pos_pPS | cqhe038 | Diabetes | 28311 | 1256 | 0.171781432 | 0.114631 | 0.228932 | 3.83E-09 | 5.75E-08 |
| ALP_Neg_pPS | cqhe038 | Diabetes | 28311 | 1256 | 0.006563587 | -0.05071 | 0.063834 | 0.822269462 | 0.928523428 |
| SHBG-LpA_pPS | cqhe038 | Diabetes | 28311 | 1256 | 0.061446917 | 0.00404 | 0.118854 | 0.035914931 | 0.158448226 |
| Obesity_pPS | cqhe038 | Diabetes | 28311 | 1256 | 0.128765553 | 0.071256 | 0.186275 | 1.14E-05 | 0.00012024 |
| VAT_Neg_pPS | cqhe038 | Diabetes | 28311 | 1256 | 0.130851101 | 0.073187 | 0.188515 | 8.69E-06 | 0.000108563 |
| Bilirubin_pPS | cqhe038 | Diabetes | 28311 | 1256 | -0.007441943 | -0.06954 | 0.054656 | 0.814298472 | 0.928523428 |
| Proinsulin_pPS | cqhe038 | Diabetes | 28311 | 1256 | 0.191943993 | 0.134867 | 0.249021 | 4.36E-11 | 9.35E-10 |
| Hyper_Insulin_pPS | cqhe038 | Diabetes | 28311 | 1256 | 0.185645191 | 0.128029 | 0.243261 | 2.70E-10 | 4.50E-09 |
| Beta_Cell_1_pPS | cqhe038 | Diabetes | 28311 | 1256 | 0.253625729 | 0.195783 | 0.311468 | 8.40E-18 | 3.15E-16 |
| Beta_Cell_2_pPS | cqhe038 | Diabetes | 28311 | 1256 | 0.292477649 | 0.234472 | 0.350483 | 4.95E-23 | 2.48E-21 |
| Total_GRS | cqhe038 | Diabetes | 28311 | 1256 | 0.476897362 | 0.418472 | 0.535323 | 1.32E-57 | 1.97E-55 |
| Cholesterol_pPS | cqhe058 | Sleep apnea | 28236 | 1266 | 0.026653101 | -0.03034 | 0.083644 | 0.359337092 | 0.69103287 |
| Lipodystrophy_1_pPS | cqhe058 | Sleep apnea | 28236 | 1266 | -0.006264623 | -0.06328 | 0.050747 | 0.829480929 | 0.928523428 |
| Lipodystrophy_2_pPS | cqhe058 | Sleep apnea | 28236 | 1266 | 0.024365452 | -0.03279 | 0.081519 | 0.403396957 | 0.736514214 |
| Liver-Lipid_pPS | cqhe058 | Sleep apnea | 28236 | 1266 | -0.00508802 | -0.06219 | 0.052018 | 0.86137087 | 0.932653041 |
| ASAT_Pos_pPS | cqhe058 | Sleep apnea | 28236 | 1266 | -0.002420449 | -0.05914 | 0.054299 | 0.933343063 | 0.958370991 |
| ALP_Neg_pPS | cqhe058 | Sleep apnea | 28236 | 1266 | -0.020835907 | -0.07782 | 0.036151 | 0.473609051 | 0.779031707 |
| SHBG-LpA_pPS | cqhe058 | Sleep apnea | 28236 | 1266 | 0.002221877 | -0.05487 | 0.059318 | 0.939203571 | 0.958370991 |
| Obesity_pPS | cqhe058 | Sleep apnea | 28236 | 1266 | 0.123430938 | 0.066373 | 0.180489 | 2.24E-05 | 0.00020964 |
| VAT_Neg_pPS | cqhe058 | Sleep apnea | 28236 | 1266 | 0.027345256 | -0.02985 | 0.084542 | 0.348738943 | 0.689288995 |
| Bilirubin_pPS | cqhe058 | Sleep apnea | 28236 | 1266 | -0.005511697 | -0.06734 | 0.056319 | 0.861304011 | 0.932653041 |
| Proinsulin_pPS | cqhe058 | Sleep apnea | 28236 | 1266 | 0.00647527 | -0.05032 | 0.063275 | 0.823195058 | 0.928523428 |
| Hyper_Insulin_pPS | cqhe058 | Sleep apnea | 28236 | 1266 | 0.043167637 | -0.01401 | 0.100346 | 0.138951008 | 0.393867342 |
| Beta_Cell_1_pPS | cqhe058 | Sleep apnea | 28236 | 1266 | 0.012252756 | -0.04497 | 0.069474 | 0.674712203 | 0.872027982 |
| Beta_Cell_2_pPS | cqhe058 | Sleep apnea | 28236 | 1266 | -0.001806536 | -0.05897 | 0.055362 | 0.95061411 | 0.963460247 |
| Total_GRS | cqhe058 | Sleep apnea | 28236 | 1266 | 0.052744978 | -0.00428 | 0.109766 | 0.069833199 | 0.249404284 |
| Cholesterol_pPS | cqhe069 | Rheumatic disease | 28236 | 1053 | -0.081286909 | -0.14335 | -0.01922 | 0.010257418 | 0.048081648 |
| Lipodystrophy_1_pPS | cqhe069 | Rheumatic disease | 28236 | 1053 | 0.015661157 | -0.04642 | 0.077743 | 0.621003306 | 0.86738823 |
| Lipodystrophy_2_pPS | cqhe069 | Rheumatic disease | 28236 | 1053 | -0.05031943 | -0.11262 | 0.011985 | 0.113432212 | 0.347241466 |
| Liver-Lipid_pPS | cqhe069 | Rheumatic disease | 28236 | 1053 | -0.054292516 | -0.11662 | 0.008036 | 0.087774857 | 0.299232467 |
| ASAT_Pos_pPS | cqhe069 | Rheumatic disease | 28236 | 1053 | 0.032466285 | -0.0295 | 0.094432 | 0.304462652 | 0.62560819 |
| ALP_Neg_pPS | cqhe069 | Rheumatic disease | 28236 | 1053 | 0.021950235 | -0.04014 | 0.084044 | 0.488401247 | 0.779363691 |
| SHBG-LpA_pPS | cqhe069 | Rheumatic disease | 28236 | 1053 | 0.015587881 | -0.04661 | 0.077784 | 0.623275877 | 0.86738823 |
| Obesity_pPS | cqhe069 | Rheumatic disease | 28236 | 1053 | -0.007641391 | -0.0697 | 0.054417 | 0.809296835 | 0.928523428 |
| VAT_Neg_pPS | cqhe069 | Rheumatic disease | 28236 | 1053 | 0.003474189 | -0.05863 | 0.065574 | 0.912686373 | 0.950714972 |
| Bilirubin_pPS | cqhe069 | Rheumatic disease | 28236 | 1053 | 0.063937381 | -0.00253 | 0.130402 | 0.059372438 | 0.227767942 |
| Proinsulin_pPS | cqhe069 | Rheumatic disease | 28236 | 1053 | 0.021082878 | -0.04049 | 0.082659 | 0.502179919 | 0.791154679 |
| Hyper_Insulin_pPS | cqhe069 | Rheumatic disease | 28236 | 1053 | -0.025944642 | -0.08796 | 0.036069 | 0.412219161 | 0.736514214 |
| Beta_Cell_1_pPS | cqhe069 | Rheumatic disease | 28236 | 1053 | 0.025426658 | -0.03683 | 0.08768 | 0.423410057 | 0.738505913 |
| Beta_Cell_2_pPS | cqhe069 | Rheumatic disease | 28236 | 1053 | -0.035253564 | -0.09744 | 0.026931 | 0.266505471 | 0.563039727 |
| Total_GRS | cqhe069 | Rheumatic disease | 28236 | 1053 | -0.014365673 | -0.07639 | 0.047663 | 0.649884422 | 0.872027982 |
| Cholesterol_pPS | cqhe073 | Cancer | 28236 | 1675 | -0.03552467 | -0.08487 | 0.01382 | 0.158237533 | 0.402298812 |
| Lipodystrophy_1_pPS | cqhe073 | Cancer | 28236 | 1675 | 0.003104564 | -0.04672 | 0.052932 | 0.902806102 | 0.946999408 |
| Lipodystrophy_2_pPS | cqhe073 | Cancer | 28236 | 1675 | -0.034536051 | -0.08455 | 0.015476 | 0.175910216 | 0.438241161 |
| Liver-Lipid_pPS | cqhe073 | Cancer | 28236 | 1675 | -0.023829414 | -0.07386 | 0.0262 | 0.350538458 | 0.689288995 |
| ASAT_Pos_pPS | cqhe073 | Cancer | 28236 | 1675 | -0.034186695 | -0.08396 | 0.015584 | 0.178218072 | 0.438241161 |
| ALP_Neg_pPS | cqhe073 | Cancer | 28236 | 1675 | -0.012443828 | -0.06227 | 0.037386 | 0.624519526 | 0.86738823 |
| SHBG-LpA_pPS | cqhe073 | Cancer | 28236 | 1675 | -0.008959179 | -0.05884 | 0.040924 | 0.724825758 | 0.90769938 |
| Obesity_pPS | cqhe073 | Cancer | 28236 | 1675 | 0.015506017 | -0.0342 | 0.065215 | 0.540943584 | 0.819611492 |
| VAT_Neg_pPS | cqhe073 | Cancer | 28236 | 1675 | -0.008618095 | -0.05845 | 0.041213 | 0.734635952 | 0.910705726 |

**Table S15: Associations between pPSs and health history in SCAPIS**

| PPS | Health_History | Health_History_Heading | Sample_Size | Cases (Positive I | coefficient | CI 2.5% | CI 97.5% | p | fdr_p_adjusted |
|---|---|---|---|---|---|---|---|---|---|
| Bilirubin_pPS | cqhe073 | Cancer | 28236 | 1675 | 0.016429184 | -0.03723 | 0.070089 | 0.548448604 | 0.822672905 |
| Proinsulin_pPS | cqhe073 | Cancer | 28236 | 1675 | -0.004180171 | -0.05373 | 0.04537 | 0.86867082 | 0.932653041 |
| Hyper_Insulin_pPS | cqhe073 | Cancer | 28236 | 1675 | 0.040879522 | -0.00897 | 0.090728 | 0.107985059 | 0.337453309 |
| Beta_Cell_1_pPS | cqhe073 | Cancer | 28236 | 1675 | -0.010506436 | -0.06043 | 0.039419 | 0.680002432 | 0.872027982 |
| Beta_Cell_2_pPS | cqhe073 | Cancer | 28236 | 1675 | -0.03157007 | -0.08147 | 0.018331 | 0.214984765 | 0.481309175 |
| Total_GRS | cqhe073 | Cancer | 28236 | 1675 | -0.03746712 | -0.0872 | 0.012265 | 0.139788128 | 0.393867342 |

**Table S16: Associations between pPSs, pMMSs and T2D-related disease outcomes in UKBB**

| Score_type | Biological_Process | Outcome | Outcome_Heading | Model_T | Sample_Size | Events | estimate | conf_low | conf_high | p | fdr_p_adjusted |
|---|---|---|---|---|---|---|---|---|---|---|---|
| genetic | Cholesterol | dm2 | type_2_diabetes | cox | 450714 | 29809 | 1.052067824 | 1.039824476 | 1.06445533 | 1.91E-17 | 9.57E-17 |
| biochemistry | Cholesterol | dm2 | type_2_diabetes | cox | 430723 | 28393 | 1.579690355 | 1.561734489 | 1.597852666 | 0 | 0 |
| metabolomics | Cholesterol | dm2 | type_2_diabetes | cox | 253406 | 17345 | 0.671787764 | 0.661767382 | 0.681959873 | 0 | 0 |
| proteomics | Cholesterol | dm2 | type_2_diabetes | cox | 40551 | 2791 | 1.014556953 | 0.977614298 | 1.052895619 | 0.445066419 | 0.856875603 |
| merged_omics | Cholesterol | dm2 | type_2_diabetes | cox | 21748 | 1504 | 1.239318833 | 1.178466457 | 1.303313437 | 6.67E-17 | 2.70E-16 |
| genetic | Lipodystrophy_1 | dm2 | type_2_diabetes | cox | 450714 | 29809 | 1.18651728 | 1.173069387 | 1.200119337 | 4.42E-190 | 1.11E-188 |
| biochemistry | Lipodystrophy_1 | dm2 | type_2_diabetes | cox | 430723 | 28393 | 1.756854636 | 1.736228114 | 1.777726203 | 0 | 0 |
| metabolomics | Lipodystrophy_1 | dm2 | type_2_diabetes | cox | 253406 | 17345 | 1.405787135 | 1.382430464 | 1.429538425 | 0 | 0 |
| proteomics | Lipodystrophy_1 | dm2 | type_2_diabetes | cox | 40551 | 2791 | 1.022261308 | 0.98503932 | 1.060889814 | 0.244644397 | 0.762749344 |
| merged_omics | Lipodystrophy_1 | dm2 | type_2_diabetes | cox | 21748 | 1504 | 1.565973868 | 1.482602173 | 1.654033833 | 4.27E-58 | 4.32E-57 |
| genetic | Lipodystrophy_2 | dm2 | type_2_diabetes | cox | 450714 | 29809 | 1.120590394 | 1.10799798 | 1.133325921 | 8.54E-87 | 1.17E-85 |
| biochemistry | Lipodystrophy_2 | dm2 | type_2_diabetes | cox | 430723 | 28393 | 2.057045314 | 2.034399454 | 2.079943255 | 0 | 0 |
| proteomics | Lipodystrophy_2 | dm2 | type_2_diabetes | cox | 40551 | 2791 | 1.013749091 | 0.976905725 | 1.05198198 | 0.469700122 | 0.856875603 |
| merged_omics | Lipodystrophy_2 | dm2 | type_2_diabetes | cox | 21748 | 1504 | 1.680728072 | 1.597343941 | 1.768464999 | 5.53E-89 | 8.96E-88 |
| genetic | Liver-Lipid | dm2 | type_2_diabetes | cox | 450714 | 29809 | 1.057117294 | 1.04509246 | 1.069280486 | 1.79E-21 | 9.97E-21 |
| biochemistry | Liver-Lipid | dm2 | type_2_diabetes | cox | 430723 | 28393 | 0.86529143 | 0.854904318 | 0.875804746 | 6.20E-122 | 1.05E-121 |
| metabolomics | Liver-Lipid | dm2 | type_2_diabetes | cox | 253406 | 17345 | 1.689478896 | 1.666950691 | 1.71231156 | 0 | 0 |
| proteomics | Liver-Lipid | dm2 | type_2_diabetes | cox | 40551 | 2791 | 1.018789497 | 0.981663809 | 1.057319247 | 0.325670657 | 0.856875603 |
| merged_omics | Liver-Lipid | dm2 | type_2_diabetes | cox | 21748 | 1504 | 0.715474333 | 0.680588065 | 0.752148837 | 2.29E-39 | 1.55E-38 |
| genetic | ASAT_Pos | dm2 | type_2_diabetes | cox | 450714 | 29809 | 1.124109149 | 1.111442928 | 1.136919717 | 4.78E-91 | 7.17E-90 |
| biochemistry | ASAT_Pos | dm2 | type_2_diabetes | cox | 430723 | 28393 | 2.152652451 | 2.12821567 | 2.177369822 | 0 | 0 |
| proteomics | ASAT_Pos | dm2 | type_2_diabetes | cox | 40551 | 2791 | 1.031342349 | 0.993833392 | 1.070266958 | 0.1025237 | 0.56782357 |
| merged_omics | ASAT_Pos | dm2 | type_2_diabetes | cox | 21748 | 1504 | 2.077076894 | 1.974249302 | 2.185260199 | 3.59E-175 | 9.70E-174 |
| genetic | ALP_Neg | dm2 | type_2_diabetes | cox | 450714 | 29809 | 1.058638602 | 1.046719719 | 1.070693203 | 5.95E-23 | 3.43E-22 |
| biochemistry | ALP_Neg | dm2 | type_2_diabetes | cox | 430723 | 28393 | 2.668351993 | 2.638178215 | 2.698870879 | 0 | 0 |
| metabolomics | ALP_Neg | dm2 | type_2_diabetes | cox | 253406 | 17345 | 0.759975852 | 0.74794637 | 0.77219881 | 3.32E-249 | 3.32E-248 |
| proteomics | ALP_Neg | dm2 | type_2_diabetes | cox | 40551 | 2791 | 0.983603737 | 0.947729118 | 1.020836326 | 0.383143409 | 0.856875603 |
| merged_omics | ALP_Neg | dm2 | type_2_diabetes | cox | 21748 | 1504 | 1.207083295 | 1.147794533 | 1.269434588 | 2.40E-13 | 7.78E-13 |
| genetic | SHBG-LpA | dm2 | type_2_diabetes | cox | 450714 | 29809 | 1.041824043 | 1.030079665 | 1.053702323 | 1.40E-12 | 5.54E-12 |
| biochemistry | SHBG-LpA | dm2 | type_2_diabetes | cox | 430723 | 28393 | 1.232282246 | 1.21798378 | 1.246748567 | 1.54E-269 | 3.84E-269 |
| genetic | Obesity | dm2 | type_2_diabetes | cox | 450714 | 29809 | 1.179078815 | 1.165795749 | 1.192513228 | 1.22E-178 | 2.61E-177 |
| biochemistry | Obesity | dm2 | type_2_diabetes | cox | 430723 | 28393 | 3.018445343 | 2.98210832 | 3.055225133 | 0 | 0 |
| genetic | VAT_Neg | dm2 | type_2_diabetes | cox | 450714 | 29809 | 1.113168293 | 1.10054328 | 1.125938135 | 8.68E-76 | 1.09E-74 |
| biochemistry | VAT_Neg | dm2 | type_2_diabetes | cox | 430723 | 28393 | 2.518567293 | 2.490951802 | 2.546488937 | 0 | 0 |
| genetic | Bilirubin | dm2 | type_2_diabetes | cox | 450714 | 29809 | 1.008042952 | 0.996682529 | 1.019532864 | 0.165948285 | 0.210951209 |
| genetic | Proinsulin | dm2 | type_2_diabetes | cox | 450714 | 29809 | 1.222128167 | 1.208284884 | 1.236130053 | 5.16E-261 | 1.94E-259 |
| biochemistry | Proinsulin | dm2 | type_2_diabetes | cox | 430723 | 28393 | 3.876364727 | 3.836031892 | 3.91712163 | 0 | 0 |
| proteomics | Proinsulin | dm2 | type_2_diabetes | cox | 40551 | 2791 | 0.962749772 | 0.92774591 | 0.99907433 | 0.044535966 | 0.356287728 |
| merged_omics | Proinsulin | dm2 | type_2_diabetes | cox | 21748 | 1504 | 1.510616148 | 1.436150287 | 1.58894314 | 1.40E-57 | 1.26E-56 |
| genetic | Hyper_Insulin | dm2 | type_2_diabetes | cox | 450714 | 29809 | 1.133937822 | 1.121129334 | 1.146892643 | 2.70E-104 | 5.07E-103 |
| biochemistry | Hyper_Insulin | dm2 | type_2_diabetes | cox | 430723 | 28393 | 2.17072876 | 2.147873417 | 2.193827305 | 0 | 0 |
| genetic | Beta_Cell_1 | dm2 | type_2_diabetes | cox | 450714 | 29809 | 1.240566957 | 1.226543443 | 1.254750807 | 2.48E-302 | 1.24E-300 |
| biochemistry | Beta_Cell_1 | dm2 | type_2_diabetes | cox | 430723 | 28393 | 3.22371927 | 3.190596046 | 3.257186364 | 0 | 0 |
| genetic | Beta_Cell_2 | dm2 | type_2_diabetes | cox | 450714 | 29809 | 1.25921501 | 1.245089481 | 1.273500792 | 0 | 0 |
| biochemistry | Beta_Cell_2 | dm2 | type_2_diabetes | cox | 430723 | 28393 | 3.248471941 | 3.213876802 | 3.283439472 | 0 | 0 |
| proteomics | Beta_Cell_2 | dm2 | type_2_diabetes | cox | 40551 | 2791 | 0.99522434 | 0.958831862 | 1.032998094 | 0.801142728 | 0.968997537 |
| merged_omics | Beta_Cell_2 | dm2 | type_2_diabetes | cox | 21748 | 1504 | 2.248047006 | 2.136228928 | 2.365718053 | 1.30E-212 | 5.28E-211 |
| genetic | Total_GRS | dm2 | type_2_diabetes | cox | 450714 | 29809 | 1.583653173 | 1.565661842 | 1.601851246 | 0 | 0 |
| biochemistry | Total_GRS | dm2 | type_2_diabetes | cox | 430723 | 28393 | 3.375922767 | 3.344296813 | 3.407847797 | 0 | 0 |
| merged_omics | Total_GRS | dm2 | type_2_diabetes | cox | 21748 | 1504 | 3.444381727 | 3.282121953 | 3.614663214 | 0 | 0 |
| genetic | Cholesterol | acmort | all_cause_mortality | cox | 458905 | 41085 | 0.941353547 | 0.932547821 | 0.950242422 | 2.01E-36 | 1.68E-35 |
| biochemistry | Cholesterol | acmort | all_cause_mortality | cox | 438513 | 39213 | 1.198072202 | 1.186360404 | 1.209899619 | 1.12E-284 | 2.95E-284 |
| metabolomics | Cholesterol | acmort | all_cause_mortality | cox | 258153 | 23285 | 0.973383979 | 0.960269735 | 0.986677321 | 9.70E-05 | 0.00011756 |
| proteomics | Cholesterol | acmort | all_cause_mortality | cox | 41377 | 4499 | 1.022852589 | 0.993378353 | 1.053201346 | 0.129860873 | 0.623332191 |
| merged_omics | Cholesterol | acmort | all_cause_mortality | cox | 22217 | 2418 | 1.131133233 | 1.087205954 | 1.176835342 | 1.08E-09 | 2.82E-09 |
| genetic | Lipodystrophy_1 | acmort | all_cause_mortality | cox | 458905 | 41085 | 1.026705804 | 1.016819926 | 1.036687796 | 9.35E-08 | 2.50E-07 |
| biochemistry | Lipodystrophy_1 | acmort | all_cause_mortality | cox | 438513 | 39213 | 0.908453799 | 0.899330789 | 0.917669355 | 1.40E-77 | 2.04E-77 |
| metabolomics | Lipodystrophy_1 | acmort | all_cause_mortality | cox | 258153 | 23285 | 0.95214998 | 0.939100454 | 0.96538084 | 3.31E-12 | 5.09E-12 |
| proteomics | Lipodystrophy_1 | acmort | all_cause_mortality | cox | 41377 | 4499 | 0.995041638 | 0.966335481 | 1.024600546 | 0.739276599 | 0.968997537 |
| merged_omics | Lipodystrophy_1 | acmort | all_cause_mortality | cox | 22217 | 2418 | 0.961203173 | 0.921785119 | 1.002306851 | 0.064004787 | 0.078551329 |
| genetic | Lipodystrophy_2 | acmort | all_cause_mortality | cox | 458905 | 41085 | 1.01182546 | 1.002074402 | 1.021671403 | 0.017339789 | 0.026814106 |
| biochemistry | Lipodystrophy_2 | acmort | all_cause_mortality | cox | 438513 | 39213 | 1.07114042 | 1.060419152 | 1.081970085 | 6.92E-41 | 9.22E-41 |
| proteomics | Lipodystrophy_2 | acmort | all_cause_mortality | cox | 41377 | 4499 | 0.983968092 | 0.955661732 | 1.013112876 | 0.277821718 | 0.800126547 |
| merged_omics | Lipodystrophy_2 | acmort | all_cause_mortality | cox | 22217 | 2418 | 1.113961753 | 1.070003173 | 1.159726268 | 1.49E-07 | 3.09E-07 |
| genetic | Liver-Lipid | acmort | all_cause_mortality | cox | 458905 | 41085 | 1.010474199 | 1.000730881 | 1.02031238 | 0.03504871 | 0.05104181 |
| biochemistry | Liver-Lipid | acmort | all_cause_mortality | cox | 438513 | 39213 | 0.87787023 | 0.868841914 | 0.886992361 | 1.16E-134 | 2.09E-134 |
| metabolomics | Liver-Lipid | acmort | all_cause_mortality | cox | 258153 | 23285 | 1.205628979 | 1.190542777 | 1.220906349 | 2.89E-186 | 1.93E-185 |
| proteomics | Liver-Lipid | acmort | all_cause_mortality | cox | 41377 | 4499 | 1.006678295 | 0.977726479 | 1.036487412 | 0.654829265 | 0.963492845 |
| merged_omics | Liver-Lipid | acmort | all_cause_mortality | cox | 22217 | 2418 | 0.951952464 | 0.914846238 | 0.99056372 | 0.015207981 | 0.021611342 |
| genetic | ASAT_Pos | acmort | all_cause_mortality | cox | 458905 | 41085 | 1.009878947 | 1.000163867 | 1.019688395 | 0.046237048 | 0.064818291 |
| biochemistry | ASAT_Pos | acmort | all_cause_mortality | cox | 438513 | 39213 | 1.020917287 | 1.010708994 | 1.031228685 | 5.40E-05 | 5.64E-05 |
| proteomics | ASAT_Pos | acmort | all_cause_mortality | cox | 41377 | 4499 | 1.011511869 | 0.982432093 | 1.041452401 | 0.441842004 | 0.856875603 |
| merged_omics | ASAT_Pos | acmort | all_cause_mortality | cox | 22217 | 2418 | 1.181258356 | 1.134433459 | 1.230015999 | 6.90E-16 | 2.66E-15 |
| genetic | ALP_Neg | acmort | all_cause_mortality | cox | 458905 | 41085 | 1.016332315 | 1.006541652 | 1.026218211 | 0.001037218 | 0.001809102 |
| biochemistry | ALP_Neg | acmort | all_cause_mortality | cox | 438513 | 39213 | 1.154292695 | 1.142757941 | 1.165943879 | 1.51E-172 | 3.01E-172 |
| metabolomics | ALP_Neg | acmort | all_cause_mortality | cox | 258153 | 23285 | 0.922695653 | 0.910053118 | 0.935513819 | 2.96E-30 | 5.38E-30 |
| proteomics | ALP_Neg | acmort | all_cause_mortality | cox | 41377 | 4499 | 0.971999338 | 0.943932419 | 1.000900799 | 0.057463306 | 0.413735801 |
| merged_omics | ALP_Neg | acmort | all_cause_mortality | cox | 22217 | 2418 | 0.981381901 | 0.942879941 | 1.021456068 | 0.357382889 | 0.391189379 |
| genetic | SHBG-LpA | acmort | all_cause_mortality | cox | 458905 | 41085 | 1.0021433 | 0.992488637 | 1.011891882 | 0.664668066 | 0.707093687 |
| biochemistry | SHBG-LpA | acmort | all_cause_mortality | cox | 438513 | 39213 | 1.022528929 | 1.012428148 | 1.032730483 | 1.09E-05 | 1.16E-05 |
| genetic | Obesity | acmort | all_cause_mortality | cox | 458905 | 41085 | 1.037541442 | 1.027557691 | 1.047622195 | 7.99E-14 | 3.24E-13 |
| biochemistry | Obesity | acmort | all_cause_mortality | cox | 438513 | 39213 | 1.289031861 | 1.275994022 | 1.302202918 | 0 | 0 |
| genetic | VAT_Neg | acmort | all_cause_mortality | cox | 458905 | 41085 | 0.990611522 | 0.981062494 | 1.000253495 | 0.056299199 | 0.077475962 |
| biochemistry | VAT_Neg | acmort | all_cause_mortality | cox | 438513 | 39213 | 1.05834081 | 1.047981073 | 1.068802957 | 1.34E-29 | 1.66E-29 |

**Table S16: Associations between pPSs, pMMSs and T2D-related disease outcomes in UKBB**

| Score_type | Biological_Process | Outcome | Outcome_Heading | Model_T | Sample_Size | Events | estimate | conf_low | conf_high | p | fdr_p_adjusted |
|---|---|---|---|---|---|---|---|---|---|---|---|
| genetic | Bilirubin | acmort | all_cause_mortality | cox | 458905 | 41085 | 0.997478478 | 0.987862901 | 1.007187651 | 0.609456105 | 0.662452288 |
| genetic | Proinsulin | acmort | all_cause_mortality | cox | 458905 | 41085 | 1.010512569 | 1.000777421 | 1.020342417 | 0.034230896 | 0.050339554 |
| biochemistry | Proinsulin | acmort | all_cause_mortality | cox | 438513 | 39213 | 1.223361008 | 1.210989797 | 1.235858601 | 0 | 0 |
| proteomics | Proinsulin | acmort | all_cause_mortality | cox | 41377 | 4499 | 0.966434896 | 0.938669065 | 0.99502204 | 0.021702477 | 0.356287728 |
| merged_omics | Proinsulin | acmort | all_cause_mortality | cox | 22217 | 2418 | 0.993592243 | 0.954584669 | 1.034193799 | 0.753070907 | 0.762484294 |
| genetic | Hyper_Insulin | acmort | all_cause_mortality | cox | 458905 | 41085 | 1.031262156 | 1.021343259 | 1.041277381 | 4.30E-10 | 1.57E-09 |
| biochemistry | Hyper_Insulin | acmort | all_cause_mortality | cox | 438513 | 39213 | 1.023157205 | 1.012981534 | 1.033435093 | 7.15E-06 | 7.70E-06 |
| genetic | Beta_Cell_1 | acmort | all_cause_mortality | cox | 458905 | 41085 | 1.00989195 | 1.000173942 | 1.01970438 | 0.046015056 | 0.064818291 |
| biochemistry | Beta_Cell_1 | acmort | all_cause_mortality | cox | 438513 | 39213 | 1.211342088 | 1.199504481 | 1.223296518 | 2.233e-320 | 6.652e-320 |
| genetic | Beta_Cell_2 | acmort | all_cause_mortality | cox | 458905 | 41085 | 0.99553478 | 0.985951386 | 1.005211324 | 0.364515685 | 0.420595021 |
| biochemistry | Beta_Cell_2 | acmort | all_cause_mortality | cox | 438513 | 39213 | 1.22276066 | 1.210810171 | 1.234829099 | 0 | 0 |
| proteomics | Beta_Cell_2 | acmort | all_cause_mortality | cox | 41377 | 4499 | 0.981786192 | 0.953295993 | 1.011127848 | 0.221161267 | 0.72380051 |
| merged_omics | Beta_Cell_2 | acmort | all_cause_mortality | cox | 22217 | 2418 | 1.067760251 | 1.026153048 | 1.111054493 | 0.001224538 | 0.001905991 |
| genetic | Total_GRS | acmort | all_cause_mortality | cox | 458905 | 41085 | 1.029848492 | 1.019931472 | 1.039861937 | 2.56E-09 | 8.35E-09 |
| biochemistry | Total_GRS | acmort | all_cause_mortality | cox | 438513 | 39213 | 1.212350143 | 1.200888192 | 1.223921493 | 0 | 0 |
| merged_omics | Total_GRS | acmort | all_cause_mortality | cox | 22217 | 2418 | 1.179347 | 1.132437573 | 1.228199576 | 1.64E-15 | 5.78E-15 |
| genetic | Cholesterol | cvdmort | cardiovascular_disease_mortality | cox | 458905 | 8491 | 0.947079229 | 0.92770753 | 0.966855434 | 2.51E-07 | 6.61E-07 |
| biochemistry | Cholesterol | cvdmort | cardiovascular_disease_mortality | cox | 438513 | 8107 | 1.258776621 | 1.23196686 | 1.28616981 | 1.78E-97 | 2.76E-97 |
| metabolomics | Cholesterol | cvdmort | cardiovascular_disease_mortality | cox | 258153 | 4858 | 0.931247201 | 0.904133694 | 0.959173797 | 2.30E-06 | 2.97E-06 |
| proteomics | Cholesterol | cvdmort | cardiovascular_disease_mortality | cox | 41377 | 929 | 1.038130712 | 0.973446494 | 1.10711311 | 0.254249781 | 0.762749344 |
| merged_omics | Cholesterol | cvdmort | cardiovascular_disease_mortality | cox | 22217 | 501 | 1.263992077 | 1.159134207 | 1.378335623 | 1.14E-07 | 2.50E-07 |
| genetic | Lipodystrophy_1 | cvdmort | cardiovascular_disease_mortality | cox | 458905 | 8491 | 1.042778799 | 1.020816993 | 1.06521309 | 0.000114715 | 0.000235715 |
| biochemistry | Lipodystrophy_1 | cvdmort | cardiovascular_disease_mortality | cox | 438513 | 8107 | 0.92780055 | 0.907663078 | 0.948384793 | 2.18E-11 | 2.44E-11 |
| metabolomics | Lipodystrophy_1 | cvdmort | cardiovascular_disease_mortality | cox | 258153 | 4858 | 1.011496338 | 0.980922449 | 1.043023169 | 0.465416827 | 0.489912449 |
| proteomics | Lipodystrophy_1 | cvdmort | cardiovascular_disease_mortality | cox | 41377 | 929 | 0.975319493 | 0.914370238 | 1.040331448 | 0.447826367 | 0.856875603 |
| merged_omics | Lipodystrophy_1 | cvdmort | cardiovascular_disease_mortality | cox | 22217 | 501 | 0.98133131 | 0.89455388 | 1.076526703 | 0.68993235 | 0.716468209 |
| genetic | Lipodystrophy_2 | cvdmort | cardiovascular_disease_mortality | cox | 458905 | 8491 | 1.017863328 | 0.996427216 | 1.039760595 | 0.103014152 | 0.134269076 |
| biochemistry | Lipodystrophy_2 | cvdmort | cardiovascular_disease_mortality | cox | 438513 | 8107 | 1.084895571 | 1.061367466 | 1.108945242 | 3.24E-13 | 3.69E-13 |
| proteomics | Lipodystrophy_2 | cvdmort | cardiovascular_disease_mortality | cox | 41377 | 929 | 1.017933974 | 0.954928402 | 1.085096614 | 0.585571634 | 0.958208128 |
| merged_omics | Lipodystrophy_2 | cvdmort | cardiovascular_disease_mortality | cox | 22217 | 501 | 1.22589787 | 1.122839881 | 1.338414865 | 5.47E-06 | 1.03E-05 |
| genetic | Liver-Lipid | cvdmort | cardiovascular_disease_mortality | cox | 458905 | 8491 | 1.008225448 | 0.98695864 | 1.029950511 | 0.451371934 | 0.50906609 |
| biochemistry | Liver-Lipid | cvdmort | cardiovascular_disease_mortality | cox | 438513 | 8107 | 0.905152329 | 0.884830794 | 0.925940581 | 7.85E-18 | 9.16E-18 |
| metabolomics | Liver-Lipid | cvdmort | cardiovascular_disease_mortality | cox | 258153 | 4858 | 1.245253374 | 1.211944671 | 1.279477522 | 1.27E-56 | 3.92E-56 |
| proteomics | Liver-Lipid | cvdmort | cardiovascular_disease_mortality | cox | 41377 | 929 | 1.01188087 | 0.948765643 | 1.079194744 | 0.719268573 | 0.968997537 |
| merged_omics | Liver-Lipid | cvdmort | cardiovascular_disease_mortality | cox | 22217 | 501 | 0.924169976 | 0.84691261 | 1.008474942 | 0.076639744 | 0.092654019 |
| genetic | ASAT_Pos | cvdmort | cardiovascular_disease_mortality | cox | 458905 | 8491 | 0.995390384 | 0.974449713 | 1.016781065 | 0.670173522 | 0.707929777 |
| biochemistry | ASAT_Pos | cvdmort | cardiovascular_disease_mortality | cox | 438513 | 8107 | 1.004162578 | 0.982464329 | 1.026340044 | 0.709371053 | 0.709371053 |
| proteomics | ASAT_Pos | cvdmort | cardiovascular_disease_mortality | cox | 41377 | 929 | 1.020141117 | 0.956688018 | 1.087802795 | 0.542782695 | 0.908845442 |
| merged_omics | ASAT_Pos | cvdmort | cardiovascular_disease_mortality | cox | 22217 | 501 | 1.359279775 | 1.245431743 | 1.48353494 | 6.07E-12 | 1.76E-11 |
| genetic | ALP_Neg | cvdmort | cardiovascular_disease_mortality | cox | 458905 | 8491 | 1.038264981 | 1.016446597 | 1.060551704 | 0.000529341 | 0.000992514 |
| biochemistry | ALP_Neg | cvdmort | cardiovascular_disease_mortality | cox | 438513 | 8107 | 1.295057209 | 1.267416491 | 1.323300734 | 5.23E-122 | 8.94E-122 |
| metabolomics | ALP_Neg | cvdmort | cardiovascular_disease_mortality | cox | 258153 | 4858 | 0.891712848 | 0.865329945 | 0.918900134 | 7.45E-14 | 1.19E-13 |
| proteomics | ALP_Neg | cvdmort | cardiovascular_disease_mortality | cox | 41377 | 929 | 0.911722856 | 0.854482474 | 0.972797677 | 0.005211381 | 0.326402977 |
| merged_omics | ALP_Neg | cvdmort | cardiovascular_disease_mortality | cox | 22217 | 501 | 0.940291816 | 0.860851118 | 1.027063426 | 0.171610714 | 0.198578112 |
| genetic | SHBG-LpA | cvdmort | cardiovascular_disease_mortality | cox | 458905 | 8491 | 1.019538503 | 0.998078737 | 1.041459677 | 0.074616481 | 0.099048426 |
| biochemistry | SHBG-LpA | cvdmort | cardiovascular_disease_mortality | cox | 438513 | 8107 | 1.152105603 | 1.127430725 | 1.177320514 | 1.29E-37 | 1.67E-37 |
| genetic | Obesity | cvdmort | cardiovascular_disease_mortality | cox | 458905 | 8491 | 1.069532573 | 1.047023936 | 1.092525095 | 5.85E-10 | 2.09E-09 |
| biochemistry | Obesity | cvdmort | cardiovascular_disease_mortality | cox | 438513 | 8107 | 1.417621185 | 1.386370369 | 1.449576441 | 9.06E-207 | 2.01E-206 |
| genetic | VAT_Neg | cvdmort | cardiovascular_disease_mortality | cox | 458905 | 8491 | 1.001936893 | 0.980785622 | 1.023544305 | 0.858915827 | 0.882447767 |
| biochemistry | VAT_Neg | cvdmort | cardiovascular_disease_mortality | cox | 438513 | 8107 | 1.137758284 | 1.113808779 | 1.162222759 | 1.33E-32 | 1.68E-32 |
| genetic | Bilirubin | cvdmort | cardiovascular_disease_mortality | cox | 458905 | 8491 | 0.997714721 | 0.976678581 | 1.019203947 | 0.833329991 | 0.862065508 |
| genetic | Proinsulin | cvdmort | cardiovascular_disease_mortality | cox | 458905 | 8491 | 1.023967369 | 1.002398066 | 1.046000794 | 0.029219527 | 0.043395337 |
| biochemistry | Proinsulin | cvdmort | cardiovascular_disease_mortality | cox | 438513 | 8107 | 1.412668864 | 1.382571062 | 1.443421878 | 5.31E-217 | 1.22E-216 |
| proteomics | Proinsulin | cvdmort | cardiovascular_disease_mortality | cox | 41377 | 929 | 0.918114873 | 0.861051643 | 0.978959771 | 0.009066749 | 0.326402977 |
| merged_omics | Proinsulin | cvdmort | cardiovascular_disease_mortality | cox | 22217 | 501 | 1.019306055 | 0.933371014 | 1.113153096 | 0.670443188 | 0.705271406 |
| genetic | Hyper_Insulin | cvdmort | cardiovascular_disease_mortality | cox | 458905 | 8491 | 1.059996398 | 1.03769879 | 1.082773127 | 7.80E-08 | 2.13E-07 |
| biochemistry | Hyper_Insulin | cvdmort | cardiovascular_disease_mortality | cox | 438513 | 8107 | 1.083939849 | 1.060896158 | 1.107484072 | 1.95E-13 | 2.24E-13 |
| genetic | Beta_Cell_1 | cvdmort | cardiovascular_disease_mortality | cox | 458905 | 8491 | 1.021574021 | 1.000066851 | 1.043543718 | 0.049280393 | 0.06844499 |
| biochemistry | Beta_Cell_1 | cvdmort | cardiovascular_disease_mortality | cox | 438513 | 8107 | 1.353441651 | 1.325597648 | 1.381870512 | 4.14E-179 | 8.52E-179 |
| genetic | Beta_Cell_2 | cvdmort | cardiovascular_disease_mortality | cox | 458905 | 8491 | 1.021920812 | 1.000439345 | 1.04386353 | 0.045443688 | 0.064818291 |
| biochemistry | Beta_Cell_2 | cvdmort | cardiovascular_disease_mortality | cox | 438513 | 8107 | 1.371342193 | 1.343162964 | 1.400112615 | 2.85E-195 | 6.04E-195 |
| proteomics | Beta_Cell_2 | cvdmort | cardiovascular_disease_mortality | cox | 41377 | 929 | 0.993488267 | 0.931074569 | 1.060085807 | 0.843552494 | 0.968997537 |
| merged_omics | Beta_Cell_2 | cvdmort | cardiovascular_disease_mortality | cox | 22217 | 501 | 1.209997666 | 1.110640231 | 1.318243577 | 1.30E-05 | 2.34E-05 |
| genetic | Total_GRS | cvdmort | cardiovascular_disease_mortality | cox | 458905 | 8491 | 1.064986514 | 1.042556894 | 1.087898686 | 6.73E-09 | 2.15E-08 |
| biochemistry | Total_GRS | cvdmort | cardiovascular_disease_mortality | cox | 438513 | 8107 | 1.343767872 | 1.317288488 | 1.370779528 | 3.67E-186 | 7.66E-186 |
| merged_omics | Total_GRS | cvdmort | cardiovascular_disease_mortality | cox | 22217 | 501 | 1.439325685 | 1.322231885 | 1.566789042 | 4.03E-17 | 1.81E-16 |
| genetic | Cholesterol | cad | coronary_artery_disease | cox | 443790 | 19980 | 0.968605889 | 0.955447723 | 0.981945265 | 4.86E-06 | 1.18E-05 |
| biochemistry | Cholesterol | cad | coronary_artery_disease | cox | 424098 | 19120 | 0.970227226 | 0.956504043 | 0.984147297 | 3.20E-05 | 3.40E-05 |
| metabolomics | Cholesterol | cad | coronary_artery_disease | cox | 249499 | 11329 | 0.76976984 | 0.75536109 | 0.784453443 | 3.17E-162 | 1.59E-161 |
| proteomics | Cholesterol | cad | coronary_artery_disease | cox | 39875 | 1905 | 0.979613235 | 0.936612948 | 1.024587683 | 0.368451764 | 0.856875603 |
| merged_omics | Cholesterol | cad | coronary_artery_disease | cox | 21390 | 997 | 0.886253573 | 0.832921409 | 0.943000608 | 0.000137067 | 0.00022658 |
| genetic | Lipodystrophy_1 | cad | coronary_artery_disease | cox | 443790 | 19980 | 1.062657877 | 1.048008607 | 1.077511918 | 9.41E-18 | 4.87E-17 |
| biochemistry | Lipodystrophy_1 | cad | coronary_artery_disease | cox | 424098 | 19120 | 1.179230635 | 1.162467614 | 1.196235382 | 8.68E-113 | 1.43E-112 |
| metabolomics | Lipodystrophy_1 | cad | coronary_artery_disease | cox | 249499 | 11329 | 1.380236667 | 1.351419378 | 1.409668448 | 6.85E-197 | 5.48E-196 |
| proteomics | Lipodystrophy_1 | cad | coronary_artery_disease | cox | 39875 | 1905 | 0.994635958 | 0.950996801 | 1.04027762 | 0.814238364 | 0.968997537 |
| merged_omics | Lipodystrophy_1 | cad | coronary_artery_disease | cox | 21390 | 997 | 1.312223679 | 1.227413729 | 1.402893698 | 1.57E-15 | 5.78E-15 |
| genetic | Lipodystrophy_2 | cad | coronary_artery_disease | cox | 443790 | 19980 | 1.02835581 | 1.014188187 | 1.042721346 | 7.80E-05 | 0.000164778 |
| biochemistry | Lipodystrophy_2 | cad | coronary_artery_disease | cox | 424098 | 19120 | 1.071777309 | 1.056417907 | 1.087360023 | 4.85E-21 | 5.75E-21 |
| proteomics | Lipodystrophy_2 | cad | coronary_artery_disease | cox | 39875 | 1905 | 0.97094474 | 0.92828472 | 1.015565235 | 0.198360188 | 0.680092075 |
| merged_omics | Lipodystrophy_2 | cad | coronary_artery_disease | cox | 21390 | 997 | 1.141039482 | 1.071484068 | 1.215110087 | 3.93E-05 | 6.63E-05 |
| genetic | Liver-Lipid | cad | coronary_artery_disease | cox | 443790 | 19980 | 0.988253515 | 0.974654805 | 1.002041958 | 0.094633376 | 0.124517599 |
| biochemistry | Liver-Lipid | cad | coronary_artery_disease | cox | 424098 | 19120 | 0.877242356 | 0.864371884 | 0.890304468 | 1.44E-67 | 2.05E-67 |
| metabolomics | Liver-Lipid | cad | coronary_artery_disease | cox | 249499 | 11329 | 1.117558381 | 1.097467963 | 1.138016577 | 3.20E-33 | 6.09E-33 |
| proteomics | Liver-Lipid | cad | coronary_artery_disease | cox | 39875 | 1905 | 1.036330146 | 0.990827456 | 1.083922498 | 0.119291813 | 0.613500753 |

**Table S16: Associations between pPSs, pMMSs and T2D-related disease outcomes in UKBB**

| Score_type | Biological_Process | Outcome | Outcome_Heading | Model_T | Sample_Size | Events | estimate | conf_low | conf_high | p | fdr_p_adjusted |
|---|---|---|---|---|---|---|---|---|---|---|---|
| merged_omics | Liver-Lipid | cad | coronary_artery_disease | cox | 21390 | 997 | 0.952682642 | 0.895461042 | 1.013560806 | 0.125080519 | 0.146833653 |
| genetic | ASAT_Pos | cad | coronary_artery_disease | cox | 443790 | 19980 | 1.027907836 | 1.013754106 | 1.042259177 | 9.98E-05 | 0.000207936 |
| biochemistry | ASAT_Pos | cad | coronary_artery_disease | cox | 424098 | 19120 | 1.030186539 | 1.015469088 | 1.045117293 | 5.10E-05 | 5.37E-05 |
| proteomics | ASAT_Pos | cad | coronary_artery_disease | cox | 39875 | 1905 | 1.048936436 | 1.002929757 | 1.097053547 | 0.03681198 | 0.356287728 |
| merged_omics | ASAT_Pos | cad | coronary_artery_disease | cox | 21390 | 997 | 1.283728852 | 1.205920088 | 1.366558018 | 4.90E-15 | 1.65E-14 |
| genetic | ALP_Neg | cad | coronary_artery_disease | cox | 443790 | 19980 | 1.017872664 | 1.003856113 | 1.032084924 | 0.012279119 | 0.020020303 |
| biochemistry | ALP_Neg | cad | coronary_artery_disease | cox | 424098 | 19120 | 1.270882502 | 1.252933213 | 1.289088928 | 2.94E-239 | 7.23E-239 |
| metabolomics | ALP_Neg | cad | coronary_artery_disease | cox | 249499 | 11329 | 0.960503188 | 0.941663647 | 0.979719645 | 6.68E-05 | 8.36E-05 |
| proteomics | ALP_Neg | cad | coronary_artery_disease | cox | 39875 | 1905 | 0.968885929 | 0.92624646 | 1.013488292 | 0.168660254 | 0.680092075 |
| merged_omics | ALP_Neg | cad | coronary_artery_disease | cox | 21390 | 997 | 0.997383692 | 0.937127514 | 1.06151427 | 0.934330799 | 0.934330799 |
| genetic | SHBG-LpA | cad | coronary_artery_disease | cox | 443790 | 19980 | 1.026567358 | 1.01243594 | 1.04089602 | 0.000209228 | 0.000412951 |
| biochemistry | SHBG-LpA | cad | coronary_artery_disease | cox | 424098 | 19120 | 1.217427241 | 1.200281415 | 1.234817993 | 9.33E-163 | 1.79E-162 |
| genetic | Obesity | cad | coronary_artery_disease | cox | 443790 | 19980 | 1.039584007 | 1.025262584 | 1.054105479 | 4.13E-08 | 1.27E-07 |
| biochemistry | Obesity | cad | coronary_artery_disease | cox | 424098 | 19120 | 1.250523498 | 1.232561279 | 1.268747482 | 1.73E-201 | 3.78E-201 |
| genetic | VAT_Neg | cad | coronary_artery_disease | cox | 443790 | 19980 | 1.02360976 | 1.009470311 | 1.037947257 | 0.001008373 | 0.001779482 |
| biochemistry | VAT_Neg | cad | coronary_artery_disease | cox | 424098 | 19120 | 1.215916512 | 1.199153565 | 1.232913789 | 1.05E-167 | 2.07E-167 |
| genetic | Bilirubin | cad | coronary_artery_disease | cox | 443790 | 19980 | 1.004292536 | 0.990473105 | 1.01830478 | 0.544577997 | 0.600637496 |
| genetic | Proinsulin | cad | coronary_artery_disease | cox | 443790 | 19980 | 1.033706656 | 1.019449178 | 1.048163531 | 2.89E-06 | 7.11E-06 |
| biochemistry | Proinsulin | cad | coronary_artery_disease | cox | 424098 | 19120 | 1.300236888 | 1.281858639 | 1.318878629 | 3.89E-286 | 1.05E-285 |
| proteomics | Proinsulin | cad | coronary_artery_disease | cox | 39875 | 1905 | 1.001371189 | 0.957484056 | 1.047269929 | 0.952214232 | 0.968997537 |
| merged_omics | Proinsulin | cad | coronary_artery_disease | cox | 21390 | 997 | 1.078068523 | 1.012836427 | 1.147501917 | 0.018249089 | 0.025053834 |
| genetic | Hyper_Insulin | cad | coronary_artery_disease | cox | 443790 | 19980 | 1.029519145 | 1.015340979 | 1.043895295 | 3.93E-05 | 8.66E-05 |
| biochemistry | Hyper_Insulin | cad | coronary_artery_disease | cox | 424098 | 19120 | 1.10425003 | 1.088811975 | 1.119906978 | 2.37E-43 | 3.19E-43 |
| genetic | Beta_Cell_1 | cad | coronary_artery_disease | cox | 443790 | 19980 | 1.046189577 | 1.031783043 | 1.060797265 | 1.74E-10 | 6.53E-10 |
| biochemistry | Beta_Cell_1 | cad | coronary_artery_disease | cox | 424098 | 19120 | 1.204071175 | 1.187422577 | 1.220953198 | 1.21E-150 | 2.26E-150 |
| genetic | Beta_Cell_2 | cad | coronary_artery_disease | cox | 443790 | 19980 | 1.02356455 | 1.009486429 | 1.037839003 | 0.00098 | 0.001749999 |
| biochemistry | Beta_Cell_2 | cad | coronary_artery_disease | cox | 424098 | 19120 | 1.186333073 | 1.169826611 | 1.203072445 | 2.94E-126 | 5.09E-126 |
| proteomics | Beta_Cell_2 | cad | coronary_artery_disease | cox | 39875 | 1905 | 0.998091072 | 0.953917324 | 1.044310406 | 0.934064617 | 0.968997537 |
| merged_omics | Beta_Cell_2 | cad | coronary_artery_disease | cox | 21390 | 997 | 1.097932394 | 1.032396971 | 1.167627934 | 0.002926398 | 0.004309787 |
| genetic | Total_GRS | cad | coronary_artery_disease | cox | 443790 | 19980 | 1.075703269 | 1.06087809 | 1.090735623 | 6.59E-25 | 4.30E-24 |
| biochemistry | Total_GRS | cad | coronary_artery_disease | cox | 424098 | 19120 | 1.234327755 | 1.217917037 | 1.250959597 | 1.06E-208 | 2.40E-208 |
| merged_omics | Total_GRS | cad | coronary_artery_disease | cox | 21390 | 997 | 1.248249318 | 1.17358011 | 1.327669364 | 1.84E-12 | 5.53E-12 |
| genetic | Cholesterol | is | ischemic_stroke | cox | 456943 | 9186 | 0.981183808 | 0.961459311 | 1.001312956 | 0.066748733 | 0.09020099 |
| biochemistry | Cholesterol | is | ischemic_stroke | cox | 436637 | 8783 | 1.1946328 | 1.170083831 | 1.21969682 | 3.02E-63 | 4.27E-63 |
| metabolomics | Cholesterol | is | ischemic_stroke | cox | 257042 | 5240 | 0.945659506 | 0.919071854 | 0.973016307 | 0.000123028 | 0.000144739 |
| proteomics | Cholesterol | is | ischemic_stroke | cox | 41179 | 903 | 1.049856837 | 0.983555583 | 1.120627444 | 0.143791672 | 0.647062526 |
| merged_omics | Cholesterol | is | ischemic_stroke | cox | 22110 | 486 | 1.102938497 | 1.009660027 | 1.204834594 | 0.029762274 | 0.04017907 |
| genetic | Lipodystrophy_1 | is | ischemic_stroke | cox | 456943 | 9186 | 1.011352278 | 0.990870169 | 1.032257769 | 0.27952841 | 0.330151665 |
| biochemistry | Lipodystrophy_1 | is | ischemic_stroke | cox | 436637 | 8783 | 0.960721469 | 0.940456366 | 0.981423247 | 0.000229668 | 0.000238174 |
| metabolomics | Lipodystrophy_1 | is | ischemic_stroke | cox | 257042 | 5240 | 0.999516492 | 0.970638118 | 1.029254053 | 0.974207349 | 0.974207349 |
| proteomics | Lipodystrophy_1 | is | ischemic_stroke | cox | 41179 | 903 | 1.022538113 | 0.957871619 | 1.091570281 | 0.50370173 | 0.86348868 |
| merged_omics | Lipodystrophy_1 | is | ischemic_stroke | cox | 22110 | 486 | 1.030416848 | 0.938124679 | 1.131788667 | 0.531405273 | 0.566366146 |
| genetic | Lipodystrophy_2 | is | ischemic_stroke | cox | 456943 | 9186 | 1.014211279 | 0.993662966 | 1.035184519 | 0.176614481 | 0.222623295 |
| biochemistry | Lipodystrophy_2 | is | ischemic_stroke | cox | 436637 | 8783 | 1.097684857 | 1.074657363 | 1.121205779 | 6.91E-18 | 8.13E-18 |
| proteomics | Lipodystrophy_2 | is | ischemic_stroke | cox | 41179 | 903 | 1.060813235 | 0.994481323 | 1.131569487 | 0.073130091 | 0.438780545 |
| merged_omics | Lipodystrophy_2 | is | ischemic_stroke | cox | 22110 | 486 | 1.142540279 | 1.044320666 | 1.249997566 | 0.00366538 | 0.00530171 |
| genetic | Liver-Lipid | is | ischemic_stroke | cox | 456943 | 9186 | 1.013579299 | 0.993013408 | 1.034571121 | 0.197176443 | 0.244433607 |
| biochemistry | Liver-Lipid | is | ischemic_stroke | cox | 436637 | 8783 | 0.933237279 | 0.913073775 | 0.953846055 | 5.64E-10 | 6.12E-10 |
| metabolomics | Liver-Lipid | is | ischemic_stroke | cox | 257042 | 5240 | 1.202385299 | 1.170907012 | 1.234709838 | 3.17E-42 | 7.05E-42 |
| proteomics | Liver-Lipid | is | ischemic_stroke | cox | 41179 | 903 | 0.983818916 | 0.921839396 | 1.049965606 | 0.623159761 | 0.963492845 |
| merged_omics | Liver-Lipid | is | ischemic_stroke | cox | 22110 | 486 | 0.950824323 | 0.869985061 | 1.039175193 | 0.265992804 | 0.299241904 |
| genetic | ASAT_Pos | is | ischemic_stroke | cox | 456943 | 9186 | 1.014574106 | 0.994043405 | 1.035528842 | 0.165379236 | 0.210951209 |
| biochemistry | ASAT_Pos | is | ischemic_stroke | cox | 436637 | 8783 | 1.028544099 | 1.00692855 | 1.050623664 | 0.009399716 | 0.00960555 |
| proteomics | ASAT_Pos | is | ischemic_stroke | cox | 41179 | 903 | 1.02559182 | 0.961042287 | 1.094476898 | 0.446117256 | 0.856875603 |
| merged_omics | ASAT_Pos | is | ischemic_stroke | cox | 22110 | 486 | 1.310428493 | 1.197900149 | 1.433527523 | 3.59E-09 | 8.56E-09 |
| genetic | ALP_Neg | is | ischemic_stroke | cox | 456943 | 9186 | 1.031987983 | 1.011107693 | 1.05329947 | 0.002534266 | 0.004271236 |
| biochemistry | ALP_Neg | is | ischemic_stroke | cox | 436637 | 8783 | 1.171856382 | 1.147265837 | 1.196974002 | 1.22E-48 | 1.67E-48 |
| metabolomics | ALP_Neg | is | ischemic_stroke | cox | 257042 | 5240 | 0.926586806 | 0.900035874 | 0.953920987 | 2.74E-07 | 3.78E-07 |
| proteomics | ALP_Neg | is | ischemic_stroke | cox | 41179 | 903 | 1.003897127 | 0.940534027 | 1.071528953 | 0.906915071 | 0.968997537 |
| merged_omics | ALP_Neg | is | ischemic_stroke | cox | 22110 | 486 | 1.09876728 | 1.005247901 | 1.200986874 | 0.037956124 | 0.050400754 |
| genetic | SHBG-LpA | is | ischemic_stroke | cox | 456943 | 9186 | 1.001759539 | 0.981457365 | 1.022481678 | 0.866355962 | 0.884036696 |
| biochemistry | SHBG-LpA | is | ischemic_stroke | cox | 436637 | 8783 | 1.079223589 | 1.056845793 | 1.102075217 | 9.90E-13 | 1.12E-12 |
| genetic | Obesity | is | ischemic_stroke | cox | 456943 | 9186 | 1.044023698 | 1.022890117 | 1.065593912 | 3.64E-05 | 8.15E-05 |
| biochemistry | Obesity | is | ischemic_stroke | cox | 436637 | 8783 | 1.303496549 | 1.275796432 | 1.33179809 | 3.15E-129 | 5.59E-129 |
| genetic | VAT_Neg | is | ischemic_stroke | cox | 456943 | 9186 | 0.998833407 | 0.978576122 | 1.019510034 | 0.911092733 | 0.918522042 |
| biochemistry | VAT_Neg | is | ischemic_stroke | cox | 436637 | 8783 | 1.116825469 | 1.093964176 | 1.140164509 | 1.18E-25 | 1.43E-25 |
| genetic | Bilirubin | is | ischemic_stroke | cox | 456943 | 9186 | 0.975250306 | 0.95532505 | 0.995591144 | 0.017334028 | 0.026814106 |
| genetic | Proinsulin | is | ischemic_stroke | cox | 456943 | 9186 | 1.035449665 | 1.014459536 | 1.056874099 | 0.000856291 | 0.001555244 |
| biochemistry | Proinsulin | is | ischemic_stroke | cox | 436637 | 8783 | 1.266811864 | 1.240011878 | 1.29419107 | 3.25E-104 | 5.23E-104 |
| proteomics | Proinsulin | is | ischemic_stroke | cox | 41179 | 903 | 0.974904689 | 0.913492958 | 1.040444968 | 0.443900794 | 0.856875603 |
| merged_omics | Proinsulin | is | ischemic_stroke | cox | 22110 | 486 | 1.060295403 | 0.969607353 | 1.159465568 | 0.199342914 | 0.227419381 |
| genetic | Hyper_Insulin | is | ischemic_stroke | cox | 456943 | 9186 | 1.024130032 | 1.003412761 | 1.045275049 | 0.022210477 | 0.033652238 |
| biochemistry | Hyper_Insulin | is | ischemic_stroke | cox | 436637 | 8783 | 1.074085797 | 1.051792805 | 1.096851295 | 2.41E-11 | 2.68E-11 |
| genetic | Beta_Cell_1 | is | ischemic_stroke | cox | 456943 | 9186 | 1.024988763 | 1.004242112 | 1.04616402 | 0.017993662 | 0.027541319 |
| biochemistry | Beta_Cell_1 | is | ischemic_stroke | cox | 436637 | 8783 | 1.225645953 | 1.200499947 | 1.251318674 | 1.79E-82 | 2.63E-82 |
| genetic | Beta_Cell_2 | is | ischemic_stroke | cox | 456943 | 9186 | 1.011494176 | 0.991030993 | 1.032379891 | 0.273079883 | 0.326186827 |
| biochemistry | Beta_Cell_2 | is | ischemic_stroke | cox | 436637 | 8783 | 1.228368719 | 1.203137003 | 1.254129586 | 4.81E-84 | 7.24E-84 |
| proteomics | Beta_Cell_2 | is | ischemic_stroke | cox | 41179 | 903 | 1.004695149 | 0.940839302 | 1.072884966 | 0.888809625 | 0.968997537 |
| merged_omics | Beta_Cell_2 | is | ischemic_stroke | cox | 22110 | 486 | 1.09216365 | 0.999559458 | 1.193347158 | 0.051146046 | 0.065759203 |
| genetic | Total_GRS | is | ischemic_stroke | cox | 456943 | 9186 | 1.058748921 | 1.037305878 | 1.080635231 | 4.54E-08 | 1.33E-07 |
| biochemistry | Total_GRS | is | ischemic_stroke | cox | 436637 | 8783 | 1.243329421 | 1.21878208 | 1.268371166 | 1.15E-101 | 1.82E-101 |
| merged_omics | Total_GRS | is | ischemic_stroke | cox | 22110 | 486 | 1.271041973 | 1.16227554 | 1.389986832 | 1.48E-07 | 3.09E-07 |
| genetic | Cholesterol | hf | heart_failure | cox | 456274 | 17352 | 0.989947682 | 0.975347883 | 1.004766022 | 0.182605134 | 0.228256418 |
| biochemistry | Cholesterol | hf | heart_failure | cox | 436022 | 16549 | 1.355834143 | 1.33556654 | 1.376409313 | 0 | 0 |

**Table S16: Associations between pPSs, pMMSs and T2D-related disease outcomes in UKBB**

| Score_type | Biological_Process | Outcome | Outcome_Heading | Model_T | Sample_Size | Events | estimate | conf_low | conf_high | p | fdr_p_adjusted |
|---|---|---|---|---|---|---|---|---|---|---|---|
| metabolomics | Cholesterol | hf | heart_failure | cox | 256628 | 10093 | 0.939598699 | 0.92045466 | 0.959140905 | 2.99E-09 | 4.27E-09 |
| proteomics | Cholesterol | hf | heart_failure | cox | 40967 | 1791 | 1.010146132 | 0.964447394 | 1.058010229 | 0.669092254 | 0.963492845 |
| merged_omics | Cholesterol | hf | heart_failure | cox | 21982 | 970 | 1.221482182 | 1.147540593 | 1.300188185 | 3.39E-10 | 9.16E-10 |
| genetic | Lipodystrophy_1 | hf | heart_failure | cox | 456274 | 17352 | 1.018325777 | 1.003284449 | 1.033592607 | 0.016761576 | 0.026465647 |
| biochemistry | Lipodystrophy_1 | hf | heart_failure | cox | 436022 | 16549 | 0.909480138 | 0.895432801 | 0.923747846 | 6.72E-33 | 8.55E-33 |
| metabolomics | Lipodystrophy_1 | hf | heart_failure | cox | 256628 | 10093 | 0.992445111 | 0.971649723 | 1.013685564 | 0.48273766 | 0.495115549 |
| proteomics | Lipodystrophy_1 | hf | heart_failure | cox | 40967 | 1791 | 1.032080938 | 0.985446223 | 1.080922569 | 0.180720909 | 0.680092075 |
| merged_omics | Lipodystrophy_1 | hf | heart_failure | cox | 21982 | 970 | 1.025357976 | 0.959380348 | 1.095872958 | 0.460533343 | 0.497376011 |
| genetic | Lipodystrophy_2 | hf | heart_failure | cox | 456274 | 17352 | 0.998230404 | 0.983469991 | 1.013212349 | 0.815736319 | 0.849725332 |
| biochemistry | Lipodystrophy_2 | hf | heart_failure | cox | 436022 | 16549 | 1.133941024 | 1.116588159 | 1.151563568 | 1.88E-57 | 2.61E-57 |
| proteomics | Lipodystrophy_2 | hf | heart_failure | cox | 40967 | 1791 | 0.992388862 | 0.947546611 | 1.039353254 | 0.74604467 | 0.968997537 |
| merged_omics | Lipodystrophy_2 | hf | heart_failure | cox | 21982 | 970 | 1.21599962 | 1.141287321 | 1.295602824 | 1.49E-09 | 3.67E-09 |
| genetic | Liver-Lipid | hf | heart_failure | cox | 456274 | 17352 | 0.999829569 | 0.985055129 | 1.014825604 | 0.982096752 | 0.982096752 |
| biochemistry | Liver-Lipid | hf | heart_failure | cox | 436022 | 16549 | 0.873761138 | 0.859903875 | 0.887841709 | 1.73E-61 | 2.43E-61 |
| metabolomics | Liver-Lipid | hf | heart_failure | cox | 256628 | 10093 | 1.311786756 | 1.287431812 | 1.336602433 | 3.25E-177 | 1.86E-176 |
| proteomics | Liver-Lipid | hf | heart_failure | cox | 40967 | 1791 | 1.004938133 | 0.959517015 | 1.052509371 | 0.834643729 | 0.968997537 |
| merged_omics | Liver-Lipid | hf | heart_failure | cox | 21982 | 970 | 0.874105185 | 0.821068796 | 0.930567425 | 2.52E-05 | 4.43E-05 |
| genetic | ASAT_Pos | hf | heart_failure | cox | 456274 | 17352 | 0.987723283 | 0.973129762 | 1.002535656 | 0.103834752 | 0.134269076 |
| biochemistry | ASAT_Pos | hf | heart_failure | cox | 436022 | 16549 | 0.992554029 | 0.977251904 | 1.00809576 | 0.345769117 | 0.350780263 |
| proteomics | ASAT_Pos | hf | heart_failure | cox | 40967 | 1791 | 1.016597327 | 0.970735758 | 1.064625586 | 0.484600001 | 0.856875603 |
| merged_omics | ASAT_Pos | hf | heart_failure | cox | 21982 | 970 | 1.328270952 | 1.246334806 | 1.415593719 | 2.36E-18 | 1.12E-17 |
| genetic | ALP_Neg | hf | heart_failure | cox | 456274 | 17352 | 1.013932427 | 0.998953052 | 1.029136419 | 0.068446157 | 0.09166896 |
| biochemistry | ALP_Neg | hf | heart_failure | cox | 436022 | 16549 | 1.241539801 | 1.222529193 | 1.260846029 | 2.94E-166 | 5.72E-166 |
| metabolomics | ALP_Neg | hf | heart_failure | cox | 256628 | 10093 | 0.859104713 | 0.84125102 | 0.877337311 | 1.34E-45 | 3.56E-45 |
| proteomics | ALP_Neg | hf | heart_failure | cox | 40967 | 1791 | 0.968507249 | 0.924537495 | 1.014568145 | 0.177054209 | 0.680092075 |
| merged_omics | ALP_Neg | hf | heart_failure | cox | 21982 | 970 | 1.011801555 | 0.949974478 | 1.077652515 | 0.715331452 | 0.733441109 |
| genetic | SHBG-LpA | hf | heart_failure | cox | 456274 | 17352 | 1.018344116 | 1.00330199 | 1.033611762 | 0.016657392 | 0.026465647 |
| biochemistry | SHBG-LpA | hf | heart_failure | cox | 436022 | 16549 | 1.097756763 | 1.081115705 | 1.114653969 | 5.26E-33 | 6.75E-33 |
| genetic | Obesity | hf | heart_failure | cox | 456274 | 17352 | 1.100488569 | 1.08426011 | 1.116959925 | 1.39E-36 | 1.23E-35 |
| biochemistry | Obesity | hf | heart_failure | cox | 436022 | 16549 | 1.473494832 | 1.450606031 | 1.49674479 | 0 | 0 |
| genetic | VAT_Neg | hf | heart_failure | cox | 456274 | 17352 | 0.999163081 | 0.984369412 | 1.014179079 | 0.912398562 | 0.918522042 |
| biochemistry | VAT_Neg | hf | heart_failure | cox | 436022 | 16549 | 1.150396024 | 1.133223701 | 1.167828568 | 1.76E-74 | 2.55E-74 |
| genetic | Bilirubin | hf | heart_failure | cox | 456274 | 17352 | 1.008882402 | 0.994018234 | 1.023968844 | 0.242912205 | 0.296234396 |
| genetic | Proinsulin | hf | heart_failure | cox | 456274 | 17352 | 1.030692981 | 1.015446633 | 1.046168243 | 7.01E-05 | 0.000150186 |
| biochemistry | Proinsulin | hf | heart_failure | cox | 436022 | 16549 | 1.353846899 | 1.333081528 | 1.374935731 | 7.4e-323 | 2.27e-322 |
| proteomics | Proinsulin | hf | heart_failure | cox | 40967 | 1791 | 0.952976853 | 0.909923362 | 0.998067442 | 0.041150244 | 0.356287728 |
| merged_omics | Proinsulin | hf | heart_failure | cox | 21982 | 970 | 1.065518877 | 1.000214351 | 1.13508717 | 0.049224511 | 0.064309442 |
| genetic | Hyper_Insulin | hf | heart_failure | cox | 456274 | 17352 | 1.027889509 | 1.012717032 | 1.043289299 | 0.000288339 | 0.000554497 |
| biochemistry | Hyper_Insulin | hf | heart_failure | cox | 436022 | 16549 | 1.091123907 | 1.074610995 | 1.107890564 | 3.69E-29 | 4.54E-29 |
| genetic | Beta_Cell_1 | hf | heart_failure | cox | 456274 | 17352 | 1.027001108 | 1.011840308 | 1.042389069 | 0.000445992 | 0.000846821 |
| biochemistry | Beta_Cell_1 | hf | heart_failure | cox | 436022 | 16549 | 1.306846208 | 1.287459497 | 1.326524846 | 7.97E-270 | 2.03E-269 |
| genetic | Beta_Cell_2 | hf | heart_failure | cox | 456274 | 17352 | 1.01859523 | 1.003564245 | 1.033851342 | 0.015137469 | 0.024415273 |
| biochemistry | Beta_Cell_2 | hf | heart_failure | cox | 436022 | 16549 | 1.330767862 | 1.311040769 | 1.350791786 | 8.62E-308 | 2.51E-307 |
| proteomics | Beta_Cell_2 | hf | heart_failure | cox | 40967 | 1791 | 0.994606719 | 0.949254287 | 1.04212595 | 0.820337225 | 0.968997537 |
| merged_omics | Beta_Cell_2 | hf | heart_failure | cox | 21982 | 970 | 1.192320541 | 1.120164053 | 1.269125061 | 3.34E-08 | 7.51E-08 |
| genetic | Total_GRS | hf | heart_failure | cox | 456274 | 17352 | 1.060193736 | 1.044528162 | 1.076094258 | 1.40E-14 | 6.02E-14 |
| biochemistry | Total_GRS | hf | heart_failure | cox | 436022 | 16549 | 1.341997874 | 1.323040556 | 1.361226823 | 0 | 0 |
| merged_omics | Total_GRS | hf | heart_failure | cox | 21982 | 970 | 1.399026883 | 1.313780965 | 1.489804063 | 1.21E-25 | 6.52E-25 |
| genetic | Cholesterol | pad | peripheral_artery_disease | cox | 455657 | 7415 | 0.987423349 | 0.965283454 | 1.010071049 | 0.273996934 | 0.326186827 |
| biochemistry | Cholesterol | pad | peripheral_artery_disease | cox | 435420 | 7056 | 1.299331903 | 1.269629704 | 1.329728965 | 3.96E-109 | 6.44E-109 |
| metabolomics | Cholesterol | pad | peripheral_artery_disease | cox | 256283 | 4341 | 0.860731851 | 0.834375399 | 0.887920858 | 3.33E-21 | 5.55E-21 |
| proteomics | Cholesterol | pad | peripheral_artery_disease | cox | 41017 | 720 | 1.070170689 | 0.994797019 | 1.151255263 | 0.068758178 | 0.438780545 |
| merged_omics | Cholesterol | pad | peripheral_artery_disease | cox | 22005 | 400 | 1.174038225 | 1.065051128 | 1.294178015 | 0.00124713 | 0.001905991 |
| genetic | Lipodystrophy_1 | pad | peripheral_artery_disease | cox | 455657 | 7415 | 1.064872156 | 1.040882991 | 1.089414197 | 6.42E-08 | 1.78E-07 |
| biochemistry | Lipodystrophy_1 | pad | peripheral_artery_disease | cox | 435420 | 7056 | 1.007490241 | 0.983847131 | 1.031701525 | 0.537951103 | 0.541821255 |
| metabolomics | Lipodystrophy_1 | pad | peripheral_artery_disease | cox | 256283 | 4341 | 1.084709431 | 1.049711773 | 1.12087392 | 1.18E-06 | 1.57E-06 |
| proteomics | Lipodystrophy_1 | pad | peripheral_artery_disease | cox | 41017 | 720 | 0.990640677 | 0.920689287 | 1.065906777 | 0.801285079 | 0.968997537 |
| merged_omics | Lipodystrophy_1 | pad | peripheral_artery_disease | cox | 22005 | 400 | 1.097123513 | 0.988686101 | 1.217454156 | 0.080862104 | 0.096321035 |
| genetic | Lipodystrophy_2 | pad | peripheral_artery_disease | cox | 455657 | 7415 | 1.037748956 | 1.014397468 | 1.061637997 | 0.001417515 | 0.002416219 |
| biochemistry | Lipodystrophy_2 | pad | peripheral_artery_disease | cox | 435420 | 7056 | 1.122839084 | 1.096617337 | 1.149687833 | 7.25E-22 | 8.68E-22 |
| proteomics | Lipodystrophy_2 | pad | peripheral_artery_disease | cox | 41017 | 720 | 1.049195425 | 0.976008361 | 1.127870502 | 0.193003378 | 0.680092075 |
| merged_omics | Lipodystrophy_2 | pad | peripheral_artery_disease | cox | 22005 | 400 | 1.230618205 | 1.11499276 | 1.35823408 | 3.75E-05 | 6.47E-05 |
| genetic | Liver-Lipid | pad | peripheral_artery_disease | cox | 455657 | 7415 | 1.00585771 | 0.983186277 | 1.029051927 | 0.615563487 | 0.664277145 |
| biochemistry | Liver-Lipid | pad | peripheral_artery_disease | cox | 435420 | 7056 | 0.837371346 | 0.817149861 | 0.858093239 | 5.89E-46 | 8.01E-46 |
| metabolomics | Liver-Lipid | pad | peripheral_artery_disease | cox | 256283 | 4341 | 1.326542248 | 1.289372811 | 1.364783188 | 1.39E-84 | 4.62E-84 |
| proteomics | Liver-Lipid | pad | peripheral_artery_disease | cox | 41017 | 720 | 0.989550474 | 0.91997148 | 1.064391845 | 0.777641213 | 0.968997537 |
| merged_omics | Liver-Lipid | pad | peripheral_artery_disease | cox | 22005 | 400 | 0.889068257 | 0.806395195 | 0.980217108 | 0.018212889 | 0.025053834 |
| genetic | ASAT_Pos | pad | peripheral_artery_disease | cox | 455657 | 7415 | 1.037883282 | 1.014542314 | 1.061761242 | 0.001354917 | 0.002336065 |
| biochemistry | ASAT_Pos | pad | peripheral_artery_disease | cox | 435420 | 7056 | 1.093161419 | 1.067599953 | 1.119334902 | 1.60E-13 | 1.85E-13 |
| proteomics | ASAT_Pos | pad | peripheral_artery_disease | cox | 41017 | 720 | 1.028387246 | 0.956068261 | 1.106176588 | 0.451805651 | 0.856875603 |
| merged_omics | ASAT_Pos | pad | peripheral_artery_disease | cox | 22005 | 400 | 1.257263793 | 1.138501513 | 1.388414708 | 6.12E-06 | 1.13E-05 |
| genetic | ALP_Neg | pad | peripheral_artery_disease | cox | 455657 | 7415 | 1.040147828 | 1.016765215 | 1.064068173 | 0.000690668 | 0.001279016 |
| biochemistry | ALP_Neg | pad | peripheral_artery_disease | cox | 435420 | 7056 | 1.353060987 | 1.32178718 | 1.38507474 | 1.07E-141 | 1.94E-141 |
| metabolomics | ALP_Neg | pad | peripheral_artery_disease | cox | 256283 | 4341 | 0.902648351 | 0.874275374 | 0.93194212 | 3.27E-10 | 4.84E-10 |
| proteomics | ALP_Neg | pad | peripheral_artery_disease | cox | 41017 | 720 | 0.994787542 | 0.924668682 | 1.070223607 | 0.888551678 | 0.968997537 |
| merged_omics | ALP_Neg | pad | peripheral_artery_disease | cox | 22005 | 400 | 1.050173803 | 0.951839476 | 1.158667028 | 0.329073922 | 0.365136817 |
| genetic | SHBG-LpA | pad | peripheral_artery_disease | cox | 455657 | 7415 | 1.011315848 | 0.988542455 | 1.034613879 | 0.332883146 | 0.387073426 |
| biochemistry | SHBG-LpA | pad | peripheral_artery_disease | cox | 435420 | 7056 | 1.16901853 | 1.14204212 | 1.196632154 | 2.87E-39 | 3.79E-39 |
| genetic | Obesity | pad | peripheral_artery_disease | cox | 455657 | 7415 | 1.073217049 | 1.049077907 | 1.097911629 | 1.14E-09 | 3.99E-09 |
| biochemistry | Obesity | pad | peripheral_artery_disease | cox | 435420 | 7056 | 1.607909492 | 1.569900337 | 1.646838895 | 0 | 0 |
| genetic | VAT_Neg | pad | peripheral_artery_disease | cox | 455657 | 7415 | 1.005452474 | 0.982770883 | 1.028657538 | 0.640427313 | 0.686172121 |
| biochemistry | VAT_Neg | pad | peripheral_artery_disease | cox | 435420 | 7056 | 1.261398988 | 1.233023651 | 1.29042732 | 4.98E-89 | 7.66E-89 |
| genetic | Bilirubin | pad | peripheral_artery_disease | cox | 455657 | 7415 | 1.011453738 | 0.988748883 | 1.03467997 | 0.325517135 | 0.381465392 |
| genetic | Proinsulin | pad | peripheral_artery_disease | cox | 455657 | 7415 | 1.050500949 | 1.026827279 | 1.074720418 | 2.27E-05 | 5.24E-05 |

**Table S16: Associations between pPSs, pMMSs and T2D-related disease outcomes in UKBB**

| Score_type | Biological_Process | Outcome | Outcome_Heading | Model_T | Sample_Size | Events | estimate | conf_low | conf_high | p | fdr_p_adjusted |
|---|---|---|---|---|---|---|---|---|---|---|---|
| biochemistry | Proinsulin | pad | peripheral_artery_disease | cox | 435420 | 7056 | 1.538652108 | 1.50357576 | 1.574546739 | 1.18E-293 | 3.31E-293 |
| proteomics | Proinsulin | pad | peripheral_artery_disease | cox | 41017 | 720 | 0.974568769 | 0.906144995 | 1.048159281 | 0.487943052 | 0.856875603 |
| merged_omics | Proinsulin | pad | peripheral_artery_disease | cox | 22005 | 400 | 1.168213455 | 1.0584912 | 1.289309422 | 0.00200416 | 0.00300624 |
| genetic | Hyper_Insulin | pad | peripheral_artery_disease | cox | 455657 | 7415 | 1.04335519 | 1.019895344 | 1.067354664 | 0.000254334 | 0.000495456 |
| biochemistry | Hyper_Insulin | pad | peripheral_artery_disease | cox | 435420 | 7056 | 1.165883627 | 1.139304328 | 1.193083004 | 6.85E-39 | 8.97E-39 |
| genetic | Beta_Cell_1 | pad | peripheral_artery_disease | cox | 455657 | 7415 | 1.053312746 | 1.029608811 | 1.0775624 | 7.73E-06 | 1.81E-05 |
| biochemistry | Beta_Cell_1 | pad | peripheral_artery_disease | cox | 435420 | 7056 | 1.438050547 | 1.406212026 | 1.470609936 | 5.77E-222 | 1.37E-221 |
| genetic | Beta_Cell_2 | pad | peripheral_artery_disease | cox | 455657 | 7415 | 1.039393541 | 1.016040499 | 1.063283338 | 0.000860568 | 0.001555244 |
| biochemistry | Beta_Cell_2 | pad | peripheral_artery_disease | cox | 435420 | 7056 | 1.455734322 | 1.423467855 | 1.48873219 | 1.80E-236 | 4.35E-236 |
| proteomics | Beta_Cell_2 | pad | peripheral_artery_disease | cox | 41017 | 720 | 1.085563515 | 1.008483375 | 1.168535024 | 0.028903118 | 0.356287728 |
| merged_omics | Beta_Cell_2 | pad | peripheral_artery_disease | cox | 22005 | 400 | 1.174195847 | 1.065231053 | 1.294306885 | 0.001230445 | 0.001905991 |
| genetic | Total_GRS | pad | peripheral_artery_disease | cox | 455657 | 7415 | 1.123289384 | 1.097988749 | 1.149173013 | 1.48E-23 | 8.90E-23 |
| biochemistry | Total_GRS | pad | peripheral_artery_disease | cox | 435420 | 7056 | 1.464735417 | 1.43410419 | 1.496020902 | 1.92E-274 | 4.97E-274 |
| merged_omics | Total_GRS | pad | peripheral_artery_disease | cox | 22005 | 400 | 1.337756923 | 1.212375277 | 1.47610531 | 6.81E-09 | 1.58E-08 |
| genetic | Cholesterol | nephrodm | diabetic_nephropathy | cox | 458786 | 692 | 1.083980661 | 1.002180443 | 1.172457595 | 0.043966639 | 0.063413422 |
| biochemistry | Cholesterol | nephrodm | diabetic_nephropathy | cox | 438398 | 658 | 2.206525321 | 2.049949349 | 2.375060629 | 1.36E-98 | 2.14E-98 |
| metabolomics | Cholesterol | nephrodm | diabetic_nephropathy | cox | 258078 | 414 | 0.58017953 | 0.527484538 | 0.63813868 | 3.79E-29 | 6.59E-29 |
| genetic | Lipodystrophy_1 | nephrodm | diabetic_nephropathy | cox | 458786 | 692 | 1.23257393 | 1.143683442 | 1.328373252 | 4.36E-08 | 1.31E-07 |
| biochemistry | Lipodystrophy_1 | nephrodm | diabetic_nephropathy | cox | 438398 | 658 | 1.556708666 | 1.442334638 | 1.680152307 | 6.09E-30 | 7.61E-30 |
| metabolomics | Lipodystrophy_1 | nephrodm | diabetic_nephropathy | cox | 258078 | 414 | 1.17218067 | 1.052474952 | 1.305501401 | 0.003845332 | 0.004157116 |
| genetic | Lipodystrophy_2 | nephrodm | diabetic_nephropathy | cox | 458786 | 692 | 1.164159728 | 1.08096495 | 1.253757464 | 5.87E-05 | 0.000127576 |
| biochemistry | Lipodystrophy_2 | nephrodm | diabetic_nephropathy | cox | 438398 | 658 | 2.019099188 | 1.881049295 | 2.167280539 | 3.15E-84 | 4.80E-84 |
| genetic | Liver-Lipid | nephrodm | diabetic_nephropathy | cox | 458786 | 692 | 1.043693813 | 0.96832142 | 1.124933057 | 0.263456775 | 0.318697711 |
| biochemistry | Liver-Lipid | nephrodm | diabetic_nephropathy | cox | 438398 | 658 | 1.288787788 | 1.191162842 | 1.394413849 | 2.74E-10 | 3.00E-10 |
| metabolomics | Liver-Lipid | nephrodm | diabetic_nephropathy | cox | 258078 | 414 | 1.838171729 | 1.690217553 | 1.99907716 | 6.97E-46 | 1.99E-45 |
| genetic | ASAT_Pos | nephrodm | diabetic_nephropathy | cox | 458786 | 692 | 1.185937776 | 1.101031343 | 1.277391798 | 6.81E-06 | 1.62E-05 |
| biochemistry | ASAT_Pos | nephrodm | diabetic_nephropathy | cox | 438398 | 658 | 1.292397759 | 1.197492399 | 1.394824694 | 4.35E-11 | 4.80E-11 |
| genetic | ALP_Neg | nephrodm | diabetic_nephropathy | cox | 458786 | 692 | 1.033374662 | 0.959216121 | 1.113266519 | 0.387547339 | 0.443756495 |
| biochemistry | ALP_Neg | nephrodm | diabetic_nephropathy | cox | 438398 | 658 | 4.690813476 | 4.411875703 | 4.987386897 | 0 | 0 |
| metabolomics | ALP_Neg | nephrodm | diabetic_nephropathy | cox | 258078 | 414 | 0.534716112 | 0.483573124 | 0.591268014 | 2.93E-34 | 5.86E-34 |
| genetic | SHBG-LpA | nephrodm | diabetic_nephropathy | cox | 458786 | 692 | 1.025313133 | 0.951663137 | 1.104662964 | 0.510991466 | 0.567768295 |
| biochemistry | SHBG-LpA | nephrodm | diabetic_nephropathy | cox | 438398 | 658 | 2.406694448 | 2.231696649 | 2.595414645 | 4.37E-115 | 7.28E-115 |
| genetic | Obesity | nephrodm | diabetic_nephropathy | cox | 458786 | 692 | 1.237575965 | 1.148962199 | 1.333024072 | 1.87E-08 | 5.86E-08 |
| biochemistry | Obesity | nephrodm | diabetic_nephropathy | cox | 438398 | 658 | 3.128895137 | 2.892634593 | 3.384452637 | 2.31E-178 | 4.68E-178 |
| genetic | VAT_Neg | nephrodm | diabetic_nephropathy | cox | 458786 | 692 | 1.119017938 | 1.038229542 | 1.206092772 | 0.003268401 | 0.005447335 |
| biochemistry | VAT_Neg | nephrodm | diabetic_nephropathy | cox | 438398 | 658 | 2.508325955 | 2.345516669 | 2.682436317 | 6.78E-159 | 1.28E-158 |
| genetic | Bilirubin | nephrodm | diabetic_nephropathy | cox | 458786 | 692 | 1.022172124 | 0.949214954 | 1.100736823 | 0.561608147 | 0.614899431 |
| genetic | Proinsulin | nephrodm | diabetic_nephropathy | cox | 458786 | 692 | 1.231397634 | 1.142718433 | 1.326958672 | 4.80E-08 | 1.38E-07 |
| biochemistry | Proinsulin | nephrodm | diabetic_nephropathy | cox | 438398 | 658 | 5.284984903 | 4.944526032 | 5.648886314 | 0 | 0 |
| genetic | Hyper_Insulin | nephrodm | diabetic_nephropathy | cox | 458786 | 692 | 1.257986476 | 1.167673974 | 1.355284102 | 1.56E-09 | 5.19E-09 |
| biochemistry | Hyper_Insulin | nephrodm | diabetic_nephropathy | cox | 438398 | 658 | 2.576733454 | 2.421736629 | 2.741650438 | 1.74E-196 | 3.74E-196 |
| genetic | Beta_Cell_1 | nephrodm | diabetic_nephropathy | cox | 458786 | 692 | 1.260067667 | 1.169505899 | 1.357642169 | 1.24E-09 | 4.23E-09 |
| biochemistry | Beta_Cell_1 | nephrodm | diabetic_nephropathy | cox | 438398 | 658 | 4.424468911 | 4.151648396 | 4.715217494 | 0 | 0 |
| genetic | Beta_Cell_2 | nephrodm | diabetic_nephropathy | cox | 458786 | 692 | 1.22888252 | 1.141172877 | 1.323333456 | 4.89E-08 | 1.38E-07 |
| biochemistry | Beta_Cell_2 | nephrodm | diabetic_nephropathy | cox | 438398 | 658 | 4.994387061 | 4.674129882 | 5.336587288 | 0 | 0 |
| genetic | Total_GRS | nephrodm | diabetic_nephropathy | cox | 458786 | 692 | 1.673741228 | 1.553473047 | 1.80332044 | 9.30E-42 | 8.72E-41 |
| biochemistry | Total_GRS | nephrodm | diabetic_nephropathy | cox | 438398 | 658 | 3.999564355 | 3.767473337 | 4.245953083 | 0 | 0 |
| genetic | Cholesterol | neurodm | diabetic_neuropathy | cox | 458500 | 1923 | 1.0199373 | 0.974674488 | 1.067302067 | 0.39399488 | 0.447721455 |
| biochemistry | Cholesterol | neurodm | diabetic_neuropathy | cox | 438131 | 1833 | 2.043829112 | 1.955220782 | 2.136453069 | 2.61E-219 | 6.10E-219 |
| metabolomics | Cholesterol | neurodm | diabetic_neuropathy | cox | 257891 | 1127 | 0.675494416 | 0.636898381 | 0.71642937 | 4.93E-39 | 1.04E-38 |
| proteomics | Cholesterol | neurodm | diabetic_neuropathy | cox | 41339 | 216 | 1.16040949 | 1.016039382 | 1.325293299 | 0.028182263 | 0.356287728 |
| merged_omics | Cholesterol | neurodm | diabetic_neuropathy | cox | 22193 | 128 | 1.582040282 | 1.331218762 | 1.880120328 | 1.90E-07 | 3.86E-07 |
| genetic | Lipodystrophy_1 | neurodm | diabetic_neuropathy | cox | 458500 | 1923 | 1.172207841 | 1.120792645 | 1.225981654 | 3.83E-12 | 1.47E-11 |
| biochemistry | Lipodystrophy_1 | neurodm | diabetic_neuropathy | cox | 438131 | 1833 | 1.79989892 | 1.719968986 | 1.883543335 | 7.00E-142 | 1.29E-141 |
| metabolomics | Lipodystrophy_1 | neurodm | diabetic_neuropathy | cox | 257891 | 1127 | 1.113658073 | 1.043903108 | 1.188074156 | 0.001106485 | 0.001264554 |
| proteomics | Lipodystrophy_1 | neurodm | diabetic_neuropathy | cox | 41339 | 216 | 0.950706107 | 0.831908197 | 1.086468562 | 0.457931723 | 0.856875603 |
| merged_omics | Lipodystrophy_1 | neurodm | diabetic_neuropathy | cox | 22193 | 128 | 1.201173729 | 0.99765325 | 1.446212227 | 0.052966384 | 0.06703558 |
| genetic | Lipodystrophy_2 | neurodm | diabetic_neuropathy | cox | 458500 | 1923 | 1.118457844 | 1.069750815 | 1.169382562 | 8.30E-07 | 2.11E-06 |
| biochemistry | Lipodystrophy_2 | neurodm | diabetic_neuropathy | cox | 438131 | 1833 | 2.391710006 | 2.295053762 | 2.49243693 | 0 | 0 |
| proteomics | Lipodystrophy_2 | neurodm | diabetic_neuropathy | cox | 41339 | 216 | 0.991963372 | 0.86808597 | 1.1335183 | 0.905623573 | 0.968997537 |
| merged_omics | Lipodystrophy_2 | neurodm | diabetic_neuropathy | cox | 22193 | 128 | 2.092823746 | 1.761626038 | 2.486288883 | 4.39E-17 | 1.87E-16 |
| genetic | Liver-Lipid | neurodm | diabetic_neuropathy | cox | 458500 | 1923 | 1.028508046 | 0.983363667 | 1.07572492 | 0.219658529 | 0.270071962 |
| biochemistry | Liver-Lipid | neurodm | diabetic_neuropathy | cox | 438131 | 1833 | 1.070986437 | 1.021241447 | 1.123154522 | 0.004710456 | 0.004848999 |
| metabolomics | Liver-Lipid | neurodm | diabetic_neuropathy | cox | 257891 | 1127 | 1.880341115 | 1.787642049 | 1.977847138 | 2.36E-132 | 9.45E-132 |
| proteomics | Liver-Lipid | neurodm | diabetic_neuropathy | cox | 41339 | 216 | 0.970749909 | 0.849818582 | 1.10889007 | 0.661870123 | 0.963492845 |
| merged_omics | Liver-Lipid | neurodm | diabetic_neuropathy | cox | 22193 | 128 | 0.648218107 | 0.546786349 | 0.768465992 | 5.93E-07 | 1.17E-06 |
| genetic | ASAT_Pos | neurodm | diabetic_neuropathy | cox | 458500 | 1923 | 1.123772363 | 1.074756366 | 1.175023814 | 2.92E-07 | 7.56E-07 |
| biochemistry | ASAT_Pos | neurodm | diabetic_neuropathy | cox | 438131 | 1833 | 2.195704795 | 2.104293029 | 2.291087543 | 9.38E-288 | 2.57E-287 |
| proteomics | ASAT_Pos | neurodm | diabetic_neuropathy | cox | 41339 | 216 | 0.997360396 | 0.872899519 | 1.139567313 | 0.968997537 | 0.968997537 |
| merged_omics | ASAT_Pos | neurodm | diabetic_neuropathy | cox | 22193 | 128 | 2.666838736 | 2.240467261 | 3.174350712 | 2.57E-28 | 1.48E-27 |
| genetic | ALP_Neg | neurodm | diabetic_neuropathy | cox | 458500 | 1923 | 1.051894443 | 1.006020296 | 1.099860435 | 0.026159757 | 0.039239635 |
| biochemistry | ALP_Neg | neurodm | diabetic_neuropathy | cox | 438131 | 1833 | 3.653731685 | 3.511955589 | 3.801231219 | 0 | 0 |
| metabolomics | ALP_Neg | neurodm | diabetic_neuropathy | cox | 257891 | 1127 | 0.644857838 | 0.606087314 | 0.686108456 | 9.89E-44 | 2.47E-43 |
| proteomics | ALP_Neg | neurodm | diabetic_neuropathy | cox | 41339 | 216 | 0.988758161 | 0.865081493 | 1.130116305 | 0.868293191 | 0.968997537 |
| merged_omics | ALP_Neg | neurodm | diabetic_neuropathy | cox | 22193 | 128 | 1.181129367 | 0.993525117 | 1.404158342 | 0.059242366 | 0.073825103 |
| genetic | SHBG-LpA | neurodm | diabetic_neuropathy | cox | 458500 | 1923 | 1.067462299 | 1.020897137 | 1.116151392 | 0.004119833 | 0.006790934 |
| biochemistry | SHBG-LpA | neurodm | diabetic_neuropathy | cox | 438131 | 1833 | 1.56573725 | 1.49602868 | 1.638693942 | 5.84E-83 | 8.69E-83 |
| genetic | Obesity | neurodm | diabetic_neuropathy | cox | 458500 | 1923 | 1.301027034 | 1.244358178 | 1.360276624 | 5.10E-31 | 3.83E-30 |
| biochemistry | Obesity | neurodm | diabetic_neuropathy | cox | 438131 | 1833 | 3.569644599 | 3.404913109 | 3.742345885 | 0 | 0 |
| genetic | VAT_Neg | neurodm | diabetic_neuropathy | cox | 458500 | 1923 | 1.099464514 | 1.051166961 | 1.149981175 | 3.52E-05 | 7.99E-05 |
| biochemistry | VAT_Neg | neurodm | diabetic_neuropathy | cox | 438131 | 1833 | 2.889020633 | 2.777049776 | 3.005506163 | 0 | 0 |
| genetic | Bilirubin | neurodm | diabetic_neuropathy | cox | 458500 | 1923 | 1.015986562 | 0.971767982 | 1.062217231 | 0.484811756 | 0.542699727 |
| genetic | Proinsulin | neurodm | diabetic_neuropathy | cox | 458500 | 1923 | 1.232116867 | 1.178105534 | 1.288604399 | 7.06E-20 | 3.78E-19 |
| biochemistry | Proinsulin | neurodm | diabetic_neuropathy | cox | 438131 | 1833 | 4.752435841 | 4.568255484 | 4.944041878 | 0 | 0 |

**Table S16: Associations between pPSs, pMMSs and T2D-related disease outcomes in UKBB**

| Score_type | Biological_Process | Outcome | Outcome_Heading | Model_T | Sample_Size | Events | estimate | conf_low | conf_high | p | fdr_p_adjusted |
|---|---|---|---|---|---|---|---|---|---|---|---|
| proteomics | Proinsulin | neurodm | diabetic_neuropathy | cox | 41339 | 216 | 0.869869423 | 0.761425188 | 0.99375858 | 0.040153892 | 0.356287728 |
| merged_omics | Proinsulin | neurodm | diabetic_neuropathy | cox | 22193 | 128 | 1.70679843 | 1.435477434 | 2.029402072 | 1.42E-09 | 3.61E-09 |
| genetic | Hyper_Insulin | neurodm | diabetic_neuropathy | cox | 458500 | 1923 | 1.207835663 | 1.155045244 | 1.263038825 | 1.22E-16 | 5.88E-16 |
| biochemistry | Hyper_Insulin | neurodm | diabetic_neuropathy | cox | 438131 | 1833 | 2.52308919 | 2.429983481 | 2.619762278 | 0 | 0 |
| genetic | Beta_Cell_1 | neurodm | diabetic_neuropathy | cox | 458500 | 1923 | 1.264474282 | 1.209166637 | 1.322311715 | 8.37E-25 | 5.23E-24 |
| biochemistry | Beta_Cell_1 | neurodm | diabetic_neuropathy | cox | 438131 | 1833 | 4.20527407 | 4.048463777 | 4.368158141 | 0 | 0 |
| genetic | Beta_Cell_2 | neurodm | diabetic_neuropathy | cox | 458500 | 1923 | 1.192709852 | 1.140871058 | 1.246904091 | 7.66E-15 | 3.48E-14 |
| biochemistry | Beta_Cell_2 | neurodm | diabetic_neuropathy | cox | 438131 | 1833 | 4.454043722 | 4.283057601 | 4.631855868 | 0 | 0 |
| proteomics | Beta_Cell_2 | neurodm | diabetic_neuropathy | cox | 41339 | 216 | 1.030932612 | 0.901346917 | 1.179148705 | 0.656681494 | 0.963492845 |
| merged_omics | Beta_Cell_2 | neurodm | diabetic_neuropathy | cox | 22193 | 128 | 3.630702169 | 3.084588893 | 4.273502467 | 3.26E-54 | 2.64E-53 |
| genetic | Total_GRS | neurodm | diabetic_neuropathy | cox | 458500 | 1923 | 1.603054573 | 1.532919437 | 1.676398578 | 5.79E-95 | 9.65E-94 |
| biochemistry | Total_GRS | neurodm | diabetic_neuropathy | cox | 438131 | 1833 | 3.993883716 | 3.85373188 | 4.139132569 | 0 | 0 |
| merged_omics | Total_GRS | neurodm | diabetic_neuropathy | cox | 22193 | 128 | 4.587510234 | 3.935150412 | 5.348016708 | 2.16E-84 | 2.92E-83 |
| genetic | Cholesterol | retidm | diabetic_retinopathy | cox | 457822 | 3782 | 1.065550978 | 1.030748314 | 1.101528736 | 0.000178579 | 0.000357157 |
| biochemistry | Cholesterol | retidm | diabetic_retinopathy | cox | 437481 | 3579 | 1.918525716 | 1.858530867 | 1.980457247 | 0 | 0 |
| metabolomics | Cholesterol | retidm | diabetic_retinopathy | cox | 257556 | 2285 | 0.745234746 | 0.714583009 | 0.777201276 | 7.44E-43 | 1.75E-42 |
| proteomics | Cholesterol | retidm | diabetic_retinopathy | cox | 41287 | 360 | 1.053564851 | 0.950309701 | 1.168039107 | 0.321433299 | 0.856875603 |
| merged_omics | Cholesterol | retidm | diabetic_retinopathy | cox | 22170 | 206 | 1.64828588 | 1.439134941 | 1.887832936 | 5.26E-13 | 1.64E-12 |
| genetic | Lipodystrophy_1 | retidm | diabetic_retinopathy | cox | 457822 | 3782 | 1.191686619 | 1.154159463 | 1.23043396 | 6.44E-27 | 4.60E-26 |
| biochemistry | Lipodystrophy_1 | retidm | diabetic_retinopathy | cox | 437481 | 3579 | 1.857480278 | 1.797881874 | 1.919054324 | 3.77E-303 | 1.08E-302 |
| metabolomics | Lipodystrophy_1 | retidm | diabetic_retinopathy | cox | 257556 | 2285 | 1.069608984 | 1.022851435 | 1.118503958 | 0.003170194 | 0.003522437 |
| proteomics | Lipodystrophy_1 | retidm | diabetic_retinopathy | cox | 41287 | 360 | 1.00263304 | 0.904285059 | 1.111677123 | 0.960184409 | 0.968997537 |
| merged_omics | Lipodystrophy_1 | retidm | diabetic_retinopathy | cox | 22170 | 206 | 1.288387745 | 1.112825383 | 1.491647304 | 0.000698085 | 0.001130898 |
| genetic | Lipodystrophy_2 | retidm | diabetic_retinopathy | cox | 457822 | 3782 | 1.141391643 | 1.105789656 | 1.17813987 | 2.84E-16 | 1.33E-15 |
| biochemistry | Lipodystrophy_2 | retidm | diabetic_retinopathy | cox | 437481 | 3579 | 2.294630911 | 2.227605056 | 2.363673491 | 0 | 0 |
| proteomics | Lipodystrophy_2 | retidm | diabetic_retinopathy | cox | 41287 | 360 | 1.009111882 | 0.910152745 | 1.118830652 | 0.863241794 | 0.968997537 |
| merged_omics | Lipodystrophy_2 | retidm | diabetic_retinopathy | cox | 22170 | 206 | 2.254452049 | 1.967013443 | 2.583893902 | 1.58E-31 | 9.83E-31 |
| genetic | Liver-Lipid | retidm | diabetic_retinopathy | cox | 457822 | 3782 | 1.083841987 | 1.049467535 | 1.119342346 | 9.77E-07 | 2.44E-06 |
| biochemistry | Liver-Lipid | retidm | diabetic_retinopathy | cox | 437481 | 3579 | 1.194287573 | 1.154384948 | 1.235569477 | 1.30E-24 | 1.57E-24 |
| metabolomics | Liver-Lipid | retidm | diabetic_retinopathy | cox | 257556 | 2285 | 1.632833965 | 1.573069831 | 1.694868661 | 1.79E-146 | 7.94E-146 |
| proteomics | Liver-Lipid | retidm | diabetic_retinopathy | cox | 41287 | 360 | 0.997951396 | 0.900038978 | 1.106515399 | 0.968952244 | 0.968997537 |
| merged_omics | Liver-Lipid | retidm | diabetic_retinopathy | cox | 22170 | 206 | 0.648231229 | 0.56694755 | 0.741168607 | 2.27E-10 | 6.34E-10 |
| genetic | ASAT_Pos | retidm | diabetic_retinopathy | cox | 457822 | 3782 | 1.128898511 | 1.093569148 | 1.165369241 | 7.79E-14 | 3.24E-13 |
| biochemistry | ASAT_Pos | retidm | diabetic_retinopathy | cox | 437481 | 3579 | 2.429871587 | 2.357539411 | 2.504423001 | 0 | 0 |
| proteomics | ASAT_Pos | retidm | diabetic_retinopathy | cox | 41287 | 360 | 1.048627425 | 0.945923281 | 1.162482729 | 0.366590969 | 0.856875603 |
| merged_omics | ASAT_Pos | retidm | diabetic_retinopathy | cox | 22170 | 206 | 2.754662424 | 2.401103789 | 3.160281995 | 2.23E-47 | 1.64E-46 |
| genetic | ALP_Neg | retidm | diabetic_retinopathy | cox | 457822 | 3782 | 1.063393558 | 1.030146838 | 1.097713276 | 0.000149001 | 0.000302028 |
| biochemistry | ALP_Neg | retidm | diabetic_retinopathy | cox | 437481 | 3579 | 3.493406453 | 3.394191836 | 3.595521184 | 0 | 0 |
| metabolomics | ALP_Neg | retidm | diabetic_retinopathy | cox | 257556 | 2285 | 0.643100879 | 0.615672134 | 0.6717516 | 1.08E-87 | 3.92E-87 |
| proteomics | ALP_Neg | retidm | diabetic_retinopathy | cox | 41287 | 360 | 0.993122822 | 0.895592416 | 1.101274331 | 0.895893903 | 0.968997537 |
| merged_omics | ALP_Neg | retidm | diabetic_retinopathy | cox | 22170 | 206 | 1.411952361 | 1.232027907 | 1.61815285 | 7.04E-07 | 1.36E-06 |
| genetic | SHBG-LpA | retidm | diabetic_retinopathy | cox | 457822 | 3782 | 1.03030927 | 0.998003629 | 1.063660654 | 0.066202204 | 0.09020099 |
| biochemistry | SHBG-LpA | retidm | diabetic_retinopathy | cox | 437481 | 3579 | 1.495007768 | 1.446913856 | 1.544700271 | 2.25E-128 | 3.93E-128 |
| genetic | Obesity | retidm | diabetic_retinopathy | cox | 457822 | 3782 | 1.208208938 | 1.170420602 | 1.247217312 | 1.89E-31 | 1.49E-30 |
| biochemistry | Obesity | retidm | diabetic_retinopathy | cox | 437481 | 3579 | 3.356064593 | 3.244727415 | 3.471222114 | 0 | 0 |
| genetic | VAT_Neg | retidm | diabetic_retinopathy | cox | 457822 | 3782 | 1.134634214 | 1.098865336 | 1.171567396 | 1.09E-14 | 4.79E-14 |
| biochemistry | VAT_Neg | retidm | diabetic_retinopathy | cox | 437481 | 3579 | 2.966911273 | 2.883767362 | 3.052452364 | 0 | 0 |
| genetic | Bilirubin | retidm | diabetic_retinopathy | cox | 457822 | 3782 | 1.006664079 | 0.975148439 | 1.039198268 | 0.682332497 | 0.715733389 |
| genetic | Proinsulin | retidm | diabetic_retinopathy | cox | 457822 | 3782 | 1.249749994 | 1.210415924 | 1.290362277 | 1.66E-42 | 1.78E-41 |
| biochemistry | Proinsulin | retidm | diabetic_retinopathy | cox | 437481 | 3579 | 4.717574103 | 4.585064426 | 4.853913348 | 0 | 0 |
| proteomics | Proinsulin | retidm | diabetic_retinopathy | cox | 41287 | 360 | 0.974667226 | 0.879112443 | 1.080608299 | 0.625971866 | 0.963492845 |
| merged_omics | Proinsulin | retidm | diabetic_retinopathy | cox | 22170 | 206 | 1.864291887 | 1.628664972 | 2.134008099 | 1.64E-19 | 8.28E-19 |
| genetic | Hyper_Insulin | retidm | diabetic_retinopathy | cox | 457822 | 3782 | 1.184940507 | 1.147794955 | 1.223288183 | 1.58E-25 | 1.08E-24 |
| biochemistry | Hyper_Insulin | retidm | diabetic_retinopathy | cox | 437481 | 3579 | 2.599313721 | 2.529920819 | 2.670609994 | 0 | 0 |
| genetic | Beta_Cell_1 | retidm | diabetic_retinopathy | cox | 457822 | 3782 | 1.284815983 | 1.244507214 | 1.326430326 | 1.40E-53 | 1.62E-52 |
| biochemistry | Beta_Cell_1 | retidm | diabetic_retinopathy | cox | 437481 | 3579 | 4.169240781 | 4.056298342 | 4.285327959 | 0 | 0 |
| genetic | Beta_Cell_2 | retidm | diabetic_retinopathy | cox | 457822 | 3782 | 1.245939285 | 1.207136642 | 1.285989213 | 2.96E-42 | 2.96E-41 |
| biochemistry | Beta_Cell_2 | retidm | diabetic_retinopathy | cox | 437481 | 3579 | 4.40031798 | 4.277034258 | 4.527155304 | 0 | 0 |
| proteomics | Beta_Cell_2 | retidm | diabetic_retinopathy | cox | 41287 | 360 | 1.040922425 | 0.938245622 | 1.154835652 | 0.449077513 | 0.856875603 |
| merged_omics | Beta_Cell_2 | retidm | diabetic_retinopathy | cox | 22170 | 206 | 3.39113816 | 2.982710506 | 3.855492512 | 1.25E-77 | 1.45E-76 |
| genetic | Total_GRS | retidm | diabetic_retinopathy | cox | 457822 | 3782 | 1.659512258 | 1.607410576 | 1.713302735 | 1.19E-212 | 3.57E-211 |
| biochemistry | Total_GRS | retidm | diabetic_retinopathy | cox | 437481 | 3579 | 3.926113265 | 3.827016311 | 4.027776239 | 0 | 0 |
| merged_omics | Total_GRS | retidm | diabetic_retinopathy | cox | 22170 | 206 | 4.622510688 | 4.087751369 | 5.227227179 | 1.45E-131 | 2.94E-130 |

**Table S17: Associations of biochemical pMMSs with T2D-related disease outcomes in UKBB, before and after adjustment for HbA1c and glocuse**

| Score_type | Biological_Process | Outcome | Outcome_Heading | Model_Ty | Sample_Size | Events | estimate | conf_low | conf_high | p | fdr_p_adjusted |
|---|---|---|---|---|---|---|---|---|---|---|---|
| biochemistry | Cholesterol | dm2 | type_2_diabetes | cox | 430723 | 28393 | 1.57969 | 1.561734 | 1.597853 | 0 | 0 |
| biochemistry (adjust for hba1c and glucose ) | Cholesterol | dm2 | type_2_diabetes | cox | 376031 | 24896 | 1.622777 | 1.603093 | 1.642702 | 0 | 0 |
| biochemistry | Lipodystrophy_1 | dm2 | type_2_diabetes | cox | 430723 | 28393 | 1.756855 | 1.736228 | 1.777726 | 0 | 0 |
| biochemistry (adjust for hba1c and glucose ) | Lipodystrophy_1 | dm2 | type_2_diabetes | cox | 376031 | 24896 | 1.354439 | 1.337266 | 1.371833 | 0 | 0 |
| biochemistry | Lipodystrophy_2 | dm2 | type_2_diabetes | cox | 430723 | 28393 | 2.057045 | 2.034399 | 2.079943 | 0 | 0 |
| biochemistry (adjust for hba1c and glucose ) | Lipodystrophy_2 | dm2 | type_2_diabetes | cox | 376031 | 24896 | 1.673655 | 1.653242 | 1.694319 | 0 | 0 |
| biochemistry | Liver-Lipid | dm2 | type_2_diabetes | cox | 430723 | 28393 | 0.865291 | 0.854904 | 0.875805 | 6.20E-122 | 1.09E-121 |
| biochemistry (adjust for hba1c and glucose ) | Liver-Lipid | dm2 | type_2_diabetes | cox | 376031 | 24896 | 0.771254 | 0.761298 | 0.78134 | 0 | 0 |
| biochemistry | ASAT_Pos | dm2 | type_2_diabetes | cox | 430723 | 28393 | 2.152652 | 2.128216 | 2.17737 | 0 | 0 |
| biochemistry (adjust for hba1c and glucose ) | ASAT_Pos | dm2 | type_2_diabetes | cox | 376031 | 24896 | 1.913848 | 1.894992 | 1.932892 | 0 | 0 |
| biochemistry | ALP_Neg | dm2 | type_2_diabetes | cox | 430723 | 28393 | 2.668352 | 2.638178 | 2.698871 | 0 | 0 |
| biochemistry (adjust for hba1c and glucose ) | ALP_Neg | dm2 | type_2_diabetes | cox | 376031 | 24896 | 2.073767 | 2.051343 | 2.096437 | 0 | 0 |
| biochemistry | SHBG-LpA | dm2 | type_2_diabetes | cox | 430723 | 28393 | 1.232282 | 1.217984 | 1.246749 | 1.54E-269 | 4.08E-269 |
| biochemistry (adjust for hba1c and glucose ) | SHBG-LpA | dm2 | type_2_diabetes | cox | 376031 | 24896 | 1.216937 | 1.201946 | 1.232114 | 1.28E-211 | 4.88E-211 |
| biochemistry | Obesity | dm2 | type_2_diabetes | cox | 430723 | 28393 | 3.018445 | 2.982108 | 3.055225 | 0 | 0 |
| biochemistry (adjust for hba1c and glucose ) | Obesity | dm2 | type_2_diabetes | cox | 376031 | 24896 | 2.030016 | 2.015914 | 2.044216 | 0 | 0 |
| biochemistry | VAT_Neg | dm2 | type_2_diabetes | cox | 430723 | 28393 | 2.518567 | 2.490952 | 2.546489 | 0 | 0 |
| biochemistry (adjust for hba1c and glucose ) | VAT_Neg | dm2 | type_2_diabetes | cox | 376031 | 24896 | 1.814481 | 1.792269 | 1.836969 | 0 | 0 |
| biochemistry | Proinsulin | dm2 | type_2_diabetes | cox | 430723 | 28393 | 3.876365 | 3.836032 | 3.917122 | 0 | 0 |
| biochemistry (adjust for hba1c and glucose ) | Proinsulin | dm2 | type_2_diabetes | cox | 376031 | 24896 | 2.843879 | 2.813155 | 2.87494 | 0 | 0 |
| biochemistry | Hyper_Insulin | dm2 | type_2_diabetes | cox | 430723 | 28393 | 2.170729 | 2.147873 | 2.193827 | 0 | 0 |
| biochemistry (adjust for hba1c and glucose ) | Hyper_Insulin | dm2 | type_2_diabetes | cox | 376031 | 24896 | 1.988113 | 1.96806 | 2.008371 | 0 | 0 |
| biochemistry | Beta_Cell_1 | dm2 | type_2_diabetes | cox | 430723 | 28393 | 3.223719 | 3.190596 | 3.257186 | 0 | 0 |
| biochemistry (adjust for hba1c and glucose ) | Beta_Cell_1 | dm2 | type_2_diabetes | cox | 376031 | 24896 | 2.370297 | 2.337594 | 2.403457 | 0 | 0 |
| biochemistry | Beta_Cell_2 | dm2 | type_2_diabetes | cox | 430723 | 28393 | 3.248472 | 3.213877 | 3.283439 | 0 | 0 |
| biochemistry (adjust for hba1c and glucose ) | Beta_Cell_2 | dm2 | type_2_diabetes | cox | 376031 | 24896 | 2.297303 | 2.266222 | 2.32881 | 0 | 0 |
| biochemistry | Cholesterol | acmort | all_cause_mortality | cox | 438513 | 39213 | 1.198072 | 1.18636 | 1.2099 | 1.12E-284 | 3.09E-284 |
| biochemistry (adjust for hba1c and glucose ) | Cholesterol | acmort | all_cause_mortality | cox | 382845 | 34360 | 1.183851 | 1.171656 | 1.196172 | 5.70E-224 | 2.32E-223 |
| biochemistry | Lipodystrophy_1 | acmort | all_cause_mortality | cox | 438513 | 39213 | 0.908454 | 0.899331 | 0.917669 | 1.40E-77 | 2.11E-77 |
| biochemistry (adjust for hba1c and glucose ) | Lipodystrophy_1 | acmort | all_cause_mortality | cox | 382845 | 34360 | 0.843077 | 0.834133 | 0.852118 | 5.66E-216 | 2.23E-215 |
| biochemistry | Lipodystrophy_2 | acmort | all_cause_mortality | cox | 438513 | 39213 | 1.07114 | 1.060419 | 1.08197 | 6.92E-41 | 9.46E-41 |
| biochemistry (adjust for hba1c and glucose ) | Lipodystrophy_2 | acmort | all_cause_mortality | cox | 382845 | 34360 | 1.006547 | 0.995763 | 1.017448 | 0.235067607 | 0.248445438 |
| biochemistry | Liver-Lipid | acmort | all_cause_mortality | cox | 438513 | 39213 | 0.87787 | 0.868842 | 0.886992 | 1.16E-134 | 2.19E-134 |
| biochemistry (adjust for hba1c and glucose ) | Liver-Lipid | acmort | all_cause_mortality | cox | 382845 | 34360 | 0.857877 | 0.848454 | 0.867403 | 5.52E-163 | 1.99E-162 |
| biochemistry | ASAT_Pos | acmort | all_cause_mortality | cox | 438513 | 39213 | 1.020917 | 1.010709 | 1.031229 | 5.40E-05 | 5.66E-05 |
| biochemistry (adjust for hba1c and glucose ) | ASAT_Pos | acmort | all_cause_mortality | cox | 382845 | 34360 | 0.98767 | 0.978445 | 0.996982 | 0.009561584 | 0.01090356 |
| biochemistry | ALP_Neg | acmort | all_cause_mortality | cox | 438513 | 39213 | 1.154293 | 1.142758 | 1.165944 | 1.51E-172 | 3.21E-172 |
| biochemistry (adjust for hba1c and glucose ) | ALP_Neg | acmort | all_cause_mortality | cox | 382845 | 34360 | 1.058511 | 1.048711 | 1.068403 | 4.42E-33 | 7.77E-33 |
| biochemistry | SHBG-LpA | acmort | all_cause_mortality | cox | 438513 | 39213 | 1.022529 | 1.012428 | 1.03273 | 1.09E-05 | 1.17E-05 |
| biochemistry (adjust for hba1c and glucose ) | SHBG-LpA | acmort | all_cause_mortality | cox | 382845 | 34360 | 1.003997 | 0.993559 | 1.014545 | 0.45434093 | 0.461440007 |
| biochemistry | Obesity | acmort | all_cause_mortality | cox | 438513 | 39213 | 1.289032 | 1.275994 | 1.302203 | 0 | 0 |
| biochemistry (adjust for hba1c and glucose ) | Obesity | acmort | all_cause_mortality | cox | 382845 | 34360 | 1.199246 | 1.188733 | 1.209853 | 0 | 0 |
| biochemistry | VAT_Neg | acmort | all_cause_mortality | cox | 438513 | 39213 | 1.058341 | 1.047981 | 1.068803 | 1.34E-29 | 1.70E-29 |
| biochemistry (adjust for hba1c and glucose ) | VAT_Neg | acmort | all_cause_mortality | cox | 382845 | 34360 | 0.95628 | 0.946496 | 0.966166 | 1.60E-17 | 2.19E-17 |
| biochemistry | Proinsulin | acmort | all_cause_mortality | cox | 438513 | 39213 | 1.223361 | 1.21099 | 1.235859 | 0 | 0 |
| biochemistry (adjust for hba1c and glucose ) | Proinsulin | acmort | all_cause_mortality | cox | 382845 | 34360 | 1.080803 | 1.072131 | 1.089545 | 1.03E-79 | 2.39E-79 |
| biochemistry | Hyper_Insulin | acmort | all_cause_mortality | cox | 438513 | 39213 | 1.023157 | 1.012982 | 1.033435 | 7.15E-06 | 7.74E-06 |
| biochemistry (adjust for hba1c and glucose ) | Hyper_Insulin | acmort | all_cause_mortality | cox | 382845 | 34360 | 0.978081 | 0.968653 | 0.987601 | 7.30E-06 | 8.71E-06 |
| biochemistry | Beta_Cell_1 | acmort | all_cause_mortality | cox | 438513 | 39213 | 1.211342 | 1.199504 | 1.223297 | 2.233e-320 | 7.081e-320 |
| biochemistry (adjust for hba1c and glucose ) | Beta_Cell_1 | acmort | all_cause_mortality | cox | 382845 | 34360 | 1.054917 | 1.044442 | 1.065497 | 8.64E-26 | 1.40E-25 |
| biochemistry | Beta_Cell_2 | acmort | all_cause_mortality | cox | 438513 | 39213 | 1.222761 | 1.21081 | 1.234829 | 0 | 0 |
| biochemistry (adjust for hba1c and glucose ) | Beta_Cell_2 | acmort | all_cause_mortality | cox | 382845 | 34360 | 1.075459 | 1.064671 | 1.086357 | 2.08E-45 | 3.92E-45 |
| biochemistry | Cholesterol | cvdmort | cardiovascular_disease_mortality | cox | 438513 | 8107 | 1.258777 | 1.231967 | 1.28617 | 1.78E-97 | 2.89E-97 |
| biochemistry (adjust for hba1c and glucose ) | Cholesterol | cvdmort | cardiovascular_disease_mortality | cox | 382845 | 7155 | 1.244168 | 1.216377 | 1.272594 | 4.00E-80 | 9.46E-80 |
| biochemistry | Lipodystrophy_1 | cvdmort | cardiovascular_disease_mortality | cox | 438513 | 8107 | 0.927801 | 0.907663 | 0.948385 | 2.18E-11 | 2.46E-11 |
| biochemistry (adjust for hba1c and glucose ) | Lipodystrophy_1 | cvdmort | cardiovascular_disease_mortality | cox | 382845 | 7155 | 0.831753 | 0.81277 | 0.85118 | 3.96E-55 | 8.17E-55 |
| biochemistry | Lipodystrophy_2 | cvdmort | cardiovascular_disease_mortality | cox | 438513 | 8107 | 1.084896 | 1.061367 | 1.108945 | 3.24E-13 | 3.73E-13 |
| biochemistry (adjust for hba1c and glucose ) | Lipodystrophy_2 | cvdmort | cardiovascular_disease_mortality | cox | 382845 | 7155 | 0.985338 | 0.962458 | 1.008761 | 0.217852679 | 0.2321381 |
| biochemistry | Liver-Lipid | cvdmort | cardiovascular_disease_mortality | cox | 438513 | 8107 | 0.905152 | 0.884831 | 0.925941 | 7.85E-18 | 9.28E-18 |
| biochemistry (adjust for hba1c and glucose ) | Liver-Lipid | cvdmort | cardiovascular_disease_mortality | cox | 382845 | 7155 | 0.86724 | 0.846491 | 0.888499 | 9.50E-31 | 1.60E-30 |
| biochemistry | ASAT_Pos | cvdmort | cardiovascular_disease_mortality | cox | 438513 | 8107 | 1.004163 | 0.982464 | 1.02634 | 0.709371053 | 0.709371053 |
| biochemistry (adjust for hba1c and glucose ) | ASAT_Pos | cvdmort | cardiovascular_disease_mortality | cox | 382845 | 7155 | 0.975311 | 0.955832 | 0.995187 | 0.015151618 | 0.017127916 |
| biochemistry | ALP_Neg | cvdmort | cardiovascular_disease_mortality | cox | 438513 | 8107 | 1.295057 | 1.267416 | 1.323301 | 5.23E-122 | 9.32E-122 |
| biochemistry (adjust for hba1c and glucose ) | ALP_Neg | cvdmort | cardiovascular_disease_mortality | cox | 382845 | 7155 | 1.144275 | 1.12152 | 1.16749 | 1.67E-39 | 3.11E-39 |
| biochemistry | SHBG-LpA | cvdmort | cardiovascular_disease_mortality | cox | 438513 | 8107 | 1.152106 | 1.127431 | 1.177321 | 1.29E-37 | 1.71E-37 |
| biochemistry (adjust for hba1c and glucose ) | SHBG-LpA | cvdmort | cardiovascular_disease_mortality | cox | 382845 | 7155 | 1.124932 | 1.099667 | 1.150777 | 3.05E-24 | 4.90E-24 |
| biochemistry | Obesity | cvdmort | cardiovascular_disease_mortality | cox | 438513 | 8107 | 1.417621 | 1.38637 | 1.449576 | 9.06E-207 | 2.14E-206 |
| biochemistry (adjust for hba1c and glucose ) | Obesity | cvdmort | cardiovascular_disease_mortality | cox | 382845 | 7155 | 1.270373 | 1.248538 | 1.292591 | 3.46E-161 | 1.22E-160 |
| biochemistry | VAT_Neg | cvdmort | cardiovascular_disease_mortality | cox | 438513 | 8107 | 1.137758 | 1.113809 | 1.162223 | 1.33E-32 | 1.71E-32 |
| biochemistry (adjust for hba1c and glucose ) | VAT_Neg | cvdmort | cardiovascular_disease_mortality | cox | 382845 | 7155 | 0.982313 | 0.960526 | 1.004594 | 0.11889517 | 0.127738612 |
| biochemistry | Proinsulin | cvdmort | cardiovascular_disease_mortality | cox | 438513 | 8107 | 1.412669 | 1.382571 | 1.443422 | 5.31E-217 | 1.28E-216 |
| biochemistry (adjust for hba1c and glucose ) | Proinsulin | cvdmort | cardiovascular_disease_mortality | cox | 382845 | 7155 | 1.182468 | 1.162075 | 1.203218 | 1.55E-79 | 3.53E-79 |
| biochemistry | Hyper_Insulin | cvdmort | cardiovascular_disease_mortality | cox | 438513 | 8107 | 1.08394 | 1.060896 | 1.107484 | 1.95E-13 | 2.27E-13 |
| biochemistry (adjust for hba1c and glucose ) | Hyper_Insulin | cvdmort | cardiovascular_disease_mortality | cox | 382845 | 7155 | 1.023575 | 1.002663 | 1.044923 | 0.026930778 | 0.029420178 |
| biochemistry | Beta_Cell_1 | cvdmort | cardiovascular_disease_mortality | cox | 438513 | 8107 | 1.353442 | 1.325598 | 1.381871 | 4.14E-179 | 9.12E-179 |
| biochemistry (adjust for hba1c and glucose ) | Beta_Cell_1 | cvdmort | cardiovascular_disease_mortality | cox | 382845 | 7155 | 1.110906 | 1.086945 | 1.135396 | 3.27E-21 | 4.99E-21 |
| biochemistry | Beta_Cell_2 | cvdmort | cardiovascular_disease_mortality | cox | 438513 | 8107 | 1.371342 | 1.343163 | 1.400113 | 2.85E-195 | 6.39E-195 |
| biochemistry (adjust for hba1c and glucose ) | Beta_Cell_2 | cvdmort | cardiovascular_disease_mortality | cox | 382845 | 7155 | 1.145433 | 1.120511 | 1.170908 | 1.08E-33 | 1.92E-33 |
| biochemistry | Cholesterol | cad | coronary_artery_disease | cox | 424098 | 19120 | 0.970227 | 0.956504 | 0.984147 | 3.20E-05 | 3.41E-05 |
| biochemistry (adjust for hba1c and glucose ) | Cholesterol | cad | coronary_artery_disease | cox | 370242 | 16817 | 0.963244 | 0.948914 | 0.97779 | 9.72E-07 | 1.20E-06 |
| biochemistry | Lipodystrophy_1 | cad | coronary_artery_disease | cox | 424098 | 19120 | 1.179231 | 1.162468 | 1.196235 | 8.68E-113 | 1.48E-112 |
| biochemistry (adjust for hba1c and glucose ) | Lipodystrophy_1 | cad | coronary_artery_disease | cox | 370242 | 16817 | 1.129827 | 1.112657 | 1.147262 | 5.09E-55 | 1.03E-54 |
| biochemistry | Lipodystrophy_2 | cad | coronary_artery_disease | cox | 424098 | 19120 | 1.071777 | 1.056418 | 1.08736 | 4.85E-21 | 5.83E-21 |
| biochemistry (adjust for hba1c and glucose ) | Lipodystrophy_2 | cad | coronary_artery_disease | cox | 370242 | 16817 | 1.020635 | 1.004999 | 1.036515 | 0.00951308 | 0.01090356 |
| biochemistry | Liver-Lipid | cad | coronary_artery_disease | cox | 424098 | 19120 | 0.877242 | 0.864372 | 0.890304 | 1.44E-67 | 2.12E-67 |
| biochemistry (adjust for hba1c and glucose ) | Liver-Lipid | cad | coronary_artery_disease | cox | 370242 | 16817 | 0.861196 | 0.847733 | 0.874873 | 3.99E-77 | 8.94E-77 |
| biochemistry | ASAT_Pos | cad | coronary_artery_disease | cox | 424098 | 19120 | 1.030187 | 1.015469 | 1.045117 | 5.10E-05 | 5.39E-05 |
| biochemistry (adjust for hba1c and glucose ) | ASAT_Pos | cad | coronary_artery_disease | cox | 370242 | 16817 | 0.9949 | 0.981704 | 1.008274 | 0.452958424 | 0.461440007 |
| biochemistry | ALP_Neg | cad | coronary_artery_disease | cox | 424098 | 19120 | 1.270883 | 1.252933 | 1.289089 | 2.94E-239 | 7.65E-239 |
| biochemistry (adjust for hba1c and glucose ) | ALP_Neg | cad | coronary_artery_disease | cox | 370242 | 16817 | 1.174344 | 1.158819 | 1.190077 | 7.59E-124 | 2.35E-123 |
| biochemistry | SHBG-LpA | cad | coronary_artery_disease | cox | 424098 | 19120 | 1.217427 | 1.200281 | 1.234818 | 9.33E-163 | 1.89E-162 |
| biochemistry (adjust for hba1c and glucose ) | SHBG-LpA | cad | coronary_artery_disease | cox | 370242 | 16817 | 1.195152 | 1.177447 | 1.213123 | 3.22E-121 | 9.74E-121 |
| biochemistry | Obesity | cad | coronary_artery_disease | cox | 424098 | 19120 | 1.250523 | 1.232561 | 1.268747 | 1.73E-201 | 4.01E-201 |
| biochemistry (adjust for hba1c and glucose ) | Obesity | cad | coronary_artery_disease | cox | 370242 | 16817 | 1.159545 | 1.145638 | 1.173621 | 9.36E-128 | 3.04E-127 |
| biochemistry | VAT_Neg | cad | coronary_artery_disease | cox | 424098 | 19120 | 1.215917 | 1.199154 | 1.232914 | 1.05E-167 | 2.21E-167 |
| biochemistry (adjust for hba1c and glucose ) | VAT_Neg | cad | coronary_artery_disease | cox | 370242 | 16817 | 1.137517 | 1.120818 | 1.154465 | 2.24E-65 | 4.86E-65 |
| biochemistry | Proinsulin | cad | coronary_artery_disease | cox | 424098 | 19120 | 1.300237 | 1.281859 | 1.318879 | 3.89E-286 | 1.10E-285 |
| biochemistry (adjust for hba1c and glucose ) | Proinsulin | cad | coronary_artery_disease | cox | 370242 | 16817 | 1.157349 | 1.14384 | 1.171018 | 1.99E-131 | 6.63E-131 |

**Table S17: Associations of biochemical pMMSs with T2D-related disease outcomes in UKBB, before and after adjustment for HbA1c and glocuse**

| Score_type | Biological_Process | Outcome | Outcome_Heading | Model_Ty | Sample_Size | Events | estimate | conf_low | conf_high | p | fdr_p_adjusted |
|---|---|---|---|---|---|---|---|---|---|---|---|
| biochemistry | Hyper_Insulin | cad | coronary_artery_disease | cox | 424098 | 19120 | 1.10425 | 1.088812 | 1.119907 | 2.37E-43 | 3.28E-43 |
| biochemistry (adjust for hba1c and glucose ) | Hyper_Insulin | cad | coronary_artery_disease | cox | 370242 | 16817 | 1.064346 | 1.049903 | 1.078987 | 3.67E-19 | 5.25E-19 |
| biochemistry | Beta_Cell_1 | cad | coronary_artery_disease | cox | 424098 | 19120 | 1.204071 | 1.187423 | 1.220953 | 1.21E-150 | 2.38E-150 |
| biochemistry (adjust for hba1c and glucose ) | Beta_Cell_1 | cad | coronary_artery_disease | cox | 370242 | 16817 | 1.087891 | 1.07259 | 1.103409 | 2.12E-31 | 3.67E-31 |
| biochemistry | Beta_Cell_2 | cad | coronary_artery_disease | cox | 424098 | 19120 | 1.186333 | 1.169827 | 1.203072 | 2.94E-126 | 5.31E-126 |
| biochemistry (adjust for hba1c and glucose ) | Beta_Cell_2 | cad | coronary_artery_disease | cox | 370242 | 16817 | 1.067568 | 1.052507 | 1.082845 | 1.89E-19 | 2.73E-19 |
| biochemistry | Cholesterol | is | ischemic_stroke | cox | 436637 | 8783 | 1.194633 | 1.170084 | 1.219697 | 3.02E-63 | 4.41E-63 |
| biochemistry (adjust for hba1c and glucose ) | Cholesterol | is | ischemic_stroke | cox | 381208 | 7693 | 1.177691 | 1.152207 | 1.203738 | 1.27E-48 | 2.43E-48 |
| biochemistry | Lipodystrophy_1 | is | ischemic_stroke | cox | 436637 | 8783 | 0.960721 | 0.940456 | 0.981423 | 0.000229668 | 0.000238855 |
| biochemistry (adjust for hba1c and glucose ) | Lipodystrophy_1 | is | ischemic_stroke | cox | 381208 | 7693 | 0.88991 | 0.870037 | 0.910237 | 4.41E-24 | 6.90E-24 |
| biochemistry | Lipodystrophy_2 | is | ischemic_stroke | cox | 436637 | 8783 | 1.097685 | 1.074657 | 1.121206 | 6.91E-18 | 8.24E-18 |
| biochemistry (adjust for hba1c and glucose ) | Lipodystrophy_2 | is | ischemic_stroke | cox | 381208 | 7693 | 1.02829 | 1.005196 | 1.051914 | 0.01607497 | 0.017861078 |
| biochemistry | Liver-Lipid | is | ischemic_stroke | cox | 436637 | 8783 | 0.933237 | 0.913074 | 0.953846 | 5.64E-10 | 6.16E-10 |
| biochemistry (adjust for hba1c and glucose ) | Liver-Lipid | is | ischemic_stroke | cox | 381208 | 7693 | 0.904355 | 0.883474 | 0.925729 | 3.30E-17 | 4.47E-17 |
| biochemistry | ASAT_Pos | is | ischemic_stroke | cox | 436637 | 8783 | 1.028544 | 1.006929 | 1.050624 | 0.009399716 | 0.009621757 |
| biochemistry (adjust for hba1c and glucose ) | ASAT_Pos | is | ischemic_stroke | cox | 381208 | 7693 | 0.99167 | 0.972269 | 1.011457 | 0.40661302 | 0.41952137 |
| biochemistry | ALP_Neg | is | ischemic_stroke | cox | 436637 | 8783 | 1.171856 | 1.147266 | 1.196974 | 1.22E-48 | 1.72E-48 |
| biochemistry (adjust for hba1c and glucose ) | ALP_Neg | is | ischemic_stroke | cox | 381208 | 7693 | 1.063942 | 1.043281 | 1.085012 | 5.83E-10 | 7.73E-10 |
| biochemistry | SHBG-LpA | is | ischemic_stroke | cox | 436637 | 8783 | 1.079224 | 1.056846 | 1.102075 | 9.90E-13 | 1.13E-12 |
| biochemistry (adjust for hba1c and glucose ) | SHBG-LpA | is | ischemic_stroke | cox | 381208 | 7693 | 1.05656 | 1.033506 | 1.080127 | 1.02E-06 | 1.25E-06 |
| biochemistry | Obesity | is | ischemic_stroke | cox | 436637 | 8783 | 1.303497 | 1.275796 | 1.331798 | 3.15E-129 | 5.86E-129 |
| biochemistry (adjust for hba1c and glucose ) | Obesity | is | ischemic_stroke | cox | 381208 | 7693 | 1.197809 | 1.176394 | 1.219613 | 1.27E-85 | 3.11E-85 |
| biochemistry | VAT_Neg | is | ischemic_stroke | cox | 436637 | 8783 | 1.116825 | 1.093964 | 1.140165 | 1.18E-25 | 1.46E-25 |
| biochemistry (adjust for hba1c and glucose ) | VAT_Neg | is | ischemic_stroke | cox | 381208 | 7693 | 1.011098 | 0.989349 | 1.033325 | 0.31982216 | 0.332615047 |
| biochemistry | Proinsulin | is | ischemic_stroke | cox | 436637 | 8783 | 1.266812 | 1.240012 | 1.294191 | 3.25E-104 | 5.42E-104 |
| biochemistry (adjust for hba1c and glucose ) | Proinsulin | is | ischemic_stroke | cox | 381208 | 7693 | 1.102955 | 1.084345 | 1.121884 | 1.52E-29 | 2.54E-29 |
| biochemistry | Hyper_Insulin | is | ischemic_stroke | cox | 436637 | 8783 | 1.074086 | 1.051793 | 1.096851 | 2.41E-11 | 2.70E-11 |
| biochemistry (adjust for hba1c and glucose ) | Hyper_Insulin | is | ischemic_stroke | cox | 381208 | 7693 | 1.017663 | 0.997217 | 1.038529 | 0.090864553 | 0.098436599 |
| biochemistry | Beta_Cell_1 | is | ischemic_stroke | cox | 436637 | 8783 | 1.225646 | 1.2005 | 1.251319 | 1.79E-82 | 2.73E-82 |
| biochemistry (adjust for hba1c and glucose ) | Beta_Cell_1 | is | ischemic_stroke | cox | 381208 | 7693 | 1.055961 | 1.03399 | 1.078398 | 3.86E-07 | 4.87E-07 |
| biochemistry | Beta_Cell_2 | is | ischemic_stroke | cox | 436637 | 8783 | 1.228369 | 1.203137 | 1.25413 | 4.81E-84 | 7.53E-84 |
| biochemistry (adjust for hba1c and glucose ) | Beta_Cell_2 | is | ischemic_stroke | cox | 381208 | 7693 | 1.068621 | 1.046191 | 1.091532 | 8.66E-10 | 1.14E-09 |
| biochemistry | Cholesterol | hf | heart_failure | cox | 436022 | 16549 | 1.355834 | 1.335567 | 1.376409 | 0 | 0 |
| biochemistry (adjust for hba1c and glucose ) | Cholesterol | hf | heart_failure | cox | 380670 | 14490 | 1.34221 | 1.321061 | 1.363697 | 7.56E-289 | 3.64E-288 |
| biochemistry | Lipodystrophy_1 | hf | heart_failure | cox | 436022 | 16549 | 0.90948 | 0.895433 | 0.923748 | 6.72E-33 | 8.74E-33 |
| biochemistry (adjust for hba1c and glucose ) | Lipodystrophy_1 | hf | heart_failure | cox | 380670 | 14490 | 0.827659 | 0.814159 | 0.841384 | 1.58E-112 | 4.57E-112 |
| biochemistry | Lipodystrophy_2 | hf | heart_failure | cox | 436022 | 16549 | 1.133941 | 1.116588 | 1.151564 | 1.88E-57 | 2.69E-57 |
| biochemistry (adjust for hba1c and glucose ) | Lipodystrophy_2 | hf | heart_failure | cox | 380670 | 14490 | 1.045926 | 1.028721 | 1.063419 | 1.12E-07 | 1.44E-07 |
| biochemistry | Liver-Lipid | hf | heart_failure | cox | 436022 | 16549 | 0.873761 | 0.859904 | 0.887842 | 1.73E-61 | 2.50E-61 |
| biochemistry (adjust for hba1c and glucose ) | Liver-Lipid | hf | heart_failure | cox | 380670 | 14490 | 0.841234 | 0.826967 | 0.855747 | 2.44E-87 | 6.23E-87 |
| biochemistry | ASAT_Pos | hf | heart_failure | cox | 436022 | 16549 | 0.992554 | 0.977252 | 1.008096 | 0.345769117 | 0.351171759 |
| biochemistry (adjust for hba1c and glucose ) | ASAT_Pos | hf | heart_failure | cox | 380670 | 14490 | 0.963971 | 0.950158 | 0.977985 | 6.26E-07 | 7.82E-07 |
| biochemistry | ALP_Neg | hf | heart_failure | cox | 436022 | 16549 | 1.24154 | 1.222529 | 1.260846 | 2.94E-166 | 6.08E-166 |
| biochemistry (adjust for hba1c and glucose ) | ALP_Neg | hf | heart_failure | cox | 380670 | 14490 | 1.116983 | 1.101139 | 1.133055 | 4.93E-52 | 9.71E-52 |
| biochemistry | SHBG-LpA | hf | heart_failure | cox | 436022 | 16549 | 1.097757 | 1.081116 | 1.114654 | 5.26E-33 | 6.90E-33 |
| biochemistry (adjust for hba1c and glucose ) | SHBG-LpA | hf | heart_failure | cox | 380670 | 14490 | 1.078638 | 1.061417 | 1.096138 | 3.00E-20 | 4.43E-20 |
| biochemistry | Obesity | hf | heart_failure | cox | 436022 | 16549 | 1.473495 | 1.450606 | 1.496745 | 0 | 0 |
| biochemistry (adjust for hba1c and glucose ) | Obesity | hf | heart_failure | cox | 380670 | 14490 | 1.318668 | 1.30144 | 1.336124 | 0 | 0 |
| biochemistry | VAT_Neg | hf | heart_failure | cox | 436022 | 16549 | 1.150396 | 1.133224 | 1.167829 | 1.76E-74 | 2.64E-74 |
| biochemistry (adjust for hba1c and glucose ) | VAT_Neg | hf | heart_failure | cox | 380670 | 14490 | 1.019819 | 1.003757 | 1.036137 | 0.015392572 | 0.017250296 |
| biochemistry | Proinsulin | hf | heart_failure | cox | 436022 | 16549 | 1.353847 | 1.333082 | 1.374936 | 7.4e-323 | 2.4e-322 |
| biochemistry (adjust for hba1c and glucose ) | Proinsulin | hf | heart_failure | cox | 380670 | 14490 | 1.15691 | 1.142723 | 1.171274 | 1.37E-118 | 4.06E-118 |
| biochemistry | Hyper_Insulin | hf | heart_failure | cox | 436022 | 16549 | 1.091124 | 1.074611 | 1.107891 | 3.69E-29 | 4.62E-29 |
| biochemistry (adjust for hba1c and glucose ) | Hyper_Insulin | hf | heart_failure | cox | 380670 | 14490 | 1.035056 | 1.019916 | 1.05042 | 4.58E-06 | 5.57E-06 |
| biochemistry | Beta_Cell_1 | hf | heart_failure | cox | 436022 | 16549 | 1.306846 | 1.287459 | 1.326525 | 7.97E-270 | 2.16E-269 |
| biochemistry (adjust for hba1c and glucose ) | Beta_Cell_1 | hf | heart_failure | cox | 380670 | 14490 | 1.103533 | 1.086573 | 1.120758 | 1.13E-35 | 2.04E-35 |
| biochemistry | Beta_Cell_2 | hf | heart_failure | cox | 436022 | 16549 | 1.330768 | 1.311041 | 1.350792 | 8.62E-308 | 2.67E-307 |
| biochemistry (adjust for hba1c and glucose ) | Beta_Cell_2 | hf | heart_failure | cox | 380670 | 14490 | 1.141299 | 1.123577 | 1.159301 | 1.53E-61 | 3.20E-61 |
| biochemistry | Cholesterol | pad | peripheral_artery_disease | cox | 435420 | 7056 | 1.299332 | 1.26963 | 1.329729 | 3.96E-109 | 6.68E-109 |
| biochemistry (adjust for hba1c and glucose ) | Cholesterol | pad | peripheral_artery_disease | cox | 380096 | 6180 | 1.292627 | 1.261494 | 1.324528 | 1.32E-94 | 3.50E-94 |
| biochemistry | Lipodystrophy_1 | pad | peripheral_artery_disease | cox | 435420 | 7056 | 1.00749 | 0.983847 | 1.031702 | 0.537951103 | 0.542121267 |
| biochemistry (adjust for hba1c and glucose ) | Lipodystrophy_1 | pad | peripheral_artery_disease | cox | 380096 | 6180 | 0.883922 | 0.861917 | 0.906489 | 8.57E-22 | 1.33E-21 |
| biochemistry | Lipodystrophy_2 | pad | peripheral_artery_disease | cox | 435420 | 7056 | 1.122839 | 1.096617 | 1.149688 | 7.25E-22 | 8.81E-22 |
| biochemistry (adjust for hba1c and glucose ) | Lipodystrophy_2 | pad | peripheral_artery_disease | cox | 380096 | 6180 | 0.995594 | 0.970538 | 1.021296 | 0.734191961 | 0.739883371 |
| biochemistry | Liver-Lipid | pad | peripheral_artery_disease | cox | 435420 | 7056 | 0.837371 | 0.81715 | 0.858093 | 5.89E-46 | 8.24E-46 |
| biochemistry (adjust for hba1c and glucose ) | Liver-Lipid | pad | peripheral_artery_disease | cox | 380096 | 6180 | 0.796998 | 0.776421 | 0.818121 | 7.98E-65 | 1.70E-64 |
| biochemistry | ASAT_Pos | pad | peripheral_artery_disease | cox | 435420 | 7056 | 1.093161 | 1.0676 | 1.119335 | 1.60E-13 | 1.87E-13 |
| biochemistry (adjust for hba1c and glucose ) | ASAT_Pos | pad | peripheral_artery_disease | cox | 380096 | 6180 | 1.061614 | 1.038774 | 1.084955 | 7.11E-08 | 9.24E-08 |
| biochemistry | ALP_Neg | pad | peripheral_artery_disease | cox | 435420 | 7056 | 1.353061 | 1.321787 | 1.385075 | 1.07E-141 | 2.04E-141 |
| biochemistry (adjust for hba1c and glucose ) | ALP_Neg | pad | peripheral_artery_disease | cox | 380096 | 6180 | 1.178441 | 1.153094 | 1.204345 | 1.46E-49 | 2.82E-49 |
| biochemistry | SHBG-LpA | pad | peripheral_artery_disease | cox | 435420 | 7056 | 1.169019 | 1.142042 | 1.196632 | 2.87E-39 | 3.89E-39 |
| biochemistry (adjust for hba1c and glucose ) | SHBG-LpA | pad | peripheral_artery_disease | cox | 380096 | 6180 | 1.141677 | 1.113918 | 1.170128 | 5.06E-26 | 8.33E-26 |
| biochemistry | Obesity | pad | peripheral_artery_disease | cox | 435420 | 7056 | 1.607909 | 1.5699 | 1.646839 | 0 | 0 |
| biochemistry (adjust for hba1c and glucose ) | Obesity | pad | peripheral_artery_disease | cox | 380096 | 6180 | 1.391749 | 1.366987 | 1.41696 | 3.28E-285 | 1.52E-284 |
| biochemistry | VAT_Neg | pad | peripheral_artery_disease | cox | 435420 | 7056 | 1.261399 | 1.233024 | 1.290427 | 4.98E-89 | 8.00E-89 |
| biochemistry (adjust for hba1c and glucose ) | VAT_Neg | pad | peripheral_artery_disease | cox | 380096 | 6180 | 1.055961 | 1.030599 | 1.081948 | 1.13E-05 | 1.34E-05 |
| biochemistry | Proinsulin | pad | peripheral_artery_disease | cox | 435420 | 7056 | 1.538652 | 1.503576 | 1.574547 | 1.18E-293 | 3.49E-293 |
| biochemistry (adjust for hba1c and glucose ) | Proinsulin | pad | peripheral_artery_disease | cox | 380096 | 6180 | 1.255806 | 1.232508 | 1.279544 | 1.27E-125 | 4.03E-125 |
| biochemistry | Hyper_Insulin | pad | peripheral_artery_disease | cox | 435420 | 7056 | 1.165884 | 1.139304 | 1.193083 | 6.85E-39 | 9.18E-39 |
| biochemistry (adjust for hba1c and glucose ) | Hyper_Insulin | pad | peripheral_artery_disease | cox | 380096 | 6180 | 1.105533 | 1.081306 | 1.130303 | 7.02E-19 | 9.93E-19 |
| biochemistry | Beta_Cell_1 | pad | peripheral_artery_disease | cox | 435420 | 7056 | 1.438051 | 1.406212 | 1.47061 | 5.77E-222 | 1.44E-221 |
| biochemistry (adjust for hba1c and glucose ) | Beta_Cell_1 | pad | peripheral_artery_disease | cox | 380096 | 6180 | 1.115717 | 1.089662 | 1.142395 | 1.06E-19 | 1.55E-19 |
| biochemistry | Beta_Cell_2 | pad | peripheral_artery_disease | cox | 435420 | 7056 | 1.455734 | 1.423468 | 1.488732 | 1.80E-236 | 4.60E-236 |
| biochemistry (adjust for hba1c and glucose ) | Beta_Cell_2 | pad | peripheral_artery_disease | cox | 380096 | 6180 | 1.151236 | 1.124102 | 1.179025 | 5.64E-31 | 9.65E-31 |
| biochemistry | Cholesterol | nephrodm | diabetic_nephropathy | cox | 438398 | 658 | 2.206525 | 2.049949 | 2.375061 | 1.36E-98 | 2.23E-98 |
| biochemistry (adjust for hba1c and glucose ) | Cholesterol | nephrodm | diabetic_nephropathy | cox | 382752 | 591 | 2.421182 | 2.240582 | 2.61634 | 1.03E-110 | 2.84E-110 |
| biochemistry | Lipodystrophy_1 | nephrodm | diabetic_nephropathy | cox | 438398 | 658 | 1.556709 | 1.442335 | 1.680152 | 6.09E-30 | 7.76E-30 |
| biochemistry (adjust for hba1c and glucose ) | Lipodystrophy_1 | nephrodm | diabetic_nephropathy | cox | 382752 | 591 | 0.680911 | 0.627883 | 0.738417 | 1.53E-20 | 2.29E-20 |
| biochemistry | Lipodystrophy_2 | nephrodm | diabetic_nephropathy | cox | 438398 | 658 | 2.019099 | 1.881049 | 2.167281 | 3.15E-84 | 5.00E-84 |
| biochemistry (adjust for hba1c and glucose ) | Lipodystrophy_2 | nephrodm | diabetic_nephropathy | cox | 382752 | 591 | 1.001379 | 0.921176 | 1.088564 | 0.974193707 | 0.974193707 |
| biochemistry | Liver-Lipid | nephrodm | diabetic_nephropathy | cox | 438398 | 658 | 1.288788 | 1.191163 | 1.394414 | 2.74E-10 | 3.02E-10 |
| biochemistry (adjust for hba1c and glucose ) | Liver-Lipid | nephrodm | diabetic_nephropathy | cox | 382752 | 591 | 0.901018 | 0.827094 | 0.981549 | 0.017014673 | 0.018744979 |
| biochemistry | ASAT_Pos | nephrodm | diabetic_nephropathy | cox | 438398 | 658 | 1.292398 | 1.197492 | 1.394825 | 4.35E-11 | 4.83E-11 |
| biochemistry (adjust for hba1c and glucose ) | ASAT_Pos | nephrodm | diabetic_nephropathy | cox | 382752 | 591 | 1.337135 | 1.25175 | 1.428343 | 6.15E-18 | 8.60E-18 |
| biochemistry | ALP_Neg | nephrodm | diabetic_nephropathy | cox | 438398 | 658 | 4.690813 | 4.411876 | 4.987387 | 0 | 0 |
| biochemistry (adjust for hba1c and glucose ) | ALP_Neg | nephrodm | diabetic_nephropathy | cox | 382752 | 591 | 3.343284 | 3.128479 | 3.572838 | 6.10E-278 | 2.73E-277 |
| biochemistry | SHBG-LpA | nephrodm | diabetic_nephropathy | cox | 438398 | 658 | 2.406694 | 2.231697 | 2.595415 | 4.37E-115 | 7.57E-115 |
| biochemistry (adjust for hba1c and glucose ) | SHBG-LpA | nephrodm | diabetic_nephropathy | cox | 382752 | 591 | 2.385674 | 2.203631 | 2.582756 | 2.97E-102 | 8.06E-102 |

**Table S17: Associations of biochemical pMMSs with T2D-related disease outcomes in UKBB, before and after adjustment for HbA1c and glocuse**

| Score_type | Biological_Process | Outcome | Outcome_Heading | Model_Ty | Sample_Size | Events | estimate | conf_low | conf_high | p | fdr_p_adjusted |
|---|---|---|---|---|---|---|---|---|---|---|---|
| biochemistry | Obesity | nephrodm | diabetic_nephropathy | cox | 438398 | 658 | 3.128895 | 2.892635 | 3.384453 | 2.31E-178 | 5.00E-178 |
| biochemistry (adjust for hba1c and glucose ) | Obesity | nephrodm | diabetic_nephropathy | cox | 382752 | 591 | 2.015564 | 1.945808 | 2.087821 | 0 | 0 |
| biochemistry | VAT_Neg | nephrodm | diabetic_nephropathy | cox | 438398 | 658 | 2.508326 | 2.345517 | 2.682436 | 6.78E-159 | 1.36E-158 |
| biochemistry (adjust for hba1c and glucose ) | VAT_Neg | nephrodm | diabetic_nephropathy | cox | 382752 | 591 | 0.882512 | 0.815363 | 0.955191 | 0.001965573 | 0.002302022 |
| biochemistry | Proinsulin | nephrodm | diabetic_nephropathy | cox | 438398 | 658 | 5.284985 | 4.944526 | 5.648886 | 0 | 0 |
| biochemistry (adjust for hba1c and glucose ) | Proinsulin | nephrodm | diabetic_nephropathy | cox | 382752 | 591 | 3.536044 | 3.279689 | 3.812437 | 2.81E-237 | 1.22E-236 |
| biochemistry | Hyper_Insulin | nephrodm | diabetic_nephropathy | cox | 438398 | 658 | 2.576733 | 2.421737 | 2.74165 | 1.74E-196 | 3.96E-196 |
| biochemistry (adjust for hba1c and glucose ) | Hyper_Insulin | nephrodm | diabetic_nephropathy | cox | 382752 | 591 | 2.176013 | 2.052528 | 2.306927 | 5.51E-150 | 1.89E-149 |
| biochemistry | Beta_Cell_1 | nephrodm | diabetic_nephropathy | cox | 438398 | 658 | 4.424469 | 4.151648 | 4.715217 | 0 | 0 |
| biochemistry (adjust for hba1c and glucose ) | Beta_Cell_1 | nephrodm | diabetic_nephropathy | cox | 382752 | 591 | 1.109185 | 1.027666 | 1.197171 | 0.007797671 | 0.009050868 |
| biochemistry | Beta_Cell_2 | nephrodm | diabetic_nephropathy | cox | 438398 | 658 | 4.994387 | 4.67413 | 5.336587 | 0 | 0 |
| biochemistry (adjust for hba1c and glucose ) | Beta_Cell_2 | nephrodm | diabetic_nephropathy | cox | 382752 | 591 | 1.524629 | 1.405272 | 1.654124 | 3.66E-24 | 5.81E-24 |
| biochemistry | Cholesterol | neurodm | diabetic_neuropathy | cox | 438131 | 1833 | 2.043829 | 1.955221 | 2.136453 | 2.61E-219 | 6.41E-219 |
| biochemistry (adjust for hba1c and glucose ) | Cholesterol | neurodm | diabetic_neuropathy | cox | 382518 | 1592 | 2.214092 | 2.111517 | 2.32165 | 1.47E-236 | 6.17E-236 |
| biochemistry | Lipodystrophy_1 | neurodm | diabetic_neuropathy | cox | 438131 | 1833 | 1.799899 | 1.719969 | 1.883543 | 7.00E-142 | 1.36E-141 |
| biochemistry (adjust for hba1c and glucose ) | Lipodystrophy_1 | neurodm | diabetic_neuropathy | cox | 382518 | 1592 | 0.876845 | 0.833958 | 0.921938 | 2.80E-07 | 3.56E-07 |
| biochemistry | Lipodystrophy_2 | neurodm | diabetic_neuropathy | cox | 438131 | 1833 | 2.39171 | 2.295054 | 2.492437 | 0 | 0 |
| biochemistry (adjust for hba1c and glucose ) | Lipodystrophy_2 | neurodm | diabetic_neuropathy | cox | 382518 | 1592 | 1.372005 | 1.305637 | 1.441747 | 7.25E-36 | 1.33E-35 |
| biochemistry | Liver-Lipid | neurodm | diabetic_neuropathy | cox | 438131 | 1833 | 1.070986 | 1.021241 | 1.123155 | 0.004710456 | 0.004859994 |
| biochemistry (adjust for hba1c and glucose ) | Liver-Lipid | neurodm | diabetic_neuropathy | cox | 382518 | 1592 | 0.780812 | 0.741204 | 0.822537 | 1.22E-20 | 1.84E-20 |
| biochemistry | ASAT_Pos | neurodm | diabetic_neuropathy | cox | 438131 | 1833 | 2.195705 | 2.104293 | 2.291088 | 9.38E-288 | 2.71E-287 |
| biochemistry (adjust for hba1c and glucose ) | ASAT_Pos | neurodm | diabetic_neuropathy | cox | 382518 | 1592 | 1.961758 | 1.894399 | 2.031512 | 1.108819134075e | 5.544095670374e-312 |
| biochemistry | ALP_Neg | neurodm | diabetic_neuropathy | cox | 438131 | 1833 | 3.653732 | 3.511956 | 3.801231 | 0 | 0 |
| biochemistry (adjust for hba1c and glucose ) | ALP_Neg | neurodm | diabetic_neuropathy | cox | 382518 | 1592 | 2.494955 | 2.392321 | 2.601992 | 0 | 0 |
| biochemistry | SHBG-LpA | neurodm | diabetic_neuropathy | cox | 438131 | 1833 | 1.565737 | 1.496029 | 1.638694 | 5.84E-83 | 9.03E-83 |
| biochemistry (adjust for hba1c and glucose ) | SHBG-LpA | neurodm | diabetic_neuropathy | cox | 382518 | 1592 | 1.557086 | 1.482794 | 1.6351 | 1.63E-70 | 3.58E-70 |
| biochemistry | Obesity | neurodm | diabetic_neuropathy | cox | 438131 | 1833 | 3.569645 | 3.404913 | 3.742346 | 0 | 0 |
| biochemistry (adjust for hba1c and glucose ) | Obesity | neurodm | diabetic_neuropathy | cox | 382518 | 1592 | 2.069913 | 2.025221 | 2.115591 | 0 | 0 |
| biochemistry | VAT_Neg | neurodm | diabetic_neuropathy | cox | 438131 | 1833 | 2.889021 | 2.77705 | 3.005506 | 0 | 0 |
| biochemistry (adjust for hba1c and glucose ) | VAT_Neg | neurodm | diabetic_neuropathy | cox | 382518 | 1592 | 1.225248 | 1.167184 | 1.2862 | 2.38E-16 | 3.19E-16 |
| biochemistry | Proinsulin | neurodm | diabetic_neuropathy | cox | 438131 | 1833 | 4.752436 | 4.568255 | 4.944042 | 0 | 0 |
| biochemistry (adjust for hba1c and glucose ) | Proinsulin | neurodm | diabetic_neuropathy | cox | 382518 | 1592 | 3.127548 | 2.993268 | 3.267852 | 0 | 0 |
| biochemistry | Hyper_Insulin | neurodm | diabetic_neuropathy | cox | 438131 | 1833 | 2.523089 | 2.429983 | 2.619762 | 0 | 0 |
| biochemistry (adjust for hba1c and glucose ) | Hyper_Insulin | neurodm | diabetic_neuropathy | cox | 382518 | 1592 | 2.180827 | 2.104565 | 2.259852 | 0 | 0 |
| biochemistry | Beta_Cell_1 | neurodm | diabetic_neuropathy | cox | 438131 | 1833 | 4.205274 | 4.048464 | 4.368158 | 0 | 0 |
| biochemistry (adjust for hba1c and glucose ) | Beta_Cell_1 | neurodm | diabetic_neuropathy | cox | 382518 | 1592 | 1.231912 | 1.174633 | 1.291984 | 9.01E-18 | 1.25E-17 |
| biochemistry | Beta_Cell_2 | neurodm | diabetic_neuropathy | cox | 438131 | 1833 | 4.454044 | 4.283058 | 4.631856 | 0 | 0 |
| biochemistry (adjust for hba1c and glucose ) | Beta_Cell_2 | neurodm | diabetic_neuropathy | cox | 382518 | 1592 | 1.471708 | 1.401045 | 1.545934 | 1.84E-53 | 3.67E-53 |
| biochemistry | Cholesterol | retidm | diabetic_retinopathy | cox | 437481 | 3579 | 1.918526 | 1.858531 | 1.980457 | 0 | 0 |
| biochemistry (adjust for hba1c and glucose ) | Cholesterol | retidm | diabetic_retinopathy | cox | 381943 | 3111 | 2.065162 | 1.996251 | 2.136453 | 0 | 0 |
| biochemistry | Lipodystrophy_1 | retidm | diabetic_retinopathy | cox | 437481 | 3579 | 1.85748 | 1.797882 | 1.919054 | 3.77E-303 | 1.14E-302 |
| biochemistry (adjust for hba1c and glucose ) | Lipodystrophy_1 | retidm | diabetic_retinopathy | cox | 381943 | 3111 | 0.98163 | 0.946744 | 1.017801 | 0.315248551 | 0.330502513 |
| biochemistry | Lipodystrophy_2 | retidm | diabetic_retinopathy | cox | 437481 | 3579 | 2.294631 | 2.227605 | 2.363673 | 0 | 0 |
| biochemistry (adjust for hba1c and glucose ) | Lipodystrophy_2 | retidm | diabetic_retinopathy | cox | 381943 | 3111 | 1.409791 | 1.361069 | 1.460257 | 1.19E-81 | 2.87E-81 |
| biochemistry | Liver-Lipid | retidm | diabetic_retinopathy | cox | 437481 | 3579 | 1.194288 | 1.154385 | 1.235569 | 1.30E-24 | 1.60E-24 |
| biochemistry (adjust for hba1c and glucose ) | Liver-Lipid | retidm | diabetic_retinopathy | cox | 381943 | 3111 | 0.91704 | 0.883491 | 0.951864 | 5.26E-06 | 6.33E-06 |
| biochemistry | ASAT_Pos | retidm | diabetic_retinopathy | cox | 437481 | 3579 | 2.429872 | 2.357539 | 2.504423 | 0 | 0 |
| biochemistry (adjust for hba1c and glucose ) | ASAT_Pos | retidm | diabetic_retinopathy | cox | 381943 | 3111 | 2.074213 | 2.023172 | 2.126542 | 0 | 0 |
| biochemistry | ALP_Neg | retidm | diabetic_retinopathy | cox | 437481 | 3579 | 3.493406 | 3.394192 | 3.595521 | 0 | 0 |
| biochemistry (adjust for hba1c and glucose ) | ALP_Neg | retidm | diabetic_retinopathy | cox | 381943 | 3111 | 2.419335 | 2.347545 | 2.49332 | 0 | 0 |
| biochemistry | SHBG-LpA | retidm | diabetic_retinopathy | cox | 437481 | 3579 | 1.495008 | 1.446914 | 1.5447 | 2.25E-128 | 4.12E-128 |
| biochemistry (adjust for hba1c and glucose ) | SHBG-LpA | retidm | diabetic_retinopathy | cox | 381943 | 3111 | 1.49447 | 1.442971 | 1.547807 | 1.12E-111 | 3.16E-111 |
| biochemistry | Obesity | retidm | diabetic_retinopathy | cox | 437481 | 3579 | 3.356065 | 3.244727 | 3.471222 | 0 | 0 |
| biochemistry (adjust for hba1c and glucose ) | Obesity | retidm | diabetic_retinopathy | cox | 381943 | 3111 | 2.052217 | 2.019477 | 2.085488 | 0 | 0 |
| biochemistry | VAT_Neg | retidm | diabetic_retinopathy | cox | 437481 | 3579 | 2.966911 | 2.883767 | 3.052452 | 0 | 0 |
| biochemistry (adjust for hba1c and glucose ) | VAT_Neg | retidm | diabetic_retinopathy | cox | 381943 | 3111 | 1.415775 | 1.367526 | 1.465727 | 5.44E-86 | 1.36E-85 |
| biochemistry | Proinsulin | retidm | diabetic_retinopathy | cox | 437481 | 3579 | 4.717574 | 4.585064 | 4.853913 | 0 | 0 |
| biochemistry (adjust for hba1c and glucose ) | Proinsulin | retidm | diabetic_retinopathy | cox | 381943 | 3111 | 3.125418 | 3.028917 | 3.224994 | 0 | 0 |
| biochemistry | Hyper_Insulin | retidm | diabetic_retinopathy | cox | 437481 | 3579 | 2.599314 | 2.529921 | 2.67061 | 0 | 0 |
| biochemistry (adjust for hba1c and glucose ) | Hyper_Insulin | retidm | diabetic_retinopathy | cox | 381943 | 3111 | 2.250608 | 2.193715 | 2.308978 | 0 | 0 |
| biochemistry | Beta_Cell_1 | retidm | diabetic_retinopathy | cox | 437481 | 3579 | 4.169241 | 4.056298 | 4.285328 | 0 | 0 |
| biochemistry (adjust for hba1c and glucose ) | Beta_Cell_1 | retidm | diabetic_retinopathy | cox | 381943 | 3111 | 1.437601 | 1.387618 | 1.489385 | 6.80E-90 | 1.77E-89 |
| biochemistry | Beta_Cell_2 | retidm | diabetic_retinopathy | cox | 437481 | 3579 | 4.400318 | 4.277034 | 4.527155 | 0 | 0 |
| biochemistry (adjust for hba1c and glucose ) | Beta_Cell_2 | retidm | diabetic_retinopathy | cox | 381943 | 3111 | 1.660894 | 1.601837 | 1.722128 | 4.40E-166 | 1.64E-165 |
